# Polygenic and familial contributions to antidepressant continuation, switching, discontinuation and augmentation in the All of Us and Pharmlines cohorts

**DOI:** 10.64898/2026.08.14.26360458

**Authors:** Alicia Walker, Xiaotong Wang, Jens Bos, Tian Lin, Frank Klont, Ilja Nolte, Harold Snieder, Roeland Broekema, Peter M. Visscher, Anjali K. Henders, Catharina Hartman, Hanna M. van Loo, Maxime Taquet, Eelko Hak, Naomi R. Wray

## Abstract

Predicting antidepressant response remains a major challenge, and it is unclear whether reported polygenic associations reflect drug-specific non-response or a broader propensity for treatment modification. We analysed participants with at least one antidepressant monotherapy episode of ≥28 days in the All of Us (AoU; n=98,357) and Pharmlines (Lifelines linked to IADB.nl; n=12,884) cohorts, comparing continuation with switching, discontinuation and augmentation (atypical antipsychotic or lithium) in relation to polygenic scores (PGS). Among individuals with recorded depression, switching, but not discontinuation, was associated with anxiety, higher depression symptom count and stress-related measures in both cohorts. Depression PGS was associated with switching in AoU (OR=1.16 per SD, 95% CI 1.12–1.20), with a concordant nominally significant estimate in Pharmlines (OR=1.11, 1.01–1.22). In AoU, depression PGS showed the strongest association with being in a higher treatment-intensity category (continuation < switching < augmentation; ordinal OR=1.18 per SD, 95% CI 1.15–1.21), whereas schizophrenia and bipolar disorder PGS were selectively associated with augmentation. There was no evidence that PGS associations with switching differed across antidepressant classes. Familial aggregation in Pharmlines was detectable for continuation, including SSRI and SNRI continuation, but not for switching or discontinuation. Antidepressant switching was associated with greater depression polygenic liability and clinical severity, with little evidence that PGS associations differed across antidepressant classes. Augmentation was additionally associated with bipolar disorder and schizophrenia polygenic liability, while familial aggregation was confined to switch-free continuation.

## Introduction

Antidepressant treatment rarely follows a uniform course. Some patients continue their initial medication, whereas others discontinue, switch antidepressants, or progress to more complex regimens such as augmentation with an atypical antipsychotic or lithium^1–5^. These prescribing events are increasingly used to infer treatment outcomes from electronic health records, but they are not interchangeable measures of antidepressant response. Switching is often treated as a proxy for inadequate response, while repeated treatment changes and augmentation can indicate increasing treatment difficulty or treatment-resistant depression (TRD)^2,6–8^. However, switching may also reflect adverse effects, adherence or patient preference^3,9^, and augmentation may additionally reflect broader psychiatric complexity or factors influencing treatment selection^6,10,11^. Variation in how these outcomes are defined has been identified as a major challenge for psychiatric pharmacogenomic research^12–14^.

Polygenic studies increasingly suggest that general psychiatric liability contributes to antidepressant prognosis. Scores derived directly from clinically assessed antidepressant-response GWAS currently have limited predictive power^15^. In contrast, higher MDD polygenic liability has shown modest but reproducible associations with non-response and non-remission across clinical antidepressant studies^16,17^. Large-scale studies have identified polygenic associations with broader treatment non-response, repeated antidepressant changes and treatment-resistant depression^10,11,18,19^. More intensive treatment-escalation phenotypes, including augmentation and electroconvulsive therapy (ECT), have additionally shown associations with bipolar disorder, schizophrenia and ADHD polygenic liability^10,11,19^. However, findings vary substantially according to how treatment resistance is defined^13,20^. Conversely, MDD PGS has not consistently predicted treatment-specific outcomes, with no significant association with SSRI switching in UK Biobank^7^ and no significant distinction between SSRI responders and non-responders in European-ancestry All of Us^18^. Current evidence therefore appears stronger for polygenic associations with prognosis and broader treatment difficulty than for differential response between antidepressants^17^.

Whether these associations depend on the treatment received remains largely unresolved. A PGS association observed across antidepressant users does not itself imply treatment-specific response. Evidence for treatment specificity requires demonstrating that the association differs between drugs or drug classes rather than observing significance in one treatment group but not another. It is also unclear whether switching, discontinuation and augmentation share the same polygenic background or represent partly distinct dimensions of treatment course. Familial influences are similarly poorly characterised, despite family history of medication response being considered clinically^21^. We therefore examined continuation, switching, discontinuation and augmentation in the US All of Us Research Program (AoU)^22^ and the Dutch Pharmlines cohort, comprising Lifelines participants linked to IADB.nl pharmacy records^23,24^. We compared their polygenic profiles, formally tested PGS × antidepressant interactions at drug and class level, and used the family structure of Pharmlines to assess familial aggregation.

## Methods

### Data sources

We analyzed medication, phenotypic and genetic data from AoU and Pharmlines. AoU is a US population cohort linking electronic health records (EHRs), surveys and genomic data. Pharmlines links participants from the population-based Lifelines cohort to community-pharmacy dispensing records from IADB.nl. The Lifelines study protocol was approved by the UMCG Medical Ethics Review Committee under reference 2007/152. Full descriptions of both cohorts, data linkage, and medication-record coverage are provided in the **Supplementary Methods**.

### Participant selection criteria

Participants were aged ≥12 years at their first recorded antidepressant and contributed at least one antidepressant monotherapy episode lasting ≥28 days. Individuals with recorded bipolar or psychotic disorders were excluded because antidepressant use in these groups may represent distinct treatment pathways. After exclusions, the analytical cohorts comprised 98,357 AoU and 12,884 Pharmlines participants. Full description of selection criteria is provided in the **Supplementary Methods**.

### Medication episodes and treatment outcomes

Details of treatment outcome eligibility rules can be found in the **Supplementary Methods**. First, antidepressant records were combined into continuous medication episodes allowing gaps of <90 days between records. Brief cross-tapers of <28 days between antidepressants were permitted at episode boundaries but excluded from monotherapy duration.

For each antidepressant, we defined three mutually exclusive monotherapy outcomes: switching, continuation and discontinuation. A switching event required ≥28 days of index-antidepressant monotherapy followed by initiation of another antidepressant with <28 days of overlap during cross-tapering or a gap of ≤90 days after index discontinuation. Among participants without a switch from that drug, continuation required at least one monotherapy episode lasting ≥90 days, whereas discontinuation required all observed monotherapy episodes to last ≥28 but <90 days. Participants could contribute outcomes for more than one antidepressant, and outcomes were analysed pooled across antidepressants and, for PGS analyses, by drug and drug class. A schematic of the treatment-outcome definitions is provided in **Supplementary Figure S1**.

Augmentation was examined separately as a marker of greater treatment intensity and was defined as ≥28 days of overlapping use of an antidepressant with lithium or an atypical antipsychotic^5^. Participants meeting augmentation criteria were excluded from the continuation reference group in augmentation-versus-continuation and discontinuation-versus-continuation analyses. In ordinal analyses, participants meeting augmentation criteria were assigned to the augmentation category, including those who also met switching or discontinuation criteria. Participants ever receiving combination therapy were excluded from continuation reference and switching groups. The antidepressants and atypical antipsychotics included in each cohort are listed in **Supplementary Table S18** and **S19**, respectively.

### Diagnostic and phenotypic measures

In AoU, recorded depression and anxiety were identified from EHR diagnostic codes, with lifetime major depressive disorder (MDD) additionally assessed in the subset completing the Composite International Diagnostic Interview–Short Form (CIDI-sf). In Pharmlines, depression was defined from Lifelines questionnaire and interview measures, principally the Mini-International Neuropsychiatric Interview and, where available, lifetime depression assessments^25–27^. Phenotypic predictors included psychiatric and medical comorbidities, depression-related measures, sociodemographic characteristics and lifestyle factors. Full phenotype definitions are provided in the **Supplementary Methods**.

In AoU, age at first antidepressant record, BMI and CIDI-sf MDD symptom count were standardized (mean=0, SD=1); all other phenotypic predictors were modelled categorically to examine dose– response relationship. When annual household income was included as a covariate in PGS analyses, it was modelled ordinally from 1 (≤US$25,000) to 8 (≥US$200,000). In Pharmlines, all continuous measures were standardized (mean=0, SD=1) prior to analysis; odds ratios represent the effect of a one standard deviation increase. Consequently, cross-cohort comparisons focused primarily on consistency of effect direction rather than direct comparison of effect magnitudes.

### Genetic data and polygenic scores

AoU genotypes were obtained from whole-genome sequence data and Pharmlines genotypes from quality-controlled and imputed array data.

We calculated PGS for major depression^28^, schizophrenia^29^, bipolar disorder^30^, ADHD^31^, migraine^32^, insomnia^32^ and BMI^33^ (**Supplementary Table S20**) using SBayesRC^34^ weights. These traits were selected to capture depression liability, broader psychiatric liability and common genetically correlated comorbidities potentially related to antidepressant treatment. The Lifelines contribution to the BMI discovery GWAS was removed using MetaSubtract^35^. Identical PGS weights were applied in both cohorts. Scores were residualized for ancestry principal components and standardised before analysis. Genotype processing, variant filtering, imputation, ancestry inference, and PGS calculation are described in the **Supplementary Methods**.

### Statistical Analyses

Primary analyses were conducted in participants of genetically inferred European ancestry; African- and Admixed/Latin-American-ancestry analyses were performed as sensitivity analyses in AoU. Associations of PGS and phenotypic predictors with switching and discontinuation were estimated using separate logistic regressions with continuation as the reference. Analyses were stratified by recorded depression status to partly address heterogeneity in treatment indication, with the full antidepressant-prescribed cohort and additional diagnostic subsets analyzed secondarily.

Augmentation was compared with continuation using analogous PGS models. To test whether polygenic liability increased with treatment intensity, we additionally fitted proportional-odds models ordered as continuation < switching < augmentation, assigning participants meeting both switching and augmentation criteria to augmentation. PGS associations with switching were also examined separately by index antidepressant and drug class. Treatment specificity was tested directly using PGS × drug and PGS × drug-class interactions rather than inferred from differences in significance between stratified analyses. For interaction models, cluster-robust standard errors were calculated at the participant level to account for individuals contributing observations for more than one antidepressant or drug class. Models were adjusted for age at first antidepressant record, sex, income and education. Multiple-testing correction was applied separately within prespecified model families using Bonferroni thresholds across the seven PGS or the phenotypic predictors tested within each cohort. Switching-window, discontinuation, diagnostic-stratum and ancestry sensitivity analyses are described in the **Supplementary Methods.**

Familial aggregation was assessed in Pharmlines using 1,152 relative pairs from 881 families. For continuation overall and by class, switching and discontinuation, we estimated recurrence-risk ratios and tetrachoric correlations of liability across parent–child, sibling and grandparent–grandchild relationships. Confidence intervals were obtained by family-level bootstrap resampling.

## Results

### Cohort and treatment characteristics

The analytical sample comprised 98,357 AoU and 12,884 Pharmlines participants. Mean age at first recorded antidepressant was 49 years (SD 16) in AoU and 44 years (SD 14) in Pharmlines; 49% and 35%, respectively, had recorded depression. AoU included substantially greater ancestral diversity, whereas Pharmlines was predominantly European ancestry.

Cohort characteristics by pooled treatment outcome are provided in **Supplementary Table S1**. Across antidepressants, 28% (N=27,389) of AoU participants and 17% (N=2,133) of Pharmlines participants recorded at least one switch (**Figure 1A-B**). Augmentation occurred in 8.3% (N=8,126) and 9.5% (N=1,223), respectively. SSRIs accounted for most initial treatments and many switches occurred within class (**Figure 1C-D**). Switching was more frequent among participants with recorded depression in both cohorts (**Supplementary Figure S2**). See the **Supplementary Results** for a complete cohort description.

**Figure 1.**
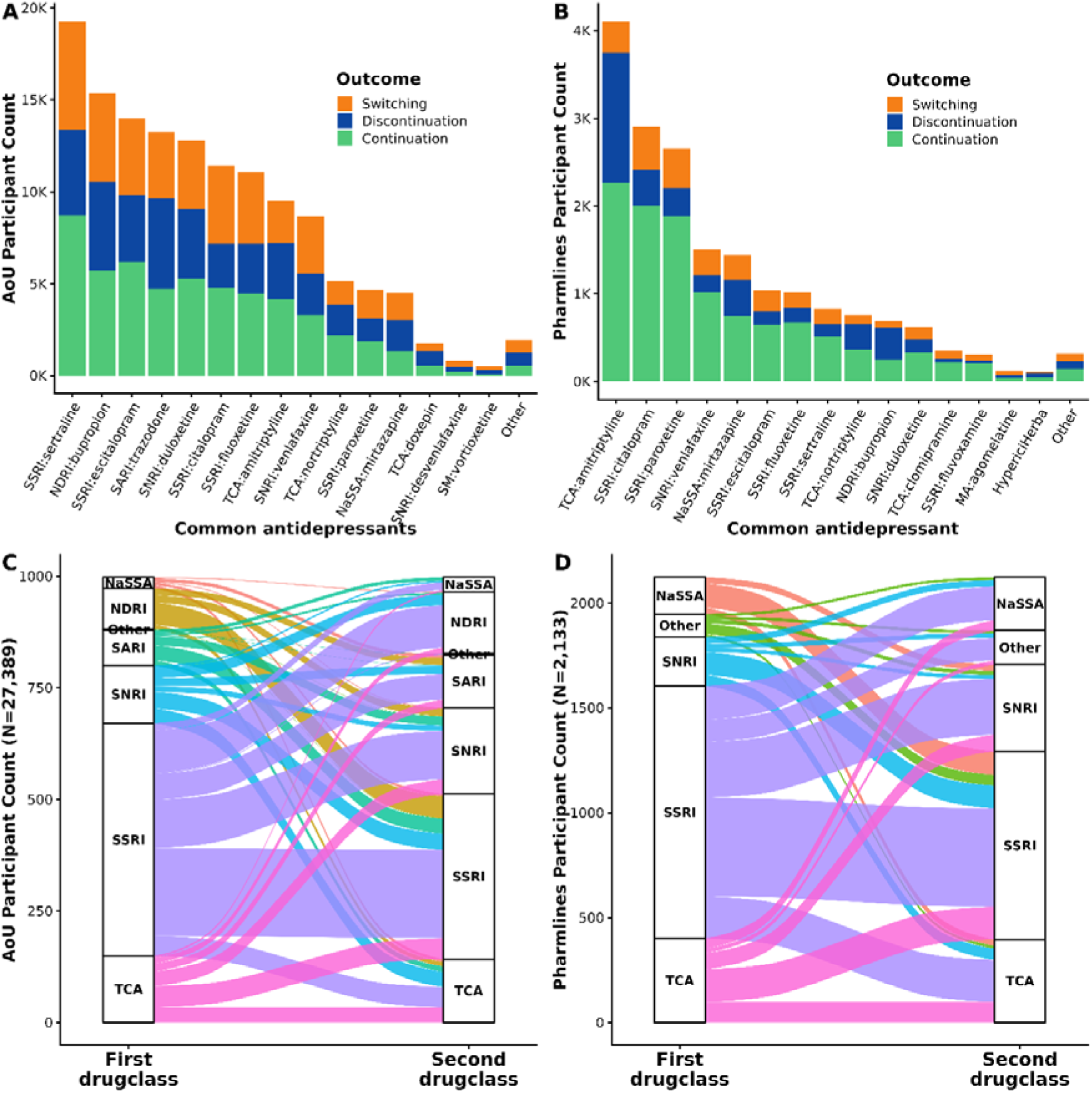
Common antidepressant treatment patterns and transitions by drug. **(A-B)** Bar plots showing frequency of the monotherapy treatment outcomes continuation, discontinuation and switching, split by common antidepressants. Plots show the 15 most common antidepressants, with the rest grouped into “Other” antidepressants. **A**. AoU. **B**. Pharmlines. To be included participants needed ≥28-days of monotherapy with the drug; participants may have an associated outcome with more than 1 antidepressant. (**C–D**) Sankey diagrams showing transitions between the first (monotherapy) and second recorded antidepressant drug classes among individuals who switched treatments. The left column represents the class of the first prescribed antidepressant and the right column represents the class of the subsequent antidepressant. Flow widths correspond to the number of individuals transitioning between drug classes. Panel C shows transitions in the AoU cohort (N = 27,389 switchers), and Panel D shows transitions in the Pharmlines cohort (N = 2,133 switchers). Antidepressant classes include tricyclic antidepressants (TCA), selective serotonin reuptake inhibitors (SSRI), serotonin–norepinephrine reuptake inhibitors (SNRI), serotonin antagonist and reuptake inhibitors (SARI), noradrenergic and specific serotonergic antidepressants (NaSSA), norepinephrine–dopamine reuptake inhibitors (NDRI), serotonin modulator (SM), melatonin agonist (MA), and other antidepressants.

### Phenotypic associations with switching and discontinuation

Among European-ancestry participants with recorded depression, switching was associated with greater psychiatric and clinical burden in both cohorts (**Figure 2; Supplementary Tables S2–S3**). Anxiety showed one of the strongest replicated associations (AoU OR=2.35, 95% CI 2.17–2.54; Pharmlines OR=1.53, 1.17–1.99), and greater depression symptom burden and stress-related measures were also associated with switching in both cohorts. Cohort-specific associations were observed for additional measures including BMI, smoking and neuroticism.

**Figure 2.**
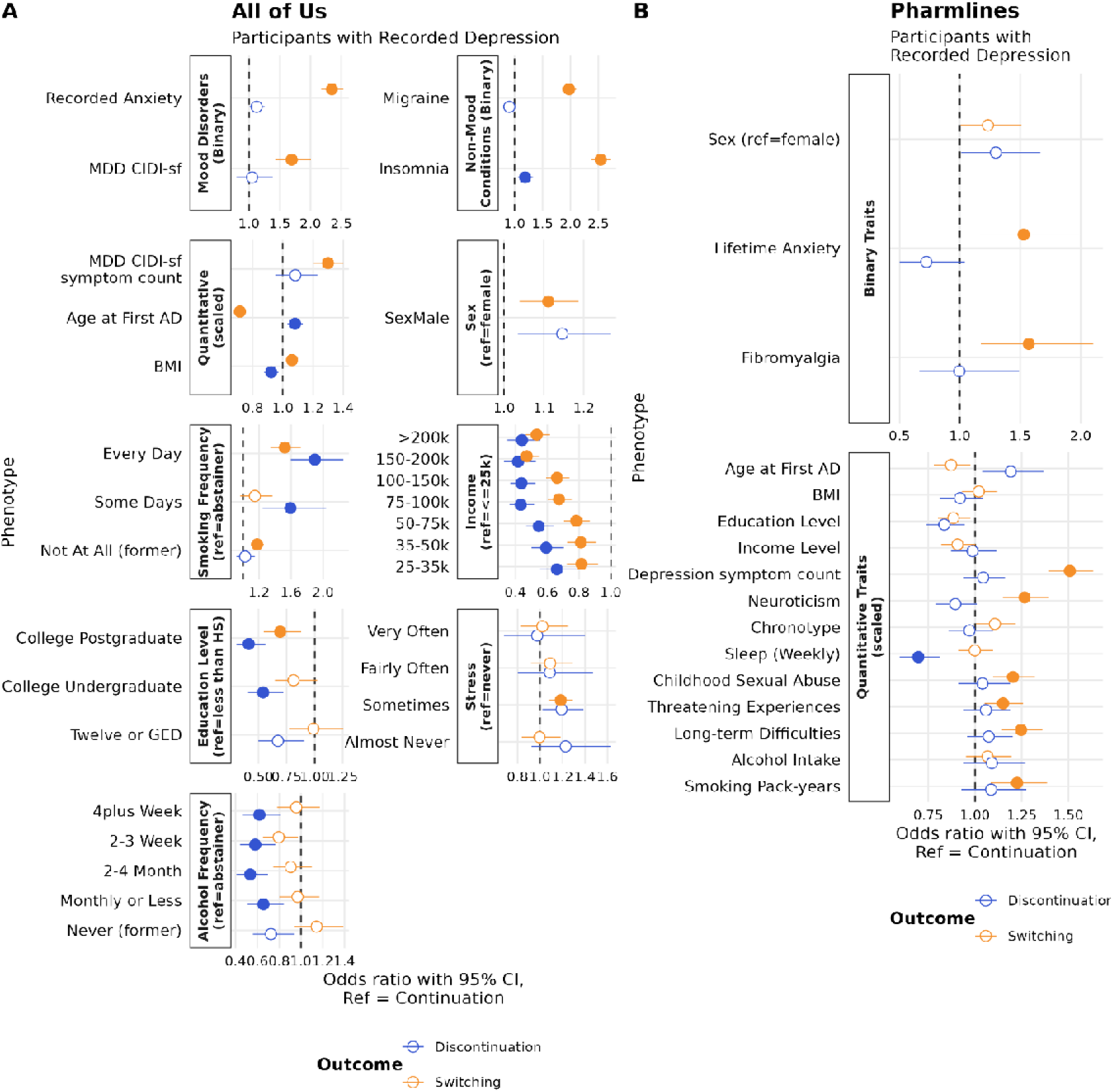
Phenotypic, demographic and lifestyle associations with antidepressant switching and discontinuation, relative to continuation, in participants of genetically inferred European ancestry with depression, across the All of Us and Pharmlines cohorts. Forest plots show adjusted odds ratios (ORs) with 95% confidence intervals from binary logistic regression models fitted separately for each exposure, contrasting switching versus continuation (orange) and discontinuation versus continuation (blue), in (**A**) All of Us participants with EHR-recorded depression and (**B**) Pharmlines participants with questionnaire-derived depression (current depression questionnaires administered to 97% of our Pharmlines participants and lifetime questionnaires administered to 49% of our Pharmlines participants). All models were adjusted for age at first antidepressant record, genetic sex/sex at birth, income, and education level. Where age or sex was the exposure of interest, the model included only age and sex, mutually adjusted. Exposures are grouped by domain: binary traits (mood and non-mood conditions in All of Us; lifetime anxiety and fibromyalgia in Pharmlines), quantitative traits (standardised to mean 0, SD 1 prior to modelling), sex (reference = female), and, in All of Us only, categorical measures, each modelled against the reference category indicated in the panel header. Because each exposure was modelled separately, model N differed across exposures. Maximum N (models in which age or sex was the exposure) was 17,576 for switching models and 10,166 for discontinuation models in All of Us, and 3,182 for switching and 3,619 for discontinuation in Pharmlines. Filled points indicate associations surviving Bonferroni correction applied within each outcome contrast (14 traits in All of Us, 17 in Pharmlines); open points are non-significant. The dashed vertical line marks OR = 1. See Supplementary Tables S2–S3 and Supplementary Methods for full details of each phenotypic exposure.

Discontinuation, defined by all observed monotherapy episodes lasting <90 days, showed a less consistent profile. In AoU, lower income and educational attainment were associated with both discontinuation and switching, with larger effects for discontinuation, whereas corresponding Pharmlines estimates were weaker. Sleep-related measures were also associated with discontinuation, including recorded insomnia in AoU and shorter sleep duration in Pharmlines. Phenotypic associations differed substantially among participants without recorded depression, supporting variation in treatment indications and in the processes represented by discontinuation (**Supplementary Figures S3–S6**). All results are further detailed in Supplemental Results.

### Polygenic associations with switching and discontinuation

Full PGS association results are provided in **Supplementary Tables S4–S9** and described in **Supplementary Results**. Among European-ancestry participants with recorded depression, depression PGS showed the clearest replicated association with switching (**Figure 3**). Each SD increase in depression PGS was associated with higher switching odds in AoU (OR=1.16, 95% CI 1.12–1.20), with a directionally concordant nominal association in Pharmlines (OR=1.11, 1.01–1.22). Insomnia, migraine and ADHD PGS were also associated with switching in AoU, with concordant effect directions in Pharmlines. The depression PGS association remained evident in AoU participants meeting CIDI-sf lifetime MDD criteria and when switching was restricted to events occurring within six months of index-treatment initiation (**Supplementary Figure S7**).

**Figure 3.**
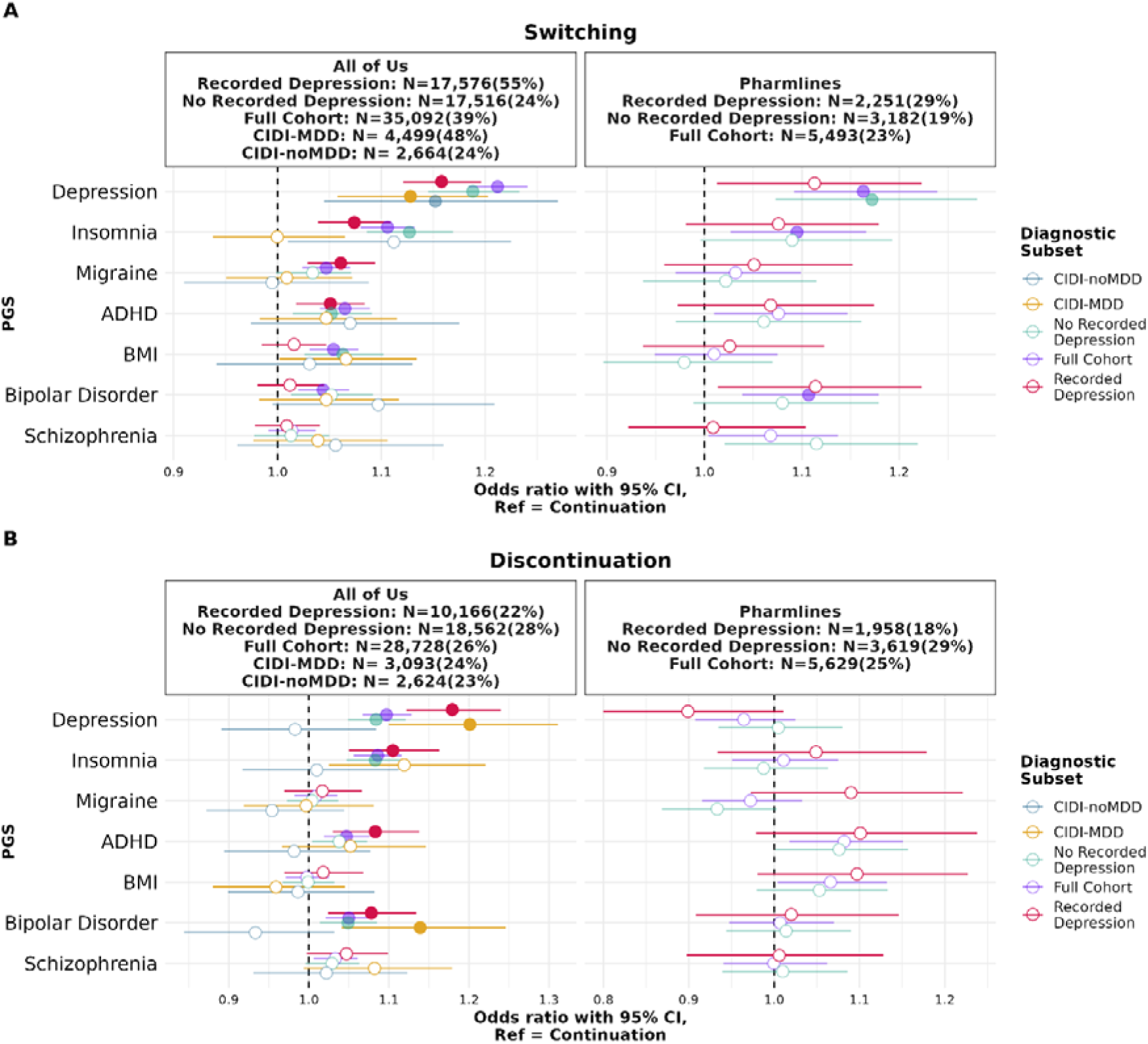
Polygenic score associations with pooled (across antidepressants) switching and discontinuation, relative to continuation, across diagnostic subsets in All of Us and Pharmlines. Forest plots show adjusted odds ratios (ORs) with 95% confidence intervals from binary logistic regression models contrasting (**A**) switching versus continuation and (**B**) discontinuation versus continuation, in European-ancestry participants from AoU (left panels) and Pharmlines (right panels). Each polygenic score (PGS) was standardised to mean 0 and SD 1 within the European-ancestry subset prior to modelling, and was modelled separately. Models included age at first recorded antidepressant, genetic sex/sex at birth, income, and education level as covariates. Results are shown separately for three diagnostic subsets: participants with a recorded depression diagnosis (red), participants without a recorded depression diagnosis (teal), and the full prescribing cohort irrespective of diagnosis (purple). Sample sizes for each subset and cohort are indicated in the panel headers; percentages in parentheses give the proportion of each modelled sample with the outcome contrasted against continuation. Filled points indicate associations significant after Bonferroni correction across 7 PGS within each stratum; open points are non-significant. The dashed vertical line marks the null (OR = 1).

PGS associations with discontinuation were less consistent between cohorts. Depression, insomnia, ADHD and bipolar disorder PGS were associated with discontinuation in AoU among participants with recorded depression, but corresponding Pharmlines estimates generally did not replicate. ADHD PGS showed the most consistent positive effect direction across cohorts. Results were largely unchanged after excluding individuals who met augmentation or combination criteria during follow-up, and in Pharmlines, the association between higher depression PGS and decreased odds of discontinuation became nominally significant (**Supplementary Figure S8**). Additional analyses among AoU participants with recorded anxiety are shown in **Supplementary Figure S9** and AoU ancestry-stratified switching and discontinuation analyses are shown in **Supplementary Figure S10**.

### Polygenic liability across treatment intensity

We next compared PGS associations across continuation, switching and augmentation. Full augmentation results across diagnostic strata are provided in **Supplementary Tables S10–S11**. Among European-ancestry AoU participants with recorded depression, the depression PGS point estimate was larger for augmentation (OR=1.29, 95% CI 1.23–1.35) than for switching (**Figure 4**). Insomnia and ADHD PGS showed a similar ordering. Schizophrenia and bipolar disorder PGS, in contrast, were associated with augmentation (schizophrenia OR=1.09, 95% CI 1.05–1.14, bipolar disorder OR=1.08, 1.03–1.13) but not switching. Depression, bipolar disorder and schizophrenia PGS associations with augmentation remained significant when analyses were restricted to participants meeting CIDI-sf lifetime MDD criteria.

**Figure 4.**
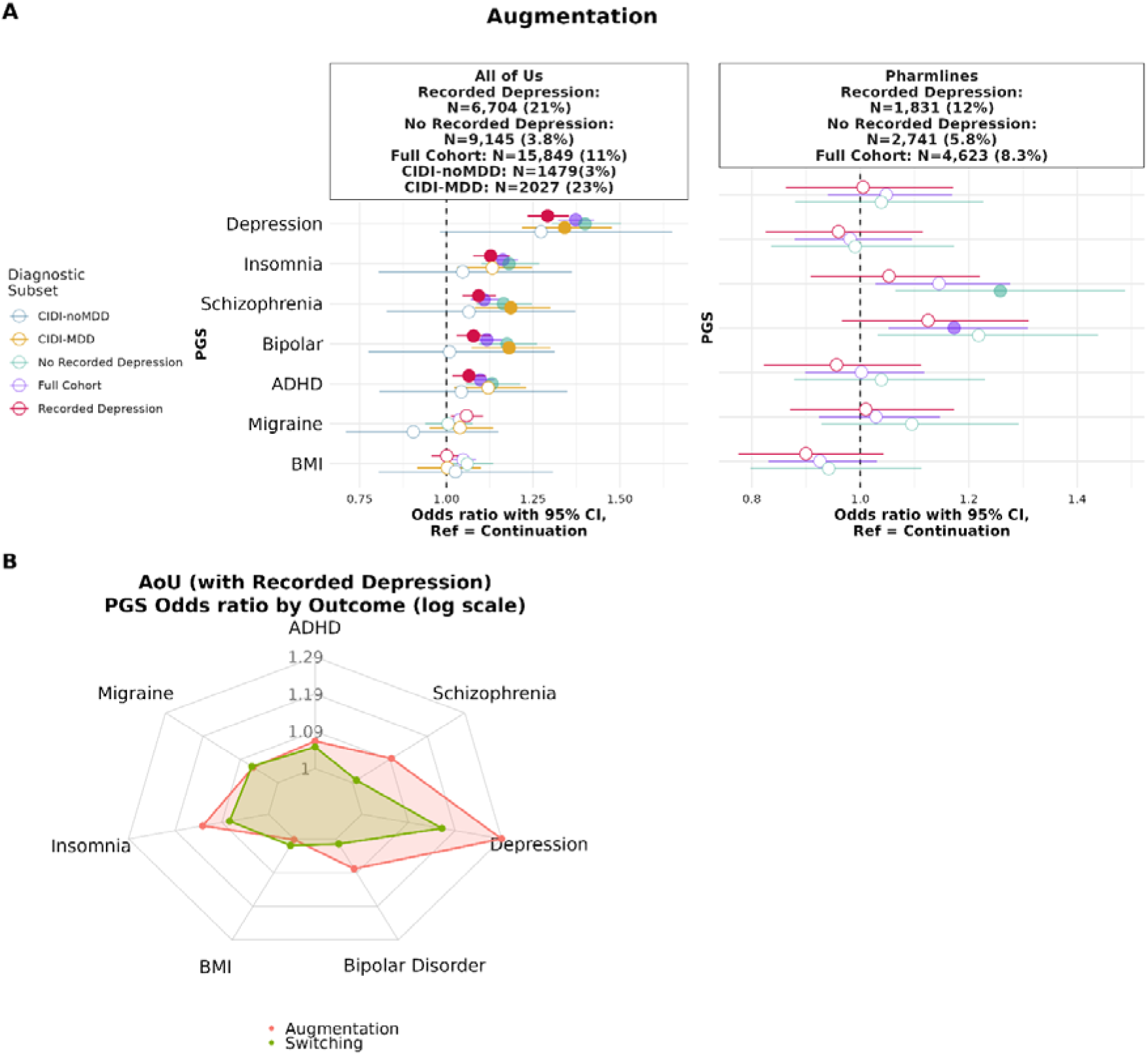
Polygenic score associations with antidepressant augmentation across diagnostic subsets, and comparison of PGS effects across treatment intensity outcomes. **(A)** Forest plots show adjusted odds ratios (ORs) with 95% confidence intervals from binary logistic regression models contrasting augmentation versus continuation in European-ancestry participants from AoU (left) and Pharmlines (right). Augmentation was defined as ≥28-day overlap between an antidepressant and an atypical antipsychotic or lithium. Each PGS was standardised to mean 0, SD 1 within the European-ancestry subset prior to modelling, and was modelled separately. Models included age at first antidepressant record, sex, income and education level as covariates. Results are shown for three diagnostic subsets: participants with a recorded depression diagnosis (red), without a recorded depression diagnosis (teal), and the full prescribing cohort (purple). Sample sizes and the proportion meeting augmentation criteria are indicated in the panel headers. Filled points indicate associations significant after Bonferroni correction across seven PGS within each stratum; open points are non-significant. The dashed vertical line marks the null (OR = 1). (**B**) Radar plot comparing PGS odds ratios for augmentation (red) and switching (green) versus continuation in AoU participants with a recorded diagnosis of depression, displayed on a logarithmic OR scale. Each axis corresponds to one of the seven PGS shown in Panel A.

We additionally fitted an ordinal model ordering continuation, switching and augmentation by increasing treatment intensity (**Supplementary Table S12** and **Supplementary Figure S11**). This ordering was intended to test whether polygenic liability increased across treatment-modification categories rather than to imply a necessary temporal progression between them. The model supported increasing polygenic liability across these categories in AoU. Depression PGS showed the strongest association with being in a higher treatment-intensity category (ordinal OR=1.18 per SD, 95% CI 1.15–1.21), with significant gradients also observed for insomnia, ADHD, migraine and schizophrenia PGS. No PGS was significantly associated with augmentation in Pharmlines among participants with recorded depression. Comprehensive results and secondary diagnostic and ancestry analyses are reported in the **Supplementary Results**, with ancestry-stratified augmentation analyses shown in **Supplementary Figure S12**.

### PGS associations with antidepressant class- and drug-specific switching

There was little evidence that PGS associations with switching differed by antidepressant (**Supplementary Tables S13–S15, Supplementary Figures S13–S15**). Among participants with recorded depression, depression PGS was positively associated with switching across all six antidepressant classes examined in AoU, reaching significance in the three largest strata (SSRI OR=1.16, 95% CI 1.12–1.21; NDRI OR=1.18, 1.10–1.27; SNRI OR=1.11, 1.04–1.18), with smaller and less precise estimates for TCA, SARI and NaSSA. However, the joint depression PGS × drug-class interaction was null in both AoU (p=0.46) and Pharmlines (p=0.44), and no PGS × drug-class or PGS × individual-drug interaction survived multiple-testing correction. Thus, although modest treatment-specific effects cannot be excluded, there was no evidence that the depression PGS association with switching differed systematically across antidepressant drugs or classes.

### Familial aggregation of antidepressant treatment outcomes

Familial aggregation in Pharmlines was confined to continuation phenotypes (**Figure 5; Supplementary Tables S16–S17**). The strongest liability correlations were observed for SNRI continuation (pooled r=0.27, 95% CI 0.09–0.42) and SSRI continuation (r=0.21, 0.12–0.31), with weaker aggregation for continuation overall (r=0.12, 0.02–0.22). Switching and discontinuation showed little evidence of familial aggregation (r≈0; recurrence-risk ratios ≈1). Corresponding RR estimates and their comparison with tetrachoric correlations are shown in **Supplementary Figures S16–S17.**

**Figure 5.**
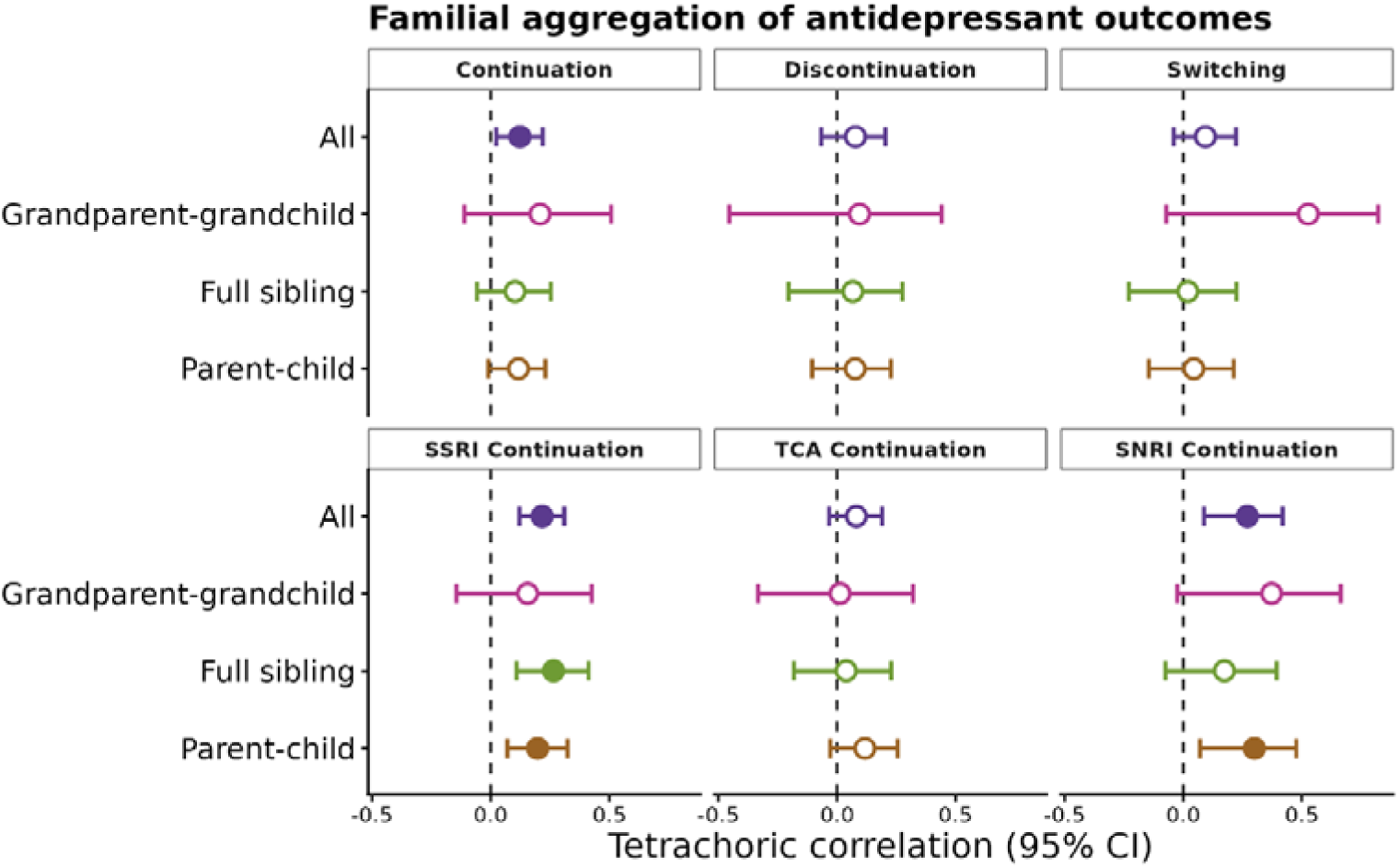
Familial aggregation of antidepressant treatment outcomes, shown as tetrachoric correlations of the underlying liability with 95% bootstrap percentile confidence intervals (family-resampled, 1000 replicates). Each panel is an outcome; rows are relationship types (parent–child, full sibling, grandparent– grandchild, and all relationships pooled ["all" analysed as yes/no for any recorded relative]). Filled points indicate intervals excluding zero (the dashed line); open points are non-significant. Colour denotes relationship type.

## Discussion

In two large cohorts, AoU and Pharmlines, switching relative to continuation showed the most reproducible polygenic association. Switchers, but not discontinuers, had greater depression severity, stress-related burden and anxiety, and higher depression PGS was associated with switching in AoU with a concordant association in Pharmlines. This differs from Lo et al., who found no association between MDD PGS and SSRI switching in UK Biobank, although switching was modestly predicted by a PGS derived from clinically assessed antidepressant non-remission^7^. In the same resource, Kamp et al. found that higher depression PGS was associated with self-reported SSRI non-response, although drug-specific associations were weaker^36^. Our findings are more consistent with Lapinska et al., who found higher depression PGS among broader treatment non-responders and individuals with TRD in AoU and ATLAS^18^. However, their European-ancestry AoU comparison of SSRI responders and non-responders was not significant^18^. In contrast, we observed a significant depression PGS association with SSRI switching in AoU, alongside significant associations in the other two largest antidepressant classes, with concordant effects across classes and in Pharmlines. Differences across studies may partly reflect variation in switching definitions and comparison groups^7,8,18^. Our definition was anchored to qualifying monotherapy episodes and their discontinuation, with explicit separation of switching from combination therapy and augmentation.

Several findings suggest that the association between depression PGS and switching primarily reflects broader depression liability associated with treatment difficulty rather than differential polygenic response between antidepressants. Depression PGS was positively associated with switching across all six antidepressant classes examined in AoU, while formal PGS × drug-class interactions were null in both cohorts and no PGS × individual-drug interaction survived multiple-testing correction. Greater CIDI-sf depression symptom burden also predicted switching, and depression PGS increased across the ordered continuation, switching and augmentation categories, consistent with previous associations between depression polygenic liability and broader TRD or difficult-to-treat depression phenotypes^10,18,19,37^. Together, these findings support depression PGS more strongly as a marker of treatment prognosis than of treatment selection. They do not imply that antidepressant response is genetically homogeneous across treatments or that treatment-specific genetic effects do not exist. Consistent with the possibility of additional genetic influences, Koch et al. found that loci and most genetic correlations for antidepressant non-response persisted after conditioning on depression^38^. Switching may therefore reflect both broader liability to a difficult treatment course and treatment-specific processes not captured by depression PGS. Distinguishing these components will require more direct information on why treatment was changed, including efficacy, tolerability and adverse-effect measures.

Augmentation showed a different polygenic profile from switching. Among AoU participants with recorded MDD, schizophrenia and bipolar disorder PGS were associated with augmentation but not switching, with similar associations among those meeting CIDI-sf lifetime MDD criteria. This may reflect broader cross-disorder psychiatric liability among individuals progressing to more intensive treatment, including subthreshold or unrecorded bipolar-spectrum or psychotic features. Participants with recorded bipolar or psychotic disorders were excluded, making treatment of those diagnosed disorders an unlikely explanation. This is consistent with evidence of greater familial schizophrenia liability in psychotic than non-psychotic forms of depression^39^, although treatment selection may also contribute. In the UK Biobank, Wang et al. found that MDD and ADHD PGS were associated primarily with antidepressant-change phenotypes, whereas bipolar disorder PGS was more strongly associated with lithium, valproate and ECT, and schizophrenia PGS with antipsychotic treatment^10^. Xu et al. similarly found no schizophrenia PGS association with their primary antidepressant-defined TRD phenotype, but an association emerged when antipsychotic treatment was incorporated into the definition^11^. Together, these findings suggest that augmentation may capture greater psychiatric complexity than antidepressant switching alone, while also partly reflecting genetic liabilities associated with the selection of particular augmentation strategies. The absence of corresponding associations in Pharmlines may partly reflect lower power and differences in treatment capture, as augmentation in the Dutch system^40^ often follows specialist referral whereas Pharmlines primarily captures community-pharmacy dispensing. Consistent with this, augmentation was less frequent among participants with recorded MDD in Pharmlines than AoU.

In contrast, most PGS associations with discontinuation, defined by antidepressant monotherapy episodes consistently lasting <90 days across the observed treatment history, showed limited cross-cohort consistency. ADHD PGS was the clearest exception, with similar positive effect estimates in both cohorts. Previous evidence linking ADHD with medication non-adherence and discontinuation^41–43^, suggests that this may reflect reduced treatment persistence rather than antidepressant-specific response^44^. Sleep-related measures also showed some cross-cohort convergence, with recorded insomnia in AoU and shorter sleep duration in Pharmlines associated with greater discontinuation. Socioeconomic factors were strongly associated with both discontinuation and switching in AoU, particularly among individuals with depression, but corresponding Pharmlines estimates were weaker. This difference may partly reflect variation between the fragmented, insurance-dependent US healthcare system^45,46^ and the greater continuity of Dutch primary care^47^, although lower power in Pharmlines is also likely. Overall, the weak reproducibility of discontinuation associations is consistent with discontinuation representing multiple processes, including remission, adverse effects, non-adherence, disengagement from care and structural barriers^48–50^. Discontinuation may consequently be less suitable than switching as a single proxy for antidepressant non-response without additional information on why treatment ended.

Familial analyses provided a complementary perspective. Continuation aggregated within Pharmlines families, whereas switching and discontinuation did not. Sustained, switch-free use may therefore capture stable family-level influences on treatment persistence, including biological factors related to tolerability as well as shared environment, healthcare context and attitudes towards medication. Family history of medication response is commonly considered clinically, as described in standard psychiatric texts^51,52^, although evidence that it predicts antidepressant response remains limited^21^. The stronger aggregation observed for SSRI and SNRI continuation is therefore of interest, but overlapping confidence intervals preclude conclusions about differences between classes. These estimates should therefore be interpreted as family-level clustering rather than direct evidence of genetic effects, as familial resemblance reflects both genetic and shared-environmental influences^53,54^.

Several limitations apply. Medication records cannot distinguish efficacy-driven from tolerability-driven switching or confirm medication ingestion and adherence. Requiring ≥28 days of monotherapy excludes very early treatment changes but cannot eliminate this ambiguity. AoU and Pharmlines also differ in data capture: AoU primarily contains EHR medication records spanning primary and specialist care, whereas Pharmlines records community-pharmacy dispensing, potentially contributing to differences in outcomes such as augmentation. Treatment indication is also imperfectly observed. Depression was primarily identified using EHR codes in AoU and questionnaire/interview measures in Pharmlines, while only a subset of AoU participants completed the CIDI-sf; absence of recorded depression therefore cannot be interpreted as absence of lifetime MDD. Medication histories are additionally left-truncated, meaning age at first antidepressant record does not necessarily represent age at treatment initiation (**Supplementary Figures S18-S19**). Finally, PGS associations were weaker and less precise in non-European ancestry groups, reflecting both smaller samples and established limitations in cross-ancestry PGS portability^55^. Our study focused on characterising the meaning of record-derived treatment phenotypes rather than estimating their genome-wide architecture; applying these definitions in larger GWAS meta-analyses represents an important next step.

In summary, antidepressant switching was reproducibly associated with greater depression polygenic liability and clinical severity across two healthcare systems. There was little evidence that the depression PGS association differed between antidepressant drugs or classes, suggesting that broader depression liability contributes to switching across treatments. How much switching additionally reflects treatment-specific pharmacological non-response remains unresolved. Augmentation was associated with additional bipolar disorder and schizophrenia liability in AoU, discontinuation showed a less reproducible association profile and appeared to capture a broader mixture of processes, and familial aggregation was confined to switch-free continuation. Different prescription-derived antidepressant outcomes therefore appear to capture distinct components of treatment course and should not be treated interchangeably as measures of antidepressant response.

## Supporting information

Supplementary_Methods_and_Results

Suppementary_Figures

## Acknowledgments

We gratefully acknowledge All of Us participants for their contributions, without whom this research would not have been possible. We also thank the National Institutes of Health’s All of Us Research Program for making available the participant data examined in this study. We also gratefully acknowledge the services of the Lifelines Cohort Study, the contributing research centres delivering data to Lifelines, and all the study participants, without whom this research would not have been possible.

## Data Availability

This study used data from the All of Us Research Program’s Controlled Tier Dataset 8 (CDRv8; C2024Q3R4), available to authorized users on the Researcher Workbench. Lifelines data may be obtained from a third party and are not publicly available. Researchers can apply to use the Lifelines data used in this study. More information about how to request Lifelines data and the conditions of use can be found on their website.

## Acknowledgment of funds

The Lifelines initiative has been made possible by subsidy from the Dutch Ministry of Health, Welfare and Sport, the Dutch Ministry of Economic Affairs, the University Medical Center Groningen (UMCG), University of Groningen and the Provinces in the North of the Netherlands (Drenthe, Friesland, Groningen). NRW acknowledges funding from the Michael Davys Trust, University of Oxford, National Health and Medical Research Council (grant numbers 1173790 and 1113400) and the Wellcome Trust (grant number 226770/Z/22/Z).

## Code Availability

The code used to analyse the cohorts is available in the GitHub repository: https://github.com/walkeralicia/PGS_of_Antidepressant_Treatment_Escalation/.

## Contributions

A.W. conceptualised the study, conducted the formal analysis, and wrote the original draft. A.W. and T.L. generated the SBayesRC weights. X.W. curated the quality-controlled All of Us genotype data. J.B., F.K. and E.H. provided access to and guidance on IADB.nl. I.N., H.S., R.B., P.M.V., A.K.H. and N.R.W. generated the quality-controlled Lifelines UGLI imputed genotype data. C.H., H.M.v.L., and M.T. provided psychiatric expertise in psychiatric interpretation, and C.H. and H.M.v.L. additionally advised on the Lifelines phenotypic data. N.R.W. conceptualised the familial aggregation analyses, and acquired funding for the Pharmlines analyses. M.T. and N.R.W. supervised the work. All authors reviewed and edited the manuscript and approved the final version.

