## Supplementary_Methods_and_Results for "Polygenic and familial contributions to antidepressant continuation, switching, discontinuation and augmentation in the All of Us and Pharmlines cohorts"

*Walker A. et al.*

### 1 Supplementary Methods

#### 1.1 Data sources and linkage

This study used data from Lifelines^1^ and the AoU Research Program^2^. Lifelines is a multi-disciplinary prospective population-based cohort study examining in a unique three-generation design the health and health-related behaviours of 167,729 persons living in the North of the Netherlands. It employs a broad range of investigative procedures in assessing the biomedical, socio-demographic, behavioural, physical and psychological factors which contribute to the health and disease of the general population, with a special focus on multi-morbidity and complex genetics. Baseline assessments took place 2007-2013 with multiple waves of follow-up assessments. The UMCG Medical Ethical Committee is under number 2007/152.

Lifelines participants were linked to community pharmacy dispensing data from the IADB.nl community pharmacy dispensing database, which contains dispensing records dating back to 1994 and covers approximately 1.5 million individuals. All participants provided informed consent for linkage of their data to external healthcare sources at enrolment. Linkage was conducted securely at Statistics Netherlands (CBS) using sex, birthdate, and postal code; approximately 66% of Lifelines participants were successfully linked. CBS linkage included dispensing records with drug names, dispense date, dose, and quantity. When Lifelines is linked with dispensing data from the IADB.nl the cohort is called Pharmlines.

AoU is a large U.S. biobank linking electronic health records (EHRs). Data from AoU were accessed on 14^th^ October 2025 using the Controlled Tier Curated Data Repository version 8 (CDRv8; C2024Q3R4) for N=633,547 participants. AoU contributed EHR-derived medication data for participants, with antidepressant medication records (AoU parent OMOP concept IDs 21604686, 21604729, and 21604788, including their descendent concepts) spanning from 13 January 1982 to 1 October 2023. Medication records included drug name, dispense date, and, where available, days' supply or explicit end dates. AoU aggregates records from multiple healthcare organizations, including academic medical centers, federally qualified health centers (primary care clinics serving underserved populations), and Veterans Health Administration facilities.

For both IADB.nl and AoU, medication records written or filled outside affiliated systems/pharmacies may not have been captured.

#### 1.2 Participant selection criteria

Briefly, following data cleaning of antidepressant records — including removal of records with implausible dates or substances not meeting the definition of an antidepressant (e.g., amino acids, nutritional supplements) — we took forward participants who had at least one antidepressant monotherapy episode lasting ≥28 days (see below) and who were ≥12 years of age at their first antidepressant record. Within AoU, we additionally excluded participants with any EHR-recorded diagnosis of bipolar disorder (OMOP concept ID 436665) or a psychotic disorder (OMOP concept ID 436073), including descendant concepts, as antidepressant prescribing in these groups reflects distinct clinical indications and treatment pathways that fall outside the depression context of this analysis. These diagnoses were not assessed via structured instruments in Lifelines, however, we also excluded those with self-report schizophrenia and bipolar disorder in Lifelines. After applying these criteria, we attained AoU N=98,357 and Pharmlines N=12,884 participants in the analytical cohorts. Because medication histories were left-truncated by data availability, age at first antidepressant record is unlikely to be age at first antidepressant medication (**Supplementary Figure S18-S19**).

#### 1.3 Defining primary outcomes for individual antidepressants

**Step 1: Identifying antidepressant medication episodes**

In the Lifelines IADB.nl database (PharmLines), antidepressant dispenses (N06AXXX) with non-specific ATC codes (i.e., N06A without further specification) were excluded, as they did not allow identification of the specific antidepressant. Likewise, in AoU, antidepressant drug names (AoU OMOP concept IDs 21604686, 21604729, and 21604788) not meeting the definition of an antidepressant (e.g., amino acids, nutritional supplements) were removed. The antidepressants considered in each cohort are listed in **Supplementary Table S18**. For the majority of antidepressant records (~95% in AoU and 99.5% in Lifelines), sufficient information was available to estimate medication record duration (PD), either through an explicit exposure end date (available in AoU) or through dosage information (available in both datasets). When dosage information was used, PD was calculated as the total prescribed dose divided by the daily dose:

$$PD =\left( \frac{Total Dose}{Daily Dose} \right)$$

Alternatively, when an explicit end date was available (AoU only), PD was calculated as the difference between the end date and the start date of the medication record. All time variables were expressed in days. Medication records with an estimated PD of less than 1 day were considered implausible and excluded, as these likely reflect substantial inconsistencies in recorded dosage or quantity fields. For records with insufficient information to calculate PD (approximately 5% in AoU and 0.05% in Lifelines), including missing dosage data or invalid daily doses (e.g., recorded as zero, for which PD could not be calculated), a fixed duration equal to the median antidepressant record duration in each dataset was assigned (30 days in both AoU and Lifelines), reflecting typical dispensing practices.

Next, for each individual, records were indexed by order of dispensing i and by drug k, such that $P_{i,k}$ denotes the i-th dispensed record of drug k. The expected end date of a record $P_{i,k}$, denoted $EED(P_{i,k})$, was calculated by adding the estimated record duration, ${PD}_{i,k}$, to the dispense date, $DD(P_{i,k}),$of the record, assuming individuals take the first dose on the day of dispense.

$$EED(P_{i,k})=DD(P_{i,k}) +{PD}_{i,k}$$

For each individual i, records for drug k were grouped into medication episodes $T_{j,k}$ over the follow-up period. The first record $P_{1,k}$, was assigned to the first episode $T_{1,k}$. Each subsequent record was assigned to an episode according to the following rule:

If the dispense date of the record i+1 occurred less than 90-days after the expected end date of the record i, it would be assigned to the same medication episode as record i (i.e., requiring a 90-day clearing period between consecutive medication episodes as done in a prior study^3^). Otherwise, if the dispense date occurred 90-days or more after the expected end date of the previous record, it would be assigned to a new medication episode.

Mathematically:

$if DD(P_{i,k}) -EED(P_{i-1,k})<90:$ $P_{i,k} \in T_{j,k}$

$$else:$$

$$P_{i,k} \in T_{j+1,k}$$

Each medication episode $T_{j,k}$, was then defined as:

- Starting on the dispense date of the first prescription in that episode, start date (SD):

$${SD(T}_{j,k})=min\left\{ DD\left( P_{i,k} \right) \in T_{j,k} \right\}$$

- Ending on the expected end date of the last prescription in that episode:

$$EE{D(T}_{j,k})= max\left\{ EED(P_{i,k}) \in T_{j,k} \right\}$$

**Step 2: Identifying antidepressant monotherapy episodes**

Monotherapy episodes were defined as periods of continuous use of a single index antidepressant, permitting brief (<28-day) cross-tapers at episode boundaries. Cross-tapers at the start of an episode were excluded from monotherapy duration by shifting the effective episode start date to the end of the overlap period. Cross-tapers at the end of an episode were permitted only if two conditions were met: (i) the overlap began at least 28 days after the effective episode start date, ensuring at least 28 days of preceding monotherapy, and (ii) the interval between overlap initiation and the medication episode end was <28 days. In these cases, the overlap period was excluded from the final monotherapy duration, $D\left( T_{j,k} \right)$. Prescription episodes that did not contain any qualifying period of monotherapy consequently had a monotherapy duration of zero.

**Step 3: Labelling individual medication episodes: Continuation or Discontinuation**

Over follow-up, individuals could have multiple medication episodes for the same antidepressant, reflecting treatment interruptions, restarts or separate treatment periods. Prescriptions separated by <90 days were combined into the same medication episode as described above.

We used 90 days of monotherapy to distinguish sustained continuation from shorter treatment episodes. This threshold was informed primarily by dispensing patterns in IADB.nl, where antidepressants were most commonly dispensed for 30 days, with 60- and 90-day durations also frequent. Dutch pharmacies typically dispense a full box for initial prescriptions, containing 30 days of medication (three 10-pill blister packages). Subsequent prescriptions commonly cover 60 or 90 days. Therefore, reaching 90 days of monotherapy generally required treatment to continue beyond an initial dispensing. The threshold is also broadly compatible with the timescale over which major depressive episodes commonly evolve, with a median episode duration of approximately 90 days reported in the Dutch general population^4^.

Accordingly, if the monotherapy duration of the *j*-th prescription episode for the *k*-th drug, $D\left( T_{j,k} \right)$ was ≥ 90-days, it was labelled as ‘continuation’, otherwise, if the duration of the episode is ≥28 to <90 days it was labelled as ‘discontinuation’. Episodes with <28 days do not become qualifying discontinuation episodes. Individuals could have multiple medication episodes with different outcomes for the same drug.

Mathematically:

$$OUTCOME\left( T_{j,k} \right)=\left\{ \begin{aligned} 'continuation^{'}, if D\left( T_{j,k} \right) \geq90 \\ 'discontinuation^{'}, otherwise \end{aligned} \right.$$

Discontinuation could occur for multiple reasons, including response followed by a shorter-than-standard treatment duration, transition from monotherapy to augmentation or combination therapy, or insufficient subsequent antidepressant exposure to meet switching criteria. Discontinuation therefore represents a residual category encompassing several distinct treatment-course scenarios.

**Step 4: Assigning antidepressant drug outcomes as switching, discontinuation or continuation**

Drug-specific treatment outcomes were defined as person-by-drug summary phenotypes derived from each individual’s complete observed treatment history rather than from the first eligible medication episode alone. This approach allowed treatment patterns across multiple episodes of the same antidepressant to contribute to classification and reduced reliance on the first observed episode, which may not represent the individual’s first lifetime treatment because medication histories are left-truncated.

For each antidepressant, an individual was classified as having continued treatment if at least one qualifying monotherapy episode met the continuation criterion of ≥90 days ($C_{k}=1)$. Thus, an isolated shorter episode did not override evidence that the individual had sustained monotherapy with that drug on another occasion. If no episode met the continuation criterion and all qualifying monotherapy episodes were classified as discontinuation, the individual was classified as having discontinued that drug ($C_{k}=0$). This definition identifies individuals whose observed use of that antidepressant was consistently limited to shorter monotherapy episodes without evidence of sustained continuation.

Mathematically:

$$C_{k}=\left\{ \begin{aligned} 1, if \sum_{j=1}^{m} 1\left( OUTCOME\left( T_{j,k} \right)=continuation \right) \geq1 \\ 0, otherwise. \end{aligned} \right.$$

Next, we determined the final outcome for each antidepressant using treatment records for both the index drug and all other antidepressants. Switching was defined as transition from an index antidepressant to a different antidepressant, including switches within or between drug classes. Each person-by-drug outcome was ultimately classified as switching, continuation or discontinuation.

Building on the Lancet definition of switching in TRD^5^ and other commonly used implementations in electronic health record studies^6-8^, we adapted the criteria to account for variation in treatment duration and prescribing delays. Previous definitions generally identify switching according to the timing of a subsequent antidepressant relative to initiation of the index treatment. We instead anchored switching to the end of the index monotherapy episode because treatment duration varied between individuals and medication end dates could be reconstructed from dispensing records. This provided a consistent post-discontinuation window for identifying subsequent treatment changes.

A qualifying switch required at least 28 days of preceding index-antidepressant monotherapy, consistent with the lower bound of the period typically used in clinical practice to assess antidepressant response, generally 4 to 8 weeks^9,10^. A different antidepressant then had to be initiated either during a brief cross-taper of <28 days at the end of the index episode or within 90 days after index discontinuation. The subsequent antidepressant did not need to form a qualifying monotherapy episode of ≥28 days, and a single subsequent antidepressant record within the switching window was sufficient to classify the index treatment as a switch. The 90-day post-episode window provided a consistent interval across individuals while allowing for delays in clinical review and dispensing. Unlike the initiation-anchored definition proposed by Koch et al.^5^, our definition also required evidence that use of the index antidepressant ended or was limited to a brief cross-taper. This reduced the possibility that sustained concurrent treatment with two antidepressants was classified as switching rather than combination therapy. The definition was therefore intended to capture clinically plausible switching while distinguishing it from sustained combination treatment and augmentation.

This definition therefore captured two switching patterns. The first comprised end-of-episode cross-tapers in which the subsequent antidepressant was initiated <28 days before the end of the index episode. The second comprised switches following discontinuation in which the subsequent antidepressant was initiated within 90 days after the index episode ended. Sustained overlap of ≥28 days was not classified as switching and was instead treated as combination therapy, as described below.

Each individual was then hierarchically assigned one mutually exclusive outcome for each antidepressant. Switching took precedence when any qualifying switch from that drug was observed. Among individuals without a qualifying switch, continuation was assigned when at least one monotherapy episode lasted ≥90 days. Discontinuation was assigned when no episode met the continuation criterion and all qualifying monotherapy episodes lasted ≥28 but <90 days. Individuals could therefore have different outcomes for different antidepressants. Formally, the antidepressant drug k was classified as

1. **Switching (**$\boldsymbol{R}_{\boldsymbol{k}}\boldsymbol{=2)}$

If there existed at least one medication episode $T_{j,k}$ such that:

a. the effective monotherapy episode duration D($T_{j,k}$) ≥ 28 days, and
b. a different antidepressant was dispensed within the switching window, defined as −28 < (dispense date of subsequent antidepressant − end date of index episode) ≤ 90 days.

2. **Continuation (**$\boldsymbol{R}_{\boldsymbol{k}}\boldsymbol{=1)}$

If the switching condition is not met, and $C_{k}=1$ (i.e., drug k was continued for ≥90 days monotherapy for at least one prescription episode).

3. **Discontinuation** $\boldsymbol{(R}_{\boldsymbol{k}}\boldsymbol{=0)}$

If the switching condition is not met, and $C_{k}=0$ (i.e., drug k was discontinued early for each prescription episode).

#### 1.4 Pooled antidepressant and drug-class specific outcomes

To increase statistical power and identify factors associated with antidepressant treatment outcomes more generally, we derived participant-level outcomes pooled across all antidepressants and separately within antidepressant drug classes.

**Pooled outcomes across all antidepressant drugs**

Let $R_{A}$ represent the overall treatment outcome for each participant. Outcomes were assigned hierarchically as follows.

1) Switching ($R_{A}=2)$

If the participant had a qualifying switch from any antidepressant (i.e., $R_{k}=2$ for any k), the pooled outcome was classified as switching ($R_{A}=2)$. The subsequent antidepressant could belong to the same or a different drug class.

2) Continuation ($R_{A}=2)$

If no qualifying switch occurred and the participant had at least one drug-specific continuation outcome, the pooled outcome was classified as continuation ($R_{A}=1)$.

3) Discontinuation ($R_{A}=0)$

- If no qualifying switch occurred and no antidepressant met the continuation criterion, with all qualifying drug-specific outcomes classified as discontinuation, the pooled outcome was classified as discontinuation ($R_{A}=0)$.

**Outcomes within antidepressant drug classes**

Let $R_{S}$ denote the treatment outcome for drug class s, based on the drug-specific outcomes $R_{k}$ for antidepressants k belonging to that class. Class-level outcomes were assigned hierarchically as follows.

1) Switching ($R_{s}=2$):

If a participant had a qualifying switch from any antidepressant within class s to a different antidepressant, the class-level outcome was classified as switching. The subsequent antidepressant could belong to the same or a different drug class.

2) Continuation ($R_{s}=0$):

If no qualifying switch from an antidepressant within class s occurred and at least one antidepressant within class s met the continuation criterion, the class-level outcome was classified as continuation.

3) Discontinuation ($R_{s}=1$):

If no qualifying switch from an antidepressant within class s occurred and no antidepressant within class s met the continuation criterion, with all qualifying drug-specific outcomes classified as discontinuation, the class-level outcome was classified as discontinuation.

#### 1.5 Exclusion criteria by analysis and augmentation therapy

Prescription episodes for atypical antipsychotics and lithium were constructed using the same algorithm described in Step 1, with a median prescription duration of 15 days used for imputation in Lifelines. No duration imputation was required in AoU. The atypical antipsychotics considered in each cohort are listed in **Supplementary Table S19**.

Following later-line treatment strategies described in Dutch secondary-care depression guidelines^11^, we defined augmentation as ≥28 days of overlap between an antidepressant episode and either lithium or an atypical antipsychotic episode. Augmentation was analysed as a separate treatment outcome. Participants meeting augmentation criteria were excluded from the continuation reference group in augmentation-versus-continuation and discontinuation-versus-continuation analyses.

Combination therapy was defined separately as ≥28 days of overlap between two different antidepressants. Combination therapy was treated as an exclusion rather than an investigated outcome because sustained concurrent use of two antidepressants could represent either an extended cross-taper or intentional combination treatment and could not be classified unambiguously as a monotherapy-to-replacement switch. Participants meeting combination-therapy criteria were therefore excluded from both switching and continuation groups.

Participants with augmentation or combination therapy remained eligible for discontinuation analyses if they otherwise met the discontinuation definition, meaning that no qualifying switch occurred and all qualifying monotherapy episodes lasted ≥28 but <90 days. Thus, augmentation or combination treatment occurring elsewhere in the observed treatment history did not itself preclude classification as monotherapy discontinuation.

#### 1.6 Switching and discontinuation definition sensitivity analyses

The primary switching definition required at least 28 days of index-antidepressant monotherapy and a subsequent antidepressant initiated either during a brief cross-taper of <28 days at the end of the index episode or within 90 days after index discontinuation. No upper limit was imposed on the duration of the index monotherapy episode, allowing switches occurring after longer periods of index treatment to qualify. However, such switches may represent a different treatment context from those occurring earlier after treatment initiation. As a sensitivity analysis for the pooled switching outcome, we therefore additionally required the subsequent antidepressant to be initiated within six months of index initiation, while retaining the same <28-day cross-taper and 90-day post-discontinuation rules. Switches occurring outside this six-month window were no longer classified as switching, and participants were reclassified according to the primary continuation and discontinuation hierarchy using their remaining qualifying treatment history.

The primary discontinuation phenotype included individuals whose qualifying monotherapy episodes were consistently short, lasting ≥28 but <90 days, even if augmentation or combination therapy occurred elsewhere in their observed treatment history. This could introduce additional treatment complexity into the discontinuation group relative to the continuation reference group, from which augmentation and combination therapy were excluded for discontinuation analyses. As a sensitivity analysis, we therefore restricted the pooled discontinuation group to individuals who met the primary discontinuation criteria and had no augmentation or combination therapy at any point during observed follow-up.

#### 1.7 Defining non-genetic exposures

We examined a set of non-genetic predictors previously implicated in antidepressant treatment outcomes. Phenotypes were harmonized conceptually across All of Us (AoU) and Lifelines but were derived from cohort-specific instruments and data sources. In AoU, exposures were obtained from survey responses, physical measurements and electronic health records. In Lifelines, exposures were derived primarily from baseline and Wave 2 assessments, with depression- and anxiety-related measures additionally incorporating later assessment waves where available. In total, 14 predictors were assessed in AoU and 17 in Lifelines.

**All of Us**

**Medication record-derived phenotypes.**

1) Age at first antidepressant record derived from date of birth (DOB) and date of first antidepressant record.

**Measurement phenotypes.**

2) BMI derived from the median value of all person-level measurements.

**Survey response phenotypes.**

Socioeconomic and demographic phenotypes were identified from survey responses.

3) Sex at birth (female or male).
4) Annual household income was captured on a 1–8 ordinal scale (1 = <$25K, 8 = ≥$200K) from the self-report question "What is your annual household income from all sources?" (OMOP concept ID 1585375).
5) Educational attainment was captured on a 1–4 ordinal scale (1 = less than a high school degree, 4 = college postgraduate degree) from the question "What is the highest grade or year of school you completed?" (OMOP concept ID 1585940).
6) Stress was captured on a 1-5 ordinal scale (1=never, 5=very often) from the self-report question “In the last month, how often have you felt nervous and "stressed"? (OMOP concept ID 40192491). Since the Social Factors survey can be taken more than once, we took the most recent response for each person.
7) Alcohol drinking frequency was captured as a 6-level factor derived from the lifetime alcohol drinking self-report question “In your entire life, have you had at least 1 drink of any kind of alcohol, not counting small tastes or sips? (By a “drink,” we mean a can or bottle of beer, a glass of wine or a wine cooler, a shot of liquor, or a mixed drink with liquor in it.)” (OMOP concept ID 1586198). Participants who answered “No” (1586200) were assigned level 1 (lifetime abstainer). Participants who answered “Yes” (1586199) received the branching question “How often did you have a drink containing alcohol in the past year?” and were assigned level 2 (“Never” 1586202), level 3 (“Monthly Or Less” 1586203), level 4 (“2-4 Per Month” 1586204), level 5 (“2-3 Per Week” 1586205), or level 6 (“>=4 Per Week” 1586206). Responses of skip or prefer not to answer at either question were set to NA.
8) Smoking status was captured as a 4-level factor derived from the lifetime cigarette self-report question "Have you smoked at least 100 cigarettes in your entire life?" (OMOP concept ID 1585857). Participants who answered "No" (1585859) were assigned level 1 (lifetime abstainer). Participants who answered "Yes" (1585858) received the branching question "Do you now smoke cigarettes every day, some days, or not at all?" and were assigned level 2 ("Not At All", 1585863), level 3 ("Some Days", 1585862), or level 4 ("Every Day", 1585861). Responses of skip, "prefer not to answer", or "don't know" at either question were set to NA.

**Recorded Medical Phenotypes.** These were derived from EHR-linked diagnosis records in the OMOP condition table. For each phenotype, we identified the relevant standard SNOMED concept and expanded it to include all descendant concepts, so that more specific child diagnoses (E.g., recurrent or single-episode major depressive disorder) were captured under the parent phenotype. Participants with at least one condition record matching the concept or any of its descendants were defined as cases, and those without as having no recorded diagnosis. These phenotypes included:

- Major depressive disorder (OMOP concept ID 4152280; SNOMED 370143000)
- Anxiety (OMOP concept ID 441542; SNOMED 48694002)
- Migraine (OMOP concept ID 318736; SNOMED 37796009)
- Insomnia (OMOP concept ID 436962; SNOMED 193462001)

**CIDI-SF major depression phenotype (All of Us)**

As a sensitivity analysis, we derived a structured major depressive disorder (MDD) phenotype in All of Us using the Composite International Diagnostic Interview Short-Form (CIDI-SF) depression items, available through the version 8 off-cycle Mental Health and Well-Being release. The CIDI-sf was administered program-wide to 97,010 AoU participants; within our analytical cohort, 15,379 (15.6%) had completed it (had scoreable symptom score)

We operationalised DSM-based criteria from the available CIDI-SF items, mapping responses to eight of the nine DSM-5 A-criterion symptoms (A1 depressed mood, A2 anhedonia, A3 appetite/weight change, A4 sleep disturbance, A6 fatigue, A7 worthlessness/guilt, A8 concentration difficulty, A9 thoughts of death; the psychomotor item A5 was not assessed in the instrument) together with the role-impairment item (criterion B).

Each symptom item was scored as present (1) or absent (0), with affirmative endorsement coded as present and negative responses coded as absent. For non-cardinal symptom items, skipped or missing responses were coded as absent. The appetite/weight item (A3) was scored as present where participants reported weight gain, weight loss, or both. We defined a total symptom score as the sum of the eight scored symptom items, ranging from 0 to 8. Participants were considered scoreable only if at least one of the two cardinal items (A1 or A2) was answered "yes" or "no". Those who skipped or were missing on both cardinal items were treated as missing for MDD (N = 1,265) and assigned no symptom score. The A-criterion was met when a participant endorsed at least one cardinal symptom (A1 or A2) and a total of at least five symptoms. The B-criterion was met when participants reported clinically significant role impairment, defined as a response of "a lot" or "somewhat" on the CIDI-SF interference item. Because the impairment item was administered only to participants endorsing depressive symptoms, missingness on criterion B among symptom-negative participants was structural and was treated as non-impaired.

MDD case status required both criteria. Participants meeting both the A- and B-criteria were classified as cases. Controls were participants who did not meet the A-criterion and showed no evidence of clinically significant impairment — either through an explicit non-impaired response or, among symptom-negative participants who were never administered the impairment item, through its structural absence. Participants who met only one criterion — endorsing the symptom threshold without reported impairment, or reporting impairment without meeting the symptom threshold — and those with insufficient impairment information to resolve case status were excluded from the binary case/control phenotype. In addition to the binary phenotype, we retained the 0–8 symptom score as a continuous measure of depressive symptom burden for scoreable participants.

**Pharmlines**

**Medication record-derived phenotypes**

1) Age at first antidepressant record was calculated as the interval between date of birth and the date of the first antidepressant record.

**Genetic-derived phenotypes**

2) Genetic sex was derived from genotype data.

**Survey-response phenotypes**

3) Income represents disposable household income (€ per month), derived from household income measured at baseline by self-report to the question "What is the net income per month?". Disposable household income was calculated as average household income divided by an equivalence factor based on the number of family members, following Statistics Netherland^12^.
4) Education was measured at baseline by self-report to the question "What is the highest level of education you have attained?", with the response mapped to estimated years of education (range 1-20)^12^ by Lifelines.
5) Chronotype was measured as mid-sleep time on free days corrected for oversleep on free days, calculated from the Munich ChronoType Questionnaire (MCTQ); higher values indicate a more evening-type, lower values a more morning-type^13^. Chronotype was attained during the Wave 1B Lifelines assessment.
6) Weekly sleep represents average sleep duration across the week for adult participants, calculated from the Munich ChronoType Questionnaire (MCTQ)^13^. Weekly sleep was attained during the Wave 1B Lifelines assessment.
7) BMI was derived from self-reported body weight and height (kg/m²), attained from the Wave 1 derivatives Lifelines assessment.
8) Alcohol intake was derived from the question "How many glasses of alcoholic drinks did you drink per day on average?". Alcohol intake was attained from the Baseline 1A – Visit 2 Lifelines assessment.
9) Smoking was measured as cumulative pack-years, attained from the Baseline 1A – Visit 2 Lifelines assessment.
10) Neuroticism was measured at baseline by self-report using the neuroticism domain of an abbreviated NEO Personality Inventory (NEO-PI-R; mental health section). The phenotype was the neuroticism sum score, with missing items imputed. Neuroticism was attained from the Baseline 1A – Visit 2 Lifelines assessment.
11) Fibromyalgia status was provided pre-defined using the 2010 American College of Rheumatology (ACR) criteria. Participants met criteria if they reported pain symptoms for ≥ 3 months and fulfilled either (WPI ≥ 7 AND SS ≥ 5) or (3 ≤ WPI ≤ 6 AND SS ≥ 9). The Widespread Pain Index (WPI) was calculated by counting the number of body areas in which the participant reported pain during the previous week. The Symptom Severity (SS) scale was derived from the Checklist Individual Strength (CIS) items and the somatization subscale of the Symptom Checklist-90 (SCL-90 SOM). The relevant symptoms were attained from the Wave 2 assessment questionnaires 2A1 and 2A2 for adults (aged 18 and older).
12) Stress was assessed over the lifetime and the past year using the List of Threatening Events (LTE) as a measure of acute stress and the Long-term Difficulties Inventory (LDI) as a measure of chronic stress^14,15^. Stress measures were attained from the Baseline 1A – Visit 2 Lifelines assessment.
    a) The LDI consists of 12 items referring to aspects of life — including housing, work, social relationships, free time, finances, health, school/study, and religion — each assessed on a 3-point scale (0 = not stressful, 1 = slightly stressful, 2 = very stressful); the total score was the sum across items.
13) The LTE comprises 12 major categories of stressful life events, assessed via self-report (mental health section), with each item scored 0 = no or 1 = yes; the total score was the sum across items.
14) Childhood sexual abuse (CSA) was assessed from five items, each scored 1 = never, 2 = rarely true, 3 = sometimes true, 4 = often true, 5 = very often true. CSA was attained from the Wave 2B Lifelines assessment. Measured as a cumulative CSA score summing responses across items a–e:
    a) During my childhood someone tried to touch me sexually or get me to touch them.
    b) During my childhood someone threatened to hurt me or tell lies about me if I refused to engage in a sexual act with them.
    c) During my childhood someone wanted me to do sexual things or let me watch sexual things.
    d) During my childhood I was molested by someone.
    e) During my childhood I believe I was abused sexually.
15) Recorded depression was classified in Lifelines following DSM-IV criteria, requiring ≥5 of nine symptom domains present within a two-week period, at least one of which was a core symptom (depressed mood or loss of interest). Symptoms were ascertained from two instruments: the MINI International Neuropsychiatric Interview, administered at three assessments and capturing current MDD (all symptom items answered by all participants but refers only to the last two weeks); and the Lifelines Depression Assessment (LIDAS)^16-18^, administered via two surveys (DEAQ and DEPQ) and supplemented by a lifetime MINI module at the third assessment, capturing lifetime MDD. A participant was classified as a current MDD case if criteria were met at any MINI assessment, and as a lifetime MDD case if criteria were met in either LIDAS survey or the lifetime MINI module. The primary recorded depression phenotype defined a case as any participant meeting criteria on either the current (MINI) or the lifetime (LIDAS/lifetime-MINI) ascertainment. Participants assessed by at least one instrument and not meeting criteria on any were classified as controls; participants with no usable data on either source were set to missing. This represents the most inclusive ("unscreened") case definition, maximizing case ascertainment by pooling cross-sectional and lifetime instruments, at the cost of a control group that was not uniformly screened for past episodes.
16) Recorded anxiety was defined as meeting criteria for lifetime generalized anxiety disorder (GAD) measured via the LIDAS DEAQ questionnaire.
17) Recorded depression symptom count was defined as the median total symptom count across all MINI and LIDAS depression assessments. Those that did not complete one assessment had their symptom count recorded as missing.

#### 1.8 AoU genetic data

Genome-wide genotypes for AoU were extracted from whole-genome sequence (WGS) data; genetically inferred ancestry classifications and global ancestry principal components were provided by AoU. Because SBayesRC weights were derived using a UK Biobank linkage disequilibrium reference, we restricted the AoU genotype data to variants represented in that reference. Using allele frequencies from unrelated European-ancestry individuals in the UK Biobank^19^ (unrelated defined as relatedness ≤ 0.05 using GCTA; European ancestry as previously defined^20^), we retained SNPs with minor allele frequency ≥ 0.01 in the UKB reference. As the UK Biobank array data are mapped to hg37 but AoU WGS data is in hg38, these SNPs were lifted to hg38 using UCSC liftOver, and the successfully converted subset was extracted from the AoU WGS Allele Count/Allele Frequency (ACAF) threshold call set using PLINK 1.9. We retained only biallelic SNPs and further restricted to variants whose tested and alternative alleles matched those in the UK Biobank array data, yielding 8,018,561 autosomal SNPs.

#### 1.9 Lifelines genetic data

Genotyping was performed using three different SNP arrays^21,22^. The first batch of ~15.500 Lifelines supposedly unrelated samples was genotyped using the Illumina human CytoSNP12 Beadchip^21^. For the second batch of ~38,000 samples the Infinium Global Screening Array® (GSA) MultiEthnic Disease Version 1.0 was used^22^. The third batch consisted of ~60,000 samples that were genotyped with the FinnGen Thermo Fisher Axiom® custom array^22^. Quality control (QC) for these batches was done separately. Genetic variants were removed from the dataset when they had a call rate < 0.95 / 0.99 / 0.98, a minor allele frequency ≤ 0.01 / 0.00 /0.0003 for the CytoSNP, GSA, or FinnGen chip, respectively, or a significant deviation from Hardy-Weinberg equilibrium (HWE; p < 10^-6^). Samples were excluded if the call rate < 0.95 / 0.99 / 0.95 for the CytoSNP, GSA, or FinnGen chip, respectively, when heterozygosity was > 4 SD from the mean or < 4SD from the mean adjusted for runs of homozygosity; if there was a gender mismatch without a possible fix based on the pedigree data; or in case of duplicate samples (non-monozygotic twins) in which case samples genotyped with the GSA or FinnGen chip were prioritized over a sample from the CytoSNP chip due to the lower SNP density of the latter chip, as well as samples with a higher call rate. Note that non-European samples were not removed at this stage.

During the QC of the FinnGen array a pedigree concordance analysis was performed to determine if samples supposed to be relatives based on the Lifelines pedigree file are indeed genetically related to the right degree, or if supposedly unrelated samples appeared to be genetically related. For this a LD-pruned SNP set consisting of overlapping autosomal genetic variants of the combined samples of the CytoSNP, GSA, and FinnGen chips was created and genetic relatedness was determined using KING version 2.2^23^. Any family relationship within families flagged as “error” that concerned a genetically calculated first-degree or “unrelated” relationship, as well as duplicates and first-degree relations between supposedly unlinked families was analysed by evaluating genetic relationships and family data from questionnaires (pseudonymised family names and birth years of the samples concerned; birth and death years of their parents; number of brothers and sisters and their birth years; report of a twin), if available. In case of an apparent sample swap, the pedigree file was updated. If family and genetic relations could not be matched, the sample was excluded from the dataset.

In addition, batch effects for the FinnGen array were checked as the first 28,000 and the second 32,000 samples were genotyped at different stages. Similar to the UKB approach^24^, a Fisher’s exact test was used on the 2x3 table of genotype counts (or 2x2 table for haploid markers) to test for array, batch, and plate effects. Markers that failed the array effects test were excluded from the full dataset, markers that failed the batch or plate effects tests were set to missing only for the samples in that failed batch or plate. After this the call rate filter as mentioned above was applied again.

Imputation of the samples in the three batches was done separately again. Previously the genotypes of the CytoSNP and GSA were genotyped using the Sanger Imputation service for genetic phasing and imputation, which makes use of the Haplotype Reference Consortium panel^25^. We also attempted to use this imputation service for the FinnGen sample but noticed a low number of SNPs with a high imputation score. This might be due to the PBWT imputation tool that the online HRC imputation service uses, which is relatively old^26^. Instead, we opted for an in-house phasing and imputation approach using a subset of ~11,000 HRC samples as a reference panel with the EAGLE2 phasing and IMPUTE5 imputation tools^25-27^.

After imputation of the three batches, we aimed to integrate the data into one dataset. For this, we selected from each of the batches the genetic variants with an imputation score > 0.99 and a minor allele frequency > 0.01 and next determined the overlapping SNP set between the three chips. This subset of variants was extracted from each of the three imputed datasets and the genotype probabilities were converted to most likely genotypes. Next these data were combined. KING was again applied to determine duplicated samples (mostly between the CytoSNP and GSA chips) and the one from the CytoSNP or with the lowest call rate was removed^24^. This cleaned dataset was then re-imputed using the same phasing and imputation pipeline as described above.

Finally, a principal component analysis using 2,504 individuals of diverse ancestries from Phase 3 of the 1000 Genomes Project as a reference population was performed to identify non-European individuals^28^. The final dataset consisted of 39,131,940 variants for 109,922 individuals, among whom 499 were non-Europeans.

#### 1.10  Polygenic Risk Scores

Polygenic scores (PGS) were derived from published genome-wide association studies conducted in European-ancestry samples (except schizophrenia, calculated from a multi-ancestry GWAS). We calculated PGS for seven psychiatric and related traits (**Supplementary Table S20**), selected to capture depression liability, broader psychiatric liability and common genetically correlated comorbidities potentially relevant to antidepressant treatment. We included major depression (Adams et al.^29^), schizophrenia (Trubetskoy et al.^30^), bipolar disorder (O’Connell et al.^31^), ADHD (Demontis et al.^32^), migraines (Hautakangas et al.^33^), insomnia (Jiang et al.^34^), and BMI (Yengo et al.^35^). Since the BMI GWAS included the Lifelines UGLI1 Cohort as a discovery cohort in the meta-analyses, we used MetaSubtract^36^ to subtract the Lifelines UGLI1 contribution from the published meta-analysis summary statistics, yielding overlap-corrected GWAS summary statistics. Scores derived from antidepressant response phenotypes (e.g., remission or non-response^6,37^) were not included because our aim was to test whether liability to psychiatric and related conditions distinguishes treatment outcomes, rather than to maximize prediction of response. Response-derived scores are designed to predict treatment response itself rather than to quantify underlying psychiatric liability, sand predictive performance in independent samples remains modest (OR 1.038, 1.015–1.062, and non-significant in three of five target cohorts^6^).

Identical SBayesRC weight files were applied in both cohorts, so the two PGS differed primarily in the proportion of weighted SNPs present in each cohort's genotype data. SNP coverage was higher in Lifelines than in AoU across all seven PGS. For example, of the 7,356,518 SNPs with depression SBayesRC weights, 7,356,466 (99.9993%) were available in the Lifelines imputed genotype data, whereas only 7,038,283 (95.7%) were available in the AoU genetic data. Prior to statistical analysis, all PGS were residualized on ancestry principal components to remove residual population structure. In AoU, all 16 global ancestry PCs provided by the Research Program were used; in Pharmlines, the first 20 within-European PCs were used to account for fine-scale population structure. PCs were not additionally included as covariates in outcome models. These residualized PGS were standardised before analysis, and ORs are reported per SD increase in PGS.

#### 1.11  Statistical analyses

**Primary analyses**

Primary genetic analyses were conducted among genetically inferred European-ancestry participants. Associations between standardised PGS or phenotypic predictors and antidepressant outcomes were estimated using logistic regression. Switching and discontinuation were each compared separately with continuation as the shared reference category. Pooled analyses combined outcomes across antidepressants, whereas PGS analyses were additionally conducted by index antidepressant and drug class. Models were adjusted for age at first antidepressant record, sex, income and educational attainment. Phenotypic and PGS predictors were entered individually rather than mutually adjusted for one another. Where age or sex was the exposure of interest, models included age and sex mutually adjusted as appropriate.

**Diagnostic stratification**

Antidepressant prescribing is vulnerable to confounding by indication. Primary analyses were therefore stratified according to recorded depression status.

Additional analyses included:

- the full antidepressant-prescribed cohort irrespective of diagnosis;
- AoU participants with recorded anxiety irrespective of depression status;
- AoU participants meeting CIDI-sf lifetime MDD criteria; and
- AoU CIDI-sf participants not meeting lifetime MDD criteria.

CIDI-stratified models were limited to European-ancestry participants because of smaller sample sizes.

**Augmentation and treatment-intensity analyses**

Augmentation was modelled as a separate binary outcome with continuation as the reference. To also assess whether PGS associations increased with treatment intensity, an ordinal model ordered participants as: continuation < switching < augmentation. Participants meeting both switching and augmentation criteria were assigned to the augmentation category for this analysis. The resulting per-step odds ratio summarises association across the ordering and should not be interpreted as requiring an identical effect between continuation-to-switching and switching-to-augmentation transitions. Binary switching-versus-continuation and augmentation-versus-continuation models were therefore retained to characterise individual thresholds.

**Drug- and class-specific analyses**

PGS associations with switching were estimated separately according to index antidepressant class and individual index antidepressant. Treatment specificity was evaluated using formal interaction terms between PGS and index drug or drug class. For these interaction models, observations were stacked across treatments and cluster-robust standard errors were calculated at the participant level to account for individuals contributing observations for more than one antidepressant or drug class. These tests assessed whether PGS effect estimates differed between treatments and avoided interpreting differences in statistical significance across separate treatment strata as evidence of interaction.

**Sensitivity analyses**

Several sensitivity analyses evaluated the robustness and interpretation of the primary prescription-derived outcomes. First, switching analyses were repeated after restricting switches to those occurring within six months of initiation of the index antidepressant. The primary requirement for ≥28 days of index monotherapy, <28-day cross-tapering and a maximum 90-day gap following index discontinuation was retained. Second, discontinuation analyses were repeated after excluding individuals with any subsequent augmentation or sustained antidepressant combination therapy during available follow-up. Third, PGS analyses were repeated in AoU participants of African and Admixed American genetically inferred ancestry. Fourth, AoU analyses using EHR-recorded depression were compared with analyses restricted to participants meeting CIDI-sf lifetime MDD criteria.

**Multiple testing**

Bonferroni correction was applied separately within prespecified analysis families. For PGS analyses, correction was applied across the seven PGS within each outcome, ancestry and diagnostic stratum. For phenotypic analyses, correction was based on the number of predictors assessed within each cohort: 14 in AoU and 17 in Pharmlines. Interaction analyses were corrected within their corresponding drug- or class-interaction model families. All-cause mortality across continuation, switching and discontinuation in Pharmlines was compared descriptively using a chi-squared test. This analysis was not included within the PGS or phenotypic-predictor multiple-testing families.

**Familial aggregation analyses**

The multigenerational structure of Lifelines allowed familial aggregation of treatment outcomes to be assessed independently of measured PGS. Analyses included 1,971 Pharmlines participants from 881 families, contributing 1,152 unique relative pairs. Parent–child, full-sibling and grandparent–grandchild relationships were examined. Familial aggregation was assessed for continuation overall, class-specific continuation, switching and discontinuation using two complementary measures: recurrence-risk ratios (λR); and tetrachoric correlations of underlying liability (r). Confidence intervals were estimated by family-level bootstrap with 1,000 replicates, resampling complete families with replacement to preserve within-family dependence. Evidence for association was defined by a 95% bootstrap confidence interval excluding zero for tetrachoric correlations or one for recurrence-risk ratios. Estimates based on fewer than 10 relative pairs were not reported.

### 2 Supplementary Results

#### 2.1 Cohort Characteristics

We identified 98,357 AoU participants and 12,884 Pharmlines participants who were ≥12 years old at their first recorded antidepressant and contributed at least one qualifying monotherapy episode of ≥28 days. Participants with bipolar disorder or psychotic disorders were excluded using cohort-specific ascertainment, based on EHR diagnoses in AoU and self-reported bipolar disorder or schizophrenia in Lifelines. AoU participants were somewhat older at first recorded antidepressant (mean 49 years [SD 16] vs 44 [14] in Pharmlines), and substantially more likely to have a recorded depression diagnosis (49% [N= 48,358] vs 35% [N=4,465]), reflecting differences in recruitment and data completeness. These proportions are lower than the ~60% with recorded depression reported in UK Biobank^8^ and the ~80% in the Estonian Biobank^38^, but are consistent with prescribing studies showing that a substantial proportion of antidepressant recipients have no recorded depression diagnosis^39^. In AoU, recorded depression was assigned from electronic health records, whereas in Pharmlines it was questionnaire-derived: the MINI, administered across three waves and completed at least once by 97% of participants, captures current depression, with lifetime depression assessed only in a subset of individuals (49%). In AoU, the CIDI-sf (Composite International Diagnostic Interview – Short Form) was also completed by 15.6% of study participants (N=15,379), providing an instrument-based MDD diagnosis to complement the EHR record; of these, 9,704 (63%) met CIDI-MDD criteria and 5,675 (37%) screened CIDI-noMDD. AoU also included greater ancestral diversity (56% [N=54,754] European, 13% [N=12,991] African, 8.8% [N=8,682] Latino/Admixed American, 1.6% [N=1,606] of other genetic ancestry, and 20.7% [N=20,324] without genetic information to infer ancestry), whereas Pharmlines was predominantly of European ancestry.

#### 2.2 Treatment patterns

Each participant was allocated a continuation, discontinuation or switching phenotype for each drug for which they had a ≥ 28-day period of monotherapy, provided they did not meet further outcome-specific exclusion criteria (See **Supplementary Figure S1** for a schema of outcomes). The prevalence of these outcomes was broadly similar across commonly prescribed antidepressants (**Figure 1A–B**). Pooled across all antidepressants, 28% (N=27,389) of the AoU cohort recorded at least one switching event compared to 17% (N=2,133) of the Pharmlines cohort. Augmentation, defined as ≥28 days of antidepressant overlap with an atypical antipsychotic or lithium, occurred in 8.3% (N=8,126) of AoU and 9.5% (N=1,223) of Pharmlines participants. Augmentation was assessed separately and could co-occur with switching and monotherapy discontinuation.

Switching patterns were broadly similar across cohorts (**Figure 1C–D**), with SSRIs accounting for most initial medications and many switches occurring within class. When serotonin–norepinephrine reuptake inhibitors (SNRIs) were the index drug, switching across classes was more frequent than when selective serotonin reuptake inhibitors (SSRIs) were the index drug. The median interval between antidepressants was short in both cohorts, though notably shorter in AoU (9 days) than Pharmlines (18 days), consistent with rapid switching or cross-tapering. Drug-specific monotherapy outcomes by depression status are summarized in **Supplementary Figure S2**. Overall, switching rates were higher among participants with recorded depression than among those without: pooled across antidepressants, 22% of those with recorded depression versus 13% of those without switched antidepressants in Pharmlines, and 39% versus 17% in AoU.

#### 2.3 Phenotypic associations with switching and discontinuation

Of the 98,357 AoU and 12,884 Pharmlines participants, 87,787 and 11,884, respectively, qualified for a primary monotherapy outcome (continuation, switching, or discontinuation) for at least one drug; outcome-specific exclusions are described in the **Supplementary Methods.** Cohort characteristics by outcome are in **Supplementary Table S1** and full association results in **Supplementary Tables S2–S3**, with significance declared after Bonferroni correction (14 traits in AoU, 17 in Pharmlines).

Among European-ancestry participants with a recorded depression diagnosis (**Figure 2**), measures associated with switching were broadly consistent across cohorts and tracked greater psychiatric and other medical burden. Recorded diagnosis of anxiety was associated with switching in both AoU (OR 2.35, 95% CI 2.17–2.54, p<2.2x10^-16^) and Pharmlines (OR 1.53, 95% CI 1.17–1.99, p=1.64x10^-3^), as were stress-related exposures ("Sometimes" vs “Never” stress in AoU; long-term difficulties, threatening experiences, and childhood sexual abuse in Pharmlines), higher depression symptom count (AoU: OR 1.30, CI 1.21-1.4, p=9.91x10^-12^; Pharmlines: OR=1.51, CI 1.39-1.63 p <2.2e-16), smoking, and cohort-specific markers of comorbidity and psychopathology (e.g., higher BMI in AoU, higher neuroticism in Pharmlines). In the AoU CIDI-sf MDD subset, recorded anxiety, migraine, insomnia, higher BMI, and higher symptom count remained significantly associated with switching (**Supplementary Figure S3**).

Sociodemographic and clinical-measurement factors, by contrast, were associated with both switching and discontinuation in AoU but not Pharmlines after multiple-testing correction, and weighed more heavily on discontinuation than switching. In AoU, lower income was associated with higher risk of both discontinuation (<25k vs 150–200k: OR=2.38, 95% CI 1.89–3.03 p=6.8x10^-13^) and switching (OR=2.13 CI 1.82-2.44, p=2.5x10^-23^). Lower educational attainment was similarly a risk factor, again most for discontinuation (Less than high school vs College Postgraduate: OR 2.44, CI 1.79-3.23, p=1.2x10^-8^). Lower alcohol frequency was a risk factor for discontinuation only. Older age at first recorded medication increased discontinuation but reduced switching, likely reflecting left-truncation of EHR records given the strong age–birth-year correlation (**Supplementary Figure S18**); male sex increased the odds of switching (OR=1.11, CI 1.04-1.19, p=0.0027). In Pharmlines, income, sex, and age were directionally similar at nominal significance (same for education specifically with switching) but none survived multiple-testing correction. The only significant protective clinical factor against discontinuation in Pharmlines was longer weekly sleep duration (OR=0.70 per SD, 95% CI 0.60–0.81, p=4.8x10^-6^), mirroring the association between recorded insomnia and increased discontinuation risk in AoU. In Pharmlines, all-cause mortality differed across the three monotherapy outcomes (p < 0.001), driven largely by higher mortality among discontinuers (7.0% vs 4.3% in continuers and 4.0% in switchers; **Supplementary Table S1**).

Among participants with unrecorded depression (**Supplementary Figure S4**), associations diverged from the depression stratum and were less consistent across cohorts. In AoU, relative to the recorded depression stratum, associations for education, smoking, and alcohol were attenuated and did not survive multiple-testing correction, while comorbidities became more prominent. Insomnia, anxiety, and migraine were each associated with a lower likelihood of monotherapy discontinuation, consistent with antidepressants being prescribed for anxiety, pain, sleep disorders, or migraine prophylaxis rather than depression alone. In contrast, lifetime MDD defined by the CIDI-sf and higher CIDI-sf symptom counts were associated with an increased likelihood of discontinuation. In a separate AoU stratum defined by recorded anxiety irrespective of depression status, associations broadly resembled those in the recorded depression stratum, with recorded depression associated with increased switching and slightly reduced discontinuation (**Supplementary Figure S5**). In Pharmlines, within those with unrecorded depression, the comorbidity profile was not associated with either outcome (fibromyalgia [a pain condition] and sleep duration attenuated to the null; BMI showed no association), and the only Bonferroni-significant signals were protective effects of higher neuroticism and education on discontinuation. AoU results without CIDI-sf MDD are shown in **Supplementary Figure S6.**

#### 2.4 Polygenic associations with switching and discontinuation

We tested whether PGS for seven traits were associated with switching and discontinuation, each relative to continuation, in models that did not distinguish between antidepressants (i.e., pooling across all antidepressants). Separate binary logistic regression models were fitted for each outcome within each ancestry and diagnostic stratum. Significance was declared after Bonferroni correction for the seven PGS tested (**Supplementary Tables S4–S5**).

**Figure 3** presents European-ancestry results. Among individuals with recorded depression, switching showed the most consistent cross-cohort signal. Higher depression PGS was associated with increased odds of switching in AoU (OR=1.16 per SD unit, 95% CI 1.12–1.20, p <2.2x10^-16^) with a concordant and nominally significant estimate in the smaller Pharmlines cohort (OR=1.11, 95% CI 1.01–1.22, p=0.026). Higher insomnia, migraine and ADHD PGS were also associated with switching in AoU, and all corresponding Pharmlines estimates were directionally concordant. Restricting analyses to AoU participants meeting lifetime MDD criteria by CIDI-sf attenuated associations for insomnia and migraine PGS towards the null, whereas associations with depression PGS remained significant and ADHD PGS estimates were largely unchanged. Associations in AoU with depression, insomnia, migraine and ADHD PGS also remained significant when switching was restricted to events occurring within six months of index antidepressant initiation. This sensitivity analysis retained the same 90-day post-discontinuation window and <28-day cross-taper allowance used in the primary switching definition (**Supplementary Table S6-7; Supplementary Figure S7**).

Discontinuation showed weaker and less consistent associations across cohorts (**Figure 3**). Among participants with recorded depression, higher depression PGS was associated with discontinuation in AoU (OR=1.18, 95% CI 1.12–1.24, p= 9.2x10^-11^), whereas the Pharmlines estimate was non-significant and opposite in sign. Insomnia, ADHD (OR=1.08, CI 1.03-1.14, p=1.7x10^-3^) and bipolar disorder PGS were also associated with monotherapy discontinuation in AoU. Of these, ADHD PGS showed the most directionally concordant estimate in Pharmlines (OR=1.10, CI 0.98-1.24), although it did not reach significance. Among AoU participants meeting CIDI-sf criteria for lifetime MDD, depression and bipolar disorder PGS remained significantly associated with discontinuation, while ADHD PGS retained a similar point estimate. Results were largely unchanged after excluding individuals who met augmentation or combination criteria during follow-up, and in Pharmlines, the association between higher depression PGS and decreased odds of discontinuation became nominally significant (**Supplementary Table S8-9, Supplementary Figure S8**).

Because antidepressants are also commonly prescribed for anxiety^39^, we additionally examined AoU participants with recorded anxiety irrespective of depression status. Higher depression PGS was associated with both switching (OR=1.20, 95% CI 1.17–1.23) and discontinuation (OR=1.14, 95% CI 1.10–1.19) in this stratum, alongside insomnia, BMI, ADHD and migraine PGS for switching, and insomnia and bipolar disorder PGS for discontinuation (**Supplementary Figure S9**). This stratum likely overlaps substantially with the recorded-depression stratum and is therefore not an independent replication.

Among AoU participants of Latin/Admixed American ancestry, effect estimates were generally similar in magnitude to those in Europeans, but most associations did not reach significance. The exception was depression PGS: higher scores were associated with increased odds of both discontinuation and switching in the full antidepressant-prescribing and recorded-anxiety cohorts, and with increased discontinuation among those with recorded depression (**Supplementary Figure S10).** Among participants of African ancestry, estimates were largely attenuated toward the null or directionally inconsistent, and no PGS was significantly associated with either outcome, except insomnia and depression PGS with switching in the full antidepressant-prescribing cohort (**Supplementary Figure S10**).

#### 2.5 Polygenic liability across treatment intensity

Having examined outcomes following antidepressant monotherapy, we next investigated augmentation as a typically later and more intensive treatment step. Augmentation was defined as ≥28 days of overlap between an antidepressant and an atypical antipsychotic or lithium. For augmentation analyses, participants meeting augmentation criteria were classified as augmentation cases regardless of whether they also met switching or discontinuation criteria. Models were run within each ancestry and adjusted and corrected as in the switching analyses (**Supplementary Tables S10–S11).**

In AoU participants with recorded depression and European ancestry (**Figure 4A**), higher depression PGS was associated with increased odds of augmentation (OR 1.29, 95% CI 1.23–1.35, p<2.2×10−16), with a larger point estimate than observed for switching (**Figure 4B**). A similar ordering of point estimates was observed for insomnia and ADHD PGS. Schizophrenia and bipolar disorder PGS were not associated with switching but were associated with augmentation (schizophrenia OR 1.09, 95% CI 1.05–1.14, bipolar disorder OR 1.08, 95% CI 1.03–1.13), indicating a different polygenic profile for augmentation. Restricting AoU participants to those meeting CIDI-SF MDD criteria, associations of depression, bipolar disorder and schizophrenia PGS with augmentation remained significant after Bonferroni correction.

We then evaluated the ordering continuation < switching < augmentation using an ordinal proportional-odds model in AoU. Participants meeting both augmentation and switching criteria were assigned to augmentation for this analysis only. Among European-ancestry participants with recorded depression (**Supplementary Table S12, Supplementary Figure S11**), depression, insomnia, ADHD, migraine and schizophrenia PGS were each significantly associated with being in a higher treatment-intensity category. The strongest association was observed for depression PGS (OR 1.18 per SD, 95% CI 1.15–1.21, p<2.2×10−16). The ordinal OR summarises the association across the ordered outcome thresholds and should not be interpreted as an identical effect for the transition from continuation to switching and from switching to augmentation. In Pharmlines, augmentation was not significantly associated with any PGS.

Among participants without recorded depression, a consistent cross-cohort signal emerged: higher bipolar disorder and schizophrenia PGS were associated with increased odds of augmentation in both AoU and Pharmlines (**Figure 4A**). Finally, among AoU participants with recorded depression, the depression and bipolar disorder PGS augmentation associations replicated in Admixed American participants, with consistent effect directions in those of African ancestry (**Supplementary Table S10; Supplementary Figure S12).**
