## Supplementary material for "Polygenic and familial contributions to antidepressant continuation, switching, discontinuation and augmentation in the All of Us and Pharmlines cohorts": Suppementary_Figures

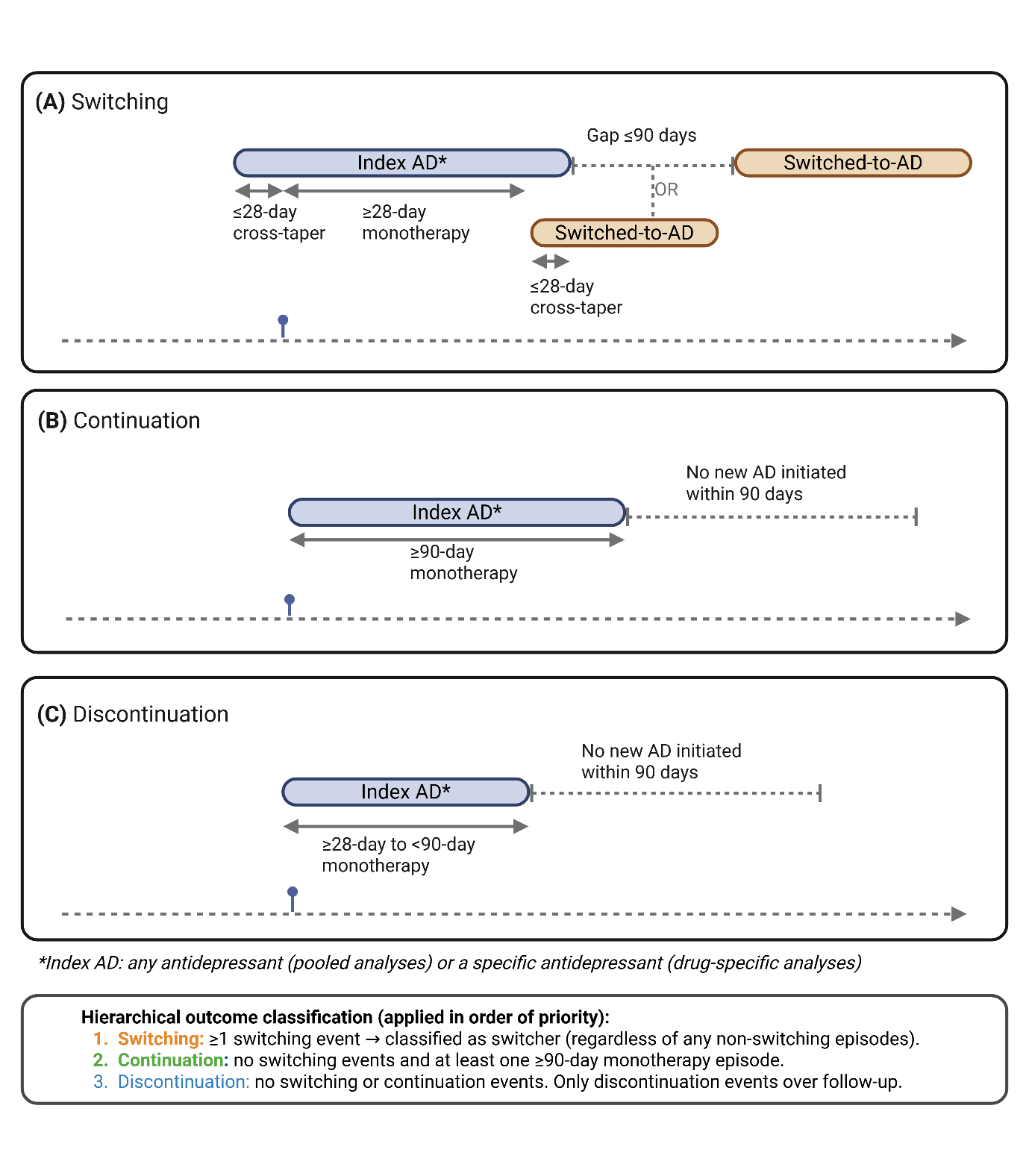
**Supplementary Figure S1. Schematic representation of switching, continuation and discontinuation following treatment with an index antidepressant.** To qualify for the study, participants were required to have at least one antidepressant monotherapy episode lasting ≥28 days. Participants could contribute outcomes for multiple antidepressants across their observed medication history. **A** Switching was defined as a qualifying monotherapy episode of ≥28 days on the index antidepressant followed by initiation of a different antidepressant either during a brief cross-taper of <28 days at the end of the index episode or within 90 days after index discontinuation. **B** Continuation was defined as at least one monotherapy episode of ≥90 days in the absence of a qualifying switch from that antidepressant. **C** Discontinuation was defined, in the absence of a qualifying switch, when all qualifying monotherapy episodes for that antidepressant lasted ≥28 but <90 days and no episode met the continuation criterion. Outcomes were assigned hierarchically at the person-by-drug level, with switching taking precedence over continuation and discontinuation. Participants meeting augmentation criteria could also meet switching or discontinuation criteria. Augmentation cases were excluded from the continuation reference group in augmentation-versus-continuation and discontinuation-versus-continuation analyses. Sustained combination therapy, defined as ≥28 days of overlap between two antidepressants, was excluded from switching and continuation groups but did not preclude classification as discontinuation when the discontinuation criteria were otherwise met. This figure was created with BioRender.com.

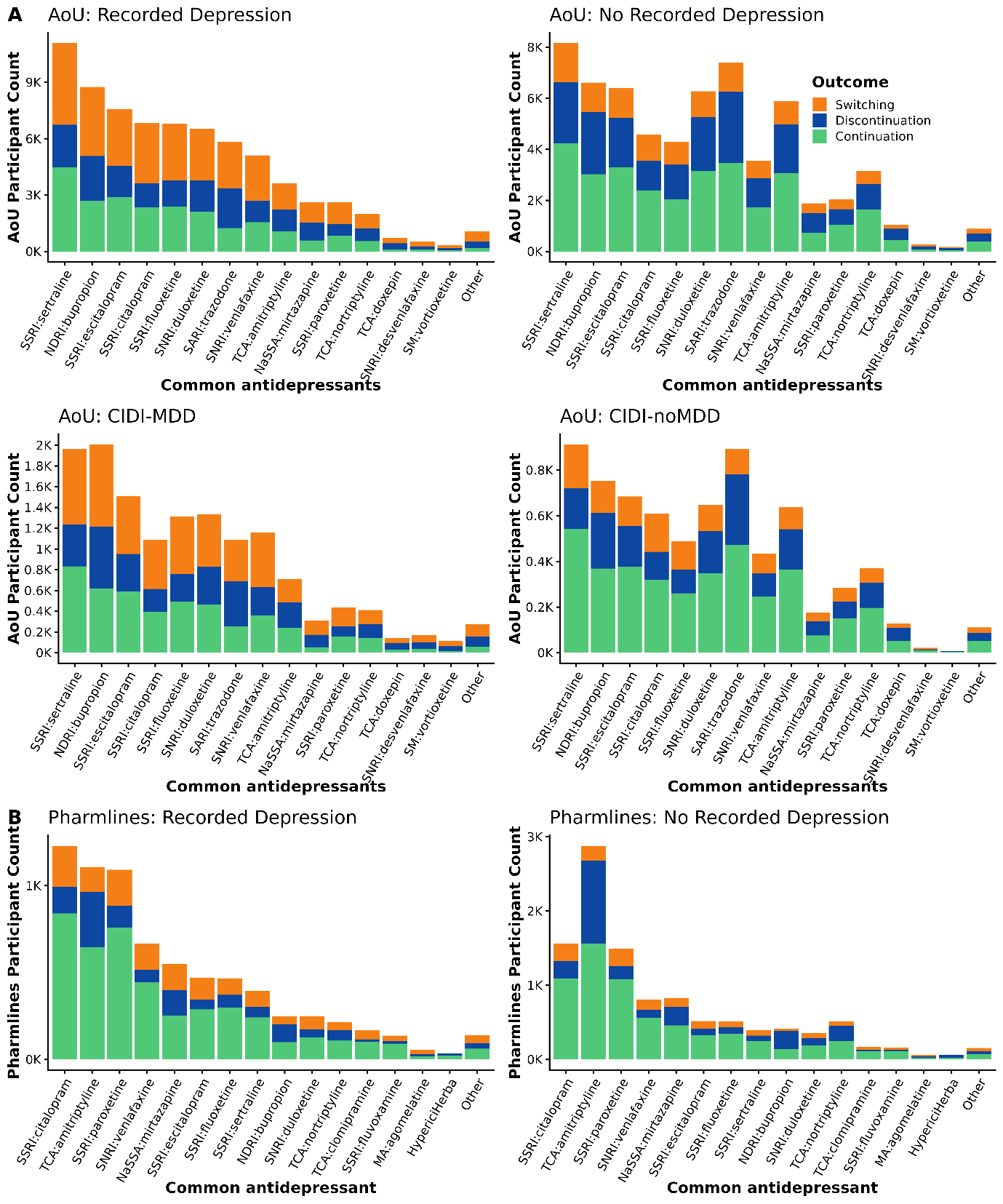

**Supplementary Figure 2. Distribution of antidepressant treatment outcomes across commonly prescribed antidepressants, stratified by cohort and recorded depression status. Stacked bars show participant counts for the most frequently prescribed antidepressants, partitioned by treatment outcome on the antidepressant: continuation (green), discontinuation (blue), and switching (orange). (A) All of Us (AoU) participants across four strata: those with versus without a recorded depression diagnosis ("Recorded Depression" / "No Recorded Depression"), and those classified as having major depressive disorder on the basis of CIDI criteria versus not ("CIDI-MDD" / "CIDI-noMDD"). (B) Pharmlines participants with versus without a recorded depression diagnosis ("Recorded Depression" / "No Recorded Depression"). Note that y-axis scales differ across panels. AoU, All of Us; LL, Pharmlines; CIDI, Composite International Diagnostic Interview; MDD, major depressive disorder.**

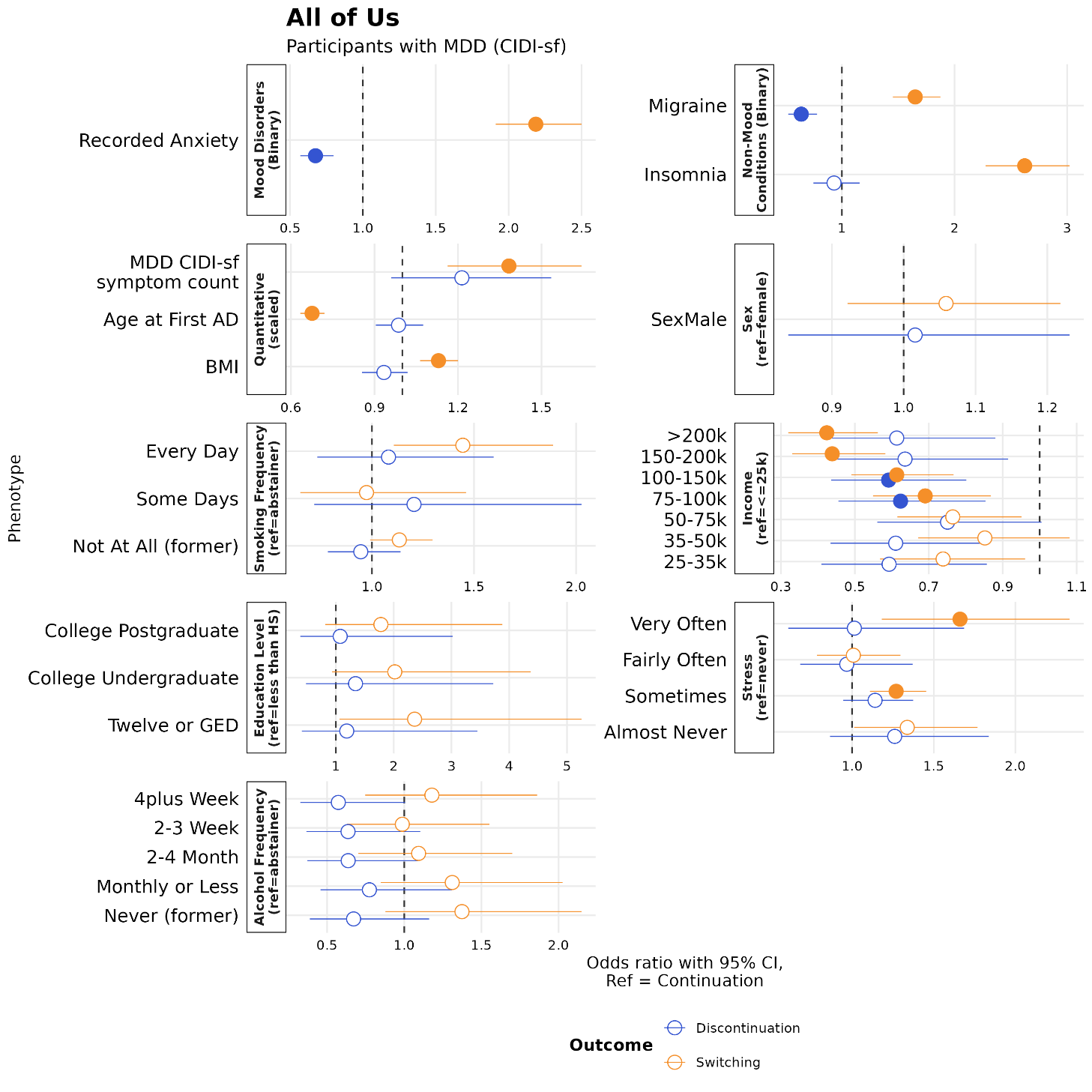

**Supplementary Figure S3. Phenotypic, demographic and lifestyle associations with antidepressant switching and discontinuation, relative to continuation, in AoU participants with lifetime MDD as measured by the CIDI-sf.** Forest plots show adjusted odds ratios (ORs) with 95% confidence intervals from two separate binary logistic regression models, contrasting switching versus continuation (orange) and discontinuation versus continuation (blue). Models included age at first antidepressant record, genetic sex, income and education level as covariates. Predictors are grouped by domain: binary traits (mood and non-mood conditions), quantitative traits (standardised to mean 0, SD 1 prior to modelling), sex (reference = female), and categorical measures, each modelled against the reference category indicated in the panel header. Filled points indicate associations significant at Bonferroni correction (for 15 traits in AoU); open points are non-significant. The dashed vertical line marks the null (OR = 1).

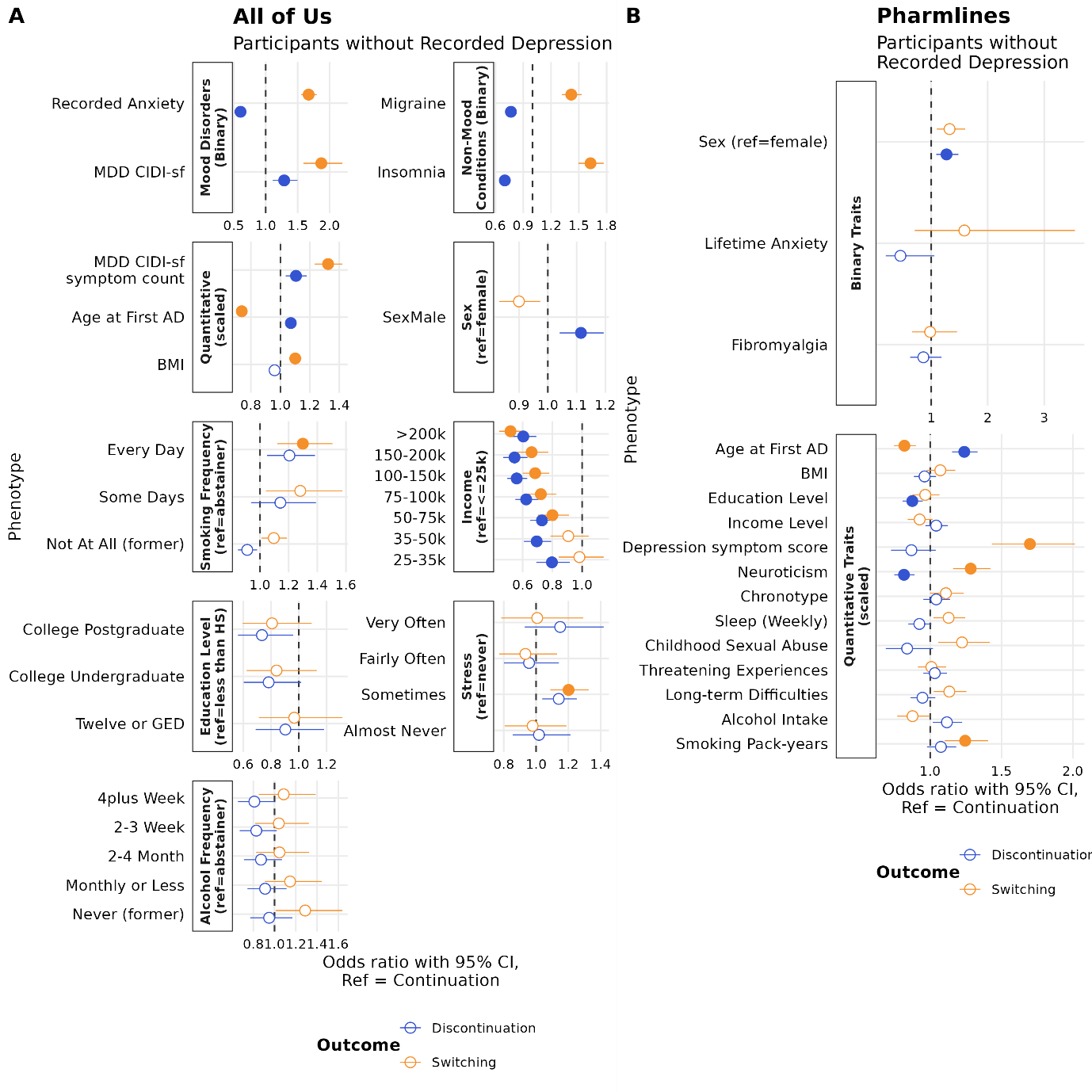

**Supplementary Figure S4. Phenotypic, demographic and lifestyle associations with antidepressant switching and discontinuation, relative to continuation, in participants without a recorded diagnosis of depression across All of Us and Pharmlines.** Forest plots show adjusted odds ratios (ORs) with 95% confidence intervals from two separate binary logistic regression models, contrasting switching versus continuation (orange) and discontinuation versus continuation (blue), in (**A**) AoU participants without a recorded diagnosis of depression and (**B**) Pharmlines participants without depression. Models included age at first antidepressant record, genetic sex, income and education level as covariates. Predictors are grouped by domain: binary traits (mood and non-mood conditions in AoU; lifetime anxiety, fibromyalgia in Pharmlines), quantitative traits (standardised to mean 0, SD 1 prior to modelling), sex (reference = female), and — in AoU only — categorical measures, each modelled against the reference category indicated in the panel header. Filled points indicate associations significant at Bonferroni correction (for 15 traits in AoU and 17 traits in Pharmlines); open points are non-significant. The dashed vertical line marks the null (OR = 1).

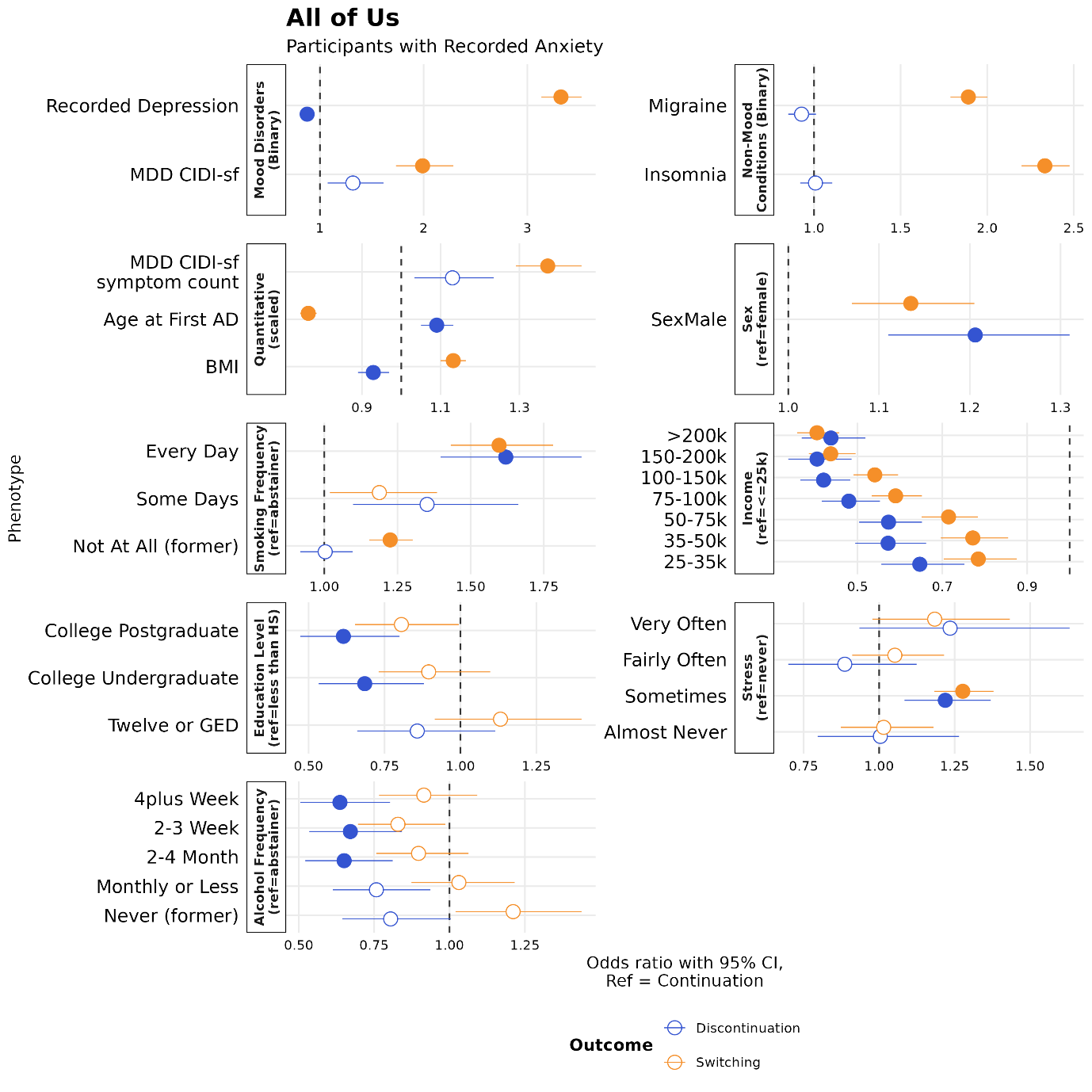

**Supplementary Figure S5. Phenotypic, demographic and lifestyle associations with antidepressant switching and discontinuation, relative to continuation, in AoU participants with recorded anxiety.** Forest plots show adjusted odds ratios (ORs) with 95% confidence intervals from two separate binary logistic regression models, contrasting switching versus continuation (orange) and discontinuation versus continuation (blue). Models included age at first antidepressant record, genetic sex, income and education level as covariates. Predictors are grouped by domain: binary traits (mood and non-mood conditions), quantitative traits (standardised to mean 0, SD 1 prior to modelling), sex (reference = female), and categorical measures, each modelled against the reference category indicated in the panel header. Filled points indicate associations significant at Bonferroni correction (for 15 traits in AoU); open points are non-significant. The dashed vertical line marks the null (OR = 1).

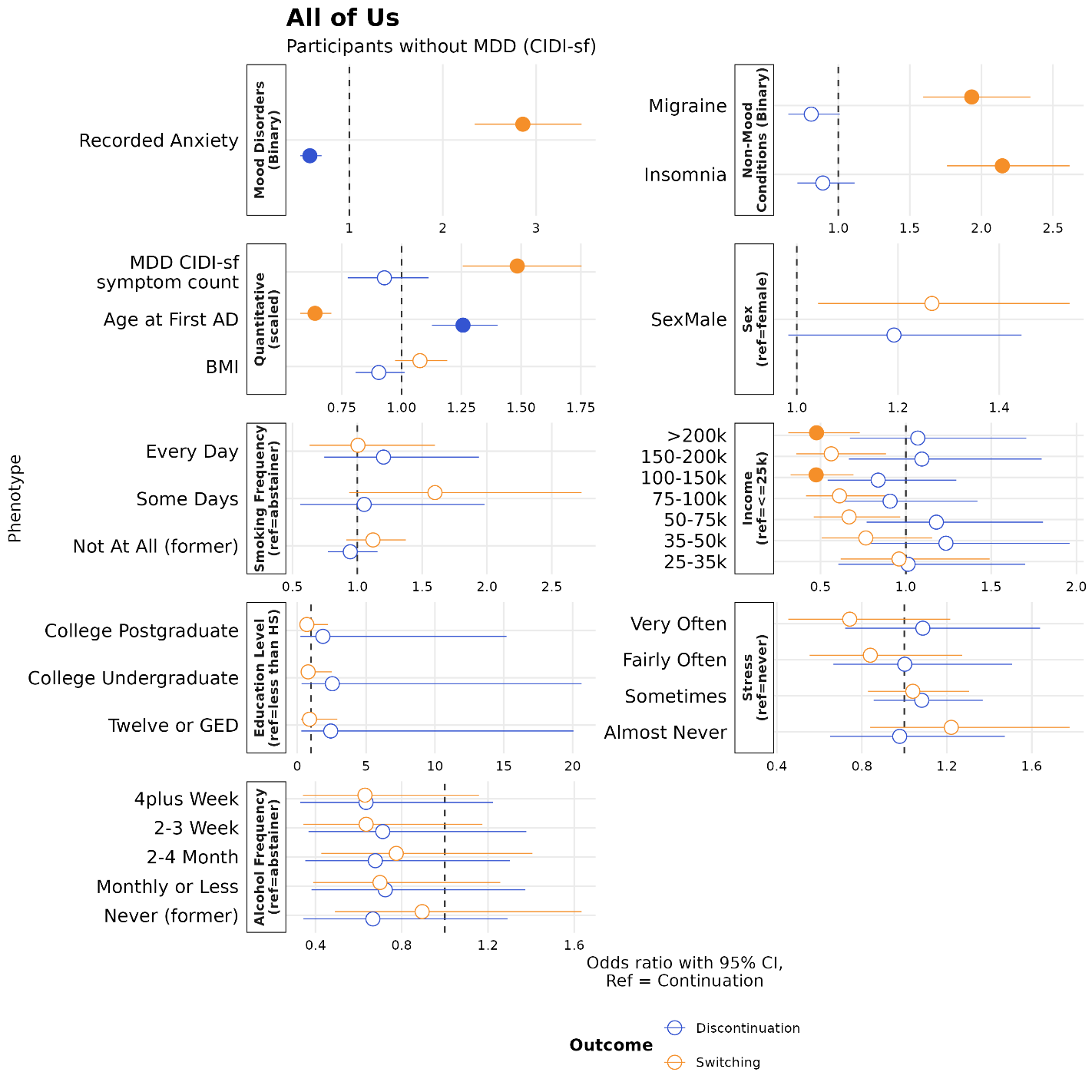

**Supplementary Figure S6. Phenotypic, demographic and lifestyle associations with antidepressant switching and discontinuation, relative to continuation, in AoU participants without lifetime MDD as measured by the CIDI-sf.** Forest plots show adjusted odds ratios (ORs) with 95% confidence intervals from two separate binary logistic regression models, contrasting switching versus continuation (orange) and discontinuation versus continuation (blue). Models included age at first antidepressant record, genetic sex, income and education level as covariates. Predictors are grouped by domain: binary traits (mood and non-mood conditions), quantitative traits (standardised to mean 0, SD 1 prior to modelling), sex (reference = female), and categorical measures, each modelled against the reference category indicated in the panel header. Filled points indicate associations significant at Bonferroni correction (for 15 traits in AoU); open points are non-significant. The dashed vertical line marks the null (OR = 1).

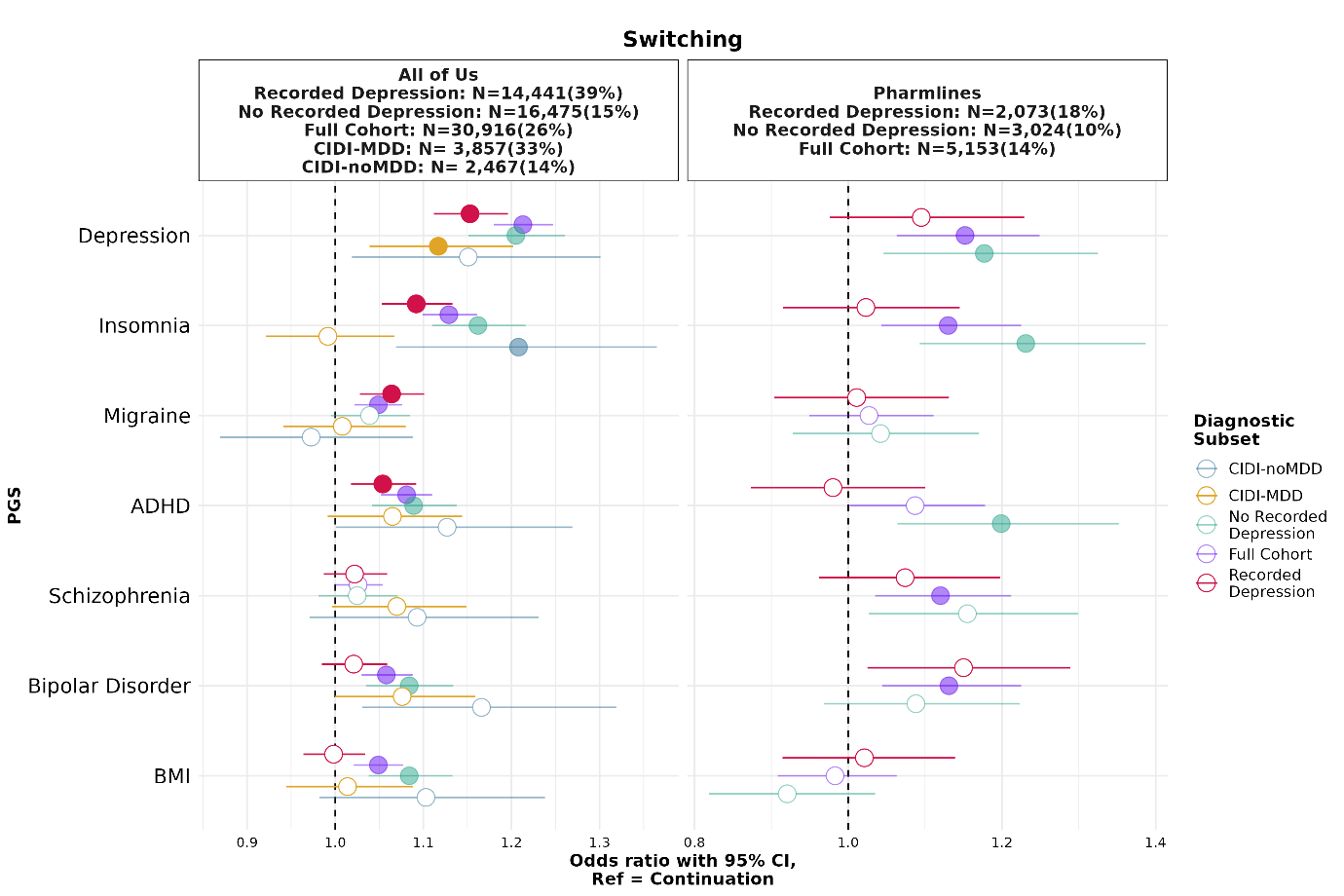

**Supplementary Figure S7. Polygenic score associations with antidepressant switching under a sensitivity definition of the switching window. Forest plot showing associations between standardised polygenic scores (PGS) and switching relative to continuation. The sensitivity definition additionally required the subsequent antidepressant to be initiated within six months of index-antidepressant initiation, while retaining the primary definition requiring ≥28 days of preceding index-antidepressant monotherapy, a post-discontinuation switching window of ≤90 days and allowance for brief cross-tapers of <28 days at the end of the index episode. Points indicate odds ratios and horizontal bars indicate 95% confidence intervals. The dashed vertical line marks the null at OR = 1. Models included age at first antidepressant record, sex, income and education as covariates. PGS were residualized for ancestry principal components and standardised before analysis. Results are shown for seven PGS across five diagnostic subsets, comprising participants with recorded depression, participants without recorded depression, the full prescribing cohort, participants meeting lifetime MDD criteria using the CIDI-SF and participants not meeting CIDI-SF lifetime MDD criteria. Filled points indicate associations significant after Bonferroni correction across seven PGS within each stratum and open points indicate non-significant associations. Sample sizes and the proportion of participants classified as switching under the sensitivity definition are shown in the panel headers. PGS abbreviations include ADHD, attention-deficit/hyperactivity disorder and BMI, body mass index.**

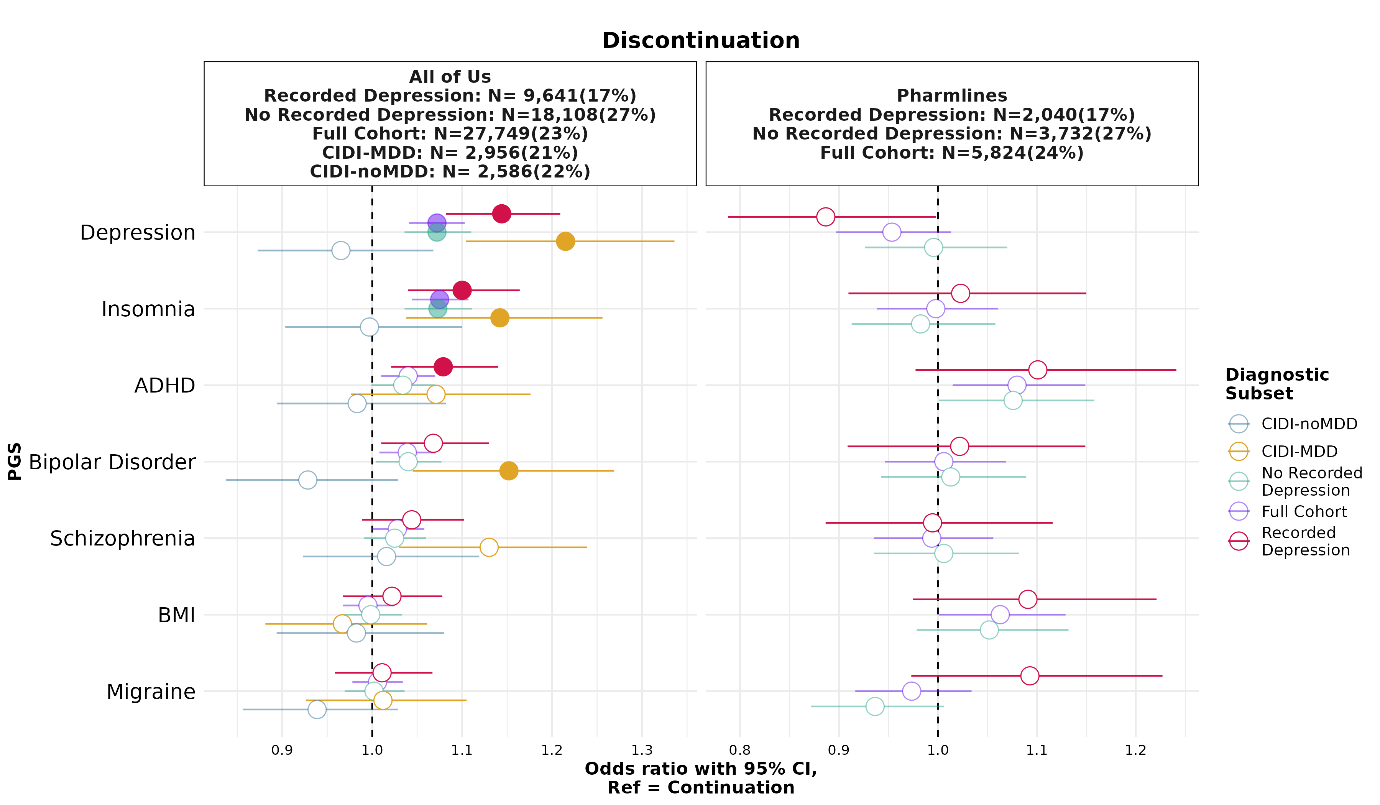

**Supplementary Figure S8. Polygenic score associations with antidepressant discontinuation under a sensitivity definition of the discontinuation phenotype.** Forest plot showing the association between standardised polygenic scores (PGS) and discontinuation, relative to continuation (reference). This analysis applies a sensitivity definition of the discontinuation phenotype in which participants with augmentation or combination treatment were removed from the discontinuation cases, restricting cases to participants who discontinued monotherapy without augmentation or combination therapy. Points indicate odds ratios and horizontal bars the 95% confidence intervals; the dashed vertical line marks the null (OR = 1). Models included age at first antidepressant record, genetic sex, income, education level and PCs (global ancestry in AoU and within-European ancestry in Pharmlines) as covariates. Results are shown for seven PGS (y-axis) across five diagnostic subsets (colour): participants with a recorded SNOMED depression diagnosis (Recorded Depression; red), participants without a recorded SNOMED depression diagnosis (No Recorded Depression; teal), all participants combined (Full Cohort; purple); participants with lifetime MDD as measured by the CIDI-sf (CIDI-MDD; orange); and participants without lifetime MDD as measured by the CIDI-sf (CIDI-noMDD; blue). Filled points denote associations significant after Bonferroni correction for 7 tested PGS traits; open points denote non-significant associations. Sample sizes and the proportion of participants classified as discontinuing under this definition are given in the header. PGS abbreviations: ADHD, attention-deficit/hyperactivity disorder; BMI, body mass index.

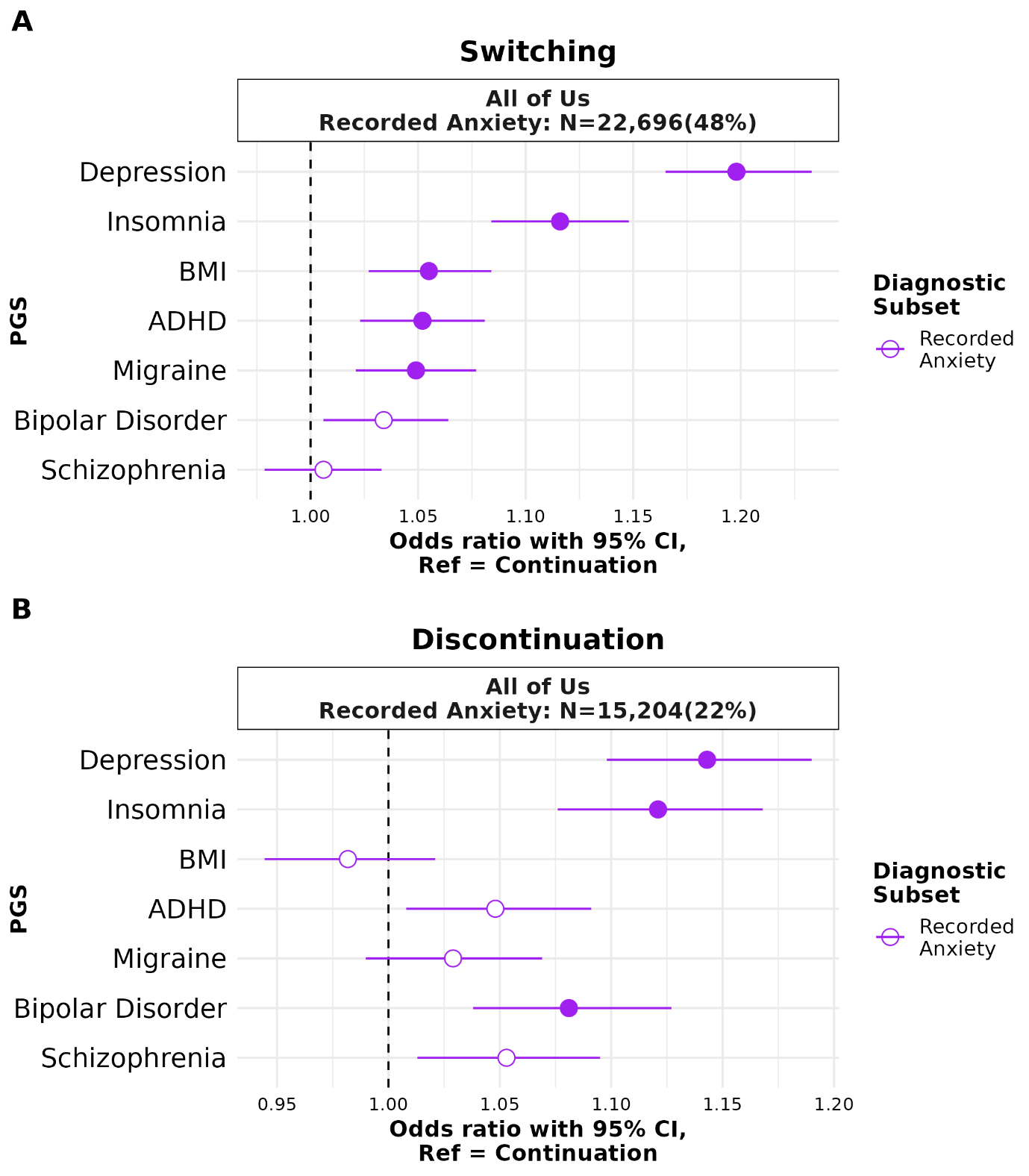

**Supplementary Figure S9. Polygenic score associations with pooled (across antidepressants) switching and monotherapy discontinuation, relative to continuation, amongst AoU participants with recorded anxiety.** Forest plots show adjusted odds ratios (ORs) with 95% confidence intervals from binary logistic regression models contrasting (A) switching versus continuation and (B) discontinuation versus continuation, in European-ancestry participants. Each polygenic score (PGS) was standardised to mean 0, SD 1 to European PGS prior to modelling, and was modelled separately. Models included age at first recorded antidepressant, genetic sex, income, education level, and the first 16 global ancestry genetic principal components as covariates. Results are shown for those with recorded anxiety, with sample size indicated in the panel headers. Filled points indicate associations significant after Bonferroni correction across 7 PGS within each stratum; open points are non-significant. The dashed vertical line marks the null (OR = 1).

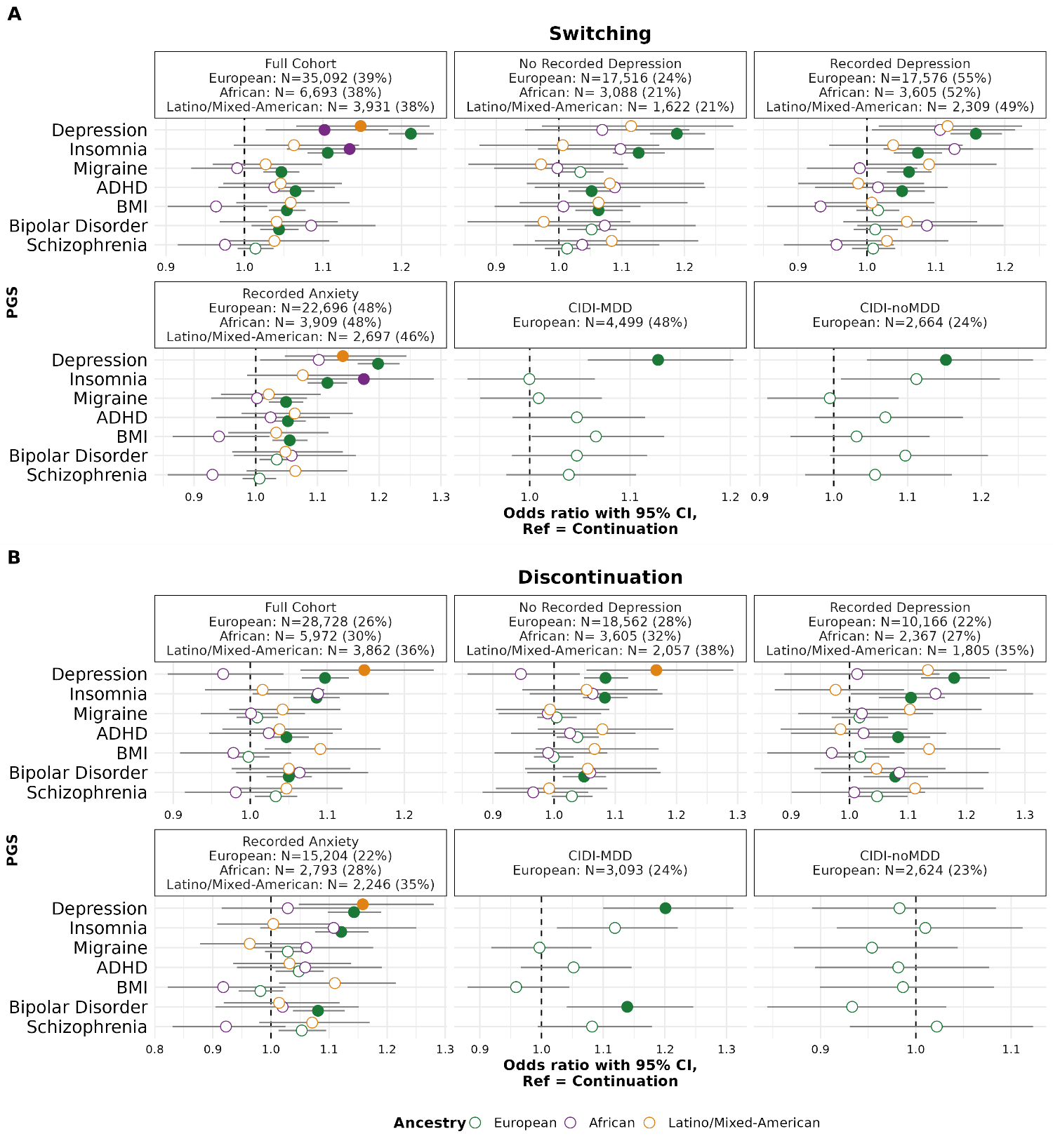

**Supplementary Figure S10. Ancestry-stratified polygenic score associations with antidepressant switching and discontinuation (All of Us).** Forest plots showing associations between standardised polygenic scores (PGS) and (**A**) switching and (**B**) discontinuation, each relative to continuation (reference), estimated separately within ancestry groups in the All of Us cohort. Within each of panels A and B, results are shown across six diagnostic subsets (sub-panels): Full Cohort, No Recorded Depression, Recorded Depression, Recorded Anxiety, CIDI-MDD, CIDI-noMDD. Points indicate odds ratios and horizontal bars the 95% confidence intervals; the dashed vertical line marks the null (OR = 1). Models included age at first antidepressant record, genetic sex, income, education level and ancestry PCs as covariates. Results are shown for seven PGS (y-axis) in three ancestry groups (colour): European (green), African (purple), and Latino/Mixed-American (orange). Filled points denote associations significant after Bonferroni correction for seven tested PGS traits; open points denote non-significant associations. Per-group sample sizes and the proportion of participants classified as switching (A) or discontinuing (B) under each diagnostic subset are given in the sub-panel headers. Note that the x-axis range differs between sub-panels. PGS abbreviations: ADHD, attention-deficit/hyperactivity disorder; BMI, body mass index.

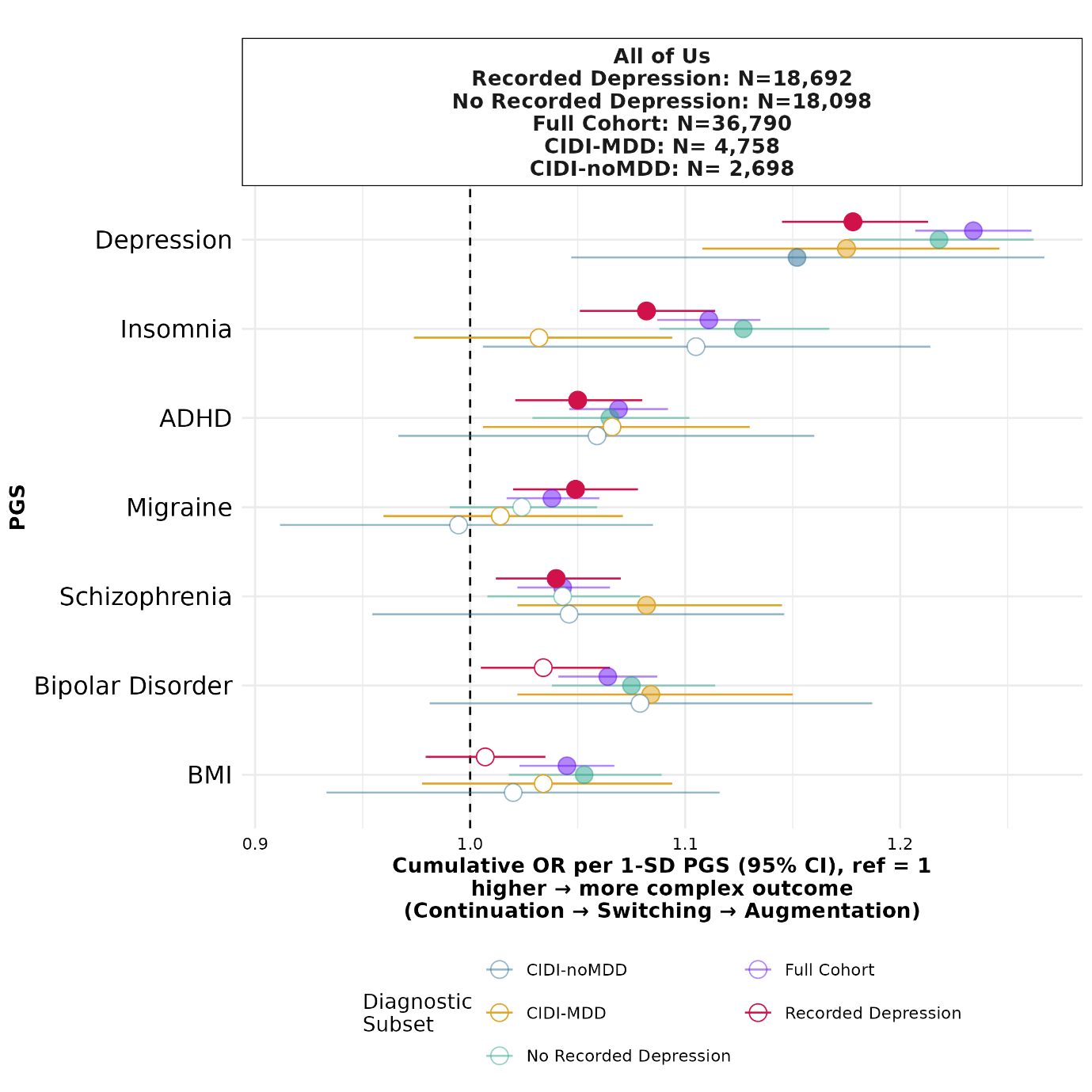

**Supplementary Figure S11. Polygenic score associations with escalating antidepressant treatment outcomes under an ordinal model (All of Us).** Forest plot showing associations between standardised polygenic scores (PGS) and an ordered three-level treatment outcome — continuation < switching < augmentation — estimated using an ordinal (proportional-odds) model in the All of Us cohort. Outcome categories were ordered by degree of treatment escalation; participants meeting criteria for both switching and augmentation were assigned to augmentation (the more escalated category). Points indicate the cumulative odds ratio per 1-SD increase in PGS and horizontal bars the 95% confidence intervals; an OR > 1 indicates that higher PGS is associated with a greater likelihood of a more escalated outcome, and the dashed vertical line marks the null (OR = 1). Models included age at first antidepressant record, genetic sex, income and education level as covariates. Results are shown for seven PGS (y-axis) across five diagnostic subsets (colour): participants with a recorded SNOMED depression diagnosis (Recorded Depression; red), participants without a recorded SNOMED depression diagnosis (No Recorded Depression; teal), all participants combined (Full Cohort; purple); participants with lifetime MDD as measured by the CIDI-sf (CIDI-MDD; orange); and participants without lifetime MDD as measured by the CIDI-sf (CIDI-noMDD; blue). Filled points denote associations significant after correction for 7 tested PGs traits; open points denote non-significant associations. Sample sizes are given in the header: PGS abbreviations: ADHD, attention-deficit/hyperactivity disorder; BMI, body mass index.

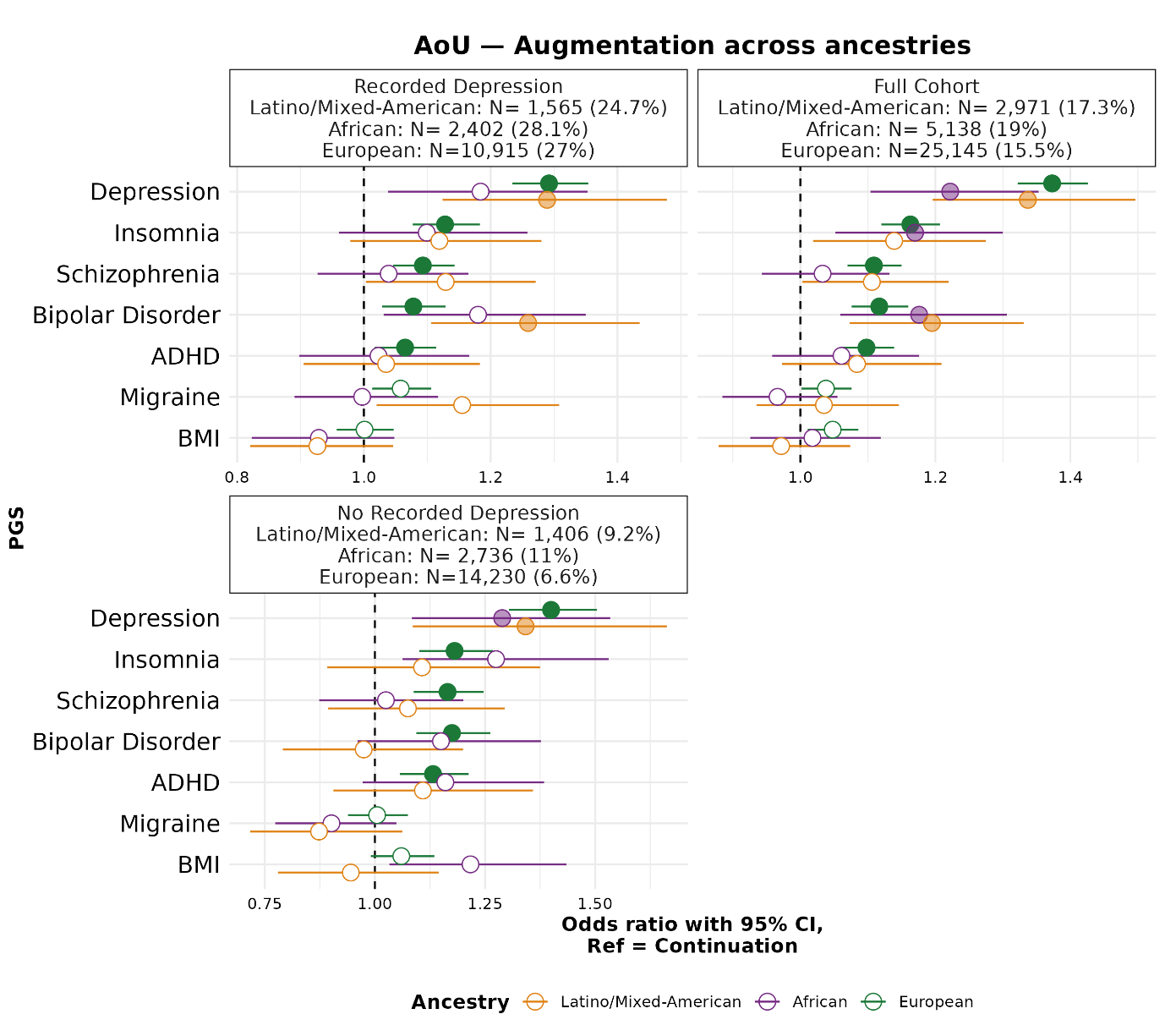

**Supplementary Figure S12. Ancestry-stratified polygenic score associations with antidepressant augmentation (All of Us).** Forest plots showing associations between standardised polygenic scores (PGS) and augmentation, relative to continuation (reference), estimated separately within ancestry groups in the All of Us cohort. Results are shown across three diagnostic subsets (sub-panels): Depression, Full Cohort, and No Depression. Points indicate odds ratios and horizontal bars the 95% confidence intervals; the dashed vertical line marks the null (OR = 1). Models included age at first antidepressant record, genetic sex, income and education level as covariates. Results are shown for seven PGS (y-axis) in three ancestry groups (colour): European (green), African (purple), and Latino/Mixed-American (orange). Filled points denote associations significant after correction for seven tests PGS traits; open points denote non-significant associations. Per-group sample sizes and the proportion of participants classified as undergoing augmentation within each diagnostic subset are given in the sub-panel headers. Note that the x-axis range differs between sub-panels. PGS abbreviations: ADHD, attention-deficit/hyperactivity disorder; BMI, body mass index.

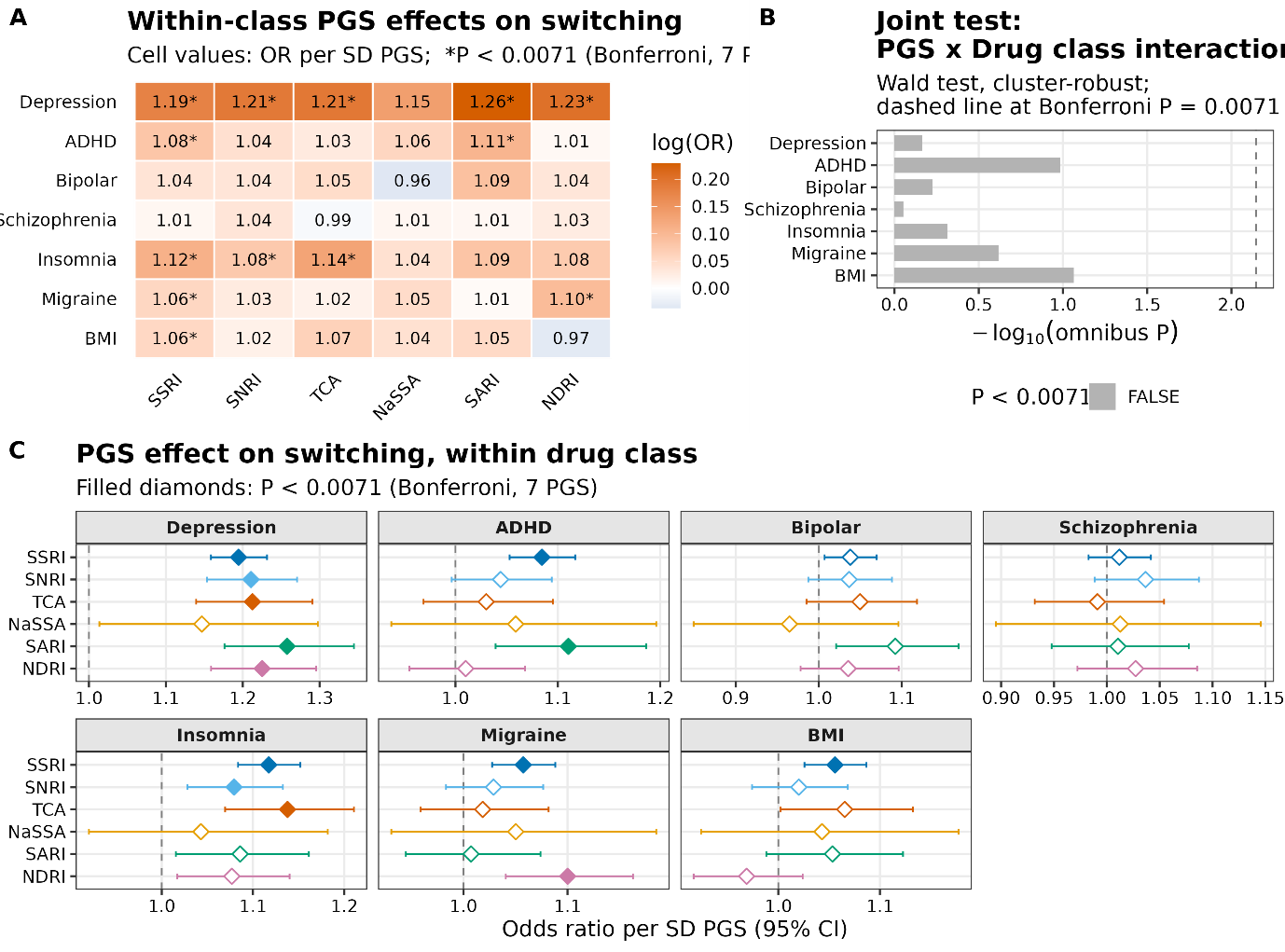

**Supplementary Figure S13. Polygenic score × antidepressant drug-class interactions for switching across the full prescribing cohort (All of Us).** Analyses were conducted across the full prescribing cohort in All of Us, with switching modelled relative to continuation (reference) and odds ratios expressed per 1-SD increase in standardised polygenic score (PGS). (**A**) Heatmap of PGS effects on switching by drug class, with cells coloured by odds ratio (orange indicating OR > 1, blue OR < 1) and asterisks marking associations significant at the Bonferroni threshold (correcting for 7 PGS). (**B**) Joint test of PGS × drug-class interaction for each PGS, shown as −log₁₀(omnibus P) from a Wald test with cluster-robust standard errors; bars extend rightward with greater interaction evidence and the dashed vertical line marks the Bonferroni threshold (P = 0.0071). (**C**) PGS effects on switching estimated within each drug class, faceted by PGS (Depression, ADHD, Bipolar, Schizophrenia, Insomnia, Migraine, BMI). Points indicate odds ratios per 1-SD PGS and horizontal bars the 95% confidence intervals; the dashed vertical line marks the null (OR = 1). Models included age at first antidepressant record, genetic sex, income and education level as covariates. Filled diamonds denote associations significant at the Bonferroni threshold (P < 0.0063); open diamonds denote non-significant associations. Drug-class abbreviations: SSRI, selective serotonin reuptake inhibitor; SNRI, serotonin–norepinephrine reuptake inhibitor; TCA, tricyclic antidepressant; NaSSA, noradrenergic and specific serotonergic antidepressant; SARI, serotonin antagonist and reuptake inhibitor; NDRI, norepinephrine–dopamine reuptake inhibitor. PGS abbreviations: ADHD, attention-deficit/hyperactivity disorder; BMI, body mass index.

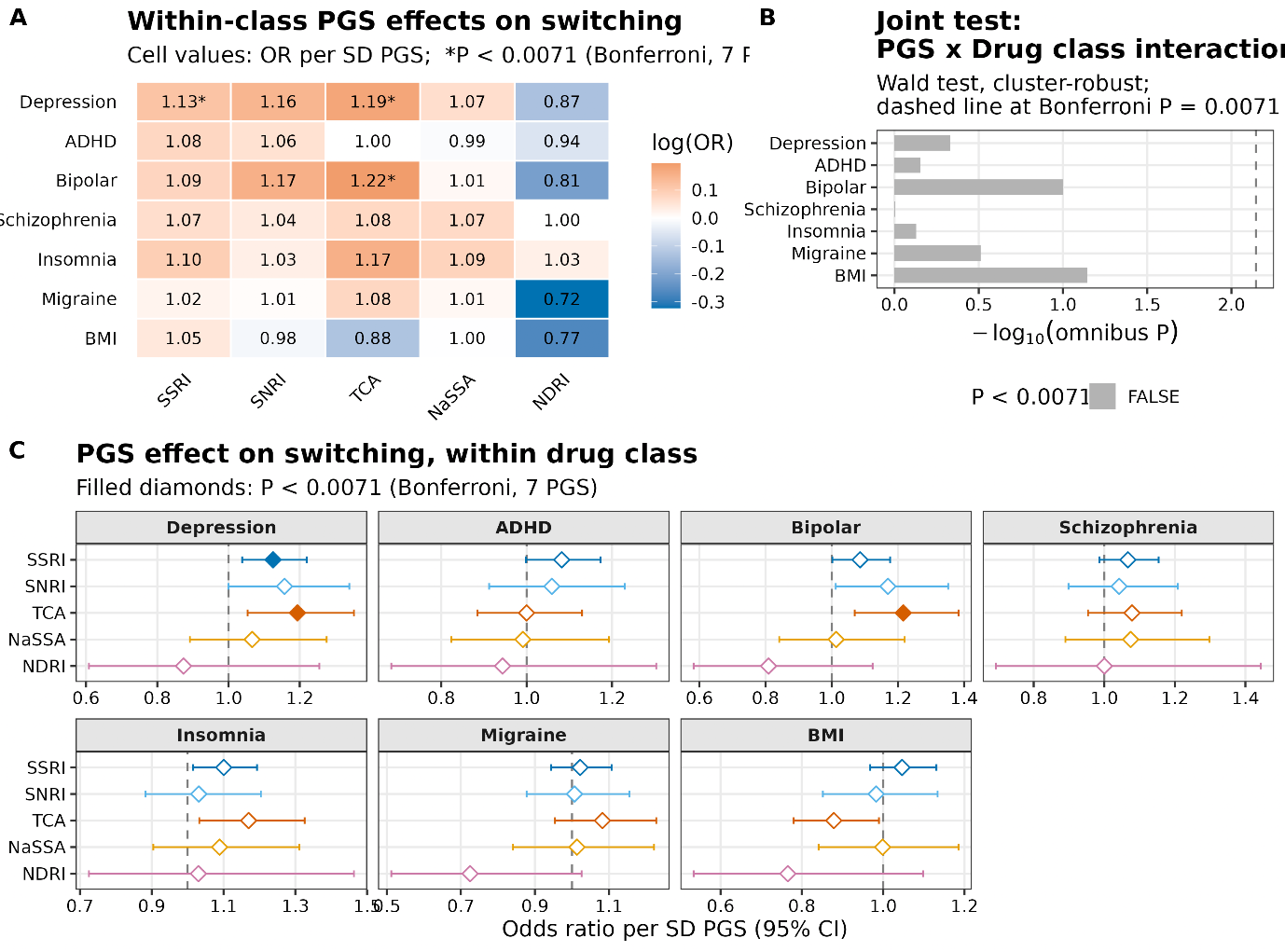

**Supplementary Figure S14. Polygenic score × antidepressant drug-class interactions for switching across the full prescribing cohort (Pharmlines).** Analyses were conducted across the full prescribing cohort in Pharmlines, with switching modelled relative to continuation (reference) and odds ratios expressed per 1-SD increase in standardised polygenic score (PGS). (**A**) Heatmap of PGS effects on switching by drug class, with cells coloured by odds ratio (orange indicating OR > 1, blue OR < 1) and asterisks marking associations significant at the Bonferroni threshold (correcting for 7 PGS). (**B**) Joint test of PGS × drug-class interaction for each PGS, shown as −log₁₀(omnibus P) from a Wald test with cluster-robust standard errors; bars extend rightward with greater interaction evidence and the dashed vertical line marks the Bonferroni threshold (P = 0.0071). (**C**) PGS effects on switching estimated within each drug class, faceted by PGS (Depression, ADHD, Bipolar, Schizophrenia, Insomnia, Migraine, BMI). Points indicate odds ratios per 1-SD PGS and horizontal bars the 95% confidence intervals; the dashed vertical line marks the null (OR = 1). Models included age at first antidepressant record, genetic sex, income and education level as covariates. Filled diamonds denote associations significant at the Bonferroni threshold (P < 0.0063); open diamonds denote non-significant associations. Drug-class abbreviations: SSRI, selective serotonin reuptake inhibitor; SNRI, serotonin–norepinephrine reuptake inhibitor; TCA, tricyclic antidepressant; NaSSA, noradrenergic and specific serotonergic antidepressant; NDRI, norepinephrine–dopamine reuptake inhibitor. PGS abbreviations: ADHD, attention-deficit/hyperactivity disorder; BMI, body mass index.

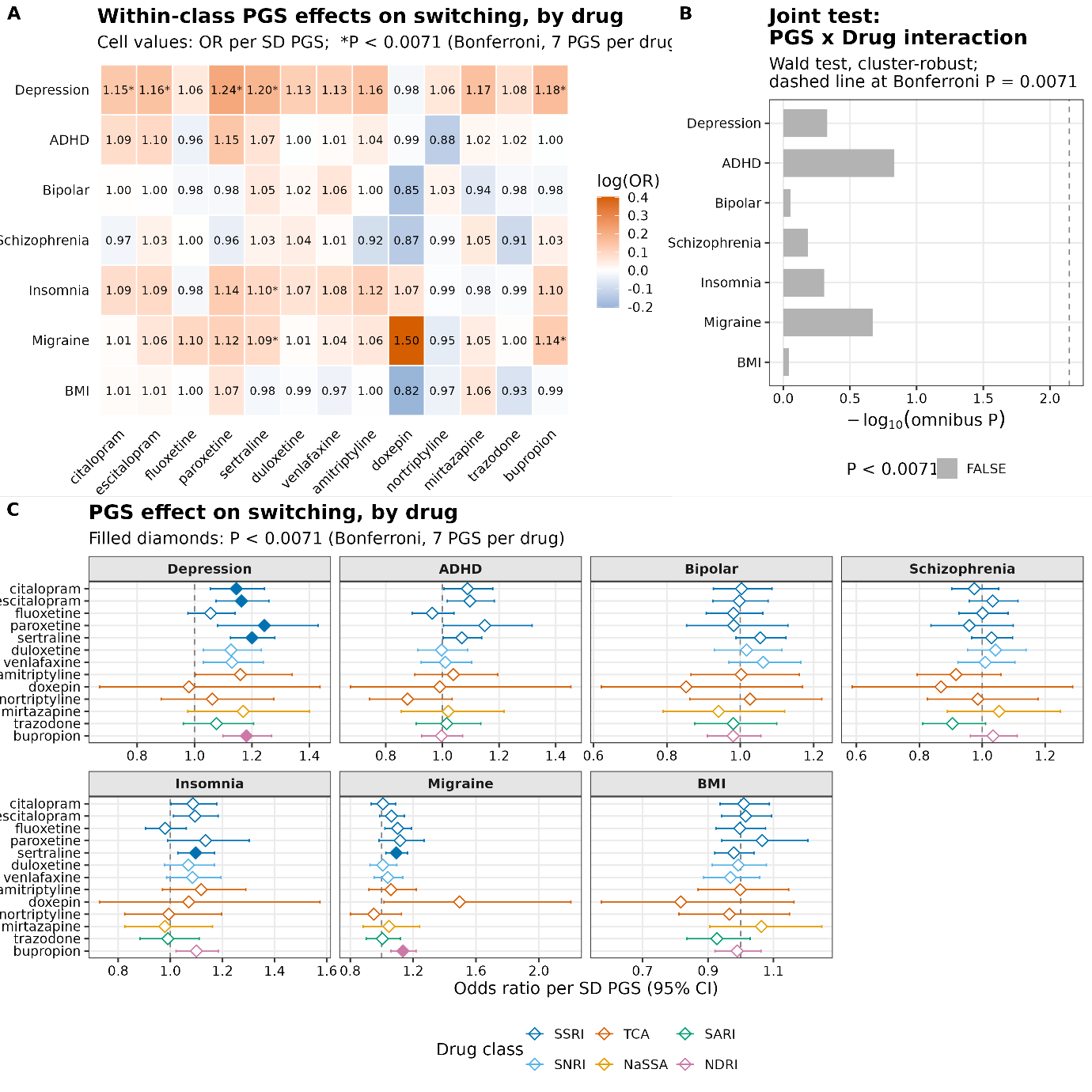

**Supplementary Figure S15. Polygenic score × antidepressant drug interactions for switching within participants with recorded depression in All of Us.** Analyses were conducted across the cohort with a recorded depression diagnosis in All of Us, with switching modelled relative to continuation (reference) and odds ratios expressed per 1-SD increase in standardised polygenic score (PGS). (**A**) Heatmap of PGS effects on switching by drug class, with cells coloured by odds ratio (orange indicating OR > 1, blue OR < 1) and asterisks marking associations significant at the Bonferroni threshold (correcting for 7 PGS). (**B**) Joint test of PGS × drug interaction for each PGS, shown as −log₁₀(omnibus P) from a Wald test with cluster-robust standard errors; bars extend rightward with greater interaction evidence and the dashed vertical line marks the Bonferroni threshold (P = 0.0071). (**C**) PGS effects on switching estimated within each drug, faceted by PGS (Depression, ADHD, Bipolar, Schizophrenia, Insomnia, Migraine, BMI). Points indicate odds ratios per 1-SD PGS and horizontal bars the 95% confidence intervals; the dashed vertical line marks the null (OR = 1). Models included age at first antidepressant record, genetic sex, income and education level as covariates. Filled diamonds denote associations significant at the Bonferroni threshold (P < 0.0071); open diamonds denote non-significant associations.

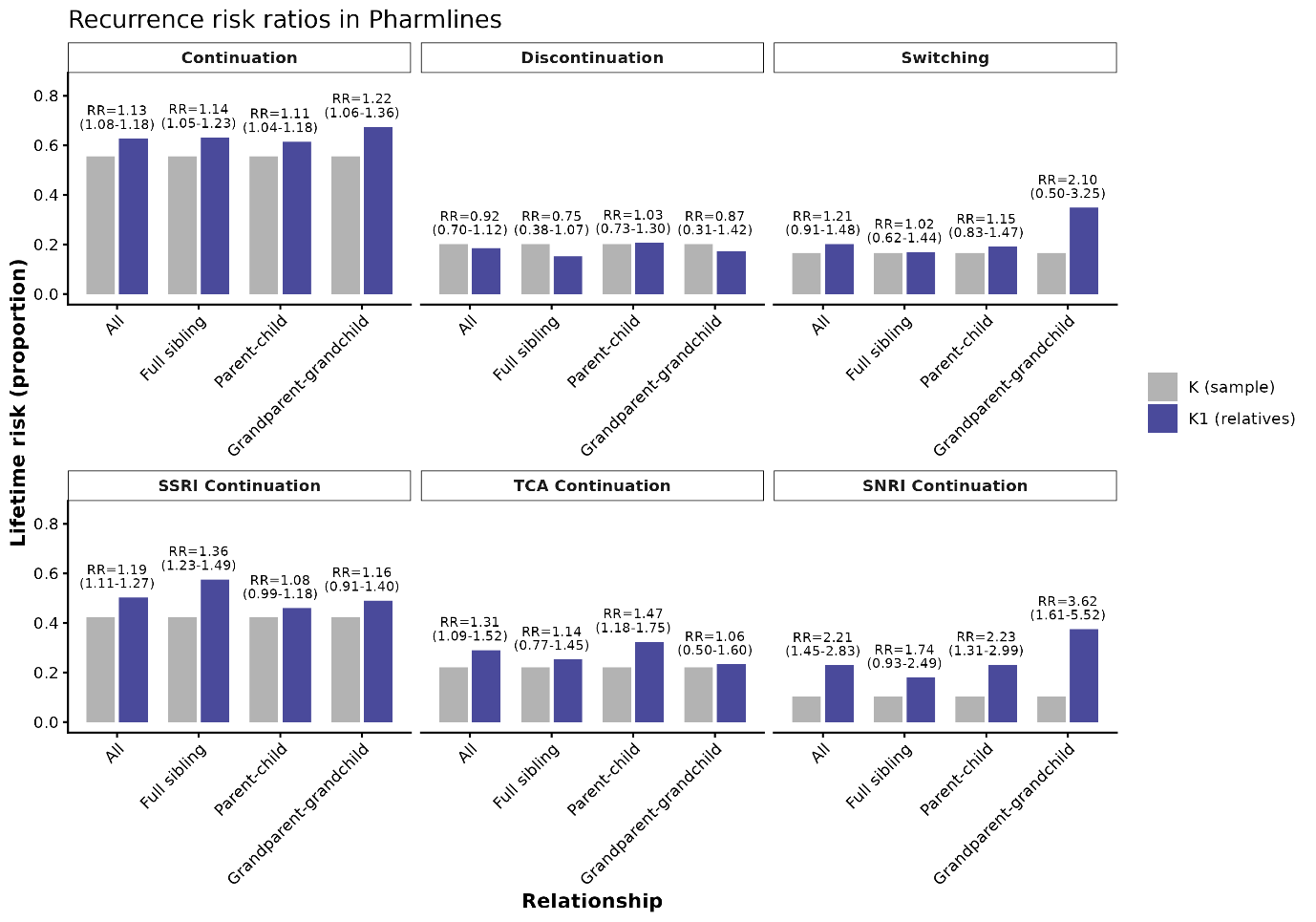

**Supplementary Figure S16. Relative risk and recurrence risk ratios of antidepressant treatment outcomes in Pharmlines.** Bars represent the population prevalence of each outcome (K; grey) and the prevalence among relatives of probands with the same outcome (K₁; blue). Recurrence risk ratios (RR) with 95% confidence intervals are shown above each comparison and were calculated as the ratio of K₁ to K, with confidence intervals derived from family-level bootstrap resampling to account for within-family clustering. Top row: pooled treatment outcomes (continuation, discontinuation, and switching) across any antidepressant. Bottom row: class-specific continuation for selective serotonin reuptake inhibitors (SSRI), tricyclic antidepressants (TCA), and serotonin–norepinephrine reuptake inhibitors (SNRI). Estimates are presented across relationship types: all relative pairs combined, full siblings, parent–child, and grandparent–grandchild pairs. Higher recurrence risk ratios indicate greater familial clustering of the outcome.

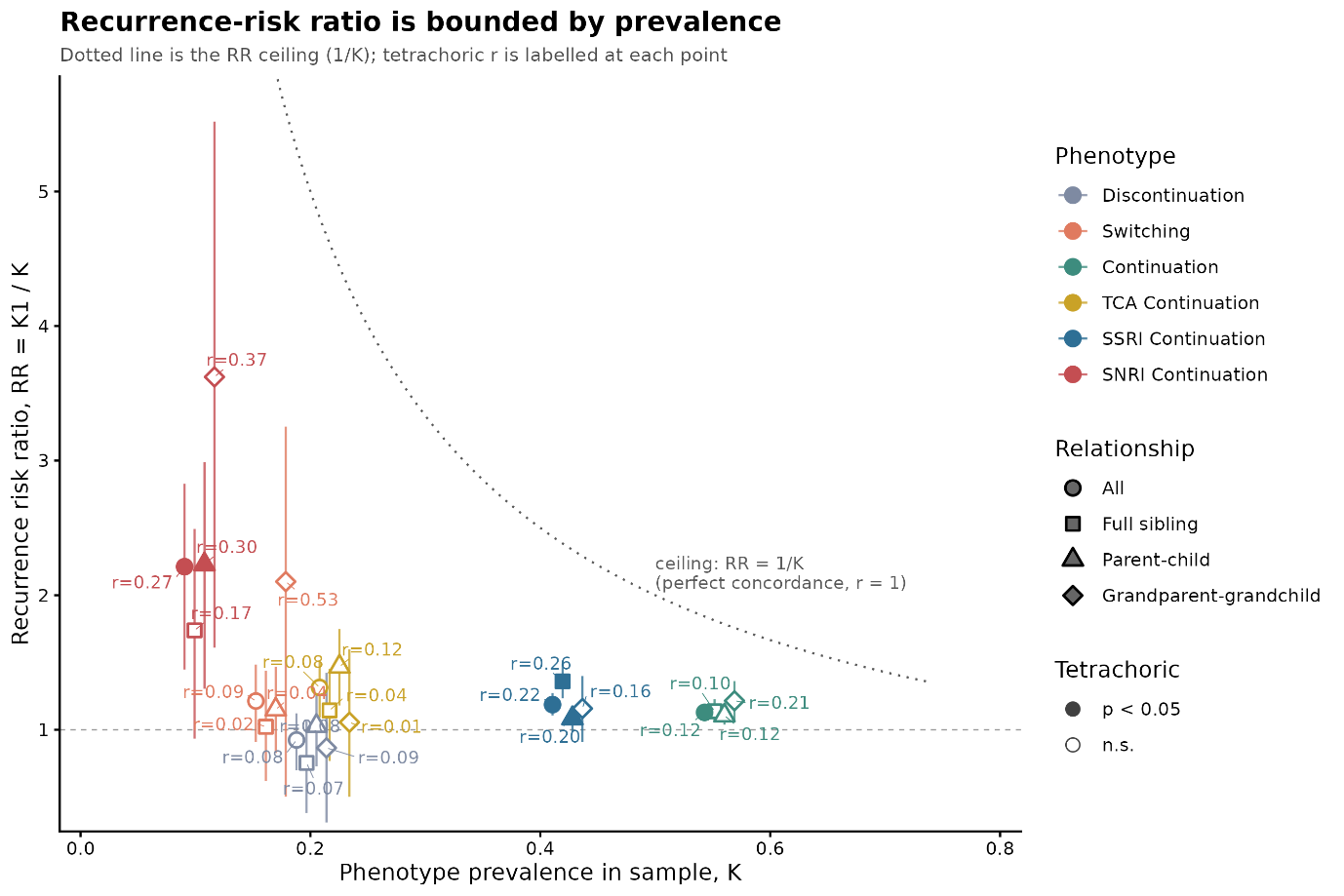
**Supplementary Figure S17. Familial aggregation of antidepressant-response phenotypes in Pharmlines.** Observed recurrence-risk ratios (RR = K₁/K, y-axis) plotted against phenotype prevalence in the sample (K, x-axis; identical across relationship types within a phenotype, points offset horizontally for clarity) for each phenotype (colour) and relationship type (symbol). Points are filled where the corresponding tetrachoric correlation is significant (p < 0.05) and open otherwise; the measured tetrachoric correlation (r) is labelled beside each point. Error bars are 95% confidence intervals on the RR (bootstrap). The dotted curved line marks the theoretical ceiling RR = 1/K, the maximum attainable recurrence-risk ratio at each prevalence. Because this ceiling rises as K falls, a given liability correlation yields a larger RR in rarer phenotypes. K₁, recurrence risk in relatives of affected probands; K, phenotype prevalence in the sample.

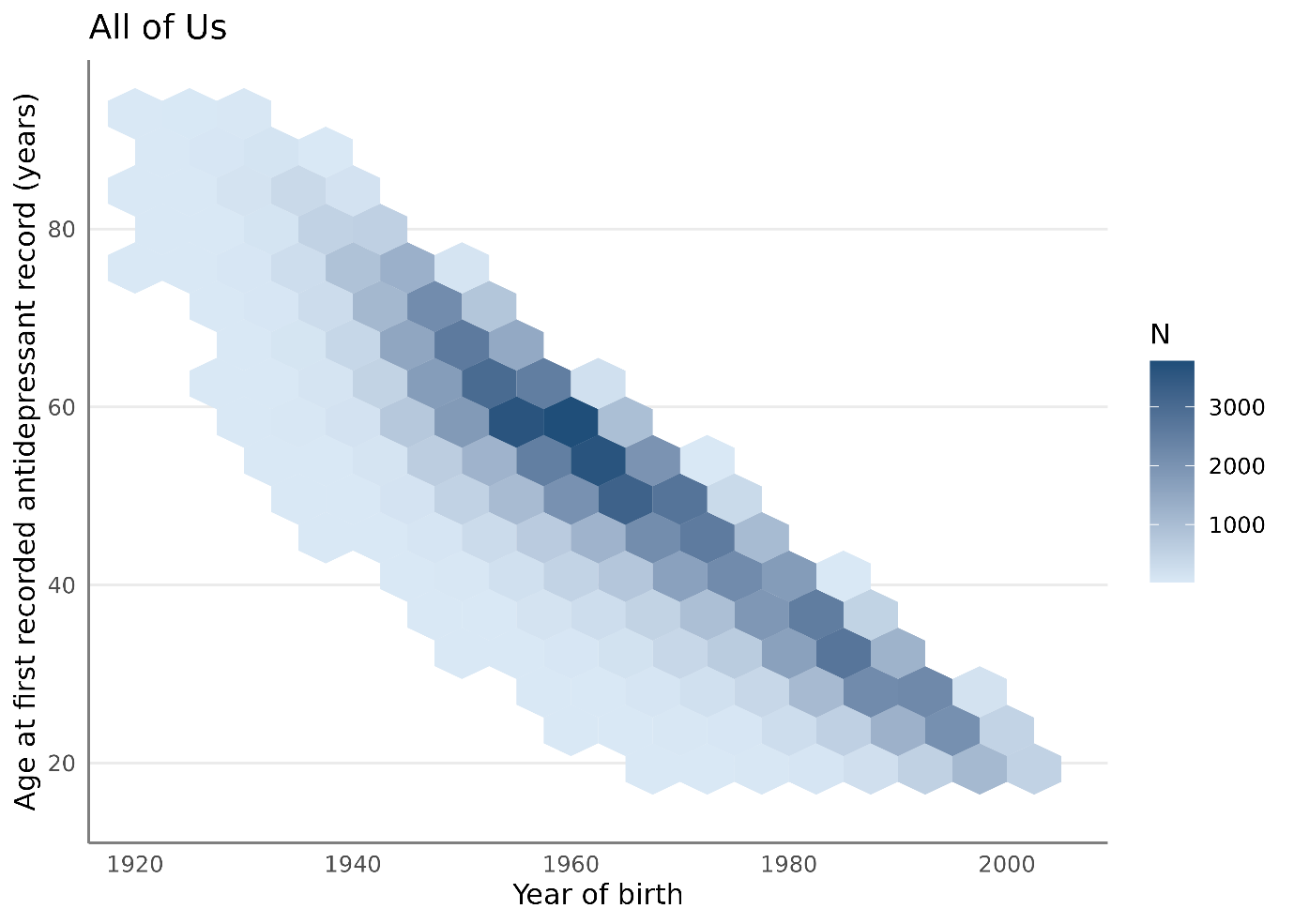

**Supplementary Figure S18. Age at first recorded antidepressant record by year of birth in the All of Us cohort, shown as a hexagonal bin plot with bin counts indicated by colour.** Age at first recorded antidepressant record is left-truncated at 12 years in All of Us due to study inclusion criteria. The strong linear relationship between year of birth and age at first medication record reflects administrative left-censoring of medication records from the 1980s onwards, rather than true variation in age at treatment initiation, and should be interpreted accordingly. Hexagonal bins containing fewer than 20 participants were suppressed per the All of Us Data and Statistics Dissemination Policy and appear as blank space.

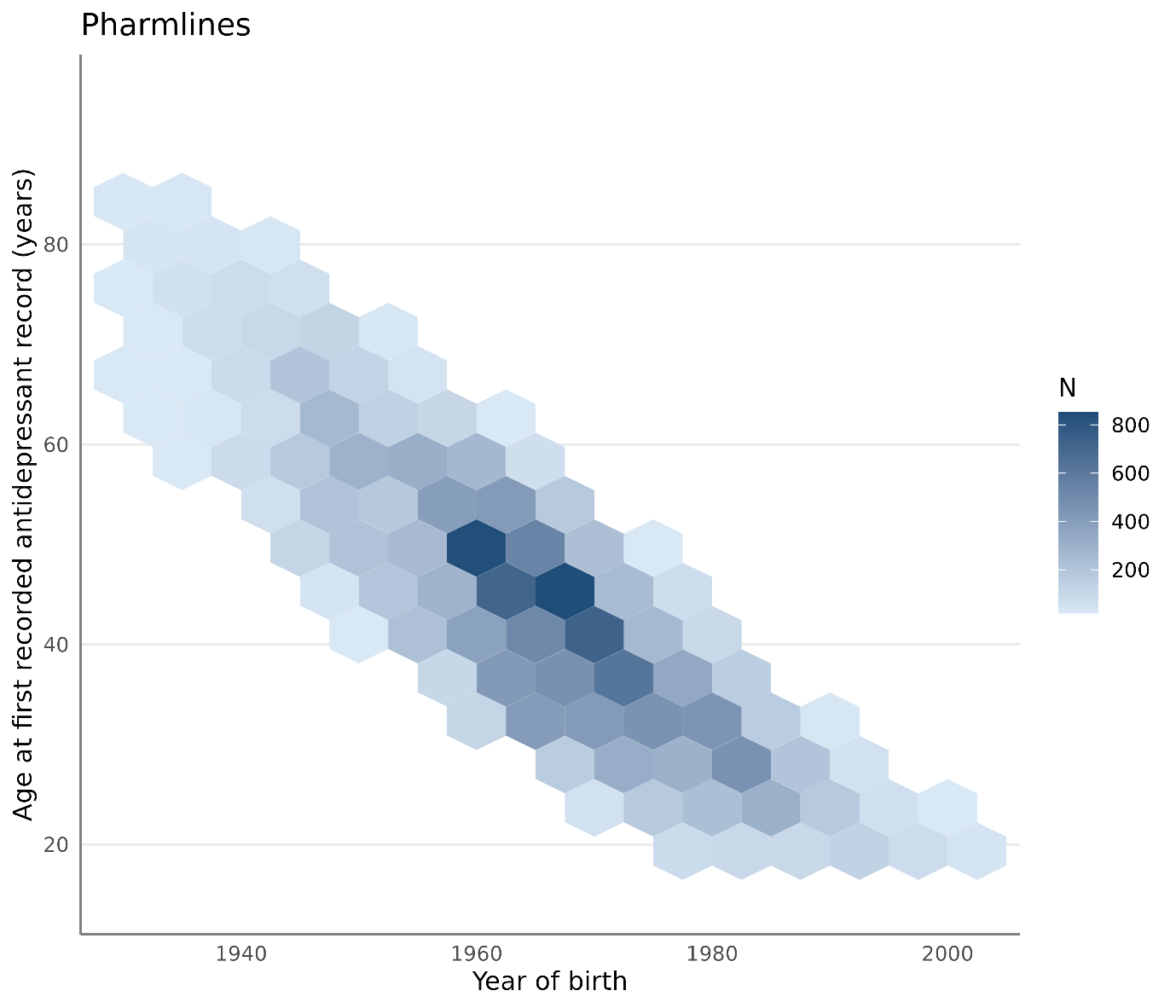

**Supplementary Figure S19. Age at first recorded antidepressant record by year of birth in the Pharmlines cohort, shown as a hexagonal bin plot with bin counts indicated by colour.** Age at first recorded antidepressant record is left-truncated at 12 years due to study inclusion criteria. The strong linear relationship between year of birth and age at first medication record reflects administrative left-censoring of medication records from 1994 onwards, rather than true variation in age at treatment initiation, and should be interpreted accordingly. Hexagonal bins containing fewer than 10 participants were suppressed per Lifelines policy and appear as blank space.
