## Supplementary_Tables for "Polygenic and familial contributions to antidepressant continuation, switching, discontinuation and augmentation in the All of Us and Pharmlines cohorts"

**README**

| Sheet Name | Analyses |
|---|---|
| Table1 | Cohort Demographics split by pooled primary treatment outcomes |
| Table2 | Phenotypic associations with pooled primary treatment outcomes in AoU |
| Table3 | Phenotypic associations with pooled primary treatment outcomes in Pharmlines |
| Table4 | PGS associations with pooled primary treatment outcomes in AoU |
| Table5 | PGS associations with pooled primary treatment outcomes in Pharmlines |
| Table6 | Pooled switching sensitivity analyses for PGS associations in AoU Europeans |
| Table7 | Pooled switching sensitivity analyses for PGS associations in Pharmlines |
| Table8 | Pooled discontinuation sensitivity analyses for PGS associations in AoU Europeans |
| Table9 | Pooled discontinuation sensitivity analyses for PGS associations in Pharmlines |
| Table10 | PGS associations with augmentation in AoU |
| Table11 | PGS associations with augmentation in Pharmlines |
| Table12 | PGS associations with ordinal treatment escalation in AoU |
| Table13 | PGS x drugclass switching analyses in AoU |
| Table14 | PGS x drugclass switching analyses in Pharmlines |
| Table 15 | PGS x drug switching analyses in AoU |
| Table16 | Recurrence risk ratios of treatment outcomes in Pharmlines |
| Table17 | Tetrachoric correlations of treatment outcomes in Pharmlines |
| Table18 | List of antidepressants considered |
| Table 19 | List of atypical antipsychotics considered |
| Table20 | GWAS summary statistics used within the study |

**Supplementary Table 1. Basic characteristics across three mutually exclusive antidepressant outcomes (continuation, switching, and discontinuation), pooled across all antidepressants in the full prescribing cohort, in two study biobanks: AoU and Pharmlines (LL).** Percentages are calculated among non-missing observations. Abbreviations: AD, antidepressant. [a] Only genetic data for LL participants of European genetic-inferred ancestry was carried forward to linkage with IADB prescription records. Ancestry is unknown for remaining LL participants; however, LL is a predominantly European-ancestry cohort. [b] Self-reported annual household income: AoU 1-8 scale (1= <$US25K, 8= ≥$US200K; derived from the question "What is your annual household income from all sources?"); LL 306-3,500 scale (Disposable household income derived from household income measured at baseline by self-report to the question "what is the net income per month?" Disposable household income equals the average household income divided by equivalence factors based on the number of family members from the Statistics Netherlands website).[c] Self-reported education: AoU 1-4 scale (Derived from the question "What is the highest grade or year of school you completed?"; 1=Less than a high school degree, 4=College Postgraduate degree); LL 1-20 point scale (total education years; measured at baseline by self-report to the question "What is the highest level of education you have attained?" [d] Depression: AoU at least one instance of a depression SNOMED diagnostic code; LL depression ascertained by both current depression (measured over 3 MINI International Neuropsychiatric Interview [MINI] assessment at baseline, second assessment and third assessment) and the lifetime depression in the 3rd MINI assessment and the LIDAS questionnaire measured on a subset of individuals.[e] Depression symptom count: AoU as the total symptom count from the MDD CIDI-sf; Lifelines as the median total symptom count across all lifetime and current depression MINI and LIDAS assessments. [f]Anxiety: AoU at least one instance of an anxiety SNOMED diagnostic code; LL lifetime anxiety. [g]Recorded deaths from Netherlands Statistics (CBS). Mortality signficantly differed by treatment pattern group, chi-squared(2, N=11,884) = 29.24, p < 0.001.

| | **All of Us (AoU; N=87,787)** | | | **Lifelines (LL; N=11,884)** | | |
|---|---|---|---|---|---|---|
| | **Continuation (N=42,197)** | **Discontinuation (N=18,201)** | **Switching (N=27,389)** | **Continuation (N=7,162)** | **Discontinuation (N=2,589)** | **Switching (N=2,133)** |
| **Age at first antidepressant record (yrs)** | 50.3 (15.7) | 50.5 (16.2) | 45.6 (14.7) | 44.2 (13.2) | 47.2 (14.8) | 41.6 (12.7) |
| **Genetic Sex** | | | | | | |
| Female | 29,299 (70%) | 12,187 (68%) | 19,550 (72%) | 3,477 (73%) | 1,116 (66%) | 986 (69%) |
| Male | 12,498 (30%) | 5,831 (32%) | 7,563 (28%) | 1,305 (27%) | 565 (34%) | 444 (31%) |
| **Ancestry**[a] | | | | | | |
| European | 24,135 (71%) | 8,865 (64%) | 15,608 (73%) | 4,784 (100%) | 1,682 (100%) | 1,430 (100%) |
| African | 5,594 (16%) | 2,538 (18%) | 3,411 (16%) | N/A | N/A | N/A |
| Latino/Admixed-American | 3,682 (11%) | 2,105 (15%) | 2,102 (9.8%) | N/A | N/A | N/A |
| Other ancestry | 788 (2.3%) | 328 (2.4%) | 336 (1.6%) | N/A | N/A | N/A |
| **Income level**[b] | 4.4 (2.2) | 4.0 (2.2) | 3.9 (2.1) | 1,549.2 (549.5) | 1,547.9 (557.3) | 1,481.1 (550.6) |
| **Education level**[c] | 3.0 (0.9) | 2.8 (0.9) | 2.9 (0.9) | 13.6 (4.2) | 12.9 (4.1) | 13.4 (4.2) |
| **Recorded Depression**[d] | 16,523 (39%) | 5,945 (33%) | 19,012 (69%) | 2,450 (35%) | 582 (23%) | 994 (48%) |
| **MDD CIDI-sf** | 3,642 (53%) | 1,414 (57%) | 3,416 (77%) | | | |
| **Depression symptom count**[e] | 4.1 (2.9) | 4.3 (3.0) | 5.4 (2.5) | 1.2 (1.8) | 1.0 (1.7) | 2.2 (2.4) |
| **Recorded Anxiety**[f] | 22,758 (54%) | 8,286 (46%) | 20,997 (77%) | 489 (26%) | 81 (13%) | 215 (39%) |
| **Recorded deaths**[g] | | | | 310 (4.3%) | 182 (7.0%) | 104 (4.9%) |

**Supplementary Table 2. Multivariable logistic regression results for phenotypic associations with antidepressant treatment outcomes across ancestry groups and clinical subgroups in All of Us.** Each model estimates the association between covariates and one of two binary outcomes — discontinuation versus continuation, or switching versus continuation — fitted separately within each combination of genetically inferred ancestry (European, African, Latino/Mixed-American), clinical group (Full Cohort; subgroups defined by recorded depression status, recorded anxiety, and CIDI-SF–derived MDD classification), and predictor. Within each model, one exposure (the "Dependent variable" column) is modelled alongside a common covariate set (age at first antidepressant, sex at birth, income band, and educational attainment); for categorical terms, each non-reference level appears on its own row and the reference level is omitted; for quantitiave terms, variables were standardised (mean 0, SD 1); each odds ratio therefore reflects the change in odds per one-standard-deviation increase.
Columns: Population = inferred genetic ancestry; ClinicalGroup = cohort or clinical subgroup; Outcome = the binary contrast modelled; Dependent variable = the exposure for that model; variable = the individual model term; odds_ratio = exponentiated coefficient; lower_95/upper_95 = 95% confidence interval bounds for the odds ratio; p_value = nominal (uncorrected) p-value; Bonf_P = Bonferroni-corrected p-value (capped at 1.00); N_Total = model sample size; N_Reference = number in the reference (continuation) group; N_Case = number in the comparison (discontinuation or switching) group. Odds ratios >1 indicate higher odds of the comparison outcome relative to continuation. Bonferroni correction was applied within each specification (ancestry × clinical group × outcome contrast) for the max 14 traits tested. P-values reported as "<2.2e-16" reflect R's floating-point reporting limit.
Reference (omitted) categories: Sex at birth: Female; Income: lowest band (<$25k); Education level: lowest band (less than high school / fewer than twelve years); Alcohol_Frequency: Abstained (Never); Smoking_Frequency: Abstained (Never); Stress: Never.

| Population | ClinicalGroup | Outcome | Dependent | variable | odds_ratio | lower_95 | upper_95 | p_value | Bonf_P | N_Total | N_Reference | N_Case |
|---|---|---|---|---|---|---|---|---|---|---|---|---|
| European | Full Cohort | Discontinuation vs Continuation | Alcohol_Frequency | (Intercept) | 0.923 | 0.731 | 1.165 | 4.99e-01 | 1.00e+00 | 28419 | 21045 | 7374 |
| European | Full Cohort | Discontinuation vs Continuation | Alcohol_Frequency | age_at_first_AD | 1.087 | 1.058 | 1.117 | 1.29e-09 | 1.81e-08 | 28419 | 21045 | 7374 |
| European | Full Cohort | Discontinuation vs Continuation | Alcohol_Frequency | sex_at_birthMale | 1.118 | 1.055 | 1.184 | 1.69e-04 | 2.37e-03 | 28419 | 21045 | 7374 |
| European | Full Cohort | Discontinuation vs Continuation | Alcohol_Frequency | Income>200k | 0.619 | 0.552 | 0.695 | 4.55e-16 | 6.36e-15 | 28419 | 21045 | 7374 |
| European | Full Cohort | Discontinuation vs Continuation | Alcohol_Frequency | Income100-150k | 0.560 | 0.507 | 0.618 | <2.2e-16 | <2.2e-16 | 28419 | 21045 | 7374 |
| European | Full Cohort | Discontinuation vs Continuation | Alcohol_Frequency | Income150-200k | 0.553 | 0.488 | 0.627 | <2.2e-16 | <2.2e-16 | 28419 | 21045 | 7374 |
| European | Full Cohort | Discontinuation vs Continuation | Alcohol_Frequency | Income25-35k | 0.753 | 0.675 | 0.842 | 5.25e-07 | 7.35e-06 | 28419 | 21045 | 7374 |
| European | Full Cohort | Discontinuation vs Continuation | Alcohol_Frequency | Income35-50k | 0.675 | 0.608 | 0.749 | 1.98e-13 | 2.77e-12 | 28419 | 21045 | 7374 |
| European | Full Cohort | Discontinuation vs Continuation | Alcohol_Frequency | Income50-75k | 0.689 | 0.627 | 0.756 | 5.56e-15 | 7.79e-14 | 28419 | 21045 | 7374 |
| European | Full Cohort | Discontinuation vs Continuation | Alcohol_Frequency | Income75-100k | 0.593 | 0.535 | 0.656 | <2.2e-16 | <2.2e-16 | 28419 | 21045 | 7374 |
| European | Full Cohort | Discontinuation vs Continuation | Alcohol_Frequency | Education_levelCollege Postgraduate | 0.604 | 0.493 | 0.741 | 1.30e-06 | 1.82e-05 | 28419 | 21045 | 7374 |
| European | Full Cohort | Discontinuation vs Continuation | Alcohol_Frequency | Education_levelCollege Undergraduate | 0.683 | 0.561 | 0.832 | 1.52e-04 | 2.12e-03 | 28419 | 21045 | 7374 |
| European | Full Cohort | Discontinuation vs Continuation | Alcohol_Frequency | Education_levelTwelve or GED | 0.806 | 0.657 | 0.989 | 3.85e-02 | 5.39e-01 | 28419 | 21045 | 7374 |
| European | Full Cohort | Discontinuation vs Continuation | Alcohol_Frequency | Alcohol_Frequency2-3 Week | 0.734 | 0.622 | 0.865 | 2.32e-04 | 3.25e-03 | 28419 | 21045 | 7374 |
| European | Full Cohort | Discontinuation vs Continuation | Alcohol_Frequency | Alcohol_Frequency2-4 Month | 0.751 | 0.640 | 0.882 | 4.84e-04 | 6.77e-03 | 28419 | 21045 | 7374 |
| European | Full Cohort | Discontinuation vs Continuation | Alcohol_Frequency | Alcohol_Frequency4plus Week | 0.736 | 0.623 | 0.869 | 3.04e-04 | 4.25e-03 | 28419 | 21045 | 7374 |
| European | Full Cohort | Discontinuation vs Continuation | Alcohol_Frequency | Alcohol_FrequencyMonthly or Less | 0.803 | 0.687 | 0.939 | 5.87e-03 | 8.22e-02 | 28419 | 21045 | 7374 |
| European | Full Cohort | Discontinuation vs Continuation | Alcohol_Frequency | Alcohol_FrequencyNever (former) | 0.848 | 0.721 | 0.997 | 4.54e-02 | 6.36e-01 | 28419 | 21045 | 7374 |
| European | Full Cohort | Switching vs Continuation | Alcohol_Frequency | (Intercept) | 1.110 | 0.902 | 1.367 | 3.24e-01 | 1.00e+00 | 34724 | 21045 | 13679 |
| European | Full Cohort | Switching vs Continuation | Alcohol_Frequency | age_at_first_AD | 0.698 | 0.682 | 0.714 | <2.2e-16 | <2.2e-16 | 34724 | 21045 | 13679 |
| European | Full Cohort | Switching vs Continuation | Alcohol_Frequency | sex_at_birthMale | 1.048 | 0.997 | 1.101 | 6.45e-02 | 9.04e-01 | 34724 | 21045 | 13679 |
| European | Full Cohort | Switching vs Continuation | Alcohol_Frequency | Income>200k | 0.432 | 0.390 | 0.479 | <2.2e-16 | <2.2e-16 | 34724 | 21045 | 13679 |
| European | Full Cohort | Switching vs Continuation | Alcohol_Frequency | Income100-150k | 0.587 | 0.541 | 0.637 | <2.2e-16 | <2.2e-16 | 34724 | 21045 | 13679 |
| European | Full Cohort | Switching vs Continuation | Alcohol_Frequency | Income150-200k | 0.474 | 0.427 | 0.527 | <2.2e-16 | <2.2e-16 | 34724 | 21045 | 13679 |
| European | Full Cohort | Switching vs Continuation | Alcohol_Frequency | Income25-35k | 0.857 | 0.782 | 0.939 | 9.39e-04 | 1.31e-02 | 34724 | 21045 | 13679 |
| European | Full Cohort | Switching vs Continuation | Alcohol_Frequency | Income35-50k | 0.806 | 0.739 | 0.878 | 7.47e-07 | 1.05e-05 | 34724 | 21045 | 13679 |
| European | Full Cohort | Switching vs Continuation | Alcohol_Frequency | Income50-75k | 0.748 | 0.692 | 0.809 | 2.90e-13 | 4.06e-12 | 34724 | 21045 | 13679 |
| European | Full Cohort | Switching vs Continuation | Alcohol_Frequency | Income75-100k | 0.635 | 0.584 | 0.690 | <2.2e-16 | <2.2e-16 | 34724 | 21045 | 13679 |
| European | Full Cohort | Switching vs Continuation | Alcohol_Frequency | Education_levelCollege Postgraduate | 0.742 | 0.619 | 0.889 | 1.21e-03 | 1.69e-02 | 34724 | 21045 | 13679 |
| European | Full Cohort | Switching vs Continuation | Alcohol_Frequency | Education_levelCollege Undergraduate | 0.808 | 0.678 | 0.964 | 1.76e-02 | 2.46e-01 | 34724 | 21045 | 13679 |
| European | Full Cohort | Switching vs Continuation | Alcohol_Frequency | Education_levelTwelve or GED | 0.939 | 0.783 | 1.127 | 5.00e-01 | 1.00e+00 | 34724 | 21045 | 13679 |
| European | Full Cohort | Switching vs Continuation | Alcohol_Frequency | Alcohol_Frequency2-3 Week | 0.866 | 0.748 | 1.001 | 5.24e-02 | 7.33e-01 | 34724 | 21045 | 13679 |
| European | Full Cohort | Switching vs Continuation | Alcohol_Frequency | Alcohol_Frequency2-4 Month | 0.929 | 0.806 | 1.071 | 3.11e-01 | 1.00e+00 | 34724 | 21045 | 13679 |
| European | Full Cohort | Switching vs Continuation | Alcohol_Frequency | Alcohol_Frequency4plus Week | 0.980 | 0.846 | 1.136 | 7.91e-01 | 1.00e+00 | 34724 | 21045 | 13679 |
| European | Full Cohort | Switching vs Continuation | Alcohol_Frequency | Alcohol_FrequencyMonthly or Less | 1.059 | 0.923 | 1.217 | 4.14e-01 | 1.00e+00 | 34724 | 21045 | 13679 |
| European | Full Cohort | Switching vs Continuation | Alcohol_Frequency | Alcohol_FrequencyNever (former) | 1.252 | 1.085 | 1.445 | 2.09e-03 | 2.92e-02 | 34724 | 21045 | 13679 |
| European | Full Cohort | Discontinuation vs Continuation | BMI | (Intercept) | 0.745 | 0.614 | 0.902 | 2.60e-03 | 3.64e-02 | 27987 | 20853 | 7134 |
| European | Full Cohort | Discontinuation vs Continuation | BMI | age_at_first_AD | 1.084 | 1.055 | 1.113 | 5.07e-09 | 7.10e-08 | 27987 | 20853 | 7134 |
| European | Full Cohort | Discontinuation vs Continuation | BMI | sex_at_birthMale | 1.135 | 1.070 | 1.203 | 2.09e-05 | 2.93e-04 | 27987 | 20853 | 7134 |
| European | Full Cohort | Discontinuation vs Continuation | BMI | Income>200k | 0.585 | 0.521 | 0.657 | <2.2e-16 | <2.2e-16 | 27987 | 20853 | 7134 |
| European | Full Cohort | Discontinuation vs Continuation | BMI | Income100-150k | 0.544 | 0.493 | 0.600 | <2.2e-16 | <2.2e-16 | 27987 | 20853 | 7134 |
| European | Full Cohort | Discontinuation vs Continuation | BMI | Income150-200k | 0.523 | 0.461 | 0.593 | <2.2e-16 | <2.2e-16 | 27987 | 20853 | 7134 |
| European | Full Cohort | Discontinuation vs Continuation | BMI | Income25-35k | 0.734 | 0.656 | 0.821 | 6.16e-08 | 8.63e-07 | 27987 | 20853 | 7134 |
| European | Full Cohort | Discontinuation vs Continuation | BMI | Income35-50k | 0.656 | 0.590 | 0.730 | 7.47e-15 | 1.05e-13 | 27987 | 20853 | 7134 |
| European | Full Cohort | Discontinuation vs Continuation | BMI | Income50-75k | 0.685 | 0.624 | 0.753 | 3.24e-15 | 4.54e-14 | 27987 | 20853 | 7134 |
| European | Full Cohort | Discontinuation vs Continuation | BMI | Income75-100k | 0.580 | 0.523 | 0.642 | <2.2e-16 | <2.2e-16 | 27987 | 20853 | 7134 |
| European | Full Cohort | Discontinuation vs Continuation | BMI | Education_levelCollege Postgraduate | 0.571 | 0.465 | 0.700 | 7.34e-08 | 1.03e-06 | 27987 | 20853 | 7134 |
| European | Full Cohort | Discontinuation vs Continuation | BMI | Education_levelCollege Undergraduate | 0.655 | 0.538 | 0.798 | 2.54e-05 | 3.55e-04 | 27987 | 20853 | 7134 |
| European | Full Cohort | Discontinuation vs Continuation | BMI | Education_levelTwelve or GED | 0.790 | 0.644 | 0.970 | 2.43e-02 | 3.40e-01 | 27987 | 20853 | 7134 |
| European | Full Cohort | Discontinuation vs Continuation | BMI | BMI | 0.933 | 0.906 | 0.962 | 5.91e-06 | 8.28e-05 | 27987 | 20853 | 7134 |
| European | Full Cohort | Switching vs Continuation | BMI | (Intercept) | 1.162 | 0.979 | 1.378 | 8.57e-02 | 1.00e+00 | 34428 | 20853 | 13575 |
| European | Full Cohort | Switching vs Continuation | BMI | age_at_first_AD | 0.707 | 0.691 | 0.723 | <2.2e-16 | <2.2e-16 | 34428 | 20853 | 13575 |
| European | Full Cohort | Switching vs Continuation | BMI | sex_at_birthMale | 1.054 | 1.003 | 1.107 | 3.67e-02 | 5.13e-01 | 34428 | 20853 | 13575 |
| European | Full Cohort | Switching vs Continuation | BMI | Income>200k | 0.429 | 0.387 | 0.475 | <2.2e-16 | <2.2e-16 | 34428 | 20853 | 13575 |
| European | Full Cohort | Switching vs Continuation | BMI | Income100-150k | 0.569 | 0.525 | 0.617 | <2.2e-16 | <2.2e-16 | 34428 | 20853 | 13575 |
| European | Full Cohort | Switching vs Continuation | BMI | Income150-200k | 0.456 | 0.410 | 0.507 | <2.2e-16 | <2.2e-16 | 34428 | 20853 | 13575 |
| European | Full Cohort | Switching vs Continuation | BMI | Income25-35k | 0.838 | 0.765 | 0.919 | 1.66e-04 | 2.32e-03 | 34428 | 20853 | 13575 |

| | | | | | | | | | | | | |
|---|---|---|---|---|---|---|---|---|---|---|---|---|
| European | Full Cohort | Switching vs Continuation | BMI | Income35-50k | 0.785 | 0.721 | 0.855 | 3.12e-08 | 4.37e-07 | 34428 | 20853 | 13575 |
| European | Full Cohort | Switching vs Continuation | BMI | Income50-75k | 0.729 | 0.674 | 0.787 | 1.29e-15 | 1.81e-14 | 34428 | 20853 | 13575 |
| European | Full Cohort | Switching vs Continuation | BMI | Income75-100k | 0.610 | 0.561 | 0.662 | <2.2e-16 | <2.2e-16 | 34428 | 20853 | 13575 |
| European | Full Cohort | Switching vs Continuation | BMI | Education_levelCollege Postgraduate | 0.743 | 0.620 | 0.890 | 1.23e-03 | 1.72e-02 | 34428 | 20853 | 13575 |
| European | Full Cohort | Switching vs Continuation | BMI | Education_levelCollege Undergraduate | 0.803 | 0.674 | 0.957 | 1.41e-02 | 1.97e-01 | 34428 | 20853 | 13575 |
| European | Full Cohort | Switching vs Continuation | BMI | Education_levelTwelve or GED | 0.944 | 0.787 | 1.132 | 5.32e-01 | 1.00e+00 | 34428 | 20853 | 13575 |
| European | Full Cohort | Switching vs Continuation | BMI | BMI | 1.127 | 1.101 | 1.153 | <2.2e-16 | <2.2e-16 | 34428 | 20853 | 13575 |
| European | Full Cohort | Discontinuation vs Continuation | Education_level | (Intercept) | 0.761 | 0.631 | 0.917 | 4.10e-03 | 5.74e-02 | 28728 | 21256 | 7472 |
| European | Full Cohort | Discontinuation vs Continuation | Education_level | age_at_first_AD | 1.089 | 1.060 | 1.118 | 3.29e-10 | 4.60e-09 | 28728 | 21256 | 7472 |
| European | Full Cohort | Discontinuation vs Continuation | Education_level | sex_at_birthMale | 1.118 | 1.056 | 1.184 | 1.33e-04 | 1.86e-03 | 28728 | 21256 | 7472 |
| European | Full Cohort | Discontinuation vs Continuation | Education_level | Income>200k | 0.595 | 0.531 | 0.666 | <2.2e-16 | <2.2e-16 | 28728 | 21256 | 7472 |
| European | Full Cohort | Discontinuation vs Continuation | Education_level | Income100-150k | 0.541 | 0.491 | 0.596 | <2.2e-16 | <2.2e-16 | 28728 | 21256 | 7472 |
| European | Full Cohort | Discontinuation vs Continuation | Education_level | Income150-200k | 0.527 | 0.466 | 0.596 | <2.2e-16 | <2.2e-16 | 28728 | 21256 | 7472 |
| European | Full Cohort | Discontinuation vs Continuation | Education_level | Income25-35k | 0.742 | 0.666 | 0.828 | 9.69e-08 | 1.36e-06 | 28728 | 21256 | 7472 |
| European | Full Cohort | Discontinuation vs Continuation | Education_level | Income35-50k | 0.662 | 0.597 | 0.735 | 6.43e-15 | 9.00e-14 | 28728 | 21256 | 7472 |
| European | Full Cohort | Discontinuation vs Continuation | Education_level | Income50-75k | 0.673 | 0.614 | 0.738 | <2.2e-16 | 5.57e-16 | 28728 | 21256 | 7472 |
| European | Full Cohort | Discontinuation vs Continuation | Education_level | Income75-100k | 0.576 | 0.521 | 0.636 | <2.2e-16 | <2.2e-16 | 28728 | 21256 | 7472 |
| European | Full Cohort | Discontinuation vs Continuation | Education_level | Education_levelCollege Postgraduate | 0.588 | 0.482 | 0.717 | 1.66e-07 | 2.32e-06 | 28728 | 21256 | 7472 |
| European | Full Cohort | Discontinuation vs Continuation | Education_level | Education_levelCollege Undergraduate | 0.666 | 0.550 | 0.807 | 3.23e-05 | 4.53e-04 | 28728 | 21256 | 7472 |
| European | Full Cohort | Discontinuation vs Continuation | Education_level | Education_levelTwelve or GED | 0.793 | 0.650 | 0.968 | 2.29e-02 | 3.20e-01 | 28728 | 21256 | 7472 |
| European | Full Cohort | Switching vs Continuation | Education_level | (Intercept) | 1.169 | 0.989 | 1.383 | 6.75e-02 | 9.44e-01 | 35092 | 21256 | 13836 |
| European | Full Cohort | Switching vs Continuation | Education_level | age_at_first_AD | 0.708 | 0.692 | 0.724 | <2.2e-16 | <2.2e-16 | 35092 | 21256 | 13836 |
| European | Full Cohort | Switching vs Continuation | Education_level | sex_at_birthMale | 1.049 | 0.999 | 1.102 | 5.35e-02 | 7.48e-01 | 35092 | 21256 | 13836 |
| European | Full Cohort | Switching vs Continuation | Education_level | Income>200k | 0.404 | 0.366 | 0.447 | <2.2e-16 | <2.2e-16 | 35092 | 21256 | 13836 |
| European | Full Cohort | Switching vs Continuation | Education_level | Income100-150k | 0.557 | 0.514 | 0.603 | <2.2e-16 | <2.2e-16 | 35092 | 21256 | 13836 |
| European | Full Cohort | Switching vs Continuation | Education_level | Income150-200k | 0.444 | 0.400 | 0.492 | <2.2e-16 | <2.2e-16 | 35092 | 21256 | 13836 |
| European | Full Cohort | Switching vs Continuation | Education_level | Income25-35k | 0.838 | 0.766 | 0.918 | 1.40e-04 | 1.96e-03 | 35092 | 21256 | 13836 |
| European | Full Cohort | Switching vs Continuation | Education_level | Income35-50k | 0.783 | 0.719 | 0.852 | 1.45e-08 | 2.04e-07 | 35092 | 21256 | 13836 |
| European | Full Cohort | Switching vs Continuation | Education_level | Income50-75k | 0.723 | 0.669 | 0.780 | <2.2e-16 | 1.51e-15 | 35092 | 21256 | 13836 |
| European | Full Cohort | Switching vs Continuation | Education_level | Income75-100k | 0.606 | 0.558 | 0.658 | <2.2e-16 | <2.2e-16 | 35092 | 21256 | 13836 |
| European | Full Cohort | Switching vs Continuation | Education_level | Education_levelCollege Postgraduate | 0.736 | 0.617 | 0.879 | 7.11e-04 | 9.95e-03 | 35092 | 21256 | 13836 |
| European | Full Cohort | Switching vs Continuation | Education_level | Education_levelCollege Undergraduate | 0.810 | 0.682 | 0.962 | 1.60e-02 | 2.25e-01 | 35092 | 21256 | 13836 |
| European | Full Cohort | Switching vs Continuation | Education_level | Education_levelTwelve or GED | 0.955 | 0.799 | 1.141 | 6.12e-01 | 1.00e+00 | 35092 | 21256 | 13836 |
| European | Full Cohort | Discontinuation vs Continuation | Income | (Intercept) | 0.761 | 0.631 | 0.917 | 4.10e-03 | 5.74e-02 | 28728 | 21256 | 7472 |
| European | Full Cohort | Discontinuation vs Continuation | Income | age_at_first_AD | 1.089 | 1.060 | 1.118 | 3.29e-10 | 4.60e-09 | 28728 | 21256 | 7472 |
| European | Full Cohort | Discontinuation vs Continuation | Income | sex_at_birthMale | 1.118 | 1.056 | 1.184 | 1.33e-04 | 1.86e-03 | 28728 | 21256 | 7472 |
| European | Full Cohort | Discontinuation vs Continuation | Income | Income>200k | 0.595 | 0.531 | 0.666 | <2.2e-16 | <2.2e-16 | 28728 | 21256 | 7472 |
| European | Full Cohort | Discontinuation vs Continuation | Income | Income100-150k | 0.541 | 0.491 | 0.596 | <2.2e-16 | <2.2e-16 | 28728 | 21256 | 7472 |
| European | Full Cohort | Discontinuation vs Continuation | Income | Income150-200k | 0.527 | 0.466 | 0.596 | <2.2e-16 | <2.2e-16 | 28728 | 21256 | 7472 |
| European | Full Cohort | Discontinuation vs Continuation | Income | Income25-35k | 0.742 | 0.666 | 0.828 | 9.69e-08 | 1.36e-06 | 28728 | 21256 | 7472 |
| European | Full Cohort | Discontinuation vs Continuation | Income | Income35-50k | 0.662 | 0.597 | 0.735 | 6.43e-15 | 9.00e-14 | 28728 | 21256 | 7472 |
| European | Full Cohort | Discontinuation vs Continuation | Income | Income50-75k | 0.673 | 0.614 | 0.738 | <2.2e-16 | 5.57e-16 | 28728 | 21256 | 7472 |
| European | Full Cohort | Discontinuation vs Continuation | Income | Income75-100k | 0.576 | 0.521 | 0.636 | <2.2e-16 | <2.2e-16 | 28728 | 21256 | 7472 |
| European | Full Cohort | Discontinuation vs Continuation | Income | Education_levelCollege Postgraduate | 0.588 | 0.482 | 0.717 | 1.66e-07 | 2.32e-06 | 28728 | 21256 | 7472 |
| European | Full Cohort | Discontinuation vs Continuation | Income | Education_levelCollege Undergraduate | 0.666 | 0.550 | 0.807 | 3.23e-05 | 4.53e-04 | 28728 | 21256 | 7472 |
| European | Full Cohort | Discontinuation vs Continuation | Income | Education_levelTwelve or GED | 0.793 | 0.650 | 0.968 | 2.29e-02 | 3.20e-01 | 28728 | 21256 | 7472 |
| European | Full Cohort | Switching vs Continuation | Income | (Intercept) | 1.169 | 0.989 | 1.383 | 6.75e-02 | 9.44e-01 | 35092 | 21256 | 13836 |
| European | Full Cohort | Switching vs Continuation | Income | age_at_first_AD | 0.708 | 0.692 | 0.724 | <2.2e-16 | <2.2e-16 | 35092 | 21256 | 13836 |
| European | Full Cohort | Switching vs Continuation | Income | sex_at_birthMale | 1.049 | 0.999 | 1.102 | 5.35e-02 | 7.48e-01 | 35092 | 21256 | 13836 |
| European | Full Cohort | Switching vs Continuation | Income | Income>200k | 0.404 | 0.366 | 0.447 | <2.2e-16 | <2.2e-16 | 35092 | 21256 | 13836 |
| European | Full Cohort | Switching vs Continuation | Income | Income100-150k | 0.557 | 0.514 | 0.603 | <2.2e-16 | <2.2e-16 | 35092 | 21256 | 13836 |
| European | Full Cohort | Switching vs Continuation | Income | Income150-200k | 0.444 | 0.400 | 0.492 | <2.2e-16 | <2.2e-16 | 35092 | 21256 | 13836 |
| European | Full Cohort | Switching vs Continuation | Income | Income25-35k | 0.838 | 0.766 | 0.918 | 1.40e-04 | 1.96e-03 | 35092 | 21256 | 13836 |
| European | Full Cohort | Switching vs Continuation | Income | Income35-50k | 0.783 | 0.719 | 0.852 | 1.45e-08 | 2.04e-07 | 35092 | 21256 | 13836 |
| European | Full Cohort | Switching vs Continuation | Income | Income50-75k | 0.723 | 0.669 | 0.780 | <2.2e-16 | 1.51e-15 | 35092 | 21256 | 13836 |
| European | Full Cohort | Switching vs Continuation | Income | Income75-100k | 0.606 | 0.558 | 0.658 | <2.2e-16 | <2.2e-16 | 35092 | 21256 | 13836 |
| European | Full Cohort | Switching vs Continuation | Income | Education_levelCollege Postgraduate | 0.736 | 0.617 | 0.879 | 7.11e-04 | 9.95e-03 | 35092 | 21256 | 13836 |
| European | Full Cohort | Switching vs Continuation | Income | Education_levelCollege Undergraduate | 0.810 | 0.682 | 0.962 | 1.60e-02 | 2.25e-01 | 35092 | 21256 | 13836 |
| European | Full Cohort | Switching vs Continuation | Income | Education_levelTwelve or GED | 0.955 | 0.799 | 1.141 | 6.12e-01 | 1.00e+00 | 35092 | 21256 | 13836 |
| European | Full Cohort | Discontinuation vs Continuation | Insomnia | (Intercept) | 0.789 | 0.654 | 0.952 | 1.33e-02 | 1.86e-01 | 28728 | 21256 | 7472 |
| European | Full Cohort | Discontinuation vs Continuation | Insomnia | age_at_first_AD | 1.097 | 1.068 | 1.127 | 8.58e-12 | 1.20e-10 | 28728 | 21256 | 7472 |
| European | Full Cohort | Discontinuation vs Continuation | Insomnia | sex_at_birthMale | 1.122 | 1.060 | 1.188 | 7.61e-05 | 1.07e-03 | 28728 | 21256 | 7472 |
| European | Full Cohort | Discontinuation vs Continuation | Insomnia | Income>200k | 0.594 | 0.530 | 0.664 | <2.2e-16 | <2.2e-16 | 28728 | 21256 | 7472 |
| European | Full Cohort | Discontinuation vs Continuation | Insomnia | Income100-150k | 0.539 | 0.490 | 0.594 | <2.2e-16 | <2.2e-16 | 28728 | 21256 | 7472 |
| European | Full Cohort | Discontinuation vs Continuation | Insomnia | Income150-200k | 0.526 | 0.465 | 0.595 | <2.2e-16 | <2.2e-16 | 28728 | 21256 | 7472 |
| European | Full Cohort | Discontinuation vs Continuation | Insomnia | Income25-35k | 0.741 | 0.664 | 0.826 | 7.76e-08 | 1.09e-06 | 28728 | 21256 | 7472 |

| | | | | | | | | | | | | |
|---|---|---|---|---|---|---|---|---|---|---|---|---|
| European | Full Cohort | Discontinuation vs Continuation | Insomnia | Income35-50k | 0.661 | 0.596 | 0.734 | 5.31e-15 | 7.43e-14 | 28728 | 21256 | 7472 |
| European | Full Cohort | Discontinuation vs Continuation | Insomnia | Income50-75k | 0.673 | 0.613 | 0.738 | <2.2e-16 | 4.86e-16 | 28728 | 21256 | 7472 |
| European | Full Cohort | Discontinuation vs Continuation | Insomnia | Income75-100k | 0.575 | 0.520 | 0.635 | <2.2e-16 | <2.2e-16 | 28728 | 21256 | 7472 |
| European | Full Cohort | Discontinuation vs Continuation | Insomnia | Education_levelCollege Postgraduate | 0.586 | 0.480 | 0.715 | 1.43e-07 | 2.00e-06 | 28728 | 21256 | 7472 |
| European | Full Cohort | Discontinuation vs Continuation | Insomnia | Education_levelCollege Undergraduate | 0.664 | 0.548 | 0.805 | 2.96e-05 | 4.15e-04 | 28728 | 21256 | 7472 |
| European | Full Cohort | Discontinuation vs Continuation | Insomnia | Education_levelTwelve or GED | 0.792 | 0.649 | 0.967 | 2.22e-02 | 3.11e-01 | 28728 | 21256 | 7472 |
| European | Full Cohort | Discontinuation vs Continuation | Insomnia | Insomnia | 0.829 | 0.774 | 0.888 | 9.71e-08 | 1.36e-06 | 28728 | 21256 | 7472 |
| European | Full Cohort | Switching vs Continuation | Insomnia | (Intercept) | 0.886 | 0.746 | 1.052 | 1.66e-01 | 1.00e+00 | 35092 | 21256 | 13836 |
| European | Full Cohort | Switching vs Continuation | Insomnia | age_at_first_AD | 0.680 | 0.665 | 0.696 | <2.2e-16 | <2.2e-16 | 35092 | 21256 | 13836 |
| European | Full Cohort | Switching vs Continuation | Insomnia | sex_at_birthMale | 1.020 | 0.970 | 1.071 | 4.42e-01 | 1.00e+00 | 35092 | 21256 | 13836 |
| European | Full Cohort | Switching vs Continuation | Insomnia | Income>200k | 0.414 | 0.374 | 0.459 | <2.2e-16 | <2.2e-16 | 35092 | 21256 | 13836 |
| European | Full Cohort | Switching vs Continuation | Insomnia | Income100-150k | 0.573 | 0.529 | 0.622 | <2.2e-16 | <2.2e-16 | 35092 | 21256 | 13836 |
| European | Full Cohort | Switching vs Continuation | Insomnia | Income150-200k | 0.458 | 0.412 | 0.509 | <2.2e-16 | <2.2e-16 | 35092 | 21256 | 13836 |
| European | Full Cohort | Switching vs Continuation | Insomnia | Income25-35k | 0.855 | 0.779 | 0.938 | 8.95e-04 | 1.25e-02 | 35092 | 21256 | 13836 |
| European | Full Cohort | Switching vs Continuation | Insomnia | Income35-50k | 0.794 | 0.728 | 0.865 | 1.53e-07 | 2.15e-06 | 35092 | 21256 | 13836 |
| European | Full Cohort | Switching vs Continuation | Insomnia | Income50-75k | 0.738 | 0.682 | 0.798 | 2.24e-14 | 3.14e-13 | 35092 | 21256 | 13836 |
| European | Full Cohort | Switching vs Continuation | Insomnia | Income75-100k | 0.620 | 0.570 | 0.674 | <2.2e-16 | <2.2e-16 | 35092 | 21256 | 13836 |
| European | Full Cohort | Switching vs Continuation | Insomnia | Education_levelCollege Postgraduate | 0.761 | 0.636 | 0.912 | 3.09e-03 | 4.33e-02 | 35092 | 21256 | 13836 |
| European | Full Cohort | Switching vs Continuation | Insomnia | Education_levelCollege Undergraduate | 0.834 | 0.700 | 0.994 | 4.21e-02 | 5.89e-01 | 35092 | 21256 | 13836 |
| European | Full Cohort | Switching vs Continuation | Insomnia | Education_levelTwelve or GED | 0.971 | 0.810 | 1.164 | 7.48e-01 | 1.00e+00 | 35092 | 21256 | 13836 |
| European | Full Cohort | Switching vs Continuation | Insomnia | Insomnia | 2.355 | 2.240 | 2.476 | <2.2e-16 | <2.2e-16 | 35092 | 21256 | 13836 |
| European | Full Cohort | Discontinuation vs Continuation | MDD CIDI-sf | (Intercept) | 0.265 | 0.107 | 0.657 | 4.14e-03 | 5.80e-02 | 5717 | 4368 | 1349 |
| European | Full Cohort | Discontinuation vs Continuation | MDD CIDI-sf | age_at_first_AD | 1.095 | 1.024 | 1.172 | 8.51e-03 | 1.19e-01 | 5717 | 4368 | 1349 |
| European | Full Cohort | Discontinuation vs Continuation | MDD CIDI-sf | sex_at_birthMale | 1.134 | 0.990 | 1.300 | 6.91e-02 | 9.68e-01 | 5717 | 4368 | 1349 |
| European | Full Cohort | Discontinuation vs Continuation | MDD CIDI-sf | Income>200k | 0.734 | 0.558 | 0.966 | 2.75e-02 | 3.85e-01 | 5717 | 4368 | 1349 |
| European | Full Cohort | Discontinuation vs Continuation | MDD CIDI-sf | Income100-150k | 0.638 | 0.500 | 0.814 | 3.04e-04 | 4.25e-03 | 5717 | 4368 | 1349 |
| European | Full Cohort | Discontinuation vs Continuation | MDD CIDI-sf | Income150-200k | 0.747 | 0.561 | 0.996 | 4.66e-02 | 6.53e-01 | 5717 | 4368 | 1349 |
| European | Full Cohort | Discontinuation vs Continuation | MDD CIDI-sf | Income25-35k | 0.705 | 0.525 | 0.948 | 2.05e-02 | 2.86e-01 | 5717 | 4368 | 1349 |
| European | Full Cohort | Discontinuation vs Continuation | MDD CIDI-sf | Income35-50k | 0.781 | 0.599 | 1.019 | 6.81e-02 | 9.53e-01 | 5717 | 4368 | 1349 |
| European | Full Cohort | Discontinuation vs Continuation | MDD CIDI-sf | Income50-75k | 0.854 | 0.675 | 1.080 | 1.86e-01 | 1.00e+00 | 5717 | 4368 | 1349 |
| European | Full Cohort | Discontinuation vs Continuation | MDD CIDI-sf | Income75-100k | 0.684 | 0.532 | 0.879 | 3.01e-03 | 4.21e-02 | 5717 | 4368 | 1349 |
| European | Full Cohort | Discontinuation vs Continuation | MDD CIDI-sf | Education_levelCollege Postgraduate | 1.174 | 0.471 | 2.925 | 7.31e-01 | 1.00e+00 | 5717 | 4368 | 1349 |
| European | Full Cohort | Discontinuation vs Continuation | MDD CIDI-sf | Education_levelCollege Undergraduate | 1.531 | 0.619 | 3.788 | 3.56e-01 | 1.00e+00 | 5717 | 4368 | 1349 |
| European | Full Cohort | Discontinuation vs Continuation | MDD CIDI-sf | Education_levelTwelve or GED | 1.405 | 0.556 | 3.554 | 4.73e-01 | 1.00e+00 | 5717 | 4368 | 1349 |
| European | Full Cohort | Discontinuation vs Continuation | MDD CIDI-sf | MDD CIDI-sf | 1.081 | 0.951 | 1.229 | 2.35e-01 | 1.00e+00 | 5717 | 4368 | 1349 |
| European | Full Cohort | Switching vs Continuation | MDD CIDI-sf | (Intercept) | 0.352 | 0.181 | 0.684 | 2.06e-03 | 2.89e-02 | 7163 | 4368 | 2795 |
| European | Full Cohort | Switching vs Continuation | MDD CIDI-sf | age_at_first_AD | 0.664 | 0.628 | 0.702 | <2.2e-16 | <2.2e-16 | 7163 | 4368 | 2795 |
| European | Full Cohort | Switching vs Continuation | MDD CIDI-sf | sex_at_birthMale | 1.171 | 1.045 | 1.314 | 6.83e-03 | 9.56e-02 | 7163 | 4368 | 2795 |
| European | Full Cohort | Switching vs Continuation | MDD CIDI-sf | Income>200k | 0.447 | 0.354 | 0.565 | 1.37e-11 | 1.92e-10 | 7163 | 4368 | 2795 |
| European | Full Cohort | Switching vs Continuation | MDD CIDI-sf | Income100-150k | 0.566 | 0.468 | 0.686 | 6.07e-09 | 8.49e-08 | 7163 | 4368 | 2795 |
| European | Full Cohort | Switching vs Continuation | MDD CIDI-sf | Income150-200k | 0.478 | 0.376 | 0.608 | 1.76e-09 | 2.46e-08 | 7163 | 4368 | 2795 |
| European | Full Cohort | Switching vs Continuation | MDD CIDI-sf | Income25-35k | 0.797 | 0.636 | 1.000 | 5.04e-02 | 7.05e-01 | 7163 | 4368 | 2795 |
| European | Full Cohort | Switching vs Continuation | MDD CIDI-sf | Income35-50k | 0.828 | 0.674 | 1.017 | 7.20e-02 | 1.00e+00 | 7163 | 4368 | 2795 |
| European | Full Cohort | Switching vs Continuation | MDD CIDI-sf | Income50-75k | 0.736 | 0.610 | 0.888 | 1.35e-03 | 1.89e-02 | 7163 | 4368 | 2795 |
| European | Full Cohort | Switching vs Continuation | MDD CIDI-sf | Income75-100k | 0.668 | 0.549 | 0.812 | 5.46e-05 | 7.64e-04 | 7163 | 4368 | 2795 |
| European | Full Cohort | Switching vs Continuation | MDD CIDI-sf | Education_levelCollege Postgraduate | 1.377 | 0.707 | 2.682 | 3.48e-01 | 1.00e+00 | 7163 | 4368 | 2795 |
| European | Full Cohort | Switching vs Continuation | MDD CIDI-sf | Education_levelCollege Undergraduate | 1.555 | 0.803 | 3.012 | 1.90e-01 | 1.00e+00 | 7163 | 4368 | 2795 |
| European | Full Cohort | Switching vs Continuation | MDD CIDI-sf | Education_levelTwelve or GED | 1.799 | 0.912 | 3.548 | 9.01e-02 | 1.00e+00 | 7163 | 4368 | 2795 |
| European | Full Cohort | Switching vs Continuation | MDD CIDI-sf | MDD CIDI-sf | 2.316 | 2.073 | 2.588 | <2.2e-16 | <2.2e-16 | 7163 | 4368 | 2795 |
| European | Full Cohort | Discontinuation vs Continuation | MDD_symptom_count | (Intercept) | 0.391 | 0.189 | 0.809 | 1.14e-02 | 1.59e-01 | 7190 | 5509 | 1681 |
| European | Full Cohort | Discontinuation vs Continuation | MDD_symptom_count | age_at_first_AD | 1.117 | 1.050 | 1.188 | 4.11e-04 | 5.76e-03 | 7190 | 5509 | 1681 |
| European | Full Cohort | Discontinuation vs Continuation | MDD_symptom_count | sex_at_birthMale | 1.122 | 0.993 | 1.268 | 6.48e-02 | 9.07e-01 | 7190 | 5509 | 1681 |
| European | Full Cohort | Discontinuation vs Continuation | MDD_symptom_count | Income>200k | 0.754 | 0.589 | 0.966 | 2.53e-02 | 3.55e-01 | 7190 | 5509 | 1681 |
| European | Full Cohort | Discontinuation vs Continuation | MDD_symptom_count | Income100-150k | 0.660 | 0.530 | 0.821 | 1.96e-04 | 2.74e-03 | 7190 | 5509 | 1681 |
| European | Full Cohort | Discontinuation vs Continuation | MDD_symptom_count | Income150-200k | 0.704 | 0.543 | 0.912 | 7.91e-03 | 1.11e-01 | 7190 | 5509 | 1681 |
| European | Full Cohort | Discontinuation vs Continuation | MDD_symptom_count | Income25-35k | 0.705 | 0.542 | 0.918 | 9.34e-03 | 1.31e-01 | 7190 | 5509 | 1681 |
| European | Full Cohort | Discontinuation vs Continuation | MDD_symptom_count | Income35-50k | 0.788 | 0.622 | 1.000 | 4.96e-02 | 6.94e-01 | 7190 | 5509 | 1681 |
| European | Full Cohort | Discontinuation vs Continuation | MDD_symptom_count | Income50-75k | 0.843 | 0.682 | 1.041 | 1.13e-01 | 1.00e+00 | 7190 | 5509 | 1681 |
| European | Full Cohort | Discontinuation vs Continuation | MDD_symptom_count | Income75-100k | 0.682 | 0.545 | 0.854 | 8.54e-04 | 1.20e-02 | 7190 | 5509 | 1681 |
| European | Full Cohort | Discontinuation vs Continuation | MDD_symptom_count | Education_levelCollege Postgraduate | 0.852 | 0.407 | 1.781 | 6.70e-01 | 1.00e+00 | 7190 | 5509 | 1681 |
| European | Full Cohort | Discontinuation vs Continuation | MDD_symptom_count | Education_levelCollege Undergraduate | 1.045 | 0.503 | 2.170 | 9.06e-01 | 1.00e+00 | 7190 | 5509 | 1681 |
| European | Full Cohort | Discontinuation vs Continuation | MDD_symptom_count | Education_levelTwelve or GED | 0.962 | 0.453 | 2.042 | 9.20e-01 | 1.00e+00 | 7190 | 5509 | 1681 |
| European | Full Cohort | Discontinuation vs Continuation | MDD_symptom_count | MDD_symptom_count | 1.035 | 0.978 | 1.095 | 2.31e-01 | 1.00e+00 | 7190 | 5509 | 1681 |
| European | Full Cohort | Switching vs Continuation | MDD_symptom_count | (Intercept) | 0.731 | 0.416 | 1.285 | 2.76e-01 | 1.00e+00 | 8962 | 5509 | 3453 |
| European | Full Cohort | Switching vs Continuation | MDD_symptom_count | age_at_first_AD | 0.665 | 0.632 | 0.699 | <2.2e-16 | <2.2e-16 | 8962 | 5509 | 3453 |
| European | Full Cohort | Switching vs Continuation | MDD_symptom_count | sex_at_birthMale | 1.214 | 1.096 | 1.345 | 2.08e-04 | 2.91e-03 | 8962 | 5509 | 3453 |

| | | | | | | | | | | | | |
|---|---|---|---|---|---|---|---|---|---|---|---|---|
| European | Full Cohort | Switching vs Continuation | MDD_symptom_count | Income>200k | 0.490 | 0.397 | 0.604 | 2.52e-11 | 3.53e-10 | 8962 | 5509 | 3453 |
| European | Full Cohort | Switching vs Continuation | MDD_symptom_count | Income100-150k | 0.610 | 0.513 | 0.725 | 2.08e-08 | 2.91e-07 | 8962 | 5509 | 3453 |
| European | Full Cohort | Switching vs Continuation | MDD_symptom_count | Income150-200k | 0.495 | 0.399 | 0.614 | 1.57e-10 | 2.20e-09 | 8962 | 5509 | 3453 |
| European | Full Cohort | Switching vs Continuation | MDD_symptom_count | Income25-35k | 0.792 | 0.647 | 0.971 | 2.48e-02 | 3.47e-01 | 8962 | 5509 | 3453 |
| European | Full Cohort | Switching vs Continuation | MDD_symptom_count | Income35-50k | 0.870 | 0.723 | 1.046 | 1.37e-01 | 1.00e+00 | 8962 | 5509 | 3453 |
| European | Full Cohort | Switching vs Continuation | MDD_symptom_count | Income50-75k | 0.764 | 0.645 | 0.905 | 1.79e-03 | 2.51e-02 | 8962 | 5509 | 3453 |
| European | Full Cohort | Switching vs Continuation | MDD_symptom_count | Income75-100k | 0.704 | 0.590 | 0.839 | 9.12e-05 | 1.28e-03 | 8962 | 5509 | 3453 |
| European | Full Cohort | Switching vs Continuation | MDD_symptom_count | Education_levelCollege Postgraduate | 1.051 | 0.593 | 1.863 | 8.65e-01 | 1.00e+00 | 8962 | 5509 | 3453 |
| European | Full Cohort | Switching vs Continuation | MDD_symptom_count | Education_levelCollege Undergraduate | 1.179 | 0.669 | 2.077 | 5.70e-01 | 1.00e+00 | 8962 | 5509 | 3453 |
| European | Full Cohort | Switching vs Continuation | MDD_symptom_count | Education_levelTwelve or GED | 1.446 | 0.807 | 2.589 | 2.15e-01 | 1.00e+00 | 8962 | 5509 | 3453 |
| European | Full Cohort | Switching vs Continuation | MDD_symptom_count | MDD_symptom_count | 1.476 | 1.405 | 1.550 | <2.2e-16 | <2.2e-16 | 8962 | 5509 | 3453 |
| European | Full Cohort | Discontinuation vs Continuation | Migraine | (Intercept) | 0.803 | 0.666 | 0.969 | 2.20e-02 | 3.08e-01 | 28728 | 21256 | 7472 |
| European | Full Cohort | Discontinuation vs Continuation | Migraine | age_at_first_AD | 1.082 | 1.054 | 1.111 | 5.79e-09 | 8.11e-08 | 28728 | 21256 | 7472 |
| European | Full Cohort | Discontinuation vs Continuation | Migraine | sex_at_birthMale | 1.093 | 1.032 | 1.158 | 2.42e-03 | 3.39e-02 | 28728 | 21256 | 7472 |
| European | Full Cohort | Discontinuation vs Continuation | Migraine | Income>200k | 0.595 | 0.531 | 0.666 | <2.2e-16 | <2.2e-16 | 28728 | 21256 | 7472 |
| European | Full Cohort | Discontinuation vs Continuation | Migraine | Income100-150k | 0.543 | 0.493 | 0.598 | <2.2e-16 | <2.2e-16 | 28728 | 21256 | 7472 |
| European | Full Cohort | Discontinuation vs Continuation | Migraine | Income150-200k | 0.529 | 0.467 | 0.598 | <2.2e-16 | <2.2e-16 | 28728 | 21256 | 7472 |
| European | Full Cohort | Discontinuation vs Continuation | Migraine | Income25-35k | 0.743 | 0.666 | 0.829 | 1.09e-07 | 1.52e-06 | 28728 | 21256 | 7472 |
| European | Full Cohort | Discontinuation vs Continuation | Migraine | Income35-50k | 0.665 | 0.599 | 0.737 | 1.19e-14 | 1.67e-13 | 28728 | 21256 | 7472 |
| European | Full Cohort | Discontinuation vs Continuation | Migraine | Income50-75k | 0.675 | 0.616 | 0.741 | <2.2e-16 | 1.03e-15 | 28728 | 21256 | 7472 |
| European | Full Cohort | Discontinuation vs Continuation | Migraine | Income75-100k | 0.577 | 0.522 | 0.637 | <2.2e-16 | <2.2e-16 | 28728 | 21256 | 7472 |
| European | Full Cohort | Discontinuation vs Continuation | Migraine | Education_levelCollege Postgraduate | 0.593 | 0.486 | 0.724 | 2.78e-07 | 3.89e-06 | 28728 | 21256 | 7472 |
| European | Full Cohort | Discontinuation vs Continuation | Migraine | Education_levelCollege Undergraduate | 0.671 | 0.553 | 0.812 | 4.51e-05 | 6.32e-04 | 28728 | 21256 | 7472 |
| European | Full Cohort | Discontinuation vs Continuation | Migraine | Education_levelTwelve or GED | 0.797 | 0.653 | 0.973 | 2.59e-02 | 3.62e-01 | 28728 | 21256 | 7472 |
| European | Full Cohort | Discontinuation vs Continuation | Migraine | Migraine | 0.799 | 0.750 | 0.851 | 3.60e-12 | 5.04e-11 | 28728 | 21256 | 7472 |
| European | Full Cohort | Switching vs Continuation | Migraine | (Intercept) | 0.954 | 0.805 | 1.130 | 5.86e-01 | 1.00e+00 | 35092 | 21256 | 13836 |
| European | Full Cohort | Switching vs Continuation | Migraine | age_at_first_AD | 0.723 | 0.707 | 0.740 | <2.2e-16 | <2.2e-16 | 35092 | 21256 | 13836 |
| European | Full Cohort | Switching vs Continuation | Migraine | sex_at_birthMale | 1.136 | 1.081 | 1.194 | 4.48e-07 | 6.28e-06 | 35092 | 21256 | 13836 |
| European | Full Cohort | Switching vs Continuation | Migraine | Income>200k | 0.410 | 0.371 | 0.454 | <2.2e-16 | <2.2e-16 | 35092 | 21256 | 13836 |
| European | Full Cohort | Switching vs Continuation | Migraine | Income100-150k | 0.559 | 0.515 | 0.606 | <2.2e-16 | <2.2e-16 | 35092 | 21256 | 13836 |
| European | Full Cohort | Switching vs Continuation | Migraine | Income150-200k | 0.448 | 0.404 | 0.498 | <2.2e-16 | <2.2e-16 | 35092 | 21256 | 13836 |
| European | Full Cohort | Switching vs Continuation | Migraine | Income25-35k | 0.836 | 0.763 | 0.916 | 1.29e-04 | 1.80e-03 | 35092 | 21256 | 13836 |
| European | Full Cohort | Switching vs Continuation | Migraine | Income35-50k | 0.782 | 0.718 | 0.852 | 1.89e-08 | 2.65e-07 | 35092 | 21256 | 13836 |
| European | Full Cohort | Switching vs Continuation | Migraine | Income50-75k | 0.725 | 0.671 | 0.784 | 4.85e-16 | 6.78e-15 | 35092 | 21256 | 13836 |
| European | Full Cohort | Switching vs Continuation | Migraine | Income75-100k | 0.607 | 0.558 | 0.659 | <2.2e-16 | <2.2e-16 | 35092 | 21256 | 13836 |
| European | Full Cohort | Switching vs Continuation | Migraine | Education_levelCollege Postgraduate | 0.707 | 0.591 | 0.846 | 1.48e-04 | 2.07e-03 | 35092 | 21256 | 13836 |
| European | Full Cohort | Switching vs Continuation | Migraine | Education_levelCollege Undergraduate | 0.779 | 0.655 | 0.927 | 4.84e-03 | 6.78e-02 | 35092 | 21256 | 13836 |
| European | Full Cohort | Switching vs Continuation | Migraine | Education_levelTwelve or GED | 0.919 | 0.768 | 1.100 | 3.58e-01 | 1.00e+00 | 35092 | 21256 | 13836 |
| European | Full Cohort | Switching vs Continuation | Migraine | Migraine | 1.869 | 1.784 | 1.959 | <2.2e-16 | <2.2e-16 | 35092 | 21256 | 13836 |
| European | Full Cohort | Discontinuation vs Continuation | Recorded Anxiety | (Intercept) | 0.981 | 0.810 | 1.187 | 8.40e-01 | 1.00e+00 | 28728 | 21256 | 7472 |
| European | Full Cohort | Discontinuation vs Continuation | Recorded Anxiety | age_at_first_AD | 1.055 | 1.027 | 1.084 | 9.34e-05 | 1.31e-03 | 28728 | 21256 | 7472 |
| European | Full Cohort | Discontinuation vs Continuation | Recorded Anxiety | sex_at_birthMale | 1.095 | 1.034 | 1.160 | 1.98e-03 | 2.77e-02 | 28728 | 21256 | 7472 |
| European | Full Cohort | Discontinuation vs Continuation | Recorded Anxiety | Income>200k | 0.574 | 0.512 | 0.643 | <2.2e-16 | <2.2e-16 | 28728 | 21256 | 7472 |
| European | Full Cohort | Discontinuation vs Continuation | Recorded Anxiety | Income100-150k | 0.531 | 0.482 | 0.585 | <2.2e-16 | <2.2e-16 | 28728 | 21256 | 7472 |
| European | Full Cohort | Discontinuation vs Continuation | Recorded Anxiety | Income150-200k | 0.514 | 0.454 | 0.582 | <2.2e-16 | <2.2e-16 | 28728 | 21256 | 7472 |
| European | Full Cohort | Discontinuation vs Continuation | Recorded Anxiety | Income25-35k | 0.744 | 0.666 | 0.830 | 1.31e-07 | 1.83e-06 | 28728 | 21256 | 7472 |
| European | Full Cohort | Discontinuation vs Continuation | Recorded Anxiety | Income35-50k | 0.657 | 0.592 | 0.729 | 2.49e-15 | 3.49e-14 | 28728 | 21256 | 7472 |
| European | Full Cohort | Discontinuation vs Continuation | Recorded Anxiety | Income50-75k | 0.667 | 0.608 | 0.732 | <2.2e-16 | <2.2e-16 | 28728 | 21256 | 7472 |
| European | Full Cohort | Discontinuation vs Continuation | Recorded Anxiety | Income75-100k | 0.566 | 0.512 | 0.626 | <2.2e-16 | <2.2e-16 | 28728 | 21256 | 7472 |
| European | Full Cohort | Discontinuation vs Continuation | Recorded Anxiety | Education_levelCollege Postgraduate | 0.572 | 0.468 | 0.699 | 4.33e-08 | 6.06e-07 | 28728 | 21256 | 7472 |
| European | Full Cohort | Discontinuation vs Continuation | Recorded Anxiety | Education_levelCollege Undergraduate | 0.650 | 0.536 | 0.788 | 1.14e-05 | 1.60e-04 | 28728 | 21256 | 7472 |
| European | Full Cohort | Discontinuation vs Continuation | Recorded Anxiety | Education_levelTwelve or GED | 0.774 | 0.633 | 0.946 | 1.21e-02 | 1.70e-01 | 28728 | 21256 | 7472 |
| European | Full Cohort | Discontinuation vs Continuation | Recorded Anxiety | Recorded Anxiety | 0.668 | 0.633 | 0.705 | <2.2e-16 | <2.2e-16 | 28728 | 21256 | 7472 |
| European | Full Cohort | Switching vs Continuation | Recorded Anxiety | (Intercept) | 0.573 | 0.481 | 0.682 | 4.14e-10 | 5.80e-09 | 35092 | 21256 | 13836 |
| European | Full Cohort | Switching vs Continuation | Recorded Anxiety | age_at_first_AD | 0.763 | 0.746 | 0.781 | <2.2e-16 | <2.2e-16 | 35092 | 21256 | 13836 |
| European | Full Cohort | Switching vs Continuation | Recorded Anxiety | sex_at_birthMale | 1.086 | 1.033 | 1.141 | 1.20e-03 | 1.68e-02 | 35092 | 21256 | 13836 |
| European | Full Cohort | Switching vs Continuation | Recorded Anxiety | Income>200k | 0.427 | 0.385 | 0.473 | <2.2e-16 | <2.2e-16 | 35092 | 21256 | 13836 |
| European | Full Cohort | Switching vs Continuation | Recorded Anxiety | Income100-150k | 0.565 | 0.521 | 0.613 | <2.2e-16 | <2.2e-16 | 35092 | 21256 | 13836 |
| European | Full Cohort | Switching vs Continuation | Recorded Anxiety | Income150-200k | 0.457 | 0.411 | 0.508 | <2.2e-16 | <2.2e-16 | 35092 | 21256 | 13836 |
| European | Full Cohort | Switching vs Continuation | Recorded Anxiety | Income25-35k | 0.826 | 0.753 | 0.906 | 5.37e-05 | 7.52e-04 | 35092 | 21256 | 13836 |
| European | Full Cohort | Switching vs Continuation | Recorded Anxiety | Income35-50k | 0.782 | 0.718 | 0.853 | 2.66e-08 | 3.72e-07 | 35092 | 21256 | 13836 |
| European | Full Cohort | Switching vs Continuation | Recorded Anxiety | Income50-75k | 0.719 | 0.665 | 0.778 | <2.2e-16 | 2.26e-15 | 35092 | 21256 | 13836 |
| European | Full Cohort | Switching vs Continuation | Recorded Anxiety | Income75-100k | 0.618 | 0.568 | 0.672 | <2.2e-16 | <2.2e-16 | 35092 | 21256 | 13836 |
| European | Full Cohort | Switching vs Continuation | Recorded Anxiety | Education_levelCollege Postgraduate | 0.772 | 0.645 | 0.925 | 5.09e-03 | 7.12e-02 | 35092 | 21256 | 13836 |
| European | Full Cohort | Switching vs Continuation | Recorded Anxiety | Education_levelCollege Undergraduate | 0.843 | 0.707 | 1.004 | 5.53e-02 | 7.74e-01 | 35092 | 21256 | 13836 |
| European | Full Cohort | Switching vs Continuation | Recorded Anxiety | Education_levelTwelve or GED | 0.981 | 0.818 | 1.177 | 8.39e-01 | 1.00e+00 | 35092 | 21256 | 13836 |

| | | | | | | | | | | | | |
|---|---|---|---|---|---|---|---|---|---|---|---|---|
| European | Full Cohort | Switching vs Continuation | Recorded Anxiety | Recorded Anxiety | 2.626 | 2.497 | 2.762 | <2.2e-16 | <2.2e-16 | 35092 | 21256 | 13836 |
| European | Full Cohort | Discontinuation vs Continuation | Recorded Depression | (Intercept) | 0.933 | 0.772 | 1.128 | 4.76e-01 | 1.00e+00 | 28728 | 21256 | 7472 |
| European | Full Cohort | Discontinuation vs Continuation | Recorded Depression | age_at_first_AD | 1.082 | 1.053 | 1.111 | 6.61e-09 | 9.26e-08 | 28728 | 21256 | 7472 |
| European | Full Cohort | Discontinuation vs Continuation | Recorded Depression | sex_at_birthMale | 1.116 | 1.054 | 1.182 | 1.73e-04 | 2.42e-03 | 28728 | 21256 | 7472 |
| European | Full Cohort | Discontinuation vs Continuation | Recorded Depression | Income>200k | 0.543 | 0.485 | 0.609 | <2.2e-16 | <2.2e-16 | 28728 | 21256 | 7472 |
| European | Full Cohort | Discontinuation vs Continuation | Recorded Depression | Income100-150k | 0.506 | 0.459 | 0.557 | <2.2e-16 | <2.2e-16 | 28728 | 21256 | 7472 |
| European | Full Cohort | Discontinuation vs Continuation | Recorded Depression | Income150-200k | 0.491 | 0.434 | 0.556 | <2.2e-16 | <2.2e-16 | 28728 | 21256 | 7472 |
| European | Full Cohort | Discontinuation vs Continuation | Recorded Depression | Income25-35k | 0.732 | 0.656 | 0.817 | 2.70e-08 | 3.78e-07 | 28728 | 21256 | 7472 |
| European | Full Cohort | Discontinuation vs Continuation | Recorded Depression | Income35-50k | 0.643 | 0.580 | 0.714 | <2.2e-16 | 1.39e-15 | 28728 | 21256 | 7472 |
| European | Full Cohort | Discontinuation vs Continuation | Recorded Depression | Income50-75k | 0.648 | 0.590 | 0.711 | <2.2e-16 | <2.2e-16 | 28728 | 21256 | 7472 |
| European | Full Cohort | Discontinuation vs Continuation | Recorded Depression | Income75-100k | 0.545 | 0.493 | 0.603 | <2.2e-16 | <2.2e-16 | 28728 | 21256 | 7472 |
| European | Full Cohort | Discontinuation vs Continuation | Recorded Depression | Education_levelCollege Postgraduate | 0.581 | 0.475 | 0.709 | 1.01e-07 | 1.41e-06 | 28728 | 21256 | 7472 |
| European | Full Cohort | Discontinuation vs Continuation | Recorded Depression | Education_levelCollege Undergraduate | 0.655 | 0.540 | 0.794 | 1.69e-05 | 2.36e-04 | 28728 | 21256 | 7472 |
| European | Full Cohort | Discontinuation vs Continuation | Recorded Depression | Education_levelTwelve or GED | 0.778 | 0.636 | 0.951 | 1.41e-02 | 1.98e-01 | 28728 | 21256 | 7472 |
| European | Full Cohort | Discontinuation vs Continuation | Recorded Depression | Recorded Depression | 0.649 | 0.612 | 0.688 | <2.2e-16 | <2.2e-16 | 28728 | 21256 | 7472 |
| European | Full Cohort | Switching vs Continuation | Recorded Depression | (Intercept) | 0.531 | 0.444 | 0.634 | 2.98e-12 | 4.17e-11 | 35092 | 21256 | 13836 |
| European | Full Cohort | Switching vs Continuation | Recorded Depression | age_at_first_AD | 0.723 | 0.706 | 0.741 | <2.2e-16 | <2.2e-16 | 35092 | 21256 | 13836 |
| European | Full Cohort | Switching vs Continuation | Recorded Depression | sex_at_birthMale | 1.027 | 0.977 | 1.081 | 2.97e-01 | 1.00e+00 | 35092 | 21256 | 13836 |
| European | Full Cohort | Switching vs Continuation | Recorded Depression | Income>200k | 0.522 | 0.470 | 0.579 | <2.2e-16 | <2.2e-16 | 35092 | 21256 | 13836 |
| European | Full Cohort | Switching vs Continuation | Recorded Depression | Income100-150k | 0.667 | 0.614 | 0.725 | <2.2e-16 | <2.2e-16 | 35092 | 21256 | 13836 |
| European | Full Cohort | Switching vs Continuation | Recorded Depression | Income150-200k | 0.553 | 0.496 | 0.617 | <2.2e-16 | <2.2e-16 | 35092 | 21256 | 13836 |
| European | Full Cohort | Switching vs Continuation | Recorded Depression | Income25-35k | 0.873 | 0.794 | 0.960 | 5.02e-03 | 7.02e-02 | 35092 | 21256 | 13836 |
| European | Full Cohort | Switching vs Continuation | Recorded Depression | Income35-50k | 0.845 | 0.774 | 0.923 | 1.92e-04 | 2.69e-03 | 35092 | 21256 | 13836 |
| European | Full Cohort | Switching vs Continuation | Recorded Depression | Income50-75k | 0.787 | 0.726 | 0.853 | 4.60e-09 | 6.44e-08 | 35092 | 21256 | 13836 |
| European | Full Cohort | Switching vs Continuation | Recorded Depression | Income75-100k | 0.692 | 0.635 | 0.754 | <2.2e-16 | 6.52e-16 | 35092 | 21256 | 13836 |
| European | Full Cohort | Switching vs Continuation | Recorded Depression | Education_levelCollege Postgraduate | 0.737 | 0.612 | 0.887 | 1.24e-03 | 1.73e-02 | 35092 | 21256 | 13836 |
| European | Full Cohort | Switching vs Continuation | Recorded Depression | Education_levelCollege Undergraduate | 0.820 | 0.685 | 0.981 | 3.04e-02 | 4.26e-01 | 35092 | 21256 | 13836 |
| European | Full Cohort | Switching vs Continuation | Recorded Depression | Education_levelTwelve or GED | 0.977 | 0.810 | 1.177 | 8.04e-01 | 1.00e+00 | 35092 | 21256 | 13836 |
| European | Full Cohort | Switching vs Continuation | Recorded Depression | Recorded Depression | 3.503 | 3.344 | 3.670 | <2.2e-16 | <2.2e-16 | 35092 | 21256 | 13836 |
| European | Full Cohort | Discontinuation vs Continuation | Smoking_Frequency | (Intercept) | 0.652 | 0.534 | 0.794 | 2.27e-05 | 3.17e-04 | 28168 | 20861 | 7307 |
| European | Full Cohort | Discontinuation vs Continuation | Smoking_Frequency | age_at_first_AD | 1.111 | 1.081 | 1.142 | 6.06e-14 | 8.48e-13 | 28168 | 20861 | 7307 |
| European | Full Cohort | Discontinuation vs Continuation | Smoking_Frequency | sex_at_birthMale | 1.114 | 1.052 | 1.181 | 2.52e-04 | 3.52e-03 | 28168 | 20861 | 7307 |
| European | Full Cohort | Discontinuation vs Continuation | Smoking_Frequency | Income>200k | 0.621 | 0.553 | 0.697 | 6.92e-16 | 9.68e-15 | 28168 | 20861 | 7307 |
| European | Full Cohort | Discontinuation vs Continuation | Smoking_Frequency | Income100-150k | 0.571 | 0.517 | 0.631 | <2.2e-16 | <2.2e-16 | 28168 | 20861 | 7307 |
| European | Full Cohort | Discontinuation vs Continuation | Smoking_Frequency | Income150-200k | 0.567 | 0.500 | 0.643 | <2.2e-16 | <2.2e-16 | 28168 | 20861 | 7307 |
| European | Full Cohort | Discontinuation vs Continuation | Smoking_Frequency | Income25-35k | 0.776 | 0.694 | 0.867 | 7.74e-06 | 1.08e-04 | 28168 | 20861 | 7307 |
| European | Full Cohort | Discontinuation vs Continuation | Smoking_Frequency | Income35-50k | 0.699 | 0.629 | 0.777 | 3.25e-11 | 4.55e-10 | 28168 | 20861 | 7307 |
| European | Full Cohort | Discontinuation vs Continuation | Smoking_Frequency | Income50-75k | 0.708 | 0.644 | 0.778 | 7.57e-13 | 1.06e-11 | 28168 | 20861 | 7307 |
| European | Full Cohort | Discontinuation vs Continuation | Smoking_Frequency | Income75-100k | 0.611 | 0.552 | 0.677 | <2.2e-16 | <2.2e-16 | 28168 | 20861 | 7307 |
| European | Full Cohort | Discontinuation vs Continuation | Smoking_Frequency | Education_levelCollege Postgraduate | 0.652 | 0.531 | 0.800 | 4.32e-05 | 6.05e-04 | 28168 | 20861 | 7307 |
| European | Full Cohort | Discontinuation vs Continuation | Smoking_Frequency | Education_levelCollege Undergraduate | 0.733 | 0.602 | 0.893 | 2.04e-03 | 2.86e-02 | 28168 | 20861 | 7307 |
| European | Full Cohort | Discontinuation vs Continuation | Smoking_Frequency | Education_levelTwelve or GED | 0.852 | 0.695 | 1.044 | 1.23e-01 | 1.00e+00 | 28168 | 20861 | 7307 |
| European | Full Cohort | Discontinuation vs Continuation | Smoking_Frequency | Smoking_FrequencyEvery Day | 1.419 | 1.275 | 1.580 | 1.56e-10 | 2.18e-09 | 28168 | 20861 | 7307 |
| European | Full Cohort | Discontinuation vs Continuation | Smoking_Frequency | Smoking_FrequencyNot At All (former) | 0.934 | 0.879 | 0.993 | 2.78e-02 | 3.90e-01 | 28168 | 20861 | 7307 |
| European | Full Cohort | Discontinuation vs Continuation | Smoking_Frequency | Smoking_FrequencySome Days | 1.267 | 1.088 | 1.477 | 2.41e-03 | 3.37e-02 | 28168 | 20861 | 7307 |
| European | Full Cohort | Switching vs Continuation | Smoking_Frequency | (Intercept) | 0.963 | 0.808 | 1.147 | 6.72e-01 | 1.00e+00 | 34389 | 20861 | 13528 |
| European | Full Cohort | Switching vs Continuation | Smoking_Frequency | age_at_first_AD | 0.701 | 0.685 | 0.718 | <2.2e-16 | <2.2e-16 | 34389 | 20861 | 13528 |
| European | Full Cohort | Switching vs Continuation | Smoking_Frequency | sex_at_birthMale | 1.032 | 0.982 | 1.085 | 2.09e-01 | 1.00e+00 | 34389 | 20861 | 13528 |
| European | Full Cohort | Switching vs Continuation | Smoking_Frequency | Income>200k | 0.423 | 0.382 | 0.469 | <2.2e-16 | <2.2e-16 | 34389 | 20861 | 13528 |
| European | Full Cohort | Switching vs Continuation | Smoking_Frequency | Income100-150k | 0.581 | 0.535 | 0.631 | <2.2e-16 | <2.2e-16 | 34389 | 20861 | 13528 |
| European | Full Cohort | Switching vs Continuation | Smoking_Frequency | Income150-200k | 0.465 | 0.418 | 0.517 | <2.2e-16 | <2.2e-16 | 34389 | 20861 | 13528 |
| European | Full Cohort | Switching vs Continuation | Smoking_Frequency | Income25-35k | 0.854 | 0.779 | 0.937 | 8.15e-04 | 1.14e-02 | 34389 | 20861 | 13528 |
| European | Full Cohort | Switching vs Continuation | Smoking_Frequency | Income35-50k | 0.809 | 0.742 | 0.882 | 1.46e-06 | 2.05e-05 | 34389 | 20861 | 13528 |
| European | Full Cohort | Switching vs Continuation | Smoking_Frequency | Income50-75k | 0.742 | 0.686 | 0.802 | 8.35e-14 | 1.17e-12 | 34389 | 20861 | 13528 |
| European | Full Cohort | Switching vs Continuation | Smoking_Frequency | Income75-100k | 0.632 | 0.581 | 0.687 | <2.2e-16 | <2.2e-16 | 34389 | 20861 | 13528 |
| European | Full Cohort | Switching vs Continuation | Smoking_Frequency | Education_levelCollege Postgraduate | 0.812 | 0.677 | 0.974 | 2.46e-02 | 3.44e-01 | 34389 | 20861 | 13528 |
| European | Full Cohort | Switching vs Continuation | Smoking_Frequency | Education_levelCollege Undergraduate | 0.872 | 0.731 | 1.039 | 1.25e-01 | 1.00e+00 | 34389 | 20861 | 13528 |
| European | Full Cohort | Switching vs Continuation | Smoking_Frequency | Education_levelTwelve or GED | 0.993 | 0.828 | 1.190 | 9.37e-01 | 1.00e+00 | 34389 | 20861 | 13528 |
| European | Full Cohort | Switching vs Continuation | Smoking_Frequency | Smoking_FrequencyEvery Day | 1.453 | 1.329 | 1.589 | 2.61e-16 | 3.66e-15 | 34389 | 20861 | 13528 |
| European | Full Cohort | Switching vs Continuation | Smoking_Frequency | Smoking_FrequencyNot At All (former) | 1.182 | 1.124 | 1.243 | 6.20e-11 | 8.68e-10 | 34389 | 20861 | 13528 |
| European | Full Cohort | Switching vs Continuation | Smoking_Frequency | Smoking_FrequencySome Days | 1.220 | 1.073 | 1.387 | 2.35e-03 | 3.29e-02 | 34389 | 20861 | 13528 |
| European | Full Cohort | Discontinuation vs Continuation | Stress | (Intercept) | 0.582 | 0.398 | 0.852 | 5.35e-03 | 7.49e-02 | 17584 | 13352 | 4232 |
| European | Full Cohort | Discontinuation vs Continuation | Stress | age_at_first_AD | 1.119 | 1.079 | 1.160 | 1.64e-09 | 2.30e-08 | 17584 | 13352 | 4232 |
| European | Full Cohort | Discontinuation vs Continuation | Stress | sex_at_birthMale | 1.039 | 0.962 | 1.122 | 3.27e-01 | 1.00e+00 | 17584 | 13352 | 4232 |
| European | Full Cohort | Discontinuation vs Continuation | Stress | Income>200k | 0.666 | 0.572 | 0.776 | 1.65e-07 | 2.32e-06 | 17584 | 13352 | 4232 |
| European | Full Cohort | Discontinuation vs Continuation | Stress | Income100-150k | 0.630 | 0.552 | 0.720 | 1.12e-11 | 1.57e-10 | 17584 | 13352 | 4232 |

| | | | | | | | | | | | | |
|---|---|---|---|---|---|---|---|---|---|---|---|---|
| European | Full Cohort | Discontinuation vs Continuation | Stress | Income150-200k | 0.623 | 0.531 | 0.732 | 7.28e-09 | 1.02e-07 | 17584 | 13352 | 4232 |
| European | Full Cohort | Discontinuation vs Continuation | Stress | Income25-35k | 0.749 | 0.639 | 0.879 | 3.79e-04 | 5.30e-03 | 17584 | 13352 | 4232 |
| European | Full Cohort | Discontinuation vs Continuation | Stress | Income35-50k | 0.695 | 0.599 | 0.805 | 1.28e-06 | 1.80e-05 | 17584 | 13352 | 4232 |
| European | Full Cohort | Discontinuation vs Continuation | Stress | Income50-75k | 0.747 | 0.656 | 0.851 | 1.12e-05 | 1.57e-04 | 17584 | 13352 | 4232 |
| European | Full Cohort | Discontinuation vs Continuation | Stress | Income75-100k | 0.650 | 0.567 | 0.746 | 8.50e-10 | 1.19e-08 | 17584 | 13352 | 4232 |
| European | Full Cohort | Discontinuation vs Continuation | Stress | Education_levelCollege Postgraduate | 0.655 | 0.445 | 0.965 | 3.21e-02 | 4.49e-01 | 17584 | 13352 | 4232 |
| European | Full Cohort | Discontinuation vs Continuation | Stress | Education_levelCollege Undergraduate | 0.746 | 0.509 | 1.093 | 1.33e-01 | 1.00e+00 | 17584 | 13352 | 4232 |
| European | Full Cohort | Discontinuation vs Continuation | Stress | Education_levelTwelve or GED | 0.728 | 0.491 | 1.079 | 1.14e-01 | 1.00e+00 | 17584 | 13352 | 4232 |
| European | Full Cohort | Discontinuation vs Continuation | Stress | StressAlmost Never | 1.051 | 0.907 | 1.218 | 5.10e-01 | 1.00e+00 | 17584 | 13352 | 4232 |
| European | Full Cohort | Discontinuation vs Continuation | Stress | StressFairly Often | 0.980 | 0.842 | 1.140 | 7.89e-01 | 1.00e+00 | 17584 | 13352 | 4232 |
| European | Full Cohort | Discontinuation vs Continuation | Stress | StressSometimes | 1.111 | 1.027 | 1.203 | 8.72e-03 | 1.22e-01 | 17584 | 13352 | 4232 |
| European | Full Cohort | Discontinuation vs Continuation | Stress | StressVery Often | 1.066 | 0.890 | 1.276 | 4.88e-01 | 1.00e+00 | 17584 | 13352 | 4232 |
| European | Full Cohort | Switching vs Continuation | Stress | (Intercept) | 1.252 | 0.927 | 1.693 | 1.43e-01 | 1.00e+00 | 21801 | 13352 | 8449 |
| European | Full Cohort | Switching vs Continuation | Stress | age_at_first_AD | 0.681 | 0.661 | 0.702 | <2.2e-16 | <2.2e-16 | 21801 | 13352 | 8449 |
| European | Full Cohort | Switching vs Continuation | Stress | sex_at_birthMale | 1.109 | 1.041 | 1.181 | 1.44e-03 | 2.02e-02 | 21801 | 13352 | 8449 |
| European | Full Cohort | Switching vs Continuation | Stress | Income>200k | 0.404 | 0.355 | 0.460 | <2.2e-16 | <2.2e-16 | 21801 | 13352 | 8449 |
| European | Full Cohort | Switching vs Continuation | Stress | Income100-150k | 0.561 | 0.504 | 0.624 | <2.2e-16 | <2.2e-16 | 21801 | 13352 | 8449 |
| European | Full Cohort | Switching vs Continuation | Stress | Income150-200k | 0.431 | 0.377 | 0.492 | <2.2e-16 | <2.2e-16 | 21801 | 13352 | 8449 |
| European | Full Cohort | Switching vs Continuation | Stress | Income25-35k | 0.803 | 0.708 | 0.910 | 5.88e-04 | 8.23e-03 | 21801 | 13352 | 8449 |
| European | Full Cohort | Switching vs Continuation | Stress | Income35-50k | 0.766 | 0.682 | 0.860 | 5.81e-06 | 8.14e-05 | 21801 | 13352 | 8449 |
| European | Full Cohort | Switching vs Continuation | Stress | Income50-75k | 0.719 | 0.648 | 0.798 | 5.16e-10 | 7.23e-09 | 21801 | 13352 | 8449 |
| European | Full Cohort | Switching vs Continuation | Stress | Income75-100k | 0.625 | 0.560 | 0.697 | <2.2e-16 | 5.71e-16 | 21801 | 13352 | 8449 |
| European | Full Cohort | Switching vs Continuation | Stress | Education_levelCollege Postgraduate | 0.629 | 0.463 | 0.856 | 3.12e-03 | 4.37e-02 | 21801 | 13352 | 8449 |
| European | Full Cohort | Switching vs Continuation | Stress | Education_levelCollege Undergraduate | 0.697 | 0.515 | 0.943 | 1.93e-02 | 2.70e-01 | 21801 | 13352 | 8449 |
| European | Full Cohort | Switching vs Continuation | Stress | Education_levelTwelve or GED | 0.804 | 0.589 | 1.098 | 1.70e-01 | 1.00e+00 | 21801 | 13352 | 8449 |
| European | Full Cohort | Switching vs Continuation | Stress | StressAlmost Never | 1.018 | 0.900 | 1.151 | 7.82e-01 | 1.00e+00 | 21801 | 13352 | 8449 |
| European | Full Cohort | Switching vs Continuation | Stress | StressFairly Often | 1.038 | 0.921 | 1.170 | 5.37e-01 | 1.00e+00 | 21801 | 13352 | 8449 |
| European | Full Cohort | Switching vs Continuation | Stress | StressSometimes | 1.274 | 1.196 | 1.356 | 3.80e-14 | 5.31e-13 | 21801 | 13352 | 8449 |
| European | Full Cohort | Switching vs Continuation | Stress | StressVery Often | 1.095 | 0.942 | 1.273 | 2.37e-01 | 1.00e+00 | 21801 | 13352 | 8449 |
| European | Full Cohort | Discontinuation vs Continuation | age_at_first_AD | (Intercept) | 0.334 | 0.323 | 0.345 | <2.2e-16 | <2.2e-16 | 28728 | 21256 | 7472 |
| European | Full Cohort | Discontinuation vs Continuation | age_at_first_AD | age_at_first_AD | 1.080 | 1.052 | 1.108 | 1.04e-08 | 1.45e-07 | 28728 | 21256 | 7472 |
| European | Full Cohort | Discontinuation vs Continuation | age_at_first_AD | sex_at_birthMale | 1.126 | 1.064 | 1.192 | 3.95e-05 | 5.52e-04 | 28728 | 21256 | 7472 |
| European | Full Cohort | Switching vs Continuation | age_at_first_AD | (Intercept) | 0.636 | 0.619 | 0.653 | <2.2e-16 | <2.2e-16 | 35092 | 21256 | 13836 |
| European | Full Cohort | Switching vs Continuation | age_at_first_AD | age_at_first_AD | 0.710 | 0.695 | 0.726 | <2.2e-16 | <2.2e-16 | 35092 | 21256 | 13836 |
| European | Full Cohort | Switching vs Continuation | age_at_first_AD | sex_at_birthMale | 1.036 | 0.988 | 1.087 | 1.47e-01 | 1.00e+00 | 35092 | 21256 | 13836 |
| European | Full Cohort | Discontinuation vs Continuation | sex_at_birth | (Intercept) | 0.334 | 0.323 | 0.345 | <2.2e-16 | <2.2e-16 | 28728 | 21256 | 7472 |
| European | Full Cohort | Discontinuation vs Continuation | sex_at_birth | age_at_first_AD | 1.080 | 1.052 | 1.108 | 1.04e-08 | 1.45e-07 | 28728 | 21256 | 7472 |
| European | Full Cohort | Discontinuation vs Continuation | sex_at_birth | sex_at_birthMale | 1.126 | 1.064 | 1.192 | 3.95e-05 | 5.52e-04 | 28728 | 21256 | 7472 |
| European | Full Cohort | Switching vs Continuation | sex_at_birth | (Intercept) | 0.636 | 0.619 | 0.653 | <2.2e-16 | <2.2e-16 | 35092 | 21256 | 13836 |
| European | Full Cohort | Switching vs Continuation | sex_at_birth | age_at_first_AD | 0.710 | 0.695 | 0.726 | <2.2e-16 | <2.2e-16 | 35092 | 21256 | 13836 |
| European | Full Cohort | Switching vs Continuation | sex_at_birth | sex_at_birthMale | 1.036 | 0.988 | 1.087 | 1.47e-01 | 1.00e+00 | 35092 | 21256 | 13836 |
| European | Recorded Depression | Discontinuation vs Continuation | Alcohol_Frequency | (Intercept) | 1.187 | 0.831 | 1.695 | 3.47e-01 | 1.00e+00 | 10060 | 7894 | 2166 |
| European | Recorded Depression | Discontinuation vs Continuation | Alcohol_Frequency | age_at_first_AD | 1.081 | 1.029 | 1.135 | 1.90e-03 | 2.66e-02 | 10060 | 7894 | 2166 |
| European | Recorded Depression | Discontinuation vs Continuation | Alcohol_Frequency | sex_at_birthMale | 1.131 | 1.018 | 1.256 | 2.18e-02 | 3.05e-01 | 10060 | 7894 | 2166 |
| European | Recorded Depression | Discontinuation vs Continuation | Alcohol_Frequency | Income>200k | 0.467 | 0.369 | 0.592 | 2.56e-10 | 3.58e-09 | 10060 | 7894 | 2166 |
| European | Recorded Depression | Discontinuation vs Continuation | Alcohol_Frequency | Income100-150k | 0.462 | 0.386 | 0.553 | <2.2e-16 | 4.76e-16 | 10060 | 7894 | 2166 |
| European | Recorded Depression | Discontinuation vs Continuation | Alcohol_Frequency | Income150-200k | 0.445 | 0.349 | 0.567 | 5.95e-11 | 8.33e-10 | 10060 | 7894 | 2166 |
| European | Recorded Depression | Discontinuation vs Continuation | Alcohol_Frequency | Income25-35k | 0.667 | 0.559 | 0.796 | 7.46e-06 | 1.04e-04 | 10060 | 7894 | 2166 |
| European | Recorded Depression | Discontinuation vs Continuation | Alcohol_Frequency | Income35-50k | 0.606 | 0.510 | 0.720 | 1.24e-08 | 1.74e-07 | 10060 | 7894 | 2166 |
| European | Recorded Depression | Discontinuation vs Continuation | Alcohol_Frequency | Income50-75k | 0.568 | 0.485 | 0.665 | 1.71e-12 | 2.39e-11 | 10060 | 7894 | 2166 |
| European | Recorded Depression | Discontinuation vs Continuation | Alcohol_Frequency | Income75-100k | 0.453 | 0.377 | 0.543 | <2.2e-16 | 2.48e-16 | 10060 | 7894 | 2166 |
| European | Recorded Depression | Discontinuation vs Continuation | Alcohol_Frequency | Education_levelCollege Postgraduate | 0.445 | 0.326 | 0.607 | 3.44e-07 | 4.81e-06 | 10060 | 7894 | 2166 |
| European | Recorded Depression | Discontinuation vs Continuation | Alcohol_Frequency | Education_levelCollege Undergraduate | 0.580 | 0.433 | 0.777 | 2.58e-04 | 3.62e-03 | 10060 | 7894 | 2166 |
| European | Recorded Depression | Discontinuation vs Continuation | Alcohol_Frequency | Education_levelTwelve or GED | 0.705 | 0.519 | 0.958 | 2.52e-02 | 3.53e-01 | 10060 | 7894 | 2166 |
| European | Recorded Depression | Discontinuation vs Continuation | Alcohol_Frequency | Alcohol_Frequency2-3 Week | 0.580 | 0.440 | 0.764 | 1.03e-04 | 1.44e-03 | 10060 | 7894 | 2166 |
| European | Recorded Depression | Discontinuation vs Continuation | Alcohol_Frequency | Alcohol_Frequency2-4 Month | 0.534 | 0.409 | 0.697 | 3.99e-06 | 5.59e-05 | 10060 | 7894 | 2166 |
| European | Recorded Depression | Discontinuation vs Continuation | Alcohol_Frequency | Alcohol_Frequency4plus Week | 0.618 | 0.468 | 0.816 | 6.90e-04 | 9.66e-03 | 10060 | 7894 | 2166 |
| European | Recorded Depression | Discontinuation vs Continuation | Alcohol_Frequency | Alcohol_FrequencyMonthly or Less | 0.655 | 0.510 | 0.842 | 9.60e-04 | 1.34e-02 | 10060 | 7894 | 2166 |
| European | Recorded Depression | Discontinuation vs Continuation | Alcohol_Frequency | Alcohol_FrequencyNever (former) | 0.725 | 0.559 | 0.939 | 1.50e-02 | 2.09e-01 | 10060 | 7894 | 2166 |
| European | Recorded Depression | Switching vs Continuation | Alcohol_Frequency | (Intercept) | 1.886 | 1.426 | 2.496 | 8.85e-06 | 1.24e-04 | 17394 | 7894 | 9500 |
| European | Recorded Depression | Switching vs Continuation | Alcohol_Frequency | age_at_first_AD | 0.701 | 0.679 | 0.725 | <2.2e-16 | <2.2e-16 | 17394 | 7894 | 9500 |
| European | Recorded Depression | Switching vs Continuation | Alcohol_Frequency | sex_at_birthMale | 1.118 | 1.045 | 1.197 | 1.24e-03 | 1.74e-02 | 17394 | 7894 | 9500 |
| European | Recorded Depression | Switching vs Continuation | Alcohol_Frequency | Income>200k | 0.558 | 0.482 | 0.646 | 4.77e-15 | 6.68e-14 | 17394 | 7894 | 9500 |
| European | Recorded Depression | Switching vs Continuation | Alcohol_Frequency | Income100-150k | 0.689 | 0.616 | 0.772 | 8.90e-11 | 1.25e-09 | 17394 | 7894 | 9500 |
| European | Recorded Depression | Switching vs Continuation | Alcohol_Frequency | Income150-200k | 0.497 | 0.428 | 0.578 | <2.2e-16 | <2.2e-16 | 17394 | 7894 | 9500 |
| European | Recorded Depression | Switching vs Continuation | Alcohol_Frequency | Income25-35k | 0.826 | 0.732 | 0.933 | 2.07e-03 | 2.90e-02 | 17394 | 7894 | 9500 |

| | | | | | | | | | | | | |
|---|---|---|---|---|---|---|---|---|---|---|---|---|
| European | Recorded Depression | Switching vs Continuation | Alcohol_Frequency | Income35-50k | 0.829 | 0.739 | 0.930 | 1.41e-03 | 1.98e-02 | 17394 | 7894 | 9500 |
| European | Recorded Depression | Switching vs Continuation | Alcohol_Frequency | Income50-75k | 0.804 | 0.724 | 0.892 | 4.05e-05 | 5.67e-04 | 17394 | 7894 | 9500 |
| European | Recorded Depression | Switching vs Continuation | Alcohol_Frequency | Income75-100k | 0.702 | 0.627 | 0.787 | 1.09e-09 | 1.53e-08 | 17394 | 7894 | 9500 |
| European | Recorded Depression | Switching vs Continuation | Alcohol_Frequency | Education_levelCollege Postgraduate | 0.712 | 0.559 | 0.907 | 5.87e-03 | 8.22e-02 | 17394 | 7894 | 9500 |
| European | Recorded Depression | Switching vs Continuation | Alcohol_Frequency | Education_levelCollege Undergraduate | 0.828 | 0.655 | 1.046 | 1.14e-01 | 1.00e+00 | 17394 | 7894 | 9500 |
| European | Recorded Depression | Switching vs Continuation | Alcohol_Frequency | Education_levelTwelve or GED | 0.996 | 0.782 | 1.270 | 9.77e-01 | 1.00e+00 | 17394 | 7894 | 9500 |
| European | Recorded Depression | Switching vs Continuation | Alcohol_Frequency | Alcohol_Frequency2-3 Week | 0.796 | 0.651 | 0.972 | 2.52e-02 | 3.52e-01 | 17394 | 7894 | 9500 |
| European | Recorded Depression | Switching vs Continuation | Alcohol_Frequency | Alcohol_Frequency2-4 Month | 0.906 | 0.746 | 1.101 | 3.22e-01 | 1.00e+00 | 17394 | 7894 | 9500 |
| European | Recorded Depression | Switching vs Continuation | Alcohol_Frequency | Alcohol_Frequency4plus Week | 0.957 | 0.782 | 1.173 | 6.74e-01 | 1.00e+00 | 17394 | 7894 | 9500 |
| European | Recorded Depression | Switching vs Continuation | Alcohol_Frequency | Alcohol_FrequencyMonthly or Less | 0.969 | 0.802 | 1.170 | 7.41e-01 | 1.00e+00 | 17394 | 7894 | 9500 |
| European | Recorded Depression | Switching vs Continuation | Alcohol_Frequency | Alcohol_FrequencyNever (former) | 1.144 | 0.941 | 1.390 | 1.77e-01 | 1.00e+00 | 17394 | 7894 | 9500 |
| European | Recorded Depression | Discontinuation vs Continuation | BMI | (Intercept) | 0.826 | 0.622 | 1.096 | 1.86e-01 | 1.00e+00 | 9832 | 7783 | 2049 |
| European | Recorded Depression | Discontinuation vs Continuation | BMI | age_at_first_AD | 1.077 | 1.025 | 1.132 | 3.08e-03 | 4.31e-02 | 9832 | 7783 | 2049 |
| European | Recorded Depression | Discontinuation vs Continuation | BMI | sex_at_birthMale | 1.149 | 1.033 | 1.278 | 1.03e-02 | 1.45e-01 | 9832 | 7783 | 2049 |
| European | Recorded Depression | Discontinuation vs Continuation | BMI | Income>200k | 0.446 | 0.352 | 0.565 | 2.48e-11 | 3.48e-10 | 9832 | 7783 | 2049 |
| European | Recorded Depression | Discontinuation vs Continuation | BMI | Income100-150k | 0.449 | 0.375 | 0.538 | <2.2e-16 | <2.2e-16 | 9832 | 7783 | 2049 |
| European | Recorded Depression | Discontinuation vs Continuation | BMI | Income150-200k | 0.412 | 0.322 | 0.527 | 2.03e-12 | 2.84e-11 | 9832 | 7783 | 2049 |
| European | Recorded Depression | Discontinuation vs Continuation | BMI | Income25-35k | 0.644 | 0.537 | 0.773 | 2.31e-06 | 3.24e-05 | 9832 | 7783 | 2049 |
| European | Recorded Depression | Discontinuation vs Continuation | BMI | Income35-50k | 0.591 | 0.496 | 0.706 | 5.83e-09 | 8.16e-08 | 9832 | 7783 | 2049 |
| European | Recorded Depression | Discontinuation vs Continuation | BMI | Income50-75k | 0.558 | 0.476 | 0.655 | 8.19e-13 | 1.15e-11 | 9832 | 7783 | 2049 |
| European | Recorded Depression | Discontinuation vs Continuation | BMI | Income75-100k | 0.439 | 0.365 | 0.528 | <2.2e-16 | <2.2e-16 | 9832 | 7783 | 2049 |
| European | Recorded Depression | Discontinuation vs Continuation | BMI | Education_levelCollege Postgraduate | 0.393 | 0.288 | 0.538 | 4.92e-09 | 6.89e-08 | 9832 | 7783 | 2049 |
| European | Recorded Depression | Discontinuation vs Continuation | BMI | Education_levelCollege Undergraduate | 0.519 | 0.387 | 0.696 | 1.18e-05 | 1.65e-04 | 9832 | 7783 | 2049 |
| European | Recorded Depression | Discontinuation vs Continuation | BMI | Education_levelTwelve or GED | 0.651 | 0.479 | 0.885 | 6.16e-03 | 8.62e-02 | 9832 | 7783 | 2049 |
| European | Recorded Depression | Discontinuation vs Continuation | BMI | BMI | 0.923 | 0.877 | 0.972 | 2.19e-03 | 3.06e-02 | 9832 | 7783 | 2049 |
| European | Recorded Depression | Switching vs Continuation | BMI | (Intercept) | 1.925 | 1.533 | 2.416 | 1.69e-08 | 2.36e-07 | 17231 | 7783 | 9448 |
| European | Recorded Depression | Switching vs Continuation | BMI | age_at_first_AD | 0.712 | 0.689 | 0.735 | <2.2e-16 | <2.2e-16 | 17231 | 7783 | 9448 |
| European | Recorded Depression | Switching vs Continuation | BMI | sex_at_birthMale | 1.127 | 1.053 | 1.206 | 5.39e-04 | 7.54e-03 | 17231 | 7783 | 9448 |
| European | Recorded Depression | Switching vs Continuation | BMI | Income>200k | 0.549 | 0.474 | 0.635 | 6.90e-16 | 9.66e-15 | 17231 | 7783 | 9448 |
| European | Recorded Depression | Switching vs Continuation | BMI | Income100-150k | 0.665 | 0.594 | 0.744 | 8.40e-13 | 1.18e-11 | 17231 | 7783 | 9448 |
| European | Recorded Depression | Switching vs Continuation | BMI | Income150-200k | 0.477 | 0.411 | 0.555 | <2.2e-16 | <2.2e-16 | 17231 | 7783 | 9448 |
| European | Recorded Depression | Switching vs Continuation | BMI | Income25-35k | 0.814 | 0.721 | 0.920 | 9.34e-04 | 1.31e-02 | 17231 | 7783 | 9448 |
| European | Recorded Depression | Switching vs Continuation | BMI | Income35-50k | 0.811 | 0.722 | 0.910 | 3.65e-04 | 5.11e-03 | 17231 | 7783 | 9448 |
| European | Recorded Depression | Switching vs Continuation | BMI | Income50-75k | 0.781 | 0.704 | 0.867 | 3.38e-06 | 4.73e-05 | 17231 | 7783 | 9448 |
| European | Recorded Depression | Switching vs Continuation | BMI | Income75-100k | 0.671 | 0.599 | 0.752 | 5.44e-12 | 7.61e-11 | 17231 | 7783 | 9448 |
| European | Recorded Depression | Switching vs Continuation | BMI | Education_levelCollege Postgraduate | 0.682 | 0.535 | 0.869 | 1.93e-03 | 2.70e-02 | 17231 | 7783 | 9448 |
| European | Recorded Depression | Switching vs Continuation | BMI | Education_levelCollege Undergraduate | 0.800 | 0.633 | 1.011 | 6.19e-02 | 8.67e-01 | 17231 | 7783 | 9448 |
| European | Recorded Depression | Switching vs Continuation | BMI | Education_levelTwelve or GED | 0.967 | 0.759 | 1.234 | 7.89e-01 | 1.00e+00 | 17231 | 7783 | 9448 |
| European | Recorded Depression | Switching vs Continuation | BMI | BMI | 1.061 | 1.028 | 1.095 | 2.68e-04 | 3.75e-03 | 17231 | 7783 | 9448 |
| European | Recorded Depression | Discontinuation vs Continuation | Education_level | (Intercept) | 0.834 | 0.633 | 1.098 | 1.96e-01 | 1.00e+00 | 10166 | 7969 | 2197 |
| European | Recorded Depression | Discontinuation vs Continuation | Education_level | age_at_first_AD | 1.094 | 1.042 | 1.148 | 2.56e-04 | 3.58e-03 | 10166 | 7969 | 2197 |
| European | Recorded Depression | Discontinuation vs Continuation | Education_level | sex_at_birthMale | 1.137 | 1.025 | 1.260 | 1.53e-02 | 2.14e-01 | 10166 | 7969 | 2197 |
| European | Recorded Depression | Discontinuation vs Continuation | Education_level | Income>200k | 0.441 | 0.350 | 0.555 | 3.91e-12 | 5.48e-11 | 10166 | 7969 | 2197 |
| European | Recorded Depression | Discontinuation vs Continuation | Education_level | Income100-150k | 0.437 | 0.367 | 0.522 | <2.2e-16 | <2.2e-16 | 10166 | 7969 | 2197 |
| European | Recorded Depression | Discontinuation vs Continuation | Education_level | Income150-200k | 0.416 | 0.328 | 0.529 | 6.75e-13 | 9.45e-12 | 10166 | 7969 | 2197 |
| European | Recorded Depression | Discontinuation vs Continuation | Education_level | Income25-35k | 0.661 | 0.555 | 0.787 | 3.32e-06 | 4.65e-05 | 10166 | 7969 | 2197 |
| European | Recorded Depression | Discontinuation vs Continuation | Education_level | Income35-50k | 0.593 | 0.500 | 0.703 | 1.86e-09 | 2.61e-08 | 10166 | 7969 | 2197 |
| European | Recorded Depression | Discontinuation vs Continuation | Education_level | Income50-75k | 0.546 | 0.468 | 0.638 | 2.25e-14 | 3.15e-13 | 10166 | 7969 | 2197 |
| European | Recorded Depression | Discontinuation vs Continuation | Education_level | Income75-100k | 0.433 | 0.362 | 0.518 | <2.2e-16 | <2.2e-16 | 10166 | 7969 | 2197 |
| European | Recorded Depression | Discontinuation vs Continuation | Education_level | Education_levelCollege Postgraduate | 0.413 | 0.305 | 0.560 | 1.19E-08 | 1.67e-07 | 10166 | 7969 | 2197 |
| European | Recorded Depression | Discontinuation vs Continuation | Education_level | Education_levelCollege Undergraduate | 0.541 | 0.407 | 0.720 | 2.47e-05 | 3.46e-04 | 10166 | 7969 | 2197 |
| European | Recorded Depression | Discontinuation vs Continuation | Education_level | Education_levelTwelve or GED | 0.673 | 0.499 | 0.907 | 9.27e-03 | 1.30e-01 | 10166 | 7969 | 2197 |
| European | Recorded Depression | Switching vs Continuation | Education_level | (Intercept) | 1.893 | 1.515 | 2.366 | 2.00e-08 | 2.80e-07 | 17576 | 7969 | 9607 |
| European | Recorded Depression | Switching vs Continuation | Education_level | age_at_first_AD | 0.713 | 0.690 | 0.736 | <2.2e-16 | <2.2e-16 | 17576 | 7969 | 9607 |
| European | Recorded Depression | Switching vs Continuation | Education_level | sex_at_birthMale | 1.125 | 1.052 | 1.203 | 5.91e-04 | 8.27e-03 | 17576 | 7969 | 9607 |
| European | Recorded Depression | Switching vs Continuation | Education_level | Income>200k | 0.534 | 0.463 | 0.616 | <2.2e-16 | <2.2e-16 | 17576 | 7969 | 9607 |
| European | Recorded Depression | Switching vs Continuation | Education_level | Income100-150k | 0.662 | 0.592 | 0.739 | 2.35e-13 | 3.28e-12 | 17576 | 7969 | 9607 |
| European | Recorded Depression | Switching vs Continuation | Education_level | Income150-200k | 0.471 | 0.406 | 0.546 | <2.2e-16 | <2.2e-16 | 17576 | 7969 | 9607 |
| European | Recorded Depression | Switching vs Continuation | Education_level | Income25-35k | 0.815 | 0.723 | 0.920 | 8.80e-04 | 1.23e-02 | 17576 | 7969 | 9607 |
| European | Recorded Depression | Switching vs Continuation | Education_level | Income35-50k | 0.811 | 0.724 | 0.909 | 3.19e-04 | 4.47e-03 | 17576 | 7969 | 9607 |
| European | Recorded Depression | Switching vs Continuation | Education_level | Income50-75k | 0.782 | 0.706 | 0.867 | 2.95e-06 | 4.13e-05 | 17576 | 7969 | 9607 |
| European | Recorded Depression | Switching vs Continuation | Education_level | Income75-100k | 0.674 | 0.602 | 0.753 | 4.64e-12 | 6.50e-11 | 17576 | 7969 | 9607 |
| European | Recorded Depression | Switching vs Continuation | Education_level | Education_levelCollege Postgraduate | 0.693 | 0.546 | 0.878 | 2.42e-03 | 3.39e-02 | 17576 | 7969 | 9607 |
| European | Recorded Depression | Switching vs Continuation | Education_level | Education_levelCollege Undergraduate | 0.812 | 0.645 | 1.021 | 7.47e-02 | 1.00e+00 | 17576 | 7969 | 9607 |
| European | Recorded Depression | Switching vs Continuation | Education_level | Education_levelTwelve or GED | 0.989 | 0.779 | 1.255 | 9.25e-01 | 1.00e+00 | 17576 | 7969 | 9607 |
| European | Recorded Depression | Discontinuation vs Continuation | Income | (Intercept) | 0.834 | 0.633 | 1.098 | 1.96e-01 | 1.00e+00 | 10166 | 7969 | 2197 |

| | | | | | | | | | | | | |
|---|---|---|---|---|---|---|---|---|---|---|---|---|
| European | Recorded Depression | Discontinuation vs Continuation | Income | age_at_first_AD | 1.094 | 1.042 | 1.148 | 2.56e-04 | 3.58e-03 | 10166 | 7969 | 2197 |
| European | Recorded Depression | Discontinuation vs Continuation | Income | sex_at_birthMale | 1.137 | 1.025 | 1.260 | 1.53e-02 | 2.14e-01 | 10166 | 7969 | 2197 |
| European | Recorded Depression | Discontinuation vs Continuation | Income | Income>200k | 0.441 | 0.350 | 0.555 | 3.91e-12 | 5.48e-11 | 10166 | 7969 | 2197 |
| European | Recorded Depression | Discontinuation vs Continuation | Income | Income100-150k | 0.437 | 0.367 | 0.522 | <2.2e-16 | <2.2e-16 | 10166 | 7969 | 2197 |
| European | Recorded Depression | Discontinuation vs Continuation | Income | Income150-200k | 0.416 | 0.328 | 0.529 | 6.75E-13 | 9.45e-12 | 10166 | 7969 | 2197 |
| European | Recorded Depression | Discontinuation vs Continuation | Income | Income25-35k | 0.661 | 0.555 | 0.787 | 3.32e-06 | 4.65e-05 | 10166 | 7969 | 2197 |
| European | Recorded Depression | Discontinuation vs Continuation | Income | Income35-50k | 0.593 | 0.500 | 0.703 | 1.86e-09 | 2.61e-08 | 10166 | 7969 | 2197 |
| European | Recorded Depression | Discontinuation vs Continuation | Income | Income50-75k | 0.546 | 0.468 | 0.638 | 2.25e-14 | 3.15e-13 | 10166 | 7969 | 2197 |
| European | Recorded Depression | Discontinuation vs Continuation | Income | Income75-100k | 0.433 | 0.362 | 0.518 | <2.2e-16 | <2.2e-16 | 10166 | 7969 | 2197 |
| European | Recorded Depression | Discontinuation vs Continuation | Income | Education_levelCollege Postgraduate | 0.413 | 0.305 | 0.560 | 1.19e-08 | 1.67e-07 | 10166 | 7969 | 2197 |
| European | Recorded Depression | Discontinuation vs Continuation | Income | Education_levelCollege Undergraduate | 0.541 | 0.407 | 0.720 | 2.47e-05 | 3.46e-04 | 10166 | 7969 | 2197 |
| European | Recorded Depression | Discontinuation vs Continuation | Income | Education_levelTwelve or GED | 0.673 | 0.499 | 0.907 | 9.27e-03 | 1.30e-01 | 10166 | 7969 | 2197 |
| European | Recorded Depression | Switching vs Continuation | Income | (Intercept) | 1.893 | 1.515 | 2.366 | 2.00e-08 | 2.80e-07 | 17576 | 7969 | 9607 |
| European | Recorded Depression | Switching vs Continuation | Income | age_at_first_AD | 0.713 | 0.690 | 0.736 | <2.2e-16 | <2.2e-16 | 17576 | 7969 | 9607 |
| European | Recorded Depression | Switching vs Continuation | Income | sex_at_birthMale | 1.125 | 1.052 | 1.203 | 5.91e-04 | 8.27e-03 | 17576 | 7969 | 9607 |
| European | Recorded Depression | Switching vs Continuation | Income | Income>200k | 0.534 | 0.463 | 0.616 | <2.2e-16 | <2.2e-16 | 17576 | 7969 | 9607 |
| European | Recorded Depression | Switching vs Continuation | Income | Income100-150k | 0.662 | 0.592 | 0.739 | 2.35e-13 | 3.28e-12 | 17576 | 7969 | 9607 |
| European | Recorded Depression | Switching vs Continuation | Income | Income150-200k | 0.471 | 0.406 | 0.546 | <2.2e-16 | <2.2e-16 | 17576 | 7969 | 9607 |
| European | Recorded Depression | Switching vs Continuation | Income | Income25-35k | 0.815 | 0.723 | 0.920 | 8.80e-04 | 1.23e-02 | 17576 | 7969 | 9607 |
| European | Recorded Depression | Switching vs Continuation | Income | Income35-50k | 0.811 | 0.724 | 0.909 | 3.19e-04 | 4.47e-03 | 17576 | 7969 | 9607 |
| European | Recorded Depression | Switching vs Continuation | Income | Income50-75k | 0.782 | 0.706 | 0.867 | 2.95e-06 | 4.13e-05 | 17576 | 7969 | 9607 |
| European | Recorded Depression | Switching vs Continuation | Income | Income75-100k | 0.674 | 0.602 | 0.753 | 4.64e-12 | 6.50e-11 | 17576 | 7969 | 9607 |
| European | Recorded Depression | Switching vs Continuation | Income | Education_levelCollege Postgraduate | 0.693 | 0.546 | 0.878 | 2.42e-03 | 3.39e-02 | 17576 | 7969 | 9607 |
| European | Recorded Depression | Switching vs Continuation | Income | Education_levelCollege Undergraduate | 0.812 | 0.645 | 1.021 | 7.47e-02 | 1.00e+00 | 17576 | 7969 | 9607 |
| European | Recorded Depression | Switching vs Continuation | Income | Education_levelTwelve or GED | 0.989 | 0.779 | 1.255 | 9.25e-01 | 1.00e+00 | 17576 | 7969 | 9607 |
| European | Recorded Depression | Discontinuation vs Continuation | Insomnia | (Intercept) | 0.798 | 0.605 | 1.053 | 1.11e-01 | 1.00e+00 | 10166 | 7969 | 2197 |
| European | Recorded Depression | Discontinuation vs Continuation | Insomnia | age_at_first_AD | 1.086 | 1.034 | 1.139 | 8.64e-04 | 1.21e-02 | 10166 | 7969 | 2197 |
| European | Recorded Depression | Discontinuation vs Continuation | Insomnia | sex_at_birthMale | 1.136 | 1.024 | 1.260 | 1.57e-02 | 2.20e-01 | 10166 | 7969 | 2197 |
| European | Recorded Depression | Discontinuation vs Continuation | Insomnia | Income>200k | 0.441 | 0.350 | 0.556 | 4.06e-12 | 5.69e-11 | 10166 | 7969 | 2197 |
| European | Recorded Depression | Discontinuation vs Continuation | Insomnia | Income100-150k | 0.440 | 0.368 | 0.524 | <2.2e-16 | <2.2e-16 | 10166 | 7969 | 2197 |
| European | Recorded Depression | Discontinuation vs Continuation | Insomnia | Income150-200k | 0.417 | 0.328 | 0.529 | 7.58e-13 | 1.06e-11 | 10166 | 7969 | 2197 |
| European | Recorded Depression | Discontinuation vs Continuation | Insomnia | Income25-35k | 0.663 | 0.556 | 0.789 | 3.95e-06 | 5.54e-05 | 10166 | 7969 | 2197 |
| European | Recorded Depression | Discontinuation vs Continuation | Insomnia | Income35-50k | 0.595 | 0.502 | 0.706 | 2.57e-09 | 3.60e-08 | 10166 | 7969 | 2197 |
| European | Recorded Depression | Discontinuation vs Continuation | Insomnia | Income50-75k | 0.549 | 0.470 | 0.641 | 3.73e-14 | 5.23e-13 | 10166 | 7969 | 2197 |
| European | Recorded Depression | Discontinuation vs Continuation | Insomnia | Income75-100k | 0.434 | 0.363 | 0.520 | <2.2e-16 | <2.2e-16 | 10166 | 7969 | 2197 |
| European | Recorded Depression | Discontinuation vs Continuation | Insomnia | Education_levelCollege Postgraduate | 0.414 | 0.306 | 0.562 | 1.36e-08 | 1.90e-07 | 10166 | 7969 | 2197 |
| European | Recorded Depression | Discontinuation vs Continuation | Insomnia | Education_levelCollege Undergraduate | 0.541 | 0.407 | 0.720 | 2.51e-05 | 3.52e-04 | 10166 | 7969 | 2197 |
| European | Recorded Depression | Discontinuation vs Continuation | Insomnia | Education_levelTwelve or GED | 0.675 | 0.500 | 0.910 | 9.90e-03 | 1.39e-01 | 10166 | 7969 | 2197 |
| European | Recorded Depression | Discontinuation vs Continuation | Insomnia | Insomnia | 1.189 | 1.063 | 1.330 | 2.54e-03 | 3.55e-02 | 10166 | 7969 | 2197 |
| European | Recorded Depression | Switching vs Continuation | Insomnia | (Intercept) | 1.364 | 1.085 | 1.715 | 7.78e-03 | 1.09e-01 | 17576 | 7969 | 9607 |
| European | Recorded Depression | Switching vs Continuation | Insomnia | age_at_first_AD | 0.686 | 0.664 | 0.709 | <2.2e-16 | <2.2e-16 | 17576 | 7969 | 9607 |
| European | Recorded Depression | Switching vs Continuation | Insomnia | sex_at_birthMale | 1.092 | 1.019 | 1.169 | 1.23e-02 | 1.73e-01 | 17576 | 7969 | 9607 |
| European | Recorded Depression | Switching vs Continuation | Insomnia | Income>200k | 0.549 | 0.474 | 0.636 | 1.03e-15 | 1.44e-14 | 17576 | 7969 | 9607 |
| European | Recorded Depression | Switching vs Continuation | Insomnia | Income100-150k | 0.696 | 0.621 | 0.779 | 2.93e-10 | 4.10e-09 | 17576 | 7969 | 9607 |
| European | Recorded Depression | Switching vs Continuation | Insomnia | Income150-200k | 0.491 | 0.422 | 0.571 | <2.2e-16 | <2.2e-16 | 17576 | 7969 | 9607 |
| European | Recorded Depression | Switching vs Continuation | Insomnia | Income25-35k | 0.837 | 0.740 | 0.946 | 4.46e-03 | 6.25e-02 | 17576 | 7969 | 9607 |
| European | Recorded Depression | Switching vs Continuation | Insomnia | Income35-50k | 0.834 | 0.742 | 0.936 | 2.17e-03 | 3.04e-02 | 17576 | 7969 | 9607 |
| European | Recorded Depression | Switching vs Continuation | Insomnia | Income50-75k | 0.818 | 0.737 | 0.909 | 1.92e-04 | 2.69e-03 | 17576 | 7969 | 9607 |
| European | Recorded Depression | Switching vs Continuation | Insomnia | Income75-100k | 0.700 | 0.624 | 0.784 | 9.40e-10 | 1.32e-08 | 17576 | 7969 | 9607 |
| European | Recorded Depression | Switching vs Continuation | Insomnia | Education_levelCollege Postgraduate | 0.708 | 0.555 | 0.902 | 5.23e-03 | 7.32e-02 | 17576 | 7969 | 9607 |
| European | Recorded Depression | Switching vs Continuation | Insomnia | Education_levelCollege Undergraduate | 0.827 | 0.654 | 1.046 | 1.12e-01 | 1.00e+00 | 17576 | 7969 | 9607 |
| European | Recorded Depression | Switching vs Continuation | Insomnia | Education_levelTwelve or GED | 1.004 | 0.787 | 1.281 | 9.72e-01 | 1.00e+00 | 17576 | 7969 | 9607 |
| European | Recorded Depression | Switching vs Continuation | Insomnia | Insomnia | 2.541 | 2.372 | 2.723 | <2.2e-16 | <2.2e-16 | 17576 | 7969 | 9607 |
| European | Recorded Depression | Discontinuation vs Continuation | MDD CIDI-sf | (Intercept) | 0.450 | 0.134 | 1.508 | 1.95e-01 | 1.00e+00 | 1888 | 1572 | 316 |
| European | Recorded Depression | Discontinuation vs Continuation | MDD CIDI-sf | age_at_first_AD | 1.044 | 0.913 | 1.195 | 5.28e-01 | 1.00e+00 | 1888 | 1572 | 316 |
| European | Recorded Depression | Discontinuation vs Continuation | MDD CIDI-sf | sex_at_birthMale | 1.150 | 0.878 | 1.507 | 3.11e-01 | 1.00e+00 | 1888 | 1572 | 316 |
| European | Recorded Depression | Discontinuation vs Continuation | MDD CIDI-sf | Income>200k | 0.584 | 0.323 | 1.056 | 7.51e-02 | 1.00e+00 | 1888 | 1572 | 316 |
| European | Recorded Depression | Discontinuation vs Continuation | MDD CIDI-sf | Income100-150k | 0.815 | 0.523 | 1.270 | 3.66e-01 | 1.00e+00 | 1888 | 1572 | 316 |
| European | Recorded Depression | Discontinuation vs Continuation | MDD CIDI-sf | Income150-200k | 0.611 | 0.342 | 1.090 | 9.54e-02 | 1.00e+00 | 1888 | 1572 | 316 |
| European | Recorded Depression | Discontinuation vs Continuation | MDD CIDI-sf | Income25-35k | 0.677 | 0.403 | 1.135 | 1.39e-01 | 1.00e+00 | 1888 | 1572 | 316 |
| European | Recorded Depression | Discontinuation vs Continuation | MDD CIDI-sf | Income35-50k | 0.848 | 0.522 | 1.377 | 5.06e-01 | 1.00e+00 | 1888 | 1572 | 316 |
| European | Recorded Depression | Discontinuation vs Continuation | MDD CIDI-sf | Income50-75k | 0.822 | 0.536 | 1.259 | 3.67e-01 | 1.00e+00 | 1888 | 1572 | 316 |
| European | Recorded Depression | Discontinuation vs Continuation | MDD CIDI-sf | Income75-100k | 0.767 | 0.487 | 1.209 | 2.53e-01 | 1.00e+00 | 1888 | 1572 | 316 |
| European | Recorded Depression | Discontinuation vs Continuation | MDD CIDI-sf | Education_levelCollege Postgraduate | 0.410 | 0.119 | 1.417 | 1.59e-01 | 1.00e+00 | 1888 | 1572 | 316 |
| European | Recorded Depression | Discontinuation vs Continuation | MDD CIDI-sf | Education_levelCollege Undergraduate | 0.583 | 0.173 | 1.966 | 3.84e-01 | 1.00e+00 | 1888 | 1572 | 316 |
| European | Recorded Depression | Discontinuation vs Continuation | MDD CIDI-sf | Education_levelTwelve or GED | 0.555 | 0.155 | 1.984 | 3.65e-01 | 1.00e+00 | 1888 | 1572 | 316 |

| | | | | | | | | | | | | |
|---|---|---|---|---|---|---|---|---|---|---|---|---|
| European | Recorded Depression | Discontinuation vs Continuation | MDD CIDI-sf | MDD CIDI-sf | 1.050 | 0.797 | 1.385 | 7.27e-01 | 1.00e+00 | 1888 | 1572 | 316 |
| European | Recorded Depression | Switching vs Continuation | MDD CIDI-sf | (Intercept) | 0.714 | 0.281 | 1.812 | 4.78e-01 | 1.00e+00 | 3466 | 1572 | 1894 |
| European | Recorded Depression | Switching vs Continuation | MDD CIDI-sf | age_at_first_AD | 0.654 | 0.604 | 0.708 | <2.2e-16 | <2.2e-16 | 3466 | 1572 | 1894 |
| European | Recorded Depression | Switching vs Continuation | MDD CIDI-sf | sex_at_birthMale | 1.206 | 1.029 | 1.412 | 2.04e-02 | 2.85e-01 | 3466 | 1572 | 1894 |
| European | Recorded Depression | Switching vs Continuation | MDD CIDI-sf | Income>200k | 0.517 | 0.371 | 0.721 | 1.01e-04 | 1.41e-03 | 3466 | 1572 | 1894 |
| European | Recorded Depression | Switching vs Continuation | MDD CIDI-sf | Income100-150k | 0.715 | 0.552 | 0.927 | 1.13e-02 | 1.58e-01 | 3466 | 1572 | 1894 |
| European | Recorded Depression | Switching vs Continuation | MDD CIDI-sf | Income150-200k | 0.501 | 0.360 | 0.697 | 4.10e-05 | 5.74e-04 | 3466 | 1572 | 1894 |
| European | Recorded Depression | Switching vs Continuation | MDD CIDI-sf | Income25-35k | 0.747 | 0.556 | 1.004 | 5.32e-02 | 7.45e-01 | 3466 | 1572 | 1894 |
| European | Recorded Depression | Switching vs Continuation | MDD CIDI-sf | Income35-50k | 1.049 | 0.796 | 1.384 | 7.33e-01 | 1.00e+00 | 3466 | 1572 | 1894 |
| European | Recorded Depression | Switching vs Continuation | MDD CIDI-sf | Income50-75k | 0.853 | 0.666 | 1.093 | 2.09e-01 | 1.00e+00 | 3466 | 1572 | 1894 |
| European | Recorded Depression | Switching vs Continuation | MDD CIDI-sf | Income75-100k | 0.737 | 0.566 | 0.959 | 2.33e-02 | 3.26e-01 | 3466 | 1572 | 1894 |
| European | Recorded Depression | Switching vs Continuation | MDD CIDI-sf | Education_levelCollege Postgraduate | 1.220 | 0.476 | 3.129 | 6.78e-01 | 1.00e+00 | 3466 | 1572 | 1894 |
| European | Recorded Depression | Switching vs Continuation | MDD CIDI-sf | Education_levelCollege Undergraduate | 1.404 | 0.552 | 3.567 | 4.76e-01 | 1.00e+00 | 3466 | 1572 | 1894 |
| European | Recorded Depression | Switching vs Continuation | MDD CIDI-sf | Education_levelTwelve or GED | 1.773 | 0.681 | 4.616 | 2.41e-01 | 1.00e+00 | 3466 | 1572 | 1894 |
| European | Recorded Depression | Switching vs Continuation | MDD CIDI-sf | MDD CIDI-sf | 1.694 | 1.431 | 2.005 | 9.15e-10 | 1.28e-08 | 3466 | 1572 | 1894 |
| European | Recorded Depression | Discontinuation vs Continuation | MDD_symptom_count | (Intercept) | 0.577 | 0.214 | 1.556 | 2.77e-01 | 1.00e+00 | 2392 | 2003 | 389 |
| European | Recorded Depression | Discontinuation vs Continuation | MDD_symptom_count | age_at_first_AD | 1.113 | 0.983 | 1.260 | 9.13e-02 | 1.00e+00 | 2392 | 2003 | 389 |
| European | Recorded Depression | Discontinuation vs Continuation | MDD_symptom_count | sex_at_birthMale | 0.985 | 0.766 | 1.266 | 9.05e-01 | 1.00e+00 | 2392 | 2003 | 389 |
| European | Recorded Depression | Discontinuation vs Continuation | MDD_symptom_count | Income>200k | 0.522 | 0.304 | 0.898 | 1.89e-02 | 2.64e-01 | 2392 | 2003 | 389 |
| European | Recorded Depression | Discontinuation vs Continuation | MDD_symptom_count | Income100-150k | 0.794 | 0.532 | 1.185 | 2.59e-01 | 1.00e+00 | 2392 | 2003 | 389 |
| European | Recorded Depression | Discontinuation vs Continuation | MDD_symptom_count | Income150-200k | 0.554 | 0.328 | 0.936 | 2.71e-02 | 3.80e-01 | 2392 | 2003 | 389 |
| European | Recorded Depression | Discontinuation vs Continuation | MDD_symptom_count | Income25-35k | 0.620 | 0.389 | 0.987 | 4.39e-02 | 6.14e-01 | 2392 | 2003 | 389 |
| European | Recorded Depression | Discontinuation vs Continuation | MDD_symptom_count | Income35-50k | 0.806 | 0.526 | 1.234 | 3.21e-01 | 1.00e+00 | 2392 | 2003 | 389 |
| European | Recorded Depression | Discontinuation vs Continuation | MDD_symptom_count | Income50-75k | 0.728 | 0.495 | 1.070 | 1.06e-01 | 1.00e+00 | 2392 | 2003 | 389 |
| European | Recorded Depression | Discontinuation vs Continuation | MDD_symptom_count | Income75-100k | 0.746 | 0.498 | 1.118 | 1.56e-01 | 1.00e+00 | 2392 | 2003 | 389 |
| European | Recorded Depression | Discontinuation vs Continuation | MDD_symptom_count | Education_levelCollege Postgraduate | 0.345 | 0.122 | 0.980 | 4.57e-02 | 6.40e-01 | 2392 | 2003 | 389 |
| European | Recorded Depression | Discontinuation vs Continuation | MDD_symptom_count | Education_levelCollege Undergraduate | 0.485 | 0.175 | 1.344 | 1.64e-01 | 1.00e+00 | 2392 | 2003 | 389 |
| European | Recorded Depression | Discontinuation vs Continuation | MDD_symptom_count | Education_levelTwelve or GED | 0.491 | 0.168 | 1.432 | 1.93e-01 | 1.00e+00 | 2392 | 2003 | 389 |
| European | Recorded Depression | Discontinuation vs Continuation | MDD_symptom_count | MDD_symptom_count | 1.083 | 0.953 | 1.230 | 2.23e-01 | 1.00e+00 | 2392 | 2003 | 389 |
| European | Recorded Depression | Switching vs Continuation | MDD_symptom_count | (Intercept) | 1.164 | 0.543 | 2.497 | 6.97e-01 | 1.00e+00 | 4326 | 2003 | 2323 |
| European | Recorded Depression | Switching vs Continuation | MDD_symptom_count | age_at_first_AD | 0.651 | 0.605 | 0.699 | <2.2e-16 | <2.2e-16 | 4326 | 2003 | 2323 |
| European | Recorded Depression | Switching vs Continuation | MDD_symptom_count | sex_at_birthMale | 1.287 | 1.117 | 1.484 | 4.89e-04 | 6.84e-03 | 4326 | 2003 | 2323 |
| European | Recorded Depression | Switching vs Continuation | MDD_symptom_count | Income>200k | 0.526 | 0.390 | 0.708 | 2.35e-05 | 3.29e-04 | 4326 | 2003 | 2323 |
| European | Recorded Depression | Switching vs Continuation | MDD_symptom_count | Income100-150k | 0.735 | 0.580 | 0.931 | 1.08e-02 | 1.51e-01 | 4326 | 2003 | 2323 |
| European | Recorded Depression | Switching vs Continuation | MDD_symptom_count | Income150-200k | 0.494 | 0.367 | 0.665 | 3.21e-06 | 4.49e-05 | 4326 | 2003 | 2323 |
| European | Recorded Depression | Switching vs Continuation | MDD_symptom_count | Income25-35k | 0.741 | 0.568 | 0.968 | 2.78e-02 | 3.89e-01 | 4326 | 2003 | 2323 |
| European | Recorded Depression | Switching vs Continuation | MDD_symptom_count | Income35-50k | 1.012 | 0.790 | 1.297 | 9.23e-01 | 1.00e+00 | 4326 | 2003 | 2323 |
| European | Recorded Depression | Switching vs Continuation | MDD_symptom_count | Income50-75k | 0.856 | 0.683 | 1.071 | 1.74e-01 | 1.00e+00 | 4326 | 2003 | 2323 |
| European | Recorded Depression | Switching vs Continuation | MDD_symptom_count | Income75-100k | 0.735 | 0.579 | 0.933 | 1.13e-02 | 1.58e-01 | 4326 | 2003 | 2323 |
| European | Recorded Depression | Switching vs Continuation | MDD_symptom_count | Education_levelCollege Postgraduate | 0.998 | 0.457 | 2.181 | 9.96e-01 | 1.00e+00 | 4326 | 2003 | 2323 |
| European | Recorded Depression | Switching vs Continuation | MDD_symptom_count | Education_levelCollege Undergraduate | 1.140 | 0.526 | 2.470 | 7.39e-01 | 1.00e+00 | 4326 | 2003 | 2323 |
| European | Recorded Depression | Switching vs Continuation | MDD_symptom_count | Education_levelTwelve or GED | 1.550 | 0.700 | 3.432 | 2.79e-01 | 1.00e+00 | 4326 | 2003 | 2323 |
| European | Recorded Depression | Switching vs Continuation | MDD_symptom_count | MDD_symptom_count | 1.299 | 1.205 | 1.401 | 9.91e-12 | 1.39e-10 | 4326 | 2003 | 2323 |
| European | Recorded Depression | Discontinuation vs Continuation | Migraine | (Intercept) | 0.860 | 0.652 | 1.136 | 2.89e-01 | 1.00e+00 | 10166 | 7969 | 2197 |
| European | Recorded Depression | Discontinuation vs Continuation | Migraine | age_at_first_AD | 1.090 | 1.038 | 1.144 | 4.72e-04 | 6.60e-03 | 10166 | 7969 | 2197 |
| European | Recorded Depression | Discontinuation vs Continuation | Migraine | sex_at_birthMale | 1.123 | 1.012 | 1.246 | 2.94e-02 | 4.11e-01 | 10166 | 7969 | 2197 |
| European | Recorded Depression | Discontinuation vs Continuation | Migraine | Income>200k | 0.440 | 0.349 | 0.554 | 3.43e-12 | 4.80e-11 | 10166 | 7969 | 2197 |
| European | Recorded Depression | Discontinuation vs Continuation | Migraine | Income100-150k | 0.438 | 0.368 | 0.523 | <2.2e-16 | <2.2e-16 | 10166 | 7969 | 2197 |
| European | Recorded Depression | Discontinuation vs Continuation | Migraine | Income150-200k | 0.416 | 0.327 | 0.528 | 6.56e-13 | 9.18e-12 | 10166 | 7969 | 2197 |
| European | Recorded Depression | Discontinuation vs Continuation | Migraine | Income25-35k | 0.660 | 0.554 | 0.786 | 3.19e-06 | 4.47e-05 | 10166 | 7969 | 2197 |
| European | Recorded Depression | Discontinuation vs Continuation | Migraine | Income35-50k | 0.594 | 0.501 | 0.704 | 2.11e-09 | 2.96e-08 | 10166 | 7969 | 2197 |
| European | Recorded Depression | Discontinuation vs Continuation | Migraine | Income50-75k | 0.546 | 0.467 | 0.637 | 2.02e-14 | 2.82e-13 | 10166 | 7969 | 2197 |
| European | Recorded Depression | Discontinuation vs Continuation | Migraine | Income75-100k | 0.432 | 0.361 | 0.516 | <2.2e-16 | <2.2e-16 | 10166 | 7969 | 2197 |
| European | Recorded Depression | Discontinuation vs Continuation | Migraine | Education_levelCollege Postgraduate | 0.414 | 0.305 | 0.561 | 1.29e-08 | 1.80e-07 | 10166 | 7969 | 2197 |
| European | Recorded Depression | Discontinuation vs Continuation | Migraine | Education_levelCollege Undergraduate | 0.542 | 0.407 | 0.721 | 2.57e-05 | 3.60e-04 | 10166 | 7969 | 2197 |
| European | Recorded Depression | Discontinuation vs Continuation | Migraine | Education_levelTwelve or GED | 0.672 | 0.498 | 0.906 | 9.09e-03 | 1.27e-01 | 10166 | 7969 | 2197 |
| European | Recorded Depression | Discontinuation vs Continuation | Migraine | Migraine | 0.906 | 0.811 | 1.011 | 7.73e-02 | 1.00e+00 | 10166 | 7969 | 2197 |
| European | Recorded Depression | Switching vs Continuation | Migraine | (Intercept) | 1.472 | 1.174 | 1.847 | 8.06e-04 | 1.13e-02 | 17576 | 7969 | 9607 |
| European | Recorded Depression | Switching vs Continuation | Migraine | age_at_first_AD | 0.732 | 0.709 | 0.756 | <2.2e-16 | <2.2e-16 | 17576 | 7969 | 9607 |
| European | Recorded Depression | Switching vs Continuation | Migraine | sex_at_birthMale | 1.236 | 1.154 | 1.324 | 1.31e-09 | 1.83e-08 | 17576 | 7969 | 9607 |
| European | Recorded Depression | Switching vs Continuation | Migraine | Income>200k | 0.546 | 0.473 | 0.632 | 3.21e-16 | 4.49e-15 | 17576 | 7969 | 9607 |
| European | Recorded Depression | Switching vs Continuation | Migraine | Income100-150k | 0.663 | 0.593 | 0.742 | 6.48e-13 | 9.07e-12 | 17576 | 7969 | 9607 |
| European | Recorded Depression | Switching vs Continuation | Migraine | Income150-200k | 0.480 | 0.413 | 0.558 | <2.2e-16 | <2.2e-16 | 17576 | 7969 | 9607 |
| European | Recorded Depression | Switching vs Continuation | Migraine | Income25-35k | 0.812 | 0.719 | 0.917 | 7.93e-04 | 1.11e-02 | 17576 | 7969 | 9607 |
| European | Recorded Depression | Switching vs Continuation | Migraine | Income35-50k | 0.809 | 0.720 | 0.907 | 3.03e-04 | 4.24e-03 | 17576 | 7969 | 9607 |
| European | Recorded Depression | Switching vs Continuation | Migraine | Income50-75k | 0.795 | 0.716 | 0.882 | 1.56e-05 | 2.19e-04 | 17576 | 7969 | 9607 |

| | | | | | | | | | | | | |
|---|---|---|---|---|---|---|---|---|---|---|---|---|
| European | Recorded Depression | Switching vs Continuation | Migraine | Income75-100k | 0.685 | 0.612 | 0.767 | 6.30e-11 | 8.82e-10 | 17576 | 7969 | 9607 |
| European | Recorded Depression | Switching vs Continuation | Migraine | Education_levelCollege Postgraduate | 0.669 | 0.526 | 0.850 | 1.02e-03 | 1.43e-02 | 17576 | 7969 | 9607 |
| European | Recorded Depression | Switching vs Continuation | Migraine | Education_levelCollege Undergraduate | 0.783 | 0.621 | 0.987 | 3.83e-02 | 5.37e-01 | 17576 | 7969 | 9607 |
| European | Recorded Depression | Switching vs Continuation | Migraine | Education_levelTwelve or GED | 0.948 | 0.745 | 1.207 | 6.65e-01 | 1.00e+00 | 17576 | 7969 | 9607 |
| European | Recorded Depression | Switching vs Continuation | Migraine | Migraine | 1.975 | 1.851 | 2.107 | <2.2e-16 | <2.2e-16 | 17576 | 7969 | 9607 |
| European | Recorded Depression | Discontinuation vs Continuation | Recorded Anxiety | (Intercept) | 0.759 | 0.569 | 1.014 | 6.20e-02 | 8.68e-01 | 10166 | 7969 | 2197 |
| European | Recorded Depression | Discontinuation vs Continuation | Recorded Anxiety | age_at_first_AD | 1.101 | 1.049 | 1.156 | 9.79e-05 | 1.37e-03 | 10166 | 7969 | 2197 |
| European | Recorded Depression | Discontinuation vs Continuation | Recorded Anxiety | sex_at_birthMale | 1.146 | 1.033 | 1.271 | 9.94e-03 | 1.39e-01 | 10166 | 7969 | 2197 |
| European | Recorded Depression | Discontinuation vs Continuation | Recorded Anxiety | Income>200k | 0.446 | 0.354 | 0.562 | 8.06e-12 | 1.13e-10 | 10166 | 7969 | 2197 |
| European | Recorded Depression | Discontinuation vs Continuation | Recorded Anxiety | Income100-150k | 0.439 | 0.368 | 0.524 | <2.2e-16 | <2.2e-16 | 10166 | 7969 | 2197 |
| European | Recorded Depression | Discontinuation vs Continuation | Recorded Anxiety | Income150-200k | 0.419 | 0.330 | 0.533 | 1.12e-12 | 1.57e-11 | 10166 | 7969 | 2197 |
| European | Recorded Depression | Discontinuation vs Continuation | Recorded Anxiety | Income25-35k | 0.661 | 0.555 | 0.787 | 3.50e-06 | 4.90e-05 | 10166 | 7969 | 2197 |
| European | Recorded Depression | Discontinuation vs Continuation | Recorded Anxiety | Income35-50k | 0.595 | 0.501 | 0.705 | 2.39e-09 | 3.35e-08 | 10166 | 7969 | 2197 |
| European | Recorded Depression | Discontinuation vs Continuation | Recorded Anxiety | Income50-75k | 0.549 | 0.470 | 0.641 | 3.60e-14 | 5.04e-13 | 10166 | 7969 | 2197 |
| European | Recorded Depression | Discontinuation vs Continuation | Recorded Anxiety | Income75-100k | 0.435 | 0.364 | 0.521 | <2.2e-16 | <2.2e-16 | 10166 | 7969 | 2197 |
| European | Recorded Depression | Discontinuation vs Continuation | Recorded Anxiety | Education_levelCollege Postgraduate | 0.415 | 0.306 | 0.562 | 1.39e-08 | 1.95e-07 | 10166 | 7969 | 2197 |
| European | Recorded Depression | Discontinuation vs Continuation | Recorded Anxiety | Education_levelCollege Undergraduate | 0.543 | 0.408 | 0.722 | 2.68e-05 | 3.75e-04 | 10166 | 7969 | 2197 |
| European | Recorded Depression | Discontinuation vs Continuation | Recorded Anxiety | Education_levelTwelve or GED | 0.675 | 0.501 | 0.910 | 9.93e-03 | 1.39e-01 | 10166 | 7969 | 2197 |
| European | Recorded Depression | Discontinuation vs Continuation | Recorded Anxiety | Recorded Anxiety | 1.123 | 1.008 | 1.253 | 3.62e-02 | 5.06e-01 | 10166 | 7969 | 2197 |
| European | Recorded Depression | Switching vs Continuation | Recorded Anxiety | (Intercept) | 0.926 | 0.733 | 1.170 | 5.20e-01 | 1.00e+00 | 17576 | 7969 | 9607 |
| European | Recorded Depression | Switching vs Continuation | Recorded Anxiety | age_at_first_AD | 0.748 | 0.724 | 0.773 | <2.2e-16 | <2.2e-16 | 17576 | 7969 | 9607 |
| European | Recorded Depression | Switching vs Continuation | Recorded Anxiety | sex_at_birthMale | 1.167 | 1.090 | 1.250 | 8.88e-06 | 1.24e-04 | 17576 | 7969 | 9607 |
| European | Recorded Depression | Switching vs Continuation | Recorded Anxiety | Income>200k | 0.570 | 0.492 | 0.659 | 3.62e-14 | 5.07e-13 | 17576 | 7969 | 9607 |
| European | Recorded Depression | Switching vs Continuation | Recorded Anxiety | Income100-150k | 0.671 | 0.600 | 0.750 | 2.78e-12 | 3.90e-11 | 17576 | 7969 | 9607 |
| European | Recorded Depression | Switching vs Continuation | Recorded Anxiety | Income150-200k | 0.490 | 0.421 | 0.569 | <2.2e-16 | <2.2e-16 | 17576 | 7969 | 9607 |
| European | Recorded Depression | Switching vs Continuation | Recorded Anxiety | Income25-35k | 0.809 | 0.717 | 0.914 | 6.63e-04 | 9.28e-03 | 17576 | 7969 | 9607 |
| European | Recorded Depression | Switching vs Continuation | Recorded Anxiety | Income35-50k | 0.820 | 0.731 | 0.921 | 7.61e-04 | 1.07e-02 | 17576 | 7969 | 9607 |
| European | Recorded Depression | Switching vs Continuation | Recorded Anxiety | Income50-75k | 0.795 | 0.716 | 0.882 | 1.63e-05 | 2.29e-04 | 17576 | 7969 | 9607 |
| European | Recorded Depression | Switching vs Continuation | Recorded Anxiety | Income75-100k | 0.699 | 0.624 | 0.783 | 6.03e-10 | 8.44e-09 | 17576 | 7969 | 9607 |
| European | Recorded Depression | Switching vs Continuation | Recorded Anxiety | Education_levelCollege Postgraduate | 0.714 | 0.561 | 0.907 | 5.84e-03 | 8.17e-02 | 17576 | 7969 | 9607 |
| European | Recorded Depression | Switching vs Continuation | Recorded Anxiety | Education_levelCollege Undergraduate | 0.823 | 0.652 | 1.037 | 9.89e-02 | 1.00e+00 | 17576 | 7969 | 9607 |
| European | Recorded Depression | Switching vs Continuation | Recorded Anxiety | Education_levelTwelve or GED | 0.987 | 0.776 | 1.256 | 9.17e-01 | 1.00e+00 | 17576 | 7969 | 9607 |
| European | Recorded Depression | Switching vs Continuation | Recorded Anxiety | Recorded Anxiety | 2.347 | 2.172 | 2.535 | <2.2e-16 | <2.2e-16 | 17576 | 7969 | 9607 |
| European | Recorded Depression | Discontinuation vs Continuation | Smoking_Frequency | (Intercept) | 0.588 | 0.436 | 0.793 | 5.00e-04 | 7.01e-03 | 9978 | 7823 | 2155 |
| European | Recorded Depression | Discontinuation vs Continuation | Smoking_Frequency | age_at_first_AD | 1.118 | 1.063 | 1.176 | 1.31e-05 | 1.84e-04 | 9978 | 7823 | 2155 |
| European | Recorded Depression | Discontinuation vs Continuation | Smoking_Frequency | sex_at_birthMale | 1.109 | 0.999 | 1.232 | 5.31e-02 | 7.43e-01 | 9978 | 7823 | 2155 |
| European | Recorded Depression | Discontinuation vs Continuation | Smoking_Frequency | Income>200k | 0.486 | 0.384 | 0.616 | 2.33e-09 | 3.26e-08 | 9978 | 7823 | 2155 |
| European | Recorded Depression | Discontinuation vs Continuation | Smoking_Frequency | Income100-150k | 0.482 | 0.402 | 0.578 | 2.90e-15 | 4.06e-14 | 9978 | 7823 | 2155 |
| European | Recorded Depression | Discontinuation vs Continuation | Smoking_Frequency | Income150-200k | 0.464 | 0.363 | 0.592 | 6.73e-10 | 9.42e-09 | 9978 | 7823 | 2155 |
| European | Recorded Depression | Discontinuation vs Continuation | Smoking_Frequency | Income25-35k | 0.706 | 0.591 | 0.843 | 1.22e-04 | 1.71e-03 | 9978 | 7823 | 2155 |
| European | Recorded Depression | Discontinuation vs Continuation | Smoking_Frequency | Income35-50k | 0.651 | 0.547 | 0.775 | 1.33e-06 | 1.86e-05 | 9978 | 7823 | 2155 |
| European | Recorded Depression | Discontinuation vs Continuation | Smoking_Frequency | Income50-75k | 0.604 | 0.515 | 0.708 | 5.05e-10 | 7.07e-09 | 9978 | 7823 | 2155 |
| European | Recorded Depression | Discontinuation vs Continuation | Smoking_Frequency | Income75-100k | 0.476 | 0.396 | 0.572 | 2.32e-15 | 3.25e-14 | 9978 | 7823 | 2155 |
| European | Recorded Depression | Discontinuation vs Continuation | Smoking_Frequency | Education_levelCollege Postgraduate | 0.521 | 0.380 | 0.715 | 5.34e-05 | 7.47e-04 | 9978 | 7823 | 2155 |
| European | Recorded Depression | Discontinuation vs Continuation | Smoking_Frequency | Education_levelCollege Undergraduate | 0.658 | 0.490 | 0.885 | 5.66e-03 | 7.92e-02 | 9978 | 7823 | 2155 |
| European | Recorded Depression | Discontinuation vs Continuation | Smoking_Frequency | Education_levelTwelve or GED | 0.779 | 0.573 | 1.060 | 1.12e-01 | 1.00e+00 | 9978 | 7823 | 2155 |
| European | Recorded Depression | Discontinuation vs Continuation | Smoking_Frequency | Smoking_FrequencyEvery Day | 1.897 | 1.597 | 2.253 | 3.00e-13 | 4.20e-12 | 9978 | 7823 | 2155 |
| European | Recorded Depression | Discontinuation vs Continuation | Smoking_Frequency | Smoking_FrequencyNot At All (former) | 1.026 | 0.919 | 1.146 | 6.47e-01 | 1.00e+00 | 9978 | 7823 | 2155 |
| European | Recorded Depression | Discontinuation vs Continuation | Smoking_Frequency | Smoking_FrequencySome Days | 1.591 | 1.244 | 2.035 | 2.16e-04 | 3.03e-03 | 9978 | 7823 | 2155 |
| European | Recorded Depression | Switching vs Continuation | Smoking_Frequency | (Intercept) | 1.549 | 1.228 | 1.954 | 2.23e-04 | 3.12e-03 | 17221 | 7823 | 9398 |
| European | Recorded Depression | Switching vs Continuation | Smoking_Frequency | age_at_first_AD | 0.706 | 0.683 | 0.730 | <2.2e-16 | <2.2e-16 | 17221 | 7823 | 9398 |
| European | Recorded Depression | Switching vs Continuation | Smoking_Frequency | sex_at_birthMale | 1.103 | 1.030 | 1.181 | 4.78e-03 | 6.70e-02 | 17221 | 7823 | 9398 |
| European | Recorded Depression | Switching vs Continuation | Smoking_Frequency | Income>200k | 0.559 | 0.483 | 0.646 | 5.23e-15 | 7.32e-14 | 17221 | 7823 | 9398 |
| European | Recorded Depression | Switching vs Continuation | Smoking_Frequency | Income100-150k | 0.686 | 0.613 | 0.768 | 6.74e-11 | 9.44e-10 | 17221 | 7823 | 9398 |
| European | Recorded Depression | Switching vs Continuation | Smoking_Frequency | Income150-200k | 0.491 | 0.422 | 0.571 | <2.2e-16 | <2.2e-16 | 17221 | 7823 | 9398 |
| European | Recorded Depression | Switching vs Continuation | Smoking_Frequency | Income25-35k | 0.824 | 0.729 | 0.931 | 1.87e-03 | 2.62e-02 | 17221 | 7823 | 9398 |
| European | Recorded Depression | Switching vs Continuation | Smoking_Frequency | Income35-50k | 0.836 | 0.745 | 0.939 | 2.54e-03 | 3.56e-02 | 17221 | 7823 | 9398 |
| European | Recorded Depression | Switching vs Continuation | Smoking_Frequency | Income50-75k | 0.799 | 0.719 | 0.888 | 3.11e-05 | 4.35e-04 | 17221 | 7823 | 9398 |
| European | Recorded Depression | Switching vs Continuation | Smoking_Frequency | Income75-100k | 0.701 | 0.625 | 0.786 | 1.18e-09 | 1.65e-08 | 17221 | 7823 | 9398 |
| European | Recorded Depression | Switching vs Continuation | Smoking_Frequency | Education_levelCollege Postgraduate | 0.775 | 0.608 | 0.987 | 3.89e-02 | 5.44e-01 | 17221 | 7823 | 9398 |
| European | Recorded Depression | Switching vs Continuation | Smoking_Frequency | Education_levelCollege Undergraduate | 0.881 | 0.698 | 1.114 | 2.90e-01 | 1.00e+00 | 17221 | 7823 | 9398 |
| European | Recorded Depression | Switching vs Continuation | Smoking_Frequency | Education_levelTwelve or GED | 1.033 | 0.811 | 1.317 | 7.90e-01 | 1.00e+00 | 17221 | 7823 | 9398 |
| European | Recorded Depression | Switching vs Continuation | Smoking_Frequency | Smoking_FrequencyEvery Day | 1.521 | 1.345 | 1.721 | 2.51e-11 | 3.51e-10 | 17221 | 7823 | 9398 |
| European | Recorded Depression | Switching vs Continuation | Smoking_Frequency | Smoking_FrequencyNot At All (former) | 1.175 | 1.097 | 1.258 | 4.22e-06 | 5.91e-05 | 17221 | 7823 | 9398 |
| European | Recorded Depression | Switching vs Continuation | Smoking_Frequency | Smoking_FrequencySome Days | 1.147 | 0.963 | 1.367 | 1.23e-01 | 1.00e+00 | 17221 | 7823 | 9398 |
| European | Recorded Depression | Discontinuation vs Continuation | Stress | (Intercept) | 0.688 | 0.389 | 1.217 | 1.99e-01 | 1.00e+00 | 5778 | 4777 | 1001 |

| | | | | | | | | | | | | |
|---|---|---|---|---|---|---|---|---|---|---|---|---|
| European | Recorded Depression | Discontinuation vs Continuation | Stress | age_at_first_AD | 1.095 | 1.020 | 1.177 | 1.25e-02 | 1.75e-01 | 5778 | 4777 | 1001 |
| European | Recorded Depression | Discontinuation vs Continuation | Stress | sex_at_birthMale | 1.013 | 0.869 | 1.180 | 8.71e-01 | 1.00e+00 | 5778 | 4777 | 1001 |
| European | Recorded Depression | Discontinuation vs Continuation | Stress | Income>200k | 0.557 | 0.405 | 0.765 | 3.08e-04 | 4.31e-03 | 5778 | 4777 | 1001 |
| European | Recorded Depression | Discontinuation vs Continuation | Stress | Income100-150k | 0.661 | 0.516 | 0.847 | 1.03e-03 | 1.45e-02 | 5778 | 4777 | 1001 |
| European | Recorded Depression | Discontinuation vs Continuation | Stress | Income150-200k | 0.581 | 0.424 | 0.796 | 7.18e-04 | 1.00e-02 | 5778 | 4777 | 1001 |
| European | Recorded Depression | Discontinuation vs Continuation | Stress | Income25-35k | 0.642 | 0.485 | 0.850 | 1.93e-03 | 2.71e-02 | 5778 | 4777 | 1001 |
| European | Recorded Depression | Discontinuation vs Continuation | Stress | Income35-50k | 0.606 | 0.464 | 0.792 | 2.39e-04 | 3.34e-03 | 5778 | 4777 | 1001 |
| European | Recorded Depression | Discontinuation vs Continuation | Stress | Income50-75k | 0.634 | 0.502 | 0.799 | 1.19e-04 | 1.66e-03 | 5778 | 4777 | 1001 |
| European | Recorded Depression | Discontinuation vs Continuation | Stress | Income75-100k | 0.558 | 0.433 | 0.719 | 6.78e-06 | 9.49e-05 | 5778 | 4777 | 1001 |
| European | Recorded Depression | Discontinuation vs Continuation | Stress | Education_levelCollege Postgraduate | 0.355 | 0.197 | 0.641 | 5.90e-04 | 8.26e-03 | 5778 | 4777 | 1001 |
| European | Recorded Depression | Discontinuation vs Continuation | Stress | Education_levelCollege Undergraduate | 0.437 | 0.246 | 0.777 | 4.83e-03 | 6.76e-02 | 5778 | 4777 | 1001 |
| European | Recorded Depression | Discontinuation vs Continuation | Stress | Education_levelTwelve or GED | 0.486 | 0.266 | 0.886 | 1.86e-02 | 2.61e-01 | 5778 | 4777 | 1001 |
| European | Recorded Depression | Discontinuation vs Continuation | Stress | StressAlmost Never | 1.228 | 0.925 | 1.630 | 1.55e-01 | 1.00e+00 | 5778 | 4777 | 1001 |
| European | Recorded Depression | Discontinuation vs Continuation | Stress | StressFairly Often | 1.087 | 0.804 | 1.471 | 5.86e-01 | 1.00e+00 | 5778 | 4777 | 1001 |
| European | Recorded Depression | Discontinuation vs Continuation | Stress | StressSometimes | 1.191 | 1.022 | 1.387 | 2.48e-02 | 3.47e-01 | 5778 | 4777 | 1001 |
| European | Recorded Depression | Discontinuation vs Continuation | Stress | StressVery Often | 0.975 | 0.680 | 1.399 | 8.92e-01 | 1.00e+00 | 5778 | 4777 | 1001 |
| European | Recorded Depression | Switching vs Continuation | Stress | (Intercept) | 2.307 | 1.526 | 3.489 | 7.36e-05 | 1.03e-03 | 10518 | 4777 | 5741 |
| European | Recorded Depression | Switching vs Continuation | Stress | age_at_first_AD | 0.693 | 0.663 | 0.723 | <2.2e-16 | <2.2e-16 | 10518 | 4777 | 5741 |
| European | Recorded Depression | Switching vs Continuation | Stress | sex_at_birthMale | 1.191 | 1.090 | 1.302 | 1.10e-04 | 1.54e-03 | 10518 | 4777 | 5741 |
| European | Recorded Depression | Switching vs Continuation | Stress | Income>200k | 0.516 | 0.428 | 0.622 | 3.98e-12 | 5.57e-11 | 10518 | 4777 | 5741 |
| European | Recorded Depression | Switching vs Continuation | Stress | Income100-150k | 0.691 | 0.595 | 0.802 | 1.14e-06 | 1.60e-05 | 10518 | 4777 | 5741 |
| European | Recorded Depression | Switching vs Continuation | Stress | Income150-200k | 0.450 | 0.371 | 0.544 | 2.25e-16 | 3.16e-15 | 10518 | 4777 | 5741 |
| European | Recorded Depression | Switching vs Continuation | Stress | Income25-35k | 0.785 | 0.664 | 0.929 | 4.71e-03 | 6.59e-02 | 10518 | 4777 | 5741 |
| European | Recorded Depression | Switching vs Continuation | Stress | Income35-50k | 0.821 | 0.702 | 0.960 | 1.36e-02 | 1.91e-01 | 10518 | 4777 | 5741 |
| European | Recorded Depression | Switching vs Continuation | Stress | Income50-75k | 0.773 | 0.672 | 0.889 | 3.14e-04 | 4.39e-03 | 10518 | 4777 | 5741 |
| European | Recorded Depression | Switching vs Continuation | Stress | Income75-100k | 0.669 | 0.576 | 0.777 | 1.43e-07 | 2.00e-06 | 10518 | 4777 | 5741 |
| European | Recorded Depression | Switching vs Continuation | Stress | Education_levelCollege Postgraduate | 0.537 | 0.351 | 0.820 | 4.01e-03 | 5.62e-02 | 10518 | 4777 | 5741 |
| European | Recorded Depression | Switching vs Continuation | Stress | Education_levelCollege Undergraduate | 0.639 | 0.421 | 0.970 | 3.56e-02 | 4.98e-01 | 10518 | 4777 | 5741 |
| European | Recorded Depression | Switching vs Continuation | Stress | Education_levelTwelve or GED | 0.758 | 0.493 | 1.164 | 2.06e-01 | 1.00e+00 | 10518 | 4777 | 5741 |
| European | Recorded Depression | Switching vs Continuation | Stress | StressAlmost Never | 0.995 | 0.837 | 1.183 | 9.59e-01 | 1.00e+00 | 10518 | 4777 | 5741 |
| European | Recorded Depression | Switching vs Continuation | Stress | StressFairly Often | 1.090 | 0.921 | 1.290 | 3.15e-01 | 1.00e+00 | 10518 | 4777 | 5741 |
| European | Recorded Depression | Switching vs Continuation | Stress | StressSometimes | 1.184 | 1.085 | 1.292 | 1.49e-04 | 2.09e-03 | 10518 | 4777 | 5741 |
| European | Recorded Depression | Switching vs Continuation | Stress | StressVery Often | 1.021 | 0.833 | 1.252 | 8.41e-01 | 1.00e+00 | 10518 | 4777 | 5741 |
| European | Recorded Depression | Discontinuation vs Continuation | age_at_first_AD | (Intercept) | 0.261 | 0.247 | 0.277 | <2.2e-16 | <2.2e-16 | 10166 | 7969 | 2197 |
| European | Recorded Depression | Discontinuation vs Continuation | age_at_first_AD | age_at_first_AD | 1.081 | 1.031 | 1.133 | 1.27e-03 | 1.77e-02 | 10166 | 7969 | 2197 |
| European | Recorded Depression | Discontinuation vs Continuation | age_at_first_AD | sex_at_birthMale | 1.146 | 1.035 | 1.268 | 8.48e-03 | 1.19e-01 | 10166 | 7969 | 2197 |
| European | Recorded Depression | Switching vs Continuation | age_at_first_AD | (Intercept) | 1.141 | 1.100 | 1.183 | 1.28e-12 | 1.79e-11 | 17576 | 7969 | 9607 |
| European | Recorded Depression | Switching vs Continuation | age_at_first_AD | age_at_first_AD | 0.716 | 0.694 | 0.739 | <2.2e-16 | <2.2e-16 | 17576 | 7969 | 9607 |
| European | Recorded Depression | Switching vs Continuation | age_at_first_AD | sex_at_birthMale | 1.111 | 1.039 | 1.187 | 1.93e-03 | 2.70e-02 | 17576 | 7969 | 9607 |
| European | Recorded Depression | Discontinuation vs Continuation | sex_at_birth | (Intercept) | 0.261 | 0.247 | 0.277 | <2.2e-16 | <2.2e-16 | 10166 | 7969 | 2197 |
| European | Recorded Depression | Discontinuation vs Continuation | sex_at_birth | age_at_first_AD | 1.081 | 1.031 | 1.133 | 1.27e-03 | 1.77e-02 | 10166 | 7969 | 2197 |
| European | Recorded Depression | Discontinuation vs Continuation | sex_at_birth | sex_at_birthMale | 1.146 | 1.035 | 1.268 | 8.48e-03 | 1.19e-01 | 10166 | 7969 | 2197 |
| European | Recorded Depression | Switching vs Continuation | sex_at_birth | (Intercept) | 1.141 | 1.100 | 1.183 | 1.28e-12 | 1.79e-11 | 17576 | 7969 | 9607 |
| European | Recorded Depression | Switching vs Continuation | sex_at_birth | age_at_first_AD | 0.716 | 0.694 | 0.739 | <2.2e-16 | <2.2e-16 | 17576 | 7969 | 9607 |
| European | Recorded Depression | Switching vs Continuation | sex_at_birth | sex_at_birthMale | 1.111 | 1.039 | 1.187 | 1.93e-03 | 2.70e-02 | 17576 | 7969 | 9607 |
| European | No Recorded Depression | Discontinuation vs Continuation | Alcohol_Frequency | (Intercept) | 0.787 | 0.577 | 1.075 | 1.32e-01 | 1.00e+00 | 18359 | 13151 | 5208 |
| European | No Recorded Depression | Discontinuation vs Continuation | Alcohol_Frequency | age_at_first_AD | 1.078 | 1.044 | 1.114 | 6.22e-06 | 8.71e-05 | 18359 | 13151 | 5208 |
| European | No Recorded Depression | Discontinuation vs Continuation | Alcohol_Frequency | sex_at_birthMale | 1.112 | 1.037 | 1.193 | 2.91e-03 | 4.08e-02 | 18359 | 13151 | 5208 |
| European | No Recorded Depression | Discontinuation vs Continuation | Alcohol_Frequency | Income>200k | 0.628 | 0.547 | 0.721 | 4.13e-11 | 5.78e-10 | 18359 | 13151 | 5208 |
| European | No Recorded Depression | Discontinuation vs Continuation | Alcohol_Frequency | Income100-150k | 0.577 | 0.511 | 0.652 | <2.2e-16 | <2.2e-16 | 18359 | 13151 | 5208 |
| European | No Recorded Depression | Discontinuation vs Continuation | Alcohol_Frequency | Income150-200k | 0.572 | 0.492 | 0.664 | 3.18e-13 | 4.45e-12 | 18359 | 13151 | 5208 |
| European | No Recorded Depression | Discontinuation vs Continuation | Alcohol_Frequency | Income25-35k | 0.812 | 0.704 | 0.937 | 4.27e-03 | 5.98e-02 | 18359 | 13151 | 5208 |
| European | No Recorded Depression | Discontinuation vs Continuation | Alcohol_Frequency | Income35-50k | 0.708 | 0.619 | 0.809 | 3.92e-07 | 5.49e-06 | 18359 | 13151 | 5208 |
| European | No Recorded Depression | Discontinuation vs Continuation | Alcohol_Frequency | Income50-75k | 0.744 | 0.661 | 0.837 | 9.41e-07 | 1.32e-05 | 18359 | 13151 | 5208 |
| European | No Recorded Depression | Discontinuation vs Continuation | Alcohol_Frequency | Income75-100k | 0.639 | 0.564 | 0.725 | 2.90e-12 | 4.06e-11 | 18359 | 13151 | 5208 |
| European | No Recorded Depression | Discontinuation vs Continuation | Alcohol_Frequency | Education_levelCollege Postgraduate | 0.732 | 0.556 | 0.964 | 2.65e-02 | 3.72e-01 | 18359 | 13151 | 5208 |
| European | No Recorded Depression | Discontinuation vs Continuation | Alcohol_Frequency | Education_levelCollege Undergraduate | 0.779 | 0.596 | 1.019 | 6.85e-02 | 9.59e-01 | 18359 | 13151 | 5208 |
| European | No Recorded Depression | Discontinuation vs Continuation | Alcohol_Frequency | Education_levelTwelve or GED | 0.893 | 0.677 | 1.178 | 4.24e-01 | 1.00e+00 | 18359 | 13151 | 5208 |
| European | No Recorded Depression | Discontinuation vs Continuation | Alcohol_Frequency | Alcohol_Frequency2-3 Week | 0.828 | 0.672 | 1.021 | 7.73e-02 | 1.00e+00 | 18359 | 13151 | 5208 |
| European | No Recorded Depression | Discontinuation vs Continuation | Alcohol_Frequency | Alcohol_Frequency2-4 Month | 0.872 | 0.711 | 1.070 | 1.90e-01 | 1.00e+00 | 18359 | 13151 | 5208 |
| European | No Recorded Depression | Discontinuation vs Continuation | Alcohol_Frequency | Alcohol_Frequency4plus Week | 0.810 | 0.656 | 1.000 | 4.97e-02 | 6.95e-01 | 18359 | 13151 | 5208 |
| European | No Recorded Depression | Discontinuation vs Continuation | Alcohol_Frequency | Alcohol_FrequencyMonthly or Less | 0.911 | 0.746 | 1.113 | 3.61e-01 | 1.00e+00 | 18359 | 13151 | 5208 |
| European | No Recorded Depression | Discontinuation vs Continuation | Alcohol_Frequency | Alcohol_FrequencyNever (former) | 0.949 | 0.771 | 1.168 | 6.21e-01 | 1.00e+00 | 18359 | 13151 | 5208 |
| European | No Recorded Depression | Switching vs Continuation | Alcohol_Frequency | (Intercept) | 0.466 | 0.327 | 0.663 | 2.30e-05 | 3.22e-04 | 17330 | 13151 | 4179 |
| European | No Recorded Depression | Switching vs Continuation | Alcohol_Frequency | age_at_first_AD | 0.732 | 0.706 | 0.759 | <2.2e-16 | <2.2e-16 | 17330 | 13151 | 4179 |
| European | No Recorded Depression | Switching vs Continuation | Alcohol_Frequency | sex_at_birthMale | 0.907 | 0.837 | 0.983 | 1.73e-02 | 2.42e-01 | 17330 | 13151 | 4179 |

| | | | | | | | | | | | | |
|---|---|---|---|---|---|---|---|---|---|---|---|---|
| European | No Recorded Depression | Switching vs Continuation | Alcohol_Frequency | Income>200k | 0.542 | 0.462 | 0.636 | 6.77e-14 | 9.48e-13 | 17330 | 13151 | 4179 |
| European | No Recorded Depression | Switching vs Continuation | Alcohol_Frequency | Income100-150k | 0.707 | 0.619 | 0.807 | 2.87e-07 | 4.02e-06 | 17330 | 13151 | 4179 |
| European | No Recorded Depression | Switching vs Continuation | Alcohol_Frequency | Income150-200k | 0.683 | 0.581 | 0.804 | 4.30e-06 | 6.02e-05 | 17330 | 13151 | 4179 |
| European | No Recorded Depression | Switching vs Continuation | Alcohol_Frequency | Income25-35k | 1.001 | 0.857 | 1.169 | 9.91e-01 | 1.00e+00 | 17330 | 13151 | 4179 |
| European | No Recorded Depression | Switching vs Continuation | Alcohol_Frequency | Income35-50k | 0.922 | 0.799 | 1.064 | 2.67e-01 | 1.00e+00 | 17330 | 13151 | 4179 |
| European | No Recorded Depression | Switching vs Continuation | Alcohol_Frequency | Income50-75k | 0.820 | 0.719 | 0.935 | 3.08e-03 | 4.31e-02 | 17330 | 13151 | 4179 |
| European | No Recorded Depression | Switching vs Continuation | Alcohol_Frequency | Income75-100k | 0.744 | 0.648 | 0.854 | 2.74e-05 | 3.83e-04 | 17330 | 13151 | 4179 |
| European | No Recorded Depression | Switching vs Continuation | Alcohol_Frequency | Education_levelCollege Postgraduate | 0.771 | 0.566 | 1.050 | 9.93e-02 | 1.00e+00 | 17330 | 13151 | 4179 |
| European | No Recorded Depression | Switching vs Continuation | Alcohol_Frequency | Education_levelCollege Undergraduate | 0.802 | 0.593 | 1.084 | 1.51e-01 | 1.00e+00 | 17330 | 13151 | 4179 |
| European | No Recorded Depression | Switching vs Continuation | Alcohol_Frequency | Education_levelTwelve or GED | 0.915 | 0.670 | 1.250 | 5.77e-01 | 1.00e+00 | 17330 | 13151 | 4179 |
| European | No Recorded Depression | Switching vs Continuation | Alcohol_Frequency | Alcohol_Frequency2-3 Week | 1.040 | 0.817 | 1.323 | 7.52e-01 | 1.00e+00 | 17330 | 13151 | 4179 |
| European | No Recorded Depression | Switching vs Continuation | Alcohol_Frequency | Alcohol_Frequency2-4 Month | 1.046 | 0.827 | 1.325 | 7.06e-01 | 1.00e+00 | 17330 | 13151 | 4179 |
| European | No Recorded Depression | Switching vs Continuation | Alcohol_Frequency | Alcohol_Frequency4plus Week | 1.086 | 0.851 | 1.387 | 5.07e-01 | 1.00e+00 | 17330 | 13151 | 4179 |
| European | No Recorded Depression | Switching vs Continuation | Alcohol_Frequency | Alcohol_FrequencyMonthly or Less | 1.146 | 0.909 | 1.445 | 2.49e-01 | 1.00e+00 | 17330 | 13151 | 4179 |
| European | No Recorded Depression | Switching vs Continuation | Alcohol_Frequency | Alcohol_FrequencyNever (former) | 1.288 | 1.013 | 1.638 | 3.88e-02 | 5.44e-01 | 17330 | 13151 | 4179 |
| European | No Recorded Depression | Discontinuation vs Continuation | BMI | (Intercept) | 0.688 | 0.529 | 0.894 | 5.09e-03 | 7.13e-02 | 18155 | 13070 | 5085 |
| European | No Recorded Depression | Discontinuation vs Continuation | BMI | age_at_first_AD | 1.075 | 1.041 | 1.111 | 1.07e-05 | 1.50e-04 | 18155 | 13070 | 5085 |
| European | No Recorded Depression | Discontinuation vs Continuation | BMI | sex_at_birthMale | 1.125 | 1.049 | 1.207 | 9.38e-04 | 1.31e-02 | 18155 | 13070 | 5085 |
| European | No Recorded Depression | Discontinuation vs Continuation | BMI | Income>200k | 0.593 | 0.516 | 0.680 | 1.07e-13 | 1.50e-12 | 18155 | 13070 | 5085 |
| European | No Recorded Depression | Discontinuation vs Continuation | BMI | Income100-150k | 0.558 | 0.495 | 0.630 | <2.2e-16 | <2.2e-16 | 18155 | 13070 | 5085 |
| European | No Recorded Depression | Discontinuation vs Continuation | BMI | Income150-200k | 0.542 | 0.466 | 0.630 | 1.44e-15 | 2.02e-14 | 18155 | 13070 | 5085 |
| European | No Recorded Depression | Discontinuation vs Continuation | BMI | Income25-35k | 0.790 | 0.684 | 0.911 | 1.24e-03 | 1.74e-02 | 18155 | 13070 | 5085 |
| European | No Recorded Depression | Discontinuation vs Continuation | BMI | Income35-50k | 0.683 | 0.598 | 0.781 | 2.60e-08 | 3.64e-07 | 18155 | 13070 | 5085 |
| European | No Recorded Depression | Discontinuation vs Continuation | BMI | Income50-75k | 0.739 | 0.656 | 0.832 | 5.29e-07 | 7.41e-06 | 18155 | 13070 | 5085 |
| European | No Recorded Depression | Discontinuation vs Continuation | BMI | Income75-100k | 0.623 | 0.550 | 0.707 | 1.52e-13 | 2.12e-12 | 18155 | 13070 | 5085 |
| European | No Recorded Depression | Discontinuation vs Continuation | BMI | Education_levelCollege Postgraduate | 0.729 | 0.554 | 0.959 | 2.38e-02 | 3.34e-01 | 18155 | 13070 | 5085 |
| European | No Recorded Depression | Discontinuation vs Continuation | BMI | Education_levelCollege Undergraduate | 0.785 | 0.601 | 1.025 | 7.54e-02 | 1.00e+00 | 18155 | 13070 | 5085 |
| European | No Recorded Depression | Discontinuation vs Continuation | BMI | Education_levelTwelve or GED | 0.919 | 0.697 | 1.211 | 5.47e-01 | 1.00e+00 | 18155 | 13070 | 5085 |
| European | No Recorded Depression | Discontinuation vs Continuation | BMI | BMI | 0.961 | 0.926 | 0.997 | 3.50e-02 | 4.90e-01 | 18155 | 13070 | 5085 |
| European | No Recorded Depression | Switching vs Continuation | BMI | (Intercept) | 0.490 | 0.363 | 0.660 | 2.71e-06 | 3.80e-05 | 17197 | 13070 | 4127 |
| European | No Recorded Depression | Switching vs Continuation | BMI | age_at_first_AD | 0.736 | 0.710 | 0.762 | <2.2e-16 | <2.2e-16 | 17197 | 13070 | 4127 |
| European | No Recorded Depression | Switching vs Continuation | BMI | sex_at_birthMale | 0.912 | 0.842 | 0.988 | 2.43e-02 | 3.41e-01 | 17197 | 13070 | 4127 |
| European | No Recorded Depression | Switching vs Continuation | BMI | Income>200k | 0.547 | 0.467 | 0.642 | 1.42e-13 | 1.99e-12 | 17197 | 13070 | 4127 |
| European | No Recorded Depression | Switching vs Continuation | BMI | Income100-150k | 0.702 | 0.615 | 0.801 | 1.44e-07 | 2.01e-06 | 17197 | 13070 | 4127 |
| European | No Recorded Depression | Switching vs Continuation | BMI | Income150-200k | 0.672 | 0.571 | 0.790 | 1.42e-06 | 1.99e-05 | 17197 | 13070 | 4127 |
| European | No Recorded Depression | Switching vs Continuation | BMI | Income25-35k | 0.984 | 0.843 | 1.150 | 8.43e-01 | 1.00e+00 | 17197 | 13070 | 4127 |
| European | No Recorded Depression | Switching vs Continuation | BMI | Income35-50k | 0.912 | 0.790 | 1.052 | 2.05e-01 | 1.00e+00 | 17197 | 13070 | 4127 |
| European | No Recorded Depression | Switching vs Continuation | BMI | Income50-75k | 0.810 | 0.710 | 0.924 | 1.68e-03 | 2.35e-02 | 17197 | 13070 | 4127 |
| European | No Recorded Depression | Switching vs Continuation | BMI | Income75-100k | 0.728 | 0.635 | 0.836 | 6.56e-06 | 9.18e-05 | 17197 | 13070 | 4127 |
| European | No Recorded Depression | Switching vs Continuation | BMI | Education_levelCollege Postgraduate | 0.834 | 0.611 | 1.138 | 2.51e-01 | 1.00e+00 | 17197 | 13070 | 4127 |
| European | No Recorded Depression | Switching vs Continuation | BMI | Education_levelCollege Undergraduate | 0.853 | 0.630 | 1.155 | 3.05e-01 | 1.00e+00 | 17197 | 13070 | 4127 |
| European | No Recorded Depression | Switching vs Continuation | BMI | Education_levelTwelve or GED | 0.991 | 0.724 | 1.356 | 9.54e-01 | 1.00e+00 | 17197 | 13070 | 4127 |
| European | No Recorded Depression | Switching vs Continuation | BMI | BMI | 1.101 | 1.060 | 1.143 | 6.22e-07 | 8.70e-06 | 17197 | 13070 | 4127 |
| European | No Recorded Depression | Discontinuation vs Continuation | Education_level | (Intercept) | 0.708 | 0.549 | 0.915 | 8.17e-03 | 1.14e-01 | 18562 | 13287 | 5275 |
| European | No Recorded Depression | Discontinuation vs Continuation | Education_level | age_at_first_AD | 1.076 | 1.042 | 1.111 | 7.54e-06 | 1.06e-04 | 18562 | 13287 | 5275 |
| European | No Recorded Depression | Discontinuation vs Continuation | Education_level | sex_at_birthMale | 1.109 | 1.035 | 1.189 | 3.22e-03 | 4.51e-02 | 18562 | 13287 | 5275 |
| European | No Recorded Depression | Discontinuation vs Continuation | Education_level | Income>200k | 0.605 | 0.528 | 0.692 | 2.49e-13 | 3.48e-12 | 18562 | 13287 | 5275 |
| European | No Recorded Depression | Discontinuation vs Continuation | Education_level | Income100-150k | 0.560 | 0.497 | 0.631 | <2.2e-16 | <2.2e-16 | 18562 | 13287 | 5275 |
| European | No Recorded Depression | Discontinuation vs Continuation | Education_level | Income150-200k | 0.546 | 0.471 | 0.633 | 9.81e-16 | 1.37e-14 | 18562 | 13287 | 5275 |
| European | No Recorded Depression | Discontinuation vs Continuation | Education_level | Income25-35k | 0.797 | 0.692 | 0.918 | 1.65e-03 | 2.31e-02 | 18562 | 13287 | 5275 |
| European | No Recorded Depression | Discontinuation vs Continuation | Education_level | Income35-50k | 0.695 | 0.609 | 0.792 | 5.91e-08 | 8.27e-07 | 18562 | 13287 | 5275 |
| European | No Recorded Depression | Discontinuation vs Continuation | Education_level | Income50-75k | 0.730 | 0.650 | 0.820 | 1.25e-07 | 1.75e-06 | 18562 | 13287 | 5275 |
| European | No Recorded Depression | Discontinuation vs Continuation | Education_level | Income75-100k | 0.624 | 0.551 | 0.706 | 7.23e-14 | 1.01e-12 | 18562 | 13287 | 5275 |
| European | No Recorded Depression | Discontinuation vs Continuation | Education_level | Education_levelCollege Postgraduate | 0.734 | 0.562 | 0.960 | 2.37e-02 | 3.32e-01 | 18562 | 13287 | 5275 |
| European | No Recorded Depression | Discontinuation vs Continuation | Education_level | Education_levelCollege Undergraduate | 0.783 | 0.603 | 1.016 | 6.52e-02 | 9.13e-01 | 18562 | 13287 | 5275 |
| European | No Recorded Depression | Discontinuation vs Continuation | Education_level | Education_levelTwelve or GED | 0.903 | 0.689 | 1.183 | 4.58e-01 | 1.00e+00 | 18562 | 13287 | 5275 |
| European | No Recorded Depression | Switching vs Continuation | Education_level | (Intercept) | 0.507 | 0.379 | 0.678 | 4.71e-06 | 6.59e-05 | 17516 | 13287 | 4229 |
| European | No Recorded Depression | Switching vs Continuation | Education_level | age_at_first_AD | 0.737 | 0.712 | 0.763 | <2.2e-16 | <2.2e-16 | 17516 | 13287 | 4229 |
| European | No Recorded Depression | Switching vs Continuation | Education_level | sex_at_birthMale | 0.906 | 0.837 | 0.981 | 1.45e-02 | 2.02e-01 | 17516 | 13287 | 4229 |
| European | No Recorded Depression | Switching vs Continuation | Education_level | Income>200k | 0.519 | 0.443 | 0.607 | 2.29e-16 | 3.21e-15 | 17516 | 13287 | 4229 |
| European | No Recorded Depression | Switching vs Continuation | Education_level | Income100-150k | 0.684 | 0.601 | 0.780 | 1.12e-08 | 1.57e-07 | 17516 | 13287 | 4229 |
| European | No Recorded Depression | Switching vs Continuation | Education_level | Income150-200k | 0.660 | 0.563 | 0.774 | 3.18e-07 | 4.45e-06 | 17516 | 13287 | 4229 |
| European | No Recorded Depression | Switching vs Continuation | Education_level | Income25-35k | 0.982 | 0.842 | 1.145 | 8.15e-01 | 1.00e+00 | 17516 | 13287 | 4229 |
| European | No Recorded Depression | Switching vs Continuation | Education_level | Income35-50k | 0.907 | 0.787 | 1.045 | 1.76e-01 | 1.00e+00 | 17516 | 13287 | 4229 |
| European | No Recorded Depression | Switching vs Continuation | Education_level | Income50-75k | 0.800 | 0.703 | 0.911 | 7.73e-04 | 1.08e-02 | 17516 | 13287 | 4229 |
| European | No Recorded Depression | Switching vs Continuation | Education_level | Income75-100k | 0.723 | 0.631 | 0.828 | 2.88e-06 | 4.03e-05 | 17516 | 13287 | 4229 |

| | | | | | | | | | | | | |
|---|---|---|---|---|---|---|---|---|---|---|---|---|
| European | No Recorded Depression | Switching vs Continuation | Education_level | Education_levelCollege Postgraduate | 0.807 | 0.596 | 1.093 | 1.65e-01 | 1.00e+00 | 17516 | 13287 | 4229 |
| European | No Recorded Depression | Switching vs Continuation | Education_level | Education_levelCollege Undergraduate | 0.840 | 0.625 | 1.129 | 2.47e-01 | 1.00e+00 | 17516 | 13287 | 4229 |
| European | No Recorded Depression | Switching vs Continuation | Education_level | Education_levelTwelve or GED | 0.967 | 0.712 | 1.314 | 8.30e-01 | 1.00e+00 | 17516 | 13287 | 4229 |
| European | No Recorded Depression | Discontinuation vs Continuation | Income | (Intercept) | 0.708 | 0.549 | 0.915 | 8.17e-03 | 1.14e-01 | 18562 | 13287 | 5275 |
| European | No Recorded Depression | Discontinuation vs Continuation | Income | age_at_first_AD | 1.076 | 1.042 | 1.111 | 7.54e-06 | 1.06e-04 | 18562 | 13287 | 5275 |
| European | No Recorded Depression | Discontinuation vs Continuation | Income | sex_at_birthMale | 1.109 | 1.035 | 1.189 | 3.22e-03 | 4.51e-02 | 18562 | 13287 | 5275 |
| European | No Recorded Depression | Discontinuation vs Continuation | Income | Income>200k | 0.605 | 0.528 | 0.692 | 2.49e-13 | 3.48e-12 | 18562 | 13287 | 5275 |
| European | No Recorded Depression | Discontinuation vs Continuation | Income | Income100-150k | 0.560 | 0.497 | 0.631 | <2.2e-16 | <2.2e-16 | 18562 | 13287 | 5275 |
| European | No Recorded Depression | Discontinuation vs Continuation | Income | Income150-200k | 0.546 | 0.471 | 0.633 | 9.81e-16 | 1.37e-14 | 18562 | 13287 | 5275 |
| European | No Recorded Depression | Discontinuation vs Continuation | Income | Income25-35k | 0.797 | 0.692 | 0.918 | 1.65e-03 | 2.31e-02 | 18562 | 13287 | 5275 |
| European | No Recorded Depression | Discontinuation vs Continuation | Income | Income35-50k | 0.695 | 0.609 | 0.792 | 5.91e-08 | 8.27e-07 | 18562 | 13287 | 5275 |
| European | No Recorded Depression | Discontinuation vs Continuation | Income | Income50-75k | 0.730 | 0.650 | 0.820 | 1.25e-07 | 1.75e-06 | 18562 | 13287 | 5275 |
| European | No Recorded Depression | Discontinuation vs Continuation | Income | Income75-100k | 0.624 | 0.551 | 0.706 | 7.23e-14 | 1.01e-12 | 18562 | 13287 | 5275 |
| European | No Recorded Depression | Discontinuation vs Continuation | Income | Education_levelCollege Postgraduate | 0.734 | 0.562 | 0.960 | 2.37e-02 | 3.32e-01 | 18562 | 13287 | 5275 |
| European | No Recorded Depression | Discontinuation vs Continuation | Income | Education_levelCollege Undergraduate | 0.783 | 0.603 | 1.016 | 6.52e-02 | 9.13e-01 | 18562 | 13287 | 5275 |
| European | No Recorded Depression | Discontinuation vs Continuation | Income | Education_levelTwelve or GED | 0.903 | 0.689 | 1.183 | 4.58e-01 | 1.00e+00 | 18562 | 13287 | 5275 |
| European | No Recorded Depression | Switching vs Continuation | Income | (Intercept) | 0.507 | 0.379 | 0.678 | 4.71e-06 | 6.59e-05 | 17516 | 13287 | 4229 |
| European | No Recorded Depression | Switching vs Continuation | Income | age_at_first_AD | 0.737 | 0.712 | 0.763 | <2.2e-16 | <2.2e-16 | 17516 | 13287 | 4229 |
| European | No Recorded Depression | Switching vs Continuation | Income | sex_at_birthMale | 0.906 | 0.837 | 0.981 | 1.45e-02 | 2.02e-01 | 17516 | 13287 | 4229 |
| European | No Recorded Depression | Switching vs Continuation | Income | Income>200k | 0.519 | 0.443 | 0.607 | 2.29e-16 | 3.21e-15 | 17516 | 13287 | 4229 |
| European | No Recorded Depression | Switching vs Continuation | Income | Income100-150k | 0.684 | 0.601 | 0.780 | 1.12e-08 | 1.57e-07 | 17516 | 13287 | 4229 |
| European | No Recorded Depression | Switching vs Continuation | Income | Income150-200k | 0.660 | 0.563 | 0.774 | 3.18e-07 | 4.45e-06 | 17516 | 13287 | 4229 |
| European | No Recorded Depression | Switching vs Continuation | Income | Income25-35k | 0.982 | 0.842 | 1.145 | 8.15e-01 | 1.00e+00 | 17516 | 13287 | 4229 |
| European | No Recorded Depression | Switching vs Continuation | Income | Income35-50k | 0.907 | 0.787 | 1.045 | 1.76e-01 | 1.00e+00 | 17516 | 13287 | 4229 |
| European | No Recorded Depression | Switching vs Continuation | Income | Income50-75k | 0.800 | 0.703 | 0.911 | 7.73e-04 | 1.08e-02 | 17516 | 13287 | 4229 |
| European | No Recorded Depression | Switching vs Continuation | Income | Income75-100k | 0.723 | 0.631 | 0.828 | 2.88e-06 | 4.03e-05 | 17516 | 13287 | 4229 |
| European | No Recorded Depression | Switching vs Continuation | Income | Education_levelCollege Postgraduate | 0.807 | 0.596 | 1.093 | 1.65e-01 | 1.00e+00 | 17516 | 13287 | 4229 |
| European | No Recorded Depression | Switching vs Continuation | Income | Education_levelCollege Undergraduate | 0.840 | 0.625 | 1.129 | 2.47e-01 | 1.00e+00 | 17516 | 13287 | 4229 |
| European | No Recorded Depression | Switching vs Continuation | Income | Education_levelTwelve or GED | 0.967 | 0.712 | 1.314 | 8.30e-01 | 1.00e+00 | 17516 | 13287 | 4229 |
| European | No Recorded Depression | Discontinuation vs Continuation | Insomnia | (Intercept) | 0.747 | 0.578 | 0.965 | 2.55e-02 | 3.57e-01 | 18562 | 13287 | 5275 |
| European | No Recorded Depression | Discontinuation vs Continuation | Insomnia | age_at_first_AD | 1.092 | 1.057 | 1.128 | 7.98e-08 | 1.12e-06 | 18562 | 13287 | 5275 |
| European | No Recorded Depression | Discontinuation vs Continuation | Insomnia | sex_at_birthMale | 1.122 | 1.047 | 1.202 | 1.12e-03 | 1.57e-02 | 18562 | 13287 | 5275 |
| European | No Recorded Depression | Discontinuation vs Continuation | Insomnia | Income>200k | 0.607 | 0.530 | 0.694 | 3.91e-13 | 5.47e-12 | 18562 | 13287 | 5275 |
| European | No Recorded Depression | Discontinuation vs Continuation | Insomnia | Income100-150k | 0.562 | 0.499 | 0.634 | <2.2e-16 | <2.2e-16 | 18562 | 13287 | 5275 |
| European | No Recorded Depression | Discontinuation vs Continuation | Insomnia | Income150-200k | 0.548 | 0.473 | 0.636 | 1.84e-15 | 2.58e-14 | 18562 | 13287 | 5275 |
| European | No Recorded Depression | Discontinuation vs Continuation | Insomnia | Income25-35k | 0.796 | 0.691 | 0.917 | 1.56e-03 | 2.18e-02 | 18562 | 13287 | 5275 |
| European | No Recorded Depression | Discontinuation vs Continuation | Insomnia | Income35-50k | 0.697 | 0.611 | 0.796 | 8.51e-08 | 1.19e-06 | 18562 | 13287 | 5275 |
| European | No Recorded Depression | Discontinuation vs Continuation | Insomnia | Income50-75k | 0.735 | 0.654 | 0.826 | 2.48e-07 | 3.47e-06 | 18562 | 13287 | 5275 |
| European | No Recorded Depression | Discontinuation vs Continuation | Insomnia | Income75-100k | 0.627 | 0.554 | 0.710 | 1.57e-13 | 2.20e-12 | 18562 | 13287 | 5275 |
| European | No Recorded Depression | Discontinuation vs Continuation | Insomnia | Education_levelCollege Postgraduate | 0.732 | 0.560 | 0.958 | 2.28e-02 | 3.19e-01 | 18562 | 13287 | 5275 |
| European | No Recorded Depression | Discontinuation vs Continuation | Insomnia | Education_levelCollege Undergraduate | 0.780 | 0.601 | 1.013 | 6.24e-02 | 8.74e-01 | 18562 | 13287 | 5275 |
| European | No Recorded Depression | Discontinuation vs Continuation | Insomnia | Education_levelTwelve or GED | 0.906 | 0.691 | 1.187 | 4.72e-01 | 1.00e+00 | 18562 | 13287 | 5275 |
| European | No Recorded Depression | Discontinuation vs Continuation | Insomnia | Insomnia | 0.701 | 0.641 | 0.765 | 3.08e-15 | 4.31e-14 | 18562 | 13287 | 5275 |
| European | No Recorded Depression | Switching vs Continuation | Insomnia | (Intercept) | 0.449 | 0.335 | 0.602 | 8.50e-08 | 1.19e-06 | 17516 | 13287 | 4229 |
| European | No Recorded Depression | Switching vs Continuation | Insomnia | age_at_first_AD | 0.719 | 0.694 | 0.745 | <2.2e-16 | <2.2e-16 | 17516 | 13287 | 4229 |
| European | No Recorded Depression | Switching vs Continuation | Insomnia | sex_at_birthMale | 0.893 | 0.825 | 0.968 | 5.62e-03 | 7.87e-02 | 17516 | 13287 | 4229 |
| European | No Recorded Depression | Switching vs Continuation | Insomnia | Income>200k | 0.515 | 0.440 | 0.603 | <2.2e-16 | 2.04e-15 | 17516 | 13287 | 4229 |
| European | No Recorded Depression | Switching vs Continuation | Insomnia | Income100-150k | 0.679 | 0.596 | 0.774 | 6.45e-09 | 9.03e-08 | 17516 | 13287 | 4229 |
| European | No Recorded Depression | Switching vs Continuation | Insomnia | Income150-200k | 0.657 | 0.560 | 0.771 | 2.60e-07 | 3.64e-06 | 17516 | 13287 | 4229 |
| European | No Recorded Depression | Switching vs Continuation | Insomnia | Income25-35k | 0.984 | 0.844 | 1.148 | 8.41e-01 | 1.00e+00 | 17516 | 13287 | 4229 |
| European | No Recorded Depression | Switching vs Continuation | Insomnia | Income35-50k | 0.900 | 0.781 | 1.038 | 1.47e-01 | 1.00e+00 | 17516 | 13287 | 4229 |
| European | No Recorded Depression | Switching vs Continuation | Insomnia | Income50-75k | 0.791 | 0.694 | 0.902 | 4.35e-04 | 6.09e-03 | 17516 | 13287 | 4229 |
| European | No Recorded Depression | Switching vs Continuation | Insomnia | Income75-100k | 0.719 | 0.627 | 0.824 | 2.15e-06 | 3.01e-05 | 17516 | 13287 | 4229 |
| European | No Recorded Depression | Switching vs Continuation | Insomnia | Education_levelCollege Postgraduate | 0.827 | 0.610 | 1.122 | 2.22e-01 | 1.00e+00 | 17516 | 13287 | 4229 |
| European | No Recorded Depression | Switching vs Continuation | Insomnia | Education_levelCollege Undergraduate | 0.858 | 0.637 | 1.154 | 3.11e-01 | 1.00e+00 | 17516 | 13287 | 4229 |
| European | No Recorded Depression | Switching vs Continuation | Insomnia | Education_levelTwelve or GED | 0.975 | 0.717 | 1.326 | 8.70e-01 | 1.00e+00 | 17516 | 13287 | 4229 |
| European | No Recorded Depression | Switching vs Continuation | Insomnia | Insomnia | 1.626 | 1.496 | 1.768 | <2.2e-16 | <2.2e-16 | 17516 | 13287 | 4229 |
| European | No Recorded Depression | Discontinuation vs Continuation | MDD CIDI-sf | (Intercept) | 0.137 | 0.031 | 0.609 | 9.04e-03 | 1.27e-01 | 3829 | 2796 | 1033 |
| European | No Recorded Depression | Discontinuation vs Continuation | MDD CIDI-sf | age_at_first_AD | 1.112 | 1.027 | 1.204 | 8.68e-03 | 1.22e-01 | 3829 | 2796 | 1033 |
| European | No Recorded Depression | Discontinuation vs Continuation | MDD CIDI-sf | sex_at_birthMale | 1.142 | 0.973 | 1.340 | 1.05e-01 | 1.00e+00 | 3829 | 2796 | 1033 |
| European | No Recorded Depression | Discontinuation vs Continuation | MDD CIDI-sf | Income>200k | 0.665 | 0.480 | 0.922 | 1.43e-02 | 2.00e-01 | 3829 | 2796 | 1033 |
| European | No Recorded Depression | Discontinuation vs Continuation | MDD CIDI-sf | Income100-150k | 0.532 | 0.395 | 0.718 | 3.56e-05 | 4.98e-04 | 3829 | 2796 | 1033 |
| European | No Recorded Depression | Discontinuation vs Continuation | MDD CIDI-sf | Income150-200k | 0.716 | 0.508 | 1.010 | 5.68e-02 | 7.95e-01 | 3829 | 2796 | 1033 |
| European | No Recorded Depression | Discontinuation vs Continuation | MDD CIDI-sf | Income25-35k | 0.714 | 0.493 | 1.032 | 7.29e-02 | 1.00e+00 | 3829 | 2796 | 1033 |
| European | No Recorded Depression | Discontinuation vs Continuation | MDD CIDI-sf | Income35-50k | 0.711 | 0.513 | 0.985 | 4.03e-02 | 5.64e-01 | 3829 | 2796 | 1033 |
| European | No Recorded Depression | Discontinuation vs Continuation | MDD CIDI-sf | Income50-75k | 0.810 | 0.606 | 1.083 | 1.56e-01 | 1.00e+00 | 3829 | 2796 | 1033 |

| | | | | | | | | | | | | |
|---|---|---|---|---|---|---|---|---|---|---|---|---|
| European | No Recorded Depression | Discontinuation vs Continuation | MDD CIDI-sf | Income75-100k | 0.605 | 0.444 | 0.824 | 1.41e-03 | 1.98e-02 | 3829 | 2796 | 1033 |
| European | No Recorded Depression | Discontinuation vs Continuation | MDD CIDI-sf | Education_levelCollege Postgraduate | 2.798 | 0.630 | 12.420 | 1.76e-01 | 1.00e+00 | 3829 | 2796 | 1033 |
| European | No Recorded Depression | Discontinuation vs Continuation | MDD CIDI-sf | Education_levelCollege Undergraduate | 3.564 | 0.807 | 15.740 | 9.34e-02 | 1.00e+00 | 3829 | 2796 | 1033 |
| European | No Recorded Depression | Discontinuation vs Continuation | MDD CIDI-sf | Education_levelTwelve or GED | 3.139 | 0.697 | 14.130 | 1.36e-01 | 1.00e+00 | 3829 | 2796 | 1033 |
| European | No Recorded Depression | Discontinuation vs Continuation | MDD CIDI-sf | MDD CIDI-sf | 1.289 | 1.109 | 1.498 | 9.22e-04 | 1.29e-02 | 3829 | 2796 | 1033 |
| European | No Recorded Depression | Switching vs Continuation | MDD CIDI-sf | (Intercept) | 0.257 | 0.095 | 0.695 | 7.47e-03 | 1.05e-01 | 3697 | 2796 | 901 |
| European | No Recorded Depression | Switching vs Continuation | MDD CIDI-sf | age_at_first_AD | 0.683 | 0.629 | 0.743 | <2.2e-16 | <2.2e-16 | 3697 | 2796 | 901 |
| European | No Recorded Depression | Switching vs Continuation | MDD CIDI-sf | sex_at_birthMale | 1.005 | 0.838 | 1.206 | 9.54e-01 | 1.00e+00 | 3697 | 2796 | 901 |
| European | No Recorded Depression | Switching vs Continuation | MDD CIDI-sf | Income>200k | 0.569 | 0.399 | 0.811 | 1.84e-03 | 2.57e-02 | 3697 | 2796 | 901 |
| European | No Recorded Depression | Switching vs Continuation | MDD CIDI-sf | Income100-150k | 0.583 | 0.427 | 0.795 | 6.63e-04 | 9.28e-03 | 3697 | 2796 | 901 |
| European | No Recorded Depression | Switching vs Continuation | MDD CIDI-sf | Income150-200k | 0.617 | 0.426 | 0.893 | 1.06e-02 | 1.48e-01 | 3697 | 2796 | 901 |
| European | No Recorded Depression | Switching vs Continuation | MDD CIDI-sf | Income25-35k | 0.970 | 0.669 | 1.405 | 8.71e-01 | 1.00e+00 | 3697 | 2796 | 901 |
| European | No Recorded Depression | Switching vs Continuation | MDD CIDI-sf | Income35-50k | 0.724 | 0.515 | 1.018 | 6.33e-02 | 8.86e-01 | 3697 | 2796 | 901 |
| European | No Recorded Depression | Switching vs Continuation | MDD CIDI-sf | Income50-75k | 0.706 | 0.517 | 0.964 | 2.87e-02 | 4.02e-01 | 3697 | 2796 | 901 |
| European | No Recorded Depression | Switching vs Continuation | MDD CIDI-sf | Income75-100k | 0.757 | 0.551 | 1.039 | 8.46e-02 | 1.00e+00 | 3697 | 2796 | 901 |
| European | No Recorded Depression | Switching vs Continuation | MDD CIDI-sf | Education_levelCollege Postgraduate | 1.230 | 0.456 | 3.316 | 6.83e-01 | 1.00e+00 | 3697 | 2796 | 901 |
| European | No Recorded Depression | Switching vs Continuation | MDD CIDI-sf | Education_levelCollege Undergraduate | 1.358 | 0.508 | 3.630 | 5.41e-01 | 1.00e+00 | 3697 | 2796 | 901 |
| European | No Recorded Depression | Switching vs Continuation | MDD CIDI-sf | Education_levelTwelve or GED | 1.432 | 0.519 | 3.952 | 4.88e-01 | 1.00e+00 | 3697 | 2796 | 901 |
| European | No Recorded Depression | Switching vs Continuation | MDD CIDI-sf | MDD CIDI-sf | 1.871 | 1.591 | 2.201 | 3.69e-14 | 5.17e-13 | 3697 | 2796 | 901 |
| European | No Recorded Depression | Discontinuation vs Continuation | MDD_symptom_count | (Intercept) | 0.270 | 0.090 | 0.815 | 2.01e-02 | 2.82e-01 | 4798 | 3506 | 1292 |
| European | No Recorded Depression | Discontinuation vs Continuation | MDD_symptom_count | age_at_first_AD | 1.116 | 1.039 | 1.198 | 2.47e-03 | 3.46e-02 | 4798 | 3506 | 1292 |
| European | No Recorded Depression | Discontinuation vs Continuation | MDD_symptom_count | sex_at_birthMale | 1.177 | 1.021 | 1.358 | 2.49e-02 | 3.49e-01 | 4798 | 3506 | 1292 |
| European | No Recorded Depression | Discontinuation vs Continuation | MDD_symptom_count | Income>200k | 0.723 | 0.539 | 0.969 | 3.00e-02 | 4.20e-01 | 4798 | 3506 | 1292 |
| European | No Recorded Depression | Discontinuation vs Continuation | MDD_symptom_count | Income100-150k | 0.574 | 0.439 | 0.750 | 4.63e-05 | 6.48e-04 | 4798 | 3506 | 1292 |
| European | No Recorded Depression | Discontinuation vs Continuation | MDD_symptom_count | Income150-200k | 0.699 | 0.513 | 0.953 | 2.34e-02 | 3.28e-01 | 4798 | 3506 | 1292 |
| European | No Recorded Depression | Discontinuation vs Continuation | MDD_symptom_count | Income25-35k | 0.753 | 0.543 | 1.044 | 8.88e-02 | 1.00e+00 | 4798 | 3506 | 1292 |
| European | No Recorded Depression | Discontinuation vs Continuation | MDD_symptom_count | Income35-50k | 0.755 | 0.564 | 1.011 | 5.95e-02 | 8.33e-01 | 4798 | 3506 | 1292 |
| European | No Recorded Depression | Discontinuation vs Continuation | MDD_symptom_count | Income50-75k | 0.846 | 0.652 | 1.098 | 2.08e-01 | 1.00e+00 | 4798 | 3506 | 1292 |
| European | No Recorded Depression | Discontinuation vs Continuation | MDD_symptom_count | Income75-100k | 0.625 | 0.474 | 0.823 | 8.47e-04 | 1.19e-02 | 4798 | 3506 | 1292 |
| European | No Recorded Depression | Discontinuation vs Continuation | MDD_symptom_count | Education_levelCollege Postgraduate | 1.631 | 0.540 | 4.930 | 3.86e-01 | 1.00e+00 | 4798 | 3506 | 1292 |
| European | No Recorded Depression | Discontinuation vs Continuation | MDD_symptom_count | Education_levelCollege Undergraduate | 1.919 | 0.639 | 5.768 | 2.46e-01 | 1.00e+00 | 4798 | 3506 | 1292 |
| European | No Recorded Depression | Discontinuation vs Continuation | MDD_symptom_count | Education_levelTwelve or GED | 1.678 | 0.546 | 5.153 | 3.66e-01 | 1.00e+00 | 4798 | 3506 | 1292 |
| European | No Recorded Depression | Discontinuation vs Continuation | MDD_symptom_count | MDD_symptom_count | 1.106 | 1.036 | 1.180 | 2.41e-03 | 3.37e-02 | 4798 | 3506 | 1292 |
| European | No Recorded Depression | Switching vs Continuation | MDD_symptom_count | (Intercept) | 0.415 | 0.171 | 1.004 | 5.11e-02 | 7.15e-01 | 4636 | 3506 | 1130 |
| European | No Recorded Depression | Switching vs Continuation | MDD_symptom_count | age_at_first_AD | 0.689 | 0.640 | 0.743 | <2.2e-16 | <2.2e-16 | 4636 | 3506 | 1130 |
| European | No Recorded Depression | Switching vs Continuation | MDD_symptom_count | sex_at_birthMale | 1.000 | 0.850 | 1.177 | 9.98e-01 | 1.00e+00 | 4636 | 3506 | 1130 |
| European | No Recorded Depression | Switching vs Continuation | MDD_symptom_count | Income>200k | 0.663 | 0.482 | 0.913 | 1.18e-02 | 1.66e-01 | 4636 | 3506 | 1130 |
| European | No Recorded Depression | Switching vs Continuation | MDD_symptom_count | Income100-150k | 0.686 | 0.519 | 0.906 | 7.83e-03 | 1.10e-01 | 4636 | 3506 | 1130 |
| European | No Recorded Depression | Switching vs Continuation | MDD_symptom_count | Income150-200k | 0.670 | 0.480 | 0.936 | 1.87e-02 | 2.62e-01 | 4636 | 3506 | 1130 |
| European | No Recorded Depression | Switching vs Continuation | MDD_symptom_count | Income25-35k | 0.962 | 0.689 | 1.343 | 8.19e-01 | 1.00e+00 | 4636 | 3506 | 1130 |
| European | No Recorded Depression | Switching vs Continuation | MDD_symptom_count | Income35-50k | 0.827 | 0.609 | 1.124 | 2.25e-01 | 1.00e+00 | 4636 | 3506 | 1130 |
| European | No Recorded Depression | Switching vs Continuation | MDD_symptom_count | Income50-75k | 0.763 | 0.576 | 1.010 | 5.89e-02 | 8.25e-01 | 4636 | 3506 | 1130 |
| European | No Recorded Depression | Switching vs Continuation | MDD_symptom_count | Income75-100k | 0.851 | 0.641 | 1.130 | 2.65e-01 | 1.00e+00 | 4636 | 3506 | 1130 |
| European | No Recorded Depression | Switching vs Continuation | MDD_symptom_count | Education_levelCollege Postgraduate | 1.005 | 0.414 | 2.441 | 9.91e-01 | 1.00e+00 | 4636 | 3506 | 1130 |
| European | No Recorded Depression | Switching vs Continuation | MDD_symptom_count | Education_levelCollege Undergraduate | 1.129 | 0.469 | 2.719 | 7.86e-01 | 1.00e+00 | 4636 | 3506 | 1130 |
| European | No Recorded Depression | Switching vs Continuation | MDD_symptom_count | Education_levelTwelve or GED | 1.190 | 0.480 | 2.949 | 7.07e-01 | 1.00e+00 | 4636 | 3506 | 1130 |
| European | No Recorded Depression | Switching vs Continuation | MDD_symptom_count | MDD_symptom_count | 1.324 | 1.233 | 1.421 | 7.64e-15 | 1.07e-13 | 4636 | 3506 | 1130 |
| European | No Recorded Depression | Discontinuation vs Continuation | Migraine | (Intercept) | 0.743 | 0.575 | 0.960 | 2.31e-02 | 3.24e-01 | 18562 | 13287 | 5275 |
| European | No Recorded Depression | Discontinuation vs Continuation | Migraine | age_at_first_AD | 1.069 | 1.036 | 1.104 | 3.92e-05 | 5.49e-04 | 18562 | 13287 | 5275 |
| European | No Recorded Depression | Discontinuation vs Continuation | Migraine | sex_at_birthMale | 1.083 | 1.010 | 1.161 | 2.42e-02 | 3.39e-01 | 18562 | 13287 | 5275 |
| European | No Recorded Depression | Discontinuation vs Continuation | Migraine | Income>200k | 0.609 | 0.532 | 0.697 | 5.51e-13 | 7.72e-12 | 18562 | 13287 | 5275 |
| European | No Recorded Depression | Discontinuation vs Continuation | Migraine | Income100-150k | 0.564 | 0.501 | 0.636 | <2.2e-16 | <2.2e-16 | 18562 | 13287 | 5275 |
| European | No Recorded Depression | Discontinuation vs Continuation | Migraine | Income150-200k | 0.552 | 0.476 | 0.640 | 3.60e-15 | 5.04e-14 | 18562 | 13287 | 5275 |
| European | No Recorded Depression | Discontinuation vs Continuation | Migraine | Income25-35k | 0.801 | 0.695 | 0.922 | 2.06e-03 | 2.88e-02 | 18562 | 13287 | 5275 |
| European | No Recorded Depression | Discontinuation vs Continuation | Migraine | Income35-50k | 0.699 | 0.613 | 0.798 | 1.04e-07 | 1.45e-06 | 18562 | 13287 | 5275 |
| European | No Recorded Depression | Discontinuation vs Continuation | Migraine | Income50-75k | 0.738 | 0.656 | 0.829 | 3.28e-07 | 4.59e-06 | 18562 | 13287 | 5275 |
| European | No Recorded Depression | Discontinuation vs Continuation | Migraine | Income75-100k | 0.630 | 0.557 | 0.713 | 2.68e-13 | 3.75e-12 | 18562 | 13287 | 5275 |
| European | No Recorded Depression | Discontinuation vs Continuation | Migraine | Education_levelCollege Postgraduate | 0.748 | 0.572 | 0.977 | 3.33e-02 | 4.67e-01 | 18562 | 13287 | 5275 |
| European | No Recorded Depression | Discontinuation vs Continuation | Migraine | Education_levelCollege Undergraduate | 0.794 | 0.612 | 1.031 | 8.38e-02 | 1.00e+00 | 18562 | 13287 | 5275 |
| European | No Recorded Depression | Discontinuation vs Continuation | Migraine | Education_levelTwelve or GED | 0.916 | 0.699 | 1.200 | 5.22e-01 | 1.00e+00 | 18562 | 13287 | 5275 |
| European | No Recorded Depression | Discontinuation vs Continuation | Migraine | Migraine | 0.767 | 0.709 | 0.829 | 2.69e-11 | 3.77e-10 | 18562 | 13287 | 5275 |
| European | No Recorded Depression | Switching vs Continuation | Migraine | (Intercept) | 0.468 | 0.350 | 0.627 | 3.42e-07 | 4.78e-06 | 17516 | 13287 | 4229 |
| European | No Recorded Depression | Switching vs Continuation | Migraine | age_at_first_AD | 0.745 | 0.719 | 0.771 | <2.2e-16 | <2.2e-16 | 17516 | 13287 | 4229 |
| European | No Recorded Depression | Switching vs Continuation | Migraine | sex_at_birthMale | 0.942 | 0.870 | 1.021 | 1.46e-01 | 1.00e+00 | 17516 | 13287 | 4229 |
| European | No Recorded Depression | Switching vs Continuation | Migraine | Income>200k | 0.515 | 0.440 | 0.603 | <2.2e-16 | 1.87e-15 | 17516 | 13287 | 4229 |
| European | No Recorded Depression | Switching vs Continuation | Migraine | Income100-150k | 0.678 | 0.595 | 0.773 | 5.56e-09 | 7.78e-08 | 17516 | 13287 | 4229 |

| | | | | | | | | | | | | |
|---|---|---|---|---|---|---|---|---|---|---|---|---|
| European | No Recorded Depression | Switching vs Continuation | Migraine | Income150-200k | 0.654 | 0.558 | 0.767 | 1.85e-07 | 2.59e-06 | 17516 | 13287 | 4229 |
| European | No Recorded Depression | Switching vs Continuation | Migraine | Income25-35k | 0.977 | 0.837 | 1.139 | 7.62e-01 | 1.00e+00 | 17516 | 13287 | 4229 |
| European | No Recorded Depression | Switching vs Continuation | Migraine | Income35-50k | 0.902 | 0.783 | 1.040 | 1.57e-01 | 1.00e+00 | 17516 | 13287 | 4229 |
| European | No Recorded Depression | Switching vs Continuation | Migraine | Income50-75k | 0.790 | 0.694 | 0.900 | 3.95e-04 | 5.52e-03 | 17516 | 13287 | 4229 |
| European | No Recorded Depression | Switching vs Continuation | Migraine | Income75-100k | 0.712 | 0.621 | 0.816 | 1.09e-06 | 1.53e-05 | 17516 | 13287 | 4229 |
| European | No Recorded Depression | Switching vs Continuation | Migraine | Education_levelCollege Postgraduate | 0.782 | 0.577 | 1.060 | 1.13e-01 | 1.00e+00 | 17516 | 13287 | 4229 |
| European | No Recorded Depression | Switching vs Continuation | Migraine | Education_levelCollege Undergraduate | 0.816 | 0.607 | 1.098 | 1.79e-01 | 1.00e+00 | 17516 | 13287 | 4229 |
| European | No Recorded Depression | Switching vs Continuation | Migraine | Education_levelTwelve or GED | 0.943 | 0.694 | 1.281 | 7.06e-01 | 1.00e+00 | 17516 | 13287 | 4229 |
| European | No Recorded Depression | Switching vs Continuation | Migraine | Migraine | 1.418 | 1.314 | 1.531 | <2.2e-16 | <2.2e-16 | 17516 | 13287 | 4229 |
| European | No Recorded Depression | Discontinuation vs Continuation | Recorded Anxiety | (Intercept) | 0.897 | 0.693 | 1.163 | 4.12e-01 | 1.00e+00 | 18562 | 13287 | 5275 |
| European | No Recorded Depression | Discontinuation vs Continuation | Recorded Anxiety | age_at_first_AD | 1.033 | 1.000 | 1.067 | 5.29e-02 | 7.41e-01 | 18562 | 13287 | 5275 |
| European | No Recorded Depression | Discontinuation vs Continuation | Recorded Anxiety | sex_at_birthMale | 1.088 | 1.015 | 1.167 | 1.70e-02 | 2.38e-01 | 18562 | 13287 | 5275 |
| European | No Recorded Depression | Discontinuation vs Continuation | Recorded Anxiety | Income>200k | 0.608 | 0.531 | 0.696 | 5.85e-13 | 8.18e-12 | 18562 | 13287 | 5275 |
| European | No Recorded Depression | Discontinuation vs Continuation | Recorded Anxiety | Income100-150k | 0.568 | 0.504 | 0.641 | <2.2e-16 | <2.2e-16 | 18562 | 13287 | 5275 |
| European | No Recorded Depression | Discontinuation vs Continuation | Recorded Anxiety | Income150-200k | 0.553 | 0.477 | 0.642 | 5.86e-15 | 8.20e-14 | 18562 | 13287 | 5275 |
| European | No Recorded Depression | Discontinuation vs Continuation | Recorded Anxiety | Income25-35k | 0.811 | 0.703 | 0.935 | 3.83e-03 | 5.36e-02 | 18562 | 13287 | 5275 |
| European | No Recorded Depression | Discontinuation vs Continuation | Recorded Anxiety | Income35-50k | 0.703 | 0.616 | 0.803 | 1.94e-07 | 2.72e-06 | 18562 | 13287 | 5275 |
| European | No Recorded Depression | Discontinuation vs Continuation | Recorded Anxiety | Income50-75k | 0.745 | 0.662 | 0.837 | 8.56e-07 | 1.20e-05 | 18562 | 13287 | 5275 |
| European | No Recorded Depression | Discontinuation vs Continuation | Recorded Anxiety | Income75-100k | 0.634 | 0.560 | 0.718 | 6.50e-13 | 9.10e-12 | 18562 | 13287 | 5275 |
| European | No Recorded Depression | Discontinuation vs Continuation | Recorded Anxiety | Education_levelCollege Postgraduate | 0.710 | 0.542 | 0.929 | 1.27e-02 | 1.78e-01 | 18562 | 13287 | 5275 |
| European | No Recorded Depression | Discontinuation vs Continuation | Recorded Anxiety | Education_levelCollege Undergraduate | 0.758 | 0.583 | 0.986 | 3.88e-02 | 5.43e-01 | 18562 | 13287 | 5275 |
| European | No Recorded Depression | Discontinuation vs Continuation | Recorded Anxiety | Education_levelTwelve or GED | 0.877 | 0.669 | 1.151 | 3.45e-01 | 1.00e+00 | 18562 | 13287 | 5275 |
| European | No Recorded Depression | Discontinuation vs Continuation | Recorded Anxiety | Recorded Anxiety | 0.605 | 0.565 | 0.647 | <2.2e-16 | <2.2e-16 | 18562 | 13287 | 5275 |
| European | No Recorded Depression | Switching vs Continuation | Recorded Anxiety | (Intercept) | 0.379 | 0.282 | 0.510 | 1.25e-10 | 1.75e-09 | 17516 | 13287 | 4229 |
| European | No Recorded Depression | Switching vs Continuation | Recorded Anxiety | age_at_first_AD | 0.774 | 0.746 | 0.802 | <2.2e-16 | <2.2e-16 | 17516 | 13287 | 4229 |
| European | No Recorded Depression | Switching vs Continuation | Recorded Anxiety | sex_at_birthMale | 0.921 | 0.851 | 0.998 | 4.45e-02 | 6.23e-01 | 17516 | 13287 | 4229 |
| European | No Recorded Depression | Switching vs Continuation | Recorded Anxiety | Income>200k | 0.504 | 0.431 | 0.590 | <2.2e-16 | 2.46e-16 | 17516 | 13287 | 4229 |
| European | No Recorded Depression | Switching vs Continuation | Recorded Anxiety | Income100-150k | 0.661 | 0.580 | 0.753 | 5.93e-10 | 8.31e-09 | 17516 | 13287 | 4229 |
| European | No Recorded Depression | Switching vs Continuation | Recorded Anxiety | Income150-200k | 0.636 | 0.542 | 0.747 | 3.15e-08 | 4.41e-07 | 17516 | 13287 | 4229 |
| European | No Recorded Depression | Switching vs Continuation | Recorded Anxiety | Income25-35k | 0.957 | 0.820 | 1.117 | 5.77e-01 | 1.00e+00 | 17516 | 13287 | 4229 |
| European | No Recorded Depression | Switching vs Continuation | Recorded Anxiety | Income35-50k | 0.880 | 0.763 | 1.015 | 7.82e-02 | 1.00e+00 | 17516 | 13287 | 4229 |
| European | No Recorded Depression | Switching vs Continuation | Recorded Anxiety | Income50-75k | 0.768 | 0.674 | 0.875 | 7.70e-05 | 1.08e-03 | 17516 | 13287 | 4229 |
| European | No Recorded Depression | Switching vs Continuation | Recorded Anxiety | Income75-100k | 0.699 | 0.609 | 0.801 | 2.91e-07 | 4.07e-06 | 17516 | 13287 | 4229 |
| European | No Recorded Depression | Switching vs Continuation | Recorded Anxiety | Education_levelCollege Postgraduate | 0.837 | 0.617 | 1.136 | 2.54e-01 | 1.00e+00 | 17516 | 13287 | 4229 |
| European | No Recorded Depression | Switching vs Continuation | Recorded Anxiety | Education_levelCollege Undergraduate | 0.870 | 0.646 | 1.171 | 3.58e-01 | 1.00e+00 | 17516 | 13287 | 4229 |
| European | No Recorded Depression | Switching vs Continuation | Recorded Anxiety | Education_levelTwelve or GED | 0.997 | 0.733 | 1.356 | 9.83e-01 | 1.00e+00 | 17516 | 13287 | 4229 |
| European | No Recorded Depression | Switching vs Continuation | Recorded Anxiety | Recorded Anxiety | 1.672 | 1.554 | 1.798 | <2.2e-16 | <2.2e-16 | 17516 | 13287 | 4229 |
| European | No Recorded Depression | Discontinuation vs Continuation | Smoking_Frequency | (Intercept) | 0.672 | 0.514 | 0.878 | 3.62e-03 | 5.06e-02 | 18190 | 13038 | 5152 |
| European | No Recorded Depression | Discontinuation vs Continuation | Smoking_Frequency | age_at_first_AD | 1.097 | 1.061 | 1.134 | 4.99e-08 | 6.98e-07 | 18190 | 13038 | 5152 |
| European | No Recorded Depression | Discontinuation vs Continuation | Smoking_Frequency | sex_at_birthMale | 1.117 | 1.041 | 1.198 | 2.00e-03 | 2.80e-02 | 18190 | 13038 | 5152 |
| European | No Recorded Depression | Discontinuation vs Continuation | Smoking_Frequency | Income>200k | 0.614 | 0.535 | 0.705 | 4.62e-12 | 6.47e-11 | 18190 | 13038 | 5152 |
| European | No Recorded Depression | Discontinuation vs Continuation | Smoking_Frequency | Income100-150k | 0.576 | 0.509 | 0.651 | <2.2e-16 | <2.2e-16 | 18190 | 13038 | 5152 |
| European | No Recorded Depression | Discontinuation vs Continuation | Smoking_Frequency | Income150-200k | 0.574 | 0.494 | 0.668 | 5.57e-13 | 7.80e-12 | 18190 | 13038 | 5152 |
| European | No Recorded Depression | Discontinuation vs Continuation | Smoking_Frequency | Income25-35k | 0.823 | 0.713 | 0.950 | 7.73e-03 | 1.08e-01 | 18190 | 13038 | 5152 |
| European | No Recorded Depression | Discontinuation vs Continuation | Smoking_Frequency | Income35-50k | 0.716 | 0.626 | 0.819 | 1.11e-06 | 1.56e-05 | 18190 | 13038 | 5152 |
| European | No Recorded Depression | Discontinuation vs Continuation | Smoking_Frequency | Income50-75k | 0.748 | 0.664 | 0.843 | 1.88e-06 | 2.64e-05 | 18190 | 13038 | 5152 |
| European | No Recorded Depression | Discontinuation vs Continuation | Smoking_Frequency | Income75-100k | 0.648 | 0.572 | 0.736 | 1.73e-11 | 2.42e-10 | 18190 | 13038 | 5152 |
| European | No Recorded Depression | Discontinuation vs Continuation | Smoking_Frequency | Education_levelCollege Postgraduate | 0.758 | 0.576 | 0.997 | 4.75e-02 | 6.65e-01 | 18190 | 13038 | 5152 |
| European | No Recorded Depression | Discontinuation vs Continuation | Smoking_Frequency | Education_levelCollege Undergraduate | 0.810 | 0.620 | 1.058 | 1.21e-01 | 1.00e+00 | 18190 | 13038 | 5152 |
| European | No Recorded Depression | Discontinuation vs Continuation | Smoking_Frequency | Education_levelTwelve or GED | 0.923 | 0.701 | 1.215 | 5.66e-01 | 1.00e+00 | 18190 | 13038 | 5152 |
| European | No Recorded Depression | Discontinuation vs Continuation | Smoking_Frequency | Smoking_FrequencyEvery Day | 1.205 | 1.049 | 1.384 | 8.37e-03 | 1.17e-01 | 18190 | 13038 | 5152 |
| European | No Recorded Depression | Discontinuation vs Continuation | Smoking_Frequency | Smoking_FrequencyNot At All (former) | 0.911 | 0.846 | 0.980 | 1.24e-02 | 1.74e-01 | 18190 | 13038 | 5152 |
| European | No Recorded Depression | Discontinuation vs Continuation | Smoking_Frequency | Smoking_FrequencySome Days | 1.142 | 0.938 | 1.390 | 1.87e-01 | 1.00e+00 | 18190 | 13038 | 5152 |
| European | No Recorded Depression | Switching vs Continuation | Smoking_Frequency | (Intercept) | 0.440 | 0.325 | 0.596 | 1.12e-07 | 1.56e-06 | 17168 | 13038 | 4130 |
| European | No Recorded Depression | Switching vs Continuation | Smoking_Frequency | age_at_first_AD | 0.736 | 0.710 | 0.763 | <2.2e-16 | <2.2e-16 | 17168 | 13038 | 4130 |
| European | No Recorded Depression | Switching vs Continuation | Smoking_Frequency | sex_at_birthMale | 0.897 | 0.827 | 0.972 | 8.25e-03 | 1.16e-01 | 17168 | 13038 | 4130 |
| European | No Recorded Depression | Switching vs Continuation | Smoking_Frequency | Income>200k | 0.536 | 0.457 | 0.630 | 2.95e-14 | 4.12e-13 | 17168 | 13038 | 4130 |
| European | No Recorded Depression | Switching vs Continuation | Smoking_Frequency | Income100-150k | 0.716 | 0.626 | 0.818 | 8.93e-07 | 1.25e-05 | 17168 | 13038 | 4130 |
| European | No Recorded Depression | Switching vs Continuation | Smoking_Frequency | Income150-200k | 0.689 | 0.585 | 0.811 | 7.62e-06 | 1.07e-04 | 17168 | 13038 | 4130 |
| European | No Recorded Depression | Switching vs Continuation | Smoking_Frequency | Income25-35k | 1.008 | 0.863 | 1.179 | 9.18e-01 | 1.00e+00 | 17168 | 13038 | 4130 |
| European | No Recorded Depression | Switching vs Continuation | Smoking_Frequency | Income35-50k | 0.936 | 0.810 | 1.081 | 3.68e-01 | 1.00e+00 | 17168 | 13038 | 4130 |
| European | No Recorded Depression | Switching vs Continuation | Smoking_Frequency | Income50-75k | 0.828 | 0.725 | 0.945 | 5.28e-03 | 7.39e-02 | 17168 | 13038 | 4130 |
| European | No Recorded Depression | Switching vs Continuation | Smoking_Frequency | Income75-100k | 0.753 | 0.655 | 0.866 | 6.48e-05 | 9.07e-04 | 17168 | 13038 | 4130 |
| European | No Recorded Depression | Switching vs Continuation | Smoking_Frequency | Education_levelCollege Postgraduate | 0.861 | 0.631 | 1.174 | 3.43e-01 | 1.00e+00 | 17168 | 13038 | 4130 |
| European | No Recorded Depression | Switching vs Continuation | Smoking_Frequency | Education_levelCollege Undergraduate | 0.887 | 0.656 | 1.200 | 4.38e-01 | 1.00e+00 | 17168 | 13038 | 4130 |
| European | No Recorded Depression | Switching vs Continuation | Smoking_Frequency | Education_levelTwelve or GED | 1.002 | 0.734 | 1.368 | 9.90e-01 | 1.00e+00 | 17168 | 13038 | 4130 |

| | | | | | | | | | | | | |
|---|---|---|---|---|---|---|---|---|---|---|---|---|
| European | No Recorded Depression | Switching vs Continuation | Smoking_Frequency | Smoking_FrequencyEvery Day | 1.299 | 1.120 | 1.506 | 5.35e-04 | 7.48e-03 | 17168 | 13038 | 4130 |
| European | No Recorded Depression | Switching vs Continuation | Smoking_Frequency | Smoking_FrequencyNot At All (former) | 1.096 | 1.010 | 1.188 | 2.72e-02 | 3.80e-01 | 17168 | 13038 | 4130 |
| European | No Recorded Depression | Switching vs Continuation | Smoking_Frequency | Smoking_FrequencySome Days | 1.281 | 1.041 | 1.575 | 1.92e-02 | 2.68e-01 | 17168 | 13038 | 4130 |
| European | No Recorded Depression | Discontinuation vs Continuation | Stress | (Intercept) | 0.512 | 0.306 | 0.858 | 1.10e-02 | 1.54e-01 | 11806 | 8575 | 3231 |
| European | No Recorded Depression | Discontinuation vs Continuation | Stress | age_at_first_AD | 1.109 | 1.063 | 1.158 | 2.00e-06 | 2.80e-05 | 11806 | 8575 | 3231 |
| European | No Recorded Depression | Discontinuation vs Continuation | Stress | sex_at_birthMale | 1.045 | 0.955 | 1.143 | 3.39e-01 | 1.00e+00 | 11806 | 8575 | 3231 |
| European | No Recorded Depression | Discontinuation vs Continuation | Stress | Income>200k | 0.617 | 0.515 | 0.738 | 1.40e-07 | 1.95e-06 | 11806 | 8575 | 3231 |
| European | No Recorded Depression | Discontinuation vs Continuation | Stress | Income100-150k | 0.568 | 0.483 | 0.668 | 7.76e-12 | 1.09e-10 | 11806 | 8575 | 3231 |
| European | No Recorded Depression | Discontinuation vs Continuation | Stress | Income150-200k | 0.579 | 0.479 | 0.701 | 2.11e-08 | 2.95e-07 | 11806 | 8575 | 3231 |
| European | No Recorded Depression | Discontinuation vs Continuation | Stress | Income25-35k | 0.789 | 0.648 | 0.960 | 1.79e-02 | 2.51e-01 | 11806 | 8575 | 3231 |
| European | No Recorded Depression | Discontinuation vs Continuation | Stress | Income35-50k | 0.702 | 0.586 | 0.841 | 1.25e-04 | 1.75e-03 | 11806 | 8575 | 3231 |
| European | No Recorded Depression | Discontinuation vs Continuation | Stress | Income50-75k | 0.764 | 0.651 | 0.897 | 1.02e-03 | 1.43e-02 | 11806 | 8575 | 3231 |
| European | No Recorded Depression | Discontinuation vs Continuation | Stress | Income75-100k | 0.643 | 0.544 | 0.760 | 2.41e-07 | 3.37e-06 | 11806 | 8575 | 3231 |
| European | No Recorded Depression | Discontinuation vs Continuation | Stress | Education_levelCollege Postgraduate | 0.941 | 0.559 | 1.584 | 8.19e-01 | 1.00e+00 | 11806 | 8575 | 3231 |
| European | No Recorded Depression | Discontinuation vs Continuation | Stress | Education_levelCollege Undergraduate | 1.036 | 0.619 | 1.734 | 8.93e-01 | 1.00e+00 | 11806 | 8575 | 3231 |
| European | No Recorded Depression | Discontinuation vs Continuation | Stress | Education_levelTwelve or GED | 0.955 | 0.562 | 1.621 | 8.64e-01 | 1.00e+00 | 11806 | 8575 | 3231 |
| European | No Recorded Depression | Discontinuation vs Continuation | Stress | StressAlmost Never | 1.018 | 0.856 | 1.212 | 8.38e-01 | 1.00e+00 | 11806 | 8575 | 3231 |
| European | No Recorded Depression | Discontinuation vs Continuation | Stress | StressFairly Often | 0.957 | 0.802 | 1.141 | 6.23e-01 | 1.00e+00 | 11806 | 8575 | 3231 |
| European | No Recorded Depression | Discontinuation vs Continuation | Stress | StressSometimes | 1.140 | 1.038 | 1.252 | 6.13e-03 | 8.58e-02 | 11806 | 8575 | 3231 |
| European | No Recorded Depression | Discontinuation vs Continuation | Stress | StressVery Often | 1.149 | 0.930 | 1.418 | 1.98e-01 | 1.00e+00 | 11806 | 8575 | 3231 |
| European | No Recorded Depression | Switching vs Continuation | Stress | (Intercept) | 0.459 | 0.269 | 0.781 | 4.12e-03 | 5.77e-02 | 11283 | 8575 | 2708 |
| European | No Recorded Depression | Switching vs Continuation | Stress | age_at_first_AD | 0.706 | 0.674 | 0.739 | <2.2e-16 | <2.2e-16 | 11283 | 8575 | 2708 |
| European | No Recorded Depression | Switching vs Continuation | Stress | sex_at_birthMale | 0.937 | 0.846 | 1.037 | 2.07e-01 | 1.00e+00 | 11283 | 8575 | 2708 |
| European | No Recorded Depression | Switching vs Continuation | Stress | Income>200k | 0.523 | 0.427 | 0.642 | 4.70e-10 | 6.58e-09 | 11283 | 8575 | 2708 |
| European | No Recorded Depression | Switching vs Continuation | Stress | Income100-150k | 0.677 | 0.569 | 0.804 | 9.73e-06 | 1.36e-04 | 11283 | 8575 | 2708 |
| European | No Recorded Depression | Switching vs Continuation | Stress | Income150-200k | 0.643 | 0.524 | 0.789 | 2.35e-05 | 3.29e-04 | 11283 | 8575 | 2708 |
| European | No Recorded Depression | Switching vs Continuation | Stress | Income25-35k | 0.950 | 0.771 | 1.170 | 6.27e-01 | 1.00e+00 | 11283 | 8575 | 2708 |
| European | No Recorded Depression | Switching vs Continuation | Stress | Income35-50k | 0.848 | 0.700 | 1.027 | 9.22e-02 | 1.00e+00 | 11283 | 8575 | 2708 |
| European | No Recorded Depression | Switching vs Continuation | Stress | Income50-75k | 0.784 | 0.658 | 0.934 | 6.31e-03 | 8.84e-02 | 11283 | 8575 | 2708 |
| European | No Recorded Depression | Switching vs Continuation | Stress | Income75-100k | 0.772 | 0.645 | 0.924 | 4.75e-03 | 6.65e-02 | 11283 | 8575 | 2708 |
| European | No Recorded Depression | Switching vs Continuation | Stress | Education_levelCollege Postgraduate | 0.863 | 0.505 | 1.473 | 5.88e-01 | 1.00e+00 | 11283 | 8575 | 2708 |
| European | No Recorded Depression | Switching vs Continuation | Stress | Education_levelCollege Undergraduate | 0.900 | 0.530 | 1.529 | 6.97e-01 | 1.00e+00 | 11283 | 8575 | 2708 |
| European | No Recorded Depression | Switching vs Continuation | Stress | Education_levelTwelve or GED | 1.030 | 0.598 | 1.773 | 9.15e-01 | 1.00e+00 | 11283 | 8575 | 2708 |
| European | No Recorded Depression | Switching vs Continuation | Stress | StressAlmost Never | 0.979 | 0.807 | 1.187 | 8.28e-01 | 1.00e+00 | 11283 | 8575 | 2708 |
| European | No Recorded Depression | Switching vs Continuation | Stress | StressFairly Often | 0.934 | 0.773 | 1.129 | 4.80e-01 | 1.00e+00 | 11283 | 8575 | 2708 |
| European | No Recorded Depression | Switching vs Continuation | Stress | StressSometimes | 1.201 | 1.088 | 1.326 | 2.79e-04 | 3.91e-03 | 11283 | 8575 | 2708 |
| European | No Recorded Depression | Switching vs Continuation | Stress | StressVery Often | 1.007 | 0.785 | 1.291 | 9.59e-01 | 1.00e+00 | 11283 | 8575 | 2708 |
| European | No Recorded Depression | Discontinuation vs Continuation | age_at_first_AD | (Intercept) | 0.378 | 0.363 | 0.393 | <2.2e-16 | <2.2e-16 | 18562 | 13287 | 5275 |
| European | No Recorded Depression | Discontinuation vs Continuation | age_at_first_AD | age_at_first_AD | 1.071 | 1.037 | 1.105 | 2.32e-05 | 3.24e-04 | 18562 | 13287 | 5275 |
| European | No Recorded Depression | Discontinuation vs Continuation | age_at_first_AD | sex_at_birthMale | 1.115 | 1.041 | 1.194 | 1.93e-03 | 2.70e-02 | 18562 | 13287 | 5275 |
| European | No Recorded Depression | Switching vs Continuation | age_at_first_AD | (Intercept) | 0.328 | 0.315 | 0.342 | <2.2e-16 | <2.2e-16 | 17516 | 13287 | 4229 |
| European | No Recorded Depression | Switching vs Continuation | age_at_first_AD | age_at_first_AD | 0.738 | 0.712 | 0.764 | <2.2e-16 | <2.2e-16 | 17516 | 13287 | 4229 |
| European | No Recorded Depression | Switching vs Continuation | age_at_first_AD | sex_at_birthMale | 0.900 | 0.831 | 0.973 | 8.56e-03 | 1.20e-01 | 17516 | 13287 | 4229 |
| European | No Recorded Depression | Discontinuation vs Continuation | sex_at_birth | (Intercept) | 0.378 | 0.363 | 0.393 | <2.2e-16 | <2.2e-16 | 18562 | 13287 | 5275 |
| European | No Recorded Depression | Discontinuation vs Continuation | sex_at_birth | age_at_first_AD | 1.071 | 1.037 | 1.105 | 2.32e-05 | 3.24e-04 | 18562 | 13287 | 5275 |
| European | No Recorded Depression | Discontinuation vs Continuation | sex_at_birth | sex_at_birthMale | 1.115 | 1.041 | 1.194 | 1.93e-03 | 2.70e-02 | 18562 | 13287 | 5275 |
| European | No Recorded Depression | Switching vs Continuation | sex_at_birth | (Intercept) | 0.328 | 0.315 | 0.342 | <2.2e-16 | <2.2e-16 | 17516 | 13287 | 4229 |
| European | No Recorded Depression | Switching vs Continuation | sex_at_birth | age_at_first_AD | 0.738 | 0.712 | 0.764 | <2.2e-16 | <2.2e-16 | 17516 | 13287 | 4229 |
| European | No Recorded Depression | Switching vs Continuation | sex_at_birth | sex_at_birthMale | 0.900 | 0.831 | 0.973 | 8.56e-03 | 1.20e-01 | 17516 | 13287 | 4229 |
| European | Recorded Anxiety | Discontinuation vs Continuation | Alcohol_Frequency | (Intercept) | 0.863 | 0.635 | 1.174 | 3.49e-01 | 1.00e+00 | 15058 | 11691 | 3367 |
| European | Recorded Anxiety | Discontinuation vs Continuation | Alcohol_Frequency | age_at_first_AD | 1.083 | 1.042 | 1.126 | 6.03e-05 | 8.45e-04 | 15058 | 11691 | 3367 |
| European | Recorded Anxiety | Discontinuation vs Continuation | Alcohol_Frequency | sex_at_birthMale | 1.219 | 1.119 | 1.328 | 5.97e-06 | 8.36e-05 | 15058 | 11691 | 3367 |
| European | Recorded Anxiety | Discontinuation vs Continuation | Alcohol_Frequency | Income>200k | 0.467 | 0.392 | 0.556 | <2.2e-16 | <2.2e-16 | 15058 | 11691 | 3367 |
| European | Recorded Anxiety | Discontinuation vs Continuation | Alcohol_Frequency | Income100-150k | 0.442 | 0.383 | 0.509 | <2.2e-16 | <2.2e-16 | 15058 | 11691 | 3367 |
| European | Recorded Anxiety | Discontinuation vs Continuation | Alcohol_Frequency | Income150-200k | 0.433 | 0.360 | 0.522 | <2.2e-16 | <2.2e-16 | 15058 | 11691 | 3367 |
| European | Recorded Anxiety | Discontinuation vs Continuation | Alcohol_Frequency | Income25-35k | 0.659 | 0.565 | 0.767 | 8.69e-08 | 1.22e-06 | 15058 | 11691 | 3367 |
| European | Recorded Anxiety | Discontinuation vs Continuation | Alcohol_Frequency | Income35-50k | 0.585 | 0.505 | 0.678 | 9.41e-13 | 1.32e-11 | 15058 | 11691 | 3367 |
| European | Recorded Anxiety | Discontinuation vs Continuation | Alcohol_Frequency | Income50-75k | 0.590 | 0.517 | 0.672 | 2.15e-15 | 3.00e-14 | 15058 | 11691 | 3367 |
| European | Recorded Anxiety | Discontinuation vs Continuation | Alcohol_Frequency | Income75-100k | 0.500 | 0.433 | 0.578 | <2.2e-16 | <2.2e-16 | 15058 | 11691 | 3367 |
| European | Recorded Anxiety | Discontinuation vs Continuation | Alcohol_Frequency | Education_levelCollege Postgraduate | 0.666 | 0.509 | 0.871 | 3.05e-03 | 4.28e-02 | 15058 | 11691 | 3367 |
| European | Recorded Anxiety | Discontinuation vs Continuation | Alcohol_Frequency | Education_levelCollege Undergraduate | 0.736 | 0.569 | 0.951 | 1.90e-02 | 2.66e-01 | 15058 | 11691 | 3367 |
| European | Recorded Anxiety | Discontinuation vs Continuation | Alcohol_Frequency | Education_levelTwelve or GED | 0.902 | 0.690 | 1.179 | 4.51e-01 | 1.00e+00 | 15058 | 11691 | 3367 |
| European | Recorded Anxiety | Discontinuation vs Continuation | Alcohol_Frequency | Alcohol_Frequency2-3 Week | 0.671 | 0.535 | 0.842 | 5.80e-04 | 8.12e-03 | 15058 | 11691 | 3367 |
| European | Recorded Anxiety | Discontinuation vs Continuation | Alcohol_Frequency | Alcohol_Frequency2-4 Month | 0.650 | 0.521 | 0.811 | 1.37e-04 | 1.91e-03 | 15058 | 11691 | 3367 |
| European | Recorded Anxiety | Discontinuation vs Continuation | Alcohol_Frequency | Alcohol_Frequency4plus Week | 0.637 | 0.505 | 0.803 | 1.35e-04 | 1.89e-03 | 15058 | 11691 | 3367 |
| European | Recorded Anxiety | Discontinuation vs Continuation | Alcohol_Frequency | Alcohol_FrequencyMonthly or Less | 0.757 | 0.613 | 0.936 | 1.02e-02 | 1.43e-01 | 15058 | 11691 | 3367 |

| | | | | | | | | | | | | |
|---|---|---|---|---|---|---|---|---|---|---|---|---|
| European | Recorded Anxiety | Discontinuation vs Continuation | Alcohol_Frequency | Alcohol_FrequencyNever (former) | 0.805 | 0.645 | 1.004 | 5.39e-02 | 7.55e-01 | 15058 | 11691 | 3367 |
| European | Recorded Anxiety | Switching vs Continuation | Alcohol_Frequency | (Intercept) | 1.361 | 1.064 | 1.741 | 1.42e-02 | 1.99e-01 | 22474 | 11691 | 10783 |
| European | Recorded Anxiety | Switching vs Continuation | Alcohol_Frequency | age_at_first_AD | 0.743 | 0.722 | 0.764 | <2.2e-16 | <2.2e-16 | 22474 | 11691 | 10783 |
| European | Recorded Anxiety | Switching vs Continuation | Alcohol_Frequency | sex_at_birthMale | 1.159 | 1.091 | 1.232 | 1.99e-06 | 2.79e-05 | 22474 | 11691 | 10783 |
| European | Recorded Anxiety | Switching vs Continuation | Alcohol_Frequency | Income>200k | 0.434 | 0.383 | 0.492 | <2.2e-16 | <2.2e-16 | 22474 | 11691 | 10783 |
| European | Recorded Anxiety | Switching vs Continuation | Alcohol_Frequency | Income100-150k | 0.571 | 0.518 | 0.630 | <2.2e-16 | <2.2e-16 | 22474 | 11691 | 10783 |
| European | Recorded Anxiety | Switching vs Continuation | Alcohol_Frequency | Income150-200k | 0.470 | 0.414 | 0.534 | <2.2e-16 | <2.2e-16 | 22474 | 11691 | 10783 |
| European | Recorded Anxiety | Switching vs Continuation | Alcohol_Frequency | Income25-35k | 0.805 | 0.720 | 0.899 | 1.16e-04 | 1.62e-03 | 22474 | 11691 | 10783 |
| European | Recorded Anxiety | Switching vs Continuation | Alcohol_Frequency | Income35-50k | 0.789 | 0.711 | 0.875 | 7.62e-06 | 1.07e-04 | 22474 | 11691 | 10783 |
| European | Recorded Anxiety | Switching vs Continuation | Alcohol_Frequency | Income50-75k | 0.739 | 0.672 | 0.812 | 2.86e-10 | 4.01e-09 | 22474 | 11691 | 10783 |
| European | Recorded Anxiety | Switching vs Continuation | Alcohol_Frequency | Income75-100k | 0.621 | 0.561 | 0.687 | <2.2e-16 | <2.2e-16 | 22474 | 11691 | 10783 |
| European | Recorded Anxiety | Switching vs Continuation | Alcohol_Frequency | Education_levelCollege Postgraduate | 0.834 | 0.673 | 1.033 | 9.62e-02 | 1.00e+00 | 22474 | 11691 | 10783 |
| European | Recorded Anxiety | Switching vs Continuation | Alcohol_Frequency | Education_levelCollege Undergraduate | 0.917 | 0.746 | 1.128 | 4.14e-01 | 1.00e+00 | 22474 | 11691 | 10783 |
| European | Recorded Anxiety | Switching vs Continuation | Alcohol_Frequency | Education_levelTwelve or GED | 1.138 | 0.918 | 1.411 | 2.39e-01 | 1.00e+00 | 22474 | 11691 | 10783 |
| European | Recorded Anxiety | Switching vs Continuation | Alcohol_Frequency | Alcohol_Frequency2-3 Week | 0.829 | 0.696 | 0.987 | 3.46e-02 | 4.85e-01 | 22474 | 11691 | 10783 |
| European | Recorded Anxiety | Switching vs Continuation | Alcohol_Frequency | Alcohol_Frequency2-4 Month | 0.897 | 0.757 | 1.063 | 2.10e-01 | 1.00e+00 | 22474 | 11691 | 10783 |
| European | Recorded Anxiety | Switching vs Continuation | Alcohol_Frequency | Alcohol_Frequency4plus Week | 0.915 | 0.766 | 1.092 | 3.24e-01 | 1.00e+00 | 22474 | 11691 | 10783 |
| European | Recorded Anxiety | Switching vs Continuation | Alcohol_Frequency | Alcohol_FrequencyMonthly or Less | 1.031 | 0.874 | 1.216 | 7.21e-01 | 1.00e+00 | 22474 | 11691 | 10783 |
| European | Recorded Anxiety | Switching vs Continuation | Alcohol_Frequency | Alcohol_FrequencyNever (former) | 1.211 | 1.020 | 1.438 | 2.86e-02 | 4.00e-01 | 22474 | 11691 | 10783 |
| European | Recorded Anxiety | Discontinuation vs Continuation | BMI | (Intercept) | 0.672 | 0.523 | 0.863 | 1.82e-03 | 2.54e-02 | 14800 | 11557 | 3243 |
| European | Recorded Anxiety | Discontinuation vs Continuation | BMI | age_at_first_AD | 1.082 | 1.041 | 1.125 | 6.91e-05 | 9.67e-04 | 14800 | 11557 | 3243 |
| European | Recorded Anxiety | Discontinuation vs Continuation | BMI | sex_at_birthMale | 1.229 | 1.127 | 1.339 | 2.71e-06 | 3.80e-05 | 14800 | 11557 | 3243 |
| European | Recorded Anxiety | Discontinuation vs Continuation | BMI | Income>200k | 0.442 | 0.371 | 0.526 | <2.2e-16 | <2.2e-16 | 14800 | 11557 | 3243 |
| European | Recorded Anxiety | Discontinuation vs Continuation | BMI | Income100-150k | 0.433 | 0.376 | 0.499 | <2.2e-16 | <2.2e-16 | 14800 | 11557 | 3243 |
| European | Recorded Anxiety | Discontinuation vs Continuation | BMI | Income150-200k | 0.412 | 0.342 | 0.497 | <2.2e-16 | <2.2e-16 | 14800 | 11557 | 3243 |
| European | Recorded Anxiety | Discontinuation vs Continuation | BMI | Income25-35k | 0.635 | 0.543 | 0.742 | 1.12e-08 | 1.56e-07 | 14800 | 11557 | 3243 |
| European | Recorded Anxiety | Discontinuation vs Continuation | BMI | Income35-50k | 0.576 | 0.496 | 0.670 | 5.58e-13 | 7.81e-12 | 14800 | 11557 | 3243 |
| European | Recorded Anxiety | Discontinuation vs Continuation | BMI | Income50-75k | 0.591 | 0.518 | 0.674 | 4.44e-15 | 6.22e-14 | 14800 | 11557 | 3243 |
| European | Recorded Anxiety | Discontinuation vs Continuation | BMI | Income75-100k | 0.492 | 0.426 | 0.569 | <2.2e-16 | <2.2e-16 | 14800 | 11557 | 3243 |
| European | Recorded Anxiety | Discontinuation vs Continuation | BMI | Education_levelCollege Postgraduate | 0.587 | 0.448 | 0.770 | 1.15e-04 | 1.61e-03 | 14800 | 11557 | 3243 |
| European | Recorded Anxiety | Discontinuation vs Continuation | BMI | Education_levelCollege Undergraduate | 0.666 | 0.515 | 0.863 | 2.06e-03 | 2.88e-02 | 14800 | 11557 | 3243 |
| European | Recorded Anxiety | Discontinuation vs Continuation | BMI | Education_levelTwelve or GED | 0.858 | 0.655 | 1.124 | 2.66e-01 | 1.00e+00 | 14800 | 11557 | 3243 |
| European | Recorded Anxiety | Discontinuation vs Continuation | BMI | BMI | 0.929 | 0.890 | 0.969 | 6.65e-04 | 9.31e-03 | 14800 | 11557 | 3243 |
| European | Recorded Anxiety | Switching vs Continuation | BMI | (Intercept) | 1.433 | 1.171 | 1.754 | 4.74e-04 | 6.63e-03 | 22286 | 11557 | 10729 |
| European | Recorded Anxiety | Switching vs Continuation | BMI | age_at_first_AD | 0.756 | 0.735 | 0.777 | <2.2e-16 | <2.2e-16 | 22286 | 11557 | 10729 |
| European | Recorded Anxiety | Switching vs Continuation | BMI | sex_at_birthMale | 1.159 | 1.091 | 1.232 | 1.78e-06 | 2.49e-05 | 22286 | 11557 | 10729 |
| European | Recorded Anxiety | Switching vs Continuation | BMI | Income>200k | 0.429 | 0.379 | 0.486 | <2.2e-16 | <2.2e-16 | 22286 | 11557 | 10729 |
| European | Recorded Anxiety | Switching vs Continuation | BMI | Income100-150k | 0.554 | 0.502 | 0.611 | <2.2e-16 | <2.2e-16 | 22286 | 11557 | 10729 |
| European | Recorded Anxiety | Switching vs Continuation | BMI | Income150-200k | 0.449 | 0.395 | 0.510 | <2.2e-16 | <2.2e-16 | 22286 | 11557 | 10729 |
| European | Recorded Anxiety | Switching vs Continuation | BMI | Income25-35k | 0.782 | 0.700 | 0.873 | 1.26e-05 | 1.76e-04 | 22286 | 11557 | 10729 |
| European | Recorded Anxiety | Switching vs Continuation | BMI | Income35-50k | 0.776 | 0.699 | 0.861 | 1.80e-06 | 2.52e-05 | 22286 | 11557 | 10729 |
| European | Recorded Anxiety | Switching vs Continuation | BMI | Income50-75k | 0.717 | 0.653 | 0.788 | 4.11e-12 | 5.76e-11 | 22286 | 11557 | 10729 |
| European | Recorded Anxiety | Switching vs Continuation | BMI | Income75-100k | 0.594 | 0.537 | 0.657 | <2.2e-16 | <2.2e-16 | 22286 | 11557 | 10729 |
| European | Recorded Anxiety | Switching vs Continuation | BMI | Education_levelCollege Postgraduate | 0.806 | 0.650 | 0.999 | 4.86e-02 | 6.80e-01 | 22286 | 11557 | 10729 |
| European | Recorded Anxiety | Switching vs Continuation | BMI | Education_levelCollege Undergraduate | 0.881 | 0.716 | 1.084 | 2.30e-01 | 1.00e+00 | 22286 | 11557 | 10729 |
| European | Recorded Anxiety | Switching vs Continuation | BMI | Education_levelTwelve or GED | 1.112 | 0.896 | 1.379 | 3.36e-01 | 1.00e+00 | 22286 | 11557 | 10729 |
| European | Recorded Anxiety | Switching vs Continuation | BMI | BMI | 1.132 | 1.100 | 1.164 | <2.2e-16 | <2.2e-16 | 22286 | 11557 | 10729 |
| European | Recorded Anxiety | Discontinuation vs Continuation | Education_level | (Intercept) | 0.688 | 0.540 | 0.877 | 2.52e-03 | 3.52e-02 | 15204 | 11798 | 3406 |
| European | Recorded Anxiety | Discontinuation vs Continuation | Education_level | age_at_first_AD | 1.088 | 1.048 | 1.131 | 1.28e-05 | 1.79e-04 | 15204 | 11798 | 3406 |
| European | Recorded Anxiety | Discontinuation vs Continuation | Education_level | sex_at_birthMale | 1.216 | 1.118 | 1.323 | 5.52e-06 | 7.72e-05 | 15204 | 11798 | 3406 |
| European | Recorded Anxiety | Discontinuation vs Continuation | Education_level | Income>200k | 0.437 | 0.369 | 0.519 | <2.2e-16 | <2.2e-16 | 15204 | 11798 | 3406 |
| European | Recorded Anxiety | Discontinuation vs Continuation | Education_level | Income100-150k | 0.420 | 0.365 | 0.483 | <2.2e-16 | <2.2e-16 | 15204 | 11798 | 3406 |
| European | Recorded Anxiety | Discontinuation vs Continuation | Education_level | Income150-200k | 0.405 | 0.337 | 0.486 | <2.2e-16 | <2.2e-16 | 15204 | 11798 | 3406 |
| European | Recorded Anxiety | Discontinuation vs Continuation | Education_level | Income25-35k | 0.647 | 0.556 | 0.752 | 1.60e-08 | 2.24e-07 | 15204 | 11798 | 3406 |
| European | Recorded Anxiety | Discontinuation vs Continuation | Education_level | Income35-50k | 0.572 | 0.495 | 0.662 | 5.70e-14 | 7.98e-13 | 15204 | 11798 | 3406 |
| European | Recorded Anxiety | Discontinuation vs Continuation | Education_level | Income50-75k | 0.573 | 0.504 | 0.652 | <2.2e-16 | 3.43e-16 | 15204 | 11798 | 3406 |
| European | Recorded Anxiety | Discontinuation vs Continuation | Education_level | Income75-100k | 0.480 | 0.416 | 0.553 | <2.2e-16 | <2.2e-16 | 15204 | 11798 | 3406 |
| European | Recorded Anxiety | Discontinuation vs Continuation | Education_level | Education_levelCollege Postgraduate | 0.615 | 0.473 | 0.800 | 2.86e-04 | 4.00e-03 | 15204 | 11798 | 3406 |
| European | Recorded Anxiety | Discontinuation vs Continuation | Education_level | Education_levelCollege Undergraduate | 0.685 | 0.533 | 0.880 | 3.07e-03 | 4.30e-02 | 15204 | 11798 | 3406 |
| European | Recorded Anxiety | Discontinuation vs Continuation | Education_level | Education_levelTwelve or GED | 0.858 | 0.660 | 1.115 | 2.51e-01 | 1.00e+00 | 15204 | 11798 | 3406 |
| European | Recorded Anxiety | Switching vs Continuation | Education_level | (Intercept) | 1.421 | 1.166 | 1.732 | 5.06e-04 | 7.09e-03 | 22696 | 11798 | 10898 |
| European | Recorded Anxiety | Switching vs Continuation | Education_level | age_at_first_AD | 0.755 | 0.735 | 0.776 | <2.2e-16 | <2.2e-16 | 22696 | 11798 | 10898 |
| European | Recorded Anxiety | Switching vs Continuation | Education_level | sex_at_birthMale | 1.159 | 1.092 | 1.231 | 1.42e-06 | 1.98e-05 | 22696 | 11798 | 10898 |
| European | Recorded Anxiety | Switching vs Continuation | Education_level | Income>200k | 0.404 | 0.358 | 0.457 | <2.2e-16 | <2.2e-16 | 22696 | 11798 | 10898 |
| European | Recorded Anxiety | Switching vs Continuation | Education_level | Income100-150k | 0.541 | 0.491 | 0.596 | <2.2e-16 | <2.2e-16 | 22696 | 11798 | 10898 |
| European | Recorded Anxiety | Switching vs Continuation | Education_level | Income150-200k | 0.437 | 0.385 | 0.496 | <2.2e-16 | <2.2e-16 | 22696 | 11798 | 10898 |

| | | | | | | | | | | | | |
|---|---|---|---|---|---|---|---|---|---|---|---|---|
| European | Recorded Anxiety | Switching vs Continuation | Education_level | Income25-35k | 0.785 | 0.703 | 0.875 | 1.40e-05 | 1.96e-04 | 22696 | 11798 | 10898 |
| European | Recorded Anxiety | Switching vs Continuation | Education_level | Income35-50k | 0.772 | 0.696 | 0.855 | 7.79e-07 | 1.09e-05 | 22696 | 11798 | 10898 |
| European | Recorded Anxiety | Switching vs Continuation | Education_level | Income50-75k | 0.715 | 0.651 | 0.784 | 1.28e-12 | 1.79e-11 | 22696 | 11798 | 10898 |
| European | Recorded Anxiety | Switching vs Continuation | Education_level | Income75-100k | 0.590 | 0.534 | 0.652 | <2.2e-16 | <2.2e-16 | 22696 | 11798 | 10898 |
| European | Recorded Anxiety | Switching vs Continuation | Education_level | Education_levelCollege Postgraduate | 0.806 | 0.653 | 0.995 | 4.42e-02 | 6.19e-01 | 22696 | 11798 | 10898 |
| European | Recorded Anxiety | Switching vs Continuation | Education_level | Education_levelCollege Undergraduate | 0.895 | 0.730 | 1.098 | 2.88e-01 | 1.00e+00 | 22696 | 11798 | 10898 |
| European | Recorded Anxiety | Switching vs Continuation | Education_level | Education_levelTwelve or GED | 1.132 | 0.916 | 1.399 | 2.53e-01 | 1.00e+00 | 22696 | 11798 | 10898 |
| European | Recorded Anxiety | Discontinuation vs Continuation | Income | (Intercept) | 0.688 | 0.540 | 0.877 | 2.52e-03 | 3.52e-02 | 15204 | 11798 | 3406 |
| European | Recorded Anxiety | Discontinuation vs Continuation | Income | age_at_first_AD | 1.088 | 1.048 | 1.131 | 1.28e-05 | 1.79e-04 | 15204 | 11798 | 3406 |
| European | Recorded Anxiety | Discontinuation vs Continuation | Income | sex_at_birthMale | 1.216 | 1.118 | 1.323 | 5.52e-06 | 7.72e-05 | 15204 | 11798 | 3406 |
| European | Recorded Anxiety | Discontinuation vs Continuation | Income | Income>200k | 0.437 | 0.369 | 0.519 | <2.2e-16 | <2.2e-16 | 15204 | 11798 | 3406 |
| European | Recorded Anxiety | Discontinuation vs Continuation | Income | Income100-150k | 0.420 | 0.365 | 0.483 | <2.2e-16 | <2.2e-16 | 15204 | 11798 | 3406 |
| European | Recorded Anxiety | Discontinuation vs Continuation | Income | Income150-200k | 0.405 | 0.337 | 0.486 | <2.2e-16 | <2.2e-16 | 15204 | 11798 | 3406 |
| European | Recorded Anxiety | Discontinuation vs Continuation | Income | Income25-35k | 0.647 | 0.556 | 0.752 | 1.60e-08 | 2.24e-07 | 15204 | 11798 | 3406 |
| European | Recorded Anxiety | Discontinuation vs Continuation | Income | Income35-50k | 0.572 | 0.495 | 0.662 | 5.70e-14 | 7.98e-13 | 15204 | 11798 | 3406 |
| European | Recorded Anxiety | Discontinuation vs Continuation | Income | Income50-75k | 0.573 | 0.504 | 0.652 | <2.2e-16 | 3.43e-16 | 15204 | 11798 | 3406 |
| European | Recorded Anxiety | Discontinuation vs Continuation | Income | Income75-100k | 0.480 | 0.416 | 0.553 | <2.2e-16 | <2.2e-16 | 15204 | 11798 | 3406 |
| European | Recorded Anxiety | Discontinuation vs Continuation | Income | Education_levelCollege Postgraduate | 0.615 | 0.473 | 0.800 | 2.86e-04 | 4.00e-03 | 15204 | 11798 | 3406 |
| European | Recorded Anxiety | Discontinuation vs Continuation | Income | Education_levelCollege Undergraduate | 0.685 | 0.533 | 0.880 | 3.07e-03 | 4.30e-02 | 15204 | 11798 | 3406 |
| European | Recorded Anxiety | Discontinuation vs Continuation | Income | Education_levelTwelve or GED | 0.858 | 0.660 | 1.115 | 2.51e-01 | 1.00e+00 | 15204 | 11798 | 3406 |
| European | Recorded Anxiety | Switching vs Continuation | Income | (Intercept) | 1.421 | 1.166 | 1.732 | 5.06e-04 | 7.09e-03 | 22696 | 11798 | 10898 |
| European | Recorded Anxiety | Switching vs Continuation | Income | age_at_first_AD | 0.755 | 0.735 | 0.776 | <2.2e-16 | <2.2e-16 | 22696 | 11798 | 10898 |
| European | Recorded Anxiety | Switching vs Continuation | Income | sex_at_birthMale | 1.159 | 1.092 | 1.231 | 1.42e-06 | 1.98e-05 | 22696 | 11798 | 10898 |
| European | Recorded Anxiety | Switching vs Continuation | Income | Income>200k | 0.404 | 0.358 | 0.457 | <2.2e-16 | <2.2e-16 | 22696 | 11798 | 10898 |
| European | Recorded Anxiety | Switching vs Continuation | Income | Income100-150k | 0.541 | 0.491 | 0.596 | <2.2e-16 | <2.2e-16 | 22696 | 11798 | 10898 |
| European | Recorded Anxiety | Switching vs Continuation | Income | Income150-200k | 0.437 | 0.385 | 0.496 | <2.2e-16 | <2.2e-16 | 22696 | 11798 | 10898 |
| European | Recorded Anxiety | Switching vs Continuation | Income | Income25-35k | 0.785 | 0.703 | 0.875 | 1.40e-05 | 1.96e-04 | 22696 | 11798 | 10898 |
| European | Recorded Anxiety | Switching vs Continuation | Income | Income35-50k | 0.772 | 0.696 | 0.855 | 7.79e-07 | 1.09e-05 | 22696 | 11798 | 10898 |
| European | Recorded Anxiety | Switching vs Continuation | Income | Income50-75k | 0.715 | 0.651 | 0.784 | 1.28e-12 | 1.79e-11 | 22696 | 11798 | 10898 |
| European | Recorded Anxiety | Switching vs Continuation | Income | Income75-100k | 0.590 | 0.534 | 0.652 | <2.2e-16 | <2.2e-16 | 22696 | 11798 | 10898 |
| European | Recorded Anxiety | Switching vs Continuation | Income | Education_levelCollege Postgraduate | 0.806 | 0.653 | 0.995 | 4.42e-02 | 6.19e-01 | 22696 | 11798 | 10898 |
| European | Recorded Anxiety | Switching vs Continuation | Income | Education_levelCollege Undergraduate | 0.895 | 0.730 | 1.098 | 2.88e-01 | 1.00e+00 | 22696 | 11798 | 10898 |
| European | Recorded Anxiety | Switching vs Continuation | Income | Education_levelTwelve or GED | 1.132 | 0.916 | 1.399 | 2.53e-01 | 1.00e+00 | 22696 | 11798 | 10898 |
| European | Recorded Anxiety | Discontinuation vs Continuation | Insomnia | (Intercept) | 0.686 | 0.538 | 0.876 | 2.50e-03 | 3.49e-02 | 15204 | 11798 | 3406 |
| European | Recorded Anxiety | Discontinuation vs Continuation | Insomnia | age_at_first_AD | 1.088 | 1.047 | 1.131 | 1.76e-05 | 2.46e-04 | 15204 | 11798 | 3406 |
| European | Recorded Anxiety | Discontinuation vs Continuation | Insomnia | sex_at_birthMale | 1.216 | 1.118 | 1.323 | 5.63e-06 | 7.88e-05 | 15204 | 11798 | 3406 |
| European | Recorded Anxiety | Discontinuation vs Continuation | Insomnia | Income>200k | 0.437 | 0.369 | 0.519 | <2.2e-16 | <2.2e-16 | 15204 | 11798 | 3406 |
| European | Recorded Anxiety | Discontinuation vs Continuation | Insomnia | Income100-150k | 0.420 | 0.365 | 0.483 | <2.2e-16 | <2.2e-16 | 15204 | 11798 | 3406 |
| European | Recorded Anxiety | Discontinuation vs Continuation | Insomnia | Income150-200k | 0.405 | 0.337 | 0.486 | <2.2e-16 | <2.2e-16 | 15204 | 11798 | 3406 |
| European | Recorded Anxiety | Discontinuation vs Continuation | Insomnia | Income25-35k | 0.647 | 0.556 | 0.753 | 1.62e-08 | 2.27e-07 | 15204 | 11798 | 3406 |
| European | Recorded Anxiety | Discontinuation vs Continuation | Insomnia | Income35-50k | 0.572 | 0.495 | 0.662 | 5.85e-14 | 8.19e-13 | 15204 | 11798 | 3406 |
| European | Recorded Anxiety | Discontinuation vs Continuation | Insomnia | Income50-75k | 0.573 | 0.504 | 0.652 | <2.2e-16 | 3.53e-16 | 15204 | 11798 | 3406 |
| European | Recorded Anxiety | Discontinuation vs Continuation | Insomnia | Income75-100k | 0.480 | 0.416 | 0.553 | <2.2e-16 | <2.2e-16 | 15204 | 11798 | 3406 |
| European | Recorded Anxiety | Discontinuation vs Continuation | Insomnia | Education_levelCollege Postgraduate | 0.615 | 0.473 | 0.800 | 2.89e-04 | 4.04e-03 | 15204 | 11798 | 3406 |
| European | Recorded Anxiety | Discontinuation vs Continuation | Insomnia | Education_levelCollege Undergraduate | 0.685 | 0.533 | 0.880 | 3.09e-03 | 4.33e-02 | 15204 | 11798 | 3406 |
| European | Recorded Anxiety | Discontinuation vs Continuation | Insomnia | Education_levelTwelve or GED | 0.858 | 0.660 | 1.115 | 2.52e-01 | 1.00e+00 | 15204 | 11798 | 3406 |
| European | Recorded Anxiety | Discontinuation vs Continuation | Insomnia | Insomnia | 1.008 | 0.920 | 1.105 | 8.57e-01 | 1.00e+00 | 15204 | 11798 | 3406 |
| European | Recorded Anxiety | Switching vs Continuation | Insomnia | (Intercept) | 1.035 | 0.845 | 1.268 | 7.40e-01 | 1.00e+00 | 22696 | 11798 | 10898 |
| European | Recorded Anxiety | Switching vs Continuation | Insomnia | age_at_first_AD | 0.719 | 0.699 | 0.739 | <2.2e-16 | <2.2e-16 | 22696 | 11798 | 10898 |
| European | Recorded Anxiety | Switching vs Continuation | Insomnia | sex_at_birthMale | 1.126 | 1.059 | 1.197 | 1.52e-04 | 2.13e-03 | 22696 | 11798 | 10898 |
| European | Recorded Anxiety | Switching vs Continuation | Insomnia | Income>200k | 0.415 | 0.366 | 0.470 | <2.2e-16 | <2.2e-16 | 22696 | 11798 | 10898 |
| European | Recorded Anxiety | Switching vs Continuation | Insomnia | Income100-150k | 0.564 | 0.511 | 0.623 | <2.2e-16 | <2.2e-16 | 22696 | 11798 | 10898 |
| European | Recorded Anxiety | Switching vs Continuation | Insomnia | Income150-200k | 0.455 | 0.400 | 0.517 | <2.2e-16 | <2.2e-16 | 22696 | 11798 | 10898 |
| European | Recorded Anxiety | Switching vs Continuation | Insomnia | Income25-35k | 0.803 | 0.718 | 0.898 | 1.18e-04 | 1.65e-03 | 22696 | 11798 | 10898 |
| European | Recorded Anxiety | Switching vs Continuation | Insomnia | Income35-50k | 0.792 | 0.713 | 0.879 | 1.23e-05 | 1.72e-04 | 22696 | 11798 | 10898 |
| European | Recorded Anxiety | Switching vs Continuation | Insomnia | Income50-75k | 0.741 | 0.674 | 0.814 | 4.93e-10 | 6.90e-09 | 22696 | 11798 | 10898 |
| European | Recorded Anxiety | Switching vs Continuation | Insomnia | Income75-100k | 0.612 | 0.553 | 0.678 | <2.2e-16 | <2.2e-16 | 22696 | 11798 | 10898 |
| European | Recorded Anxiety | Switching vs Continuation | Insomnia | Education_levelCollege Postgraduate | 0.834 | 0.672 | 1.033 | 9.67e-02 | 1.00e+00 | 22696 | 11798 | 10898 |
| European | Recorded Anxiety | Switching vs Continuation | Insomnia | Education_levelCollege Undergraduate | 0.920 | 0.748 | 1.133 | 4.34e-01 | 1.00e+00 | 22696 | 11798 | 10898 |
| European | Recorded Anxiety | Switching vs Continuation | Insomnia | Education_levelTwelve or GED | 1.155 | 0.931 | 1.434 | 1.91e-01 | 1.00e+00 | 22696 | 11798 | 10898 |
| European | Recorded Anxiety | Switching vs Continuation | Insomnia | Insomnia | 2.333 | 2.198 | 2.476 | <2.2e-16 | <2.2e-16 | 22696 | 11798 | 10898 |
| European | Recorded Anxiety | Discontinuation vs Continuation | MDD CIDI-sf | (Intercept) | 0.117 | 0.027 | 0.518 | 4.70e-03 | 6.58e-02 | 2806 | 2260 | 546 |
| European | Recorded Anxiety | Discontinuation vs Continuation | MDD CIDI-sf | age_at_first_AD | 1.120 | 1.014 | 1.238 | 2.61e-02 | 3.66e-01 | 2806 | 2260 | 546 |
| European | Recorded Anxiety | Discontinuation vs Continuation | MDD CIDI-sf | sex_at_birthMale | 1.119 | 0.901 | 1.389 | 3.09e-01 | 1.00e+00 | 2806 | 2260 | 546 |
| European | Recorded Anxiety | Discontinuation vs Continuation | MDD CIDI-sf | Income>200k | 0.590 | 0.384 | 0.908 | 1.64e-02 | 2.30e-01 | 2806 | 2260 | 546 |
| European | Recorded Anxiety | Discontinuation vs Continuation | MDD CIDI-sf | Income100-150k | 0.610 | 0.429 | 0.867 | 5.91e-03 | 8.27e-02 | 2806 | 2260 | 546 |

| | | | | | | | | | | | | |
|---|---|---|---|---|---|---|---|---|---|---|---|---|
| European | Recorded Anxiety | Discontinuation vs Continuation | MDD CIDI-sf | Income150-200k | 0.618 | 0.403 | 0.946 | 2.67e-02 | 3.73e-01 | 2806 | 2260 | 546 |
| European | Recorded Anxiety | Discontinuation vs Continuation | MDD CIDI-sf | Income25-35k | 0.638 | 0.415 | 0.981 | 4.08e-02 | 5.71e-01 | 2806 | 2260 | 546 |
| European | Recorded Anxiety | Discontinuation vs Continuation | MDD CIDI-sf | Income35-50k | 0.695 | 0.471 | 1.027 | 6.78e-02 | 9.49e-01 | 2806 | 2260 | 546 |
| European | Recorded Anxiety | Discontinuation vs Continuation | MDD CIDI-sf | Income50-75k | 0.793 | 0.566 | 1.111 | 1.77e-01 | 1.00e+00 | 2806 | 2260 | 546 |
| European | Recorded Anxiety | Discontinuation vs Continuation | MDD CIDI-sf | Income75-100k | 0.603 | 0.416 | 0.873 | 7.42e-03 | 1.04e-01 | 2806 | 2260 | 546 |
| European | Recorded Anxiety | Discontinuation vs Continuation | MDD CIDI-sf | Education_levelCollege Postgraduate | 2.201 | 0.493 | 9.824 | 3.01e-01 | 1.00e+00 | 2806 | 2260 | 546 |
| European | Recorded Anxiety | Discontinuation vs Continuation | MDD CIDI-sf | Education_levelCollege Undergraduate | 2.573 | 0.582 | 11.370 | 2.12e-01 | 1.00e+00 | 2806 | 2260 | 546 |
| European | Recorded Anxiety | Discontinuation vs Continuation | MDD CIDI-sf | Education_levelTwelve or GED | 2.204 | 0.483 | 10.050 | 3.07e-01 | 1.00e+00 | 2806 | 2260 | 546 |
| European | Recorded Anxiety | Discontinuation vs Continuation | MDD CIDI-sf | MDD CIDI-sf | 1.318 | 1.075 | 1.614 | 7.81e-03 | 1.09e-01 | 2806 | 2260 | 546 |
| European | Recorded Anxiety | Switching vs Continuation | MDD CIDI-sf | (Intercept) | 0.385 | 0.175 | 0.847 | 1.77e-02 | 2.48e-01 | 4388 | 2260 | 2128 |
| European | Recorded Anxiety | Switching vs Continuation | MDD CIDI-sf | age_at_first_AD | 0.691 | 0.645 | 0.740 | <2.2e-16 | <2.2e-16 | 4388 | 2260 | 2128 |
| European | Recorded Anxiety | Switching vs Continuation | MDD CIDI-sf | sex_at_birthMale | 1.255 | 1.087 | 1.450 | 2.02e-03 | 2.83e-02 | 4388 | 2260 | 2128 |
| European | Recorded Anxiety | Switching vs Continuation | MDD CIDI-sf | Income>200k | 0.501 | 0.375 | 0.670 | 3.05e-06 | 4.27e-05 | 4388 | 2260 | 2128 |
| European | Recorded Anxiety | Switching vs Continuation | MDD CIDI-sf | Income100-150k | 0.575 | 0.455 | 0.727 | 3.87e-06 | 5.42e-05 | 4388 | 2260 | 2128 |
| European | Recorded Anxiety | Switching vs Continuation | MDD CIDI-sf | Income150-200k | 0.461 | 0.344 | 0.618 | 2.16e-07 | 3.03e-06 | 4388 | 2260 | 2128 |
| European | Recorded Anxiety | Switching vs Continuation | MDD CIDI-sf | Income25-35k | 0.758 | 0.575 | 0.999 | 4.94e-02 | 6.91e-01 | 4388 | 2260 | 2128 |
| European | Recorded Anxiety | Switching vs Continuation | MDD CIDI-sf | Income35-50k | 0.851 | 0.661 | 1.095 | 2.10e-01 | 1.00e+00 | 4388 | 2260 | 2128 |
| European | Recorded Anxiety | Switching vs Continuation | MDD CIDI-sf | Income50-75k | 0.772 | 0.614 | 0.969 | 2.58e-02 | 3.62e-01 | 4388 | 2260 | 2128 |
| European | Recorded Anxiety | Switching vs Continuation | MDD CIDI-sf | Income75-100k | 0.667 | 0.524 | 0.850 | 1.03e-03 | 1.44e-02 | 4388 | 2260 | 2128 |
| European | Recorded Anxiety | Switching vs Continuation | MDD CIDI-sf | Education_levelCollege Postgraduate | 1.785 | 0.805 | 3.960 | 1.54e-01 | 1.00e+00 | 4388 | 2260 | 2128 |
| European | Recorded Anxiety | Switching vs Continuation | MDD CIDI-sf | Education_levelCollege Undergraduate | 2.094 | 0.952 | 4.605 | 6.62e-02 | 9.27e-01 | 4388 | 2260 | 2128 |
| European | Recorded Anxiety | Switching vs Continuation | MDD CIDI-sf | Education_levelTwelve or GED | 2.275 | 1.012 | 5.114 | 4.68e-02 | 6.55e-01 | 4388 | 2260 | 2128 |
| European | Recorded Anxiety | Switching vs Continuation | MDD CIDI-sf | MDD CIDI-sf | 1.990 | 1.733 | 2.286 | <2.2e-16 | <2.2e-16 | 4388 | 2260 | 2128 |
| European | Recorded Anxiety | Discontinuation vs Continuation | MDD_symptom_count | (Intercept) | 0.223 | 0.076 | 0.656 | 6.42e-03 | 8.99e-02 | 3523 | 2850 | 673 |
| European | Recorded Anxiety | Discontinuation vs Continuation | MDD_symptom_count | age_at_first_AD | 1.125 | 1.027 | 1.233 | 1.12e-02 | 1.56e-01 | 3523 | 2850 | 673 |
| European | Recorded Anxiety | Discontinuation vs Continuation | MDD_symptom_count | sex_at_birthMale | 1.094 | 0.899 | 1.331 | 3.70e-01 | 1.00e+00 | 3523 | 2850 | 673 |
| European | Recorded Anxiety | Discontinuation vs Continuation | MDD_symptom_count | Income>200k | 0.616 | 0.418 | 0.907 | 1.42e-02 | 1.99e-01 | 3523 | 2850 | 673 |
| European | Recorded Anxiety | Discontinuation vs Continuation | MDD_symptom_count | Income100-150k | 0.610 | 0.444 | 0.837 | 2.23e-03 | 3.12e-02 | 3523 | 2850 | 673 |
| European | Recorded Anxiety | Discontinuation vs Continuation | MDD_symptom_count | Income150-200k | 0.620 | 0.423 | 0.910 | 1.46e-02 | 2.05e-01 | 3523 | 2850 | 673 |
| European | Recorded Anxiety | Discontinuation vs Continuation | MDD_symptom_count | Income25-35k | 0.569 | 0.385 | 0.841 | 4.64e-03 | 6.50e-02 | 3523 | 2850 | 673 |
| European | Recorded Anxiety | Discontinuation vs Continuation | MDD_symptom_count | Income35-50k | 0.727 | 0.514 | 1.029 | 7.24e-02 | 1.00e+00 | 3523 | 2850 | 673 |
| European | Recorded Anxiety | Discontinuation vs Continuation | MDD_symptom_count | Income50-75k | 0.760 | 0.560 | 1.031 | 7.76e-02 | 1.00e+00 | 3523 | 2850 | 673 |
| European | Recorded Anxiety | Discontinuation vs Continuation | MDD_symptom_count | Income75-100k | 0.598 | 0.429 | 0.836 | 2.58e-03 | 3.61e-02 | 3523 | 2850 | 673 |
| European | Recorded Anxiety | Discontinuation vs Continuation | MDD_symptom_count | Education_levelCollege Postgraduate | 1.394 | 0.464 | 4.190 | 5.54e-01 | 1.00e+00 | 3523 | 2850 | 673 |
| European | Recorded Anxiety | Discontinuation vs Continuation | MDD_symptom_count | Education_levelCollege Undergraduate | 1.555 | 0.523 | 4.620 | 4.27e-01 | 1.00e+00 | 3523 | 2850 | 673 |
| European | Recorded Anxiety | Discontinuation vs Continuation | MDD_symptom_count | Education_levelTwelve or GED | 1.412 | 0.460 | 4.339 | 5.47e-01 | 1.00e+00 | 3523 | 2850 | 673 |
| European | Recorded Anxiety | Discontinuation vs Continuation | MDD_symptom_count | MDD_symptom_count | 1.130 | 1.033 | 1.235 | 7.43e-03 | 1.04e-01 | 3523 | 2850 | 673 |
| European | Recorded Anxiety | Switching vs Continuation | MDD_symptom_count | (Intercept) | 0.747 | 0.394 | 1.417 | 3.72e-01 | 1.00e+00 | 5474 | 2850 | 2624 |
| European | Recorded Anxiety | Switching vs Continuation | MDD_symptom_count | age_at_first_AD | 0.701 | 0.659 | 0.746 | <2.2e-16 | <2.2e-16 | 5474 | 2850 | 2624 |
| European | Recorded Anxiety | Switching vs Continuation | MDD_symptom_count | sex_at_birthMale | 1.307 | 1.149 | 1.487 | 4.84e-05 | 6.77e-04 | 5474 | 2850 | 2624 |
| European | Recorded Anxiety | Switching vs Continuation | MDD_symptom_count | Income>200k | 0.544 | 0.420 | 0.707 | 4.87e-06 | 6.82e-05 | 5474 | 2850 | 2624 |
| European | Recorded Anxiety | Switching vs Continuation | MDD_symptom_count | Income100-150k | 0.601 | 0.486 | 0.743 | 2.36e-06 | 3.30e-05 | 5474 | 2850 | 2624 |
| European | Recorded Anxiety | Switching vs Continuation | MDD_symptom_count | Income150-200k | 0.490 | 0.376 | 0.637 | 1.01e-07 | 1.41e-06 | 5474 | 2850 | 2624 |
| European | Recorded Anxiety | Switching vs Continuation | MDD_symptom_count | Income25-35k | 0.732 | 0.573 | 0.936 | 1.27e-02 | 1.78e-01 | 5474 | 2850 | 2624 |
| European | Recorded Anxiety | Switching vs Continuation | MDD_symptom_count | Income35-50k | 0.904 | 0.721 | 1.134 | 3.84e-01 | 1.00e+00 | 5474 | 2850 | 2624 |
| European | Recorded Anxiety | Switching vs Continuation | MDD_symptom_count | Income50-75k | 0.795 | 0.648 | 0.977 | 2.88e-02 | 4.03e-01 | 5474 | 2850 | 2624 |
| European | Recorded Anxiety | Switching vs Continuation | MDD_symptom_count | Income75-100k | 0.704 | 0.566 | 0.874 | 1.47e-03 | 2.06e-02 | 5474 | 2850 | 2624 |
| European | Recorded Anxiety | Switching vs Continuation | MDD_symptom_count | Education_levelCollege Postgraduate | 1.329 | 0.690 | 2.557 | 3.95e-01 | 1.00e+00 | 5474 | 2850 | 2624 |
| European | Recorded Anxiety | Switching vs Continuation | MDD_symptom_count | Education_levelCollege Undergraduate | 1.546 | 0.810 | 2.951 | 1.86e-01 | 1.00e+00 | 5474 | 2850 | 2624 |
| European | Recorded Anxiety | Switching vs Continuation | MDD_symptom_count | Education_levelTwelve or GED | 1.881 | 0.966 | 3.664 | 6.32e-02 | 8.84e-01 | 5474 | 2850 | 2624 |
| European | Recorded Anxiety | Switching vs Continuation | MDD_symptom_count | MDD_symptom_count | 1.372 | 1.291 | 1.458 | <2.2e-16 | <2.2e-16 | 5474 | 2850 | 2624 |
| European | Recorded Anxiety | Discontinuation vs Continuation | Migraine | (Intercept) | 0.704 | 0.552 | 0.899 | 4.85e-03 | 6.79e-02 | 15204 | 11798 | 3406 |
| European | Recorded Anxiety | Discontinuation vs Continuation | Migraine | age_at_first_AD | 1.087 | 1.046 | 1.129 | 1.85e-05 | 2.59e-04 | 15204 | 11798 | 3406 |
| European | Recorded Anxiety | Discontinuation vs Continuation | Migraine | sex_at_birthMale | 1.205 | 1.106 | 1.312 | 1.79e-05 | 2.51e-04 | 15204 | 11798 | 3406 |
| European | Recorded Anxiety | Discontinuation vs Continuation | Migraine | Income>200k | 0.437 | 0.369 | 0.519 | <2.2e-16 | <2.2e-16 | 15204 | 11798 | 3406 |
| European | Recorded Anxiety | Discontinuation vs Continuation | Migraine | Income100-150k | 0.420 | 0.366 | 0.483 | <2.2e-16 | <2.2e-16 | 15204 | 11798 | 3406 |
| European | Recorded Anxiety | Discontinuation vs Continuation | Migraine | Income150-200k | 0.405 | 0.337 | 0.486 | <2.2e-16 | <2.2e-16 | 15204 | 11798 | 3406 |
| European | Recorded Anxiety | Discontinuation vs Continuation | Migraine | Income25-35k | 0.647 | 0.557 | 0.753 | 1.71e-08 | 2.40e-07 | 15204 | 11798 | 3406 |
| European | Recorded Anxiety | Discontinuation vs Continuation | Migraine | Income35-50k | 0.572 | 0.495 | 0.662 | 5.98e-14 | 8.37e-13 | 15204 | 11798 | 3406 |
| European | Recorded Anxiety | Discontinuation vs Continuation | Migraine | Income50-75k | 0.574 | 0.505 | 0.653 | <2.2e-16 | 4.30e-16 | 15204 | 11798 | 3406 |
| European | Recorded Anxiety | Discontinuation vs Continuation | Migraine | Income75-100k | 0.479 | 0.416 | 0.553 | <2.2e-16 | <2.2e-16 | 15204 | 11798 | 3406 |
| European | Recorded Anxiety | Discontinuation vs Continuation | Migraine | Education_levelCollege Postgraduate | 0.616 | 0.474 | 0.801 | 3.06e-04 | 4.28e-03 | 15204 | 11798 | 3406 |
| European | Recorded Anxiety | Discontinuation vs Continuation | Migraine | Education_levelCollege Undergraduate | 0.686 | 0.534 | 0.881 | 3.19e-03 | 4.47e-02 | 15204 | 11798 | 3406 |
| European | Recorded Anxiety | Discontinuation vs Continuation | Migraine | Education_levelTwelve or GED | 0.858 | 0.660 | 1.115 | 2.52e-01 | 1.00e+00 | 15204 | 11798 | 3406 |
| European | Recorded Anxiety | Discontinuation vs Continuation | Migraine | Migraine | 0.928 | 0.851 | 1.011 | 8.82e-02 | 1.00e+00 | 15204 | 11798 | 3406 |
| European | Recorded Anxiety | Switching vs Continuation | Migraine | (Intercept) | 1.123 | 0.919 | 1.373 | 2.58e-01 | 1.00e+00 | 22696 | 11798 | 10898 |

| | | | | | | | | | | | | |
|---|---|---|---|---|---|---|---|---|---|---|---|---|
| European | Recorded Anxiety | Switching vs Continuation | Migraine | age_at_first_AD | 0.767 | 0.746 | 0.789 | <2.2e-16 | <2.2e-16 | 22696 | 11798 | 10898 |
| European | Recorded Anxiety | Switching vs Continuation | Migraine | sex_at_birthMale | 1.268 | 1.193 | 1.348 | 3.00e-14 | 4.20e-13 | 22696 | 11798 | 10898 |
| European | Recorded Anxiety | Switching vs Continuation | Migraine | Income>200k | 0.413 | 0.365 | 0.468 | <2.2e-16 | <2.2e-16 | 22696 | 11798 | 10898 |
| European | Recorded Anxiety | Switching vs Continuation | Migraine | Income100-150k | 0.548 | 0.497 | 0.604 | <2.2e-16 | <2.2e-16 | 22696 | 11798 | 10898 |
| European | Recorded Anxiety | Switching vs Continuation | Migraine | Income150-200k | 0.449 | 0.395 | 0.510 | <2.2e-16 | <2.2e-16 | 22696 | 11798 | 10898 |
| European | Recorded Anxiety | Switching vs Continuation | Migraine | Income25-35k | 0.780 | 0.698 | 0.871 | 1.04e-05 | 1.45e-04 | 22696 | 11798 | 10898 |
| European | Recorded Anxiety | Switching vs Continuation | Migraine | Income35-50k | 0.775 | 0.699 | 0.860 | 1.62e-06 | 2.27e-05 | 22696 | 11798 | 10898 |
| European | Recorded Anxiety | Switching vs Continuation | Migraine | Income50-75k | 0.721 | 0.656 | 0.792 | 8.06e-12 | 1.13e-10 | 22696 | 11798 | 10898 |
| European | Recorded Anxiety | Switching vs Continuation | Migraine | Income75-100k | 0.598 | 0.540 | 0.662 | <2.2e-16 | <2.2e-16 | 22696 | 11798 | 10898 |
| European | Recorded Anxiety | Switching vs Continuation | Migraine | Education_levelCollege Postgraduate | 0.772 | 0.624 | 0.955 | 1.70e-02 | 2.38e-01 | 22696 | 11798 | 10898 |
| European | Recorded Anxiety | Switching vs Continuation | Migraine | Education_levelCollege Undergraduate | 0.860 | 0.700 | 1.056 | 1.50e-01 | 1.00e+00 | 22696 | 11798 | 10898 |
| European | Recorded Anxiety | Switching vs Continuation | Migraine | Education_levelTwelve or GED | 1.088 | 0.878 | 1.348 | 4.39e-01 | 1.00e+00 | 22696 | 11798 | 10898 |
| European | Recorded Anxiety | Switching vs Continuation | Migraine | Migraine | 1.891 | 1.788 | 2.000 | <2.2e-16 | <2.2e-16 | 22696 | 11798 | 10898 |
| European | Recorded Anxiety | Discontinuation vs Continuation | Recorded Depression | (Intercept) | 0.745 | 0.582 | 0.955 | 1.98e-02 | 2.78e-01 | 15204 | 11798 | 3406 |
| European | Recorded Anxiety | Discontinuation vs Continuation | Recorded Depression | age_at_first_AD | 1.090 | 1.049 | 1.132 | 9.65e-06 | 1.35e-04 | 15204 | 11798 | 3406 |
| European | Recorded Anxiety | Discontinuation vs Continuation | Recorded Depression | sex_at_birthMale | 1.216 | 1.118 | 1.323 | 5.54e-06 | 7.75e-05 | 15204 | 11798 | 3406 |
| European | Recorded Anxiety | Discontinuation vs Continuation | Recorded Depression | Income>200k | 0.422 | 0.355 | 0.501 | <2.2e-16 | <2.2e-16 | 15204 | 11798 | 3406 |
| European | Recorded Anxiety | Discontinuation vs Continuation | Recorded Depression | Income100-150k | 0.409 | 0.356 | 0.471 | <2.2e-16 | <2.2e-16 | 15204 | 11798 | 3406 |
| European | Recorded Anxiety | Discontinuation vs Continuation | Recorded Depression | Income150-200k | 0.393 | 0.327 | 0.473 | <2.2e-16 | <2.2e-16 | 15204 | 11798 | 3406 |
| European | Recorded Anxiety | Discontinuation vs Continuation | Recorded Depression | Income25-35k | 0.642 | 0.552 | 0.747 | 9.67e-09 | 1.35e-07 | 15204 | 11798 | 3406 |
| European | Recorded Anxiety | Discontinuation vs Continuation | Recorded Depression | Income35-50k | 0.565 | 0.488 | 0.654 | 1.79e-14 | 2.51e-13 | 15204 | 11798 | 3406 |
| European | Recorded Anxiety | Discontinuation vs Continuation | Recorded Depression | Income50-75k | 0.564 | 0.495 | 0.641 | <2.2e-16 | <2.2e-16 | 15204 | 11798 | 3406 |
| European | Recorded Anxiety | Discontinuation vs Continuation | Recorded Depression | Income75-100k | 0.469 | 0.407 | 0.541 | <2.2e-16 | <2.2e-16 | 15204 | 11798 | 3406 |
| European | Recorded Anxiety | Discontinuation vs Continuation | Recorded Depression | Education_levelCollege Postgraduate | 0.615 | 0.472 | 0.800 | 2.90e-04 | 4.05e-03 | 15204 | 11798 | 3406 |
| European | Recorded Anxiety | Discontinuation vs Continuation | Recorded Depression | Education_levelCollege Undergraduate | 0.684 | 0.532 | 0.879 | 2.98e-03 | 4.17e-02 | 15204 | 11798 | 3406 |
| European | Recorded Anxiety | Discontinuation vs Continuation | Recorded Depression | Education_levelTwelve or GED | 0.855 | 0.658 | 1.112 | 2.43e-01 | 1.00e+00 | 15204 | 11798 | 3406 |
| European | Recorded Anxiety | Discontinuation vs Continuation | Recorded Depression | Recorded Depression | 0.877 | 0.810 | 0.948 | 1.03e-03 | 1.45e-02 | 15204 | 11798 | 3406 |
| European | Recorded Anxiety | Switching vs Continuation | Recorded Depression | (Intercept) | 0.600 | 0.486 | 0.740 | 1.75e-06 | 2.45e-05 | 22696 | 11798 | 10898 |
| European | Recorded Anxiety | Switching vs Continuation | Recorded Depression | age_at_first_AD | 0.748 | 0.727 | 0.770 | <2.2e-16 | <2.2e-16 | 22696 | 11798 | 10898 |
| European | Recorded Anxiety | Switching vs Continuation | Recorded Depression | sex_at_birthMale | 1.135 | 1.066 | 1.208 | 6.92e-05 | 9.68e-04 | 22696 | 11798 | 10898 |
| European | Recorded Anxiety | Switching vs Continuation | Recorded Depression | Income>200k | 0.538 | 0.473 | 0.612 | <2.2e-16 | <2.2e-16 | 22696 | 11798 | 10898 |
| European | Recorded Anxiety | Switching vs Continuation | Recorded Depression | Income100-150k | 0.658 | 0.594 | 0.727 | 3.53e-16 | 4.94e-15 | 22696 | 11798 | 10898 |
| European | Recorded Anxiety | Switching vs Continuation | Recorded Depression | Income150-200k | 0.563 | 0.494 | 0.643 | <2.2e-16 | <2.2e-16 | 22696 | 11798 | 10898 |
| European | Recorded Anxiety | Switching vs Continuation | Recorded Depression | Income25-35k | 0.827 | 0.739 | 0.927 | 1.04e-03 | 1.45e-02 | 22696 | 11798 | 10898 |
| European | Recorded Anxiety | Switching vs Continuation | Recorded Depression | Income35-50k | 0.852 | 0.766 | 0.948 | 3.36e-03 | 4.70e-02 | 22696 | 11798 | 10898 |
| European | Recorded Anxiety | Switching vs Continuation | Recorded Depression | Income50-75k | 0.805 | 0.731 | 0.886 | 9.69e-06 | 1.36e-04 | 22696 | 11798 | 10898 |
| European | Recorded Anxiety | Switching vs Continuation | Recorded Depression | Income75-100k | 0.693 | 0.624 | 0.769 | 4.81e-12 | 6.74e-11 | 22696 | 11798 | 10898 |
| European | Recorded Anxiety | Switching vs Continuation | Recorded Depression | Education_levelCollege Postgraduate | 0.791 | 0.636 | 0.984 | 3.55e-02 | 4.97e-01 | 22696 | 11798 | 10898 |
| European | Recorded Anxiety | Switching vs Continuation | Recorded Depression | Education_levelCollege Undergraduate | 0.882 | 0.714 | 1.090 | 2.46e-01 | 1.00e+00 | 22696 | 11798 | 10898 |
| European | Recorded Anxiety | Switching vs Continuation | Recorded Depression | Education_levelTwelve or GED | 1.130 | 0.907 | 1.408 | 2.78e-01 | 1.00e+00 | 22696 | 11798 | 10898 |
| European | Recorded Anxiety | Switching vs Continuation | Recorded Depression | Recorded Depression | 3.324 | 3.135 | 3.523 | <2.2e-16 | <2.2e-16 | 22696 | 11798 | 10898 |
| European | Recorded Anxiety | Discontinuation vs Continuation | Smoking_Frequency | (Intercept) | 0.526 | 0.406 | 0.682 | 1.21e-06 | 1.69e-05 | 14887 | 11556 | 3331 |
| European | Recorded Anxiety | Discontinuation vs Continuation | Smoking_Frequency | age_at_first_AD | 1.109 | 1.066 | 1.154 | 3.05e-07 | 4.27e-06 | 14887 | 11556 | 3331 |
| European | Recorded Anxiety | Discontinuation vs Continuation | Smoking_Frequency | sex_at_birthMale | 1.187 | 1.089 | 1.293 | 9.37e-05 | 1.31e-03 | 14887 | 11556 | 3331 |
| European | Recorded Anxiety | Discontinuation vs Continuation | Smoking_Frequency | Income>200k | 0.469 | 0.394 | 0.559 | <2.2e-16 | 4.16e-16 | 14887 | 11556 | 3331 |
| European | Recorded Anxiety | Discontinuation vs Continuation | Smoking_Frequency | Income100-150k | 0.453 | 0.393 | 0.523 | <2.2e-16 | <2.2e-16 | 14887 | 11556 | 3331 |
| European | Recorded Anxiety | Discontinuation vs Continuation | Smoking_Frequency | Income150-200k | 0.449 | 0.372 | 0.541 | <2.2e-16 | 5.06e-16 | 14887 | 11556 | 3331 |
| European | Recorded Anxiety | Discontinuation vs Continuation | Smoking_Frequency | Income25-35k | 0.686 | 0.588 | 0.800 | 1.46e-06 | 2.04e-05 | 14887 | 11556 | 3331 |
| European | Recorded Anxiety | Discontinuation vs Continuation | Smoking_Frequency | Income35-50k | 0.613 | 0.528 | 0.711 | 1.09e-10 | 1.53e-09 | 14887 | 11556 | 3331 |
| European | Recorded Anxiety | Discontinuation vs Continuation | Smoking_Frequency | Income50-75k | 0.613 | 0.537 | 0.699 | 3.77e-13 | 5.28e-12 | 14887 | 11556 | 3331 |
| European | Recorded Anxiety | Discontinuation vs Continuation | Smoking_Frequency | Income75-100k | 0.517 | 0.447 | 0.599 | <2.2e-16 | <2.2e-16 | 14887 | 11556 | 3331 |
| European | Recorded Anxiety | Discontinuation vs Continuation | Smoking_Frequency | Education_levelCollege Postgraduate | 0.737 | 0.562 | 0.967 | 2.79e-02 | 3.90e-01 | 14887 | 11556 | 3331 |
| European | Recorded Anxiety | Discontinuation vs Continuation | Smoking_Frequency | Education_levelCollege Undergraduate | 0.810 | 0.626 | 1.049 | 1.10e-01 | 1.00e+00 | 14887 | 11556 | 3331 |
| European | Recorded Anxiety | Discontinuation vs Continuation | Smoking_Frequency | Education_levelTwelve or GED | 0.981 | 0.750 | 1.283 | 8.88e-01 | 1.00e+00 | 14887 | 11556 | 3331 |
| European | Recorded Anxiety | Discontinuation vs Continuation | Smoking_Frequency | Smoking_FrequencyEvery Day | 1.620 | 1.397 | 1.879 | 1.67e-10 | 2.34e-09 | 14887 | 11556 | 3331 |
| European | Recorded Anxiety | Discontinuation vs Continuation | Smoking_Frequency | Smoking_FrequencyNot At All (former) | 1.003 | 0.918 | 1.097 | 9.39e-01 | 1.00e+00 | 14887 | 11556 | 3331 |
| European | Recorded Anxiety | Discontinuation vs Continuation | Smoking_Frequency | Smoking_FrequencySome Days | 1.351 | 1.098 | 1.663 | 4.54e-03 | 6.36e-02 | 14887 | 11556 | 3331 |
| European | Recorded Anxiety | Switching vs Continuation | Smoking_Frequency | (Intercept) | 1.108 | 0.902 | 1.361 | 3.27e-01 | 1.00e+00 | 22221 | 11556 | 10665 |
| European | Recorded Anxiety | Switching vs Continuation | Smoking_Frequency | age_at_first_AD | 0.746 | 0.725 | 0.768 | <2.2e-16 | <2.2e-16 | 22221 | 11556 | 10665 |
| European | Recorded Anxiety | Switching vs Continuation | Smoking_Frequency | sex_at_birthMale | 1.132 | 1.065 | 1.203 | 7.08e-05 | 9.92e-04 | 22221 | 11556 | 10665 |
| European | Recorded Anxiety | Switching vs Continuation | Smoking_Frequency | Income>200k | 0.422 | 0.373 | 0.479 | <2.2e-16 | <2.2e-16 | 22221 | 11556 | 10665 |
| European | Recorded Anxiety | Switching vs Continuation | Smoking_Frequency | Income100-150k | 0.568 | 0.514 | 0.627 | <2.2e-16 | <2.2e-16 | 22221 | 11556 | 10665 |
| European | Recorded Anxiety | Switching vs Continuation | Smoking_Frequency | Income150-200k | 0.464 | 0.408 | 0.528 | <2.2e-16 | <2.2e-16 | 22221 | 11556 | 10665 |
| European | Recorded Anxiety | Switching vs Continuation | Smoking_Frequency | Income25-35k | 0.800 | 0.716 | 0.895 | 9.02e-05 | 1.26e-03 | 22221 | 11556 | 10665 |
| European | Recorded Anxiety | Switching vs Continuation | Smoking_Frequency | Income35-50k | 0.803 | 0.723 | 0.892 | 4.10e-05 | 5.75e-04 | 22221 | 11556 | 10665 |
| European | Recorded Anxiety | Switching vs Continuation | Smoking_Frequency | Income50-75k | 0.737 | 0.671 | 0.811 | 3.30e-10 | 4.62e-09 | 22221 | 11556 | 10665 |

| | | | | | | | | | | | | |
|---|---|---|---|---|---|---|---|---|---|---|---|---|
| European | Recorded Anxiety | Switching vs Continuation | Smoking_Frequency | Income75-100k | 0.618 | 0.558 | 0.685 | <2.2e-16 | <2.2e-16 | 22221 | 11556 | 10665 |
| European | Recorded Anxiety | Switching vs Continuation | Smoking_Frequency | Education_levelCollege Postgraduate | 0.926 | 0.747 | 1.147 | 4.80e-01 | 1.00e+00 | 22221 | 11556 | 10665 |
| European | Recorded Anxiety | Switching vs Continuation | Smoking_Frequency | Education_levelCollege Undergraduate | 0.998 | 0.811 | 1.227 | 9.83e-01 | 1.00e+00 | 22221 | 11556 | 10665 |
| European | Recorded Anxiety | Switching vs Continuation | Smoking_Frequency | Education_levelTwelve or GED | 1.215 | 0.981 | 1.506 | 7.47e-02 | 1.00e+00 | 22221 | 11556 | 10665 |
| European | Recorded Anxiety | Switching vs Continuation | Smoking_Frequency | Smoking_FrequencyEvery Day | 1.597 | 1.432 | 1.782 | <2.2e-16 | 7.45e-16 | 22221 | 11556 | 10665 |
| European | Recorded Anxiety | Switching vs Continuation | Smoking_Frequency | Smoking_FrequencyNot At All (former) | 1.225 | 1.153 | 1.302 | 6.15e-11 | 8.61e-10 | 22221 | 11556 | 10665 |
| European | Recorded Anxiety | Switching vs Continuation | Smoking_Frequency | Smoking_FrequencySome Days | 1.188 | 1.019 | 1.385 | 2.76e-02 | 3.86e-01 | 22221 | 11556 | 10665 |
| European | Recorded Anxiety | Discontinuation vs Continuation | Stress | (Intercept) | 0.431 | 0.258 | 0.720 | 1.32e-03 | 1.84e-02 | 8894 | 7158 | 1736 |
| European | Recorded Anxiety | Discontinuation vs Continuation | Stress | age_at_first_AD | 1.115 | 1.056 | 1.176 | 7.66e-05 | 1.07e-03 | 8894 | 7158 | 1736 |
| European | Recorded Anxiety | Discontinuation vs Continuation | Stress | sex_at_birthMale | 1.093 | 0.970 | 1.232 | 1.46e-01 | 1.00e+00 | 8894 | 7158 | 1736 |
| European | Recorded Anxiety | Discontinuation vs Continuation | Stress | Income>200k | 0.503 | 0.397 | 0.638 | 1.28e-08 | 1.80e-07 | 8894 | 7158 | 1736 |
| European | Recorded Anxiety | Discontinuation vs Continuation | Stress | Income100-150k | 0.542 | 0.445 | 0.660 | 1.04e-09 | 1.46e-08 | 8894 | 7158 | 1736 |
| European | Recorded Anxiety | Discontinuation vs Continuation | Stress | Income150-200k | 0.510 | 0.400 | 0.649 | 4.37e-08 | 6.12e-07 | 8894 | 7158 | 1736 |
| European | Recorded Anxiety | Discontinuation vs Continuation | Stress | Income25-35k | 0.647 | 0.513 | 0.816 | 2.31e-04 | 3.23e-03 | 8894 | 7158 | 1736 |
| European | Recorded Anxiety | Discontinuation vs Continuation | Stress | Income35-50k | 0.601 | 0.483 | 0.747 | 4.93e-06 | 6.90e-05 | 8894 | 7158 | 1736 |
| European | Recorded Anxiety | Discontinuation vs Continuation | Stress | Income50-75k | 0.670 | 0.555 | 0.809 | 3.23e-05 | 4.52e-04 | 8894 | 7158 | 1736 |
| European | Recorded Anxiety | Discontinuation vs Continuation | Stress | Income75-100k | 0.573 | 0.467 | 0.702 | 8.16e-08 | 1.14e-06 | 8894 | 7158 | 1736 |
| European | Recorded Anxiety | Discontinuation vs Continuation | Stress | Education_levelCollege Postgraduate | 0.792 | 0.468 | 1.340 | 3.84e-01 | 1.00e+00 | 8894 | 7158 | 1736 |
| European | Recorded Anxiety | Discontinuation vs Continuation | Stress | Education_levelCollege Undergraduate | 0.835 | 0.498 | 1.401 | 4.95e-01 | 1.00e+00 | 8894 | 7158 | 1736 |
| European | Recorded Anxiety | Discontinuation vs Continuation | Stress | Education_levelTwelve or GED | 0.824 | 0.482 | 1.410 | 4.80e-01 | 1.00e+00 | 8894 | 7158 | 1736 |
| European | Recorded Anxiety | Discontinuation vs Continuation | Stress | StressAlmost Never | 1.004 | 0.796 | 1.265 | 9.76e-01 | 1.00e+00 | 8894 | 7158 | 1736 |
| European | Recorded Anxiety | Discontinuation vs Continuation | Stress | StressFairly Often | 0.886 | 0.699 | 1.124 | 3.18e-01 | 1.00e+00 | 8894 | 7158 | 1736 |
| European | Recorded Anxiety | Discontinuation vs Continuation | Stress | StressSometimes | 1.219 | 1.084 | 1.370 | 9.22e-04 | 1.29e-02 | 8894 | 7158 | 1736 |
| European | Recorded Anxiety | Discontinuation vs Continuation | Stress | StressVery Often | 1.235 | 0.935 | 1.631 | 1.38e-01 | 1.00e+00 | 8894 | 7158 | 1736 |
| European | Recorded Anxiety | Switching vs Continuation | Stress | (Intercept) | 1.384 | 0.974 | 1.967 | 6.97e-02 | 9.76e-01 | 13672 | 7158 | 6514 |
| European | Recorded Anxiety | Switching vs Continuation | Stress | age_at_first_AD | 0.736 | 0.709 | 0.763 | <2.2e-16 | <2.2e-16 | 13672 | 7158 | 6514 |
| European | Recorded Anxiety | Switching vs Continuation | Stress | sex_at_birthMale | 1.229 | 1.135 | 1.330 | 3.41e-07 | 4.77e-06 | 13672 | 7158 | 6514 |
| European | Recorded Anxiety | Switching vs Continuation | Stress | Income>200k | 0.398 | 0.338 | 0.468 | <2.2e-16 | <2.2e-16 | 13672 | 7158 | 6514 |
| European | Recorded Anxiety | Switching vs Continuation | Stress | Income100-150k | 0.534 | 0.469 | 0.610 | <2.2e-16 | <2.2e-16 | 13672 | 7158 | 6514 |
| European | Recorded Anxiety | Switching vs Continuation | Stress | Income150-200k | 0.400 | 0.340 | 0.472 | <2.2e-16 | <2.2e-16 | 13672 | 7158 | 6514 |
| European | Recorded Anxiety | Switching vs Continuation | Stress | Income25-35k | 0.737 | 0.632 | 0.859 | 9.05e-05 | 1.27e-03 | 13672 | 7158 | 6514 |
| European | Recorded Anxiety | Switching vs Continuation | Stress | Income35-50k | 0.749 | 0.649 | 0.863 | 6.84e-05 | 9.58e-04 | 13672 | 7158 | 6514 |
| European | Recorded Anxiety | Switching vs Continuation | Stress | Income50-75k | 0.696 | 0.612 | 0.790 | 2.45e-08 | 3.44e-07 | 13672 | 7158 | 6514 |
| European | Recorded Anxiety | Switching vs Continuation | Stress | Income75-100k | 0.600 | 0.524 | 0.687 | 1.44e-13 | 2.02e-12 | 13672 | 7158 | 6514 |
| European | Recorded Anxiety | Switching vs Continuation | Stress | Education_levelCollege Postgraduate | 0.774 | 0.540 | 1.109 | 1.63e-01 | 1.00e+00 | 13672 | 7158 | 6514 |
| European | Recorded Anxiety | Switching vs Continuation | Stress | Education_levelCollege Undergraduate | 0.874 | 0.614 | 1.246 | 4.57e-01 | 1.00e+00 | 13672 | 7158 | 6514 |
| European | Recorded Anxiety | Switching vs Continuation | Stress | Education_levelTwelve or GED | 1.021 | 0.708 | 1.471 | 9.13e-01 | 1.00e+00 | 13672 | 7158 | 6514 |
| European | Recorded Anxiety | Switching vs Continuation | Stress | StressAlmost Never | 1.015 | 0.873 | 1.180 | 8.47e-01 | 1.00e+00 | 13672 | 7158 | 6514 |
| European | Recorded Anxiety | Switching vs Continuation | Stress | StressFairly Often | 1.052 | 0.911 | 1.215 | 4.91e-01 | 1.00e+00 | 13672 | 7158 | 6514 |
| European | Recorded Anxiety | Switching vs Continuation | Stress | StressSometimes | 1.277 | 1.183 | 1.379 | 3.75e-10 | 5.25e-09 | 13672 | 7158 | 6514 |
| European | Recorded Anxiety | Switching vs Continuation | Stress | StressVery Often | 1.184 | 0.978 | 1.433 | 8.41e-02 | 1.00e+00 | 13672 | 7158 | 6514 |
| European | Recorded Anxiety | Discontinuation vs Continuation | age_at_first_AD | (Intercept) | 0.273 | 0.260 | 0.285 | <2.2e-16 | <2.2e-16 | 15204 | 11798 | 3406 |
| European | Recorded Anxiety | Discontinuation vs Continuation | age_at_first_AD | age_at_first_AD | 1.090 | 1.050 | 1.132 | 7.79e-06 | 1.09e-04 | 15204 | 11798 | 3406 |
| European | Recorded Anxiety | Discontinuation vs Continuation | age_at_first_AD | sex_at_birthMale | 1.206 | 1.110 | 1.310 | 1.04e-05 | 1.46e-04 | 15204 | 11798 | 3406 |
| European | Recorded Anxiety | Switching vs Continuation | age_at_first_AD | (Intercept) | 0.853 | 0.827 | 0.881 | <2.2e-16 | <2.2e-16 | 22696 | 11798 | 10898 |
| European | Recorded Anxiety | Switching vs Continuation | age_at_first_AD | age_at_first_AD | 0.764 | 0.743 | 0.785 | <2.2e-16 | <2.2e-16 | 22696 | 11798 | 10898 |
| European | Recorded Anxiety | Switching vs Continuation | age_at_first_AD | sex_at_birthMale | 1.135 | 1.070 | 1.205 | 2.66e-05 | 3.72e-04 | 22696 | 11798 | 10898 |
| European | Recorded Anxiety | Discontinuation vs Continuation | sex_at_birth | (Intercept) | 0.273 | 0.260 | 0.285 | <2.2e-16 | <2.2e-16 | 15204 | 11798 | 3406 |
| European | Recorded Anxiety | Discontinuation vs Continuation | sex_at_birth | age_at_first_AD | 1.090 | 1.050 | 1.132 | 7.79e-06 | 1.09e-04 | 15204 | 11798 | 3406 |
| European | Recorded Anxiety | Discontinuation vs Continuation | sex_at_birth | sex_at_birthMale | 1.206 | 1.110 | 1.310 | 1.04e-05 | 1.46e-04 | 15204 | 11798 | 3406 |
| European | Recorded Anxiety | Switching vs Continuation | sex_at_birth | (Intercept) | 0.853 | 0.827 | 0.881 | <2.2e-16 | <2.2e-16 | 22696 | 11798 | 10898 |
| European | Recorded Anxiety | Switching vs Continuation | sex_at_birth | age_at_first_AD | 0.764 | 0.743 | 0.785 | <2.2e-16 | <2.2e-16 | 22696 | 11798 | 10898 |
| European | Recorded Anxiety | Switching vs Continuation | sex_at_birth | sex_at_birthMale | 1.135 | 1.070 | 1.205 | 2.66e-05 | 3.72e-04 | 22696 | 11798 | 10898 |
| European | CIDI-MDD | Discontinuation vs Continuation | Alcohol_Frequency | (Intercept) | 0.546 | 0.176 | 1.695 | 2.95e-01 | 1.00e+00 | 3085 | 2344 | 741 |
| European | CIDI-MDD | Discontinuation vs Continuation | Alcohol_Frequency | age_at_first_AD | 1.016 | 0.929 | 1.110 | 7.31e-01 | 1.00e+00 | 3085 | 2344 | 741 |
| European | CIDI-MDD | Discontinuation vs Continuation | Alcohol_Frequency | sex_at_birthMale | 1.048 | 0.862 | 1.274 | 6.38e-01 | 1.00e+00 | 3085 | 2344 | 741 |
| European | CIDI-MDD | Discontinuation vs Continuation | Alcohol_Frequency | Income>200k | 0.668 | 0.462 | 0.965 | 3.14e-02 | 4.40e-01 | 3085 | 2344 | 741 |
| European | CIDI-MDD | Discontinuation vs Continuation | Alcohol_Frequency | Income100-150k | 0.621 | 0.456 | 0.846 | 2.53e-03 | 3.54e-02 | 3085 | 2344 | 741 |
| European | CIDI-MDD | Discontinuation vs Continuation | Alcohol_Frequency | Income150-200k | 0.675 | 0.467 | 0.976 | 3.66e-02 | 5.13e-01 | 3085 | 2344 | 741 |
| European | CIDI-MDD | Discontinuation vs Continuation | Alcohol_Frequency | Income25-35k | 0.613 | 0.423 | 0.889 | 9.78e-03 | 1.37e-01 | 3085 | 2344 | 741 |
| European | CIDI-MDD | Discontinuation vs Continuation | Alcohol_Frequency | Income35-50k | 0.628 | 0.446 | 0.885 | 7.85e-03 | 1.10e-01 | 3085 | 2344 | 741 |
| European | CIDI-MDD | Discontinuation vs Continuation | Alcohol_Frequency | Income50-75k | 0.772 | 0.575 | 1.037 | 8.57e-02 | 1.00e+00 | 3085 | 2344 | 741 |
| European | CIDI-MDD | Discontinuation vs Continuation | Alcohol_Frequency | Income75-100k | 0.657 | 0.478 | 0.903 | 9.72e-03 | 1.36e-01 | 3085 | 2344 | 741 |
| European | CIDI-MDD | Discontinuation vs Continuation | Alcohol_Frequency | Education_levelCollege Postgraduate | 1.051 | 0.370 | 2.987 | 9.25e-01 | 1.00e+00 | 3085 | 2344 | 741 |
| European | CIDI-MDD | Discontinuation vs Continuation | Alcohol_Frequency | Education_levelCollege Undergraduate | 1.289 | 0.459 | 3.618 | 6.30e-01 | 1.00e+00 | 3085 | 2344 | 741 |
| European | CIDI-MDD | Discontinuation vs Continuation | Alcohol_Frequency | Education_levelTwelve or GED | 1.106 | 0.377 | 3.247 | 8.54e-01 | 1.00e+00 | 3085 | 2344 | 741 |

| | | | | | | | | | | | | |
|---|---|---|---|---|---|---|---|---|---|---|---|---|
| European | CIDI-MDD | Discontinuation vs Continuation | Alcohol_Frequency | Alcohol_Frequency2-3 Week | 0.636 | 0.366 | 1.104 | 1.08e-01 | 1.00e+00 | 3085 | 2344 | 741 |
| European | CIDI-MDD | Discontinuation vs Continuation | Alcohol_Frequency | Alcohol_Frequency2-4 Month | 0.637 | 0.372 | 1.090 | 1.00e-01 | 1.00e+00 | 3085 | 2344 | 741 |
| European | CIDI-MDD | Discontinuation vs Continuation | Alcohol_Frequency | Alcohol_Frequency4plus Week | 0.573 | 0.326 | 1.006 | 5.26e-02 | 7.37e-01 | 3085 | 2344 | 741 |
| European | CIDI-MDD | Discontinuation vs Continuation | Alcohol_Frequency | Alcohol_FrequencyMonthly or Less | 0.774 | 0.458 | 1.310 | 3.40e-01 | 1.00e+00 | 3085 | 2344 | 741 |
| European | CIDI-MDD | Discontinuation vs Continuation | Alcohol_Frequency | Alcohol_FrequencyNever (former) | 0.672 | 0.389 | 1.161 | 1.54e-01 | 1.00e+00 | 3085 | 2344 | 741 |
| European | CIDI-MDD | Switching vs Continuation | Alcohol_Frequency | (Intercept) | 0.533 | 0.224 | 1.269 | 1.55e-01 | 1.00e+00 | 4488 | 2344 | 2144 |
| European | CIDI-MDD | Switching vs Continuation | Alcohol_Frequency | age_at_first_AD | 0.670 | 0.627 | 0.717 | <2.2e-16 | <2.2e-16 | 4488 | 2344 | 2144 |
| European | CIDI-MDD | Switching vs Continuation | Alcohol_Frequency | sex_at_birthMale | 1.103 | 0.957 | 1.272 | 1.75e-01 | 1.00e+00 | 4488 | 2344 | 2144 |
| European | CIDI-MDD | Switching vs Continuation | Alcohol_Frequency | Income>200k | 0.456 | 0.342 | 0.607 | 7.72e-08 | 1.08e-06 | 4488 | 2344 | 2144 |
| European | CIDI-MDD | Switching vs Continuation | Alcohol_Frequency | Income100-150k | 0.640 | 0.510 | 0.802 | 1.09e-04 | 1.52e-03 | 4488 | 2344 | 2144 |
| European | CIDI-MDD | Switching vs Continuation | Alcohol_Frequency | Income150-200k | 0.461 | 0.346 | 0.613 | 1.07e-07 | 1.50e-06 | 4488 | 2344 | 2144 |
| European | CIDI-MDD | Switching vs Continuation | Alcohol_Frequency | Income25-35k | 0.751 | 0.577 | 0.978 | 3.36e-02 | 4.70e-01 | 4488 | 2344 | 2144 |
| European | CIDI-MDD | Switching vs Continuation | Alcohol_Frequency | Income35-50k | 0.874 | 0.688 | 1.110 | 2.69e-01 | 1.00e+00 | 4488 | 2344 | 2144 |
| European | CIDI-MDD | Switching vs Continuation | Alcohol_Frequency | Income50-75k | 0.783 | 0.629 | 0.975 | 2.86e-02 | 4.00e-01 | 4488 | 2344 | 2144 |
| European | CIDI-MDD | Switching vs Continuation | Alcohol_Frequency | Income75-100k | 0.721 | 0.573 | 0.909 | 5.59e-03 | 7.83e-02 | 4488 | 2344 | 2144 |
| European | CIDI-MDD | Switching vs Continuation | Alcohol_Frequency | Education_levelCollege Postgraduate | 1.726 | 0.782 | 3.810 | 1.77e-01 | 1.00e+00 | 4488 | 2344 | 2144 |
| European | CIDI-MDD | Switching vs Continuation | Alcohol_Frequency | Education_levelCollege Undergraduate | 1.924 | 0.879 | 4.214 | 1.02e-01 | 1.00e+00 | 4488 | 2344 | 2144 |
| European | CIDI-MDD | Switching vs Continuation | Alcohol_Frequency | Education_levelTwelve or GED | 2.223 | 0.989 | 4.996 | 5.31e-02 | 7.44e-01 | 4488 | 2344 | 2144 |
| European | CIDI-MDD | Switching vs Continuation | Alcohol_Frequency | Alcohol_Frequency2-3 Week | 0.988 | 0.628 | 1.552 | 9.57e-01 | 1.00e+00 | 4488 | 2344 | 2144 |
| European | CIDI-MDD | Switching vs Continuation | Alcohol_Frequency | Alcohol_Frequency2-4 Month | 1.093 | 0.702 | 1.700 | 6.94e-01 | 1.00e+00 | 4488 | 2344 | 2144 |
| European | CIDI-MDD | Switching vs Continuation | Alcohol_Frequency | Alcohol_Frequency4plus Week | 1.179 | 0.747 | 1.861 | 4.79e-01 | 1.00e+00 | 4488 | 2344 | 2144 |
| European | CIDI-MDD | Switching vs Continuation | Alcohol_Frequency | Alcohol_FrequencyMonthly or Less | 1.311 | 0.848 | 2.026 | 2.23e-01 | 1.00e+00 | 4488 | 2344 | 2144 |
| European | CIDI-MDD | Switching vs Continuation | Alcohol_Frequency | Alcohol_FrequencyNever (former) | 1.374 | 0.879 | 2.149 | 1.64e-01 | 1.00e+00 | 4488 | 2344 | 2144 |
| European | CIDI-MDD | Discontinuation vs Continuation | BMI | (Intercept) | 0.310 | 0.102 | 0.941 | 3.87e-02 | 5.41e-01 | 3059 | 2329 | 730 |
| European | CIDI-MDD | Discontinuation vs Continuation | BMI | age_at_first_AD | 1.002 | 0.917 | 1.093 | 9.72e-01 | 1.00e+00 | 3059 | 2329 | 730 |
| European | CIDI-MDD | Discontinuation vs Continuation | BMI | sex_at_birthMale | 1.039 | 0.855 | 1.263 | 6.98e-01 | 1.00e+00 | 3059 | 2329 | 730 |
| European | CIDI-MDD | Discontinuation vs Continuation | BMI | Income>200k | 0.591 | 0.409 | 0.853 | 5.00e-03 | 7.00e-02 | 3059 | 2329 | 730 |
| European | CIDI-MDD | Discontinuation vs Continuation | BMI | Income100-150k | 0.564 | 0.414 | 0.767 | 2.66e-04 | 3.73e-03 | 3059 | 2329 | 730 |
| European | CIDI-MDD | Discontinuation vs Continuation | BMI | Income150-200k | 0.599 | 0.414 | 0.868 | 6.76e-03 | 9.46e-02 | 3059 | 2329 | 730 |
| European | CIDI-MDD | Discontinuation vs Continuation | BMI | Income25-35k | 0.569 | 0.391 | 0.827 | 3.12e-03 | 4.36e-02 | 3059 | 2329 | 730 |
| European | CIDI-MDD | Discontinuation vs Continuation | BMI | Income35-50k | 0.604 | 0.429 | 0.850 | 3.88e-03 | 5.44e-02 | 3059 | 2329 | 730 |
| European | CIDI-MDD | Discontinuation vs Continuation | BMI | Income50-75k | 0.755 | 0.563 | 1.012 | 6.04e-02 | 8.46e-01 | 3059 | 2329 | 730 |
| European | CIDI-MDD | Discontinuation vs Continuation | BMI | Income75-100k | 0.609 | 0.444 | 0.836 | 2.15e-03 | 3.00e-02 | 3059 | 2329 | 730 |
| European | CIDI-MDD | Discontinuation vs Continuation | BMI | Education_levelCollege Postgraduate | 1.285 | 0.417 | 3.958 | 6.62e-01 | 1.00e+00 | 3059 | 2329 | 730 |
| European | CIDI-MDD | Discontinuation vs Continuation | BMI | Education_levelCollege Undergraduate | 1.656 | 0.544 | 5.045 | 3.75e-01 | 1.00e+00 | 3059 | 2329 | 730 |
| European | CIDI-MDD | Discontinuation vs Continuation | BMI | Education_levelTwelve or GED | 1.396 | 0.439 | 4.439 | 5.71e-01 | 1.00e+00 | 3059 | 2329 | 730 |
| European | CIDI-MDD | Discontinuation vs Continuation | BMI | BMI | 0.934 | 0.856 | 1.019 | 1.26e-01 | 1.00e+00 | 3059 | 2329 | 730 |
| European | CIDI-MDD | Switching vs Continuation | BMI | (Intercept) | 0.571 | 0.259 | 1.260 | 1.65e-01 | 1.00e+00 | 4460 | 2329 | 2131 |
| European | CIDI-MDD | Switching vs Continuation | BMI | age_at_first_AD | 0.680 | 0.636 | 0.726 | <2.2e-16 | <2.2e-16 | 4460 | 2329 | 2131 |
| European | CIDI-MDD | Switching vs Continuation | BMI | sex_at_birthMale | 1.098 | 0.953 | 1.265 | 1.97e-01 | 1.00e+00 | 4460 | 2329 | 2131 |
| European | CIDI-MDD | Switching vs Continuation | BMI | Income>200k | 0.457 | 0.344 | 0.608 | 7.27e-08 | 1.02e-06 | 4460 | 2329 | 2131 |
| European | CIDI-MDD | Switching vs Continuation | BMI | Income100-150k | 0.620 | 0.495 | 0.776 | 3.16e-05 | 4.42e-04 | 4460 | 2329 | 2131 |
| European | CIDI-MDD | Switching vs Continuation | BMI | Income150-200k | 0.461 | 0.347 | 0.614 | 1.07e-07 | 1.50e-06 | 4460 | 2329 | 2131 |
| European | CIDI-MDD | Switching vs Continuation | BMI | Income25-35k | 0.749 | 0.575 | 0.975 | 3.17e-02 | 4.44e-01 | 4460 | 2329 | 2131 |
| European | CIDI-MDD | Switching vs Continuation | BMI | Income35-50k | 0.843 | 0.664 | 1.072 | 1.64e-01 | 1.00e+00 | 4460 | 2329 | 2131 |
| European | CIDI-MDD | Switching vs Continuation | BMI | Income50-75k | 0.773 | 0.621 | 0.963 | 2.16e-02 | 3.02e-01 | 4460 | 2329 | 2131 |
| European | CIDI-MDD | Switching vs Continuation | BMI | Income75-100k | 0.696 | 0.553 | 0.877 | 2.08e-03 | 2.91e-02 | 4460 | 2329 | 2131 |
| European | CIDI-MDD | Switching vs Continuation | BMI | Education_levelCollege Postgraduate | 1.965 | 0.882 | 4.381 | 9.86e-02 | 1.00e+00 | 4460 | 2329 | 2131 |
| European | CIDI-MDD | Switching vs Continuation | BMI | Education_levelCollege Undergraduate | 2.186 | 0.988 | 4.836 | 5.34e-02 | 7.48e-01 | 4460 | 2329 | 2131 |
| European | CIDI-MDD | Switching vs Continuation | BMI | Education_levelTwelve or GED | 2.531 | 1.115 | 5.746 | 2.65e-02 | 3.71e-01 | 4460 | 2329 | 2131 |
| European | CIDI-MDD | Switching vs Continuation | BMI | BMI | 1.130 | 1.064 | 1.200 | 7.38e-05 | 1.03e-03 | 4460 | 2329 | 2131 |
| European | CIDI-MDD | Discontinuation vs Continuation | Education_level | (Intercept) | 0.376 | 0.136 | 1.039 | 5.92e-02 | 8.28e-01 | 3093 | 2349 | 744 |
| European | CIDI-MDD | Discontinuation vs Continuation | Education_level | age_at_first_AD | 1.000 | 0.917 | 1.092 | 9.93e-01 | 1.00e+00 | 3093 | 2349 | 744 |
| European | CIDI-MDD | Discontinuation vs Continuation | Education_level | sex_at_birthMale | 1.033 | 0.852 | 1.253 | 7.43e-01 | 1.00e+00 | 3093 | 2349 | 744 |
| European | CIDI-MDD | Discontinuation vs Continuation | Education_level | Income>200k | 0.613 | 0.427 | 0.880 | 8.02e-03 | 1.12e-01 | 3093 | 2349 | 744 |
| European | CIDI-MDD | Discontinuation vs Continuation | Education_level | Income100-150k | 0.591 | 0.436 | 0.802 | 7.10e-04 | 9.94e-03 | 3093 | 2349 | 744 |
| European | CIDI-MDD | Discontinuation vs Continuation | Education_level | Income150-200k | 0.636 | 0.442 | 0.915 | 1.47e-02 | 2.06e-01 | 3093 | 2349 | 744 |
| European | CIDI-MDD | Discontinuation vs Continuation | Education_level | Income25-35k | 0.593 | 0.410 | 0.857 | 5.42e-03 | 7.58e-02 | 3093 | 2349 | 744 |
| European | CIDI-MDD | Discontinuation vs Continuation | Education_level | Income35-50k | 0.610 | 0.435 | 0.858 | 4.43e-03 | 6.20e-02 | 3093 | 2349 | 744 |
| European | CIDI-MDD | Discontinuation vs Continuation | Education_level | Income50-75k | 0.751 | 0.561 | 1.005 | 5.42e-02 | 7.58e-01 | 3093 | 2349 | 744 |
| European | CIDI-MDD | Discontinuation vs Continuation | Education_level | Income75-100k | 0.624 | 0.456 | 0.854 | 3.21e-03 | 4.49e-02 | 3093 | 2349 | 744 |
| European | CIDI-MDD | Discontinuation vs Continuation | Education_level | Education_levelCollege Postgraduate | 1.074 | 0.382 | 3.022 | 8.92e-01 | 1.00e+00 | 3093 | 2349 | 744 |
| European | CIDI-MDD | Discontinuation vs Continuation | Education_level | Education_levelCollege Undergraduate | 1.339 | 0.482 | 3.724 | 5.76e-01 | 1.00e+00 | 3093 | 2349 | 744 |
| European | CIDI-MDD | Discontinuation vs Continuation | Education_level | Education_levelTwelve or GED | 1.187 | 0.409 | 3.451 | 7.52e-01 | 1.00e+00 | 3093 | 2349 | 744 |
| European | CIDI-MDD | Switching vs Continuation | Education_level | (Intercept) | 0.632 | 0.293 | 1.366 | 2.43e-01 | 1.00e+00 | 4499 | 2349 | 2150 |
| European | CIDI-MDD | Switching vs Continuation | Education_level | age_at_first_AD | 0.677 | 0.634 | 0.724 | <2.2e-16 | <2.2e-16 | 4499 | 2349 | 2150 |

| | | | | | | | | | | | | |
|---|---|---|---|---|---|---|---|---|---|---|---|---|
| European | CIDI-MDD | Switching vs Continuation | Education_level | sex_at_birthMale | 1.104 | 0.959 | 1.271 | 1.69e-01 | 1.00e+00 | 4499 | 2349 | 2150 |
| European | CIDI-MDD | Switching vs Continuation | Education_level | Income>200k | 0.424 | 0.320 | 0.562 | 2.46e-09 | 3.45e-08 | 4499 | 2349 | 2150 |
| European | CIDI-MDD | Switching vs Continuation | Education_level | Income100-150k | 0.613 | 0.490 | 0.767 | 1.82e-05 | 2.55e-04 | 4499 | 2349 | 2150 |
| European | CIDI-MDD | Switching vs Continuation | Education_level | Income150-200k | 0.439 | 0.330 | 0.583 | 1.22e-08 | 1.71e-07 | 4499 | 2349 | 2150 |
| European | CIDI-MDD | Switching vs Continuation | Education_level | Income25-35k | 0.739 | 0.568 | 0.960 | 2.37e-02 | 3.32e-01 | 4499 | 2349 | 2150 |
| European | CIDI-MDD | Switching vs Continuation | Education_level | Income35-50k | 0.852 | 0.671 | 1.081 | 1.86e-01 | 1.00e+00 | 4499 | 2349 | 2150 |
| European | CIDI-MDD | Switching vs Continuation | Education_level | Income50-75k | 0.765 | 0.615 | 0.951 | 1.59e-02 | 2.22e-01 | 4499 | 2349 | 2150 |
| European | CIDI-MDD | Switching vs Continuation | Education_level | Income75-100k | 0.690 | 0.549 | 0.868 | 1.51e-03 | 2.11e-02 | 4499 | 2349 | 2150 |
| European | CIDI-MDD | Switching vs Continuation | Education_level | Education_levelCollege Postgraduate | 1.778 | 0.815 | 3.882 | 1.48e-01 | 1.00e+00 | 4499 | 2349 | 2150 |
| European | CIDI-MDD | Switching vs Continuation | Education_level | Education_levelCollege Undergraduate | 2.019 | 0.932 | 4.373 | 7.49e-02 | 1.00e+00 | 4499 | 2349 | 2150 |
| European | CIDI-MDD | Switching vs Continuation | Education_level | Education_levelTwelve or GED | 2.362 | 1.062 | 5.253 | 3.51e-02 | 4.91e-01 | 4499 | 2349 | 2150 |
| European | CIDI-MDD | Discontinuation vs Continuation | Income | (Intercept) | 0.376 | 0.136 | 1.039 | 5.92e-02 | 8.28e-01 | 3093 | 2349 | 744 |
| European | CIDI-MDD | Discontinuation vs Continuation | Income | age_at_first_AD | 1.000 | 0.917 | 1.092 | 9.93e-01 | 1.00e+00 | 3093 | 2349 | 744 |
| European | CIDI-MDD | Discontinuation vs Continuation | Income | sex_at_birthMale | 1.033 | 0.852 | 1.253 | 7.43e-01 | 1.00e+00 | 3093 | 2349 | 744 |
| European | CIDI-MDD | Discontinuation vs Continuation | Income | Income>200k | 0.613 | 0.427 | 0.880 | 8.02e-03 | 1.12e-01 | 3093 | 2349 | 744 |
| European | CIDI-MDD | Discontinuation vs Continuation | Income | Income100-150k | 0.591 | 0.436 | 0.802 | 7.10e-04 | 9.94e-03 | 3093 | 2349 | 744 |
| European | CIDI-MDD | Discontinuation vs Continuation | Income | Income150-200k | 0.636 | 0.442 | 0.915 | 1.47e-02 | 2.06e-01 | 3093 | 2349 | 744 |
| European | CIDI-MDD | Discontinuation vs Continuation | Income | Income25-35k | 0.593 | 0.410 | 0.857 | 5.42e-03 | 7.58e-02 | 3093 | 2349 | 744 |
| European | CIDI-MDD | Discontinuation vs Continuation | Income | Income35-50k | 0.610 | 0.435 | 0.858 | 4.43e-03 | 6.20e-02 | 3093 | 2349 | 744 |
| European | CIDI-MDD | Discontinuation vs Continuation | Income | Income50-75k | 0.751 | 0.561 | 1.005 | 5.42e-02 | 7.58e-01 | 3093 | 2349 | 744 |
| European | CIDI-MDD | Discontinuation vs Continuation | Income | Income75-100k | 0.624 | 0.456 | 0.854 | 3.21e-03 | 4.49e-02 | 3093 | 2349 | 744 |
| European | CIDI-MDD | Discontinuation vs Continuation | Income | Education_levelCollege Postgraduate | 1.074 | 0.382 | 3.022 | 8.92e-01 | 1.00e+00 | 3093 | 2349 | 744 |
| European | CIDI-MDD | Discontinuation vs Continuation | Income | Education_levelCollege Undergraduate | 1.339 | 0.482 | 3.724 | 5.76e-01 | 1.00e+00 | 3093 | 2349 | 744 |
| European | CIDI-MDD | Discontinuation vs Continuation | Income | Education_levelTwelve or GED | 1.187 | 0.409 | 3.451 | 7.52e-01 | 1.00e+00 | 3093 | 2349 | 744 |
| European | CIDI-MDD | Switching vs Continuation | Income | (Intercept) | 0.632 | 0.293 | 1.366 | 2.43e-01 | 1.00e+00 | 4499 | 2349 | 2150 |
| European | CIDI-MDD | Switching vs Continuation | Income | age_at_first_AD | 0.677 | 0.634 | 0.724 | <2.2e-16 | <2.2e-16 | 4499 | 2349 | 2150 |
| European | CIDI-MDD | Switching vs Continuation | Income | sex_at_birthMale | 1.104 | 0.959 | 1.271 | 1.69e-01 | 1.00e+00 | 4499 | 2349 | 2150 |
| European | CIDI-MDD | Switching vs Continuation | Income | Income>200k | 0.424 | 0.320 | 0.562 | 2.46e-09 | 3.45e-08 | 4499 | 2349 | 2150 |
| European | CIDI-MDD | Switching vs Continuation | Income | Income100-150k | 0.613 | 0.490 | 0.767 | 1.82e-05 | 2.55e-04 | 4499 | 2349 | 2150 |
| European | CIDI-MDD | Switching vs Continuation | Income | Income150-200k | 0.439 | 0.330 | 0.583 | 1.22e-08 | 1.71e-07 | 4499 | 2349 | 2150 |
| European | CIDI-MDD | Switching vs Continuation | Income | Income25-35k | 0.739 | 0.568 | 0.960 | 2.37e-02 | 3.32e-01 | 4499 | 2349 | 2150 |
| European | CIDI-MDD | Switching vs Continuation | Income | Income35-50k | 0.852 | 0.671 | 1.081 | 1.86e-01 | 1.00e+00 | 4499 | 2349 | 2150 |
| European | CIDI-MDD | Switching vs Continuation | Income | Income50-75k | 0.765 | 0.615 | 0.951 | 1.59e-02 | 2.22e-01 | 4499 | 2349 | 2150 |
| European | CIDI-MDD | Switching vs Continuation | Income | Income75-100k | 0.690 | 0.549 | 0.868 | 1.51e-03 | 2.11e-02 | 4499 | 2349 | 2150 |
| European | CIDI-MDD | Switching vs Continuation | Income | Education_levelCollege Postgraduate | 1.778 | 0.815 | 3.882 | 1.48e-01 | 1.00e+00 | 4499 | 2349 | 2150 |
| European | CIDI-MDD | Switching vs Continuation | Income | Education_levelCollege Undergraduate | 2.019 | 0.932 | 4.373 | 7.49e-02 | 1.00e+00 | 4499 | 2349 | 2150 |
| European | CIDI-MDD | Switching vs Continuation | Income | Education_levelTwelve or GED | 2.362 | 1.062 | 5.253 | 3.51e-02 | 4.91e-01 | 4499 | 2349 | 2150 |
| European | CIDI-MDD | Discontinuation vs Continuation | Insomnia | (Intercept) | 0.384 | 0.139 | 1.064 | 6.56e-02 | 9.18e-01 | 3093 | 2349 | 744 |
| European | CIDI-MDD | Discontinuation vs Continuation | Insomnia | age_at_first_AD | 1.004 | 0.919 | 1.095 | 9.37e-01 | 1.00e+00 | 3093 | 2349 | 744 |
| European | CIDI-MDD | Discontinuation vs Continuation | Insomnia | sex_at_birthMale | 1.032 | 0.850 | 1.251 | 7.53e-01 | 1.00e+00 | 3093 | 2349 | 744 |
| European | CIDI-MDD | Discontinuation vs Continuation | Insomnia | Income>200k | 0.613 | 0.427 | 0.879 | 7.89e-03 | 1.10e-01 | 3093 | 2349 | 744 |
| European | CIDI-MDD | Discontinuation vs Continuation | Insomnia | Income100-150k | 0.592 | 0.437 | 0.802 | 7.25e-04 | 1.01e-02 | 3093 | 2349 | 744 |
| European | CIDI-MDD | Discontinuation vs Continuation | Insomnia | Income150-200k | 0.636 | 0.442 | 0.916 | 1.49e-02 | 2.08e-01 | 3093 | 2349 | 744 |
| European | CIDI-MDD | Discontinuation vs Continuation | Insomnia | Income25-35k | 0.591 | 0.409 | 0.855 | 5.26e-03 | 7.37e-02 | 3093 | 2349 | 744 |
| European | CIDI-MDD | Discontinuation vs Continuation | Insomnia | Income35-50k | 0.610 | 0.434 | 0.857 | 4.36e-03 | 6.11e-02 | 3093 | 2349 | 744 |
| European | CIDI-MDD | Discontinuation vs Continuation | Insomnia | Income50-75k | 0.750 | 0.560 | 1.004 | 5.36e-02 | 7.50e-01 | 3093 | 2349 | 744 |
| European | CIDI-MDD | Discontinuation vs Continuation | Insomnia | Income75-100k | 0.624 | 0.456 | 0.854 | 3.23e-03 | 4.52e-02 | 3093 | 2349 | 744 |
| European | CIDI-MDD | Discontinuation vs Continuation | Insomnia | Education_levelCollege Postgraduate | 1.065 | 0.378 | 3.000 | 9.04e-01 | 1.00e+00 | 3093 | 2349 | 744 |
| European | CIDI-MDD | Discontinuation vs Continuation | Insomnia | Education_levelCollege Undergraduate | 1.328 | 0.477 | 3.696 | 5.87e-01 | 1.00e+00 | 3093 | 2349 | 744 |
| European | CIDI-MDD | Discontinuation vs Continuation | Insomnia | Education_levelTwelve or GED | 1.177 | 0.405 | 3.423 | 7.65e-01 | 1.00e+00 | 3093 | 2349 | 744 |
| European | CIDI-MDD | Discontinuation vs Continuation | Insomnia | Insomnia | 0.930 | 0.747 | 1.158 | 5.17e-01 | 1.00e+00 | 3093 | 2349 | 744 |
| European | CIDI-MDD | Switching vs Continuation | Insomnia | (Intercept) | 0.471 | 0.214 | 1.036 | 6.13e-02 | 8.58e-01 | 4499 | 2349 | 2150 |
| European | CIDI-MDD | Switching vs Continuation | Insomnia | age_at_first_AD | 0.655 | 0.612 | 0.701 | <2.2e-16 | <2.2e-16 | 4499 | 2349 | 2150 |
| European | CIDI-MDD | Switching vs Continuation | Insomnia | sex_at_birthMale | 1.092 | 0.946 | 1.262 | 2.30e-01 | 1.00e+00 | 4499 | 2349 | 2150 |
| European | CIDI-MDD | Switching vs Continuation | Insomnia | Income>200k | 0.431 | 0.323 | 0.575 | 1.05e-08 | 1.47e-07 | 4499 | 2349 | 2150 |
| European | CIDI-MDD | Switching vs Continuation | Insomnia | Income100-150k | 0.605 | 0.481 | 0.760 | 1.58e-05 | 2.21e-04 | 4499 | 2349 | 2150 |
| European | CIDI-MDD | Switching vs Continuation | Insomnia | Income150-200k | 0.442 | 0.331 | 0.590 | 3.10e-08 | 4.35e-07 | 4499 | 2349 | 2150 |
| European | CIDI-MDD | Switching vs Continuation | Insomnia | Income25-35k | 0.736 | 0.563 | 0.964 | 2.58e-02 | 3.61e-01 | 4499 | 2349 | 2150 |
| European | CIDI-MDD | Switching vs Continuation | Insomnia | Income35-50k | 0.858 | 0.673 | 1.094 | 2.16e-01 | 1.00e+00 | 4499 | 2349 | 2150 |
| European | CIDI-MDD | Switching vs Continuation | Insomnia | Income50-75k | 0.768 | 0.615 | 0.960 | 2.04e-02 | 2.86e-01 | 4499 | 2349 | 2150 |
| European | CIDI-MDD | Switching vs Continuation | Insomnia | Income75-100k | 0.697 | 0.551 | 0.880 | 2.43e-03 | 3.40e-02 | 4499 | 2349 | 2150 |
| European | CIDI-MDD | Switching vs Continuation | Insomnia | Education_levelCollege Postgraduate | 1.837 | 0.828 | 4.076 | 1.35e-01 | 1.00e+00 | 4499 | 2349 | 2150 |
| European | CIDI-MDD | Switching vs Continuation | Insomnia | Education_levelCollege Undergraduate | 2.104 | 0.956 | 4.632 | 6.47e-02 | 9.06e-01 | 4499 | 2349 | 2150 |
| European | CIDI-MDD | Switching vs Continuation | Insomnia | Education_levelTwelve or GED | 2.435 | 1.076 | 5.510 | 3.26e-02 | 4.57e-01 | 4499 | 2349 | 2150 |
| European | CIDI-MDD | Switching vs Continuation | Insomnia | Insomnia | 2.623 | 2.277 | 3.022 | <2.2e-16 | <2.2e-16 | 4499 | 2349 | 2150 |
| European | CIDI-MDD | Discontinuation vs Continuation | MDD_symptom_count | (Intercept) | 0.324 | 0.115 | 0.911 | 3.26e-02 | 4.57e-01 | 3093 | 2349 | 744 |

| | | | | | | | | | | | | |
|---|---|---|---|---|---|---|---|---|---|---|---|---|
| European | CIDI-MDD | Discontinuation vs Continuation | MDD_symptom_count | age_at_first_AD | 1.014 | 0.928 | 1.108 | 7.63e-01 | 1.00e+00 | 3093 | 2349 | 744 |
| European | CIDI-MDD | Discontinuation vs Continuation | MDD_symptom_count | sex_at_birthMale | 1.036 | 0.854 | 1.256 | 7.22e-01 | 1.00e+00 | 3093 | 2349 | 744 |
| European | CIDI-MDD | Discontinuation vs Continuation | MDD_symptom_count | Income>200k | 0.626 | 0.436 | 0.900 | 1.14e-02 | 1.59e-01 | 3093 | 2349 | 744 |
| European | CIDI-MDD | Discontinuation vs Continuation | MDD_symptom_count | Income100-150k | 0.603 | 0.444 | 0.818 | 1.15e-03 | 1.62e-02 | 3093 | 2349 | 744 |
| European | CIDI-MDD | Discontinuation vs Continuation | MDD_symptom_count | Income150-200k | 0.643 | 0.447 | 0.925 | 1.75e-02 | 2.45e-01 | 3093 | 2349 | 744 |
| European | CIDI-MDD | Discontinuation vs Continuation | MDD_symptom_count | Income25-35k | 0.602 | 0.416 | 0.871 | 7.12e-03 | 9.97e-02 | 3093 | 2349 | 744 |
| European | CIDI-MDD | Discontinuation vs Continuation | MDD_symptom_count | Income35-50k | 0.622 | 0.442 | 0.874 | 6.25e-03 | 8.74e-02 | 3093 | 2349 | 744 |
| European | CIDI-MDD | Discontinuation vs Continuation | MDD_symptom_count | Income50-75k | 0.760 | 0.568 | 1.018 | 6.59e-02 | 9.23e-01 | 3093 | 2349 | 744 |
| European | CIDI-MDD | Discontinuation vs Continuation | MDD_symptom_count | Income75-100k | 0.635 | 0.463 | 0.870 | 4.68e-03 | 6.55e-02 | 3093 | 2349 | 744 |
| European | CIDI-MDD | Discontinuation vs Continuation | MDD_symptom_count | Education_levelCollege Postgraduate | 1.068 | 0.379 | 3.007 | 9.01e-01 | 1.00e+00 | 3093 | 2349 | 744 |
| European | CIDI-MDD | Discontinuation vs Continuation | MDD_symptom_count | Education_levelCollege Undergraduate | 1.327 | 0.477 | 3.694 | 5.88e-01 | 1.00e+00 | 3093 | 2349 | 744 |
| European | CIDI-MDD | Discontinuation vs Continuation | MDD_symptom_count | Education_levelTwelve or GED | 1.187 | 0.408 | 3.452 | 7.53e-01 | 1.00e+00 | 3093 | 2349 | 744 |
| European | CIDI-MDD | Discontinuation vs Continuation | MDD_symptom_count | MDD_symptom_count | 1.214 | 0.960 | 1.535 | 1.05e-01 | 1.00e+00 | 3093 | 2349 | 744 |
| European | CIDI-MDD | Switching vs Continuation | MDD_symptom_count | (Intercept) | 0.505 | 0.231 | 1.106 | 8.78e-02 | 1.00e+00 | 4499 | 2349 | 2150 |
| European | CIDI-MDD | Switching vs Continuation | MDD_symptom_count | age_at_first_AD | 0.693 | 0.648 | 0.742 | <2.2e-16 | <2.2e-16 | 4499 | 2349 | 2150 |
| European | CIDI-MDD | Switching vs Continuation | MDD_symptom_count | sex_at_birthMale | 1.100 | 0.955 | 1.266 | 1.88e-01 | 1.00e+00 | 4499 | 2349 | 2150 |
| European | CIDI-MDD | Switching vs Continuation | MDD_symptom_count | Income>200k | 0.441 | 0.332 | 0.584 | 1.33e-08 | 1.87e-07 | 4499 | 2349 | 2150 |
| European | CIDI-MDD | Switching vs Continuation | MDD_symptom_count | Income100-150k | 0.634 | 0.506 | 0.793 | 6.76e-05 | 9.47e-04 | 4499 | 2349 | 2150 |
| European | CIDI-MDD | Switching vs Continuation | MDD_symptom_count | Income150-200k | 0.449 | 0.338 | 0.597 | 3.42e-08 | 4.79e-07 | 4499 | 2349 | 2150 |
| European | CIDI-MDD | Switching vs Continuation | MDD_symptom_count | Income25-35k | 0.754 | 0.579 | 0.980 | 3.50e-02 | 4.90e-01 | 4499 | 2349 | 2150 |
| European | CIDI-MDD | Switching vs Continuation | MDD_symptom_count | Income35-50k | 0.872 | 0.687 | 1.106 | 2.59e-01 | 1.00e+00 | 4499 | 2349 | 2150 |
| European | CIDI-MDD | Switching vs Continuation | MDD_symptom_count | Income50-75k | 0.780 | 0.627 | 0.970 | 2.56e-02 | 3.59e-01 | 4499 | 2349 | 2150 |
| European | CIDI-MDD | Switching vs Continuation | MDD_symptom_count | Income75-100k | 0.708 | 0.563 | 0.891 | 3.20e-03 | 4.48e-02 | 4499 | 2349 | 2150 |
| European | CIDI-MDD | Switching vs Continuation | MDD_symptom_count | Education_levelCollege Postgraduate | 1.719 | 0.784 | 3.769 | 1.76e-01 | 1.00e+00 | 4499 | 2349 | 2150 |
| European | CIDI-MDD | Switching vs Continuation | MDD_symptom_count | Education_levelCollege Undergraduate | 1.952 | 0.897 | 4.246 | 9.18e-02 | 1.00e+00 | 4499 | 2349 | 2150 |
| European | CIDI-MDD | Switching vs Continuation | MDD_symptom_count | Education_levelTwelve or GED | 2.286 | 1.023 | 5.105 | 4.37e-02 | 6.12e-01 | 4499 | 2349 | 2150 |
| European | CIDI-MDD | Switching vs Continuation | MDD_symptom_count | MDD_symptom_count | 1.383 | 1.163 | 1.644 | 2.41e-04 | 3.38e-03 | 4499 | 2349 | 2150 |
| European | CIDI-MDD | Discontinuation vs Continuation | Migraine | (Intercept) | 0.413 | 0.149 | 1.146 | 8.96e-02 | 1.00e+00 | 3093 | 2349 | 744 |
| European | CIDI-MDD | Discontinuation vs Continuation | Migraine | age_at_first_AD | 0.982 | 0.899 | 1.072 | 6.81e-01 | 1.00e+00 | 3093 | 2349 | 744 |
| European | CIDI-MDD | Discontinuation vs Continuation | Migraine | sex_at_birthMale | 0.989 | 0.815 | 1.202 | 9.13e-01 | 1.00e+00 | 3093 | 2349 | 744 |
| European | CIDI-MDD | Discontinuation vs Continuation | Migraine | Income>200k | 0.616 | 0.429 | 0.886 | 8.86e-03 | 1.24e-01 | 3093 | 2349 | 744 |
| European | CIDI-MDD | Discontinuation vs Continuation | Migraine | Income100-150k | 0.609 | 0.449 | 0.826 | 1.44e-03 | 2.01e-02 | 3093 | 2349 | 744 |
| European | CIDI-MDD | Discontinuation vs Continuation | Migraine | Income150-200k | 0.654 | 0.454 | 0.942 | 2.27e-02 | 3.17e-01 | 3093 | 2349 | 744 |
| European | CIDI-MDD | Discontinuation vs Continuation | Migraine | Income25-35k | 0.598 | 0.413 | 0.865 | 6.41e-03 | 8.98e-02 | 3093 | 2349 | 744 |
| European | CIDI-MDD | Discontinuation vs Continuation | Migraine | Income35-50k | 0.620 | 0.441 | 0.872 | 6.02e-03 | 8.43e-02 | 3093 | 2349 | 744 |
| European | CIDI-MDD | Discontinuation vs Continuation | Migraine | Income50-75k | 0.767 | 0.573 | 1.028 | 7.63e-02 | 1.00e+00 | 3093 | 2349 | 744 |
| European | CIDI-MDD | Discontinuation vs Continuation | Migraine | Income75-100k | 0.637 | 0.464 | 0.872 | 4.95e-03 | 6.93e-02 | 3093 | 2349 | 744 |
| European | CIDI-MDD | Discontinuation vs Continuation | Migraine | Education_levelCollege Postgraduate | 1.087 | 0.385 | 3.066 | 8.75e-01 | 1.00e+00 | 3093 | 2349 | 744 |
| European | CIDI-MDD | Discontinuation vs Continuation | Migraine | Education_levelCollege Undergraduate | 1.359 | 0.488 | 3.789 | 5.58e-01 | 1.00e+00 | 3093 | 2349 | 744 |
| European | CIDI-MDD | Discontinuation vs Continuation | Migraine | Education_levelTwelve or GED | 1.197 | 0.411 | 3.489 | 7.41e-01 | 1.00e+00 | 3093 | 2349 | 744 |
| European | CIDI-MDD | Discontinuation vs Continuation | Migraine | Migraine | 0.639 | 0.524 | 0.780 | 1.05e-05 | 1.47e-04 | 3093 | 2349 | 744 |
| European | CIDI-MDD | Switching vs Continuation | Migraine | (Intercept) | 0.558 | 0.257 | 1.213 | 1.41e-01 | 1.00e+00 | 4499 | 2349 | 2150 |
| European | CIDI-MDD | Switching vs Continuation | Migraine | age_at_first_AD | 0.697 | 0.652 | 0.745 | <2.2e-16 | <2.2e-16 | 4499 | 2349 | 2150 |
| European | CIDI-MDD | Switching vs Continuation | Migraine | sex_at_birthMale | 1.179 | 1.022 | 1.360 | 2.39e-02 | 3.34e-01 | 4499 | 2349 | 2150 |
| European | CIDI-MDD | Switching vs Continuation | Migraine | Income>200k | 0.429 | 0.323 | 0.570 | 5.15e-09 | 7.21e-08 | 4499 | 2349 | 2150 |
| European | CIDI-MDD | Switching vs Continuation | Migraine | Income100-150k | 0.612 | 0.488 | 0.766 | 1.88e-05 | 2.63e-04 | 4499 | 2349 | 2150 |
| European | CIDI-MDD | Switching vs Continuation | Migraine | Income150-200k | 0.444 | 0.334 | 0.591 | 2.43e-08 | 3.40e-07 | 4499 | 2349 | 2150 |
| European | CIDI-MDD | Switching vs Continuation | Migraine | Income25-35k | 0.744 | 0.571 | 0.969 | 2.83e-02 | 3.96e-01 | 4499 | 2349 | 2150 |
| European | CIDI-MDD | Switching vs Continuation | Migraine | Income35-50k | 0.855 | 0.673 | 1.087 | 2.01e-01 | 1.00e+00 | 4499 | 2349 | 2150 |
| European | CIDI-MDD | Switching vs Continuation | Migraine | Income50-75k | 0.761 | 0.611 | 0.948 | 1.49e-02 | 2.08e-01 | 4499 | 2349 | 2150 |
| European | CIDI-MDD | Switching vs Continuation | Migraine | Income75-100k | 0.691 | 0.549 | 0.871 | 1.70e-03 | 2.39e-02 | 4499 | 2349 | 2150 |
| European | CIDI-MDD | Switching vs Continuation | Migraine | Education_levelCollege Postgraduate | 1.653 | 0.753 | 3.630 | 2.10e-01 | 1.00e+00 | 4499 | 2349 | 2150 |
| European | CIDI-MDD | Switching vs Continuation | Migraine | Education_levelCollege Undergraduate | 1.877 | 0.862 | 4.090 | 1.13e-01 | 1.00e+00 | 4499 | 2349 | 2150 |
| European | CIDI-MDD | Switching vs Continuation | Migraine | Education_levelTwelve or GED | 2.200 | 0.984 | 4.922 | 5.49e-02 | 7.68e-01 | 4499 | 2349 | 2150 |
| European | CIDI-MDD | Switching vs Continuation | Migraine | Migraine | 1.651 | 1.453 | 1.875 | 1.28e-14 | 1.79e-13 | 4499 | 2349 | 2150 |
| European | CIDI-MDD | Discontinuation vs Continuation | Recorded Anxiety | (Intercept) | 0.471 | 0.169 | 1.312 | 1.50e-01 | 1.00e+00 | 3093 | 2349 | 744 |
| European | CIDI-MDD | Discontinuation vs Continuation | Recorded Anxiety | age_at_first_AD | 0.973 | 0.891 | 1.063 | 5.41e-01 | 1.00e+00 | 3093 | 2349 | 744 |
| European | CIDI-MDD | Discontinuation vs Continuation | Recorded Anxiety | sex_at_birthMale | 1.019 | 0.839 | 1.237 | 8.52e-01 | 1.00e+00 | 3093 | 2349 | 744 |
| European | CIDI-MDD | Discontinuation vs Continuation | Recorded Anxiety | Income>200k | 0.590 | 0.410 | 0.848 | 4.35e-03 | 6.09e-02 | 3093 | 2349 | 744 |
| European | CIDI-MDD | Discontinuation vs Continuation | Recorded Anxiety | Income100-150k | 0.590 | 0.435 | 0.801 | 7.16e-04 | 1.00e-02 | 3093 | 2349 | 744 |
| European | CIDI-MDD | Discontinuation vs Continuation | Recorded Anxiety | Income150-200k | 0.635 | 0.441 | 0.915 | 1.49e-02 | 2.08e-01 | 3093 | 2349 | 744 |
| European | CIDI-MDD | Discontinuation vs Continuation | Recorded Anxiety | Income25-35k | 0.597 | 0.412 | 0.864 | 6.23e-03 | 8.72e-02 | 3093 | 2349 | 744 |
| European | CIDI-MDD | Discontinuation vs Continuation | Recorded Anxiety | Income35-50k | 0.614 | 0.436 | 0.863 | 5.04e-03 | 7.06e-02 | 3093 | 2349 | 744 |
| European | CIDI-MDD | Discontinuation vs Continuation | Recorded Anxiety | Income50-75k | 0.743 | 0.554 | 0.996 | 4.69e-02 | 6.57e-01 | 3093 | 2349 | 744 |
| European | CIDI-MDD | Discontinuation vs Continuation | Recorded Anxiety | Income75-100k | 0.612 | 0.446 | 0.839 | 2.27e-03 | 3.18e-02 | 3093 | 2349 | 744 |
| European | CIDI-MDD | Discontinuation vs Continuation | Recorded Anxiety | Education_levelCollege Postgraduate | 1.071 | 0.379 | 3.027 | 8.98e-01 | 1.00e+00 | 3093 | 2349 | 744 |

| | | | | | | | | | | | | |
|---|---|---|---|---|---|---|---|---|---|---|---|---|
| European | CIDI-MDD | Discontinuation vs Continuation | Recorded Anxiety | Education_levelCollege Undergraduate | 1.332 | 0.477 | 3.721 | 5.85e-01 | 1.00e+00 | 3093 | 2349 | 744 |
| European | CIDI-MDD | Discontinuation vs Continuation | Recorded Anxiety | Education_levelTwelve or GED | 1.188 | 0.407 | 3.470 | 7.52e-01 | 1.00e+00 | 3093 | 2349 | 744 |
| European | CIDI-MDD | Discontinuation vs Continuation | Recorded Anxiety | Recorded Anxiety | 0.675 | 0.570 | 0.799 | 4.72e-06 | 6.61e-05 | 3093 | 2349 | 744 |
| European | CIDI-MDD | Switching vs Continuation | Recorded Anxiety | (Intercept) | 0.375 | 0.171 | 0.823 | 1.44e-02 | 2.02e-01 | 4499 | 2349 | 2150 |
| European | CIDI-MDD | Switching vs Continuation | Recorded Anxiety | age_at_first_AD | 0.729 | 0.681 | 0.780 | <2.2e-16 | <2.2e-16 | 4499 | 2349 | 2150 |
| European | CIDI-MDD | Switching vs Continuation | Recorded Anxiety | sex_at_birthMale | 1.105 | 0.958 | 1.276 | 1.70e-01 | 1.00e+00 | 4499 | 2349 | 2150 |
| European | CIDI-MDD | Switching vs Continuation | Recorded Anxiety | Income>200k | 0.447 | 0.335 | 0.595 | 3.40e-08 | 4.77e-07 | 4499 | 2349 | 2150 |
| European | CIDI-MDD | Switching vs Continuation | Recorded Anxiety | Income100-150k | 0.622 | 0.495 | 0.780 | 4.04e-05 | 5.66e-04 | 4499 | 2349 | 2150 |
| European | CIDI-MDD | Switching vs Continuation | Recorded Anxiety | Income150-200k | 0.447 | 0.335 | 0.596 | 4.08e-08 | 5.71e-07 | 4499 | 2349 | 2150 |
| European | CIDI-MDD | Switching vs Continuation | Recorded Anxiety | Income25-35k | 0.731 | 0.560 | 0.954 | 2.13e-02 | 2.98e-01 | 4499 | 2349 | 2150 |
| European | CIDI-MDD | Switching vs Continuation | Recorded Anxiety | Income35-50k | 0.846 | 0.665 | 1.077 | 1.75e-01 | 1.00e+00 | 4499 | 2349 | 2150 |
| European | CIDI-MDD | Switching vs Continuation | Recorded Anxiety | Income50-75k | 0.772 | 0.619 | 0.963 | 2.20e-02 | 3.08e-01 | 4499 | 2349 | 2150 |
| European | CIDI-MDD | Switching vs Continuation | Recorded Anxiety | Income75-100k | 0.710 | 0.563 | 0.896 | 3.95e-03 | 5.53e-02 | 4499 | 2349 | 2150 |
| European | CIDI-MDD | Switching vs Continuation | Recorded Anxiety | Education_levelCollege Postgraduate | 1.767 | 0.802 | 3.894 | 1.58e-01 | 1.00e+00 | 4499 | 2349 | 2150 |
| European | CIDI-MDD | Switching vs Continuation | Recorded Anxiety | Education_levelCollege Undergraduate | 1.983 | 0.907 | 4.334 | 8.62e-02 | 1.00e+00 | 4499 | 2349 | 2150 |
| European | CIDI-MDD | Switching vs Continuation | Recorded Anxiety | Education_levelTwelve or GED | 2.269 | 1.011 | 5.095 | 4.70e-02 | 6.59e-01 | 4499 | 2349 | 2150 |
| European | CIDI-MDD | Switching vs Continuation | Recorded Anxiety | Recorded Anxiety | 2.186 | 1.911 | 2.500 | <2.2e-16 | <2.2e-16 | 4499 | 2349 | 2150 |
| European | CIDI-MDD | Discontinuation vs Continuation | Smoking_Frequency | (Intercept) | 0.363 | 0.129 | 1.028 | 5.64e-02 | 7.89e-01 | 3050 | 2322 | 728 |
| European | CIDI-MDD | Discontinuation vs Continuation | Smoking_Frequency | age_at_first_AD | 1.009 | 0.922 | 1.105 | 8.43e-01 | 1.00e+00 | 3050 | 2322 | 728 |
| European | CIDI-MDD | Discontinuation vs Continuation | Smoking_Frequency | sex_at_birthMale | 1.048 | 0.862 | 1.273 | 6.40e-01 | 1.00e+00 | 3050 | 2322 | 728 |
| European | CIDI-MDD | Discontinuation vs Continuation | Smoking_Frequency | Income>200k | 0.619 | 0.429 | 0.893 | 1.04e-02 | 1.45e-01 | 3050 | 2322 | 728 |
| European | CIDI-MDD | Discontinuation vs Continuation | Smoking_Frequency | Income100-150k | 0.590 | 0.433 | 0.804 | 8.41e-04 | 1.18e-02 | 3050 | 2322 | 728 |
| European | CIDI-MDD | Discontinuation vs Continuation | Smoking_Frequency | Income150-200k | 0.657 | 0.455 | 0.951 | 2.58e-02 | 3.61e-01 | 3050 | 2322 | 728 |
| European | CIDI-MDD | Discontinuation vs Continuation | Smoking_Frequency | Income25-35k | 0.578 | 0.397 | 0.840 | 4.01e-03 | 5.62e-02 | 3050 | 2322 | 728 |
| European | CIDI-MDD | Discontinuation vs Continuation | Smoking_Frequency | Income35-50k | 0.596 | 0.421 | 0.842 | 3.34e-03 | 4.68e-02 | 3050 | 2322 | 728 |
| European | CIDI-MDD | Discontinuation vs Continuation | Smoking_Frequency | Income50-75k | 0.745 | 0.554 | 1.002 | 5.16e-02 | 7.22e-01 | 3050 | 2322 | 728 |
| European | CIDI-MDD | Discontinuation vs Continuation | Smoking_Frequency | Income75-100k | 0.633 | 0.460 | 0.869 | 4.74e-03 | 6.64e-02 | 3050 | 2322 | 728 |
| European | CIDI-MDD | Discontinuation vs Continuation | Smoking_Frequency | Education_levelCollege Postgraduate | 1.111 | 0.390 | 3.170 | 8.43e-01 | 1.00e+00 | 3050 | 2322 | 728 |
| European | CIDI-MDD | Discontinuation vs Continuation | Smoking_Frequency | Education_levelCollege Undergraduate | 1.375 | 0.489 | 3.869 | 5.46e-01 | 1.00e+00 | 3050 | 2322 | 728 |
| European | CIDI-MDD | Discontinuation vs Continuation | Smoking_Frequency | Education_levelTwelve or GED | 1.251 | 0.427 | 3.664 | 6.83e-01 | 1.00e+00 | 3050 | 2322 | 728 |
| European | CIDI-MDD | Discontinuation vs Continuation | Smoking_Frequency | Smoking_FrequencyEvery Day | 1.081 | 0.732 | 1.596 | 6.96e-01 | 1.00e+00 | 3050 | 2322 | 728 |
| European | CIDI-MDD | Discontinuation vs Continuation | Smoking_Frequency | Smoking_FrequencyNot At All (former) | 0.945 | 0.784 | 1.140 | 5.54e-01 | 1.00e+00 | 3050 | 2322 | 728 |
| European | CIDI-MDD | Discontinuation vs Continuation | Smoking_Frequency | Smoking_FrequencySome Days | 1.206 | 0.717 | 2.027 | 4.80e-01 | 1.00e+00 | 3050 | 2322 | 728 |
| European | CIDI-MDD | Switching vs Continuation | Smoking_Frequency | (Intercept) | 0.509 | 0.232 | 1.114 | 9.08e-02 | 1.00e+00 | 4431 | 2322 | 2109 |
| European | CIDI-MDD | Switching vs Continuation | Smoking_Frequency | age_at_first_AD | 0.668 | 0.624 | 0.715 | <2.2e-16 | <2.2e-16 | 4431 | 2322 | 2109 |
| European | CIDI-MDD | Switching vs Continuation | Smoking_Frequency | sex_at_birthMale | 1.096 | 0.950 | 1.264 | 2.09e-01 | 1.00e+00 | 4431 | 2322 | 2109 |
| European | CIDI-MDD | Switching vs Continuation | Smoking_Frequency | Income>200k | 0.441 | 0.332 | 0.586 | 1.73e-08 | 2.42e-07 | 4431 | 2322 | 2109 |
| European | CIDI-MDD | Switching vs Continuation | Smoking_Frequency | Income100-150k | 0.621 | 0.495 | 0.780 | 3.95e-05 | 5.53e-04 | 4431 | 2322 | 2109 |
| European | CIDI-MDD | Switching vs Continuation | Smoking_Frequency | Income150-200k | 0.444 | 0.333 | 0.593 | 3.43e-08 | 4.80e-07 | 4431 | 2322 | 2109 |
| European | CIDI-MDD | Switching vs Continuation | Smoking_Frequency | Income25-35k | 0.728 | 0.558 | 0.949 | 1.89e-02 | 2.65e-01 | 4431 | 2322 | 2109 |
| European | CIDI-MDD | Switching vs Continuation | Smoking_Frequency | Income35-50k | 0.856 | 0.673 | 1.089 | 2.07e-01 | 1.00e+00 | 4431 | 2322 | 2109 |
| European | CIDI-MDD | Switching vs Continuation | Smoking_Frequency | Income50-75k | 0.768 | 0.616 | 0.958 | 1.92e-02 | 2.69e-01 | 4431 | 2322 | 2109 |
| European | CIDI-MDD | Switching vs Continuation | Smoking_Frequency | Income75-100k | 0.699 | 0.554 | 0.881 | 2.45e-03 | 3.43e-02 | 4431 | 2322 | 2109 |
| European | CIDI-MDD | Switching vs Continuation | Smoking_Frequency | Education_levelCollege Postgraduate | 2.089 | 0.950 | 4.595 | 6.70e-02 | 9.38e-01 | 4431 | 2322 | 2109 |
| European | CIDI-MDD | Switching vs Continuation | Smoking_Frequency | Education_levelCollege Undergraduate | 2.299 | 1.055 | 5.008 | 3.61e-02 | 5.05e-01 | 4431 | 2322 | 2109 |
| European | CIDI-MDD | Switching vs Continuation | Smoking_Frequency | Education_levelTwelve or GED | 2.593 | 1.161 | 5.789 | 2.01e-02 | 2.81e-01 | 4431 | 2322 | 2109 |
| European | CIDI-MDD | Switching vs Continuation | Smoking_Frequency | Smoking_FrequencyEvery Day | 1.445 | 1.107 | 1.887 | 6.81e-03 | 9.54e-02 | 4431 | 2322 | 2109 |
| European | CIDI-MDD | Switching vs Continuation | Smoking_Frequency | Smoking_FrequencyNot At All (former) | 1.134 | 0.991 | 1.297 | 6.80e-02 | 9.52e-01 | 4431 | 2322 | 2109 |
| European | CIDI-MDD | Switching vs Continuation | Smoking_Frequency | Smoking_FrequencySome Days | 0.973 | 0.649 | 1.461 | 8.96e-01 | 1.00e+00 | 4431 | 2322 | 2109 |
| European | CIDI-MDD | Discontinuation vs Continuation | Stress | (Intercept) | 0.359 | 0.129 | 0.997 | 4.93e-02 | 6.90e-01 | 2979 | 2260 | 719 |
| European | CIDI-MDD | Discontinuation vs Continuation | Stress | age_at_first_AD | 1.006 | 0.920 | 1.101 | 8.94e-01 | 1.00e+00 | 2979 | 2260 | 719 |
| European | CIDI-MDD | Discontinuation vs Continuation | Stress | sex_at_birthMale | 1.047 | 0.860 | 1.275 | 6.45e-01 | 1.00e+00 | 2979 | 2260 | 719 |
| European | CIDI-MDD | Discontinuation vs Continuation | Stress | Income>200k | 0.610 | 0.420 | 0.886 | 9.43e-03 | 1.32e-01 | 2979 | 2260 | 719 |
| European | CIDI-MDD | Discontinuation vs Continuation | Stress | Income100-150k | 0.622 | 0.456 | 0.850 | 2.81e-03 | 3.93e-02 | 2979 | 2260 | 719 |
| European | CIDI-MDD | Discontinuation vs Continuation | Stress | Income150-200k | 0.643 | 0.442 | 0.934 | 2.05e-02 | 2.87e-01 | 2979 | 2260 | 719 |
| European | CIDI-MDD | Discontinuation vs Continuation | Stress | Income25-35k | 0.601 | 0.413 | 0.875 | 7.93e-03 | 1.11e-01 | 2979 | 2260 | 719 |
| European | CIDI-MDD | Discontinuation vs Continuation | Stress | Income35-50k | 0.609 | 0.429 | 0.864 | 5.40e-03 | 7.56e-02 | 2979 | 2260 | 719 |
| European | CIDI-MDD | Discontinuation vs Continuation | Stress | Income50-75k | 0.792 | 0.588 | 1.068 | 1.26e-01 | 1.00e+00 | 2979 | 2260 | 719 |
| European | CIDI-MDD | Discontinuation vs Continuation | Stress | Income75-100k | 0.641 | 0.465 | 0.884 | 6.73e-03 | 9.42e-02 | 2979 | 2260 | 719 |
| European | CIDI-MDD | Discontinuation vs Continuation | Stress | Education_levelCollege Postgraduate | 1.031 | 0.366 | 2.909 | 9.54e-01 | 1.00e+00 | 2979 | 2260 | 719 |
| European | CIDI-MDD | Discontinuation vs Continuation | Stress | Education_levelCollege Undergraduate | 1.288 | 0.462 | 3.590 | 6.29e-01 | 1.00e+00 | 2979 | 2260 | 719 |
| European | CIDI-MDD | Discontinuation vs Continuation | Stress | Education_levelTwelve or GED | 1.211 | 0.415 | 3.530 | 7.26e-01 | 1.00e+00 | 2979 | 2260 | 719 |
| European | CIDI-MDD | Discontinuation vs Continuation | Stress | StressAlmost Never | 1.261 | 0.865 | 1.837 | 2.28e-01 | 1.00e+00 | 2979 | 2260 | 719 |
| European | CIDI-MDD | Discontinuation vs Continuation | Stress | StressFairly Often | 0.968 | 0.683 | 1.371 | 8.52e-01 | 1.00e+00 | 2979 | 2260 | 719 |
| European | CIDI-MDD | Discontinuation vs Continuation | Stress | StressSometimes | 1.141 | 0.947 | 1.374 | 1.66e-01 | 1.00e+00 | 2979 | 2260 | 719 |
| European | CIDI-MDD | Discontinuation vs Continuation | Stress | StressVery Often | 1.014 | 0.610 | 1.686 | 9.58e-01 | 1.00e+00 | 2979 | 2260 | 719 |

| | | | | | | | | | | | | |
|---|---|---|---|---|---|---|---|---|---|---|---|---|
| European | CIDI-MDD | Switching vs Continuation | Stress | (Intercept) | 0.503 | 0.226 | 1.117 | 9.13e-02 | 1.00e+00 | 4348 | 2260 | 2088 |
| European | CIDI-MDD | Switching vs Continuation | Stress | age_at_first_AD | 0.681 | 0.636 | 0.728 | <2.2e-16 | <2.2e-16 | 4348 | 2260 | 2088 |
| European | CIDI-MDD | Switching vs Continuation | Stress | sex_at_birthMale | 1.115 | 0.965 | 1.287 | 1.39e-01 | 1.00e+00 | 4348 | 2260 | 2088 |
| European | CIDI-MDD | Switching vs Continuation | Stress | Income>200k | 0.442 | 0.332 | 0.590 | 2.84e-08 | 3.97e-07 | 4348 | 2260 | 2088 |
| European | CIDI-MDD | Switching vs Continuation | Stress | Income100-150k | 0.632 | 0.503 | 0.795 | 8.95e-05 | 1.25e-03 | 4348 | 2260 | 2088 |
| European | CIDI-MDD | Switching vs Continuation | Stress | Income150-200k | 0.452 | 0.338 | 0.603 | 7.23e-08 | 1.01e-06 | 4348 | 2260 | 2088 |
| European | CIDI-MDD | Switching vs Continuation | Stress | Income25-35k | 0.753 | 0.576 | 0.984 | 3.77e-02 | 5.27e-01 | 4348 | 2260 | 2088 |
| European | CIDI-MDD | Switching vs Continuation | Stress | Income35-50k | 0.851 | 0.666 | 1.086 | 1.94e-01 | 1.00e+00 | 4348 | 2260 | 2088 |
| European | CIDI-MDD | Switching vs Continuation | Stress | Income50-75k | 0.781 | 0.625 | 0.976 | 3.01e-02 | 4.21e-01 | 4348 | 2260 | 2088 |
| European | CIDI-MDD | Switching vs Continuation | Stress | Income75-100k | 0.711 | 0.562 | 0.898 | 4.28e-03 | 6.00e-02 | 4348 | 2260 | 2088 |
| European | CIDI-MDD | Switching vs Continuation | Stress | Education_levelCollege Postgraduate | 1.943 | 0.869 | 4.346 | 1.06e-01 | 1.00e+00 | 4348 | 2260 | 2088 |
| European | CIDI-MDD | Switching vs Continuation | Stress | Education_levelCollege Undergraduate | 2.214 | 0.998 | 4.913 | 5.06e-02 | 7.08e-01 | 4348 | 2260 | 2088 |
| European | CIDI-MDD | Switching vs Continuation | Stress | Education_levelTwelve or GED | 2.515 | 1.103 | 5.736 | 2.84e-02 | 3.97e-01 | 4348 | 2260 | 2088 |
| European | CIDI-MDD | Switching vs Continuation | Stress | StressAlmost Never | 1.338 | 1.013 | 1.768 | 4.00e-02 | 5.60e-01 | 4348 | 2260 | 2088 |
| European | CIDI-MDD | Switching vs Continuation | Stress | StressFairly Often | 1.008 | 0.785 | 1.296 | 9.47e-01 | 1.00e+00 | 4348 | 2260 | 2088 |
| European | CIDI-MDD | Switching vs Continuation | Stress | StressSometimes | 1.270 | 1.109 | 1.454 | 5.30e-04 | 7.42e-03 | 4348 | 2260 | 2088 |
| European | CIDI-MDD | Switching vs Continuation | Stress | StressVery Often | 1.661 | 1.182 | 2.332 | 3.42e-03 | 4.78e-02 | 4348 | 2260 | 2088 |
| European | CIDI-MDD | Discontinuation vs Continuation | age_at_first_AD | (Intercept) | 0.316 | 0.287 | 0.347 | <2.2e-16 | <2.2e-16 | 3093 | 2349 | 744 |
| European | CIDI-MDD | Discontinuation vs Continuation | age_at_first_AD | age_at_first_AD | 0.986 | 0.905 | 1.075 | 7.57e-01 | 1.00e+00 | 3093 | 2349 | 744 |
| European | CIDI-MDD | Discontinuation vs Continuation | age_at_first_AD | sex_at_birthMale | 1.016 | 0.839 | 1.231 | 8.68e-01 | 1.00e+00 | 3093 | 2349 | 744 |
| European | CIDI-MDD | Switching vs Continuation | age_at_first_AD | (Intercept) | 0.866 | 0.808 | 0.929 | 5.04e-05 | 7.06e-04 | 4499 | 2349 | 2150 |
| European | CIDI-MDD | Switching vs Continuation | age_at_first_AD | age_at_first_AD | 0.676 | 0.633 | 0.721 | <2.2e-16 | <2.2e-16 | 4499 | 2349 | 2150 |
| European | CIDI-MDD | Switching vs Continuation | age_at_first_AD | sex_at_birthMale | 1.059 | 0.922 | 1.218 | 4.16e-01 | 1.00e+00 | 4499 | 2349 | 2150 |
| European | CIDI-MDD | Discontinuation vs Continuation | sex_at_birth | (Intercept) | 0.316 | 0.287 | 0.347 | <2.2e-16 | <2.2e-16 | 3093 | 2349 | 744 |
| European | CIDI-MDD | Discontinuation vs Continuation | sex_at_birth | age_at_first_AD | 0.986 | 0.905 | 1.075 | 7.57e-01 | 1.00e+00 | 3093 | 2349 | 744 |
| European | CIDI-MDD | Discontinuation vs Continuation | sex_at_birth | sex_at_birthMale | 1.016 | 0.839 | 1.231 | 8.68e-01 | 1.00e+00 | 3093 | 2349 | 744 |
| European | CIDI-MDD | Switching vs Continuation | sex_at_birth | (Intercept) | 0.866 | 0.808 | 0.929 | 5.04e-05 | 7.06e-04 | 4499 | 2349 | 2150 |
| European | CIDI-MDD | Switching vs Continuation | sex_at_birth | age_at_first_AD | 0.676 | 0.633 | 0.721 | <2.2e-16 | <2.2e-16 | 4499 | 2349 | 2150 |
| European | CIDI-MDD | Switching vs Continuation | sex_at_birth | sex_at_birthMale | 1.059 | 0.922 | 1.218 | 4.16e-01 | 1.00e+00 | 4499 | 2349 | 2150 |
| European | CIDI-noMDD | Discontinuation vs Continuation | Alcohol_Frequency | (Intercept) | 0.145 | 0.017 | 1.261 | 8.02e-02 | 1.00e+00 | 2614 | 2012 | 602 |
| European | CIDI-noMDD | Discontinuation vs Continuation | Alcohol_Frequency | age_at_first_AD | 1.269 | 1.135 | 1.420 | 2.98e-05 | 4.17e-04 | 2614 | 2012 | 602 |
| European | CIDI-noMDD | Discontinuation vs Continuation | Alcohol_Frequency | sex_at_birthMale | 1.241 | 1.020 | 1.509 | 3.11e-02 | 4.35e-01 | 2614 | 2012 | 602 |
| European | CIDI-noMDD | Discontinuation vs Continuation | Alcohol_Frequency | Income>200k | 1.113 | 0.689 | 1.798 | 6.63e-01 | 1.00e+00 | 2614 | 2012 | 602 |
| European | CIDI-noMDD | Discontinuation vs Continuation | Alcohol_Frequency | Income100-150k | 0.861 | 0.551 | 1.345 | 5.11e-01 | 1.00e+00 | 2614 | 2012 | 602 |
| European | CIDI-noMDD | Discontinuation vs Continuation | Alcohol_Frequency | Income150-200k | 1.136 | 0.684 | 1.888 | 6.22e-01 | 1.00e+00 | 2614 | 2012 | 602 |
| European | CIDI-noMDD | Discontinuation vs Continuation | Alcohol_Frequency | Income25-35k | 1.031 | 0.612 | 1.736 | 9.09e-01 | 1.00e+00 | 2614 | 2012 | 602 |
| European | CIDI-noMDD | Discontinuation vs Continuation | Alcohol_Frequency | Income35-50k | 1.252 | 0.786 | 1.996 | 3.45e-01 | 1.00e+00 | 2614 | 2012 | 602 |
| European | CIDI-noMDD | Discontinuation vs Continuation | Alcohol_Frequency | Income50-75k | 1.198 | 0.778 | 1.844 | 4.12e-01 | 1.00e+00 | 2614 | 2012 | 602 |
| European | CIDI-noMDD | Discontinuation vs Continuation | Alcohol_Frequency | Income75-100k | 0.937 | 0.596 | 1.474 | 7.79e-01 | 1.00e+00 | 2614 | 2012 | 602 |
| European | CIDI-noMDD | Discontinuation vs Continuation | Alcohol_Frequency | Education_levelCollege Postgraduate | 1.901 | 0.233 | 15.500 | 5.49e-01 | 1.00e+00 | 2614 | 2012 | 602 |
| European | CIDI-noMDD | Discontinuation vs Continuation | Alcohol_Frequency | Education_levelCollege Undergraduate | 2.581 | 0.319 | 20.910 | 3.74e-01 | 1.00e+00 | 2614 | 2012 | 602 |
| European | CIDI-noMDD | Discontinuation vs Continuation | Alcohol_Frequency | Education_levelTwelve or GED | 2.506 | 0.304 | 20.670 | 3.93e-01 | 1.00e+00 | 2614 | 2012 | 602 |
| European | CIDI-noMDD | Discontinuation vs Continuation | Alcohol_Frequency | Alcohol_Frequency2-3 Week | 0.712 | 0.368 | 1.378 | 3.13e-01 | 1.00e+00 | 2614 | 2012 | 602 |
| European | CIDI-noMDD | Discontinuation vs Continuation | Alcohol_Frequency | Alcohol_Frequency2-4 Month | 0.678 | 0.353 | 1.302 | 2.42e-01 | 1.00e+00 | 2614 | 2012 | 602 |
| European | CIDI-noMDD | Discontinuation vs Continuation | Alcohol_Frequency | Alcohol_Frequency4plus Week | 0.635 | 0.329 | 1.223 | 1.74e-01 | 1.00e+00 | 2614 | 2012 | 602 |
| European | CIDI-noMDD | Discontinuation vs Continuation | Alcohol_Frequency | Alcohol_FrequencyMonthly or Less | 0.724 | 0.381 | 1.373 | 3.23e-01 | 1.00e+00 | 2614 | 2012 | 602 |
| European | CIDI-noMDD | Discontinuation vs Continuation | Alcohol_Frequency | Alcohol_FrequencyNever (former) | 0.666 | 0.344 | 1.291 | 2.29e-01 | 1.00e+00 | 2614 | 2012 | 602 |
| European | CIDI-noMDD | Switching vs Continuation | Alcohol_Frequency | (Intercept) | 0.845 | 0.247 | 2.886 | 7.88e-01 | 1.00e+00 | 2654 | 2012 | 642 |
| European | CIDI-noMDD | Switching vs Continuation | Alcohol_Frequency | age_at_first_AD | 0.634 | 0.571 | 0.704 | <2.2e-16 | <2.2e-16 | 2654 | 2012 | 642 |
| European | CIDI-noMDD | Switching vs Continuation | Alcohol_Frequency | sex_at_birthMale | 1.323 | 1.083 | 1.615 | 6.02e-03 | 8.42e-02 | 2654 | 2012 | 642 |
| European | CIDI-noMDD | Switching vs Continuation | Alcohol_Frequency | Income>200k | 0.531 | 0.343 | 0.823 | 4.65e-03 | 6.51e-02 | 2654 | 2012 | 642 |
| European | CIDI-noMDD | Switching vs Continuation | Alcohol_Frequency | Income100-150k | 0.509 | 0.346 | 0.749 | 6.14e-04 | 8.60e-03 | 2654 | 2012 | 642 |
| European | CIDI-noMDD | Switching vs Continuation | Alcohol_Frequency | Income150-200k | 0.616 | 0.389 | 0.975 | 3.86e-02 | 5.40e-01 | 2654 | 2012 | 642 |
| European | CIDI-noMDD | Switching vs Continuation | Alcohol_Frequency | Income25-35k | 0.994 | 0.639 | 1.547 | 9.80e-01 | 1.00e+00 | 2654 | 2012 | 642 |
| European | CIDI-noMDD | Switching vs Continuation | Alcohol_Frequency | Income35-50k | 0.798 | 0.528 | 1.206 | 2.85e-01 | 1.00e+00 | 2654 | 2012 | 642 |
| European | CIDI-noMDD | Switching vs Continuation | Alcohol_Frequency | Income50-75k | 0.699 | 0.480 | 1.017 | 6.12e-02 | 8.57e-01 | 2654 | 2012 | 642 |
| European | CIDI-noMDD | Switching vs Continuation | Alcohol_Frequency | Income75-100k | 0.655 | 0.443 | 0.971 | 3.49e-02 | 4.89e-01 | 2654 | 2012 | 642 |
| European | CIDI-noMDD | Switching vs Continuation | Alcohol_Frequency | Education_levelCollege Postgraduate | 0.748 | 0.232 | 2.407 | 6.26e-01 | 1.00e+00 | 2654 | 2012 | 642 |
| European | CIDI-noMDD | Switching vs Continuation | Alcohol_Frequency | Education_levelCollege Undergraduate | 0.851 | 0.267 | 2.709 | 7.85e-01 | 1.00e+00 | 2654 | 2012 | 642 |
| European | CIDI-noMDD | Switching vs Continuation | Alcohol_Frequency | Education_levelTwelve or GED | 0.941 | 0.289 | 3.072 | 9.20e-01 | 1.00e+00 | 2654 | 2012 | 642 |
| European | CIDI-noMDD | Switching vs Continuation | Alcohol_Frequency | Alcohol_Frequency2-3 Week | 0.635 | 0.344 | 1.174 | 1.47e-01 | 1.00e+00 | 2654 | 2012 | 642 |
| European | CIDI-noMDD | Switching vs Continuation | Alcohol_Frequency | Alcohol_Frequency2-4 Month | 0.775 | 0.427 | 1.406 | 4.02e-01 | 1.00e+00 | 2654 | 2012 | 642 |
| European | CIDI-noMDD | Switching vs Continuation | Alcohol_Frequency | Alcohol_Frequency4plus Week | 0.630 | 0.342 | 1.158 | 1.37e-01 | 1.00e+00 | 2654 | 2012 | 642 |
| European | CIDI-noMDD | Switching vs Continuation | Alcohol_Frequency | Alcohol_FrequencyMonthly or Less | 0.699 | 0.389 | 1.257 | 2.32e-01 | 1.00e+00 | 2654 | 2012 | 642 |
| European | CIDI-noMDD | Switching vs Continuation | Alcohol_Frequency | Alcohol_FrequencyNever (former) | 0.895 | 0.490 | 1.634 | 7.18e-01 | 1.00e+00 | 2654 | 2012 | 642 |
| European | CIDI-noMDD | Discontinuation vs Continuation | BMI | (Intercept) | 0.107 | 0.013 | 0.873 | 3.69e-02 | 5.16e-01 | 2596 | 2000 | 596 |

| | | | | | | | | | | | | |
|---|---|---|---|---|---|---|---|---|---|---|---|---|
| European | CIDI-noMDD | Discontinuation vs Continuation | BMI | age_at_first_AD | 1.256 | 1.123 | 1.405 | 6.69e-05 | 9.37e-04 | 2596 | 2000 | 596 |
| European | CIDI-noMDD | Discontinuation vs Continuation | BMI | sex_at_birthMale | 1.250 | 1.027 | 1.522 | 2.63e-02 | 3.68e-01 | 2596 | 2000 | 596 |
| European | CIDI-noMDD | Discontinuation vs Continuation | BMI | Income>200k | 1.003 | 0.626 | 1.607 | 9.89e-01 | 1.00e+00 | 2596 | 2000 | 596 |
| European | CIDI-noMDD | Discontinuation vs Continuation | BMI | Income100-150k | 0.803 | 0.518 | 1.245 | 3.26e-01 | 1.00e+00 | 2596 | 2000 | 596 |
| European | CIDI-noMDD | Discontinuation vs Continuation | BMI | Income150-200k | 1.071 | 0.650 | 1.766 | 7.87e-01 | 1.00e+00 | 2596 | 2000 | 596 |
| European | CIDI-noMDD | Discontinuation vs Continuation | BMI | Income25-35k | 0.998 | 0.594 | 1.678 | 9.93e-01 | 1.00e+00 | 2596 | 2000 | 596 |
| European | CIDI-noMDD | Discontinuation vs Continuation | BMI | Income35-50k | 1.177 | 0.739 | 1.873 | 4.93e-01 | 1.00e+00 | 2596 | 2000 | 596 |
| European | CIDI-noMDD | Discontinuation vs Continuation | BMI | Income50-75k | 1.153 | 0.752 | 1.768 | 5.13e-01 | 1.00e+00 | 2596 | 2000 | 596 |
| European | CIDI-noMDD | Discontinuation vs Continuation | BMI | Income75-100k | 0.865 | 0.552 | 1.356 | 5.28e-01 | 1.00e+00 | 2596 | 2000 | 596 |
| European | CIDI-noMDD | Discontinuation vs Continuation | BMI | Education_levelCollege Postgraduate | 1.797 | 0.220 | 14.680 | 5.84e-01 | 1.00e+00 | 2596 | 2000 | 596 |
| European | CIDI-noMDD | Discontinuation vs Continuation | BMI | Education_levelCollege Undergraduate | 2.535 | 0.313 | 20.560 | 3.84e-01 | 1.00e+00 | 2596 | 2000 | 596 |
| European | CIDI-noMDD | Discontinuation vs Continuation | BMI | Education_levelTwelve or GED | 2.467 | 0.299 | 20.360 | 4.02e-01 | 1.00e+00 | 2596 | 2000 | 596 |
| European | CIDI-noMDD | Discontinuation vs Continuation | BMI | BMI | 0.905 | 0.808 | 1.013 | 8.25e-02 | 1.00e+00 | 2596 | 2000 | 596 |
| European | CIDI-noMDD | Switching vs Continuation | BMI | (Intercept) | 0.698 | 0.219 | 2.219 | 5.42e-01 | 1.00e+00 | 2636 | 2000 | 636 |
| European | CIDI-noMDD | Switching vs Continuation | BMI | age_at_first_AD | 0.638 | 0.575 | 0.709 | <2.2e-16 | 5.10e-16 | 2636 | 2000 | 636 |
| European | CIDI-noMDD | Switching vs Continuation | BMI | sex_at_birthMale | 1.311 | 1.075 | 1.599 | 7.58e-03 | 1.06e-01 | 2636 | 2000 | 636 |
| European | CIDI-noMDD | Switching vs Continuation | BMI | Income>200k | 0.484 | 0.313 | 0.748 | 1.10e-03 | 1.54e-02 | 2636 | 2000 | 636 |
| European | CIDI-noMDD | Switching vs Continuation | BMI | Income100-150k | 0.490 | 0.334 | 0.719 | 2.66e-04 | 3.73e-03 | 2636 | 2000 | 636 |
| European | CIDI-noMDD | Switching vs Continuation | BMI | Income150-200k | 0.590 | 0.374 | 0.932 | 2.37e-02 | 3.32e-01 | 2636 | 2000 | 636 |
| European | CIDI-noMDD | Switching vs Continuation | BMI | Income25-35k | 0.969 | 0.621 | 1.513 | 8.91e-01 | 1.00e+00 | 2636 | 2000 | 636 |
| European | CIDI-noMDD | Switching vs Continuation | BMI | Income35-50k | 0.768 | 0.508 | 1.163 | 2.13e-01 | 1.00e+00 | 2636 | 2000 | 636 |
| European | CIDI-noMDD | Switching vs Continuation | BMI | Income50-75k | 0.677 | 0.465 | 0.985 | 4.13e-02 | 5.79e-01 | 2636 | 2000 | 636 |
| European | CIDI-noMDD | Switching vs Continuation | BMI | Income75-100k | 0.624 | 0.423 | 0.922 | 1.78e-02 | 2.49e-01 | 2636 | 2000 | 636 |
| European | CIDI-noMDD | Switching vs Continuation | BMI | Education_levelCollege Postgraduate | 0.691 | 0.215 | 2.220 | 5.35e-01 | 1.00e+00 | 2636 | 2000 | 636 |
| European | CIDI-noMDD | Switching vs Continuation | BMI | Education_levelCollege Undergraduate | 0.782 | 0.246 | 2.483 | 6.76e-01 | 1.00e+00 | 2636 | 2000 | 636 |
| European | CIDI-noMDD | Switching vs Continuation | BMI | Education_levelTwelve or GED | 0.909 | 0.279 | 2.961 | 8.75e-01 | 1.00e+00 | 2636 | 2000 | 636 |
| European | CIDI-noMDD | Switching vs Continuation | BMI | BMI | 1.077 | 0.973 | 1.191 | 1.53e-01 | 1.00e+00 | 2636 | 2000 | 636 |
| European | CIDI-noMDD | Discontinuation vs Continuation | Education_level | (Intercept) | 0.106 | 0.013 | 0.862 | 3.58e-02 | 5.01e-01 | 2624 | 2019 | 605 |
| European | CIDI-noMDD | Discontinuation vs Continuation | Education_level | age_at_first_AD | 1.256 | 1.124 | 1.403 | 5.49e-05 | 7.68e-04 | 2624 | 2019 | 605 |
| European | CIDI-noMDD | Discontinuation vs Continuation | Education_level | sex_at_birthMale | 1.217 | 1.002 | 1.479 | 4.73e-02 | 6.63e-01 | 2624 | 2019 | 605 |
| European | CIDI-noMDD | Discontinuation vs Continuation | Education_level | Income>200k | 1.070 | 0.672 | 1.706 | 7.75e-01 | 1.00e+00 | 2624 | 2019 | 605 |
| European | CIDI-noMDD | Discontinuation vs Continuation | Education_level | Income100-150k | 0.838 | 0.542 | 1.296 | 4.26e-01 | 1.00e+00 | 2624 | 2019 | 605 |
| European | CIDI-noMDD | Discontinuation vs Continuation | Education_level | Income150-200k | 1.094 | 0.666 | 1.796 | 7.23e-01 | 1.00e+00 | 2624 | 2019 | 605 |
| European | CIDI-noMDD | Discontinuation vs Continuation | Education_level | Income25-35k | 1.013 | 0.604 | 1.699 | 9.61e-01 | 1.00e+00 | 2624 | 2019 | 605 |
| European | CIDI-noMDD | Discontinuation vs Continuation | Education_level | Income35-50k | 1.235 | 0.778 | 1.960 | 3.71e-01 | 1.00e+00 | 2624 | 2019 | 605 |
| European | CIDI-noMDD | Discontinuation vs Continuation | Education_level | Income50-75k | 1.179 | 0.770 | 1.804 | 4.49e-01 | 1.00e+00 | 2624 | 2019 | 605 |
| European | CIDI-noMDD | Discontinuation vs Continuation | Education_level | Income75-100k | 0.908 | 0.581 | 1.419 | 6.71e-01 | 1.00e+00 | 2624 | 2019 | 605 |
| European | CIDI-noMDD | Discontinuation vs Continuation | Education_level | Education_levelCollege Postgraduate | 1.864 | 0.229 | 15.200 | 5.61e-01 | 1.00e+00 | 2624 | 2019 | 605 |
| European | CIDI-noMDD | Discontinuation vs Continuation | Education_level | Education_levelCollege Undergraduate | 2.548 | 0.315 | 20.650 | 3.81e-01 | 1.00e+00 | 2624 | 2019 | 605 |
| European | CIDI-noMDD | Discontinuation vs Continuation | Education_level | Education_levelTwelve or GED | 2.433 | 0.295 | 20.060 | 4.09e-01 | 1.00e+00 | 2624 | 2019 | 605 |
| European | CIDI-noMDD | Switching vs Continuation | Education_level | (Intercept) | 0.703 | 0.222 | 2.232 | 5.50e-01 | 1.00e+00 | 2664 | 2019 | 645 |
| European | CIDI-noMDD | Switching vs Continuation | Education_level | age_at_first_AD | 0.630 | 0.569 | 0.699 | <2.2e-16 | <2.2e-16 | 2664 | 2019 | 645 |
| European | CIDI-noMDD | Switching vs Continuation | Education_level | sex_at_birthMale | 1.309 | 1.074 | 1.594 | 7.53e-03 | 1.05e-01 | 2664 | 2019 | 645 |
| European | CIDI-noMDD | Switching vs Continuation | Education_level | Income>200k | 0.476 | 0.311 | 0.730 | 6.56e-04 | 9.19e-03 | 2664 | 2019 | 645 |
| European | CIDI-noMDD | Switching vs Continuation | Education_level | Income100-150k | 0.474 | 0.324 | 0.693 | 1.16e-04 | 1.62e-03 | 2664 | 2019 | 645 |
| European | CIDI-noMDD | Switching vs Continuation | Education_level | Income150-200k | 0.563 | 0.358 | 0.883 | 1.24e-02 | 1.74e-01 | 2664 | 2019 | 645 |
| European | CIDI-noMDD | Switching vs Continuation | Education_level | Income25-35k | 0.960 | 0.618 | 1.491 | 8.56e-01 | 1.00e+00 | 2664 | 2019 | 645 |
| European | CIDI-noMDD | Switching vs Continuation | Education_level | Income35-50k | 0.765 | 0.508 | 1.154 | 2.02e-01 | 1.00e+00 | 2664 | 2019 | 645 |
| European | CIDI-noMDD | Switching vs Continuation | Education_level | Income50-75k | 0.667 | 0.460 | 0.967 | 3.27e-02 | 4.58e-01 | 2664 | 2019 | 645 |
| European | CIDI-noMDD | Switching vs Continuation | Education_level | Income75-100k | 0.611 | 0.415 | 0.899 | 1.25e-02 | 1.75e-01 | 2664 | 2019 | 645 |
| European | CIDI-noMDD | Switching vs Continuation | Education_level | Education_levelCollege Postgraduate | 0.698 | 0.218 | 2.236 | 5.45e-01 | 1.00e+00 | 2664 | 2019 | 645 |
| European | CIDI-noMDD | Switching vs Continuation | Education_level | Education_levelCollege Undergraduate | 0.791 | 0.249 | 2.506 | 6.90e-01 | 1.00e+00 | 2664 | 2019 | 645 |
| European | CIDI-noMDD | Switching vs Continuation | Education_level | Education_levelTwelve or GED | 0.896 | 0.276 | 2.910 | 8.55e-01 | 1.00e+00 | 2664 | 2019 | 645 |
| European | CIDI-noMDD | Discontinuation vs Continuation | Income | (Intercept) | 0.106 | 0.013 | 0.862 | 3.58e-02 | 5.01e-01 | 2624 | 2019 | 605 |
| European | CIDI-noMDD | Discontinuation vs Continuation | Income | age_at_first_AD | 1.256 | 1.124 | 1.403 | 5.49e-05 | 7.68e-04 | 2624 | 2019 | 605 |
| European | CIDI-noMDD | Discontinuation vs Continuation | Income | sex_at_birthMale | 1.217 | 1.002 | 1.479 | 4.73e-02 | 6.63e-01 | 2624 | 2019 | 605 |
| European | CIDI-noMDD | Discontinuation vs Continuation | Income | Income>200k | 1.070 | 0.672 | 1.706 | 7.75e-01 | 1.00e+00 | 2624 | 2019 | 605 |
| European | CIDI-noMDD | Discontinuation vs Continuation | Income | Income100-150k | 0.838 | 0.542 | 1.296 | 4.26e-01 | 1.00e+00 | 2624 | 2019 | 605 |
| European | CIDI-noMDD | Discontinuation vs Continuation | Income | Income150-200k | 1.094 | 0.666 | 1.796 | 7.23e-01 | 1.00e+00 | 2624 | 2019 | 605 |
| European | CIDI-noMDD | Discontinuation vs Continuation | Income | Income25-35k | 1.013 | 0.604 | 1.699 | 9.61e-01 | 1.00e+00 | 2624 | 2019 | 605 |
| European | CIDI-noMDD | Discontinuation vs Continuation | Income | Income35-50k | 1.235 | 0.778 | 1.960 | 3.71e-01 | 1.00e+00 | 2624 | 2019 | 605 |
| European | CIDI-noMDD | Discontinuation vs Continuation | Income | Income50-75k | 1.179 | 0.770 | 1.804 | 4.49e-01 | 1.00e+00 | 2624 | 2019 | 605 |
| European | CIDI-noMDD | Discontinuation vs Continuation | Income | Income75-100k | 0.908 | 0.581 | 1.419 | 6.71e-01 | 1.00e+00 | 2624 | 2019 | 605 |
| European | CIDI-noMDD | Discontinuation vs Continuation | Income | Education_levelCollege Postgraduate | 1.864 | 0.229 | 15.200 | 5.61e-01 | 1.00e+00 | 2624 | 2019 | 605 |
| European | CIDI-noMDD | Discontinuation vs Continuation | Income | Education_levelCollege Undergraduate | 2.548 | 0.315 | 20.650 | 3.81e-01 | 1.00e+00 | 2624 | 2019 | 605 |
| European | CIDI-noMDD | Discontinuation vs Continuation | Income | Education_levelTwelve or GED | 2.433 | 0.295 | 20.060 | 4.09e-01 | 1.00e+00 | 2624 | 2019 | 605 |

| | | | | | | | | | | | | |
|---|---|---|---|---|---|---|---|---|---|---|---|---|
| European | CIDI-noMDD | Switching vs Continuation | Income | (Intercept) | 0.703 | 0.222 | 2.232 | 5.50e-01 | 1.00e+00 | 2664 | 2019 | 645 |
| European | CIDI-noMDD | Switching vs Continuation | Income | age_at_first_AD | 0.630 | 0.569 | 0.699 | <2.2e-16 | <2.2e-16 | 2664 | 2019 | 645 |
| European | CIDI-noMDD | Switching vs Continuation | Income | sex_at_birthMale | 1.309 | 1.074 | 1.594 | 7.53e-03 | 1.05e-01 | 2664 | 2019 | 645 |
| European | CIDI-noMDD | Switching vs Continuation | Income | Income>200k | 0.476 | 0.311 | 0.730 | 6.56e-04 | 9.19e-03 | 2664 | 2019 | 645 |
| European | CIDI-noMDD | Switching vs Continuation | Income | Income100-150k | 0.474 | 0.324 | 0.693 | 1.16e-04 | 1.62e-03 | 2664 | 2019 | 645 |
| European | CIDI-noMDD | Switching vs Continuation | Income | Income150-200k | 0.563 | 0.358 | 0.883 | 1.24e-02 | 1.74e-01 | 2664 | 2019 | 645 |
| European | CIDI-noMDD | Switching vs Continuation | Income | Income25-35k | 0.960 | 0.618 | 1.491 | 8.56e-01 | 1.00e+00 | 2664 | 2019 | 645 |
| European | CIDI-noMDD | Switching vs Continuation | Income | Income35-50k | 0.765 | 0.508 | 1.154 | 2.02e-01 | 1.00e+00 | 2664 | 2019 | 645 |
| European | CIDI-noMDD | Switching vs Continuation | Income | Income50-75k | 0.667 | 0.460 | 0.967 | 3.27e-02 | 4.58e-01 | 2664 | 2019 | 645 |
| European | CIDI-noMDD | Switching vs Continuation | Income | Income75-100k | 0.611 | 0.415 | 0.899 | 1.25e-02 | 1.75e-01 | 2664 | 2019 | 645 |
| European | CIDI-noMDD | Switching vs Continuation | Income | Education_levelCollege Postgraduate | 0.698 | 0.218 | 2.236 | 5.45e-01 | 1.00e+00 | 2664 | 2019 | 645 |
| European | CIDI-noMDD | Switching vs Continuation | Income | Education_levelCollege Undergraduate | 0.791 | 0.249 | 2.506 | 6.90e-01 | 1.00e+00 | 2664 | 2019 | 645 |
| European | CIDI-noMDD | Switching vs Continuation | Income | Education_levelTwelve or GED | 0.896 | 0.276 | 2.910 | 8.55e-01 | 1.00e+00 | 2664 | 2019 | 645 |
| European | CIDI-noMDD | Discontinuation vs Continuation | Insomnia | (Intercept) | 0.108 | 0.013 | 0.883 | 3.79e-02 | 5.31e-01 | 2624 | 2019 | 605 |
| European | CIDI-noMDD | Discontinuation vs Continuation | Insomnia | age_at_first_AD | 1.264 | 1.130 | 1.412 | 3.78e-05 | 5.30e-04 | 2624 | 2019 | 605 |
| European | CIDI-noMDD | Discontinuation vs Continuation | Insomnia | sex_at_birthMale | 1.218 | 1.003 | 1.480 | 4.66e-02 | 6.52e-01 | 2624 | 2019 | 605 |
| European | CIDI-noMDD | Discontinuation vs Continuation | Insomnia | Income>200k | 1.069 | 0.671 | 1.704 | 7.80e-01 | 1.00e+00 | 2624 | 2019 | 605 |
| European | CIDI-noMDD | Discontinuation vs Continuation | Insomnia | Income100-150k | 0.839 | 0.542 | 1.298 | 4.30e-01 | 1.00e+00 | 2624 | 2019 | 605 |
| European | CIDI-noMDD | Discontinuation vs Continuation | Insomnia | Income150-200k | 1.098 | 0.669 | 1.803 | 7.12e-01 | 1.00e+00 | 2624 | 2019 | 605 |
| European | CIDI-noMDD | Discontinuation vs Continuation | Insomnia | Income25-35k | 1.008 | 0.601 | 1.691 | 9.76e-01 | 1.00e+00 | 2624 | 2019 | 605 |
| European | CIDI-noMDD | Discontinuation vs Continuation | Insomnia | Income35-50k | 1.240 | 0.781 | 1.969 | 3.62e-01 | 1.00e+00 | 2624 | 2019 | 605 |
| European | CIDI-noMDD | Discontinuation vs Continuation | Insomnia | Income50-75k | 1.179 | 0.770 | 1.805 | 4.48e-01 | 1.00e+00 | 2624 | 2019 | 605 |
| European | CIDI-noMDD | Discontinuation vs Continuation | Insomnia | Income75-100k | 0.910 | 0.582 | 1.422 | 6.79e-01 | 1.00e+00 | 2624 | 2019 | 605 |
| European | CIDI-noMDD | Discontinuation vs Continuation | Insomnia | Education_levelCollege Postgraduate | 1.859 | 0.228 | 15.170 | 5.62e-01 | 1.00e+00 | 2624 | 2019 | 605 |
| European | CIDI-noMDD | Discontinuation vs Continuation | Insomnia | Education_levelCollege Undergraduate | 2.535 | 0.313 | 20.540 | 3.84e-01 | 1.00e+00 | 2624 | 2019 | 605 |
| European | CIDI-noMDD | Discontinuation vs Continuation | Insomnia | Education_levelTwelve or GED | 2.439 | 0.296 | 20.100 | 4.07e-01 | 1.00e+00 | 2624 | 2019 | 605 |
| European | CIDI-noMDD | Discontinuation vs Continuation | Insomnia | Insomnia | 0.892 | 0.714 | 1.114 | 3.14e-01 | 1.00e+00 | 2624 | 2019 | 605 |
| European | CIDI-noMDD | Switching vs Continuation | Insomnia | (Intercept) | 0.523 | 0.162 | 1.693 | 2.80e-01 | 1.00e+00 | 2664 | 2019 | 645 |
| European | CIDI-noMDD | Switching vs Continuation | Insomnia | age_at_first_AD | 0.608 | 0.547 | 0.675 | <2.2e-16 | <2.2e-16 | 2664 | 2019 | 645 |
| European | CIDI-noMDD | Switching vs Continuation | Insomnia | sex_at_birthMale | 1.296 | 1.061 | 1.582 | 1.10e-02 | 1.54e-01 | 2664 | 2019 | 645 |
| European | CIDI-noMDD | Switching vs Continuation | Insomnia | Income>200k | 0.500 | 0.325 | 0.770 | 1.67e-03 | 2.34e-02 | 2664 | 2019 | 645 |
| European | CIDI-noMDD | Switching vs Continuation | Insomnia | Income100-150k | 0.474 | 0.322 | 0.696 | 1.40e-04 | 1.96e-03 | 2664 | 2019 | 645 |
| European | CIDI-noMDD | Switching vs Continuation | Insomnia | Income150-200k | 0.571 | 0.361 | 0.903 | 1.64e-02 | 2.30e-01 | 2664 | 2019 | 645 |
| European | CIDI-noMDD | Switching vs Continuation | Insomnia | Income25-35k | 0.981 | 0.628 | 1.535 | 9.34e-01 | 1.00e+00 | 2664 | 2019 | 645 |
| European | CIDI-noMDD | Switching vs Continuation | Insomnia | Income35-50k | 0.745 | 0.491 | 1.130 | 1.65e-01 | 1.00e+00 | 2664 | 2019 | 645 |
| European | CIDI-noMDD | Switching vs Continuation | Insomnia | Income50-75k | 0.664 | 0.455 | 0.967 | 3.29e-02 | 4.60e-01 | 2664 | 2019 | 645 |
| European | CIDI-noMDD | Switching vs Continuation | Insomnia | Income75-100k | 0.601 | 0.406 | 0.889 | 1.08e-02 | 1.52e-01 | 2664 | 2019 | 645 |
| European | CIDI-noMDD | Switching vs Continuation | Insomnia | Education_levelCollege Postgraduate | 0.743 | 0.228 | 2.421 | 6.22e-01 | 1.00e+00 | 2664 | 2019 | 645 |
| European | CIDI-noMDD | Switching vs Continuation | Insomnia | Education_levelCollege Undergraduate | 0.873 | 0.271 | 2.815 | 8.20e-01 | 1.00e+00 | 2664 | 2019 | 645 |
| European | CIDI-noMDD | Switching vs Continuation | Insomnia | Education_levelTwelve or GED | 0.939 | 0.284 | 3.103 | 9.18e-01 | 1.00e+00 | 2664 | 2019 | 645 |
| European | CIDI-noMDD | Switching vs Continuation | Insomnia | Insomnia | 2.146 | 1.760 | 2.616 | 4.12e-14 | 5.76e-13 | 2664 | 2019 | 645 |
| European | CIDI-noMDD | Discontinuation vs Continuation | MDD_symptom_count | (Intercept) | 0.094 | 0.011 | 0.781 | 2.86e-02 | 4.01e-01 | 2624 | 2019 | 605 |
| European | CIDI-noMDD | Discontinuation vs Continuation | MDD_symptom_count | age_at_first_AD | 1.258 | 1.126 | 1.405 | 4.85e-05 | 6.79e-04 | 2624 | 2019 | 605 |
| European | CIDI-noMDD | Discontinuation vs Continuation | MDD_symptom_count | sex_at_birthMale | 1.215 | 1.001 | 1.476 | 4.94e-02 | 6.91e-01 | 2624 | 2019 | 605 |
| European | CIDI-noMDD | Discontinuation vs Continuation | MDD_symptom_count | Income>200k | 1.064 | 0.667 | 1.696 | 7.96e-01 | 1.00e+00 | 2624 | 2019 | 605 |
| European | CIDI-noMDD | Discontinuation vs Continuation | MDD_symptom_count | Income100-150k | 0.836 | 0.540 | 1.293 | 4.20e-01 | 1.00e+00 | 2624 | 2019 | 605 |
| European | CIDI-noMDD | Discontinuation vs Continuation | MDD_symptom_count | Income150-200k | 1.083 | 0.659 | 1.780 | 7.52e-01 | 1.00e+00 | 2624 | 2019 | 605 |
| European | CIDI-noMDD | Discontinuation vs Continuation | MDD_symptom_count | Income25-35k | 1.012 | 0.603 | 1.698 | 9.64e-01 | 1.00e+00 | 2624 | 2019 | 605 |
| European | CIDI-noMDD | Discontinuation vs Continuation | MDD_symptom_count | Income35-50k | 1.233 | 0.777 | 1.957 | 3.74e-01 | 1.00e+00 | 2624 | 2019 | 605 |
| European | CIDI-noMDD | Discontinuation vs Continuation | MDD_symptom_count | Income50-75k | 1.180 | 0.771 | 1.806 | 4.47e-01 | 1.00e+00 | 2624 | 2019 | 605 |
| European | CIDI-noMDD | Discontinuation vs Continuation | MDD_symptom_count | Income75-100k | 0.904 | 0.578 | 1.413 | 6.57e-01 | 1.00e+00 | 2624 | 2019 | 605 |
| European | CIDI-noMDD | Discontinuation vs Continuation | MDD_symptom_count | Education_levelCollege Postgraduate | 1.909 | 0.234 | 15.580 | 5.46e-01 | 1.00e+00 | 2624 | 2019 | 605 |
| European | CIDI-noMDD | Discontinuation vs Continuation | MDD_symptom_count | Education_levelCollege Undergraduate | 2.607 | 0.322 | 21.140 | 3.70e-01 | 1.00e+00 | 2624 | 2019 | 605 |
| European | CIDI-noMDD | Discontinuation vs Continuation | MDD_symptom_count | Education_levelTwelve or GED | 2.480 | 0.301 | 20.450 | 3.99e-01 | 1.00e+00 | 2624 | 2019 | 605 |
| European | CIDI-noMDD | Discontinuation vs Continuation | MDD_symptom_count | MDD_symptom_count | 0.928 | 0.775 | 1.113 | 4.21e-01 | 1.00e+00 | 2624 | 2019 | 605 |
| European | CIDI-noMDD | Switching vs Continuation | MDD_symptom_count | (Intercept) | 1.271 | 0.388 | 4.165 | 6.93e-01 | 1.00e+00 | 2664 | 2019 | 645 |
| European | CIDI-noMDD | Switching vs Continuation | MDD_symptom_count | age_at_first_AD | 0.616 | 0.555 | 0.684 | <2.2e-16 | <2.2e-16 | 2664 | 2019 | 645 |
| European | CIDI-noMDD | Switching vs Continuation | MDD_symptom_count | sex_at_birthMale | 1.333 | 1.093 | 1.626 | 4.52e-03 | 6.33e-02 | 2664 | 2019 | 645 |
| European | CIDI-noMDD | Switching vs Continuation | MDD_symptom_count | Income>200k | 0.485 | 0.316 | 0.745 | 9.37e-04 | 1.31e-02 | 2664 | 2019 | 645 |
| European | CIDI-noMDD | Switching vs Continuation | MDD_symptom_count | Income100-150k | 0.477 | 0.326 | 0.698 | 1.37e-04 | 1.92e-03 | 2664 | 2019 | 645 |
| European | CIDI-noMDD | Switching vs Continuation | MDD_symptom_count | Income150-200k | 0.591 | 0.376 | 0.929 | 2.27e-02 | 3.17e-01 | 2664 | 2019 | 645 |
| European | CIDI-noMDD | Switching vs Continuation | MDD_symptom_count | Income25-35k | 0.968 | 0.622 | 1.506 | 8.86e-01 | 1.00e+00 | 2664 | 2019 | 645 |
| European | CIDI-noMDD | Switching vs Continuation | MDD_symptom_count | Income35-50k | 0.757 | 0.502 | 1.143 | 1.85e-01 | 1.00e+00 | 2664 | 2019 | 645 |
| European | CIDI-noMDD | Switching vs Continuation | MDD_symptom_count | Income50-75k | 0.653 | 0.450 | 0.948 | 2.51e-02 | 3.51e-01 | 2664 | 2019 | 645 |
| European | CIDI-noMDD | Switching vs Continuation | MDD_symptom_count | Income75-100k | 0.612 | 0.415 | 0.903 | 1.33e-02 | 1.86e-01 | 2664 | 2019 | 645 |
| European | CIDI-noMDD | Switching vs Continuation | MDD_symptom_count | Education_levelCollege Postgraduate | 0.621 | 0.193 | 2.006 | 4.26e-01 | 1.00e+00 | 2664 | 2019 | 645 |

| | | | | | | | | | | | | |
|---|---|---|---|---|---|---|---|---|---|---|---|---|
| European | CIDI-noMDD | Switching vs Continuation | MDD_symptom_count | Education_levelCollege Undergraduate | 0.718 | 0.225 | 2.290 | 5.75e-01 | 1.00e+00 | 2664 | 2019 | 645 |
| European | CIDI-noMDD | Switching vs Continuation | MDD_symptom_count | Education_levelTwelve or GED | 0.833 | 0.255 | 2.723 | 7.62e-01 | 1.00e+00 | 2664 | 2019 | 645 |
| European | CIDI-noMDD | Switching vs Continuation | MDD_symptom_count | MDD_symptom_count | 1.484 | 1.256 | 1.753 | 3.59e-06 | 5.03e-05 | 2664 | 2019 | 645 |
| European | CIDI-noMDD | Discontinuation vs Continuation | Migraine | (Intercept) | 0.110 | 0.013 | 0.892 | 3.88e-02 | 5.43e-01 | 2624 | 2019 | 605 |
| European | CIDI-noMDD | Discontinuation vs Continuation | Migraine | age_at_first_AD | 1.252 | 1.121 | 1.399 | 6.99e-05 | 9.79e-04 | 2624 | 2019 | 605 |
| European | CIDI-noMDD | Discontinuation vs Continuation | Migraine | sex_at_birthMale | 1.190 | 0.978 | 1.448 | 8.16e-02 | 1.00e+00 | 2624 | 2019 | 605 |
| European | CIDI-noMDD | Discontinuation vs Continuation | Migraine | Income>200k | 1.081 | 0.678 | 1.724 | 7.44e-01 | 1.00e+00 | 2624 | 2019 | 605 |
| European | CIDI-noMDD | Discontinuation vs Continuation | Migraine | Income100-150k | 0.843 | 0.545 | 1.305 | 4.43e-01 | 1.00e+00 | 2624 | 2019 | 605 |
| European | CIDI-noMDD | Discontinuation vs Continuation | Migraine | Income150-200k | 1.110 | 0.676 | 1.824 | 6.81e-01 | 1.00e+00 | 2624 | 2019 | 605 |
| European | CIDI-noMDD | Discontinuation vs Continuation | Migraine | Income25-35k | 1.014 | 0.604 | 1.702 | 9.58e-01 | 1.00e+00 | 2624 | 2019 | 605 |
| European | CIDI-noMDD | Discontinuation vs Continuation | Migraine | Income35-50k | 1.238 | 0.779 | 1.965 | 3.66e-01 | 1.00e+00 | 2624 | 2019 | 605 |
| European | CIDI-noMDD | Discontinuation vs Continuation | Migraine | Income50-75k | 1.184 | 0.773 | 1.814 | 4.37e-01 | 1.00e+00 | 2624 | 2019 | 605 |
| European | CIDI-noMDD | Discontinuation vs Continuation | Migraine | Income75-100k | 0.910 | 0.582 | 1.424 | 6.81e-01 | 1.00e+00 | 2624 | 2019 | 605 |
| European | CIDI-noMDD | Discontinuation vs Continuation | Migraine | Education_levelCollege Postgraduate | 1.909 | 0.234 | 15.580 | 5.46e-01 | 1.00e+00 | 2624 | 2019 | 605 |
| European | CIDI-noMDD | Discontinuation vs Continuation | Migraine | Education_levelCollege Undergraduate | 2.594 | 0.320 | 21.030 | 3.72e-01 | 1.00e+00 | 2624 | 2019 | 605 |
| European | CIDI-noMDD | Discontinuation vs Continuation | Migraine | Education_levelTwelve or GED | 2.494 | 0.302 | 20.570 | 3.96e-01 | 1.00e+00 | 2624 | 2019 | 605 |
| European | CIDI-noMDD | Discontinuation vs Continuation | Migraine | Migraine | 0.811 | 0.651 | 1.011 | 6.28e-02 | 8.80e-01 | 2624 | 2019 | 605 |
| European | CIDI-noMDD | Switching vs Continuation | Migraine | (Intercept) | 0.599 | 0.188 | 1.910 | 3.86e-01 | 1.00e+00 | 2664 | 2019 | 645 |
| European | CIDI-noMDD | Switching vs Continuation | Migraine | age_at_first_AD | 0.642 | 0.578 | 0.713 | <2.2e-16 | 1.51e-15 | 2664 | 2019 | 645 |
| European | CIDI-noMDD | Switching vs Continuation | Migraine | sex_at_birthMale | 1.438 | 1.176 | 1.759 | 4.08e-04 | 5.72e-03 | 2664 | 2019 | 645 |
| European | CIDI-noMDD | Switching vs Continuation | Migraine | Income>200k | 0.483 | 0.314 | 0.743 | 9.29e-04 | 1.30e-02 | 2664 | 2019 | 645 |
| European | CIDI-noMDD | Switching vs Continuation | Migraine | Income100-150k | 0.476 | 0.325 | 0.698 | 1.46e-04 | 2.05e-03 | 2664 | 2019 | 645 |
| European | CIDI-noMDD | Switching vs Continuation | Migraine | Income150-200k | 0.563 | 0.357 | 0.887 | 1.33e-02 | 1.86e-01 | 2664 | 2019 | 645 |
| European | CIDI-noMDD | Switching vs Continuation | Migraine | Income25-35k | 0.943 | 0.604 | 1.473 | 7.97e-01 | 1.00e+00 | 2664 | 2019 | 645 |
| European | CIDI-noMDD | Switching vs Continuation | Migraine | Income35-50k | 0.761 | 0.502 | 1.152 | 1.97e-01 | 1.00e+00 | 2664 | 2019 | 645 |
| European | CIDI-noMDD | Switching vs Continuation | Migraine | Income50-75k | 0.669 | 0.460 | 0.973 | 3.56e-02 | 4.98e-01 | 2664 | 2019 | 645 |
| European | CIDI-noMDD | Switching vs Continuation | Migraine | Income75-100k | 0.619 | 0.419 | 0.915 | 1.60e-02 | 2.24e-01 | 2664 | 2019 | 645 |
| European | CIDI-noMDD | Switching vs Continuation | Migraine | Education_levelCollege Postgraduate | 0.619 | 0.192 | 1.994 | 4.22e-01 | 1.00e+00 | 2664 | 2019 | 645 |
| European | CIDI-noMDD | Switching vs Continuation | Migraine | Education_levelCollege Undergraduate | 0.719 | 0.226 | 2.289 | 5.77e-01 | 1.00e+00 | 2664 | 2019 | 645 |
| European | CIDI-noMDD | Switching vs Continuation | Migraine | Education_levelTwelve or GED | 0.815 | 0.250 | 2.658 | 7.34e-01 | 1.00e+00 | 2664 | 2019 | 645 |
| European | CIDI-noMDD | Switching vs Continuation | Migraine | Migraine | 1.932 | 1.593 | 2.342 | 2.05e-11 | 2.87e-10 | 2664 | 2019 | 645 |
| European | CIDI-noMDD | Discontinuation vs Continuation | Recorded Anxiety | (Intercept) | 0.147 | 0.018 | 1.212 | 7.48e-02 | 1.00e+00 | 2624 | 2019 | 605 |
| European | CIDI-noMDD | Discontinuation vs Continuation | Recorded Anxiety | age_at_first_AD | 1.213 | 1.084 | 1.357 | 7.47e-04 | 1.05e-02 | 2624 | 2019 | 605 |
| European | CIDI-noMDD | Discontinuation vs Continuation | Recorded Anxiety | sex_at_birthMale | 1.170 | 0.962 | 1.424 | 1.16e-01 | 1.00e+00 | 2624 | 2019 | 605 |
| European | CIDI-noMDD | Discontinuation vs Continuation | Recorded Anxiety | Income>200k | 0.989 | 0.618 | 1.581 | 9.62e-01 | 1.00e+00 | 2624 | 2019 | 605 |
| European | CIDI-noMDD | Discontinuation vs Continuation | Recorded Anxiety | Income100-150k | 0.790 | 0.509 | 1.225 | 2.92e-01 | 1.00e+00 | 2624 | 2019 | 605 |
| European | CIDI-noMDD | Discontinuation vs Continuation | Recorded Anxiety | Income150-200k | 1.042 | 0.633 | 1.716 | 8.71e-01 | 1.00e+00 | 2624 | 2019 | 605 |
| European | CIDI-noMDD | Discontinuation vs Continuation | Recorded Anxiety | Income25-35k | 0.971 | 0.577 | 1.635 | 9.13e-01 | 1.00e+00 | 2624 | 2019 | 605 |
| European | CIDI-noMDD | Discontinuation vs Continuation | Recorded Anxiety | Income35-50k | 1.155 | 0.725 | 1.839 | 5.44e-01 | 1.00e+00 | 2624 | 2019 | 605 |
| European | CIDI-noMDD | Discontinuation vs Continuation | Recorded Anxiety | Income50-75k | 1.150 | 0.750 | 1.766 | 5.22e-01 | 1.00e+00 | 2624 | 2019 | 605 |
| European | CIDI-noMDD | Discontinuation vs Continuation | Recorded Anxiety | Income75-100k | 0.867 | 0.553 | 1.358 | 5.32e-01 | 1.00e+00 | 2624 | 2019 | 605 |
| European | CIDI-noMDD | Discontinuation vs Continuation | Recorded Anxiety | Education_levelCollege Postgraduate | 1.804 | 0.220 | 14.800 | 5.83e-01 | 1.00e+00 | 2624 | 2019 | 605 |
| European | CIDI-noMDD | Discontinuation vs Continuation | Recorded Anxiety | Education_levelCollege Undergraduate | 2.447 | 0.300 | 19.960 | 4.03e-01 | 1.00e+00 | 2624 | 2019 | 605 |
| European | CIDI-noMDD | Discontinuation vs Continuation | Recorded Anxiety | Education_levelTwelve or GED | 2.377 | 0.286 | 19.730 | 4.23e-01 | 1.00e+00 | 2624 | 2019 | 605 |
| European | CIDI-noMDD | Discontinuation vs Continuation | Recorded Anxiety | Recorded Anxiety | 0.577 | 0.474 | 0.703 | 4.53e-08 | 6.34e-07 | 2624 | 2019 | 605 |
| European | CIDI-noMDD | Switching vs Continuation | Recorded Anxiety | (Intercept) | 0.318 | 0.095 | 1.069 | 6.40e-02 | 8.96e-01 | 2664 | 2019 | 645 |
| European | CIDI-noMDD | Switching vs Continuation | Recorded Anxiety | age_at_first_AD | 0.685 | 0.616 | 0.761 | 2.46e-12 | 3.45e-11 | 2664 | 2019 | 645 |
| European | CIDI-noMDD | Switching vs Continuation | Recorded Anxiety | sex_at_birthMale | 1.411 | 1.152 | 1.728 | 8.73e-04 | 1.22e-02 | 2664 | 2019 | 645 |
| European | CIDI-noMDD | Switching vs Continuation | Recorded Anxiety | Income>200k | 0.548 | 0.354 | 0.849 | 7.07e-03 | 9.90e-02 | 2664 | 2019 | 645 |
| European | CIDI-noMDD | Switching vs Continuation | Recorded Anxiety | Income100-150k | 0.527 | 0.357 | 0.777 | 1.23e-03 | 1.72e-02 | 2664 | 2019 | 645 |
| European | CIDI-noMDD | Switching vs Continuation | Recorded Anxiety | Income150-200k | 0.598 | 0.377 | 0.949 | 2.92e-02 | 4.08e-01 | 2664 | 2019 | 645 |
| European | CIDI-noMDD | Switching vs Continuation | Recorded Anxiety | Income25-35k | 1.025 | 0.652 | 1.612 | 9.15e-01 | 1.00e+00 | 2664 | 2019 | 645 |
| European | CIDI-noMDD | Switching vs Continuation | Recorded Anxiety | Income35-50k | 0.866 | 0.568 | 1.320 | 5.03e-01 | 1.00e+00 | 2664 | 2019 | 645 |
| European | CIDI-noMDD | Switching vs Continuation | Recorded Anxiety | Income50-75k | 0.685 | 0.468 | 1.002 | 5.10e-02 | 7.15e-01 | 2664 | 2019 | 645 |
| European | CIDI-noMDD | Switching vs Continuation | Recorded Anxiety | Income75-100k | 0.672 | 0.452 | 0.999 | 4.94e-02 | 6.91e-01 | 2664 | 2019 | 645 |
| European | CIDI-noMDD | Switching vs Continuation | Recorded Anxiety | Education_levelCollege Postgraduate | 0.722 | 0.215 | 2.431 | 5.99e-01 | 1.00e+00 | 2664 | 2019 | 645 |
| European | CIDI-noMDD | Switching vs Continuation | Recorded Anxiety | Education_levelCollege Undergraduate | 0.829 | 0.249 | 2.760 | 7.60e-01 | 1.00e+00 | 2664 | 2019 | 645 |
| European | CIDI-noMDD | Switching vs Continuation | Recorded Anxiety | Education_levelTwelve or GED | 0.932 | 0.273 | 3.176 | 9.10e-01 | 1.00e+00 | 2664 | 2019 | 645 |
| European | CIDI-noMDD | Switching vs Continuation | Recorded Anxiety | Recorded Anxiety | 2.857 | 2.342 | 3.486 | <2.2e-16 | <2.2e-16 | 2664 | 2019 | 645 |
| European | CIDI-noMDD | Discontinuation vs Continuation | Smoking_Frequency | (Intercept) | 0.100 | 0.012 | 0.831 | 3.31e-02 | 4.64e-01 | 2585 | 1988 | 597 |
| European | CIDI-noMDD | Discontinuation vs Continuation | Smoking_Frequency | age_at_first_AD | 1.266 | 1.131 | 1.419 | 4.49e-05 | 6.28e-04 | 2585 | 1988 | 597 |
| European | CIDI-noMDD | Discontinuation vs Continuation | Smoking_Frequency | sex_at_birthMale | 1.235 | 1.014 | 1.503 | 3.54e-02 | 4.96e-01 | 2585 | 1988 | 597 |
| European | CIDI-noMDD | Discontinuation vs Continuation | Smoking_Frequency | Income>200k | 1.088 | 0.681 | 1.740 | 7.24e-01 | 1.00e+00 | 2585 | 1988 | 597 |
| European | CIDI-noMDD | Discontinuation vs Continuation | Smoking_Frequency | Income100-150k | 0.814 | 0.525 | 1.264 | 3.60e-01 | 1.00e+00 | 2585 | 1988 | 597 |
| European | CIDI-noMDD | Discontinuation vs Continuation | Smoking_Frequency | Income150-200k | 1.087 | 0.659 | 1.793 | 7.44e-01 | 1.00e+00 | 2585 | 1988 | 597 |
| European | CIDI-noMDD | Discontinuation vs Continuation | Smoking_Frequency | Income25-35k | 1.014 | 0.604 | 1.702 | 9.58e-01 | 1.00e+00 | 2585 | 1988 | 597 |

| | | | | | | | | | | | | |
|---|---|---|---|---|---|---|---|---|---|---|---|---|
| European | CIDI-noMDD | Discontinuation vs Continuation | Smoking_Frequency | Income35-50k | 1.257 | 0.791 | 1.998 | 3.34e-01 | 1.00e+00 | 2585 | 1988 | 597 |
| European | CIDI-noMDD | Discontinuation vs Continuation | Smoking_Frequency | Income50-75k | 1.178 | 0.768 | 1.808 | 4.53e-01 | 1.00e+00 | 2585 | 1988 | 597 |
| European | CIDI-noMDD | Discontinuation vs Continuation | Smoking_Frequency | Income75-100k | 0.918 | 0.586 | 1.440 | 7.10e-01 | 1.00e+00 | 2585 | 1988 | 597 |
| European | CIDI-noMDD | Discontinuation vs Continuation | Smoking_Frequency | Education_levelCollege Postgraduate | 2.009 | 0.242 | 16.690 | 5.18e-01 | 1.00e+00 | 2585 | 1988 | 597 |
| European | CIDI-noMDD | Discontinuation vs Continuation | Smoking_Frequency | Education_levelCollege Undergraduate | 2.719 | 0.330 | 22.400 | 3.53e-01 | 1.00e+00 | 2585 | 1988 | 597 |
| European | CIDI-noMDD | Discontinuation vs Continuation | Smoking_Frequency | Education_levelTwelve or GED | 2.489 | 0.297 | 20.860 | 4.01e-01 | 1.00e+00 | 2585 | 1988 | 597 |
| European | CIDI-noMDD | Discontinuation vs Continuation | Smoking_Frequency | Smoking_FrequencyEvery Day | 1.202 | 0.745 | 1.938 | 4.51e-01 | 1.00e+00 | 2585 | 1988 | 597 |
| European | CIDI-noMDD | Discontinuation vs Continuation | Smoking_Frequency | Smoking_FrequencyNot At All (former) | 0.946 | 0.773 | 1.156 | 5.86e-01 | 1.00e+00 | 2585 | 1988 | 597 |
| European | CIDI-noMDD | Discontinuation vs Continuation | Smoking_Frequency | Smoking_FrequencySome Days | 1.053 | 0.560 | 1.981 | 8.73e-01 | 1.00e+00 | 2585 | 1988 | 597 |
| European | CIDI-noMDD | Switching vs Continuation | Smoking_Frequency | (Intercept) | 0.546 | 0.166 | 1.795 | 3.19e-01 | 1.00e+00 | 2618 | 1988 | 630 |
| European | CIDI-noMDD | Switching vs Continuation | Smoking_Frequency | age_at_first_AD | 0.624 | 0.561 | 0.695 | <2.2e-16 | <2.2e-16 | 2618 | 1988 | 630 |
| European | CIDI-noMDD | Switching vs Continuation | Smoking_Frequency | sex_at_birthMale | 1.298 | 1.062 | 1.586 | 1.07e-02 | 1.50e-01 | 2618 | 1988 | 630 |
| European | CIDI-noMDD | Switching vs Continuation | Smoking_Frequency | Income>200k | 0.523 | 0.338 | 0.809 | 3.61e-03 | 5.06e-02 | 2618 | 1988 | 630 |
| European | CIDI-noMDD | Switching vs Continuation | Smoking_Frequency | Income100-150k | 0.522 | 0.354 | 0.769 | 1.02e-03 | 1.43e-02 | 2618 | 1988 | 630 |
| European | CIDI-noMDD | Switching vs Continuation | Smoking_Frequency | Income150-200k | 0.634 | 0.401 | 1.004 | 5.20e-02 | 7.29e-01 | 2618 | 1988 | 630 |
| European | CIDI-noMDD | Switching vs Continuation | Smoking_Frequency | Income25-35k | 1.007 | 0.642 | 1.581 | 9.75e-01 | 1.00e+00 | 2618 | 1988 | 630 |
| European | CIDI-noMDD | Switching vs Continuation | Smoking_Frequency | Income35-50k | 0.861 | 0.567 | 1.306 | 4.81e-01 | 1.00e+00 | 2618 | 1988 | 630 |
| European | CIDI-noMDD | Switching vs Continuation | Smoking_Frequency | Income50-75k | 0.728 | 0.498 | 1.064 | 1.01e-01 | 1.00e+00 | 2618 | 1988 | 630 |
| European | CIDI-noMDD | Switching vs Continuation | Smoking_Frequency | Income75-100k | 0.665 | 0.448 | 0.989 | 4.38e-02 | 6.13e-01 | 2618 | 1988 | 630 |
| European | CIDI-noMDD | Switching vs Continuation | Smoking_Frequency | Education_levelCollege Postgraduate | 0.793 | 0.243 | 2.593 | 7.01e-01 | 1.00e+00 | 2618 | 1988 | 630 |
| European | CIDI-noMDD | Switching vs Continuation | Smoking_Frequency | Education_levelCollege Undergraduate | 0.880 | 0.272 | 2.840 | 8.30e-01 | 1.00e+00 | 2618 | 1988 | 630 |
| European | CIDI-noMDD | Switching vs Continuation | Smoking_Frequency | Education_levelTwelve or GED | 0.992 | 0.300 | 3.279 | 9.89e-01 | 1.00e+00 | 2618 | 1988 | 630 |
| European | CIDI-noMDD | Switching vs Continuation | Smoking_Frequency | Smoking_FrequencyEvery Day | 1.005 | 0.632 | 1.599 | 9.82e-01 | 1.00e+00 | 2618 | 1988 | 630 |
| European | CIDI-noMDD | Switching vs Continuation | Smoking_Frequency | Smoking_FrequencyNot At All (former) | 1.121 | 0.914 | 1.374 | 2.74e-01 | 1.00e+00 | 2618 | 1988 | 630 |
| European | CIDI-noMDD | Switching vs Continuation | Smoking_Frequency | Smoking_FrequencySome Days | 1.599 | 0.937 | 2.730 | 8.53e-02 | 1.00e+00 | 2618 | 1988 | 630 |
| European | CIDI-noMDD | Discontinuation vs Continuation | Stress | (Intercept) | 0.116 | 0.014 | 0.965 | 4.62e-02 | 6.47e-01 | 2535 | 1949 | 586 |
| European | CIDI-noMDD | Discontinuation vs Continuation | Stress | age_at_first_AD | 1.248 | 1.114 | 1.398 | 1.32e-04 | 1.85e-03 | 2535 | 1949 | 586 |
| European | CIDI-noMDD | Discontinuation vs Continuation | Stress | sex_at_birthMale | 1.231 | 1.010 | 1.501 | 3.98e-02 | 5.58e-01 | 2535 | 1949 | 586 |
| European | CIDI-noMDD | Discontinuation vs Continuation | Stress | Income>200k | 1.055 | 0.656 | 1.695 | 8.26e-01 | 1.00e+00 | 2535 | 1949 | 586 |
| European | CIDI-noMDD | Discontinuation vs Continuation | Stress | Income100-150k | 0.829 | 0.532 | 1.291 | 4.06e-01 | 1.00e+00 | 2535 | 1949 | 586 |
| European | CIDI-noMDD | Discontinuation vs Continuation | Stress | Income150-200k | 1.097 | 0.664 | 1.813 | 7.18e-01 | 1.00e+00 | 2535 | 1949 | 586 |
| European | CIDI-noMDD | Discontinuation vs Continuation | Stress | Income25-35k | 1.061 | 0.629 | 1.788 | 8.25e-01 | 1.00e+00 | 2535 | 1949 | 586 |
| European | CIDI-noMDD | Discontinuation vs Continuation | Stress | Income35-50k | 1.262 | 0.789 | 2.018 | 3.31e-01 | 1.00e+00 | 2535 | 1949 | 586 |
| European | CIDI-noMDD | Discontinuation vs Continuation | Stress | Income50-75k | 1.199 | 0.778 | 1.846 | 4.11e-01 | 1.00e+00 | 2535 | 1949 | 586 |
| European | CIDI-noMDD | Discontinuation vs Continuation | Stress | Income75-100k | 0.925 | 0.588 | 1.454 | 7.35e-01 | 1.00e+00 | 2535 | 1949 | 586 |
| European | CIDI-noMDD | Discontinuation vs Continuation | Stress | Education_levelCollege Postgraduate | 1.710 | 0.206 | 14.220 | 6.20e-01 | 1.00e+00 | 2535 | 1949 | 586 |
| European | CIDI-noMDD | Discontinuation vs Continuation | Stress | Education_levelCollege Undergraduate | 2.222 | 0.269 | 18.360 | 4.59e-01 | 1.00e+00 | 2535 | 1949 | 586 |
| European | CIDI-noMDD | Discontinuation vs Continuation | Stress | Education_levelTwelve or GED | 2.139 | 0.254 | 18.000 | 4.84e-01 | 1.00e+00 | 2535 | 1949 | 586 |
| European | CIDI-noMDD | Discontinuation vs Continuation | Stress | StressAlmost Never | 0.978 | 0.650 | 1.473 | 9.16e-01 | 1.00e+00 | 2535 | 1949 | 586 |
| European | CIDI-noMDD | Discontinuation vs Continuation | Stress | StressFairly Often | 1.002 | 0.666 | 1.507 | 9.93e-01 | 1.00e+00 | 2535 | 1949 | 586 |
| European | CIDI-noMDD | Discontinuation vs Continuation | Stress | StressSometimes | 1.082 | 0.856 | 1.369 | 5.10e-01 | 1.00e+00 | 2535 | 1949 | 586 |
| European | CIDI-noMDD | Discontinuation vs Continuation | Stress | StressVery Often | 1.087 | 0.721 | 1.639 | 6.89e-01 | 1.00e+00 | 2535 | 1949 | 586 |
| European | CIDI-noMDD | Switching vs Continuation | Stress | (Intercept) | 0.639 | 0.178 | 2.293 | 4.92e-01 | 1.00e+00 | 2571 | 1949 | 622 |
| European | CIDI-noMDD | Switching vs Continuation | Stress | age_at_first_AD | 0.633 | 0.569 | 0.704 | <2.2e-16 | 5.63e-16 | 2571 | 1949 | 622 |
| European | CIDI-noMDD | Switching vs Continuation | Stress | sex_at_birthMale | 1.311 | 1.071 | 1.604 | 8.53e-03 | 1.19e-01 | 2571 | 1949 | 622 |
| European | CIDI-noMDD | Switching vs Continuation | Stress | Income>200k | 0.439 | 0.283 | 0.680 | 2.34e-04 | 3.28e-03 | 2571 | 1949 | 622 |
| European | CIDI-noMDD | Switching vs Continuation | Stress | Income100-150k | 0.466 | 0.318 | 0.685 | 9.94e-05 | 1.39e-03 | 2571 | 1949 | 622 |
| European | CIDI-noMDD | Switching vs Continuation | Stress | Income150-200k | 0.542 | 0.343 | 0.857 | 8.80e-03 | 1.23e-01 | 2571 | 1949 | 622 |
| European | CIDI-noMDD | Switching vs Continuation | Stress | Income25-35k | 0.964 | 0.617 | 1.507 | 8.72e-01 | 1.00e+00 | 2571 | 1949 | 622 |
| European | CIDI-noMDD | Switching vs Continuation | Stress | Income35-50k | 0.769 | 0.508 | 1.166 | 2.17e-01 | 1.00e+00 | 2571 | 1949 | 622 |
| European | CIDI-noMDD | Switching vs Continuation | Stress | Income50-75k | 0.670 | 0.460 | 0.977 | 3.72e-02 | 5.21e-01 | 2571 | 1949 | 622 |
| European | CIDI-noMDD | Switching vs Continuation | Stress | Income75-100k | 0.627 | 0.424 | 0.926 | 1.90e-02 | 2.66e-01 | 2571 | 1949 | 622 |
| European | CIDI-noMDD | Switching vs Continuation | Stress | Education_levelCollege Postgraduate | 0.803 | 0.222 | 2.897 | 7.37e-01 | 1.00e+00 | 2571 | 1949 | 622 |
| European | CIDI-noMDD | Switching vs Continuation | Stress | Education_levelCollege Undergraduate | 0.858 | 0.240 | 3.065 | 8.13e-01 | 1.00e+00 | 2571 | 1949 | 622 |
| European | CIDI-noMDD | Switching vs Continuation | Stress | Education_levelTwelve or GED | 0.989 | 0.270 | 3.618 | 9.87e-01 | 1.00e+00 | 2571 | 1949 | 622 |
| European | CIDI-noMDD | Switching vs Continuation | Stress | StressAlmost Never | 1.221 | 0.839 | 1.778 | 2.96e-01 | 1.00e+00 | 2571 | 1949 | 622 |
| European | CIDI-noMDD | Switching vs Continuation | Stress | StressFairly Often | 0.840 | 0.554 | 1.272 | 4.10e-01 | 1.00e+00 | 2571 | 1949 | 622 |
| European | CIDI-noMDD | Switching vs Continuation | Stress | StressSometimes | 1.040 | 0.829 | 1.305 | 7.36e-01 | 1.00e+00 | 2571 | 1949 | 622 |
| European | CIDI-noMDD | Switching vs Continuation | Stress | StressVery Often | 0.743 | 0.454 | 1.216 | 2.37e-01 | 1.00e+00 | 2571 | 1949 | 622 |
| European | CIDI-noMDD | Discontinuation vs Continuation | age_at_first_AD | (Intercept) | 0.247 | 0.217 | 0.280 | <2.2e-16 | <2.2e-16 | 2624 | 2019 | 605 |
| European | CIDI-noMDD | Discontinuation vs Continuation | age_at_first_AD | age_at_first_AD | 1.257 | 1.127 | 1.402 | 4.11e-05 | 5.75e-04 | 2624 | 2019 | 605 |
| European | CIDI-noMDD | Discontinuation vs Continuation | age_at_first_AD | sex_at_birthMale | 1.192 | 0.983 | 1.444 | 7.39e-02 | 1.00e+00 | 2624 | 2019 | 605 |
| European | CIDI-noMDD | Switching vs Continuation | age_at_first_AD | (Intercept) | 0.341 | 0.305 | 0.381 | <2.2e-16 | <2.2e-16 | 2664 | 2019 | 645 |
| European | CIDI-noMDD | Switching vs Continuation | age_at_first_AD | age_at_first_AD | 0.638 | 0.576 | 0.706 | <2.2e-16 | <2.2e-16 | 2664 | 2019 | 645 |
| European | CIDI-noMDD | Switching vs Continuation | age_at_first_AD | sex_at_birthMale | 1.267 | 1.042 | 1.539 | 1.74e-02 | 2.44e-01 | 2664 | 2019 | 645 |
| European | CIDI-noMDD | Discontinuation vs Continuation | sex_at_birth | (Intercept) | 0.247 | 0.217 | 0.280 | <2.2e-16 | <2.2e-16 | 2624 | 2019 | 605 |

| | | | | | | | | | | | | |
|---|---|---|---|---|---|---|---|---|---|---|---|---|
| European | CIDI-noMDD | Discontinuation vs Continuation | sex_at_birth | age_at_first_AD | 1.257 | 1.127 | 1.402 | 4.11e-05 | 5.75e-04 | 2624 | 2019 | 605 |
| European | CIDI-noMDD | Discontinuation vs Continuation | sex_at_birth | sex_at_birthMale | 1.192 | 0.983 | 1.444 | 7.39e-02 | 1.00e+00 | 2624 | 2019 | 605 |
| European | CIDI-noMDD | Switching vs Continuation | sex_at_birth | (Intercept) | 0.341 | 0.305 | 0.381 | <2.2e-16 | <2.2e-16 | 2664 | 2019 | 645 |
| European | CIDI-noMDD | Switching vs Continuation | sex_at_birth | age_at_first_AD | 0.638 | 0.576 | 0.706 | <2.2e-16 | <2.2e-16 | 2664 | 2019 | 645 |
| European | CIDI-noMDD | Switching vs Continuation | sex_at_birth | sex_at_birthMale | 1.267 | 1.042 | 1.539 | 1.74e-02 | 2.44e-01 | 2664 | 2019 | 645 |
| African | Full Cohort | Discontinuation vs Continuation | Alcohol_Frequency | (Intercept) | 0.399 | 0.320 | 0.498 | 3.91e-16 | 5.47e-15 | 5806 | 4050 | 1756 |
| African | Full Cohort | Discontinuation vs Continuation | Alcohol_Frequency | age_at_first_AD | 1.002 | 0.939 | 1.069 | 9.60e-01 | 1.00e+00 | 5806 | 4050 | 1756 |
| African | Full Cohort | Discontinuation vs Continuation | Alcohol_Frequency | sex_at_birthMale | 1.204 | 1.059 | 1.370 | 4.57e-03 | 6.39e-02 | 5806 | 4050 | 1756 |
| African | Full Cohort | Discontinuation vs Continuation | Alcohol_Frequency | Income>200k | 0.820 | 0.463 | 1.451 | 4.95e-01 | 1.00e+00 | 5806 | 4050 | 1756 |
| African | Full Cohort | Discontinuation vs Continuation | Alcohol_Frequency | Income100-150k | 0.789 | 0.580 | 1.074 | 1.32e-01 | 1.00e+00 | 5806 | 4050 | 1756 |
| African | Full Cohort | Discontinuation vs Continuation | Alcohol_Frequency | Income150-200k | 1.304 | 0.806 | 2.112 | 2.80e-01 | 1.00e+00 | 5806 | 4050 | 1756 |
| African | Full Cohort | Discontinuation vs Continuation | Alcohol_Frequency | Income25-35k | 1.162 | 0.969 | 1.394 | 1.06e-01 | 1.00e+00 | 5806 | 4050 | 1756 |
| African | Full Cohort | Discontinuation vs Continuation | Alcohol_Frequency | Income35-50k | 0.976 | 0.803 | 1.186 | 8.09e-01 | 1.00e+00 | 5806 | 4050 | 1756 |
| African | Full Cohort | Discontinuation vs Continuation | Alcohol_Frequency | Income50-75k | 0.990 | 0.808 | 1.213 | 9.23e-01 | 1.00e+00 | 5806 | 4050 | 1756 |
| African | Full Cohort | Discontinuation vs Continuation | Alcohol_Frequency | Income75-100k | 0.804 | 0.599 | 1.079 | 1.46e-01 | 1.00e+00 | 5806 | 4050 | 1756 |
| African | Full Cohort | Discontinuation vs Continuation | Alcohol_Frequency | Education_levelCollege Postgraduate | 0.953 | 0.720 | 1.261 | 7.37e-01 | 1.00e+00 | 5806 | 4050 | 1756 |
| African | Full Cohort | Discontinuation vs Continuation | Alcohol_Frequency | Education_levelCollege Undergraduate | 1.065 | 0.870 | 1.303 | 5.43e-01 | 1.00e+00 | 5806 | 4050 | 1756 |
| African | Full Cohort | Discontinuation vs Continuation | Alcohol_Frequency | Education_levelTwelve or GED | 1.081 | 0.880 | 1.327 | 4.60e-01 | 1.00e+00 | 5806 | 4050 | 1756 |
| African | Full Cohort | Discontinuation vs Continuation | Alcohol_Frequency | Alcohol_Frequency2-3 Week | 0.957 | 0.751 | 1.220 | 7.23e-01 | 1.00e+00 | 5806 | 4050 | 1756 |
| African | Full Cohort | Discontinuation vs Continuation | Alcohol_Frequency | Alcohol_Frequency2-4 Month | 1.031 | 0.842 | 1.261 | 7.70e-01 | 1.00e+00 | 5806 | 4050 | 1756 |
| African | Full Cohort | Discontinuation vs Continuation | Alcohol_Frequency | Alcohol_Frequency4plus Week | 1.014 | 0.774 | 1.330 | 9.18e-01 | 1.00e+00 | 5806 | 4050 | 1756 |
| African | Full Cohort | Discontinuation vs Continuation | Alcohol_Frequency | Alcohol_FrequencyMonthly or Less | 0.987 | 0.824 | 1.182 | 8.87e-01 | 1.00e+00 | 5806 | 4050 | 1756 |
| African | Full Cohort | Discontinuation vs Continuation | Alcohol_Frequency | Alcohol_FrequencyNever (former) | 0.936 | 0.761 | 1.151 | 5.31e-01 | 1.00e+00 | 5806 | 4050 | 1756 |
| African | Full Cohort | Switching vs Continuation | Alcohol_Frequency | (Intercept) | 0.658 | 0.541 | 0.801 | 2.91e-05 | 4.08e-04 | 6513 | 4050 | 2463 |
| African | Full Cohort | Switching vs Continuation | Alcohol_Frequency | age_at_first_AD | 0.727 | 0.684 | 0.774 | <2.2e-16 | <2.2e-16 | 6513 | 4050 | 2463 |
| African | Full Cohort | Switching vs Continuation | Alcohol_Frequency | sex_at_birthMale | 0.920 | 0.814 | 1.039 | 1.78e-01 | 1.00e+00 | 6513 | 4050 | 2463 |
| African | Full Cohort | Switching vs Continuation | Alcohol_Frequency | Income>200k | 0.462 | 0.262 | 0.815 | 7.62e-03 | 1.07e-01 | 6513 | 4050 | 2463 |
| African | Full Cohort | Switching vs Continuation | Alcohol_Frequency | Income100-150k | 0.618 | 0.468 | 0.816 | 6.74e-04 | 9.44e-03 | 6513 | 4050 | 2463 |
| African | Full Cohort | Switching vs Continuation | Alcohol_Frequency | Income150-200k | 0.523 | 0.306 | 0.893 | 1.75e-02 | 2.45e-01 | 6513 | 4050 | 2463 |
| African | Full Cohort | Switching vs Continuation | Alcohol_Frequency | Income25-35k | 0.741 | 0.622 | 0.883 | 7.89e-04 | 1.10e-02 | 6513 | 4050 | 2463 |
| African | Full Cohort | Switching vs Continuation | Alcohol_Frequency | Income35-50k | 0.805 | 0.675 | 0.959 | 1.50e-02 | 2.10e-01 | 6513 | 4050 | 2463 |
| African | Full Cohort | Switching vs Continuation | Alcohol_Frequency | Income50-75k | 0.682 | 0.563 | 0.826 | 8.64e-05 | 1.21e-03 | 6513 | 4050 | 2463 |
| African | Full Cohort | Switching vs Continuation | Alcohol_Frequency | Income75-100k | 0.653 | 0.502 | 0.850 | 1.51e-03 | 2.11e-02 | 6513 | 4050 | 2463 |
| African | Full Cohort | Switching vs Continuation | Alcohol_Frequency | Education_levelCollege Postgraduate | 1.092 | 0.852 | 1.399 | 4.87e-01 | 1.00e+00 | 6513 | 4050 | 2463 |
| African | Full Cohort | Switching vs Continuation | Alcohol_Frequency | Education_levelCollege Undergraduate | 1.021 | 0.854 | 1.221 | 8.18e-01 | 1.00e+00 | 6513 | 4050 | 2463 |
| African | Full Cohort | Switching vs Continuation | Alcohol_Frequency | Education_levelTwelve or GED | 0.945 | 0.786 | 1.136 | 5.46e-01 | 1.00e+00 | 6513 | 4050 | 2463 |
| African | Full Cohort | Switching vs Continuation | Alcohol_Frequency | Alcohol_Frequency2-3 Week | 0.979 | 0.784 | 1.222 | 8.52e-01 | 1.00e+00 | 6513 | 4050 | 2463 |
| African | Full Cohort | Switching vs Continuation | Alcohol_Frequency | Alcohol_Frequency2-4 Month | 0.955 | 0.794 | 1.149 | 6.27e-01 | 1.00e+00 | 6513 | 4050 | 2463 |
| African | Full Cohort | Switching vs Continuation | Alcohol_Frequency | Alcohol_Frequency4plus Week | 1.040 | 0.810 | 1.335 | 7.61e-01 | 1.00e+00 | 6513 | 4050 | 2463 |
| African | Full Cohort | Switching vs Continuation | Alcohol_Frequency | Alcohol_FrequencyMonthly or Less | 1.016 | 0.864 | 1.196 | 8.44e-01 | 1.00e+00 | 6513 | 4050 | 2463 |
| African | Full Cohort | Switching vs Continuation | Alcohol_Frequency | Alcohol_FrequencyNever (former) | 1.282 | 1.068 | 1.539 | 7.70e-03 | 1.08e-01 | 6513 | 4050 | 2463 |
| African | Full Cohort | Discontinuation vs Continuation | BMI | (Intercept) | 0.389 | 0.324 | 0.467 | <2.2e-16 | <2.2e-16 | 5888 | 4123 | 1765 |
| African | Full Cohort | Discontinuation vs Continuation | BMI | age_at_first_AD | 0.988 | 0.927 | 1.053 | 7.12e-01 | 1.00e+00 | 5888 | 4123 | 1765 |
| African | Full Cohort | Discontinuation vs Continuation | BMI | sex_at_birthMale | 1.191 | 1.047 | 1.355 | 7.93e-03 | 1.11e-01 | 5888 | 4123 | 1765 |
| African | Full Cohort | Discontinuation vs Continuation | BMI | Income>200k | 0.815 | 0.461 | 1.442 | 4.82e-01 | 1.00e+00 | 5888 | 4123 | 1765 |
| African | Full Cohort | Discontinuation vs Continuation | BMI | Income100-150k | 0.810 | 0.595 | 1.101 | 1.79e-01 | 1.00e+00 | 5888 | 4123 | 1765 |
| African | Full Cohort | Discontinuation vs Continuation | BMI | Income150-200k | 1.264 | 0.777 | 2.057 | 3.45e-01 | 1.00e+00 | 5888 | 4123 | 1765 |
| African | Full Cohort | Discontinuation vs Continuation | BMI | Income25-35k | 1.163 | 0.970 | 1.394 | 1.03e-01 | 1.00e+00 | 5888 | 4123 | 1765 |
| African | Full Cohort | Discontinuation vs Continuation | BMI | Income35-50k | 0.977 | 0.804 | 1.186 | 8.10e-01 | 1.00e+00 | 5888 | 4123 | 1765 |
| African | Full Cohort | Discontinuation vs Continuation | BMI | Income50-75k | 1.001 | 0.819 | 1.225 | 9.90e-01 | 1.00e+00 | 5888 | 4123 | 1765 |
| African | Full Cohort | Discontinuation vs Continuation | BMI | Income75-100k | 0.781 | 0.581 | 1.051 | 1.02e-01 | 1.00e+00 | 5888 | 4123 | 1765 |
| African | Full Cohort | Discontinuation vs Continuation | BMI | Education_levelCollege Postgraduate | 0.965 | 0.731 | 1.275 | 8.04e-01 | 1.00e+00 | 5888 | 4123 | 1765 |
| African | Full Cohort | Discontinuation vs Continuation | BMI | Education_levelCollege Undergraduate | 1.072 | 0.878 | 1.309 | 4.95e-01 | 1.00e+00 | 5888 | 4123 | 1765 |
| African | Full Cohort | Discontinuation vs Continuation | BMI | Education_levelTwelve or GED | 1.100 | 0.897 | 1.349 | 3.60e-01 | 1.00e+00 | 5888 | 4123 | 1765 |
| African | Full Cohort | Discontinuation vs Continuation | BMI | BMI | 0.969 | 0.920 | 1.020 | 2.31e-01 | 1.00e+00 | 5888 | 4123 | 1765 |
| African | Full Cohort | Switching vs Continuation | BMI | (Intercept) | 0.685 | 0.584 | 0.803 | 3.06e-06 | 4.29e-05 | 6625 | 4123 | 2502 |
| African | Full Cohort | Switching vs Continuation | BMI | age_at_first_AD | 0.743 | 0.700 | 0.789 | <2.2e-16 | <2.2e-16 | 6625 | 4123 | 2502 |
| African | Full Cohort | Switching vs Continuation | BMI | sex_at_birthMale | 0.937 | 0.830 | 1.057 | 2.89e-01 | 1.00e+00 | 6625 | 4123 | 2502 |
| African | Full Cohort | Switching vs Continuation | BMI | Income>200k | 0.480 | 0.275 | 0.836 | 9.59e-03 | 1.34e-01 | 6625 | 4123 | 2502 |
| African | Full Cohort | Switching vs Continuation | BMI | Income100-150k | 0.606 | 0.459 | 0.801 | 4.33e-04 | 6.06e-03 | 6625 | 4123 | 2502 |
| African | Full Cohort | Switching vs Continuation | BMI | Income150-200k | 0.581 | 0.347 | 0.975 | 3.96e-02 | 5.54e-01 | 6625 | 4123 | 2502 |
| African | Full Cohort | Switching vs Continuation | BMI | Income25-35k | 0.763 | 0.642 | 0.907 | 2.17e-03 | 3.04e-02 | 6625 | 4123 | 2502 |
| African | Full Cohort | Switching vs Continuation | BMI | Income35-50k | 0.803 | 0.675 | 0.956 | 1.34e-02 | 1.88e-01 | 6625 | 4123 | 2502 |
| African | Full Cohort | Switching vs Continuation | BMI | Income50-75k | 0.674 | 0.558 | 0.815 | 4.57e-05 | 6.40e-04 | 6625 | 4123 | 2502 |
| African | Full Cohort | Switching vs Continuation | BMI | Income75-100k | 0.651 | 0.502 | 0.845 | 1.26e-03 | 1.77e-02 | 6625 | 4123 | 2502 |
| African | Full Cohort | Switching vs Continuation | BMI | Education_levelCollege Postgraduate | 1.103 | 0.865 | 1.407 | 4.30e-01 | 1.00e+00 | 6625 | 4123 | 2502 |

| | | | | | | | | | | | | |
|---|---|---|---|---|---|---|---|---|---|---|---|---|
| African | Full Cohort | Switching vs Continuation | BMI | Education_levelCollege Undergraduate | 1.017 | 0.854 | 1.212 | 8.49e-01 | 1.00e+00 | 6625 | 4123 | 2502 |
| African | Full Cohort | Switching vs Continuation | BMI | Education_levelTwelve or GED | 0.940 | 0.785 | 1.125 | 4.98e-01 | 1.00e+00 | 6625 | 4123 | 2502 |
| African | Full Cohort | Switching vs Continuation | BMI | BMI | 1.002 | 0.958 | 1.048 | 9.46e-01 | 1.00e+00 | 6625 | 4123 | 2502 |
| African | Full Cohort | Discontinuation vs Continuation | Education_level | (Intercept) | 0.394 | 0.330 | 0.472 | <2.2e-16 | <2.2e-16 | 5972 | 4164 | 1808 |
| African | Full Cohort | Discontinuation vs Continuation | Education_level | age_at_first_AD | 0.988 | 0.928 | 1.053 | 7.17e-01 | 1.00e+00 | 5972 | 4164 | 1808 |
| African | Full Cohort | Discontinuation vs Continuation | Education_level | sex_at_birthMale | 1.218 | 1.076 | 1.380 | 1.87e-03 | 2.62e-02 | 5972 | 4164 | 1808 |
| African | Full Cohort | Discontinuation vs Continuation | Education_level | Income>200k | 0.805 | 0.456 | 1.421 | 4.54e-01 | 1.00e+00 | 5972 | 4164 | 1808 |
| African | Full Cohort | Discontinuation vs Continuation | Education_level | Income100-150k | 0.796 | 0.586 | 1.082 | 1.46e-01 | 1.00e+00 | 5972 | 4164 | 1808 |
| African | Full Cohort | Discontinuation vs Continuation | Education_level | Income150-200k | 1.406 | 0.878 | 2.252 | 1.56e-01 | 1.00e+00 | 5972 | 4164 | 1808 |
| African | Full Cohort | Discontinuation vs Continuation | Education_level | Income25-35k | 1.169 | 0.977 | 1.400 | 8.87e-02 | 1.00e+00 | 5972 | 4164 | 1808 |
| African | Full Cohort | Discontinuation vs Continuation | Education_level | Income35-50k | 0.994 | 0.821 | 1.204 | 9.50e-01 | 1.00e+00 | 5972 | 4164 | 1808 |
| African | Full Cohort | Discontinuation vs Continuation | Education_level | Income50-75k | 1.016 | 0.833 | 1.241 | 8.72e-01 | 1.00e+00 | 5972 | 4164 | 1808 |
| African | Full Cohort | Discontinuation vs Continuation | Education_level | Income75-100k | 0.816 | 0.610 | 1.091 | 1.69e-01 | 1.00e+00 | 5972 | 4164 | 1808 |
| African | Full Cohort | Discontinuation vs Continuation | Education_level | Education_levelCollege Postgraduate | 0.944 | 0.716 | 1.243 | 6.80e-01 | 1.00e+00 | 5972 | 4164 | 1808 |
| African | Full Cohort | Discontinuation vs Continuation | Education_level | Education_levelCollege Undergraduate | 1.045 | 0.858 | 1.273 | 6.63e-01 | 1.00e+00 | 5972 | 4164 | 1808 |
| African | Full Cohort | Discontinuation vs Continuation | Education_level | Education_levelTwelve or GED | 1.075 | 0.879 | 1.316 | 4.79e-01 | 1.00e+00 | 5972 | 4164 | 1808 |
| African | Full Cohort | Switching vs Continuation | Education_level | (Intercept) | 0.688 | 0.588 | 0.806 | 3.37e-06 | 4.72e-05 | 6693 | 4164 | 2529 |
| African | Full Cohort | Switching vs Continuation | Education_level | age_at_first_AD | 0.742 | 0.699 | 0.787 | <2.2e-16 | <2.2e-16 | 6693 | 4164 | 2529 |
| African | Full Cohort | Switching vs Continuation | Education_level | sex_at_birthMale | 0.933 | 0.829 | 1.050 | 2.48e-01 | 1.00e+00 | 6693 | 4164 | 2529 |
| African | Full Cohort | Switching vs Continuation | Education_level | Income>200k | 0.471 | 0.270 | 0.820 | 7.75e-03 | 1.09e-01 | 6693 | 4164 | 2529 |
| African | Full Cohort | Switching vs Continuation | Education_level | Income100-150k | 0.613 | 0.465 | 0.808 | 5.20e-04 | 7.29e-03 | 6693 | 4164 | 2529 |
| African | Full Cohort | Switching vs Continuation | Education_level | Income150-200k | 0.582 | 0.347 | 0.975 | 3.99e-02 | 5.59e-01 | 6693 | 4164 | 2529 |
| African | Full Cohort | Switching vs Continuation | Education_level | Income25-35k | 0.760 | 0.640 | 0.903 | 1.83e-03 | 2.56e-02 | 6693 | 4164 | 2529 |
| African | Full Cohort | Switching vs Continuation | Education_level | Income35-50k | 0.806 | 0.678 | 0.958 | 1.43e-02 | 2.01e-01 | 6693 | 4164 | 2529 |
| African | Full Cohort | Switching vs Continuation | Education_level | Income50-75k | 0.676 | 0.559 | 0.816 | 4.71e-05 | 6.59e-04 | 6693 | 4164 | 2529 |
| African | Full Cohort | Switching vs Continuation | Education_level | Income75-100k | 0.650 | 0.501 | 0.842 | 1.12e-03 | 1.57e-02 | 6693 | 4164 | 2529 |
| African | Full Cohort | Switching vs Continuation | Education_level | Education_levelCollege Postgraduate | 1.092 | 0.857 | 1.393 | 4.76e-01 | 1.00e+00 | 6693 | 4164 | 2529 |
| African | Full Cohort | Switching vs Continuation | Education_level | Education_levelCollege Undergraduate | 1.017 | 0.854 | 1.210 | 8.51e-01 | 1.00e+00 | 6693 | 4164 | 2529 |
| African | Full Cohort | Switching vs Continuation | Education_level | Education_levelTwelve or GED | 0.930 | 0.777 | 1.113 | 4.26e-01 | 1.00e+00 | 6693 | 4164 | 2529 |
| African | Full Cohort | Discontinuation vs Continuation | Income | (Intercept) | 0.394 | 0.330 | 0.472 | <2.2e-16 | <2.2e-16 | 5972 | 4164 | 1808 |
| African | Full Cohort | Discontinuation vs Continuation | Income | age_at_first_AD | 0.988 | 0.928 | 1.053 | 7.17e-01 | 1.00e+00 | 5972 | 4164 | 1808 |
| African | Full Cohort | Discontinuation vs Continuation | Income | sex_at_birthMale | 1.218 | 1.076 | 1.380 | 1.87e-03 | 2.62e-02 | 5972 | 4164 | 1808 |
| African | Full Cohort | Discontinuation vs Continuation | Income | Income>200k | 0.805 | 0.456 | 1.421 | 4.54e-01 | 1.00e+00 | 5972 | 4164 | 1808 |
| African | Full Cohort | Discontinuation vs Continuation | Income | Income100-150k | 0.796 | 0.586 | 1.082 | 1.46e-01 | 1.00e+00 | 5972 | 4164 | 1808 |
| African | Full Cohort | Discontinuation vs Continuation | Income | Income150-200k | 1.406 | 0.878 | 2.252 | 1.56e-01 | 1.00e+00 | 5972 | 4164 | 1808 |
| African | Full Cohort | Discontinuation vs Continuation | Income | Income25-35k | 1.169 | 0.977 | 1.400 | 8.87e-02 | 1.00e+00 | 5972 | 4164 | 1808 |
| African | Full Cohort | Discontinuation vs Continuation | Income | Income35-50k | 0.994 | 0.821 | 1.204 | 9.50e-01 | 1.00e+00 | 5972 | 4164 | 1808 |
| African | Full Cohort | Discontinuation vs Continuation | Income | Income50-75k | 1.016 | 0.833 | 1.241 | 8.72e-01 | 1.00e+00 | 5972 | 4164 | 1808 |
| African | Full Cohort | Discontinuation vs Continuation | Income | Income75-100k | 0.816 | 0.610 | 1.091 | 1.69e-01 | 1.00e+00 | 5972 | 4164 | 1808 |
| African | Full Cohort | Discontinuation vs Continuation | Income | Education_levelCollege Postgraduate | 0.944 | 0.716 | 1.243 | 6.80e-01 | 1.00e+00 | 5972 | 4164 | 1808 |
| African | Full Cohort | Discontinuation vs Continuation | Income | Education_levelCollege Undergraduate | 1.045 | 0.858 | 1.273 | 6.63e-01 | 1.00e+00 | 5972 | 4164 | 1808 |
| African | Full Cohort | Discontinuation vs Continuation | Income | Education_levelTwelve or GED | 1.075 | 0.879 | 1.316 | 4.79e-01 | 1.00e+00 | 5972 | 4164 | 1808 |
| African | Full Cohort | Switching vs Continuation | Income | (Intercept) | 0.688 | 0.588 | 0.806 | 3.37e-06 | 4.72e-05 | 6693 | 4164 | 2529 |
| African | Full Cohort | Switching vs Continuation | Income | age_at_first_AD | 0.742 | 0.699 | 0.787 | <2.2e-16 | <2.2e-16 | 6693 | 4164 | 2529 |
| African | Full Cohort | Switching vs Continuation | Income | sex_at_birthMale | 0.933 | 0.829 | 1.050 | 2.48e-01 | 1.00e+00 | 6693 | 4164 | 2529 |
| African | Full Cohort | Switching vs Continuation | Income | Income>200k | 0.471 | 0.270 | 0.820 | 7.75e-03 | 1.09e-01 | 6693 | 4164 | 2529 |
| African | Full Cohort | Switching vs Continuation | Income | Income100-150k | 0.613 | 0.465 | 0.808 | 5.20e-04 | 7.29e-03 | 6693 | 4164 | 2529 |
| African | Full Cohort | Switching vs Continuation | Income | Income150-200k | 0.582 | 0.347 | 0.975 | 3.99e-02 | 5.59e-01 | 6693 | 4164 | 2529 |
| African | Full Cohort | Switching vs Continuation | Income | Income25-35k | 0.760 | 0.640 | 0.903 | 1.83e-03 | 2.56e-02 | 6693 | 4164 | 2529 |
| African | Full Cohort | Switching vs Continuation | Income | Income35-50k | 0.806 | 0.678 | 0.958 | 1.43e-02 | 2.01e-01 | 6693 | 4164 | 2529 |
| African | Full Cohort | Switching vs Continuation | Income | Income50-75k | 0.676 | 0.559 | 0.816 | 4.71e-05 | 6.59e-04 | 6693 | 4164 | 2529 |
| African | Full Cohort | Switching vs Continuation | Income | Income75-100k | 0.650 | 0.501 | 0.842 | 1.12e-03 | 1.57e-02 | 6693 | 4164 | 2529 |
| African | Full Cohort | Switching vs Continuation | Income | Education_levelCollege Postgraduate | 1.092 | 0.857 | 1.393 | 4.76e-01 | 1.00e+00 | 6693 | 4164 | 2529 |
| African | Full Cohort | Switching vs Continuation | Income | Education_levelCollege Undergraduate | 1.017 | 0.854 | 1.210 | 8.51e-01 | 1.00e+00 | 6693 | 4164 | 2529 |
| African | Full Cohort | Switching vs Continuation | Income | Education_levelTwelve or GED | 0.930 | 0.777 | 1.113 | 4.26e-01 | 1.00e+00 | 6693 | 4164 | 2529 |
| African | Full Cohort | Discontinuation vs Continuation | Insomnia | (Intercept) | 0.411 | 0.343 | 0.492 | <2.2e-16 | <2.2e-16 | 5972 | 4164 | 1808 |
| African | Full Cohort | Discontinuation vs Continuation | Insomnia | age_at_first_AD | 0.995 | 0.934 | 1.060 | 8.73e-01 | 1.00e+00 | 5972 | 4164 | 1808 |
| African | Full Cohort | Discontinuation vs Continuation | Insomnia | sex_at_birthMale | 1.214 | 1.072 | 1.375 | 2.25e-03 | 3.15e-02 | 5972 | 4164 | 1808 |
| African | Full Cohort | Discontinuation vs Continuation | Insomnia | Income>200k | 0.808 | 0.457 | 1.427 | 4.62e-01 | 1.00e+00 | 5972 | 4164 | 1808 |
| African | Full Cohort | Discontinuation vs Continuation | Insomnia | Income100-150k | 0.803 | 0.591 | 1.092 | 1.63e-01 | 1.00e+00 | 5972 | 4164 | 1808 |
| African | Full Cohort | Discontinuation vs Continuation | Insomnia | Income150-200k | 1.422 | 0.887 | 2.279 | 1.43e-01 | 1.00e+00 | 5972 | 4164 | 1808 |
| African | Full Cohort | Discontinuation vs Continuation | Insomnia | Income25-35k | 1.167 | 0.975 | 1.398 | 9.22e-02 | 1.00e+00 | 5972 | 4164 | 1808 |
| African | Full Cohort | Discontinuation vs Continuation | Insomnia | Income35-50k | 1.003 | 0.828 | 1.215 | 9.75e-01 | 1.00e+00 | 5972 | 4164 | 1808 |
| African | Full Cohort | Discontinuation vs Continuation | Insomnia | Income50-75k | 1.021 | 0.836 | 1.246 | 8.40e-01 | 1.00e+00 | 5972 | 4164 | 1808 |
| African | Full Cohort | Discontinuation vs Continuation | Insomnia | Income75-100k | 0.821 | 0.614 | 1.097 | 1.82e-01 | 1.00e+00 | 5972 | 4164 | 1808 |
| African | Full Cohort | Discontinuation vs Continuation | Insomnia | Education_levelCollege Postgraduate | 0.941 | 0.714 | 1.240 | 6.66e-01 | 1.00e+00 | 5972 | 4164 | 1808 |

| | | | | | | | | | | | | |
|---|---|---|---|---|---|---|---|---|---|---|---|---|
| African | Full Cohort | Discontinuation vs Continuation | Insomnia | Education_levelCollege Undergraduate | 1.051 | 0.863 | 1.281 | 6.20e-01 | 1.00e+00 | 5972 | 4164 | 1808 |
| African | Full Cohort | Discontinuation vs Continuation | Insomnia | Education_levelTwelve or GED | 1.077 | 0.881 | 1.318 | 4.68e-01 | 1.00e+00 | 5972 | 4164 | 1808 |
| African | Full Cohort | Discontinuation vs Continuation | Insomnia | Insomnia | 0.804 | 0.700 | 0.924 | 2.16e-03 | 3.02e-02 | 5972 | 4164 | 1808 |
| African | Full Cohort | Switching vs Continuation | Insomnia | (Intercept) | 0.512 | 0.435 | 0.604 | 1.87e-15 | 2.62e-14 | 6693 | 4164 | 2529 |
| African | Full Cohort | Switching vs Continuation | Insomnia | age_at_first_AD | 0.721 | 0.678 | 0.766 | <2.2e-16 | <2.2e-16 | 6693 | 4164 | 2529 |
| African | Full Cohort | Switching vs Continuation | Insomnia | sex_at_birthMale | 0.938 | 0.832 | 1.058 | 2.96e-01 | 1.00e+00 | 6693 | 4164 | 2529 |
| African | Full Cohort | Switching vs Continuation | Insomnia | Income>200k | 0.491 | 0.280 | 0.863 | 1.35e-02 | 1.89e-01 | 6693 | 4164 | 2529 |
| African | Full Cohort | Switching vs Continuation | Insomnia | Income100-150k | 0.581 | 0.438 | 0.771 | 1.64e-04 | 2.29e-03 | 6693 | 4164 | 2529 |
| African | Full Cohort | Switching vs Continuation | Insomnia | Income150-200k | 0.564 | 0.333 | 0.956 | 3.35e-02 | 4.68e-01 | 6693 | 4164 | 2529 |
| African | Full Cohort | Switching vs Continuation | Insomnia | Income25-35k | 0.771 | 0.647 | 0.919 | 3.78e-03 | 5.29e-02 | 6693 | 4164 | 2529 |
| African | Full Cohort | Switching vs Continuation | Insomnia | Income35-50k | 0.786 | 0.659 | 0.938 | 7.56e-03 | 1.06e-01 | 6693 | 4164 | 2529 |
| African | Full Cohort | Switching vs Continuation | Insomnia | Income50-75k | 0.672 | 0.554 | 0.815 | 5.27e-05 | 7.38e-04 | 6693 | 4164 | 2529 |
| African | Full Cohort | Switching vs Continuation | Insomnia | Income75-100k | 0.642 | 0.492 | 0.838 | 1.10e-03 | 1.54e-02 | 6693 | 4164 | 2529 |
| African | Full Cohort | Switching vs Continuation | Insomnia | Education_levelCollege Postgraduate | 1.096 | 0.855 | 1.405 | 4.70e-01 | 1.00e+00 | 6693 | 4164 | 2529 |
| African | Full Cohort | Switching vs Continuation | Insomnia | Education_levelCollege Undergraduate | 0.997 | 0.834 | 1.192 | 9.76e-01 | 1.00e+00 | 6693 | 4164 | 2529 |
| African | Full Cohort | Switching vs Continuation | Insomnia | Education_levelTwelve or GED | 0.950 | 0.791 | 1.142 | 5.86e-01 | 1.00e+00 | 6693 | 4164 | 2529 |
| African | Full Cohort | Switching vs Continuation | Insomnia | Insomnia | 2.560 | 2.295 | 2.855 | <2.2e-16 | <2.2e-16 | 6693 | 4164 | 2529 |
| African | Full Cohort | Discontinuation vs Continuation | MDD CIDI-sf | (Intercept) | 0.565 | 0.090 | 3.533 | 5.41e-01 | 1.00e+00 | 325 | 234 | 91 |
| African | Full Cohort | Discontinuation vs Continuation | MDD CIDI-sf | age_at_first_AD | 0.964 | 0.724 | 1.283 | 8.01e-01 | 1.00e+00 | 325 | 234 | 91 |
| African | Full Cohort | Discontinuation vs Continuation | MDD CIDI-sf | sex_at_birthMale | 1.534 | 0.843 | 2.791 | 1.61e-01 | 1.00e+00 | 325 | 234 | 91 |
| African | Full Cohort | Discontinuation vs Continuation | MDD CIDI-sf | Income>200k | 1.749 | 0.436 | 7.009 | 4.30e-01 | 1.00e+00 | 325 | 234 | 91 |
| African | Full Cohort | Discontinuation vs Continuation | MDD CIDI-sf | Income100-150k | 1.383 | 0.571 | 3.348 | 4.72e-01 | 1.00e+00 | 325 | 234 | 91 |
| African | Full Cohort | Discontinuation vs Continuation | MDD CIDI-sf | Income150-200k | 1.748 | 0.268 | 11.390 | 5.59e-01 | 1.00e+00 | 325 | 234 | 91 |
| African | Full Cohort | Discontinuation vs Continuation | MDD CIDI-sf | Income25-35k | 0.818 | 0.324 | 2.065 | 6.71e-01 | 1.00e+00 | 325 | 234 | 91 |
| African | Full Cohort | Discontinuation vs Continuation | MDD CIDI-sf | Income35-50k | 1.312 | 0.606 | 2.838 | 4.91e-01 | 1.00e+00 | 325 | 234 | 91 |
| African | Full Cohort | Discontinuation vs Continuation | MDD CIDI-sf | Income50-75k | 1.011 | 0.441 | 2.315 | 9.80e-01 | 1.00e+00 | 325 | 234 | 91 |
| African | Full Cohort | Discontinuation vs Continuation | MDD CIDI-sf | Income75-100k | 2.074 | 0.799 | 5.380 | 1.34e-01 | 1.00e+00 | 325 | 234 | 91 |
| African | Full Cohort | Discontinuation vs Continuation | MDD CIDI-sf | Education_levelCollege Postgraduate | 0.329 | 0.046 | 2.349 | 2.68e-01 | 1.00e+00 | 325 | 234 | 91 |
| African | Full Cohort | Discontinuation vs Continuation | MDD CIDI-sf | Education_levelCollege Undergraduate | 0.419 | 0.064 | 2.728 | 3.63e-01 | 1.00e+00 | 325 | 234 | 91 |
| African | Full Cohort | Discontinuation vs Continuation | MDD CIDI-sf | Education_levelTwelve or GED | 0.469 | 0.065 | 3.374 | 4.52e-01 | 1.00e+00 | 325 | 234 | 91 |
| African | Full Cohort | Discontinuation vs Continuation | MDD CIDI-sf | MDD CIDI-sf | 1.671 | 0.997 | 2.800 | 5.13e-02 | 7.18e-01 | 325 | 234 | 91 |
| African | Full Cohort | Discontinuation vs Continuation | MDD_symptom_count | (Intercept) | 1.126 | 0.217 | 5.848 | 8.87e-01 | 1.00e+00 | 384 | 279 | 105 |
| African | Full Cohort | Discontinuation vs Continuation | MDD_symptom_count | age_at_first_AD | 0.969 | 0.738 | 1.271 | 8.20e-01 | 1.00e+00 | 384 | 279 | 105 |
| African | Full Cohort | Discontinuation vs Continuation | MDD_symptom_count | sex_at_birthMale | 1.437 | 0.815 | 2.536 | 2.10e-01 | 1.00e+00 | 384 | 279 | 105 |
| African | Full Cohort | Discontinuation vs Continuation | MDD_symptom_count | Income>200k | 1.098 | 0.304 | 3.971 | 8.87e-01 | 1.00e+00 | 384 | 279 | 105 |
| African | Full Cohort | Discontinuation vs Continuation | MDD_symptom_count | Income100-150k | 1.240 | 0.547 | 2.812 | 6.07e-01 | 1.00e+00 | 384 | 279 | 105 |
| African | Full Cohort | Discontinuation vs Continuation | MDD_symptom_count | Income150-200k | 1.969 | 0.398 | 9.741 | 4.06e-01 | 1.00e+00 | 384 | 279 | 105 |
| African | Full Cohort | Discontinuation vs Continuation | MDD_symptom_count | Income25-35k | 0.937 | 0.391 | 2.248 | 8.84e-01 | 1.00e+00 | 384 | 279 | 105 |
| African | Full Cohort | Discontinuation vs Continuation | MDD_symptom_count | Income35-50k | 1.356 | 0.652 | 2.819 | 4.14e-01 | 1.00e+00 | 384 | 279 | 105 |
| African | Full Cohort | Discontinuation vs Continuation | MDD_symptom_count | Income50-75k | 1.258 | 0.593 | 2.669 | 5.51e-01 | 1.00e+00 | 384 | 279 | 105 |
| African | Full Cohort | Discontinuation vs Continuation | MDD_symptom_count | Income75-100k | 1.759 | 0.766 | 4.037 | 1.83e-01 | 1.00e+00 | 384 | 279 | 105 |
| African | Full Cohort | Discontinuation vs Continuation | MDD_symptom_count | Education_levelCollege Postgraduate | 0.258 | 0.044 | 1.510 | 1.33e-01 | 1.00e+00 | 384 | 279 | 105 |
| African | Full Cohort | Discontinuation vs Continuation | MDD_symptom_count | Education_levelCollege Undergraduate | 0.259 | 0.048 | 1.400 | 1.16e-01 | 1.00e+00 | 384 | 279 | 105 |
| African | Full Cohort | Discontinuation vs Continuation | MDD_symptom_count | Education_levelTwelve or GED | 0.291 | 0.049 | 1.738 | 1.76e-01 | 1.00e+00 | 384 | 279 | 105 |
| African | Full Cohort | Discontinuation vs Continuation | MDD_symptom_count | MDD_symptom_count | 1.190 | 0.956 | 1.481 | 1.19e-01 | 1.00e+00 | 384 | 279 | 105 |
| African | Full Cohort | Discontinuation vs Continuation | Migraine | (Intercept) | 0.413 | 0.344 | 0.495 | <2.2e-16 | <2.2e-16 | 5972 | 4164 | 1808 |
| African | Full Cohort | Discontinuation vs Continuation | Migraine | age_at_first_AD | 0.978 | 0.917 | 1.042 | 4.91e-01 | 1.00e+00 | 5972 | 4164 | 1808 |
| African | Full Cohort | Discontinuation vs Continuation | Migraine | sex_at_birthMale | 1.191 | 1.050 | 1.350 | 6.45e-03 | 9.03e-02 | 5972 | 4164 | 1808 |
| African | Full Cohort | Discontinuation vs Continuation | Migraine | Income>200k | 0.802 | 0.454 | 1.416 | 4.46e-01 | 1.00e+00 | 5972 | 4164 | 1808 |
| African | Full Cohort | Discontinuation vs Continuation | Migraine | Income100-150k | 0.802 | 0.589 | 1.090 | 1.58e-01 | 1.00e+00 | 5972 | 4164 | 1808 |
| African | Full Cohort | Discontinuation vs Continuation | Migraine | Income150-200k | 1.413 | 0.882 | 2.264 | 1.51e-01 | 1.00e+00 | 5972 | 4164 | 1808 |
| African | Full Cohort | Discontinuation vs Continuation | Migraine | Income25-35k | 1.169 | 0.977 | 1.400 | 8.88e-02 | 1.00e+00 | 5972 | 4164 | 1808 |
| African | Full Cohort | Discontinuation vs Continuation | Migraine | Income35-50k | 1.003 | 0.828 | 1.216 | 9.73e-01 | 1.00e+00 | 5972 | 4164 | 1808 |
| African | Full Cohort | Discontinuation vs Continuation | Migraine | Income50-75k | 1.026 | 0.840 | 1.253 | 8.00e-01 | 1.00e+00 | 5972 | 4164 | 1808 |
| African | Full Cohort | Discontinuation vs Continuation | Migraine | Income75-100k | 0.820 | 0.613 | 1.097 | 1.81e-01 | 1.00e+00 | 5972 | 4164 | 1808 |
| African | Full Cohort | Discontinuation vs Continuation | Migraine | Education_levelCollege Postgraduate | 0.944 | 0.717 | 1.244 | 6.81e-01 | 1.00e+00 | 5972 | 4164 | 1808 |
| African | Full Cohort | Discontinuation vs Continuation | Migraine | Education_levelCollege Undergraduate | 1.051 | 0.862 | 1.280 | 6.25e-01 | 1.00e+00 | 5972 | 4164 | 1808 |
| African | Full Cohort | Discontinuation vs Continuation | Migraine | Education_levelTwelve or GED | 1.077 | 0.880 | 1.317 | 4.73e-01 | 1.00e+00 | 5972 | 4164 | 1808 |
| African | Full Cohort | Discontinuation vs Continuation | Migraine | Migraine | 0.831 | 0.728 | 0.949 | 6.37e-03 | 8.92e-02 | 5972 | 4164 | 1808 |
| African | Full Cohort | Switching vs Continuation | Migraine | (Intercept) | 0.575 | 0.489 | 0.676 | 2.09e-11 | 2.93e-10 | 6693 | 4164 | 2529 |
| African | Full Cohort | Switching vs Continuation | Migraine | age_at_first_AD | 0.777 | 0.731 | 0.826 | 3.85e-16 | 5.39e-15 | 6693 | 4164 | 2529 |
| African | Full Cohort | Switching vs Continuation | Migraine | sex_at_birthMale | 1.016 | 0.902 | 1.145 | 7.91e-01 | 1.00e+00 | 6693 | 4164 | 2529 |
| African | Full Cohort | Switching vs Continuation | Migraine | Income>200k | 0.467 | 0.267 | 0.817 | 7.63e-03 | 1.07e-01 | 6693 | 4164 | 2529 |
| African | Full Cohort | Switching vs Continuation | Migraine | Income100-150k | 0.593 | 0.449 | 0.784 | 2.40e-04 | 3.36e-03 | 6693 | 4164 | 2529 |
| African | Full Cohort | Switching vs Continuation | Migraine | Income150-200k | 0.590 | 0.350 | 0.993 | 4.71e-02 | 6.59e-01 | 6693 | 4164 | 2529 |
| African | Full Cohort | Switching vs Continuation | Migraine | Income25-35k | 0.764 | 0.642 | 0.909 | 2.42e-03 | 3.38e-02 | 6693 | 4164 | 2529 |

| | | | | | | | | | | | | |
|---|---|---|---|---|---|---|---|---|---|---|---|---|
| African | Full Cohort | Switching vs Continuation | Migraine | Income35-50k | 0.770 | 0.646 | 0.916 | 3.28e-03 | 4.59e-02 | 6693 | 4164 | 2529 |
| African | Full Cohort | Switching vs Continuation | Migraine | Income50-75k | 0.668 | 0.552 | 0.808 | 3.37e-05 | 4.72e-04 | 6693 | 4164 | 2529 |
| African | Full Cohort | Switching vs Continuation | Migraine | Income75-100k | 0.635 | 0.489 | 0.826 | 6.93e-04 | 9.70e-03 | 6693 | 4164 | 2529 |
| African | Full Cohort | Switching vs Continuation | Migraine | Education_levelCollege Postgraduate | 1.064 | 0.833 | 1.359 | 6.21e-01 | 1.00e+00 | 6693 | 4164 | 2529 |
| African | Full Cohort | Switching vs Continuation | Migraine | Education_levelCollege Undergraduate | 0.975 | 0.818 | 1.163 | 7.81e-01 | 1.00e+00 | 6693 | 4164 | 2529 |
| African | Full Cohort | Switching vs Continuation | Migraine | Education_levelTwelve or GED | 0.912 | 0.761 | 1.092 | 3.16e-01 | 1.00e+00 | 6693 | 4164 | 2529 |
| African | Full Cohort | Switching vs Continuation | Migraine | Migraine | 1.831 | 1.642 | 2.041 | <2.2e-16 | <2.2e-16 | 6693 | 4164 | 2529 |
| African | Full Cohort | Discontinuation vs Continuation | Recorded Anxiety | (Intercept) | 0.428 | 0.356 | 0.514 | <2.2e-16 | <2.2e-16 | 5972 | 4164 | 1808 |
| African | Full Cohort | Discontinuation vs Continuation | Recorded Anxiety | age_at_first_AD | 0.974 | 0.914 | 1.038 | 4.15e-01 | 1.00e+00 | 5972 | 4164 | 1808 |
| African | Full Cohort | Discontinuation vs Continuation | Recorded Anxiety | sex_at_birthMale | 1.198 | 1.057 | 1.357 | 4.61e-03 | 6.46e-02 | 5972 | 4164 | 1808 |
| African | Full Cohort | Discontinuation vs Continuation | Recorded Anxiety | Income>200k | 0.789 | 0.447 | 1.395 | 4.15e-01 | 1.00e+00 | 5972 | 4164 | 1808 |
| African | Full Cohort | Discontinuation vs Continuation | Recorded Anxiety | Income100-150k | 0.799 | 0.588 | 1.087 | 1.53e-01 | 1.00e+00 | 5972 | 4164 | 1808 |
| African | Full Cohort | Discontinuation vs Continuation | Recorded Anxiety | Income150-200k | 1.401 | 0.874 | 2.245 | 1.62e-01 | 1.00e+00 | 5972 | 4164 | 1808 |
| African | Full Cohort | Discontinuation vs Continuation | Recorded Anxiety | Income25-35k | 1.173 | 0.979 | 1.404 | 8.34e-02 | 1.00e+00 | 5972 | 4164 | 1808 |
| African | Full Cohort | Discontinuation vs Continuation | Recorded Anxiety | Income35-50k | 1.003 | 0.827 | 1.215 | 9.79e-01 | 1.00e+00 | 5972 | 4164 | 1808 |
| African | Full Cohort | Discontinuation vs Continuation | Recorded Anxiety | Income50-75k | 1.027 | 0.841 | 1.253 | 7.97e-01 | 1.00e+00 | 5972 | 4164 | 1808 |
| African | Full Cohort | Discontinuation vs Continuation | Recorded Anxiety | Income75-100k | 0.825 | 0.617 | 1.103 | 1.95e-01 | 1.00e+00 | 5972 | 4164 | 1808 |
| African | Full Cohort | Discontinuation vs Continuation | Recorded Anxiety | Education_levelCollege Postgraduate | 0.967 | 0.734 | 1.275 | 8.13e-01 | 1.00e+00 | 5972 | 4164 | 1808 |
| African | Full Cohort | Discontinuation vs Continuation | Recorded Anxiety | Education_levelCollege Undergraduate | 1.061 | 0.871 | 1.294 | 5.55e-01 | 1.00e+00 | 5972 | 4164 | 1808 |
| African | Full Cohort | Discontinuation vs Continuation | Recorded Anxiety | Education_levelTwelve or GED | 1.085 | 0.887 | 1.328 | 4.27e-01 | 1.00e+00 | 5972 | 4164 | 1808 |
| African | Full Cohort | Discontinuation vs Continuation | Recorded Anxiety | Recorded Anxiety | 0.818 | 0.730 | 0.916 | 5.06e-04 | 7.09e-03 | 5972 | 4164 | 1808 |
| African | Full Cohort | Switching vs Continuation | Recorded Anxiety | (Intercept) | 0.368 | 0.309 | 0.439 | <2.2e-16 | <2.2e-16 | 6693 | 4164 | 2529 |
| African | Full Cohort | Switching vs Continuation | Recorded Anxiety | age_at_first_AD | 0.800 | 0.752 | 0.851 | 2.00e-12 | 2.79e-11 | 6693 | 4164 | 2529 |
| African | Full Cohort | Switching vs Continuation | Recorded Anxiety | sex_at_birthMale | 1.025 | 0.907 | 1.158 | 6.97e-01 | 1.00e+00 | 6693 | 4164 | 2529 |
| African | Full Cohort | Switching vs Continuation | Recorded Anxiety | Income>200k | 0.487 | 0.275 | 0.862 | 1.35e-02 | 1.89e-01 | 6693 | 4164 | 2529 |
| African | Full Cohort | Switching vs Continuation | Recorded Anxiety | Income100-150k | 0.569 | 0.429 | 0.756 | 1.00e-04 | 1.41e-03 | 6693 | 4164 | 2529 |
| African | Full Cohort | Switching vs Continuation | Recorded Anxiety | Income150-200k | 0.558 | 0.329 | 0.946 | 3.04e-02 | 4.25e-01 | 6693 | 4164 | 2529 |
| African | Full Cohort | Switching vs Continuation | Recorded Anxiety | Income25-35k | 0.737 | 0.617 | 0.880 | 7.70e-04 | 1.08e-02 | 6693 | 4164 | 2529 |
| African | Full Cohort | Switching vs Continuation | Recorded Anxiety | Income35-50k | 0.747 | 0.626 | 0.893 | 1.30e-03 | 1.82e-02 | 6693 | 4164 | 2529 |
| African | Full Cohort | Switching vs Continuation | Recorded Anxiety | Income50-75k | 0.642 | 0.529 | 0.780 | 8.10e-06 | 1.13e-04 | 6693 | 4164 | 2529 |
| African | Full Cohort | Switching vs Continuation | Recorded Anxiety | Income75-100k | 0.605 | 0.463 | 0.790 | 2.22e-04 | 3.10e-03 | 6693 | 4164 | 2529 |
| African | Full Cohort | Switching vs Continuation | Recorded Anxiety | Education_levelCollege Postgraduate | 0.958 | 0.745 | 1.231 | 7.37e-01 | 1.00e+00 | 6693 | 4164 | 2529 |
| African | Full Cohort | Switching vs Continuation | Recorded Anxiety | Education_levelCollege Undergraduate | 0.917 | 0.765 | 1.099 | 3.47e-01 | 1.00e+00 | 6693 | 4164 | 2529 |
| African | Full Cohort | Switching vs Continuation | Recorded Anxiety | Education_levelTwelve or GED | 0.881 | 0.731 | 1.062 | 1.84e-01 | 1.00e+00 | 6693 | 4164 | 2529 |
| African | Full Cohort | Switching vs Continuation | Recorded Anxiety | Recorded Anxiety | 3.150 | 2.818 | 3.521 | <2.2e-16 | <2.2e-16 | 6693 | 4164 | 2529 |
| African | Full Cohort | Discontinuation vs Continuation | Recorded Depression | (Intercept) | 0.436 | 0.362 | 0.524 | <2.2e-16 | <2.2e-16 | 5972 | 4164 | 1808 |
| African | Full Cohort | Discontinuation vs Continuation | Recorded Depression | age_at_first_AD | 0.972 | 0.912 | 1.036 | 3.86e-01 | 1.00e+00 | 5972 | 4164 | 1808 |
| African | Full Cohort | Discontinuation vs Continuation | Recorded Depression | sex_at_birthMale | 1.206 | 1.065 | 1.366 | 3.21e-03 | 4.49e-02 | 5972 | 4164 | 1808 |
| African | Full Cohort | Discontinuation vs Continuation | Recorded Depression | Income>200k | 0.776 | 0.439 | 1.372 | 3.83e-01 | 1.00e+00 | 5972 | 4164 | 1808 |
| African | Full Cohort | Discontinuation vs Continuation | Recorded Depression | Income100-150k | 0.785 | 0.577 | 1.067 | 1.22e-01 | 1.00e+00 | 5972 | 4164 | 1808 |
| African | Full Cohort | Discontinuation vs Continuation | Recorded Depression | Income150-200k | 1.372 | 0.856 | 2.200 | 1.89e-01 | 1.00e+00 | 5972 | 4164 | 1808 |
| African | Full Cohort | Discontinuation vs Continuation | Recorded Depression | Income25-35k | 1.160 | 0.969 | 1.390 | 1.06e-01 | 1.00e+00 | 5972 | 4164 | 1808 |
| African | Full Cohort | Discontinuation vs Continuation | Recorded Depression | Income35-50k | 0.988 | 0.815 | 1.197 | 9.02e-01 | 1.00e+00 | 5972 | 4164 | 1808 |
| African | Full Cohort | Discontinuation vs Continuation | Recorded Depression | Income50-75k | 1.007 | 0.824 | 1.229 | 9.48e-01 | 1.00e+00 | 5972 | 4164 | 1808 |
| African | Full Cohort | Discontinuation vs Continuation | Recorded Depression | Income75-100k | 0.802 | 0.599 | 1.072 | 1.37e-01 | 1.00e+00 | 5972 | 4164 | 1808 |
| African | Full Cohort | Discontinuation vs Continuation | Recorded Depression | Education_levelCollege Postgraduate | 0.950 | 0.721 | 1.252 | 7.18e-01 | 1.00e+00 | 5972 | 4164 | 1808 |
| African | Full Cohort | Discontinuation vs Continuation | Recorded Depression | Education_levelCollege Undergraduate | 1.057 | 0.867 | 1.289 | 5.81e-01 | 1.00e+00 | 5972 | 4164 | 1808 |
| African | Full Cohort | Discontinuation vs Continuation | Recorded Depression | Education_levelTwelve or GED | 1.085 | 0.887 | 1.328 | 4.26e-01 | 1.00e+00 | 5972 | 4164 | 1808 |
| African | Full Cohort | Discontinuation vs Continuation | Recorded Depression | Recorded Depression | 0.768 | 0.684 | 0.862 | 7.43e-06 | 1.04e-04 | 5972 | 4164 | 1808 |
| African | Full Cohort | Switching vs Continuation | Recorded Depression | (Intercept) | 0.328 | 0.274 | 0.392 | <2.2e-16 | <2.2e-16 | 6693 | 4164 | 2529 |
| African | Full Cohort | Switching vs Continuation | Recorded Depression | age_at_first_AD | 0.789 | 0.741 | 0.840 | 1.47e-13 | 2.06e-12 | 6693 | 4164 | 2529 |
| African | Full Cohort | Switching vs Continuation | Recorded Depression | sex_at_birthMale | 0.971 | 0.858 | 1.099 | 6.42e-01 | 1.00e+00 | 6693 | 4164 | 2529 |
| African | Full Cohort | Switching vs Continuation | Recorded Depression | Income>200k | 0.593 | 0.332 | 1.057 | 7.62e-02 | 1.00e+00 | 6693 | 4164 | 2529 |
| African | Full Cohort | Switching vs Continuation | Recorded Depression | Income100-150k | 0.694 | 0.520 | 0.928 | 1.36e-02 | 1.91e-01 | 6693 | 4164 | 2529 |
| African | Full Cohort | Switching vs Continuation | Recorded Depression | Income150-200k | 0.614 | 0.358 | 1.053 | 7.62e-02 | 1.00e+00 | 6693 | 4164 | 2529 |
| African | Full Cohort | Switching vs Continuation | Recorded Depression | Income25-35k | 0.753 | 0.628 | 0.902 | 2.06e-03 | 2.88e-02 | 6693 | 4164 | 2529 |
| African | Full Cohort | Switching vs Continuation | Recorded Depression | Income35-50k | 0.802 | 0.669 | 0.960 | 1.64e-02 | 2.30e-01 | 6693 | 4164 | 2529 |
| African | Full Cohort | Switching vs Continuation | Recorded Depression | Income50-75k | 0.704 | 0.577 | 0.858 | 4.96e-04 | 6.94e-03 | 6693 | 4164 | 2529 |
| African | Full Cohort | Switching vs Continuation | Recorded Depression | Income75-100k | 0.696 | 0.530 | 0.914 | 9.13e-03 | 1.28e-01 | 6693 | 4164 | 2529 |
| African | Full Cohort | Switching vs Continuation | Recorded Depression | Education_levelCollege Postgraduate | 1.028 | 0.797 | 1.327 | 8.32e-01 | 1.00e+00 | 6693 | 4164 | 2529 |
| African | Full Cohort | Switching vs Continuation | Recorded Depression | Education_levelCollege Undergraduate | 0.922 | 0.768 | 1.108 | 3.88e-01 | 1.00e+00 | 6693 | 4164 | 2529 |
| African | Full Cohort | Switching vs Continuation | Recorded Depression | Education_levelTwelve or GED | 0.880 | 0.728 | 1.064 | 1.86e-01 | 1.00e+00 | 6693 | 4164 | 2529 |
| African | Full Cohort | Switching vs Continuation | Recorded Depression | Recorded Depression | 3.904 | 3.499 | 4.356 | <2.2e-16 | <2.2e-16 | 6693 | 4164 | 2529 |
| African | Full Cohort | Discontinuation vs Continuation | Smoking_Frequency | (Intercept) | 0.382 | 0.315 | 0.462 | <2.2e-16 | <2.2e-16 | 5834 | 4076 | 1758 |
| African | Full Cohort | Discontinuation vs Continuation | Smoking_Frequency | age_at_first_AD | 1.010 | 0.946 | 1.079 | 7.58e-01 | 1.00e+00 | 5834 | 4076 | 1758 |
| African | Full Cohort | Discontinuation vs Continuation | Smoking_Frequency | sex_at_birthMale | 1.217 | 1.069 | 1.384 | 2.87e-03 | 4.01e-02 | 5834 | 4076 | 1758 |

| | | | | | | | | | | | | |
|---|---|---|---|---|---|---|---|---|---|---|---|---|
| African | Full Cohort | Discontinuation vs Continuation | Smoking_Frequency | Income>200k | 0.822 | 0.458 | 1.476 | 5.12e-01 | 1.00e+00 | 5834 | 4076 | 1758 |
| African | Full Cohort | Discontinuation vs Continuation | Smoking_Frequency | Income100-150k | 0.833 | 0.611 | 1.135 | 2.46e-01 | 1.00e+00 | 5834 | 4076 | 1758 |
| African | Full Cohort | Discontinuation vs Continuation | Smoking_Frequency | Income150-200k | 1.296 | 0.796 | 2.110 | 2.98e-01 | 1.00e+00 | 5834 | 4076 | 1758 |
| African | Full Cohort | Discontinuation vs Continuation | Smoking_Frequency | Income25-35k | 1.188 | 0.989 | 1.427 | 6.60e-02 | 9.23e-01 | 5834 | 4076 | 1758 |
| African | Full Cohort | Discontinuation vs Continuation | Smoking_Frequency | Income35-50k | 1.017 | 0.836 | 1.236 | 8.68e-01 | 1.00e+00 | 5834 | 4076 | 1758 |
| African | Full Cohort | Discontinuation vs Continuation | Smoking_Frequency | Income50-75k | 1.033 | 0.842 | 1.267 | 7.55e-01 | 1.00e+00 | 5834 | 4076 | 1758 |
| African | Full Cohort | Discontinuation vs Continuation | Smoking_Frequency | Income75-100k | 0.853 | 0.635 | 1.144 | 2.88e-01 | 1.00e+00 | 5834 | 4076 | 1758 |
| African | Full Cohort | Discontinuation vs Continuation | Smoking_Frequency | Education_levelCollege Postgraduate | 0.963 | 0.727 | 1.275 | 7.91e-01 | 1.00e+00 | 5834 | 4076 | 1758 |
| African | Full Cohort | Discontinuation vs Continuation | Smoking_Frequency | Education_levelCollege Undergraduate | 1.077 | 0.881 | 1.316 | 4.72e-01 | 1.00e+00 | 5834 | 4076 | 1758 |
| African | Full Cohort | Discontinuation vs Continuation | Smoking_Frequency | Education_levelTwelve or GED | 1.081 | 0.881 | 1.326 | 4.57e-01 | 1.00e+00 | 5834 | 4076 | 1758 |
| African | Full Cohort | Discontinuation vs Continuation | Smoking_Frequency | Smoking_FrequencyEvery Day | 1.236 | 1.052 | 1.452 | 9.93e-03 | 1.39e-01 | 5834 | 4076 | 1758 |
| African | Full Cohort | Discontinuation vs Continuation | Smoking_Frequency | Smoking_FrequencyNot At All (former) | 0.830 | 0.709 | 0.971 | 2.02e-02 | 2.83e-01 | 5834 | 4076 | 1758 |
| African | Full Cohort | Discontinuation vs Continuation | Smoking_Frequency | Smoking_FrequencySome Days | 0.993 | 0.811 | 1.217 | 9.49e-01 | 1.00e+00 | 5834 | 4076 | 1758 |
| African | Full Cohort | Switching vs Continuation | Smoking_Frequency | (Intercept) | 0.605 | 0.510 | 0.718 | 9.39e-09 | 1.32e-07 | 6515 | 4076 | 2439 |
| African | Full Cohort | Switching vs Continuation | Smoking_Frequency | age_at_first_AD | 0.730 | 0.686 | 0.777 | <2.2e-16 | <2.2e-16 | 6515 | 4076 | 2439 |
| African | Full Cohort | Switching vs Continuation | Smoking_Frequency | sex_at_birthMale | 0.884 | 0.782 | 0.999 | 4.80e-02 | 6.73e-01 | 6515 | 4076 | 2439 |
| African | Full Cohort | Switching vs Continuation | Smoking_Frequency | Income>200k | 0.461 | 0.257 | 0.825 | 9.16e-03 | 1.28e-01 | 6515 | 4076 | 2439 |
| African | Full Cohort | Switching vs Continuation | Smoking_Frequency | Income100-150k | 0.636 | 0.481 | 0.841 | 1.48e-03 | 2.08e-02 | 6515 | 4076 | 2439 |
| African | Full Cohort | Switching vs Continuation | Smoking_Frequency | Income150-200k | 0.578 | 0.342 | 0.979 | 4.13e-02 | 5.78e-01 | 6515 | 4076 | 2439 |
| African | Full Cohort | Switching vs Continuation | Smoking_Frequency | Income25-35k | 0.774 | 0.649 | 0.922 | 4.22e-03 | 5.90e-02 | 6515 | 4076 | 2439 |
| African | Full Cohort | Switching vs Continuation | Smoking_Frequency | Income35-50k | 0.822 | 0.690 | 0.980 | 2.89e-02 | 4.05e-01 | 6515 | 4076 | 2439 |
| African | Full Cohort | Switching vs Continuation | Smoking_Frequency | Income50-75k | 0.693 | 0.571 | 0.840 | 1.89e-04 | 2.64e-03 | 6515 | 4076 | 2439 |
| African | Full Cohort | Switching vs Continuation | Smoking_Frequency | Income75-100k | 0.655 | 0.502 | 0.854 | 1.75e-03 | 2.45e-02 | 6515 | 4076 | 2439 |
| African | Full Cohort | Switching vs Continuation | Smoking_Frequency | Education_levelCollege Postgraduate | 1.165 | 0.909 | 1.494 | 2.29e-01 | 1.00e+00 | 6515 | 4076 | 2439 |
| African | Full Cohort | Switching vs Continuation | Smoking_Frequency | Education_levelCollege Undergraduate | 1.050 | 0.878 | 1.255 | 5.94e-01 | 1.00e+00 | 6515 | 4076 | 2439 |
| African | Full Cohort | Switching vs Continuation | Smoking_Frequency | Education_levelTwelve or GED | 0.954 | 0.794 | 1.146 | 6.14e-01 | 1.00e+00 | 6515 | 4076 | 2439 |
| African | Full Cohort | Switching vs Continuation | Smoking_Frequency | Smoking_FrequencyEvery Day | 1.346 | 1.162 | 1.560 | 7.60e-05 | 1.06e-03 | 6515 | 4076 | 2439 |
| African | Full Cohort | Switching vs Continuation | Smoking_Frequency | Smoking_FrequencyNot At All (former) | 1.188 | 1.034 | 1.364 | 1.48e-02 | 2.07e-01 | 6515 | 4076 | 2439 |
| African | Full Cohort | Switching vs Continuation | Smoking_Frequency | Smoking_FrequencySome Days | 1.050 | 0.872 | 1.266 | 6.06e-01 | 1.00e+00 | 6515 | 4076 | 2439 |
| African | Full Cohort | Discontinuation vs Continuation | Stress | (Intercept) | 0.341 | 0.206 | 0.564 | 2.79e-05 | 3.91e-04 | 1745 | 1239 | 506 |
| African | Full Cohort | Discontinuation vs Continuation | Stress | age_at_first_AD | 1.146 | 1.015 | 1.294 | 2.79e-02 | 3.90e-01 | 1745 | 1239 | 506 |
| African | Full Cohort | Discontinuation vs Continuation | Stress | sex_at_birthMale | 1.170 | 0.909 | 1.505 | 2.22e-01 | 1.00e+00 | 1745 | 1239 | 506 |
| African | Full Cohort | Discontinuation vs Continuation | Stress | Income>200k | 0.990 | 0.458 | 2.140 | 9.79e-01 | 1.00e+00 | 1745 | 1239 | 506 |
| African | Full Cohort | Discontinuation vs Continuation | Stress | Income100-150k | 0.964 | 0.628 | 1.480 | 8.68e-01 | 1.00e+00 | 1745 | 1239 | 506 |
| African | Full Cohort | Discontinuation vs Continuation | Stress | Income150-200k | 1.861 | 0.979 | 3.538 | 5.83e-02 | 8.16e-01 | 1745 | 1239 | 506 |
| African | Full Cohort | Discontinuation vs Continuation | Stress | Income25-35k | 1.230 | 0.860 | 1.760 | 2.58e-01 | 1.00e+00 | 1745 | 1239 | 506 |
| African | Full Cohort | Discontinuation vs Continuation | Stress | Income35-50k | 1.089 | 0.773 | 1.535 | 6.25e-01 | 1.00e+00 | 1745 | 1239 | 506 |
| African | Full Cohort | Discontinuation vs Continuation | Stress | Income50-75k | 1.159 | 0.835 | 1.610 | 3.77e-01 | 1.00e+00 | 1745 | 1239 | 506 |
| African | Full Cohort | Discontinuation vs Continuation | Stress | Income75-100k | 0.821 | 0.521 | 1.293 | 3.95e-01 | 1.00e+00 | 1745 | 1239 | 506 |
| African | Full Cohort | Discontinuation vs Continuation | Stress | Education_levelCollege Postgraduate | 0.857 | 0.477 | 1.540 | 6.07e-01 | 1.00e+00 | 1745 | 1239 | 506 |
| African | Full Cohort | Discontinuation vs Continuation | Stress | Education_levelCollege Undergraduate | 0.902 | 0.538 | 1.511 | 6.95e-01 | 1.00e+00 | 1745 | 1239 | 506 |
| African | Full Cohort | Discontinuation vs Continuation | Stress | Education_levelTwelve or GED | 1.017 | 0.592 | 1.746 | 9.51e-01 | 1.00e+00 | 1745 | 1239 | 506 |
| African | Full Cohort | Discontinuation vs Continuation | Stress | StressAlmost Never | 1.660 | 1.011 | 2.724 | 4.50e-02 | 6.30e-01 | 1745 | 1239 | 506 |
| African | Full Cohort | Discontinuation vs Continuation | Stress | StressFairly Often | 1.060 | 0.656 | 1.714 | 8.11e-01 | 1.00e+00 | 1745 | 1239 | 506 |
| African | Full Cohort | Discontinuation vs Continuation | Stress | StressSometimes | 1.342 | 1.068 | 1.687 | 1.17e-02 | 1.63e-01 | 1745 | 1239 | 506 |
| African | Full Cohort | Discontinuation vs Continuation | Stress | StressVery Often | 1.413 | 0.836 | 2.387 | 1.96e-01 | 1.00e+00 | 1745 | 1239 | 506 |
| African | Full Cohort | Switching vs Continuation | Stress | (Intercept) | 0.810 | 0.536 | 1.223 | 3.16e-01 | 1.00e+00 | 1984 | 1239 | 745 |
| African | Full Cohort | Switching vs Continuation | Stress | age_at_first_AD | 0.636 | 0.567 | 0.712 | 6.94e-15 | 9.72e-14 | 1984 | 1239 | 745 |
| African | Full Cohort | Switching vs Continuation | Stress | sex_at_birthMale | 1.061 | 0.838 | 1.344 | 6.22e-01 | 1.00e+00 | 1984 | 1239 | 745 |
| African | Full Cohort | Switching vs Continuation | Stress | Income>200k | 0.339 | 0.143 | 0.803 | 1.39e-02 | 1.95e-01 | 1984 | 1239 | 745 |
| African | Full Cohort | Switching vs Continuation | Stress | Income100-150k | 0.651 | 0.439 | 0.964 | 3.23e-02 | 4.52e-01 | 1984 | 1239 | 745 |
| African | Full Cohort | Switching vs Continuation | Stress | Income150-200k | 0.651 | 0.321 | 1.320 | 2.34e-01 | 1.00e+00 | 1984 | 1239 | 745 |
| African | Full Cohort | Switching vs Continuation | Stress | Income25-35k | 0.816 | 0.588 | 1.133 | 2.25e-01 | 1.00e+00 | 1984 | 1239 | 745 |
| African | Full Cohort | Switching vs Continuation | Stress | Income35-50k | 0.818 | 0.603 | 1.111 | 1.98e-01 | 1.00e+00 | 1984 | 1239 | 745 |
| African | Full Cohort | Switching vs Continuation | Stress | Income50-75k | 0.763 | 0.563 | 1.033 | 8.03e-02 | 1.00e+00 | 1984 | 1239 | 745 |
| African | Full Cohort | Switching vs Continuation | Stress | Income75-100k | 0.704 | 0.477 | 1.037 | 7.56e-02 | 1.00e+00 | 1984 | 1239 | 745 |
| African | Full Cohort | Switching vs Continuation | Stress | Education_levelCollege Postgraduate | 0.763 | 0.467 | 1.245 | 2.79e-01 | 1.00e+00 | 1984 | 1239 | 745 |
| African | Full Cohort | Switching vs Continuation | Stress | Education_levelCollege Undergraduate | 0.656 | 0.429 | 1.004 | 5.23e-02 | 7.32e-01 | 1984 | 1239 | 745 |
| African | Full Cohort | Switching vs Continuation | Stress | Education_levelTwelve or GED | 0.643 | 0.409 | 1.009 | 5.47e-02 | 7.65e-01 | 1984 | 1239 | 745 |
| African | Full Cohort | Switching vs Continuation | Stress | StressAlmost Never | 2.041 | 1.327 | 3.138 | 1.16e-03 | 1.63e-02 | 1984 | 1239 | 745 |
| African | Full Cohort | Switching vs Continuation | Stress | StressFairly Often | 1.100 | 0.723 | 1.674 | 6.55e-01 | 1.00e+00 | 1984 | 1239 | 745 |
| African | Full Cohort | Switching vs Continuation | Stress | StressSometimes | 1.352 | 1.102 | 1.660 | 3.84e-03 | 5.37e-02 | 1984 | 1239 | 745 |
| African | Full Cohort | Switching vs Continuation | Stress | StressVery Often | 1.433 | 0.907 | 2.264 | 1.24e-01 | 1.00e+00 | 1984 | 1239 | 745 |
| African | Full Cohort | Discontinuation vs Continuation | age_at_first_AD | (Intercept) | 0.410 | 0.384 | 0.438 | <2.2e-16 | <2.2e-16 | 5972 | 4164 | 1808 |
| African | Full Cohort | Discontinuation vs Continuation | age_at_first_AD | age_at_first_AD | 0.984 | 0.924 | 1.048 | 6.20e-01 | 1.00e+00 | 5972 | 4164 | 1808 |
| African | Full Cohort | Discontinuation vs Continuation | age_at_first_AD | sex_at_birthMale | 1.223 | 1.081 | 1.382 | 1.34e-03 | 1.87e-02 | 5972 | 4164 | 1808 |

| | | | | | | | | | | | | |
|---|---|---|---|---|---|---|---|---|---|---|---|---|
| African | Full Cohort | Switching vs Continuation | age_at_first_AD | (Intercept) | 0.598 | 0.564 | 0.634 | <2.2e-16 | <2.2e-16 | 6693 | 4164 | 2529 |
| African | Full Cohort | Switching vs Continuation | age_at_first_AD | age_at_first_AD | 0.751 | 0.708 | 0.797 | <2.2e-16 | <2.2e-16 | 6693 | 4164 | 2529 |
| African | Full Cohort | Switching vs Continuation | age_at_first_AD | sex_at_birthMale | 0.937 | 0.834 | 1.052 | 2.71e-01 | 1.00e+00 | 6693 | 4164 | 2529 |
| African | Full Cohort | Discontinuation vs Continuation | sex_at_birth | (Intercept) | 0.410 | 0.384 | 0.438 | <2.2e-16 | <2.2e-16 | 5972 | 4164 | 1808 |
| African | Full Cohort | Discontinuation vs Continuation | sex_at_birth | age_at_first_AD | 0.984 | 0.924 | 1.048 | 6.20e-01 | 1.00e+00 | 5972 | 4164 | 1808 |
| African | Full Cohort | Discontinuation vs Continuation | sex_at_birth | sex_at_birthMale | 1.223 | 1.081 | 1.382 | 1.34e-03 | 1.87e-02 | 5972 | 4164 | 1808 |
| African | Full Cohort | Switching vs Continuation | sex_at_birth | (Intercept) | 0.598 | 0.564 | 0.634 | <2.2e-16 | <2.2e-16 | 6693 | 4164 | 2529 |
| African | Full Cohort | Switching vs Continuation | sex_at_birth | age_at_first_AD | 0.751 | 0.708 | 0.797 | <2.2e-16 | <2.2e-16 | 6693 | 4164 | 2529 |
| African | Full Cohort | Switching vs Continuation | sex_at_birth | sex_at_birthMale | 0.937 | 0.834 | 1.052 | 2.71e-01 | 1.00e+00 | 6693 | 4164 | 2529 |
| African | Recorded Depression | Discontinuation vs Continuation | Alcohol_Frequency | (Intercept) | 0.399 | 0.277 | 0.575 | 8.49e-07 | 1.19e-05 | 2295 | 1675 | 620 |
| African | Recorded Depression | Discontinuation vs Continuation | Alcohol_Frequency | age_at_first_AD | 0.909 | 0.818 | 1.011 | 7.77e-02 | 1.00e+00 | 2295 | 1675 | 620 |
| African | Recorded Depression | Discontinuation vs Continuation | Alcohol_Frequency | sex_at_birthMale | 1.210 | 0.973 | 1.505 | 8.73e-02 | 1.00e+00 | 2295 | 1675 | 620 |
| African | Recorded Depression | Discontinuation vs Continuation | Alcohol_Frequency | Income>200k | 0.608 | 0.194 | 1.905 | 3.93e-01 | 1.00e+00 | 2295 | 1675 | 620 |
| African | Recorded Depression | Discontinuation vs Continuation | Alcohol_Frequency | Income100-150k | 0.752 | 0.448 | 1.263 | 2.81e-01 | 1.00e+00 | 2295 | 1675 | 620 |
| African | Recorded Depression | Discontinuation vs Continuation | Alcohol_Frequency | Income150-200k | 0.629 | 0.229 | 1.728 | 3.68e-01 | 1.00e+00 | 2295 | 1675 | 620 |
| African | Recorded Depression | Discontinuation vs Continuation | Alcohol_Frequency | Income25-35k | 0.960 | 0.710 | 1.297 | 7.88e-01 | 1.00e+00 | 2295 | 1675 | 620 |
| African | Recorded Depression | Discontinuation vs Continuation | Alcohol_Frequency | Income35-50k | 0.745 | 0.536 | 1.036 | 7.98e-02 | 1.00e+00 | 2295 | 1675 | 620 |
| African | Recorded Depression | Discontinuation vs Continuation | Alcohol_Frequency | Income50-75k | 0.852 | 0.604 | 1.201 | 3.61e-01 | 1.00e+00 | 2295 | 1675 | 620 |
| African | Recorded Depression | Discontinuation vs Continuation | Alcohol_Frequency | Income75-100k | 0.510 | 0.294 | 0.886 | 1.68e-02 | 2.35e-01 | 2295 | 1675 | 620 |
| African | Recorded Depression | Discontinuation vs Continuation | Alcohol_Frequency | Education_levelCollege Postgraduate | 1.126 | 0.697 | 1.818 | 6.28e-01 | 1.00e+00 | 2295 | 1675 | 620 |
| African | Recorded Depression | Discontinuation vs Continuation | Alcohol_Frequency | Education_levelCollege Undergraduate | 1.089 | 0.773 | 1.534 | 6.25e-01 | 1.00e+00 | 2295 | 1675 | 620 |
| African | Recorded Depression | Discontinuation vs Continuation | Alcohol_Frequency | Education_levelTwelve or GED | 1.104 | 0.780 | 1.562 | 5.78e-01 | 1.00e+00 | 2295 | 1675 | 620 |
| African | Recorded Depression | Discontinuation vs Continuation | Alcohol_Frequency | Alcohol_Frequency2-3 Week | 0.818 | 0.544 | 1.229 | 3.32e-01 | 1.00e+00 | 2295 | 1675 | 620 |
| African | Recorded Depression | Discontinuation vs Continuation | Alcohol_Frequency | Alcohol_Frequency2-4 Month | 1.013 | 0.726 | 1.413 | 9.40e-01 | 1.00e+00 | 2295 | 1675 | 620 |
| African | Recorded Depression | Discontinuation vs Continuation | Alcohol_Frequency | Alcohol_Frequency4plus Week | 0.890 | 0.568 | 1.396 | 6.12e-01 | 1.00e+00 | 2295 | 1675 | 620 |
| African | Recorded Depression | Discontinuation vs Continuation | Alcohol_Frequency | Alcohol_FrequencyMonthly or Less | 0.768 | 0.569 | 1.036 | 8.36e-02 | 1.00e+00 | 2295 | 1675 | 620 |
| African | Recorded Depression | Discontinuation vs Continuation | Alcohol_Frequency | Alcohol_FrequencyNever (former) | 0.935 | 0.666 | 1.312 | 6.96e-01 | 1.00e+00 | 2295 | 1675 | 620 |
| African | Recorded Depression | Switching vs Continuation | Alcohol_Frequency | (Intercept) | 1.214 | 0.929 | 1.588 | 1.56e-01 | 1.00e+00 | 3511 | 1675 | 1836 |
| African | Recorded Depression | Switching vs Continuation | Alcohol_Frequency | age_at_first_AD | 0.823 | 0.758 | 0.893 | 3.17e-06 | 4.43e-05 | 3511 | 1675 | 1836 |
| African | Recorded Depression | Switching vs Continuation | Alcohol_Frequency | sex_at_birthMale | 1.003 | 0.852 | 1.181 | 9.71e-01 | 1.00e+00 | 3511 | 1675 | 1836 |
| African | Recorded Depression | Switching vs Continuation | Alcohol_Frequency | Income>200k | 0.428 | 0.184 | 0.992 | 4.78e-02 | 6.70e-01 | 3511 | 1675 | 1836 |
| African | Recorded Depression | Switching vs Continuation | Alcohol_Frequency | Income100-150k | 0.588 | 0.402 | 0.861 | 6.31e-03 | 8.83e-02 | 3511 | 1675 | 1836 |
| African | Recorded Depression | Switching vs Continuation | Alcohol_Frequency | Income150-200k | 0.560 | 0.279 | 1.124 | 1.03e-01 | 1.00e+00 | 3511 | 1675 | 1836 |
| African | Recorded Depression | Switching vs Continuation | Alcohol_Frequency | Income25-35k | 0.790 | 0.632 | 0.988 | 3.85e-02 | 5.38e-01 | 3511 | 1675 | 1836 |
| African | Recorded Depression | Switching vs Continuation | Alcohol_Frequency | Income35-50k | 0.817 | 0.653 | 1.022 | 7.69e-02 | 1.00e+00 | 3511 | 1675 | 1836 |
| African | Recorded Depression | Switching vs Continuation | Alcohol_Frequency | Income50-75k | 0.698 | 0.543 | 0.897 | 4.91e-03 | 6.87e-02 | 3511 | 1675 | 1836 |
| African | Recorded Depression | Switching vs Continuation | Alcohol_Frequency | Income75-100k | 0.655 | 0.464 | 0.926 | 1.66e-02 | 2.32e-01 | 3511 | 1675 | 1836 |
| African | Recorded Depression | Switching vs Continuation | Alcohol_Frequency | Education_levelCollege Postgraduate | 1.192 | 0.853 | 1.665 | 3.03e-01 | 1.00e+00 | 3511 | 1675 | 1836 |
| African | Recorded Depression | Switching vs Continuation | Alcohol_Frequency | Education_levelCollege Undergraduate | 1.052 | 0.824 | 1.344 | 6.82e-01 | 1.00e+00 | 3511 | 1675 | 1836 |
| African | Recorded Depression | Switching vs Continuation | Alcohol_Frequency | Education_levelTwelve or GED | 0.915 | 0.712 | 1.177 | 4.90e-01 | 1.00e+00 | 3511 | 1675 | 1836 |
| African | Recorded Depression | Switching vs Continuation | Alcohol_Frequency | Alcohol_Frequency2-3 Week | 0.897 | 0.667 | 1.207 | 4.73e-01 | 1.00e+00 | 3511 | 1675 | 1836 |
| African | Recorded Depression | Switching vs Continuation | Alcohol_Frequency | Alcohol_Frequency2-4 Month | 0.954 | 0.744 | 1.224 | 7.14e-01 | 1.00e+00 | 3511 | 1675 | 1836 |
| African | Recorded Depression | Switching vs Continuation | Alcohol_Frequency | Alcohol_Frequency4plus Week | 0.946 | 0.676 | 1.324 | 7.46e-01 | 1.00e+00 | 3511 | 1675 | 1836 |
| African | Recorded Depression | Switching vs Continuation | Alcohol_Frequency | Alcohol_FrequencyMonthly or Less | 0.885 | 0.710 | 1.103 | 2.75e-01 | 1.00e+00 | 3511 | 1675 | 1836 |
| African | Recorded Depression | Switching vs Continuation | Alcohol_Frequency | Alcohol_FrequencyNever (former) | 1.217 | 0.950 | 1.559 | 1.21e-01 | 1.00e+00 | 3511 | 1675 | 1836 |
| African | Recorded Depression | Discontinuation vs Continuation | BMI | (Intercept) | 0.352 | 0.258 | 0.481 | 5.19e-11 | 7.26e-10 | 2329 | 1708 | 621 |
| African | Recorded Depression | Discontinuation vs Continuation | BMI | age_at_first_AD | 0.903 | 0.813 | 1.003 | 5.57e-02 | 7.80e-01 | 2329 | 1708 | 621 |
| African | Recorded Depression | Discontinuation vs Continuation | BMI | sex_at_birthMale | 1.201 | 0.965 | 1.493 | 1.01e-01 | 1.00e+00 | 2329 | 1708 | 621 |
| African | Recorded Depression | Discontinuation vs Continuation | BMI | Income>200k | 0.618 | 0.196 | 1.946 | 4.11e-01 | 1.00e+00 | 2329 | 1708 | 621 |
| African | Recorded Depression | Discontinuation vs Continuation | BMI | Income100-150k | 0.759 | 0.452 | 1.274 | 2.96e-01 | 1.00e+00 | 2329 | 1708 | 621 |
| African | Recorded Depression | Discontinuation vs Continuation | BMI | Income150-200k | 0.727 | 0.283 | 1.868 | 5.08e-01 | 1.00e+00 | 2329 | 1708 | 621 |
| African | Recorded Depression | Discontinuation vs Continuation | BMI | Income25-35k | 0.938 | 0.693 | 1.269 | 6.77e-01 | 1.00e+00 | 2329 | 1708 | 621 |
| African | Recorded Depression | Discontinuation vs Continuation | BMI | Income35-50k | 0.726 | 0.523 | 1.008 | 5.57e-02 | 7.79e-01 | 2329 | 1708 | 621 |
| African | Recorded Depression | Discontinuation vs Continuation | BMI | Income50-75k | 0.803 | 0.571 | 1.131 | 2.10e-01 | 1.00e+00 | 2329 | 1708 | 621 |
| African | Recorded Depression | Discontinuation vs Continuation | BMI | Income75-100k | 0.459 | 0.257 | 0.819 | 8.36e-03 | 1.17e-01 | 2329 | 1708 | 621 |
| African | Recorded Depression | Discontinuation vs Continuation | BMI | Education_levelCollege Postgraduate | 1.167 | 0.726 | 1.876 | 5.23e-01 | 1.00e+00 | 2329 | 1708 | 621 |
| African | Recorded Depression | Discontinuation vs Continuation | BMI | Education_levelCollege Undergraduate | 1.095 | 0.782 | 1.535 | 5.96e-01 | 1.00e+00 | 2329 | 1708 | 621 |
| African | Recorded Depression | Discontinuation vs Continuation | BMI | Education_levelTwelve or GED | 1.098 | 0.778 | 1.549 | 5.95e-01 | 1.00e+00 | 2329 | 1708 | 621 |
| African | Recorded Depression | Discontinuation vs Continuation | BMI | BMI | 0.960 | 0.885 | 1.041 | 3.25e-01 | 1.00e+00 | 2329 | 1708 | 621 |
| African | Recorded Depression | Switching vs Continuation | BMI | (Intercept) | 1.179 | 0.947 | 1.468 | 1.41e-01 | 1.00e+00 | 3567 | 1708 | 1859 |
| African | Recorded Depression | Switching vs Continuation | BMI | age_at_first_AD | 0.836 | 0.771 | 0.906 | 1.23e-05 | 1.73e-04 | 3567 | 1708 | 1859 |
| African | Recorded Depression | Switching vs Continuation | BMI | sex_at_birthMale | 1.037 | 0.883 | 1.219 | 6.54e-01 | 1.00e+00 | 3567 | 1708 | 1859 |
| African | Recorded Depression | Switching vs Continuation | BMI | Income>200k | 0.449 | 0.192 | 1.050 | 6.48e-02 | 9.07e-01 | 3567 | 1708 | 1859 |
| African | Recorded Depression | Switching vs Continuation | BMI | Income100-150k | 0.580 | 0.395 | 0.850 | 5.20e-03 | 7.27e-02 | 3567 | 1708 | 1859 |
| African | Recorded Depression | Switching vs Continuation | BMI | Income150-200k | 0.661 | 0.338 | 1.295 | 2.28e-01 | 1.00e+00 | 3567 | 1708 | 1859 |
| African | Recorded Depression | Switching vs Continuation | BMI | Income25-35k | 0.821 | 0.658 | 1.024 | 8.01e-02 | 1.00e+00 | 3567 | 1708 | 1859 |

| | | | | | | | | | | | | |
|---|---|---|---|---|---|---|---|---|---|---|---|---|
| African | Recorded Depression | Switching vs Continuation | BMI | Income35-50k | 0.821 | 0.657 | 1.026 | 8.31e-02 | 1.00e+00 | 3567 | 1708 | 1859 |
| African | Recorded Depression | Switching vs Continuation | BMI | Income50-75k | 0.684 | 0.534 | 0.876 | 2.58e-03 | 3.62e-02 | 3567 | 1708 | 1859 |
| African | Recorded Depression | Switching vs Continuation | BMI | Income75-100k | 0.646 | 0.457 | 0.913 | 1.33e-02 | 1.87e-01 | 3567 | 1708 | 1859 |
| African | Recorded Depression | Switching vs Continuation | BMI | Education_levelCollege Postgraduate | 1.192 | 0.858 | 1.656 | 2.95e-01 | 1.00e+00 | 3567 | 1708 | 1859 |
| African | Recorded Depression | Switching vs Continuation | BMI | Education_levelCollege Undergraduate | 1.033 | 0.814 | 1.312 | 7.88e-01 | 1.00e+00 | 3567 | 1708 | 1859 |
| African | Recorded Depression | Switching vs Continuation | BMI | Education_levelTwelve or GED | 0.898 | 0.702 | 1.149 | 3.93e-01 | 1.00e+00 | 3567 | 1708 | 1859 |
| African | Recorded Depression | Switching vs Continuation | BMI | BMI | 1.000 | 0.944 | 1.059 | 9.98e-01 | 1.00e+00 | 3567 | 1708 | 1859 |
| African | Recorded Depression | Discontinuation vs Continuation | Education_level | (Intercept) | 0.360 | 0.265 | 0.488 | 4.58e-11 | 6.41e-10 | 2367 | 1728 | 639 |
| African | Recorded Depression | Discontinuation vs Continuation | Education_level | age_at_first_AD | 0.900 | 0.812 | 0.998 | 4.66e-02 | 6.52e-01 | 2367 | 1728 | 639 |
| African | Recorded Depression | Discontinuation vs Continuation | Education_level | sex_at_birthMale | 1.247 | 1.010 | 1.540 | 4.03e-02 | 5.64e-01 | 2367 | 1728 | 639 |
| African | Recorded Depression | Discontinuation vs Continuation | Education_level | Income>200k | 0.613 | 0.196 | 1.914 | 3.99e-01 | 1.00e+00 | 2367 | 1728 | 639 |
| African | Recorded Depression | Discontinuation vs Continuation | Education_level | Income100-150k | 0.753 | 0.449 | 1.262 | 2.81e-01 | 1.00e+00 | 2367 | 1728 | 639 |
| African | Recorded Depression | Discontinuation vs Continuation | Education_level | Income150-200k | 0.751 | 0.293 | 1.928 | 5.52e-01 | 1.00e+00 | 2367 | 1728 | 639 |
| African | Recorded Depression | Discontinuation vs Continuation | Education_level | Income25-35k | 0.946 | 0.701 | 1.275 | 7.15e-01 | 1.00e+00 | 2367 | 1728 | 639 |
| African | Recorded Depression | Discontinuation vs Continuation | Education_level | Income35-50k | 0.758 | 0.550 | 1.046 | 9.14e-02 | 1.00e+00 | 2367 | 1728 | 639 |
| African | Recorded Depression | Discontinuation vs Continuation | Education_level | Income50-75k | 0.846 | 0.605 | 1.184 | 3.30e-01 | 1.00e+00 | 2367 | 1728 | 639 |
| African | Recorded Depression | Discontinuation vs Continuation | Education_level | Income75-100k | 0.517 | 0.298 | 0.897 | 1.89e-02 | 2.65e-01 | 2367 | 1728 | 639 |
| African | Recorded Depression | Discontinuation vs Continuation | Education_level | Education_levelCollege Postgraduate | 1.080 | 0.675 | 1.728 | 7.47e-01 | 1.00e+00 | 2367 | 1728 | 639 |
| African | Recorded Depression | Discontinuation vs Continuation | Education_level | Education_levelCollege Undergraduate | 1.039 | 0.746 | 1.447 | 8.21e-01 | 1.00e+00 | 2367 | 1728 | 639 |
| African | Recorded Depression | Discontinuation vs Continuation | Education_level | Education_levelTwelve or GED | 1.069 | 0.762 | 1.499 | 7.01e-01 | 1.00e+00 | 2367 | 1728 | 639 |
| African | Recorded Depression | Switching vs Continuation | Education_level | (Intercept) | 1.173 | 0.943 | 1.458 | 1.52e-01 | 1.00e+00 | 3605 | 1728 | 1877 |
| African | Recorded Depression | Switching vs Continuation | Education_level | age_at_first_AD | 0.834 | 0.771 | 0.904 | 8.14e-06 | 1.14e-04 | 3605 | 1728 | 1877 |
| African | Recorded Depression | Switching vs Continuation | Education_level | sex_at_birthMale | 1.036 | 0.885 | 1.213 | 6.56e-01 | 1.00e+00 | 3605 | 1728 | 1877 |
| African | Recorded Depression | Switching vs Continuation | Education_level | Income>200k | 0.420 | 0.181 | 0.972 | 4.27e-02 | 5.97e-01 | 3605 | 1728 | 1877 |
| African | Recorded Depression | Switching vs Continuation | Education_level | Income100-150k | 0.581 | 0.398 | 0.849 | 5.05e-03 | 7.08e-02 | 3605 | 1728 | 1877 |
| African | Recorded Depression | Switching vs Continuation | Education_level | Income150-200k | 0.661 | 0.338 | 1.293 | 2.27e-01 | 1.00e+00 | 3605 | 1728 | 1877 |
| African | Recorded Depression | Switching vs Continuation | Education_level | Income25-35k | 0.811 | 0.650 | 1.010 | 6.16e-02 | 8.62e-01 | 3605 | 1728 | 1877 |
| African | Recorded Depression | Switching vs Continuation | Education_level | Income35-50k | 0.823 | 0.660 | 1.026 | 8.29e-02 | 1.00e+00 | 3605 | 1728 | 1877 |
| African | Recorded Depression | Switching vs Continuation | Education_level | Income50-75k | 0.680 | 0.531 | 0.870 | 2.14e-03 | 3.00e-02 | 3605 | 1728 | 1877 |
| African | Recorded Depression | Switching vs Continuation | Education_level | Income75-100k | 0.645 | 0.458 | 0.910 | 1.26e-02 | 1.76e-01 | 3605 | 1728 | 1877 |
| African | Recorded Depression | Switching vs Continuation | Education_level | Education_levelCollege Postgraduate | 1.199 | 0.864 | 1.665 | 2.78e-01 | 1.00e+00 | 3605 | 1728 | 1877 |
| African | Recorded Depression | Switching vs Continuation | Education_level | Education_levelCollege Undergraduate | 1.040 | 0.820 | 1.318 | 7.49e-01 | 1.00e+00 | 3605 | 1728 | 1877 |
| African | Recorded Depression | Switching vs Continuation | Education_level | Education_levelTwelve or GED | 0.903 | 0.706 | 1.154 | 4.14e-01 | 1.00e+00 | 3605 | 1728 | 1877 |
| African | Recorded Depression | Discontinuation vs Continuation | Income | (Intercept) | 0.360 | 0.265 | 0.488 | 4.58e-11 | 6.41e-10 | 2367 | 1728 | 639 |
| African | Recorded Depression | Discontinuation vs Continuation | Income | age_at_first_AD | 0.900 | 0.812 | 0.998 | 4.66e-02 | 6.52e-01 | 2367 | 1728 | 639 |
| African | Recorded Depression | Discontinuation vs Continuation | Income | sex_at_birthMale | 1.247 | 1.010 | 1.540 | 4.03e-02 | 5.64e-01 | 2367 | 1728 | 639 |
| African | Recorded Depression | Discontinuation vs Continuation | Income | Income>200k | 0.613 | 0.196 | 1.914 | 3.99e-01 | 1.00e+00 | 2367 | 1728 | 639 |
| African | Recorded Depression | Discontinuation vs Continuation | Income | Income100-150k | 0.753 | 0.449 | 1.262 | 2.81e-01 | 1.00e+00 | 2367 | 1728 | 639 |
| African | Recorded Depression | Discontinuation vs Continuation | Income | Income150-200k | 0.751 | 0.293 | 1.928 | 5.52e-01 | 1.00e+00 | 2367 | 1728 | 639 |
| African | Recorded Depression | Discontinuation vs Continuation | Income | Income25-35k | 0.946 | 0.701 | 1.275 | 7.15e-01 | 1.00e+00 | 2367 | 1728 | 639 |
| African | Recorded Depression | Discontinuation vs Continuation | Income | Income35-50k | 0.758 | 0.550 | 1.046 | 9.14e-02 | 1.00e+00 | 2367 | 1728 | 639 |
| African | Recorded Depression | Discontinuation vs Continuation | Income | Income50-75k | 0.846 | 0.605 | 1.184 | 3.30e-01 | 1.00e+00 | 2367 | 1728 | 639 |
| African | Recorded Depression | Discontinuation vs Continuation | Income | Income75-100k | 0.517 | 0.298 | 0.897 | 1.89e-02 | 2.65e-01 | 2367 | 1728 | 639 |
| African | Recorded Depression | Discontinuation vs Continuation | Income | Education_levelCollege Postgraduate | 1.080 | 0.675 | 1.728 | 7.47e-01 | 1.00e+00 | 2367 | 1728 | 639 |
| African | Recorded Depression | Discontinuation vs Continuation | Income | Education_levelCollege Undergraduate | 1.039 | 0.746 | 1.447 | 8.21e-01 | 1.00e+00 | 2367 | 1728 | 639 |
| African | Recorded Depression | Discontinuation vs Continuation | Income | Education_levelTwelve or GED | 1.069 | 0.762 | 1.499 | 7.01e-01 | 1.00e+00 | 2367 | 1728 | 639 |
| African | Recorded Depression | Switching vs Continuation | Income | (Intercept) | 1.173 | 0.943 | 1.458 | 1.52e-01 | 1.00e+00 | 3605 | 1728 | 1877 |
| African | Recorded Depression | Switching vs Continuation | Income | age_at_first_AD | 0.834 | 0.771 | 0.904 | 8.14e-06 | 1.14e-04 | 3605 | 1728 | 1877 |
| African | Recorded Depression | Switching vs Continuation | Income | sex_at_birthMale | 1.036 | 0.885 | 1.213 | 6.56e-01 | 1.00e+00 | 3605 | 1728 | 1877 |
| African | Recorded Depression | Switching vs Continuation | Income | Income>200k | 0.420 | 0.181 | 0.972 | 4.27e-02 | 5.97e-01 | 3605 | 1728 | 1877 |
| African | Recorded Depression | Switching vs Continuation | Income | Income100-150k | 0.581 | 0.398 | 0.849 | 5.05e-03 | 7.08e-02 | 3605 | 1728 | 1877 |
| African | Recorded Depression | Switching vs Continuation | Income | Income150-200k | 0.661 | 0.338 | 1.293 | 2.27e-01 | 1.00e+00 | 3605 | 1728 | 1877 |
| African | Recorded Depression | Switching vs Continuation | Income | Income25-35k | 0.811 | 0.650 | 1.010 | 6.16e-02 | 8.62e-01 | 3605 | 1728 | 1877 |
| African | Recorded Depression | Switching vs Continuation | Income | Income35-50k | 0.823 | 0.660 | 1.026 | 8.29e-02 | 1.00e+00 | 3605 | 1728 | 1877 |
| African | Recorded Depression | Switching vs Continuation | Income | Income50-75k | 0.680 | 0.531 | 0.870 | 2.14e-03 | 3.00e-02 | 3605 | 1728 | 1877 |
| African | Recorded Depression | Switching vs Continuation | Income | Income75-100k | 0.645 | 0.458 | 0.910 | 1.26e-02 | 1.76e-01 | 3605 | 1728 | 1877 |
| African | Recorded Depression | Switching vs Continuation | Income | Education_levelCollege Postgraduate | 1.199 | 0.864 | 1.665 | 2.78e-01 | 1.00e+00 | 3605 | 1728 | 1877 |
| African | Recorded Depression | Switching vs Continuation | Income | Education_levelCollege Undergraduate | 1.040 | 0.820 | 1.318 | 7.49e-01 | 1.00e+00 | 3605 | 1728 | 1877 |
| African | Recorded Depression | Switching vs Continuation | Income | Education_levelTwelve or GED | 0.903 | 0.706 | 1.154 | 4.14e-01 | 1.00e+00 | 3605 | 1728 | 1877 |
| African | Recorded Depression | Discontinuation vs Continuation | Insomnia | (Intercept) | 0.367 | 0.270 | 0.499 | 1.81e-10 | 2.53e-09 | 2367 | 1728 | 639 |
| African | Recorded Depression | Discontinuation vs Continuation | Insomnia | age_at_first_AD | 0.903 | 0.814 | 1.002 | 5.39e-02 | 7.55e-01 | 2367 | 1728 | 639 |
| African | Recorded Depression | Discontinuation vs Continuation | Insomnia | sex_at_birthMale | 1.246 | 1.009 | 1.538 | 4.13e-02 | 5.78e-01 | 2367 | 1728 | 639 |
| African | Recorded Depression | Discontinuation vs Continuation | Insomnia | Income>200k | 0.608 | 0.195 | 1.899 | 3.92e-01 | 1.00e+00 | 2367 | 1728 | 639 |
| African | Recorded Depression | Discontinuation vs Continuation | Insomnia | Income100-150k | 0.751 | 0.448 | 1.259 | 2.77e-01 | 1.00e+00 | 2367 | 1728 | 639 |
| African | Recorded Depression | Discontinuation vs Continuation | Insomnia | Income150-200k | 0.756 | 0.295 | 1.939 | 5.60e-01 | 1.00e+00 | 2367 | 1728 | 639 |
| African | Recorded Depression | Discontinuation vs Continuation | Insomnia | Income25-35k | 0.946 | 0.701 | 1.276 | 7.15e-01 | 1.00e+00 | 2367 | 1728 | 639 |

| | | | | | | | | | | | | |
|---|---|---|---|---|---|---|---|---|---|---|---|---|
| African | Recorded Depression | Discontinuation vs Continuation | Insomnia | Income35-50k | 0.760 | 0.551 | 1.049 | 9.52e-02 | 1.00e+00 | 2367 | 1728 | 639 |
| African | Recorded Depression | Discontinuation vs Continuation | Insomnia | Income50-75k | 0.847 | 0.605 | 1.184 | 3.31e-01 | 1.00e+00 | 2367 | 1728 | 639 |
| African | Recorded Depression | Discontinuation vs Continuation | Insomnia | Income75-100k | 0.521 | 0.300 | 0.904 | 2.03e-02 | 2.84e-01 | 2367 | 1728 | 639 |
| African | Recorded Depression | Discontinuation vs Continuation | Insomnia | Education_levelCollege Postgraduate | 1.081 | 0.676 | 1.730 | 7.44e-01 | 1.00e+00 | 2367 | 1728 | 639 |
| African | Recorded Depression | Discontinuation vs Continuation | Insomnia | Education_levelCollege Undergraduate | 1.042 | 0.748 | 1.452 | 8.06e-01 | 1.00e+00 | 2367 | 1728 | 639 |
| African | Recorded Depression | Discontinuation vs Continuation | Insomnia | Education_levelTwelve or GED | 1.070 | 0.763 | 1.501 | 6.94e-01 | 1.00e+00 | 2367 | 1728 | 639 |
| African | Recorded Depression | Discontinuation vs Continuation | Insomnia | Insomnia | 0.922 | 0.748 | 1.136 | 4.45e-01 | 1.00e+00 | 2367 | 1728 | 639 |
| African | Recorded Depression | Switching vs Continuation | Insomnia | (Intercept) | 0.865 | 0.689 | 1.085 | 2.09e-01 | 1.00e+00 | 3605 | 1728 | 1877 |
| African | Recorded Depression | Switching vs Continuation | Insomnia | age_at_first_AD | 0.811 | 0.747 | 0.879 | 4.01e-07 | 5.61e-06 | 3605 | 1728 | 1877 |
| African | Recorded Depression | Switching vs Continuation | Insomnia | sex_at_birthMale | 1.027 | 0.875 | 1.207 | 7.43e-01 | 1.00e+00 | 3605 | 1728 | 1877 |
| African | Recorded Depression | Switching vs Continuation | Insomnia | Income>200k | 0.429 | 0.182 | 1.007 | 5.20e-02 | 7.28e-01 | 3605 | 1728 | 1877 |
| African | Recorded Depression | Switching vs Continuation | Insomnia | Income100-150k | 0.586 | 0.398 | 0.863 | 6.84e-03 | 9.58e-02 | 3605 | 1728 | 1877 |
| African | Recorded Depression | Switching vs Continuation | Insomnia | Income150-200k | 0.616 | 0.310 | 1.226 | 1.67e-01 | 1.00e+00 | 3605 | 1728 | 1877 |
| African | Recorded Depression | Switching vs Continuation | Insomnia | Income25-35k | 0.812 | 0.649 | 1.017 | 6.99e-02 | 9.78e-01 | 3605 | 1728 | 1877 |
| African | Recorded Depression | Switching vs Continuation | Insomnia | Income35-50k | 0.812 | 0.648 | 1.017 | 7.01e-02 | 9.82e-01 | 3605 | 1728 | 1877 |
| African | Recorded Depression | Switching vs Continuation | Insomnia | Income50-75k | 0.687 | 0.534 | 0.883 | 3.36e-03 | 4.71e-02 | 3605 | 1728 | 1877 |
| African | Recorded Depression | Switching vs Continuation | Insomnia | Income75-100k | 0.615 | 0.433 | 0.875 | 6.87e-03 | 9.62e-02 | 3605 | 1728 | 1877 |
| African | Recorded Depression | Switching vs Continuation | Insomnia | Education_levelCollege Postgraduate | 1.198 | 0.857 | 1.674 | 2.91e-01 | 1.00e+00 | 3605 | 1728 | 1877 |
| African | Recorded Depression | Switching vs Continuation | Insomnia | Education_levelCollege Undergraduate | 1.029 | 0.808 | 1.312 | 8.14e-01 | 1.00e+00 | 3605 | 1728 | 1877 |
| African | Recorded Depression | Switching vs Continuation | Insomnia | Education_levelTwelve or GED | 0.910 | 0.708 | 1.169 | 4.60e-01 | 1.00e+00 | 3605 | 1728 | 1877 |
| African | Recorded Depression | Switching vs Continuation | Insomnia | Insomnia | 2.327 | 2.021 | 2.678 | <2.2e-16 | <2.2e-16 | 3605 | 1728 | 1877 |
| African | Recorded Depression | Discontinuation vs Continuation | Migraine | (Intercept) | 0.373 | 0.274 | 0.509 | 4.72e-10 | 6.60e-09 | 2367 | 1728 | 639 |
| African | Recorded Depression | Discontinuation vs Continuation | Migraine | age_at_first_AD | 0.895 | 0.806 | 0.993 | 3.57e-02 | 4.99e-01 | 2367 | 1728 | 639 |
| African | Recorded Depression | Discontinuation vs Continuation | Migraine | sex_at_birthMale | 1.224 | 0.989 | 1.515 | 6.38e-02 | 8.93e-01 | 2367 | 1728 | 639 |
| African | Recorded Depression | Discontinuation vs Continuation | Migraine | Income>200k | 0.606 | 0.194 | 1.892 | 3.88e-01 | 1.00e+00 | 2367 | 1728 | 639 |
| African | Recorded Depression | Discontinuation vs Continuation | Migraine | Income100-150k | 0.750 | 0.447 | 1.257 | 2.75e-01 | 1.00e+00 | 2367 | 1728 | 639 |
| African | Recorded Depression | Discontinuation vs Continuation | Migraine | Income150-200k | 0.744 | 0.290 | 1.910 | 5.39e-01 | 1.00e+00 | 2367 | 1728 | 639 |
| African | Recorded Depression | Discontinuation vs Continuation | Migraine | Income25-35k | 0.947 | 0.702 | 1.276 | 7.19e-01 | 1.00e+00 | 2367 | 1728 | 639 |
| African | Recorded Depression | Discontinuation vs Continuation | Migraine | Income35-50k | 0.765 | 0.554 | 1.055 | 1.03e-01 | 1.00e+00 | 2367 | 1728 | 639 |
| African | Recorded Depression | Discontinuation vs Continuation | Migraine | Income50-75k | 0.852 | 0.609 | 1.192 | 3.51e-01 | 1.00e+00 | 2367 | 1728 | 639 |
| African | Recorded Depression | Discontinuation vs Continuation | Migraine | Income75-100k | 0.522 | 0.301 | 0.905 | 2.06e-02 | 2.89e-01 | 2367 | 1728 | 639 |
| African | Recorded Depression | Discontinuation vs Continuation | Migraine | Education_levelCollege Postgraduate | 1.081 | 0.676 | 1.730 | 7.45e-01 | 1.00e+00 | 2367 | 1728 | 639 |
| African | Recorded Depression | Discontinuation vs Continuation | Migraine | Education_levelCollege Undergraduate | 1.042 | 0.748 | 1.451 | 8.09e-01 | 1.00e+00 | 2367 | 1728 | 639 |
| African | Recorded Depression | Discontinuation vs Continuation | Migraine | Education_levelTwelve or GED | 1.071 | 0.764 | 1.502 | 6.91e-01 | 1.00e+00 | 2367 | 1728 | 639 |
| African | Recorded Depression | Discontinuation vs Continuation | Migraine | Migraine | 0.886 | 0.721 | 1.087 | 2.45e-01 | 1.00e+00 | 2367 | 1728 | 639 |
| African | Recorded Depression | Switching vs Continuation | Migraine | (Intercept) | 0.978 | 0.782 | 1.223 | 8.44e-01 | 1.00e+00 | 3605 | 1728 | 1877 |
| African | Recorded Depression | Switching vs Continuation | Migraine | age_at_first_AD | 0.866 | 0.798 | 0.938 | 4.41e-04 | 6.18e-03 | 3605 | 1728 | 1877 |
| African | Recorded Depression | Switching vs Continuation | Migraine | sex_at_birthMale | 1.126 | 0.960 | 1.322 | 1.46e-01 | 1.00e+00 | 3605 | 1728 | 1877 |
| African | Recorded Depression | Switching vs Continuation | Migraine | Income>200k | 0.418 | 0.180 | 0.973 | 4.31e-02 | 6.03e-01 | 3605 | 1728 | 1877 |
| African | Recorded Depression | Switching vs Continuation | Migraine | Income100-150k | 0.580 | 0.395 | 0.850 | 5.20e-03 | 7.27e-02 | 3605 | 1728 | 1877 |
| African | Recorded Depression | Switching vs Continuation | Migraine | Income150-200k | 0.704 | 0.358 | 1.386 | 3.10e-01 | 1.00e+00 | 3605 | 1728 | 1877 |
| African | Recorded Depression | Switching vs Continuation | Migraine | Income25-35k | 0.809 | 0.648 | 1.010 | 6.09e-02 | 8.53e-01 | 3605 | 1728 | 1877 |
| African | Recorded Depression | Switching vs Continuation | Migraine | Income35-50k | 0.778 | 0.622 | 0.973 | 2.76e-02 | 3.86e-01 | 3605 | 1728 | 1877 |
| African | Recorded Depression | Switching vs Continuation | Migraine | Income50-75k | 0.678 | 0.529 | 0.869 | 2.19e-03 | 3.07e-02 | 3605 | 1728 | 1877 |
| African | Recorded Depression | Switching vs Continuation | Migraine | Income75-100k | 0.618 | 0.437 | 0.873 | 6.43e-03 | 9.00e-02 | 3605 | 1728 | 1877 |
| African | Recorded Depression | Switching vs Continuation | Migraine | Education_levelCollege Postgraduate | 1.183 | 0.850 | 1.646 | 3.20e-01 | 1.00e+00 | 3605 | 1728 | 1877 |
| African | Recorded Depression | Switching vs Continuation | Migraine | Education_levelCollege Undergraduate | 0.999 | 0.787 | 1.269 | 9.95e-01 | 1.00e+00 | 3605 | 1728 | 1877 |
| African | Recorded Depression | Switching vs Continuation | Migraine | Education_levelTwelve or GED | 0.872 | 0.680 | 1.117 | 2.77e-01 | 1.00e+00 | 3605 | 1728 | 1877 |
| African | Recorded Depression | Switching vs Continuation | Migraine | Migraine | 1.740 | 1.512 | 2.003 | 1.03e-14 | 1.44e-13 | 3605 | 1728 | 1877 |
| African | Recorded Depression | Discontinuation vs Continuation | Recorded Anxiety | (Intercept) | 0.372 | 0.269 | 0.515 | 2.63e-09 | 3.68e-08 | 2367 | 1728 | 639 |
| African | Recorded Depression | Discontinuation vs Continuation | Recorded Anxiety | age_at_first_AD | 0.898 | 0.810 | 0.996 | 4.22e-02 | 5.91e-01 | 2367 | 1728 | 639 |
| African | Recorded Depression | Discontinuation vs Continuation | Recorded Anxiety | sex_at_birthMale | 1.244 | 1.007 | 1.537 | 4.26e-02 | 5.97e-01 | 2367 | 1728 | 639 |
| African | Recorded Depression | Discontinuation vs Continuation | Recorded Anxiety | Income>200k | 0.605 | 0.194 | 1.892 | 3.88e-01 | 1.00e+00 | 2367 | 1728 | 639 |
| African | Recorded Depression | Discontinuation vs Continuation | Recorded Anxiety | Income100-150k | 0.752 | 0.449 | 1.261 | 2.80e-01 | 1.00e+00 | 2367 | 1728 | 639 |
| African | Recorded Depression | Discontinuation vs Continuation | Recorded Anxiety | Income150-200k | 0.756 | 0.294 | 1.941 | 5.61e-01 | 1.00e+00 | 2367 | 1728 | 639 |
| African | Recorded Depression | Discontinuation vs Continuation | Recorded Anxiety | Income25-35k | 0.946 | 0.702 | 1.276 | 7.16e-01 | 1.00e+00 | 2367 | 1728 | 639 |
| African | Recorded Depression | Discontinuation vs Continuation | Recorded Anxiety | Income35-50k | 0.760 | 0.551 | 1.048 | 9.43e-02 | 1.00e+00 | 2367 | 1728 | 639 |
| African | Recorded Depression | Discontinuation vs Continuation | Recorded Anxiety | Income50-75k | 0.850 | 0.608 | 1.190 | 3.45e-01 | 1.00e+00 | 2367 | 1728 | 639 |
| African | Recorded Depression | Discontinuation vs Continuation | Recorded Anxiety | Income75-100k | 0.520 | 0.300 | 0.903 | 2.01e-02 | 2.81e-01 | 2367 | 1728 | 639 |
| African | Recorded Depression | Discontinuation vs Continuation | Recorded Anxiety | Education_levelCollege Postgraduate | 1.088 | 0.680 | 1.742 | 7.26e-01 | 1.00e+00 | 2367 | 1728 | 639 |
| African | Recorded Depression | Discontinuation vs Continuation | Recorded Anxiety | Education_levelCollege Undergraduate | 1.043 | 0.749 | 1.453 | 8.04e-01 | 1.00e+00 | 2367 | 1728 | 639 |
| African | Recorded Depression | Discontinuation vs Continuation | Recorded Anxiety | Education_levelTwelve or GED | 1.070 | 0.763 | 1.501 | 6.95e-01 | 1.00e+00 | 2367 | 1728 | 639 |
| African | Recorded Depression | Discontinuation vs Continuation | Recorded Anxiety | Recorded Anxiety | 0.945 | 0.778 | 1.147 | 5.67e-01 | 1.00e+00 | 2367 | 1728 | 639 |
| African | Recorded Depression | Switching vs Continuation | Recorded Anxiety | (Intercept) | 0.619 | 0.483 | 0.794 | 1.64e-04 | 2.29e-03 | 3605 | 1728 | 1877 |
| African | Recorded Depression | Switching vs Continuation | Recorded Anxiety | age_at_first_AD | 0.868 | 0.800 | 0.942 | 6.45e-04 | 9.03e-03 | 3605 | 1728 | 1877 |
| African | Recorded Depression | Switching vs Continuation | Recorded Anxiety | sex_at_birthMale | 1.104 | 0.940 | 1.297 | 2.29e-01 | 1.00e+00 | 3605 | 1728 | 1877 |

| | | | | | | | | | | | | |
|---|---|---|---|---|---|---|---|---|---|---|---|---|
| African | Recorded Depression | Switching vs Continuation | Recorded Anxiety | Income>200k | 0.416 | 0.178 | 0.975 | 4.36e-02 | 6.11e-01 | 3605 | 1728 | 1877 |
| African | Recorded Depression | Switching vs Continuation | Recorded Anxiety | Income100-150k | 0.551 | 0.375 | 0.809 | 2.40e-03 | 3.36e-02 | 3605 | 1728 | 1877 |
| African | Recorded Depression | Switching vs Continuation | Recorded Anxiety | Income150-200k | 0.604 | 0.306 | 1.190 | 1.45e-01 | 1.00e+00 | 3605 | 1728 | 1877 |
| African | Recorded Depression | Switching vs Continuation | Recorded Anxiety | Income25-35k | 0.806 | 0.644 | 1.008 | 5.93e-02 | 8.30e-01 | 3605 | 1728 | 1877 |
| African | Recorded Depression | Switching vs Continuation | Recorded Anxiety | Income35-50k | 0.783 | 0.625 | 0.980 | 3.24e-02 | 4.53e-01 | 3605 | 1728 | 1877 |
| African | Recorded Depression | Switching vs Continuation | Recorded Anxiety | Income50-75k | 0.643 | 0.500 | 0.827 | 5.65e-04 | 7.91e-03 | 3605 | 1728 | 1877 |
| African | Recorded Depression | Switching vs Continuation | Recorded Anxiety | Income75-100k | 0.592 | 0.418 | 0.838 | 3.15e-03 | 4.42e-02 | 3605 | 1728 | 1877 |
| African | Recorded Depression | Switching vs Continuation | Recorded Anxiety | Education_levelCollege Postgraduate | 1.138 | 0.815 | 1.590 | 4.48e-01 | 1.00e+00 | 3605 | 1728 | 1877 |
| African | Recorded Depression | Switching vs Continuation | Recorded Anxiety | Education_levelCollege Undergraduate | 0.986 | 0.774 | 1.258 | 9.12e-01 | 1.00e+00 | 3605 | 1728 | 1877 |
| African | Recorded Depression | Switching vs Continuation | Recorded Anxiety | Education_levelTwelve or GED | 0.877 | 0.682 | 1.128 | 3.06e-01 | 1.00e+00 | 3605 | 1728 | 1877 |
| African | Recorded Depression | Switching vs Continuation | Recorded Anxiety | Recorded Anxiety | 2.486 | 2.119 | 2.918 | <2.2e-16 | <2.2e-16 | 3605 | 1728 | 1877 |
| African | Recorded Depression | Discontinuation vs Continuation | Smoking_Frequency | (Intercept) | 0.338 | 0.244 | 0.468 | 7.16e-11 | 1.00e-09 | 2306 | 1685 | 621 |
| African | Recorded Depression | Discontinuation vs Continuation | Smoking_Frequency | age_at_first_AD | 0.913 | 0.820 | 1.017 | 9.89e-02 | 1.00e+00 | 2306 | 1685 | 621 |
| African | Recorded Depression | Discontinuation vs Continuation | Smoking_Frequency | sex_at_birthMale | 1.246 | 1.001 | 1.551 | 4.88e-02 | 6.83e-01 | 2306 | 1685 | 621 |
| African | Recorded Depression | Discontinuation vs Continuation | Smoking_Frequency | Income>200k | 0.681 | 0.215 | 2.156 | 5.14e-01 | 1.00e+00 | 2306 | 1685 | 621 |
| African | Recorded Depression | Discontinuation vs Continuation | Smoking_Frequency | Income100-150k | 0.789 | 0.469 | 1.327 | 3.71e-01 | 1.00e+00 | 2306 | 1685 | 621 |
| African | Recorded Depression | Discontinuation vs Continuation | Smoking_Frequency | Income150-200k | 0.629 | 0.229 | 1.729 | 3.69e-01 | 1.00e+00 | 2306 | 1685 | 621 |
| African | Recorded Depression | Discontinuation vs Continuation | Smoking_Frequency | Income25-35k | 0.981 | 0.724 | 1.329 | 9.00e-01 | 1.00e+00 | 2306 | 1685 | 621 |
| African | Recorded Depression | Discontinuation vs Continuation | Smoking_Frequency | Income35-50k | 0.760 | 0.546 | 1.057 | 1.03e-01 | 1.00e+00 | 2306 | 1685 | 621 |
| African | Recorded Depression | Discontinuation vs Continuation | Smoking_Frequency | Income50-75k | 0.905 | 0.642 | 1.275 | 5.68e-01 | 1.00e+00 | 2306 | 1685 | 621 |
| African | Recorded Depression | Discontinuation vs Continuation | Smoking_Frequency | Income75-100k | 0.549 | 0.315 | 0.956 | 3.41e-02 | 4.77e-01 | 2306 | 1685 | 621 |
| African | Recorded Depression | Discontinuation vs Continuation | Smoking_Frequency | Education_levelCollege Postgraduate | 1.138 | 0.706 | 1.835 | 5.96e-01 | 1.00e+00 | 2306 | 1685 | 621 |
| African | Recorded Depression | Discontinuation vs Continuation | Smoking_Frequency | Education_levelCollege Undergraduate | 1.067 | 0.762 | 1.496 | 7.05e-01 | 1.00e+00 | 2306 | 1685 | 621 |
| African | Recorded Depression | Discontinuation vs Continuation | Smoking_Frequency | Education_levelTwelve or GED | 1.061 | 0.753 | 1.495 | 7.36e-01 | 1.00e+00 | 2306 | 1685 | 621 |
| African | Recorded Depression | Discontinuation vs Continuation | Smoking_Frequency | Smoking_FrequencyEvery Day | 1.405 | 1.087 | 1.814 | 9.26e-03 | 1.30e-01 | 2306 | 1685 | 621 |
| African | Recorded Depression | Discontinuation vs Continuation | Smoking_Frequency | Smoking_FrequencyNot At All (former) | 0.918 | 0.707 | 1.190 | 5.17e-01 | 1.00e+00 | 2306 | 1685 | 621 |
| African | Recorded Depression | Discontinuation vs Continuation | Smoking_Frequency | Smoking_FrequencySome Days | 0.860 | 0.606 | 1.221 | 4.00e-01 | 1.00e+00 | 2306 | 1685 | 621 |
| African | Recorded Depression | Switching vs Continuation | Smoking_Frequency | (Intercept) | 1.056 | 0.835 | 1.335 | 6.48e-01 | 1.00e+00 | 3498 | 1685 | 1813 |
| African | Recorded Depression | Switching vs Continuation | Smoking_Frequency | age_at_first_AD | 0.824 | 0.759 | 0.895 | 4.37e-06 | 6.12e-05 | 3498 | 1685 | 1813 |
| African | Recorded Depression | Switching vs Continuation | Smoking_Frequency | sex_at_birthMale | 0.973 | 0.826 | 1.146 | 7.43e-01 | 1.00e+00 | 3498 | 1685 | 1813 |
| African | Recorded Depression | Switching vs Continuation | Smoking_Frequency | Income>200k | 0.467 | 0.199 | 1.097 | 8.07e-02 | 1.00e+00 | 3498 | 1685 | 1813 |
| African | Recorded Depression | Switching vs Continuation | Smoking_Frequency | Income100-150k | 0.605 | 0.412 | 0.889 | 1.04e-02 | 1.46e-01 | 3498 | 1685 | 1813 |
| African | Recorded Depression | Switching vs Continuation | Smoking_Frequency | Income150-200k | 0.640 | 0.323 | 1.268 | 2.01e-01 | 1.00e+00 | 3498 | 1685 | 1813 |
| African | Recorded Depression | Switching vs Continuation | Smoking_Frequency | Income25-35k | 0.820 | 0.655 | 1.026 | 8.31e-02 | 1.00e+00 | 3498 | 1685 | 1813 |
| African | Recorded Depression | Switching vs Continuation | Smoking_Frequency | Income35-50k | 0.835 | 0.667 | 1.045 | 1.16e-01 | 1.00e+00 | 3498 | 1685 | 1813 |
| African | Recorded Depression | Switching vs Continuation | Smoking_Frequency | Income50-75k | 0.703 | 0.546 | 0.905 | 6.21e-03 | 8.69e-02 | 3498 | 1685 | 1813 |
| African | Recorded Depression | Switching vs Continuation | Smoking_Frequency | Income75-100k | 0.657 | 0.463 | 0.933 | 1.88e-02 | 2.64e-01 | 3498 | 1685 | 1813 |
| African | Recorded Depression | Switching vs Continuation | Smoking_Frequency | Education_levelCollege Postgraduate | 1.244 | 0.890 | 1.739 | 2.01e-01 | 1.00e+00 | 3498 | 1685 | 1813 |
| African | Recorded Depression | Switching vs Continuation | Smoking_Frequency | Education_levelCollege Undergraduate | 1.064 | 0.835 | 1.356 | 6.16e-01 | 1.00e+00 | 3498 | 1685 | 1813 |
| African | Recorded Depression | Switching vs Continuation | Smoking_Frequency | Education_levelTwelve or GED | 0.916 | 0.713 | 1.176 | 4.91e-01 | 1.00e+00 | 3498 | 1685 | 1813 |
| African | Recorded Depression | Switching vs Continuation | Smoking_Frequency | Smoking_FrequencyEvery Day | 1.304 | 1.074 | 1.585 | 7.39e-03 | 1.04e-01 | 3498 | 1685 | 1813 |
| African | Recorded Depression | Switching vs Continuation | Smoking_Frequency | Smoking_FrequencyNot At All (former) | 1.167 | 0.975 | 1.397 | 9.15e-02 | 1.00e+00 | 3498 | 1685 | 1813 |
| African | Recorded Depression | Switching vs Continuation | Smoking_Frequency | Smoking_FrequencySome Days | 1.044 | 0.818 | 1.333 | 7.27e-01 | 1.00e+00 | 3498 | 1685 | 1813 |
| African | Recorded Depression | Discontinuation vs Continuation | Stress | (Intercept) | 0.252 | 0.105 | 0.607 | 2.13e-03 | 2.99e-02 | 653 | 491 | 162 |
| African | Recorded Depression | Discontinuation vs Continuation | Stress | age_at_first_AD | 0.973 | 0.784 | 1.207 | 8.03e-01 | 1.00e+00 | 653 | 491 | 162 |
| African | Recorded Depression | Discontinuation vs Continuation | Stress | sex_at_birthMale | 0.968 | 0.613 | 1.528 | 8.89e-01 | 1.00e+00 | 653 | 491 | 162 |
| African | Recorded Depression | Discontinuation vs Continuation | Stress | Income>200k | 0.443 | 0.085 | 2.307 | 3.33e-01 | 1.00e+00 | 653 | 491 | 162 |
| African | Recorded Depression | Discontinuation vs Continuation | Stress | Income100-150k | 0.789 | 0.383 | 1.623 | 5.19e-01 | 1.00e+00 | 653 | 491 | 162 |
| African | Recorded Depression | Discontinuation vs Continuation | Stress | Income150-200k | 0.514 | 0.104 | 2.535 | 4.13e-01 | 1.00e+00 | 653 | 491 | 162 |
| African | Recorded Depression | Discontinuation vs Continuation | Stress | Income25-35k | 0.724 | 0.381 | 1.376 | 3.25e-01 | 1.00e+00 | 653 | 491 | 162 |
| African | Recorded Depression | Discontinuation vs Continuation | Stress | Income35-50k | 0.597 | 0.319 | 1.118 | 1.07e-01 | 1.00e+00 | 653 | 491 | 162 |
| African | Recorded Depression | Discontinuation vs Continuation | Stress | Income50-75k | 0.914 | 0.523 | 1.598 | 7.53e-01 | 1.00e+00 | 653 | 491 | 162 |
| African | Recorded Depression | Discontinuation vs Continuation | Stress | Income75-100k | 0.320 | 0.118 | 0.871 | 2.57e-02 | 3.59e-01 | 653 | 491 | 162 |
| African | Recorded Depression | Discontinuation vs Continuation | Stress | Education_levelCollege Postgraduate | 1.621 | 0.576 | 4.567 | 3.60e-01 | 1.00e+00 | 653 | 491 | 162 |
| African | Recorded Depression | Discontinuation vs Continuation | Stress | Education_levelCollege Undergraduate | 1.279 | 0.516 | 3.173 | 5.95e-01 | 1.00e+00 | 653 | 491 | 162 |
| African | Recorded Depression | Discontinuation vs Continuation | Stress | Education_levelTwelve or GED | 1.211 | 0.471 | 3.114 | 6.92e-01 | 1.00e+00 | 653 | 491 | 162 |
| African | Recorded Depression | Discontinuation vs Continuation | Stress | StressAlmost Never | 2.777 | 1.231 | 6.267 | 1.39e-02 | 1.95e-01 | 653 | 491 | 162 |
| African | Recorded Depression | Discontinuation vs Continuation | Stress | StressFairly Often | 1.106 | 0.523 | 2.339 | 7.92e-01 | 1.00e+00 | 653 | 491 | 162 |
| African | Recorded Depression | Discontinuation vs Continuation | Stress | StressSometimes | 1.385 | 0.925 | 2.075 | 1.14e-01 | 1.00e+00 | 653 | 491 | 162 |
| African | Recorded Depression | Discontinuation vs Continuation | Stress | StressVery Often | 1.671 | 0.715 | 3.908 | 2.36e-01 | 1.00e+00 | 653 | 491 | 162 |
| African | Recorded Depression | Switching vs Continuation | Stress | (Intercept) | 1.244 | 0.714 | 2.167 | 4.41e-01 | 1.00e+00 | 1071 | 491 | 580 |
| African | Recorded Depression | Switching vs Continuation | Stress | age_at_first_AD | 0.701 | 0.601 | 0.818 | 6.76e-06 | 9.46e-05 | 1071 | 491 | 580 |
| African | Recorded Depression | Switching vs Continuation | Stress | sex_at_birthMale | 1.113 | 0.818 | 1.514 | 4.96e-01 | 1.00e+00 | 1071 | 491 | 580 |
| African | Recorded Depression | Switching vs Continuation | Stress | Income>200k | 0.224 | 0.057 | 0.873 | 3.11e-02 | 4.36e-01 | 1071 | 491 | 580 |
| African | Recorded Depression | Switching vs Continuation | Stress | Income100-150k | 0.558 | 0.328 | 0.951 | 3.19e-02 | 4.46e-01 | 1071 | 491 | 580 |
| African | Recorded Depression | Switching vs Continuation | Stress | Income150-200k | 0.734 | 0.283 | 1.904 | 5.24e-01 | 1.00e+00 | 1071 | 491 | 580 |

| | | | | | | | | | | | | |
|---|---|---|---|---|---|---|---|---|---|---|---|---|
| African | Recorded Depression | Switching vs Continuation | Stress | Income25-35k | 0.910 | 0.600 | 1.382 | 6.60e-01 | 1.00e+00 | 1071 | 491 | 580 |
| African | Recorded Depression | Switching vs Continuation | Stress | Income35-50k | 0.725 | 0.487 | 1.081 | 1.15e-01 | 1.00e+00 | 1071 | 491 | 580 |
| African | Recorded Depression | Switching vs Continuation | Stress | Income50-75k | 0.812 | 0.542 | 1.215 | 3.11e-01 | 1.00e+00 | 1071 | 491 | 580 |
| African | Recorded Depression | Switching vs Continuation | Stress | Income75-100k | 0.650 | 0.386 | 1.095 | 1.05e-01 | 1.00e+00 | 1071 | 491 | 580 |
| African | Recorded Depression | Switching vs Continuation | Stress | Education_levelCollege Postgraduate | 1.095 | 0.562 | 2.134 | 7.90e-01 | 1.00e+00 | 1071 | 491 | 580 |
| African | Recorded Depression | Switching vs Continuation | Stress | Education_levelCollege Undergraduate | 0.898 | 0.505 | 1.598 | 7.15e-01 | 1.00e+00 | 1071 | 491 | 580 |
| African | Recorded Depression | Switching vs Continuation | Stress | Education_levelTwelve or GED | 0.815 | 0.444 | 1.498 | 5.10e-01 | 1.00e+00 | 1071 | 491 | 580 |
| African | Recorded Depression | Switching vs Continuation | Stress | StressAlmost Never | 2.104 | 1.124 | 3.936 | 2.00e-02 | 2.80e-01 | 1071 | 491 | 580 |
| African | Recorded Depression | Switching vs Continuation | Stress | StressFairly Often | 0.838 | 0.502 | 1.400 | 5.00e-01 | 1.00e+00 | 1071 | 491 | 580 |
| African | Recorded Depression | Switching vs Continuation | Stress | StressSometimes | 1.221 | 0.932 | 1.600 | 1.47e-01 | 1.00e+00 | 1071 | 491 | 580 |
| African | Recorded Depression | Switching vs Continuation | Stress | StressVery Often | 1.510 | 0.832 | 2.743 | 1.76e-01 | 1.00e+00 | 1071 | 491 | 580 |
| African | Recorded Depression | Discontinuation vs Continuation | age_at_first_AD | (Intercept) | 0.343 | 0.308 | 0.382 | <2.2e-16 | <2.2e-16 | 2367 | 1728 | 639 |
| African | Recorded Depression | Discontinuation vs Continuation | age_at_first_AD | age_at_first_AD | 0.903 | 0.815 | 1.000 | 5.05e-02 | 7.06e-01 | 2367 | 1728 | 639 |
| African | Recorded Depression | Discontinuation vs Continuation | age_at_first_AD | sex_at_birthMale | 1.261 | 1.025 | 1.551 | 2.84e-02 | 3.97e-01 | 2367 | 1728 | 639 |
| African | Recorded Depression | Switching vs Continuation | age_at_first_AD | (Intercept) | 1.049 | 0.972 | 1.133 | 2.19e-01 | 1.00e+00 | 3605 | 1728 | 1877 |
| African | Recorded Depression | Switching vs Continuation | age_at_first_AD | age_at_first_AD | 0.843 | 0.779 | 0.912 | 2.23e-05 | 3.12e-04 | 3605 | 1728 | 1877 |
| African | Recorded Depression | Switching vs Continuation | age_at_first_AD | sex_at_birthMale | 1.021 | 0.874 | 1.192 | 7.97e-01 | 1.00e+00 | 3605 | 1728 | 1877 |
| African | Recorded Depression | Discontinuation vs Continuation | sex_at_birth | (Intercept) | 0.343 | 0.308 | 0.382 | <2.2e-16 | <2.2e-16 | 2367 | 1728 | 639 |
| African | Recorded Depression | Discontinuation vs Continuation | sex_at_birth | age_at_first_AD | 0.903 | 0.815 | 1.000 | 5.05e-02 | 7.06e-01 | 2367 | 1728 | 639 |
| African | Recorded Depression | Discontinuation vs Continuation | sex_at_birth | sex_at_birthMale | 1.261 | 1.025 | 1.551 | 2.84e-02 | 3.97e-01 | 2367 | 1728 | 639 |
| African | Recorded Depression | Switching vs Continuation | sex_at_birth | (Intercept) | 1.049 | 0.972 | 1.133 | 2.19e-01 | 1.00e+00 | 3605 | 1728 | 1877 |
| African | Recorded Depression | Switching vs Continuation | sex_at_birth | age_at_first_AD | 0.843 | 0.779 | 0.912 | 2.23e-05 | 3.12e-04 | 3605 | 1728 | 1877 |
| African | Recorded Depression | Switching vs Continuation | sex_at_birth | sex_at_birthMale | 1.021 | 0.874 | 1.192 | 7.97e-01 | 1.00e+00 | 3605 | 1728 | 1877 |
| African | No Recorded Depression | Discontinuation vs Continuation | Alcohol_Frequency | (Intercept) | 0.389 | 0.294 | 0.515 | 3.72e-11 | 5.20e-10 | 3511 | 2375 | 1136 |
| African | No Recorded Depression | Discontinuation vs Continuation | Alcohol_Frequency | age_at_first_AD | 1.029 | 0.946 | 1.120 | 5.04e-01 | 1.00e+00 | 3511 | 2375 | 1136 |
| African | No Recorded Depression | Discontinuation vs Continuation | Alcohol_Frequency | sex_at_birthMale | 1.181 | 1.006 | 1.387 | 4.23e-02 | 5.93e-01 | 3511 | 2375 | 1136 |
| African | No Recorded Depression | Discontinuation vs Continuation | Alcohol_Frequency | Income>200k | 0.892 | 0.459 | 1.732 | 7.35e-01 | 1.00e+00 | 3511 | 2375 | 1136 |
| African | No Recorded Depression | Discontinuation vs Continuation | Alcohol_Frequency | Income100-150k | 0.796 | 0.541 | 1.169 | 2.45e-01 | 1.00e+00 | 3511 | 2375 | 1136 |
| African | No Recorded Depression | Discontinuation vs Continuation | Alcohol_Frequency | Income150-200k | 1.635 | 0.927 | 2.885 | 8.96e-02 | 1.00e+00 | 3511 | 2375 | 1136 |
| African | No Recorded Depression | Discontinuation vs Continuation | Alcohol_Frequency | Income25-35k | 1.281 | 1.017 | 1.613 | 3.54e-02 | 4.95e-01 | 3511 | 2375 | 1136 |
| African | No Recorded Depression | Discontinuation vs Continuation | Alcohol_Frequency | Income35-50k | 1.145 | 0.896 | 1.464 | 2.79e-01 | 1.00e+00 | 3511 | 2375 | 1136 |
| African | No Recorded Depression | Discontinuation vs Continuation | Alcohol_Frequency | Income50-75k | 1.062 | 0.824 | 1.369 | 6.43e-01 | 1.00e+00 | 3511 | 2375 | 1136 |
| African | No Recorded Depression | Discontinuation vs Continuation | Alcohol_Frequency | Income75-100k | 0.975 | 0.684 | 1.391 | 8.91e-01 | 1.00e+00 | 3511 | 2375 | 1136 |
| African | No Recorded Depression | Discontinuation vs Continuation | Alcohol_Frequency | Education_levelCollege Postgraduate | 0.877 | 0.619 | 1.243 | 4.62e-01 | 1.00e+00 | 3511 | 2375 | 1136 |
| African | No Recorded Depression | Discontinuation vs Continuation | Alcohol_Frequency | Education_levelCollege Undergraduate | 1.071 | 0.833 | 1.378 | 5.92e-01 | 1.00e+00 | 3511 | 2375 | 1136 |
| African | No Recorded Depression | Discontinuation vs Continuation | Alcohol_Frequency | Education_levelTwelve or GED | 1.087 | 0.841 | 1.404 | 5.26e-01 | 1.00e+00 | 3511 | 2375 | 1136 |
| African | No Recorded Depression | Discontinuation vs Continuation | Alcohol_Frequency | Alcohol_Frequency2-3 Week | 1.070 | 0.789 | 1.452 | 6.64e-01 | 1.00e+00 | 3511 | 2375 | 1136 |
| African | No Recorded Depression | Discontinuation vs Continuation | Alcohol_Frequency | Alcohol_Frequency2-4 Month | 1.045 | 0.810 | 1.348 | 7.35e-01 | 1.00e+00 | 3511 | 2375 | 1136 |
| African | No Recorded Depression | Discontinuation vs Continuation | Alcohol_Frequency | Alcohol_Frequency4plus Week | 1.104 | 0.785 | 1.554 | 5.69e-01 | 1.00e+00 | 3511 | 2375 | 1136 |
| African | No Recorded Depression | Discontinuation vs Continuation | Alcohol_Frequency | Alcohol_FrequencyMonthly or Less | 1.160 | 0.925 | 1.456 | 1.99e-01 | 1.00e+00 | 3511 | 2375 | 1136 |
| African | No Recorded Depression | Discontinuation vs Continuation | Alcohol_Frequency | Alcohol_FrequencyNever (former) | 0.959 | 0.738 | 1.248 | 7.57e-01 | 1.00e+00 | 3511 | 2375 | 1136 |
| African | No Recorded Depression | Switching vs Continuation | Alcohol_Frequency | (Intercept) | 0.363 | 0.264 | 0.500 | 5.85e-10 | 8.19e-09 | 3002 | 2375 | 627 |
| African | No Recorded Depression | Switching vs Continuation | Alcohol_Frequency | age_at_first_AD | 0.712 | 0.639 | 0.793 | 5.36e-10 | 7.50e-09 | 3002 | 2375 | 627 |
| African | No Recorded Depression | Switching vs Continuation | Alcohol_Frequency | sex_at_birthMale | 0.891 | 0.722 | 1.100 | 2.82e-01 | 1.00e+00 | 3002 | 2375 | 627 |
| African | No Recorded Depression | Switching vs Continuation | Alcohol_Frequency | Income>200k | 0.797 | 0.357 | 1.783 | 5.81e-01 | 1.00e+00 | 3002 | 2375 | 627 |
| African | No Recorded Depression | Switching vs Continuation | Alcohol_Frequency | Income100-150k | 0.934 | 0.602 | 1.448 | 7.61e-01 | 1.00e+00 | 3002 | 2375 | 627 |
| African | No Recorded Depression | Switching vs Continuation | Alcohol_Frequency | Income150-200k | 0.548 | 0.207 | 1.454 | 2.27e-01 | 1.00e+00 | 3002 | 2375 | 627 |
| African | No Recorded Depression | Switching vs Continuation | Alcohol_Frequency | Income25-35k | 0.640 | 0.457 | 0.896 | 9.35e-03 | 1.31e-01 | 3002 | 2375 | 627 |
| African | No Recorded Depression | Switching vs Continuation | Alcohol_Frequency | Income35-50k | 0.778 | 0.562 | 1.078 | 1.31e-01 | 1.00e+00 | 3002 | 2375 | 627 |
| African | No Recorded Depression | Switching vs Continuation | Alcohol_Frequency | Income50-75k | 0.754 | 0.539 | 1.056 | 1.00e-01 | 1.00e+00 | 3002 | 2375 | 627 |
| African | No Recorded Depression | Switching vs Continuation | Alcohol_Frequency | Income75-100k | 0.798 | 0.507 | 1.257 | 3.30e-01 | 1.00e+00 | 3002 | 2375 | 627 |
| African | No Recorded Depression | Switching vs Continuation | Alcohol_Frequency | Education_levelCollege Postgraduate | 0.795 | 0.522 | 1.211 | 2.86e-01 | 1.00e+00 | 3002 | 2375 | 627 |
| African | No Recorded Depression | Switching vs Continuation | Alcohol_Frequency | Education_levelCollege Undergraduate | 0.769 | 0.571 | 1.034 | 8.25e-02 | 1.00e+00 | 3002 | 2375 | 627 |
| African | No Recorded Depression | Switching vs Continuation | Alcohol_Frequency | Education_levelTwelve or GED | 0.876 | 0.650 | 1.181 | 3.85e-01 | 1.00e+00 | 3002 | 2375 | 627 |
| African | No Recorded Depression | Switching vs Continuation | Alcohol_Frequency | Alcohol_Frequency2-3 Week | 0.955 | 0.654 | 1.394 | 8.09e-01 | 1.00e+00 | 3002 | 2375 | 627 |
| African | No Recorded Depression | Switching vs Continuation | Alcohol_Frequency | Alcohol_Frequency2-4 Month | 0.841 | 0.612 | 1.157 | 2.88e-01 | 1.00e+00 | 3002 | 2375 | 627 |
| African | No Recorded Depression | Switching vs Continuation | Alcohol_Frequency | Alcohol_Frequency4plus Week | 1.046 | 0.686 | 1.595 | 8.36e-01 | 1.00e+00 | 3002 | 2375 | 627 |
| African | No Recorded Depression | Switching vs Continuation | Alcohol_Frequency | Alcohol_FrequencyMonthly or Less | 1.062 | 0.809 | 1.395 | 6.63e-01 | 1.00e+00 | 3002 | 2375 | 627 |
| African | No Recorded Depression | Switching vs Continuation | Alcohol_Frequency | Alcohol_FrequencyNever (former) | 1.071 | 0.781 | 1.468 | 6.71e-01 | 1.00e+00 | 3002 | 2375 | 627 |
| African | No Recorded Depression | Discontinuation vs Continuation | BMI | (Intercept) | 0.411 | 0.327 | 0.515 | 1.25e-14 | 1.75e-13 | 3559 | 2415 | 1144 |
| African | No Recorded Depression | Discontinuation vs Continuation | BMI | age_at_first_AD | 1.013 | 0.933 | 1.100 | 7.55e-01 | 1.00e+00 | 3559 | 2415 | 1144 |
| African | No Recorded Depression | Discontinuation vs Continuation | BMI | sex_at_birthMale | 1.171 | 0.996 | 1.375 | 5.53e-02 | 7.74e-01 | 3559 | 2415 | 1144 |
| African | No Recorded Depression | Discontinuation vs Continuation | BMI | Income>200k | 0.882 | 0.456 | 1.708 | 7.10e-01 | 1.00e+00 | 3559 | 2415 | 1144 |
| African | No Recorded Depression | Discontinuation vs Continuation | BMI | Income100-150k | 0.829 | 0.564 | 1.218 | 3.39e-01 | 1.00e+00 | 3559 | 2415 | 1144 |
| African | No Recorded Depression | Discontinuation vs Continuation | BMI | Income150-200k | 1.579 | 0.884 | 2.819 | 1.23e-01 | 1.00e+00 | 3559 | 2415 | 1144 |
| African | No Recorded Depression | Discontinuation vs Continuation | BMI | Income25-35k | 1.302 | 1.036 | 1.638 | 2.38e-02 | 3.34e-01 | 3559 | 2415 | 1144 |

| | | | | | | | | | | | | |
|---|---|---|---|---|---|---|---|---|---|---|---|---|
| African | No Recorded Depression | Discontinuation vs Continuation | BMI | Income35-50k | 1.154 | 0.904 | 1.474 | 2.51e-01 | 1.00e+00 | 3559 | 2415 | 1144 |
| African | No Recorded Depression | Discontinuation vs Continuation | BMI | Income50-75k | 1.122 | 0.873 | 1.443 | 3.69e-01 | 1.00e+00 | 3559 | 2415 | 1144 |
| African | No Recorded Depression | Discontinuation vs Continuation | BMI | Income75-100k | 0.970 | 0.682 | 1.380 | 8.66e-01 | 1.00e+00 | 3559 | 2415 | 1144 |
| African | No Recorded Depression | Discontinuation vs Continuation | BMI | Education_levelCollege Postgraduate | 0.878 | 0.621 | 1.241 | 4.61e-01 | 1.00e+00 | 3559 | 2415 | 1144 |
| African | No Recorded Depression | Discontinuation vs Continuation | BMI | Education_levelCollege Undergraduate | 1.067 | 0.832 | 1.369 | 6.10e-01 | 1.00e+00 | 3559 | 2415 | 1144 |
| African | No Recorded Depression | Discontinuation vs Continuation | BMI | Education_levelTwelve or GED | 1.110 | 0.861 | 1.431 | 4.20e-01 | 1.00e+00 | 3559 | 2415 | 1144 |
| African | No Recorded Depression | Discontinuation vs Continuation | BMI | BMI | 0.975 | 0.911 | 1.044 | 4.74e-01 | 1.00e+00 | 3559 | 2415 | 1144 |
| African | No Recorded Depression | Switching vs Continuation | BMI | (Intercept) | 0.373 | 0.289 | 0.481 | 4.03e-14 | 5.64e-13 | 3058 | 2415 | 643 |
| African | No Recorded Depression | Switching vs Continuation | BMI | age_at_first_AD | 0.720 | 0.649 | 0.799 | 5.85e-10 | 8.19e-09 | 3058 | 2415 | 643 |
| African | No Recorded Depression | Switching vs Continuation | BMI | sex_at_birthMale | 0.868 | 0.705 | 1.070 | 1.85e-01 | 1.00e+00 | 3058 | 2415 | 643 |
| African | No Recorded Depression | Switching vs Continuation | BMI | Income>200k | 0.817 | 0.379 | 1.760 | 6.05e-01 | 1.00e+00 | 3058 | 2415 | 643 |
| African | No Recorded Depression | Switching vs Continuation | BMI | Income100-150k | 0.874 | 0.562 | 1.360 | 5.51e-01 | 1.00e+00 | 3058 | 2415 | 643 |
| African | No Recorded Depression | Switching vs Continuation | BMI | Income150-200k | 0.526 | 0.198 | 1.394 | 1.96e-01 | 1.00e+00 | 3058 | 2415 | 643 |
| African | No Recorded Depression | Switching vs Continuation | BMI | Income25-35k | 0.643 | 0.461 | 0.898 | 9.44e-03 | 1.32e-01 | 3058 | 2415 | 643 |
| African | No Recorded Depression | Switching vs Continuation | BMI | Income35-50k | 0.747 | 0.540 | 1.035 | 7.95e-02 | 1.00e+00 | 3058 | 2415 | 643 |
| African | No Recorded Depression | Switching vs Continuation | BMI | Income50-75k | 0.728 | 0.520 | 1.020 | 6.48e-02 | 9.07e-01 | 3058 | 2415 | 643 |
| African | No Recorded Depression | Switching vs Continuation | BMI | Income75-100k | 0.806 | 0.519 | 1.251 | 3.36e-01 | 1.00e+00 | 3058 | 2415 | 643 |
| African | No Recorded Depression | Switching vs Continuation | BMI | Education_levelCollege Postgraduate | 0.829 | 0.548 | 1.252 | 3.71e-01 | 1.00e+00 | 3058 | 2415 | 643 |
| African | No Recorded Depression | Switching vs Continuation | BMI | Education_levelCollege Undergraduate | 0.775 | 0.580 | 1.037 | 8.59e-02 | 1.00e+00 | 3058 | 2415 | 643 |
| African | No Recorded Depression | Switching vs Continuation | BMI | Education_levelTwelve or GED | 0.894 | 0.667 | 1.198 | 4.52e-01 | 1.00e+00 | 3058 | 2415 | 643 |
| African | No Recorded Depression | Switching vs Continuation | BMI | BMI | 0.953 | 0.877 | 1.036 | 2.56e-01 | 1.00e+00 | 3058 | 2415 | 643 |
| African | No Recorded Depression | Discontinuation vs Continuation | Education_level | (Intercept) | 0.415 | 0.332 | 0.518 | 8.36e-15 | 1.17e-13 | 3605 | 2436 | 1169 |
| African | No Recorded Depression | Discontinuation vs Continuation | Education_level | age_at_first_AD | 1.016 | 0.937 | 1.102 | 7.01e-01 | 1.00e+00 | 3605 | 2436 | 1169 |
| African | No Recorded Depression | Discontinuation vs Continuation | Education_level | sex_at_birthMale | 1.186 | 1.016 | 1.385 | 3.08e-02 | 4.31e-01 | 3605 | 2436 | 1169 |
| African | No Recorded Depression | Discontinuation vs Continuation | Education_level | Income>200k | 0.872 | 0.451 | 1.688 | 6.85e-01 | 1.00e+00 | 3605 | 2436 | 1169 |
| African | No Recorded Depression | Discontinuation vs Continuation | Education_level | Income100-150k | 0.809 | 0.551 | 1.187 | 2.79e-01 | 1.00e+00 | 3605 | 2436 | 1169 |
| African | No Recorded Depression | Discontinuation vs Continuation | Education_level | Income150-200k | 1.773 | 1.014 | 3.100 | 4.47e-02 | 6.26e-01 | 3605 | 2436 | 1169 |
| African | No Recorded Depression | Discontinuation vs Continuation | Education_level | Income25-35k | 1.310 | 1.043 | 1.644 | 2.01e-02 | 2.82e-01 | 3605 | 2436 | 1169 |
| African | No Recorded Depression | Discontinuation vs Continuation | Education_level | Income35-50k | 1.159 | 0.910 | 1.476 | 2.31e-01 | 1.00e+00 | 3605 | 2436 | 1169 |
| African | No Recorded Depression | Discontinuation vs Continuation | Education_level | Income50-75k | 1.118 | 0.871 | 1.435 | 3.82e-01 | 1.00e+00 | 3605 | 2436 | 1169 |
| African | No Recorded Depression | Discontinuation vs Continuation | Education_level | Income75-100k | 0.987 | 0.697 | 1.398 | 9.40e-01 | 1.00e+00 | 3605 | 2436 | 1169 |
| African | No Recorded Depression | Discontinuation vs Continuation | Education_level | Education_levelCollege Postgraduate | 0.882 | 0.626 | 1.242 | 4.71e-01 | 1.00e+00 | 3605 | 2436 | 1169 |
| African | No Recorded Depression | Discontinuation vs Continuation | Education_level | Education_levelCollege Undergraduate | 1.056 | 0.825 | 1.352 | 6.67e-01 | 1.00e+00 | 3605 | 2436 | 1169 |
| African | No Recorded Depression | Discontinuation vs Continuation | Education_level | Education_levelTwelve or GED | 1.086 | 0.845 | 1.397 | 5.19e-01 | 1.00e+00 | 3605 | 2436 | 1169 |
| African | No Recorded Depression | Switching vs Continuation | Education_level | (Intercept) | 0.375 | 0.291 | 0.482 | 2.16e-14 | 3.02e-13 | 3088 | 2436 | 652 |
| African | No Recorded Depression | Switching vs Continuation | Education_level | age_at_first_AD | 0.719 | 0.649 | 0.797 | 3.93e-10 | 5.50e-09 | 3088 | 2436 | 652 |
| African | No Recorded Depression | Switching vs Continuation | Education_level | sex_at_birthMale | 0.880 | 0.718 | 1.078 | 2.17e-01 | 1.00e+00 | 3088 | 2436 | 652 |
| African | No Recorded Depression | Switching vs Continuation | Education_level | Income>200k | 0.834 | 0.387 | 1.795 | 6.43e-01 | 1.00e+00 | 3088 | 2436 | 652 |
| African | No Recorded Depression | Switching vs Continuation | Education_level | Income100-150k | 0.904 | 0.584 | 1.398 | 6.50e-01 | 1.00e+00 | 3088 | 2436 | 652 |
| African | No Recorded Depression | Switching vs Continuation | Education_level | Income150-200k | 0.534 | 0.201 | 1.413 | 2.06e-01 | 1.00e+00 | 3088 | 2436 | 652 |
| African | No Recorded Depression | Switching vs Continuation | Education_level | Income25-35k | 0.648 | 0.466 | 0.902 | 1.01e-02 | 1.41e-01 | 3088 | 2436 | 652 |
| African | No Recorded Depression | Switching vs Continuation | Education_level | Income35-50k | 0.762 | 0.553 | 1.050 | 9.70e-02 | 1.00e+00 | 3088 | 2436 | 652 |
| African | No Recorded Depression | Switching vs Continuation | Education_level | Income50-75k | 0.752 | 0.539 | 1.048 | 9.21e-02 | 1.00e+00 | 3088 | 2436 | 652 |
| African | No Recorded Depression | Switching vs Continuation | Education_level | Income75-100k | 0.806 | 0.519 | 1.251 | 3.36e-01 | 1.00e+00 | 3088 | 2436 | 652 |
| African | No Recorded Depression | Switching vs Continuation | Education_level | Education_levelCollege Postgraduate | 0.795 | 0.527 | 1.198 | 2.73e-01 | 1.00e+00 | 3088 | 2436 | 652 |
| African | No Recorded Depression | Switching vs Continuation | Education_level | Education_levelCollege Undergraduate | 0.759 | 0.569 | 1.013 | 6.10e-02 | 8.54e-01 | 3088 | 2436 | 652 |
| African | No Recorded Depression | Switching vs Continuation | Education_level | Education_levelTwelve or GED | 0.859 | 0.642 | 1.149 | 3.05e-01 | 1.00e+00 | 3088 | 2436 | 652 |
| African | No Recorded Depression | Discontinuation vs Continuation | Income | (Intercept) | 0.415 | 0.332 | 0.518 | 8.36e-15 | 1.17e-13 | 3605 | 2436 | 1169 |
| African | No Recorded Depression | Discontinuation vs Continuation | Income | age_at_first_AD | 1.016 | 0.937 | 1.102 | 7.01e-01 | 1.00e+00 | 3605 | 2436 | 1169 |
| African | No Recorded Depression | Discontinuation vs Continuation | Income | sex_at_birthMale | 1.186 | 1.016 | 1.385 | 3.08e-02 | 4.31e-01 | 3605 | 2436 | 1169 |
| African | No Recorded Depression | Discontinuation vs Continuation | Income | Income>200k | 0.872 | 0.451 | 1.688 | 6.85e-01 | 1.00e+00 | 3605 | 2436 | 1169 |
| African | No Recorded Depression | Discontinuation vs Continuation | Income | Income100-150k | 0.809 | 0.551 | 1.187 | 2.79e-01 | 1.00e+00 | 3605 | 2436 | 1169 |
| African | No Recorded Depression | Discontinuation vs Continuation | Income | Income150-200k | 1.773 | 1.014 | 3.100 | 4.47e-02 | 6.26e-01 | 3605 | 2436 | 1169 |
| African | No Recorded Depression | Discontinuation vs Continuation | Income | Income25-35k | 1.310 | 1.043 | 1.644 | 2.01e-02 | 2.82e-01 | 3605 | 2436 | 1169 |
| African | No Recorded Depression | Discontinuation vs Continuation | Income | Income35-50k | 1.159 | 0.910 | 1.476 | 2.31e-01 | 1.00e+00 | 3605 | 2436 | 1169 |
| African | No Recorded Depression | Discontinuation vs Continuation | Income | Income50-75k | 1.118 | 0.871 | 1.435 | 3.82e-01 | 1.00e+00 | 3605 | 2436 | 1169 |
| African | No Recorded Depression | Discontinuation vs Continuation | Income | Income75-100k | 0.987 | 0.697 | 1.398 | 9.40e-01 | 1.00e+00 | 3605 | 2436 | 1169 |
| African | No Recorded Depression | Discontinuation vs Continuation | Income | Education_levelCollege Postgraduate | 0.882 | 0.626 | 1.242 | 4.71e-01 | 1.00e+00 | 3605 | 2436 | 1169 |
| African | No Recorded Depression | Discontinuation vs Continuation | Income | Education_levelCollege Undergraduate | 1.056 | 0.825 | 1.352 | 6.67e-01 | 1.00e+00 | 3605 | 2436 | 1169 |
| African | No Recorded Depression | Discontinuation vs Continuation | Income | Education_levelTwelve or GED | 1.086 | 0.845 | 1.397 | 5.19e-01 | 1.00e+00 | 3605 | 2436 | 1169 |
| African | No Recorded Depression | Switching vs Continuation | Income | (Intercept) | 0.375 | 0.291 | 0.482 | 2.16e-14 | 3.02e-13 | 3088 | 2436 | 652 |
| African | No Recorded Depression | Switching vs Continuation | Income | age_at_first_AD | 0.719 | 0.649 | 0.797 | 3.93e-10 | 5.50e-09 | 3088 | 2436 | 652 |
| African | No Recorded Depression | Switching vs Continuation | Income | sex_at_birthMale | 0.880 | 0.718 | 1.078 | 2.17e-01 | 1.00e+00 | 3088 | 2436 | 652 |
| African | No Recorded Depression | Switching vs Continuation | Income | Income>200k | 0.834 | 0.387 | 1.795 | 6.43e-01 | 1.00e+00 | 3088 | 2436 | 652 |
| African | No Recorded Depression | Switching vs Continuation | Income | Income100-150k | 0.904 | 0.584 | 1.398 | 6.50e-01 | 1.00e+00 | 3088 | 2436 | 652 |
| African | No Recorded Depression | Switching vs Continuation | Income | Income150-200k | 0.534 | 0.201 | 1.413 | 2.06e-01 | 1.00e+00 | 3088 | 2436 | 652 |

| | | | | | | | | | | | | |
|---|---|---|---|---|---|---|---|---|---|---|---|---|
| African | No Recorded Depression | Switching vs Continuation | Income | Income25-35k | 0.648 | 0.466 | 0.902 | 1.01e-02 | 1.41e-01 | 3088 | 2436 | 652 |
| African | No Recorded Depression | Switching vs Continuation | Income | Income35-50k | 0.762 | 0.553 | 1.050 | 9.70e-02 | 1.00e+00 | 3088 | 2436 | 652 |
| African | No Recorded Depression | Switching vs Continuation | Income | Income50-75k | 0.752 | 0.539 | 1.048 | 9.21e-02 | 1.00e+00 | 3088 | 2436 | 652 |
| African | No Recorded Depression | Switching vs Continuation | Income | Income75-100k | 0.806 | 0.519 | 1.251 | 3.36e-01 | 1.00e+00 | 3088 | 2436 | 652 |
| African | No Recorded Depression | Switching vs Continuation | Income | Education_levelCollege Postgraduate | 0.795 | 0.527 | 1.198 | 2.73e-01 | 1.00e+00 | 3088 | 2436 | 652 |
| African | No Recorded Depression | Switching vs Continuation | Income | Education_levelCollege Undergraduate | 0.759 | 0.569 | 1.013 | 6.10e-02 | 8.54e-01 | 3088 | 2436 | 652 |
| African | No Recorded Depression | Switching vs Continuation | Income | Education_levelTwelve or GED | 0.859 | 0.642 | 1.149 | 3.05e-01 | 1.00e+00 | 3088 | 2436 | 652 |
| African | No Recorded Depression | Discontinuation vs Continuation | Insomnia | (Intercept) | 0.432 | 0.345 | 0.541 | 2.15e-13 | 3.01e-12 | 3605 | 2436 | 1169 |
| African | No Recorded Depression | Discontinuation vs Continuation | Insomnia | age_at_first_AD | 1.026 | 0.945 | 1.113 | 5.42e-01 | 1.00e+00 | 3605 | 2436 | 1169 |
| African | No Recorded Depression | Discontinuation vs Continuation | Insomnia | sex_at_birthMale | 1.183 | 1.013 | 1.382 | 3.36e-02 | 4.70e-01 | 3605 | 2436 | 1169 |
| African | No Recorded Depression | Discontinuation vs Continuation | Insomnia | Income>200k | 0.891 | 0.460 | 1.725 | 7.32e-01 | 1.00e+00 | 3605 | 2436 | 1169 |
| African | No Recorded Depression | Discontinuation vs Continuation | Insomnia | Income100-150k | 0.828 | 0.564 | 1.216 | 3.36e-01 | 1.00e+00 | 3605 | 2436 | 1169 |
| African | No Recorded Depression | Discontinuation vs Continuation | Insomnia | Income150-200k | 1.801 | 1.029 | 3.151 | 3.93e-02 | 5.50e-01 | 3605 | 2436 | 1169 |
| African | No Recorded Depression | Discontinuation vs Continuation | Insomnia | Income25-35k | 1.307 | 1.041 | 1.641 | 2.12e-02 | 2.96e-01 | 3605 | 2436 | 1169 |
| African | No Recorded Depression | Discontinuation vs Continuation | Insomnia | Income35-50k | 1.175 | 0.922 | 1.497 | 1.93e-01 | 1.00e+00 | 3605 | 2436 | 1169 |
| African | No Recorded Depression | Discontinuation vs Continuation | Insomnia | Income50-75k | 1.128 | 0.879 | 1.449 | 3.44e-01 | 1.00e+00 | 3605 | 2436 | 1169 |
| African | No Recorded Depression | Discontinuation vs Continuation | Insomnia | Income75-100k | 0.989 | 0.698 | 1.401 | 9.49e-01 | 1.00e+00 | 3605 | 2436 | 1169 |
| African | No Recorded Depression | Discontinuation vs Continuation | Insomnia | Education_levelCollege Postgraduate | 0.876 | 0.621 | 1.233 | 4.47e-01 | 1.00e+00 | 3605 | 2436 | 1169 |
| African | No Recorded Depression | Discontinuation vs Continuation | Insomnia | Education_levelCollege Undergraduate | 1.060 | 0.828 | 1.358 | 6.43e-01 | 1.00e+00 | 3605 | 2436 | 1169 |
| African | No Recorded Depression | Discontinuation vs Continuation | Insomnia | Education_levelTwelve or GED | 1.086 | 0.844 | 1.397 | 5.23e-01 | 1.00e+00 | 3605 | 2436 | 1169 |
| African | No Recorded Depression | Discontinuation vs Continuation | Insomnia | Insomnia | 0.767 | 0.635 | 0.927 | 6.11e-03 | 8.56e-02 | 3605 | 2436 | 1169 |
| African | No Recorded Depression | Switching vs Continuation | Insomnia | (Intercept) | 0.320 | 0.247 | 0.415 | <2.2e-16 | <2.2e-16 | 3088 | 2436 | 652 |
| African | No Recorded Depression | Switching vs Continuation | Insomnia | age_at_first_AD | 0.704 | 0.634 | 0.781 | 4.55e-11 | 6.37e-10 | 3088 | 2436 | 652 |
| African | No Recorded Depression | Switching vs Continuation | Insomnia | sex_at_birthMale | 0.890 | 0.725 | 1.092 | 2.65e-01 | 1.00e+00 | 3088 | 2436 | 652 |
| African | No Recorded Depression | Switching vs Continuation | Insomnia | Income>200k | 0.841 | 0.389 | 1.818 | 6.60e-01 | 1.00e+00 | 3088 | 2436 | 652 |
| African | No Recorded Depression | Switching vs Continuation | Insomnia | Income100-150k | 0.830 | 0.534 | 1.288 | 4.05e-01 | 1.00e+00 | 3088 | 2436 | 652 |
| African | No Recorded Depression | Switching vs Continuation | Insomnia | Income150-200k | 0.543 | 0.205 | 1.440 | 2.20e-01 | 1.00e+00 | 3088 | 2436 | 652 |
| African | No Recorded Depression | Switching vs Continuation | Insomnia | Income25-35k | 0.666 | 0.478 | 0.928 | 1.61e-02 | 2.26e-01 | 3088 | 2436 | 652 |
| African | No Recorded Depression | Switching vs Continuation | Insomnia | Income35-50k | 0.742 | 0.537 | 1.026 | 7.08e-02 | 9.92e-01 | 3088 | 2436 | 652 |
| African | No Recorded Depression | Switching vs Continuation | Insomnia | Income50-75k | 0.736 | 0.527 | 1.028 | 7.20e-02 | 1.00e+00 | 3088 | 2436 | 652 |
| African | No Recorded Depression | Switching vs Continuation | Insomnia | Income75-100k | 0.820 | 0.527 | 1.276 | 3.79e-01 | 1.00e+00 | 3088 | 2436 | 652 |
| African | No Recorded Depression | Switching vs Continuation | Insomnia | Education_levelCollege Postgraduate | 0.805 | 0.533 | 1.217 | 3.03e-01 | 1.00e+00 | 3088 | 2436 | 652 |
| African | No Recorded Depression | Switching vs Continuation | Insomnia | Education_levelCollege Undergraduate | 0.753 | 0.563 | 1.006 | 5.51e-02 | 7.71e-01 | 3088 | 2436 | 652 |
| African | No Recorded Depression | Switching vs Continuation | Insomnia | Education_levelTwelve or GED | 0.884 | 0.659 | 1.185 | 4.10e-01 | 1.00e+00 | 3088 | 2436 | 652 |
| African | No Recorded Depression | Switching vs Continuation | Insomnia | Insomnia | 1.876 | 1.535 | 2.292 | 7.80e-10 | 1.09e-08 | 3088 | 2436 | 652 |
| African | No Recorded Depression | Discontinuation vs Continuation | MDD CIDI-sf | (Intercept) | 0.822 | 0.104 | 6.490 | 8.52e-01 | 1.00e+00 | 203 | 143 | 60 |
| African | No Recorded Depression | Discontinuation vs Continuation | MDD CIDI-sf | age_at_first_AD | 0.873 | 0.600 | 1.271 | 4.80e-01 | 1.00e+00 | 203 | 143 | 60 |
| African | No Recorded Depression | Discontinuation vs Continuation | MDD CIDI-sf | sex_at_birthMale | 3.000 | 1.406 | 6.401 | 4.49e-03 | 6.29e-02 | 203 | 143 | 60 |
| African | No Recorded Depression | Discontinuation vs Continuation | MDD CIDI-sf | Income>200k | 2.080 | 0.360 | 12.010 | 4.13e-01 | 1.00e+00 | 203 | 143 | 60 |
| African | No Recorded Depression | Discontinuation vs Continuation | MDD CIDI-sf | Income100-150k | 0.952 | 0.291 | 3.113 | 9.35e-01 | 1.00e+00 | 203 | 143 | 60 |
| African | No Recorded Depression | Discontinuation vs Continuation | MDD CIDI-sf | Income150-200k | 1.344 | 0.095 | 19.080 | 8.27e-01 | 1.00e+00 | 203 | 143 | 60 |
| African | No Recorded Depression | Discontinuation vs Continuation | MDD CIDI-sf | Income25-35k | 0.909 | 0.300 | 2.752 | 8.66e-01 | 1.00e+00 | 203 | 143 | 60 |
| African | No Recorded Depression | Discontinuation vs Continuation | MDD CIDI-sf | Income35-50k | 1.572 | 0.548 | 4.512 | 4.00e-01 | 1.00e+00 | 203 | 143 | 60 |
| African | No Recorded Depression | Discontinuation vs Continuation | MDD CIDI-sf | Income50-75k | 1.165 | 0.404 | 3.359 | 7.78e-01 | 1.00e+00 | 203 | 143 | 60 |
| African | No Recorded Depression | Discontinuation vs Continuation | MDD CIDI-sf | Income75-100k | 3.915 | 1.148 | 13.350 | 2.92e-02 | 4.09e-01 | 203 | 143 | 60 |
| African | No Recorded Depression | Discontinuation vs Continuation | MDD CIDI-sf | Education_levelCollege Postgraduate | 0.198 | 0.020 | 1.945 | 1.65e-01 | 1.00e+00 | 203 | 143 | 60 |
| African | No Recorded Depression | Discontinuation vs Continuation | MDD CIDI-sf | Education_levelCollege Undergraduate | 0.249 | 0.029 | 2.126 | 2.04e-01 | 1.00e+00 | 203 | 143 | 60 |
| African | No Recorded Depression | Discontinuation vs Continuation | MDD CIDI-sf | Education_levelTwelve or GED | 0.164 | 0.015 | 1.813 | 1.40e-01 | 1.00e+00 | 203 | 143 | 60 |
| African | No Recorded Depression | Discontinuation vs Continuation | MDD CIDI-sf | MDD CIDI-sf | 1.903 | 0.962 | 3.765 | 6.44e-02 | 9.02e-01 | 203 | 143 | 60 |
| African | No Recorded Depression | Discontinuation vs Continuation | MDD_symptom_count | (Intercept) | 1.864 | 0.280 | 12.400 | 5.20e-01 | 1.00e+00 | 235 | 166 | 69 |
| African | No Recorded Depression | Discontinuation vs Continuation | MDD_symptom_count | age_at_first_AD | 0.955 | 0.665 | 1.370 | 8.01e-01 | 1.00e+00 | 235 | 166 | 69 |
| African | No Recorded Depression | Discontinuation vs Continuation | MDD_symptom_count | sex_at_birthMale | 2.421 | 1.189 | 4.932 | 1.48e-02 | 2.08e-01 | 235 | 166 | 69 |
| African | No Recorded Depression | Discontinuation vs Continuation | MDD_symptom_count | Income>200k | 1.525 | 0.307 | 7.571 | 6.06e-01 | 1.00e+00 | 235 | 166 | 69 |
| African | No Recorded Depression | Discontinuation vs Continuation | MDD_symptom_count | Income100-150k | 0.814 | 0.266 | 2.485 | 7.17e-01 | 1.00e+00 | 235 | 166 | 69 |
| African | No Recorded Depression | Discontinuation vs Continuation | MDD_symptom_count | Income150-200k | 2.212 | 0.292 | 16.760 | 4.42e-01 | 1.00e+00 | 235 | 166 | 69 |
| African | No Recorded Depression | Discontinuation vs Continuation | MDD_symptom_count | Income25-35k | 0.957 | 0.326 | 2.811 | 9.37e-01 | 1.00e+00 | 235 | 166 | 69 |
| African | No Recorded Depression | Discontinuation vs Continuation | MDD_symptom_count | Income35-50k | 1.566 | 0.583 | 4.212 | 3.74e-01 | 1.00e+00 | 235 | 166 | 69 |
| African | No Recorded Depression | Discontinuation vs Continuation | MDD_symptom_count | Income50-75k | 1.246 | 0.474 | 3.271 | 6.56e-01 | 1.00e+00 | 235 | 166 | 69 |
| African | No Recorded Depression | Discontinuation vs Continuation | MDD_symptom_count | Income75-100k | 3.333 | 1.149 | 9.667 | 2.67e-02 | 3.74e-01 | 235 | 166 | 69 |
| African | No Recorded Depression | Discontinuation vs Continuation | MDD_symptom_count | Education_levelCollege Postgraduate | 0.141 | 0.017 | 1.136 | 6.57e-02 | 9.20e-01 | 235 | 166 | 69 |
| African | No Recorded Depression | Discontinuation vs Continuation | MDD_symptom_count | Education_levelCollege Undergraduate | 0.166 | 0.024 | 1.168 | 7.13e-02 | 9.98e-01 | 235 | 166 | 69 |
| African | No Recorded Depression | Discontinuation vs Continuation | MDD_symptom_count | Education_levelTwelve or GED | 0.085 | 0.009 | 0.796 | 3.08e-02 | 4.31e-01 | 235 | 166 | 69 |
| African | No Recorded Depression | Discontinuation vs Continuation | MDD_symptom_count | MDD_symptom_count | 1.293 | 0.975 | 1.716 | 7.43e-02 | 1.00e+00 | 235 | 166 | 69 |
| African | No Recorded Depression | Discontinuation vs Continuation | Migraine | (Intercept) | 0.432 | 0.345 | 0.542 | 2.99e-13 | 4.19e-12 | 3605 | 2436 | 1169 |
| African | No Recorded Depression | Discontinuation vs Continuation | Migraine | age_at_first_AD | 1.006 | 0.926 | 1.092 | 8.94e-01 | 1.00e+00 | 3605 | 2436 | 1169 |
| African | No Recorded Depression | Discontinuation vs Continuation | Migraine | sex_at_birthMale | 1.163 | 0.995 | 1.360 | 5.75e-02 | 8.05e-01 | 3605 | 2436 | 1169 |

| | | | | | | | | | | | | |
|---|---|---|---|---|---|---|---|---|---|---|---|---|
| African | No Recorded Depression | Discontinuation vs Continuation | Migraine | Income>200k | 0.875 | 0.452 | 1.694 | 6.92e-01 | 1.00e+00 | 3605 | 2436 | 1169 |
| African | No Recorded Depression | Discontinuation vs Continuation | Migraine | Income100-150k | 0.821 | 0.559 | 1.206 | 3.16e-01 | 1.00e+00 | 3605 | 2436 | 1169 |
| African | No Recorded Depression | Discontinuation vs Continuation | Migraine | Income150-200k | 1.800 | 1.029 | 3.150 | 3.94e-02 | 5.52e-01 | 3605 | 2436 | 1169 |
| African | No Recorded Depression | Discontinuation vs Continuation | Migraine | Income25-35k | 1.310 | 1.043 | 1.645 | 2.02e-02 | 2.82e-01 | 3605 | 2436 | 1169 |
| African | No Recorded Depression | Discontinuation vs Continuation | Migraine | Income35-50k | 1.168 | 0.917 | 1.488 | 2.07e-01 | 1.00e+00 | 3605 | 2436 | 1169 |
| African | No Recorded Depression | Discontinuation vs Continuation | Migraine | Income50-75k | 1.129 | 0.879 | 1.450 | 3.41e-01 | 1.00e+00 | 3605 | 2436 | 1169 |
| African | No Recorded Depression | Discontinuation vs Continuation | Migraine | Income75-100k | 0.989 | 0.698 | 1.401 | 9.50e-01 | 1.00e+00 | 3605 | 2436 | 1169 |
| African | No Recorded Depression | Discontinuation vs Continuation | Migraine | Education_levelCollege Postgraduate | 0.882 | 0.626 | 1.242 | 4.71e-01 | 1.00e+00 | 3605 | 2436 | 1169 |
| African | No Recorded Depression | Discontinuation vs Continuation | Migraine | Education_levelCollege Undergraduate | 1.062 | 0.829 | 1.360 | 6.34e-01 | 1.00e+00 | 3605 | 2436 | 1169 |
| African | No Recorded Depression | Discontinuation vs Continuation | Migraine | Education_levelTwelve or GED | 1.085 | 0.844 | 1.396 | 5.24e-01 | 1.00e+00 | 3605 | 2436 | 1169 |
| African | No Recorded Depression | Discontinuation vs Continuation | Migraine | Migraine | 0.826 | 0.692 | 0.986 | 3.41e-02 | 4.78e-01 | 3605 | 2436 | 1169 |
| African | No Recorded Depression | Switching vs Continuation | Migraine | (Intercept) | 0.344 | 0.266 | 0.445 | 5.44e-16 | 7.61e-15 | 3088 | 2436 | 652 |
| African | No Recorded Depression | Switching vs Continuation | Migraine | age_at_first_AD | 0.739 | 0.666 | 0.821 | 1.58e-08 | 2.21e-07 | 3088 | 2436 | 652 |
| African | No Recorded Depression | Switching vs Continuation | Migraine | sex_at_birthMale | 0.916 | 0.746 | 1.125 | 4.01e-01 | 1.00e+00 | 3088 | 2436 | 652 |
| African | No Recorded Depression | Switching vs Continuation | Migraine | Income>200k | 0.825 | 0.383 | 1.780 | 6.24e-01 | 1.00e+00 | 3088 | 2436 | 652 |
| African | No Recorded Depression | Switching vs Continuation | Migraine | Income100-150k | 0.873 | 0.563 | 1.352 | 5.41e-01 | 1.00e+00 | 3088 | 2436 | 652 |
| African | No Recorded Depression | Switching vs Continuation | Migraine | Income150-200k | 0.520 | 0.196 | 1.379 | 1.89e-01 | 1.00e+00 | 3088 | 2436 | 652 |
| African | No Recorded Depression | Switching vs Continuation | Migraine | Income25-35k | 0.654 | 0.470 | 0.910 | 1.17e-02 | 1.63e-01 | 3088 | 2436 | 652 |
| African | No Recorded Depression | Switching vs Continuation | Migraine | Income35-50k | 0.752 | 0.545 | 1.038 | 8.29e-02 | 1.00e+00 | 3088 | 2436 | 652 |
| African | No Recorded Depression | Switching vs Continuation | Migraine | Income50-75k | 0.743 | 0.533 | 1.036 | 7.94e-02 | 1.00e+00 | 3088 | 2436 | 652 |
| African | No Recorded Depression | Switching vs Continuation | Migraine | Income75-100k | 0.804 | 0.517 | 1.249 | 3.31e-01 | 1.00e+00 | 3088 | 2436 | 652 |
| African | No Recorded Depression | Switching vs Continuation | Migraine | Education_levelCollege Postgraduate | 0.781 | 0.517 | 1.178 | 2.39e-01 | 1.00e+00 | 3088 | 2436 | 652 |
| African | No Recorded Depression | Switching vs Continuation | Migraine | Education_levelCollege Undergraduate | 0.746 | 0.559 | 0.996 | 4.71e-02 | 6.59e-01 | 3088 | 2436 | 652 |
| African | No Recorded Depression | Switching vs Continuation | Migraine | Education_levelTwelve or GED | 0.863 | 0.644 | 1.154 | 3.20e-01 | 1.00e+00 | 3088 | 2436 | 652 |
| African | No Recorded Depression | Switching vs Continuation | Migraine | Migraine | 1.393 | 1.141 | 1.700 | 1.11e-03 | 1.56e-02 | 3088 | 2436 | 652 |
| African | No Recorded Depression | Discontinuation vs Continuation | Recorded Anxiety | (Intercept) | 0.437 | 0.348 | 0.548 | 7.37e-13 | 1.03e-11 | 3605 | 2436 | 1169 |
| African | No Recorded Depression | Discontinuation vs Continuation | Recorded Anxiety | age_at_first_AD | 1.005 | 0.926 | 1.091 | 8.97e-01 | 1.00e+00 | 3605 | 2436 | 1169 |
| African | No Recorded Depression | Discontinuation vs Continuation | Recorded Anxiety | sex_at_birthMale | 1.167 | 0.999 | 1.364 | 5.12e-02 | 7.17e-01 | 3605 | 2436 | 1169 |
| African | No Recorded Depression | Discontinuation vs Continuation | Recorded Anxiety | Income>200k | 0.874 | 0.451 | 1.692 | 6.89e-01 | 1.00e+00 | 3605 | 2436 | 1169 |
| African | No Recorded Depression | Discontinuation vs Continuation | Recorded Anxiety | Income100-150k | 0.818 | 0.557 | 1.200 | 3.04e-01 | 1.00e+00 | 3605 | 2436 | 1169 |
| African | No Recorded Depression | Discontinuation vs Continuation | Recorded Anxiety | Income150-200k | 1.762 | 1.007 | 3.083 | 4.71e-02 | 6.60e-01 | 3605 | 2436 | 1169 |
| African | No Recorded Depression | Discontinuation vs Continuation | Recorded Anxiety | Income25-35k | 1.318 | 1.050 | 1.655 | 1.74e-02 | 2.44e-01 | 3605 | 2436 | 1169 |
| African | No Recorded Depression | Discontinuation vs Continuation | Recorded Anxiety | Income35-50k | 1.171 | 0.919 | 1.492 | 2.01e-01 | 1.00e+00 | 3605 | 2436 | 1169 |
| African | No Recorded Depression | Discontinuation vs Continuation | Recorded Anxiety | Income50-75k | 1.127 | 0.878 | 1.447 | 3.48e-01 | 1.00e+00 | 3605 | 2436 | 1169 |
| African | No Recorded Depression | Discontinuation vs Continuation | Recorded Anxiety | Income75-100k | 0.998 | 0.704 | 1.414 | 9.90e-01 | 1.00e+00 | 3605 | 2436 | 1169 |
| African | No Recorded Depression | Discontinuation vs Continuation | Recorded Anxiety | Education_levelCollege Postgraduate | 0.899 | 0.638 | 1.267 | 5.44e-01 | 1.00e+00 | 3605 | 2436 | 1169 |
| African | No Recorded Depression | Discontinuation vs Continuation | Recorded Anxiety | Education_levelCollege Undergraduate | 1.067 | 0.833 | 1.367 | 6.07e-01 | 1.00e+00 | 3605 | 2436 | 1169 |
| African | No Recorded Depression | Discontinuation vs Continuation | Recorded Anxiety | Education_levelTwelve or GED | 1.093 | 0.850 | 1.407 | 4.87e-01 | 1.00e+00 | 3605 | 2436 | 1169 |
| African | No Recorded Depression | Discontinuation vs Continuation | Recorded Anxiety | Recorded Anxiety | 0.837 | 0.719 | 0.975 | 2.21e-02 | 3.10e-01 | 3605 | 2436 | 1169 |
| African | No Recorded Depression | Switching vs Continuation | Recorded Anxiety | (Intercept) | 0.298 | 0.229 | 0.387 | <2.2e-16 | <2.2e-16 | 3088 | 2436 | 652 |
| African | No Recorded Depression | Switching vs Continuation | Recorded Anxiety | age_at_first_AD | 0.752 | 0.677 | 0.834 | 8.57e-08 | 1.20e-06 | 3088 | 2436 | 652 |
| African | No Recorded Depression | Switching vs Continuation | Recorded Anxiety | sex_at_birthMale | 0.925 | 0.754 | 1.137 | 4.60e-01 | 1.00e+00 | 3088 | 2436 | 652 |
| African | No Recorded Depression | Switching vs Continuation | Recorded Anxiety | Income>200k | 0.824 | 0.380 | 1.788 | 6.24e-01 | 1.00e+00 | 3088 | 2436 | 652 |
| African | No Recorded Depression | Switching vs Continuation | Recorded Anxiety | Income100-150k | 0.836 | 0.538 | 1.299 | 4.25e-01 | 1.00e+00 | 3088 | 2436 | 652 |
| African | No Recorded Depression | Switching vs Continuation | Recorded Anxiety | Income150-200k | 0.532 | 0.200 | 1.418 | 2.07e-01 | 1.00e+00 | 3088 | 2436 | 652 |
| African | No Recorded Depression | Switching vs Continuation | Recorded Anxiety | Income25-35k | 0.628 | 0.450 | 0.875 | 6.07e-03 | 8.50e-02 | 3088 | 2436 | 652 |
| African | No Recorded Depression | Switching vs Continuation | Recorded Anxiety | Income35-50k | 0.725 | 0.525 | 1.001 | 5.09e-02 | 7.12e-01 | 3088 | 2436 | 652 |
| African | No Recorded Depression | Switching vs Continuation | Recorded Anxiety | Income50-75k | 0.731 | 0.523 | 1.022 | 6.66e-02 | 9.33e-01 | 3088 | 2436 | 652 |
| African | No Recorded Depression | Switching vs Continuation | Recorded Anxiety | Income75-100k | 0.774 | 0.497 | 1.207 | 2.58e-01 | 1.00e+00 | 3088 | 2436 | 652 |
| African | No Recorded Depression | Switching vs Continuation | Recorded Anxiety | Education_levelCollege Postgraduate | 0.726 | 0.479 | 1.099 | 1.30e-01 | 1.00e+00 | 3088 | 2436 | 652 |
| African | No Recorded Depression | Switching vs Continuation | Recorded Anxiety | Education_levelCollege Undergraduate | 0.723 | 0.540 | 0.967 | 2.90e-02 | 4.06e-01 | 3088 | 2436 | 652 |
| African | No Recorded Depression | Switching vs Continuation | Recorded Anxiety | Education_levelTwelve or GED | 0.842 | 0.628 | 1.129 | 2.51e-01 | 1.00e+00 | 3088 | 2436 | 652 |
| African | No Recorded Depression | Switching vs Continuation | Recorded Anxiety | Recorded Anxiety | 1.890 | 1.578 | 2.263 | 4.53e-12 | 6.35e-11 | 3088 | 2436 | 652 |
| African | No Recorded Depression | Discontinuation vs Continuation | Smoking_Frequency | (Intercept) | 0.403 | 0.318 | 0.512 | 8.44e-14 | 1.18e-12 | 3528 | 2391 | 1137 |
| African | No Recorded Depression | Discontinuation vs Continuation | Smoking_Frequency | age_at_first_AD | 1.043 | 0.958 | 1.135 | 3.31e-01 | 1.00e+00 | 3528 | 2391 | 1137 |
| African | No Recorded Depression | Discontinuation vs Continuation | Smoking_Frequency | sex_at_birthMale | 1.189 | 1.012 | 1.396 | 3.47e-02 | 4.86e-01 | 3528 | 2391 | 1137 |
| African | No Recorded Depression | Discontinuation vs Continuation | Smoking_Frequency | Income>200k | 0.852 | 0.430 | 1.685 | 6.44e-01 | 1.00e+00 | 3528 | 2391 | 1137 |
| African | No Recorded Depression | Discontinuation vs Continuation | Smoking_Frequency | Income100-150k | 0.845 | 0.574 | 1.243 | 3.91e-01 | 1.00e+00 | 3528 | 2391 | 1137 |
| African | No Recorded Depression | Discontinuation vs Continuation | Smoking_Frequency | Income150-200k | 1.685 | 0.949 | 2.993 | 7.49e-02 | 1.00e+00 | 3528 | 2391 | 1137 |
| African | No Recorded Depression | Discontinuation vs Continuation | Smoking_Frequency | Income25-35k | 1.314 | 1.042 | 1.657 | 2.10e-02 | 2.94e-01 | 3528 | 2391 | 1137 |
| African | No Recorded Depression | Discontinuation vs Continuation | Smoking_Frequency | Income35-50k | 1.195 | 0.935 | 1.527 | 1.54e-01 | 1.00e+00 | 3528 | 2391 | 1137 |
| African | No Recorded Depression | Discontinuation vs Continuation | Smoking_Frequency | Income50-75k | 1.107 | 0.857 | 1.429 | 4.37e-01 | 1.00e+00 | 3528 | 2391 | 1137 |
| African | No Recorded Depression | Discontinuation vs Continuation | Smoking_Frequency | Income75-100k | 1.022 | 0.718 | 1.455 | 9.04e-01 | 1.00e+00 | 3528 | 2391 | 1137 |
| African | No Recorded Depression | Discontinuation vs Continuation | Smoking_Frequency | Education_levelCollege Postgraduate | 0.889 | 0.626 | 1.261 | 5.09e-01 | 1.00e+00 | 3528 | 2391 | 1137 |
| African | No Recorded Depression | Discontinuation vs Continuation | Smoking_Frequency | Education_levelCollege Undergraduate | 1.097 | 0.853 | 1.410 | 4.73e-01 | 1.00e+00 | 3528 | 2391 | 1137 |
| African | No Recorded Depression | Discontinuation vs Continuation | Smoking_Frequency | Education_levelTwelve or GED | 1.105 | 0.855 | 1.428 | 4.45e-01 | 1.00e+00 | 3528 | 2391 | 1137 |

| | | | | | | | | | | | | |
|---|---|---|---|---|---|---|---|---|---|---|---|---|
| African | No Recorded Depression | Discontinuation vs Continuation | Smoking_Frequency | Smoking_FrequencyEvery Day | 1.152 | 0.935 | 1.420 | 1.83e-01 | 1.00e+00 | 3528 | 2391 | 1137 |
| African | No Recorded Depression | Discontinuation vs Continuation | Smoking_Frequency | Smoking_FrequencyNot At All (former) | 0.796 | 0.652 | 0.970 | 2.40e-02 | 3.37e-01 | 3528 | 2391 | 1137 |
| African | No Recorded Depression | Discontinuation vs Continuation | Smoking_Frequency | Smoking_FrequencySome Days | 1.076 | 0.837 | 1.383 | 5.70e-01 | 1.00e+00 | 3528 | 2391 | 1137 |
| African | No Recorded Depression | Switching vs Continuation | Smoking_Frequency | (Intercept) | 0.338 | 0.256 | 0.447 | 2.66e-14 | 3.72e-13 | 3017 | 2391 | 626 |
| African | No Recorded Depression | Switching vs Continuation | Smoking_Frequency | age_at_first_AD | 0.718 | 0.644 | 0.801 | 2.33e-09 | 3.27e-08 | 3017 | 2391 | 626 |
| African | No Recorded Depression | Switching vs Continuation | Smoking_Frequency | sex_at_birthMale | 0.865 | 0.700 | 1.069 | 1.80e-01 | 1.00e+00 | 3017 | 2391 | 626 |
| African | No Recorded Depression | Switching vs Continuation | Smoking_Frequency | Income>200k | 0.687 | 0.295 | 1.599 | 3.83e-01 | 1.00e+00 | 3017 | 2391 | 626 |
| African | No Recorded Depression | Switching vs Continuation | Smoking_Frequency | Income100-150k | 0.928 | 0.597 | 1.440 | 7.38e-01 | 1.00e+00 | 3017 | 2391 | 626 |
| African | No Recorded Depression | Switching vs Continuation | Smoking_Frequency | Income150-200k | 0.547 | 0.206 | 1.451 | 2.25e-01 | 1.00e+00 | 3017 | 2391 | 626 |
| African | No Recorded Depression | Switching vs Continuation | Smoking_Frequency | Income25-35k | 0.672 | 0.481 | 0.939 | 1.99e-02 | 2.79e-01 | 3017 | 2391 | 626 |
| African | No Recorded Depression | Switching vs Continuation | Smoking_Frequency | Income35-50k | 0.774 | 0.558 | 1.074 | 1.25e-01 | 1.00e+00 | 3017 | 2391 | 626 |
| African | No Recorded Depression | Switching vs Continuation | Smoking_Frequency | Income50-75k | 0.754 | 0.537 | 1.058 | 1.03e-01 | 1.00e+00 | 3017 | 2391 | 626 |
| African | No Recorded Depression | Switching vs Continuation | Smoking_Frequency | Income75-100k | 0.783 | 0.497 | 1.233 | 2.91e-01 | 1.00e+00 | 3017 | 2391 | 626 |
| African | No Recorded Depression | Switching vs Continuation | Smoking_Frequency | Education_levelCollege Postgraduate | 0.870 | 0.570 | 1.326 | 5.16e-01 | 1.00e+00 | 3017 | 2391 | 626 |
| African | No Recorded Depression | Switching vs Continuation | Smoking_Frequency | Education_levelCollege Undergraduate | 0.800 | 0.594 | 1.076 | 1.40e-01 | 1.00e+00 | 3017 | 2391 | 626 |
| African | No Recorded Depression | Switching vs Continuation | Smoking_Frequency | Education_levelTwelve or GED | 0.884 | 0.655 | 1.192 | 4.18e-01 | 1.00e+00 | 3017 | 2391 | 626 |
| African | No Recorded Depression | Switching vs Continuation | Smoking_Frequency | Smoking_FrequencyEvery Day | 1.284 | 0.995 | 1.657 | 5.50e-02 | 7.70e-01 | 3017 | 2391 | 626 |
| African | No Recorded Depression | Switching vs Continuation | Smoking_Frequency | Smoking_FrequencyNot At All (former) | 1.017 | 0.791 | 1.308 | 8.96e-01 | 1.00e+00 | 3017 | 2391 | 626 |
| African | No Recorded Depression | Switching vs Continuation | Smoking_Frequency | Smoking_FrequencySome Days | 0.952 | 0.682 | 1.331 | 7.75e-01 | 1.00e+00 | 3017 | 2391 | 626 |
| African | No Recorded Depression | Discontinuation vs Continuation | Stress | (Intercept) | 0.393 | 0.210 | 0.736 | 3.51e-03 | 4.91e-02 | 1092 | 748 | 344 |
| African | No Recorded Depression | Discontinuation vs Continuation | Stress | age_at_first_AD | 1.189 | 1.022 | 1.382 | 2.46e-02 | 3.45e-01 | 1092 | 748 | 344 |
| African | No Recorded Depression | Discontinuation vs Continuation | Stress | sex_at_birthMale | 1.280 | 0.940 | 1.742 | 1.17e-01 | 1.00e+00 | 1092 | 748 | 344 |
| African | No Recorded Depression | Discontinuation vs Continuation | Stress | Income>200k | 1.229 | 0.506 | 2.981 | 6.49e-01 | 1.00e+00 | 1092 | 748 | 344 |
| African | No Recorded Depression | Discontinuation vs Continuation | Stress | Income100-150k | 1.036 | 0.605 | 1.775 | 8.97e-01 | 1.00e+00 | 1092 | 748 | 344 |
| African | No Recorded Depression | Discontinuation vs Continuation | Stress | Income150-200k | 2.639 | 1.256 | 5.543 | 1.04e-02 | 1.46e-01 | 1092 | 748 | 344 |
| African | No Recorded Depression | Discontinuation vs Continuation | Stress | Income25-35k | 1.566 | 1.008 | 2.433 | 4.61e-02 | 6.46e-01 | 1092 | 748 | 344 |
| African | No Recorded Depression | Discontinuation vs Continuation | Stress | Income35-50k | 1.431 | 0.940 | 2.179 | 9.47e-02 | 1.00e+00 | 1092 | 748 | 344 |
| African | No Recorded Depression | Discontinuation vs Continuation | Stress | Income50-75k | 1.291 | 0.856 | 1.947 | 2.23e-01 | 1.00e+00 | 1092 | 748 | 344 |
| African | No Recorded Depression | Discontinuation vs Continuation | Stress | Income75-100k | 1.149 | 0.675 | 1.955 | 6.09e-01 | 1.00e+00 | 1092 | 748 | 344 |
| African | No Recorded Depression | Discontinuation vs Continuation | Stress | Education_levelCollege Postgraduate | 0.644 | 0.313 | 1.327 | 2.33e-01 | 1.00e+00 | 1092 | 748 | 344 |
| African | No Recorded Depression | Discontinuation vs Continuation | Stress | Education_levelCollege Undergraduate | 0.754 | 0.398 | 1.428 | 3.86e-01 | 1.00e+00 | 1092 | 748 | 344 |
| African | No Recorded Depression | Discontinuation vs Continuation | Stress | Education_levelTwelve or GED | 0.927 | 0.474 | 1.815 | 8.25e-01 | 1.00e+00 | 1092 | 748 | 344 |
| African | No Recorded Depression | Discontinuation vs Continuation | Stress | StressAlmost Never | 1.206 | 0.638 | 2.282 | 5.64e-01 | 1.00e+00 | 1092 | 748 | 344 |
| African | No Recorded Depression | Discontinuation vs Continuation | Stress | StressFairly Often | 1.064 | 0.558 | 2.027 | 8.51e-01 | 1.00e+00 | 1092 | 748 | 344 |
| African | No Recorded Depression | Discontinuation vs Continuation | Stress | StressSometimes | 1.336 | 1.008 | 1.772 | 4.41e-02 | 6.17e-01 | 1092 | 748 | 344 |
| African | No Recorded Depression | Discontinuation vs Continuation | Stress | StressVery Often | 1.324 | 0.675 | 2.599 | 4.15e-01 | 1.00e+00 | 1092 | 748 | 344 |
| African | No Recorded Depression | Switching vs Continuation | Stress | (Intercept) | 0.456 | 0.232 | 0.893 | 2.21e-02 | 3.10e-01 | 913 | 748 | 165 |
| African | No Recorded Depression | Switching vs Continuation | Stress | age_at_first_AD | 0.645 | 0.525 | 0.793 | 3.17e-05 | 4.44e-04 | 913 | 748 | 165 |
| African | No Recorded Depression | Switching vs Continuation | Stress | sex_at_birthMale | 0.800 | 0.498 | 1.285 | 3.57e-01 | 1.00e+00 | 913 | 748 | 165 |
| African | No Recorded Depression | Switching vs Continuation | Stress | Income>200k | 0.943 | 0.297 | 2.995 | 9.20e-01 | 1.00e+00 | 913 | 748 | 165 |
| African | No Recorded Depression | Switching vs Continuation | Stress | Income100-150k | 1.246 | 0.642 | 2.418 | 5.16e-01 | 1.00e+00 | 913 | 748 | 165 |
| African | No Recorded Depression | Switching vs Continuation | Stress | Income150-200k | 0.744 | 0.200 | 2.774 | 6.60e-01 | 1.00e+00 | 913 | 748 | 165 |
| African | No Recorded Depression | Switching vs Continuation | Stress | Income25-35k | 0.536 | 0.257 | 1.120 | 9.72e-02 | 1.00e+00 | 913 | 748 | 165 |
| African | No Recorded Depression | Switching vs Continuation | Stress | Income35-50k | 1.185 | 0.681 | 2.059 | 5.48e-01 | 1.00e+00 | 913 | 748 | 165 |
| African | No Recorded Depression | Switching vs Continuation | Stress | Income50-75k | 0.862 | 0.485 | 1.530 | 6.12e-01 | 1.00e+00 | 913 | 748 | 165 |
| African | No Recorded Depression | Switching vs Continuation | Stress | Income75-100k | 1.054 | 0.531 | 2.090 | 8.81e-01 | 1.00e+00 | 913 | 748 | 165 |
| African | No Recorded Depression | Switching vs Continuation | Stress | Education_levelCollege Postgraduate | 0.398 | 0.178 | 0.889 | 2.47e-02 | 3.46e-01 | 913 | 748 | 165 |
| African | No Recorded Depression | Switching vs Continuation | Stress | Education_levelCollege Undergraduate | 0.321 | 0.161 | 0.642 | 1.30e-03 | 1.83e-02 | 913 | 748 | 165 |
| African | No Recorded Depression | Switching vs Continuation | Stress | Education_levelTwelve or GED | 0.373 | 0.176 | 0.789 | 9.91e-03 | 1.39e-01 | 913 | 748 | 165 |
| African | No Recorded Depression | Switching vs Continuation | Stress | StressAlmost Never | 2.681 | 1.316 | 5.464 | 6.61e-03 | 9.26e-02 | 913 | 748 | 165 |
| African | No Recorded Depression | Switching vs Continuation | Stress | StressFairly Often | 1.186 | 0.494 | 2.845 | 7.03e-01 | 1.00e+00 | 913 | 748 | 165 |
| African | No Recorded Depression | Switching vs Continuation | Stress | StressSometimes | 1.771 | 1.208 | 2.597 | 3.43e-03 | 4.81e-02 | 913 | 748 | 165 |
| African | No Recorded Depression | Switching vs Continuation | Stress | StressVery Often | 0.939 | 0.341 | 2.586 | 9.03e-01 | 1.00e+00 | 913 | 748 | 165 |
| African | No Recorded Depression | Discontinuation vs Continuation | age_at_first_AD | (Intercept) | 0.456 | 0.420 | 0.496 | <2.2e-16 | <2.2e-16 | 3605 | 2436 | 1169 |
| African | No Recorded Depression | Discontinuation vs Continuation | age_at_first_AD | age_at_first_AD | 1.012 | 0.933 | 1.097 | 7.79e-01 | 1.00e+00 | 3605 | 2436 | 1169 |
| African | No Recorded Depression | Discontinuation vs Continuation | age_at_first_AD | sex_at_birthMale | 1.181 | 1.013 | 1.376 | 3.32e-02 | 4.65e-01 | 3605 | 2436 | 1169 |
| African | No Recorded Depression | Switching vs Continuation | age_at_first_AD | (Intercept) | 0.270 | 0.244 | 0.299 | <2.2e-16 | <2.2e-16 | 3088 | 2436 | 652 |
| African | No Recorded Depression | Switching vs Continuation | age_at_first_AD | age_at_first_AD | 0.734 | 0.663 | 0.812 | 2.50e-09 | 3.50e-08 | 3088 | 2436 | 652 |
| African | No Recorded Depression | Switching vs Continuation | age_at_first_AD | sex_at_birthMale | 0.922 | 0.754 | 1.127 | 4.27e-01 | 1.00e+00 | 3088 | 2436 | 652 |
| African | No Recorded Depression | Discontinuation vs Continuation | sex_at_birth | (Intercept) | 0.456 | 0.420 | 0.496 | <2.2e-16 | <2.2e-16 | 3605 | 2436 | 1169 |
| African | No Recorded Depression | Discontinuation vs Continuation | sex_at_birth | age_at_first_AD | 1.012 | 0.933 | 1.097 | 7.79e-01 | 1.00e+00 | 3605 | 2436 | 1169 |
| African | No Recorded Depression | Discontinuation vs Continuation | sex_at_birth | sex_at_birthMale | 1.181 | 1.013 | 1.376 | 3.32e-02 | 4.65e-01 | 3605 | 2436 | 1169 |
| African | No Recorded Depression | Switching vs Continuation | sex_at_birth | (Intercept) | 0.270 | 0.244 | 0.299 | <2.2e-16 | <2.2e-16 | 3088 | 2436 | 652 |
| African | No Recorded Depression | Switching vs Continuation | sex_at_birth | age_at_first_AD | 0.734 | 0.663 | 0.812 | 2.50e-09 | 3.50e-08 | 3088 | 2436 | 652 |
| African | No Recorded Depression | Switching vs Continuation | sex_at_birth | sex_at_birthMale | 0.922 | 0.754 | 1.127 | 4.27e-01 | 1.00e+00 | 3088 | 2436 | 652 |
| African | Recorded Anxiety | Discontinuation vs Continuation | Alcohol_Frequency | (Intercept) | 0.424 | 0.298 | 0.602 | 1.75e-06 | 2.45e-05 | 2721 | 1961 | 760 |

| | | | | | | | | | | | | |
|---|---|---|---|---|---|---|---|---|---|---|---|---|
| African | Recorded Anxiety | Discontinuation vs Continuation | Alcohol_Frequency | age_at_first_AD | 0.958 | 0.869 | 1.055 | 3.82e-01 | 1.00e+00 | 2721 | 1961 | 760 |
| African | Recorded Anxiety | Discontinuation vs Continuation | Alcohol_Frequency | sex_at_birthMale | 1.226 | 0.999 | 1.503 | 5.07e-02 | 7.09e-01 | 2721 | 1961 | 760 |
| African | Recorded Anxiety | Discontinuation vs Continuation | Alcohol_Frequency | Income>200k | 0.582 | 0.214 | 1.581 | 2.89e-01 | 1.00e+00 | 2721 | 1961 | 760 |
| African | Recorded Anxiety | Discontinuation vs Continuation | Alcohol_Frequency | Income100-150k | 0.482 | 0.292 | 0.798 | 4.51e-03 | 6.32e-02 | 2721 | 1961 | 760 |
| African | Recorded Anxiety | Discontinuation vs Continuation | Alcohol_Frequency | Income150-200k | 0.836 | 0.384 | 1.820 | 6.51e-01 | 1.00e+00 | 2721 | 1961 | 760 |
| African | Recorded Anxiety | Discontinuation vs Continuation | Alcohol_Frequency | Income25-35k | 1.116 | 0.854 | 1.459 | 4.21e-01 | 1.00e+00 | 2721 | 1961 | 760 |
| African | Recorded Anxiety | Discontinuation vs Continuation | Alcohol_Frequency | Income35-50k | 0.715 | 0.534 | 0.958 | 2.44e-02 | 3.42e-01 | 2721 | 1961 | 760 |
| African | Recorded Anxiety | Discontinuation vs Continuation | Alcohol_Frequency | Income50-75k | 1.036 | 0.777 | 1.380 | 8.11e-01 | 1.00e+00 | 2721 | 1961 | 760 |
| African | Recorded Anxiety | Discontinuation vs Continuation | Alcohol_Frequency | Income75-100k | 0.615 | 0.397 | 0.951 | 2.90e-02 | 4.06e-01 | 2721 | 1961 | 760 |
| African | Recorded Anxiety | Discontinuation vs Continuation | Alcohol_Frequency | Education_levelCollege Postgraduate | 0.876 | 0.573 | 1.338 | 5.39e-01 | 1.00e+00 | 2721 | 1961 | 760 |
| African | Recorded Anxiety | Discontinuation vs Continuation | Alcohol_Frequency | Education_levelCollege Undergraduate | 0.964 | 0.699 | 1.329 | 8.22e-01 | 1.00e+00 | 2721 | 1961 | 760 |
| African | Recorded Anxiety | Discontinuation vs Continuation | Alcohol_Frequency | Education_levelTwelve or GED | 0.975 | 0.701 | 1.356 | 8.80e-01 | 1.00e+00 | 2721 | 1961 | 760 |
| African | Recorded Anxiety | Discontinuation vs Continuation | Alcohol_Frequency | Alcohol_Frequency2-3 Week | 0.942 | 0.651 | 1.363 | 7.51e-01 | 1.00e+00 | 2721 | 1961 | 760 |
| African | Recorded Anxiety | Discontinuation vs Continuation | Alcohol_Frequency | Alcohol_Frequency2-4 Month | 0.983 | 0.726 | 1.332 | 9.13e-01 | 1.00e+00 | 2721 | 1961 | 760 |
| African | Recorded Anxiety | Discontinuation vs Continuation | Alcohol_Frequency | Alcohol_Frequency4plus Week | 0.792 | 0.517 | 1.212 | 2.83e-01 | 1.00e+00 | 2721 | 1961 | 760 |
| African | Recorded Anxiety | Discontinuation vs Continuation | Alcohol_Frequency | Alcohol_FrequencyMonthly or Less | 1.004 | 0.766 | 1.316 | 9.77e-01 | 1.00e+00 | 2721 | 1961 | 760 |
| African | Recorded Anxiety | Discontinuation vs Continuation | Alcohol_Frequency | Alcohol_FrequencyNever (former) | 0.996 | 0.726 | 1.367 | 9.81e-01 | 1.00e+00 | 2721 | 1961 | 760 |
| African | Recorded Anxiety | Switching vs Continuation | Alcohol_Frequency | (Intercept) | 1.156 | 0.887 | 1.507 | 2.83e-01 | 1.00e+00 | 3810 | 1961 | 1849 |
| African | Recorded Anxiety | Switching vs Continuation | Alcohol_Frequency | age_at_first_AD | 0.786 | 0.727 | 0.851 | 2.35e-09 | 3.29e-08 | 3810 | 1961 | 1849 |
| African | Recorded Anxiety | Switching vs Continuation | Alcohol_Frequency | sex_at_birthMale | 1.048 | 0.892 | 1.233 | 5.68e-01 | 1.00e+00 | 3810 | 1961 | 1849 |
| African | Recorded Anxiety | Switching vs Continuation | Alcohol_Frequency | Income>200k | 0.586 | 0.292 | 1.175 | 1.32e-01 | 1.00e+00 | 3810 | 1961 | 1849 |
| African | Recorded Anxiety | Switching vs Continuation | Alcohol_Frequency | Income100-150k | 0.591 | 0.424 | 0.822 | 1.80e-03 | 2.52e-02 | 3810 | 1961 | 1849 |
| African | Recorded Anxiety | Switching vs Continuation | Alcohol_Frequency | Income150-200k | 0.524 | 0.277 | 0.991 | 4.68e-02 | 6.55e-01 | 3810 | 1961 | 1849 |
| African | Recorded Anxiety | Switching vs Continuation | Alcohol_Frequency | Income25-35k | 0.740 | 0.595 | 0.919 | 6.54e-03 | 9.16e-02 | 3810 | 1961 | 1849 |
| African | Recorded Anxiety | Switching vs Continuation | Alcohol_Frequency | Income35-50k | 0.731 | 0.591 | 0.904 | 3.76e-03 | 5.26e-02 | 3810 | 1961 | 1849 |
| African | Recorded Anxiety | Switching vs Continuation | Alcohol_Frequency | Income50-75k | 0.674 | 0.534 | 0.851 | 9.05e-04 | 1.27e-02 | 3810 | 1961 | 1849 |
| African | Recorded Anxiety | Switching vs Continuation | Alcohol_Frequency | Income75-100k | 0.597 | 0.436 | 0.817 | 1.28e-03 | 1.79e-02 | 3810 | 1961 | 1849 |
| African | Recorded Anxiety | Switching vs Continuation | Alcohol_Frequency | Education_levelCollege Postgraduate | 0.926 | 0.673 | 1.275 | 6.39e-01 | 1.00e+00 | 3810 | 1961 | 1849 |
| African | Recorded Anxiety | Switching vs Continuation | Alcohol_Frequency | Education_levelCollege Undergraduate | 0.944 | 0.739 | 1.206 | 6.45e-01 | 1.00e+00 | 3810 | 1961 | 1849 |
| African | Recorded Anxiety | Switching vs Continuation | Alcohol_Frequency | Education_levelTwelve or GED | 0.852 | 0.662 | 1.098 | 2.16e-01 | 1.00e+00 | 3810 | 1961 | 1849 |
| African | Recorded Anxiety | Switching vs Continuation | Alcohol_Frequency | Alcohol_Frequency2-3 Week | 0.939 | 0.708 | 1.246 | 6.64e-01 | 1.00e+00 | 3810 | 1961 | 1849 |
| African | Recorded Anxiety | Switching vs Continuation | Alcohol_Frequency | Alcohol_Frequency2-4 Month | 0.889 | 0.703 | 1.124 | 3.27e-01 | 1.00e+00 | 3810 | 1961 | 1849 |
| African | Recorded Anxiety | Switching vs Continuation | Alcohol_Frequency | Alcohol_Frequency4plus Week | 0.903 | 0.658 | 1.239 | 5.26e-01 | 1.00e+00 | 3810 | 1961 | 1849 |
| African | Recorded Anxiety | Switching vs Continuation | Alcohol_Frequency | Alcohol_FrequencyMonthly or Less | 0.946 | 0.768 | 1.166 | 6.03e-01 | 1.00e+00 | 3810 | 1961 | 1849 |
| African | Recorded Anxiety | Switching vs Continuation | Alcohol_Frequency | Alcohol_FrequencyNever (former) | 1.271 | 1.003 | 1.612 | 4.76e-02 | 6.67e-01 | 3810 | 1961 | 1849 |
| African | Recorded Anxiety | Discontinuation vs Continuation | BMI | (Intercept) | 0.401 | 0.297 | 0.541 | 2.27e-09 | 3.18e-08 | 2745 | 1991 | 754 |
| African | Recorded Anxiety | Discontinuation vs Continuation | BMI | age_at_first_AD | 0.934 | 0.849 | 1.028 | 1.61e-01 | 1.00e+00 | 2745 | 1991 | 754 |
| African | Recorded Anxiety | Discontinuation vs Continuation | BMI | sex_at_birthMale | 1.151 | 0.938 | 1.412 | 1.79e-01 | 1.00e+00 | 2745 | 1991 | 754 |
| African | Recorded Anxiety | Discontinuation vs Continuation | BMI | Income>200k | 0.533 | 0.197 | 1.443 | 2.16e-01 | 1.00e+00 | 2745 | 1991 | 754 |
| African | Recorded Anxiety | Discontinuation vs Continuation | BMI | Income100-150k | 0.494 | 0.299 | 0.818 | 6.15e-03 | 8.61e-02 | 2745 | 1991 | 754 |
| African | Recorded Anxiety | Discontinuation vs Continuation | BMI | Income150-200k | 0.910 | 0.430 | 1.926 | 8.06e-01 | 1.00e+00 | 2745 | 1991 | 754 |
| African | Recorded Anxiety | Discontinuation vs Continuation | BMI | Income25-35k | 1.108 | 0.846 | 1.450 | 4.56e-01 | 1.00e+00 | 2745 | 1991 | 754 |
| African | Recorded Anxiety | Discontinuation vs Continuation | BMI | Income35-50k | 0.707 | 0.528 | 0.947 | 2.01e-02 | 2.82e-01 | 2745 | 1991 | 754 |
| African | Recorded Anxiety | Discontinuation vs Continuation | BMI | Income50-75k | 1.002 | 0.752 | 1.335 | 9.89e-01 | 1.00e+00 | 2745 | 1991 | 754 |
| African | Recorded Anxiety | Discontinuation vs Continuation | BMI | Income75-100k | 0.552 | 0.351 | 0.868 | 1.01e-02 | 1.41e-01 | 2745 | 1991 | 754 |
| African | Recorded Anxiety | Discontinuation vs Continuation | BMI | Education_levelCollege Postgraduate | 0.923 | 0.602 | 1.415 | 7.12e-01 | 1.00e+00 | 2745 | 1991 | 754 |
| African | Recorded Anxiety | Discontinuation vs Continuation | BMI | Education_levelCollege Undergraduate | 1.025 | 0.742 | 1.416 | 8.80e-01 | 1.00e+00 | 2745 | 1991 | 754 |
| African | Recorded Anxiety | Discontinuation vs Continuation | BMI | Education_levelTwelve or GED | 1.046 | 0.750 | 1.458 | 7.91e-01 | 1.00e+00 | 2745 | 1991 | 754 |
| African | Recorded Anxiety | Discontinuation vs Continuation | BMI | BMI | 0.920 | 0.851 | 0.994 | 3.56e-02 | 4.98e-01 | 2745 | 1991 | 754 |
| African | Recorded Anxiety | Switching vs Continuation | BMI | (Intercept) | 1.140 | 0.914 | 1.420 | 2.45e-01 | 1.00e+00 | 3866 | 1991 | 1875 |
| African | Recorded Anxiety | Switching vs Continuation | BMI | age_at_first_AD | 0.796 | 0.737 | 0.860 | 6.99e-09 | 9.79e-08 | 3866 | 1991 | 1875 |
| African | Recorded Anxiety | Switching vs Continuation | BMI | sex_at_birthMale | 1.074 | 0.917 | 1.259 | 3.76e-01 | 1.00e+00 | 3866 | 1991 | 1875 |
| African | Recorded Anxiety | Switching vs Continuation | BMI | Income>200k | 0.538 | 0.270 | 1.072 | 7.82e-02 | 1.00e+00 | 3866 | 1991 | 1875 |
| African | Recorded Anxiety | Switching vs Continuation | BMI | Income100-150k | 0.570 | 0.408 | 0.795 | 9.26e-04 | 1.30e-02 | 3866 | 1991 | 1875 |
| African | Recorded Anxiety | Switching vs Continuation | BMI | Income150-200k | 0.593 | 0.321 | 1.096 | 9.53e-02 | 1.00e+00 | 3866 | 1991 | 1875 |
| African | Recorded Anxiety | Switching vs Continuation | BMI | Income25-35k | 0.762 | 0.614 | 0.946 | 1.37e-02 | 1.92e-01 | 3866 | 1991 | 1875 |
| African | Recorded Anxiety | Switching vs Continuation | BMI | Income35-50k | 0.734 | 0.595 | 0.906 | 4.01e-03 | 5.61e-02 | 3866 | 1991 | 1875 |
| African | Recorded Anxiety | Switching vs Continuation | BMI | Income50-75k | 0.656 | 0.521 | 0.826 | 3.41e-04 | 4.77e-03 | 3866 | 1991 | 1875 |
| African | Recorded Anxiety | Switching vs Continuation | BMI | Income75-100k | 0.569 | 0.416 | 0.779 | 4.30e-04 | 6.02e-03 | 3866 | 1991 | 1875 |
| African | Recorded Anxiety | Switching vs Continuation | BMI | Education_levelCollege Postgraduate | 0.937 | 0.685 | 1.283 | 6.85e-01 | 1.00e+00 | 3866 | 1991 | 1875 |
| African | Recorded Anxiety | Switching vs Continuation | BMI | Education_levelCollege Undergraduate | 0.945 | 0.744 | 1.199 | 6.39e-01 | 1.00e+00 | 3866 | 1991 | 1875 |
| African | Recorded Anxiety | Switching vs Continuation | BMI | Education_levelTwelve or GED | 0.859 | 0.670 | 1.101 | 2.29e-01 | 1.00e+00 | 3866 | 1991 | 1875 |
| African | Recorded Anxiety | Switching vs Continuation | BMI | BMI | 0.996 | 0.941 | 1.054 | 8.91e-01 | 1.00e+00 | 3866 | 1991 | 1875 |
| African | Recorded Anxiety | Discontinuation vs Continuation | Education_level | (Intercept) | 0.407 | 0.304 | 0.544 | 1.42e-09 | 1.99e-08 | 2793 | 2014 | 779 |
| African | Recorded Anxiety | Discontinuation vs Continuation | Education_level | age_at_first_AD | 0.943 | 0.858 | 1.036 | 2.24e-01 | 1.00e+00 | 2793 | 2014 | 779 |
| African | Recorded Anxiety | Discontinuation vs Continuation | Education_level | sex_at_birthMale | 1.228 | 1.008 | 1.497 | 4.11e-02 | 5.75e-01 | 2793 | 2014 | 779 |

| | | | | | | | | | | | | |
|---|---|---|---|---|---|---|---|---|---|---|---|---|
| African | Recorded Anxiety | Discontinuation vs Continuation | Education_level | Income>200k | 0.554 | 0.205 | 1.497 | 2.44e-01 | 1.00e+00 | 2793 | 2014 | 779 |
| African | Recorded Anxiety | Discontinuation vs Continuation | Education_level | Income100-150k | 0.484 | 0.293 | 0.800 | 4.61e-03 | 6.46e-02 | 2793 | 2014 | 779 |
| African | Recorded Anxiety | Discontinuation vs Continuation | Education_level | Income150-200k | 0.942 | 0.445 | 1.991 | 8.75e-01 | 1.00e+00 | 2793 | 2014 | 779 |
| African | Recorded Anxiety | Discontinuation vs Continuation | Education_level | Income25-35k | 1.115 | 0.855 | 1.454 | 4.23e-01 | 1.00e+00 | 2793 | 2014 | 779 |
| African | Recorded Anxiety | Discontinuation vs Continuation | Education_level | Income35-50k | 0.742 | 0.557 | 0.988 | 4.10e-02 | 5.74e-01 | 2793 | 2014 | 779 |
| African | Recorded Anxiety | Discontinuation vs Continuation | Education_level | Income50-75k | 1.037 | 0.782 | 1.375 | 8.01e-01 | 1.00e+00 | 2793 | 2014 | 779 |
| African | Recorded Anxiety | Discontinuation vs Continuation | Education_level | Income75-100k | 0.626 | 0.407 | 0.961 | 3.24e-02 | 4.53e-01 | 2793 | 2014 | 779 |
| African | Recorded Anxiety | Discontinuation vs Continuation | Education_level | Education_levelCollege Postgraduate | 0.872 | 0.573 | 1.327 | 5.21e-01 | 1.00e+00 | 2793 | 2014 | 779 |
| African | Recorded Anxiety | Discontinuation vs Continuation | Education_level | Education_levelCollege Undergraduate | 0.968 | 0.705 | 1.328 | 8.38e-01 | 1.00e+00 | 2793 | 2014 | 779 |
| African | Recorded Anxiety | Discontinuation vs Continuation | Education_level | Education_levelTwelve or GED | 0.997 | 0.720 | 1.380 | 9.85e-01 | 1.00e+00 | 2793 | 2014 | 779 |
| African | Recorded Anxiety | Switching vs Continuation | Education_level | (Intercept) | 1.145 | 0.920 | 1.424 | 2.25e-01 | 1.00e+00 | 3909 | 2014 | 1895 |
| African | Recorded Anxiety | Switching vs Continuation | Education_level | age_at_first_AD | 0.801 | 0.742 | 0.865 | 1.31e-08 | 1.83e-07 | 3909 | 2014 | 1895 |
| African | Recorded Anxiety | Switching vs Continuation | Education_level | sex_at_birthMale | 1.074 | 0.919 | 1.255 | 3.70e-01 | 1.00e+00 | 3909 | 2014 | 1895 |
| African | Recorded Anxiety | Switching vs Continuation | Education_level | Income>200k | 0.542 | 0.272 | 1.079 | 8.14e-02 | 1.00e+00 | 3909 | 2014 | 1895 |
| African | Recorded Anxiety | Switching vs Continuation | Education_level | Income100-150k | 0.576 | 0.415 | 0.800 | 1.01e-03 | 1.42e-02 | 3909 | 2014 | 1895 |
| African | Recorded Anxiety | Switching vs Continuation | Education_level | Income150-200k | 0.595 | 0.322 | 1.099 | 9.71e-02 | 1.00e+00 | 3909 | 2014 | 1895 |
| African | Recorded Anxiety | Switching vs Continuation | Education_level | Income25-35k | 0.760 | 0.614 | 0.942 | 1.23e-02 | 1.72e-01 | 3909 | 2014 | 1895 |
| African | Recorded Anxiety | Switching vs Continuation | Education_level | Income35-50k | 0.738 | 0.599 | 0.909 | 4.34e-03 | 6.07e-02 | 3909 | 2014 | 1895 |
| African | Recorded Anxiety | Switching vs Continuation | Education_level | Income50-75k | 0.655 | 0.520 | 0.824 | 3.04e-04 | 4.26e-03 | 3909 | 2014 | 1895 |
| African | Recorded Anxiety | Switching vs Continuation | Education_level | Income75-100k | 0.567 | 0.415 | 0.774 | 3.56e-04 | 4.98e-03 | 3909 | 2014 | 1895 |
| African | Recorded Anxiety | Switching vs Continuation | Education_level | Education_levelCollege Postgraduate | 0.927 | 0.678 | 1.268 | 6.36e-01 | 1.00e+00 | 3909 | 2014 | 1895 |
| African | Recorded Anxiety | Switching vs Continuation | Education_level | Education_levelCollege Undergraduate | 0.941 | 0.742 | 1.194 | 6.16e-01 | 1.00e+00 | 3909 | 2014 | 1895 |
| African | Recorded Anxiety | Switching vs Continuation | Education_level | Education_levelTwelve or GED | 0.846 | 0.661 | 1.084 | 1.86e-01 | 1.00e+00 | 3909 | 2014 | 1895 |
| African | Recorded Anxiety | Discontinuation vs Continuation | Income | (Intercept) | 0.407 | 0.304 | 0.544 | 1.42e-09 | 1.99e-08 | 2793 | 2014 | 779 |
| African | Recorded Anxiety | Discontinuation vs Continuation | Income | age_at_first_AD | 0.943 | 0.858 | 1.036 | 2.24e-01 | 1.00e+00 | 2793 | 2014 | 779 |
| African | Recorded Anxiety | Discontinuation vs Continuation | Income | sex_at_birthMale | 1.228 | 1.008 | 1.497 | 4.11e-02 | 5.75e-01 | 2793 | 2014 | 779 |
| African | Recorded Anxiety | Discontinuation vs Continuation | Income | Income>200k | 0.554 | 0.205 | 1.497 | 2.44e-01 | 1.00e+00 | 2793 | 2014 | 779 |
| African | Recorded Anxiety | Discontinuation vs Continuation | Income | Income100-150k | 0.484 | 0.293 | 0.800 | 4.61e-03 | 6.46e-02 | 2793 | 2014 | 779 |
| African | Recorded Anxiety | Discontinuation vs Continuation | Income | Income150-200k | 0.942 | 0.445 | 1.991 | 8.75e-01 | 1.00e+00 | 2793 | 2014 | 779 |
| African | Recorded Anxiety | Discontinuation vs Continuation | Income | Income25-35k | 1.115 | 0.855 | 1.454 | 4.23e-01 | 1.00e+00 | 2793 | 2014 | 779 |
| African | Recorded Anxiety | Discontinuation vs Continuation | Income | Income35-50k | 0.742 | 0.557 | 0.988 | 4.10e-02 | 5.74e-01 | 2793 | 2014 | 779 |
| African | Recorded Anxiety | Discontinuation vs Continuation | Income | Income50-75k | 1.037 | 0.782 | 1.375 | 8.01e-01 | 1.00e+00 | 2793 | 2014 | 779 |
| African | Recorded Anxiety | Discontinuation vs Continuation | Income | Income75-100k | 0.626 | 0.407 | 0.961 | 3.24e-02 | 4.53e-01 | 2793 | 2014 | 779 |
| African | Recorded Anxiety | Discontinuation vs Continuation | Income | Education_levelCollege Postgraduate | 0.872 | 0.573 | 1.327 | 5.21e-01 | 1.00e+00 | 2793 | 2014 | 779 |
| African | Recorded Anxiety | Discontinuation vs Continuation | Income | Education_levelCollege Undergraduate | 0.968 | 0.705 | 1.328 | 8.38e-01 | 1.00e+00 | 2793 | 2014 | 779 |
| African | Recorded Anxiety | Discontinuation vs Continuation | Income | Education_levelTwelve or GED | 0.997 | 0.720 | 1.380 | 9.85e-01 | 1.00e+00 | 2793 | 2014 | 779 |
| African | Recorded Anxiety | Switching vs Continuation | Income | (Intercept) | 1.145 | 0.920 | 1.424 | 2.25e-01 | 1.00e+00 | 3909 | 2014 | 1895 |
| African | Recorded Anxiety | Switching vs Continuation | Income | age_at_first_AD | 0.801 | 0.742 | 0.865 | 1.31e-08 | 1.83e-07 | 3909 | 2014 | 1895 |
| African | Recorded Anxiety | Switching vs Continuation | Income | sex_at_birthMale | 1.074 | 0.919 | 1.255 | 3.70e-01 | 1.00e+00 | 3909 | 2014 | 1895 |
| African | Recorded Anxiety | Switching vs Continuation | Income | Income>200k | 0.542 | 0.272 | 1.079 | 8.14e-02 | 1.00e+00 | 3909 | 2014 | 1895 |
| African | Recorded Anxiety | Switching vs Continuation | Income | Income100-150k | 0.576 | 0.415 | 0.800 | 1.01e-03 | 1.42e-02 | 3909 | 2014 | 1895 |
| African | Recorded Anxiety | Switching vs Continuation | Income | Income150-200k | 0.595 | 0.322 | 1.099 | 9.71e-02 | 1.00e+00 | 3909 | 2014 | 1895 |
| African | Recorded Anxiety | Switching vs Continuation | Income | Income25-35k | 0.760 | 0.614 | 0.942 | 1.23e-02 | 1.72e-01 | 3909 | 2014 | 1895 |
| African | Recorded Anxiety | Switching vs Continuation | Income | Income35-50k | 0.738 | 0.599 | 0.909 | 4.34e-03 | 6.07e-02 | 3909 | 2014 | 1895 |
| African | Recorded Anxiety | Switching vs Continuation | Income | Income50-75k | 0.655 | 0.520 | 0.824 | 3.04e-04 | 4.26e-03 | 3909 | 2014 | 1895 |
| African | Recorded Anxiety | Switching vs Continuation | Income | Income75-100k | 0.567 | 0.415 | 0.774 | 3.56e-04 | 4.98e-03 | 3909 | 2014 | 1895 |
| African | Recorded Anxiety | Switching vs Continuation | Income | Education_levelCollege Postgraduate | 0.927 | 0.678 | 1.268 | 6.36e-01 | 1.00e+00 | 3909 | 2014 | 1895 |
| African | Recorded Anxiety | Switching vs Continuation | Income | Education_levelCollege Undergraduate | 0.941 | 0.742 | 1.194 | 6.16e-01 | 1.00e+00 | 3909 | 2014 | 1895 |
| African | Recorded Anxiety | Switching vs Continuation | Income | Education_levelTwelve or GED | 0.846 | 0.661 | 1.084 | 1.86e-01 | 1.00e+00 | 3909 | 2014 | 1895 |
| African | Recorded Anxiety | Discontinuation vs Continuation | Insomnia | (Intercept) | 0.415 | 0.309 | 0.558 | 5.52e-09 | 7.72e-08 | 2793 | 2014 | 779 |
| African | Recorded Anxiety | Discontinuation vs Continuation | Insomnia | age_at_first_AD | 0.946 | 0.861 | 1.040 | 2.52e-01 | 1.00e+00 | 2793 | 2014 | 779 |
| African | Recorded Anxiety | Discontinuation vs Continuation | Insomnia | sex_at_birthMale | 1.228 | 1.008 | 1.496 | 4.15e-02 | 5.82e-01 | 2793 | 2014 | 779 |
| African | Recorded Anxiety | Discontinuation vs Continuation | Insomnia | Income>200k | 0.554 | 0.205 | 1.497 | 2.44e-01 | 1.00e+00 | 2793 | 2014 | 779 |
| African | Recorded Anxiety | Discontinuation vs Continuation | Insomnia | Income100-150k | 0.485 | 0.294 | 0.802 | 4.76e-03 | 6.66e-02 | 2793 | 2014 | 779 |
| African | Recorded Anxiety | Discontinuation vs Continuation | Insomnia | Income150-200k | 0.946 | 0.447 | 1.999 | 8.84e-01 | 1.00e+00 | 2793 | 2014 | 779 |
| African | Recorded Anxiety | Discontinuation vs Continuation | Insomnia | Income25-35k | 1.112 | 0.853 | 1.451 | 4.32e-01 | 1.00e+00 | 2793 | 2014 | 779 |
| African | Recorded Anxiety | Discontinuation vs Continuation | Insomnia | Income35-50k | 0.746 | 0.560 | 0.993 | 4.46e-02 | 6.24e-01 | 2793 | 2014 | 779 |
| African | Recorded Anxiety | Discontinuation vs Continuation | Insomnia | Income50-75k | 1.037 | 0.782 | 1.375 | 8.02e-01 | 1.00e+00 | 2793 | 2014 | 779 |
| African | Recorded Anxiety | Discontinuation vs Continuation | Insomnia | Income75-100k | 0.629 | 0.409 | 0.966 | 3.43e-02 | 4.80e-01 | 2793 | 2014 | 779 |
| African | Recorded Anxiety | Discontinuation vs Continuation | Insomnia | Education_levelCollege Postgraduate | 0.870 | 0.571 | 1.324 | 5.15e-01 | 1.00e+00 | 2793 | 2014 | 779 |
| African | Recorded Anxiety | Discontinuation vs Continuation | Insomnia | Education_levelCollege Undergraduate | 0.969 | 0.706 | 1.330 | 8.44e-01 | 1.00e+00 | 2793 | 2014 | 779 |
| African | Recorded Anxiety | Discontinuation vs Continuation | Insomnia | Education_levelTwelve or GED | 0.996 | 0.719 | 1.379 | 9.79e-01 | 1.00e+00 | 2793 | 2014 | 779 |
| African | Recorded Anxiety | Discontinuation vs Continuation | Insomnia | Insomnia | 0.927 | 0.768 | 1.121 | 4.35e-01 | 1.00e+00 | 2793 | 2014 | 779 |
| African | Recorded Anxiety | Switching vs Continuation | Insomnia | (Intercept) | 0.804 | 0.640 | 1.012 | 6.30e-02 | 8.82e-01 | 3909 | 2014 | 1895 |
| African | Recorded Anxiety | Switching vs Continuation | Insomnia | age_at_first_AD | 0.771 | 0.713 | 0.834 | 8.65e-11 | 1.21e-09 | 3909 | 2014 | 1895 |
| African | Recorded Anxiety | Switching vs Continuation | Insomnia | sex_at_birthMale | 1.060 | 0.904 | 1.244 | 4.70e-01 | 1.00e+00 | 3909 | 2014 | 1895 |

| | | | | | | | | | | | | |
|---|---|---|---|---|---|---|---|---|---|---|---|---|
| African | Recorded Anxiety | Switching vs Continuation | Insomnia | Income>200k | 0.580 | 0.288 | 1.167 | 1.27e-01 | 1.00e+00 | 3909 | 2014 | 1895 |
| African | Recorded Anxiety | Switching vs Continuation | Insomnia | Income100-150k | 0.552 | 0.394 | 0.772 | 5.29e-04 | 7.41e-03 | 3909 | 2014 | 1895 |
| African | Recorded Anxiety | Switching vs Continuation | Insomnia | Income150-200k | 0.583 | 0.311 | 1.094 | 9.29e-02 | 1.00e+00 | 3909 | 2014 | 1895 |
| African | Recorded Anxiety | Switching vs Continuation | Insomnia | Income25-35k | 0.772 | 0.620 | 0.962 | 2.09e-02 | 2.92e-01 | 3909 | 2014 | 1895 |
| African | Recorded Anxiety | Switching vs Continuation | Insomnia | Income35-50k | 0.723 | 0.584 | 0.895 | 2.92e-03 | 4.09e-02 | 3909 | 2014 | 1895 |
| African | Recorded Anxiety | Switching vs Continuation | Insomnia | Income50-75k | 0.652 | 0.515 | 0.824 | 3.55e-04 | 4.97e-03 | 3909 | 2014 | 1895 |
| African | Recorded Anxiety | Switching vs Continuation | Insomnia | Income75-100k | 0.553 | 0.402 | 0.761 | 2.79e-04 | 3.90e-03 | 3909 | 2014 | 1895 |
| African | Recorded Anxiety | Switching vs Continuation | Insomnia | Education_levelCollege Postgraduate | 0.962 | 0.699 | 1.326 | 8.14e-01 | 1.00e+00 | 3909 | 2014 | 1895 |
| African | Recorded Anxiety | Switching vs Continuation | Insomnia | Education_levelCollege Undergraduate | 0.948 | 0.743 | 1.209 | 6.64e-01 | 1.00e+00 | 3909 | 2014 | 1895 |
| African | Recorded Anxiety | Switching vs Continuation | Insomnia | Education_levelTwelve or GED | 0.878 | 0.681 | 1.130 | 3.12e-01 | 1.00e+00 | 3909 | 2014 | 1895 |
| African | Recorded Anxiety | Switching vs Continuation | Insomnia | Insomnia | 2.429 | 2.123 | 2.780 | <2.2e-16 | <2.2e-16 | 3909 | 2014 | 1895 |
| African | Recorded Anxiety | Discontinuation vs Continuation | MDD CIDI-sf | (Intercept) | 0.458 | 0.038 | 5.526 | 5.39e-01 | 1.00e+00 | 161 | 117 | 44 |
| African | Recorded Anxiety | Discontinuation vs Continuation | MDD CIDI-sf | age_at_first_AD | 0.967 | 0.641 | 1.459 | 8.74e-01 | 1.00e+00 | 161 | 117 | 44 |
| African | Recorded Anxiety | Discontinuation vs Continuation | MDD CIDI-sf | sex_at_birthMale | 0.759 | 0.296 | 1.947 | 5.66e-01 | 1.00e+00 | 161 | 117 | 44 |
| African | Recorded Anxiety | Discontinuation vs Continuation | MDD CIDI-sf | Income>200k | 4.157 | 0.414 | 41.730 | 2.26e-01 | 1.00e+00 | 161 | 117 | 44 |
| African | Recorded Anxiety | Discontinuation vs Continuation | MDD CIDI-sf | Income100-150k | 1.525 | 0.390 | 5.969 | 5.44e-01 | 1.00e+00 | 161 | 117 | 44 |
| African | Recorded Anxiety | Discontinuation vs Continuation | MDD CIDI-sf | Income150-200k | 3.914 | 0.191 | 80.280 | 3.76e-01 | 1.00e+00 | 161 | 117 | 44 |
| African | Recorded Anxiety | Discontinuation vs Continuation | MDD CIDI-sf | Income25-35k | 0.570 | 0.108 | 2.992 | 5.06e-01 | 1.00e+00 | 161 | 117 | 44 |
| African | Recorded Anxiety | Discontinuation vs Continuation | MDD CIDI-sf | Income35-50k | 1.632 | 0.539 | 4.941 | 3.86e-01 | 1.00e+00 | 161 | 117 | 44 |
| African | Recorded Anxiety | Discontinuation vs Continuation | MDD CIDI-sf | Income50-75k | 1.174 | 0.382 | 3.616 | 7.79e-01 | 1.00e+00 | 161 | 117 | 44 |
| African | Recorded Anxiety | Discontinuation vs Continuation | MDD CIDI-sf | Income75-100k | 1.792 | 0.431 | 7.451 | 4.22e-01 | 1.00e+00 | 161 | 117 | 44 |
| African | Recorded Anxiety | Discontinuation vs Continuation | MDD CIDI-sf | Education_levelCollege Postgraduate | 0.461 | 0.033 | 6.513 | 5.66e-01 | 1.00e+00 | 161 | 117 | 44 |
| African | Recorded Anxiety | Discontinuation vs Continuation | MDD CIDI-sf | Education_levelCollege Undergraduate | 0.463 | 0.037 | 5.781 | 5.50e-01 | 1.00e+00 | 161 | 117 | 44 |
| African | Recorded Anxiety | Discontinuation vs Continuation | MDD CIDI-sf | Education_levelTwelve or GED | 1.245 | 0.092 | 16.800 | 8.69e-01 | 1.00e+00 | 161 | 117 | 44 |
| African | Recorded Anxiety | Discontinuation vs Continuation | MDD CIDI-sf | MDD CIDI-sf | 1.463 | 0.685 | 3.125 | 3.26e-01 | 1.00e+00 | 161 | 117 | 44 |
| African | Recorded Anxiety | Discontinuation vs Continuation | MDD_symptom_count | (Intercept) | 0.485 | 0.041 | 5.736 | 5.66e-01 | 1.00e+00 | 186 | 137 | 49 |
| African | Recorded Anxiety | Discontinuation vs Continuation | MDD_symptom_count | age_at_first_AD | 0.969 | 0.657 | 1.427 | 8.71e-01 | 1.00e+00 | 186 | 137 | 49 |
| African | Recorded Anxiety | Discontinuation vs Continuation | MDD_symptom_count | sex_at_birthMale | 0.836 | 0.347 | 2.010 | 6.88e-01 | 1.00e+00 | 186 | 137 | 49 |
| African | Recorded Anxiety | Discontinuation vs Continuation | MDD_symptom_count | Income>200k | 2.346 | 0.291 | 18.900 | 4.23e-01 | 1.00e+00 | 186 | 137 | 49 |
| African | Recorded Anxiety | Discontinuation vs Continuation | MDD_symptom_count | Income100-150k | 1.271 | 0.355 | 4.547 | 7.12e-01 | 1.00e+00 | 186 | 137 | 49 |
| African | Recorded Anxiety | Discontinuation vs Continuation | MDD_symptom_count | Income150-200k | 3.421 | 0.175 | 66.730 | 4.17e-01 | 1.00e+00 | 186 | 137 | 49 |
| African | Recorded Anxiety | Discontinuation vs Continuation | MDD_symptom_count | Income25-35k | 0.885 | 0.211 | 3.714 | 8.67e-01 | 1.00e+00 | 186 | 137 | 49 |
| African | Recorded Anxiety | Discontinuation vs Continuation | MDD_symptom_count | Income35-50k | 1.559 | 0.535 | 4.545 | 4.16e-01 | 1.00e+00 | 186 | 137 | 49 |
| African | Recorded Anxiety | Discontinuation vs Continuation | MDD_symptom_count | Income50-75k | 1.613 | 0.579 | 4.498 | 3.61e-01 | 1.00e+00 | 186 | 137 | 49 |
| African | Recorded Anxiety | Discontinuation vs Continuation | MDD_symptom_count | Income75-100k | 1.285 | 0.336 | 4.911 | 7.14e-01 | 1.00e+00 | 186 | 137 | 49 |
| African | Recorded Anxiety | Discontinuation vs Continuation | MDD_symptom_count | Education_levelCollege Postgraduate | 0.613 | 0.045 | 8.326 | 7.13e-01 | 1.00e+00 | 186 | 137 | 49 |
| African | Recorded Anxiety | Discontinuation vs Continuation | MDD_symptom_count | Education_levelCollege Undergraduate | 0.471 | 0.038 | 5.829 | 5.58e-01 | 1.00e+00 | 186 | 137 | 49 |
| African | Recorded Anxiety | Discontinuation vs Continuation | MDD_symptom_count | Education_levelTwelve or GED | 1.376 | 0.104 | 18.160 | 8.08e-01 | 1.00e+00 | 186 | 137 | 49 |
| African | Recorded Anxiety | Discontinuation vs Continuation | MDD_symptom_count | MDD_symptom_count | 1.092 | 0.787 | 1.515 | 5.99e-01 | 1.00e+00 | 186 | 137 | 49 |
| African | Recorded Anxiety | Discontinuation vs Continuation | Migraine | (Intercept) | 0.416 | 0.309 | 0.560 | 7.74e-09 | 1.08e-07 | 2793 | 2014 | 779 |
| African | Recorded Anxiety | Discontinuation vs Continuation | Migraine | age_at_first_AD | 0.941 | 0.856 | 1.034 | 2.04e-01 | 1.00e+00 | 2793 | 2014 | 779 |
| African | Recorded Anxiety | Discontinuation vs Continuation | Migraine | sex_at_birthMale | 1.218 | 0.998 | 1.486 | 5.25e-02 | 7.34e-01 | 2793 | 2014 | 779 |
| African | Recorded Anxiety | Discontinuation vs Continuation | Migraine | Income>200k | 0.553 | 0.204 | 1.494 | 2.42e-01 | 1.00e+00 | 2793 | 2014 | 779 |
| African | Recorded Anxiety | Discontinuation vs Continuation | Migraine | Income100-150k | 0.484 | 0.293 | 0.800 | 4.67e-03 | 6.54e-02 | 2793 | 2014 | 779 |
| African | Recorded Anxiety | Discontinuation vs Continuation | Migraine | Income150-200k | 0.942 | 0.446 | 1.991 | 8.75e-01 | 1.00e+00 | 2793 | 2014 | 779 |
| African | Recorded Anxiety | Discontinuation vs Continuation | Migraine | Income25-35k | 1.113 | 0.853 | 1.451 | 4.31e-01 | 1.00e+00 | 2793 | 2014 | 779 |
| African | Recorded Anxiety | Discontinuation vs Continuation | Migraine | Income35-50k | 0.744 | 0.559 | 0.991 | 4.31e-02 | 6.04e-01 | 2793 | 2014 | 779 |
| African | Recorded Anxiety | Discontinuation vs Continuation | Migraine | Income50-75k | 1.040 | 0.784 | 1.379 | 7.86e-01 | 1.00e+00 | 2793 | 2014 | 779 |
| African | Recorded Anxiety | Discontinuation vs Continuation | Migraine | Income75-100k | 0.628 | 0.408 | 0.965 | 3.36e-02 | 4.70e-01 | 2793 | 2014 | 779 |
| African | Recorded Anxiety | Discontinuation vs Continuation | Migraine | Education_levelCollege Postgraduate | 0.869 | 0.570 | 1.322 | 5.11e-01 | 1.00e+00 | 2793 | 2014 | 779 |
| African | Recorded Anxiety | Discontinuation vs Continuation | Migraine | Education_levelCollege Undergraduate | 0.968 | 0.705 | 1.328 | 8.39e-01 | 1.00e+00 | 2793 | 2014 | 779 |
| African | Recorded Anxiety | Discontinuation vs Continuation | Migraine | Education_levelTwelve or GED | 0.996 | 0.719 | 1.379 | 9.79e-01 | 1.00e+00 | 2793 | 2014 | 779 |
| African | Recorded Anxiety | Discontinuation vs Continuation | Migraine | Migraine | 0.938 | 0.783 | 1.125 | 4.91e-01 | 1.00e+00 | 2793 | 2014 | 779 |
| African | Recorded Anxiety | Switching vs Continuation | Migraine | (Intercept) | 0.934 | 0.746 | 1.170 | 5.54e-01 | 1.00e+00 | 3909 | 2014 | 1895 |
| African | Recorded Anxiety | Switching vs Continuation | Migraine | age_at_first_AD | 0.830 | 0.768 | 0.896 | 2.26e-06 | 3.16e-05 | 3909 | 2014 | 1895 |
| African | Recorded Anxiety | Switching vs Continuation | Migraine | sex_at_birthMale | 1.163 | 0.993 | 1.362 | 6.13e-02 | 8.58e-01 | 3909 | 2014 | 1895 |
| African | Recorded Anxiety | Switching vs Continuation | Migraine | Income>200k | 0.531 | 0.265 | 1.063 | 7.37e-02 | 1.00e+00 | 3909 | 2014 | 1895 |
| African | Recorded Anxiety | Switching vs Continuation | Migraine | Income100-150k | 0.565 | 0.405 | 0.788 | 7.61e-04 | 1.07e-02 | 3909 | 2014 | 1895 |
| African | Recorded Anxiety | Switching vs Continuation | Migraine | Income150-200k | 0.618 | 0.333 | 1.148 | 1.28e-01 | 1.00e+00 | 3909 | 2014 | 1895 |
| African | Recorded Anxiety | Switching vs Continuation | Migraine | Income25-35k | 0.774 | 0.623 | 0.960 | 1.99e-02 | 2.79e-01 | 3909 | 2014 | 1895 |
| African | Recorded Anxiety | Switching vs Continuation | Migraine | Income35-50k | 0.705 | 0.571 | 0.870 | 1.16e-03 | 1.62e-02 | 3909 | 2014 | 1895 |
| African | Recorded Anxiety | Switching vs Continuation | Migraine | Income50-75k | 0.649 | 0.514 | 0.819 | 2.63e-04 | 3.68e-03 | 3909 | 2014 | 1895 |
| African | Recorded Anxiety | Switching vs Continuation | Migraine | Income75-100k | 0.554 | 0.404 | 0.758 | 2.25e-04 | 3.15e-03 | 3909 | 2014 | 1895 |
| African | Recorded Anxiety | Switching vs Continuation | Migraine | Education_levelCollege Postgraduate | 0.917 | 0.669 | 1.257 | 5.88e-01 | 1.00e+00 | 3909 | 2014 | 1895 |
| African | Recorded Anxiety | Switching vs Continuation | Migraine | Education_levelCollege Undergraduate | 0.910 | 0.716 | 1.157 | 4.41e-01 | 1.00e+00 | 3909 | 2014 | 1895 |
| African | Recorded Anxiety | Switching vs Continuation | Migraine | Education_levelTwelve or GED | 0.831 | 0.647 | 1.066 | 1.44e-01 | 1.00e+00 | 3909 | 2014 | 1895 |

| | | | | | | | | | | | | |
|---|---|---|---|---|---|---|---|---|---|---|---|---|
| African | Recorded Anxiety | Switching vs Continuation | Migraine | Migraine | 1.760 | 1.540 | 2.010 | <2.2e-16 | 1.22e-15 | 3909 | 2014 | 1895 |
| African | Recorded Anxiety | Discontinuation vs Continuation | Recorded Depression | (Intercept) | 0.453 | 0.333 | 0.615 | 3.92e-07 | 5.49e-06 | 2793 | 2014 | 779 |
| African | Recorded Anxiety | Discontinuation vs Continuation | Recorded Depression | age_at_first_AD | 0.936 | 0.852 | 1.029 | 1.72e-01 | 1.00e+00 | 2793 | 2014 | 779 |
| African | Recorded Anxiety | Discontinuation vs Continuation | Recorded Depression | sex_at_birthMale | 1.233 | 1.012 | 1.502 | 3.81e-02 | 5.34e-01 | 2793 | 2014 | 779 |
| African | Recorded Anxiety | Discontinuation vs Continuation | Recorded Depression | Income>200k | 0.529 | 0.195 | 1.433 | 2.11e-01 | 1.00e+00 | 2793 | 2014 | 779 |
| African | Recorded Anxiety | Discontinuation vs Continuation | Recorded Depression | Income100-150k | 0.474 | 0.287 | 0.783 | 3.60e-03 | 5.04e-02 | 2793 | 2014 | 779 |
| African | Recorded Anxiety | Discontinuation vs Continuation | Recorded Depression | Income150-200k | 0.936 | 0.442 | 1.979 | 8.62e-01 | 1.00e+00 | 2793 | 2014 | 779 |
| African | Recorded Anxiety | Discontinuation vs Continuation | Recorded Depression | Income25-35k | 1.103 | 0.845 | 1.439 | 4.71e-01 | 1.00e+00 | 2793 | 2014 | 779 |
| African | Recorded Anxiety | Discontinuation vs Continuation | Recorded Depression | Income35-50k | 0.735 | 0.552 | 0.979 | 3.50e-02 | 4.90e-01 | 2793 | 2014 | 779 |
| African | Recorded Anxiety | Discontinuation vs Continuation | Recorded Depression | Income50-75k | 1.028 | 0.775 | 1.364 | 8.49e-01 | 1.00e+00 | 2793 | 2014 | 779 |
| African | Recorded Anxiety | Discontinuation vs Continuation | Recorded Depression | Income75-100k | 0.616 | 0.401 | 0.947 | 2.71e-02 | 3.80e-01 | 2793 | 2014 | 779 |
| African | Recorded Anxiety | Discontinuation vs Continuation | Recorded Depression | Education_levelCollege Postgraduate | 0.869 | 0.570 | 1.323 | 5.12e-01 | 1.00e+00 | 2793 | 2014 | 779 |
| African | Recorded Anxiety | Discontinuation vs Continuation | Recorded Depression | Education_levelCollege Undergraduate | 0.971 | 0.707 | 1.334 | 8.57e-01 | 1.00e+00 | 2793 | 2014 | 779 |
| African | Recorded Anxiety | Discontinuation vs Continuation | Recorded Depression | Education_levelTwelve or GED | 0.999 | 0.721 | 1.384 | 9.95e-01 | 1.00e+00 | 2793 | 2014 | 779 |
| African | Recorded Anxiety | Discontinuation vs Continuation | Recorded Depression | Recorded Depression | 0.825 | 0.698 | 0.977 | 2.52e-02 | 3.53e-01 | 2793 | 2014 | 779 |
| African | Recorded Anxiety | Switching vs Continuation | Recorded Depression | (Intercept) | 0.487 | 0.379 | 0.625 | 1.72e-08 | 2.41e-07 | 3909 | 2014 | 1895 |
| African | Recorded Anxiety | Switching vs Continuation | Recorded Depression | age_at_first_AD | 0.817 | 0.755 | 0.885 | 6.45e-07 | 9.03e-06 | 3909 | 2014 | 1895 |
| African | Recorded Anxiety | Switching vs Continuation | Recorded Depression | sex_at_birthMale | 1.077 | 0.916 | 1.266 | 3.70e-01 | 1.00e+00 | 3909 | 2014 | 1895 |
| African | Recorded Anxiety | Switching vs Continuation | Recorded Depression | Income>200k | 0.679 | 0.333 | 1.385 | 2.87e-01 | 1.00e+00 | 3909 | 2014 | 1895 |
| African | Recorded Anxiety | Switching vs Continuation | Recorded Depression | Income100-150k | 0.675 | 0.479 | 0.950 | 2.42e-02 | 3.39e-01 | 3909 | 2014 | 1895 |
| African | Recorded Anxiety | Switching vs Continuation | Recorded Depression | Income150-200k | 0.605 | 0.320 | 1.141 | 1.20e-01 | 1.00e+00 | 3909 | 2014 | 1895 |
| African | Recorded Anxiety | Switching vs Continuation | Recorded Depression | Income25-35k | 0.775 | 0.621 | 0.968 | 2.47e-02 | 3.46e-01 | 3909 | 2014 | 1895 |
| African | Recorded Anxiety | Switching vs Continuation | Recorded Depression | Income35-50k | 0.754 | 0.607 | 0.936 | 1.03e-02 | 1.45e-01 | 3909 | 2014 | 1895 |
| African | Recorded Anxiety | Switching vs Continuation | Recorded Depression | Income50-75k | 0.682 | 0.537 | 0.866 | 1.65e-03 | 2.30e-02 | 3909 | 2014 | 1895 |
| African | Recorded Anxiety | Switching vs Continuation | Recorded Depression | Income75-100k | 0.597 | 0.432 | 0.824 | 1.70e-03 | 2.38e-02 | 3909 | 2014 | 1895 |
| African | Recorded Anxiety | Switching vs Continuation | Recorded Depression | Education_levelCollege Postgraduate | 0.945 | 0.683 | 1.307 | 7.31e-01 | 1.00e+00 | 3909 | 2014 | 1895 |
| African | Recorded Anxiety | Switching vs Continuation | Recorded Depression | Education_levelCollege Undergraduate | 0.901 | 0.704 | 1.153 | 4.08e-01 | 1.00e+00 | 3909 | 2014 | 1895 |
| African | Recorded Anxiety | Switching vs Continuation | Recorded Depression | Education_levelTwelve or GED | 0.822 | 0.636 | 1.062 | 1.34e-01 | 1.00e+00 | 3909 | 2014 | 1895 |
| African | Recorded Anxiety | Switching vs Continuation | Recorded Depression | Recorded Depression | 3.392 | 2.922 | 3.937 | <2.2e-16 | <2.2e-16 | 3909 | 2014 | 1895 |
| African | Recorded Anxiety | Discontinuation vs Continuation | Smoking_Frequency | (Intercept) | 0.394 | 0.289 | 0.537 | 3.67e-09 | 5.14e-08 | 2732 | 1973 | 759 |
| African | Recorded Anxiety | Discontinuation vs Continuation | Smoking_Frequency | age_at_first_AD | 0.969 | 0.879 | 1.069 | 5.34e-01 | 1.00e+00 | 2732 | 1973 | 759 |
| African | Recorded Anxiety | Discontinuation vs Continuation | Smoking_Frequency | sex_at_birthMale | 1.225 | 1.000 | 1.502 | 5.04e-02 | 7.06e-01 | 2732 | 1973 | 759 |
| African | Recorded Anxiety | Discontinuation vs Continuation | Smoking_Frequency | Income>200k | 0.619 | 0.228 | 1.682 | 3.47e-01 | 1.00e+00 | 2732 | 1973 | 759 |
| African | Recorded Anxiety | Discontinuation vs Continuation | Smoking_Frequency | Income100-150k | 0.507 | 0.306 | 0.840 | 8.35e-03 | 1.17e-01 | 2732 | 1973 | 759 |
| African | Recorded Anxiety | Discontinuation vs Continuation | Smoking_Frequency | Income150-200k | 0.864 | 0.397 | 1.884 | 7.14e-01 | 1.00e+00 | 2732 | 1973 | 759 |
| African | Recorded Anxiety | Discontinuation vs Continuation | Smoking_Frequency | Income25-35k | 1.166 | 0.891 | 1.527 | 2.64e-01 | 1.00e+00 | 2732 | 1973 | 759 |
| African | Recorded Anxiety | Discontinuation vs Continuation | Smoking_Frequency | Income35-50k | 0.780 | 0.583 | 1.043 | 9.34e-02 | 1.00e+00 | 2732 | 1973 | 759 |
| African | Recorded Anxiety | Discontinuation vs Continuation | Smoking_Frequency | Income50-75k | 1.075 | 0.803 | 1.440 | 6.26e-01 | 1.00e+00 | 2732 | 1973 | 759 |
| African | Recorded Anxiety | Discontinuation vs Continuation | Smoking_Frequency | Income75-100k | 0.677 | 0.439 | 1.045 | 7.81e-02 | 1.00e+00 | 2732 | 1973 | 759 |
| African | Recorded Anxiety | Discontinuation vs Continuation | Smoking_Frequency | Education_levelCollege Postgraduate | 0.876 | 0.571 | 1.343 | 5.43e-01 | 1.00e+00 | 2732 | 1973 | 759 |
| African | Recorded Anxiety | Discontinuation vs Continuation | Smoking_Frequency | Education_levelCollege Undergraduate | 0.987 | 0.716 | 1.362 | 9.38e-01 | 1.00e+00 | 2732 | 1973 | 759 |
| African | Recorded Anxiety | Discontinuation vs Continuation | Smoking_Frequency | Education_levelTwelve or GED | 0.986 | 0.709 | 1.371 | 9.34e-01 | 1.00e+00 | 2732 | 1973 | 759 |
| African | Recorded Anxiety | Discontinuation vs Continuation | Smoking_Frequency | Smoking_FrequencyEvery Day | 1.360 | 1.062 | 1.742 | 1.47e-02 | 2.06e-01 | 2732 | 1973 | 759 |
| African | Recorded Anxiety | Discontinuation vs Continuation | Smoking_Frequency | Smoking_FrequencyNot At All (former) | 0.822 | 0.648 | 1.043 | 1.06e-01 | 1.00e+00 | 2732 | 1973 | 759 |
| African | Recorded Anxiety | Discontinuation vs Continuation | Smoking_Frequency | Smoking_FrequencySome Days | 0.919 | 0.669 | 1.263 | 6.03e-01 | 1.00e+00 | 2732 | 1973 | 759 |
| African | Recorded Anxiety | Switching vs Continuation | Smoking_Frequency | (Intercept) | 0.994 | 0.785 | 1.257 | 9.56e-01 | 1.00e+00 | 3804 | 1973 | 1831 |
| African | Recorded Anxiety | Switching vs Continuation | Smoking_Frequency | age_at_first_AD | 0.790 | 0.729 | 0.855 | 6.01e-09 | 8.41e-08 | 3804 | 1973 | 1831 |
| African | Recorded Anxiety | Switching vs Continuation | Smoking_Frequency | sex_at_birthMale | 1.012 | 0.861 | 1.189 | 8.89e-01 | 1.00e+00 | 3804 | 1973 | 1831 |
| African | Recorded Anxiety | Switching vs Continuation | Smoking_Frequency | Income>200k | 0.603 | 0.300 | 1.209 | 1.54e-01 | 1.00e+00 | 3804 | 1973 | 1831 |
| African | Recorded Anxiety | Switching vs Continuation | Smoking_Frequency | Income100-150k | 0.592 | 0.424 | 0.825 | 2.00e-03 | 2.80e-02 | 3804 | 1973 | 1831 |
| African | Recorded Anxiety | Switching vs Continuation | Smoking_Frequency | Income150-200k | 0.582 | 0.311 | 1.088 | 8.99e-02 | 1.00e+00 | 3804 | 1973 | 1831 |
| African | Recorded Anxiety | Switching vs Continuation | Smoking_Frequency | Income25-35k | 0.774 | 0.623 | 0.963 | 2.15e-02 | 3.00e-01 | 3804 | 1973 | 1831 |
| African | Recorded Anxiety | Switching vs Continuation | Smoking_Frequency | Income35-50k | 0.749 | 0.605 | 0.926 | 7.61e-03 | 1.07e-01 | 3804 | 1973 | 1831 |
| African | Recorded Anxiety | Switching vs Continuation | Smoking_Frequency | Income50-75k | 0.688 | 0.544 | 0.870 | 1.83e-03 | 2.57e-02 | 3804 | 1973 | 1831 |
| African | Recorded Anxiety | Switching vs Continuation | Smoking_Frequency | Income75-100k | 0.583 | 0.425 | 0.800 | 8.38e-04 | 1.17e-02 | 3804 | 1973 | 1831 |
| African | Recorded Anxiety | Switching vs Continuation | Smoking_Frequency | Education_levelCollege Postgraduate | 1.007 | 0.732 | 1.385 | 9.68e-01 | 1.00e+00 | 3804 | 1973 | 1831 |
| African | Recorded Anxiety | Switching vs Continuation | Smoking_Frequency | Education_levelCollege Undergraduate | 0.989 | 0.775 | 1.262 | 9.31e-01 | 1.00e+00 | 3804 | 1973 | 1831 |
| African | Recorded Anxiety | Switching vs Continuation | Smoking_Frequency | Education_levelTwelve or GED | 0.864 | 0.672 | 1.112 | 2.57e-01 | 1.00e+00 | 3804 | 1973 | 1831 |
| African | Recorded Anxiety | Switching vs Continuation | Smoking_Frequency | Smoking_FrequencyEvery Day | 1.426 | 1.173 | 1.732 | 3.60e-04 | 5.04e-03 | 3804 | 1973 | 1831 |
| African | Recorded Anxiety | Switching vs Continuation | Smoking_Frequency | Smoking_FrequencyNot At All (former) | 1.165 | 0.980 | 1.385 | 8.36e-02 | 1.00e+00 | 3804 | 1973 | 1831 |
| African | Recorded Anxiety | Switching vs Continuation | Smoking_Frequency | Smoking_FrequencySome Days | 1.023 | 0.804 | 1.303 | 8.51e-01 | 1.00e+00 | 3804 | 1973 | 1831 |
| African | Recorded Anxiety | Discontinuation vs Continuation | Stress | (Intercept) | 0.235 | 0.104 | 0.530 | 4.88e-04 | 6.83e-03 | 854 | 626 | 228 |
| African | Recorded Anxiety | Discontinuation vs Continuation | Stress | age_at_first_AD | 1.059 | 0.883 | 1.270 | 5.35e-01 | 1.00e+00 | 854 | 626 | 228 |
| African | Recorded Anxiety | Discontinuation vs Continuation | Stress | sex_at_birthMale | 1.001 | 0.677 | 1.482 | 9.95e-01 | 1.00e+00 | 854 | 626 | 228 |
| African | Recorded Anxiety | Discontinuation vs Continuation | Stress | Income>200k | 0.758 | 0.230 | 2.500 | 6.49e-01 | 1.00e+00 | 854 | 626 | 228 |
| African | Recorded Anxiety | Discontinuation vs Continuation | Stress | Income100-150k | 0.513 | 0.253 | 1.040 | 6.41e-02 | 8.98e-01 | 854 | 626 | 228 |

| | | | | | | | | | | | | |
|---|---|---|---|---|---|---|---|---|---|---|---|---|
| African | Recorded Anxiety | Discontinuation vs Continuation | Stress | Income150-200k | 1.671 | 0.606 | 4.607 | 3.21e-01 | 1.00e+00 | 854 | 626 | 228 |
| African | Recorded Anxiety | Discontinuation vs Continuation | Stress | Income25-35k | 1.492 | 0.894 | 2.489 | 1.26e-01 | 1.00e+00 | 854 | 626 | 228 |
| African | Recorded Anxiety | Discontinuation vs Continuation | Stress | Income35-50k | 0.692 | 0.416 | 1.149 | 1.55e-01 | 1.00e+00 | 854 | 626 | 228 |
| African | Recorded Anxiety | Discontinuation vs Continuation | Stress | Income50-75k | 1.155 | 0.729 | 1.830 | 5.38e-01 | 1.00e+00 | 854 | 626 | 228 |
| African | Recorded Anxiety | Discontinuation vs Continuation | Stress | Income75-100k | 0.621 | 0.310 | 1.242 | 1.78e-01 | 1.00e+00 | 854 | 626 | 228 |
| African | Recorded Anxiety | Discontinuation vs Continuation | Stress | Education_levelCollege Postgraduate | 1.429 | 0.571 | 3.579 | 4.46e-01 | 1.00e+00 | 854 | 626 | 228 |
| African | Recorded Anxiety | Discontinuation vs Continuation | Stress | Education_levelCollege Undergraduate | 1.266 | 0.551 | 2.914 | 5.78e-01 | 1.00e+00 | 854 | 626 | 228 |
| African | Recorded Anxiety | Discontinuation vs Continuation | Stress | Education_levelTwelve or GED | 1.682 | 0.705 | 4.016 | 2.41e-01 | 1.00e+00 | 854 | 626 | 228 |
| African | Recorded Anxiety | Discontinuation vs Continuation | Stress | StressAlmost Never | 2.108 | 1.064 | 4.179 | 3.26e-02 | 4.56e-01 | 854 | 626 | 228 |
| African | Recorded Anxiety | Discontinuation vs Continuation | Stress | StressFairly Often | 1.108 | 0.546 | 2.246 | 7.77e-01 | 1.00e+00 | 854 | 626 | 228 |
| African | Recorded Anxiety | Discontinuation vs Continuation | Stress | StressSometimes | 1.363 | 0.967 | 1.921 | 7.69e-02 | 1.00e+00 | 854 | 626 | 228 |
| African | Recorded Anxiety | Discontinuation vs Continuation | Stress | StressVery Often | 1.355 | 0.659 | 2.788 | 4.09e-01 | 1.00e+00 | 854 | 626 | 228 |
| African | Recorded Anxiety | Switching vs Continuation | Stress | (Intercept) | 1.005 | 0.589 | 1.717 | 9.84e-01 | 1.00e+00 | 1207 | 626 | 581 |
| African | Recorded Anxiety | Switching vs Continuation | Stress | age_at_first_AD | 0.677 | 0.585 | 0.783 | 1.42e-07 | 1.99e-06 | 1207 | 626 | 581 |
| African | Recorded Anxiety | Switching vs Continuation | Stress | sex_at_birthMale | 1.122 | 0.836 | 1.507 | 4.43e-01 | 1.00e+00 | 1207 | 626 | 581 |
| African | Recorded Anxiety | Switching vs Continuation | Stress | Income>200k | 0.285 | 0.098 | 0.828 | 2.11e-02 | 2.95e-01 | 1207 | 626 | 581 |
| African | Recorded Anxiety | Switching vs Continuation | Stress | Income100-150k | 0.569 | 0.353 | 0.917 | 2.04e-02 | 2.86e-01 | 1207 | 626 | 581 |
| African | Recorded Anxiety | Switching vs Continuation | Stress | Income150-200k | 0.837 | 0.340 | 2.064 | 7.00e-01 | 1.00e+00 | 1207 | 626 | 581 |
| African | Recorded Anxiety | Switching vs Continuation | Stress | Income25-35k | 0.908 | 0.603 | 1.368 | 6.46e-01 | 1.00e+00 | 1207 | 626 | 581 |
| African | Recorded Anxiety | Switching vs Continuation | Stress | Income35-50k | 0.617 | 0.428 | 0.891 | 9.87e-03 | 1.38e-01 | 1207 | 626 | 581 |
| African | Recorded Anxiety | Switching vs Continuation | Stress | Income50-75k | 0.693 | 0.479 | 1.002 | 5.13e-02 | 7.18e-01 | 1207 | 626 | 581 |
| African | Recorded Anxiety | Switching vs Continuation | Stress | Income75-100k | 0.614 | 0.381 | 0.990 | 4.51e-02 | 6.32e-01 | 1207 | 626 | 581 |
| African | Recorded Anxiety | Switching vs Continuation | Stress | Education_levelCollege Postgraduate | 1.070 | 0.574 | 1.995 | 8.31e-01 | 1.00e+00 | 1207 | 626 | 581 |
| African | Recorded Anxiety | Switching vs Continuation | Stress | Education_levelCollege Undergraduate | 0.879 | 0.507 | 1.524 | 6.46e-01 | 1.00e+00 | 1207 | 626 | 581 |
| African | Recorded Anxiety | Switching vs Continuation | Stress | Education_levelTwelve or GED | 1.024 | 0.570 | 1.842 | 9.36e-01 | 1.00e+00 | 1207 | 626 | 581 |
| African | Recorded Anxiety | Switching vs Continuation | Stress | StressAlmost Never | 1.513 | 0.866 | 2.645 | 1.46e-01 | 1.00e+00 | 1207 | 626 | 581 |
| African | Recorded Anxiety | Switching vs Continuation | Stress | StressFairly Often | 1.146 | 0.693 | 1.897 | 5.95e-01 | 1.00e+00 | 1207 | 626 | 581 |
| African | Recorded Anxiety | Switching vs Continuation | Stress | StressSometimes | 1.227 | 0.952 | 1.583 | 1.15e-01 | 1.00e+00 | 1207 | 626 | 581 |
| African | Recorded Anxiety | Switching vs Continuation | Stress | StressVery Often | 1.134 | 0.654 | 1.965 | 6.54e-01 | 1.00e+00 | 1207 | 626 | 581 |
| African | Recorded Anxiety | Discontinuation vs Continuation | age_at_first_AD | (Intercept) | 0.365 | 0.331 | 0.402 | <2.2e-16 | <2.2e-16 | 2793 | 2014 | 779 |
| African | Recorded Anxiety | Discontinuation vs Continuation | age_at_first_AD | age_at_first_AD | 0.942 | 0.858 | 1.035 | 2.14e-01 | 1.00e+00 | 2793 | 2014 | 779 |
| African | Recorded Anxiety | Discontinuation vs Continuation | age_at_first_AD | sex_at_birthMale | 1.239 | 1.020 | 1.506 | 3.07e-02 | 4.30e-01 | 2793 | 2014 | 779 |
| African | Recorded Anxiety | Switching vs Continuation | age_at_first_AD | (Intercept) | 0.889 | 0.826 | 0.957 | 1.77e-03 | 2.48e-02 | 3909 | 2014 | 1895 |
| African | Recorded Anxiety | Switching vs Continuation | age_at_first_AD | age_at_first_AD | 0.812 | 0.753 | 0.875 | 6.25e-08 | 8.75e-07 | 3909 | 2014 | 1895 |
| African | Recorded Anxiety | Switching vs Continuation | age_at_first_AD | sex_at_birthMale | 1.070 | 0.917 | 1.248 | 3.90e-01 | 1.00e+00 | 3909 | 2014 | 1895 |
| African | Recorded Anxiety | Discontinuation vs Continuation | sex_at_birth | (Intercept) | 0.365 | 0.331 | 0.402 | <2.2e-16 | <2.2e-16 | 2793 | 2014 | 779 |
| African | Recorded Anxiety | Discontinuation vs Continuation | sex_at_birth | age_at_first_AD | 0.942 | 0.858 | 1.035 | 2.14e-01 | 1.00e+00 | 2793 | 2014 | 779 |
| African | Recorded Anxiety | Discontinuation vs Continuation | sex_at_birth | sex_at_birthMale | 1.239 | 1.020 | 1.506 | 3.07e-02 | 4.30e-01 | 2793 | 2014 | 779 |
| African | Recorded Anxiety | Switching vs Continuation | sex_at_birth | (Intercept) | 0.889 | 0.826 | 0.957 | 1.77e-03 | 2.48e-02 | 3909 | 2014 | 1895 |
| African | Recorded Anxiety | Switching vs Continuation | sex_at_birth | age_at_first_AD | 0.812 | 0.753 | 0.875 | 6.25e-08 | 8.75e-07 | 3909 | 2014 | 1895 |
| African | Recorded Anxiety | Switching vs Continuation | sex_at_birth | sex_at_birthMale | 1.070 | 0.917 | 1.248 | 3.90e-01 | 1.00e+00 | 3909 | 2014 | 1895 |
| African | CIDI-noMDD | Discontinuation vs Continuation | Alcohol_Frequency | (Intercept) | 1.498 | 0.137 | 16.340 | 7.40e-01 | 1.00e+00 | 159 | 122 | 37 |
| African | CIDI-noMDD | Discontinuation vs Continuation | Alcohol_Frequency | age_at_first_AD | 0.968 | 0.585 | 1.604 | 9.01e-01 | 1.00e+00 | 159 | 122 | 37 |
| African | CIDI-noMDD | Discontinuation vs Continuation | Alcohol_Frequency | sex_at_birthMale | 4.475 | 1.603 | 12.490 | 4.23e-03 | 5.92e-02 | 159 | 122 | 37 |
| African | CIDI-noMDD | Discontinuation vs Continuation | Alcohol_Frequency | Income>200k | 5.797 | 0.793 | 42.390 | 8.34e-02 | 1.00e+00 | 159 | 122 | 37 |
| African | CIDI-noMDD | Discontinuation vs Continuation | Alcohol_Frequency | Income100-150k | 1.200 | 0.254 | 5.669 | 8.18e-01 | 1.00e+00 | 159 | 122 | 37 |
| African | CIDI-noMDD | Discontinuation vs Continuation | Alcohol_Frequency | Income150-200k | 1.026 | 0.064 | 16.350 | 9.86e-01 | 1.00e+00 | 159 | 122 | 37 |
| African | CIDI-noMDD | Discontinuation vs Continuation | Alcohol_Frequency | Income25-35k | 2.527 | 0.657 | 9.722 | 1.77e-01 | 1.00e+00 | 159 | 122 | 37 |
| African | CIDI-noMDD | Discontinuation vs Continuation | Alcohol_Frequency | Income35-50k | 1.377 | 0.282 | 6.734 | 6.93e-01 | 1.00e+00 | 159 | 122 | 37 |
| African | CIDI-noMDD | Discontinuation vs Continuation | Alcohol_Frequency | Income50-75k | 1.415 | 0.371 | 5.393 | 6.11e-01 | 1.00e+00 | 159 | 122 | 37 |
| African | CIDI-noMDD | Discontinuation vs Continuation | Alcohol_Frequency | Income75-100k | 3.640 | 0.857 | 15.470 | 8.01e-02 | 1.00e+00 | 159 | 122 | 37 |
| African | CIDI-noMDD | Discontinuation vs Continuation | Alcohol_Frequency | Education_levelCollege Postgraduate | 0.238 | 0.021 | 2.695 | 2.46e-01 | 1.00e+00 | 159 | 122 | 37 |
| African | CIDI-noMDD | Discontinuation vs Continuation | Alcohol_Frequency | Education_levelCollege Undergraduate | 0.251 | 0.027 | 2.339 | 2.25e-01 | 1.00e+00 | 159 | 122 | 37 |
| African | CIDI-noMDD | Discontinuation vs Continuation | Alcohol_Frequency | Education_levelTwelve or GED | 0.142 | 0.010 | 2.027 | 1.50e-01 | 1.00e+00 | 159 | 122 | 37 |
| African | CIDI-noMDD | Discontinuation vs Continuation | Alcohol_Frequency | Alcohol_Frequency2-3 Week | 0.147 | 0.020 | 1.059 | 5.70e-02 | 7.98e-01 | 159 | 122 | 37 |
| African | CIDI-noMDD | Discontinuation vs Continuation | Alcohol_Frequency | Alcohol_Frequency2-4 Month | 0.519 | 0.090 | 2.989 | 4.63e-01 | 1.00e+00 | 159 | 122 | 37 |
| African | CIDI-noMDD | Discontinuation vs Continuation | Alcohol_Frequency | Alcohol_Frequency4plus Week | 0.283 | 0.029 | 2.810 | 2.81e-01 | 1.00e+00 | 159 | 122 | 37 |
| African | CIDI-noMDD | Discontinuation vs Continuation | Alcohol_Frequency | Alcohol_FrequencyMonthly or Less | 0.315 | 0.067 | 1.479 | 1.43e-01 | 1.00e+00 | 159 | 122 | 37 |
| African | CIDI-noMDD | Discontinuation vs Continuation | Alcohol_Frequency | Alcohol_FrequencyNever (former) | 0.390 | 0.067 | 2.254 | 2.93e-01 | 1.00e+00 | 159 | 122 | 37 |
| African | CIDI-noMDD | Discontinuation vs Continuation | BMI | (Intercept) | 0.897 | 0.100 | 8.044 | 9.22e-01 | 1.00e+00 | 158 | 121 | 37 |
| African | CIDI-noMDD | Discontinuation vs Continuation | BMI | age_at_first_AD | 0.984 | 0.607 | 1.595 | 9.49e-01 | 1.00e+00 | 158 | 121 | 37 |
| African | CIDI-noMDD | Discontinuation vs Continuation | BMI | sex_at_birthMale | 2.967 | 1.103 | 7.978 | 3.12e-02 | 4.36e-01 | 158 | 121 | 37 |
| African | CIDI-noMDD | Discontinuation vs Continuation | BMI | Income>200k | 6.471 | 0.804 | 52.060 | 7.92e-02 | 1.00e+00 | 158 | 121 | 37 |
| African | CIDI-noMDD | Discontinuation vs Continuation | BMI | Income100-150k | 0.807 | 0.176 | 3.694 | 7.82e-01 | 1.00e+00 | 158 | 121 | 37 |
| African | CIDI-noMDD | Discontinuation vs Continuation | BMI | Income150-200k | 0.936 | 0.066 | 13.230 | 9.61e-01 | 1.00e+00 | 158 | 121 | 37 |
| African | CIDI-noMDD | Discontinuation vs Continuation | BMI | Income25-35k | 2.043 | 0.550 | 7.591 | 2.86e-01 | 1.00e+00 | 158 | 121 | 37 |

| | | | | | | | | | | | | |
|---|---|---|---|---|---|---|---|---|---|---|---|---|
| African | CIDI-noMDD | Discontinuation vs Continuation | BMI | Income35-50k | 0.934 | 0.195 | 4.461 | 9.32e-01 | 1.00e+00 | 158 | 121 | 37 |
| African | CIDI-noMDD | Discontinuation vs Continuation | BMI | Income50-75k | 1.155 | 0.310 | 4.300 | 8.29e-01 | 1.00e+00 | 158 | 121 | 37 |
| African | CIDI-noMDD | Discontinuation vs Continuation | BMI | Income75-100k | 3.027 | 0.736 | 12.450 | 1.25e-01 | 1.00e+00 | 158 | 121 | 37 |
| African | CIDI-noMDD | Discontinuation vs Continuation | BMI | Education_levelCollege Postgraduate | 0.224 | 0.019 | 2.615 | 2.33e-01 | 1.00e+00 | 158 | 121 | 37 |
| African | CIDI-noMDD | Discontinuation vs Continuation | BMI | Education_levelCollege Undergraduate | 0.207 | 0.022 | 1.981 | 1.72e-01 | 1.00e+00 | 158 | 121 | 37 |
| African | CIDI-noMDD | Discontinuation vs Continuation | BMI | Education_levelTwelve or GED | 0.125 | 0.008 | 1.874 | 1.32e-01 | 1.00e+00 | 158 | 121 | 37 |
| African | CIDI-noMDD | Discontinuation vs Continuation | BMI | BMI | 0.688 | 0.430 | 1.100 | 1.19e-01 | 1.00e+00 | 158 | 121 | 37 |
| African | CIDI-noMDD | Discontinuation vs Continuation | Insomnia | (Intercept) | 0.566 | 0.068 | 4.698 | 5.98e-01 | 1.00e+00 | 160 | 123 | 37 |
| African | CIDI-noMDD | Discontinuation vs Continuation | Insomnia | age_at_first_AD | 0.950 | 0.578 | 1.562 | 8.40e-01 | 1.00e+00 | 160 | 123 | 37 |
| African | CIDI-noMDD | Discontinuation vs Continuation | Insomnia | sex_at_birthMale | 3.756 | 1.439 | 9.803 | 6.85e-03 | 9.60e-02 | 160 | 123 | 37 |
| African | CIDI-noMDD | Discontinuation vs Continuation | Insomnia | Income>200k | 4.019 | 0.616 | 26.240 | 1.46e-01 | 1.00e+00 | 160 | 123 | 37 |
| African | CIDI-noMDD | Discontinuation vs Continuation | Insomnia | Income100-150k | 0.860 | 0.189 | 3.907 | 8.45e-01 | 1.00e+00 | 160 | 123 | 37 |
| African | CIDI-noMDD | Discontinuation vs Continuation | Insomnia | Income150-200k | 0.886 | 0.060 | 13.080 | 9.30e-01 | 1.00e+00 | 160 | 123 | 37 |
| African | CIDI-noMDD | Discontinuation vs Continuation | Insomnia | Income25-35k | 1.867 | 0.510 | 6.833 | 3.46e-01 | 1.00e+00 | 160 | 123 | 37 |
| African | CIDI-noMDD | Discontinuation vs Continuation | Insomnia | Income35-50k | 0.959 | 0.201 | 4.563 | 9.58e-01 | 1.00e+00 | 160 | 123 | 37 |
| African | CIDI-noMDD | Discontinuation vs Continuation | Insomnia | Income50-75k | 0.988 | 0.250 | 3.912 | 9.87e-01 | 1.00e+00 | 160 | 123 | 37 |
| African | CIDI-noMDD | Discontinuation vs Continuation | Insomnia | Income75-100k | 3.055 | 0.753 | 12.400 | 1.18e-01 | 1.00e+00 | 160 | 123 | 37 |
| African | CIDI-noMDD | Discontinuation vs Continuation | Insomnia | Education_levelCollege Postgraduate | 0.251 | 0.024 | 2.668 | 2.52e-01 | 1.00e+00 | 160 | 123 | 37 |
| African | CIDI-noMDD | Discontinuation vs Continuation | Insomnia | Education_levelCollege Undergraduate | 0.261 | 0.029 | 2.303 | 2.26e-01 | 1.00e+00 | 160 | 123 | 37 |
| African | CIDI-noMDD | Discontinuation vs Continuation | Insomnia | Education_levelTwelve or GED | 0.153 | 0.011 | 2.074 | 1.58e-01 | 1.00e+00 | 160 | 123 | 37 |
| African | CIDI-noMDD | Discontinuation vs Continuation | Insomnia | Insomnia | 1.769 | 0.678 | 4.611 | 2.44e-01 | 1.00e+00 | 160 | 123 | 37 |
| African | CIDI-noMDD | Discontinuation vs Continuation | MDD_symptom_count | (Intercept) | 0.018 | 0.000 | 0.861 | 4.18e-02 | 5.85e-01 | 160 | 123 | 37 |
| African | CIDI-noMDD | Discontinuation vs Continuation | MDD_symptom_count | age_at_first_AD | 1.025 | 0.626 | 1.680 | 9.21e-01 | 1.00e+00 | 160 | 123 | 37 |
| African | CIDI-noMDD | Discontinuation vs Continuation | MDD_symptom_count | sex_at_birthMale | 5.041 | 1.793 | 14.170 | 2.16e-03 | 3.03e-02 | 160 | 123 | 37 |
| African | CIDI-noMDD | Discontinuation vs Continuation | MDD_symptom_count | Income>200k | 3.182 | 0.449 | 22.580 | 2.47e-01 | 1.00e+00 | 160 | 123 | 37 |
| African | CIDI-noMDD | Discontinuation vs Continuation | MDD_symptom_count | Income100-150k | 0.665 | 0.140 | 3.157 | 6.08e-01 | 1.00e+00 | 160 | 123 | 37 |
| African | CIDI-noMDD | Discontinuation vs Continuation | MDD_symptom_count | Income150-200k | 0.503 | 0.030 | 8.503 | 6.34e-01 | 1.00e+00 | 160 | 123 | 37 |
| African | CIDI-noMDD | Discontinuation vs Continuation | MDD_symptom_count | Income25-35k | 1.550 | 0.405 | 5.937 | 5.23e-01 | 1.00e+00 | 160 | 123 | 37 |
| African | CIDI-noMDD | Discontinuation vs Continuation | MDD_symptom_count | Income35-50k | 1.062 | 0.224 | 5.034 | 9.40e-01 | 1.00e+00 | 160 | 123 | 37 |
| African | CIDI-noMDD | Discontinuation vs Continuation | MDD_symptom_count | Income50-75k | 0.994 | 0.260 | 3.798 | 9.93e-01 | 1.00e+00 | 160 | 123 | 37 |
| African | CIDI-noMDD | Discontinuation vs Continuation | MDD_symptom_count | Income75-100k | 2.287 | 0.551 | 9.486 | 2.54e-01 | 1.00e+00 | 160 | 123 | 37 |
| African | CIDI-noMDD | Discontinuation vs Continuation | MDD_symptom_count | Education_levelCollege Postgraduate | 0.426 | 0.037 | 4.850 | 4.91e-01 | 1.00e+00 | 160 | 123 | 37 |
| African | CIDI-noMDD | Discontinuation vs Continuation | MDD_symptom_count | Education_levelCollege Undergraduate | 0.305 | 0.033 | 2.806 | 2.95e-01 | 1.00e+00 | 160 | 123 | 37 |
| African | CIDI-noMDD | Discontinuation vs Continuation | MDD_symptom_count | Education_levelTwelve or GED | 0.173 | 0.012 | 2.448 | 1.94e-01 | 1.00e+00 | 160 | 123 | 37 |
| African | CIDI-noMDD | Discontinuation vs Continuation | MDD_symptom_count | MDD_symptom_count | 0.105 | 0.015 | 0.754 | 2.50e-02 | 3.50e-01 | 160 | 123 | 37 |
| African | CIDI-noMDD | Discontinuation vs Continuation | Migraine | (Intercept) | 0.640 | 0.076 | 5.394 | 6.82e-01 | 1.00e+00 | 160 | 123 | 37 |
| African | CIDI-noMDD | Discontinuation vs Continuation | Migraine | age_at_first_AD | 1.013 | 0.621 | 1.653 | 9.57e-01 | 1.00e+00 | 160 | 123 | 37 |
| African | CIDI-noMDD | Discontinuation vs Continuation | Migraine | sex_at_birthMale | 3.952 | 1.493 | 10.460 | 5.66e-03 | 7.92e-02 | 160 | 123 | 37 |
| African | CIDI-noMDD | Discontinuation vs Continuation | Migraine | Income>200k | 4.135 | 0.626 | 27.330 | 1.41e-01 | 1.00e+00 | 160 | 123 | 37 |
| African | CIDI-noMDD | Discontinuation vs Continuation | Migraine | Income100-150k | 0.964 | 0.216 | 4.313 | 9.62e-01 | 1.00e+00 | 160 | 123 | 37 |
| African | CIDI-noMDD | Discontinuation vs Continuation | Migraine | Income150-200k | 0.966 | 0.067 | 13.860 | 9.79e-01 | 1.00e+00 | 160 | 123 | 37 |
| African | CIDI-noMDD | Discontinuation vs Continuation | Migraine | Income25-35k | 1.989 | 0.544 | 7.278 | 2.99e-01 | 1.00e+00 | 160 | 123 | 37 |
| African | CIDI-noMDD | Discontinuation vs Continuation | Migraine | Income35-50k | 1.154 | 0.250 | 5.335 | 8.54e-01 | 1.00e+00 | 160 | 123 | 37 |
| African | CIDI-noMDD | Discontinuation vs Continuation | Migraine | Income50-75k | 1.207 | 0.324 | 4.496 | 7.79e-01 | 1.00e+00 | 160 | 123 | 37 |
| African | CIDI-noMDD | Discontinuation vs Continuation | Migraine | Income75-100k | 3.284 | 0.822 | 13.130 | 9.26e-02 | 1.00e+00 | 160 | 123 | 37 |
| African | CIDI-noMDD | Discontinuation vs Continuation | Migraine | Education_levelCollege Postgraduate | 0.223 | 0.021 | 2.375 | 2.14e-01 | 1.00e+00 | 160 | 123 | 37 |
| African | CIDI-noMDD | Discontinuation vs Continuation | Migraine | Education_levelCollege Undergraduate | 0.220 | 0.025 | 1.941 | 1.73e-01 | 1.00e+00 | 160 | 123 | 37 |
| African | CIDI-noMDD | Discontinuation vs Continuation | Migraine | Education_levelTwelve or GED | 0.133 | 0.010 | 1.799 | 1.29e-01 | 1.00e+00 | 160 | 123 | 37 |
| African | CIDI-noMDD | Discontinuation vs Continuation | Migraine | Migraine | 1.257 | 0.482 | 3.278 | 6.40e-01 | 1.00e+00 | 160 | 123 | 37 |
| African | CIDI-noMDD | Discontinuation vs Continuation | Recorded Anxiety | (Intercept) | 0.669 | 0.078 | 5.721 | 7.13e-01 | 1.00e+00 | 160 | 123 | 37 |
| African | CIDI-noMDD | Discontinuation vs Continuation | Recorded Anxiety | age_at_first_AD | 1.007 | 0.616 | 1.646 | 9.77e-01 | 1.00e+00 | 160 | 123 | 37 |
| African | CIDI-noMDD | Discontinuation vs Continuation | Recorded Anxiety | sex_at_birthMale | 3.761 | 1.446 | 9.783 | 6.62e-03 | 9.26e-02 | 160 | 123 | 37 |
| African | CIDI-noMDD | Discontinuation vs Continuation | Recorded Anxiety | Income>200k | 4.204 | 0.637 | 27.760 | 1.36e-01 | 1.00e+00 | 160 | 123 | 37 |
| African | CIDI-noMDD | Discontinuation vs Continuation | Recorded Anxiety | Income100-150k | 0.934 | 0.209 | 4.182 | 9.29e-01 | 1.00e+00 | 160 | 123 | 37 |
| African | CIDI-noMDD | Discontinuation vs Continuation | Recorded Anxiety | Income150-200k | 1.002 | 0.071 | 14.090 | 9.99e-01 | 1.00e+00 | 160 | 123 | 37 |
| African | CIDI-noMDD | Discontinuation vs Continuation | Recorded Anxiety | Income25-35k | 2.018 | 0.551 | 7.389 | 2.89e-01 | 1.00e+00 | 160 | 123 | 37 |
| African | CIDI-noMDD | Discontinuation vs Continuation | Recorded Anxiety | Income35-50k | 1.117 | 0.240 | 5.195 | 8.87e-01 | 1.00e+00 | 160 | 123 | 37 |
| African | CIDI-noMDD | Discontinuation vs Continuation | Recorded Anxiety | Income50-75k | 1.217 | 0.326 | 4.547 | 7.70e-01 | 1.00e+00 | 160 | 123 | 37 |
| African | CIDI-noMDD | Discontinuation vs Continuation | Recorded Anxiety | Income75-100k | 3.259 | 0.808 | 13.150 | 9.69e-02 | 1.00e+00 | 160 | 123 | 37 |
| African | CIDI-noMDD | Discontinuation vs Continuation | Recorded Anxiety | Education_levelCollege Postgraduate | 0.229 | 0.021 | 2.451 | 2.23e-01 | 1.00e+00 | 160 | 123 | 37 |
| African | CIDI-noMDD | Discontinuation vs Continuation | Recorded Anxiety | Education_levelCollege Undergraduate | 0.218 | 0.025 | 1.931 | 1.71e-01 | 1.00e+00 | 160 | 123 | 37 |
| African | CIDI-noMDD | Discontinuation vs Continuation | Recorded Anxiety | Education_levelTwelve or GED | 0.132 | 0.010 | 1.792 | 1.28e-01 | 1.00e+00 | 160 | 123 | 37 |
| African | CIDI-noMDD | Discontinuation vs Continuation | Recorded Anxiety | Recorded Anxiety | 1.067 | 0.469 | 2.429 | 8.77e-01 | 1.00e+00 | 160 | 123 | 37 |
| African | CIDI-noMDD | Discontinuation vs Continuation | age_at_first_AD | (Intercept) | 0.225 | 0.142 | 0.355 | 1.70e-10 | 2.38e-09 | 160 | 123 | 37 |
| African | CIDI-noMDD | Discontinuation vs Continuation | age_at_first_AD | age_at_first_AD | 0.996 | 0.627 | 1.581 | 9.86e-01 | 1.00e+00 | 160 | 123 | 37 |
| African | CIDI-noMDD | Discontinuation vs Continuation | age_at_first_AD | sex_at_birthMale | 3.621 | 1.537 | 8.528 | 3.24e-03 | 4.54e-02 | 160 | 123 | 37 |

| | | | | | | | | | | | | |
|---|---|---|---|---|---|---|---|---|---|---|---|---|
| African | CIDI-noMDD | Discontinuation vs Continuation | sex_at_birth | (Intercept) | 0.225 | 0.142 | 0.355 | 1.70e-10 | 2.38e-09 | 160 | 123 | 37 |
| African | CIDI-noMDD | Discontinuation vs Continuation | sex_at_birth | age_at_first_AD | 0.996 | 0.627 | 1.581 | 9.86e-01 | 1.00e+00 | 160 | 123 | 37 |
| African | CIDI-noMDD | Discontinuation vs Continuation | sex_at_birth | sex_at_birthMale | 3.621 | 1.537 | 8.528 | 3.24e-03 | 4.54e-02 | 160 | 123 | 37 |
| Latino/Mixed-American | Full Cohort | Discontinuation vs Continuation | Alcohol_Frequency | (Intercept) | 0.450 | 0.362 | 0.559 | 5.09e-13 | 7.12e-12 | 3772 | 2405 | 1367 |
| Latino/Mixed-American | Full Cohort | Discontinuation vs Continuation | Alcohol_Frequency | age_at_first_AD | 0.988 | 0.919 | 1.063 | 7.54e-01 | 1.00e+00 | 3772 | 2405 | 1367 |
| Latino/Mixed-American | Full Cohort | Discontinuation vs Continuation | Alcohol_Frequency | sex_at_birthMale | 1.204 | 1.036 | 1.400 | 1.53e-02 | 2.14e-01 | 3772 | 2405 | 1367 |
| Latino/Mixed-American | Full Cohort | Discontinuation vs Continuation | Alcohol_Frequency | Income>200k | 0.607 | 0.379 | 0.971 | 3.74e-02 | 5.24e-01 | 3772 | 2405 | 1367 |
| Latino/Mixed-American | Full Cohort | Discontinuation vs Continuation | Alcohol_Frequency | Income100-150k | 0.686 | 0.512 | 0.921 | 1.21e-02 | 1.70e-01 | 3772 | 2405 | 1367 |
| Latino/Mixed-American | Full Cohort | Discontinuation vs Continuation | Alcohol_Frequency | Income150-200k | 0.716 | 0.468 | 1.094 | 1.22e-01 | 1.00e+00 | 3772 | 2405 | 1367 |
| Latino/Mixed-American | Full Cohort | Discontinuation vs Continuation | Alcohol_Frequency | Income25-35k | 0.810 | 0.649 | 1.012 | 6.32e-02 | 8.85e-01 | 3772 | 2405 | 1367 |
| Latino/Mixed-American | Full Cohort | Discontinuation vs Continuation | Alcohol_Frequency | Income35-50k | 1.056 | 0.839 | 1.331 | 6.41e-01 | 1.00e+00 | 3772 | 2405 | 1367 |
| Latino/Mixed-American | Full Cohort | Discontinuation vs Continuation | Alcohol_Frequency | Income50-75k | 0.843 | 0.669 | 1.061 | 1.45e-01 | 1.00e+00 | 3772 | 2405 | 1367 |
| Latino/Mixed-American | Full Cohort | Discontinuation vs Continuation | Alcohol_Frequency | Income75-100k | 0.753 | 0.552 | 1.027 | 7.28e-02 | 1.00e+00 | 3772 | 2405 | 1367 |
| Latino/Mixed-American | Full Cohort | Discontinuation vs Continuation | Alcohol_Frequency | Education_levelCollege Postgraduate | 0.944 | 0.690 | 1.291 | 7.18e-01 | 1.00e+00 | 3772 | 2405 | 1367 |
| Latino/Mixed-American | Full Cohort | Discontinuation vs Continuation | Alcohol_Frequency | Education_levelCollege Undergraduate | 1.347 | 1.099 | 1.651 | 4.13e-03 | 5.78e-02 | 3772 | 2405 | 1367 |
| Latino/Mixed-American | Full Cohort | Discontinuation vs Continuation | Alcohol_Frequency | Education_levelTwelve or GED | 1.276 | 1.027 | 1.585 | 2.77e-02 | 3.88e-01 | 3772 | 2405 | 1367 |
| Latino/Mixed-American | Full Cohort | Discontinuation vs Continuation | Alcohol_Frequency | Alcohol_Frequency2-3 Week | 0.978 | 0.714 | 1.340 | 8.91e-01 | 1.00e+00 | 3772 | 2405 | 1367 |
| Latino/Mixed-American | Full Cohort | Discontinuation vs Continuation | Alcohol_Frequency | Alcohol_Frequency2-4 Month | 0.913 | 0.704 | 1.184 | 4.92e-01 | 1.00e+00 | 3772 | 2405 | 1367 |
| Latino/Mixed-American | Full Cohort | Discontinuation vs Continuation | Alcohol_Frequency | Alcohol_Frequency4plus Week | 1.492 | 1.043 | 2.134 | 2.85e-02 | 3.99e-01 | 3772 | 2405 | 1367 |
| Latino/Mixed-American | Full Cohort | Discontinuation vs Continuation | Alcohol_Frequency | Alcohol_FrequencyMonthly or Less | 1.156 | 0.927 | 1.441 | 1.99e-01 | 1.00e+00 | 3772 | 2405 | 1367 |
| Latino/Mixed-American | Full Cohort | Discontinuation vs Continuation | Alcohol_Frequency | Alcohol_FrequencyNever (former) | 1.167 | 0.920 | 1.481 | 2.04e-01 | 1.00e+00 | 3772 | 2405 | 1367 |
| Latino/Mixed-American | Full Cohort | Switching vs Continuation | Alcohol_Frequency | (Intercept) | 0.610 | 0.497 | 0.748 | 2.03e-06 | 2.84e-05 | 3852 | 2405 | 1447 |
| Latino/Mixed-American | Full Cohort | Switching vs Continuation | Alcohol_Frequency | age_at_first_AD | 0.779 | 0.722 | 0.840 | 9.11e-11 | 1.28e-09 | 3852 | 2405 | 1447 |
| Latino/Mixed-American | Full Cohort | Switching vs Continuation | Alcohol_Frequency | sex_at_birthMale | 1.039 | 0.892 | 1.210 | 6.26e-01 | 1.00e+00 | 3852 | 2405 | 1447 |
| Latino/Mixed-American | Full Cohort | Switching vs Continuation | Alcohol_Frequency | Income>200k | 0.509 | 0.316 | 0.820 | 5.49e-03 | 7.68e-02 | 3852 | 2405 | 1447 |
| Latino/Mixed-American | Full Cohort | Switching vs Continuation | Alcohol_Frequency | Income100-150k | 0.773 | 0.584 | 1.025 | 7.33e-02 | 1.00e+00 | 3852 | 2405 | 1447 |
| Latino/Mixed-American | Full Cohort | Switching vs Continuation | Alcohol_Frequency | Income150-200k | 0.678 | 0.446 | 1.032 | 6.97e-02 | 9.76e-01 | 3852 | 2405 | 1447 |
| Latino/Mixed-American | Full Cohort | Switching vs Continuation | Alcohol_Frequency | Income25-35k | 0.817 | 0.657 | 1.017 | 7.02e-02 | 9.82e-01 | 3852 | 2405 | 1447 |
| Latino/Mixed-American | Full Cohort | Switching vs Continuation | Alcohol_Frequency | Income35-50k | 1.048 | 0.834 | 1.318 | 6.87e-01 | 1.00e+00 | 3852 | 2405 | 1447 |
| Latino/Mixed-American | Full Cohort | Switching vs Continuation | Alcohol_Frequency | Income50-75k | 0.789 | 0.627 | 0.994 | 4.41e-02 | 6.18e-01 | 3852 | 2405 | 1447 |
| Latino/Mixed-American | Full Cohort | Switching vs Continuation | Alcohol_Frequency | Income75-100k | 0.929 | 0.693 | 1.245 | 6.20e-01 | 1.00e+00 | 3852 | 2405 | 1447 |
| Latino/Mixed-American | Full Cohort | Switching vs Continuation | Alcohol_Frequency | Education_levelCollege Postgraduate | 1.083 | 0.809 | 1.450 | 5.93e-01 | 1.00e+00 | 3852 | 2405 | 1447 |
| Latino/Mixed-American | Full Cohort | Switching vs Continuation | Alcohol_Frequency | Education_levelCollege Undergraduate | 1.140 | 0.933 | 1.392 | 2.00e-01 | 1.00e+00 | 3852 | 2405 | 1447 |
| Latino/Mixed-American | Full Cohort | Switching vs Continuation | Alcohol_Frequency | Education_levelTwelve or GED | 0.989 | 0.797 | 1.227 | 9.18e-01 | 1.00e+00 | 3852 | 2405 | 1447 |
| Latino/Mixed-American | Full Cohort | Switching vs Continuation | Alcohol_Frequency | Alcohol_Frequency2-3 Week | 0.860 | 0.636 | 1.163 | 3.27e-01 | 1.00e+00 | 3852 | 2405 | 1447 |
| Latino/Mixed-American | Full Cohort | Switching vs Continuation | Alcohol_Frequency | Alcohol_Frequency2-4 Month | 0.724 | 0.562 | 0.932 | 1.23e-02 | 1.72e-01 | 3852 | 2405 | 1447 |
| Latino/Mixed-American | Full Cohort | Switching vs Continuation | Alcohol_Frequency | Alcohol_Frequency4plus Week | 1.219 | 0.857 | 1.735 | 2.71e-01 | 1.00e+00 | 3852 | 2405 | 1447 |
| Latino/Mixed-American | Full Cohort | Switching vs Continuation | Alcohol_Frequency | Alcohol_FrequencyMonthly or Less | 0.892 | 0.720 | 1.104 | 2.93e-01 | 1.00e+00 | 3852 | 2405 | 1447 |
| Latino/Mixed-American | Full Cohort | Switching vs Continuation | Alcohol_Frequency | Alcohol_FrequencyNever (former) | 1.086 | 0.864 | 1.366 | 4.80e-01 | 1.00e+00 | 3852 | 2405 | 1447 |
| Latino/Mixed-American | Full Cohort | Discontinuation vs Continuation | BMI | (Intercept) | 0.465 | 0.394 | 0.549 | <2.2e-16 | <2.2e-16 | 3685 | 2390 | 1295 |
| Latino/Mixed-American | Full Cohort | Discontinuation vs Continuation | BMI | age_at_first_AD | 0.983 | 0.913 | 1.058 | 6.47e-01 | 1.00e+00 | 3685 | 2390 | 1295 |
| Latino/Mixed-American | Full Cohort | Discontinuation vs Continuation | BMI | sex_at_birthMale | 1.265 | 1.088 | 1.471 | 2.25e-03 | 3.15e-02 | 3685 | 2390 | 1295 |
| Latino/Mixed-American | Full Cohort | Discontinuation vs Continuation | BMI | Income>200k | 0.607 | 0.377 | 0.975 | 3.90e-02 | 5.46e-01 | 3685 | 2390 | 1295 |
| Latino/Mixed-American | Full Cohort | Discontinuation vs Continuation | BMI | Income100-150k | 0.676 | 0.502 | 0.908 | 9.43e-03 | 1.32e-01 | 3685 | 2390 | 1295 |
| Latino/Mixed-American | Full Cohort | Discontinuation vs Continuation | BMI | Income150-200k | 0.710 | 0.462 | 1.091 | 1.18e-01 | 1.00e+00 | 3685 | 2390 | 1295 |
| Latino/Mixed-American | Full Cohort | Discontinuation vs Continuation | BMI | Income25-35k | 0.787 | 0.627 | 0.988 | 3.86e-02 | 5.40e-01 | 3685 | 2390 | 1295 |
| Latino/Mixed-American | Full Cohort | Discontinuation vs Continuation | BMI | Income35-50k | 1.092 | 0.867 | 1.376 | 4.53e-01 | 1.00e+00 | 3685 | 2390 | 1295 |
| Latino/Mixed-American | Full Cohort | Discontinuation vs Continuation | BMI | Income50-75k | 0.803 | 0.635 | 1.015 | 6.65e-02 | 9.30e-01 | 3685 | 2390 | 1295 |
| Latino/Mixed-American | Full Cohort | Discontinuation vs Continuation | BMI | Income75-100k | 0.701 | 0.510 | 0.962 | 2.77e-02 | 3.88e-01 | 3685 | 2390 | 1295 |
| Latino/Mixed-American | Full Cohort | Discontinuation vs Continuation | BMI | Education_levelCollege Postgraduate | 0.935 | 0.681 | 1.284 | 6.80e-01 | 1.00e+00 | 3685 | 2390 | 1295 |
| Latino/Mixed-American | Full Cohort | Discontinuation vs Continuation | BMI | Education_levelCollege Undergraduate | 1.360 | 1.108 | 1.669 | 3.22e-03 | 4.51e-02 | 3685 | 2390 | 1295 |
| Latino/Mixed-American | Full Cohort | Discontinuation vs Continuation | BMI | Education_levelTwelve or GED | 1.257 | 1.010 | 1.564 | 4.08e-02 | 5.72e-01 | 3685 | 2390 | 1295 |
| Latino/Mixed-American | Full Cohort | Discontinuation vs Continuation | BMI | BMI | 1.011 | 0.944 | 1.082 | 7.55e-01 | 1.00e+00 | 3685 | 2390 | 1295 |
| Latino/Mixed-American | Full Cohort | Switching vs Continuation | BMI | (Intercept) | 0.577 | 0.494 | 0.674 | 3.88e-12 | 5.43e-11 | 3832 | 2390 | 1442 |
| Latino/Mixed-American | Full Cohort | Switching vs Continuation | BMI | age_at_first_AD | 0.804 | 0.746 | 0.866 | 8.57e-09 | 1.20e-07 | 3832 | 2390 | 1442 |
| Latino/Mixed-American | Full Cohort | Switching vs Continuation | BMI | sex_at_birthMale | 1.063 | 0.914 | 1.237 | 4.28e-01 | 1.00e+00 | 3832 | 2390 | 1442 |
| Latino/Mixed-American | Full Cohort | Switching vs Continuation | BMI | Income>200k | 0.527 | 0.331 | 0.838 | 6.82e-03 | 9.55e-02 | 3832 | 2390 | 1442 |
| Latino/Mixed-American | Full Cohort | Switching vs Continuation | BMI | Income100-150k | 0.752 | 0.569 | 0.993 | 4.48e-02 | 6.27e-01 | 3832 | 2390 | 1442 |
| Latino/Mixed-American | Full Cohort | Switching vs Continuation | BMI | Income150-200k | 0.662 | 0.435 | 1.009 | 5.50e-02 | 7.69e-01 | 3832 | 2390 | 1442 |
| Latino/Mixed-American | Full Cohort | Switching vs Continuation | BMI | Income25-35k | 0.824 | 0.662 | 1.025 | 8.15e-02 | 1.00e+00 | 3832 | 2390 | 1442 |
| Latino/Mixed-American | Full Cohort | Switching vs Continuation | BMI | Income35-50k | 1.046 | 0.834 | 1.312 | 6.98e-01 | 1.00e+00 | 3832 | 2390 | 1442 |
| Latino/Mixed-American | Full Cohort | Switching vs Continuation | BMI | Income50-75k | 0.777 | 0.618 | 0.977 | 3.06e-02 | 4.29e-01 | 3832 | 2390 | 1442 |
| Latino/Mixed-American | Full Cohort | Switching vs Continuation | BMI | Income75-100k | 0.869 | 0.651 | 1.161 | 3.43e-01 | 1.00e+00 | 3832 | 2390 | 1442 |
| Latino/Mixed-American | Full Cohort | Switching vs Continuation | BMI | Education_levelCollege Postgraduate | 1.037 | 0.776 | 1.385 | 8.06e-01 | 1.00e+00 | 3832 | 2390 | 1442 |
| Latino/Mixed-American | Full Cohort | Switching vs Continuation | BMI | Education_levelCollege Undergraduate | 1.117 | 0.917 | 1.360 | 2.73e-01 | 1.00e+00 | 3832 | 2390 | 1442 |
| Latino/Mixed-American | Full Cohort | Switching vs Continuation | BMI | Education_levelTwelve or GED | 0.995 | 0.803 | 1.232 | 9.61e-01 | 1.00e+00 | 3832 | 2390 | 1442 |

| | | | | | | | | | | | | |
|---|---|---|---|---|---|---|---|---|---|---|---|---|
| Latino/Mixed-American | Full Cohort | Switching vs Continuation | BMI | BMI | 1.037 | 0.969 | 1.110 | 2.99e-01 | 1.00e+00 | 3832 | 2390 | 1442 |
| Latino/Mixed-American | Full Cohort | Discontinuation vs Continuation | Education_level | (Intercept) | 0.497 | 0.423 | 0.584 | <2.2e-16 | 2.78e-16 | 3862 | 2456 | 1406 |
| Latino/Mixed-American | Full Cohort | Discontinuation vs Continuation | Education_level | age_at_first_AD | 0.994 | 0.926 | 1.067 | 8.75e-01 | 1.00e+00 | 3862 | 2456 | 1406 |
| Latino/Mixed-American | Full Cohort | Discontinuation vs Continuation | Education_level | sex_at_birthMale | 1.225 | 1.059 | 1.418 | 6.39e-03 | 8.94e-02 | 3862 | 2456 | 1406 |
| Latino/Mixed-American | Full Cohort | Discontinuation vs Continuation | Education_level | Income>200k | 0.620 | 0.392 | 0.982 | 4.18e-02 | 5.85e-01 | 3862 | 2456 | 1406 |
| Latino/Mixed-American | Full Cohort | Discontinuation vs Continuation | Education_level | Income100-150k | 0.681 | 0.511 | 0.908 | 8.86e-03 | 1.24e-01 | 3862 | 2456 | 1406 |
| Latino/Mixed-American | Full Cohort | Discontinuation vs Continuation | Education_level | Income150-200k | 0.700 | 0.461 | 1.063 | 9.46e-02 | 1.00e+00 | 3862 | 2456 | 1406 |
| Latino/Mixed-American | Full Cohort | Discontinuation vs Continuation | Education_level | Income25-35k | 0.820 | 0.659 | 1.019 | 7.39e-02 | 1.00e+00 | 3862 | 2456 | 1406 |
| Latino/Mixed-American | Full Cohort | Discontinuation vs Continuation | Education_level | Income35-50k | 1.069 | 0.854 | 1.338 | 5.59e-01 | 1.00e+00 | 3862 | 2456 | 1406 |
| Latino/Mixed-American | Full Cohort | Discontinuation vs Continuation | Education_level | Income50-75k | 0.832 | 0.664 | 1.042 | 1.09e-01 | 1.00e+00 | 3862 | 2456 | 1406 |
| Latino/Mixed-American | Full Cohort | Discontinuation vs Continuation | Education_level | Income75-100k | 0.759 | 0.561 | 1.028 | 7.49e-02 | 1.00e+00 | 3862 | 2456 | 1406 |
| Latino/Mixed-American | Full Cohort | Discontinuation vs Continuation | Education_level | Education_levelCollege Postgraduate | 0.909 | 0.669 | 1.237 | 5.45e-01 | 1.00e+00 | 3862 | 2456 | 1406 |
| Latino/Mixed-American | Full Cohort | Discontinuation vs Continuation | Education_level | Education_levelCollege Undergraduate | 1.336 | 1.095 | 1.629 | 4.27e-03 | 5.98e-02 | 3862 | 2456 | 1406 |
| Latino/Mixed-American | Full Cohort | Discontinuation vs Continuation | Education_level | Education_levelTwelve or GED | 1.259 | 1.018 | 1.557 | 3.36e-02 | 4.71e-01 | 3862 | 2456 | 1406 |
| Latino/Mixed-American | Full Cohort | Switching vs Continuation | Education_level | (Intercept) | 0.591 | 0.507 | 0.689 | 1.89e-11 | 2.65e-10 | 3931 | 2456 | 1475 |
| Latino/Mixed-American | Full Cohort | Switching vs Continuation | Education_level | age_at_first_AD | 0.806 | 0.749 | 0.867 | 7.66e-09 | 1.07e-07 | 3931 | 2456 | 1475 |
| Latino/Mixed-American | Full Cohort | Switching vs Continuation | Education_level | sex_at_birthMale | 1.056 | 0.910 | 1.225 | 4.76e-01 | 1.00e+00 | 3931 | 2456 | 1475 |
| Latino/Mixed-American | Full Cohort | Switching vs Continuation | Education_level | Income>200k | 0.515 | 0.324 | 0.817 | 4.80e-03 | 6.72e-02 | 3931 | 2456 | 1475 |
| Latino/Mixed-American | Full Cohort | Switching vs Continuation | Education_level | Income100-150k | 0.769 | 0.584 | 1.011 | 6.01e-02 | 8.42e-01 | 3931 | 2456 | 1475 |
| Latino/Mixed-American | Full Cohort | Switching vs Continuation | Education_level | Income150-200k | 0.676 | 0.448 | 1.020 | 6.18e-02 | 8.65e-01 | 3931 | 2456 | 1475 |
| Latino/Mixed-American | Full Cohort | Switching vs Continuation | Education_level | Income25-35k | 0.821 | 0.662 | 1.018 | 7.25e-02 | 1.00e+00 | 3931 | 2456 | 1475 |
| Latino/Mixed-American | Full Cohort | Switching vs Continuation | Education_level | Income35-50k | 1.043 | 0.835 | 1.303 | 7.13e-01 | 1.00e+00 | 3931 | 2456 | 1475 |
| Latino/Mixed-American | Full Cohort | Switching vs Continuation | Education_level | Income50-75k | 0.770 | 0.614 | 0.965 | 2.31e-02 | 3.23e-01 | 3931 | 2456 | 1475 |
| Latino/Mixed-American | Full Cohort | Switching vs Continuation | Education_level | Income75-100k | 0.885 | 0.664 | 1.178 | 4.02e-01 | 1.00e+00 | 3931 | 2456 | 1475 |
| Latino/Mixed-American | Full Cohort | Switching vs Continuation | Education_level | Education_levelCollege Postgraduate | 1.017 | 0.765 | 1.352 | 9.09e-01 | 1.00e+00 | 3931 | 2456 | 1475 |
| Latino/Mixed-American | Full Cohort | Switching vs Continuation | Education_level | Education_levelCollege Undergraduate | 1.085 | 0.893 | 1.318 | 4.13e-01 | 1.00e+00 | 3931 | 2456 | 1475 |
| Latino/Mixed-American | Full Cohort | Switching vs Continuation | Education_level | Education_levelTwelve or GED | 0.957 | 0.774 | 1.183 | 6.84e-01 | 1.00e+00 | 3931 | 2456 | 1475 |
| Latino/Mixed-American | Full Cohort | Discontinuation vs Continuation | Income | (Intercept) | 0.497 | 0.423 | 0.584 | <2.2e-16 | 2.78e-16 | 3862 | 2456 | 1406 |
| Latino/Mixed-American | Full Cohort | Discontinuation vs Continuation | Income | age_at_first_AD | 0.994 | 0.926 | 1.067 | 8.75e-01 | 1.00e+00 | 3862 | 2456 | 1406 |
| Latino/Mixed-American | Full Cohort | Discontinuation vs Continuation | Income | sex_at_birthMale | 1.225 | 1.059 | 1.418 | 6.39e-03 | 8.94e-02 | 3862 | 2456 | 1406 |
| Latino/Mixed-American | Full Cohort | Discontinuation vs Continuation | Income | Income>200k | 0.620 | 0.392 | 0.982 | 4.18e-02 | 5.85e-01 | 3862 | 2456 | 1406 |
| Latino/Mixed-American | Full Cohort | Discontinuation vs Continuation | Income | Income100-150k | 0.681 | 0.511 | 0.908 | 8.86e-03 | 1.24e-01 | 3862 | 2456 | 1406 |
| Latino/Mixed-American | Full Cohort | Discontinuation vs Continuation | Income | Income150-200k | 0.700 | 0.461 | 1.063 | 9.46e-02 | 1.00e+00 | 3862 | 2456 | 1406 |
| Latino/Mixed-American | Full Cohort | Discontinuation vs Continuation | Income | Income25-35k | 0.820 | 0.659 | 1.019 | 7.39e-02 | 1.00e+00 | 3862 | 2456 | 1406 |
| Latino/Mixed-American | Full Cohort | Discontinuation vs Continuation | Income | Income35-50k | 1.069 | 0.854 | 1.338 | 5.59e-01 | 1.00e+00 | 3862 | 2456 | 1406 |
| Latino/Mixed-American | Full Cohort | Discontinuation vs Continuation | Income | Income50-75k | 0.832 | 0.664 | 1.042 | 1.09e-01 | 1.00e+00 | 3862 | 2456 | 1406 |
| Latino/Mixed-American | Full Cohort | Discontinuation vs Continuation | Income | Income75-100k | 0.759 | 0.561 | 1.028 | 7.49e-02 | 1.00e+00 | 3862 | 2456 | 1406 |
| Latino/Mixed-American | Full Cohort | Discontinuation vs Continuation | Income | Education_levelCollege Postgraduate | 0.909 | 0.669 | 1.237 | 5.45e-01 | 1.00e+00 | 3862 | 2456 | 1406 |
| Latino/Mixed-American | Full Cohort | Discontinuation vs Continuation | Income | Education_levelCollege Undergraduate | 1.336 | 1.095 | 1.629 | 4.27e-03 | 5.98e-02 | 3862 | 2456 | 1406 |
| Latino/Mixed-American | Full Cohort | Discontinuation vs Continuation | Income | Education_levelTwelve or GED | 1.259 | 1.018 | 1.557 | 3.36e-02 | 4.71e-01 | 3862 | 2456 | 1406 |
| Latino/Mixed-American | Full Cohort | Switching vs Continuation | Income | (Intercept) | 0.591 | 0.507 | 0.689 | 1.89e-11 | 2.65e-10 | 3931 | 2456 | 1475 |
| Latino/Mixed-American | Full Cohort | Switching vs Continuation | Income | age_at_first_AD | 0.806 | 0.749 | 0.867 | 7.66e-09 | 1.07e-07 | 3931 | 2456 | 1475 |
| Latino/Mixed-American | Full Cohort | Switching vs Continuation | Income | sex_at_birthMale | 1.056 | 0.910 | 1.225 | 4.76e-01 | 1.00e+00 | 3931 | 2456 | 1475 |
| Latino/Mixed-American | Full Cohort | Switching vs Continuation | Income | Income>200k | 0.515 | 0.324 | 0.817 | 4.80e-03 | 6.72e-02 | 3931 | 2456 | 1475 |
| Latino/Mixed-American | Full Cohort | Switching vs Continuation | Income | Income100-150k | 0.769 | 0.584 | 1.011 | 6.01e-02 | 8.42e-01 | 3931 | 2456 | 1475 |
| Latino/Mixed-American | Full Cohort | Switching vs Continuation | Income | Income150-200k | 0.676 | 0.448 | 1.020 | 6.18e-02 | 8.65e-01 | 3931 | 2456 | 1475 |
| Latino/Mixed-American | Full Cohort | Switching vs Continuation | Income | Income25-35k | 0.821 | 0.662 | 1.018 | 7.25e-02 | 1.00e+00 | 3931 | 2456 | 1475 |
| Latino/Mixed-American | Full Cohort | Switching vs Continuation | Income | Income35-50k | 1.043 | 0.835 | 1.303 | 7.13e-01 | 1.00e+00 | 3931 | 2456 | 1475 |
| Latino/Mixed-American | Full Cohort | Switching vs Continuation | Income | Income50-75k | 0.770 | 0.614 | 0.965 | 2.31e-02 | 3.23e-01 | 3931 | 2456 | 1475 |
| Latino/Mixed-American | Full Cohort | Switching vs Continuation | Income | Income75-100k | 0.885 | 0.664 | 1.178 | 4.02e-01 | 1.00e+00 | 3931 | 2456 | 1475 |
| Latino/Mixed-American | Full Cohort | Switching vs Continuation | Income | Education_levelCollege Postgraduate | 1.017 | 0.765 | 1.352 | 9.09e-01 | 1.00e+00 | 3931 | 2456 | 1475 |
| Latino/Mixed-American | Full Cohort | Switching vs Continuation | Income | Education_levelCollege Undergraduate | 1.085 | 0.893 | 1.318 | 4.13e-01 | 1.00e+00 | 3931 | 2456 | 1475 |
| Latino/Mixed-American | Full Cohort | Switching vs Continuation | Income | Education_levelTwelve or GED | 0.957 | 0.774 | 1.183 | 6.84e-01 | 1.00e+00 | 3931 | 2456 | 1475 |
| Latino/Mixed-American | Full Cohort | Discontinuation vs Continuation | Insomnia | (Intercept) | 0.531 | 0.450 | 0.626 | 5.76e-14 | 8.07e-13 | 3862 | 2456 | 1406 |
| Latino/Mixed-American | Full Cohort | Discontinuation vs Continuation | Insomnia | age_at_first_AD | 1.008 | 0.939 | 1.083 | 8.24e-01 | 1.00e+00 | 3862 | 2456 | 1406 |
| Latino/Mixed-American | Full Cohort | Discontinuation vs Continuation | Insomnia | sex_at_birthMale | 1.240 | 1.071 | 1.435 | 4.05e-03 | 5.66e-02 | 3862 | 2456 | 1406 |
| Latino/Mixed-American | Full Cohort | Discontinuation vs Continuation | Insomnia | Income>200k | 0.611 | 0.385 | 0.968 | 3.60e-02 | 5.04e-01 | 3862 | 2456 | 1406 |
| Latino/Mixed-American | Full Cohort | Discontinuation vs Continuation | Insomnia | Income100-150k | 0.680 | 0.509 | 0.907 | 8.65e-03 | 1.21e-01 | 3862 | 2456 | 1406 |
| Latino/Mixed-American | Full Cohort | Discontinuation vs Continuation | Insomnia | Income150-200k | 0.692 | 0.455 | 1.052 | 8.49e-02 | 1.00e+00 | 3862 | 2456 | 1406 |
| Latino/Mixed-American | Full Cohort | Discontinuation vs Continuation | Insomnia | Income25-35k | 0.818 | 0.657 | 1.018 | 7.12e-02 | 9.97e-01 | 3862 | 2456 | 1406 |
| Latino/Mixed-American | Full Cohort | Discontinuation vs Continuation | Insomnia | Income35-50k | 1.055 | 0.843 | 1.321 | 6.40e-01 | 1.00e+00 | 3862 | 2456 | 1406 |
| Latino/Mixed-American | Full Cohort | Discontinuation vs Continuation | Insomnia | Income50-75k | 0.829 | 0.662 | 1.039 | 1.04e-01 | 1.00e+00 | 3862 | 2456 | 1406 |
| Latino/Mixed-American | Full Cohort | Discontinuation vs Continuation | Insomnia | Income75-100k | 0.756 | 0.558 | 1.024 | 7.08e-02 | 9.91e-01 | 3862 | 2456 | 1406 |
| Latino/Mixed-American | Full Cohort | Discontinuation vs Continuation | Insomnia | Education_levelCollege Postgraduate | 0.901 | 0.663 | 1.227 | 5.09e-01 | 1.00e+00 | 3862 | 2456 | 1406 |
| Latino/Mixed-American | Full Cohort | Discontinuation vs Continuation | Insomnia | Education_levelCollege Undergraduate | 1.342 | 1.100 | 1.637 | 3.75e-03 | 5.25e-02 | 3862 | 2456 | 1406 |
| Latino/Mixed-American | Full Cohort | Discontinuation vs Continuation | Insomnia | Education_levelTwelve or GED | 1.265 | 1.022 | 1.566 | 3.05e-02 | 4.28e-01 | 3862 | 2456 | 1406 |

| | | | | | | | | | | | | |
|---|---|---|---|---|---|---|---|---|---|---|---|---|
| Latino/Mixed-American | Full Cohort | Discontinuation vs Continuation | Insomnia | Insomnia | 0.736 | 0.624 | 0.868 | 2.64e-04 | 3.69e-03 | 3862 | 2456 | 1406 |
| Latino/Mixed-American | Full Cohort | Switching vs Continuation | Insomnia | (Intercept) | 0.431 | 0.365 | 0.508 | <2.2e-16 | <2.2e-16 | 3931 | 2456 | 1475 |
| Latino/Mixed-American | Full Cohort | Switching vs Continuation | Insomnia | age_at_first_AD | 0.759 | 0.704 | 0.819 | 1.19e-12 | 1.67e-11 | 3931 | 2456 | 1475 |
| Latino/Mixed-American | Full Cohort | Switching vs Continuation | Insomnia | sex_at_birthMale | 1.018 | 0.874 | 1.186 | 8.18e-01 | 1.00e+00 | 3931 | 2456 | 1475 |
| Latino/Mixed-American | Full Cohort | Switching vs Continuation | Insomnia | Income>200k | 0.537 | 0.336 | 0.860 | 9.70e-03 | 1.36e-01 | 3931 | 2456 | 1475 |
| Latino/Mixed-American | Full Cohort | Switching vs Continuation | Insomnia | Income100-150k | 0.752 | 0.567 | 0.996 | 4.67e-02 | 6.53e-01 | 3931 | 2456 | 1475 |
| Latino/Mixed-American | Full Cohort | Switching vs Continuation | Insomnia | Income150-200k | 0.717 | 0.472 | 1.091 | 1.20e-01 | 1.00e+00 | 3931 | 2456 | 1475 |
| Latino/Mixed-American | Full Cohort | Switching vs Continuation | Insomnia | Income25-35k | 0.827 | 0.663 | 1.031 | 9.07e-02 | 1.00e+00 | 3931 | 2456 | 1475 |
| Latino/Mixed-American | Full Cohort | Switching vs Continuation | Insomnia | Income35-50k | 1.030 | 0.820 | 1.294 | 8.00e-01 | 1.00e+00 | 3931 | 2456 | 1475 |
| Latino/Mixed-American | Full Cohort | Switching vs Continuation | Insomnia | Income50-75k | 0.773 | 0.613 | 0.975 | 2.94e-02 | 4.12e-01 | 3931 | 2456 | 1475 |
| Latino/Mixed-American | Full Cohort | Switching vs Continuation | Insomnia | Income75-100k | 0.883 | 0.658 | 1.184 | 4.04e-01 | 1.00e+00 | 3931 | 2456 | 1475 |
| Latino/Mixed-American | Full Cohort | Switching vs Continuation | Insomnia | Education_levelCollege Postgraduate | 1.041 | 0.778 | 1.394 | 7.87e-01 | 1.00e+00 | 3931 | 2456 | 1475 |
| Latino/Mixed-American | Full Cohort | Switching vs Continuation | Insomnia | Education_levelCollege Undergraduate | 1.063 | 0.870 | 1.297 | 5.52e-01 | 1.00e+00 | 3931 | 2456 | 1475 |
| Latino/Mixed-American | Full Cohort | Switching vs Continuation | Insomnia | Education_levelTwelve or GED | 0.925 | 0.745 | 1.150 | 4.85e-01 | 1.00e+00 | 3931 | 2456 | 1475 |
| Latino/Mixed-American | Full Cohort | Switching vs Continuation | Insomnia | Insomnia | 2.625 | 2.278 | 3.026 | <2.2e-16 | <2.2e-16 | 3931 | 2456 | 1475 |
| Latino/Mixed-American | Full Cohort | Discontinuation vs Continuation | MDD CIDI-sf | (Intercept) | 0.628 | 0.127 | 3.118 | 5.70e-01 | 1.00e+00 | 294 | 209 | 85 |
| Latino/Mixed-American | Full Cohort | Discontinuation vs Continuation | MDD CIDI-sf | age_at_first_AD | 0.698 | 0.503 | 0.967 | 3.08e-02 | 4.31e-01 | 294 | 209 | 85 |
| Latino/Mixed-American | Full Cohort | Discontinuation vs Continuation | MDD CIDI-sf | sex_at_birthMale | 1.488 | 0.809 | 2.734 | 2.01e-01 | 1.00e+00 | 294 | 209 | 85 |
| Latino/Mixed-American | Full Cohort | Discontinuation vs Continuation | MDD CIDI-sf | Income>200k | 1.251 | 0.372 | 4.213 | 7.18e-01 | 1.00e+00 | 294 | 209 | 85 |
| Latino/Mixed-American | Full Cohort | Discontinuation vs Continuation | MDD CIDI-sf | Income100-150k | 1.351 | 0.547 | 3.338 | 5.14e-01 | 1.00e+00 | 294 | 209 | 85 |
| Latino/Mixed-American | Full Cohort | Discontinuation vs Continuation | MDD CIDI-sf | Income150-200k | 0.828 | 0.219 | 3.127 | 7.80e-01 | 1.00e+00 | 294 | 209 | 85 |
| Latino/Mixed-American | Full Cohort | Discontinuation vs Continuation | MDD CIDI-sf | Income25-35k | 1.501 | 0.603 | 3.740 | 3.83e-01 | 1.00e+00 | 294 | 209 | 85 |
| Latino/Mixed-American | Full Cohort | Discontinuation vs Continuation | MDD CIDI-sf | Income35-50k | 1.480 | 0.580 | 3.779 | 4.12e-01 | 1.00e+00 | 294 | 209 | 85 |
| Latino/Mixed-American | Full Cohort | Discontinuation vs Continuation | MDD CIDI-sf | Income50-75k | 0.573 | 0.225 | 1.461 | 2.44e-01 | 1.00e+00 | 294 | 209 | 85 |
| Latino/Mixed-American | Full Cohort | Discontinuation vs Continuation | MDD CIDI-sf | Income75-100k | 0.561 | 0.189 | 1.662 | 2.96e-01 | 1.00e+00 | 294 | 209 | 85 |
| Latino/Mixed-American | Full Cohort | Discontinuation vs Continuation | MDD CIDI-sf | Education_levelCollege Postgraduate | 0.331 | 0.062 | 1.763 | 1.95e-01 | 1.00e+00 | 294 | 209 | 85 |
| Latino/Mixed-American | Full Cohort | Discontinuation vs Continuation | MDD CIDI-sf | Education_levelCollege Undergraduate | 0.438 | 0.090 | 2.142 | 3.08e-01 | 1.00e+00 | 294 | 209 | 85 |
| Latino/Mixed-American | Full Cohort | Discontinuation vs Continuation | MDD CIDI-sf | Education_levelTwelve or GED | 0.599 | 0.112 | 3.201 | 5.48e-01 | 1.00e+00 | 294 | 209 | 85 |
| Latino/Mixed-American | Full Cohort | Discontinuation vs Continuation | MDD CIDI-sf | MDD CIDI-sf | 1.292 | 0.707 | 2.361 | 4.05e-01 | 1.00e+00 | 294 | 209 | 85 |
| Latino/Mixed-American | Full Cohort | Switching vs Continuation | MDD CIDI-sf | (Intercept) | 0.636 | 0.125 | 3.226 | 5.85e-01 | 1.00e+00 | 354 | 209 | 145 |
| Latino/Mixed-American | Full Cohort | Switching vs Continuation | MDD CIDI-sf | age_at_first_AD | 0.685 | 0.516 | 0.910 | 9.03e-03 | 1.26e-01 | 354 | 209 | 145 |
| Latino/Mixed-American | Full Cohort | Switching vs Continuation | MDD CIDI-sf | sex_at_birthMale | 1.387 | 0.807 | 2.383 | 2.36e-01 | 1.00e+00 | 354 | 209 | 145 |
| Latino/Mixed-American | Full Cohort | Switching vs Continuation | MDD CIDI-sf | Income>200k | 0.501 | 0.166 | 1.517 | 2.21e-01 | 1.00e+00 | 354 | 209 | 145 |
| Latino/Mixed-American | Full Cohort | Switching vs Continuation | MDD CIDI-sf | Income100-150k | 0.584 | 0.258 | 1.319 | 1.96e-01 | 1.00e+00 | 354 | 209 | 145 |
| Latino/Mixed-American | Full Cohort | Switching vs Continuation | MDD CIDI-sf | Income150-200k | 0.704 | 0.250 | 1.985 | 5.07e-01 | 1.00e+00 | 354 | 209 | 145 |
| Latino/Mixed-American | Full Cohort | Switching vs Continuation | MDD CIDI-sf | Income25-35k | 0.660 | 0.289 | 1.506 | 3.24e-01 | 1.00e+00 | 354 | 209 | 145 |
| Latino/Mixed-American | Full Cohort | Switching vs Continuation | MDD CIDI-sf | Income35-50k | 0.583 | 0.242 | 1.404 | 2.29e-01 | 1.00e+00 | 354 | 209 | 145 |
| Latino/Mixed-American | Full Cohort | Switching vs Continuation | MDD CIDI-sf | Income50-75k | 0.643 | 0.323 | 1.282 | 2.10e-01 | 1.00e+00 | 354 | 209 | 145 |
| Latino/Mixed-American | Full Cohort | Switching vs Continuation | MDD CIDI-sf | Income75-100k | 0.615 | 0.277 | 1.365 | 2.32e-01 | 1.00e+00 | 354 | 209 | 145 |
| Latino/Mixed-American | Full Cohort | Switching vs Continuation | MDD CIDI-sf | Education_levelCollege Postgraduate | 0.477 | 0.086 | 2.649 | 3.97e-01 | 1.00e+00 | 354 | 209 | 145 |
| Latino/Mixed-American | Full Cohort | Switching vs Continuation | MDD CIDI-sf | Education_levelCollege Undergraduate | 0.796 | 0.154 | 4.117 | 7.85e-01 | 1.00e+00 | 354 | 209 | 145 |
| Latino/Mixed-American | Full Cohort | Switching vs Continuation | MDD CIDI-sf | Education_levelTwelve or GED | 0.851 | 0.152 | 4.758 | 8.54e-01 | 1.00e+00 | 354 | 209 | 145 |
| Latino/Mixed-American | Full Cohort | Switching vs Continuation | MDD CIDI-sf | MDD CIDI-sf | 2.225 | 1.303 | 3.800 | 3.41e-03 | 4.77e-02 | 354 | 209 | 145 |
| Latino/Mixed-American | Full Cohort | Discontinuation vs Continuation | MDD_symptom_count | (Intercept) | 0.438 | 0.105 | 1.821 | 2.56e-01 | 1.00e+00 | 358 | 251 | 107 |
| Latino/Mixed-American | Full Cohort | Discontinuation vs Continuation | MDD_symptom_count | age_at_first_AD | 0.735 | 0.553 | 0.977 | 3.40e-02 | 4.76e-01 | 358 | 251 | 107 |
| Latino/Mixed-American | Full Cohort | Discontinuation vs Continuation | MDD_symptom_count | sex_at_birthMale | 1.712 | 0.999 | 2.933 | 5.04e-02 | 7.05e-01 | 358 | 251 | 107 |
| Latino/Mixed-American | Full Cohort | Discontinuation vs Continuation | MDD_symptom_count | Income>200k | 1.207 | 0.402 | 3.624 | 7.37e-01 | 1.00e+00 | 358 | 251 | 107 |
| Latino/Mixed-American | Full Cohort | Discontinuation vs Continuation | MDD_symptom_count | Income100-150k | 1.721 | 0.772 | 3.839 | 1.84e-01 | 1.00e+00 | 358 | 251 | 107 |
| Latino/Mixed-American | Full Cohort | Discontinuation vs Continuation | MDD_symptom_count | Income150-200k | 0.934 | 0.278 | 3.135 | 9.12e-01 | 1.00e+00 | 358 | 251 | 107 |
| Latino/Mixed-American | Full Cohort | Discontinuation vs Continuation | MDD_symptom_count | Income25-35k | 1.475 | 0.622 | 3.500 | 3.78e-01 | 1.00e+00 | 358 | 251 | 107 |
| Latino/Mixed-American | Full Cohort | Discontinuation vs Continuation | MDD_symptom_count | Income35-50k | 1.341 | 0.572 | 3.145 | 4.99e-01 | 1.00e+00 | 358 | 251 | 107 |
| Latino/Mixed-American | Full Cohort | Discontinuation vs Continuation | MDD_symptom_count | Income50-75k | 0.608 | 0.263 | 1.407 | 2.45e-01 | 1.00e+00 | 358 | 251 | 107 |
| Latino/Mixed-American | Full Cohort | Discontinuation vs Continuation | MDD_symptom_count | Income75-100k | 0.859 | 0.342 | 2.156 | 7.46e-01 | 1.00e+00 | 358 | 251 | 107 |
| Latino/Mixed-American | Full Cohort | Discontinuation vs Continuation | MDD_symptom_count | Education_levelCollege Postgraduate | 0.489 | 0.105 | 2.283 | 3.63e-01 | 1.00e+00 | 358 | 251 | 107 |
| Latino/Mixed-American | Full Cohort | Discontinuation vs Continuation | MDD_symptom_count | Education_levelCollege Undergraduate | 0.727 | 0.169 | 3.132 | 6.69e-01 | 1.00e+00 | 358 | 251 | 107 |
| Latino/Mixed-American | Full Cohort | Discontinuation vs Continuation | MDD_symptom_count | Education_levelTwelve or GED | 1.029 | 0.222 | 4.757 | 9.71e-01 | 1.00e+00 | 358 | 251 | 107 |
| Latino/Mixed-American | Full Cohort | Discontinuation vs Continuation | MDD_symptom_count | MDD_symptom_count | 1.084 | 0.845 | 1.390 | 5.24e-01 | 1.00e+00 | 358 | 251 | 107 |
| Latino/Mixed-American | Full Cohort | Switching vs Continuation | MDD_symptom_count | (Intercept) | 1.092 | 0.329 | 3.619 | 8.86e-01 | 1.00e+00 | 418 | 251 | 167 |
| Latino/Mixed-American | Full Cohort | Switching vs Continuation | MDD_symptom_count | age_at_first_AD | 0.657 | 0.508 | 0.849 | 1.34e-03 | 1.88e-02 | 418 | 251 | 167 |
| Latino/Mixed-American | Full Cohort | Switching vs Continuation | MDD_symptom_count | sex_at_birthMale | 1.488 | 0.913 | 2.426 | 1.11e-01 | 1.00e+00 | 418 | 251 | 167 |
| Latino/Mixed-American | Full Cohort | Switching vs Continuation | MDD_symptom_count | Income>200k | 0.456 | 0.166 | 1.253 | 1.28e-01 | 1.00e+00 | 418 | 251 | 167 |
| Latino/Mixed-American | Full Cohort | Switching vs Continuation | MDD_symptom_count | Income100-150k | 0.628 | 0.300 | 1.317 | 2.19e-01 | 1.00e+00 | 418 | 251 | 167 |
| Latino/Mixed-American | Full Cohort | Switching vs Continuation | MDD_symptom_count | Income150-200k | 0.676 | 0.255 | 1.795 | 4.32e-01 | 1.00e+00 | 418 | 251 | 167 |
| Latino/Mixed-American | Full Cohort | Switching vs Continuation | MDD_symptom_count | Income25-35k | 0.646 | 0.296 | 1.408 | 2.72e-01 | 1.00e+00 | 418 | 251 | 167 |
| Latino/Mixed-American | Full Cohort | Switching vs Continuation | MDD_symptom_count | Income35-50k | 0.533 | 0.243 | 1.173 | 1.18e-01 | 1.00e+00 | 418 | 251 | 167 |
| Latino/Mixed-American | Full Cohort | Switching vs Continuation | MDD_symptom_count | Income50-75k | 0.566 | 0.300 | 1.071 | 8.02e-02 | 1.00e+00 | 418 | 251 | 167 |

| | | | | | | | | | | | | |
|---|---|---|---|---|---|---|---|---|---|---|---|---|
| Latino/Mixed-American | Full Cohort | Switching vs Continuation | MDD_symptom_count | Income75-100k | 0.611 | 0.290 | 1.286 | 1.94e-01 | 1.00e+00 | 418 | 251 | 167 |
| Latino/Mixed-American | Full Cohort | Switching vs Continuation | MDD_symptom_count | Education_levelCollege Postgraduate | 0.453 | 0.119 | 1.728 | 2.46e-01 | 1.00e+00 | 418 | 251 | 167 |
| Latino/Mixed-American | Full Cohort | Switching vs Continuation | MDD_symptom_count | Education_levelCollege Undergraduate | 0.771 | 0.219 | 2.719 | 6.86e-01 | 1.00e+00 | 418 | 251 | 167 |
| Latino/Mixed-American | Full Cohort | Switching vs Continuation | MDD_symptom_count | Education_levelTwelve or GED | 0.743 | 0.194 | 2.846 | 6.65e-01 | 1.00e+00 | 418 | 251 | 167 |
| Latino/Mixed-American | Full Cohort | Switching vs Continuation | MDD_symptom_count | MDD_symptom_count | 1.264 | 1.012 | 1.577 | 3.85e-02 | 5.39e-01 | 418 | 251 | 167 |
| Latino/Mixed-American | Full Cohort | Discontinuation vs Continuation | Migraine | (Intercept) | 0.549 | 0.464 | 0.648 | 1.91e-12 | 2.67e-11 | 3862 | 2456 | 1406 |
| Latino/Mixed-American | Full Cohort | Discontinuation vs Continuation | Migraine | age_at_first_AD | 0.982 | 0.914 | 1.054 | 6.14e-01 | 1.00e+00 | 3862 | 2456 | 1406 |
| Latino/Mixed-American | Full Cohort | Discontinuation vs Continuation | Migraine | sex_at_birthMale | 1.178 | 1.017 | 1.365 | 2.92e-02 | 4.09e-01 | 3862 | 2456 | 1406 |
| Latino/Mixed-American | Full Cohort | Discontinuation vs Continuation | Migraine | Income>200k | 0.625 | 0.394 | 0.991 | 4.55e-02 | 6.37e-01 | 3862 | 2456 | 1406 |
| Latino/Mixed-American | Full Cohort | Discontinuation vs Continuation | Migraine | Income100-150k | 0.691 | 0.518 | 0.922 | 1.21e-02 | 1.69e-01 | 3862 | 2456 | 1406 |
| Latino/Mixed-American | Full Cohort | Discontinuation vs Continuation | Migraine | Income150-200k | 0.698 | 0.459 | 1.062 | 9.31e-02 | 1.00e+00 | 3862 | 2456 | 1406 |
| Latino/Mixed-American | Full Cohort | Discontinuation vs Continuation | Migraine | Income25-35k | 0.825 | 0.663 | 1.026 | 8.44e-02 | 1.00e+00 | 3862 | 2456 | 1406 |
| Latino/Mixed-American | Full Cohort | Discontinuation vs Continuation | Migraine | Income35-50k | 1.074 | 0.858 | 1.345 | 5.32e-01 | 1.00e+00 | 3862 | 2456 | 1406 |
| Latino/Mixed-American | Full Cohort | Discontinuation vs Continuation | Migraine | Income50-75k | 0.834 | 0.665 | 1.045 | 1.14e-01 | 1.00e+00 | 3862 | 2456 | 1406 |
| Latino/Mixed-American | Full Cohort | Discontinuation vs Continuation | Migraine | Income75-100k | 0.768 | 0.567 | 1.041 | 8.90e-02 | 1.00e+00 | 3862 | 2456 | 1406 |
| Latino/Mixed-American | Full Cohort | Discontinuation vs Continuation | Migraine | Education_levelCollege Postgraduate | 0.895 | 0.657 | 1.217 | 4.79e-01 | 1.00e+00 | 3862 | 2456 | 1406 |
| Latino/Mixed-American | Full Cohort | Discontinuation vs Continuation | Migraine | Education_levelCollege Undergraduate | 1.332 | 1.092 | 1.626 | 4.70e-03 | 6.58e-02 | 3862 | 2456 | 1406 |
| Latino/Mixed-American | Full Cohort | Discontinuation vs Continuation | Migraine | Education_levelTwelve or GED | 1.261 | 1.019 | 1.560 | 3.31e-02 | 4.63e-01 | 3862 | 2456 | 1406 |
| Latino/Mixed-American | Full Cohort | Discontinuation vs Continuation | Migraine | Migraine | 0.715 | 0.614 | 0.833 | 1.58e-05 | 2.22e-04 | 3862 | 2456 | 1406 |
| Latino/Mixed-American | Full Cohort | Switching vs Continuation | Migraine | (Intercept) | 0.444 | 0.376 | 0.525 | <2.2e-16 | <2.2e-16 | 3931 | 2456 | 1475 |
| Latino/Mixed-American | Full Cohort | Switching vs Continuation | Migraine | age_at_first_AD | 0.834 | 0.774 | 0.898 | 1.72e-06 | 2.41e-05 | 3931 | 2456 | 1475 |
| Latino/Mixed-American | Full Cohort | Switching vs Continuation | Migraine | sex_at_birthMale | 1.175 | 1.008 | 1.368 | 3.86e-02 | 5.41e-01 | 3931 | 2456 | 1475 |
| Latino/Mixed-American | Full Cohort | Switching vs Continuation | Migraine | Income>200k | 0.518 | 0.325 | 0.825 | 5.66e-03 | 7.93e-02 | 3931 | 2456 | 1475 |
| Latino/Mixed-American | Full Cohort | Switching vs Continuation | Migraine | Income100-150k | 0.756 | 0.573 | 0.997 | 4.78e-02 | 6.70e-01 | 3931 | 2456 | 1475 |
| Latino/Mixed-American | Full Cohort | Switching vs Continuation | Migraine | Income150-200k | 0.703 | 0.464 | 1.065 | 9.66e-02 | 1.00e+00 | 3931 | 2456 | 1475 |
| Latino/Mixed-American | Full Cohort | Switching vs Continuation | Migraine | Income25-35k | 0.820 | 0.659 | 1.020 | 7.44e-02 | 1.00e+00 | 3931 | 2456 | 1475 |
| Latino/Mixed-American | Full Cohort | Switching vs Continuation | Migraine | Income35-50k | 1.039 | 0.829 | 1.302 | 7.41e-01 | 1.00e+00 | 3931 | 2456 | 1475 |
| Latino/Mixed-American | Full Cohort | Switching vs Continuation | Migraine | Income50-75k | 0.777 | 0.618 | 0.977 | 3.09e-02 | 4.33e-01 | 3931 | 2456 | 1475 |
| Latino/Mixed-American | Full Cohort | Switching vs Continuation | Migraine | Income75-100k | 0.870 | 0.651 | 1.163 | 3.48e-01 | 1.00e+00 | 3931 | 2456 | 1475 |
| Latino/Mixed-American | Full Cohort | Switching vs Continuation | Migraine | Education_levelCollege Postgraduate | 1.062 | 0.796 | 1.417 | 6.83e-01 | 1.00e+00 | 3931 | 2456 | 1475 |
| Latino/Mixed-American | Full Cohort | Switching vs Continuation | Migraine | Education_levelCollege Undergraduate | 1.092 | 0.897 | 1.330 | 3.79e-01 | 1.00e+00 | 3931 | 2456 | 1475 |
| Latino/Mixed-American | Full Cohort | Switching vs Continuation | Migraine | Education_levelTwelve or GED | 0.952 | 0.768 | 1.180 | 6.55e-01 | 1.00e+00 | 3931 | 2456 | 1475 |
| Latino/Mixed-American | Full Cohort | Switching vs Continuation | Migraine | Migraine | 1.972 | 1.720 | 2.261 | <2.2e-16 | <2.2e-16 | 3931 | 2456 | 1475 |
| Latino/Mixed-American | Full Cohort | Discontinuation vs Continuation | Recorded Anxiety | (Intercept) | 0.551 | 0.461 | 0.658 | 4.74e-11 | 6.64e-10 | 3862 | 2456 | 1406 |
| Latino/Mixed-American | Full Cohort | Discontinuation vs Continuation | Recorded Anxiety | age_at_first_AD | 0.981 | 0.913 | 1.054 | 5.96e-01 | 1.00e+00 | 3862 | 2456 | 1406 |
| Latino/Mixed-American | Full Cohort | Discontinuation vs Continuation | Recorded Anxiety | sex_at_birthMale | 1.224 | 1.058 | 1.417 | 6.63e-03 | 9.28e-02 | 3862 | 2456 | 1406 |
| Latino/Mixed-American | Full Cohort | Discontinuation vs Continuation | Recorded Anxiety | Income>200k | 0.601 | 0.379 | 0.952 | 3.02e-02 | 4.22e-01 | 3862 | 2456 | 1406 |
| Latino/Mixed-American | Full Cohort | Discontinuation vs Continuation | Recorded Anxiety | Income100-150k | 0.676 | 0.507 | 0.902 | 7.79e-03 | 1.09e-01 | 3862 | 2456 | 1406 |
| Latino/Mixed-American | Full Cohort | Discontinuation vs Continuation | Recorded Anxiety | Income150-200k | 0.688 | 0.452 | 1.045 | 7.96e-02 | 1.00e+00 | 3862 | 2456 | 1406 |
| Latino/Mixed-American | Full Cohort | Discontinuation vs Continuation | Recorded Anxiety | Income25-35k | 0.817 | 0.657 | 1.016 | 6.91e-02 | 9.67e-01 | 3862 | 2456 | 1406 |
| Latino/Mixed-American | Full Cohort | Discontinuation vs Continuation | Recorded Anxiety | Income35-50k | 1.063 | 0.849 | 1.330 | 5.96e-01 | 1.00e+00 | 3862 | 2456 | 1406 |
| Latino/Mixed-American | Full Cohort | Discontinuation vs Continuation | Recorded Anxiety | Income50-75k | 0.831 | 0.663 | 1.042 | 1.08e-01 | 1.00e+00 | 3862 | 2456 | 1406 |
| Latino/Mixed-American | Full Cohort | Discontinuation vs Continuation | Recorded Anxiety | Income75-100k | 0.760 | 0.561 | 1.029 | 7.55e-02 | 1.00e+00 | 3862 | 2456 | 1406 |
| Latino/Mixed-American | Full Cohort | Discontinuation vs Continuation | Recorded Anxiety | Education_levelCollege Postgraduate | 0.909 | 0.668 | 1.237 | 5.44e-01 | 1.00e+00 | 3862 | 2456 | 1406 |
| Latino/Mixed-American | Full Cohort | Discontinuation vs Continuation | Recorded Anxiety | Education_levelCollege Undergraduate | 1.339 | 1.098 | 1.634 | 3.98e-03 | 5.57e-02 | 3862 | 2456 | 1406 |
| Latino/Mixed-American | Full Cohort | Discontinuation vs Continuation | Recorded Anxiety | Education_levelTwelve or GED | 1.275 | 1.030 | 1.578 | 2.55e-02 | 3.57e-01 | 3862 | 2456 | 1406 |
| Latino/Mixed-American | Full Cohort | Discontinuation vs Continuation | Recorded Anxiety | Recorded Anxiety | 0.830 | 0.725 | 0.951 | 7.11e-03 | 9.95e-02 | 3862 | 2456 | 1406 |
| Latino/Mixed-American | Full Cohort | Switching vs Continuation | Recorded Anxiety | (Intercept) | 0.256 | 0.210 | 0.312 | <2.2e-16 | <2.2e-16 | 3931 | 2456 | 1475 |
| Latino/Mixed-American | Full Cohort | Switching vs Continuation | Recorded Anxiety | age_at_first_AD | 0.869 | 0.806 | 0.938 | 3.03e-04 | 4.25e-03 | 3931 | 2456 | 1475 |
| Latino/Mixed-American | Full Cohort | Switching vs Continuation | Recorded Anxiety | sex_at_birthMale | 1.062 | 0.910 | 1.238 | 4.47e-01 | 1.00e+00 | 3931 | 2456 | 1475 |
| Latino/Mixed-American | Full Cohort | Switching vs Continuation | Recorded Anxiety | Income>200k | 0.586 | 0.364 | 0.944 | 2.80e-02 | 3.92e-01 | 3931 | 2456 | 1475 |
| Latino/Mixed-American | Full Cohort | Switching vs Continuation | Recorded Anxiety | Income100-150k | 0.765 | 0.577 | 1.015 | 6.35e-02 | 8.90e-01 | 3931 | 2456 | 1475 |
| Latino/Mixed-American | Full Cohort | Switching vs Continuation | Recorded Anxiety | Income150-200k | 0.757 | 0.495 | 1.158 | 2.00e-01 | 1.00e+00 | 3931 | 2456 | 1475 |
| Latino/Mixed-American | Full Cohort | Switching vs Continuation | Recorded Anxiety | Income25-35k | 0.827 | 0.662 | 1.033 | 9.36e-02 | 1.00e+00 | 3931 | 2456 | 1475 |
| Latino/Mixed-American | Full Cohort | Switching vs Continuation | Recorded Anxiety | Income35-50k | 1.021 | 0.811 | 1.284 | 8.62e-01 | 1.00e+00 | 3931 | 2456 | 1475 |
| Latino/Mixed-American | Full Cohort | Switching vs Continuation | Recorded Anxiety | Income50-75k | 0.737 | 0.584 | 0.929 | 9.96e-03 | 1.39e-01 | 3931 | 2456 | 1475 |
| Latino/Mixed-American | Full Cohort | Switching vs Continuation | Recorded Anxiety | Income75-100k | 0.870 | 0.647 | 1.170 | 3.58e-01 | 1.00e+00 | 3931 | 2456 | 1475 |
| Latino/Mixed-American | Full Cohort | Switching vs Continuation | Recorded Anxiety | Education_levelCollege Postgraduate | 1.025 | 0.764 | 1.376 | 8.68e-01 | 1.00e+00 | 3931 | 2456 | 1475 |
| Latino/Mixed-American | Full Cohort | Switching vs Continuation | Recorded Anxiety | Education_levelCollege Undergraduate | 1.072 | 0.877 | 1.311 | 4.96e-01 | 1.00e+00 | 3931 | 2456 | 1475 |
| Latino/Mixed-American | Full Cohort | Switching vs Continuation | Recorded Anxiety | Education_levelTwelve or GED | 0.902 | 0.725 | 1.122 | 3.52e-01 | 1.00e+00 | 3931 | 2456 | 1475 |
| Latino/Mixed-American | Full Cohort | Switching vs Continuation | Recorded Anxiety | Recorded Anxiety | 3.355 | 2.853 | 3.946 | <2.2e-16 | <2.2e-16 | 3931 | 2456 | 1475 |
| Latino/Mixed-American | Full Cohort | Discontinuation vs Continuation | Recorded Depression | (Intercept) | 0.547 | 0.459 | 0.652 | 1.55e-11 | 2.17e-10 | 3862 | 2456 | 1406 |
| Latino/Mixed-American | Full Cohort | Discontinuation vs Continuation | Recorded Depression | age_at_first_AD | 0.989 | 0.921 | 1.062 | 7.55e-01 | 1.00e+00 | 3862 | 2456 | 1406 |
| Latino/Mixed-American | Full Cohort | Discontinuation vs Continuation | Recorded Depression | sex_at_birthMale | 1.227 | 1.060 | 1.420 | 6.05e-03 | 8.47e-02 | 3862 | 2456 | 1406 |
| Latino/Mixed-American | Full Cohort | Discontinuation vs Continuation | Recorded Depression | Income>200k | 0.600 | 0.379 | 0.952 | 3.01e-02 | 4.21e-01 | 3862 | 2456 | 1406 |
| Latino/Mixed-American | Full Cohort | Discontinuation vs Continuation | Recorded Depression | Income100-150k | 0.667 | 0.500 | 0.890 | 5.87e-03 | 8.22e-02 | 3862 | 2456 | 1406 |

| | | | | | | | | | | | | |
|---|---|---|---|---|---|---|---|---|---|---|---|---|
| Latino/Mixed-American | Full Cohort | Discontinuation vs Continuation | Recorded Depression | Income150-200k | 0.668 | 0.439 | 1.017 | 6.01e-02 | 8.42e-01 | 3862 | 2456 | 1406 |
| Latino/Mixed-American | Full Cohort | Discontinuation vs Continuation | Recorded Depression | Income25-35k | 0.811 | 0.652 | 1.009 | 6.05e-02 | 8.46e-01 | 3862 | 2456 | 1406 |
| Latino/Mixed-American | Full Cohort | Discontinuation vs Continuation | Recorded Depression | Income35-50k | 1.051 | 0.839 | 1.315 | 6.67e-01 | 1.00e+00 | 3862 | 2456 | 1406 |
| Latino/Mixed-American | Full Cohort | Discontinuation vs Continuation | Recorded Depression | Income50-75k | 0.818 | 0.652 | 1.025 | 8.08e-02 | 1.00e+00 | 3862 | 2456 | 1406 |
| Latino/Mixed-American | Full Cohort | Discontinuation vs Continuation | Recorded Depression | Income75-100k | 0.743 | 0.549 | 1.007 | 5.54e-02 | 7.76e-01 | 3862 | 2456 | 1406 |
| Latino/Mixed-American | Full Cohort | Discontinuation vs Continuation | Recorded Depression | Education_levelCollege Postgraduate | 0.899 | 0.661 | 1.224 | 5.00e-01 | 1.00e+00 | 3862 | 2456 | 1406 |
| Latino/Mixed-American | Full Cohort | Discontinuation vs Continuation | Recorded Depression | Education_levelCollege Undergraduate | 1.329 | 1.090 | 1.622 | 4.97e-03 | 6.96e-02 | 3862 | 2456 | 1406 |
| Latino/Mixed-American | Full Cohort | Discontinuation vs Continuation | Recorded Depression | Education_levelTwelve or GED | 1.266 | 1.023 | 1.566 | 3.00e-02 | 4.20e-01 | 3862 | 2456 | 1406 |
| Latino/Mixed-American | Full Cohort | Discontinuation vs Continuation | Recorded Depression | Recorded Depression | 0.833 | 0.728 | 0.953 | 7.57e-03 | 1.06e-01 | 3862 | 2456 | 1406 |
| Latino/Mixed-American | Full Cohort | Switching vs Continuation | Recorded Depression | (Intercept) | 0.251 | 0.207 | 0.304 | <2.2e-16 | <2.2e-16 | 3931 | 2456 | 1475 |
| Latino/Mixed-American | Full Cohort | Switching vs Continuation | Recorded Depression | age_at_first_AD | 0.828 | 0.767 | 0.894 | 1.25e-06 | 1.75e-05 | 3931 | 2456 | 1475 |
| Latino/Mixed-American | Full Cohort | Switching vs Continuation | Recorded Depression | sex_at_birthMale | 1.033 | 0.884 | 1.206 | 6.85e-01 | 1.00e+00 | 3931 | 2456 | 1475 |
| Latino/Mixed-American | Full Cohort | Switching vs Continuation | Recorded Depression | Income>200k | 0.655 | 0.404 | 1.059 | 8.45e-02 | 1.00e+00 | 3931 | 2456 | 1475 |
| Latino/Mixed-American | Full Cohort | Switching vs Continuation | Recorded Depression | Income100-150k | 0.904 | 0.678 | 1.205 | 4.91e-01 | 1.00e+00 | 3931 | 2456 | 1475 |
| Latino/Mixed-American | Full Cohort | Switching vs Continuation | Recorded Depression | Income150-200k | 0.941 | 0.612 | 1.445 | 7.81e-01 | 1.00e+00 | 3931 | 2456 | 1475 |
| Latino/Mixed-American | Full Cohort | Switching vs Continuation | Recorded Depression | Income25-35k | 0.878 | 0.701 | 1.098 | 2.54e-01 | 1.00e+00 | 3931 | 2456 | 1475 |
| Latino/Mixed-American | Full Cohort | Switching vs Continuation | Recorded Depression | Income35-50k | 1.162 | 0.921 | 1.467 | 2.05e-01 | 1.00e+00 | 3931 | 2456 | 1475 |
| Latino/Mixed-American | Full Cohort | Switching vs Continuation | Recorded Depression | Income50-75k | 0.861 | 0.681 | 1.090 | 2.15e-01 | 1.00e+00 | 3931 | 2456 | 1475 |
| Latino/Mixed-American | Full Cohort | Switching vs Continuation | Recorded Depression | Income75-100k | 1.056 | 0.782 | 1.426 | 7.21e-01 | 1.00e+00 | 3931 | 2456 | 1475 |
| Latino/Mixed-American | Full Cohort | Switching vs Continuation | Recorded Depression | Education_levelCollege Postgraduate | 1.041 | 0.774 | 1.400 | 7.91e-01 | 1.00e+00 | 3931 | 2456 | 1475 |
| Latino/Mixed-American | Full Cohort | Switching vs Continuation | Recorded Depression | Education_levelCollege Undergraduate | 1.115 | 0.911 | 1.365 | 2.92e-01 | 1.00e+00 | 3931 | 2456 | 1475 |
| Latino/Mixed-American | Full Cohort | Switching vs Continuation | Recorded Depression | Education_levelTwelve or GED | 0.899 | 0.722 | 1.121 | 3.45e-01 | 1.00e+00 | 3931 | 2456 | 1475 |
| Latino/Mixed-American | Full Cohort | Switching vs Continuation | Recorded Depression | Recorded Depression | 3.529 | 3.047 | 4.088 | <2.2e-16 | <2.2e-16 | 3931 | 2456 | 1475 |
| Latino/Mixed-American | Full Cohort | Discontinuation vs Continuation | Smoking_Frequency | (Intercept) | 0.449 | 0.378 | 0.532 | <2.2e-16 | <2.2e-16 | 3751 | 2378 | 1373 |
| Latino/Mixed-American | Full Cohort | Discontinuation vs Continuation | Smoking_Frequency | age_at_first_AD | 0.992 | 0.922 | 1.067 | 8.22e-01 | 1.00e+00 | 3751 | 2378 | 1373 |
| Latino/Mixed-American | Full Cohort | Discontinuation vs Continuation | Smoking_Frequency | sex_at_birthMale | 1.151 | 0.989 | 1.339 | 6.92e-02 | 9.69e-01 | 3751 | 2378 | 1373 |
| Latino/Mixed-American | Full Cohort | Discontinuation vs Continuation | Smoking_Frequency | Income>200k | 0.672 | 0.422 | 1.068 | 9.25e-02 | 1.00e+00 | 3751 | 2378 | 1373 |
| Latino/Mixed-American | Full Cohort | Discontinuation vs Continuation | Smoking_Frequency | Income100-150k | 0.721 | 0.538 | 0.967 | 2.90e-02 | 4.05e-01 | 3751 | 2378 | 1373 |
| Latino/Mixed-American | Full Cohort | Discontinuation vs Continuation | Smoking_Frequency | Income150-200k | 0.747 | 0.490 | 1.139 | 1.76e-01 | 1.00e+00 | 3751 | 2378 | 1373 |
| Latino/Mixed-American | Full Cohort | Discontinuation vs Continuation | Smoking_Frequency | Income25-35k | 0.854 | 0.685 | 1.066 | 1.63e-01 | 1.00e+00 | 3751 | 2378 | 1373 |
| Latino/Mixed-American | Full Cohort | Discontinuation vs Continuation | Smoking_Frequency | Income35-50k | 1.074 | 0.855 | 1.350 | 5.39e-01 | 1.00e+00 | 3751 | 2378 | 1373 |
| Latino/Mixed-American | Full Cohort | Discontinuation vs Continuation | Smoking_Frequency | Income50-75k | 0.867 | 0.689 | 1.091 | 2.25e-01 | 1.00e+00 | 3751 | 2378 | 1373 |
| Latino/Mixed-American | Full Cohort | Discontinuation vs Continuation | Smoking_Frequency | Income75-100k | 0.788 | 0.579 | 1.073 | 1.30e-01 | 1.00e+00 | 3751 | 2378 | 1373 |
| Latino/Mixed-American | Full Cohort | Discontinuation vs Continuation | Smoking_Frequency | Education_levelCollege Postgraduate | 0.951 | 0.695 | 1.300 | 7.52e-01 | 1.00e+00 | 3751 | 2378 | 1373 |
| Latino/Mixed-American | Full Cohort | Discontinuation vs Continuation | Smoking_Frequency | Education_levelCollege Undergraduate | 1.363 | 1.114 | 1.666 | 2.57e-03 | 3.60e-02 | 3751 | 2378 | 1373 |
| Latino/Mixed-American | Full Cohort | Discontinuation vs Continuation | Smoking_Frequency | Education_levelTwelve or GED | 1.296 | 1.044 | 1.608 | 1.87e-02 | 2.61e-01 | 3751 | 2378 | 1373 |
| Latino/Mixed-American | Full Cohort | Discontinuation vs Continuation | Smoking_Frequency | Smoking_FrequencyEvery Day | 1.657 | 1.280 | 2.145 | 1.27e-04 | 1.78e-03 | 3751 | 2378 | 1373 |
| Latino/Mixed-American | Full Cohort | Discontinuation vs Continuation | Smoking_Frequency | Smoking_FrequencyNot At All (former) | 1.155 | 0.974 | 1.370 | 9.80e-02 | 1.00e+00 | 3751 | 2378 | 1373 |
| Latino/Mixed-American | Full Cohort | Discontinuation vs Continuation | Smoking_Frequency | Smoking_FrequencySome Days | 1.386 | 1.032 | 1.861 | 2.99e-02 | 4.19e-01 | 3751 | 2378 | 1373 |
| Latino/Mixed-American | Full Cohort | Switching vs Continuation | Smoking_Frequency | (Intercept) | 0.520 | 0.442 | 0.613 | 5.56e-15 | 7.78e-14 | 3801 | 2378 | 1423 |
| Latino/Mixed-American | Full Cohort | Switching vs Continuation | Smoking_Frequency | age_at_first_AD | 0.794 | 0.736 | 0.856 | 2.35e-09 | 3.30e-08 | 3801 | 2378 | 1423 |
| Latino/Mixed-American | Full Cohort | Switching vs Continuation | Smoking_Frequency | sex_at_birthMale | 0.962 | 0.824 | 1.123 | 6.23e-01 | 1.00e+00 | 3801 | 2378 | 1423 |
| Latino/Mixed-American | Full Cohort | Switching vs Continuation | Smoking_Frequency | Income>200k | 0.500 | 0.310 | 0.807 | 4.48e-03 | 6.27e-02 | 3801 | 2378 | 1423 |
| Latino/Mixed-American | Full Cohort | Switching vs Continuation | Smoking_Frequency | Income100-150k | 0.786 | 0.594 | 1.040 | 9.25e-02 | 1.00e+00 | 3801 | 2378 | 1423 |
| Latino/Mixed-American | Full Cohort | Switching vs Continuation | Smoking_Frequency | Income150-200k | 0.693 | 0.457 | 1.050 | 8.37e-02 | 1.00e+00 | 3801 | 2378 | 1423 |
| Latino/Mixed-American | Full Cohort | Switching vs Continuation | Smoking_Frequency | Income25-35k | 0.834 | 0.669 | 1.040 | 1.07e-01 | 1.00e+00 | 3801 | 2378 | 1423 |
| Latino/Mixed-American | Full Cohort | Switching vs Continuation | Smoking_Frequency | Income35-50k | 1.021 | 0.813 | 1.283 | 8.58e-01 | 1.00e+00 | 3801 | 2378 | 1423 |
| Latino/Mixed-American | Full Cohort | Switching vs Continuation | Smoking_Frequency | Income50-75k | 0.772 | 0.613 | 0.972 | 2.77e-02 | 3.88e-01 | 3801 | 2378 | 1423 |
| Latino/Mixed-American | Full Cohort | Switching vs Continuation | Smoking_Frequency | Income75-100k | 0.875 | 0.652 | 1.173 | 3.71e-01 | 1.00e+00 | 3801 | 2378 | 1423 |
| Latino/Mixed-American | Full Cohort | Switching vs Continuation | Smoking_Frequency | Education_levelCollege Postgraduate | 1.102 | 0.824 | 1.474 | 5.13e-01 | 1.00e+00 | 3801 | 2378 | 1423 |
| Latino/Mixed-American | Full Cohort | Switching vs Continuation | Smoking_Frequency | Education_levelCollege Undergraduate | 1.118 | 0.917 | 1.363 | 2.70e-01 | 1.00e+00 | 3801 | 2378 | 1423 |
| Latino/Mixed-American | Full Cohort | Switching vs Continuation | Smoking_Frequency | Education_levelTwelve or GED | 0.989 | 0.797 | 1.228 | 9.20e-01 | 1.00e+00 | 3801 | 2378 | 1423 |
| Latino/Mixed-American | Full Cohort | Switching vs Continuation | Smoking_Frequency | Smoking_FrequencyEvery Day | 1.723 | 1.332 | 2.229 | 3.43e-05 | 4.80e-04 | 3801 | 2378 | 1423 |
| Latino/Mixed-American | Full Cohort | Switching vs Continuation | Smoking_Frequency | Smoking_FrequencyNot At All (former) | 1.348 | 1.140 | 1.594 | 4.76e-04 | 6.67e-03 | 3801 | 2378 | 1423 |
| Latino/Mixed-American | Full Cohort | Switching vs Continuation | Smoking_Frequency | Smoking_FrequencySome Days | 1.191 | 0.878 | 1.617 | 2.61e-01 | 1.00e+00 | 3801 | 2378 | 1423 |
| Latino/Mixed-American | Full Cohort | Discontinuation vs Continuation | Stress | (Intercept) | 0.379 | 0.257 | 0.558 | 9.02e-07 | 1.26e-05 | 1364 | 930 | 434 |
| Latino/Mixed-American | Full Cohort | Discontinuation vs Continuation | Stress | age_at_first_AD | 0.964 | 0.848 | 1.095 | 5.69e-01 | 1.00e+00 | 1364 | 930 | 434 |
| Latino/Mixed-American | Full Cohort | Discontinuation vs Continuation | Stress | sex_at_birthMale | 1.046 | 0.801 | 1.366 | 7.40e-01 | 1.00e+00 | 1364 | 930 | 434 |
| Latino/Mixed-American | Full Cohort | Discontinuation vs Continuation | Stress | Income>200k | 0.844 | 0.455 | 1.567 | 5.91e-01 | 1.00e+00 | 1364 | 930 | 434 |
| Latino/Mixed-American | Full Cohort | Discontinuation vs Continuation | Stress | Income100-150k | 0.861 | 0.565 | 1.312 | 4.85e-01 | 1.00e+00 | 1364 | 930 | 434 |
| Latino/Mixed-American | Full Cohort | Discontinuation vs Continuation | Stress | Income150-200k | 0.806 | 0.444 | 1.462 | 4.78e-01 | 1.00e+00 | 1364 | 930 | 434 |
| Latino/Mixed-American | Full Cohort | Discontinuation vs Continuation | Stress | Income25-35k | 0.888 | 0.592 | 1.332 | 5.67e-01 | 1.00e+00 | 1364 | 930 | 434 |
| Latino/Mixed-American | Full Cohort | Discontinuation vs Continuation | Stress | Income35-50k | 1.289 | 0.863 | 1.926 | 2.14e-01 | 1.00e+00 | 1364 | 930 | 434 |
| Latino/Mixed-American | Full Cohort | Discontinuation vs Continuation | Stress | Income50-75k | 0.850 | 0.587 | 1.231 | 3.91e-01 | 1.00e+00 | 1364 | 930 | 434 |
| Latino/Mixed-American | Full Cohort | Discontinuation vs Continuation | Stress | Income75-100k | 0.687 | 0.418 | 1.127 | 1.37e-01 | 1.00e+00 | 1364 | 930 | 434 |
| Latino/Mixed-American | Full Cohort | Discontinuation vs Continuation | Stress | Education_levelCollege Postgraduate | 0.965 | 0.563 | 1.655 | 8.97e-01 | 1.00e+00 | 1364 | 930 | 434 |

| | | | | | | | | | | | | |
|---|---|---|---|---|---|---|---|---|---|---|---|---|
| Latino/Mixed-American | Full Cohort | Discontinuation vs Continuation | Stress | Education_levelCollege Undergraduate | 1.524 | 0.986 | 2.355 | 5.78e-02 | 8.09e-01 | 1364 | 930 | 434 |
| Latino/Mixed-American | Full Cohort | Discontinuation vs Continuation | Stress | Education_levelTwelve or GED | 1.461 | 0.908 | 2.349 | 1.18e-01 | 1.00e+00 | 1364 | 930 | 434 |
| Latino/Mixed-American | Full Cohort | Discontinuation vs Continuation | Stress | StressAlmost Never | 0.992 | 0.583 | 1.687 | 9.76e-01 | 1.00e+00 | 1364 | 930 | 434 |
| Latino/Mixed-American | Full Cohort | Discontinuation vs Continuation | Stress | StressFairly Often | 1.147 | 0.707 | 1.862 | 5.79e-01 | 1.00e+00 | 1364 | 930 | 434 |
| Latino/Mixed-American | Full Cohort | Discontinuation vs Continuation | Stress | StressSometimes | 0.921 | 0.712 | 1.191 | 5.31e-01 | 1.00e+00 | 1364 | 930 | 434 |
| Latino/Mixed-American | Full Cohort | Discontinuation vs Continuation | Stress | StressVery Often | 0.594 | 0.285 | 1.238 | 1.65e-01 | 1.00e+00 | 1364 | 930 | 434 |
| Latino/Mixed-American | Full Cohort | Switching vs Continuation | Stress | (Intercept) | 0.587 | 0.423 | 0.815 | 1.44e-03 | 2.01e-02 | 1563 | 930 | 633 |
| Latino/Mixed-American | Full Cohort | Switching vs Continuation | Stress | age_at_first_AD | 0.752 | 0.668 | 0.846 | 2.20e-06 | 3.08e-05 | 1563 | 930 | 633 |
| Latino/Mixed-American | Full Cohort | Switching vs Continuation | Stress | sex_at_birthMale | 1.248 | 0.986 | 1.579 | 6.51e-02 | 9.12e-01 | 1563 | 930 | 633 |
| Latino/Mixed-American | Full Cohort | Switching vs Continuation | Stress | Income>200k | 0.586 | 0.324 | 1.059 | 7.69e-02 | 1.00e+00 | 1563 | 930 | 633 |
| Latino/Mixed-American | Full Cohort | Switching vs Continuation | Stress | Income100-150k | 0.766 | 0.519 | 1.129 | 1.77e-01 | 1.00e+00 | 1563 | 930 | 633 |
| Latino/Mixed-American | Full Cohort | Switching vs Continuation | Stress | Income150-200k | 0.697 | 0.403 | 1.206 | 1.97e-01 | 1.00e+00 | 1563 | 930 | 633 |
| Latino/Mixed-American | Full Cohort | Switching vs Continuation | Stress | Income25-35k | 0.965 | 0.677 | 1.376 | 8.44e-01 | 1.00e+00 | 1563 | 930 | 633 |
| Latino/Mixed-American | Full Cohort | Switching vs Continuation | Stress | Income35-50k | 1.262 | 0.875 | 1.820 | 2.13e-01 | 1.00e+00 | 1563 | 930 | 633 |
| Latino/Mixed-American | Full Cohort | Switching vs Continuation | Stress | Income50-75k | 0.803 | 0.574 | 1.123 | 1.99e-01 | 1.00e+00 | 1563 | 930 | 633 |
| Latino/Mixed-American | Full Cohort | Switching vs Continuation | Stress | Income75-100k | 1.043 | 0.698 | 1.556 | 8.38e-01 | 1.00e+00 | 1563 | 930 | 633 |
| Latino/Mixed-American | Full Cohort | Switching vs Continuation | Stress | Education_levelCollege Postgraduate | 0.846 | 0.535 | 1.339 | 4.76e-01 | 1.00e+00 | 1563 | 930 | 633 |
| Latino/Mixed-American | Full Cohort | Switching vs Continuation | Stress | Education_levelCollege Undergraduate | 1.103 | 0.760 | 1.602 | 6.05e-01 | 1.00e+00 | 1563 | 930 | 633 |
| Latino/Mixed-American | Full Cohort | Switching vs Continuation | Stress | Education_levelTwelve or GED | 0.953 | 0.626 | 1.450 | 8.22e-01 | 1.00e+00 | 1563 | 930 | 633 |
| Latino/Mixed-American | Full Cohort | Switching vs Continuation | Stress | StressAlmost Never | 1.208 | 0.757 | 1.927 | 4.27e-01 | 1.00e+00 | 1563 | 930 | 633 |
| Latino/Mixed-American | Full Cohort | Switching vs Continuation | Stress | StressFairly Often | 1.129 | 0.725 | 1.758 | 5.91e-01 | 1.00e+00 | 1563 | 930 | 633 |
| Latino/Mixed-American | Full Cohort | Switching vs Continuation | Stress | StressSometimes | 1.059 | 0.844 | 1.330 | 6.19e-01 | 1.00e+00 | 1563 | 930 | 633 |
| Latino/Mixed-American | Full Cohort | Switching vs Continuation | Stress | StressVery Often | 0.789 | 0.437 | 1.426 | 4.33e-01 | 1.00e+00 | 1563 | 930 | 633 |
| Latino/Mixed-American | Full Cohort | Discontinuation vs Continuation | age_at_first_AD | (Intercept) | 0.536 | 0.494 | 0.581 | <2.2e-16 | <2.2e-16 | 3862 | 2456 | 1406 |
| Latino/Mixed-American | Full Cohort | Discontinuation vs Continuation | age_at_first_AD | age_at_first_AD | 0.970 | 0.906 | 1.039 | 3.84e-01 | 1.00e+00 | 3862 | 2456 | 1406 |
| Latino/Mixed-American | Full Cohort | Discontinuation vs Continuation | age_at_first_AD | sex_at_birthMale | 1.226 | 1.061 | 1.417 | 5.85e-03 | 8.19e-02 | 3862 | 2456 | 1406 |
| Latino/Mixed-American | Full Cohort | Switching vs Continuation | age_at_first_AD | (Intercept) | 0.551 | 0.508 | 0.598 | <2.2e-16 | <2.2e-16 | 3931 | 2456 | 1475 |
| Latino/Mixed-American | Full Cohort | Switching vs Continuation | age_at_first_AD | age_at_first_AD | 0.806 | 0.751 | 0.865 | 1.95e-09 | 2.73e-08 | 3931 | 2456 | 1475 |
| Latino/Mixed-American | Full Cohort | Switching vs Continuation | age_at_first_AD | sex_at_birthMale | 1.047 | 0.903 | 1.213 | 5.46e-01 | 1.00e+00 | 3931 | 2456 | 1475 |
| Latino/Mixed-American | Full Cohort | Discontinuation vs Continuation | sex_at_birth | (Intercept) | 0.536 | 0.494 | 0.581 | <2.2e-16 | <2.2e-16 | 3862 | 2456 | 1406 |
| Latino/Mixed-American | Full Cohort | Discontinuation vs Continuation | sex_at_birth | age_at_first_AD | 0.970 | 0.906 | 1.039 | 3.84e-01 | 1.00e+00 | 3862 | 2456 | 1406 |
| Latino/Mixed-American | Full Cohort | Discontinuation vs Continuation | sex_at_birth | sex_at_birthMale | 1.226 | 1.061 | 1.417 | 5.85e-03 | 8.19e-02 | 3862 | 2456 | 1406 |
| Latino/Mixed-American | Full Cohort | Switching vs Continuation | sex_at_birth | (Intercept) | 0.551 | 0.508 | 0.598 | <2.2e-16 | <2.2e-16 | 3931 | 2456 | 1475 |
| Latino/Mixed-American | Full Cohort | Switching vs Continuation | sex_at_birth | age_at_first_AD | 0.806 | 0.751 | 0.865 | 1.95e-09 | 2.73e-08 | 3931 | 2456 | 1475 |
| Latino/Mixed-American | Full Cohort | Switching vs Continuation | sex_at_birth | sex_at_birthMale | 1.047 | 0.903 | 1.213 | 5.46e-01 | 1.00e+00 | 3931 | 2456 | 1475 |
| Latino/Mixed-American | Recorded Depression | Discontinuation vs Continuation | Alcohol_Frequency | (Intercept) | 0.469 | 0.344 | 0.639 | 1.65e-06 | 2.30e-05 | 1764 | 1155 | 609 |
| Latino/Mixed-American | Recorded Depression | Discontinuation vs Continuation | Alcohol_Frequency | age_at_first_AD | 0.974 | 0.873 | 1.087 | 6.42e-01 | 1.00e+00 | 1764 | 1155 | 609 |
| Latino/Mixed-American | Recorded Depression | Discontinuation vs Continuation | Alcohol_Frequency | sex_at_birthMale | 1.127 | 0.902 | 1.408 | 2.94e-01 | 1.00e+00 | 1764 | 1155 | 609 |
| Latino/Mixed-American | Recorded Depression | Discontinuation vs Continuation | Alcohol_Frequency | Income>200k | 0.553 | 0.237 | 1.293 | 1.72e-01 | 1.00e+00 | 1764 | 1155 | 609 |
| Latino/Mixed-American | Recorded Depression | Discontinuation vs Continuation | Alcohol_Frequency | Income100-150k | 0.608 | 0.377 | 0.980 | 4.11e-02 | 5.75e-01 | 1764 | 1155 | 609 |
| Latino/Mixed-American | Recorded Depression | Discontinuation vs Continuation | Alcohol_Frequency | Income150-200k | 1.157 | 0.527 | 2.539 | 7.16e-01 | 1.00e+00 | 1764 | 1155 | 609 |
| Latino/Mixed-American | Recorded Depression | Discontinuation vs Continuation | Alcohol_Frequency | Income25-35k | 0.894 | 0.650 | 1.229 | 4.91e-01 | 1.00e+00 | 1764 | 1155 | 609 |
| Latino/Mixed-American | Recorded Depression | Discontinuation vs Continuation | Alcohol_Frequency | Income35-50k | 1.097 | 0.776 | 1.550 | 6.02e-01 | 1.00e+00 | 1764 | 1155 | 609 |
| Latino/Mixed-American | Recorded Depression | Discontinuation vs Continuation | Alcohol_Frequency | Income50-75k | 0.829 | 0.582 | 1.181 | 2.99e-01 | 1.00e+00 | 1764 | 1155 | 609 |
| Latino/Mixed-American | Recorded Depression | Discontinuation vs Continuation | Alcohol_Frequency | Income75-100k | 0.942 | 0.585 | 1.516 | 8.05e-01 | 1.00e+00 | 1764 | 1155 | 609 |
| Latino/Mixed-American | Recorded Depression | Discontinuation vs Continuation | Alcohol_Frequency | Education_levelCollege Postgraduate | 0.832 | 0.504 | 1.371 | 4.70e-01 | 1.00e+00 | 1764 | 1155 | 609 |
| Latino/Mixed-American | Recorded Depression | Discontinuation vs Continuation | Alcohol_Frequency | Education_levelCollege Undergraduate | 1.308 | 0.980 | 1.745 | 6.83e-02 | 9.57e-01 | 1764 | 1155 | 609 |
| Latino/Mixed-American | Recorded Depression | Discontinuation vs Continuation | Alcohol_Frequency | Education_levelTwelve or GED | 1.019 | 0.750 | 1.385 | 9.05e-01 | 1.00e+00 | 1764 | 1155 | 609 |
| Latino/Mixed-American | Recorded Depression | Discontinuation vs Continuation | Alcohol_Frequency | Alcohol_Frequency2-3 Week | 0.993 | 0.625 | 1.576 | 9.75e-01 | 1.00e+00 | 1764 | 1155 | 609 |
| Latino/Mixed-American | Recorded Depression | Discontinuation vs Continuation | Alcohol_Frequency | Alcohol_Frequency2-4 Month | 0.824 | 0.562 | 1.210 | 3.24e-01 | 1.00e+00 | 1764 | 1155 | 609 |
| Latino/Mixed-American | Recorded Depression | Discontinuation vs Continuation | Alcohol_Frequency | Alcohol_Frequency4plus Week | 1.601 | 0.926 | 2.768 | 9.20e-02 | 1.00e+00 | 1764 | 1155 | 609 |
| Latino/Mixed-American | Recorded Depression | Discontinuation vs Continuation | Alcohol_Frequency | Alcohol_FrequencyMonthly or Less | 1.059 | 0.773 | 1.450 | 7.23e-01 | 1.00e+00 | 1764 | 1155 | 609 |
| Latino/Mixed-American | Recorded Depression | Discontinuation vs Continuation | Alcohol_Frequency | Alcohol_FrequencyNever (former) | 1.044 | 0.740 | 1.472 | 8.08e-01 | 1.00e+00 | 1764 | 1155 | 609 |
| Latino/Mixed-American | Recorded Depression | Switching vs Continuation | Alcohol_Frequency | (Intercept) | 1.019 | 0.794 | 1.309 | 8.81e-01 | 1.00e+00 | 2265 | 1155 | 1110 |
| Latino/Mixed-American | Recorded Depression | Switching vs Continuation | Alcohol_Frequency | age_at_first_AD | 0.821 | 0.746 | 0.904 | 6.01e-05 | 8.41e-04 | 2265 | 1155 | 1110 |
| Latino/Mixed-American | Recorded Depression | Switching vs Continuation | Alcohol_Frequency | sex_at_birthMale | 0.934 | 0.769 | 1.133 | 4.86e-01 | 1.00e+00 | 2265 | 1155 | 1110 |
| Latino/Mixed-American | Recorded Depression | Switching vs Continuation | Alcohol_Frequency | Income>200k | 0.551 | 0.283 | 1.072 | 7.92e-02 | 1.00e+00 | 2265 | 1155 | 1110 |
| Latino/Mixed-American | Recorded Depression | Switching vs Continuation | Alcohol_Frequency | Income100-150k | 0.740 | 0.508 | 1.079 | 1.17e-01 | 1.00e+00 | 2265 | 1155 | 1110 |
| Latino/Mixed-American | Recorded Depression | Switching vs Continuation | Alcohol_Frequency | Income150-200k | 1.364 | 0.723 | 2.574 | 3.37e-01 | 1.00e+00 | 2265 | 1155 | 1110 |
| Latino/Mixed-American | Recorded Depression | Switching vs Continuation | Alcohol_Frequency | Income25-35k | 0.927 | 0.706 | 1.216 | 5.82e-01 | 1.00e+00 | 2265 | 1155 | 1110 |
| Latino/Mixed-American | Recorded Depression | Switching vs Continuation | Alcohol_Frequency | Income35-50k | 1.240 | 0.925 | 1.664 | 1.51e-01 | 1.00e+00 | 2265 | 1155 | 1110 |
| Latino/Mixed-American | Recorded Depression | Switching vs Continuation | Alcohol_Frequency | Income50-75k | 0.891 | 0.663 | 1.197 | 4.42e-01 | 1.00e+00 | 2265 | 1155 | 1110 |
| Latino/Mixed-American | Recorded Depression | Switching vs Continuation | Alcohol_Frequency | Income75-100k | 1.123 | 0.755 | 1.670 | 5.68e-01 | 1.00e+00 | 2265 | 1155 | 1110 |
| Latino/Mixed-American | Recorded Depression | Switching vs Continuation | Alcohol_Frequency | Education_levelCollege Postgraduate | 1.258 | 0.864 | 1.832 | 2.31e-01 | 1.00e+00 | 2265 | 1155 | 1110 |
| Latino/Mixed-American | Recorded Depression | Switching vs Continuation | Alcohol_Frequency | Education_levelCollege Undergraduate | 1.164 | 0.909 | 1.489 | 2.28e-01 | 1.00e+00 | 2265 | 1155 | 1110 |
| Latino/Mixed-American | Recorded Depression | Switching vs Continuation | Alcohol_Frequency | Education_levelTwelve or GED | 0.864 | 0.665 | 1.122 | 2.73e-01 | 1.00e+00 | 2265 | 1155 | 1110 |

| | | | | | | | | | | | | |
|---|---|---|---|---|---|---|---|---|---|---|---|---|
| Latino/Mixed-American | Recorded Depression | Switching vs Continuation | Alcohol_Frequency | Alcohol_Frequency2-3 Week | 0.728 | 0.493 | 1.074 | 1.10e-01 | 1.00e+00 | 2265 | 1155 | 1110 |
| Latino/Mixed-American | Recorded Depression | Switching vs Continuation | Alcohol_Frequency | Alcohol_Frequency2-4 Month | 0.663 | 0.482 | 0.910 | 1.10e-02 | 1.54e-01 | 2265 | 1155 | 1110 |
| Latino/Mixed-American | Recorded Depression | Switching vs Continuation | Alcohol_Frequency | Alcohol_Frequency4plus Week | 1.370 | 0.859 | 2.186 | 1.87e-01 | 1.00e+00 | 2265 | 1155 | 1110 |
| Latino/Mixed-American | Recorded Depression | Switching vs Continuation | Alcohol_Frequency | Alcohol_FrequencyMonthly or Less | 0.813 | 0.625 | 1.057 | 1.23e-01 | 1.00e+00 | 2265 | 1155 | 1110 |
| Latino/Mixed-American | Recorded Depression | Switching vs Continuation | Alcohol_Frequency | Alcohol_FrequencyNever (former) | 0.981 | 0.738 | 1.303 | 8.92e-01 | 1.00e+00 | 2265 | 1155 | 1110 |
| Latino/Mixed-American | Recorded Depression | Discontinuation vs Continuation | BMI | (Intercept) | 0.458 | 0.362 | 0.579 | 7.97e-11 | 1.12e-09 | 1706 | 1137 | 569 |
| Latino/Mixed-American | Recorded Depression | Discontinuation vs Continuation | BMI | age_at_first_AD | 0.937 | 0.837 | 1.048 | 2.54e-01 | 1.00e+00 | 1706 | 1137 | 569 |
| Latino/Mixed-American | Recorded Depression | Discontinuation vs Continuation | BMI | sex_at_birthMale | 1.188 | 0.947 | 1.489 | 1.36e-01 | 1.00e+00 | 1706 | 1137 | 569 |
| Latino/Mixed-American | Recorded Depression | Discontinuation vs Continuation | BMI | Income>200k | 0.637 | 0.273 | 1.485 | 2.96e-01 | 1.00e+00 | 1706 | 1137 | 569 |
| Latino/Mixed-American | Recorded Depression | Discontinuation vs Continuation | BMI | Income100-150k | 0.616 | 0.381 | 0.999 | 4.94e-02 | 6.92e-01 | 1706 | 1137 | 569 |
| Latino/Mixed-American | Recorded Depression | Discontinuation vs Continuation | BMI | Income150-200k | 1.105 | 0.493 | 2.476 | 8.09e-01 | 1.00e+00 | 1706 | 1137 | 569 |
| Latino/Mixed-American | Recorded Depression | Discontinuation vs Continuation | BMI | Income25-35k | 0.897 | 0.645 | 1.248 | 5.18e-01 | 1.00e+00 | 1706 | 1137 | 569 |
| Latino/Mixed-American | Recorded Depression | Discontinuation vs Continuation | BMI | Income35-50k | 1.107 | 0.781 | 1.569 | 5.68e-01 | 1.00e+00 | 1706 | 1137 | 569 |
| Latino/Mixed-American | Recorded Depression | Discontinuation vs Continuation | BMI | Income50-75k | 0.759 | 0.526 | 1.094 | 1.39e-01 | 1.00e+00 | 1706 | 1137 | 569 |
| Latino/Mixed-American | Recorded Depression | Discontinuation vs Continuation | BMI | Income75-100k | 0.888 | 0.547 | 1.443 | 6.32e-01 | 1.00e+00 | 1706 | 1137 | 569 |
| Latino/Mixed-American | Recorded Depression | Discontinuation vs Continuation | BMI | Education_levelCollege Postgraduate | 0.744 | 0.443 | 1.251 | 2.64e-01 | 1.00e+00 | 1706 | 1137 | 569 |
| Latino/Mixed-American | Recorded Depression | Discontinuation vs Continuation | BMI | Education_levelCollege Undergraduate | 1.291 | 0.961 | 1.733 | 8.96e-02 | 1.00e+00 | 1706 | 1137 | 569 |
| Latino/Mixed-American | Recorded Depression | Discontinuation vs Continuation | BMI | Education_levelTwelve or GED | 0.987 | 0.722 | 1.349 | 9.35e-01 | 1.00e+00 | 1706 | 1137 | 569 |
| Latino/Mixed-American | Recorded Depression | Discontinuation vs Continuation | BMI | BMI | 0.966 | 0.875 | 1.065 | 4.86e-01 | 1.00e+00 | 1706 | 1137 | 569 |
| Latino/Mixed-American | Recorded Depression | Switching vs Continuation | BMI | (Intercept) | 0.906 | 0.750 | 1.094 | 3.05e-01 | 1.00e+00 | 2236 | 1137 | 1099 |
| Latino/Mixed-American | Recorded Depression | Switching vs Continuation | BMI | age_at_first_AD | 0.851 | 0.774 | 0.936 | 8.49e-04 | 1.19e-02 | 2236 | 1137 | 1099 |
| Latino/Mixed-American | Recorded Depression | Switching vs Continuation | BMI | sex_at_birthMale | 0.955 | 0.788 | 1.158 | 6.41e-01 | 1.00e+00 | 2236 | 1137 | 1099 |
| Latino/Mixed-American | Recorded Depression | Switching vs Continuation | BMI | Income>200k | 0.578 | 0.302 | 1.107 | 9.84e-02 | 1.00e+00 | 2236 | 1137 | 1099 |
| Latino/Mixed-American | Recorded Depression | Switching vs Continuation | BMI | Income100-150k | 0.713 | 0.490 | 1.037 | 7.66e-02 | 1.00e+00 | 2236 | 1137 | 1099 |
| Latino/Mixed-American | Recorded Depression | Switching vs Continuation | BMI | Income150-200k | 1.223 | 0.643 | 2.326 | 5.39e-01 | 1.00e+00 | 2236 | 1137 | 1099 |
| Latino/Mixed-American | Recorded Depression | Switching vs Continuation | BMI | Income25-35k | 0.973 | 0.741 | 1.279 | 8.46e-01 | 1.00e+00 | 2236 | 1137 | 1099 |
| Latino/Mixed-American | Recorded Depression | Switching vs Continuation | BMI | Income35-50k | 1.219 | 0.909 | 1.633 | 1.86e-01 | 1.00e+00 | 2236 | 1137 | 1099 |
| Latino/Mixed-American | Recorded Depression | Switching vs Continuation | BMI | Income50-75k | 0.858 | 0.640 | 1.150 | 3.04e-01 | 1.00e+00 | 2236 | 1137 | 1099 |
| Latino/Mixed-American | Recorded Depression | Switching vs Continuation | BMI | Income75-100k | 1.049 | 0.710 | 1.551 | 8.09e-01 | 1.00e+00 | 2236 | 1137 | 1099 |
| Latino/Mixed-American | Recorded Depression | Switching vs Continuation | BMI | Education_levelCollege Postgraduate | 1.210 | 0.833 | 1.757 | 3.18e-01 | 1.00e+00 | 2236 | 1137 | 1099 |
| Latino/Mixed-American | Recorded Depression | Switching vs Continuation | BMI | Education_levelCollege Undergraduate | 1.143 | 0.894 | 1.460 | 2.87e-01 | 1.00e+00 | 2236 | 1137 | 1099 |
| Latino/Mixed-American | Recorded Depression | Switching vs Continuation | BMI | Education_levelTwelve or GED | 0.874 | 0.674 | 1.134 | 3.12e-01 | 1.00e+00 | 2236 | 1137 | 1099 |
| Latino/Mixed-American | Recorded Depression | Switching vs Continuation | BMI | BMI | 0.973 | 0.893 | 1.060 | 5.25e-01 | 1.00e+00 | 2236 | 1137 | 1099 |
| Latino/Mixed-American | Recorded Depression | Discontinuation vs Continuation | Education_level | (Intercept) | 0.491 | 0.392 | 0.615 | 6.39e-10 | 8.95e-09 | 1805 | 1179 | 626 |
| Latino/Mixed-American | Recorded Depression | Discontinuation vs Continuation | Education_level | age_at_first_AD | 0.968 | 0.870 | 1.077 | 5.48e-01 | 1.00e+00 | 1805 | 1179 | 626 |
| Latino/Mixed-American | Recorded Depression | Discontinuation vs Continuation | Education_level | sex_at_birthMale | 1.140 | 0.917 | 1.417 | 2.37e-01 | 1.00e+00 | 1805 | 1179 | 626 |
| Latino/Mixed-American | Recorded Depression | Discontinuation vs Continuation | Education_level | Income>200k | 0.582 | 0.252 | 1.344 | 2.05e-01 | 1.00e+00 | 1805 | 1179 | 626 |
| Latino/Mixed-American | Recorded Depression | Discontinuation vs Continuation | Education_level | Income100-150k | 0.607 | 0.379 | 0.971 | 3.71e-02 | 5.19e-01 | 1805 | 1179 | 626 |
| Latino/Mixed-American | Recorded Depression | Discontinuation vs Continuation | Education_level | Income150-200k | 1.092 | 0.503 | 2.370 | 8.24e-01 | 1.00e+00 | 1805 | 1179 | 626 |
| Latino/Mixed-American | Recorded Depression | Discontinuation vs Continuation | Education_level | Income25-35k | 0.921 | 0.673 | 1.259 | 6.04e-01 | 1.00e+00 | 1805 | 1179 | 626 |
| Latino/Mixed-American | Recorded Depression | Discontinuation vs Continuation | Education_level | Income35-50k | 1.110 | 0.794 | 1.552 | 5.41e-01 | 1.00e+00 | 1805 | 1179 | 626 |
| Latino/Mixed-American | Recorded Depression | Discontinuation vs Continuation | Education_level | Income50-75k | 0.808 | 0.571 | 1.144 | 2.29e-01 | 1.00e+00 | 1805 | 1179 | 626 |
| Latino/Mixed-American | Recorded Depression | Discontinuation vs Continuation | Education_level | Income75-100k | 0.941 | 0.591 | 1.501 | 8.00e-01 | 1.00e+00 | 1805 | 1179 | 626 |
| Latino/Mixed-American | Recorded Depression | Discontinuation vs Continuation | Education_level | Education_levelCollege Postgraduate | 0.775 | 0.473 | 1.269 | 3.11e-01 | 1.00e+00 | 1805 | 1179 | 626 |
| Latino/Mixed-American | Recorded Depression | Discontinuation vs Continuation | Education_level | Education_levelCollege Undergraduate | 1.265 | 0.953 | 1.677 | 1.03e-01 | 1.00e+00 | 1805 | 1179 | 626 |
| Latino/Mixed-American | Recorded Depression | Discontinuation vs Continuation | Education_level | Education_levelTwelve or GED | 0.994 | 0.735 | 1.343 | 9.66e-01 | 1.00e+00 | 1805 | 1179 | 626 |
| Latino/Mixed-American | Recorded Depression | Switching vs Continuation | Education_level | (Intercept) | 0.920 | 0.764 | 1.108 | 3.81e-01 | 1.00e+00 | 2309 | 1179 | 1130 |
| Latino/Mixed-American | Recorded Depression | Switching vs Continuation | Education_level | age_at_first_AD | 0.849 | 0.773 | 0.931 | 5.53e-04 | 7.74e-03 | 2309 | 1179 | 1130 |
| Latino/Mixed-American | Recorded Depression | Switching vs Continuation | Education_level | sex_at_birthMale | 0.958 | 0.793 | 1.157 | 6.55e-01 | 1.00e+00 | 2309 | 1179 | 1130 |
| Latino/Mixed-American | Recorded Depression | Switching vs Continuation | Education_level | Income>200k | 0.568 | 0.299 | 1.080 | 8.43e-02 | 1.00e+00 | 2309 | 1179 | 1130 |
| Latino/Mixed-American | Recorded Depression | Switching vs Continuation | Education_level | Income100-150k | 0.741 | 0.513 | 1.070 | 1.10e-01 | 1.00e+00 | 2309 | 1179 | 1130 |
| Latino/Mixed-American | Recorded Depression | Switching vs Continuation | Education_level | Income150-200k | 1.271 | 0.682 | 2.369 | 4.50e-01 | 1.00e+00 | 2309 | 1179 | 1130 |
| Latino/Mixed-American | Recorded Depression | Switching vs Continuation | Education_level | Income25-35k | 0.940 | 0.719 | 1.229 | 6.50e-01 | 1.00e+00 | 2309 | 1179 | 1130 |
| Latino/Mixed-American | Recorded Depression | Switching vs Continuation | Education_level | Income35-50k | 1.222 | 0.917 | 1.627 | 1.71e-01 | 1.00e+00 | 2309 | 1179 | 1130 |
| Latino/Mixed-American | Recorded Depression | Switching vs Continuation | Education_level | Income50-75k | 0.860 | 0.645 | 1.148 | 3.07e-01 | 1.00e+00 | 2309 | 1179 | 1130 |
| Latino/Mixed-American | Recorded Depression | Switching vs Continuation | Education_level | Income75-100k | 1.075 | 0.731 | 1.582 | 7.14e-01 | 1.00e+00 | 2309 | 1179 | 1130 |
| Latino/Mixed-American | Recorded Depression | Switching vs Continuation | Education_level | Education_levelCollege Postgraduate | 1.181 | 0.818 | 1.704 | 3.74e-01 | 1.00e+00 | 2309 | 1179 | 1130 |
| Latino/Mixed-American | Recorded Depression | Switching vs Continuation | Education_level | Education_levelCollege Undergraduate | 1.097 | 0.862 | 1.396 | 4.51e-01 | 1.00e+00 | 2309 | 1179 | 1130 |
| Latino/Mixed-American | Recorded Depression | Switching vs Continuation | Education_level | Education_levelTwelve or GED | 0.832 | 0.643 | 1.076 | 1.61e-01 | 1.00e+00 | 2309 | 1179 | 1130 |
| Latino/Mixed-American | Recorded Depression | Discontinuation vs Continuation | Income | (Intercept) | 0.491 | 0.392 | 0.615 | 6.39e-10 | 8.95e-09 | 1805 | 1179 | 626 |
| Latino/Mixed-American | Recorded Depression | Discontinuation vs Continuation | Income | age_at_first_AD | 0.968 | 0.870 | 1.077 | 5.48e-01 | 1.00e+00 | 1805 | 1179 | 626 |
| Latino/Mixed-American | Recorded Depression | Discontinuation vs Continuation | Income | sex_at_birthMale | 1.140 | 0.917 | 1.417 | 2.37e-01 | 1.00e+00 | 1805 | 1179 | 626 |
| Latino/Mixed-American | Recorded Depression | Discontinuation vs Continuation | Income | Income>200k | 0.582 | 0.252 | 1.344 | 2.05e-01 | 1.00e+00 | 1805 | 1179 | 626 |
| Latino/Mixed-American | Recorded Depression | Discontinuation vs Continuation | Income | Income100-150k | 0.607 | 0.379 | 0.971 | 3.71e-02 | 5.19e-01 | 1805 | 1179 | 626 |
| Latino/Mixed-American | Recorded Depression | Discontinuation vs Continuation | Income | Income150-200k | 1.092 | 0.503 | 2.370 | 8.24e-01 | 1.00e+00 | 1805 | 1179 | 626 |
| Latino/Mixed-American | Recorded Depression | Discontinuation vs Continuation | Income | Income25-35k | 0.921 | 0.673 | 1.259 | 6.04e-01 | 1.00e+00 | 1805 | 1179 | 626 |

| | | | | | | | | | | | | |
|---|---|---|---|---|---|---|---|---|---|---|---|---|
| Latino/Mixed-American | Recorded Depression | Discontinuation vs Continuation | Income | Income35-50k | 1.110 | 0.794 | 1.552 | 5.41e-01 | 1.00e+00 | 1805 | 1179 | 626 |
| Latino/Mixed-American | Recorded Depression | Discontinuation vs Continuation | Income | Income50-75k | 0.808 | 0.571 | 1.144 | 2.29e-01 | 1.00e+00 | 1805 | 1179 | 626 |
| Latino/Mixed-American | Recorded Depression | Discontinuation vs Continuation | Income | Income75-100k | 0.941 | 0.591 | 1.501 | 8.00e-01 | 1.00e+00 | 1805 | 1179 | 626 |
| Latino/Mixed-American | Recorded Depression | Discontinuation vs Continuation | Income | Education_levelCollege Postgraduate | 0.775 | 0.473 | 1.269 | 3.11e-01 | 1.00e+00 | 1805 | 1179 | 626 |
| Latino/Mixed-American | Recorded Depression | Discontinuation vs Continuation | Income | Education_levelCollege Undergraduate | 1.265 | 0.953 | 1.677 | 1.03e-01 | 1.00e+00 | 1805 | 1179 | 626 |
| Latino/Mixed-American | Recorded Depression | Discontinuation vs Continuation | Income | Education_levelTwelve or GED | 0.994 | 0.735 | 1.343 | 9.66e-01 | 1.00e+00 | 1805 | 1179 | 626 |
| Latino/Mixed-American | Recorded Depression | Switching vs Continuation | Income | (Intercept) | 0.920 | 0.764 | 1.108 | 3.81e-01 | 1.00e+00 | 2309 | 1179 | 1130 |
| Latino/Mixed-American | Recorded Depression | Switching vs Continuation | Income | age_at_first_AD | 0.849 | 0.773 | 0.931 | 5.53e-04 | 7.74e-03 | 2309 | 1179 | 1130 |
| Latino/Mixed-American | Recorded Depression | Switching vs Continuation | Income | sex_at_birthMale | 0.958 | 0.793 | 1.157 | 6.55e-01 | 1.00e+00 | 2309 | 1179 | 1130 |
| Latino/Mixed-American | Recorded Depression | Switching vs Continuation | Income | Income>200k | 0.568 | 0.299 | 1.080 | 8.43e-02 | 1.00e+00 | 2309 | 1179 | 1130 |
| Latino/Mixed-American | Recorded Depression | Switching vs Continuation | Income | Income100-150k | 0.741 | 0.513 | 1.070 | 1.10e-01 | 1.00e+00 | 2309 | 1179 | 1130 |
| Latino/Mixed-American | Recorded Depression | Switching vs Continuation | Income | Income150-200k | 1.271 | 0.682 | 2.369 | 4.50e-01 | 1.00e+00 | 2309 | 1179 | 1130 |
| Latino/Mixed-American | Recorded Depression | Switching vs Continuation | Income | Income25-35k | 0.940 | 0.719 | 1.229 | 6.50e-01 | 1.00e+00 | 2309 | 1179 | 1130 |
| Latino/Mixed-American | Recorded Depression | Switching vs Continuation | Income | Income35-50k | 1.222 | 0.917 | 1.627 | 1.71e-01 | 1.00e+00 | 2309 | 1179 | 1130 |
| Latino/Mixed-American | Recorded Depression | Switching vs Continuation | Income | Income50-75k | 0.860 | 0.645 | 1.148 | 3.07e-01 | 1.00e+00 | 2309 | 1179 | 1130 |
| Latino/Mixed-American | Recorded Depression | Switching vs Continuation | Income | Income75-100k | 1.075 | 0.731 | 1.582 | 7.14e-01 | 1.00e+00 | 2309 | 1179 | 1130 |
| Latino/Mixed-American | Recorded Depression | Switching vs Continuation | Income | Education_levelCollege Postgraduate | 1.181 | 0.818 | 1.704 | 3.74e-01 | 1.00e+00 | 2309 | 1179 | 1130 |
| Latino/Mixed-American | Recorded Depression | Switching vs Continuation | Income | Education_levelCollege Undergraduate | 1.097 | 0.862 | 1.396 | 4.51e-01 | 1.00e+00 | 2309 | 1179 | 1130 |
| Latino/Mixed-American | Recorded Depression | Switching vs Continuation | Income | Education_levelTwelve or GED | 0.832 | 0.643 | 1.076 | 1.61e-01 | 1.00e+00 | 2309 | 1179 | 1130 |
| Latino/Mixed-American | Recorded Depression | Discontinuation vs Continuation | Insomnia | (Intercept) | 0.505 | 0.400 | 0.637 | 8.05e-09 | 1.13e-07 | 1805 | 1179 | 626 |
| Latino/Mixed-American | Recorded Depression | Discontinuation vs Continuation | Insomnia | age_at_first_AD | 0.975 | 0.875 | 1.085 | 6.38e-01 | 1.00e+00 | 1805 | 1179 | 626 |
| Latino/Mixed-American | Recorded Depression | Discontinuation vs Continuation | Insomnia | sex_at_birthMale | 1.145 | 0.921 | 1.423 | 2.22e-01 | 1.00e+00 | 1805 | 1179 | 626 |
| Latino/Mixed-American | Recorded Depression | Discontinuation vs Continuation | Insomnia | Income>200k | 0.587 | 0.254 | 1.355 | 2.12e-01 | 1.00e+00 | 1805 | 1179 | 626 |
| Latino/Mixed-American | Recorded Depression | Discontinuation vs Continuation | Insomnia | Income100-150k | 0.598 | 0.373 | 0.957 | 3.22e-02 | 4.51e-01 | 1805 | 1179 | 626 |
| Latino/Mixed-American | Recorded Depression | Discontinuation vs Continuation | Insomnia | Income150-200k | 1.084 | 0.499 | 2.354 | 8.38e-01 | 1.00e+00 | 1805 | 1179 | 626 |
| Latino/Mixed-American | Recorded Depression | Discontinuation vs Continuation | Insomnia | Income25-35k | 0.918 | 0.671 | 1.255 | 5.90e-01 | 1.00e+00 | 1805 | 1179 | 626 |
| Latino/Mixed-American | Recorded Depression | Discontinuation vs Continuation | Insomnia | Income35-50k | 1.103 | 0.788 | 1.542 | 5.68e-01 | 1.00e+00 | 1805 | 1179 | 626 |
| Latino/Mixed-American | Recorded Depression | Discontinuation vs Continuation | Insomnia | Income50-75k | 0.808 | 0.571 | 1.143 | 2.28e-01 | 1.00e+00 | 1805 | 1179 | 626 |
| Latino/Mixed-American | Recorded Depression | Discontinuation vs Continuation | Insomnia | Income75-100k | 0.935 | 0.586 | 1.491 | 7.76e-01 | 1.00e+00 | 1805 | 1179 | 626 |
| Latino/Mixed-American | Recorded Depression | Discontinuation vs Continuation | Insomnia | Education_levelCollege Postgraduate | 0.776 | 0.474 | 1.272 | 3.14e-01 | 1.00e+00 | 1805 | 1179 | 626 |
| Latino/Mixed-American | Recorded Depression | Discontinuation vs Continuation | Insomnia | Education_levelCollege Undergraduate | 1.275 | 0.961 | 1.692 | 9.23e-02 | 1.00e+00 | 1805 | 1179 | 626 |
| Latino/Mixed-American | Recorded Depression | Discontinuation vs Continuation | Insomnia | Education_levelTwelve or GED | 0.999 | 0.739 | 1.350 | 9.95e-01 | 1.00e+00 | 1805 | 1179 | 626 |
| Latino/Mixed-American | Recorded Depression | Discontinuation vs Continuation | Insomnia | Insomnia | 0.891 | 0.712 | 1.115 | 3.14e-01 | 1.00e+00 | 1805 | 1179 | 626 |
| Latino/Mixed-American | Recorded Depression | Switching vs Continuation | Insomnia | (Intercept) | 0.661 | 0.541 | 0.808 | 5.10e-05 | 7.15e-04 | 2309 | 1179 | 1130 |
| Latino/Mixed-American | Recorded Depression | Switching vs Continuation | Insomnia | age_at_first_AD | 0.800 | 0.727 | 0.881 | 5.93e-06 | 8.31e-05 | 2309 | 1179 | 1130 |
| Latino/Mixed-American | Recorded Depression | Switching vs Continuation | Insomnia | sex_at_birthMale | 0.934 | 0.769 | 1.134 | 4.92e-01 | 1.00e+00 | 2309 | 1179 | 1130 |
| Latino/Mixed-American | Recorded Depression | Switching vs Continuation | Insomnia | Income>200k | 0.558 | 0.290 | 1.075 | 8.10e-02 | 1.00e+00 | 2309 | 1179 | 1130 |
| Latino/Mixed-American | Recorded Depression | Switching vs Continuation | Insomnia | Income100-150k | 0.778 | 0.534 | 1.134 | 1.91e-01 | 1.00e+00 | 2309 | 1179 | 1130 |
| Latino/Mixed-American | Recorded Depression | Switching vs Continuation | Insomnia | Income150-200k | 1.334 | 0.708 | 2.513 | 3.72e-01 | 1.00e+00 | 2309 | 1179 | 1130 |
| Latino/Mixed-American | Recorded Depression | Switching vs Continuation | Insomnia | Income25-35k | 0.938 | 0.713 | 1.234 | 6.46e-01 | 1.00e+00 | 2309 | 1179 | 1130 |
| Latino/Mixed-American | Recorded Depression | Switching vs Continuation | Insomnia | Income35-50k | 1.197 | 0.893 | 1.606 | 2.30e-01 | 1.00e+00 | 2309 | 1179 | 1130 |
| Latino/Mixed-American | Recorded Depression | Switching vs Continuation | Insomnia | Income50-75k | 0.865 | 0.644 | 1.162 | 3.35e-01 | 1.00e+00 | 2309 | 1179 | 1130 |
| Latino/Mixed-American | Recorded Depression | Switching vs Continuation | Insomnia | Income75-100k | 1.051 | 0.707 | 1.562 | 8.06e-01 | 1.00e+00 | 2309 | 1179 | 1130 |
| Latino/Mixed-American | Recorded Depression | Switching vs Continuation | Insomnia | Education_levelCollege Postgraduate | 1.170 | 0.804 | 1.703 | 4.12e-01 | 1.00e+00 | 2309 | 1179 | 1130 |
| Latino/Mixed-American | Recorded Depression | Switching vs Continuation | Insomnia | Education_levelCollege Undergraduate | 1.051 | 0.821 | 1.345 | 6.95e-01 | 1.00e+00 | 2309 | 1179 | 1130 |
| Latino/Mixed-American | Recorded Depression | Switching vs Continuation | Insomnia | Education_levelTwelve or GED | 0.802 | 0.616 | 1.045 | 1.02e-01 | 1.00e+00 | 2309 | 1179 | 1130 |
| Latino/Mixed-American | Recorded Depression | Switching vs Continuation | Insomnia | Insomnia | 2.527 | 2.118 | 3.015 | <2.2e-16 | <2.2e-16 | 2309 | 1179 | 1130 |
| Latino/Mixed-American | Recorded Depression | Switching vs Continuation | MDD CIDI-sf | (Intercept) | 1.047 | 0.154 | 7.137 | 9.63e-01 | 1.00e+00 | 176 | 78 | 98 |
| Latino/Mixed-American | Recorded Depression | Switching vs Continuation | MDD CIDI-sf | age_at_first_AD | 0.883 | 0.597 | 1.305 | 5.33e-01 | 1.00e+00 | 176 | 78 | 98 |
| Latino/Mixed-American | Recorded Depression | Switching vs Continuation | MDD CIDI-sf | sex_at_birthMale | 0.889 | 0.432 | 1.826 | 7.48e-01 | 1.00e+00 | 176 | 78 | 98 |
| Latino/Mixed-American | Recorded Depression | Switching vs Continuation | MDD CIDI-sf | Income>200k | 0.927 | 0.155 | 5.554 | 9.34e-01 | 1.00e+00 | 176 | 78 | 98 |
| Latino/Mixed-American | Recorded Depression | Switching vs Continuation | MDD CIDI-sf | Income100-150k | 0.765 | 0.252 | 2.317 | 6.35e-01 | 1.00e+00 | 176 | 78 | 98 |
| Latino/Mixed-American | Recorded Depression | Switching vs Continuation | MDD CIDI-sf | Income150-200k | 1.559 | 0.325 | 7.469 | 5.79e-01 | 1.00e+00 | 176 | 78 | 98 |
| Latino/Mixed-American | Recorded Depression | Switching vs Continuation | MDD CIDI-sf | Income25-35k | 0.654 | 0.225 | 1.898 | 4.35e-01 | 1.00e+00 | 176 | 78 | 98 |
| Latino/Mixed-American | Recorded Depression | Switching vs Continuation | MDD CIDI-sf | Income35-50k | 0.678 | 0.190 | 2.413 | 5.48e-01 | 1.00e+00 | 176 | 78 | 98 |
| Latino/Mixed-American | Recorded Depression | Switching vs Continuation | MDD CIDI-sf | Income50-75k | 1.048 | 0.418 | 2.626 | 9.21e-01 | 1.00e+00 | 176 | 78 | 98 |
| Latino/Mixed-American | Recorded Depression | Switching vs Continuation | MDD CIDI-sf | Income75-100k | 1.008 | 0.327 | 3.107 | 9.89e-01 | 1.00e+00 | 176 | 78 | 98 |
| Latino/Mixed-American | Recorded Depression | Switching vs Continuation | MDD CIDI-sf | Education_levelCollege Postgraduate | 0.491 | 0.062 | 3.898 | 5.01e-01 | 1.00e+00 | 176 | 78 | 98 |
| Latino/Mixed-American | Recorded Depression | Switching vs Continuation | MDD CIDI-sf | Education_levelCollege Undergraduate | 0.803 | 0.114 | 5.650 | 8.25e-01 | 1.00e+00 | 176 | 78 | 98 |
| Latino/Mixed-American | Recorded Depression | Switching vs Continuation | MDD CIDI-sf | Education_levelTwelve or GED | 0.587 | 0.075 | 4.565 | 6.10e-01 | 1.00e+00 | 176 | 78 | 98 |
| Latino/Mixed-American | Recorded Depression | Switching vs Continuation | MDD CIDI-sf | MDD CIDI-sf | 2.133 | 0.933 | 4.877 | 7.27e-02 | 1.00e+00 | 176 | 78 | 98 |
| Latino/Mixed-American | Recorded Depression | Switching vs Continuation | MDD_symptom_count | (Intercept) | 1.211 | 0.260 | 5.634 | 8.07e-01 | 1.00e+00 | 203 | 93 | 110 |
| Latino/Mixed-American | Recorded Depression | Switching vs Continuation | MDD_symptom_count | age_at_first_AD | 0.832 | 0.585 | 1.185 | 3.08e-01 | 1.00e+00 | 203 | 93 | 110 |
| Latino/Mixed-American | Recorded Depression | Switching vs Continuation | MDD_symptom_count | sex_at_birthMale | 0.788 | 0.404 | 1.539 | 4.86e-01 | 1.00e+00 | 203 | 93 | 110 |
| Latino/Mixed-American | Recorded Depression | Switching vs Continuation | MDD_symptom_count | Income>200k | 0.754 | 0.170 | 3.344 | 7.10e-01 | 1.00e+00 | 203 | 93 | 110 |
| Latino/Mixed-American | Recorded Depression | Switching vs Continuation | MDD_symptom_count | Income100-150k | 0.849 | 0.303 | 2.373 | 7.54e-01 | 1.00e+00 | 203 | 93 | 110 |

| | | | | | | | | | | | | |
|---|---|---|---|---|---|---|---|---|---|---|---|---|
| Latino/Mixed-American | Recorded Depression | Switching vs Continuation | MDD_symptom_count | Income150-200k | 2.102 | 0.455 | 9.708 | 3.41e-01 | 1.00e+00 | 203 | 93 | 110 |
| Latino/Mixed-American | Recorded Depression | Switching vs Continuation | MDD_symptom_count | Income25-35k | 0.740 | 0.266 | 2.061 | 5.64e-01 | 1.00e+00 | 203 | 93 | 110 |
| Latino/Mixed-American | Recorded Depression | Switching vs Continuation | MDD_symptom_count | Income35-50k | 0.539 | 0.176 | 1.650 | 2.79e-01 | 1.00e+00 | 203 | 93 | 110 |
| Latino/Mixed-American | Recorded Depression | Switching vs Continuation | MDD_symptom_count | Income50-75k | 1.034 | 0.436 | 2.449 | 9.40e-01 | 1.00e+00 | 203 | 93 | 110 |
| Latino/Mixed-American | Recorded Depression | Switching vs Continuation | MDD_symptom_count | Income75-100k | 1.088 | 0.375 | 3.155 | 8.77e-01 | 1.00e+00 | 203 | 93 | 110 |
| Latino/Mixed-American | Recorded Depression | Switching vs Continuation | MDD_symptom_count | Education_levelCollege Postgraduate | 0.687 | 0.119 | 3.977 | 6.75e-01 | 1.00e+00 | 203 | 93 | 110 |
| Latino/Mixed-American | Recorded Depression | Switching vs Continuation | MDD_symptom_count | Education_levelCollege Undergraduate | 1.158 | 0.227 | 5.904 | 8.60e-01 | 1.00e+00 | 203 | 93 | 110 |
| Latino/Mixed-American | Recorded Depression | Switching vs Continuation | MDD_symptom_count | Education_levelTwelve or GED | 0.773 | 0.136 | 4.386 | 7.71e-01 | 1.00e+00 | 203 | 93 | 110 |
| Latino/Mixed-American | Recorded Depression | Switching vs Continuation | MDD_symptom_count | MDD_symptom_count | 1.196 | 0.835 | 1.714 | 3.29e-01 | 1.00e+00 | 203 | 93 | 110 |
| Latino/Mixed-American | Recorded Depression | Discontinuation vs Continuation | Migraine | (Intercept) | 0.544 | 0.429 | 0.689 | 4.54e-07 | 6.35e-06 | 1805 | 1179 | 626 |
| Latino/Mixed-American | Recorded Depression | Discontinuation vs Continuation | Migraine | age_at_first_AD | 0.958 | 0.860 | 1.066 | 4.27e-01 | 1.00e+00 | 1805 | 1179 | 626 |
| Latino/Mixed-American | Recorded Depression | Discontinuation vs Continuation | Migraine | sex_at_birthMale | 1.110 | 0.893 | 1.381 | 3.48e-01 | 1.00e+00 | 1805 | 1179 | 626 |
| Latino/Mixed-American | Recorded Depression | Discontinuation vs Continuation | Migraine | Income>200k | 0.602 | 0.260 | 1.391 | 2.35e-01 | 1.00e+00 | 1805 | 1179 | 626 |
| Latino/Mixed-American | Recorded Depression | Discontinuation vs Continuation | Migraine | Income100-150k | 0.611 | 0.382 | 0.978 | 4.02e-02 | 5.63e-01 | 1805 | 1179 | 626 |
| Latino/Mixed-American | Recorded Depression | Discontinuation vs Continuation | Migraine | Income150-200k | 1.068 | 0.491 | 2.322 | 8.68e-01 | 1.00e+00 | 1805 | 1179 | 626 |
| Latino/Mixed-American | Recorded Depression | Discontinuation vs Continuation | Migraine | Income25-35k | 0.924 | 0.675 | 1.264 | 6.19e-01 | 1.00e+00 | 1805 | 1179 | 626 |
| Latino/Mixed-American | Recorded Depression | Discontinuation vs Continuation | Migraine | Income35-50k | 1.104 | 0.789 | 1.545 | 5.65e-01 | 1.00e+00 | 1805 | 1179 | 626 |
| Latino/Mixed-American | Recorded Depression | Discontinuation vs Continuation | Migraine | Income50-75k | 0.818 | 0.578 | 1.158 | 2.58e-01 | 1.00e+00 | 1805 | 1179 | 626 |
| Latino/Mixed-American | Recorded Depression | Discontinuation vs Continuation | Migraine | Income75-100k | 0.937 | 0.587 | 1.495 | 7.84e-01 | 1.00e+00 | 1805 | 1179 | 626 |
| Latino/Mixed-American | Recorded Depression | Discontinuation vs Continuation | Migraine | Education_levelCollege Postgraduate | 0.749 | 0.456 | 1.229 | 2.52e-01 | 1.00e+00 | 1805 | 1179 | 626 |
| Latino/Mixed-American | Recorded Depression | Discontinuation vs Continuation | Migraine | Education_levelCollege Undergraduate | 1.250 | 0.942 | 1.659 | 1.23e-01 | 1.00e+00 | 1805 | 1179 | 626 |
| Latino/Mixed-American | Recorded Depression | Discontinuation vs Continuation | Migraine | Education_levelTwelve or GED | 0.992 | 0.733 | 1.341 | 9.56e-01 | 1.00e+00 | 1805 | 1179 | 626 |
| Latino/Mixed-American | Recorded Depression | Discontinuation vs Continuation | Migraine | Migraine | 0.739 | 0.595 | 0.918 | 6.38e-03 | 8.93e-02 | 1805 | 1179 | 626 |
| Latino/Mixed-American | Recorded Depression | Switching vs Continuation | Migraine | (Intercept) | 0.688 | 0.562 | 0.843 | 3.08e-04 | 4.31e-03 | 2309 | 1179 | 1130 |
| Latino/Mixed-American | Recorded Depression | Switching vs Continuation | Migraine | age_at_first_AD | 0.881 | 0.801 | 0.969 | 8.83e-03 | 1.24e-01 | 2309 | 1179 | 1130 |
| Latino/Mixed-American | Recorded Depression | Switching vs Continuation | Migraine | sex_at_birthMale | 1.032 | 0.852 | 1.251 | 7.46e-01 | 1.00e+00 | 2309 | 1179 | 1130 |
| Latino/Mixed-American | Recorded Depression | Switching vs Continuation | Migraine | Income>200k | 0.593 | 0.310 | 1.134 | 1.14e-01 | 1.00e+00 | 2309 | 1179 | 1130 |
| Latino/Mixed-American | Recorded Depression | Switching vs Continuation | Migraine | Income100-150k | 0.735 | 0.507 | 1.066 | 1.05e-01 | 1.00e+00 | 2309 | 1179 | 1130 |
| Latino/Mixed-American | Recorded Depression | Switching vs Continuation | Migraine | Income150-200k | 1.348 | 0.720 | 2.525 | 3.51e-01 | 1.00e+00 | 2309 | 1179 | 1130 |
| Latino/Mixed-American | Recorded Depression | Switching vs Continuation | Migraine | Income25-35k | 0.934 | 0.712 | 1.225 | 6.21e-01 | 1.00e+00 | 2309 | 1179 | 1130 |
| Latino/Mixed-American | Recorded Depression | Switching vs Continuation | Migraine | Income35-50k | 1.219 | 0.912 | 1.630 | 1.81e-01 | 1.00e+00 | 2309 | 1179 | 1130 |
| Latino/Mixed-American | Recorded Depression | Switching vs Continuation | Migraine | Income50-75k | 0.858 | 0.640 | 1.149 | 3.04e-01 | 1.00e+00 | 2309 | 1179 | 1130 |
| Latino/Mixed-American | Recorded Depression | Switching vs Continuation | Migraine | Income75-100k | 1.079 | 0.730 | 1.596 | 7.03e-01 | 1.00e+00 | 2309 | 1179 | 1130 |
| Latino/Mixed-American | Recorded Depression | Switching vs Continuation | Migraine | Education_levelCollege Postgraduate | 1.261 | 0.870 | 1.829 | 2.21e-01 | 1.00e+00 | 2309 | 1179 | 1130 |
| Latino/Mixed-American | Recorded Depression | Switching vs Continuation | Migraine | Education_levelCollege Undergraduate | 1.115 | 0.874 | 1.424 | 3.81e-01 | 1.00e+00 | 2309 | 1179 | 1130 |
| Latino/Mixed-American | Recorded Depression | Switching vs Continuation | Migraine | Education_levelTwelve or GED | 0.829 | 0.639 | 1.076 | 1.58e-01 | 1.00e+00 | 2309 | 1179 | 1130 |
| Latino/Mixed-American | Recorded Depression | Switching vs Continuation | Migraine | Migraine | 1.935 | 1.629 | 2.297 | 4.88e-14 | 6.83e-13 | 2309 | 1179 | 1130 |
| Latino/Mixed-American | Recorded Depression | Discontinuation vs Continuation | Recorded Anxiety | (Intercept) | 0.444 | 0.333 | 0.592 | 3.16e-08 | 4.43e-07 | 1805 | 1179 | 626 |
| Latino/Mixed-American | Recorded Depression | Discontinuation vs Continuation | Recorded Anxiety | age_at_first_AD | 0.977 | 0.877 | 1.088 | 6.67e-01 | 1.00e+00 | 1805 | 1179 | 626 |
| Latino/Mixed-American | Recorded Depression | Discontinuation vs Continuation | Recorded Anxiety | sex_at_birthMale | 1.148 | 0.923 | 1.427 | 2.15e-01 | 1.00e+00 | 1805 | 1179 | 626 |
| Latino/Mixed-American | Recorded Depression | Discontinuation vs Continuation | Recorded Anxiety | Income>200k | 0.582 | 0.252 | 1.342 | 2.04e-01 | 1.00e+00 | 1805 | 1179 | 626 |
| Latino/Mixed-American | Recorded Depression | Discontinuation vs Continuation | Recorded Anxiety | Income100-150k | 0.613 | 0.383 | 0.980 | 4.11e-02 | 5.75e-01 | 1805 | 1179 | 626 |
| Latino/Mixed-American | Recorded Depression | Discontinuation vs Continuation | Recorded Anxiety | Income150-200k | 1.100 | 0.506 | 2.390 | 8.10e-01 | 1.00e+00 | 1805 | 1179 | 626 |
| Latino/Mixed-American | Recorded Depression | Discontinuation vs Continuation | Recorded Anxiety | Income25-35k | 0.923 | 0.675 | 1.263 | 6.17e-01 | 1.00e+00 | 1805 | 1179 | 626 |
| Latino/Mixed-American | Recorded Depression | Discontinuation vs Continuation | Recorded Anxiety | Income35-50k | 1.114 | 0.797 | 1.558 | 5.27e-01 | 1.00e+00 | 1805 | 1179 | 626 |
| Latino/Mixed-American | Recorded Depression | Discontinuation vs Continuation | Recorded Anxiety | Income50-75k | 0.807 | 0.570 | 1.142 | 2.27e-01 | 1.00e+00 | 1805 | 1179 | 626 |
| Latino/Mixed-American | Recorded Depression | Discontinuation vs Continuation | Recorded Anxiety | Income75-100k | 0.935 | 0.586 | 1.491 | 7.77e-01 | 1.00e+00 | 1805 | 1179 | 626 |
| Latino/Mixed-American | Recorded Depression | Discontinuation vs Continuation | Recorded Anxiety | Education_levelCollege Postgraduate | 0.783 | 0.478 | 1.284 | 3.33e-01 | 1.00e+00 | 1805 | 1179 | 626 |
| Latino/Mixed-American | Recorded Depression | Discontinuation vs Continuation | Recorded Anxiety | Education_levelCollege Undergraduate | 1.264 | 0.953 | 1.677 | 1.04e-01 | 1.00e+00 | 1805 | 1179 | 626 |
| Latino/Mixed-American | Recorded Depression | Discontinuation vs Continuation | Recorded Anxiety | Education_levelTwelve or GED | 0.990 | 0.733 | 1.338 | 9.49e-01 | 1.00e+00 | 1805 | 1179 | 626 |
| Latino/Mixed-American | Recorded Depression | Discontinuation vs Continuation | Recorded Anxiety | Recorded Anxiety | 1.142 | 0.904 | 1.443 | 2.67e-01 | 1.00e+00 | 1805 | 1179 | 626 |
| Latino/Mixed-American | Recorded Depression | Switching vs Continuation | Recorded Anxiety | (Intercept) | 0.341 | 0.258 | 0.452 | 7.37e-14 | 1.03e-12 | 2309 | 1179 | 1130 |
| Latino/Mixed-American | Recorded Depression | Switching vs Continuation | Recorded Anxiety | age_at_first_AD | 0.902 | 0.820 | 0.993 | 3.56e-02 | 4.98e-01 | 2309 | 1179 | 1130 |
| Latino/Mixed-American | Recorded Depression | Switching vs Continuation | Recorded Anxiety | sex_at_birthMale | 1.002 | 0.825 | 1.217 | 9.81e-01 | 1.00e+00 | 2309 | 1179 | 1130 |
| Latino/Mixed-American | Recorded Depression | Switching vs Continuation | Recorded Anxiety | Income>200k | 0.570 | 0.295 | 1.098 | 9.30e-02 | 1.00e+00 | 2309 | 1179 | 1130 |
| Latino/Mixed-American | Recorded Depression | Switching vs Continuation | Recorded Anxiety | Income100-150k | 0.796 | 0.545 | 1.162 | 2.37e-01 | 1.00e+00 | 2309 | 1179 | 1130 |
| Latino/Mixed-American | Recorded Depression | Switching vs Continuation | Recorded Anxiety | Income150-200k | 1.287 | 0.679 | 2.441 | 4.39e-01 | 1.00e+00 | 2309 | 1179 | 1130 |
| Latino/Mixed-American | Recorded Depression | Switching vs Continuation | Recorded Anxiety | Income25-35k | 0.944 | 0.718 | 1.241 | 6.80e-01 | 1.00e+00 | 2309 | 1179 | 1130 |
| Latino/Mixed-American | Recorded Depression | Switching vs Continuation | Recorded Anxiety | Income35-50k | 1.207 | 0.901 | 1.618 | 2.07e-01 | 1.00e+00 | 2309 | 1179 | 1130 |
| Latino/Mixed-American | Recorded Depression | Switching vs Continuation | Recorded Anxiety | Income50-75k | 0.851 | 0.634 | 1.143 | 2.84e-01 | 1.00e+00 | 2309 | 1179 | 1130 |
| Latino/Mixed-American | Recorded Depression | Switching vs Continuation | Recorded Anxiety | Income75-100k | 1.070 | 0.721 | 1.587 | 7.38e-01 | 1.00e+00 | 2309 | 1179 | 1130 |
| Latino/Mixed-American | Recorded Depression | Switching vs Continuation | Recorded Anxiety | Education_levelCollege Postgraduate | 1.217 | 0.835 | 1.774 | 3.06e-01 | 1.00e+00 | 2309 | 1179 | 1130 |
| Latino/Mixed-American | Recorded Depression | Switching vs Continuation | Recorded Anxiety | Education_levelCollege Undergraduate | 1.068 | 0.834 | 1.368 | 6.01e-01 | 1.00e+00 | 2309 | 1179 | 1130 |
| Latino/Mixed-American | Recorded Depression | Switching vs Continuation | Recorded Anxiety | Education_levelTwelve or GED | 0.791 | 0.608 | 1.029 | 8.11e-02 | 1.00e+00 | 2309 | 1179 | 1130 |
| Latino/Mixed-American | Recorded Depression | Switching vs Continuation | Recorded Anxiety | Recorded Anxiety | 3.369 | 2.633 | 4.311 | <2.2e-16 | <2.2e-16 | 2309 | 1179 | 1130 |
| Latino/Mixed-American | Recorded Depression | Discontinuation vs Continuation | Smoking_Frequency | (Intercept) | 0.419 | 0.328 | 0.536 | 3.59e-12 | 5.03e-11 | 1753 | 1143 | 610 |

| | | | | | | | | | | | | |
|---|---|---|---|---|---|---|---|---|---|---|---|---|
| Latino/Mixed-American | Recorded Depression | Discontinuation vs Continuation | Smoking_Frequency | age_at_first_AD | 0.959 | 0.859 | 1.071 | 4.61e-01 | 1.00e+00 | 1753 | 1143 | 610 |
| Latino/Mixed-American | Recorded Depression | Discontinuation vs Continuation | Smoking_Frequency | sex_at_birthMale | 1.068 | 0.853 | 1.336 | 5.67e-01 | 1.00e+00 | 1753 | 1143 | 610 |
| Latino/Mixed-American | Recorded Depression | Discontinuation vs Continuation | Smoking_Frequency | Income>200k | 0.617 | 0.266 | 1.431 | 2.60e-01 | 1.00e+00 | 1753 | 1143 | 610 |
| Latino/Mixed-American | Recorded Depression | Discontinuation vs Continuation | Smoking_Frequency | Income100-150k | 0.622 | 0.385 | 1.007 | 5.32e-02 | 7.44e-01 | 1753 | 1143 | 610 |
| Latino/Mixed-American | Recorded Depression | Discontinuation vs Continuation | Smoking_Frequency | Income150-200k | 1.105 | 0.507 | 2.409 | 8.02e-01 | 1.00e+00 | 1753 | 1143 | 610 |
| Latino/Mixed-American | Recorded Depression | Discontinuation vs Continuation | Smoking_Frequency | Income25-35k | 0.928 | 0.675 | 1.276 | 6.46e-01 | 1.00e+00 | 1753 | 1143 | 610 |
| Latino/Mixed-American | Recorded Depression | Discontinuation vs Continuation | Smoking_Frequency | Income35-50k | 1.091 | 0.775 | 1.537 | 6.18e-01 | 1.00e+00 | 1753 | 1143 | 610 |
| Latino/Mixed-American | Recorded Depression | Discontinuation vs Continuation | Smoking_Frequency | Income50-75k | 0.825 | 0.581 | 1.173 | 2.85e-01 | 1.00e+00 | 1753 | 1143 | 610 |
| Latino/Mixed-American | Recorded Depression | Discontinuation vs Continuation | Smoking_Frequency | Income75-100k | 0.906 | 0.564 | 1.455 | 6.83e-01 | 1.00e+00 | 1753 | 1143 | 610 |
| Latino/Mixed-American | Recorded Depression | Discontinuation vs Continuation | Smoking_Frequency | Education_levelCollege Postgraduate | 0.883 | 0.535 | 1.457 | 6.26e-01 | 1.00e+00 | 1753 | 1143 | 610 |
| Latino/Mixed-American | Recorded Depression | Discontinuation vs Continuation | Smoking_Frequency | Education_levelCollege Undergraduate | 1.323 | 0.993 | 1.763 | 5.58e-02 | 7.82e-01 | 1753 | 1143 | 610 |
| Latino/Mixed-American | Recorded Depression | Discontinuation vs Continuation | Smoking_Frequency | Education_levelTwelve or GED | 1.030 | 0.757 | 1.401 | 8.51e-01 | 1.00e+00 | 1753 | 1143 | 610 |
| Latino/Mixed-American | Recorded Depression | Discontinuation vs Continuation | Smoking_Frequency | Smoking_FrequencyEvery Day | 1.589 | 1.115 | 2.265 | 1.04e-02 | 1.46e-01 | 1753 | 1143 | 610 |
| Latino/Mixed-American | Recorded Depression | Discontinuation vs Continuation | Smoking_Frequency | Smoking_FrequencyNot At All (former) | 1.326 | 1.032 | 1.703 | 2.71e-02 | 3.80e-01 | 1753 | 1143 | 610 |
| Latino/Mixed-American | Recorded Depression | Discontinuation vs Continuation | Smoking_Frequency | Smoking_FrequencySome Days | 1.721 | 1.135 | 2.610 | 1.05e-02 | 1.48e-01 | 1753 | 1143 | 610 |
| Latino/Mixed-American | Recorded Depression | Switching vs Continuation | Smoking_Frequency | (Intercept) | 0.848 | 0.695 | 1.035 | 1.05e-01 | 1.00e+00 | 2239 | 1143 | 1096 |
| Latino/Mixed-American | Recorded Depression | Switching vs Continuation | Smoking_Frequency | age_at_first_AD | 0.843 | 0.765 | 0.928 | 4.78e-04 | 6.69e-03 | 2239 | 1143 | 1096 |
| Latino/Mixed-American | Recorded Depression | Switching vs Continuation | Smoking_Frequency | sex_at_birthMale | 0.889 | 0.731 | 1.081 | 2.38e-01 | 1.00e+00 | 2239 | 1143 | 1096 |
| Latino/Mixed-American | Recorded Depression | Switching vs Continuation | Smoking_Frequency | Income>200k | 0.558 | 0.289 | 1.080 | 8.34e-02 | 1.00e+00 | 2239 | 1143 | 1096 |
| Latino/Mixed-American | Recorded Depression | Switching vs Continuation | Smoking_Frequency | Income100-150k | 0.758 | 0.521 | 1.103 | 1.48e-01 | 1.00e+00 | 2239 | 1143 | 1096 |
| Latino/Mixed-American | Recorded Depression | Switching vs Continuation | Smoking_Frequency | Income150-200k | 1.242 | 0.664 | 2.324 | 4.98e-01 | 1.00e+00 | 2239 | 1143 | 1096 |
| Latino/Mixed-American | Recorded Depression | Switching vs Continuation | Smoking_Frequency | Income25-35k | 0.939 | 0.715 | 1.232 | 6.48e-01 | 1.00e+00 | 2239 | 1143 | 1096 |
| Latino/Mixed-American | Recorded Depression | Switching vs Continuation | Smoking_Frequency | Income35-50k | 1.179 | 0.880 | 1.581 | 2.71e-01 | 1.00e+00 | 2239 | 1143 | 1096 |
| Latino/Mixed-American | Recorded Depression | Switching vs Continuation | Smoking_Frequency | Income50-75k | 0.826 | 0.615 | 1.109 | 2.03e-01 | 1.00e+00 | 2239 | 1143 | 1096 |
| Latino/Mixed-American | Recorded Depression | Switching vs Continuation | Smoking_Frequency | Income75-100k | 0.980 | 0.661 | 1.455 | 9.21e-01 | 1.00e+00 | 2239 | 1143 | 1096 |
| Latino/Mixed-American | Recorded Depression | Switching vs Continuation | Smoking_Frequency | Education_levelCollege Postgraduate | 1.273 | 0.875 | 1.852 | 2.06e-01 | 1.00e+00 | 2239 | 1143 | 1096 |
| Latino/Mixed-American | Recorded Depression | Switching vs Continuation | Smoking_Frequency | Education_levelCollege Undergraduate | 1.142 | 0.894 | 1.458 | 2.88e-01 | 1.00e+00 | 2239 | 1143 | 1096 |
| Latino/Mixed-American | Recorded Depression | Switching vs Continuation | Smoking_Frequency | Education_levelTwelve or GED | 0.857 | 0.660 | 1.113 | 2.47e-01 | 1.00e+00 | 2239 | 1143 | 1096 |
| Latino/Mixed-American | Recorded Depression | Switching vs Continuation | Smoking_Frequency | Smoking_FrequencyEvery Day | 1.501 | 1.102 | 2.044 | 9.92e-03 | 1.39e-01 | 2239 | 1143 | 1096 |
| Latino/Mixed-American | Recorded Depression | Switching vs Continuation | Smoking_Frequency | Smoking_FrequencyNot At All (former) | 1.224 | 0.989 | 1.515 | 6.31e-02 | 8.83e-01 | 2239 | 1143 | 1096 |
| Latino/Mixed-American | Recorded Depression | Switching vs Continuation | Smoking_Frequency | Smoking_FrequencySome Days | 1.051 | 0.706 | 1.564 | 8.07e-01 | 1.00e+00 | 2239 | 1143 | 1096 |
| Latino/Mixed-American | Recorded Depression | Discontinuation vs Continuation | Stress | (Intercept) | 0.207 | 0.105 | 0.410 | 6.33e-06 | 8.86e-05 | 542 | 395 | 147 |
| Latino/Mixed-American | Recorded Depression | Discontinuation vs Continuation | Stress | age_at_first_AD | 0.837 | 0.673 | 1.040 | 1.09e-01 | 1.00e+00 | 542 | 395 | 147 |
| Latino/Mixed-American | Recorded Depression | Discontinuation vs Continuation | Stress | sex_at_birthMale | 0.857 | 0.547 | 1.343 | 5.00e-01 | 1.00e+00 | 542 | 395 | 147 |
| Latino/Mixed-American | Recorded Depression | Discontinuation vs Continuation | Stress | Income>200k | 0.553 | 0.147 | 2.080 | 3.81e-01 | 1.00e+00 | 542 | 395 | 147 |
| Latino/Mixed-American | Recorded Depression | Discontinuation vs Continuation | Stress | Income100-150k | 0.762 | 0.360 | 1.615 | 4.78e-01 | 1.00e+00 | 542 | 395 | 147 |
| Latino/Mixed-American | Recorded Depression | Discontinuation vs Continuation | Stress | Income150-200k | 1.384 | 0.434 | 4.411 | 5.83e-01 | 1.00e+00 | 542 | 395 | 147 |
| Latino/Mixed-American | Recorded Depression | Discontinuation vs Continuation | Stress | Income25-35k | 1.209 | 0.645 | 2.268 | 5.54e-01 | 1.00e+00 | 542 | 395 | 147 |
| Latino/Mixed-American | Recorded Depression | Discontinuation vs Continuation | Stress | Income35-50k | 1.867 | 0.982 | 3.550 | 5.67e-02 | 7.94e-01 | 542 | 395 | 147 |
| Latino/Mixed-American | Recorded Depression | Discontinuation vs Continuation | Stress | Income50-75k | 0.928 | 0.495 | 1.739 | 8.15e-01 | 1.00e+00 | 542 | 395 | 147 |
| Latino/Mixed-American | Recorded Depression | Discontinuation vs Continuation | Stress | Income75-100k | 1.036 | 0.465 | 2.304 | 9.32e-01 | 1.00e+00 | 542 | 395 | 147 |
| Latino/Mixed-American | Recorded Depression | Discontinuation vs Continuation | Stress | Education_levelCollege Postgraduate | 1.272 | 0.491 | 3.299 | 6.20e-01 | 1.00e+00 | 542 | 395 | 147 |
| Latino/Mixed-American | Recorded Depression | Discontinuation vs Continuation | Stress | Education_levelCollege Undergraduate | 1.693 | 0.793 | 3.612 | 1.73e-01 | 1.00e+00 | 542 | 395 | 147 |
| Latino/Mixed-American | Recorded Depression | Discontinuation vs Continuation | Stress | Education_levelTwelve or GED | 1.693 | 0.759 | 3.780 | 1.98e-01 | 1.00e+00 | 542 | 395 | 147 |
| Latino/Mixed-American | Recorded Depression | Discontinuation vs Continuation | Stress | StressAlmost Never | 1.661 | 0.729 | 3.782 | 2.27e-01 | 1.00e+00 | 542 | 395 | 147 |
| Latino/Mixed-American | Recorded Depression | Discontinuation vs Continuation | Stress | StressFairly Often | 1.348 | 0.627 | 2.898 | 4.45e-01 | 1.00e+00 | 542 | 395 | 147 |
| Latino/Mixed-American | Recorded Depression | Discontinuation vs Continuation | Stress | StressSometimes | 1.057 | 0.683 | 1.638 | 8.03e-01 | 1.00e+00 | 542 | 395 | 147 |
| Latino/Mixed-American | Recorded Depression | Discontinuation vs Continuation | Stress | StressVery Often | 0.707 | 0.218 | 2.289 | 5.63e-01 | 1.00e+00 | 542 | 395 | 147 |
| Latino/Mixed-American | Recorded Depression | Switching vs Continuation | Stress | (Intercept) | 0.983 | 0.652 | 1.483 | 9.36e-01 | 1.00e+00 | 867 | 395 | 472 |
| Latino/Mixed-American | Recorded Depression | Switching vs Continuation | Stress | age_at_first_AD | 0.797 | 0.679 | 0.935 | 5.31e-03 | 7.43e-02 | 867 | 395 | 472 |
| Latino/Mixed-American | Recorded Depression | Switching vs Continuation | Stress | sex_at_birthMale | 1.055 | 0.774 | 1.440 | 7.33e-01 | 1.00e+00 | 867 | 395 | 472 |
| Latino/Mixed-American | Recorded Depression | Switching vs Continuation | Stress | Income>200k | 0.614 | 0.265 | 1.421 | 2.55e-01 | 1.00e+00 | 867 | 395 | 472 |
| Latino/Mixed-American | Recorded Depression | Switching vs Continuation | Stress | Income100-150k | 0.705 | 0.417 | 1.191 | 1.91e-01 | 1.00e+00 | 867 | 395 | 472 |
| Latino/Mixed-American | Recorded Depression | Switching vs Continuation | Stress | Income150-200k | 1.361 | 0.576 | 3.219 | 4.83e-01 | 1.00e+00 | 867 | 395 | 472 |
| Latino/Mixed-American | Recorded Depression | Switching vs Continuation | Stress | Income25-35k | 1.222 | 0.778 | 1.919 | 3.85e-01 | 1.00e+00 | 867 | 395 | 472 |
| Latino/Mixed-American | Recorded Depression | Switching vs Continuation | Stress | Income35-50k | 1.819 | 1.111 | 2.980 | 1.75e-02 | 2.45e-01 | 867 | 395 | 472 |
| Latino/Mixed-American | Recorded Depression | Switching vs Continuation | Stress | Income50-75k | 0.980 | 0.631 | 1.522 | 9.29e-01 | 1.00e+00 | 867 | 395 | 472 |
| Latino/Mixed-American | Recorded Depression | Switching vs Continuation | Stress | Income75-100k | 1.314 | 0.756 | 2.286 | 3.33e-01 | 1.00e+00 | 867 | 395 | 472 |
| Latino/Mixed-American | Recorded Depression | Switching vs Continuation | Stress | Education_levelCollege Postgraduate | 0.998 | 0.545 | 1.828 | 9.94e-01 | 1.00e+00 | 867 | 395 | 472 |
| Latino/Mixed-American | Recorded Depression | Switching vs Continuation | Stress | Education_levelCollege Undergraduate | 0.996 | 0.618 | 1.605 | 9.88e-01 | 1.00e+00 | 867 | 395 | 472 |
| Latino/Mixed-American | Recorded Depression | Switching vs Continuation | Stress | Education_levelTwelve or GED | 0.779 | 0.458 | 1.323 | 3.55e-01 | 1.00e+00 | 867 | 395 | 472 |
| Latino/Mixed-American | Recorded Depression | Switching vs Continuation | Stress | StressAlmost Never | 1.043 | 0.541 | 2.012 | 9.00e-01 | 1.00e+00 | 867 | 395 | 472 |
| Latino/Mixed-American | Recorded Depression | Switching vs Continuation | Stress | StressFairly Often | 1.166 | 0.667 | 2.038 | 5.90e-01 | 1.00e+00 | 867 | 395 | 472 |
| Latino/Mixed-American | Recorded Depression | Switching vs Continuation | Stress | StressSometimes | 1.129 | 0.836 | 1.527 | 4.29e-01 | 1.00e+00 | 867 | 395 | 472 |
| Latino/Mixed-American | Recorded Depression | Switching vs Continuation | Stress | StressVery Often | 0.968 | 0.463 | 2.024 | 9.31e-01 | 1.00e+00 | 867 | 395 | 472 |
| Latino/Mixed-American | Recorded Depression | Discontinuation vs Continuation | age_at_first_AD | (Intercept) | 0.506 | 0.448 | 0.571 | <2.2e-16 | <2.2e-16 | 1805 | 1179 | 626 |

| | | | | | | | | | | | | |
|---|---|---|---|---|---|---|---|---|---|---|---|---|
| Latino/Mixed-American | Recorded Depression | Discontinuation vs Continuation | age_at_first_AD | age_at_first_AD | 0.954 | 0.862 | 1.057 | 3.70e-01 | 1.00e+00 | 1805 | 1179 | 626 |
| Latino/Mixed-American | Recorded Depression | Discontinuation vs Continuation | age_at_first_AD | sex_at_birthMale | 1.124 | 0.907 | 1.393 | 2.84e-01 | 1.00e+00 | 1805 | 1179 | 626 |
| Latino/Mixed-American | Recorded Depression | Switching vs Continuation | age_at_first_AD | (Intercept) | 0.913 | 0.824 | 1.012 | 8.26e-02 | 1.00e+00 | 2309 | 1179 | 1130 |
| Latino/Mixed-American | Recorded Depression | Switching vs Continuation | age_at_first_AD | age_at_first_AD | 0.848 | 0.775 | 0.927 | 2.90e-04 | 4.06e-03 | 2309 | 1179 | 1130 |
| Latino/Mixed-American | Recorded Depression | Switching vs Continuation | age_at_first_AD | sex_at_birthMale | 0.957 | 0.795 | 1.152 | 6.41e-01 | 1.00e+00 | 2309 | 1179 | 1130 |
| Latino/Mixed-American | Recorded Depression | Discontinuation vs Continuation | sex_at_birth | (Intercept) | 0.506 | 0.448 | 0.571 | <2.2e-16 | <2.2e-16 | 1805 | 1179 | 626 |
| Latino/Mixed-American | Recorded Depression | Discontinuation vs Continuation | sex_at_birth | age_at_first_AD | 0.954 | 0.862 | 1.057 | 3.70e-01 | 1.00e+00 | 1805 | 1179 | 626 |
| Latino/Mixed-American | Recorded Depression | Discontinuation vs Continuation | sex_at_birth | sex_at_birthMale | 1.124 | 0.907 | 1.393 | 2.84e-01 | 1.00e+00 | 1805 | 1179 | 626 |
| Latino/Mixed-American | Recorded Depression | Switching vs Continuation | sex_at_birth | (Intercept) | 0.913 | 0.824 | 1.012 | 8.26e-02 | 1.00e+00 | 2309 | 1179 | 1130 |
| Latino/Mixed-American | Recorded Depression | Switching vs Continuation | sex_at_birth | age_at_first_AD | 0.848 | 0.775 | 0.927 | 2.90e-04 | 4.06e-03 | 2309 | 1179 | 1130 |
| Latino/Mixed-American | Recorded Depression | Switching vs Continuation | sex_at_birth | sex_at_birthMale | 0.957 | 0.795 | 1.152 | 6.41e-01 | 1.00e+00 | 2309 | 1179 | 1130 |
| Latino/Mixed-American | No Recorded Depression | Discontinuation vs Continuation | Alcohol_Frequency | (Intercept) | 0.436 | 0.321 | 0.591 | 9.21e-08 | 1.29e-06 | 2008 | 1250 | 758 |
| Latino/Mixed-American | No Recorded Depression | Discontinuation vs Continuation | Alcohol_Frequency | age_at_first_AD | 0.986 | 0.894 | 1.088 | 7.81e-01 | 1.00e+00 | 2008 | 1250 | 758 |
| Latino/Mixed-American | No Recorded Depression | Discontinuation vs Continuation | Alcohol_Frequency | sex_at_birthMale | 1.289 | 1.049 | 1.583 | 1.56e-02 | 2.18e-01 | 2008 | 1250 | 758 |
| Latino/Mixed-American | No Recorded Depression | Discontinuation vs Continuation | Alcohol_Frequency | Income>200k | 0.602 | 0.339 | 1.067 | 8.24e-02 | 1.00e+00 | 2008 | 1250 | 758 |
| Latino/Mixed-American | No Recorded Depression | Discontinuation vs Continuation | Alcohol_Frequency | Income100-150k | 0.700 | 0.479 | 1.023 | 6.53e-02 | 9.14e-01 | 2008 | 1250 | 758 |
| Latino/Mixed-American | No Recorded Depression | Discontinuation vs Continuation | Alcohol_Frequency | Income150-200k | 0.569 | 0.340 | 0.952 | 3.16e-02 | 4.43e-01 | 2008 | 1250 | 758 |
| Latino/Mixed-American | No Recorded Depression | Discontinuation vs Continuation | Alcohol_Frequency | Income25-35k | 0.716 | 0.524 | 0.977 | 3.53e-02 | 4.95e-01 | 2008 | 1250 | 758 |
| Latino/Mixed-American | No Recorded Depression | Discontinuation vs Continuation | Alcohol_Frequency | Income35-50k | 0.988 | 0.722 | 1.353 | 9.42e-01 | 1.00e+00 | 2008 | 1250 | 758 |
| Latino/Mixed-American | No Recorded Depression | Discontinuation vs Continuation | Alcohol_Frequency | Income50-75k | 0.822 | 0.604 | 1.118 | 2.12e-01 | 1.00e+00 | 2008 | 1250 | 758 |
| Latino/Mixed-American | No Recorded Depression | Discontinuation vs Continuation | Alcohol_Frequency | Income75-100k | 0.624 | 0.413 | 0.945 | 2.57e-02 | 3.60e-01 | 2008 | 1250 | 758 |
| Latino/Mixed-American | No Recorded Depression | Discontinuation vs Continuation | Alcohol_Frequency | Education_levelCollege Postgraduate | 1.031 | 0.683 | 1.557 | 8.83e-01 | 1.00e+00 | 2008 | 1250 | 758 |
| Latino/Mixed-American | No Recorded Depression | Discontinuation vs Continuation | Alcohol_Frequency | Education_levelCollege Undergraduate | 1.399 | 1.048 | 1.867 | 2.28e-02 | 3.20e-01 | 2008 | 1250 | 758 |
| Latino/Mixed-American | No Recorded Depression | Discontinuation vs Continuation | Alcohol_Frequency | Education_levelTwelve or GED | 1.624 | 1.190 | 2.216 | 2.22e-03 | 3.11e-02 | 2008 | 1250 | 758 |
| Latino/Mixed-American | No Recorded Depression | Discontinuation vs Continuation | Alcohol_Frequency | Alcohol_Frequency2-3 Week | 0.987 | 0.641 | 1.521 | 9.53e-01 | 1.00e+00 | 2008 | 1250 | 758 |
| Latino/Mixed-American | No Recorded Depression | Discontinuation vs Continuation | Alcohol_Frequency | Alcohol_Frequency2-4 Month | 0.993 | 0.695 | 1.421 | 9.71e-01 | 1.00e+00 | 2008 | 1250 | 758 |
| Latino/Mixed-American | No Recorded Depression | Discontinuation vs Continuation | Alcohol_Frequency | Alcohol_Frequency4plus Week | 1.410 | 0.872 | 2.279 | 1.61e-01 | 1.00e+00 | 2008 | 1250 | 758 |
| Latino/Mixed-American | No Recorded Depression | Discontinuation vs Continuation | Alcohol_Frequency | Alcohol_FrequencyMonthly or Less | 1.255 | 0.919 | 1.714 | 1.54e-01 | 1.00e+00 | 2008 | 1250 | 758 |
| Latino/Mixed-American | No Recorded Depression | Discontinuation vs Continuation | Alcohol_Frequency | Alcohol_FrequencyNever (former) | 1.301 | 0.933 | 1.812 | 1.20e-01 | 1.00e+00 | 2008 | 1250 | 758 |
| Latino/Mixed-American | No Recorded Depression | Switching vs Continuation | Alcohol_Frequency | (Intercept) | 0.191 | 0.124 | 0.293 | 3.51e-14 | 4.91e-13 | 1587 | 1250 | 337 |
| Latino/Mixed-American | No Recorded Depression | Switching vs Continuation | Alcohol_Frequency | age_at_first_AD | 0.751 | 0.653 | 0.863 | 5.86e-05 | 8.20e-04 | 1587 | 1250 | 337 |
| Latino/Mixed-American | No Recorded Depression | Switching vs Continuation | Alcohol_Frequency | sex_at_birthMale | 1.231 | 0.932 | 1.626 | 1.44e-01 | 1.00e+00 | 1587 | 1250 | 337 |
| Latino/Mixed-American | No Recorded Depression | Switching vs Continuation | Alcohol_Frequency | Income>200k | 0.809 | 0.386 | 1.696 | 5.74e-01 | 1.00e+00 | 1587 | 1250 | 337 |
| Latino/Mixed-American | No Recorded Depression | Switching vs Continuation | Alcohol_Frequency | Income100-150k | 1.148 | 0.722 | 1.825 | 5.61e-01 | 1.00e+00 | 1587 | 1250 | 337 |
| Latino/Mixed-American | No Recorded Depression | Switching vs Continuation | Alcohol_Frequency | Income150-200k | 0.679 | 0.341 | 1.349 | 2.69e-01 | 1.00e+00 | 1587 | 1250 | 337 |
| Latino/Mixed-American | No Recorded Depression | Switching vs Continuation | Alcohol_Frequency | Income25-35k | 0.728 | 0.473 | 1.120 | 1.49e-01 | 1.00e+00 | 1587 | 1250 | 337 |
| Latino/Mixed-American | No Recorded Depression | Switching vs Continuation | Alcohol_Frequency | Income35-50k | 0.988 | 0.648 | 1.506 | 9.55e-01 | 1.00e+00 | 1587 | 1250 | 337 |
| Latino/Mixed-American | No Recorded Depression | Switching vs Continuation | Alcohol_Frequency | Income50-75k | 0.835 | 0.550 | 1.270 | 4.00e-01 | 1.00e+00 | 1587 | 1250 | 337 |
| Latino/Mixed-American | No Recorded Depression | Switching vs Continuation | Alcohol_Frequency | Income75-100k | 1.055 | 0.642 | 1.732 | 8.34e-01 | 1.00e+00 | 1587 | 1250 | 337 |
| Latino/Mixed-American | No Recorded Depression | Switching vs Continuation | Alcohol_Frequency | Education_levelCollege Postgraduate | 0.949 | 0.548 | 1.642 | 8.52e-01 | 1.00e+00 | 1587 | 1250 | 337 |
| Latino/Mixed-American | No Recorded Depression | Switching vs Continuation | Alcohol_Frequency | Education_levelCollege Undergraduate | 1.251 | 0.841 | 1.862 | 2.69e-01 | 1.00e+00 | 1587 | 1250 | 337 |
| Latino/Mixed-American | No Recorded Depression | Switching vs Continuation | Alcohol_Frequency | Education_levelTwelve or GED | 1.237 | 0.799 | 1.916 | 3.41e-01 | 1.00e+00 | 1587 | 1250 | 337 |
| Latino/Mixed-American | No Recorded Depression | Switching vs Continuation | Alcohol_Frequency | Alcohol_Frequency2-3 Week | 1.360 | 0.789 | 2.342 | 2.68e-01 | 1.00e+00 | 1587 | 1250 | 337 |
| Latino/Mixed-American | No Recorded Depression | Switching vs Continuation | Alcohol_Frequency | Alcohol_Frequency2-4 Month | 0.981 | 0.603 | 1.597 | 9.40e-01 | 1.00e+00 | 1587 | 1250 | 337 |
| Latino/Mixed-American | No Recorded Depression | Switching vs Continuation | Alcohol_Frequency | Alcohol_Frequency4plus Week | 1.201 | 0.619 | 2.331 | 5.88e-01 | 1.00e+00 | 1587 | 1250 | 337 |
| Latino/Mixed-American | No Recorded Depression | Switching vs Continuation | Alcohol_Frequency | Alcohol_FrequencyMonthly or Less | 1.052 | 0.681 | 1.625 | 8.19e-01 | 1.00e+00 | 1587 | 1250 | 337 |
| Latino/Mixed-American | No Recorded Depression | Switching vs Continuation | Alcohol_Frequency | Alcohol_FrequencyNever (former) | 1.349 | 0.854 | 2.131 | 1.99e-01 | 1.00e+00 | 1587 | 1250 | 337 |
| Latino/Mixed-American | No Recorded Depression | Discontinuation vs Continuation | BMI | (Intercept) | 0.479 | 0.379 | 0.607 | 9.35e-10 | 1.31e-08 | 1979 | 1253 | 726 |
| Latino/Mixed-American | No Recorded Depression | Discontinuation vs Continuation | BMI | age_at_first_AD | 1.006 | 0.912 | 1.110 | 9.07e-01 | 1.00e+00 | 1979 | 1253 | 726 |
| Latino/Mixed-American | No Recorded Depression | Discontinuation vs Continuation | BMI | sex_at_birthMale | 1.340 | 1.092 | 1.645 | 5.11e-03 | 7.15e-02 | 1979 | 1253 | 726 |
| Latino/Mixed-American | No Recorded Depression | Discontinuation vs Continuation | BMI | Income>200k | 0.557 | 0.312 | 0.994 | 4.77e-02 | 6.67e-01 | 1979 | 1253 | 726 |
| Latino/Mixed-American | No Recorded Depression | Discontinuation vs Continuation | BMI | Income100-150k | 0.662 | 0.453 | 0.968 | 3.32e-02 | 4.64e-01 | 1979 | 1253 | 726 |
| Latino/Mixed-American | No Recorded Depression | Discontinuation vs Continuation | BMI | Income150-200k | 0.574 | 0.342 | 0.964 | 3.57e-02 | 5.00e-01 | 1979 | 1253 | 726 |
| Latino/Mixed-American | No Recorded Depression | Discontinuation vs Continuation | BMI | Income25-35k | 0.666 | 0.486 | 0.914 | 1.17e-02 | 1.64e-01 | 1979 | 1253 | 726 |
| Latino/Mixed-American | No Recorded Depression | Discontinuation vs Continuation | BMI | Income35-50k | 1.036 | 0.759 | 1.413 | 8.24e-01 | 1.00e+00 | 1979 | 1253 | 726 |
| Latino/Mixed-American | No Recorded Depression | Discontinuation vs Continuation | BMI | Income50-75k | 0.796 | 0.585 | 1.084 | 1.48e-01 | 1.00e+00 | 1979 | 1253 | 726 |
| Latino/Mixed-American | No Recorded Depression | Discontinuation vs Continuation | BMI | Income75-100k | 0.568 | 0.372 | 0.865 | 8.43e-03 | 1.18e-01 | 1979 | 1253 | 726 |
| Latino/Mixed-American | No Recorded Depression | Discontinuation vs Continuation | BMI | Education_levelCollege Postgraduate | 1.091 | 0.724 | 1.643 | 6.78e-01 | 1.00e+00 | 1979 | 1253 | 726 |
| Latino/Mixed-American | No Recorded Depression | Discontinuation vs Continuation | BMI | Education_levelCollege Undergraduate | 1.438 | 1.080 | 1.915 | 1.28e-02 | 1.79e-01 | 1979 | 1253 | 726 |
| Latino/Mixed-American | No Recorded Depression | Discontinuation vs Continuation | BMI | Education_levelTwelve or GED | 1.607 | 1.178 | 2.193 | 2.74e-03 | 3.83e-02 | 1979 | 1253 | 726 |
| Latino/Mixed-American | No Recorded Depression | Discontinuation vs Continuation | BMI | BMI | 1.069 | 0.971 | 1.178 | 1.74e-01 | 1.00e+00 | 1979 | 1253 | 726 |
| Latino/Mixed-American | No Recorded Depression | Switching vs Continuation | BMI | (Intercept) | 0.226 | 0.165 | 0.309 | <2.2e-16 | <2.2e-16 | 1596 | 1253 | 343 |
| Latino/Mixed-American | No Recorded Depression | Switching vs Continuation | BMI | age_at_first_AD | 0.768 | 0.670 | 0.880 | 1.46e-04 | 2.04e-03 | 1596 | 1253 | 343 |
| Latino/Mixed-American | No Recorded Depression | Switching vs Continuation | BMI | sex_at_birthMale | 1.250 | 0.952 | 1.642 | 1.09e-01 | 1.00e+00 | 1596 | 1253 | 343 |
| Latino/Mixed-American | No Recorded Depression | Switching vs Continuation | BMI | Income>200k | 0.826 | 0.399 | 1.708 | 6.06e-01 | 1.00e+00 | 1596 | 1253 | 343 |
| Latino/Mixed-American | No Recorded Depression | Switching vs Continuation | BMI | Income100-150k | 1.159 | 0.737 | 1.825 | 5.23e-01 | 1.00e+00 | 1596 | 1253 | 343 |

| | | | | | | | | | | | | |
|---|---|---|---|---|---|---|---|---|---|---|---|---|
| Latino/Mixed-American | No Recorded Depression | Switching vs Continuation | BMI | Income150-200k | 0.757 | 0.390 | 1.468 | 4.10e-01 | 1.00e+00 | 1596 | 1253 | 343 |
| Latino/Mixed-American | No Recorded Depression | Switching vs Continuation | BMI | Income25-35k | 0.709 | 0.463 | 1.088 | 1.15e-01 | 1.00e+00 | 1596 | 1253 | 343 |
| Latino/Mixed-American | No Recorded Depression | Switching vs Continuation | BMI | Income35-50k | 1.054 | 0.701 | 1.585 | 8.00e-01 | 1.00e+00 | 1596 | 1253 | 343 |
| Latino/Mixed-American | No Recorded Depression | Switching vs Continuation | BMI | Income50-75k | 0.862 | 0.571 | 1.301 | 4.80e-01 | 1.00e+00 | 1596 | 1253 | 343 |
| Latino/Mixed-American | No Recorded Depression | Switching vs Continuation | BMI | Income75-100k | 1.034 | 0.635 | 1.684 | 8.92e-01 | 1.00e+00 | 1596 | 1253 | 343 |
| Latino/Mixed-American | No Recorded Depression | Switching vs Continuation | BMI | Education_levelCollege Postgraduate | 0.863 | 0.505 | 1.473 | 5.88e-01 | 1.00e+00 | 1596 | 1253 | 343 |
| Latino/Mixed-American | No Recorded Depression | Switching vs Continuation | BMI | Education_levelCollege Undergraduate | 1.188 | 0.811 | 1.741 | 3.75e-01 | 1.00e+00 | 1596 | 1253 | 343 |
| Latino/Mixed-American | No Recorded Depression | Switching vs Continuation | BMI | Education_levelTwelve or GED | 1.166 | 0.761 | 1.786 | 4.81e-01 | 1.00e+00 | 1596 | 1253 | 343 |
| Latino/Mixed-American | No Recorded Depression | Switching vs Continuation | BMI | BMI | 1.029 | 0.906 | 1.170 | 6.58e-01 | 1.00e+00 | 1596 | 1253 | 343 |
| Latino/Mixed-American | No Recorded Depression | Discontinuation vs Continuation | Education_level | (Intercept) | 0.506 | 0.402 | 0.637 | 7.28e-09 | 1.02e-07 | 2057 | 1277 | 780 |
| Latino/Mixed-American | No Recorded Depression | Discontinuation vs Continuation | Education_level | age_at_first_AD | 0.999 | 0.907 | 1.099 | 9.76e-01 | 1.00e+00 | 2057 | 1277 | 780 |
| Latino/Mixed-American | No Recorded Depression | Discontinuation vs Continuation | Education_level | sex_at_birthMale | 1.312 | 1.075 | 1.601 | 7.53e-03 | 1.05e-01 | 2057 | 1277 | 780 |
| Latino/Mixed-American | No Recorded Depression | Discontinuation vs Continuation | Education_level | Income>200k | 0.592 | 0.339 | 1.034 | 6.54e-02 | 9.16e-01 | 2057 | 1277 | 780 |
| Latino/Mixed-American | No Recorded Depression | Discontinuation vs Continuation | Education_level | Income100-150k | 0.682 | 0.472 | 0.987 | 4.25e-02 | 5.95e-01 | 2057 | 1277 | 780 |
| Latino/Mixed-American | No Recorded Depression | Discontinuation vs Continuation | Education_level | Income150-200k | 0.553 | 0.334 | 0.918 | 2.19e-02 | 3.07e-01 | 2057 | 1277 | 780 |
| Latino/Mixed-American | No Recorded Depression | Discontinuation vs Continuation | Education_level | Income25-35k | 0.711 | 0.524 | 0.964 | 2.81e-02 | 3.94e-01 | 2057 | 1277 | 780 |
| Latino/Mixed-American | No Recorded Depression | Discontinuation vs Continuation | Education_level | Income35-50k | 0.991 | 0.731 | 1.344 | 9.55e-01 | 1.00e+00 | 2057 | 1277 | 780 |
| Latino/Mixed-American | No Recorded Depression | Discontinuation vs Continuation | Education_level | Income50-75k | 0.812 | 0.601 | 1.096 | 1.74e-01 | 1.00e+00 | 2057 | 1277 | 780 |
| Latino/Mixed-American | No Recorded Depression | Discontinuation vs Continuation | Education_level | Income75-100k | 0.629 | 0.421 | 0.940 | 2.36e-02 | 3.30e-01 | 2057 | 1277 | 780 |
| Latino/Mixed-American | No Recorded Depression | Discontinuation vs Continuation | Education_level | Education_levelCollege Postgraduate | 1.027 | 0.687 | 1.534 | 8.98e-01 | 1.00e+00 | 2057 | 1277 | 780 |
| Latino/Mixed-American | No Recorded Depression | Discontinuation vs Continuation | Education_level | Education_levelCollege Undergraduate | 1.427 | 1.079 | 1.889 | 1.28e-02 | 1.79e-01 | 2057 | 1277 | 780 |
| Latino/Mixed-American | No Recorded Depression | Discontinuation vs Continuation | Education_level | Education_levelTwelve or GED | 1.624 | 1.199 | 2.199 | 1.74e-03 | 2.44e-02 | 2057 | 1277 | 780 |
| Latino/Mixed-American | No Recorded Depression | Switching vs Continuation | Education_level | (Intercept) | 0.224 | 0.164 | 0.307 | <2.2e-16 | <2.2e-16 | 1622 | 1277 | 345 |
| Latino/Mixed-American | No Recorded Depression | Switching vs Continuation | Education_level | age_at_first_AD | 0.769 | 0.672 | 0.881 | 1.48e-04 | 2.08e-03 | 1622 | 1277 | 345 |
| Latino/Mixed-American | No Recorded Depression | Switching vs Continuation | Education_level | sex_at_birthMale | 1.243 | 0.948 | 1.629 | 1.15e-01 | 1.00e+00 | 1622 | 1277 | 345 |
| Latino/Mixed-American | No Recorded Depression | Switching vs Continuation | Education_level | Income>200k | 0.828 | 0.402 | 1.707 | 6.09e-01 | 1.00e+00 | 1622 | 1277 | 345 |
| Latino/Mixed-American | No Recorded Depression | Switching vs Continuation | Education_level | Income100-150k | 1.196 | 0.763 | 1.875 | 4.35e-01 | 1.00e+00 | 1622 | 1277 | 345 |
| Latino/Mixed-American | No Recorded Depression | Switching vs Continuation | Education_level | Income150-200k | 0.742 | 0.384 | 1.437 | 3.77e-01 | 1.00e+00 | 1622 | 1277 | 345 |
| Latino/Mixed-American | No Recorded Depression | Switching vs Continuation | Education_level | Income25-35k | 0.743 | 0.487 | 1.135 | 1.69e-01 | 1.00e+00 | 1622 | 1277 | 345 |
| Latino/Mixed-American | No Recorded Depression | Switching vs Continuation | Education_level | Income35-50k | 1.036 | 0.690 | 1.555 | 8.64e-01 | 1.00e+00 | 1622 | 1277 | 345 |
| Latino/Mixed-American | No Recorded Depression | Switching vs Continuation | Education_level | Income50-75k | 0.852 | 0.565 | 1.284 | 4.43e-01 | 1.00e+00 | 1622 | 1277 | 345 |
| Latino/Mixed-American | No Recorded Depression | Switching vs Continuation | Education_level | Income75-100k | 1.047 | 0.644 | 1.704 | 8.52e-01 | 1.00e+00 | 1622 | 1277 | 345 |
| Latino/Mixed-American | No Recorded Depression | Switching vs Continuation | Education_level | Education_levelCollege Postgraduate | 0.864 | 0.508 | 1.471 | 5.91e-01 | 1.00e+00 | 1622 | 1277 | 345 |
| Latino/Mixed-American | No Recorded Depression | Switching vs Continuation | Education_level | Education_levelCollege Undergraduate | 1.170 | 0.800 | 1.712 | 4.19e-01 | 1.00e+00 | 1622 | 1277 | 345 |
| Latino/Mixed-American | No Recorded Depression | Switching vs Continuation | Education_level | Education_levelTwelve or GED | 1.145 | 0.748 | 1.752 | 5.32e-01 | 1.00e+00 | 1622 | 1277 | 345 |
| Latino/Mixed-American | No Recorded Depression | Discontinuation vs Continuation | Income | (Intercept) | 0.506 | 0.402 | 0.637 | 7.28e-09 | 1.02e-07 | 2057 | 1277 | 780 |
| Latino/Mixed-American | No Recorded Depression | Discontinuation vs Continuation | Income | age_at_first_AD | 0.999 | 0.907 | 1.099 | 9.76e-01 | 1.00e+00 | 2057 | 1277 | 780 |
| Latino/Mixed-American | No Recorded Depression | Discontinuation vs Continuation | Income | sex_at_birthMale | 1.312 | 1.075 | 1.601 | 7.53e-03 | 1.05e-01 | 2057 | 1277 | 780 |
| Latino/Mixed-American | No Recorded Depression | Discontinuation vs Continuation | Income | Income>200k | 0.592 | 0.339 | 1.034 | 6.54e-02 | 9.16e-01 | 2057 | 1277 | 780 |
| Latino/Mixed-American | No Recorded Depression | Discontinuation vs Continuation | Income | Income100-150k | 0.682 | 0.472 | 0.987 | 4.25e-02 | 5.95e-01 | 2057 | 1277 | 780 |
| Latino/Mixed-American | No Recorded Depression | Discontinuation vs Continuation | Income | Income150-200k | 0.553 | 0.334 | 0.918 | 2.19e-02 | 3.07e-01 | 2057 | 1277 | 780 |
| Latino/Mixed-American | No Recorded Depression | Discontinuation vs Continuation | Income | Income25-35k | 0.711 | 0.524 | 0.964 | 2.81e-02 | 3.94e-01 | 2057 | 1277 | 780 |
| Latino/Mixed-American | No Recorded Depression | Discontinuation vs Continuation | Income | Income35-50k | 0.991 | 0.731 | 1.344 | 9.55e-01 | 1.00e+00 | 2057 | 1277 | 780 |
| Latino/Mixed-American | No Recorded Depression | Discontinuation vs Continuation | Income | Income50-75k | 0.812 | 0.601 | 1.096 | 1.74e-01 | 1.00e+00 | 2057 | 1277 | 780 |
| Latino/Mixed-American | No Recorded Depression | Discontinuation vs Continuation | Income | Income75-100k | 0.629 | 0.421 | 0.940 | 2.36e-02 | 3.30e-01 | 2057 | 1277 | 780 |
| Latino/Mixed-American | No Recorded Depression | Discontinuation vs Continuation | Income | Education_levelCollege Postgraduate | 1.027 | 0.687 | 1.534 | 8.98e-01 | 1.00e+00 | 2057 | 1277 | 780 |
| Latino/Mixed-American | No Recorded Depression | Discontinuation vs Continuation | Income | Education_levelCollege Undergraduate | 1.427 | 1.079 | 1.889 | 1.28e-02 | 1.79e-01 | 2057 | 1277 | 780 |
| Latino/Mixed-American | No Recorded Depression | Discontinuation vs Continuation | Income | Education_levelTwelve or GED | 1.624 | 1.199 | 2.199 | 1.74e-03 | 2.44e-02 | 2057 | 1277 | 780 |
| Latino/Mixed-American | No Recorded Depression | Switching vs Continuation | Income | (Intercept) | 0.224 | 0.164 | 0.307 | <2.2e-16 | <2.2e-16 | 1622 | 1277 | 345 |
| Latino/Mixed-American | No Recorded Depression | Switching vs Continuation | Income | age_at_first_AD | 0.769 | 0.672 | 0.881 | 1.48e-04 | 2.08e-03 | 1622 | 1277 | 345 |
| Latino/Mixed-American | No Recorded Depression | Switching vs Continuation | Income | sex_at_birthMale | 1.243 | 0.948 | 1.629 | 1.15e-01 | 1.00e+00 | 1622 | 1277 | 345 |
| Latino/Mixed-American | No Recorded Depression | Switching vs Continuation | Income | Income>200k | 0.828 | 0.402 | 1.707 | 6.09e-01 | 1.00e+00 | 1622 | 1277 | 345 |
| Latino/Mixed-American | No Recorded Depression | Switching vs Continuation | Income | Income100-150k | 1.196 | 0.763 | 1.875 | 4.35e-01 | 1.00e+00 | 1622 | 1277 | 345 |
| Latino/Mixed-American | No Recorded Depression | Switching vs Continuation | Income | Income150-200k | 0.742 | 0.384 | 1.437 | 3.77e-01 | 1.00e+00 | 1622 | 1277 | 345 |
| Latino/Mixed-American | No Recorded Depression | Switching vs Continuation | Income | Income25-35k | 0.743 | 0.487 | 1.135 | 1.69e-01 | 1.00e+00 | 1622 | 1277 | 345 |
| Latino/Mixed-American | No Recorded Depression | Switching vs Continuation | Income | Income35-50k | 1.036 | 0.690 | 1.555 | 8.64e-01 | 1.00e+00 | 1622 | 1277 | 345 |
| Latino/Mixed-American | No Recorded Depression | Switching vs Continuation | Income | Income50-75k | 0.852 | 0.565 | 1.284 | 4.43e-01 | 1.00e+00 | 1622 | 1277 | 345 |
| Latino/Mixed-American | No Recorded Depression | Switching vs Continuation | Income | Income75-100k | 1.047 | 0.644 | 1.704 | 8.52e-01 | 1.00e+00 | 1622 | 1277 | 345 |
| Latino/Mixed-American | No Recorded Depression | Switching vs Continuation | Income | Education_levelCollege Postgraduate | 0.864 | 0.508 | 1.471 | 5.91e-01 | 1.00e+00 | 1622 | 1277 | 345 |
| Latino/Mixed-American | No Recorded Depression | Switching vs Continuation | Income | Education_levelCollege Undergraduate | 1.170 | 0.800 | 1.712 | 4.19e-01 | 1.00e+00 | 1622 | 1277 | 345 |
| Latino/Mixed-American | No Recorded Depression | Switching vs Continuation | Income | Education_levelTwelve or GED | 1.145 | 0.748 | 1.752 | 5.32e-01 | 1.00e+00 | 1622 | 1277 | 345 |
| Latino/Mixed-American | No Recorded Depression | Discontinuation vs Continuation | Insomnia | (Intercept) | 0.553 | 0.437 | 0.699 | 7.97e-07 | 1.12e-05 | 2057 | 1277 | 780 |
| Latino/Mixed-American | No Recorded Depression | Discontinuation vs Continuation | Insomnia | age_at_first_AD | 1.017 | 0.923 | 1.120 | 7.33e-01 | 1.00e+00 | 2057 | 1277 | 780 |
| Latino/Mixed-American | No Recorded Depression | Discontinuation vs Continuation | Insomnia | sex_at_birthMale | 1.333 | 1.092 | 1.629 | 4.84e-03 | 6.77e-02 | 2057 | 1277 | 780 |
| Latino/Mixed-American | No Recorded Depression | Discontinuation vs Continuation | Insomnia | Income>200k | 0.575 | 0.329 | 1.007 | 5.28e-02 | 7.39e-01 | 2057 | 1277 | 780 |
| Latino/Mixed-American | No Recorded Depression | Discontinuation vs Continuation | Insomnia | Income100-150k | 0.716 | 0.494 | 1.037 | 7.74e-02 | 1.00e+00 | 2057 | 1277 | 780 |

| | | | | | | | | | | | | |
|---|---|---|---|---|---|---|---|---|---|---|---|---|
| Latino/Mixed-American | No Recorded Depression | Discontinuation vs Continuation | Insomnia | Income150-200k | 0.558 | 0.336 | 0.927 | 2.44e-02 | 3.41e-01 | 2057 | 1277 | 780 |
| Latino/Mixed-American | No Recorded Depression | Discontinuation vs Continuation | Insomnia | Income25-35k | 0.718 | 0.529 | 0.976 | 3.46e-02 | 4.84e-01 | 2057 | 1277 | 780 |
| Latino/Mixed-American | No Recorded Depression | Discontinuation vs Continuation | Insomnia | Income35-50k | 0.988 | 0.728 | 1.340 | 9.36e-01 | 1.00e+00 | 2057 | 1277 | 780 |
| Latino/Mixed-American | No Recorded Depression | Discontinuation vs Continuation | Insomnia | Income50-75k | 0.816 | 0.604 | 1.102 | 1.85e-01 | 1.00e+00 | 2057 | 1277 | 780 |
| Latino/Mixed-American | No Recorded Depression | Discontinuation vs Continuation | Insomnia | Income75-100k | 0.643 | 0.430 | 0.962 | 3.19e-02 | 4.46e-01 | 2057 | 1277 | 780 |
| Latino/Mixed-American | No Recorded Depression | Discontinuation vs Continuation | Insomnia | Education_levelCollege Postgraduate | 0.995 | 0.665 | 1.490 | 9.82e-01 | 1.00e+00 | 2057 | 1277 | 780 |
| Latino/Mixed-American | No Recorded Depression | Discontinuation vs Continuation | Insomnia | Education_levelCollege Undergraduate | 1.404 | 1.060 | 1.861 | 1.81e-02 | 2.53e-01 | 2057 | 1277 | 780 |
| Latino/Mixed-American | No Recorded Depression | Discontinuation vs Continuation | Insomnia | Education_levelTwelve or GED | 1.606 | 1.184 | 2.178 | 2.32e-03 | 3.25e-02 | 2057 | 1277 | 780 |
| Latino/Mixed-American | No Recorded Depression | Discontinuation vs Continuation | Insomnia | Insomnia | 0.616 | 0.480 | 0.792 | 1.57e-04 | 2.19e-03 | 2057 | 1277 | 780 |
| Latino/Mixed-American | No Recorded Depression | Switching vs Continuation | Insomnia | (Intercept) | 0.196 | 0.141 | 0.271 | <2.2e-16 | <2.2e-16 | 1622 | 1277 | 345 |
| Latino/Mixed-American | No Recorded Depression | Switching vs Continuation | Insomnia | age_at_first_AD | 0.745 | 0.649 | 0.855 | 2.77e-05 | 3.88e-04 | 1622 | 1277 | 345 |
| Latino/Mixed-American | No Recorded Depression | Switching vs Continuation | Insomnia | sex_at_birthMale | 1.204 | 0.917 | 1.582 | 1.81e-01 | 1.00e+00 | 1622 | 1277 | 345 |
| Latino/Mixed-American | No Recorded Depression | Switching vs Continuation | Insomnia | Income>200k | 0.843 | 0.407 | 1.743 | 6.45e-01 | 1.00e+00 | 1622 | 1277 | 345 |
| Latino/Mixed-American | No Recorded Depression | Switching vs Continuation | Insomnia | Income100-150k | 1.103 | 0.700 | 1.737 | 6.73e-01 | 1.00e+00 | 1622 | 1277 | 345 |
| Latino/Mixed-American | No Recorded Depression | Switching vs Continuation | Insomnia | Income150-200k | 0.744 | 0.384 | 1.444 | 3.82e-01 | 1.00e+00 | 1622 | 1277 | 345 |
| Latino/Mixed-American | No Recorded Depression | Switching vs Continuation | Insomnia | Income25-35k | 0.750 | 0.491 | 1.146 | 1.83e-01 | 1.00e+00 | 1622 | 1277 | 345 |
| Latino/Mixed-American | No Recorded Depression | Switching vs Continuation | Insomnia | Income35-50k | 1.023 | 0.680 | 1.540 | 9.12e-01 | 1.00e+00 | 1622 | 1277 | 345 |
| Latino/Mixed-American | No Recorded Depression | Switching vs Continuation | Insomnia | Income50-75k | 0.840 | 0.556 | 1.269 | 4.07e-01 | 1.00e+00 | 1622 | 1277 | 345 |
| Latino/Mixed-American | No Recorded Depression | Switching vs Continuation | Insomnia | Income75-100k | 1.036 | 0.636 | 1.688 | 8.86e-01 | 1.00e+00 | 1622 | 1277 | 345 |
| Latino/Mixed-American | No Recorded Depression | Switching vs Continuation | Insomnia | Education_levelCollege Postgraduate | 0.904 | 0.530 | 1.544 | 7.12e-01 | 1.00e+00 | 1622 | 1277 | 345 |
| Latino/Mixed-American | No Recorded Depression | Switching vs Continuation | Insomnia | Education_levelCollege Undergraduate | 1.183 | 0.807 | 1.733 | 3.90e-01 | 1.00e+00 | 1622 | 1277 | 345 |
| Latino/Mixed-American | No Recorded Depression | Switching vs Continuation | Insomnia | Education_levelTwelve or GED | 1.139 | 0.743 | 1.746 | 5.50e-01 | 1.00e+00 | 1622 | 1277 | 345 |
| Latino/Mixed-American | No Recorded Depression | Switching vs Continuation | Insomnia | Insomnia | 1.732 | 1.312 | 2.286 | 1.05e-04 | 1.47e-03 | 1622 | 1277 | 345 |
| Latino/Mixed-American | No Recorded Depression | Discontinuation vs Continuation | MDD CIDI-sf | (Intercept) | 0.458 | 0.036 | 5.864 | 5.48e-01 | 1.00e+00 | 189 | 131 | 58 |
| Latino/Mixed-American | No Recorded Depression | Discontinuation vs Continuation | MDD CIDI-sf | age_at_first_AD | 0.602 | 0.393 | 0.922 | 1.97e-02 | 2.75e-01 | 189 | 131 | 58 |
| Latino/Mixed-American | No Recorded Depression | Discontinuation vs Continuation | MDD CIDI-sf | sex_at_birthMale | 2.786 | 1.239 | 6.268 | 1.32e-02 | 1.85e-01 | 189 | 131 | 58 |
| Latino/Mixed-American | No Recorded Depression | Discontinuation vs Continuation | MDD CIDI-sf | Income>200k | 1.344 | 0.332 | 5.439 | 6.79e-01 | 1.00e+00 | 189 | 131 | 58 |
| Latino/Mixed-American | No Recorded Depression | Discontinuation vs Continuation | MDD CIDI-sf | Income100-150k | 1.686 | 0.527 | 5.393 | 3.78e-01 | 1.00e+00 | 189 | 131 | 58 |
| Latino/Mixed-American | No Recorded Depression | Discontinuation vs Continuation | MDD CIDI-sf | Income150-200k | 0.891 | 0.171 | 4.658 | 8.91e-01 | 1.00e+00 | 189 | 131 | 58 |
| Latino/Mixed-American | No Recorded Depression | Discontinuation vs Continuation | MDD CIDI-sf | Income25-35k | 1.916 | 0.554 | 6.629 | 3.04e-01 | 1.00e+00 | 189 | 131 | 58 |
| Latino/Mixed-American | No Recorded Depression | Discontinuation vs Continuation | MDD CIDI-sf | Income35-50k | 1.398 | 0.383 | 5.112 | 6.12e-01 | 1.00e+00 | 189 | 131 | 58 |
| Latino/Mixed-American | No Recorded Depression | Discontinuation vs Continuation | MDD CIDI-sf | Income50-75k | 0.570 | 0.170 | 1.912 | 3.63e-01 | 1.00e+00 | 189 | 131 | 58 |
| Latino/Mixed-American | No Recorded Depression | Discontinuation vs Continuation | MDD CIDI-sf | Income75-100k | 0.534 | 0.132 | 2.167 | 3.80e-01 | 1.00e+00 | 189 | 131 | 58 |
| Latino/Mixed-American | No Recorded Depression | Discontinuation vs Continuation | MDD CIDI-sf | Education_levelCollege Postgraduate | 0.332 | 0.024 | 4.647 | 4.12e-01 | 1.00e+00 | 189 | 131 | 58 |
| Latino/Mixed-American | No Recorded Depression | Discontinuation vs Continuation | MDD CIDI-sf | Education_levelCollege Undergraduate | 0.392 | 0.030 | 5.175 | 4.77e-01 | 1.00e+00 | 189 | 131 | 58 |
| Latino/Mixed-American | No Recorded Depression | Discontinuation vs Continuation | MDD CIDI-sf | Education_levelTwelve or GED | 0.416 | 0.027 | 6.335 | 5.28e-01 | 1.00e+00 | 189 | 131 | 58 |
| Latino/Mixed-American | No Recorded Depression | Discontinuation vs Continuation | MDD CIDI-sf | MDD CIDI-sf | 2.093 | 0.946 | 4.632 | 6.83e-02 | 9.57e-01 | 189 | 131 | 58 |
| Latino/Mixed-American | No Recorded Depression | Discontinuation vs Continuation | MDD_symptom_count | (Intercept) | 0.313 | 0.029 | 3.321 | 3.35e-01 | 1.00e+00 | 229 | 158 | 71 |
| Latino/Mixed-American | No Recorded Depression | Discontinuation vs Continuation | MDD_symptom_count | age_at_first_AD | 0.616 | 0.424 | 0.896 | 1.12e-02 | 1.57e-01 | 229 | 158 | 71 |
| Latino/Mixed-American | No Recorded Depression | Discontinuation vs Continuation | MDD_symptom_count | sex_at_birthMale | 3.303 | 1.626 | 6.712 | 9.57e-04 | 1.34e-02 | 229 | 158 | 71 |
| Latino/Mixed-American | No Recorded Depression | Discontinuation vs Continuation | MDD_symptom_count | Income>200k | 1.600 | 0.440 | 5.822 | 4.76e-01 | 1.00e+00 | 229 | 158 | 71 |
| Latino/Mixed-American | No Recorded Depression | Discontinuation vs Continuation | MDD_symptom_count | Income100-150k | 2.199 | 0.780 | 6.203 | 1.36e-01 | 1.00e+00 | 229 | 158 | 71 |
| Latino/Mixed-American | No Recorded Depression | Discontinuation vs Continuation | MDD_symptom_count | Income150-200k | 0.916 | 0.206 | 4.072 | 9.09e-01 | 1.00e+00 | 229 | 158 | 71 |
| Latino/Mixed-American | No Recorded Depression | Discontinuation vs Continuation | MDD_symptom_count | Income25-35k | 1.718 | 0.530 | 5.570 | 3.67e-01 | 1.00e+00 | 229 | 158 | 71 |
| Latino/Mixed-American | No Recorded Depression | Discontinuation vs Continuation | MDD_symptom_count | Income35-50k | 1.312 | 0.398 | 4.328 | 6.56e-01 | 1.00e+00 | 229 | 158 | 71 |
| Latino/Mixed-American | No Recorded Depression | Discontinuation vs Continuation | MDD_symptom_count | Income50-75k | 0.597 | 0.199 | 1.791 | 3.57e-01 | 1.00e+00 | 229 | 158 | 71 |
| Latino/Mixed-American | No Recorded Depression | Discontinuation vs Continuation | MDD_symptom_count | Income75-100k | 0.657 | 0.181 | 2.385 | 5.24e-01 | 1.00e+00 | 229 | 158 | 71 |
| Latino/Mixed-American | No Recorded Depression | Discontinuation vs Continuation | MDD_symptom_count | Education_levelCollege Postgraduate | 0.607 | 0.051 | 7.196 | 6.92e-01 | 1.00e+00 | 229 | 158 | 71 |
| Latino/Mixed-American | No Recorded Depression | Discontinuation vs Continuation | MDD_symptom_count | Education_levelCollege Undergraduate | 0.869 | 0.078 | 9.644 | 9.09e-01 | 1.00e+00 | 229 | 158 | 71 |
| Latino/Mixed-American | No Recorded Depression | Discontinuation vs Continuation | MDD_symptom_count | Education_levelTwelve or GED | 0.923 | 0.075 | 11.440 | 9.50e-01 | 1.00e+00 | 229 | 158 | 71 |
| Latino/Mixed-American | No Recorded Depression | Discontinuation vs Continuation | MDD_symptom_count | MDD_symptom_count | 1.194 | 0.876 | 1.629 | 2.62e-01 | 1.00e+00 | 229 | 158 | 71 |
| Latino/Mixed-American | No Recorded Depression | Switching vs Continuation | MDD_symptom_count | (Intercept) | 1.351 | 0.198 | 9.195 | 7.59e-01 | 1.00e+00 | 215 | 158 | 57 |
| Latino/Mixed-American | No Recorded Depression | Switching vs Continuation | MDD_symptom_count | age_at_first_AD | 0.482 | 0.311 | 0.747 | 1.09e-03 | 1.53e-02 | 215 | 158 | 57 |
| Latino/Mixed-American | No Recorded Depression | Switching vs Continuation | MDD_symptom_count | sex_at_birthMale | 2.235 | 1.008 | 4.956 | 4.77e-02 | 6.68e-01 | 215 | 158 | 57 |
| Latino/Mixed-American | No Recorded Depression | Switching vs Continuation | MDD_symptom_count | Income>200k | 0.352 | 0.079 | 1.563 | 1.70e-01 | 1.00e+00 | 215 | 158 | 57 |
| Latino/Mixed-American | No Recorded Depression | Switching vs Continuation | MDD_symptom_count | Income100-150k | 0.501 | 0.161 | 1.562 | 2.34e-01 | 1.00e+00 | 215 | 158 | 57 |
| Latino/Mixed-American | No Recorded Depression | Switching vs Continuation | MDD_symptom_count | Income150-200k | 0.262 | 0.046 | 1.482 | 1.30e-01 | 1.00e+00 | 215 | 158 | 57 |
| Latino/Mixed-American | No Recorded Depression | Switching vs Continuation | MDD_symptom_count | Income25-35k | 0.539 | 0.151 | 1.923 | 3.41e-01 | 1.00e+00 | 215 | 158 | 57 |
| Latino/Mixed-American | No Recorded Depression | Switching vs Continuation | MDD_symptom_count | Income35-50k | 0.628 | 0.202 | 1.955 | 4.22e-01 | 1.00e+00 | 215 | 158 | 57 |
| Latino/Mixed-American | No Recorded Depression | Switching vs Continuation | MDD_symptom_count | Income50-75k | 0.236 | 0.078 | 0.712 | 1.04e-02 | 1.46e-01 | 215 | 158 | 57 |
| Latino/Mixed-American | No Recorded Depression | Switching vs Continuation | MDD_symptom_count | Income75-100k | 0.429 | 0.134 | 1.378 | 1.55e-01 | 1.00e+00 | 215 | 158 | 57 |
| Latino/Mixed-American | No Recorded Depression | Switching vs Continuation | MDD_symptom_count | Education_levelCollege Postgraduate | 0.233 | 0.028 | 1.980 | 1.82e-01 | 1.00e+00 | 215 | 158 | 57 |
| Latino/Mixed-American | No Recorded Depression | Switching vs Continuation | MDD_symptom_count | Education_levelCollege Undergraduate | 0.392 | 0.052 | 2.933 | 3.62e-01 | 1.00e+00 | 215 | 158 | 57 |
| Latino/Mixed-American | No Recorded Depression | Switching vs Continuation | MDD_symptom_count | Education_levelTwelve or GED | 0.491 | 0.057 | 4.210 | 5.16e-01 | 1.00e+00 | 215 | 158 | 57 |
| Latino/Mixed-American | No Recorded Depression | Switching vs Continuation | MDD_symptom_count | MDD_symptom_count | 1.079 | 0.779 | 1.495 | 6.48e-01 | 1.00e+00 | 215 | 158 | 57 |
| Latino/Mixed-American | No Recorded Depression | Discontinuation vs Continuation | Migraine | (Intercept) | 0.550 | 0.434 | 0.697 | 7.38e-07 | 1.03e-05 | 2057 | 1277 | 780 |

| | | | | | | | | | | | | |
|---|---|---|---|---|---|---|---|---|---|---|---|---|
| Latino/Mixed-American | No Recorded Depression | Discontinuation vs Continuation | Migraine | age_at_first_AD | 0.987 | 0.896 | 1.086 | 7.82e-01 | 1.00e+00 | 2057 | 1277 | 780 |
| Latino/Mixed-American | No Recorded Depression | Discontinuation vs Continuation | Migraine | sex_at_birthMale | 1.251 | 1.022 | 1.530 | 2.97e-02 | 4.16e-01 | 2057 | 1277 | 780 |
| Latino/Mixed-American | No Recorded Depression | Discontinuation vs Continuation | Migraine | Income>200k | 0.591 | 0.338 | 1.033 | 6.50e-02 | 9.11e-01 | 2057 | 1277 | 780 |
| Latino/Mixed-American | No Recorded Depression | Discontinuation vs Continuation | Migraine | Income100-150k | 0.700 | 0.483 | 1.013 | 5.88e-02 | 8.23e-01 | 2057 | 1277 | 780 |
| Latino/Mixed-American | No Recorded Depression | Discontinuation vs Continuation | Migraine | Income150-200k | 0.558 | 0.336 | 0.927 | 2.43e-02 | 3.41e-01 | 2057 | 1277 | 780 |
| Latino/Mixed-American | No Recorded Depression | Discontinuation vs Continuation | Migraine | Income25-35k | 0.719 | 0.529 | 0.976 | 3.44e-02 | 4.82e-01 | 2057 | 1277 | 780 |
| Latino/Mixed-American | No Recorded Depression | Discontinuation vs Continuation | Migraine | Income35-50k | 1.008 | 0.743 | 1.368 | 9.61e-01 | 1.00e+00 | 2057 | 1277 | 780 |
| Latino/Mixed-American | No Recorded Depression | Discontinuation vs Continuation | Migraine | Income50-75k | 0.810 | 0.600 | 1.094 | 1.70e-01 | 1.00e+00 | 2057 | 1277 | 780 |
| Latino/Mixed-American | No Recorded Depression | Discontinuation vs Continuation | Migraine | Income75-100k | 0.646 | 0.432 | 0.966 | 3.31e-02 | 4.64e-01 | 2057 | 1277 | 780 |
| Latino/Mixed-American | No Recorded Depression | Discontinuation vs Continuation | Migraine | Education_levelCollege Postgraduate | 1.027 | 0.687 | 1.535 | 8.98e-01 | 1.00e+00 | 2057 | 1277 | 780 |
| Latino/Mixed-American | No Recorded Depression | Discontinuation vs Continuation | Migraine | Education_levelCollege Undergraduate | 1.437 | 1.085 | 1.903 | 1.14e-02 | 1.60e-01 | 2057 | 1277 | 780 |
| Latino/Mixed-American | No Recorded Depression | Discontinuation vs Continuation | Migraine | Education_levelTwelve or GED | 1.629 | 1.202 | 2.208 | 1.66e-03 | 2.32e-02 | 2057 | 1277 | 780 |
| Latino/Mixed-American | No Recorded Depression | Discontinuation vs Continuation | Migraine | Migraine | 0.717 | 0.578 | 0.889 | 2.48e-03 | 3.47e-02 | 2057 | 1277 | 780 |
| Latino/Mixed-American | No Recorded Depression | Switching vs Continuation | Migraine | (Intercept) | 0.184 | 0.132 | 0.256 | <2.2e-16 | <2.2e-16 | 1622 | 1277 | 345 |
| Latino/Mixed-American | No Recorded Depression | Switching vs Continuation | Migraine | age_at_first_AD | 0.783 | 0.682 | 0.898 | 4.55e-04 | 6.37e-03 | 1622 | 1277 | 345 |
| Latino/Mixed-American | No Recorded Depression | Switching vs Continuation | Migraine | sex_at_birthMale | 1.402 | 1.060 | 1.853 | 1.78e-02 | 2.49e-01 | 1622 | 1277 | 345 |
| Latino/Mixed-American | No Recorded Depression | Switching vs Continuation | Migraine | Income>200k | 0.811 | 0.392 | 1.681 | 5.74e-01 | 1.00e+00 | 1622 | 1277 | 345 |
| Latino/Mixed-American | No Recorded Depression | Switching vs Continuation | Migraine | Income100-150k | 1.165 | 0.742 | 1.829 | 5.08e-01 | 1.00e+00 | 1622 | 1277 | 345 |
| Latino/Mixed-American | No Recorded Depression | Switching vs Continuation | Migraine | Income150-200k | 0.749 | 0.386 | 1.453 | 3.93e-01 | 1.00e+00 | 1622 | 1277 | 345 |
| Latino/Mixed-American | No Recorded Depression | Switching vs Continuation | Migraine | Income25-35k | 0.743 | 0.486 | 1.136 | 1.70e-01 | 1.00e+00 | 1622 | 1277 | 345 |
| Latino/Mixed-American | No Recorded Depression | Switching vs Continuation | Migraine | Income35-50k | 1.021 | 0.679 | 1.536 | 9.21e-01 | 1.00e+00 | 1622 | 1277 | 345 |
| Latino/Mixed-American | No Recorded Depression | Switching vs Continuation | Migraine | Income50-75k | 0.863 | 0.571 | 1.304 | 4.84e-01 | 1.00e+00 | 1622 | 1277 | 345 |
| Latino/Mixed-American | No Recorded Depression | Switching vs Continuation | Migraine | Income75-100k | 1.012 | 0.621 | 1.650 | 9.61e-01 | 1.00e+00 | 1622 | 1277 | 345 |
| Latino/Mixed-American | No Recorded Depression | Switching vs Continuation | Migraine | Education_levelCollege Postgraduate | 0.870 | 0.509 | 1.485 | 6.10e-01 | 1.00e+00 | 1622 | 1277 | 345 |
| Latino/Mixed-American | No Recorded Depression | Switching vs Continuation | Migraine | Education_levelCollege Undergraduate | 1.160 | 0.792 | 1.699 | 4.46e-01 | 1.00e+00 | 1622 | 1277 | 345 |
| Latino/Mixed-American | No Recorded Depression | Switching vs Continuation | Migraine | Education_levelTwelve or GED | 1.146 | 0.747 | 1.758 | 5.32e-01 | 1.00e+00 | 1622 | 1277 | 345 |
| Latino/Mixed-American | No Recorded Depression | Switching vs Continuation | Migraine | Migraine | 1.706 | 1.319 | 2.206 | 4.67e-05 | 6.54e-04 | 1622 | 1277 | 345 |
| Latino/Mixed-American | No Recorded Depression | Discontinuation vs Continuation | Recorded Anxiety | (Intercept) | 0.565 | 0.445 | 0.718 | 2.85e-06 | 3.99e-05 | 2057 | 1277 | 780 |
| Latino/Mixed-American | No Recorded Depression | Discontinuation vs Continuation | Recorded Anxiety | age_at_first_AD | 0.980 | 0.889 | 1.079 | 6.77e-01 | 1.00e+00 | 2057 | 1277 | 780 |
| Latino/Mixed-American | No Recorded Depression | Discontinuation vs Continuation | Recorded Anxiety | sex_at_birthMale | 1.326 | 1.086 | 1.620 | 5.61e-03 | 7.85e-02 | 2057 | 1277 | 780 |
| Latino/Mixed-American | No Recorded Depression | Discontinuation vs Continuation | Recorded Anxiety | Income>200k | 0.559 | 0.319 | 0.979 | 4.20e-02 | 5.88e-01 | 2057 | 1277 | 780 |
| Latino/Mixed-American | No Recorded Depression | Discontinuation vs Continuation | Recorded Anxiety | Income100-150k | 0.694 | 0.479 | 1.006 | 5.38e-02 | 7.53e-01 | 2057 | 1277 | 780 |
| Latino/Mixed-American | No Recorded Depression | Discontinuation vs Continuation | Recorded Anxiety | Income150-200k | 0.553 | 0.333 | 0.918 | 2.20e-02 | 3.08e-01 | 2057 | 1277 | 780 |
| Latino/Mixed-American | No Recorded Depression | Discontinuation vs Continuation | Recorded Anxiety | Income25-35k | 0.714 | 0.526 | 0.970 | 3.13e-02 | 4.38e-01 | 2057 | 1277 | 780 |
| Latino/Mixed-American | No Recorded Depression | Discontinuation vs Continuation | Recorded Anxiety | Income35-50k | 0.998 | 0.736 | 1.354 | 9.90e-01 | 1.00e+00 | 2057 | 1277 | 780 |
| Latino/Mixed-American | No Recorded Depression | Discontinuation vs Continuation | Recorded Anxiety | Income50-75k | 0.825 | 0.610 | 1.115 | 2.10e-01 | 1.00e+00 | 2057 | 1277 | 780 |
| Latino/Mixed-American | No Recorded Depression | Discontinuation vs Continuation | Recorded Anxiety | Income75-100k | 0.636 | 0.425 | 0.951 | 2.75e-02 | 3.85e-01 | 2057 | 1277 | 780 |
| Latino/Mixed-American | No Recorded Depression | Discontinuation vs Continuation | Recorded Anxiety | Education_levelCollege Postgraduate | 1.057 | 0.706 | 1.581 | 7.88e-01 | 1.00e+00 | 2057 | 1277 | 780 |
| Latino/Mixed-American | No Recorded Depression | Discontinuation vs Continuation | Recorded Anxiety | Education_levelCollege Undergraduate | 1.449 | 1.094 | 1.919 | 9.78e-03 | 1.37e-01 | 2057 | 1277 | 780 |
| Latino/Mixed-American | No Recorded Depression | Discontinuation vs Continuation | Recorded Anxiety | Education_levelTwelve or GED | 1.674 | 1.234 | 2.270 | 9.29e-04 | 1.30e-02 | 2057 | 1277 | 780 |
| Latino/Mixed-American | No Recorded Depression | Discontinuation vs Continuation | Recorded Anxiety | Recorded Anxiety | 0.719 | 0.597 | 0.866 | 5.18e-04 | 7.25e-03 | 2057 | 1277 | 780 |
| Latino/Mixed-American | No Recorded Depression | Switching vs Continuation | Recorded Anxiety | (Intercept) | 0.182 | 0.130 | 0.254 | <2.2e-16 | <2.2e-16 | 1622 | 1277 | 345 |
| Latino/Mixed-American | No Recorded Depression | Switching vs Continuation | Recorded Anxiety | age_at_first_AD | 0.797 | 0.694 | 0.914 | 1.19e-03 | 1.67e-02 | 1622 | 1277 | 345 |
| Latino/Mixed-American | No Recorded Depression | Switching vs Continuation | Recorded Anxiety | sex_at_birthMale | 1.212 | 0.923 | 1.591 | 1.66e-01 | 1.00e+00 | 1622 | 1277 | 345 |
| Latino/Mixed-American | No Recorded Depression | Switching vs Continuation | Recorded Anxiety | Income>200k | 0.863 | 0.417 | 1.784 | 6.91e-01 | 1.00e+00 | 1622 | 1277 | 345 |
| Latino/Mixed-American | No Recorded Depression | Switching vs Continuation | Recorded Anxiety | Income100-150k | 1.098 | 0.697 | 1.728 | 6.88e-01 | 1.00e+00 | 1622 | 1277 | 345 |
| Latino/Mixed-American | No Recorded Depression | Switching vs Continuation | Recorded Anxiety | Income150-200k | 0.743 | 0.383 | 1.442 | 3.80e-01 | 1.00e+00 | 1622 | 1277 | 345 |
| Latino/Mixed-American | No Recorded Depression | Switching vs Continuation | Recorded Anxiety | Income25-35k | 0.730 | 0.477 | 1.116 | 1.46e-01 | 1.00e+00 | 1622 | 1277 | 345 |
| Latino/Mixed-American | No Recorded Depression | Switching vs Continuation | Recorded Anxiety | Income35-50k | 0.978 | 0.648 | 1.474 | 9.14e-01 | 1.00e+00 | 1622 | 1277 | 345 |
| Latino/Mixed-American | No Recorded Depression | Switching vs Continuation | Recorded Anxiety | Income50-75k | 0.786 | 0.519 | 1.190 | 2.55e-01 | 1.00e+00 | 1622 | 1277 | 345 |
| Latino/Mixed-American | No Recorded Depression | Switching vs Continuation | Recorded Anxiety | Income75-100k | 0.979 | 0.599 | 1.601 | 9.33e-01 | 1.00e+00 | 1622 | 1277 | 345 |
| Latino/Mixed-American | No Recorded Depression | Switching vs Continuation | Recorded Anxiety | Education_levelCollege Postgraduate | 0.856 | 0.501 | 1.462 | 5.70e-01 | 1.00e+00 | 1622 | 1277 | 345 |
| Latino/Mixed-American | No Recorded Depression | Switching vs Continuation | Recorded Anxiety | Education_levelCollege Undergraduate | 1.168 | 0.797 | 1.713 | 4.26e-01 | 1.00e+00 | 1622 | 1277 | 345 |
| Latino/Mixed-American | No Recorded Depression | Switching vs Continuation | Recorded Anxiety | Education_levelTwelve or GED | 1.133 | 0.739 | 1.737 | 5.68e-01 | 1.00e+00 | 1622 | 1277 | 345 |
| Latino/Mixed-American | No Recorded Depression | Switching vs Continuation | Recorded Anxiety | Recorded Anxiety | 1.650 | 1.287 | 2.116 | 7.77e-05 | 1.09e-03 | 1622 | 1277 | 345 |
| Latino/Mixed-American | No Recorded Depression | Discontinuation vs Continuation | Smoking_Frequency | (Intercept) | 0.473 | 0.372 | 0.602 | 1.07e-09 | 1.50e-08 | 1998 | 1235 | 763 |
| Latino/Mixed-American | No Recorded Depression | Discontinuation vs Continuation | Smoking_Frequency | age_at_first_AD | 0.999 | 0.906 | 1.103 | 9.91e-01 | 1.00e+00 | 1998 | 1235 | 763 |
| Latino/Mixed-American | No Recorded Depression | Discontinuation vs Continuation | Smoking_Frequency | sex_at_birthMale | 1.237 | 1.004 | 1.523 | 4.53e-02 | 6.35e-01 | 1998 | 1235 | 763 |
| Latino/Mixed-American | No Recorded Depression | Discontinuation vs Continuation | Smoking_Frequency | Income>200k | 0.663 | 0.377 | 1.165 | 1.53e-01 | 1.00e+00 | 1998 | 1235 | 763 |
| Latino/Mixed-American | No Recorded Depression | Discontinuation vs Continuation | Smoking_Frequency | Income100-150k | 0.737 | 0.506 | 1.074 | 1.12e-01 | 1.00e+00 | 1998 | 1235 | 763 |
| Latino/Mixed-American | No Recorded Depression | Discontinuation vs Continuation | Smoking_Frequency | Income150-200k | 0.616 | 0.369 | 1.027 | 6.32e-02 | 8.85e-01 | 1998 | 1235 | 763 |
| Latino/Mixed-American | No Recorded Depression | Discontinuation vs Continuation | Smoking_Frequency | Income25-35k | 0.771 | 0.565 | 1.052 | 1.01e-01 | 1.00e+00 | 1998 | 1235 | 763 |
| Latino/Mixed-American | No Recorded Depression | Discontinuation vs Continuation | Smoking_Frequency | Income35-50k | 1.025 | 0.751 | 1.398 | 8.77e-01 | 1.00e+00 | 1998 | 1235 | 763 |
| Latino/Mixed-American | No Recorded Depression | Discontinuation vs Continuation | Smoking_Frequency | Income50-75k | 0.865 | 0.637 | 1.176 | 3.55e-01 | 1.00e+00 | 1998 | 1235 | 763 |
| Latino/Mixed-American | No Recorded Depression | Discontinuation vs Continuation | Smoking_Frequency | Income75-100k | 0.692 | 0.459 | 1.042 | 7.78e-02 | 1.00e+00 | 1998 | 1235 | 763 |
| Latino/Mixed-American | No Recorded Depression | Discontinuation vs Continuation | Smoking_Frequency | Education_levelCollege Postgraduate | 1.015 | 0.674 | 1.527 | 9.44e-01 | 1.00e+00 | 1998 | 1235 | 763 |

| | | | | | | | | | | | | |
|---|---|---|---|---|---|---|---|---|---|---|---|---|
| Latino/Mixed-American | No Recorded Depression | Discontinuation vs Continuation | Smoking_Frequency | Education_levelCollege Undergraduate | 1.427 | 1.074 | 1.895 | 1.42e-02 | 1.98e-01 | 1998 | 1235 | 763 |
| Latino/Mixed-American | No Recorded Depression | Discontinuation vs Continuation | Smoking_Frequency | Education_levelTwelve or GED | 1.679 | 1.234 | 2.283 | 9.68e-04 | 1.36e-02 | 1998 | 1235 | 763 |
| Latino/Mixed-American | No Recorded Depression | Discontinuation vs Continuation | Smoking_Frequency | Smoking_FrequencyEvery Day | 1.771 | 1.204 | 2.603 | 3.66e-03 | 5.13e-02 | 1998 | 1235 | 763 |
| Latino/Mixed-American | No Recorded Depression | Discontinuation vs Continuation | Smoking_Frequency | Smoking_FrequencyNot At All (former) | 1.036 | 0.818 | 1.311 | 7.69e-01 | 1.00e+00 | 1998 | 1235 | 763 |
| Latino/Mixed-American | No Recorded Depression | Discontinuation vs Continuation | Smoking_Frequency | Smoking_FrequencySome Days | 1.093 | 0.718 | 1.665 | 6.78e-01 | 1.00e+00 | 1998 | 1235 | 763 |
| Latino/Mixed-American | No Recorded Depression | Switching vs Continuation | Smoking_Frequency | (Intercept) | 0.178 | 0.126 | 0.250 | <2.2e-16 | <2.2e-16 | 1562 | 1235 | 327 |
| Latino/Mixed-American | No Recorded Depression | Switching vs Continuation | Smoking_Frequency | age_at_first_AD | 0.742 | 0.644 | 0.856 | 4.09e-05 | 5.73e-04 | 1562 | 1235 | 327 |
| Latino/Mixed-American | No Recorded Depression | Switching vs Continuation | Smoking_Frequency | sex_at_birthMale | 1.116 | 0.840 | 1.483 | 4.48e-01 | 1.00e+00 | 1562 | 1235 | 327 |
| Latino/Mixed-American | No Recorded Depression | Switching vs Continuation | Smoking_Frequency | Income>200k | 0.798 | 0.374 | 1.703 | 5.59e-01 | 1.00e+00 | 1562 | 1235 | 327 |
| Latino/Mixed-American | No Recorded Depression | Switching vs Continuation | Smoking_Frequency | Income100-150k | 1.252 | 0.787 | 1.991 | 3.43e-01 | 1.00e+00 | 1562 | 1235 | 327 |
| Latino/Mixed-American | No Recorded Depression | Switching vs Continuation | Smoking_Frequency | Income150-200k | 0.809 | 0.414 | 1.580 | 5.34e-01 | 1.00e+00 | 1562 | 1235 | 327 |
| Latino/Mixed-American | No Recorded Depression | Switching vs Continuation | Smoking_Frequency | Income25-35k | 0.754 | 0.485 | 1.173 | 2.11e-01 | 1.00e+00 | 1562 | 1235 | 327 |
| Latino/Mixed-American | No Recorded Depression | Switching vs Continuation | Smoking_Frequency | Income35-50k | 1.027 | 0.673 | 1.567 | 9.01e-01 | 1.00e+00 | 1562 | 1235 | 327 |
| Latino/Mixed-American | No Recorded Depression | Switching vs Continuation | Smoking_Frequency | Income50-75k | 0.928 | 0.611 | 1.411 | 7.28e-01 | 1.00e+00 | 1562 | 1235 | 327 |
| Latino/Mixed-American | No Recorded Depression | Switching vs Continuation | Smoking_Frequency | Income75-100k | 1.168 | 0.712 | 1.918 | 5.39e-01 | 1.00e+00 | 1562 | 1235 | 327 |
| Latino/Mixed-American | No Recorded Depression | Switching vs Continuation | Smoking_Frequency | Education_levelCollege Postgraduate | 0.965 | 0.559 | 1.667 | 9.00e-01 | 1.00e+00 | 1562 | 1235 | 327 |
| Latino/Mixed-American | No Recorded Depression | Switching vs Continuation | Smoking_Frequency | Education_levelCollege Undergraduate | 1.192 | 0.801 | 1.772 | 3.86e-01 | 1.00e+00 | 1562 | 1235 | 327 |
| Latino/Mixed-American | No Recorded Depression | Switching vs Continuation | Smoking_Frequency | Education_levelTwelve or GED | 1.210 | 0.779 | 1.881 | 3.96e-01 | 1.00e+00 | 1562 | 1235 | 327 |
| Latino/Mixed-American | No Recorded Depression | Switching vs Continuation | Smoking_Frequency | Smoking_FrequencyEvery Day | 1.755 | 1.030 | 2.992 | 3.86e-02 | 5.41e-01 | 1562 | 1235 | 327 |
| Latino/Mixed-American | No Recorded Depression | Switching vs Continuation | Smoking_Frequency | Smoking_FrequencyNot At All (former) | 1.574 | 1.161 | 2.133 | 3.44e-03 | 4.82e-02 | 1562 | 1235 | 327 |
| Latino/Mixed-American | No Recorded Depression | Switching vs Continuation | Smoking_Frequency | Smoking_FrequencySome Days | 1.754 | 1.055 | 2.916 | 3.02e-02 | 4.22e-01 | 1562 | 1235 | 327 |
| Latino/Mixed-American | No Recorded Depression | Discontinuation vs Continuation | Stress | (Intercept) | 0.537 | 0.330 | 0.872 | 1.21e-02 | 1.69e-01 | 822 | 535 | 287 |
| Latino/Mixed-American | No Recorded Depression | Discontinuation vs Continuation | Stress | age_at_first_AD | 1.004 | 0.854 | 1.181 | 9.58e-01 | 1.00e+00 | 822 | 535 | 287 |
| Latino/Mixed-American | No Recorded Depression | Discontinuation vs Continuation | Stress | sex_at_birthMale | 1.195 | 0.849 | 1.682 | 3.07e-01 | 1.00e+00 | 822 | 535 | 287 |
| Latino/Mixed-American | No Recorded Depression | Discontinuation vs Continuation | Stress | Income>200k | 0.786 | 0.378 | 1.633 | 5.19e-01 | 1.00e+00 | 822 | 535 | 287 |
| Latino/Mixed-American | No Recorded Depression | Discontinuation vs Continuation | Stress | Income100-150k | 0.781 | 0.462 | 1.321 | 3.56e-01 | 1.00e+00 | 822 | 535 | 287 |
| Latino/Mixed-American | No Recorded Depression | Discontinuation vs Continuation | Stress | Income150-200k | 0.542 | 0.266 | 1.104 | 9.13e-02 | 1.00e+00 | 822 | 535 | 287 |
| Latino/Mixed-American | No Recorded Depression | Discontinuation vs Continuation | Stress | Income25-35k | 0.678 | 0.395 | 1.163 | 1.58e-01 | 1.00e+00 | 822 | 535 | 287 |
| Latino/Mixed-American | No Recorded Depression | Discontinuation vs Continuation | Stress | Income35-50k | 0.911 | 0.539 | 1.538 | 7.26e-01 | 1.00e+00 | 822 | 535 | 287 |
| Latino/Mixed-American | No Recorded Depression | Discontinuation vs Continuation | Stress | Income50-75k | 0.716 | 0.447 | 1.147 | 1.65e-01 | 1.00e+00 | 822 | 535 | 287 |
| Latino/Mixed-American | No Recorded Depression | Discontinuation vs Continuation | Stress | Income75-100k | 0.472 | 0.248 | 0.899 | 2.23e-02 | 3.12e-01 | 822 | 535 | 287 |
| Latino/Mixed-American | No Recorded Depression | Discontinuation vs Continuation | Stress | Education_levelCollege Postgraduate | 0.898 | 0.460 | 1.753 | 7.53e-01 | 1.00e+00 | 822 | 535 | 287 |
| Latino/Mixed-American | No Recorded Depression | Discontinuation vs Continuation | Stress | Education_levelCollege Undergraduate | 1.564 | 0.904 | 2.707 | 1.10e-01 | 1.00e+00 | 822 | 535 | 287 |
| Latino/Mixed-American | No Recorded Depression | Discontinuation vs Continuation | Stress | Education_levelTwelve or GED | 1.410 | 0.766 | 2.596 | 2.70e-01 | 1.00e+00 | 822 | 535 | 287 |
| Latino/Mixed-American | No Recorded Depression | Discontinuation vs Continuation | Stress | StressAlmost Never | 0.705 | 0.345 | 1.443 | 3.39e-01 | 1.00e+00 | 822 | 535 | 287 |
| Latino/Mixed-American | No Recorded Depression | Discontinuation vs Continuation | Stress | StressFairly Often | 1.100 | 0.579 | 2.089 | 7.71e-01 | 1.00e+00 | 822 | 535 | 287 |
| Latino/Mixed-American | No Recorded Depression | Discontinuation vs Continuation | Stress | StressSometimes | 0.851 | 0.613 | 1.179 | 3.32e-01 | 1.00e+00 | 822 | 535 | 287 |
| Latino/Mixed-American | No Recorded Depression | Discontinuation vs Continuation | Stress | StressVery Often | 0.520 | 0.199 | 1.363 | 1.84e-01 | 1.00e+00 | 822 | 535 | 287 |
| Latino/Mixed-American | No Recorded Depression | Switching vs Continuation | Stress | (Intercept) | 0.246 | 0.129 | 0.467 | 1.89e-05 | 2.65e-04 | 696 | 535 | 161 |
| Latino/Mixed-American | No Recorded Depression | Switching vs Continuation | Stress | age_at_first_AD | 0.721 | 0.586 | 0.887 | 2.00e-03 | 2.79e-02 | 696 | 535 | 161 |
| Latino/Mixed-American | No Recorded Depression | Switching vs Continuation | Stress | sex_at_birthMale | 1.392 | 0.919 | 2.108 | 1.19e-01 | 1.00e+00 | 696 | 535 | 161 |
| Latino/Mixed-American | No Recorded Depression | Switching vs Continuation | Stress | Income>200k | 0.839 | 0.336 | 2.100 | 7.08e-01 | 1.00e+00 | 696 | 535 | 161 |
| Latino/Mixed-American | No Recorded Depression | Switching vs Continuation | Stress | Income100-150k | 1.107 | 0.588 | 2.087 | 7.52e-01 | 1.00e+00 | 696 | 535 | 161 |
| Latino/Mixed-American | No Recorded Depression | Switching vs Continuation | Stress | Income150-200k | 0.682 | 0.289 | 1.611 | 3.83e-01 | 1.00e+00 | 696 | 535 | 161 |
| Latino/Mixed-American | No Recorded Depression | Switching vs Continuation | Stress | Income25-35k | 0.670 | 0.334 | 1.344 | 2.60e-01 | 1.00e+00 | 696 | 535 | 161 |
| Latino/Mixed-American | No Recorded Depression | Switching vs Continuation | Stress | Income35-50k | 0.792 | 0.401 | 1.567 | 5.04e-01 | 1.00e+00 | 696 | 535 | 161 |
| Latino/Mixed-American | No Recorded Depression | Switching vs Continuation | Stress | Income50-75k | 0.727 | 0.399 | 1.323 | 2.96e-01 | 1.00e+00 | 696 | 535 | 161 |
| Latino/Mixed-American | No Recorded Depression | Switching vs Continuation | Stress | Income75-100k | 1.020 | 0.518 | 2.006 | 9.55e-01 | 1.00e+00 | 696 | 535 | 161 |
| Latino/Mixed-American | No Recorded Depression | Switching vs Continuation | Stress | Education_levelCollege Postgraduate | 0.787 | 0.334 | 1.851 | 5.82e-01 | 1.00e+00 | 696 | 535 | 161 |
| Latino/Mixed-American | No Recorded Depression | Switching vs Continuation | Stress | Education_levelCollege Undergraduate | 1.491 | 0.734 | 3.030 | 2.70e-01 | 1.00e+00 | 696 | 535 | 161 |
| Latino/Mixed-American | No Recorded Depression | Switching vs Continuation | Stress | Education_levelTwelve or GED | 1.114 | 0.488 | 2.542 | 7.98e-01 | 1.00e+00 | 696 | 535 | 161 |
| Latino/Mixed-American | No Recorded Depression | Switching vs Continuation | Stress | StressAlmost Never | 1.561 | 0.768 | 3.172 | 2.19e-01 | 1.00e+00 | 696 | 535 | 161 |
| Latino/Mixed-American | No Recorded Depression | Switching vs Continuation | Stress | StressFairly Often | 0.667 | 0.260 | 1.710 | 3.99e-01 | 1.00e+00 | 696 | 535 | 161 |
| Latino/Mixed-American | No Recorded Depression | Switching vs Continuation | Stress | StressSometimes | 0.841 | 0.562 | 1.258 | 3.99e-01 | 1.00e+00 | 696 | 535 | 161 |
| Latino/Mixed-American | No Recorded Depression | Switching vs Continuation | Stress | StressVery Often | 0.310 | 0.069 | 1.385 | 1.25e-01 | 1.00e+00 | 696 | 535 | 161 |
| Latino/Mixed-American | No Recorded Depression | Discontinuation vs Continuation | age_at_first_AD | (Intercept) | 0.561 | 0.503 | 0.626 | <2.2e-16 | <2.2e-16 | 2057 | 1277 | 780 |
| Latino/Mixed-American | No Recorded Depression | Discontinuation vs Continuation | age_at_first_AD | age_at_first_AD | 0.976 | 0.890 | 1.070 | 6.01e-01 | 1.00e+00 | 2057 | 1277 | 780 |
| Latino/Mixed-American | No Recorded Depression | Discontinuation vs Continuation | age_at_first_AD | sex_at_birthMale | 1.321 | 1.085 | 1.609 | 5.60e-03 | 7.84e-02 | 2057 | 1277 | 780 |
| Latino/Mixed-American | No Recorded Depression | Switching vs Continuation | age_at_first_AD | (Intercept) | 0.233 | 0.200 | 0.272 | <2.2e-16 | <2.2e-16 | 1622 | 1277 | 345 |
| Latino/Mixed-American | No Recorded Depression | Switching vs Continuation | age_at_first_AD | age_at_first_AD | 0.763 | 0.670 | 0.869 | 4.57e-05 | 6.40e-04 | 1622 | 1277 | 345 |
| Latino/Mixed-American | No Recorded Depression | Switching vs Continuation | age_at_first_AD | sex_at_birthMale | 1.240 | 0.948 | 1.624 | 1.17e-01 | 1.00e+00 | 1622 | 1277 | 345 |
| Latino/Mixed-American | No Recorded Depression | Discontinuation vs Continuation | sex_at_birth | (Intercept) | 0.561 | 0.503 | 0.626 | <2.2e-16 | <2.2e-16 | 2057 | 1277 | 780 |
| Latino/Mixed-American | No Recorded Depression | Discontinuation vs Continuation | sex_at_birth | age_at_first_AD | 0.976 | 0.890 | 1.070 | 6.01e-01 | 1.00e+00 | 2057 | 1277 | 780 |
| Latino/Mixed-American | No Recorded Depression | Discontinuation vs Continuation | sex_at_birth | sex_at_birthMale | 1.321 | 1.085 | 1.609 | 5.60e-03 | 7.84e-02 | 2057 | 1277 | 780 |
| Latino/Mixed-American | No Recorded Depression | Switching vs Continuation | sex_at_birth | (Intercept) | 0.233 | 0.200 | 0.272 | <2.2e-16 | <2.2e-16 | 1622 | 1277 | 345 |
| Latino/Mixed-American | No Recorded Depression | Switching vs Continuation | sex_at_birth | age_at_first_AD | 0.763 | 0.670 | 0.869 | 4.57e-05 | 6.40e-04 | 1622 | 1277 | 345 |

| | | | | | | | | | | | | |
|---|---|---|---|---|---|---|---|---|---|---|---|---|
| Latino/Mixed-American | No Recorded Depression | Switching vs Continuation | sex_at_birth | sex_at_birthMale | 1.240 | 0.948 | 1.624 | 1.17e-01 | 1.00e+00 | 1622 | 1277 | 345 |
| Latino/Mixed-American | Recorded Anxiety | Discontinuation vs Continuation | Alcohol_Frequency | (Intercept) | 0.415 | 0.305 | 0.565 | 2.35e-08 | 3.29e-07 | 2193 | 1434 | 759 |
| Latino/Mixed-American | Recorded Anxiety | Discontinuation vs Continuation | Alcohol_Frequency | age_at_first_AD | 1.027 | 0.930 | 1.134 | 6.00e-01 | 1.00e+00 | 2193 | 1434 | 759 |
| Latino/Mixed-American | Recorded Anxiety | Discontinuation vs Continuation | Alcohol_Frequency | sex_at_birthMale | 1.255 | 1.028 | 1.533 | 2.59e-02 | 3.63e-01 | 2193 | 1434 | 759 |
| Latino/Mixed-American | Recorded Anxiety | Discontinuation vs Continuation | Alcohol_Frequency | Income>200k | 0.557 | 0.268 | 1.158 | 1.17e-01 | 1.00e+00 | 2193 | 1434 | 759 |
| Latino/Mixed-American | Recorded Anxiety | Discontinuation vs Continuation | Alcohol_Frequency | Income100-150k | 0.587 | 0.391 | 0.881 | 1.00e-02 | 1.41e-01 | 2193 | 1434 | 759 |
| Latino/Mixed-American | Recorded Anxiety | Discontinuation vs Continuation | Alcohol_Frequency | Income150-200k | 0.851 | 0.476 | 1.519 | 5.85e-01 | 1.00e+00 | 2193 | 1434 | 759 |
| Latino/Mixed-American | Recorded Anxiety | Discontinuation vs Continuation | Alcohol_Frequency | Income25-35k | 0.832 | 0.623 | 1.110 | 2.11e-01 | 1.00e+00 | 2193 | 1434 | 759 |
| Latino/Mixed-American | Recorded Anxiety | Discontinuation vs Continuation | Alcohol_Frequency | Income35-50k | 0.884 | 0.648 | 1.206 | 4.38e-01 | 1.00e+00 | 2193 | 1434 | 759 |
| Latino/Mixed-American | Recorded Anxiety | Discontinuation vs Continuation | Alcohol_Frequency | Income50-75k | 0.761 | 0.562 | 1.030 | 7.66e-02 | 1.00e+00 | 2193 | 1434 | 759 |
| Latino/Mixed-American | Recorded Anxiety | Discontinuation vs Continuation | Alcohol_Frequency | Income75-100k | 0.822 | 0.551 | 1.227 | 3.38e-01 | 1.00e+00 | 2193 | 1434 | 759 |
| Latino/Mixed-American | Recorded Anxiety | Discontinuation vs Continuation | Alcohol_Frequency | Education_levelCollege Postgraduate | 0.994 | 0.651 | 1.518 | 9.78e-01 | 1.00e+00 | 2193 | 1434 | 759 |
| Latino/Mixed-American | Recorded Anxiety | Discontinuation vs Continuation | Alcohol_Frequency | Education_levelCollege Undergraduate | 1.279 | 0.975 | 1.677 | 7.54e-02 | 1.00e+00 | 2193 | 1434 | 759 |
| Latino/Mixed-American | Recorded Anxiety | Discontinuation vs Continuation | Alcohol_Frequency | Education_levelTwelve or GED | 1.162 | 0.873 | 1.547 | 3.02e-01 | 1.00e+00 | 2193 | 1434 | 759 |
| Latino/Mixed-American | Recorded Anxiety | Discontinuation vs Continuation | Alcohol_Frequency | Alcohol_Frequency2-3 Week | 1.047 | 0.681 | 1.610 | 8.35e-01 | 1.00e+00 | 2193 | 1434 | 759 |
| Latino/Mixed-American | Recorded Anxiety | Discontinuation vs Continuation | Alcohol_Frequency | Alcohol_Frequency2-4 Month | 1.040 | 0.729 | 1.483 | 8.31e-01 | 1.00e+00 | 2193 | 1434 | 759 |
| Latino/Mixed-American | Recorded Anxiety | Discontinuation vs Continuation | Alcohol_Frequency | Alcohol_Frequency4plus Week | 1.649 | 1.019 | 2.668 | 4.16e-02 | 5.83e-01 | 2193 | 1434 | 759 |
| Latino/Mixed-American | Recorded Anxiety | Discontinuation vs Continuation | Alcohol_Frequency | Alcohol_FrequencyMonthly or Less | 1.248 | 0.922 | 1.689 | 1.52e-01 | 1.00e+00 | 2193 | 1434 | 759 |
| Latino/Mixed-American | Recorded Anxiety | Discontinuation vs Continuation | Alcohol_Frequency | Alcohol_FrequencyNever (former) | 1.241 | 0.895 | 1.722 | 1.96e-01 | 1.00e+00 | 2193 | 1434 | 759 |
| Latino/Mixed-American | Recorded Anxiety | Switching vs Continuation | Alcohol_Frequency | (Intercept) | 0.916 | 0.715 | 1.175 | 4.90e-01 | 1.00e+00 | 2645 | 1434 | 1211 |
| Latino/Mixed-American | Recorded Anxiety | Switching vs Continuation | Alcohol_Frequency | age_at_first_AD | 0.861 | 0.787 | 0.942 | 1.16e-03 | 1.62e-02 | 2645 | 1434 | 1211 |
| Latino/Mixed-American | Recorded Anxiety | Switching vs Continuation | Alcohol_Frequency | sex_at_birthMale | 1.065 | 0.890 | 1.275 | 4.93e-01 | 1.00e+00 | 2645 | 1434 | 1211 |
| Latino/Mixed-American | Recorded Anxiety | Switching vs Continuation | Alcohol_Frequency | Income>200k | 0.721 | 0.404 | 1.286 | 2.68e-01 | 1.00e+00 | 2645 | 1434 | 1211 |
| Latino/Mixed-American | Recorded Anxiety | Switching vs Continuation | Alcohol_Frequency | Income100-150k | 0.805 | 0.579 | 1.121 | 2.00e-01 | 1.00e+00 | 2645 | 1434 | 1211 |
| Latino/Mixed-American | Recorded Anxiety | Switching vs Continuation | Alcohol_Frequency | Income150-200k | 0.872 | 0.523 | 1.454 | 5.98e-01 | 1.00e+00 | 2645 | 1434 | 1211 |
| Latino/Mixed-American | Recorded Anxiety | Switching vs Continuation | Alcohol_Frequency | Income25-35k | 0.899 | 0.697 | 1.159 | 4.12e-01 | 1.00e+00 | 2645 | 1434 | 1211 |
| Latino/Mixed-American | Recorded Anxiety | Switching vs Continuation | Alcohol_Frequency | Income35-50k | 1.119 | 0.856 | 1.461 | 4.11e-01 | 1.00e+00 | 2645 | 1434 | 1211 |
| Latino/Mixed-American | Recorded Anxiety | Switching vs Continuation | Alcohol_Frequency | Income50-75k | 0.859 | 0.660 | 1.118 | 2.59e-01 | 1.00e+00 | 2645 | 1434 | 1211 |
| Latino/Mixed-American | Recorded Anxiety | Switching vs Continuation | Alcohol_Frequency | Income75-100k | 1.054 | 0.748 | 1.486 | 7.63e-01 | 1.00e+00 | 2645 | 1434 | 1211 |
| Latino/Mixed-American | Recorded Anxiety | Switching vs Continuation | Alcohol_Frequency | Education_levelCollege Postgraduate | 1.069 | 0.757 | 1.508 | 7.06e-01 | 1.00e+00 | 2645 | 1434 | 1211 |
| Latino/Mixed-American | Recorded Anxiety | Switching vs Continuation | Alcohol_Frequency | Education_levelCollege Undergraduate | 1.069 | 0.843 | 1.356 | 5.82e-01 | 1.00e+00 | 2645 | 1434 | 1211 |
| Latino/Mixed-American | Recorded Anxiety | Switching vs Continuation | Alcohol_Frequency | Education_levelTwelve or GED | 0.849 | 0.659 | 1.094 | 2.06e-01 | 1.00e+00 | 2645 | 1434 | 1211 |
| Latino/Mixed-American | Recorded Anxiety | Switching vs Continuation | Alcohol_Frequency | Alcohol_Frequency2-3 Week | 0.854 | 0.598 | 1.219 | 3.84e-01 | 1.00e+00 | 2645 | 1434 | 1211 |
| Latino/Mixed-American | Recorded Anxiety | Switching vs Continuation | Alcohol_Frequency | Alcohol_Frequency2-4 Month | 0.716 | 0.531 | 0.967 | 2.91e-02 | 4.08e-01 | 2645 | 1434 | 1211 |
| Latino/Mixed-American | Recorded Anxiety | Switching vs Continuation | Alcohol_Frequency | Alcohol_Frequency4plus Week | 1.139 | 0.748 | 1.736 | 5.44e-01 | 1.00e+00 | 2645 | 1434 | 1211 |
| Latino/Mixed-American | Recorded Anxiety | Switching vs Continuation | Alcohol_Frequency | Alcohol_FrequencyMonthly or Less | 0.823 | 0.637 | 1.062 | 1.34e-01 | 1.00e+00 | 2645 | 1434 | 1211 |
| Latino/Mixed-American | Recorded Anxiety | Switching vs Continuation | Alcohol_Frequency | Alcohol_FrequencyNever (former) | 1.049 | 0.797 | 1.380 | 7.34e-01 | 1.00e+00 | 2645 | 1434 | 1211 |
| Latino/Mixed-American | Recorded Anxiety | Discontinuation vs Continuation | BMI | (Intercept) | 0.447 | 0.356 | 0.560 | 3.40e-12 | 4.76e-11 | 2137 | 1419 | 718 |
| Latino/Mixed-American | Recorded Anxiety | Discontinuation vs Continuation | BMI | age_at_first_AD | 1.021 | 0.924 | 1.128 | 6.88e-01 | 1.00e+00 | 2137 | 1419 | 718 |
| Latino/Mixed-American | Recorded Anxiety | Discontinuation vs Continuation | BMI | sex_at_birthMale | 1.305 | 1.068 | 1.595 | 9.18e-03 | 1.29e-01 | 2137 | 1419 | 718 |
| Latino/Mixed-American | Recorded Anxiety | Discontinuation vs Continuation | BMI | Income>200k | 0.587 | 0.284 | 1.212 | 1.50e-01 | 1.00e+00 | 2137 | 1419 | 718 |
| Latino/Mixed-American | Recorded Anxiety | Discontinuation vs Continuation | BMI | Income100-150k | 0.594 | 0.395 | 0.894 | 1.25e-02 | 1.75e-01 | 2137 | 1419 | 718 |
| Latino/Mixed-American | Recorded Anxiety | Discontinuation vs Continuation | BMI | Income150-200k | 0.850 | 0.467 | 1.548 | 5.96e-01 | 1.00e+00 | 2137 | 1419 | 718 |
| Latino/Mixed-American | Recorded Anxiety | Discontinuation vs Continuation | BMI | Income25-35k | 0.832 | 0.618 | 1.119 | 2.24e-01 | 1.00e+00 | 2137 | 1419 | 718 |
| Latino/Mixed-American | Recorded Anxiety | Discontinuation vs Continuation | BMI | Income35-50k | 0.960 | 0.707 | 1.304 | 7.96e-01 | 1.00e+00 | 2137 | 1419 | 718 |
| Latino/Mixed-American | Recorded Anxiety | Discontinuation vs Continuation | BMI | Income50-75k | 0.732 | 0.536 | 0.999 | 4.92e-02 | 6.89e-01 | 2137 | 1419 | 718 |
| Latino/Mixed-American | Recorded Anxiety | Discontinuation vs Continuation | BMI | Income75-100k | 0.768 | 0.511 | 1.155 | 2.05e-01 | 1.00e+00 | 2137 | 1419 | 718 |
| Latino/Mixed-American | Recorded Anxiety | Discontinuation vs Continuation | BMI | Education_levelCollege Postgraduate | 1.041 | 0.675 | 1.605 | 8.55e-01 | 1.00e+00 | 2137 | 1419 | 718 |
| Latino/Mixed-American | Recorded Anxiety | Discontinuation vs Continuation | BMI | Education_levelCollege Undergraduate | 1.316 | 0.999 | 1.734 | 5.07e-02 | 7.09e-01 | 2137 | 1419 | 718 |
| Latino/Mixed-American | Recorded Anxiety | Discontinuation vs Continuation | BMI | Education_levelTwelve or GED | 1.201 | 0.899 | 1.605 | 2.14e-01 | 1.00e+00 | 2137 | 1419 | 718 |
| Latino/Mixed-American | Recorded Anxiety | Discontinuation vs Continuation | BMI | BMI | 1.020 | 0.933 | 1.115 | 6.71e-01 | 1.00e+00 | 2137 | 1419 | 718 |
| Latino/Mixed-American | Recorded Anxiety | Switching vs Continuation | BMI | (Intercept) | 0.832 | 0.691 | 1.002 | 5.20e-02 | 7.28e-01 | 2623 | 1419 | 1204 |
| Latino/Mixed-American | Recorded Anxiety | Switching vs Continuation | BMI | age_at_first_AD | 0.899 | 0.822 | 0.982 | 1.81e-02 | 2.54e-01 | 2623 | 1419 | 1204 |
| Latino/Mixed-American | Recorded Anxiety | Switching vs Continuation | BMI | sex_at_birthMale | 1.080 | 0.904 | 1.291 | 3.97e-01 | 1.00e+00 | 2623 | 1419 | 1204 |
| Latino/Mixed-American | Recorded Anxiety | Switching vs Continuation | BMI | Income>200k | 0.722 | 0.410 | 1.269 | 2.57e-01 | 1.00e+00 | 2623 | 1419 | 1204 |
| Latino/Mixed-American | Recorded Anxiety | Switching vs Continuation | BMI | Income100-150k | 0.772 | 0.556 | 1.071 | 1.21e-01 | 1.00e+00 | 2623 | 1419 | 1204 |
| Latino/Mixed-American | Recorded Anxiety | Switching vs Continuation | BMI | Income150-200k | 0.839 | 0.498 | 1.415 | 5.11e-01 | 1.00e+00 | 2623 | 1419 | 1204 |
| Latino/Mixed-American | Recorded Anxiety | Switching vs Continuation | BMI | Income25-35k | 0.929 | 0.720 | 1.199 | 5.73e-01 | 1.00e+00 | 2623 | 1419 | 1204 |
| Latino/Mixed-American | Recorded Anxiety | Switching vs Continuation | BMI | Income35-50k | 1.114 | 0.856 | 1.452 | 4.22e-01 | 1.00e+00 | 2623 | 1419 | 1204 |
| Latino/Mixed-American | Recorded Anxiety | Switching vs Continuation | BMI | Income50-75k | 0.866 | 0.666 | 1.125 | 2.81e-01 | 1.00e+00 | 2623 | 1419 | 1204 |
| Latino/Mixed-American | Recorded Anxiety | Switching vs Continuation | BMI | Income75-100k | 0.984 | 0.702 | 1.379 | 9.26e-01 | 1.00e+00 | 2623 | 1419 | 1204 |
| Latino/Mixed-American | Recorded Anxiety | Switching vs Continuation | BMI | Education_levelCollege Postgraduate | 1.054 | 0.747 | 1.488 | 7.64e-01 | 1.00e+00 | 2623 | 1419 | 1204 |
| Latino/Mixed-American | Recorded Anxiety | Switching vs Continuation | BMI | Education_levelCollege Undergraduate | 1.049 | 0.828 | 1.328 | 6.93e-01 | 1.00e+00 | 2623 | 1419 | 1204 |
| Latino/Mixed-American | Recorded Anxiety | Switching vs Continuation | BMI | Education_levelTwelve or GED | 0.862 | 0.670 | 1.109 | 2.49e-01 | 1.00e+00 | 2623 | 1419 | 1204 |
| Latino/Mixed-American | Recorded Anxiety | Switching vs Continuation | BMI | BMI | 1.023 | 0.945 | 1.108 | 5.69e-01 | 1.00e+00 | 2623 | 1419 | 1204 |
| Latino/Mixed-American | Recorded Anxiety | Discontinuation vs Continuation | Education_level | (Intercept) | 0.492 | 0.396 | 0.612 | 1.79e-10 | 2.51e-09 | 2246 | 1462 | 784 |

| | | | | | | | | | | | | |
|---|---|---|---|---|---|---|---|---|---|---|---|---|
| Latino/Mixed-American | Recorded Anxiety | Discontinuation vs Continuation | Education_level | age_at_first_AD | 1.026 | 0.933 | 1.130 | 5.95e-01 | 1.00e+00 | 2246 | 1462 | 784 |
| Latino/Mixed-American | Recorded Anxiety | Discontinuation vs Continuation | Education_level | sex_at_birthMale | 1.292 | 1.065 | 1.568 | 9.49e-03 | 1.33e-01 | 2246 | 1462 | 784 |
| Latino/Mixed-American | Recorded Anxiety | Discontinuation vs Continuation | Education_level | Income>200k | 0.556 | 0.270 | 1.144 | 1.11e-01 | 1.00e+00 | 2246 | 1462 | 784 |
| Latino/Mixed-American | Recorded Anxiety | Discontinuation vs Continuation | Education_level | Income100-150k | 0.580 | 0.390 | 0.862 | 7.15e-03 | 1.00e-01 | 2246 | 1462 | 784 |
| Latino/Mixed-American | Recorded Anxiety | Discontinuation vs Continuation | Education_level | Income150-200k | 0.844 | 0.476 | 1.497 | 5.62e-01 | 1.00e+00 | 2246 | 1462 | 784 |
| Latino/Mixed-American | Recorded Anxiety | Discontinuation vs Continuation | Education_level | Income25-35k | 0.843 | 0.635 | 1.119 | 2.38e-01 | 1.00e+00 | 2246 | 1462 | 784 |
| Latino/Mixed-American | Recorded Anxiety | Discontinuation vs Continuation | Education_level | Income35-50k | 0.917 | 0.681 | 1.236 | 5.71e-01 | 1.00e+00 | 2246 | 1462 | 784 |
| Latino/Mixed-American | Recorded Anxiety | Discontinuation vs Continuation | Education_level | Income50-75k | 0.750 | 0.557 | 1.010 | 5.79e-02 | 8.11e-01 | 2246 | 1462 | 784 |
| Latino/Mixed-American | Recorded Anxiety | Discontinuation vs Continuation | Education_level | Income75-100k | 0.830 | 0.563 | 1.225 | 3.49e-01 | 1.00e+00 | 2246 | 1462 | 784 |
| Latino/Mixed-American | Recorded Anxiety | Discontinuation vs Continuation | Education_level | Education_levelCollege Postgraduate | 0.990 | 0.652 | 1.503 | 9.63e-01 | 1.00e+00 | 2246 | 1462 | 784 |
| Latino/Mixed-American | Recorded Anxiety | Discontinuation vs Continuation | Education_level | Education_levelCollege Undergraduate | 1.269 | 0.974 | 1.655 | 7.78e-02 | 1.00e+00 | 2246 | 1462 | 784 |
| Latino/Mixed-American | Recorded Anxiety | Discontinuation vs Continuation | Education_level | Education_levelTwelve or GED | 1.147 | 0.867 | 1.517 | 3.37e-01 | 1.00e+00 | 2246 | 1462 | 784 |
| Latino/Mixed-American | Recorded Anxiety | Switching vs Continuation | Education_level | (Intercept) | 0.854 | 0.712 | 1.026 | 9.19e-02 | 1.00e+00 | 2697 | 1462 | 1235 |
| Latino/Mixed-American | Recorded Anxiety | Switching vs Continuation | Education_level | age_at_first_AD | 0.897 | 0.822 | 0.979 | 1.46e-02 | 2.04e-01 | 2697 | 1462 | 1235 |
| Latino/Mixed-American | Recorded Anxiety | Switching vs Continuation | Education_level | sex_at_birthMale | 1.088 | 0.913 | 1.297 | 3.45e-01 | 1.00e+00 | 2697 | 1462 | 1235 |
| Latino/Mixed-American | Recorded Anxiety | Switching vs Continuation | Education_level | Income>200k | 0.713 | 0.406 | 1.252 | 2.39e-01 | 1.00e+00 | 2697 | 1462 | 1235 |
| Latino/Mixed-American | Recorded Anxiety | Switching vs Continuation | Education_level | Income100-150k | 0.789 | 0.572 | 1.088 | 1.48e-01 | 1.00e+00 | 2697 | 1462 | 1235 |
| Latino/Mixed-American | Recorded Anxiety | Switching vs Continuation | Education_level | Income150-200k | 0.850 | 0.512 | 1.412 | 5.31e-01 | 1.00e+00 | 2697 | 1462 | 1235 |
| Latino/Mixed-American | Recorded Anxiety | Switching vs Continuation | Education_level | Income25-35k | 0.899 | 0.700 | 1.156 | 4.08e-01 | 1.00e+00 | 2697 | 1462 | 1235 |
| Latino/Mixed-American | Recorded Anxiety | Switching vs Continuation | Education_level | Income35-50k | 1.109 | 0.855 | 1.438 | 4.35e-01 | 1.00e+00 | 2697 | 1462 | 1235 |
| Latino/Mixed-American | Recorded Anxiety | Switching vs Continuation | Education_level | Income50-75k | 0.842 | 0.650 | 1.090 | 1.92e-01 | 1.00e+00 | 2697 | 1462 | 1235 |
| Latino/Mixed-American | Recorded Anxiety | Switching vs Continuation | Education_level | Income75-100k | 0.994 | 0.711 | 1.388 | 9.71e-01 | 1.00e+00 | 2697 | 1462 | 1235 |
| Latino/Mixed-American | Recorded Anxiety | Switching vs Continuation | Education_level | Education_levelCollege Postgraduate | 1.035 | 0.738 | 1.452 | 8.43e-01 | 1.00e+00 | 2697 | 1462 | 1235 |
| Latino/Mixed-American | Recorded Anxiety | Switching vs Continuation | Education_level | Education_levelCollege Undergraduate | 1.021 | 0.809 | 1.287 | 8.64e-01 | 1.00e+00 | 2697 | 1462 | 1235 |
| Latino/Mixed-American | Recorded Anxiety | Switching vs Continuation | Education_level | Education_levelTwelve or GED | 0.824 | 0.642 | 1.057 | 1.28e-01 | 1.00e+00 | 2697 | 1462 | 1235 |
| Latino/Mixed-American | Recorded Anxiety | Discontinuation vs Continuation | Income | (Intercept) | 0.492 | 0.396 | 0.612 | 1.79e-10 | 2.51e-09 | 2246 | 1462 | 784 |
| Latino/Mixed-American | Recorded Anxiety | Discontinuation vs Continuation | Income | age_at_first_AD | 1.026 | 0.933 | 1.130 | 5.95e-01 | 1.00e+00 | 2246 | 1462 | 784 |
| Latino/Mixed-American | Recorded Anxiety | Discontinuation vs Continuation | Income | sex_at_birthMale | 1.292 | 1.065 | 1.568 | 9.49e-03 | 1.33e-01 | 2246 | 1462 | 784 |
| Latino/Mixed-American | Recorded Anxiety | Discontinuation vs Continuation | Income | Income>200k | 0.556 | 0.270 | 1.144 | 1.11e-01 | 1.00e+00 | 2246 | 1462 | 784 |
| Latino/Mixed-American | Recorded Anxiety | Discontinuation vs Continuation | Income | Income100-150k | 0.580 | 0.390 | 0.862 | 7.15e-03 | 1.00e-01 | 2246 | 1462 | 784 |
| Latino/Mixed-American | Recorded Anxiety | Discontinuation vs Continuation | Income | Income150-200k | 0.844 | 0.476 | 1.497 | 5.62e-01 | 1.00e+00 | 2246 | 1462 | 784 |
| Latino/Mixed-American | Recorded Anxiety | Discontinuation vs Continuation | Income | Income25-35k | 0.843 | 0.635 | 1.119 | 2.38e-01 | 1.00e+00 | 2246 | 1462 | 784 |
| Latino/Mixed-American | Recorded Anxiety | Discontinuation vs Continuation | Income | Income35-50k | 0.917 | 0.681 | 1.236 | 5.71e-01 | 1.00e+00 | 2246 | 1462 | 784 |
| Latino/Mixed-American | Recorded Anxiety | Discontinuation vs Continuation | Income | Income50-75k | 0.750 | 0.557 | 1.010 | 5.79e-02 | 8.11e-01 | 2246 | 1462 | 784 |
| Latino/Mixed-American | Recorded Anxiety | Discontinuation vs Continuation | Income | Income75-100k | 0.830 | 0.563 | 1.225 | 3.49e-01 | 1.00e+00 | 2246 | 1462 | 784 |
| Latino/Mixed-American | Recorded Anxiety | Discontinuation vs Continuation | Income | Education_levelCollege Postgraduate | 0.990 | 0.652 | 1.503 | 9.63e-01 | 1.00e+00 | 2246 | 1462 | 784 |
| Latino/Mixed-American | Recorded Anxiety | Discontinuation vs Continuation | Income | Education_levelCollege Undergraduate | 1.269 | 0.974 | 1.655 | 7.78e-02 | 1.00e+00 | 2246 | 1462 | 784 |
| Latino/Mixed-American | Recorded Anxiety | Discontinuation vs Continuation | Income | Education_levelTwelve or GED | 1.147 | 0.867 | 1.517 | 3.37e-01 | 1.00e+00 | 2246 | 1462 | 784 |
| Latino/Mixed-American | Recorded Anxiety | Switching vs Continuation | Income | (Intercept) | 0.854 | 0.712 | 1.026 | 9.19e-02 | 1.00e+00 | 2697 | 1462 | 1235 |
| Latino/Mixed-American | Recorded Anxiety | Switching vs Continuation | Income | age_at_first_AD | 0.897 | 0.822 | 0.979 | 1.46e-02 | 2.04e-01 | 2697 | 1462 | 1235 |
| Latino/Mixed-American | Recorded Anxiety | Switching vs Continuation | Income | sex_at_birthMale | 1.088 | 0.913 | 1.297 | 3.45e-01 | 1.00e+00 | 2697 | 1462 | 1235 |
| Latino/Mixed-American | Recorded Anxiety | Switching vs Continuation | Income | Income>200k | 0.713 | 0.406 | 1.252 | 2.39e-01 | 1.00e+00 | 2697 | 1462 | 1235 |
| Latino/Mixed-American | Recorded Anxiety | Switching vs Continuation | Income | Income100-150k | 0.789 | 0.572 | 1.088 | 1.48e-01 | 1.00e+00 | 2697 | 1462 | 1235 |
| Latino/Mixed-American | Recorded Anxiety | Switching vs Continuation | Income | Income150-200k | 0.850 | 0.512 | 1.412 | 5.31e-01 | 1.00e+00 | 2697 | 1462 | 1235 |
| Latino/Mixed-American | Recorded Anxiety | Switching vs Continuation | Income | Income25-35k | 0.899 | 0.700 | 1.156 | 4.08e-01 | 1.00e+00 | 2697 | 1462 | 1235 |
| Latino/Mixed-American | Recorded Anxiety | Switching vs Continuation | Income | Income35-50k | 1.109 | 0.855 | 1.438 | 4.35e-01 | 1.00e+00 | 2697 | 1462 | 1235 |
| Latino/Mixed-American | Recorded Anxiety | Switching vs Continuation | Income | Income50-75k | 0.842 | 0.650 | 1.090 | 1.92e-01 | 1.00e+00 | 2697 | 1462 | 1235 |
| Latino/Mixed-American | Recorded Anxiety | Switching vs Continuation | Income | Income75-100k | 0.994 | 0.711 | 1.388 | 9.71e-01 | 1.00e+00 | 2697 | 1462 | 1235 |
| Latino/Mixed-American | Recorded Anxiety | Switching vs Continuation | Income | Education_levelCollege Postgraduate | 1.035 | 0.738 | 1.452 | 8.43e-01 | 1.00e+00 | 2697 | 1462 | 1235 |
| Latino/Mixed-American | Recorded Anxiety | Switching vs Continuation | Income | Education_levelCollege Undergraduate | 1.021 | 0.809 | 1.287 | 8.64e-01 | 1.00e+00 | 2697 | 1462 | 1235 |
| Latino/Mixed-American | Recorded Anxiety | Switching vs Continuation | Income | Education_levelTwelve or GED | 0.824 | 0.642 | 1.057 | 1.28e-01 | 1.00e+00 | 2697 | 1462 | 1235 |
| Latino/Mixed-American | Recorded Anxiety | Discontinuation vs Continuation | Insomnia | (Intercept) | 0.505 | 0.403 | 0.631 | 2.22e-09 | 3.11e-08 | 2246 | 1462 | 784 |
| Latino/Mixed-American | Recorded Anxiety | Discontinuation vs Continuation | Insomnia | age_at_first_AD | 1.033 | 0.938 | 1.138 | 5.14e-01 | 1.00e+00 | 2246 | 1462 | 784 |
| Latino/Mixed-American | Recorded Anxiety | Discontinuation vs Continuation | Insomnia | sex_at_birthMale | 1.296 | 1.067 | 1.573 | 8.79e-03 | 1.23e-01 | 2246 | 1462 | 784 |
| Latino/Mixed-American | Recorded Anxiety | Discontinuation vs Continuation | Insomnia | Income>200k | 0.556 | 0.270 | 1.144 | 1.11e-01 | 1.00e+00 | 2246 | 1462 | 784 |
| Latino/Mixed-American | Recorded Anxiety | Discontinuation vs Continuation | Insomnia | Income100-150k | 0.578 | 0.389 | 0.861 | 6.93e-03 | 9.70e-02 | 2246 | 1462 | 784 |
| Latino/Mixed-American | Recorded Anxiety | Discontinuation vs Continuation | Insomnia | Income150-200k | 0.834 | 0.470 | 1.479 | 5.34e-01 | 1.00e+00 | 2246 | 1462 | 784 |
| Latino/Mixed-American | Recorded Anxiety | Discontinuation vs Continuation | Insomnia | Income25-35k | 0.841 | 0.633 | 1.117 | 2.31e-01 | 1.00e+00 | 2246 | 1462 | 784 |
| Latino/Mixed-American | Recorded Anxiety | Discontinuation vs Continuation | Insomnia | Income35-50k | 0.914 | 0.678 | 1.232 | 5.55e-01 | 1.00e+00 | 2246 | 1462 | 784 |
| Latino/Mixed-American | Recorded Anxiety | Discontinuation vs Continuation | Insomnia | Income50-75k | 0.750 | 0.557 | 1.010 | 5.80e-02 | 8.13e-01 | 2246 | 1462 | 784 |
| Latino/Mixed-American | Recorded Anxiety | Discontinuation vs Continuation | Insomnia | Income75-100k | 0.829 | 0.562 | 1.223 | 3.44e-01 | 1.00e+00 | 2246 | 1462 | 784 |
| Latino/Mixed-American | Recorded Anxiety | Discontinuation vs Continuation | Insomnia | Education_levelCollege Postgraduate | 0.989 | 0.651 | 1.501 | 9.58e-01 | 1.00e+00 | 2246 | 1462 | 784 |
| Latino/Mixed-American | Recorded Anxiety | Discontinuation vs Continuation | Insomnia | Education_levelCollege Undergraduate | 1.274 | 0.978 | 1.662 | 7.32e-02 | 1.00e+00 | 2246 | 1462 | 784 |
| Latino/Mixed-American | Recorded Anxiety | Discontinuation vs Continuation | Insomnia | Education_levelTwelve or GED | 1.150 | 0.869 | 1.521 | 3.29e-01 | 1.00e+00 | 2246 | 1462 | 784 |
| Latino/Mixed-American | Recorded Anxiety | Discontinuation vs Continuation | Insomnia | Insomnia | 0.913 | 0.746 | 1.116 | 3.73e-01 | 1.00e+00 | 2246 | 1462 | 784 |
| Latino/Mixed-American | Recorded Anxiety | Switching vs Continuation | Insomnia | (Intercept) | 0.609 | 0.500 | 0.742 | 7.89e-07 | 1.10e-05 | 2697 | 1462 | 1235 |

| | | | | | | | | | | | | |
|---|---|---|---|---|---|---|---|---|---|---|---|---|
| Latino/Mixed-American | Recorded Anxiety | Switching vs Continuation | Insomnia | age_at_first_AD | 0.836 | 0.763 | 0.915 | 1.02e-04 | 1.43e-03 | 2697 | 1462 | 1235 |
| Latino/Mixed-American | Recorded Anxiety | Switching vs Continuation | Insomnia | sex_at_birthMale | 1.055 | 0.881 | 1.263 | 5.59e-01 | 1.00e+00 | 2697 | 1462 | 1235 |
| Latino/Mixed-American | Recorded Anxiety | Switching vs Continuation | Insomnia | Income>200k | 0.727 | 0.409 | 1.292 | 2.77e-01 | 1.00e+00 | 2697 | 1462 | 1235 |
| Latino/Mixed-American | Recorded Anxiety | Switching vs Continuation | Insomnia | Income100-150k | 0.761 | 0.547 | 1.058 | 1.04e-01 | 1.00e+00 | 2697 | 1462 | 1235 |
| Latino/Mixed-American | Recorded Anxiety | Switching vs Continuation | Insomnia | Income150-200k | 0.934 | 0.556 | 1.567 | 7.95e-01 | 1.00e+00 | 2697 | 1462 | 1235 |
| Latino/Mixed-American | Recorded Anxiety | Switching vs Continuation | Insomnia | Income25-35k | 0.909 | 0.703 | 1.176 | 4.67e-01 | 1.00e+00 | 2697 | 1462 | 1235 |
| Latino/Mixed-American | Recorded Anxiety | Switching vs Continuation | Insomnia | Income35-50k | 1.097 | 0.840 | 1.431 | 4.97e-01 | 1.00e+00 | 2697 | 1462 | 1235 |
| Latino/Mixed-American | Recorded Anxiety | Switching vs Continuation | Insomnia | Income50-75k | 0.849 | 0.651 | 1.106 | 2.25e-01 | 1.00e+00 | 2697 | 1462 | 1235 |
| Latino/Mixed-American | Recorded Anxiety | Switching vs Continuation | Insomnia | Income75-100k | 1.003 | 0.712 | 1.412 | 9.87e-01 | 1.00e+00 | 2697 | 1462 | 1235 |
| Latino/Mixed-American | Recorded Anxiety | Switching vs Continuation | Insomnia | Education_levelCollege Postgraduate | 1.051 | 0.743 | 1.486 | 7.78e-01 | 1.00e+00 | 2697 | 1462 | 1235 |
| Latino/Mixed-American | Recorded Anxiety | Switching vs Continuation | Insomnia | Education_levelCollege Undergraduate | 0.982 | 0.774 | 1.246 | 8.79e-01 | 1.00e+00 | 2697 | 1462 | 1235 |
| Latino/Mixed-American | Recorded Anxiety | Switching vs Continuation | Insomnia | Education_levelTwelve or GED | 0.790 | 0.612 | 1.020 | 7.07e-02 | 9.89e-01 | 2697 | 1462 | 1235 |
| Latino/Mixed-American | Recorded Anxiety | Switching vs Continuation | Insomnia | Insomnia | 2.539 | 2.155 | 2.991 | <2.2e-16 | <2.2e-16 | 2697 | 1462 | 1235 |
| Latino/Mixed-American | Recorded Anxiety | Discontinuation vs Continuation | MDD CIDI-sf | (Intercept) | 0.232 | 0.020 | 2.741 | 2.46e-01 | 1.00e+00 | 160 | 117 | 43 |
| Latino/Mixed-American | Recorded Anxiety | Discontinuation vs Continuation | MDD CIDI-sf | age_at_first_AD | 0.915 | 0.564 | 1.483 | 7.17e-01 | 1.00e+00 | 160 | 117 | 43 |
| Latino/Mixed-American | Recorded Anxiety | Discontinuation vs Continuation | MDD CIDI-sf | sex_at_birthMale | 1.559 | 0.654 | 3.714 | 3.16e-01 | 1.00e+00 | 160 | 117 | 43 |
| Latino/Mixed-American | Recorded Anxiety | Discontinuation vs Continuation | MDD CIDI-sf | Income>200k | 2.825 | 0.373 | 21.410 | 3.15e-01 | 1.00e+00 | 160 | 117 | 43 |
| Latino/Mixed-American | Recorded Anxiety | Discontinuation vs Continuation | MDD CIDI-sf | Income100-150k | 0.903 | 0.261 | 3.119 | 8.71e-01 | 1.00e+00 | 160 | 117 | 43 |
| Latino/Mixed-American | Recorded Anxiety | Discontinuation vs Continuation | MDD CIDI-sf | Income150-200k | 0.432 | 0.042 | 4.445 | 4.80e-01 | 1.00e+00 | 160 | 117 | 43 |
| Latino/Mixed-American | Recorded Anxiety | Discontinuation vs Continuation | MDD CIDI-sf | Income25-35k | 1.506 | 0.449 | 5.055 | 5.08e-01 | 1.00e+00 | 160 | 117 | 43 |
| Latino/Mixed-American | Recorded Anxiety | Discontinuation vs Continuation | MDD CIDI-sf | Income35-50k | 1.118 | 0.301 | 4.149 | 8.67e-01 | 1.00e+00 | 160 | 117 | 43 |
| Latino/Mixed-American | Recorded Anxiety | Discontinuation vs Continuation | MDD CIDI-sf | Income50-75k | 0.550 | 0.152 | 1.984 | 3.61e-01 | 1.00e+00 | 160 | 117 | 43 |
| Latino/Mixed-American | Recorded Anxiety | Discontinuation vs Continuation | MDD CIDI-sf | Income75-100k | 0.482 | 0.107 | 2.168 | 3.42e-01 | 1.00e+00 | 160 | 117 | 43 |
| Latino/Mixed-American | Recorded Anxiety | Discontinuation vs Continuation | MDD CIDI-sf | Education_levelCollege Postgraduate | 0.912 | 0.075 | 11.080 | 9.42e-01 | 1.00e+00 | 160 | 117 | 43 |
| Latino/Mixed-American | Recorded Anxiety | Discontinuation vs Continuation | MDD CIDI-sf | Education_levelCollege Undergraduate | 0.888 | 0.079 | 9.995 | 9.23e-01 | 1.00e+00 | 160 | 117 | 43 |
| Latino/Mixed-American | Recorded Anxiety | Discontinuation vs Continuation | MDD CIDI-sf | Education_levelTwelve or GED | 1.800 | 0.156 | 20.800 | 6.38e-01 | 1.00e+00 | 160 | 117 | 43 |
| Latino/Mixed-American | Recorded Anxiety | Discontinuation vs Continuation | MDD CIDI-sf | MDD CIDI-sf | 1.720 | 0.692 | 4.274 | 2.43e-01 | 1.00e+00 | 160 | 117 | 43 |
| Latino/Mixed-American | Recorded Anxiety | Switching vs Continuation | MDD CIDI-sf | (Intercept) | 0.855 | 0.148 | 4.927 | 8.60e-01 | 1.00e+00 | 237 | 117 | 120 |
| Latino/Mixed-American | Recorded Anxiety | Switching vs Continuation | MDD CIDI-sf | age_at_first_AD | 0.759 | 0.539 | 1.068 | 1.14e-01 | 1.00e+00 | 237 | 117 | 120 |
| Latino/Mixed-American | Recorded Anxiety | Switching vs Continuation | MDD CIDI-sf | sex_at_birthMale | 1.479 | 0.751 | 2.914 | 2.58e-01 | 1.00e+00 | 237 | 117 | 120 |
| Latino/Mixed-American | Recorded Anxiety | Switching vs Continuation | MDD CIDI-sf | Income>200k | 1.630 | 0.323 | 8.230 | 5.55e-01 | 1.00e+00 | 237 | 117 | 120 |
| Latino/Mixed-American | Recorded Anxiety | Switching vs Continuation | MDD CIDI-sf | Income100-150k | 0.550 | 0.214 | 1.414 | 2.15e-01 | 1.00e+00 | 237 | 117 | 120 |
| Latino/Mixed-American | Recorded Anxiety | Switching vs Continuation | MDD CIDI-sf | Income150-200k | 0.750 | 0.197 | 2.864 | 6.74e-01 | 1.00e+00 | 237 | 117 | 120 |
| Latino/Mixed-American | Recorded Anxiety | Switching vs Continuation | MDD CIDI-sf | Income25-35k | 0.588 | 0.215 | 1.606 | 3.00e-01 | 1.00e+00 | 237 | 117 | 120 |
| Latino/Mixed-American | Recorded Anxiety | Switching vs Continuation | MDD CIDI-sf | Income35-50k | 0.645 | 0.229 | 1.815 | 4.06e-01 | 1.00e+00 | 237 | 117 | 120 |
| Latino/Mixed-American | Recorded Anxiety | Switching vs Continuation | MDD CIDI-sf | Income50-75k | 0.661 | 0.291 | 1.504 | 3.24e-01 | 1.00e+00 | 237 | 117 | 120 |
| Latino/Mixed-American | Recorded Anxiety | Switching vs Continuation | MDD CIDI-sf | Income75-100k | 0.704 | 0.280 | 1.773 | 4.57e-01 | 1.00e+00 | 237 | 117 | 120 |
| Latino/Mixed-American | Recorded Anxiety | Switching vs Continuation | MDD CIDI-sf | Education_levelCollege Postgraduate | 0.481 | 0.074 | 3.123 | 4.43e-01 | 1.00e+00 | 237 | 117 | 120 |
| Latino/Mixed-American | Recorded Anxiety | Switching vs Continuation | MDD CIDI-sf | Education_levelCollege Undergraduate | 0.851 | 0.143 | 5.055 | 8.59e-01 | 1.00e+00 | 237 | 117 | 120 |
| Latino/Mixed-American | Recorded Anxiety | Switching vs Continuation | MDD CIDI-sf | Education_levelTwelve or GED | 0.822 | 0.126 | 5.353 | 8.38e-01 | 1.00e+00 | 237 | 117 | 120 |
| Latino/Mixed-American | Recorded Anxiety | Switching vs Continuation | MDD CIDI-sf | MDD CIDI-sf | 2.113 | 1.097 | 4.070 | 2.54e-02 | 3.55e-01 | 237 | 117 | 120 |
| Latino/Mixed-American | Recorded Anxiety | Discontinuation vs Continuation | MDD_symptom_count | (Intercept) | 0.213 | 0.024 | 1.913 | 1.67e-01 | 1.00e+00 | 198 | 143 | 55 |
| Latino/Mixed-American | Recorded Anxiety | Discontinuation vs Continuation | MDD_symptom_count | age_at_first_AD | 0.807 | 0.531 | 1.226 | 3.15e-01 | 1.00e+00 | 198 | 143 | 55 |
| Latino/Mixed-American | Recorded Anxiety | Discontinuation vs Continuation | MDD_symptom_count | sex_at_birthMale | 1.932 | 0.898 | 4.156 | 9.21e-02 | 1.00e+00 | 198 | 143 | 55 |
| Latino/Mixed-American | Recorded Anxiety | Discontinuation vs Continuation | MDD_symptom_count | Income>200k | 1.222 | 0.217 | 6.896 | 8.20e-01 | 1.00e+00 | 198 | 143 | 55 |
| Latino/Mixed-American | Recorded Anxiety | Discontinuation vs Continuation | MDD_symptom_count | Income100-150k | 0.947 | 0.313 | 2.864 | 9.23e-01 | 1.00e+00 | 198 | 143 | 55 |
| Latino/Mixed-American | Recorded Anxiety | Discontinuation vs Continuation | MDD_symptom_count | Income150-200k | 0.740 | 0.116 | 4.740 | 7.51e-01 | 1.00e+00 | 198 | 143 | 55 |
| Latino/Mixed-American | Recorded Anxiety | Discontinuation vs Continuation | MDD_symptom_count | Income25-35k | 1.352 | 0.443 | 4.126 | 5.96e-01 | 1.00e+00 | 198 | 143 | 55 |
| Latino/Mixed-American | Recorded Anxiety | Discontinuation vs Continuation | MDD_symptom_count | Income35-50k | 0.689 | 0.203 | 2.332 | 5.49e-01 | 1.00e+00 | 198 | 143 | 55 |
| Latino/Mixed-American | Recorded Anxiety | Discontinuation vs Continuation | MDD_symptom_count | Income50-75k | 0.489 | 0.159 | 1.499 | 2.10e-01 | 1.00e+00 | 198 | 143 | 55 |
| Latino/Mixed-American | Recorded Anxiety | Discontinuation vs Continuation | MDD_symptom_count | Income75-100k | 0.687 | 0.209 | 2.253 | 5.35e-01 | 1.00e+00 | 198 | 143 | 55 |
| Latino/Mixed-American | Recorded Anxiety | Discontinuation vs Continuation | MDD_symptom_count | Education_levelCollege Postgraduate | 1.397 | 0.134 | 14.610 | 7.80e-01 | 1.00e+00 | 198 | 143 | 55 |
| Latino/Mixed-American | Recorded Anxiety | Discontinuation vs Continuation | MDD_symptom_count | Education_levelCollege Undergraduate | 1.381 | 0.146 | 13.110 | 7.79e-01 | 1.00e+00 | 198 | 143 | 55 |
| Latino/Mixed-American | Recorded Anxiety | Discontinuation vs Continuation | MDD_symptom_count | Education_levelTwelve or GED | 2.608 | 0.262 | 25.980 | 4.14e-01 | 1.00e+00 | 198 | 143 | 55 |
| Latino/Mixed-American | Recorded Anxiety | Discontinuation vs Continuation | MDD_symptom_count | MDD_symptom_count | 1.318 | 0.896 | 1.938 | 1.61e-01 | 1.00e+00 | 198 | 143 | 55 |
| Latino/Mixed-American | Recorded Anxiety | Switching vs Continuation | MDD_symptom_count | (Intercept) | 0.994 | 0.270 | 3.659 | 9.92e-01 | 1.00e+00 | 281 | 143 | 138 |
| Latino/Mixed-American | Recorded Anxiety | Switching vs Continuation | MDD_symptom_count | age_at_first_AD | 0.696 | 0.512 | 0.947 | 2.09e-02 | 2.92e-01 | 281 | 143 | 138 |
| Latino/Mixed-American | Recorded Anxiety | Switching vs Continuation | MDD_symptom_count | sex_at_birthMale | 1.574 | 0.855 | 2.897 | 1.45e-01 | 1.00e+00 | 281 | 143 | 138 |
| Latino/Mixed-American | Recorded Anxiety | Switching vs Continuation | MDD_symptom_count | Income>200k | 0.973 | 0.264 | 3.585 | 9.67e-01 | 1.00e+00 | 281 | 143 | 138 |
| Latino/Mixed-American | Recorded Anxiety | Switching vs Continuation | MDD_symptom_count | Income100-150k | 0.643 | 0.269 | 1.536 | 3.20e-01 | 1.00e+00 | 281 | 143 | 138 |
| Latino/Mixed-American | Recorded Anxiety | Switching vs Continuation | MDD_symptom_count | Income150-200k | 0.885 | 0.246 | 3.183 | 8.51e-01 | 1.00e+00 | 281 | 143 | 138 |
| Latino/Mixed-American | Recorded Anxiety | Switching vs Continuation | MDD_symptom_count | Income25-35k | 0.605 | 0.237 | 1.547 | 2.94e-01 | 1.00e+00 | 281 | 143 | 138 |
| Latino/Mixed-American | Recorded Anxiety | Switching vs Continuation | MDD_symptom_count | Income35-50k | 0.563 | 0.221 | 1.438 | 2.30e-01 | 1.00e+00 | 281 | 143 | 138 |
| Latino/Mixed-American | Recorded Anxiety | Switching vs Continuation | MDD_symptom_count | Income50-75k | 0.612 | 0.286 | 1.307 | 2.04e-01 | 1.00e+00 | 281 | 143 | 138 |
| Latino/Mixed-American | Recorded Anxiety | Switching vs Continuation | MDD_symptom_count | Income75-100k | 0.682 | 0.290 | 1.603 | 3.80e-01 | 1.00e+00 | 281 | 143 | 138 |
| Latino/Mixed-American | Recorded Anxiety | Switching vs Continuation | MDD_symptom_count | Education_levelCollege Postgraduate | 0.653 | 0.148 | 2.883 | 5.73e-01 | 1.00e+00 | 281 | 143 | 138 |

| | | | | | | | | | | | | |
|---|---|---|---|---|---|---|---|---|---|---|---|---|
| Latino/Mixed-American | Recorded Anxiety | Switching vs Continuation | MDD_symptom_count | Education_levelCollege Undergraduate | 1.124 | 0.281 | 4.500 | 8.68e-01 | 1.00e+00 | 281 | 143 | 138 |
| Latino/Mixed-American | Recorded Anxiety | Switching vs Continuation | MDD_symptom_count | Education_levelTwelve or GED | 0.964 | 0.218 | 4.268 | 9.62e-01 | 1.00e+00 | 281 | 143 | 138 |
| Latino/Mixed-American | Recorded Anxiety | Switching vs Continuation | MDD_symptom_count | MDD_symptom_count | 1.234 | 0.947 | 1.609 | 1.20e-01 | 1.00e+00 | 281 | 143 | 138 |
| Latino/Mixed-American | Recorded Anxiety | Discontinuation vs Continuation | Migraine | (Intercept) | 0.530 | 0.422 | 0.664 | 3.55e-08 | 4.97e-07 | 2246 | 1462 | 784 |
| Latino/Mixed-American | Recorded Anxiety | Discontinuation vs Continuation | Migraine | age_at_first_AD | 1.022 | 0.928 | 1.125 | 6.58e-01 | 1.00e+00 | 2246 | 1462 | 784 |
| Latino/Mixed-American | Recorded Anxiety | Discontinuation vs Continuation | Migraine | sex_at_birthMale | 1.258 | 1.035 | 1.529 | 2.12e-02 | 2.97e-01 | 2246 | 1462 | 784 |
| Latino/Mixed-American | Recorded Anxiety | Discontinuation vs Continuation | Migraine | Income>200k | 0.563 | 0.273 | 1.158 | 1.19e-01 | 1.00e+00 | 2246 | 1462 | 784 |
| Latino/Mixed-American | Recorded Anxiety | Discontinuation vs Continuation | Migraine | Income100-150k | 0.590 | 0.396 | 0.878 | 9.32e-03 | 1.31e-01 | 2246 | 1462 | 784 |
| Latino/Mixed-American | Recorded Anxiety | Discontinuation vs Continuation | Migraine | Income150-200k | 0.822 | 0.463 | 1.459 | 5.03e-01 | 1.00e+00 | 2246 | 1462 | 784 |
| Latino/Mixed-American | Recorded Anxiety | Discontinuation vs Continuation | Migraine | Income25-35k | 0.847 | 0.638 | 1.125 | 2.52e-01 | 1.00e+00 | 2246 | 1462 | 784 |
| Latino/Mixed-American | Recorded Anxiety | Discontinuation vs Continuation | Migraine | Income35-50k | 0.923 | 0.685 | 1.244 | 5.98e-01 | 1.00e+00 | 2246 | 1462 | 784 |
| Latino/Mixed-American | Recorded Anxiety | Discontinuation vs Continuation | Migraine | Income50-75k | 0.756 | 0.561 | 1.018 | 6.52e-02 | 9.13e-01 | 2246 | 1462 | 784 |
| Latino/Mixed-American | Recorded Anxiety | Discontinuation vs Continuation | Migraine | Income75-100k | 0.840 | 0.569 | 1.239 | 3.78e-01 | 1.00e+00 | 2246 | 1462 | 784 |
| Latino/Mixed-American | Recorded Anxiety | Discontinuation vs Continuation | Migraine | Education_levelCollege Postgraduate | 0.983 | 0.647 | 1.493 | 9.36e-01 | 1.00e+00 | 2246 | 1462 | 784 |
| Latino/Mixed-American | Recorded Anxiety | Discontinuation vs Continuation | Migraine | Education_levelCollege Undergraduate | 1.270 | 0.973 | 1.656 | 7.82e-02 | 1.00e+00 | 2246 | 1462 | 784 |
| Latino/Mixed-American | Recorded Anxiety | Discontinuation vs Continuation | Migraine | Education_levelTwelve or GED | 1.151 | 0.869 | 1.523 | 3.26e-01 | 1.00e+00 | 2246 | 1462 | 784 |
| Latino/Mixed-American | Recorded Anxiety | Discontinuation vs Continuation | Migraine | Migraine | 0.796 | 0.657 | 0.965 | 2.03e-02 | 2.84e-01 | 2246 | 1462 | 784 |
| Latino/Mixed-American | Recorded Anxiety | Switching vs Continuation | Migraine | (Intercept) | 0.632 | 0.518 | 0.771 | 6.12e-06 | 8.56e-05 | 2697 | 1462 | 1235 |
| Latino/Mixed-American | Recorded Anxiety | Switching vs Continuation | Migraine | age_at_first_AD | 0.918 | 0.840 | 1.003 | 5.83e-02 | 8.16e-01 | 2697 | 1462 | 1235 |
| Latino/Mixed-American | Recorded Anxiety | Switching vs Continuation | Migraine | sex_at_birthMale | 1.210 | 1.011 | 1.448 | 3.73e-02 | 5.22e-01 | 2697 | 1462 | 1235 |
| Latino/Mixed-American | Recorded Anxiety | Switching vs Continuation | Migraine | Income>200k | 0.724 | 0.409 | 1.280 | 2.66e-01 | 1.00e+00 | 2697 | 1462 | 1235 |
| Latino/Mixed-American | Recorded Anxiety | Switching vs Continuation | Migraine | Income100-150k | 0.765 | 0.552 | 1.060 | 1.07e-01 | 1.00e+00 | 2697 | 1462 | 1235 |
| Latino/Mixed-American | Recorded Anxiety | Switching vs Continuation | Migraine | Income150-200k | 0.950 | 0.569 | 1.586 | 8.43e-01 | 1.00e+00 | 2697 | 1462 | 1235 |
| Latino/Mixed-American | Recorded Anxiety | Switching vs Continuation | Migraine | Income25-35k | 0.901 | 0.699 | 1.162 | 4.22e-01 | 1.00e+00 | 2697 | 1462 | 1235 |
| Latino/Mixed-American | Recorded Anxiety | Switching vs Continuation | Migraine | Income35-50k | 1.102 | 0.847 | 1.433 | 4.71e-01 | 1.00e+00 | 2697 | 1462 | 1235 |
| Latino/Mixed-American | Recorded Anxiety | Switching vs Continuation | Migraine | Income50-75k | 0.849 | 0.653 | 1.103 | 2.20e-01 | 1.00e+00 | 2697 | 1462 | 1235 |
| Latino/Mixed-American | Recorded Anxiety | Switching vs Continuation | Migraine | Income75-100k | 0.989 | 0.705 | 1.388 | 9.48e-01 | 1.00e+00 | 2697 | 1462 | 1235 |
| Latino/Mixed-American | Recorded Anxiety | Switching vs Continuation | Migraine | Education_levelCollege Postgraduate | 1.071 | 0.760 | 1.510 | 6.94e-01 | 1.00e+00 | 2697 | 1462 | 1235 |
| Latino/Mixed-American | Recorded Anxiety | Switching vs Continuation | Migraine | Education_levelCollege Undergraduate | 1.021 | 0.807 | 1.293 | 8.60e-01 | 1.00e+00 | 2697 | 1462 | 1235 |
| Latino/Mixed-American | Recorded Anxiety | Switching vs Continuation | Migraine | Education_levelTwelve or GED | 0.808 | 0.628 | 1.041 | 9.86e-02 | 1.00e+00 | 2697 | 1462 | 1235 |
| Latino/Mixed-American | Recorded Anxiety | Switching vs Continuation | Migraine | Migraine | 1.972 | 1.682 | 2.311 | <2.2e-16 | 7.14e-16 | 2697 | 1462 | 1235 |
| Latino/Mixed-American | Recorded Anxiety | Discontinuation vs Continuation | Recorded Depression | (Intercept) | 0.478 | 0.371 | 0.617 | 1.20e-08 | 1.69e-07 | 2246 | 1462 | 784 |
| Latino/Mixed-American | Recorded Anxiety | Discontinuation vs Continuation | Recorded Depression | age_at_first_AD | 1.027 | 0.933 | 1.130 | 5.86e-01 | 1.00e+00 | 2246 | 1462 | 784 |
| Latino/Mixed-American | Recorded Anxiety | Discontinuation vs Continuation | Recorded Depression | sex_at_birthMale | 1.293 | 1.066 | 1.570 | 9.23e-03 | 1.29e-01 | 2246 | 1462 | 784 |
| Latino/Mixed-American | Recorded Anxiety | Discontinuation vs Continuation | Recorded Depression | Income>200k | 0.557 | 0.270 | 1.146 | 1.12e-01 | 1.00e+00 | 2246 | 1462 | 784 |
| Latino/Mixed-American | Recorded Anxiety | Discontinuation vs Continuation | Recorded Depression | Income100-150k | 0.583 | 0.392 | 0.869 | 8.01e-03 | 1.12e-01 | 2246 | 1462 | 784 |
| Latino/Mixed-American | Recorded Anxiety | Discontinuation vs Continuation | Recorded Depression | Income150-200k | 0.854 | 0.480 | 1.518 | 5.91e-01 | 1.00e+00 | 2246 | 1462 | 784 |
| Latino/Mixed-American | Recorded Anxiety | Discontinuation vs Continuation | Recorded Depression | Income25-35k | 0.846 | 0.637 | 1.123 | 2.46e-01 | 1.00e+00 | 2246 | 1462 | 784 |
| Latino/Mixed-American | Recorded Anxiety | Discontinuation vs Continuation | Recorded Depression | Income35-50k | 0.922 | 0.684 | 1.242 | 5.92e-01 | 1.00e+00 | 2246 | 1462 | 784 |
| Latino/Mixed-American | Recorded Anxiety | Discontinuation vs Continuation | Recorded Depression | Income50-75k | 0.753 | 0.559 | 1.015 | 6.25e-02 | 8.75e-01 | 2246 | 1462 | 784 |
| Latino/Mixed-American | Recorded Anxiety | Discontinuation vs Continuation | Recorded Depression | Income75-100k | 0.834 | 0.565 | 1.231 | 3.62e-01 | 1.00e+00 | 2246 | 1462 | 784 |
| Latino/Mixed-American | Recorded Anxiety | Discontinuation vs Continuation | Recorded Depression | Education_levelCollege Postgraduate | 0.996 | 0.655 | 1.513 | 9.84e-01 | 1.00e+00 | 2246 | 1462 | 784 |
| Latino/Mixed-American | Recorded Anxiety | Discontinuation vs Continuation | Recorded Depression | Education_levelCollege Undergraduate | 1.272 | 0.976 | 1.658 | 7.55e-02 | 1.00e+00 | 2246 | 1462 | 784 |
| Latino/Mixed-American | Recorded Anxiety | Discontinuation vs Continuation | Recorded Depression | Education_levelTwelve or GED | 1.147 | 0.867 | 1.518 | 3.36e-01 | 1.00e+00 | 2246 | 1462 | 784 |
| Latino/Mixed-American | Recorded Anxiety | Discontinuation vs Continuation | Recorded Depression | Recorded Depression | 1.041 | 0.868 | 1.249 | 6.66e-01 | 1.00e+00 | 2246 | 1462 | 784 |
| Latino/Mixed-American | Recorded Anxiety | Switching vs Continuation | Recorded Depression | (Intercept) | 0.316 | 0.248 | 0.403 | <2.2e-16 | <2.2e-16 | 2697 | 1462 | 1235 |
| Latino/Mixed-American | Recorded Anxiety | Switching vs Continuation | Recorded Depression | age_at_first_AD | 0.905 | 0.827 | 0.991 | 3.08e-02 | 4.31e-01 | 2697 | 1462 | 1235 |
| Latino/Mixed-American | Recorded Anxiety | Switching vs Continuation | Recorded Depression | sex_at_birthMale | 1.100 | 0.917 | 1.320 | 3.04e-01 | 1.00e+00 | 2697 | 1462 | 1235 |
| Latino/Mixed-American | Recorded Anxiety | Switching vs Continuation | Recorded Depression | Income>200k | 0.842 | 0.468 | 1.514 | 5.66e-01 | 1.00e+00 | 2697 | 1462 | 1235 |
| Latino/Mixed-American | Recorded Anxiety | Switching vs Continuation | Recorded Depression | Income100-150k | 1.016 | 0.724 | 1.424 | 9.28e-01 | 1.00e+00 | 2697 | 1462 | 1235 |
| Latino/Mixed-American | Recorded Anxiety | Switching vs Continuation | Recorded Depression | Income150-200k | 1.138 | 0.669 | 1.935 | 6.33e-01 | 1.00e+00 | 2697 | 1462 | 1235 |
| Latino/Mixed-American | Recorded Anxiety | Switching vs Continuation | Recorded Depression | Income25-35k | 0.968 | 0.746 | 1.255 | 8.04e-01 | 1.00e+00 | 2697 | 1462 | 1235 |
| Latino/Mixed-American | Recorded Anxiety | Switching vs Continuation | Recorded Depression | Income35-50k | 1.273 | 0.972 | 1.667 | 7.98e-02 | 1.00e+00 | 2697 | 1462 | 1235 |
| Latino/Mixed-American | Recorded Anxiety | Switching vs Continuation | Recorded Depression | Income50-75k | 0.998 | 0.762 | 1.306 | 9.87e-01 | 1.00e+00 | 2697 | 1462 | 1235 |
| Latino/Mixed-American | Recorded Anxiety | Switching vs Continuation | Recorded Depression | Income75-100k | 1.238 | 0.873 | 1.756 | 2.31e-01 | 1.00e+00 | 2697 | 1462 | 1235 |
| Latino/Mixed-American | Recorded Anxiety | Switching vs Continuation | Recorded Depression | Education_levelCollege Postgraduate | 1.080 | 0.760 | 1.533 | 6.69e-01 | 1.00e+00 | 2697 | 1462 | 1235 |
| Latino/Mixed-American | Recorded Anxiety | Switching vs Continuation | Recorded Depression | Education_levelCollege Undergraduate | 1.039 | 0.818 | 1.321 | 7.54e-01 | 1.00e+00 | 2697 | 1462 | 1235 |
| Latino/Mixed-American | Recorded Anxiety | Switching vs Continuation | Recorded Depression | Education_levelTwelve or GED | 0.779 | 0.603 | 1.007 | 5.65e-02 | 7.91e-01 | 2697 | 1462 | 1235 |
| Latino/Mixed-American | Recorded Anxiety | Switching vs Continuation | Recorded Depression | Recorded Depression | 3.465 | 2.873 | 4.180 | <2.2e-16 | <2.2e-16 | 2697 | 1462 | 1235 |
| Latino/Mixed-American | Recorded Anxiety | Discontinuation vs Continuation | Smoking_Frequency | (Intercept) | 0.432 | 0.342 | 0.547 | 2.48e-12 | 3.48e-11 | 2185 | 1420 | 765 |
| Latino/Mixed-American | Recorded Anxiety | Discontinuation vs Continuation | Smoking_Frequency | age_at_first_AD | 1.028 | 0.932 | 1.134 | 5.82e-01 | 1.00e+00 | 2185 | 1420 | 765 |
| Latino/Mixed-American | Recorded Anxiety | Discontinuation vs Continuation | Smoking_Frequency | sex_at_birthMale | 1.211 | 0.991 | 1.479 | 6.15e-02 | 8.60e-01 | 2185 | 1420 | 765 |
| Latino/Mixed-American | Recorded Anxiety | Discontinuation vs Continuation | Smoking_Frequency | Income>200k | 0.598 | 0.289 | 1.234 | 1.64e-01 | 1.00e+00 | 2185 | 1420 | 765 |
| Latino/Mixed-American | Recorded Anxiety | Discontinuation vs Continuation | Smoking_Frequency | Income100-150k | 0.619 | 0.413 | 0.927 | 2.01e-02 | 2.81e-01 | 2185 | 1420 | 765 |
| Latino/Mixed-American | Recorded Anxiety | Discontinuation vs Continuation | Smoking_Frequency | Income150-200k | 0.885 | 0.498 | 1.573 | 6.76e-01 | 1.00e+00 | 2185 | 1420 | 765 |
| Latino/Mixed-American | Recorded Anxiety | Discontinuation vs Continuation | Smoking_Frequency | Income25-35k | 0.861 | 0.646 | 1.149 | 3.09e-01 | 1.00e+00 | 2185 | 1420 | 765 |

| | | | | | | | | | | | | |
|---|---|---|---|---|---|---|---|---|---|---|---|---|
| Latino/Mixed-American | Recorded Anxiety | Discontinuation vs Continuation | Smoking_Frequency | Income35-50k | 0.912 | 0.673 | 1.237 | 5.53e-01 | 1.00e+00 | 2185 | 1420 | 765 |
| Latino/Mixed-American | Recorded Anxiety | Discontinuation vs Continuation | Smoking_Frequency | Income50-75k | 0.784 | 0.580 | 1.061 | 1.15e-01 | 1.00e+00 | 2185 | 1420 | 765 |
| Latino/Mixed-American | Recorded Anxiety | Discontinuation vs Continuation | Smoking_Frequency | Income75-100k | 0.863 | 0.581 | 1.283 | 4.67e-01 | 1.00e+00 | 2185 | 1420 | 765 |
| Latino/Mixed-American | Recorded Anxiety | Discontinuation vs Continuation | Smoking_Frequency | Education_levelCollege Postgraduate | 1.054 | 0.690 | 1.611 | 8.07e-01 | 1.00e+00 | 2185 | 1420 | 765 |
| Latino/Mixed-American | Recorded Anxiety | Discontinuation vs Continuation | Smoking_Frequency | Education_levelCollege Undergraduate | 1.318 | 1.007 | 1.726 | 4.45e-02 | 6.23e-01 | 2185 | 1420 | 765 |
| Latino/Mixed-American | Recorded Anxiety | Discontinuation vs Continuation | Smoking_Frequency | Education_levelTwelve or GED | 1.224 | 0.920 | 1.628 | 1.66e-01 | 1.00e+00 | 2185 | 1420 | 765 |
| Latino/Mixed-American | Recorded Anxiety | Discontinuation vs Continuation | Smoking_Frequency | Smoking_FrequencyEvery Day | 1.531 | 1.094 | 2.142 | 1.30e-02 | 1.82e-01 | 2185 | 1420 | 765 |
| Latino/Mixed-American | Recorded Anxiety | Discontinuation vs Continuation | Smoking_Frequency | Smoking_FrequencyNot At All (former) | 1.178 | 0.940 | 1.475 | 1.54e-01 | 1.00e+00 | 2185 | 1420 | 765 |
| Latino/Mixed-American | Recorded Anxiety | Discontinuation vs Continuation | Smoking_Frequency | Smoking_FrequencySome Days | 1.626 | 1.109 | 2.384 | 1.28e-02 | 1.79e-01 | 2185 | 1420 | 765 |
| Latino/Mixed-American | Recorded Anxiety | Switching vs Continuation | Smoking_Frequency | (Intercept) | 0.759 | 0.624 | 0.923 | 5.74e-03 | 8.04e-02 | 2610 | 1420 | 1190 |
| Latino/Mixed-American | Recorded Anxiety | Switching vs Continuation | Smoking_Frequency | age_at_first_AD | 0.891 | 0.814 | 0.975 | 1.16e-02 | 1.62e-01 | 2610 | 1420 | 1190 |
| Latino/Mixed-American | Recorded Anxiety | Switching vs Continuation | Smoking_Frequency | sex_at_birthMale | 1.000 | 0.834 | 1.199 | 9.97e-01 | 1.00e+00 | 2610 | 1420 | 1190 |
| Latino/Mixed-American | Recorded Anxiety | Switching vs Continuation | Smoking_Frequency | Income>200k | 0.684 | 0.382 | 1.224 | 2.01e-01 | 1.00e+00 | 2610 | 1420 | 1190 |
| Latino/Mixed-American | Recorded Anxiety | Switching vs Continuation | Smoking_Frequency | Income100-150k | 0.824 | 0.593 | 1.145 | 2.48e-01 | 1.00e+00 | 2610 | 1420 | 1190 |
| Latino/Mixed-American | Recorded Anxiety | Switching vs Continuation | Smoking_Frequency | Income150-200k | 0.879 | 0.528 | 1.464 | 6.21e-01 | 1.00e+00 | 2610 | 1420 | 1190 |
| Latino/Mixed-American | Recorded Anxiety | Switching vs Continuation | Smoking_Frequency | Income25-35k | 0.921 | 0.713 | 1.189 | 5.26e-01 | 1.00e+00 | 2610 | 1420 | 1190 |
| Latino/Mixed-American | Recorded Anxiety | Switching vs Continuation | Smoking_Frequency | Income35-50k | 1.079 | 0.827 | 1.407 | 5.76e-01 | 1.00e+00 | 2610 | 1420 | 1190 |
| Latino/Mixed-American | Recorded Anxiety | Switching vs Continuation | Smoking_Frequency | Income50-75k | 0.845 | 0.649 | 1.100 | 2.10e-01 | 1.00e+00 | 2610 | 1420 | 1190 |
| Latino/Mixed-American | Recorded Anxiety | Switching vs Continuation | Smoking_Frequency | Income75-100k | 0.990 | 0.702 | 1.397 | 9.55e-01 | 1.00e+00 | 2610 | 1420 | 1190 |
| Latino/Mixed-American | Recorded Anxiety | Switching vs Continuation | Smoking_Frequency | Education_levelCollege Postgraduate | 1.100 | 0.779 | 1.554 | 5.87e-01 | 1.00e+00 | 2610 | 1420 | 1190 |
| Latino/Mixed-American | Recorded Anxiety | Switching vs Continuation | Smoking_Frequency | Education_levelCollege Undergraduate | 1.059 | 0.836 | 1.341 | 6.34e-01 | 1.00e+00 | 2610 | 1420 | 1190 |
| Latino/Mixed-American | Recorded Anxiety | Switching vs Continuation | Smoking_Frequency | Education_levelTwelve or GED | 0.859 | 0.666 | 1.108 | 2.43e-01 | 1.00e+00 | 2610 | 1420 | 1190 |
| Latino/Mixed-American | Recorded Anxiety | Switching vs Continuation | Smoking_Frequency | Smoking_FrequencyEvery Day | 1.596 | 1.183 | 2.153 | 2.23e-03 | 3.12e-02 | 2610 | 1420 | 1190 |
| Latino/Mixed-American | Recorded Anxiety | Switching vs Continuation | Smoking_Frequency | Smoking_FrequencyNot At All (former) | 1.244 | 1.023 | 1.514 | 2.88e-02 | 4.03e-01 | 2610 | 1420 | 1190 |
| Latino/Mixed-American | Recorded Anxiety | Switching vs Continuation | Smoking_Frequency | Smoking_FrequencySome Days | 1.220 | 0.848 | 1.756 | 2.84e-01 | 1.00e+00 | 2610 | 1420 | 1190 |
| Latino/Mixed-American | Recorded Anxiety | Discontinuation vs Continuation | Stress | (Intercept) | 0.359 | 0.209 | 0.617 | 2.12e-04 | 2.97e-03 | 754 | 540 | 214 |
| Latino/Mixed-American | Recorded Anxiety | Discontinuation vs Continuation | Stress | age_at_first_AD | 0.934 | 0.782 | 1.116 | 4.51e-01 | 1.00e+00 | 754 | 540 | 214 |
| Latino/Mixed-American | Recorded Anxiety | Discontinuation vs Continuation | Stress | sex_at_birthMale | 1.166 | 0.806 | 1.689 | 4.15e-01 | 1.00e+00 | 754 | 540 | 214 |
| Latino/Mixed-American | Recorded Anxiety | Discontinuation vs Continuation | Stress | Income>200k | 0.655 | 0.242 | 1.772 | 4.05e-01 | 1.00e+00 | 754 | 540 | 214 |
| Latino/Mixed-American | Recorded Anxiety | Discontinuation vs Continuation | Stress | Income100-150k | 0.704 | 0.376 | 1.319 | 2.73e-01 | 1.00e+00 | 754 | 540 | 214 |
| Latino/Mixed-American | Recorded Anxiety | Discontinuation vs Continuation | Stress | Income150-200k | 1.189 | 0.530 | 2.669 | 6.74e-01 | 1.00e+00 | 754 | 540 | 214 |
| Latino/Mixed-American | Recorded Anxiety | Discontinuation vs Continuation | Stress | Income25-35k | 1.086 | 0.636 | 1.855 | 7.62e-01 | 1.00e+00 | 754 | 540 | 214 |
| Latino/Mixed-American | Recorded Anxiety | Discontinuation vs Continuation | Stress | Income35-50k | 1.334 | 0.772 | 2.306 | 3.02e-01 | 1.00e+00 | 754 | 540 | 214 |
| Latino/Mixed-American | Recorded Anxiety | Discontinuation vs Continuation | Stress | Income50-75k | 0.850 | 0.507 | 1.423 | 5.35e-01 | 1.00e+00 | 754 | 540 | 214 |
| Latino/Mixed-American | Recorded Anxiety | Discontinuation vs Continuation | Stress | Income75-100k | 0.751 | 0.390 | 1.445 | 3.91e-01 | 1.00e+00 | 754 | 540 | 214 |
| Latino/Mixed-American | Recorded Anxiety | Discontinuation vs Continuation | Stress | Education_levelCollege Postgraduate | 1.092 | 0.520 | 2.295 | 8.15e-01 | 1.00e+00 | 754 | 540 | 214 |
| Latino/Mixed-American | Recorded Anxiety | Discontinuation vs Continuation | Stress | Education_levelCollege Undergraduate | 1.156 | 0.633 | 2.113 | 6.37e-01 | 1.00e+00 | 754 | 540 | 214 |
| Latino/Mixed-American | Recorded Anxiety | Discontinuation vs Continuation | Stress | Education_levelTwelve or GED | 1.294 | 0.683 | 2.452 | 4.28e-01 | 1.00e+00 | 754 | 540 | 214 |
| Latino/Mixed-American | Recorded Anxiety | Discontinuation vs Continuation | Stress | StressAlmost Never | 0.559 | 0.248 | 1.263 | 1.62e-01 | 1.00e+00 | 754 | 540 | 214 |
| Latino/Mixed-American | Recorded Anxiety | Discontinuation vs Continuation | Stress | StressFairly Often | 0.957 | 0.499 | 1.834 | 8.93e-01 | 1.00e+00 | 754 | 540 | 214 |
| Latino/Mixed-American | Recorded Anxiety | Discontinuation vs Continuation | Stress | StressSometimes | 0.952 | 0.667 | 1.359 | 7.85e-01 | 1.00e+00 | 754 | 540 | 214 |
| Latino/Mixed-American | Recorded Anxiety | Discontinuation vs Continuation | Stress | StressVery Often | 0.730 | 0.296 | 1.801 | 4.94e-01 | 1.00e+00 | 754 | 540 | 214 |
| Latino/Mixed-American | Recorded Anxiety | Switching vs Continuation | Stress | (Intercept) | 0.952 | 0.639 | 1.419 | 8.09e-01 | 1.00e+00 | 1072 | 540 | 532 |
| Latino/Mixed-American | Recorded Anxiety | Switching vs Continuation | Stress | age_at_first_AD | 0.783 | 0.678 | 0.904 | 8.21e-04 | 1.15e-02 | 1072 | 540 | 532 |
| Latino/Mixed-American | Recorded Anxiety | Switching vs Continuation | Stress | sex_at_birthMale | 1.342 | 1.013 | 1.778 | 4.07e-02 | 5.69e-01 | 1072 | 540 | 532 |
| Latino/Mixed-American | Recorded Anxiety | Switching vs Continuation | Stress | Income>200k | 0.686 | 0.332 | 1.416 | 3.08e-01 | 1.00e+00 | 1072 | 540 | 532 |
| Latino/Mixed-American | Recorded Anxiety | Switching vs Continuation | Stress | Income100-150k | 0.735 | 0.462 | 1.170 | 1.95e-01 | 1.00e+00 | 1072 | 540 | 532 |
| Latino/Mixed-American | Recorded Anxiety | Switching vs Continuation | Stress | Income150-200k | 0.809 | 0.412 | 1.588 | 5.37e-01 | 1.00e+00 | 1072 | 540 | 532 |
| Latino/Mixed-American | Recorded Anxiety | Switching vs Continuation | Stress | Income25-35k | 1.003 | 0.660 | 1.525 | 9.89e-01 | 1.00e+00 | 1072 | 540 | 532 |
| Latino/Mixed-American | Recorded Anxiety | Switching vs Continuation | Stress | Income35-50k | 1.387 | 0.894 | 2.151 | 1.44e-01 | 1.00e+00 | 1072 | 540 | 532 |
| Latino/Mixed-American | Recorded Anxiety | Switching vs Continuation | Stress | Income50-75k | 0.847 | 0.571 | 1.256 | 4.08e-01 | 1.00e+00 | 1072 | 540 | 532 |
| Latino/Mixed-American | Recorded Anxiety | Switching vs Continuation | Stress | Income75-100k | 1.005 | 0.629 | 1.605 | 9.83e-01 | 1.00e+00 | 1072 | 540 | 532 |
| Latino/Mixed-American | Recorded Anxiety | Switching vs Continuation | Stress | Education_levelCollege Postgraduate | 0.856 | 0.493 | 1.487 | 5.81e-01 | 1.00e+00 | 1072 | 540 | 532 |
| Latino/Mixed-American | Recorded Anxiety | Switching vs Continuation | Stress | Education_levelCollege Undergraduate | 0.961 | 0.611 | 1.511 | 8.64e-01 | 1.00e+00 | 1072 | 540 | 532 |
| Latino/Mixed-American | Recorded Anxiety | Switching vs Continuation | Stress | Education_levelTwelve or GED | 0.702 | 0.425 | 1.158 | 1.66e-01 | 1.00e+00 | 1072 | 540 | 532 |
| Latino/Mixed-American | Recorded Anxiety | Switching vs Continuation | Stress | StressAlmost Never | 0.888 | 0.514 | 1.532 | 6.69e-01 | 1.00e+00 | 1072 | 540 | 532 |
| Latino/Mixed-American | Recorded Anxiety | Switching vs Continuation | Stress | StressFairly Often | 0.949 | 0.580 | 1.554 | 8.36e-01 | 1.00e+00 | 1072 | 540 | 532 |
| Latino/Mixed-American | Recorded Anxiety | Switching vs Continuation | Stress | StressSometimes | 1.052 | 0.803 | 1.378 | 7.15e-01 | 1.00e+00 | 1072 | 540 | 532 |
| Latino/Mixed-American | Recorded Anxiety | Switching vs Continuation | Stress | StressVery Often | 0.547 | 0.264 | 1.137 | 1.06e-01 | 1.00e+00 | 1072 | 540 | 532 |
| Latino/Mixed-American | Recorded Anxiety | Discontinuation vs Continuation | age_at_first_AD | (Intercept) | 0.501 | 0.448 | 0.559 | <2.2e-16 | <2.2e-16 | 2246 | 1462 | 784 |
| Latino/Mixed-American | Recorded Anxiety | Discontinuation vs Continuation | age_at_first_AD | age_at_first_AD | 1.012 | 0.922 | 1.110 | 8.05e-01 | 1.00e+00 | 2246 | 1462 | 784 |
| Latino/Mixed-American | Recorded Anxiety | Discontinuation vs Continuation | age_at_first_AD | sex_at_birthMale | 1.295 | 1.069 | 1.569 | 8.36e-03 | 1.17e-01 | 2246 | 1462 | 784 |
| Latino/Mixed-American | Recorded Anxiety | Switching vs Continuation | age_at_first_AD | (Intercept) | 0.791 | 0.718 | 0.872 | 2.35e-06 | 3.29e-05 | 2697 | 1462 | 1235 |
| Latino/Mixed-American | Recorded Anxiety | Switching vs Continuation | age_at_first_AD | age_at_first_AD | 0.898 | 0.825 | 0.976 | 1.17e-02 | 1.63e-01 | 2697 | 1462 | 1235 |
| Latino/Mixed-American | Recorded Anxiety | Switching vs Continuation | age_at_first_AD | sex_at_birthMale | 1.077 | 0.906 | 1.281 | 4.01e-01 | 1.00e+00 | 2697 | 1462 | 1235 |
| Latino/Mixed-American | Recorded Anxiety | Discontinuation vs Continuation | sex_at_birth | (Intercept) | 0.501 | 0.448 | 0.559 | <2.2e-16 | <2.2e-16 | 2246 | 1462 | 784 |

| | | | | | | | | | | | | |
|---|---|---|---|---|---|---|---|---|---|---|---|---|
| Latino/Mixed-American | Recorded Anxiety | Discontinuation vs Continuation | sex_at_birth | age_at_first_AD | 1.012 | 0.922 | 1.110 | 8.05e-01 | 1.00e+00 | 2246 | 1462 | 784 |
| Latino/Mixed-American | Recorded Anxiety | Discontinuation vs Continuation | sex_at_birth | sex_at_birthMale | 1.295 | 1.069 | 1.569 | 8.36e-03 | 1.17e-01 | 2246 | 1462 | 784 |
| Latino/Mixed-American | Recorded Anxiety | Switching vs Continuation | sex_at_birth | (Intercept) | 0.791 | 0.718 | 0.872 | 2.35e-06 | 3.29e-05 | 2697 | 1462 | 1235 |
| Latino/Mixed-American | Recorded Anxiety | Switching vs Continuation | sex_at_birth | age_at_first_AD | 0.898 | 0.825 | 0.976 | 1.17e-02 | 1.63e-01 | 2697 | 1462 | 1235 |
| Latino/Mixed-American | Recorded Anxiety | Switching vs Continuation | sex_at_birth | sex_at_birthMale | 1.077 | 0.906 | 1.281 | 4.01e-01 | 1.00e+00 | 2697 | 1462 | 1235 |
| Latino/Mixed-American | CIDI-MDD | Discontinuation vs Continuation | BMI | (Intercept) | 1.684 | 0.147 | 19.320 | 6.75e-01 | 1.00e+00 | 186 | 128 | 58 |
| Latino/Mixed-American | CIDI-MDD | Discontinuation vs Continuation | BMI | age_at_first_AD | 0.732 | 0.488 | 1.096 | 1.30e-01 | 1.00e+00 | 186 | 128 | 58 |
| Latino/Mixed-American | CIDI-MDD | Discontinuation vs Continuation | BMI | sex_at_birthMale | 1.148 | 0.499 | 2.643 | 7.45e-01 | 1.00e+00 | 186 | 128 | 58 |
| Latino/Mixed-American | CIDI-MDD | Discontinuation vs Continuation | BMI | Income>200k | 1.961 | 0.465 | 8.274 | 3.59e-01 | 1.00e+00 | 186 | 128 | 58 |
| Latino/Mixed-American | CIDI-MDD | Discontinuation vs Continuation | BMI | Income100-150k | 1.588 | 0.502 | 5.020 | 4.31e-01 | 1.00e+00 | 186 | 128 | 58 |
| Latino/Mixed-American | CIDI-MDD | Discontinuation vs Continuation | BMI | Income150-200k | 1.568 | 0.233 | 10.540 | 6.44e-01 | 1.00e+00 | 186 | 128 | 58 |
| Latino/Mixed-American | CIDI-MDD | Discontinuation vs Continuation | BMI | Income25-35k | 1.296 | 0.405 | 4.146 | 6.63e-01 | 1.00e+00 | 186 | 128 | 58 |
| Latino/Mixed-American | CIDI-MDD | Discontinuation vs Continuation | BMI | Income35-50k | 1.893 | 0.625 | 5.730 | 2.59e-01 | 1.00e+00 | 186 | 128 | 58 |
| Latino/Mixed-American | CIDI-MDD | Discontinuation vs Continuation | BMI | Income50-75k | 0.524 | 0.163 | 1.689 | 2.79e-01 | 1.00e+00 | 186 | 128 | 58 |
| Latino/Mixed-American | CIDI-MDD | Discontinuation vs Continuation | BMI | Income75-100k | 0.883 | 0.247 | 3.156 | 8.48e-01 | 1.00e+00 | 186 | 128 | 58 |
| Latino/Mixed-American | CIDI-MDD | Discontinuation vs Continuation | BMI | Education_levelCollege Postgraduate | 0.115 | 0.008 | 1.608 | 1.08e-01 | 1.00e+00 | 186 | 128 | 58 |
| Latino/Mixed-American | CIDI-MDD | Discontinuation vs Continuation | BMI | Education_levelCollege Undergraduate | 0.211 | 0.017 | 2.619 | 2.26e-01 | 1.00e+00 | 186 | 128 | 58 |
| Latino/Mixed-American | CIDI-MDD | Discontinuation vs Continuation | BMI | Education_levelTwelve or GED | 0.265 | 0.020 | 3.582 | 3.17e-01 | 1.00e+00 | 186 | 128 | 58 |
| Latino/Mixed-American | CIDI-MDD | Discontinuation vs Continuation | BMI | BMI | 1.081 | 0.768 | 1.523 | 6.55e-01 | 1.00e+00 | 186 | 128 | 58 |
| Latino/Mixed-American | CIDI-MDD | Switching vs Continuation | BMI | (Intercept) | 1.631 | 0.145 | 18.400 | 6.92e-01 | 1.00e+00 | 245 | 128 | 117 |
| Latino/Mixed-American | CIDI-MDD | Switching vs Continuation | BMI | age_at_first_AD | 0.661 | 0.472 | 0.926 | 1.59e-02 | 2.23e-01 | 245 | 128 | 117 |
| Latino/Mixed-American | CIDI-MDD | Switching vs Continuation | BMI | sex_at_birthMale | 1.381 | 0.718 | 2.655 | 3.34e-01 | 1.00e+00 | 245 | 128 | 117 |
| Latino/Mixed-American | CIDI-MDD | Switching vs Continuation | BMI | Income>200k | 0.577 | 0.149 | 2.243 | 4.28e-01 | 1.00e+00 | 245 | 128 | 117 |
| Latino/Mixed-American | CIDI-MDD | Switching vs Continuation | BMI | Income100-150k | 0.798 | 0.298 | 2.133 | 6.53e-01 | 1.00e+00 | 245 | 128 | 117 |
| Latino/Mixed-American | CIDI-MDD | Switching vs Continuation | BMI | Income150-200k | 2.184 | 0.554 | 8.612 | 2.65e-01 | 1.00e+00 | 245 | 128 | 117 |
| Latino/Mixed-American | CIDI-MDD | Switching vs Continuation | BMI | Income25-35k | 0.655 | 0.243 | 1.764 | 4.02e-01 | 1.00e+00 | 245 | 128 | 117 |
| Latino/Mixed-American | CIDI-MDD | Switching vs Continuation | BMI | Income35-50k | 0.742 | 0.267 | 2.063 | 5.67e-01 | 1.00e+00 | 245 | 128 | 117 |
| Latino/Mixed-American | CIDI-MDD | Switching vs Continuation | BMI | Income50-75k | 0.679 | 0.303 | 1.521 | 3.47e-01 | 1.00e+00 | 245 | 128 | 117 |
| Latino/Mixed-American | CIDI-MDD | Switching vs Continuation | BMI | Income75-100k | 0.824 | 0.320 | 2.122 | 6.89e-01 | 1.00e+00 | 245 | 128 | 117 |
| Latino/Mixed-American | CIDI-MDD | Switching vs Continuation | BMI | Education_levelCollege Postgraduate | 0.336 | 0.026 | 4.410 | 4.06e-01 | 1.00e+00 | 245 | 128 | 117 |
| Latino/Mixed-American | CIDI-MDD | Switching vs Continuation | BMI | Education_levelCollege Undergraduate | 0.551 | 0.045 | 6.708 | 6.40e-01 | 1.00e+00 | 245 | 128 | 117 |
| Latino/Mixed-American | CIDI-MDD | Switching vs Continuation | BMI | Education_levelTwelve or GED | 0.575 | 0.044 | 7.480 | 6.72e-01 | 1.00e+00 | 245 | 128 | 117 |
| Latino/Mixed-American | CIDI-MDD | Switching vs Continuation | BMI | BMI | 1.329 | 0.971 | 1.820 | 7.61e-02 | 1.00e+00 | 245 | 128 | 117 |
| Latino/Mixed-American | CIDI-MDD | Discontinuation vs Continuation | Income | (Intercept) | 1.628 | 0.141 | 18.850 | 6.96e-01 | 1.00e+00 | 191 | 131 | 60 |
| Latino/Mixed-American | CIDI-MDD | Discontinuation vs Continuation | Income | age_at_first_AD | 0.742 | 0.498 | 1.106 | 1.43e-01 | 1.00e+00 | 191 | 131 | 60 |
| Latino/Mixed-American | CIDI-MDD | Discontinuation vs Continuation | Income | sex_at_birthMale | 1.118 | 0.500 | 2.502 | 7.85e-01 | 1.00e+00 | 191 | 131 | 60 |
| Latino/Mixed-American | CIDI-MDD | Discontinuation vs Continuation | Income | Income>200k | 1.987 | 0.482 | 8.189 | 3.42e-01 | 1.00e+00 | 191 | 131 | 60 |
| Latino/Mixed-American | CIDI-MDD | Discontinuation vs Continuation | Income | Income100-150k | 1.634 | 0.523 | 5.106 | 3.98e-01 | 1.00e+00 | 191 | 131 | 60 |
| Latino/Mixed-American | CIDI-MDD | Discontinuation vs Continuation | Income | Income150-200k | 1.660 | 0.248 | 11.110 | 6.01e-01 | 1.00e+00 | 191 | 131 | 60 |
| Latino/Mixed-American | CIDI-MDD | Discontinuation vs Continuation | Income | Income25-35k | 1.413 | 0.449 | 4.451 | 5.55e-01 | 1.00e+00 | 191 | 131 | 60 |
| Latino/Mixed-American | CIDI-MDD | Discontinuation vs Continuation | Income | Income35-50k | 2.093 | 0.708 | 6.190 | 1.82e-01 | 1.00e+00 | 191 | 131 | 60 |
| Latino/Mixed-American | CIDI-MDD | Discontinuation vs Continuation | Income | Income50-75k | 0.656 | 0.214 | 2.012 | 4.61e-01 | 1.00e+00 | 191 | 131 | 60 |
| Latino/Mixed-American | CIDI-MDD | Discontinuation vs Continuation | Income | Income75-100k | 0.824 | 0.234 | 2.902 | 7.63e-01 | 1.00e+00 | 191 | 131 | 60 |
| Latino/Mixed-American | CIDI-MDD | Discontinuation vs Continuation | Income | Education_levelCollege Postgraduate | 0.112 | 0.008 | 1.574 | 1.04e-01 | 1.00e+00 | 191 | 131 | 60 |
| Latino/Mixed-American | CIDI-MDD | Discontinuation vs Continuation | Income | Education_levelCollege Undergraduate | 0.214 | 0.017 | 2.689 | 2.32e-01 | 1.00e+00 | 191 | 131 | 60 |
| Latino/Mixed-American | CIDI-MDD | Discontinuation vs Continuation | Income | Education_levelTwelve or GED | 0.270 | 0.020 | 3.690 | 3.27e-01 | 1.00e+00 | 191 | 131 | 60 |
| Latino/Mixed-American | CIDI-MDD | Switching vs Continuation | Income | (Intercept) | 1.821 | 0.162 | 20.480 | 6.28e-01 | 1.00e+00 | 249 | 131 | 118 |
| Latino/Mixed-American | CIDI-MDD | Switching vs Continuation | Income | age_at_first_AD | 0.651 | 0.468 | 0.908 | 1.14e-02 | 1.59e-01 | 249 | 131 | 118 |
| Latino/Mixed-American | CIDI-MDD | Switching vs Continuation | Income | sex_at_birthMale | 1.377 | 0.723 | 2.622 | 3.30e-01 | 1.00e+00 | 249 | 131 | 118 |
| Latino/Mixed-American | CIDI-MDD | Switching vs Continuation | Income | Income>200k | 0.480 | 0.127 | 1.813 | 2.79e-01 | 1.00e+00 | 249 | 131 | 118 |
| Latino/Mixed-American | CIDI-MDD | Switching vs Continuation | Income | Income100-150k | 0.741 | 0.283 | 1.939 | 5.41e-01 | 1.00e+00 | 249 | 131 | 118 |
| Latino/Mixed-American | CIDI-MDD | Switching vs Continuation | Income | Income150-200k | 2.047 | 0.530 | 7.905 | 2.99e-01 | 1.00e+00 | 249 | 131 | 118 |
| Latino/Mixed-American | CIDI-MDD | Switching vs Continuation | Income | Income25-35k | 0.802 | 0.305 | 2.106 | 6.53e-01 | 1.00e+00 | 249 | 131 | 118 |
| Latino/Mixed-American | CIDI-MDD | Switching vs Continuation | Income | Income35-50k | 0.690 | 0.253 | 1.886 | 4.70e-01 | 1.00e+00 | 249 | 131 | 118 |
| Latino/Mixed-American | CIDI-MDD | Switching vs Continuation | Income | Income50-75k | 0.743 | 0.337 | 1.638 | 4.61e-01 | 1.00e+00 | 249 | 131 | 118 |
| Latino/Mixed-American | CIDI-MDD | Switching vs Continuation | Income | Income75-100k | 0.841 | 0.338 | 2.092 | 7.09e-01 | 1.00e+00 | 249 | 131 | 118 |
| Latino/Mixed-American | CIDI-MDD | Switching vs Continuation | Income | Education_levelCollege Postgraduate | 0.311 | 0.024 | 4.059 | 3.73e-01 | 1.00e+00 | 249 | 131 | 118 |
| Latino/Mixed-American | CIDI-MDD | Switching vs Continuation | Income | Education_levelCollege Undergraduate | 0.491 | 0.040 | 5.956 | 5.76e-01 | 1.00e+00 | 249 | 131 | 118 |
| Latino/Mixed-American | CIDI-MDD | Switching vs Continuation | Income | Education_levelTwelve or GED | 0.533 | 0.041 | 6.914 | 6.30e-01 | 1.00e+00 | 249 | 131 | 118 |
| Latino/Mixed-American | CIDI-MDD | Discontinuation vs Continuation | Insomnia | (Intercept) | 1.509 | 0.129 | 17.660 | 7.43e-01 | 1.00e+00 | 191 | 131 | 60 |
| Latino/Mixed-American | CIDI-MDD | Discontinuation vs Continuation | Insomnia | age_at_first_AD | 0.743 | 0.498 | 1.107 | 1.44e-01 | 1.00e+00 | 191 | 131 | 60 |
| Latino/Mixed-American | CIDI-MDD | Discontinuation vs Continuation | Insomnia | sex_at_birthMale | 1.082 | 0.478 | 2.453 | 8.50e-01 | 1.00e+00 | 191 | 131 | 60 |
| Latino/Mixed-American | CIDI-MDD | Discontinuation vs Continuation | Insomnia | Income>200k | 2.026 | 0.490 | 8.381 | 3.30e-01 | 1.00e+00 | 191 | 131 | 60 |
| Latino/Mixed-American | CIDI-MDD | Discontinuation vs Continuation | Insomnia | Income100-150k | 1.645 | 0.526 | 5.150 | 3.92e-01 | 1.00e+00 | 191 | 131 | 60 |
| Latino/Mixed-American | CIDI-MDD | Discontinuation vs Continuation | Insomnia | Income150-200k | 1.673 | 0.249 | 11.240 | 5.97e-01 | 1.00e+00 | 191 | 131 | 60 |
| Latino/Mixed-American | CIDI-MDD | Discontinuation vs Continuation | Insomnia | Income25-35k | 1.428 | 0.453 | 4.502 | 5.43e-01 | 1.00e+00 | 191 | 131 | 60 |

| | | | | | | | | | | | | |
|---|---|---|---|---|---|---|---|---|---|---|---|---|
| Latino/Mixed-American | CIDI-MDD | Discontinuation vs Continuation | Insomnia | Income35-50k | 2.104 | 0.711 | 6.223 | 1.79e-01 | 1.00e+00 | 191 | 131 | 60 |
| Latino/Mixed-American | CIDI-MDD | Discontinuation vs Continuation | Insomnia | Income50-75k | 0.645 | 0.210 | 1.986 | 4.45e-01 | 1.00e+00 | 191 | 131 | 60 |
| Latino/Mixed-American | CIDI-MDD | Discontinuation vs Continuation | Insomnia | Income75-100k | 0.835 | 0.237 | 2.946 | 7.79e-01 | 1.00e+00 | 191 | 131 | 60 |
| Latino/Mixed-American | CIDI-MDD | Discontinuation vs Continuation | Insomnia | Education_levelCollege Postgraduate | 0.115 | 0.008 | 1.601 | 1.07e-01 | 1.00e+00 | 191 | 131 | 60 |
| Latino/Mixed-American | CIDI-MDD | Discontinuation vs Continuation | Insomnia | Education_levelCollege Undergraduate | 0.226 | 0.018 | 2.834 | 2.49e-01 | 1.00e+00 | 191 | 131 | 60 |
| Latino/Mixed-American | CIDI-MDD | Discontinuation vs Continuation | Insomnia | Education_levelTwelve or GED | 0.278 | 0.021 | 3.760 | 3.35e-01 | 1.00e+00 | 191 | 131 | 60 |
| Latino/Mixed-American | CIDI-MDD | Discontinuation vs Continuation | Insomnia | Insomnia | 1.225 | 0.519 | 2.888 | 6.43e-01 | 1.00e+00 | 191 | 131 | 60 |
| Latino/Mixed-American | CIDI-MDD | Switching vs Continuation | Insomnia | (Intercept) | 1.244 | 0.109 | 14.220 | 8.60e-01 | 1.00e+00 | 249 | 131 | 118 |
| Latino/Mixed-American | CIDI-MDD | Switching vs Continuation | Insomnia | age_at_first_AD | 0.631 | 0.448 | 0.889 | 8.44e-03 | 1.18e-01 | 249 | 131 | 118 |
| Latino/Mixed-American | CIDI-MDD | Switching vs Continuation | Insomnia | sex_at_birthMale | 1.175 | 0.602 | 2.293 | 6.37e-01 | 1.00e+00 | 249 | 131 | 118 |
| Latino/Mixed-American | CIDI-MDD | Switching vs Continuation | Insomnia | Income>200k | 0.437 | 0.112 | 1.709 | 2.34e-01 | 1.00e+00 | 249 | 131 | 118 |
| Latino/Mixed-American | CIDI-MDD | Switching vs Continuation | Insomnia | Income100-150k | 0.673 | 0.248 | 1.829 | 4.38e-01 | 1.00e+00 | 249 | 131 | 118 |
| Latino/Mixed-American | CIDI-MDD | Switching vs Continuation | Insomnia | Income150-200k | 2.211 | 0.553 | 8.841 | 2.62e-01 | 1.00e+00 | 249 | 131 | 118 |
| Latino/Mixed-American | CIDI-MDD | Switching vs Continuation | Insomnia | Income25-35k | 0.926 | 0.345 | 2.489 | 8.79e-01 | 1.00e+00 | 249 | 131 | 118 |
| Latino/Mixed-American | CIDI-MDD | Switching vs Continuation | Insomnia | Income35-50k | 0.665 | 0.238 | 1.853 | 4.35e-01 | 1.00e+00 | 249 | 131 | 118 |
| Latino/Mixed-American | CIDI-MDD | Switching vs Continuation | Insomnia | Income50-75k | 0.704 | 0.310 | 1.600 | 4.02e-01 | 1.00e+00 | 249 | 131 | 118 |
| Latino/Mixed-American | CIDI-MDD | Switching vs Continuation | Insomnia | Income75-100k | 0.847 | 0.332 | 2.160 | 7.28e-01 | 1.00e+00 | 249 | 131 | 118 |
| Latino/Mixed-American | CIDI-MDD | Switching vs Continuation | Insomnia | Education_levelCollege Postgraduate | 0.345 | 0.026 | 4.557 | 4.19e-01 | 1.00e+00 | 249 | 131 | 118 |
| Latino/Mixed-American | CIDI-MDD | Switching vs Continuation | Insomnia | Education_levelCollege Undergraduate | 0.577 | 0.047 | 7.089 | 6.68e-01 | 1.00e+00 | 249 | 131 | 118 |
| Latino/Mixed-American | CIDI-MDD | Switching vs Continuation | Insomnia | Education_levelTwelve or GED | 0.520 | 0.039 | 6.848 | 6.19e-01 | 1.00e+00 | 249 | 131 | 118 |
| Latino/Mixed-American | CIDI-MDD | Switching vs Continuation | Insomnia | Insomnia | 3.105 | 1.646 | 5.857 | 4.68e-04 | 6.55e-03 | 249 | 131 | 118 |
| Latino/Mixed-American | CIDI-MDD | Discontinuation vs Continuation | MDD_symptom_count | (Intercept) | 2.315 | 0.177 | 30.300 | 5.22e-01 | 1.00e+00 | 191 | 131 | 60 |
| Latino/Mixed-American | CIDI-MDD | Discontinuation vs Continuation | MDD_symptom_count | age_at_first_AD | 0.704 | 0.464 | 1.069 | 9.98e-02 | 1.00e+00 | 191 | 131 | 60 |
| Latino/Mixed-American | CIDI-MDD | Discontinuation vs Continuation | MDD_symptom_count | sex_at_birthMale | 1.117 | 0.499 | 2.501 | 7.88e-01 | 1.00e+00 | 191 | 131 | 60 |
| Latino/Mixed-American | CIDI-MDD | Discontinuation vs Continuation | MDD_symptom_count | Income>200k | 2.059 | 0.498 | 8.511 | 3.19e-01 | 1.00e+00 | 191 | 131 | 60 |
| Latino/Mixed-American | CIDI-MDD | Discontinuation vs Continuation | MDD_symptom_count | Income100-150k | 1.688 | 0.538 | 5.300 | 3.70e-01 | 1.00e+00 | 191 | 131 | 60 |
| Latino/Mixed-American | CIDI-MDD | Discontinuation vs Continuation | MDD_symptom_count | Income150-200k | 1.831 | 0.271 | 12.390 | 5.35e-01 | 1.00e+00 | 191 | 131 | 60 |
| Latino/Mixed-American | CIDI-MDD | Discontinuation vs Continuation | MDD_symptom_count | Income25-35k | 1.313 | 0.411 | 4.189 | 6.46e-01 | 1.00e+00 | 191 | 131 | 60 |
| Latino/Mixed-American | CIDI-MDD | Discontinuation vs Continuation | MDD_symptom_count | Income35-50k | 2.253 | 0.748 | 6.781 | 1.49e-01 | 1.00e+00 | 191 | 131 | 60 |
| Latino/Mixed-American | CIDI-MDD | Discontinuation vs Continuation | MDD_symptom_count | Income50-75k | 0.675 | 0.219 | 2.079 | 4.93e-01 | 1.00e+00 | 191 | 131 | 60 |
| Latino/Mixed-American | CIDI-MDD | Discontinuation vs Continuation | MDD_symptom_count | Income75-100k | 0.802 | 0.226 | 2.845 | 7.32e-01 | 1.00e+00 | 191 | 131 | 60 |
| Latino/Mixed-American | CIDI-MDD | Discontinuation vs Continuation | MDD_symptom_count | Education_levelCollege Postgraduate | 0.107 | 0.008 | 1.511 | 9.80e-02 | 1.00e+00 | 191 | 131 | 60 |
| Latino/Mixed-American | CIDI-MDD | Discontinuation vs Continuation | MDD_symptom_count | Education_levelCollege Undergraduate | 0.202 | 0.016 | 2.540 | 2.15e-01 | 1.00e+00 | 191 | 131 | 60 |
| Latino/Mixed-American | CIDI-MDD | Discontinuation vs Continuation | MDD_symptom_count | Education_levelTwelve or GED | 0.272 | 0.020 | 3.700 | 3.28e-01 | 1.00e+00 | 191 | 131 | 60 |
| Latino/Mixed-American | CIDI-MDD | Discontinuation vs Continuation | MDD_symptom_count | MDD_symptom_count | 0.650 | 0.247 | 1.713 | 3.84e-01 | 1.00e+00 | 191 | 131 | 60 |
| Latino/Mixed-American | CIDI-MDD | Switching vs Continuation | MDD_symptom_count | (Intercept) | 2.037 | 0.170 | 24.430 | 5.75e-01 | 1.00e+00 | 249 | 131 | 118 |
| Latino/Mixed-American | CIDI-MDD | Switching vs Continuation | MDD_symptom_count | age_at_first_AD | 0.641 | 0.456 | 0.902 | 1.07e-02 | 1.50e-01 | 249 | 131 | 118 |
| Latino/Mixed-American | CIDI-MDD | Switching vs Continuation | MDD_symptom_count | sex_at_birthMale | 1.393 | 0.730 | 2.661 | 3.15e-01 | 1.00e+00 | 249 | 131 | 118 |
| Latino/Mixed-American | CIDI-MDD | Switching vs Continuation | MDD_symptom_count | Income>200k | 0.480 | 0.127 | 1.815 | 2.80e-01 | 1.00e+00 | 249 | 131 | 118 |
| Latino/Mixed-American | CIDI-MDD | Switching vs Continuation | MDD_symptom_count | Income100-150k | 0.740 | 0.283 | 1.937 | 5.40e-01 | 1.00e+00 | 249 | 131 | 118 |
| Latino/Mixed-American | CIDI-MDD | Switching vs Continuation | MDD_symptom_count | Income150-200k | 2.097 | 0.540 | 8.141 | 2.85e-01 | 1.00e+00 | 249 | 131 | 118 |
| Latino/Mixed-American | CIDI-MDD | Switching vs Continuation | MDD_symptom_count | Income25-35k | 0.798 | 0.304 | 2.097 | 6.47e-01 | 1.00e+00 | 249 | 131 | 118 |
| Latino/Mixed-American | CIDI-MDD | Switching vs Continuation | MDD_symptom_count | Income35-50k | 0.704 | 0.256 | 1.934 | 4.96e-01 | 1.00e+00 | 249 | 131 | 118 |
| Latino/Mixed-American | CIDI-MDD | Switching vs Continuation | MDD_symptom_count | Income50-75k | 0.746 | 0.338 | 1.646 | 4.68e-01 | 1.00e+00 | 249 | 131 | 118 |
| Latino/Mixed-American | CIDI-MDD | Switching vs Continuation | MDD_symptom_count | Income75-100k | 0.843 | 0.338 | 2.099 | 7.13e-01 | 1.00e+00 | 249 | 131 | 118 |
| Latino/Mixed-American | CIDI-MDD | Switching vs Continuation | MDD_symptom_count | Education_levelCollege Postgraduate | 0.311 | 0.024 | 4.064 | 3.73e-01 | 1.00e+00 | 249 | 131 | 118 |
| Latino/Mixed-American | CIDI-MDD | Switching vs Continuation | MDD_symptom_count | Education_levelCollege Undergraduate | 0.490 | 0.040 | 5.952 | 5.75e-01 | 1.00e+00 | 249 | 131 | 118 |
| Latino/Mixed-American | CIDI-MDD | Switching vs Continuation | MDD_symptom_count | Education_levelTwelve or GED | 0.531 | 0.041 | 6.901 | 6.29e-01 | 1.00e+00 | 249 | 131 | 118 |
| Latino/Mixed-American | CIDI-MDD | Switching vs Continuation | MDD_symptom_count | MDD_symptom_count | 0.858 | 0.403 | 1.824 | 6.90e-01 | 1.00e+00 | 249 | 131 | 118 |
| Latino/Mixed-American | CIDI-MDD | Discontinuation vs Continuation | Migraine | (Intercept) | 1.749 | 0.151 | 20.230 | 6.54e-01 | 1.00e+00 | 191 | 131 | 60 |
| Latino/Mixed-American | CIDI-MDD | Discontinuation vs Continuation | Migraine | age_at_first_AD | 0.740 | 0.497 | 1.103 | 1.40e-01 | 1.00e+00 | 191 | 131 | 60 |
| Latino/Mixed-American | CIDI-MDD | Discontinuation vs Continuation | Migraine | sex_at_birthMale | 1.125 | 0.503 | 2.519 | 7.74e-01 | 1.00e+00 | 191 | 131 | 60 |
| Latino/Mixed-American | CIDI-MDD | Discontinuation vs Continuation | Migraine | Income>200k | 1.976 | 0.479 | 8.149 | 3.46e-01 | 1.00e+00 | 191 | 131 | 60 |
| Latino/Mixed-American | CIDI-MDD | Discontinuation vs Continuation | Migraine | Income100-150k | 1.728 | 0.545 | 5.480 | 3.53e-01 | 1.00e+00 | 191 | 131 | 60 |
| Latino/Mixed-American | CIDI-MDD | Discontinuation vs Continuation | Migraine | Income150-200k | 1.777 | 0.261 | 12.080 | 5.57e-01 | 1.00e+00 | 191 | 131 | 60 |
| Latino/Mixed-American | CIDI-MDD | Discontinuation vs Continuation | Migraine | Income25-35k | 1.399 | 0.443 | 4.419 | 5.67e-01 | 1.00e+00 | 191 | 131 | 60 |
| Latino/Mixed-American | CIDI-MDD | Discontinuation vs Continuation | Migraine | Income35-50k | 2.098 | 0.709 | 6.210 | 1.81e-01 | 1.00e+00 | 191 | 131 | 60 |
| Latino/Mixed-American | CIDI-MDD | Discontinuation vs Continuation | Migraine | Income50-75k | 0.674 | 0.219 | 2.071 | 4.90e-01 | 1.00e+00 | 191 | 131 | 60 |
| Latino/Mixed-American | CIDI-MDD | Discontinuation vs Continuation | Migraine | Income75-100k | 0.846 | 0.239 | 2.991 | 7.95e-01 | 1.00e+00 | 191 | 131 | 60 |
| Latino/Mixed-American | CIDI-MDD | Discontinuation vs Continuation | Migraine | Education_levelCollege Postgraduate | 0.110 | 0.008 | 1.526 | 9.99e-02 | 1.00e+00 | 191 | 131 | 60 |
| Latino/Mixed-American | CIDI-MDD | Discontinuation vs Continuation | Migraine | Education_levelCollege Undergraduate | 0.211 | 0.017 | 2.610 | 2.25e-01 | 1.00e+00 | 191 | 131 | 60 |
| Latino/Mixed-American | CIDI-MDD | Discontinuation vs Continuation | Migraine | Education_levelTwelve or GED | 0.256 | 0.019 | 3.474 | 3.06e-01 | 1.00e+00 | 191 | 131 | 60 |
| Latino/Mixed-American | CIDI-MDD | Discontinuation vs Continuation | Migraine | Migraine | 0.781 | 0.373 | 1.636 | 5.13e-01 | 1.00e+00 | 191 | 131 | 60 |
| Latino/Mixed-American | CIDI-MDD | Switching vs Continuation | Migraine | (Intercept) | 1.558 | 0.132 | 18.370 | 7.25e-01 | 1.00e+00 | 249 | 131 | 118 |
| Latino/Mixed-American | CIDI-MDD | Switching vs Continuation | Migraine | age_at_first_AD | 0.664 | 0.475 | 0.928 | 1.66e-02 | 2.32e-01 | 249 | 131 | 118 |
| Latino/Mixed-American | CIDI-MDD | Switching vs Continuation | Migraine | sex_at_birthMale | 1.500 | 0.781 | 2.879 | 2.23e-01 | 1.00e+00 | 249 | 131 | 118 |

| | | | | | | | | | | | | |
|---|---|---|---|---|---|---|---|---|---|---|---|---|
| Latino/Mixed-American | CIDI-MDD | Switching vs Continuation | Migraine | Income>200k | 0.509 | 0.133 | 1.950 | 3.24e-01 | 1.00e+00 | 249 | 131 | 118 |
| Latino/Mixed-American | CIDI-MDD | Switching vs Continuation | Migraine | Income100-150k | 0.683 | 0.259 | 1.800 | 4.40e-01 | 1.00e+00 | 249 | 131 | 118 |
| Latino/Mixed-American | CIDI-MDD | Switching vs Continuation | Migraine | Income150-200k | 1.867 | 0.485 | 7.184 | 3.64e-01 | 1.00e+00 | 249 | 131 | 118 |
| Latino/Mixed-American | CIDI-MDD | Switching vs Continuation | Migraine | Income25-35k | 0.866 | 0.326 | 2.301 | 7.73e-01 | 1.00e+00 | 249 | 131 | 118 |
| Latino/Mixed-American | CIDI-MDD | Switching vs Continuation | Migraine | Income35-50k | 0.732 | 0.266 | 2.012 | 5.45e-01 | 1.00e+00 | 249 | 131 | 118 |
| Latino/Mixed-American | CIDI-MDD | Switching vs Continuation | Migraine | Income50-75k | 0.774 | 0.347 | 1.726 | 5.31e-01 | 1.00e+00 | 249 | 131 | 118 |
| Latino/Mixed-American | CIDI-MDD | Switching vs Continuation | Migraine | Income75-100k | 0.912 | 0.361 | 2.303 | 8.46e-01 | 1.00e+00 | 249 | 131 | 118 |
| Latino/Mixed-American | CIDI-MDD | Switching vs Continuation | Migraine | Education_levelCollege Postgraduate | 0.272 | 0.020 | 3.726 | 3.30e-01 | 1.00e+00 | 249 | 131 | 118 |
| Latino/Mixed-American | CIDI-MDD | Switching vs Continuation | Migraine | Education_levelCollege Undergraduate | 0.448 | 0.035 | 5.690 | 5.35e-01 | 1.00e+00 | 249 | 131 | 118 |
| Latino/Mixed-American | CIDI-MDD | Switching vs Continuation | Migraine | Education_levelTwelve or GED | 0.518 | 0.038 | 7.026 | 6.21e-01 | 1.00e+00 | 249 | 131 | 118 |
| Latino/Mixed-American | CIDI-MDD | Switching vs Continuation | Migraine | Migraine | 1.804 | 1.034 | 3.148 | 3.76e-02 | 5.27e-01 | 249 | 131 | 118 |
| Latino/Mixed-American | CIDI-MDD | Discontinuation vs Continuation | Recorded Anxiety | (Intercept) | 1.974 | 0.162 | 24.040 | 5.94e-01 | 1.00e+00 | 191 | 131 | 60 |
| Latino/Mixed-American | CIDI-MDD | Discontinuation vs Continuation | Recorded Anxiety | age_at_first_AD | 0.725 | 0.484 | 1.085 | 1.18e-01 | 1.00e+00 | 191 | 131 | 60 |
| Latino/Mixed-American | CIDI-MDD | Discontinuation vs Continuation | Recorded Anxiety | sex_at_birthMale | 1.131 | 0.505 | 2.533 | 7.64e-01 | 1.00e+00 | 191 | 131 | 60 |
| Latino/Mixed-American | CIDI-MDD | Discontinuation vs Continuation | Recorded Anxiety | Income>200k | 1.794 | 0.425 | 7.571 | 4.26e-01 | 1.00e+00 | 191 | 131 | 60 |
| Latino/Mixed-American | CIDI-MDD | Discontinuation vs Continuation | Recorded Anxiety | Income100-150k | 1.697 | 0.539 | 5.344 | 3.66e-01 | 1.00e+00 | 191 | 131 | 60 |
| Latino/Mixed-American | CIDI-MDD | Discontinuation vs Continuation | Recorded Anxiety | Income150-200k | 1.619 | 0.242 | 10.820 | 6.19e-01 | 1.00e+00 | 191 | 131 | 60 |
| Latino/Mixed-American | CIDI-MDD | Discontinuation vs Continuation | Recorded Anxiety | Income25-35k | 1.406 | 0.445 | 4.443 | 5.62e-01 | 1.00e+00 | 191 | 131 | 60 |
| Latino/Mixed-American | CIDI-MDD | Discontinuation vs Continuation | Recorded Anxiety | Income35-50k | 2.084 | 0.703 | 6.174 | 1.85e-01 | 1.00e+00 | 191 | 131 | 60 |
| Latino/Mixed-American | CIDI-MDD | Discontinuation vs Continuation | Recorded Anxiety | Income50-75k | 0.660 | 0.215 | 2.027 | 4.68e-01 | 1.00e+00 | 191 | 131 | 60 |
| Latino/Mixed-American | CIDI-MDD | Discontinuation vs Continuation | Recorded Anxiety | Income75-100k | 0.814 | 0.231 | 2.875 | 7.50e-01 | 1.00e+00 | 191 | 131 | 60 |
| Latino/Mixed-American | CIDI-MDD | Discontinuation vs Continuation | Recorded Anxiety | Education_levelCollege Postgraduate | 0.110 | 0.008 | 1.554 | 1.02e-01 | 1.00e+00 | 191 | 131 | 60 |
| Latino/Mixed-American | CIDI-MDD | Discontinuation vs Continuation | Recorded Anxiety | Education_levelCollege Undergraduate | 0.203 | 0.016 | 2.582 | 2.19e-01 | 1.00e+00 | 191 | 131 | 60 |
| Latino/Mixed-American | CIDI-MDD | Discontinuation vs Continuation | Recorded Anxiety | Education_levelTwelve or GED | 0.267 | 0.019 | 3.670 | 3.23e-01 | 1.00e+00 | 191 | 131 | 60 |
| Latino/Mixed-American | CIDI-MDD | Discontinuation vs Continuation | Recorded Anxiety | Recorded Anxiety | 0.753 | 0.386 | 1.468 | 4.04e-01 | 1.00e+00 | 191 | 131 | 60 |
| Latino/Mixed-American | CIDI-MDD | Switching vs Continuation | Recorded Anxiety | (Intercept) | 0.585 | 0.048 | 7.080 | 6.73e-01 | 1.00e+00 | 249 | 131 | 118 |
| Latino/Mixed-American | CIDI-MDD | Switching vs Continuation | Recorded Anxiety | age_at_first_AD | 0.713 | 0.506 | 1.004 | 5.28e-02 | 7.39e-01 | 249 | 131 | 118 |
| Latino/Mixed-American | CIDI-MDD | Switching vs Continuation | Recorded Anxiety | sex_at_birthMale | 1.410 | 0.726 | 2.742 | 3.11e-01 | 1.00e+00 | 249 | 131 | 118 |
| Latino/Mixed-American | CIDI-MDD | Switching vs Continuation | Recorded Anxiety | Income>200k | 0.659 | 0.166 | 2.616 | 5.53e-01 | 1.00e+00 | 249 | 131 | 118 |
| Latino/Mixed-American | CIDI-MDD | Switching vs Continuation | Recorded Anxiety | Income100-150k | 0.682 | 0.256 | 1.817 | 4.44e-01 | 1.00e+00 | 249 | 131 | 118 |
| Latino/Mixed-American | CIDI-MDD | Switching vs Continuation | Recorded Anxiety | Income150-200k | 2.235 | 0.541 | 9.239 | 2.67e-01 | 1.00e+00 | 249 | 131 | 118 |
| Latino/Mixed-American | CIDI-MDD | Switching vs Continuation | Recorded Anxiety | Income25-35k | 0.838 | 0.308 | 2.281 | 7.29e-01 | 1.00e+00 | 249 | 131 | 118 |
| Latino/Mixed-American | CIDI-MDD | Switching vs Continuation | Recorded Anxiety | Income35-50k | 0.610 | 0.218 | 1.710 | 3.47e-01 | 1.00e+00 | 249 | 131 | 118 |
| Latino/Mixed-American | CIDI-MDD | Switching vs Continuation | Recorded Anxiety | Income50-75k | 0.739 | 0.328 | 1.663 | 4.65e-01 | 1.00e+00 | 249 | 131 | 118 |
| Latino/Mixed-American | CIDI-MDD | Switching vs Continuation | Recorded Anxiety | Income75-100k | 0.782 | 0.307 | 1.994 | 6.07e-01 | 1.00e+00 | 249 | 131 | 118 |
| Latino/Mixed-American | CIDI-MDD | Switching vs Continuation | Recorded Anxiety | Education_levelCollege Postgraduate | 0.448 | 0.034 | 5.865 | 5.40e-01 | 1.00e+00 | 249 | 131 | 118 |
| Latino/Mixed-American | CIDI-MDD | Switching vs Continuation | Recorded Anxiety | Education_levelCollege Undergraduate | 0.685 | 0.056 | 8.324 | 7.66e-01 | 1.00e+00 | 249 | 131 | 118 |
| Latino/Mixed-American | CIDI-MDD | Switching vs Continuation | Recorded Anxiety | Education_levelTwelve or GED | 0.748 | 0.057 | 9.792 | 8.25e-01 | 1.00e+00 | 249 | 131 | 118 |
| Latino/Mixed-American | CIDI-MDD | Switching vs Continuation | Recorded Anxiety | Recorded Anxiety | 3.170 | 1.677 | 5.989 | 3.80e-04 | 5.33e-03 | 249 | 131 | 118 |
| Latino/Mixed-American | CIDI-MDD | Discontinuation vs Continuation | Smoking_Frequency | (Intercept) | 2.232 | 0.169 | 29.550 | 5.42e-01 | 1.00e+00 | 185 | 126 | 59 |
| Latino/Mixed-American | CIDI-MDD | Discontinuation vs Continuation | Smoking_Frequency | age_at_first_AD | 0.756 | 0.496 | 1.152 | 1.93e-01 | 1.00e+00 | 185 | 126 | 59 |
| Latino/Mixed-American | CIDI-MDD | Discontinuation vs Continuation | Smoking_Frequency | sex_at_birthMale | 1.245 | 0.534 | 2.905 | 6.12e-01 | 1.00e+00 | 185 | 126 | 59 |
| Latino/Mixed-American | CIDI-MDD | Discontinuation vs Continuation | Smoking_Frequency | Income>200k | 2.268 | 0.532 | 9.677 | 2.69e-01 | 1.00e+00 | 185 | 126 | 59 |
| Latino/Mixed-American | CIDI-MDD | Discontinuation vs Continuation | Smoking_Frequency | Income100-150k | 1.850 | 0.569 | 6.018 | 3.07e-01 | 1.00e+00 | 185 | 126 | 59 |
| Latino/Mixed-American | CIDI-MDD | Discontinuation vs Continuation | Smoking_Frequency | Income150-200k | 1.759 | 0.256 | 12.100 | 5.66e-01 | 1.00e+00 | 185 | 126 | 59 |
| Latino/Mixed-American | CIDI-MDD | Discontinuation vs Continuation | Smoking_Frequency | Income25-35k | 1.699 | 0.500 | 5.775 | 3.96e-01 | 1.00e+00 | 185 | 126 | 59 |
| Latino/Mixed-American | CIDI-MDD | Discontinuation vs Continuation | Smoking_Frequency | Income35-50k | 2.155 | 0.701 | 6.624 | 1.80e-01 | 1.00e+00 | 185 | 126 | 59 |
| Latino/Mixed-American | CIDI-MDD | Discontinuation vs Continuation | Smoking_Frequency | Income50-75k | 0.575 | 0.176 | 1.881 | 3.60e-01 | 1.00e+00 | 185 | 126 | 59 |
| Latino/Mixed-American | CIDI-MDD | Discontinuation vs Continuation | Smoking_Frequency | Income75-100k | 0.802 | 0.223 | 2.893 | 7.37e-01 | 1.00e+00 | 185 | 126 | 59 |
| Latino/Mixed-American | CIDI-MDD | Discontinuation vs Continuation | Smoking_Frequency | Education_levelCollege Postgraduate | 0.075 | 0.005 | 1.190 | 6.62e-02 | 9.27e-01 | 185 | 126 | 59 |
| Latino/Mixed-American | CIDI-MDD | Discontinuation vs Continuation | Smoking_Frequency | Education_levelCollege Undergraduate | 0.168 | 0.012 | 2.331 | 1.84e-01 | 1.00e+00 | 185 | 126 | 59 |
| Latino/Mixed-American | CIDI-MDD | Discontinuation vs Continuation | Smoking_Frequency | Education_levelTwelve or GED | 0.181 | 0.011 | 2.840 | 2.23e-01 | 1.00e+00 | 185 | 126 | 59 |
| Latino/Mixed-American | CIDI-MDD | Discontinuation vs Continuation | Smoking_Frequency | Smoking_FrequencyEvery Day | 3.373 | 0.442 | 25.750 | 2.41e-01 | 1.00e+00 | 185 | 126 | 59 |
| Latino/Mixed-American | CIDI-MDD | Discontinuation vs Continuation | Smoking_Frequency | Smoking_FrequencyNot At All (former) | 0.792 | 0.349 | 1.798 | 5.78e-01 | 1.00e+00 | 185 | 126 | 59 |
| Latino/Mixed-American | CIDI-MDD | Discontinuation vs Continuation | Smoking_Frequency | Smoking_FrequencySome Days | 0.490 | 0.076 | 3.149 | 4.52e-01 | 1.00e+00 | 185 | 126 | 59 |
| Latino/Mixed-American | CIDI-MDD | Switching vs Continuation | Smoking_Frequency | (Intercept) | 1.514 | 0.130 | 17.680 | 7.41e-01 | 1.00e+00 | 239 | 126 | 113 |
| Latino/Mixed-American | CIDI-MDD | Switching vs Continuation | Smoking_Frequency | age_at_first_AD | 0.597 | 0.417 | 0.857 | 5.10e-03 | 7.14e-02 | 239 | 126 | 113 |
| Latino/Mixed-American | CIDI-MDD | Switching vs Continuation | Smoking_Frequency | sex_at_birthMale | 1.242 | 0.631 | 2.445 | 5.30e-01 | 1.00e+00 | 239 | 126 | 113 |
| Latino/Mixed-American | CIDI-MDD | Switching vs Continuation | Smoking_Frequency | Income>200k | 0.388 | 0.089 | 1.693 | 2.08e-01 | 1.00e+00 | 239 | 126 | 113 |
| Latino/Mixed-American | CIDI-MDD | Switching vs Continuation | Smoking_Frequency | Income100-150k | 0.649 | 0.233 | 1.809 | 4.09e-01 | 1.00e+00 | 239 | 126 | 113 |
| Latino/Mixed-American | CIDI-MDD | Switching vs Continuation | Smoking_Frequency | Income150-200k | 1.875 | 0.475 | 7.404 | 3.70e-01 | 1.00e+00 | 239 | 126 | 113 |
| Latino/Mixed-American | CIDI-MDD | Switching vs Continuation | Smoking_Frequency | Income25-35k | 0.781 | 0.285 | 2.138 | 6.30e-01 | 1.00e+00 | 239 | 126 | 113 |
| Latino/Mixed-American | CIDI-MDD | Switching vs Continuation | Smoking_Frequency | Income35-50k | 0.563 | 0.197 | 1.610 | 2.84e-01 | 1.00e+00 | 239 | 126 | 113 |
| Latino/Mixed-American | CIDI-MDD | Switching vs Continuation | Smoking_Frequency | Income50-75k | 0.633 | 0.278 | 1.444 | 2.77e-01 | 1.00e+00 | 239 | 126 | 113 |
| Latino/Mixed-American | CIDI-MDD | Switching vs Continuation | Smoking_Frequency | Income75-100k | 0.718 | 0.278 | 1.855 | 4.94e-01 | 1.00e+00 | 239 | 126 | 113 |
| Latino/Mixed-American | CIDI-MDD | Switching vs Continuation | Smoking_Frequency | Education_levelCollege Postgraduate | 0.379 | 0.028 | 5.130 | 4.65e-01 | 1.00e+00 | 239 | 126 | 113 |

| | | | | | | | | | | | | |
|---|---|---|---|---|---|---|---|---|---|---|---|---|
| Latino/Mixed-American | CIDI-MDD | Switching vs Continuation | Smoking_Frequency | Education_levelCollege Undergraduate | 0.506 | 0.040 | 6.350 | 5.97e-01 | 1.00e+00 | 239 | 126 | 113 |
| Latino/Mixed-American | CIDI-MDD | Switching vs Continuation | Smoking_Frequency | Education_levelTwelve or GED | 0.556 | 0.041 | 7.550 | 6.59e-01 | 1.00e+00 | 239 | 126 | 113 |
| Latino/Mixed-American | CIDI-MDD | Switching vs Continuation | Smoking_Frequency | Smoking_FrequencyEvery Day | 4.991 | 0.958 | 26.000 | 5.63e-02 | 7.88e-01 | 239 | 126 | 113 |
| Latino/Mixed-American | CIDI-MDD | Switching vs Continuation | Smoking_Frequency | Smoking_FrequencyNot At All (former) | 1.696 | 0.906 | 3.175 | 9.85e-02 | 1.00e+00 | 239 | 126 | 113 |
| Latino/Mixed-American | CIDI-MDD | Switching vs Continuation | Smoking_Frequency | Smoking_FrequencySome Days | 0.501 | 0.091 | 2.753 | 4.27e-01 | 1.00e+00 | 239 | 126 | 113 |
| Latino/Mixed-American | CIDI-MDD | Discontinuation vs Continuation | age_at_first_AD | (Intercept) | 0.397 | 0.266 | 0.592 | 6.05e-06 | 8.47e-05 | 191 | 131 | 60 |
| Latino/Mixed-American | CIDI-MDD | Discontinuation vs Continuation | age_at_first_AD | age_at_first_AD | 0.746 | 0.515 | 1.083 | 1.23e-01 | 1.00e+00 | 191 | 131 | 60 |
| Latino/Mixed-American | CIDI-MDD | Discontinuation vs Continuation | age_at_first_AD | sex_at_birthMale | 1.038 | 0.479 | 2.245 | 9.26e-01 | 1.00e+00 | 191 | 131 | 60 |
| Latino/Mixed-American | CIDI-MDD | Switching vs Continuation | age_at_first_AD | (Intercept) | 0.683 | 0.485 | 0.961 | 2.84e-02 | 3.98e-01 | 249 | 131 | 118 |
| Latino/Mixed-American | CIDI-MDD | Switching vs Continuation | age_at_first_AD | age_at_first_AD | 0.665 | 0.484 | 0.912 | 1.14e-02 | 1.60e-01 | 249 | 131 | 118 |
| Latino/Mixed-American | CIDI-MDD | Switching vs Continuation | age_at_first_AD | sex_at_birthMale | 1.419 | 0.761 | 2.644 | 2.71e-01 | 1.00e+00 | 249 | 131 | 118 |
| Latino/Mixed-American | CIDI-MDD | Discontinuation vs Continuation | sex_at_birth | (Intercept) | 0.397 | 0.266 | 0.592 | 6.05e-06 | 8.47e-05 | 191 | 131 | 60 |
| Latino/Mixed-American | CIDI-MDD | Discontinuation vs Continuation | sex_at_birth | age_at_first_AD | 0.746 | 0.515 | 1.083 | 1.23e-01 | 1.00e+00 | 191 | 131 | 60 |
| Latino/Mixed-American | CIDI-MDD | Discontinuation vs Continuation | sex_at_birth | sex_at_birthMale | 1.038 | 0.479 | 2.245 | 9.26e-01 | 1.00e+00 | 191 | 131 | 60 |
| Latino/Mixed-American | CIDI-MDD | Switching vs Continuation | sex_at_birth | (Intercept) | 0.683 | 0.485 | 0.961 | 2.84e-02 | 3.98e-01 | 249 | 131 | 118 |
| Latino/Mixed-American | CIDI-MDD | Switching vs Continuation | sex_at_birth | age_at_first_AD | 0.665 | 0.484 | 0.912 | 1.14e-02 | 1.60e-01 | 249 | 131 | 118 |
| Latino/Mixed-American | CIDI-MDD | Switching vs Continuation | sex_at_birth | sex_at_birthMale | 1.419 | 0.761 | 2.644 | 2.71e-01 | 1.00e+00 | 249 | 131 | 118 |
| Latino/Mixed-American | CIDI-noMDD | Discontinuation vs Continuation | Insomnia | (Intercept) | 0.324 | 0.025 | 4.135 | 3.85e-01 | 1.00e+00 | 103 | 78 | 25 |
| Latino/Mixed-American | CIDI-noMDD | Discontinuation vs Continuation | Insomnia | age_at_first_AD | 0.577 | 0.297 | 1.122 | 1.05e-01 | 1.00e+00 | 103 | 78 | 25 |
| Latino/Mixed-American | CIDI-noMDD | Discontinuation vs Continuation | Insomnia | sex_at_birthMale | 2.994 | 1.033 | 8.676 | 4.33e-02 | 6.07e-01 | 103 | 78 | 25 |
| Latino/Mixed-American | CIDI-noMDD | Discontinuation vs Continuation | Insomnia | Income>200k | 0.280 | 0.018 | 4.418 | 3.66e-01 | 1.00e+00 | 103 | 78 | 25 |
| Latino/Mixed-American | CIDI-noMDD | Discontinuation vs Continuation | Insomnia | Income100-150k | 0.763 | 0.153 | 3.815 | 7.42e-01 | 1.00e+00 | 103 | 78 | 25 |
| Latino/Mixed-American | CIDI-noMDD | Discontinuation vs Continuation | Insomnia | Income150-200k | 0.289 | 0.034 | 2.445 | 2.55e-01 | 1.00e+00 | 103 | 78 | 25 |
| Latino/Mixed-American | CIDI-noMDD | Discontinuation vs Continuation | Insomnia | Income25-35k | 1.418 | 0.289 | 6.972 | 6.67e-01 | 1.00e+00 | 103 | 78 | 25 |
| Latino/Mixed-American | CIDI-noMDD | Discontinuation vs Continuation | Insomnia | Income35-50k | 0.430 | 0.037 | 5.004 | 5.01e-01 | 1.00e+00 | 103 | 78 | 25 |
| Latino/Mixed-American | CIDI-noMDD | Discontinuation vs Continuation | Insomnia | Income50-75k | 0.434 | 0.070 | 2.691 | 3.70e-01 | 1.00e+00 | 103 | 78 | 25 |
| Latino/Mixed-American | CIDI-noMDD | Discontinuation vs Continuation | Insomnia | Income75-100k | 0.147 | 0.012 | 1.737 | 1.28e-01 | 1.00e+00 | 103 | 78 | 25 |
| Latino/Mixed-American | CIDI-noMDD | Discontinuation vs Continuation | Insomnia | Education_levelCollege Postgraduate | 1.217 | 0.090 | 16.380 | 8.82e-01 | 1.00e+00 | 103 | 78 | 25 |
| Latino/Mixed-American | CIDI-noMDD | Discontinuation vs Continuation | Insomnia | Education_levelCollege Undergraduate | 0.843 | 0.069 | 10.300 | 8.93e-01 | 1.00e+00 | 103 | 78 | 25 |
| Latino/Mixed-American | CIDI-noMDD | Discontinuation vs Continuation | Insomnia | Education_levelTwelve or GED | 1.192 | 0.082 | 17.390 | 8.98e-01 | 1.00e+00 | 103 | 78 | 25 |
| Latino/Mixed-American | CIDI-noMDD | Discontinuation vs Continuation | Insomnia | Insomnia | 1.480 | 0.398 | 5.499 | 5.58e-01 | 1.00e+00 | 103 | 78 | 25 |
| Latino/Mixed-American | CIDI-noMDD | Discontinuation vs Continuation | MDD_symptom_count | (Intercept) | 0.321 | 0.021 | 4.817 | 4.11e-01 | 1.00e+00 | 103 | 78 | 25 |
| Latino/Mixed-American | CIDI-noMDD | Discontinuation vs Continuation | MDD_symptom_count | age_at_first_AD | 0.581 | 0.300 | 1.125 | 1.07e-01 | 1.00e+00 | 103 | 78 | 25 |
| Latino/Mixed-American | CIDI-noMDD | Discontinuation vs Continuation | MDD_symptom_count | sex_at_birthMale | 2.840 | 0.996 | 8.104 | 5.10e-02 | 7.13e-01 | 103 | 78 | 25 |
| Latino/Mixed-American | CIDI-noMDD | Discontinuation vs Continuation | MDD_symptom_count | Income>200k | 0.314 | 0.021 | 4.660 | 4.00e-01 | 1.00e+00 | 103 | 78 | 25 |
| Latino/Mixed-American | CIDI-noMDD | Discontinuation vs Continuation | MDD_symptom_count | Income100-150k | 0.776 | 0.156 | 3.849 | 7.56e-01 | 1.00e+00 | 103 | 78 | 25 |
| Latino/Mixed-American | CIDI-noMDD | Discontinuation vs Continuation | MDD_symptom_count | Income150-200k | 0.278 | 0.033 | 2.330 | 2.38e-01 | 1.00e+00 | 103 | 78 | 25 |
| Latino/Mixed-American | CIDI-noMDD | Discontinuation vs Continuation | MDD_symptom_count | Income25-35k | 1.362 | 0.274 | 6.760 | 7.06e-01 | 1.00e+00 | 103 | 78 | 25 |
| Latino/Mixed-American | CIDI-noMDD | Discontinuation vs Continuation | MDD_symptom_count | Income35-50k | 0.383 | 0.033 | 4.517 | 4.46e-01 | 1.00e+00 | 103 | 78 | 25 |
| Latino/Mixed-American | CIDI-noMDD | Discontinuation vs Continuation | MDD_symptom_count | Income50-75k | 0.410 | 0.064 | 2.617 | 3.46e-01 | 1.00e+00 | 103 | 78 | 25 |
| Latino/Mixed-American | CIDI-noMDD | Discontinuation vs Continuation | MDD_symptom_count | Income75-100k | 0.159 | 0.014 | 1.824 | 1.39e-01 | 1.00e+00 | 103 | 78 | 25 |
| Latino/Mixed-American | CIDI-noMDD | Discontinuation vs Continuation | MDD_symptom_count | Education_levelCollege Postgraduate | 1.240 | 0.092 | 16.800 | 8.71e-01 | 1.00e+00 | 103 | 78 | 25 |
| Latino/Mixed-American | CIDI-noMDD | Discontinuation vs Continuation | MDD_symptom_count | Education_levelCollege Undergraduate | 0.915 | 0.077 | 10.930 | 9.44e-01 | 1.00e+00 | 103 | 78 | 25 |
| Latino/Mixed-American | CIDI-noMDD | Discontinuation vs Continuation | MDD_symptom_count | Education_levelTwelve or GED | 1.324 | 0.094 | 18.640 | 8.35e-01 | 1.00e+00 | 103 | 78 | 25 |
| Latino/Mixed-American | CIDI-noMDD | Discontinuation vs Continuation | MDD_symptom_count | MDD_symptom_count | 0.960 | 0.384 | 2.399 | 9.30e-01 | 1.00e+00 | 103 | 78 | 25 |
| Latino/Mixed-American | CIDI-noMDD | Discontinuation vs Continuation | Migraine | (Intercept) | 0.319 | 0.026 | 3.984 | 3.75e-01 | 1.00e+00 | 103 | 78 | 25 |
| Latino/Mixed-American | CIDI-noMDD | Discontinuation vs Continuation | Migraine | age_at_first_AD | 0.611 | 0.316 | 1.180 | 1.42e-01 | 1.00e+00 | 103 | 78 | 25 |
| Latino/Mixed-American | CIDI-noMDD | Discontinuation vs Continuation | Migraine | sex_at_birthMale | 2.781 | 0.984 | 7.858 | 5.36e-02 | 7.51e-01 | 103 | 78 | 25 |
| Latino/Mixed-American | CIDI-noMDD | Discontinuation vs Continuation | Migraine | Income>200k | 0.264 | 0.017 | 4.129 | 3.43e-01 | 1.00e+00 | 103 | 78 | 25 |
| Latino/Mixed-American | CIDI-noMDD | Discontinuation vs Continuation | Migraine | Income100-150k | 0.737 | 0.149 | 3.647 | 7.08e-01 | 1.00e+00 | 103 | 78 | 25 |
| Latino/Mixed-American | CIDI-noMDD | Discontinuation vs Continuation | Migraine | Income150-200k | 0.302 | 0.037 | 2.449 | 2.62e-01 | 1.00e+00 | 103 | 78 | 25 |
| Latino/Mixed-American | CIDI-noMDD | Discontinuation vs Continuation | Migraine | Income25-35k | 1.210 | 0.238 | 6.147 | 8.18e-01 | 1.00e+00 | 103 | 78 | 25 |
| Latino/Mixed-American | CIDI-noMDD | Discontinuation vs Continuation | Migraine | Income35-50k | 0.358 | 0.030 | 4.279 | 4.17e-01 | 1.00e+00 | 103 | 78 | 25 |
| Latino/Mixed-American | CIDI-noMDD | Discontinuation vs Continuation | Migraine | Income50-75k | 0.399 | 0.065 | 2.439 | 3.20e-01 | 1.00e+00 | 103 | 78 | 25 |
| Latino/Mixed-American | CIDI-noMDD | Discontinuation vs Continuation | Migraine | Income75-100k | 0.152 | 0.013 | 1.764 | 1.32e-01 | 1.00e+00 | 103 | 78 | 25 |
| Latino/Mixed-American | CIDI-noMDD | Discontinuation vs Continuation | Migraine | Education_levelCollege Postgraduate | 1.151 | 0.088 | 15.030 | 9.15e-01 | 1.00e+00 | 103 | 78 | 25 |
| Latino/Mixed-American | CIDI-noMDD | Discontinuation vs Continuation | Migraine | Education_levelCollege Undergraduate | 0.896 | 0.077 | 10.450 | 9.30e-01 | 1.00e+00 | 103 | 78 | 25 |
| Latino/Mixed-American | CIDI-noMDD | Discontinuation vs Continuation | Migraine | Education_levelTwelve or GED | 1.281 | 0.093 | 17.710 | 8.53e-01 | 1.00e+00 | 103 | 78 | 25 |
| Latino/Mixed-American | CIDI-noMDD | Discontinuation vs Continuation | Migraine | Migraine | 1.711 | 0.559 | 5.235 | 3.46e-01 | 1.00e+00 | 103 | 78 | 25 |
| Latino/Mixed-American | CIDI-noMDD | Discontinuation vs Continuation | Recorded Anxiety | (Intercept) | 0.403 | 0.030 | 5.417 | 4.93e-01 | 1.00e+00 | 103 | 78 | 25 |
| Latino/Mixed-American | CIDI-noMDD | Discontinuation vs Continuation | Recorded Anxiety | age_at_first_AD | 0.595 | 0.306 | 1.157 | 1.26e-01 | 1.00e+00 | 103 | 78 | 25 |
| Latino/Mixed-American | CIDI-noMDD | Discontinuation vs Continuation | Recorded Anxiety | sex_at_birthMale | 2.639 | 0.921 | 7.558 | 7.07e-02 | 9.89e-01 | 103 | 78 | 25 |
| Latino/Mixed-American | CIDI-noMDD | Discontinuation vs Continuation | Recorded Anxiety | Income>200k | 0.309 | 0.021 | 4.491 | 3.90e-01 | 1.00e+00 | 103 | 78 | 25 |
| Latino/Mixed-American | CIDI-noMDD | Discontinuation vs Continuation | Recorded Anxiety | Income100-150k | 0.731 | 0.145 | 3.678 | 7.04e-01 | 1.00e+00 | 103 | 78 | 25 |
| Latino/Mixed-American | CIDI-noMDD | Discontinuation vs Continuation | Recorded Anxiety | Income150-200k | 0.267 | 0.031 | 2.290 | 2.29e-01 | 1.00e+00 | 103 | 78 | 25 |
| Latino/Mixed-American | CIDI-noMDD | Discontinuation vs Continuation | Recorded Anxiety | Income25-35k | 1.386 | 0.281 | 6.844 | 6.88e-01 | 1.00e+00 | 103 | 78 | 25 |

| | | | | | | | | | | | | |
|---|---|---|---|---|---|---|---|---|---|---|---|---|
| Latino/Mixed-American | CIDI-noMDD | Discontinuation vs Continuation | Recorded Anxiety | Income35-50k | 0.365 | 0.032 | 4.119 | 4.15e-01 | 1.00e+00 | 103 | 78 | 25 |
| Latino/Mixed-American | CIDI-noMDD | Discontinuation vs Continuation | Recorded Anxiety | Income50-75k | 0.422 | 0.068 | 2.620 | 3.54e-01 | 1.00e+00 | 103 | 78 | 25 |
| Latino/Mixed-American | CIDI-noMDD | Discontinuation vs Continuation | Recorded Anxiety | Income75-100k | 0.176 | 0.015 | 2.024 | 1.63e-01 | 1.00e+00 | 103 | 78 | 25 |
| Latino/Mixed-American | CIDI-noMDD | Discontinuation vs Continuation | Recorded Anxiety | Education_levelCollege Postgraduate | 1.262 | 0.094 | 17.020 | 8.61e-01 | 1.00e+00 | 103 | 78 | 25 |
| Latino/Mixed-American | CIDI-noMDD | Discontinuation vs Continuation | Recorded Anxiety | Education_levelCollege Undergraduate | 0.909 | 0.076 | 10.940 | 9.40e-01 | 1.00e+00 | 103 | 78 | 25 |
| Latino/Mixed-American | CIDI-noMDD | Discontinuation vs Continuation | Recorded Anxiety | Education_levelTwelve or GED | 1.505 | 0.103 | 22.040 | 7.65e-01 | 1.00e+00 | 103 | 78 | 25 |
| Latino/Mixed-American | CIDI-noMDD | Discontinuation vs Continuation | Recorded Anxiety | Recorded Anxiety | 0.696 | 0.240 | 2.019 | 5.05e-01 | 1.00e+00 | 103 | 78 | 25 |
| Latino/Mixed-American | CIDI-noMDD | Discontinuation vs Continuation | age_at_first_AD | (Intercept) | 0.237 | 0.127 | 0.443 | 6.29e-06 | 8.81e-05 | 103 | 78 | 25 |
| Latino/Mixed-American | CIDI-noMDD | Discontinuation vs Continuation | age_at_first_AD | age_at_first_AD | 0.662 | 0.377 | 1.160 | 1.49e-01 | 1.00e+00 | 103 | 78 | 25 |
| Latino/Mixed-American | CIDI-noMDD | Discontinuation vs Continuation | age_at_first_AD | sex_at_birthMale | 2.182 | 0.837 | 5.688 | 1.10e-01 | 1.00e+00 | 103 | 78 | 25 |
| Latino/Mixed-American | CIDI-noMDD | Discontinuation vs Continuation | sex_at_birth | (Intercept) | 0.237 | 0.127 | 0.443 | 6.29e-06 | 8.81e-05 | 103 | 78 | 25 |
| Latino/Mixed-American | CIDI-noMDD | Discontinuation vs Continuation | sex_at_birth | age_at_first_AD | 0.662 | 0.377 | 1.160 | 1.49e-01 | 1.00e+00 | 103 | 78 | 25 |
| Latino/Mixed-American | CIDI-noMDD | Discontinuation vs Continuation | sex_at_birth | sex_at_birthMale | 2.182 | 0.837 | 5.688 | 1.10e-01 | 1.00e+00 | 103 | 78 | 25 |

**Supplementary Table 3. Multivariable logistic regression results for phenotypic associations with antidepressant treatment outcomes in the Pharmlines cohort (European-ancestry participants).** Each model estimates the association between participant-level exposures and one of two binary outcomes — switching versus continuation, or discontinuation versus continuation — fitted separately within each combination of diagnostic subset (Full Cohort; Recorded Depression; No Recorded Depression) and each phenotypic exposure. Within each model, one phenotypic exposure (the "Dependent" column) is modelled alongside a common covariate set (age at first antidepressant, sex at birth, income level, and education level); focal exposures span demographic, socioeconomic, clinical, and psychosocial measures (BMI, recorded depression, lifetime anxiety, fibromyalgia, childhood sexual abuse, neuroticism, long-term difficulties, threatening experiences, smoking pack-years, alcohol frequency, chronotype, and weekly sleep). The covariate-only rows (Dependent = age_at_first_AD or sex_at_birth) report the base model.

Columns: Population = genetic ancestry (European); Diagnostic Subset = cohort or clinical subset; Outcome = the binary contrast modelled; Dependent = the focal exposure for that model; variable = the individual model term; odds_ratio = exponentiated coefficient; lower_95/upper_95 = 95% confidence interval bounds for the odds ratio; p_value = nominal (uncorrected) p-value; Bonf_p = Bonferroni-corrected p-value; N_Reference = number in the reference (continuation) group; N_Case = number in the comparison (switching or discontinuation) group. Odds ratios >1 indicate higher odds of the comparison outcome relative to continuation. P-values reported as "<2.2e-16" reflect R's floating-point reporting limit; where a corrected value still falls below this limit it is likewise shown as "<2.2e-16".

Bonferroni correction was applied for 17 tested traits within each specification (diagnostic subset × outcome contrast).

Variable coding and reference categories:

All quantitative variables were standardised (mean 0, SD 1); each odds ratio therefore reflects the change in odds per one-standard-deviation increase. This applies to BMI, Income_level, Education_level, Neuroticism, Longterm_difficulties, Threatening_experiences, Smoking_Packyears, Alcohol_frequency, Chronotype, Sleep_weekly, Childhood_sexual_abuse, and age_at_first_AD.

sex_at_birth: binary term (sex_at_birthMale); reference = Female.

Depression, Lifetime_anxiety, Fibromyalgia: binary terms (level "1" shown); reference = absence of the condition (coded 0).

| Population | ClinicalGroup | Outcome | Dependent | variable | odds_ratio | lower_95 | upper_95 | p_value | Bonf_p | N_Total | N_Referer | N_Case |
|---|---|---|---|---|---|---|---|---|---|---|---|---|
| European | Full Cohort | Switching vs Continuation | age_at_first_AD | (Intercept) | 0.282 | 0.261 | 0.304 | <2.2e-16 | <2.2e-16 | 5493 | 4208 | 1285 |
| European | Full Cohort | Switching vs Continuation | age_at_first_AD | age_at_first_AD | 0.810 | 0.755 | 0.868 | 3.12e-09 | 5.31e-08 | 5493 | 4208 | 1285 |
| European | Full Cohort | Switching vs Continuation | age_at_first_AD | sex_at_birthMale | 1.229 | 1.072 | 1.409 | 3.06e-03 | 5.21e-02 | 5493 | 4208 | 1285 |
| European | Full Cohort | Discontinuation vs Continuation | age_at_first_AD | (Intercept) | 0.304 | 0.283 | 0.328 | <2.2e-16 | <2.2e-16 | 5629 | 4208 | 1421 |
| European | Full Cohort | Discontinuation vs Continuation | age_at_first_AD | age_at_first_AD | 1.263 | 1.186 | 1.345 | 3.81e-13 | 6.48e-12 | 5629 | 4208 | 1421 |
| European | Full Cohort | Discontinuation vs Continuation | age_at_first_AD | sex_at_birthMale | 1.297 | 1.139 | 1.476 | 8.33e-05 | 1.42e-03 | 5629 | 4208 | 1421 |
| European | Full Cohort | Switching vs Continuation | sex_at_birth | (Intercept) | 0.282 | 0.261 | 0.304 | <2.2e-16 | <2.2e-16 | 5493 | 4208 | 1285 |
| European | Full Cohort | Switching vs Continuation | sex_at_birth | age_at_first_AD | 0.810 | 0.755 | 0.868 | 3.12e-09 | 5.31e-08 | 5493 | 4208 | 1285 |
| European | Full Cohort | Switching vs Continuation | sex_at_birth | sex_at_birthMale | 1.229 | 1.072 | 1.409 | 3.06e-03 | 5.21e-02 | 5493 | 4208 | 1285 |
| European | Full Cohort | Discontinuation vs Continuation | sex_at_birth | (Intercept) | 0.304 | 0.283 | 0.328 | <2.2e-16 | <2.2e-16 | 5629 | 4208 | 1421 |
| European | Full Cohort | Discontinuation vs Continuation | sex_at_birth | age_at_first_AD | 1.263 | 1.186 | 1.345 | 3.81e-13 | 6.48e-12 | 5629 | 4208 | 1421 |
| European | Full Cohort | Discontinuation vs Continuation | sex_at_birth | sex_at_birthMale | 1.297 | 1.139 | 1.476 | 8.33e-05 | 1.42e-03 | 5629 | 4208 | 1421 |
| European | Full Cohort | Switching vs Continuation | Education_level | (Intercept) | 0.282 | 0.261 | 0.304 | <2.2e-16 | <2.2e-16 | 5493 | 4208 | 1285 |
| European | Full Cohort | Switching vs Continuation | Education_level | age_at_first_AD | 0.809 | 0.752 | 0.869 | 7.03e-09 | 1.20e-07 | 5493 | 4208 | 1285 |
| European | Full Cohort | Switching vs Continuation | Education_level | sex_at_birthMale | 1.254 | 1.093 | 1.439 | 1.21e-03 | 2.06e-02 | 5493 | 4208 | 1285 |
| European | Full Cohort | Switching vs Continuation | Education_level | Income_level | 0.914 | 0.854 | 0.979 | 9.88e-03 | 1.68e-01 | 5493 | 4208 | 1285 |
| European | Full Cohort | Switching vs Continuation | Education_level | Education_level | 0.943 | 0.881 | 1.010 | 9.52e-02 | 1.00e+00 | 5493 | 4208 | 1285 |
| European | Full Cohort | Discontinuation vs Continuation | Education_level | (Intercept) | 0.306 | 0.284 | 0.330 | <2.2e-16 | <2.2e-16 | 5629 | 4208 | 1421 |
| European | Full Cohort | Discontinuation vs Continuation | Education_level | age_at_first_AD | 1.218 | 1.141 | 1.300 | 2.75e-09 | 4.68e-08 | 5629 | 4208 | 1421 |
| European | Full Cohort | Discontinuation vs Continuation | Education_level | sex_at_birthMale | 1.299 | 1.141 | 1.480 | 8.09e-05 | 1.38e-03 | 5629 | 4208 | 1421 |
| European | Full Cohort | Discontinuation vs Continuation | Education_level | Income_level | 1.033 | 0.969 | 1.102 | 3.16e-01 | 1.00e+00 | 5629 | 4208 | 1421 |
| European | Full Cohort | Discontinuation vs Continuation | Education_level | Education_level | 0.847 | 0.792 | 0.905 | 9.92e-07 | 1.69e-05 | 5629 | 4208 | 1421 |
| European | Full Cohort | Switching vs Continuation | BMI | (Intercept) | 0.282 | 0.261 | 0.304 | <2.2e-16 | <2.2e-16 | 5487 | 4203 | 1284 |
| European | Full Cohort | Switching vs Continuation | BMI | age_at_first_AD | 0.805 | 0.749 | 0.866 | 4.65e-09 | 7.90e-08 | 5487 | 4203 | 1284 |
| European | Full Cohort | Switching vs Continuation | BMI | sex_at_birthMale | 1.254 | 1.093 | 1.438 | 1.26e-03 | 2.14e-02 | 5487 | 4203 | 1284 |
| European | Full Cohort | Switching vs Continuation | BMI | Income_level | 0.917 | 0.856 | 0.981 | 1.24e-02 | 2.11e-01 | 5487 | 4203 | 1284 |
| European | Full Cohort | Switching vs Continuation | BMI | Education_level | 0.950 | 0.887 | 1.018 | 1.44e-01 | 1.00e+00 | 5487 | 4203 | 1284 |
| European | Full Cohort | Switching vs Continuation | BMI | BMI | 1.043 | 0.978 | 1.113 | 2.03e-01 | 1.00e+00 | 5487 | 4203 | 1284 |
| European | Full Cohort | Discontinuation vs Continuation | BMI | (Intercept) | 0.305 | 0.283 | 0.329 | <2.2e-16 | <2.2e-16 | 5624 | 4203 | 1421 |
| European | Full Cohort | Discontinuation vs Continuation | BMI | age_at_first_AD | 1.225 | 1.148 | 1.308 | 1.08e-09 | 1.83e-08 | 5624 | 4203 | 1421 |
| European | Full Cohort | Discontinuation vs Continuation | BMI | sex_at_birthMale | 1.309 | 1.149 | 1.492 | 5.12e-05 | 8.70e-04 | 5624 | 4203 | 1421 |
| European | Full Cohort | Discontinuation vs Continuation | BMI | Income_level | 1.031 | 0.967 | 1.099 | 3.53e-01 | 1.00e+00 | 5624 | 4203 | 1421 |
| European | Full Cohort | Discontinuation vs Continuation | BMI | Education_level | 0.841 | 0.786 | 0.900 | 4.45e-07 | 7.56e-06 | 5624 | 4203 | 1421 |

| | | | | | | | | | | | | |
|---|---|---|---|---|---|---|---|---|---|---|---|---|
| European | Full Cohort | Discontinuation vs Continuation | BMI | BMI | 0.945 | 0.884 | 1.010 | 9.59e-02 | 1.00e+00 | 5624 | 4203 | 1421 |
| European | Full Cohort | Switching vs Continuation | Income_level | (Intercept) | 0.282 | 0.261 | 0.304 | <2.2e-16 | <2.2e-16 | 5493 | 4208 | 1285 |
| European | Full Cohort | Switching vs Continuation | Income_level | age_at_first_AD | 0.809 | 0.752 | 0.869 | 7.03e-09 | 1.20e-07 | 5493 | 4208 | 1285 |
| European | Full Cohort | Switching vs Continuation | Income_level | sex_at_birthMale | 1.254 | 1.093 | 1.439 | 1.21e-03 | 2.06e-02 | 5493 | 4208 | 1285 |
| European | Full Cohort | Switching vs Continuation | Income_level | Income_level | 0.914 | 0.854 | 0.979 | 9.88e-03 | 1.68e-01 | 5493 | 4208 | 1285 |
| European | Full Cohort | Switching vs Continuation | Income_level | Education_level | 0.943 | 0.881 | 1.010 | 9.52e-02 | 1.00e+00 | 5493 | 4208 | 1285 |
| European | Full Cohort | Discontinuation vs Continuation | Income_level | (Intercept) | 0.306 | 0.284 | 0.330 | <2.2e-16 | <2.2e-16 | 5629 | 4208 | 1421 |
| European | Full Cohort | Discontinuation vs Continuation | Income_level | age_at_first_AD | 1.218 | 1.141 | 1.300 | 2.75e-09 | 4.68e-08 | 5629 | 4208 | 1421 |
| European | Full Cohort | Discontinuation vs Continuation | Income_level | sex_at_birthMale | 1.299 | 1.141 | 1.480 | 8.09e-05 | 1.38e-03 | 5629 | 4208 | 1421 |
| European | Full Cohort | Discontinuation vs Continuation | Income_level | Income_level | 1.033 | 0.969 | 1.102 | 3.16e-01 | 1.00e+00 | 5629 | 4208 | 1421 |
| European | Full Cohort | Discontinuation vs Continuation | Income_level | Education_level | 0.847 | 0.792 | 0.905 | 9.92e-07 | 1.69e-05 | 5629 | 4208 | 1421 |
| European | Full Cohort | Switching vs Continuation | Depression | (Intercept) | 0.219 | 0.198 | 0.242 | <2.2e-16 | <2.2e-16 | 5433 | 4168 | 1265 |
| European | Full Cohort | Switching vs Continuation | Depression | age_at_first_AD | 0.837 | 0.777 | 0.900 | 2.02e-06 | 3.43e-05 | 5433 | 4168 | 1265 |
| European | Full Cohort | Switching vs Continuation | Depression | sex_at_birthMale | 1.310 | 1.140 | 1.505 | 1.43e-04 | 2.42e-03 | 5433 | 4168 | 1265 |
| European | Full Cohort | Switching vs Continuation | Depression | Income_level | 0.918 | 0.856 | 0.983 | 1.47e-02 | 2.51e-01 | 5433 | 4168 | 1265 |
| European | Full Cohort | Switching vs Continuation | Depression | Education_level | 0.925 | 0.863 | 0.992 | 2.79e-02 | 4.74e-01 | 5433 | 4168 | 1265 |
| European | Full Cohort | Switching vs Continuation | Depression | Depression1 | 1.707 | 1.501 | 1.941 | 3.28e-16 | 5.57e-15 | 5433 | 4168 | 1265 |
| European | Full Cohort | Discontinuation vs Continuation | Depression | (Intercept) | 0.361 | 0.332 | 0.393 | <2.2e-16 | <2.2e-16 | 5577 | 4168 | 1409 |
| European | Full Cohort | Discontinuation vs Continuation | Depression | age_at_first_AD | 1.191 | 1.115 | 1.272 | 1.89e-07 | 3.21e-06 | 5577 | 4168 | 1409 |
| European | Full Cohort | Discontinuation vs Continuation | Depression | sex_at_birthMale | 1.281 | 1.123 | 1.461 | 2.25e-04 | 3.82e-03 | 5577 | 4168 | 1409 |
| European | Full Cohort | Discontinuation vs Continuation | Depression | Income_level | 1.027 | 0.963 | 1.096 | 4.13e-01 | 1.00e+00 | 5577 | 4168 | 1409 |
| European | Full Cohort | Discontinuation vs Continuation | Depression | Education_level | 0.863 | 0.806 | 0.923 | 1.74e-05 | 2.96e-04 | 5577 | 4168 | 1409 |
| European | Full Cohort | Discontinuation vs Continuation | Depression | Depression1 | 0.609 | 0.531 | 0.698 | 1.32e-12 | 2.25e-11 | 5577 | 4168 | 1409 |
| European | Full Cohort | Switching vs Continuation | Dep_sum_score | (Intercept) | 0.260 | 0.240 | 0.282 | <2.2e-16 | <2.2e-16 | 5433 | 4168 | 1265 |
| European | Full Cohort | Switching vs Continuation | Dep_sum_score | age_at_first_AD | 0.859 | 0.797 | 0.925 | 6.45e-05 | 1.10e-03 | 5433 | 4168 | 1265 |
| European | Full Cohort | Switching vs Continuation | Dep_sum_score | sex_at_birthMale | 1.302 | 1.131 | 1.500 | 2.44e-04 | 4.14e-03 | 5433 | 4168 | 1265 |
| European | Full Cohort | Switching vs Continuation | Dep_sum_score | Income_level | 0.944 | 0.880 | 1.013 | 1.10e-01 | 1.00e+00 | 5433 | 4168 | 1265 |
| European | Full Cohort | Switching vs Continuation | Dep_sum_score | Education_level | 0.971 | 0.905 | 1.041 | 4.07e-01 | 1.00e+00 | 5433 | 4168 | 1265 |
| European | Full Cohort | Switching vs Continuation | Dep_sum_score | Dep_sum_score | 1.540 | 1.453 | 1.633 | <2.2e-16 | <2.2e-16 | 5433 | 4168 | 1265 |
| European | Full Cohort | Discontinuation vs Continuation | Dep_sum_score | (Intercept) | 0.301 | 0.279 | 0.324 | <2.2e-16 | <2.2e-16 | 5577 | 4168 | 1409 |
| European | Full Cohort | Discontinuation vs Continuation | Dep_sum_score | age_at_first_AD | 1.207 | 1.130 | 1.289 | 2.06e-08 | 3.51e-07 | 5577 | 4168 | 1409 |
| European | Full Cohort | Discontinuation vs Continuation | Dep_sum_score | sex_at_birthMale | 1.300 | 1.141 | 1.482 | 8.57e-05 | 1.46e-03 | 5577 | 4168 | 1409 |
| European | Full Cohort | Discontinuation vs Continuation | Dep_sum_score | Income_level | 1.022 | 0.958 | 1.090 | 5.17e-01 | 1.00e+00 | 5577 | 4168 | 1409 |
| European | Full Cohort | Discontinuation vs Continuation | Dep_sum_score | Education_level | 0.844 | 0.789 | 0.903 | 7.22e-07 | 1.23e-05 | 5577 | 4168 | 1409 |
| European | Full Cohort | Discontinuation vs Continuation | Dep_sum_score | Dep_sum_score | 0.862 | 0.799 | 0.929 | 1.06e-04 | 1.81e-03 | 5577 | 4168 | 1409 |
| European | Full Cohort | Switching vs Continuation | Lifetime_anxiety | (Intercept) | 0.241 | 0.205 | 0.284 | <2.2e-16 | <2.2e-16 | 1705 | 1310 | 395 |
| European | Full Cohort | Switching vs Continuation | Lifetime_anxiety | age_at_first_AD | 0.858 | 0.744 | 0.988 | 3.34e-02 | 5.68e-01 | 1705 | 1310 | 395 |
| European | Full Cohort | Switching vs Continuation | Lifetime_anxiety | sex_at_birthMale | 1.216 | 0.947 | 1.562 | 1.26e-01 | 1.00e+00 | 1705 | 1310 | 395 |
| European | Full Cohort | Switching vs Continuation | Lifetime_anxiety | Income_level | 0.913 | 0.809 | 1.031 | 1.42e-01 | 1.00e+00 | 1705 | 1310 | 395 |
| European | Full Cohort | Switching vs Continuation | Lifetime_anxiety | Education_level | 0.905 | 0.801 | 1.022 | 1.07e-01 | 1.00e+00 | 1705 | 1310 | 395 |
| European | Full Cohort | Switching vs Continuation | Lifetime_anxiety | Lifetime_anxiety1 | 1.846 | 1.454 | 2.344 | 4.86e-07 | 8.26e-06 | 1705 | 1310 | 395 |
| European | Full Cohort | Discontinuation vs Continuation | Lifetime_anxiety | (Intercept) | 0.342 | 0.295 | 0.397 | <2.2e-16 | <2.2e-16 | 1729 | 1310 | 419 |
| European | Full Cohort | Discontinuation vs Continuation | Lifetime_anxiety | age_at_first_AD | 1.309 | 1.147 | 1.494 | 6.42e-05 | 1.09e-03 | 1729 | 1310 | 419 |
| European | Full Cohort | Discontinuation vs Continuation | Lifetime_anxiety | sex_at_birthMale | 1.181 | 0.926 | 1.506 | 1.79e-01 | 1.00e+00 | 1729 | 1310 | 419 |
| European | Full Cohort | Discontinuation vs Continuation | Lifetime_anxiety | Income_level | 0.943 | 0.838 | 1.061 | 3.28e-01 | 1.00e+00 | 1729 | 1310 | 419 |
| European | Full Cohort | Discontinuation vs Continuation | Lifetime_anxiety | Education_level | 0.850 | 0.752 | 0.960 | 8.65e-03 | 1.47e-01 | 1729 | 1310 | 419 |
| European | Full Cohort | Discontinuation vs Continuation | Lifetime_anxiety | Lifetime_anxiety1 | 0.477 | 0.350 | 0.652 | 3.28e-06 | 5.57e-05 | 1729 | 1310 | 419 |
| European | Full Cohort | Switching vs Continuation | Fibromyalgia | (Intercept) | 0.269 | 0.240 | 0.301 | <2.2e-16 | <2.2e-16 | 2963 | 2281 | 682 |
| European | Full Cohort | Switching vs Continuation | Fibromyalgia | age_at_first_AD | 0.821 | 0.741 | 0.909 | 1.52e-04 | 2.59e-03 | 2963 | 2281 | 682 |
| European | Full Cohort | Switching vs Continuation | Fibromyalgia | sex_at_birthMale | 1.228 | 1.016 | 1.485 | 3.36e-02 | 5.71e-01 | 2963 | 2281 | 682 |
| European | Full Cohort | Switching vs Continuation | Fibromyalgia | Income_level | 0.965 | 0.879 | 1.059 | 4.53e-01 | 1.00e+00 | 2963 | 2281 | 682 |

| | | | | | | | | | | | | |
|---|---|---|---|---|---|---|---|---|---|---|---|---|
| European | Full Cohort | Switching vs Continuation | Fibromyalgia | Education_level | 0.934 | 0.850 | 1.026 | 1.56e-01 | 1.00e+00 | 2963 | 2281 | 682 |
| European | Full Cohort | Switching vs Continuation | Fibromyalgia | Fibromyalgia1 | 1.379 | 1.097 | 1.735 | 5.96e-03 | 1.01e-01 | 2963 | 2281 | 682 |
| European | Full Cohort | Discontinuation vs Continuation | Fibromyalgia | (Intercept) | 0.310 | 0.279 | 0.346 | <2.2e-16 | <2.2e-16 | 3049 | 2281 | 768 |
| European | Full Cohort | Discontinuation vs Continuation | Fibromyalgia | age_at_first_AD | 1.209 | 1.103 | 1.326 | 5.21e-05 | 8.85e-04 | 3049 | 2281 | 768 |
| European | Full Cohort | Discontinuation vs Continuation | Fibromyalgia | sex_at_birthMale | 1.264 | 1.058 | 1.512 | 1.00e-02 | 1.70e-01 | 3049 | 2281 | 768 |
| European | Full Cohort | Discontinuation vs Continuation | Fibromyalgia | Income_level | 1.030 | 0.945 | 1.124 | 4.97e-01 | 1.00e+00 | 3049 | 2281 | 768 |
| European | Full Cohort | Discontinuation vs Continuation | Fibromyalgia | Education_level | 0.834 | 0.762 | 0.914 | 9.09e-05 | 1.55e-03 | 3049 | 2281 | 768 |
| European | Full Cohort | Discontinuation vs Continuation | Fibromyalgia | Fibromyalgia1 | 0.879 | 0.686 | 1.126 | 3.08e-01 | 1.00e+00 | 3049 | 2281 | 768 |
| European | Full Cohort | Switching vs Continuation | Childhood_sexual_abuse | (Intercept) | 0.288 | 0.257 | 0.323 | <2.2e-16 | <2.2e-16 | 2438 | 1860 | 578 |
| European | Full Cohort | Switching vs Continuation | Childhood_sexual_abuse | age_at_first_AD | 0.840 | 0.753 | 0.937 | 1.79e-03 | 3.04e-02 | 2438 | 1860 | 578 |
| European | Full Cohort | Switching vs Continuation | Childhood_sexual_abuse | sex_at_birthMale | 1.281 | 1.040 | 1.576 | 1.96e-02 | 3.32e-01 | 2438 | 1860 | 578 |
| European | Full Cohort | Switching vs Continuation | Childhood_sexual_abuse | Income_level | 0.959 | 0.866 | 1.061 | 4.12e-01 | 1.00e+00 | 2438 | 1860 | 578 |
| European | Full Cohort | Switching vs Continuation | Childhood_sexual_abuse | Education_level | 0.927 | 0.839 | 1.025 | 1.42e-01 | 1.00e+00 | 2438 | 1860 | 578 |
| European | Full Cohort | Switching vs Continuation | Childhood_sexual_abuse | Childhood_sexual_a | 1.234 | 1.141 | 1.333 | 1.19e-07 | 2.03e-06 | 2438 | 1860 | 578 |
| European | Full Cohort | Discontinuation vs Continuation | Childhood_sexual_abuse | (Intercept) | 0.296 | 0.264 | 0.332 | <2.2e-16 | <2.2e-16 | 2512 | 1860 | 652 |
| European | Full Cohort | Discontinuation vs Continuation | Childhood_sexual_abuse | age_at_first_AD | 1.369 | 1.240 | 1.512 | 5.54e-10 | 9.42e-09 | 2512 | 1860 | 652 |
| European | Full Cohort | Discontinuation vs Continuation | Childhood_sexual_abuse | sex_at_birthMale | 1.299 | 1.069 | 1.577 | 8.35e-03 | 1.42e-01 | 2512 | 1860 | 652 |
| European | Full Cohort | Discontinuation vs Continuation | Childhood_sexual_abuse | Income_level | 0.982 | 0.894 | 1.079 | 7.06e-01 | 1.00e+00 | 2512 | 1860 | 652 |
| European | Full Cohort | Discontinuation vs Continuation | Childhood_sexual_abuse | Education_level | 0.870 | 0.789 | 0.959 | 5.00e-03 | 8.50e-02 | 2512 | 1860 | 652 |
| European | Full Cohort | Discontinuation vs Continuation | Childhood_sexual_abuse | Childhood_sexual_a | 0.935 | 0.837 | 1.043 | 2.28e-01 | 1.00e+00 | 2512 | 1860 | 652 |
| European | Full Cohort | Switching vs Continuation | Neuroticism | (Intercept) | 0.265 | 0.244 | 0.288 | <2.2e-16 | <2.2e-16 | 5089 | 3892 | 1197 |
| European | Full Cohort | Switching vs Continuation | Neuroticism | age_at_first_AD | 0.812 | 0.747 | 0.882 | 7.94e-07 | 1.35e-05 | 5089 | 3892 | 1197 |
| European | Full Cohort | Switching vs Continuation | Neuroticism | sex_at_birthMale | 1.317 | 1.139 | 1.522 | 1.98e-04 | 3.37e-03 | 5089 | 3892 | 1197 |
| European | Full Cohort | Switching vs Continuation | Neuroticism | Income_level | 0.950 | 0.885 | 1.020 | 1.60e-01 | 1.00e+00 | 5089 | 3892 | 1197 |
| European | Full Cohort | Switching vs Continuation | Neuroticism | Education_level | 0.958 | 0.891 | 1.029 | 2.34e-01 | 1.00e+00 | 5089 | 3892 | 1197 |
| European | Full Cohort | Switching vs Continuation | Neuroticism | Neuroticism | 1.338 | 1.250 | 1.432 | <2.2e-16 | 6.05e-16 | 5089 | 3892 | 1197 |
| European | Full Cohort | Discontinuation vs Continuation | Neuroticism | (Intercept) | 0.305 | 0.282 | 0.330 | <2.2e-16 | <2.2e-16 | 5168 | 3892 | 1276 |
| European | Full Cohort | Discontinuation vs Continuation | Neuroticism | age_at_first_AD | 1.141 | 1.054 | 1.234 | 1.09e-03 | 1.86e-02 | 5168 | 3892 | 1276 |
| European | Full Cohort | Discontinuation vs Continuation | Neuroticism | sex_at_birthMale | 1.223 | 1.066 | 1.405 | 4.23e-03 | 7.20e-02 | 5168 | 3892 | 1276 |
| European | Full Cohort | Discontinuation vs Continuation | Neuroticism | Income_level | 1.028 | 0.960 | 1.099 | 4.30e-01 | 1.00e+00 | 5168 | 3892 | 1276 |
| European | Full Cohort | Discontinuation vs Continuation | Neuroticism | Education_level | 0.826 | 0.770 | 0.887 | 1.18e-07 | 2.00e-06 | 5168 | 3892 | 1276 |
| European | Full Cohort | Discontinuation vs Continuation | Neuroticism | Neuroticism | 0.810 | 0.756 | 0.867 | 1.89e-09 | 3.22e-08 | 5168 | 3892 | 1276 |
| European | Full Cohort | Switching vs Continuation | Longterm_difficulties | (Intercept) | 0.273 | 0.252 | 0.296 | <2.2e-16 | <2.2e-16 | 5317 | 4074 | 1243 |
| European | Full Cohort | Switching vs Continuation | Longterm_difficulties | age_at_first_AD | 0.857 | 0.793 | 0.925 | 7.70e-05 | 1.31e-03 | 5317 | 4074 | 1243 |
| European | Full Cohort | Switching vs Continuation | Longterm_difficulties | sex_at_birthMale | 1.288 | 1.119 | 1.482 | 4.19e-04 | 7.12e-03 | 5317 | 4074 | 1243 |
| European | Full Cohort | Switching vs Continuation | Longterm_difficulties | Income_level | 0.959 | 0.894 | 1.029 | 2.48e-01 | 1.00e+00 | 5317 | 4074 | 1243 |
| European | Full Cohort | Switching vs Continuation | Longterm_difficulties | Education_level | 0.937 | 0.874 | 1.005 | 6.86e-02 | 1.00e+00 | 5317 | 4074 | 1243 |
| European | Full Cohort | Switching vs Continuation | Longterm_difficulties | Longterm_difficultie | 1.254 | 1.177 | 1.336 | 2.97e-12 | 5.06e-11 | 5317 | 4074 | 1243 |
| European | Full Cohort | Discontinuation vs Continuation | Longterm_difficulties | (Intercept) | 0.310 | 0.288 | 0.334 | <2.2e-16 | <2.2e-16 | 5459 | 4074 | 1385 |
| European | Full Cohort | Discontinuation vs Continuation | Longterm_difficulties | age_at_first_AD | 1.214 | 1.133 | 1.301 | 3.48e-08 | 5.91e-07 | 5459 | 4074 | 1385 |
| European | Full Cohort | Discontinuation vs Continuation | Longterm_difficulties | sex_at_birthMale | 1.265 | 1.108 | 1.444 | 4.97e-04 | 8.45e-03 | 5459 | 4074 | 1385 |
| European | Full Cohort | Discontinuation vs Continuation | Longterm_difficulties | Income_level | 1.031 | 0.966 | 1.101 | 3.62e-01 | 1.00e+00 | 5459 | 4074 | 1385 |
| European | Full Cohort | Discontinuation vs Continuation | Longterm_difficulties | Education_level | 0.854 | 0.798 | 0.914 | 5.14e-06 | 8.75e-05 | 5459 | 4074 | 1385 |
| European | Full Cohort | Discontinuation vs Continuation | Longterm_difficulties | Longterm_difficultie | 0.950 | 0.886 | 1.018 | 1.43e-01 | 1.00e+00 | 5459 | 4074 | 1385 |
| European | Full Cohort | Switching vs Continuation | Threatening_experiences | (Intercept) | 0.280 | 0.259 | 0.303 | <2.2e-16 | <2.2e-16 | 5380 | 4123 | 1257 |
| European | Full Cohort | Switching vs Continuation | Threatening_experiences | age_at_first_AD | 0.806 | 0.750 | 0.868 | 8.06e-09 | 1.37e-07 | 5380 | 4123 | 1257 |
| European | Full Cohort | Switching vs Continuation | Threatening_experiences | sex_at_birthMale | 1.260 | 1.097 | 1.448 | 1.09e-03 | 1.85e-02 | 5380 | 4123 | 1257 |
| European | Full Cohort | Switching vs Continuation | Threatening_experiences | Income_level | 0.937 | 0.874 | 1.004 | 6.45e-02 | 1.00e+00 | 5380 | 4123 | 1257 |
| European | Full Cohort | Switching vs Continuation | Threatening_experiences | Education_level | 0.956 | 0.892 | 1.024 | 1.99e-01 | 1.00e+00 | 5380 | 4123 | 1257 |
| European | Full Cohort | Switching vs Continuation | Threatening_experiences | Threatening_experie | 1.103 | 1.035 | 1.176 | 2.70e-03 | 4.59e-02 | 5380 | 4123 | 1257 |
| European | Full Cohort | Discontinuation vs Continuation | Threatening_experiences | (Intercept) | 0.307 | 0.285 | 0.331 | <2.2e-16 | <2.2e-16 | 5515 | 4123 | 1392 |

| | | | | | | | | | | | | |
|---|---|---|---|---|---|---|---|---|---|---|---|---|
| European | Full Cohort | Discontinuation vs Continuation | Threatening_experiences | age_at_first_AD | 1.226 | 1.148 | 1.310 | 1.54e-09 | 2.61e-08 | 5515 | 4123 | 1392 |
| European | Full Cohort | Discontinuation vs Continuation | Threatening_experiences | sex_at_birthMale | 1.289 | 1.130 | 1.470 | 1.53e-04 | 2.60e-03 | 5515 | 4123 | 1392 |
| European | Full Cohort | Discontinuation vs Continuation | Threatening_experiences | Income_level | 1.034 | 0.969 | 1.103 | 3.17e-01 | 1.00e+00 | 5515 | 4123 | 1392 |
| European | Full Cohort | Discontinuation vs Continuation | Threatening_experiences | Education_level | 0.856 | 0.800 | 0.915 | 5.54e-06 | 9.42e-05 | 5515 | 4123 | 1392 |
| European | Full Cohort | Discontinuation vs Continuation | Threatening_experiences | Threatening_experie | 1.028 | 0.962 | 1.098 | 4.14e-01 | 1.00e+00 | 5515 | 4123 | 1392 |
| European | Full Cohort | Switching vs Continuation | Smoking_Packyears | (Intercept) | 0.301 | 0.272 | 0.333 | <2.2e-16 | <2.2e-16 | 3171 | 2401 | 770 |
| European | Full Cohort | Switching vs Continuation | Smoking_Packyears | age_at_first_AD | 0.688 | 0.623 | 0.761 | 2.51e-13 | 4.28e-12 | 3171 | 2401 | 770 |
| European | Full Cohort | Switching vs Continuation | Smoking_Packyears | sex_at_birthMale | 1.132 | 0.946 | 1.355 | 1.75e-01 | 1.00e+00 | 3171 | 2401 | 770 |
| European | Full Cohort | Switching vs Continuation | Smoking_Packyears | Income_level | 0.968 | 0.886 | 1.057 | 4.65e-01 | 1.00e+00 | 3171 | 2401 | 770 |
| European | Full Cohort | Switching vs Continuation | Smoking_Packyears | Education_level | 0.940 | 0.859 | 1.029 | 1.78e-01 | 1.00e+00 | 3171 | 2401 | 770 |
| European | Full Cohort | Switching vs Continuation | Smoking_Packyears | Smoking_Packyears | 1.251 | 1.149 | 1.363 | 2.49e-07 | 4.22e-06 | 3171 | 2401 | 770 |
| European | Full Cohort | Discontinuation vs Continuation | Smoking_Packyears | (Intercept) | 0.317 | 0.287 | 0.350 | <2.2e-16 | <2.2e-16 | 3281 | 2401 | 880 |
| European | Full Cohort | Discontinuation vs Continuation | Smoking_Packyears | age_at_first_AD | 1.216 | 1.115 | 1.326 | 1.07e-05 | 1.82e-04 | 3281 | 2401 | 880 |
| European | Full Cohort | Discontinuation vs Continuation | Smoking_Packyears | sex_at_birthMale | 1.361 | 1.153 | 1.607 | 2.72e-04 | 4.62e-03 | 3281 | 2401 | 880 |
| European | Full Cohort | Discontinuation vs Continuation | Smoking_Packyears | Income_level | 1.031 | 0.949 | 1.119 | 4.74e-01 | 1.00e+00 | 3281 | 2401 | 880 |
| European | Full Cohort | Discontinuation vs Continuation | Smoking_Packyears | Education_level | 0.847 | 0.777 | 0.923 | 1.59e-04 | 2.71e-03 | 3281 | 2401 | 880 |
| European | Full Cohort | Discontinuation vs Continuation | Smoking_Packyears | Smoking_Packyears | 1.072 | 0.989 | 1.163 | 9.16e-02 | 1.00e+00 | 3281 | 2401 | 880 |
| European | Full Cohort | Switching vs Continuation | Alcohol_intake | (Intercept) | 0.276 | 0.250 | 0.306 | <2.2e-16 | <2.2e-16 | 3508 | 2710 | 798 |
| European | Full Cohort | Switching vs Continuation | Alcohol_intake | age_at_first_AD | 0.802 | 0.733 | 0.878 | 1.75e-06 | 2.98e-05 | 3508 | 2710 | 798 |
| European | Full Cohort | Switching vs Continuation | Alcohol_intake | sex_at_birthMale | 1.224 | 1.026 | 1.460 | 2.51e-02 | 4.26e-01 | 3508 | 2710 | 798 |
| European | Full Cohort | Switching vs Continuation | Alcohol_intake | Income_level | 0.947 | 0.870 | 1.030 | 2.02e-01 | 1.00e+00 | 3508 | 2710 | 798 |
| European | Full Cohort | Switching vs Continuation | Alcohol_intake | Education_level | 0.914 | 0.839 | 0.995 | 3.78e-02 | 6.42e-01 | 3508 | 2710 | 798 |
| European | Full Cohort | Switching vs Continuation | Alcohol_intake | Alcohol_intake | 0.978 | 0.898 | 1.064 | 6.03e-01 | 1.00e+00 | 3508 | 2710 | 798 |
| European | Full Cohort | Discontinuation vs Continuation | Alcohol_intake | (Intercept) | 0.312 | 0.283 | 0.344 | <2.2e-16 | <2.2e-16 | 3606 | 2710 | 896 |
| European | Full Cohort | Discontinuation vs Continuation | Alcohol_intake | age_at_first_AD | 1.270 | 1.171 | 1.378 | 7.82e-09 | 1.33e-07 | 3606 | 2710 | 896 |
| European | Full Cohort | Discontinuation vs Continuation | Alcohol_intake | sex_at_birthMale | 1.141 | 0.964 | 1.350 | 1.25e-01 | 1.00e+00 | 3606 | 2710 | 896 |
| European | Full Cohort | Discontinuation vs Continuation | Alcohol_intake | Income_level | 1.041 | 0.961 | 1.127 | 3.23e-01 | 1.00e+00 | 3606 | 2710 | 896 |
| European | Full Cohort | Discontinuation vs Continuation | Alcohol_intake | Education_level | 0.840 | 0.773 | 0.912 | 3.54e-05 | 6.01e-04 | 3606 | 2710 | 896 |
| European | Full Cohort | Discontinuation vs Continuation | Alcohol_intake | Alcohol_intake | 1.108 | 1.026 | 1.197 | 9.14e-03 | 1.55e-01 | 3606 | 2710 | 896 |
| European | Full Cohort | Switching vs Continuation | Chronotype | (Intercept) | 0.277 | 0.253 | 0.302 | <2.2e-16 | <2.2e-16 | 4223 | 3248 | 975 |
| European | Full Cohort | Switching vs Continuation | Chronotype | age_at_first_AD | 0.823 | 0.757 | 0.895 | 5.40e-06 | 9.17e-05 | 4223 | 3248 | 975 |
| European | Full Cohort | Switching vs Continuation | Chronotype | sex_at_birthMale | 1.284 | 1.097 | 1.501 | 1.81e-03 | 3.07e-02 | 4223 | 3248 | 975 |
| European | Full Cohort | Switching vs Continuation | Chronotype | Income_level | 0.889 | 0.822 | 0.962 | 3.27e-03 | 5.56e-02 | 4223 | 3248 | 975 |
| European | Full Cohort | Switching vs Continuation | Chronotype | Education_level | 0.949 | 0.878 | 1.025 | 1.84e-01 | 1.00e+00 | 4223 | 3248 | 975 |
| European | Full Cohort | Switching vs Continuation | Chronotype | Chronotype | 1.118 | 1.042 | 1.200 | 1.97e-03 | 3.35e-02 | 4223 | 3248 | 975 |
| European | Full Cohort | Discontinuation vs Continuation | Chronotype | (Intercept) | 0.308 | 0.283 | 0.336 | <2.2e-16 | <2.2e-16 | 4347 | 3248 | 1099 |
| European | Full Cohort | Discontinuation vs Continuation | Chronotype | age_at_first_AD | 1.236 | 1.146 | 1.334 | 4.27e-08 | 7.26e-07 | 4347 | 3248 | 1099 |
| European | Full Cohort | Discontinuation vs Continuation | Chronotype | sex_at_birthMale | 1.246 | 1.074 | 1.445 | 3.61e-03 | 6.13e-02 | 4347 | 3248 | 1099 |
| European | Full Cohort | Discontinuation vs Continuation | Chronotype | Income_level | 1.032 | 0.960 | 1.110 | 3.94e-01 | 1.00e+00 | 4347 | 3248 | 1099 |
| European | Full Cohort | Discontinuation vs Continuation | Chronotype | Education_level | 0.877 | 0.813 | 0.945 | 5.65e-04 | 9.60e-03 | 4347 | 3248 | 1099 |
| European | Full Cohort | Discontinuation vs Continuation | Chronotype | Chronotype | 1.007 | 0.937 | 1.081 | 8.57e-01 | 1.00e+00 | 4347 | 3248 | 1099 |
| European | Full Cohort | Switching vs Continuation | Sleep_weekly | (Intercept) | 0.276 | 0.254 | 0.300 | <2.2e-16 | <2.2e-16 | 4859 | 3734 | 1125 |
| European | Full Cohort | Switching vs Continuation | Sleep_weekly | age_at_first_AD | 0.816 | 0.754 | 0.882 | 3.11e-07 | 5.28e-06 | 4859 | 3734 | 1125 |
| European | Full Cohort | Switching vs Continuation | Sleep_weekly | sex_at_birthMale | 1.301 | 1.122 | 1.508 | 4.89e-04 | 8.31e-03 | 4859 | 3734 | 1125 |
| European | Full Cohort | Switching vs Continuation | Sleep_weekly | Income_level | 0.893 | 0.831 | 0.961 | 2.37e-03 | 4.04e-02 | 4859 | 3734 | 1125 |
| European | Full Cohort | Switching vs Continuation | Sleep_weekly | Education_level | 0.945 | 0.879 | 1.016 | 1.26e-01 | 1.00e+00 | 4859 | 3734 | 1125 |
| European | Full Cohort | Switching vs Continuation | Sleep_weekly | Sleep_weekly | 1.061 | 0.992 | 1.134 | 8.41e-02 | 1.00e+00 | 4859 | 3734 | 1125 |
| European | Full Cohort | Discontinuation vs Continuation | Sleep_weekly | (Intercept) | 0.305 | 0.281 | 0.330 | <2.2e-16 | <2.2e-16 | 4974 | 3734 | 1240 |
| European | Full Cohort | Discontinuation vs Continuation | Sleep_weekly | age_at_first_AD | 1.187 | 1.105 | 1.276 | 2.65e-06 | 4.51e-05 | 4974 | 3734 | 1240 |
| European | Full Cohort | Discontinuation vs Continuation | Sleep_weekly | sex_at_birthMale | 1.244 | 1.080 | 1.431 | 2.37e-03 | 4.03e-02 | 4974 | 3734 | 1240 |
| European | Full Cohort | Discontinuation vs Continuation | Sleep_weekly | Income_level | 1.026 | 0.958 | 1.098 | 4.66e-01 | 1.00e+00 | 4974 | 3734 | 1240 |

| | | | | | | | | | | | | |
|---|---|---|---|---|---|---|---|---|---|---|---|---|
| European | Full Cohort | Discontinuation vs Continuation | Sleep_weekly | Education_level | 0.861 | 0.802 | 0.924 | 3.41e-05 | 5.80e-04 | 4974 | 3734 | 1240 |
| European | Full Cohort | Discontinuation vs Continuation | Sleep_weekly | Sleep_weekly | 0.857 | 0.794 | 0.925 | 7.53e-05 | 1.28e-03 | 4974 | 3734 | 1240 |
| European | Recorded Depression | Switching vs Continuation | age_at_first_AD | (Intercept) | 0.377 | 0.338 | 0.422 | <2.2e-16 | <2.2e-16 | 2251 | 1596 | 655 |
| European | Recorded Depression | Switching vs Continuation | age_at_first_AD | age_at_first_AD | 0.872 | 0.779 | 0.976 | 1.75e-02 | 2.97e-01 | 2251 | 1596 | 655 |
| European | Recorded Depression | Switching vs Continuation | age_at_first_AD | sex_at_birthMale | 1.232 | 1.005 | 1.510 | 4.47e-02 | 7.60e-01 | 2251 | 1596 | 655 |
| European | Recorded Depression | Discontinuation vs Continuation | age_at_first_AD | (Intercept) | 0.214 | 0.187 | 0.246 | <2.2e-16 | <2.2e-16 | 1958 | 1596 | 362 |
| European | Recorded Depression | Discontinuation vs Continuation | age_at_first_AD | age_at_first_AD | 1.191 | 1.038 | 1.366 | 1.26e-02 | 2.15e-01 | 1958 | 1596 | 362 |
| European | Recorded Depression | Discontinuation vs Continuation | age_at_first_AD | sex_at_birthMale | 1.298 | 1.010 | 1.667 | 4.12e-02 | 7.00e-01 | 1958 | 1596 | 362 |
| European | Recorded Depression | Switching vs Continuation | sex_at_birth | (Intercept) | 0.377 | 0.338 | 0.422 | <2.2e-16 | <2.2e-16 | 2251 | 1596 | 655 |
| European | Recorded Depression | Switching vs Continuation | sex_at_birth | age_at_first_AD | 0.872 | 0.779 | 0.976 | 1.75e-02 | 2.97e-01 | 2251 | 1596 | 655 |
| European | Recorded Depression | Switching vs Continuation | sex_at_birth | sex_at_birthMale | 1.232 | 1.005 | 1.510 | 4.47e-02 | 7.60e-01 | 2251 | 1596 | 655 |
| European | Recorded Depression | Discontinuation vs Continuation | sex_at_birth | (Intercept) | 0.214 | 0.187 | 0.246 | <2.2e-16 | <2.2e-16 | 1958 | 1596 | 362 |
| European | Recorded Depression | Discontinuation vs Continuation | sex_at_birth | age_at_first_AD | 1.191 | 1.038 | 1.366 | 1.26e-02 | 2.15e-01 | 1958 | 1596 | 362 |
| European | Recorded Depression | Discontinuation vs Continuation | sex_at_birth | sex_at_birthMale | 1.298 | 1.010 | 1.667 | 4.12e-02 | 7.00e-01 | 1958 | 1596 | 362 |
| European | Recorded Depression | Switching vs Continuation | Education_level | (Intercept) | 0.383 | 0.342 | 0.428 | <2.2e-16 | <2.2e-16 | 2251 | 1596 | 655 |
| European | Recorded Depression | Switching vs Continuation | Education_level | age_at_first_AD | 0.871 | 0.776 | 0.977 | 1.81e-02 | 3.08e-01 | 2251 | 1596 | 655 |
| European | Recorded Depression | Switching vs Continuation | Education_level | sex_at_birthMale | 1.269 | 1.033 | 1.558 | 2.34e-02 | 3.98e-01 | 2251 | 1596 | 655 |
| European | Recorded Depression | Switching vs Continuation | Education_level | Income_level | 0.907 | 0.820 | 1.003 | 5.72e-02 | 9.73e-01 | 2251 | 1596 | 655 |
| European | Recorded Depression | Switching vs Continuation | Education_level | Education_level | 0.884 | 0.801 | 0.976 | 1.43e-02 | 2.42e-01 | 2251 | 1596 | 655 |
| European | Recorded Depression | Discontinuation vs Continuation | Education_level | (Intercept) | 0.220 | 0.191 | 0.252 | <2.2e-16 | <2.2e-16 | 1958 | 1596 | 362 |
| European | Recorded Depression | Discontinuation vs Continuation | Education_level | age_at_first_AD | 1.169 | 1.016 | 1.345 | 2.88e-02 | 4.90e-01 | 1958 | 1596 | 362 |
| European | Recorded Depression | Discontinuation vs Continuation | Education_level | sex_at_birthMale | 1.307 | 1.016 | 1.681 | 3.75e-02 | 6.37e-01 | 1958 | 1596 | 362 |
| European | Recorded Depression | Discontinuation vs Continuation | Education_level | Income_level | 0.987 | 0.872 | 1.116 | 8.31e-01 | 1.00e+00 | 1958 | 1596 | 362 |
| European | Recorded Depression | Discontinuation vs Continuation | Education_level | Education_level | 0.836 | 0.739 | 0.945 | 4.27e-03 | 7.26e-02 | 1958 | 1596 | 362 |
| European | Recorded Depression | Switching vs Continuation | BMI | (Intercept) | 0.382 | 0.341 | 0.427 | <2.2e-16 | <2.2e-16 | 2249 | 1595 | 654 |
| European | Recorded Depression | Switching vs Continuation | BMI | age_at_first_AD | 0.870 | 0.775 | 0.976 | 1.75e-02 | 2.98e-01 | 2249 | 1595 | 654 |
| European | Recorded Depression | Switching vs Continuation | BMI | sex_at_birthMale | 1.272 | 1.035 | 1.563 | 2.20e-02 | 3.74e-01 | 2249 | 1595 | 654 |
| European | Recorded Depression | Switching vs Continuation | BMI | Income_level | 0.906 | 0.819 | 1.002 | 5.45e-02 | 9.27e-01 | 2249 | 1595 | 654 |
| European | Recorded Depression | Switching vs Continuation | BMI | Education_level | 0.889 | 0.805 | 0.982 | 2.02e-02 | 3.44e-01 | 2249 | 1595 | 654 |
| European | Recorded Depression | Switching vs Continuation | BMI | BMI | 1.020 | 0.930 | 1.120 | 6.72e-01 | 1.00e+00 | 2249 | 1595 | 654 |
| European | Recorded Depression | Discontinuation vs Continuation | BMI | (Intercept) | 0.218 | 0.190 | 0.251 | <2.2e-16 | <2.2e-16 | 1957 | 1595 | 362 |
| European | Recorded Depression | Discontinuation vs Continuation | BMI | age_at_first_AD | 1.176 | 1.023 | 1.353 | 2.29e-02 | 3.89e-01 | 1957 | 1595 | 362 |
| European | Recorded Depression | Discontinuation vs Continuation | BMI | sex_at_birthMale | 1.321 | 1.027 | 1.701 | 3.05e-02 | 5.18e-01 | 1957 | 1595 | 362 |
| European | Recorded Depression | Discontinuation vs Continuation | BMI | Income_level | 0.983 | 0.869 | 1.112 | 7.82e-01 | 1.00e+00 | 1957 | 1595 | 362 |
| European | Recorded Depression | Discontinuation vs Continuation | BMI | Education_level | 0.828 | 0.731 | 0.937 | 2.83e-03 | 4.82e-02 | 1957 | 1595 | 362 |
| European | Recorded Depression | Discontinuation vs Continuation | BMI | BMI | 0.920 | 0.812 | 1.042 | 1.89e-01 | 1.00e+00 | 1957 | 1595 | 362 |
| European | Recorded Depression | Switching vs Continuation | Income_level | (Intercept) | 0.383 | 0.342 | 0.428 | <2.2e-16 | <2.2e-16 | 2251 | 1596 | 655 |
| European | Recorded Depression | Switching vs Continuation | Income_level | age_at_first_AD | 0.871 | 0.776 | 0.977 | 1.81e-02 | 3.08e-01 | 2251 | 1596 | 655 |
| European | Recorded Depression | Switching vs Continuation | Income_level | sex_at_birthMale | 1.269 | 1.033 | 1.558 | 2.34e-02 | 3.98e-01 | 2251 | 1596 | 655 |
| European | Recorded Depression | Switching vs Continuation | Income_level | Income_level | 0.907 | 0.820 | 1.003 | 5.72e-02 | 9.73e-01 | 2251 | 1596 | 655 |
| European | Recorded Depression | Switching vs Continuation | Income_level | Education_level | 0.884 | 0.801 | 0.976 | 1.43e-02 | 2.42e-01 | 2251 | 1596 | 655 |
| European | Recorded Depression | Discontinuation vs Continuation | Income_level | (Intercept) | 0.220 | 0.191 | 0.252 | <2.2e-16 | <2.2e-16 | 1958 | 1596 | 362 |
| European | Recorded Depression | Discontinuation vs Continuation | Income_level | age_at_first_AD | 1.169 | 1.016 | 1.345 | 2.88e-02 | 4.90e-01 | 1958 | 1596 | 362 |
| European | Recorded Depression | Discontinuation vs Continuation | Income_level | sex_at_birthMale | 1.307 | 1.016 | 1.681 | 3.75e-02 | 6.37e-01 | 1958 | 1596 | 362 |
| European | Recorded Depression | Discontinuation vs Continuation | Income_level | Income_level | 0.987 | 0.872 | 1.116 | 8.31e-01 | 1.00e+00 | 1958 | 1596 | 362 |
| European | Recorded Depression | Discontinuation vs Continuation | Income_level | Education_level | 0.836 | 0.739 | 0.945 | 4.27e-03 | 7.26e-02 | 1958 | 1596 | 362 |
| European | Recorded Depression | Switching vs Continuation | Dep_sum_score | (Intercept) | 0.273 | 0.238 | 0.313 | <2.2e-16 | <2.2e-16 | 2251 | 1596 | 655 |
| European | Recorded Depression | Switching vs Continuation | Dep_sum_score | age_at_first_AD | 0.902 | 0.802 | 1.015 | 8.80e-02 | 1.00e+00 | 2251 | 1596 | 655 |
| European | Recorded Depression | Switching vs Continuation | Dep_sum_score | sex_at_birthMale | 1.236 | 1.001 | 1.526 | 4.91e-02 | 8.35e-01 | 2251 | 1596 | 655 |
| European | Recorded Depression | Switching vs Continuation | Dep_sum_score | Income_level | 0.954 | 0.860 | 1.059 | 3.78e-01 | 1.00e+00 | 2251 | 1596 | 655 |
| European | Recorded Depression | Switching vs Continuation | Dep_sum_score | Education_level | 0.950 | 0.858 | 1.051 | 3.21e-01 | 1.00e+00 | 2251 | 1596 | 655 |

| | | | | | | | | | | | | |
|---|---|---|---|---|---|---|---|---|---|---|---|---|
| European | Recorded Depression | Switching vs Continuation | Dep_sum_score | Dep_sum_score | 1.507 | 1.392 | 1.632 | <2.2e-16 | <2.2e-16 | 2251 | 1596 | 655 |
| European | Recorded Depression | Discontinuation vs Continuation | Dep_sum_score | (Intercept) | 0.214 | 0.184 | 0.249 | <2.2e-16 | <2.2e-16 | 1958 | 1596 | 362 |
| European | Recorded Depression | Discontinuation vs Continuation | Dep_sum_score | age_at_first_AD | 1.173 | 1.020 | 1.350 | 2.55e-02 | 4.34e-01 | 1958 | 1596 | 362 |
| European | Recorded Depression | Discontinuation vs Continuation | Dep_sum_score | sex_at_birthMale | 1.298 | 1.009 | 1.671 | 4.27e-02 | 7.27e-01 | 1958 | 1596 | 362 |
| European | Recorded Depression | Discontinuation vs Continuation | Dep_sum_score | Income_level | 0.994 | 0.878 | 1.125 | 9.21e-01 | 1.00e+00 | 1958 | 1596 | 362 |
| European | Recorded Depression | Discontinuation vs Continuation | Dep_sum_score | Education_level | 0.841 | 0.743 | 0.952 | 6.13e-03 | 1.04e-01 | 1958 | 1596 | 362 |
| European | Recorded Depression | Discontinuation vs Continuation | Dep_sum_score | Dep_sum_score | 1.044 | 0.940 | 1.160 | 4.21e-01 | 1.00e+00 | 1958 | 1596 | 362 |
| European | Recorded Depression | Switching vs Continuation | Lifetime_anxiety | (Intercept) | 0.311 | 0.255 | 0.378 | <2.2e-16 | <2.2e-16 | 1137 | 825 | 312 |
| European | Recorded Depression | Switching vs Continuation | Lifetime_anxiety | age_at_first_AD | 0.920 | 0.779 | 1.087 | 3.27e-01 | 1.00e+00 | 1137 | 825 | 312 |
| European | Recorded Depression | Switching vs Continuation | Lifetime_anxiety | sex_at_birthMale | 1.164 | 0.866 | 1.564 | 3.14e-01 | 1.00e+00 | 1137 | 825 | 312 |
| European | Recorded Depression | Switching vs Continuation | Lifetime_anxiety | Income_level | 0.920 | 0.798 | 1.059 | 2.46e-01 | 1.00e+00 | 1137 | 825 | 312 |
| European | Recorded Depression | Switching vs Continuation | Lifetime_anxiety | Education_level | 0.900 | 0.783 | 1.034 | 1.38e-01 | 1.00e+00 | 1137 | 825 | 312 |
| European | Recorded Depression | Switching vs Continuation | Lifetime_anxiety | Lifetime_anxiety1 | 1.528 | 1.174 | 1.990 | 1.64e-03 | 2.79e-02 | 1137 | 825 | 312 |
| European | Recorded Depression | Discontinuation vs Continuation | Lifetime_anxiety | (Intercept) | 0.216 | 0.171 | 0.274 | <2.2e-16 | <2.2e-16 | 987 | 825 | 162 |
| European | Recorded Depression | Discontinuation vs Continuation | Lifetime_anxiety | age_at_first_AD | 1.261 | 1.021 | 1.559 | 3.16e-02 | 5.37e-01 | 987 | 825 | 162 |
| European | Recorded Depression | Discontinuation vs Continuation | Lifetime_anxiety | sex_at_birthMale | 1.193 | 0.819 | 1.736 | 3.58e-01 | 1.00e+00 | 987 | 825 | 162 |
| European | Recorded Depression | Discontinuation vs Continuation | Lifetime_anxiety | Income_level | 0.883 | 0.735 | 1.060 | 1.82e-01 | 1.00e+00 | 987 | 825 | 162 |
| European | Recorded Depression | Discontinuation vs Continuation | Lifetime_anxiety | Education_level | 0.880 | 0.736 | 1.052 | 1.60e-01 | 1.00e+00 | 987 | 825 | 162 |
| European | Recorded Depression | Discontinuation vs Continuation | Lifetime_anxiety | Lifetime_anxiety1 | 0.722 | 0.500 | 1.041 | 8.08e-02 | 1.00e+00 | 987 | 825 | 162 |
| European | Recorded Depression | Switching vs Continuation | Fibromyalgia | (Intercept) | 0.346 | 0.296 | 0.403 | <2.2e-16 | <2.2e-16 | 1437 | 1029 | 408 |
| European | Recorded Depression | Switching vs Continuation | Fibromyalgia | age_at_first_AD | 0.911 | 0.784 | 1.057 | 2.18e-01 | 1.00e+00 | 1437 | 1029 | 408 |
| European | Recorded Depression | Switching vs Continuation | Fibromyalgia | sex_at_birthMale | 1.232 | 0.948 | 1.602 | 1.19e-01 | 1.00e+00 | 1437 | 1029 | 408 |
| European | Recorded Depression | Switching vs Continuation | Fibromyalgia | Income_level | 0.917 | 0.807 | 1.040 | 1.78e-01 | 1.00e+00 | 1437 | 1029 | 408 |
| European | Recorded Depression | Switching vs Continuation | Fibromyalgia | Education_level | 0.929 | 0.820 | 1.052 | 2.46e-01 | 1.00e+00 | 1437 | 1029 | 408 |
| European | Recorded Depression | Switching vs Continuation | Fibromyalgia | Fibromyalgia1 | 1.571 | 1.172 | 2.105 | 2.50e-03 | 4.26e-02 | 1437 | 1029 | 408 |
| European | Recorded Depression | Discontinuation vs Continuation | Fibromyalgia | (Intercept) | 0.216 | 0.179 | 0.260 | <2.2e-16 | <2.2e-16 | 1254 | 1029 | 225 |
| European | Recorded Depression | Discontinuation vs Continuation | Fibromyalgia | age_at_first_AD | 1.082 | 0.901 | 1.298 | 4.00e-01 | 1.00e+00 | 1254 | 1029 | 225 |
| European | Recorded Depression | Discontinuation vs Continuation | Fibromyalgia | sex_at_birthMale | 1.178 | 0.852 | 1.630 | 3.22e-01 | 1.00e+00 | 1254 | 1029 | 225 |
| European | Recorded Depression | Discontinuation vs Continuation | Fibromyalgia | Income_level | 1.001 | 0.859 | 1.167 | 9.88e-01 | 1.00e+00 | 1254 | 1029 | 225 |
| European | Recorded Depression | Discontinuation vs Continuation | Fibromyalgia | Education_level | 0.881 | 0.755 | 1.027 | 1.05e-01 | 1.00e+00 | 1254 | 1029 | 225 |
| European | Recorded Depression | Discontinuation vs Continuation | Fibromyalgia | Fibromyalgia1 | 0.995 | 0.665 | 1.490 | 9.80e-01 | 1.00e+00 | 1254 | 1029 | 225 |
| European | Recorded Depression | Switching vs Continuation | Childhood_sexual_abuse | (Intercept) | 0.361 | 0.311 | 0.420 | <2.2e-16 | <2.2e-16 | 1284 | 917 | 367 |
| European | Recorded Depression | Switching vs Continuation | Childhood_sexual_abuse | age_at_first_AD | 0.919 | 0.789 | 1.069 | 2.73e-01 | 1.00e+00 | 1284 | 917 | 367 |
| European | Recorded Depression | Switching vs Continuation | Childhood_sexual_abuse | sex_at_birthMale | 1.326 | 1.007 | 1.745 | 4.42e-02 | 7.52e-01 | 1284 | 917 | 367 |
| European | Recorded Depression | Switching vs Continuation | Childhood_sexual_abuse | Income_level | 0.950 | 0.832 | 1.086 | 4.54e-01 | 1.00e+00 | 1284 | 917 | 367 |
| European | Recorded Depression | Switching vs Continuation | Childhood_sexual_abuse | Education_level | 0.905 | 0.796 | 1.030 | 1.31e-01 | 1.00e+00 | 1284 | 917 | 367 |
| European | Recorded Depression | Switching vs Continuation | Childhood_sexual_abuse | Childhood_sexual_a | 1.203 | 1.097 | 1.320 | 8.91e-05 | 1.51e-03 | 1284 | 917 | 367 |
| European | Recorded Depression | Discontinuation vs Continuation | Childhood_sexual_abuse | (Intercept) | 0.221 | 0.184 | 0.265 | <2.2e-16 | <2.2e-16 | 1126 | 917 | 209 |
| European | Recorded Depression | Discontinuation vs Continuation | Childhood_sexual_abuse | age_at_first_AD | 1.309 | 1.087 | 1.576 | 4.55e-03 | 7.74e-02 | 1126 | 917 | 209 |
| European | Recorded Depression | Discontinuation vs Continuation | Childhood_sexual_abuse | sex_at_birthMale | 1.200 | 0.855 | 1.683 | 2.92e-01 | 1.00e+00 | 1126 | 917 | 209 |
| European | Recorded Depression | Discontinuation vs Continuation | Childhood_sexual_abuse | Income_level | 0.939 | 0.801 | 1.102 | 4.44e-01 | 1.00e+00 | 1126 | 917 | 209 |
| European | Recorded Depression | Discontinuation vs Continuation | Childhood_sexual_abuse | Education_level | 0.849 | 0.724 | 0.997 | 4.58e-02 | 7.79e-01 | 1126 | 917 | 209 |
| European | Recorded Depression | Discontinuation vs Continuation | Childhood_sexual_abuse | Childhood_sexual_a | 1.040 | 0.911 | 1.187 | 5.64e-01 | 1.00e+00 | 1126 | 917 | 209 |
| European | Recorded Depression | Switching vs Continuation | Neuroticism | (Intercept) | 0.352 | 0.311 | 0.398 | <2.2e-16 | <2.2e-16 | 2158 | 1529 | 629 |
| European | Recorded Depression | Switching vs Continuation | Neuroticism | age_at_first_AD | 0.878 | 0.776 | 0.994 | 3.97e-02 | 6.74e-01 | 2158 | 1529 | 629 |
| European | Recorded Depression | Switching vs Continuation | Neuroticism | sex_at_birthMale | 1.277 | 1.032 | 1.580 | 2.45e-02 | 4.17e-01 | 2158 | 1529 | 629 |
| European | Recorded Depression | Switching vs Continuation | Neuroticism | Income_level | 0.942 | 0.850 | 1.045 | 2.62e-01 | 1.00e+00 | 2158 | 1529 | 629 |
| European | Recorded Depression | Switching vs Continuation | Neuroticism | Education_level | 0.910 | 0.822 | 1.007 | 6.70e-02 | 1.00e+00 | 2158 | 1529 | 629 |
| European | Recorded Depression | Switching vs Continuation | Neuroticism | Neuroticism | 1.265 | 1.150 | 1.391 | 1.43e-06 | 2.42e-05 | 2158 | 1529 | 629 |
| European | Recorded Depression | Discontinuation vs Continuation | Neuroticism | (Intercept) | 0.229 | 0.198 | 0.264 | <2.2e-16 | <2.2e-16 | 1879 | 1529 | 350 |
| European | Recorded Depression | Discontinuation vs Continuation | Neuroticism | age_at_first_AD | 1.157 | 0.994 | 1.346 | 5.91e-02 | 1.00e+00 | 1879 | 1529 | 350 |

| | | | | | | | | | | | | |
|---|---|---|---|---|---|---|---|---|---|---|---|---|
| European | Recorded Depression | Discontinuation vs Continuation | Neuroticism | sex_at_birthMale | 1.285 | 0.993 | 1.662 | 5.64e-02 | 9.58e-01 | 1879 | 1529 | 350 |
| European | Recorded Depression | Discontinuation vs Continuation | Neuroticism | Income_level | 0.992 | 0.875 | 1.126 | 9.04e-01 | 1.00e+00 | 1879 | 1529 | 350 |
| European | Recorded Depression | Discontinuation vs Continuation | Neuroticism | Education_level | 0.812 | 0.714 | 0.923 | 1.39e-03 | 2.37e-02 | 1879 | 1529 | 350 |
| European | Recorded Depression | Discontinuation vs Continuation | Neuroticism | Neuroticism | 0.895 | 0.792 | 1.010 | 7.26e-02 | 1.00e+00 | 1879 | 1529 | 350 |
| European | Recorded Depression | Switching vs Continuation | Longterm_difficulties | (Intercept) | 0.356 | 0.315 | 0.401 | <2.2e-16 | <2.2e-16 | 2180 | 1544 | 636 |
| European | Recorded Depression | Switching vs Continuation | Longterm_difficulties | age_at_first_AD | 0.917 | 0.813 | 1.035 | 1.63e-01 | 1.00e+00 | 2180 | 1544 | 636 |
| European | Recorded Depression | Switching vs Continuation | Longterm_difficulties | sex_at_birthMale | 1.282 | 1.038 | 1.582 | 2.08e-02 | 3.54e-01 | 2180 | 1544 | 636 |
| European | Recorded Depression | Switching vs Continuation | Longterm_difficulties | Income_level | 0.955 | 0.860 | 1.060 | 3.88e-01 | 1.00e+00 | 2180 | 1544 | 636 |
| European | Recorded Depression | Switching vs Continuation | Longterm_difficulties | Education_level | 0.889 | 0.805 | 0.983 | 2.17e-02 | 3.69e-01 | 2180 | 1544 | 636 |
| European | Recorded Depression | Switching vs Continuation | Longterm_difficulties | Longterm_difficultie | 1.246 | 1.142 | 1.360 | 7.70e-07 | 1.31e-05 | 2180 | 1544 | 636 |
| European | Recorded Depression | Discontinuation vs Continuation | Longterm_difficulties | (Intercept) | 0.219 | 0.189 | 0.252 | <2.2e-16 | <2.2e-16 | 1896 | 1544 | 352 |
| European | Recorded Depression | Discontinuation vs Continuation | Longterm_difficulties | age_at_first_AD | 1.199 | 1.035 | 1.388 | 1.53e-02 | 2.60e-01 | 1896 | 1544 | 352 |
| European | Recorded Depression | Discontinuation vs Continuation | Longterm_difficulties | sex_at_birthMale | 1.270 | 0.982 | 1.642 | 6.80e-02 | 1.00e+00 | 1896 | 1544 | 352 |
| European | Recorded Depression | Discontinuation vs Continuation | Longterm_difficulties | Income_level | 1.019 | 0.898 | 1.157 | 7.70e-01 | 1.00e+00 | 1896 | 1544 | 352 |
| European | Recorded Depression | Discontinuation vs Continuation | Longterm_difficulties | Education_level | 0.838 | 0.740 | 0.950 | 5.71e-03 | 9.70e-02 | 1896 | 1544 | 352 |
| European | Recorded Depression | Discontinuation vs Continuation | Longterm_difficulties | Longterm_difficultie | 1.073 | 0.957 | 1.202 | 2.30e-01 | 1.00e+00 | 1896 | 1544 | 352 |
| European | Recorded Depression | Switching vs Continuation | Threatening_experiences | (Intercept) | 0.379 | 0.338 | 0.425 | <2.2e-16 | <2.2e-16 | 2201 | 1561 | 640 |
| European | Recorded Depression | Switching vs Continuation | Threatening_experiences | age_at_first_AD | 0.875 | 0.779 | 0.984 | 2.52e-02 | 4.28e-01 | 2201 | 1561 | 640 |
| European | Recorded Depression | Switching vs Continuation | Threatening_experiences | sex_at_birthMale | 1.260 | 1.023 | 1.553 | 2.98e-02 | 5.06e-01 | 2201 | 1561 | 640 |
| European | Recorded Depression | Switching vs Continuation | Threatening_experiences | Income_level | 0.936 | 0.845 | 1.038 | 2.09e-01 | 1.00e+00 | 2201 | 1561 | 640 |
| European | Recorded Depression | Switching vs Continuation | Threatening_experiences | Education_level | 0.906 | 0.820 | 1.001 | 5.34e-02 | 9.07e-01 | 2201 | 1561 | 640 |
| European | Recorded Depression | Switching vs Continuation | Threatening_experiences | Threatening_experie | 1.150 | 1.051 | 1.257 | 2.31e-03 | 3.94e-02 | 2201 | 1561 | 640 |
| European | Recorded Depression | Discontinuation vs Continuation | Threatening_experiences | (Intercept) | 0.217 | 0.189 | 0.250 | <2.2e-16 | <2.2e-16 | 1914 | 1561 | 353 |
| European | Recorded Depression | Discontinuation vs Continuation | Threatening_experiences | age_at_first_AD | 1.172 | 1.017 | 1.351 | 2.84e-02 | 4.82e-01 | 1914 | 1561 | 353 |
| European | Recorded Depression | Discontinuation vs Continuation | Threatening_experiences | sex_at_birthMale | 1.326 | 1.029 | 1.710 | 2.95e-02 | 5.01e-01 | 1914 | 1561 | 353 |
| European | Recorded Depression | Discontinuation vs Continuation | Threatening_experiences | Income_level | 0.997 | 0.879 | 1.129 | 9.57e-01 | 1.00e+00 | 1914 | 1561 | 353 |
| European | Recorded Depression | Discontinuation vs Continuation | Threatening_experiences | Education_level | 0.850 | 0.751 | 0.963 | 1.08e-02 | 1.83e-01 | 1914 | 1561 | 353 |
| European | Recorded Depression | Discontinuation vs Continuation | Threatening_experiences | Threatening_experie | 1.058 | 0.940 | 1.191 | 3.50e-01 | 1.00e+00 | 1914 | 1561 | 353 |
| European | Recorded Depression | Switching vs Continuation | Smoking_Packyears | (Intercept) | 0.409 | 0.353 | 0.473 | <2.2e-16 | <2.2e-16 | 1296 | 894 | 402 |
| European | Recorded Depression | Switching vs Continuation | Smoking_Packyears | age_at_first_AD | 0.753 | 0.646 | 0.877 | 2.74e-04 | 4.65e-03 | 1296 | 894 | 402 |
| European | Recorded Depression | Switching vs Continuation | Smoking_Packyears | sex_at_birthMale | 1.225 | 0.941 | 1.595 | 1.31e-01 | 1.00e+00 | 1296 | 894 | 402 |
| European | Recorded Depression | Switching vs Continuation | Smoking_Packyears | Income_level | 0.966 | 0.849 | 1.100 | 6.06e-01 | 1.00e+00 | 1296 | 894 | 402 |
| European | Recorded Depression | Switching vs Continuation | Smoking_Packyears | Education_level | 0.888 | 0.779 | 1.013 | 7.82e-02 | 1.00e+00 | 1296 | 894 | 402 |
| European | Recorded Depression | Switching vs Continuation | Smoking_Packyears | Smoking_Packyears | 1.225 | 1.084 | 1.385 | 1.15e-03 | 1.96e-02 | 1296 | 894 | 402 |
| European | Recorded Depression | Discontinuation vs Continuation | Smoking_Packyears | (Intercept) | 0.235 | 0.196 | 0.281 | <2.2e-16 | <2.2e-16 | 1121 | 894 | 227 |
| European | Recorded Depression | Discontinuation vs Continuation | Smoking_Packyears | age_at_first_AD | 1.112 | 0.928 | 1.334 | 2.51e-01 | 1.00e+00 | 1121 | 894 | 227 |
| European | Recorded Depression | Discontinuation vs Continuation | Smoking_Packyears | sex_at_birthMale | 1.348 | 0.980 | 1.854 | 6.62e-02 | 1.00e+00 | 1121 | 894 | 227 |
| European | Recorded Depression | Discontinuation vs Continuation | Smoking_Packyears | Income_level | 1.017 | 0.870 | 1.188 | 8.36e-01 | 1.00e+00 | 1121 | 894 | 227 |
| European | Recorded Depression | Discontinuation vs Continuation | Smoking_Packyears | Education_level | 0.834 | 0.711 | 0.978 | 2.53e-02 | 4.31e-01 | 1121 | 894 | 227 |
| European | Recorded Depression | Discontinuation vs Continuation | Smoking_Packyears | Smoking_Packyears | 1.086 | 0.926 | 1.274 | 3.09e-01 | 1.00e+00 | 1121 | 894 | 227 |
| European | Recorded Depression | Switching vs Continuation | Alcohol_intake | (Intercept) | 0.363 | 0.313 | 0.422 | <2.2e-16 | <2.2e-16 | 1431 | 1041 | 390 |
| European | Recorded Depression | Switching vs Continuation | Alcohol_intake | age_at_first_AD | 0.837 | 0.724 | 0.968 | 1.66e-02 | 2.83e-01 | 1431 | 1041 | 390 |
| European | Recorded Depression | Switching vs Continuation | Alcohol_intake | sex_at_birthMale | 1.101 | 0.842 | 1.441 | 4.81e-01 | 1.00e+00 | 1431 | 1041 | 390 |
| European | Recorded Depression | Switching vs Continuation | Alcohol_intake | Income_level | 1.004 | 0.886 | 1.138 | 9.50e-01 | 1.00e+00 | 1431 | 1041 | 390 |
| European | Recorded Depression | Switching vs Continuation | Alcohol_intake | Education_level | 0.860 | 0.759 | 0.975 | 1.80e-02 | 3.07e-01 | 1431 | 1041 | 390 |
| European | Recorded Depression | Switching vs Continuation | Alcohol_intake | Alcohol_intake | 1.067 | 0.952 | 1.195 | 2.68e-01 | 1.00e+00 | 1431 | 1041 | 390 |
| European | Recorded Depression | Discontinuation vs Continuation | Alcohol_intake | (Intercept) | 0.224 | 0.187 | 0.268 | <2.2e-16 | <2.2e-16 | 1256 | 1041 | 215 |
| European | Recorded Depression | Discontinuation vs Continuation | Alcohol_intake | age_at_first_AD | 1.269 | 1.062 | 1.516 | 8.76e-03 | 1.49e-01 | 1256 | 1041 | 215 |
| European | Recorded Depression | Discontinuation vs Continuation | Alcohol_intake | sex_at_birthMale | 0.912 | 0.648 | 1.285 | 5.98e-01 | 1.00e+00 | 1256 | 1041 | 215 |
| European | Recorded Depression | Discontinuation vs Continuation | Alcohol_intake | Income_level | 1.044 | 0.892 | 1.221 | 5.92e-01 | 1.00e+00 | 1256 | 1041 | 215 |
| European | Recorded Depression | Discontinuation vs Continuation | Alcohol_intake | Education_level | 0.861 | 0.737 | 1.005 | 5.85e-02 | 9.95e-01 | 1256 | 1041 | 215 |

| | | | | | | | | | | | | |
|---|---|---|---|---|---|---|---|---|---|---|---|---|
| European | Recorded Depression | Discontinuation vs Continuation | Alcohol_intake | Alcohol_intake | 1.090 | 0.940 | 1.265 | 2.56e-01 | 1.00e+00 | 1256 | 1041 | 215 |
| European | Recorded Depression | Switching vs Continuation | Chronotype | (Intercept) | 0.364 | 0.320 | 0.414 | <2.2e-16 | <2.2e-16 | 1757 | 1263 | 494 |
| European | Recorded Depression | Switching vs Continuation | Chronotype | age_at_first_AD | 0.912 | 0.798 | 1.043 | 1.78e-01 | 1.00e+00 | 1757 | 1263 | 494 |
| European | Recorded Depression | Switching vs Continuation | Chronotype | sex_at_birthMale | 1.267 | 1.002 | 1.601 | 4.80e-02 | 8.17e-01 | 1757 | 1263 | 494 |
| European | Recorded Depression | Switching vs Continuation | Chronotype | Income_level | 0.847 | 0.755 | 0.952 | 5.16e-03 | 8.77e-02 | 1757 | 1263 | 494 |
| European | Recorded Depression | Switching vs Continuation | Chronotype | Education_level | 0.913 | 0.816 | 1.021 | 1.11e-01 | 1.00e+00 | 1757 | 1263 | 494 |
| European | Recorded Depression | Switching vs Continuation | Chronotype | Chronotype | 1.106 | 1.005 | 1.217 | 3.99e-02 | 6.79e-01 | 1757 | 1263 | 494 |
| European | Recorded Depression | Discontinuation vs Continuation | Chronotype | (Intercept) | 0.230 | 0.197 | 0.268 | <2.2e-16 | <2.2e-16 | 1556 | 1263 | 293 |
| European | Recorded Depression | Discontinuation vs Continuation | Chronotype | age_at_first_AD | 1.212 | 1.035 | 1.419 | 1.71e-02 | 2.91e-01 | 1556 | 1263 | 293 |
| European | Recorded Depression | Discontinuation vs Continuation | Chronotype | sex_at_birthMale | 1.172 | 0.883 | 1.555 | 2.72e-01 | 1.00e+00 | 1556 | 1263 | 293 |
| European | Recorded Depression | Discontinuation vs Continuation | Chronotype | Income_level | 0.968 | 0.844 | 1.110 | 6.40e-01 | 1.00e+00 | 1556 | 1263 | 293 |
| European | Recorded Depression | Discontinuation vs Continuation | Chronotype | Education_level | 0.892 | 0.779 | 1.022 | 1.00e-01 | 1.00e+00 | 1556 | 1263 | 293 |
| European | Recorded Depression | Discontinuation vs Continuation | Chronotype | Chronotype | 0.970 | 0.857 | 1.098 | 6.33e-01 | 1.00e+00 | 1556 | 1263 | 293 |
| European | Recorded Depression | Switching vs Continuation | Sleep_weekly | (Intercept) | 0.377 | 0.335 | 0.425 | <2.2e-16 | <2.2e-16 | 2037 | 1453 | 584 |
| European | Recorded Depression | Switching vs Continuation | Sleep_weekly | age_at_first_AD | 0.882 | 0.780 | 0.998 | 4.66e-02 | 7.92e-01 | 2037 | 1453 | 584 |
| European | Recorded Depression | Switching vs Continuation | Sleep_weekly | sex_at_birthMale | 1.268 | 1.018 | 1.579 | 3.40e-02 | 5.77e-01 | 2037 | 1453 | 584 |
| European | Recorded Depression | Switching vs Continuation | Sleep_weekly | Income_level | 0.873 | 0.784 | 0.971 | 1.25e-02 | 2.12e-01 | 2037 | 1453 | 584 |
| European | Recorded Depression | Switching vs Continuation | Sleep_weekly | Education_level | 0.900 | 0.811 | 0.998 | 4.52e-02 | 7.68e-01 | 2037 | 1453 | 584 |
| European | Recorded Depression | Switching vs Continuation | Sleep_weekly | Sleep_weekly | 0.999 | 0.909 | 1.097 | 9.82e-01 | 1.00e+00 | 2037 | 1453 | 584 |
| European | Recorded Depression | Discontinuation vs Continuation | Sleep_weekly | (Intercept) | 0.220 | 0.190 | 0.255 | <2.2e-16 | <2.2e-16 | 1779 | 1453 | 326 |
| European | Recorded Depression | Discontinuation vs Continuation | Sleep_weekly | age_at_first_AD | 1.130 | 0.972 | 1.314 | 1.12e-01 | 1.00e+00 | 1779 | 1453 | 326 |
| European | Recorded Depression | Discontinuation vs Continuation | Sleep_weekly | sex_at_birthMale | 1.123 | 0.855 | 1.474 | 4.05e-01 | 1.00e+00 | 1779 | 1453 | 326 |
| European | Recorded Depression | Discontinuation vs Continuation | Sleep_weekly | Income_level | 0.992 | 0.871 | 1.130 | 9.04e-01 | 1.00e+00 | 1779 | 1453 | 326 |
| European | Recorded Depression | Discontinuation vs Continuation | Sleep_weekly | Education_level | 0.861 | 0.757 | 0.981 | 2.40e-02 | 4.09e-01 | 1779 | 1453 | 326 |
| European | Recorded Depression | Discontinuation vs Continuation | Sleep_weekly | Sleep_weekly | 0.697 | 0.597 | 0.814 | 4.81e-06 | 8.17e-05 | 1779 | 1453 | 326 |
| European | No Recorded Depression | Switching vs Continuation | age_at_first_AD | (Intercept) | 0.216 | 0.194 | 0.241 | <2.2e-16 | <2.2e-16 | 3182 | 2572 | 610 |
| European | No Recorded Depression | Switching vs Continuation | age_at_first_AD | age_at_first_AD | 0.819 | 0.746 | 0.899 | 2.61e-05 | 4.43e-04 | 3182 | 2572 | 610 |
| European | No Recorded Depression | Switching vs Continuation | age_at_first_AD | sex_at_birthMale | 1.325 | 1.098 | 1.599 | 3.33e-03 | 5.66e-02 | 3182 | 2572 | 610 |
| European | No Recorded Depression | Discontinuation vs Continuation | age_at_first_AD | (Intercept) | 0.361 | 0.330 | 0.395 | <2.2e-16 | <2.2e-16 | 3619 | 2572 | 1047 |
| European | No Recorded Depression | Discontinuation vs Continuation | age_at_first_AD | age_at_first_AD | 1.239 | 1.153 | 1.332 | 5.11e-09 | 8.69e-08 | 3619 | 2572 | 1047 |
| European | No Recorded Depression | Discontinuation vs Continuation | age_at_first_AD | sex_at_birthMale | 1.271 | 1.090 | 1.482 | 2.19e-03 | 3.72e-02 | 3619 | 2572 | 1047 |
| European | No Recorded Depression | Switching vs Continuation | sex_at_birth | (Intercept) | 0.216 | 0.194 | 0.241 | <2.2e-16 | <2.2e-16 | 3182 | 2572 | 610 |
| European | No Recorded Depression | Switching vs Continuation | sex_at_birth | age_at_first_AD | 0.819 | 0.746 | 0.899 | 2.61e-05 | 4.43e-04 | 3182 | 2572 | 610 |
| European | No Recorded Depression | Switching vs Continuation | sex_at_birth | sex_at_birthMale | 1.325 | 1.098 | 1.599 | 3.33e-03 | 5.66e-02 | 3182 | 2572 | 610 |
| European | No Recorded Depression | Discontinuation vs Continuation | sex_at_birth | (Intercept) | 0.361 | 0.330 | 0.395 | <2.2e-16 | <2.2e-16 | 3619 | 2572 | 1047 |
| European | No Recorded Depression | Discontinuation vs Continuation | sex_at_birth | age_at_first_AD | 1.239 | 1.153 | 1.332 | 5.11e-09 | 8.69e-08 | 3619 | 2572 | 1047 |
| European | No Recorded Depression | Discontinuation vs Continuation | sex_at_birth | sex_at_birthMale | 1.271 | 1.090 | 1.482 | 2.19e-03 | 3.72e-02 | 3619 | 2572 | 1047 |
| European | No Recorded Depression | Switching vs Continuation | Education_level | (Intercept) | 0.216 | 0.194 | 0.241 | <2.2e-16 | <2.2e-16 | 3182 | 2572 | 610 |
| European | No Recorded Depression | Switching vs Continuation | Education_level | age_at_first_AD | 0.819 | 0.744 | 0.902 | 5.13e-05 | 8.71e-04 | 3182 | 2572 | 610 |
| European | No Recorded Depression | Switching vs Continuation | Education_level | sex_at_birthMale | 1.345 | 1.114 | 1.625 | 2.07e-03 | 3.52e-02 | 3182 | 2572 | 610 |
| European | No Recorded Depression | Switching vs Continuation | Education_level | Income_level | 0.926 | 0.842 | 1.018 | 1.13e-01 | 1.00e+00 | 3182 | 2572 | 610 |
| European | No Recorded Depression | Switching vs Continuation | Education_level | Education_level | 0.965 | 0.875 | 1.065 | 4.78e-01 | 1.00e+00 | 3182 | 2572 | 610 |
| European | No Recorded Depression | Discontinuation vs Continuation | Education_level | (Intercept) | 0.361 | 0.330 | 0.395 | <2.2e-16 | <2.2e-16 | 3619 | 2572 | 1047 |
| European | No Recorded Depression | Discontinuation vs Continuation | Education_level | age_at_first_AD | 1.200 | 1.113 | 1.293 | 1.81e-06 | 3.08e-05 | 3619 | 2572 | 1047 |
| European | No Recorded Depression | Discontinuation vs Continuation | Education_level | sex_at_birthMale | 1.271 | 1.090 | 1.483 | 2.26e-03 | 3.85e-02 | 3619 | 2572 | 1047 |
| European | No Recorded Depression | Discontinuation vs Continuation | Education_level | Income_level | 1.042 | 0.966 | 1.124 | 2.87e-01 | 1.00e+00 | 3619 | 2572 | 1047 |
| European | No Recorded Depression | Discontinuation vs Continuation | Education_level | Education_level | 0.875 | 0.807 | 0.948 | 1.13e-03 | 1.93e-02 | 3619 | 2572 | 1047 |
| European | No Recorded Depression | Switching vs Continuation | BMI | (Intercept) | 0.217 | 0.194 | 0.242 | <2.2e-16 | <2.2e-16 | 3178 | 2568 | 610 |
| European | No Recorded Depression | Switching vs Continuation | BMI | age_at_first_AD | 0.813 | 0.737 | 0.896 | 3.01e-05 | 5.11e-04 | 3178 | 2568 | 610 |
| European | No Recorded Depression | Switching vs Continuation | BMI | sex_at_birthMale | 1.340 | 1.110 | 1.619 | 2.38e-03 | 4.05e-02 | 3178 | 2568 | 610 |
| European | No Recorded Depression | Switching vs Continuation | BMI | Income_level | 0.933 | 0.848 | 1.026 | 1.52e-01 | 1.00e+00 | 3178 | 2568 | 610 |

| | | | | | | | | | | | | |
|---|---|---|---|---|---|---|---|---|---|---|---|---|
| European | No Recorded Depression | Switching vs Continuation | BMI | Education_level | 0.974 | 0.882 | 1.075 | 6.00e-01 | 1.00e+00 | 3178 | 2568 | 610 |
| European | No Recorded Depression | Switching vs Continuation | BMI | BMI | 1.071 | 0.976 | 1.176 | 1.49e-01 | 1.00e+00 | 3178 | 2568 | 610 |
| European | No Recorded Depression | Discontinuation vs Continuation | BMI | (Intercept) | 0.360 | 0.329 | 0.394 | <2.2e-16 | <2.2e-16 | 3615 | 2568 | 1047 |
| European | No Recorded Depression | Discontinuation vs Continuation | BMI | age_at_first_AD | 1.206 | 1.118 | 1.300 | 1.15e-06 | 1.95e-05 | 3615 | 2568 | 1047 |
| European | No Recorded Depression | Discontinuation vs Continuation | BMI | sex_at_birthMale | 1.279 | 1.096 | 1.492 | 1.81e-03 | 3.08e-02 | 3615 | 2568 | 1047 |
| European | No Recorded Depression | Discontinuation vs Continuation | BMI | Income_level | 1.041 | 0.965 | 1.123 | 3.02e-01 | 1.00e+00 | 3615 | 2568 | 1047 |
| European | No Recorded Depression | Discontinuation vs Continuation | BMI | Education_level | 0.870 | 0.802 | 0.944 | 8.26e-04 | 1.40e-02 | 3615 | 2568 | 1047 |
| European | No Recorded Depression | Discontinuation vs Continuation | BMI | BMI | 0.960 | 0.886 | 1.041 | 3.25e-01 | 1.00e+00 | 3615 | 2568 | 1047 |
| European | No Recorded Depression | Switching vs Continuation | Income_level | (Intercept) | 0.216 | 0.194 | 0.241 | <2.2e-16 | <2.2e-16 | 3182 | 2572 | 610 |
| European | No Recorded Depression | Switching vs Continuation | Income_level | age_at_first_AD | 0.819 | 0.744 | 0.902 | 5.13e-05 | 8.71e-04 | 3182 | 2572 | 610 |
| European | No Recorded Depression | Switching vs Continuation | Income_level | sex_at_birthMale | 1.345 | 1.114 | 1.625 | 2.07e-03 | 3.52e-02 | 3182 | 2572 | 610 |
| European | No Recorded Depression | Switching vs Continuation | Income_level | Income_level | 0.926 | 0.842 | 1.018 | 1.13e-01 | 1.00e+00 | 3182 | 2572 | 610 |
| European | No Recorded Depression | Switching vs Continuation | Income_level | Education_level | 0.965 | 0.875 | 1.065 | 4.78e-01 | 1.00e+00 | 3182 | 2572 | 610 |
| European | No Recorded Depression | Discontinuation vs Continuation | Income_level | (Intercept) | 0.361 | 0.330 | 0.395 | <2.2e-16 | <2.2e-16 | 3619 | 2572 | 1047 |
| European | No Recorded Depression | Discontinuation vs Continuation | Income_level | age_at_first_AD | 1.200 | 1.113 | 1.293 | 1.81e-06 | 3.08e-05 | 3619 | 2572 | 1047 |
| European | No Recorded Depression | Discontinuation vs Continuation | Income_level | sex_at_birthMale | 1.271 | 1.090 | 1.483 | 2.26e-03 | 3.85e-02 | 3619 | 2572 | 1047 |
| European | No Recorded Depression | Discontinuation vs Continuation | Income_level | Income_level | 1.042 | 0.966 | 1.124 | 2.87e-01 | 1.00e+00 | 3619 | 2572 | 1047 |
| European | No Recorded Depression | Discontinuation vs Continuation | Income_level | Education_level | 0.875 | 0.807 | 0.948 | 1.13e-03 | 1.93e-02 | 3619 | 2572 | 1047 |
| European | No Recorded Depression | Switching vs Continuation | Dep_sum_score | (Intercept) | 0.264 | 0.233 | 0.300 | <2.2e-16 | <2.2e-16 | 3182 | 2572 | 610 |
| European | No Recorded Depression | Switching vs Continuation | Dep_sum_score | age_at_first_AD | 0.836 | 0.758 | 0.921 | 3.07e-04 | 5.23e-03 | 3182 | 2572 | 610 |
| European | No Recorded Depression | Switching vs Continuation | Dep_sum_score | sex_at_birthMale | 1.361 | 1.126 | 1.647 | 1.46e-03 | 2.49e-02 | 3182 | 2572 | 610 |
| European | No Recorded Depression | Switching vs Continuation | Dep_sum_score | Income_level | 0.937 | 0.851 | 1.030 | 1.77e-01 | 1.00e+00 | 3182 | 2572 | 610 |
| European | No Recorded Depression | Switching vs Continuation | Dep_sum_score | Education_level | 0.989 | 0.896 | 1.091 | 8.23e-01 | 1.00e+00 | 3182 | 2572 | 610 |
| European | No Recorded Depression | Switching vs Continuation | Dep_sum_score | Dep_sum_score | 1.699 | 1.432 | 2.015 | 1.16e-09 | 1.97e-08 | 3182 | 2572 | 610 |
| European | No Recorded Depression | Discontinuation vs Continuation | Dep_sum_score | (Intercept) | 0.339 | 0.300 | 0.383 | <2.2e-16 | <2.2e-16 | 3619 | 2572 | 1047 |
| European | No Recorded Depression | Discontinuation vs Continuation | Dep_sum_score | age_at_first_AD | 1.195 | 1.109 | 1.288 | 3.18e-06 | 5.40e-05 | 3619 | 2572 | 1047 |
| European | No Recorded Depression | Discontinuation vs Continuation | Dep_sum_score | sex_at_birthMale | 1.267 | 1.086 | 1.478 | 2.66e-03 | 4.53e-02 | 3619 | 2572 | 1047 |
| European | No Recorded Depression | Discontinuation vs Continuation | Dep_sum_score | Income_level | 1.040 | 0.964 | 1.122 | 3.11e-01 | 1.00e+00 | 3619 | 2572 | 1047 |
| European | No Recorded Depression | Discontinuation vs Continuation | Dep_sum_score | Education_level | 0.871 | 0.803 | 0.945 | 8.30e-04 | 1.41e-02 | 3619 | 2572 | 1047 |
| European | No Recorded Depression | Discontinuation vs Continuation | Dep_sum_score | Dep_sum_score | 0.868 | 0.726 | 1.039 | 1.23e-01 | 1.00e+00 | 3619 | 2572 | 1047 |
| European | No Recorded Depression | Switching vs Continuation | Lifetime_anxiety | (Intercept) | 0.152 | 0.110 | 0.209 | <2.2e-16 | <2.2e-16 | 568 | 485 | 83 |
| European | No Recorded Depression | Switching vs Continuation | Lifetime_anxiety | age_at_first_AD | 0.844 | 0.630 | 1.130 | 2.54e-01 | 1.00e+00 | 568 | 485 | 83 |
| European | No Recorded Depression | Switching vs Continuation | Lifetime_anxiety | sex_at_birthMale | 1.550 | 0.953 | 2.521 | 7.76e-02 | 1.00e+00 | 568 | 485 | 83 |
| European | No Recorded Depression | Switching vs Continuation | Lifetime_anxiety | Income_level | 0.941 | 0.736 | 1.203 | 6.28e-01 | 1.00e+00 | 568 | 485 | 83 |
| European | No Recorded Depression | Switching vs Continuation | Lifetime_anxiety | Education_level | 0.879 | 0.680 | 1.136 | 3.26e-01 | 1.00e+00 | 568 | 485 | 83 |
| European | No Recorded Depression | Switching vs Continuation | Lifetime_anxiety | Lifetime_anxiety1 | 1.587 | 0.710 | 3.545 | 2.60e-01 | 1.00e+00 | 568 | 485 | 83 |
| European | No Recorded Depression | Discontinuation vs Continuation | Lifetime_anxiety | (Intercept) | 0.507 | 0.414 | 0.622 | 6.15e-11 | 1.05e-09 | 742 | 485 | 257 |
| European | No Recorded Depression | Discontinuation vs Continuation | Lifetime_anxiety | age_at_first_AD | 1.166 | 0.977 | 1.392 | 8.86e-02 | 1.00e+00 | 742 | 485 | 257 |
| European | No Recorded Depression | Discontinuation vs Continuation | Lifetime_anxiety | sex_at_birthMale | 1.115 | 0.805 | 1.545 | 5.12e-01 | 1.00e+00 | 742 | 485 | 257 |
| European | No Recorded Depression | Discontinuation vs Continuation | Lifetime_anxiety | Income_level | 0.961 | 0.820 | 1.126 | 6.22e-01 | 1.00e+00 | 742 | 485 | 257 |
| European | No Recorded Depression | Discontinuation vs Continuation | Lifetime_anxiety | Education_level | 0.857 | 0.721 | 1.019 | 8.07e-02 | 1.00e+00 | 742 | 485 | 257 |
| European | No Recorded Depression | Discontinuation vs Continuation | Lifetime_anxiety | Lifetime_anxiety1 | 0.456 | 0.196 | 1.057 | 6.73e-02 | 1.00e+00 | 742 | 485 | 257 |
| European | No Recorded Depression | Switching vs Continuation | Fibromyalgia | (Intercept) | 0.205 | 0.172 | 0.243 | <2.2e-16 | <2.2e-16 | 1520 | 1248 | 272 |
| European | No Recorded Depression | Switching vs Continuation | Fibromyalgia | age_at_first_AD | 0.812 | 0.699 | 0.942 | 5.94e-03 | 1.01e-01 | 1520 | 1248 | 272 |
| European | No Recorded Depression | Switching vs Continuation | Fibromyalgia | sex_at_birthMale | 1.318 | 0.995 | 1.745 | 5.41e-02 | 9.20e-01 | 1520 | 1248 | 272 |
| European | No Recorded Depression | Switching vs Continuation | Fibromyalgia | Income_level | 1.052 | 0.914 | 1.210 | 4.81e-01 | 1.00e+00 | 1520 | 1248 | 272 |
| European | No Recorded Depression | Switching vs Continuation | Fibromyalgia | Education_level | 0.894 | 0.771 | 1.037 | 1.38e-01 | 1.00e+00 | 1520 | 1248 | 272 |
| European | No Recorded Depression | Switching vs Continuation | Fibromyalgia | Fibromyalgia1 | 0.981 | 0.661 | 1.456 | 9.23e-01 | 1.00e+00 | 1520 | 1248 | 272 |
| European | No Recorded Depression | Discontinuation vs Continuation | Fibromyalgia | (Intercept) | 0.387 | 0.338 | 0.444 | <2.2e-16 | <2.2e-16 | 1791 | 1248 | 543 |
| European | No Recorded Depression | Discontinuation vs Continuation | Fibromyalgia | age_at_first_AD | 1.167 | 1.046 | 1.302 | 5.51e-03 | 9.37e-02 | 1791 | 1248 | 543 |
| European | No Recorded Depression | Discontinuation vs Continuation | Fibromyalgia | sex_at_birthMale | 1.256 | 1.011 | 1.561 | 3.97e-02 | 6.75e-01 | 1791 | 1248 | 543 |

| | | | | | | | | | | | | |
|---|---|---|---|---|---|---|---|---|---|---|---|---|
| European | No Recorded Depression | Discontinuation vs Continuation | Fibromyalgia | Income_level | 1.041 | 0.936 | 1.158 | 4.55e-01 | 1.00e+00 | 1791 | 1248 | 543 |
| European | No Recorded Depression | Discontinuation vs Continuation | Fibromyalgia | Education_level | 0.843 | 0.752 | 0.945 | 3.45e-03 | 5.86e-02 | 1791 | 1248 | 543 |
| European | No Recorded Depression | Discontinuation vs Continuation | Fibromyalgia | Fibromyalgia1 | 0.862 | 0.627 | 1.183 | 3.57e-01 | 1.00e+00 | 1791 | 1248 | 543 |
| European | No Recorded Depression | Switching vs Continuation | Childhood_sexual_abuse | (Intercept) | 0.217 | 0.180 | 0.262 | <2.2e-16 | <2.2e-16 | 1149 | 938 | 211 |
| European | No Recorded Depression | Switching vs Continuation | Childhood_sexual_abuse | age_at_first_AD | 0.844 | 0.712 | 1.000 | 5.02e-02 | 8.53e-01 | 1149 | 938 | 211 |
| European | No Recorded Depression | Switching vs Continuation | Childhood_sexual_abuse | sex_at_birthMale | 1.298 | 0.939 | 1.795 | 1.14e-01 | 1.00e+00 | 1149 | 938 | 211 |
| European | No Recorded Depression | Switching vs Continuation | Childhood_sexual_abuse | Income_level | 0.981 | 0.837 | 1.150 | 8.12e-01 | 1.00e+00 | 1149 | 938 | 211 |
| European | No Recorded Depression | Switching vs Continuation | Childhood_sexual_abuse | Education_level | 0.901 | 0.764 | 1.063 | 2.17e-01 | 1.00e+00 | 1149 | 938 | 211 |
| European | No Recorded Depression | Switching vs Continuation | Childhood_sexual_abuse | Childhood_sexual_a | 1.223 | 1.056 | 1.417 | 7.33e-03 | 1.25e-01 | 1149 | 938 | 211 |
| European | No Recorded Depression | Discontinuation vs Continuation | Childhood_sexual_abuse | (Intercept) | 0.374 | 0.320 | 0.437 | <2.2e-16 | <2.2e-16 | 1381 | 938 | 443 |
| European | No Recorded Depression | Discontinuation vs Continuation | Childhood_sexual_abuse | age_at_first_AD | 1.283 | 1.136 | 1.449 | 5.81e-05 | 9.88e-04 | 1381 | 938 | 443 |
| European | No Recorded Depression | Discontinuation vs Continuation | Childhood_sexual_abuse | sex_at_birthMale | 1.284 | 1.008 | 1.635 | 4.29e-02 | 7.29e-01 | 1381 | 938 | 443 |
| European | No Recorded Depression | Discontinuation vs Continuation | Childhood_sexual_abuse | Income_level | 0.994 | 0.883 | 1.120 | 9.24e-01 | 1.00e+00 | 1381 | 938 | 443 |
| European | No Recorded Depression | Discontinuation vs Continuation | Childhood_sexual_abuse | Education_level | 0.918 | 0.810 | 1.041 | 1.82e-01 | 1.00e+00 | 1381 | 938 | 443 |
| European | No Recorded Depression | Discontinuation vs Continuation | Childhood_sexual_abuse | Childhood_sexual_a | 0.838 | 0.689 | 1.020 | 7.83e-02 | 1.00e+00 | 1381 | 938 | 443 |
| European | No Recorded Depression | Switching vs Continuation | Neuroticism | (Intercept) | 0.206 | 0.183 | 0.232 | <2.2e-16 | <2.2e-16 | 2878 | 2329 | 549 |
| European | No Recorded Depression | Switching vs Continuation | Neuroticism | age_at_first_AD | 0.777 | 0.692 | 0.872 | 1.73e-05 | 2.94e-04 | 2878 | 2329 | 549 |
| European | No Recorded Depression | Switching vs Continuation | Neuroticism | sex_at_birthMale | 1.437 | 1.175 | 1.759 | 4.23e-04 | 7.20e-03 | 2878 | 2329 | 549 |
| European | No Recorded Depression | Switching vs Continuation | Neuroticism | Income_level | 0.957 | 0.865 | 1.059 | 3.97e-01 | 1.00e+00 | 2878 | 2329 | 549 |
| European | No Recorded Depression | Switching vs Continuation | Neuroticism | Education_level | 0.968 | 0.873 | 1.075 | 5.44e-01 | 1.00e+00 | 2878 | 2329 | 549 |
| European | No Recorded Depression | Switching vs Continuation | Neuroticism | Neuroticism | 1.284 | 1.160 | 1.422 | 1.48e-06 | 2.51e-05 | 2878 | 2329 | 549 |
| European | No Recorded Depression | Discontinuation vs Continuation | Neuroticism | (Intercept) | 0.351 | 0.319 | 0.387 | <2.2e-16 | <2.2e-16 | 3243 | 2329 | 914 |
| European | No Recorded Depression | Discontinuation vs Continuation | Neuroticism | age_at_first_AD | 1.124 | 1.024 | 1.235 | 1.38e-02 | 2.35e-01 | 3243 | 2329 | 914 |
| European | No Recorded Depression | Discontinuation vs Continuation | Neuroticism | sex_at_birthMale | 1.192 | 1.009 | 1.407 | 3.84e-02 | 6.53e-01 | 3243 | 2329 | 914 |
| European | No Recorded Depression | Discontinuation vs Continuation | Neuroticism | Income_level | 1.036 | 0.955 | 1.123 | 3.94e-01 | 1.00e+00 | 3243 | 2329 | 914 |
| European | No Recorded Depression | Discontinuation vs Continuation | Neuroticism | Education_level | 0.859 | 0.788 | 0.937 | 5.72e-04 | 9.72e-03 | 3243 | 2329 | 914 |
| European | No Recorded Depression | Discontinuation vs Continuation | Neuroticism | Neuroticism | 0.816 | 0.748 | 0.891 | 5.08e-06 | 8.64e-05 | 3243 | 2329 | 914 |
| European | No Recorded Depression | Switching vs Continuation | Longterm_difficulties | (Intercept) | 0.216 | 0.193 | 0.241 | <2.2e-16 | <2.2e-16 | 3082 | 2494 | 588 |
| European | No Recorded Depression | Switching vs Continuation | Longterm_difficulties | age_at_first_AD | 0.837 | 0.755 | 0.928 | 7.12e-04 | 1.21e-02 | 3082 | 2494 | 588 |
| European | No Recorded Depression | Switching vs Continuation | Longterm_difficulties | sex_at_birthMale | 1.372 | 1.131 | 1.663 | 1.30e-03 | 2.21e-02 | 3082 | 2494 | 588 |
| European | No Recorded Depression | Switching vs Continuation | Longterm_difficulties | Income_level | 0.954 | 0.865 | 1.052 | 3.46e-01 | 1.00e+00 | 3082 | 2494 | 588 |
| European | No Recorded Depression | Switching vs Continuation | Longterm_difficulties | Education_level | 0.965 | 0.873 | 1.066 | 4.84e-01 | 1.00e+00 | 3082 | 2494 | 588 |
| European | No Recorded Depression | Switching vs Continuation | Longterm_difficulties | Longterm_difficultie | 1.134 | 1.025 | 1.254 | 1.47e-02 | 2.51e-01 | 3082 | 2494 | 588 |
| European | No Recorded Depression | Discontinuation vs Continuation | Longterm_difficulties | (Intercept) | 0.362 | 0.330 | 0.397 | <2.2e-16 | <2.2e-16 | 3515 | 2494 | 1021 |
| European | No Recorded Depression | Discontinuation vs Continuation | Longterm_difficulties | age_at_first_AD | 1.198 | 1.107 | 1.297 | 7.50e-06 | 1.28e-04 | 3515 | 2494 | 1021 |
| European | No Recorded Depression | Discontinuation vs Continuation | Longterm_difficulties | sex_at_birthMale | 1.244 | 1.063 | 1.455 | 6.43e-03 | 1.09e-01 | 3515 | 2494 | 1021 |
| European | No Recorded Depression | Discontinuation vs Continuation | Longterm_difficulties | Income_level | 1.034 | 0.957 | 1.118 | 3.95e-01 | 1.00e+00 | 3515 | 2494 | 1021 |
| European | No Recorded Depression | Discontinuation vs Continuation | Longterm_difficulties | Education_level | 0.887 | 0.817 | 0.963 | 4.25e-03 | 7.23e-02 | 3515 | 2494 | 1021 |
| European | No Recorded Depression | Discontinuation vs Continuation | Longterm_difficulties | Longterm_difficultie | 0.946 | 0.864 | 1.036 | 2.30e-01 | 1.00e+00 | 3515 | 2494 | 1021 |
| European | No Recorded Depression | Switching vs Continuation | Threatening_experiences | (Intercept) | 0.214 | 0.192 | 0.240 | <2.2e-16 | <2.2e-16 | 3125 | 2527 | 598 |
| European | No Recorded Depression | Switching vs Continuation | Threatening_experiences | age_at_first_AD | 0.808 | 0.732 | 0.891 | 2.04e-05 | 3.46e-04 | 3125 | 2527 | 598 |
| European | No Recorded Depression | Switching vs Continuation | Threatening_experiences | sex_at_birthMale | 1.362 | 1.126 | 1.648 | 1.49e-03 | 2.53e-02 | 3125 | 2527 | 598 |
| European | No Recorded Depression | Switching vs Continuation | Threatening_experiences | Income_level | 0.938 | 0.852 | 1.033 | 1.96e-01 | 1.00e+00 | 3125 | 2527 | 598 |
| European | No Recorded Depression | Switching vs Continuation | Threatening_experiences | Education_level | 0.972 | 0.881 | 1.073 | 5.73e-01 | 1.00e+00 | 3125 | 2527 | 598 |
| European | No Recorded Depression | Switching vs Continuation | Threatening_experiences | Threatening_experie | 1.008 | 0.914 | 1.111 | 8.81e-01 | 1.00e+00 | 3125 | 2527 | 598 |
| European | No Recorded Depression | Discontinuation vs Continuation | Threatening_experiences | (Intercept) | 0.363 | 0.332 | 0.398 | <2.2e-16 | <2.2e-16 | 3554 | 2527 | 1027 |
| European | No Recorded Depression | Discontinuation vs Continuation | Threatening_experiences | age_at_first_AD | 1.206 | 1.118 | 1.301 | 1.30e-06 | 2.22e-05 | 3554 | 2527 | 1027 |
| European | No Recorded Depression | Discontinuation vs Continuation | Threatening_experiences | sex_at_birthMale | 1.256 | 1.075 | 1.468 | 4.16e-03 | 7.07e-02 | 3554 | 2527 | 1027 |
| European | No Recorded Depression | Discontinuation vs Continuation | Threatening_experiences | Income_level | 1.042 | 0.965 | 1.126 | 2.95e-01 | 1.00e+00 | 3554 | 2527 | 1027 |
| European | No Recorded Depression | Discontinuation vs Continuation | Threatening_experiences | Education_level | 0.884 | 0.815 | 0.958 | 2.80e-03 | 4.77e-02 | 3554 | 2527 | 1027 |
| European | No Recorded Depression | Discontinuation vs Continuation | Threatening_experiences | Threatening_experie | 1.032 | 0.952 | 1.118 | 4.51e-01 | 1.00e+00 | 3554 | 2527 | 1027 |

| | | | | | | | | | | | | |
|---|---|---|---|---|---|---|---|---|---|---|---|---|
| European | No Recorded Depression | Switching vs Continuation | Smoking_Packyears | (Intercept) | 0.225 | 0.194 | 0.260 | <2.2e-16 | <2.2e-16 | 1831 | 1479 | 352 |
| European | No Recorded Depression | Switching vs Continuation | Smoking_Packyears | age_at_first_AD | 0.692 | 0.602 | 0.794 | 1.81e-07 | 3.08e-06 | 1831 | 1479 | 352 |
| European | No Recorded Depression | Switching vs Continuation | Smoking_Packyears | sex_at_birthMale | 1.189 | 0.924 | 1.530 | 1.79e-01 | 1.00e+00 | 1831 | 1479 | 352 |
| European | No Recorded Depression | Switching vs Continuation | Smoking_Packyears | Income_level | 0.987 | 0.871 | 1.119 | 8.42e-01 | 1.00e+00 | 1831 | 1479 | 352 |
| European | No Recorded Depression | Switching vs Continuation | Smoking_Packyears | Education_level | 0.969 | 0.852 | 1.103 | 6.33e-01 | 1.00e+00 | 1831 | 1479 | 352 |
| European | No Recorded Depression | Switching vs Continuation | Smoking_Packyears | Smoking_Packyears | 1.245 | 1.102 | 1.407 | 4.22e-04 | 7.17e-03 | 1831 | 1479 | 352 |
| European | No Recorded Depression | Discontinuation vs Continuation | Smoking_Packyears | (Intercept) | 0.370 | 0.328 | 0.417 | <2.2e-16 | <2.2e-16 | 2126 | 1479 | 647 |
| European | No Recorded Depression | Discontinuation vs Continuation | Smoking_Packyears | age_at_first_AD | 1.210 | 1.094 | 1.338 | 2.16e-04 | 3.68e-03 | 2126 | 1479 | 647 |
| European | No Recorded Depression | Discontinuation vs Continuation | Smoking_Packyears | sex_at_birthMale | 1.338 | 1.099 | 1.629 | 3.78e-03 | 6.43e-02 | 2126 | 1479 | 647 |
| European | No Recorded Depression | Discontinuation vs Continuation | Smoking_Packyears | Income_level | 1.023 | 0.927 | 1.128 | 6.54e-01 | 1.00e+00 | 2126 | 1479 | 647 |
| European | No Recorded Depression | Discontinuation vs Continuation | Smoking_Packyears | Education_level | 0.869 | 0.783 | 0.965 | 8.52e-03 | 1.45e-01 | 2126 | 1479 | 647 |
| European | No Recorded Depression | Discontinuation vs Continuation | Smoking_Packyears | Smoking_Packyears | 1.075 | 0.978 | 1.182 | 1.35e-01 | 1.00e+00 | 2126 | 1479 | 647 |
| European | No Recorded Depression | Switching vs Continuation | Alcohol_intake | (Intercept) | 0.221 | 0.192 | 0.255 | <2.2e-16 | <2.2e-16 | 2074 | 1667 | 407 |
| European | No Recorded Depression | Switching vs Continuation | Alcohol_intake | age_at_first_AD | 0.804 | 0.714 | 0.906 | 3.27e-04 | 5.56e-03 | 2074 | 1667 | 407 |
| European | No Recorded Depression | Switching vs Continuation | Alcohol_intake | sex_at_birthMale | 1.387 | 1.094 | 1.757 | 6.87e-03 | 1.17e-01 | 2074 | 1667 | 407 |
| European | No Recorded Depression | Switching vs Continuation | Alcohol_intake | Income_level | 0.909 | 0.810 | 1.019 | 1.01e-01 | 1.00e+00 | 2074 | 1667 | 407 |
| European | No Recorded Depression | Switching vs Continuation | Alcohol_intake | Education_level | 0.933 | 0.829 | 1.050 | 2.48e-01 | 1.00e+00 | 2074 | 1667 | 407 |
| European | No Recorded Depression | Switching vs Continuation | Alcohol_intake | Alcohol_intake | 0.876 | 0.769 | 0.998 | 4.65e-02 | 7.90e-01 | 2074 | 1667 | 407 |
| European | No Recorded Depression | Discontinuation vs Continuation | Alcohol_intake | (Intercept) | 0.367 | 0.327 | 0.412 | <2.2e-16 | <2.2e-16 | 2346 | 1667 | 679 |
| European | No Recorded Depression | Discontinuation vs Continuation | Alcohol_intake | age_at_first_AD | 1.226 | 1.118 | 1.345 | 1.53e-05 | 2.61e-04 | 2346 | 1667 | 679 |
| European | No Recorded Depression | Discontinuation vs Continuation | Alcohol_intake | sex_at_birthMale | 1.197 | 0.984 | 1.456 | 7.27e-02 | 1.00e+00 | 2346 | 1667 | 679 |
| European | No Recorded Depression | Discontinuation vs Continuation | Alcohol_intake | Income_level | 1.035 | 0.943 | 1.137 | 4.70e-01 | 1.00e+00 | 2346 | 1667 | 679 |
| European | No Recorded Depression | Discontinuation vs Continuation | Alcohol_intake | Education_level | 0.857 | 0.777 | 0.947 | 2.29e-03 | 3.89e-02 | 2346 | 1667 | 679 |
| European | No Recorded Depression | Discontinuation vs Continuation | Alcohol_intake | Alcohol_intake | 1.117 | 1.018 | 1.224 | 1.91e-02 | 3.24e-01 | 2346 | 1667 | 679 |
| European | No Recorded Depression | Switching vs Continuation | Chronotype | (Intercept) | 0.221 | 0.195 | 0.251 | <2.2e-16 | <2.2e-16 | 2441 | 1966 | 475 |
| European | No Recorded Depression | Switching vs Continuation | Chronotype | age_at_first_AD | 0.803 | 0.718 | 0.898 | 1.17e-04 | 2.00e-03 | 2441 | 1966 | 475 |
| European | No Recorded Depression | Switching vs Continuation | Chronotype | sex_at_birthMale | 1.364 | 1.100 | 1.690 | 4.64e-03 | 7.89e-02 | 2441 | 1966 | 475 |
| European | No Recorded Depression | Switching vs Continuation | Chronotype | Income_level | 0.930 | 0.835 | 1.036 | 1.86e-01 | 1.00e+00 | 2441 | 1966 | 475 |
| European | No Recorded Depression | Switching vs Continuation | Chronotype | Education_level | 0.949 | 0.850 | 1.060 | 3.56e-01 | 1.00e+00 | 2441 | 1966 | 475 |
| European | No Recorded Depression | Switching vs Continuation | Chronotype | Chronotype | 1.110 | 0.998 | 1.234 | 5.41e-02 | 9.19e-01 | 2441 | 1966 | 475 |
| European | No Recorded Depression | Discontinuation vs Continuation | Chronotype | (Intercept) | 0.361 | 0.326 | 0.401 | <2.2e-16 | <2.2e-16 | 2768 | 1966 | 802 |
| European | No Recorded Depression | Discontinuation vs Continuation | Chronotype | age_at_first_AD | 1.208 | 1.106 | 1.319 | 2.59e-05 | 4.40e-04 | 2768 | 1966 | 802 |
| European | No Recorded Depression | Discontinuation vs Continuation | Chronotype | sex_at_birthMale | 1.251 | 1.049 | 1.492 | 1.25e-02 | 2.13e-01 | 2768 | 1966 | 802 |
| European | No Recorded Depression | Discontinuation vs Continuation | Chronotype | Income_level | 1.053 | 0.966 | 1.147 | 2.43e-01 | 1.00e+00 | 2768 | 1966 | 802 |
| European | No Recorded Depression | Discontinuation vs Continuation | Chronotype | Education_level | 0.894 | 0.816 | 0.979 | 1.60e-02 | 2.71e-01 | 2768 | 1966 | 802 |
| European | No Recorded Depression | Discontinuation vs Continuation | Chronotype | Chronotype | 1.042 | 0.953 | 1.139 | 3.70e-01 | 1.00e+00 | 2768 | 1966 | 802 |
| European | No Recorded Depression | Switching vs Continuation | Sleep_weekly | (Intercept) | 0.212 | 0.189 | 0.239 | <2.2e-16 | <2.2e-16 | 2796 | 2261 | 535 |
| European | No Recorded Depression | Switching vs Continuation | Sleep_weekly | age_at_first_AD | 0.813 | 0.732 | 0.903 | 1.07e-04 | 1.82e-03 | 2796 | 2261 | 535 |
| European | No Recorded Depression | Switching vs Continuation | Sleep_weekly | sex_at_birthMale | 1.409 | 1.149 | 1.728 | 1.00e-03 | 1.70e-02 | 2796 | 2261 | 535 |
| European | No Recorded Depression | Switching vs Continuation | Sleep_weekly | Income_level | 0.919 | 0.830 | 1.017 | 1.02e-01 | 1.00e+00 | 2796 | 2261 | 535 |
| European | No Recorded Depression | Switching vs Continuation | Sleep_weekly | Education_level | 0.954 | 0.860 | 1.059 | 3.76e-01 | 1.00e+00 | 2796 | 2261 | 535 |
| European | No Recorded Depression | Switching vs Continuation | Sleep_weekly | Sleep_weekly | 1.129 | 1.023 | 1.246 | 1.57e-02 | 2.67e-01 | 2796 | 2261 | 535 |
| European | No Recorded Depression | Discontinuation vs Continuation | Sleep_weekly | (Intercept) | 0.360 | 0.327 | 0.396 | <2.2e-16 | <2.2e-16 | 3171 | 2261 | 910 |
| European | No Recorded Depression | Discontinuation vs Continuation | Sleep_weekly | age_at_first_AD | 1.168 | 1.075 | 1.269 | 2.27e-04 | 3.86e-03 | 3171 | 2261 | 910 |
| European | No Recorded Depression | Discontinuation vs Continuation | Sleep_weekly | sex_at_birthMale | 1.261 | 1.068 | 1.489 | 6.14e-03 | 1.04e-01 | 3171 | 2261 | 910 |
| European | No Recorded Depression | Discontinuation vs Continuation | Sleep_weekly | Income_level | 1.034 | 0.954 | 1.121 | 4.15e-01 | 1.00e+00 | 3171 | 2261 | 910 |
| European | No Recorded Depression | Discontinuation vs Continuation | Sleep_weekly | Education_level | 0.883 | 0.811 | 0.962 | 4.44e-03 | 7.55e-02 | 3171 | 2261 | 910 |
| European | No Recorded Depression | Discontinuation vs Continuation | Sleep_weekly | Sleep_weekly | 0.924 | 0.846 | 1.009 | 7.80e-02 | 1.00e+00 | 3171 | 2261 | 910 |

**Supplementary Table 4. Multivariable logistic regression results for PGS associations with antidepressant treatment outcomes across ancestry groups and clinical subgroups in All of Us.** Each model estimates the association between 7 PGS traits and one of two binary outcomes — discontinuation versus continuation, or switching versus continuation — fitted separately within each combination of genetically inferred ancestry (European, African, Latino/Mixed-American), clinical group (Full Cohort; subgroups defined by recorded depression status, recorded anxiety, and CIDI-SF–derived MDD classification), and PGS exposure. Within each model, one PGS exposure is modelled alongside a common covariate set (age at first antidepressant, sex at birth, income, and educational attainment).All PGS were residualised on the top 16 global genetic ancestry principal components and the residuals standardised to mean 0, SD 1; each odds ratio therefore reflects the change in odds per one-standard-deviation increase in the PC-adjusted PGS. Because ancestry PCs were regressed out at this stage, they are not included in the model covariate set.

Columns: Population = inferred genetic ancestry; ClinicalGroup = cohort or clinical subgroup; Outcome = the binary contrast modelled; PGS = the PGS trait for that model; variable = the individual model term; odds_ratio = exponentiated coefficient; lower_95/upper_95 = 95% confidence interval bounds for the odds ratio; p_value = nominal (uncorrected) p-value; Bonf_P = Bonferroni-corrected p-value (capped at 1.00); N_Total = model sample size; N_Reference = number in the reference (continuation) group; N_Case = number in the comparison (discontinuation or switching) group. Odds ratios >1 indicate higher odds of the comparison outcome relative to continuation. Bonferroni correction was applied within each specification (ancestry × clinical group × outcome contrast) for the max 7 PGS traits tested. P-values reported as "<2.2e-16" reflect R's floating-point reporting limit.

Reference (omitted) categories: Sex at birth: Female; Income: lowest band (<$25k); Education level: lowest band (less than high school / fewer than twelve years).

| Population | ClinicalGroup | Outcome | PGS | variable | odds_ratio | lower_95 | upper_95 | p_value | Bonf_P | N_Total | N_Reference | N_Case |
|---|---|---|---|---|---|---|---|---|---|---|---|---|
| European | Full Cohort | Discontinuation vs Continuation | ADHD | (Intercept) | 0.332 | 0.293 | 0.377 | <2.2e-16 | NA | 28728 | 21256 | 7472 |
| European | Full Cohort | Discontinuation vs Continuation | ADHD | age_at_first_AD | 1.005 | 1.003 | 1.007 | 2.55e-09 | NA | 28728 | 21256 | 7472 |
| European | Full Cohort | Discontinuation vs Continuation | ADHD | sex_at_birthMale | 1.129 | 1.066 | 1.195 | 3.08e-05 | NA | 28728 | 21256 | 7472 |
| European | Full Cohort | Discontinuation vs Continuation | ADHD | Income_quantitative | 0.921 | 0.909 | 0.934 | <2.2e-16 | NA | 28728 | 21256 | 7472 |
| European | Full Cohort | Discontinuation vs Continuation | ADHD | Education_levelCollege Undergraduate | 1.107 | 1.038 | 1.181 | 2.04e-03 | NA | 28728 | 21256 | 7472 |
| European | Full Cohort | Discontinuation vs Continuation | ADHD | Education_levelLess than a high school degree | 1.774 | 1.455 | 2.162 | 1.43e-08 | NA | 28728 | 21256 | 7472 |
| European | Full Cohort | Discontinuation vs Continuation | ADHD | Education_levelTwelve or GED | 1.346 | 1.223 | 1.482 | 1.37e-09 | NA | 28728 | 21256 | 7472 |
| European | Full Cohort | Discontinuation vs Continuation | ADHD | ADHD | 1.047 | 1.019 | 1.076 | 1.03e-03 | 7.20e-03 | 28728 | 21256 | 7472 |
| European | Full Cohort | Switching vs Continuation | ADHD | (Intercept) | 2.864 | 2.585 | 3.173 | <2.2e-16 | NA | 35092 | 21256 | 13836 |
| European | Full Cohort | Switching vs Continuation | ADHD | age_at_first_AD | 0.978 | 0.977 | 0.980 | <2.2e-16 | NA | 35092 | 21256 | 13836 |
| European | Full Cohort | Switching vs Continuation | ADHD | sex_at_birthMale | 1.052 | 1.002 | 1.104 | 4.25e-02 | NA | 35092 | 21256 | 13836 |
| European | Full Cohort | Switching vs Continuation | ADHD | Income_quantitative | 0.884 | 0.875 | 0.894 | <2.2e-16 | NA | 35092 | 21256 | 13836 |
| European | Full Cohort | Switching vs Continuation | ADHD | Education_levelCollege Undergraduate | 1.091 | 1.034 | 1.151 | 1.41e-03 | NA | 35092 | 21256 | 13836 |
| European | Full Cohort | Switching vs Continuation | ADHD | Education_levelLess than a high school degree | 1.311 | 1.098 | 1.564 | 2.69e-03 | NA | 35092 | 21256 | 13836 |
| European | Full Cohort | Switching vs Continuation | ADHD | Education_levelTwelve or GED | 1.273 | 1.174 | 1.381 | 5.36e-09 | NA | 35092 | 21256 | 13836 |
| European | Full Cohort | Switching vs Continuation | ADHD | ADHD | 1.065 | 1.041 | 1.089 | 6.21e-08 | 4.35e-07 | 35092 | 21256 | 13836 |
| European | Full Cohort | Discontinuation vs Continuation | BMI | (Intercept) | 0.335 | 0.295 | 0.380 | <2.2e-16 | NA | 28728 | 21256 | 7472 |
| European | Full Cohort | Discontinuation vs Continuation | BMI | age_at_first_AD | 1.005 | 1.003 | 1.007 | 5.02e-09 | NA | 28728 | 21256 | 7472 |
| European | Full Cohort | Discontinuation vs Continuation | BMI | sex_at_birthMale | 1.127 | 1.064 | 1.193 | 4.07e-05 | NA | 28728 | 21256 | 7472 |
| European | Full Cohort | Discontinuation vs Continuation | BMI | Income_quantitative | 0.919 | 0.907 | 0.932 | <2.2e-16 | NA | 28728 | 21256 | 7472 |
| European | Full Cohort | Discontinuation vs Continuation | BMI | Education_levelCollege Undergraduate | 1.115 | 1.046 | 1.190 | 9.33e-04 | NA | 28728 | 21256 | 7472 |
| European | Full Cohort | Discontinuation vs Continuation | BMI | Education_levelLess than a high school degree | 1.820 | 1.494 | 2.218 | 2.84e-09 | NA | 28728 | 21256 | 7472 |
| European | Full Cohort | Discontinuation vs Continuation | BMI | Education_levelTwelve or GED | 1.364 | 1.239 | 1.501 | 2.45e-10 | NA | 28728 | 21256 | 7472 |
| European | Full Cohort | Discontinuation vs Continuation | BMI | BMI | 0.998 | 0.971 | 1.025 | 8.65e-01 | 1.00e+00 | 28728 | 21256 | 7472 |
| European | Full Cohort | Switching vs Continuation | BMI | (Intercept) | 2.858 | 2.579 | 3.166 | <2.2e-16 | NA | 35092 | 21256 | 13836 |
| European | Full Cohort | Switching vs Continuation | BMI | age_at_first_AD | 0.978 | 0.977 | 0.980 | <2.2e-16 | NA | 35092 | 21256 | 13836 |
| European | Full Cohort | Switching vs Continuation | BMI | sex_at_birthMale | 1.050 | 1.000 | 1.102 | 5.19e-02 | NA | 35092 | 21256 | 13836 |
| European | Full Cohort | Switching vs Continuation | BMI | Income_quantitative | 0.884 | 0.874 | 0.894 | <2.2e-16 | NA | 35092 | 21256 | 13836 |
| European | Full Cohort | Switching vs Continuation | BMI | Education_levelCollege Undergraduate | 1.095 | 1.038 | 1.156 | 8.46e-04 | NA | 35092 | 21256 | 13836 |
| European | Full Cohort | Switching vs Continuation | BMI | Education_levelLess than a high school degree | 1.333 | 1.118 | 1.590 | 1.40e-03 | NA | 35092 | 21256 | 13836 |
| European | Full Cohort | Switching vs Continuation | BMI | Education_levelTwelve or GED | 1.281 | 1.181 | 1.389 | 2.06e-09 | NA | 35092 | 21256 | 13836 |
| European | Full Cohort | Switching vs Continuation | BMI | BMI | 1.054 | 1.031 | 1.078 | 3.54e-06 | 2.48e-05 | 35092 | 21256 | 13836 |
| European | Full Cohort | Discontinuation vs Continuation | Bipolar Disorder | (Intercept) | 0.334 | 0.294 | 0.379 | <2.2e-16 | NA | 28728 | 21256 | 7472 |
| European | Full Cohort | Discontinuation vs Continuation | Bipolar Disorder | age_at_first_AD | 1.005 | 1.003 | 1.007 | 6.09e-09 | NA | 28728 | 21256 | 7472 |
| European | Full Cohort | Discontinuation vs Continuation | Bipolar Disorder | sex_at_birthMale | 1.126 | 1.063 | 1.192 | 4.70e-05 | NA | 28728 | 21256 | 7472 |
| European | Full Cohort | Discontinuation vs Continuation | Bipolar Disorder | Income_quantitative | 0.920 | 0.908 | 0.932 | <2.2e-16 | NA | 28728 | 21256 | 7472 |
| European | Full Cohort | Discontinuation vs Continuation | Bipolar Disorder | Education_levelCollege Undergraduate | 1.118 | 1.048 | 1.193 | 7.07e-04 | NA | 28728 | 21256 | 7472 |
| European | Full Cohort | Discontinuation vs Continuation | Bipolar Disorder | Education_levelLess than a high school degree | 1.824 | 1.497 | 2.223 | 2.44e-09 | NA | 28728 | 21256 | 7472 |
| European | Full Cohort | Discontinuation vs Continuation | Bipolar Disorder | Education_levelTwelve or GED | 1.371 | 1.246 | 1.509 | 1.16e-10 | NA | 28728 | 21256 | 7472 |
| European | Full Cohort | Discontinuation vs Continuation | Bipolar Disorder | Bipolar Disorder | 1.050 | 1.021 | 1.080 | 6.16e-04 | 4.31e-03 | 28728 | 21256 | 7472 |
| European | Full Cohort | Switching vs Continuation | Bipolar Disorder | (Intercept) | 2.887 | 2.606 | 3.199 | <2.2e-16 | NA | 35092 | 21256 | 13836 |
| European | Full Cohort | Switching vs Continuation | Bipolar Disorder | age_at_first_AD | 0.978 | 0.977 | 0.980 | <2.2e-16 | NA | 35092 | 21256 | 13836 |
| European | Full Cohort | Switching vs Continuation | Bipolar Disorder | sex_at_birthMale | 1.049 | 0.999 | 1.101 | 5.66e-02 | NA | 35092 | 21256 | 13836 |
| European | Full Cohort | Switching vs Continuation | Bipolar Disorder | Income_quantitative | 0.882 | 0.873 | 0.892 | <2.2e-16 | NA | 35092 | 21256 | 13836 |
| European | Full Cohort | Switching vs Continuation | Bipolar Disorder | Education_levelCollege Undergraduate | 1.104 | 1.047 | 1.165 | 2.78e-04 | NA | 35092 | 21256 | 13836 |

| | | | | | | | | | | | | |
|---|---|---|---|---|---|---|---|---|---|---|---|---|
| European | Full Cohort | Switching vs Continuation | Bipolar Disorder | Education_levelLess than a high school degree | 1.354 | 1.135 | 1.615 | 7.54e-04 | NA | 35092 | 21256 | 13836 |
| European | Full Cohort | Switching vs Continuation | Bipolar Disorder | Education_levelTwelve or GED | 1.301 | 1.200 | 1.411 | 1.76e-10 | NA | 35092 | 21256 | 13836 |
| European | Full Cohort | Switching vs Continuation | Bipolar Disorder | Bipolar Disorder | 1.044 | 1.020 | 1.069 | 2.76e-04 | 1.93e-03 | 35092 | 21256 | 13836 |
| European | Full Cohort | Discontinuation vs Continuation | Depression | (Intercept) | 0.330 | 0.290 | 0.374 | <2.2e-16 | NA | 28728 | 21256 | 7472 |
| European | Full Cohort | Discontinuation vs Continuation | Depression | age_at_first_AD | 1.005 | 1.004 | 1.007 | 1.27e-09 | NA | 28728 | 21256 | 7472 |
| European | Full Cohort | Discontinuation vs Continuation | Depression | sex_at_birthMale | 1.124 | 1.062 | 1.190 | 5.95e-05 | NA | 28728 | 21256 | 7472 |
| European | Full Cohort | Discontinuation vs Continuation | Depression | Income_quantitative | 0.922 | 0.910 | 0.935 | <2.2e-16 | NA | 28728 | 21256 | 7472 |
| European | Full Cohort | Discontinuation vs Continuation | Depression | Education_levelCollege Undergraduate | 1.109 | 1.040 | 1.183 | 1.66e-03 | NA | 28728 | 21256 | 7472 |
| European | Full Cohort | Discontinuation vs Continuation | Depression | Education_levelLess than a high school degree | 1.761 | 1.445 | 2.147 | 2.06e-08 | NA | 28728 | 21256 | 7472 |
| European | Full Cohort | Discontinuation vs Continuation | Depression | Education_levelTwelve or GED | 1.353 | 1.229 | 1.489 | 7.03e-10 | NA | 28728 | 21256 | 7472 |
| European | Full Cohort | Discontinuation vs Continuation | Depression | Depression | 1.097 | 1.067 | 1.128 | 5.03e-11 | 3.52e-10 | 28728 | 21256 | 7472 |
| European | Full Cohort | Switching vs Continuation | Depression | (Intercept) | 2.799 | 2.525 | 3.102 | <2.2e-16 | NA | 35092 | 21256 | 13836 |
| European | Full Cohort | Switching vs Continuation | Depression | age_at_first_AD | 0.978 | 0.977 | 0.980 | <2.2e-16 | NA | 35092 | 21256 | 13836 |
| European | Full Cohort | Switching vs Continuation | Depression | sex_at_birthMale | 1.040 | 0.991 | 1.092 | 1.14e-01 | NA | 35092 | 21256 | 13836 |
| European | Full Cohort | Switching vs Continuation | Depression | Income_quantitative | 0.888 | 0.878 | 0.898 | <2.2e-16 | NA | 35092 | 21256 | 13836 |
| European | Full Cohort | Switching vs Continuation | Depression | Education_levelCollege Undergraduate | 1.091 | 1.034 | 1.152 | 1.38e-03 | NA | 35092 | 21256 | 13836 |
| European | Full Cohort | Switching vs Continuation | Depression | Education_levelLess than a high school degree | 1.264 | 1.058 | 1.509 | 9.64e-03 | NA | 35092 | 21256 | 13836 |
| European | Full Cohort | Switching vs Continuation | Depression | Education_levelTwelve or GED | 1.278 | 1.179 | 1.386 | 3.02e-09 | NA | 35092 | 21256 | 13836 |
| European | Full Cohort | Switching vs Continuation | Depression | Depression | 1.212 | 1.184 | 1.241 | <2.2e-16 | <2.2e-16 | 35092 | 21256 | 13836 |
| European | Full Cohort | Discontinuation vs Continuation | Insomnia | (Intercept) | 0.330 | 0.291 | 0.375 | <2.2e-16 | NA | 28728 | 21256 | 7472 |
| European | Full Cohort | Discontinuation vs Continuation | Insomnia | age_at_first_AD | 1.005 | 1.004 | 1.007 | 1.65e-09 | NA | 28728 | 21256 | 7472 |
| European | Full Cohort | Discontinuation vs Continuation | Insomnia | sex_at_birthMale | 1.127 | 1.064 | 1.193 | 4.20e-05 | NA | 28728 | 21256 | 7472 |
| European | Full Cohort | Discontinuation vs Continuation | Insomnia | Income_quantitative | 0.922 | 0.910 | 0.934 | <2.2e-16 | NA | 28728 | 21256 | 7472 |
| European | Full Cohort | Discontinuation vs Continuation | Insomnia | Education_levelCollege Undergraduate | 1.108 | 1.038 | 1.182 | 1.92e-03 | NA | 28728 | 21256 | 7472 |
| European | Full Cohort | Discontinuation vs Continuation | Insomnia | Education_levelLess than a high school degree | 1.765 | 1.448 | 2.151 | 1.86e-08 | NA | 28728 | 21256 | 7472 |
| European | Full Cohort | Discontinuation vs Continuation | Insomnia | Education_levelTwelve or GED | 1.348 | 1.224 | 1.484 | 1.12e-09 | NA | 28728 | 21256 | 7472 |
| European | Full Cohort | Discontinuation vs Continuation | Insomnia | Insomnia | 1.086 | 1.056 | 1.116 | 7.53e-09 | 5.27e-08 | 28728 | 21256 | 7472 |
| European | Full Cohort | Switching vs Continuation | Insomnia | (Intercept) | 2.842 | 2.565 | 3.149 | <2.2e-16 | NA | 35092 | 21256 | 13836 |
| European | Full Cohort | Switching vs Continuation | Insomnia | age_at_first_AD | 0.978 | 0.977 | 0.980 | <2.2e-16 | NA | 35092 | 21256 | 13836 |
| European | Full Cohort | Switching vs Continuation | Insomnia | sex_at_birthMale | 1.047 | 0.998 | 1.100 | 6.23e-02 | NA | 35092 | 21256 | 13836 |
| European | Full Cohort | Switching vs Continuation | Insomnia | Income_quantitative | 0.885 | 0.875 | 0.895 | <2.2e-16 | NA | 35092 | 21256 | 13836 |
| European | Full Cohort | Switching vs Continuation | Insomnia | Education_levelCollege Undergraduate | 1.094 | 1.037 | 1.155 | 9.48e-04 | NA | 35092 | 21256 | 13836 |
| European | Full Cohort | Switching vs Continuation | Insomnia | Education_levelLess than a high school degree | 1.303 | 1.092 | 1.555 | 3.31e-03 | NA | 35092 | 21256 | 13836 |
| European | Full Cohort | Switching vs Continuation | Insomnia | Education_levelTwelve or GED | 1.279 | 1.180 | 1.387 | 2.54e-09 | NA | 35092 | 21256 | 13836 |
| European | Full Cohort | Switching vs Continuation | Insomnia | Insomnia | 1.106 | 1.080 | 1.132 | <2.2e-16 | <2.2e-16 | 35092 | 21256 | 13836 |
| European | Full Cohort | Discontinuation vs Continuation | Migraine | (Intercept) | 0.335 | 0.295 | 0.380 | <2.2e-16 | NA | 28728 | 21256 | 7472 |
| European | Full Cohort | Discontinuation vs Continuation | Migraine | age_at_first_AD | 1.005 | 1.003 | 1.007 | 4.59e-09 | NA | 28728 | 21256 | 7472 |
| European | Full Cohort | Discontinuation vs Continuation | Migraine | sex_at_birthMale | 1.127 | 1.064 | 1.193 | 4.06e-05 | NA | 28728 | 21256 | 7472 |
| European | Full Cohort | Discontinuation vs Continuation | Migraine | Income_quantitative | 0.920 | 0.907 | 0.932 | <2.2e-16 | NA | 28728 | 21256 | 7472 |
| European | Full Cohort | Discontinuation vs Continuation | Migraine | Education_levelCollege Undergraduate | 1.115 | 1.045 | 1.189 | 9.86e-04 | NA | 28728 | 21256 | 7472 |
| European | Full Cohort | Discontinuation vs Continuation | Migraine | Education_levelLess than a high school degree | 1.819 | 1.493 | 2.216 | 2.92e-09 | NA | 28728 | 21256 | 7472 |
| European | Full Cohort | Discontinuation vs Continuation | Migraine | Education_levelTwelve or GED | 1.363 | 1.238 | 1.500 | 2.61e-10 | NA | 28728 | 21256 | 7472 |
| European | Full Cohort | Discontinuation vs Continuation | Migraine | Migraine | 1.009 | 0.982 | 1.036 | 5.28e-01 | 1.00e+00 | 28728 | 21256 | 7472 |
| European | Full Cohort | Switching vs Continuation | Migraine | (Intercept) | 2.880 | 2.599 | 3.191 | <2.2e-16 | NA | 35092 | 21256 | 13836 |
| European | Full Cohort | Switching vs Continuation | Migraine | age_at_first_AD | 0.978 | 0.977 | 0.980 | <2.2e-16 | NA | 35092 | 21256 | 13836 |
| European | Full Cohort | Switching vs Continuation | Migraine | sex_at_birthMale | 1.049 | 0.999 | 1.101 | 5.64e-02 | NA | 35092 | 21256 | 13836 |
| European | Full Cohort | Switching vs Continuation | Migraine | Income_quantitative | 0.882 | 0.873 | 0.892 | <2.2e-16 | NA | 35092 | 21256 | 13836 |
| European | Full Cohort | Switching vs Continuation | Migraine | Education_levelCollege Undergraduate | 1.101 | 1.044 | 1.161 | 4.26e-04 | NA | 35092 | 21256 | 13836 |
| European | Full Cohort | Switching vs Continuation | Migraine | Education_levelLess than a high school degree | 1.348 | 1.130 | 1.608 | 8.89e-04 | NA | 35092 | 21256 | 13836 |
| European | Full Cohort | Switching vs Continuation | Migraine | Education_levelTwelve or GED | 1.294 | 1.193 | 1.402 | 4.36e-10 | NA | 35092 | 21256 | 13836 |
| European | Full Cohort | Switching vs Continuation | Migraine | Migraine | 1.047 | 1.024 | 1.070 | 4.82e-05 | 3.37e-04 | 35092 | 21256 | 13836 |
| European | Full Cohort | Discontinuation vs Continuation | Schizophrenia | (Intercept) | 0.333 | 0.294 | 0.378 | <2.2e-16 | NA | 28728 | 21256 | 7472 |
| European | Full Cohort | Discontinuation vs Continuation | Schizophrenia | age_at_first_AD | 1.005 | 1.003 | 1.007 | 4.08e-09 | NA | 28728 | 21256 | 7472 |
| European | Full Cohort | Discontinuation vs Continuation | Schizophrenia | sex_at_birthMale | 1.127 | 1.064 | 1.193 | 4.24e-05 | NA | 28728 | 21256 | 7472 |
| European | Full Cohort | Discontinuation vs Continuation | Schizophrenia | Income_quantitative | 0.920 | 0.908 | 0.932 | <2.2e-16 | NA | 28728 | 21256 | 7472 |
| European | Full Cohort | Discontinuation vs Continuation | Schizophrenia | Education_levelCollege Undergraduate | 1.116 | 1.046 | 1.191 | 8.49e-04 | NA | 28728 | 21256 | 7472 |
| European | Full Cohort | Discontinuation vs Continuation | Schizophrenia | Education_levelLess than a high school degree | 1.811 | 1.486 | 2.207 | 3.79e-09 | NA | 28728 | 21256 | 7472 |

| | | | | | | | | | | | | |
|---|---|---|---|---|---|---|---|---|---|---|---|---|
| European | Full Cohort | Discontinuation vs Continuation | Schizophrenia | Education_levelTwelve or GED | 1.366 | 1.241 | 1.503 | 1.90e-10 | NA | 28728 | 21256 | 7472 |
| European | Full Cohort | Discontinuation vs Continuation | Schizophrenia | Schizophrenia | 1.033 | 1.006 | 1.061 | 1.82e-02 | 1.28e-01 | 28728 | 21256 | 7472 |
| European | Full Cohort | Switching vs Continuation | Schizophrenia | (Intercept) | 2.881 | 2.600 | 3.192 | <2.2e-16 | NA | 35092 | 21256 | 13836 |
| European | Full Cohort | Switching vs Continuation | Schizophrenia | age_at_first_AD | 0.978 | 0.977 | 0.980 | <2.2e-16 | NA | 35092 | 21256 | 13836 |
| European | Full Cohort | Switching vs Continuation | Schizophrenia | sex_at_birthMale | 1.049 | 0.999 | 1.101 | 5.40e-02 | NA | 35092 | 21256 | 13836 |
| European | Full Cohort | Switching vs Continuation | Schizophrenia | Income_quantitative | 0.882 | 0.873 | 0.892 | <2.2e-16 | NA | 35092 | 21256 | 13836 |
| European | Full Cohort | Switching vs Continuation | Schizophrenia | Education_levelCollege Undergraduate | 1.103 | 1.045 | 1.163 | 3.36e-04 | NA | 35092 | 21256 | 13836 |
| European | Full Cohort | Switching vs Continuation | Schizophrenia | Education_levelLess than a high school degree | 1.350 | 1.132 | 1.610 | 8.44e-04 | NA | 35092 | 21256 | 13836 |
| European | Full Cohort | Switching vs Continuation | Schizophrenia | Education_levelTwelve or GED | 1.297 | 1.196 | 1.406 | 3.01e-10 | NA | 35092 | 21256 | 13836 |
| European | Full Cohort | Switching vs Continuation | Schizophrenia | Schizophrenia | 1.014 | 0.991 | 1.037 | 2.34e-01 | 1.00e+00 | 35092 | 21256 | 13836 |
| European | Recorded Depression | Discontinuation vs Continuation | ADHD | (Intercept) | 0.269 | 0.213 | 0.338 | <2.2e-16 | NA | 10166 | 7969 | 2197 |
| European | Recorded Depression | Discontinuation vs Continuation | ADHD | age_at_first_AD | 1.005 | 1.002 | 1.008 | 7.88e-04 | NA | 10166 | 7969 | 2197 |
| European | Recorded Depression | Discontinuation vs Continuation | ADHD | sex_at_birthMale | 1.155 | 1.042 | 1.281 | 6.18e-03 | NA | 10166 | 7969 | 2197 |
| European | Recorded Depression | Discontinuation vs Continuation | ADHD | Income_quantitative | 0.874 | 0.852 | 0.896 | <2.2e-16 | NA | 10166 | 7969 | 2197 |
| European | Recorded Depression | Discontinuation vs Continuation | ADHD | Education_levelCollege Undergraduate | 1.261 | 1.112 | 1.429 | 2.89e-04 | NA | 10166 | 7969 | 2197 |
| European | Recorded Depression | Discontinuation vs Continuation | ADHD | Education_levelLess than a high school degree | 2.478 | 1.830 | 3.355 | 4.38e-09 | NA | 10166 | 7969 | 2197 |
| European | Recorded Depression | Discontinuation vs Continuation | ADHD | Education_levelTwelve or GED | 1.604 | 1.351 | 1.904 | 6.95e-08 | NA | 10166 | 7969 | 2197 |
| European | Recorded Depression | Discontinuation vs Continuation | ADHD | ADHD | 1.083 | 1.030 | 1.138 | 1.65e-03 | 1.15e-02 | 10166 | 7969 | 2197 |
| European | Recorded Depression | Switching vs Continuation | ADHD | (Intercept) | 4.041 | 3.499 | 4.667 | <2.2e-16 | NA | 17576 | 7969 | 9607 |
| European | Recorded Depression | Switching vs Continuation | ADHD | age_at_first_AD | 0.979 | 0.977 | 0.981 | <2.2e-16 | NA | 17576 | 7969 | 9607 |
| European | Recorded Depression | Switching vs Continuation | ADHD | sex_at_birthMale | 1.129 | 1.056 | 1.207 | 3.84e-04 | NA | 17576 | 7969 | 9607 |
| European | Recorded Depression | Switching vs Continuation | ADHD | Income_quantitative | 0.915 | 0.901 | 0.929 | <2.2e-16 | NA | 17576 | 7969 | 9607 |
| European | Recorded Depression | Switching vs Continuation | ADHD | Education_levelCollege Undergraduate | 1.163 | 1.079 | 1.253 | 7.37e-05 | NA | 17576 | 7969 | 9607 |
| European | Recorded Depression | Switching vs Continuation | ADHD | Education_levelLess than a high school degree | 1.425 | 1.125 | 1.805 | 3.30e-03 | NA | 17576 | 7969 | 9607 |
| European | Recorded Depression | Switching vs Continuation | ADHD | Education_levelTwelve or GED | 1.408 | 1.259 | 1.575 | 2.07e-09 | NA | 17576 | 7969 | 9607 |
| European | Recorded Depression | Switching vs Continuation | ADHD | ADHD | 1.051 | 1.018 | 1.084 | 2.04e-03 | 1.43e-02 | 17576 | 7969 | 9607 |
| European | Recorded Depression | Discontinuation vs Continuation | BMI | (Intercept) | 0.270 | 0.215 | 0.340 | <2.2e-16 | NA | 10166 | 7969 | 2197 |
| European | Recorded Depression | Discontinuation vs Continuation | BMI | age_at_first_AD | 1.005 | 1.002 | 1.008 | 8.94e-04 | NA | 10166 | 7969 | 2197 |
| European | Recorded Depression | Discontinuation vs Continuation | BMI | sex_at_birthMale | 1.149 | 1.036 | 1.273 | 8.39e-03 | NA | 10166 | 7969 | 2197 |
| European | Recorded Depression | Discontinuation vs Continuation | BMI | Income_quantitative | 0.872 | 0.850 | 0.894 | <2.2e-16 | NA | 10166 | 7969 | 2197 |
| European | Recorded Depression | Discontinuation vs Continuation | BMI | Education_levelCollege Undergraduate | 1.276 | 1.126 | 1.446 | 1.38e-04 | NA | 10166 | 7969 | 2197 |
| European | Recorded Depression | Discontinuation vs Continuation | BMI | Education_levelLess than a high school degree | 2.574 | 1.903 | 3.482 | 8.49e-10 | NA | 10166 | 7969 | 2197 |
| European | Recorded Depression | Discontinuation vs Continuation | BMI | Education_levelTwelve or GED | 1.634 | 1.376 | 1.939 | 1.98e-08 | NA | 10166 | 7969 | 2197 |
| European | Recorded Depression | Discontinuation vs Continuation | BMI | BMI | 1.018 | 0.969 | 1.068 | 4.83e-01 | 1.00e+00 | 10166 | 7969 | 2197 |
| European | Recorded Depression | Switching vs Continuation | BMI | (Intercept) | 4.042 | 3.500 | 4.669 | <2.2e-16 | NA | 17576 | 7969 | 9607 |
| European | Recorded Depression | Switching vs Continuation | BMI | age_at_first_AD | 0.979 | 0.977 | 0.981 | <2.2e-16 | NA | 17576 | 7969 | 9607 |
| European | Recorded Depression | Switching vs Continuation | BMI | sex_at_birthMale | 1.127 | 1.054 | 1.205 | 4.60e-04 | NA | 17576 | 7969 | 9607 |
| European | Recorded Depression | Switching vs Continuation | BMI | Income_quantitative | 0.913 | 0.899 | 0.928 | <2.2e-16 | NA | 17576 | 7969 | 9607 |
| European | Recorded Depression | Switching vs Continuation | BMI | Education_levelCollege Undergraduate | 1.171 | 1.087 | 1.261 | 3.20e-05 | NA | 17576 | 7969 | 9607 |
| European | Recorded Depression | Switching vs Continuation | BMI | Education_levelLess than a high school degree | 1.451 | 1.146 | 1.837 | 1.99e-03 | NA | 17576 | 7969 | 9607 |
| European | Recorded Depression | Switching vs Continuation | BMI | Education_levelTwelve or GED | 1.424 | 1.273 | 1.592 | 5.62e-10 | NA | 17576 | 7969 | 9607 |
| European | Recorded Depression | Switching vs Continuation | BMI | BMI | 1.016 | 0.985 | 1.047 | 3.24e-01 | 1.00e+00 | 17576 | 7969 | 9607 |
| European | Recorded Depression | Discontinuation vs Continuation | Bipolar Disorder | (Intercept) | 0.272 | 0.216 | 0.342 | <2.2e-16 | NA | 10166 | 7969 | 2197 |
| European | Recorded Depression | Discontinuation vs Continuation | Bipolar Disorder | age_at_first_AD | 1.005 | 1.002 | 1.008 | 1.16e-03 | NA | 10166 | 7969 | 2197 |
| European | Recorded Depression | Discontinuation vs Continuation | Bipolar Disorder | sex_at_birthMale | 1.148 | 1.036 | 1.273 | 8.64e-03 | NA | 10166 | 7969 | 2197 |
| European | Recorded Depression | Discontinuation vs Continuation | Bipolar Disorder | Income_quantitative | 0.871 | 0.849 | 0.893 | <2.2e-16 | NA | 10166 | 7969 | 2197 |
| European | Recorded Depression | Discontinuation vs Continuation | Bipolar Disorder | Education_levelCollege Undergraduate | 1.280 | 1.130 | 1.451 | 1.08e-04 | NA | 10166 | 7969 | 2197 |
| European | Recorded Depression | Discontinuation vs Continuation | Bipolar Disorder | Education_levelLess than a high school degree | 2.593 | 1.917 | 3.507 | 6.28e-10 | NA | 10166 | 7969 | 2197 |
| European | Recorded Depression | Discontinuation vs Continuation | Bipolar Disorder | Education_levelTwelve or GED | 1.649 | 1.390 | 1.957 | 1.01e-08 | NA | 10166 | 7969 | 2197 |
| European | Recorded Depression | Discontinuation vs Continuation | Bipolar Disorder | Bipolar Disorder | 1.078 | 1.024 | 1.134 | 3.90e-03 | 2.73e-02 | 10166 | 7969 | 2197 |
| European | Recorded Depression | Switching vs Continuation | Bipolar Disorder | (Intercept) | 4.058 | 3.513 | 4.686 | <2.2e-16 | NA | 17576 | 7969 | 9607 |
| European | Recorded Depression | Switching vs Continuation | Bipolar Disorder | age_at_first_AD | 0.979 | 0.977 | 0.981 | <2.2e-16 | NA | 17576 | 7969 | 9607 |
| European | Recorded Depression | Switching vs Continuation | Bipolar Disorder | sex_at_birthMale | 1.127 | 1.054 | 1.205 | 4.60e-04 | NA | 17576 | 7969 | 9607 |
| European | Recorded Depression | Switching vs Continuation | Bipolar Disorder | Income_quantitative | 0.913 | 0.899 | 0.927 | <2.2e-16 | NA | 17576 | 7969 | 9607 |
| European | Recorded Depression | Switching vs Continuation | Bipolar Disorder | Education_levelCollege Undergraduate | 1.173 | 1.089 | 1.264 | 2.51e-05 | NA | 17576 | 7969 | 9607 |
| European | Recorded Depression | Switching vs Continuation | Bipolar Disorder | Education_levelLess than a high school degree | 1.457 | 1.151 | 1.844 | 1.75e-03 | NA | 17576 | 7969 | 9607 |
| European | Recorded Depression | Switching vs Continuation | Bipolar Disorder | Education_levelTwelve or GED | 1.430 | 1.279 | 1.598 | 3.38e-10 | NA | 17576 | 7969 | 9607 |

| | | | | | | | | | | | | |
|---|---|---|---|---|---|---|---|---|---|---|---|---|
| European | Recorded Depression | Switching vs Continuation | Bipolar Disorder | Bipolar Disorder | 1.012 | 0.981 | 1.045 | 4.48e-01 | 1.00e+00 | 17576 | 7969 | 9607 |
| European | Recorded Depression | Discontinuation vs Continuation | Depression | (Intercept) | 0.267 | 0.212 | 0.336 | <2.2e-16 | NA | 10166 | 7969 | 2197 |
| European | Recorded Depression | Discontinuation vs Continuation | Depression | age_at_first_AD | 1.005 | 1.002 | 1.008 | 7.19e-04 | NA | 10166 | 7969 | 2197 |
| European | Recorded Depression | Discontinuation vs Continuation | Depression | sex_at_birthMale | 1.148 | 1.035 | 1.273 | 8.94e-03 | NA | 10166 | 7969 | 2197 |
| European | Recorded Depression | Discontinuation vs Continuation | Depression | Income_quantitative | 0.875 | 0.853 | 0.897 | <2.2e-16 | NA | 10166 | 7969 | 2197 |
| European | Recorded Depression | Discontinuation vs Continuation | Depression | Education_levelCollege Undergraduate | 1.265 | 1.116 | 1.434 | 2.39e-04 | NA | 10166 | 7969 | 2197 |
| European | Recorded Depression | Discontinuation vs Continuation | Depression | Education_levelLess than a high school degree | 2.469 | 1.824 | 3.342 | 4.91e-09 | NA | 10166 | 7969 | 2197 |
| European | Recorded Depression | Discontinuation vs Continuation | Depression | Education_levelTwelve or GED | 1.606 | 1.353 | 1.906 | 6.18e-08 | NA | 10166 | 7969 | 2197 |
| European | Recorded Depression | Discontinuation vs Continuation | Depression | Depression | 1.179 | 1.122 | 1.240 | 9.23E-11 | 6.46e-10 | 10166 | 7969 | 2197 |
| European | Recorded Depression | Switching vs Continuation | Depression | (Intercept) | 4.021 | 3.481 | 4.645 | <2.2e-16 | NA | 17576 | 7969 | 9607 |
| European | Recorded Depression | Switching vs Continuation | Depression | age_at_first_AD | 0.979 | 0.977 | 0.981 | <2.2e-16 | NA | 17576 | 7969 | 9607 |
| European | Recorded Depression | Switching vs Continuation | Depression | sex_at_birthMale | 1.120 | 1.047 | 1.197 | 9.77e-04 | NA | 17576 | 7969 | 9607 |
| European | Recorded Depression | Switching vs Continuation | Depression | Income_quantitative | 0.917 | 0.903 | 0.931 | <2.2e-16 | NA | 17576 | 7969 | 9607 |
| European | Recorded Depression | Switching vs Continuation | Depression | Education_levelCollege Undergraduate | 1.165 | 1.082 | 1.255 | 5.69e-05 | NA | 17576 | 7969 | 9607 |
| European | Recorded Depression | Switching vs Continuation | Depression | Education_levelLess than a high school degree | 1.382 | 1.091 | 1.750 | 7.35e-03 | NA | 17576 | 7969 | 9607 |
| European | Recorded Depression | Switching vs Continuation | Depression | Education_levelTwelve or GED | 1.415 | 1.266 | 1.583 | 1.13e-09 | NA | 17576 | 7969 | 9607 |
| European | Recorded Depression | Switching vs Continuation | Depression | Depression | 1.158 | 1.121 | 1.196 | <2.2e-16 | <2.2e-16 | 17576 | 7969 | 9607 |
| European | Recorded Depression | Discontinuation vs Continuation | Insomnia | (Intercept) | 0.268 | 0.213 | 0.338 | <2.2e-16 | NA | 10166 | 7969 | 2197 |
| European | Recorded Depression | Discontinuation vs Continuation | Insomnia | age_at_first_AD | 1.005 | 1.002 | 1.008 | 7.30e-04 | NA | 10166 | 7969 | 2197 |
| European | Recorded Depression | Discontinuation vs Continuation | Insomnia | sex_at_birthMale | 1.149 | 1.036 | 1.273 | 8.53e-03 | NA | 10166 | 7969 | 2197 |
| European | Recorded Depression | Discontinuation vs Continuation | Insomnia | Income_quantitative | 0.874 | 0.852 | 0.896 | <2.2e-16 | NA | 10166 | 7969 | 2197 |
| European | Recorded Depression | Discontinuation vs Continuation | Insomnia | Education_levelCollege Undergraduate | 1.269 | 1.119 | 1.438 | 1.96e-04 | NA | 10166 | 7969 | 2197 |
| European | Recorded Depression | Discontinuation vs Continuation | Insomnia | Education_levelLess than a high school degree | 2.479 | 1.831 | 3.356 | 4.22e-09 | NA | 10166 | 7969 | 2197 |
| European | Recorded Depression | Discontinuation vs Continuation | Insomnia | Education_levelTwelve or GED | 1.611 | 1.357 | 1.913 | 4.93e-08 | NA | 10166 | 7969 | 2197 |
| European | Recorded Depression | Discontinuation vs Continuation | Insomnia | Insomnia | 1.105 | 1.050 | 1.163 | 1.13e-04 | 7.93e-04 | 10166 | 7969 | 2197 |
| European | Recorded Depression | Switching vs Continuation | Insomnia | (Intercept) | 4.012 | 3.474 | 4.634 | <2.2e-16 | NA | 17576 | 7969 | 9607 |
| European | Recorded Depression | Switching vs Continuation | Insomnia | age_at_first_AD | 0.979 | 0.977 | 0.981 | <2.2e-16 | NA | 17576 | 7969 | 9607 |
| European | Recorded Depression | Switching vs Continuation | Insomnia | sex_at_birthMale | 1.125 | 1.052 | 1.203 | 5.82e-04 | NA | 17576 | 7969 | 9607 |
| European | Recorded Depression | Switching vs Continuation | Insomnia | Income_quantitative | 0.915 | 0.901 | 0.930 | <2.2e-16 | NA | 17576 | 7969 | 9607 |
| European | Recorded Depression | Switching vs Continuation | Insomnia | Education_levelCollege Undergraduate | 1.167 | 1.084 | 1.257 | 4.50e-05 | NA | 17576 | 7969 | 9607 |
| European | Recorded Depression | Switching vs Continuation | Insomnia | Education_levelLess than a high school degree | 1.418 | 1.119 | 1.795 | 3.77e-03 | NA | 17576 | 7969 | 9607 |
| European | Recorded Depression | Switching vs Continuation | Insomnia | Education_levelTwelve or GED | 1.415 | 1.266 | 1.583 | 1.08e-09 | NA | 17576 | 7969 | 9607 |
| European | Recorded Depression | Switching vs Continuation | Insomnia | Insomnia | 1.074 | 1.039 | 1.109 | 1.66e-05 | 1.16e-04 | 17576 | 7969 | 9607 |
| European | Recorded Depression | Discontinuation vs Continuation | Migraine | (Intercept) | 0.271 | 0.215 | 0.341 | <2.2e-16 | NA | 10166 | 7969 | 2197 |
| European | Recorded Depression | Discontinuation vs Continuation | Migraine | age_at_first_AD | 1.005 | 1.002 | 1.008 | 9.88e-04 | NA | 10166 | 7969 | 2197 |
| European | Recorded Depression | Discontinuation vs Continuation | Migraine | sex_at_birthMale | 1.149 | 1.037 | 1.274 | 8.11e-03 | NA | 10166 | 7969 | 2197 |
| European | Recorded Depression | Discontinuation vs Continuation | Migraine | Income_quantitative | 0.871 | 0.850 | 0.893 | <2.2e-16 | NA | 10166 | 7969 | 2197 |
| European | Recorded Depression | Discontinuation vs Continuation | Migraine | Education_levelCollege Undergraduate | 1.277 | 1.127 | 1.447 | 1.28e-04 | NA | 10166 | 7969 | 2197 |
| European | Recorded Depression | Discontinuation vs Continuation | Migraine | Education_levelLess than a high school degree | 2.585 | 1.912 | 3.496 | 6.84e-10 | NA | 10166 | 7969 | 2197 |
| European | Recorded Depression | Discontinuation vs Continuation | Migraine | Education_levelTwelve or GED | 1.639 | 1.382 | 1.945 | 1.48e-08 | NA | 10166 | 7969 | 2197 |
| European | Recorded Depression | Discontinuation vs Continuation | Migraine | Migraine | 1.017 | 0.969 | 1.066 | 4.98e-01 | 1.00e+00 | 10166 | 7969 | 2197 |
| European | Recorded Depression | Switching vs Continuation | Migraine | (Intercept) | 4.046 | 3.504 | 4.673 | <2.2e-16 | NA | 17576 | 7969 | 9607 |
| European | Recorded Depression | Switching vs Continuation | Migraine | age_at_first_AD | 0.979 | 0.977 | 0.981 | <2.2e-16 | NA | 17576 | 7969 | 9607 |
| European | Recorded Depression | Switching vs Continuation | Migraine | sex_at_birthMale | 1.127 | 1.054 | 1.205 | 4.82e-04 | NA | 17576 | 7969 | 9607 |
| European | Recorded Depression | Switching vs Continuation | Migraine | Income_quantitative | 0.913 | 0.899 | 0.928 | <2.2e-16 | NA | 17576 | 7969 | 9607 |
| European | Recorded Depression | Switching vs Continuation | Migraine | Education_levelCollege Undergraduate | 1.172 | 1.088 | 1.262 | 2.93e-05 | NA | 17576 | 7969 | 9607 |
| European | Recorded Depression | Switching vs Continuation | Migraine | Education_levelLess than a high school degree | 1.459 | 1.152 | 1.847 | 1.70e-03 | NA | 17576 | 7969 | 9607 |
| European | Recorded Depression | Switching vs Continuation | Migraine | Education_levelTwelve or GED | 1.428 | 1.277 | 1.597 | 3.89e-10 | NA | 17576 | 7969 | 9607 |
| European | Recorded Depression | Switching vs Continuation | Migraine | Migraine | 1.061 | 1.029 | 1.094 | 1.52e-04 | 1.06e-03 | 17576 | 7969 | 9607 |
| European | Recorded Depression | Discontinuation vs Continuation | Schizophrenia | (Intercept) | 0.271 | 0.215 | 0.341 | <2.2e-16 | NA | 10166 | 7969 | 2197 |
| European | Recorded Depression | Discontinuation vs Continuation | Schizophrenia | age_at_first_AD | 1.005 | 1.002 | 1.008 | 9.46e-04 | NA | 10166 | 7969 | 2197 |
| European | Recorded Depression | Discontinuation vs Continuation | Schizophrenia | sex_at_birthMale | 1.149 | 1.037 | 1.274 | 8.09e-03 | NA | 10166 | 7969 | 2197 |
| European | Recorded Depression | Discontinuation vs Continuation | Schizophrenia | Income_quantitative | 0.871 | 0.850 | 0.893 | <2.2e-16 | NA | 10166 | 7969 | 2197 |
| European | Recorded Depression | Discontinuation vs Continuation | Schizophrenia | Education_levelCollege Undergraduate | 1.278 | 1.127 | 1.448 | 1.23e-04 | NA | 10166 | 7969 | 2197 |
| European | Recorded Depression | Discontinuation vs Continuation | Schizophrenia | Education_levelLess than a high school degree | 2.564 | 1.896 | 3.467 | 9.87e-10 | NA | 10166 | 7969 | 2197 |
| European | Recorded Depression | Discontinuation vs Continuation | Schizophrenia | Education_levelTwelve or GED | 1.641 | 1.383 | 1.947 | 1.40e-08 | NA | 10166 | 7969 | 2197 |
| European | Recorded Depression | Discontinuation vs Continuation | Schizophrenia | Schizophrenia | 1.047 | 0.998 | 1.099 | 6.32e-02 | 4.43e-01 | 10166 | 7969 | 2197 |

| | | | | | | | | | | | | |
|---|---|---|---|---|---|---|---|---|---|---|---|---|
| European | Recorded Depression | Switching vs Continuation | Schizophrenia | (Intercept) | 4.051 | 3.508 | 4.679 | <2.2e-16 | NA | 17576 | 7969 | 9607 |
| European | Recorded Depression | Switching vs Continuation | Schizophrenia | age_at_first_AD | 0.979 | 0.977 | 0.981 | <2.2e-16 | NA | 17576 | 7969 | 9607 |
| European | Recorded Depression | Switching vs Continuation | Schizophrenia | sex_at_birthMale | 1.127 | 1.054 | 1.205 | 4.65e-04 | NA | 17576 | 7969 | 9607 |
| European | Recorded Depression | Switching vs Continuation | Schizophrenia | Income_quantitative | 0.913 | 0.899 | 0.927 | <2.2e-16 | NA | 17576 | 7969 | 9607 |
| European | Recorded Depression | Switching vs Continuation | Schizophrenia | Education_levelCollege Undergraduate | 1.173 | 1.089 | 1.263 | 2.58e-05 | NA | 17576 | 7969 | 9607 |
| European | Recorded Depression | Switching vs Continuation | Schizophrenia | Education_levelLess than a high school degree | 1.456 | 1.150 | 1.843 | 1.80e-03 | NA | 17576 | 7969 | 9607 |
| European | Recorded Depression | Switching vs Continuation | Schizophrenia | Education_levelTwelve or GED | 1.429 | 1.278 | 1.597 | 3.58e-10 | NA | 17576 | 7969 | 9607 |
| European | Recorded Depression | Switching vs Continuation | Schizophrenia | Schizophrenia | 1.009 | 0.978 | 1.041 | 5.71e-01 | 1.00e+00 | 17576 | 7969 | 9607 |
| European | No Recorded Depression | Discontinuation vs Continuation | ADHD | (Intercept) | 0.408 | 0.350 | 0.475 | <2.2e-16 | NA | 18562 | 13287 | 5275 |
| European | No Recorded Depression | Discontinuation vs Continuation | ADHD | age_at_first_AD | 1.004 | 1.002 | 1.007 | 1.52e-05 | NA | 18562 | 13287 | 5275 |
| European | No Recorded Depression | Discontinuation vs Continuation | ADHD | sex_at_birthMale | 1.118 | 1.043 | 1.198 | 1.54e-03 | NA | 18562 | 13287 | 5275 |
| European | No Recorded Depression | Discontinuation vs Continuation | ADHD | Income_quantitative | 0.925 | 0.910 | 0.940 | <2.2e-16 | NA | 18562 | 13287 | 5275 |
| European | No Recorded Depression | Discontinuation vs Continuation | ADHD | Education_levelCollege Undergraduate | 1.049 | 0.972 | 1.132 | 2.20e-01 | NA | 18562 | 13287 | 5275 |
| European | No Recorded Depression | Discontinuation vs Continuation | ADHD | Education_levelLess than a high school degree | 1.403 | 1.075 | 1.832 | 1.28e-02 | NA | 18562 | 13287 | 5275 |
| European | No Recorded Depression | Discontinuation vs Continuation | ADHD | Education_levelTwelve or GED | 1.227 | 1.091 | 1.381 | 6.49e-04 | NA | 18562 | 13287 | 5275 |
| European | No Recorded Depression | Discontinuation vs Continuation | ADHD | ADHD | 1.038 | 1.004 | 1.073 | 2.72e-02 | 1.90e-01 | 18562 | 13287 | 5275 |
| European | No Recorded Depression | Switching vs Continuation | ADHD | (Intercept) | 1.197 | 1.019 | 1.406 | 2.84e-02 | NA | 17516 | 13287 | 4229 |
| European | No Recorded Depression | Switching vs Continuation | ADHD | age_at_first_AD | 0.981 | 0.979 | 0.983 | <2.2e-16 | NA | 17516 | 13287 | 4229 |
| European | No Recorded Depression | Switching vs Continuation | ADHD | sex_at_birthMale | 0.907 | 0.837 | 0.981 | 1.54e-02 | NA | 17516 | 13287 | 4229 |
| European | No Recorded Depression | Switching vs Continuation | ADHD | Income_quantitative | 0.917 | 0.901 | 0.934 | <2.2e-16 | NA | 17516 | 13287 | 4229 |
| European | No Recorded Depression | Switching vs Continuation | ADHD | Education_levelCollege Undergraduate | 1.036 | 0.953 | 1.127 | 4.08e-01 | NA | 17516 | 13287 | 4229 |
| European | No Recorded Depression | Switching vs Continuation | ADHD | Education_levelLess than a high school degree | 1.182 | 0.873 | 1.600 | 2.79e-01 | NA | 17516 | 13287 | 4229 |
| European | No Recorded Depression | Switching vs Continuation | ADHD | Education_levelTwelve or GED | 1.179 | 1.035 | 1.344 | 1.34e-02 | NA | 17516 | 13287 | 4229 |
| European | No Recorded Depression | Switching vs Continuation | ADHD | ADHD | 1.052 | 1.015 | 1.091 | 5.86e-03 | 4.11e-02 | 17516 | 13287 | 4229 |
| European | No Recorded Depression | Discontinuation vs Continuation | BMI | (Intercept) | 0.410 | 0.352 | 0.478 | <2.2e-16 | NA | 18562 | 13287 | 5275 |
| European | No Recorded Depression | Discontinuation vs Continuation | BMI | age_at_first_AD | 1.004 | 1.002 | 1.006 | 2.23e-05 | NA | 18562 | 13287 | 5275 |
| European | No Recorded Depression | Discontinuation vs Continuation | BMI | sex_at_birthMale | 1.117 | 1.042 | 1.196 | 1.69e-03 | NA | 18562 | 13287 | 5275 |
| European | No Recorded Depression | Discontinuation vs Continuation | BMI | Income_quantitative | 0.924 | 0.909 | 0.939 | <2.2e-16 | NA | 18562 | 13287 | 5275 |
| European | No Recorded Depression | Discontinuation vs Continuation | BMI | Education_levelCollege Undergraduate | 1.055 | 0.978 | 1.139 | 1.69e-01 | NA | 18562 | 13287 | 5275 |
| European | No Recorded Depression | Discontinuation vs Continuation | BMI | Education_levelLess than a high school degree | 1.433 | 1.098 | 1.870 | 8.06e-03 | NA | 18562 | 13287 | 5275 |
| European | No Recorded Depression | Discontinuation vs Continuation | BMI | Education_levelTwelve or GED | 1.240 | 1.102 | 1.394 | 3.39e-04 | NA | 18562 | 13287 | 5275 |
| European | No Recorded Depression | Discontinuation vs Continuation | BMI | BMI | 0.999 | 0.967 | 1.032 | 9.59e-01 | 1.00e+00 | 18562 | 13287 | 5275 |
| European | No Recorded Depression | Switching vs Continuation | BMI | (Intercept) | 1.195 | 1.017 | 1.404 | 3.03e-02 | NA | 17516 | 13287 | 4229 |
| European | No Recorded Depression | Switching vs Continuation | BMI | age_at_first_AD | 0.981 | 0.979 | 0.983 | <2.2e-16 | NA | 17516 | 13287 | 4229 |
| European | No Recorded Depression | Switching vs Continuation | BMI | sex_at_birthMale | 0.905 | 0.836 | 0.979 | 1.31e-02 | NA | 17516 | 13287 | 4229 |
| European | No Recorded Depression | Switching vs Continuation | BMI | Income_quantitative | 0.918 | 0.902 | 0.934 | <2.2e-16 | NA | 17516 | 13287 | 4229 |
| European | No Recorded Depression | Switching vs Continuation | BMI | Education_levelCollege Undergraduate | 1.035 | 0.952 | 1.126 | 4.20e-01 | NA | 17516 | 13287 | 4229 |
| European | No Recorded Depression | Switching vs Continuation | BMI | Education_levelLess than a high school degree | 1.199 | 0.886 | 1.622 | 2.40e-01 | NA | 17516 | 13287 | 4229 |
| European | No Recorded Depression | Switching vs Continuation | BMI | Education_levelTwelve or GED | 1.179 | 1.034 | 1.343 | 1.36e-02 | NA | 17516 | 13287 | 4229 |
| European | No Recorded Depression | Switching vs Continuation | BMI | BMI | 1.063 | 1.026 | 1.102 | 7.29e-04 | 5.10e-03 | 17516 | 13287 | 4229 |
| European | No Recorded Depression | Discontinuation vs Continuation | Bipolar Disorder | (Intercept) | 0.409 | 0.351 | 0.477 | <2.2e-16 | NA | 18562 | 13287 | 5275 |
| European | No Recorded Depression | Discontinuation vs Continuation | Bipolar Disorder | age_at_first_AD | 1.004 | 1.002 | 1.006 | 2.50e-05 | NA | 18562 | 13287 | 5275 |
| European | No Recorded Depression | Discontinuation vs Continuation | Bipolar Disorder | sex_at_birthMale | 1.115 | 1.041 | 1.195 | 1.92e-03 | NA | 18562 | 13287 | 5275 |
| European | No Recorded Depression | Discontinuation vs Continuation | Bipolar Disorder | Income_quantitative | 0.924 | 0.910 | 0.939 | <2.2e-16 | NA | 18562 | 13287 | 5275 |
| European | No Recorded Depression | Discontinuation vs Continuation | Bipolar Disorder | Education_levelCollege Undergraduate | 1.058 | 0.981 | 1.142 | 1.46e-01 | NA | 18562 | 13287 | 5275 |
| European | No Recorded Depression | Discontinuation vs Continuation | Bipolar Disorder | Education_levelLess than a high school degree | 1.438 | 1.102 | 1.876 | 7.49e-03 | NA | 18562 | 13287 | 5275 |
| European | No Recorded Depression | Discontinuation vs Continuation | Bipolar Disorder | Education_levelTwelve or GED | 1.248 | 1.110 | 1.403 | 2.21e-04 | NA | 18562 | 13287 | 5275 |
| European | No Recorded Depression | Discontinuation vs Continuation | Bipolar Disorder | Bipolar Disorder | 1.049 | 1.014 | 1.085 | 6.30e-03 | 4.41e-02 | 18562 | 13287 | 5275 |
| European | No Recorded Depression | Switching vs Continuation | Bipolar Disorder | (Intercept) | 1.204 | 1.025 | 1.414 | 2.39e-02 | NA | 17516 | 13287 | 4229 |
| European | No Recorded Depression | Switching vs Continuation | Bipolar Disorder | age_at_first_AD | 0.981 | 0.979 | 0.983 | <2.2e-16 | NA | 17516 | 13287 | 4229 |
| European | No Recorded Depression | Switching vs Continuation | Bipolar Disorder | sex_at_birthMale | 0.903 | 0.834 | 0.977 | 1.15e-02 | NA | 17516 | 13287 | 4229 |
| European | No Recorded Depression | Switching vs Continuation | Bipolar Disorder | Income_quantitative | 0.916 | 0.900 | 0.933 | <2.2e-16 | NA | 17516 | 13287 | 4229 |
| European | No Recorded Depression | Switching vs Continuation | Bipolar Disorder | Education_levelCollege Undergraduate | 1.046 | 0.962 | 1.138 | 2.91e-01 | NA | 17516 | 13287 | 4229 |
| European | No Recorded Depression | Switching vs Continuation | Bipolar Disorder | Education_levelLess than a high school degree | 1.220 | 0.902 | 1.651 | 1.97e-01 | NA | 17516 | 13287 | 4229 |
| European | No Recorded Depression | Switching vs Continuation | Bipolar Disorder | Education_levelTwelve or GED | 1.204 | 1.056 | 1.371 | 5.38e-03 | NA | 17516 | 13287 | 4229 |
| European | No Recorded Depression | Switching vs Continuation | Bipolar Disorder | Bipolar Disorder | 1.052 | 1.013 | 1.092 | 7.93e-03 | 5.55e-02 | 17516 | 13287 | 4229 |
| European | No Recorded Depression | Discontinuation vs Continuation | Depression | (Intercept) | 0.404 | 0.346 | 0.470 | <2.2e-16 | NA | 18562 | 13287 | 5275 |

| | | | | | | | | | | | | |
|---|---|---|---|---|---|---|---|---|---|---|---|---|
| European | No Recorded Depression | Discontinuation vs Continuation | Depression | age_at_first_AD | 1.005 | 1.003 | 1.007 | 8.99e-06 | NA | 18562 | 13287 | 5275 |
| European | No Recorded Depression | Discontinuation vs Continuation | Depression | sex_at_birthMale | 1.113 | 1.039 | 1.193 | 2.31e-03 | NA | 18562 | 13287 | 5275 |
| European | No Recorded Depression | Discontinuation vs Continuation | Depression | Income_quantitative | 0.926 | 0.911 | 0.941 | <2.2e-16 | NA | 18562 | 13287 | 5275 |
| European | No Recorded Depression | Discontinuation vs Continuation | Depression | Education_levelCollege Undergraduate | 1.050 | 0.973 | 1.133 | 2.11e-01 | NA | 18562 | 13287 | 5275 |
| European | No Recorded Depression | Discontinuation vs Continuation | Depression | Education_levelLess than a high school degree | 1.388 | 1.063 | 1.812 | 1.60e-02 | NA | 18562 | 13287 | 5275 |
| European | No Recorded Depression | Discontinuation vs Continuation | Depression | Education_levelTwelve or GED | 1.233 | 1.097 | 1.387 | 4.69e-04 | NA | 18562 | 13287 | 5275 |
| European | No Recorded Depression | Discontinuation vs Continuation | Depression | Depression | 1.084 | 1.049 | 1.121 | 2.14e-06 | 1.49e-05 | 18562 | 13287 | 5275 |
| European | No Recorded Depression | Switching vs Continuation | Depression | (Intercept) | 1.164 | 0.991 | 1.369 | 6.49e-02 | NA | 17516 | 13287 | 4229 |
| European | No Recorded Depression | Switching vs Continuation | Depression | age_at_first_AD | 0.981 | 0.979 | 0.983 | <2.2e-16 | NA | 17516 | 13287 | 4229 |
| European | No Recorded Depression | Switching vs Continuation | Depression | sex_at_birthMale | 0.898 | 0.829 | 0.972 | 7.85e-03 | NA | 17516 | 13287 | 4229 |
| European | No Recorded Depression | Switching vs Continuation | Depression | Income_quantitative | 0.920 | 0.904 | 0.937 | <2.2e-16 | NA | 17516 | 13287 | 4229 |
| European | No Recorded Depression | Switching vs Continuation | Depression | Education_levelCollege Undergraduate | 1.033 | 0.950 | 1.124 | 4.49e-01 | NA | 17516 | 13287 | 4229 |
| European | No Recorded Depression | Switching vs Continuation | Depression | Education_levelLess than a high school degree | 1.145 | 0.845 | 1.552 | 3.81e-01 | NA | 17516 | 13287 | 4229 |
| European | No Recorded Depression | Switching vs Continuation | Depression | Education_levelTwelve or GED | 1.180 | 1.035 | 1.344 | 1.33e-02 | NA | 17516 | 13287 | 4229 |
| European | No Recorded Depression | Switching vs Continuation | Depression | Depression | 1.188 | 1.145 | 1.233 | <2.2e-16 | <2.2e-16 | 17516 | 13287 | 4229 |
| European | No Recorded Depression | Discontinuation vs Continuation | Insomnia | (Intercept) | 0.404 | 0.347 | 0.471 | <2.2e-16 | NA | 18562 | 13287 | 5275 |
| European | No Recorded Depression | Discontinuation vs Continuation | Insomnia | age_at_first_AD | 1.005 | 1.003 | 1.007 | 1.09e-05 | NA | 18562 | 13287 | 5275 |
| European | No Recorded Depression | Discontinuation vs Continuation | Insomnia | sex_at_birthMale | 1.117 | 1.042 | 1.196 | 1.71e-03 | NA | 18562 | 13287 | 5275 |
| European | No Recorded Depression | Discontinuation vs Continuation | Insomnia | Income_quantitative | 0.926 | 0.911 | 0.941 | <2.2e-16 | NA | 18562 | 13287 | 5275 |
| European | No Recorded Depression | Discontinuation vs Continuation | Insomnia | Education_levelCollege Undergraduate | 1.048 | 0.971 | 1.131 | 2.33e-01 | NA | 18562 | 13287 | 5275 |
| European | No Recorded Depression | Discontinuation vs Continuation | Insomnia | Education_levelLess than a high school degree | 1.397 | 1.070 | 1.823 | 1.39e-02 | NA | 18562 | 13287 | 5275 |
| European | No Recorded Depression | Discontinuation vs Continuation | Insomnia | Education_levelTwelve or GED | 1.228 | 1.092 | 1.381 | 6.10e-04 | NA | 18562 | 13287 | 5275 |
| European | No Recorded Depression | Discontinuation vs Continuation | Insomnia | Insomnia | 1.083 | 1.047 | 1.120 | 3.44e-06 | 2.40e-05 | 18562 | 13287 | 5275 |
| European | No Recorded Depression | Switching vs Continuation | Insomnia | (Intercept) | 1.186 | 1.010 | 1.394 | 3.77e-02 | NA | 17516 | 13287 | 4229 |
| European | No Recorded Depression | Switching vs Continuation | Insomnia | age_at_first_AD | 0.981 | 0.979 | 0.983 | <2.2e-16 | NA | 17516 | 13287 | 4229 |
| European | No Recorded Depression | Switching vs Continuation | Insomnia | sex_at_birthMale | 0.904 | 0.835 | 0.979 | 1.28e-02 | NA | 17516 | 13287 | 4229 |
| European | No Recorded Depression | Switching vs Continuation | Insomnia | Income_quantitative | 0.919 | 0.903 | 0.935 | <2.2e-16 | NA | 17516 | 13287 | 4229 |
| European | No Recorded Depression | Switching vs Continuation | Insomnia | Education_levelCollege Undergraduate | 1.034 | 0.951 | 1.125 | 4.33e-01 | NA | 17516 | 13287 | 4229 |
| European | No Recorded Depression | Switching vs Continuation | Insomnia | Education_levelLess than a high school degree | 1.169 | 0.863 | 1.583 | 3.13e-01 | NA | 17516 | 13287 | 4229 |
| European | No Recorded Depression | Switching vs Continuation | Insomnia | Education_levelTwelve or GED | 1.178 | 1.034 | 1.343 | 1.39e-02 | NA | 17516 | 13287 | 4229 |
| European | No Recorded Depression | Switching vs Continuation | Insomnia | Insomnia | 1.127 | 1.086 | 1.169 | 2.58e-10 | 1.81e-09 | 17516 | 13287 | 4229 |
| European | No Recorded Depression | Discontinuation vs Continuation | Migraine | (Intercept) | 0.410 | 0.352 | 0.478 | <2.2e-16 | NA | 18562 | 13287 | 5275 |
| European | No Recorded Depression | Discontinuation vs Continuation | Migraine | age_at_first_AD | 1.004 | 1.002 | 1.006 | 2.16e-05 | NA | 18562 | 13287 | 5275 |
| European | No Recorded Depression | Discontinuation vs Continuation | Migraine | sex_at_birthMale | 1.117 | 1.042 | 1.196 | 1.70e-03 | NA | 18562 | 13287 | 5275 |
| European | No Recorded Depression | Discontinuation vs Continuation | Migraine | Income_quantitative | 0.924 | 0.909 | 0.939 | <2.2e-16 | NA | 18562 | 13287 | 5275 |
| European | No Recorded Depression | Discontinuation vs Continuation | Migraine | Education_levelCollege Undergraduate | 1.055 | 0.977 | 1.138 | 1.71e-01 | NA | 18562 | 13287 | 5275 |
| European | No Recorded Depression | Discontinuation vs Continuation | Migraine | Education_levelLess than a high school degree | 1.433 | 1.098 | 1.869 | 8.07e-03 | NA | 18562 | 13287 | 5275 |
| European | No Recorded Depression | Discontinuation vs Continuation | Migraine | Education_levelTwelve or GED | 1.239 | 1.102 | 1.393 | 3.44e-04 | NA | 18562 | 13287 | 5275 |
| European | No Recorded Depression | Discontinuation vs Continuation | Migraine | Migraine | 1.004 | 0.972 | 1.037 | 8.03e-01 | 1.00e+00 | 18562 | 13287 | 5275 |
| European | No Recorded Depression | Switching vs Continuation | Migraine | (Intercept) | 1.203 | 1.024 | 1.413 | 2.42e-02 | NA | 17516 | 13287 | 4229 |
| European | No Recorded Depression | Switching vs Continuation | Migraine | age_at_first_AD | 0.981 | 0.979 | 0.983 | <2.2e-16 | NA | 17516 | 13287 | 4229 |
| European | No Recorded Depression | Switching vs Continuation | Migraine | sex_at_birthMale | 0.904 | 0.835 | 0.978 | 1.24e-02 | NA | 17516 | 13287 | 4229 |
| European | No Recorded Depression | Switching vs Continuation | Migraine | Income_quantitative | 0.916 | 0.900 | 0.932 | <2.2e-16 | NA | 17516 | 13287 | 4229 |
| European | No Recorded Depression | Switching vs Continuation | Migraine | Education_levelCollege Undergraduate | 1.043 | 0.959 | 1.134 | 3.30e-01 | NA | 17516 | 13287 | 4229 |
| European | No Recorded Depression | Switching vs Continuation | Migraine | Education_levelLess than a high school degree | 1.211 | 0.895 | 1.638 | 2.14e-01 | NA | 17516 | 13287 | 4229 |
| European | No Recorded Depression | Switching vs Continuation | Migraine | Education_levelTwelve or GED | 1.193 | 1.047 | 1.359 | 8.04e-03 | NA | 17516 | 13287 | 4229 |
| European | No Recorded Depression | Switching vs Continuation | Migraine | Migraine | 1.034 | 0.999 | 1.071 | 6.07e-02 | 4.25e-01 | 17516 | 13287 | 4229 |
| European | No Recorded Depression | Discontinuation vs Continuation | Schizophrenia | (Intercept) | 0.408 | 0.350 | 0.476 | <2.2e-16 | NA | 18562 | 13287 | 5275 |
| European | No Recorded Depression | Discontinuation vs Continuation | Schizophrenia | age_at_first_AD | 1.004 | 1.002 | 1.006 | 1.98e-05 | NA | 18562 | 13287 | 5275 |
| European | No Recorded Depression | Discontinuation vs Continuation | Schizophrenia | sex_at_birthMale | 1.116 | 1.042 | 1.196 | 1.79e-03 | NA | 18562 | 13287 | 5275 |
| European | No Recorded Depression | Discontinuation vs Continuation | Schizophrenia | Income_quantitative | 0.925 | 0.910 | 0.940 | <2.2e-16 | NA | 18562 | 13287 | 5275 |
| European | No Recorded Depression | Discontinuation vs Continuation | Schizophrenia | Education_levelCollege Undergraduate | 1.056 | 0.979 | 1.140 | 1.60e-01 | NA | 18562 | 13287 | 5275 |
| European | No Recorded Depression | Discontinuation vs Continuation | Schizophrenia | Education_levelLess than a high school degree | 1.429 | 1.095 | 1.864 | 8.59e-03 | NA | 18562 | 13287 | 5275 |
| European | No Recorded Depression | Discontinuation vs Continuation | Schizophrenia | Education_levelTwelve or GED | 1.243 | 1.105 | 1.397 | 2.88e-04 | NA | 18562 | 13287 | 5275 |
| European | No Recorded Depression | Discontinuation vs Continuation | Schizophrenia | Schizophrenia | 1.029 | 0.996 | 1.063 | 8.57e-02 | 6.00e-01 | 18562 | 13287 | 5275 |
| European | No Recorded Depression | Switching vs Continuation | Schizophrenia | (Intercept) | 1.203 | 1.024 | 1.413 | 2.43e-02 | NA | 17516 | 13287 | 4229 |
| European | No Recorded Depression | Switching vs Continuation | Schizophrenia | age_at_first_AD | 0.981 | 0.979 | 0.983 | <2.2e-16 | NA | 17516 | 13287 | 4229 |

| | | | | | | | | | | | | |
|---|---|---|---|---|---|---|---|---|---|---|---|---|
| European | No Recorded Depression | Switching vs Continuation | Schizophrenia | sex_at_birthMale | 0.904 | 0.835 | 0.979 | 1.28e-02 | NA | 17516 | 13287 | 4229 |
| European | No Recorded Depression | Switching vs Continuation | Schizophrenia | Income_quantitative | 0.916 | 0.900 | 0.932 | <2.2e-16 | NA | 17516 | 13287 | 4229 |
| European | No Recorded Depression | Switching vs Continuation | Schizophrenia | Education_levelCollege Undergraduate | 1.044 | 0.960 | 1.136 | 3.11e-01 | NA | 17516 | 13287 | 4229 |
| European | No Recorded Depression | Switching vs Continuation | Schizophrenia | Education_levelLess than a high school degree | 1.216 | 0.899 | 1.645 | 2.04e-01 | NA | 17516 | 13287 | 4229 |
| European | No Recorded Depression | Switching vs Continuation | Schizophrenia | Education_levelTwelve or GED | 1.197 | 1.050 | 1.363 | 6.96e-03 | NA | 17516 | 13287 | 4229 |
| European | No Recorded Depression | Switching vs Continuation | Schizophrenia | Schizophrenia | 1.013 | 0.978 | 1.050 | 4.77e-01 | 1.00e+00 | 17516 | 13287 | 4229 |
| European | Recorded Anxiety | Discontinuation vs Continuation | ADHD | (Intercept) | 0.323 | 0.270 | 0.387 | <2.2e-16 | NA | 15204 | 11798 | 3406 |
| European | Recorded Anxiety | Discontinuation vs Continuation | ADHD | age_at_first_AD | 1.005 | 1.003 | 1.007 | 4.29e-05 | NA | 15204 | 11798 | 3406 |
| European | Recorded Anxiety | Discontinuation vs Continuation | ADHD | sex_at_birthMale | 1.226 | 1.127 | 1.334 | 2.05e-06 | NA | 15204 | 11798 | 3406 |
| European | Recorded Anxiety | Discontinuation vs Continuation | ADHD | Income_quantitative | 0.880 | 0.863 | 0.898 | <2.2e-16 | NA | 15204 | 11798 | 3406 |
| European | Recorded Anxiety | Discontinuation vs Continuation | ADHD | Education_levelCollege Undergraduate | 1.088 | 0.987 | 1.198 | 8.94e-02 | NA | 15204 | 11798 | 3406 |
| European | Recorded Anxiety | Discontinuation vs Continuation | ADHD | Education_levelLess than a high school degree | 1.720 | 1.324 | 2.233 | 4.74e-05 | NA | 15204 | 11798 | 3406 |
| European | Recorded Anxiety | Discontinuation vs Continuation | ADHD | Education_levelTwelve or GED | 1.395 | 1.215 | 1.602 | 2.31e-06 | NA | 15204 | 11798 | 3406 |
| European | Recorded Anxiety | Discontinuation vs Continuation | ADHD | ADHD | 1.048 | 1.008 | 1.091 | 1.97e-02 | 1.38e-01 | 15204 | 11798 | 3406 |
| European | Recorded Anxiety | Switching vs Continuation | ADHD | (Intercept) | 3.057 | 2.702 | 3.458 | <2.2e-16 | NA | 22696 | 11798 | 10898 |
| European | Recorded Anxiety | Switching vs Continuation | ADHD | age_at_first_AD | 0.982 | 0.981 | 0.984 | <2.2e-16 | NA | 22696 | 11798 | 10898 |
| European | Recorded Anxiety | Switching vs Continuation | ADHD | sex_at_birthMale | 1.162 | 1.094 | 1.234 | 1.02e-06 | NA | 22696 | 11798 | 10898 |
| European | Recorded Anxiety | Switching vs Continuation | ADHD | Income_quantitative | 0.884 | 0.872 | 0.896 | <2.2e-16 | NA | 22696 | 11798 | 10898 |
| European | Recorded Anxiety | Switching vs Continuation | ADHD | Education_levelCollege Undergraduate | 1.104 | 1.034 | 1.178 | 3.00e-03 | NA | 22696 | 11798 | 10898 |
| European | Recorded Anxiety | Switching vs Continuation | ADHD | Education_levelLess than a high school degree | 1.218 | 0.988 | 1.502 | 6.47e-02 | NA | 22696 | 11798 | 10898 |
| European | Recorded Anxiety | Switching vs Continuation | ADHD | Education_levelTwelve or GED | 1.388 | 1.258 | 1.531 | 6.97e-11 | NA | 22696 | 11798 | 10898 |
| European | Recorded Anxiety | Switching vs Continuation | ADHD | ADHD | 1.052 | 1.023 | 1.081 | 3.59e-04 | 2.51e-03 | 22696 | 11798 | 10898 |
| European | Recorded Anxiety | Discontinuation vs Continuation | BMI | (Intercept) | 0.325 | 0.272 | 0.389 | <2.2e-16 | NA | 15204 | 11798 | 3406 |
| European | Recorded Anxiety | Discontinuation vs Continuation | BMI | age_at_first_AD | 1.005 | 1.003 | 1.007 | 5.96e-05 | NA | 15204 | 11798 | 3406 |
| European | Recorded Anxiety | Discontinuation vs Continuation | BMI | sex_at_birthMale | 1.224 | 1.125 | 1.331 | 2.60e-06 | NA | 15204 | 11798 | 3406 |
| European | Recorded Anxiety | Discontinuation vs Continuation | BMI | Income_quantitative | 0.878 | 0.861 | 0.895 | <2.2e-16 | NA | 15204 | 11798 | 3406 |
| European | Recorded Anxiety | Discontinuation vs Continuation | BMI | Education_levelCollege Undergraduate | 1.099 | 0.998 | 1.211 | 5.57e-02 | NA | 15204 | 11798 | 3406 |
| European | Recorded Anxiety | Discontinuation vs Continuation | BMI | Education_levelLess than a high school degree | 1.772 | 1.365 | 2.300 | 1.71e-05 | NA | 15204 | 11798 | 3406 |
| European | Recorded Anxiety | Discontinuation vs Continuation | BMI | Education_levelTwelve or GED | 1.420 | 1.237 | 1.630 | 6.56e-07 | NA | 15204 | 11798 | 3406 |
| European | Recorded Anxiety | Discontinuation vs Continuation | BMI | BMI | 0.982 | 0.944 | 1.021 | 3.55e-01 | 1.00e+00 | 15204 | 11798 | 3406 |
| European | Recorded Anxiety | Switching vs Continuation | BMI | (Intercept) | 3.046 | 2.693 | 3.446 | <2.2e-16 | NA | 22696 | 11798 | 10898 |
| European | Recorded Anxiety | Switching vs Continuation | BMI | age_at_first_AD | 0.982 | 0.981 | 0.984 | <2.2e-16 | NA | 22696 | 11798 | 10898 |
| European | Recorded Anxiety | Switching vs Continuation | BMI | sex_at_birthMale | 1.160 | 1.092 | 1.232 | 1.29e-06 | NA | 22696 | 11798 | 10898 |
| European | Recorded Anxiety | Switching vs Continuation | BMI | Income_quantitative | 0.884 | 0.872 | 0.896 | <2.2e-16 | NA | 22696 | 11798 | 10898 |
| European | Recorded Anxiety | Switching vs Continuation | BMI | Education_levelCollege Undergraduate | 1.105 | 1.035 | 1.180 | 2.66e-03 | NA | 22696 | 11798 | 10898 |
| European | Recorded Anxiety | Switching vs Continuation | BMI | Education_levelLess than a high school degree | 1.230 | 0.998 | 1.516 | 5.21e-02 | NA | 22696 | 11798 | 10898 |
| European | Recorded Anxiety | Switching vs Continuation | BMI | Education_levelTwelve or GED | 1.389 | 1.259 | 1.533 | 5.79e-11 | NA | 22696 | 11798 | 10898 |
| European | Recorded Anxiety | Switching vs Continuation | BMI | BMI | 1.055 | 1.027 | 1.084 | 1.10e-04 | 7.73e-04 | 22696 | 11798 | 10898 |
| European | Recorded Anxiety | Discontinuation vs Continuation | Bipolar Disorder | (Intercept) | 0.324 | 0.271 | 0.387 | <2.2e-16 | NA | 15204 | 11798 | 3406 |
| European | Recorded Anxiety | Discontinuation vs Continuation | Bipolar Disorder | age_at_first_AD | 1.005 | 1.002 | 1.007 | 7.27e-05 | NA | 15204 | 11798 | 3406 |
| European | Recorded Anxiety | Discontinuation vs Continuation | Bipolar Disorder | sex_at_birthMale | 1.221 | 1.122 | 1.328 | 3.54e-06 | NA | 15204 | 11798 | 3406 |
| European | Recorded Anxiety | Discontinuation vs Continuation | Bipolar Disorder | Income_quantitative | 0.879 | 0.862 | 0.896 | <2.2e-16 | NA | 15204 | 11798 | 3406 |
| European | Recorded Anxiety | Discontinuation vs Continuation | Bipolar Disorder | Education_levelCollege Undergraduate | 1.101 | 1.000 | 1.213 | 5.07e-02 | NA | 15204 | 11798 | 3406 |
| European | Recorded Anxiety | Discontinuation vs Continuation | Bipolar Disorder | Education_levelLess than a high school degree | 1.773 | 1.366 | 2.301 | 1.66e-05 | NA | 15204 | 11798 | 3406 |
| European | Recorded Anxiety | Discontinuation vs Continuation | Bipolar Disorder | Education_levelTwelve or GED | 1.425 | 1.242 | 1.636 | 4.75e-07 | NA | 15204 | 11798 | 3406 |
| European | Recorded Anxiety | Discontinuation vs Continuation | Bipolar Disorder | Bipolar Disorder | 1.081 | 1.038 | 1.127 | 1.90e-04 | 1.33e-03 | 15204 | 11798 | 3406 |
| European | Recorded Anxiety | Switching vs Continuation | Bipolar Disorder | (Intercept) | 3.072 | 2.716 | 3.475 | <2.2e-16 | NA | 22696 | 11798 | 10898 |
| European | Recorded Anxiety | Switching vs Continuation | Bipolar Disorder | age_at_first_AD | 0.982 | 0.980 | 0.984 | <2.2e-16 | NA | 22696 | 11798 | 10898 |
| European | Recorded Anxiety | Switching vs Continuation | Bipolar Disorder | sex_at_birthMale | 1.159 | 1.091 | 1.231 | 1.48e-06 | NA | 22696 | 11798 | 10898 |
| European | Recorded Anxiety | Switching vs Continuation | Bipolar Disorder | Income_quantitative | 0.883 | 0.871 | 0.895 | <2.2e-16 | NA | 22696 | 11798 | 10898 |
| European | Recorded Anxiety | Switching vs Continuation | Bipolar Disorder | Education_levelCollege Undergraduate | 1.115 | 1.044 | 1.189 | 1.09e-03 | NA | 22696 | 11798 | 10898 |
| European | Recorded Anxiety | Switching vs Continuation | Bipolar Disorder | Education_levelLess than a high school degree | 1.250 | 1.014 | 1.541 | 3.63e-02 | NA | 22696 | 11798 | 10898 |
| European | Recorded Anxiety | Switching vs Continuation | Bipolar Disorder | Education_levelTwelve or GED | 1.410 | 1.279 | 1.556 | 6.65e-12 | NA | 22696 | 11798 | 10898 |
| European | Recorded Anxiety | Switching vs Continuation | Bipolar Disorder | Bipolar Disorder | 1.034 | 1.006 | 1.064 | 1.89e-02 | 1.33e-01 | 22696 | 11798 | 10898 |
| European | Recorded Anxiety | Discontinuation vs Continuation | Depression | (Intercept) | 0.321 | 0.268 | 0.384 | <2.2e-16 | NA | 15204 | 11798 | 3406 |
| European | Recorded Anxiety | Discontinuation vs Continuation | Depression | age_at_first_AD | 1.005 | 1.003 | 1.008 | 3.40e-05 | NA | 15204 | 11798 | 3406 |
| European | Recorded Anxiety | Discontinuation vs Continuation | Depression | sex_at_birthMale | 1.219 | 1.121 | 1.327 | 4.03e-06 | NA | 15204 | 11798 | 3406 |

| | | | | | | | | | | | | |
|---|---|---|---|---|---|---|---|---|---|---|---|---|
| European | Recorded Anxiety | Discontinuation vs Continuation | Depression | Income_quantitative | 0.883 | 0.865 | 0.900 | <2.2e-16 | NA | 15204 | 11798 | 3406 |
| European | Recorded Anxiety | Discontinuation vs Continuation | Depression | Education_levelCollege Undergraduate | 1.084 | 0.984 | 1.195 | 1.01e-01 | NA | 15204 | 11798 | 3406 |
| European | Recorded Anxiety | Discontinuation vs Continuation | Depression | Education_levelLess than a high school degree | 1.692 | 1.303 | 2.198 | 7.85e-05 | NA | 15204 | 11798 | 3406 |
| European | Recorded Anxiety | Discontinuation vs Continuation | Depression | Education_levelTwelve or GED | 1.391 | 1.212 | 1.597 | 2.76e-06 | NA | 15204 | 11798 | 3406 |
| European | Recorded Anxiety | Discontinuation vs Continuation | Depression | Depression | 1.143 | 1.098 | 1.190 | 8.16e-11 | 5.71e-10 | 15204 | 11798 | 3406 |
| European | Recorded Anxiety | Switching vs Continuation | Depression | (Intercept) | 3.015 | 2.664 | 3.412 | <2.2e-16 | NA | 22696 | 11798 | 10898 |
| European | Recorded Anxiety | Switching vs Continuation | Depression | age_at_first_AD | 0.982 | 0.981 | 0.984 | <2.2e-16 | NA | 22696 | 11798 | 10898 |
| European | Recorded Anxiety | Switching vs Continuation | Depression | sex_at_birthMale | 1.149 | 1.081 | 1.220 | 6.68e-06 | NA | 22696 | 11798 | 10898 |
| European | Recorded Anxiety | Switching vs Continuation | Depression | Income_quantitative | 0.888 | 0.876 | 0.900 | <2.2e-16 | NA | 22696 | 11798 | 10898 |
| European | Recorded Anxiety | Switching vs Continuation | Depression | Education_levelCollege Undergraduate | 1.102 | 1.033 | 1.177 | 3.47e-03 | NA | 22696 | 11798 | 10898 |
| European | Recorded Anxiety | Switching vs Continuation | Depression | Education_levelLess than a high school degree | 1.176 | 0.954 | 1.451 | 1.30e-01 | NA | 22696 | 11798 | 10898 |
| European | Recorded Anxiety | Switching vs Continuation | Depression | Education_levelTwelve or GED | 1.387 | 1.256 | 1.530 | 7.97e-11 | NA | 22696 | 11798 | 10898 |
| European | Recorded Anxiety | Switching vs Continuation | Depression | Depression | 1.198 | 1.165 | 1.233 | <2.2e-16 | <2.2e-16 | 22696 | 11798 | 10898 |
| European | Recorded Anxiety | Discontinuation vs Continuation | Insomnia | (Intercept) | 0.322 | 0.269 | 0.385 | <2.2e-16 | NA | 15204 | 11798 | 3406 |
| European | Recorded Anxiety | Discontinuation vs Continuation | Insomnia | age_at_first_AD | 1.005 | 1.003 | 1.008 | 3.10e-05 | NA | 15204 | 11798 | 3406 |
| European | Recorded Anxiety | Discontinuation vs Continuation | Insomnia | sex_at_birthMale | 1.222 | 1.124 | 1.330 | 3.03e-06 | NA | 15204 | 11798 | 3406 |
| European | Recorded Anxiety | Discontinuation vs Continuation | Insomnia | Income_quantitative | 0.881 | 0.864 | 0.899 | <2.2e-16 | NA | 15204 | 11798 | 3406 |
| European | Recorded Anxiety | Discontinuation vs Continuation | Insomnia | Education_levelCollege Undergraduate | 1.084 | 0.984 | 1.194 | 1.03e-01 | NA | 15204 | 11798 | 3406 |
| European | Recorded Anxiety | Discontinuation vs Continuation | Insomnia | Education_levelLess than a high school degree | 1.687 | 1.299 | 2.190 | 8.79e-05 | NA | 15204 | 11798 | 3406 |
| European | Recorded Anxiety | Discontinuation vs Continuation | Insomnia | Education_levelTwelve or GED | 1.384 | 1.206 | 1.590 | 3.93e-06 | NA | 15204 | 11798 | 3406 |
| European | Recorded Anxiety | Discontinuation vs Continuation | Insomnia | Insomnia | 1.121 | 1.076 | 1.168 | 4.69e-08 | 3.28e-07 | 15204 | 11798 | 3406 |
| European | Recorded Anxiety | Switching vs Continuation | Insomnia | (Intercept) | 3.033 | 2.681 | 3.431 | <2.2e-16 | NA | 22696 | 11798 | 10898 |
| European | Recorded Anxiety | Switching vs Continuation | Insomnia | age_at_first_AD | 0.982 | 0.981 | 0.984 | <2.2e-16 | NA | 22696 | 11798 | 10898 |
| European | Recorded Anxiety | Switching vs Continuation | Insomnia | sex_at_birthMale | 1.157 | 1.089 | 1.229 | 2.01e-06 | NA | 22696 | 11798 | 10898 |
| European | Recorded Anxiety | Switching vs Continuation | Insomnia | Income_quantitative | 0.886 | 0.874 | 0.898 | <2.2e-16 | NA | 22696 | 11798 | 10898 |
| European | Recorded Anxiety | Switching vs Continuation | Insomnia | Education_levelCollege Undergraduate | 1.103 | 1.033 | 1.177 | 3.31e-03 | NA | 22696 | 11798 | 10898 |
| European | Recorded Anxiety | Switching vs Continuation | Insomnia | Education_levelLess than a high school degree | 1.199 | 0.973 | 1.479 | 8.88e-02 | NA | 22696 | 11798 | 10898 |
| European | Recorded Anxiety | Switching vs Continuation | Insomnia | Education_levelTwelve or GED | 1.385 | 1.255 | 1.528 | 8.51e-11 | NA | 22696 | 11798 | 10898 |
| European | Recorded Anxiety | Switching vs Continuation | Insomnia | Insomnia | 1.116 | 1.084 | 1.148 | 4.01e-14 | 2.81e-13 | 22696 | 11798 | 10898 |
| European | Recorded Anxiety | Discontinuation vs Continuation | Migraine | (Intercept) | 0.325 | 0.272 | 0.388 | <2.2e-16 | NA | 15204 | 11798 | 3406 |
| European | Recorded Anxiety | Discontinuation vs Continuation | Migraine | age_at_first_AD | 1.005 | 1.003 | 1.007 | 5.18e-05 | NA | 15204 | 11798 | 3406 |
| European | Recorded Anxiety | Discontinuation vs Continuation | Migraine | sex_at_birthMale | 1.224 | 1.125 | 1.331 | 2.62e-06 | NA | 15204 | 11798 | 3406 |
| European | Recorded Anxiety | Discontinuation vs Continuation | Migraine | Income_quantitative | 0.879 | 0.862 | 0.896 | <2.2e-16 | NA | 15204 | 11798 | 3406 |
| European | Recorded Anxiety | Discontinuation vs Continuation | Migraine | Education_levelCollege Undergraduate | 1.095 | 0.994 | 1.206 | 6.67e-02 | NA | 15204 | 11798 | 3406 |
| European | Recorded Anxiety | Discontinuation vs Continuation | Migraine | Education_levelLess than a high school degree | 1.756 | 1.353 | 2.279 | 2.27e-05 | NA | 15204 | 11798 | 3406 |
| European | Recorded Anxiety | Discontinuation vs Continuation | Migraine | Education_levelTwelve or GED | 1.412 | 1.230 | 1.620 | 9.40e-07 | NA | 15204 | 11798 | 3406 |
| European | Recorded Anxiety | Discontinuation vs Continuation | Migraine | Migraine | 1.029 | 0.990 | 1.069 | 1.51e-01 | 1.00e+00 | 15204 | 11798 | 3406 |
| European | Recorded Anxiety | Switching vs Continuation | Migraine | (Intercept) | 3.065 | 2.709 | 3.467 | <2.2e-16 | NA | 22696 | 11798 | 10898 |
| European | Recorded Anxiety | Switching vs Continuation | Migraine | age_at_first_AD | 0.982 | 0.981 | 0.984 | <2.2e-16 | NA | 22696 | 11798 | 10898 |
| European | Recorded Anxiety | Switching vs Continuation | Migraine | sex_at_birthMale | 1.158 | 1.091 | 1.230 | 1.62e-06 | NA | 22696 | 11798 | 10898 |
| European | Recorded Anxiety | Switching vs Continuation | Migraine | Income_quantitative | 0.883 | 0.871 | 0.895 | <2.2e-16 | NA | 22696 | 11798 | 10898 |
| European | Recorded Anxiety | Switching vs Continuation | Migraine | Education_levelCollege Undergraduate | 1.111 | 1.041 | 1.186 | 1.47e-03 | NA | 22696 | 11798 | 10898 |
| European | Recorded Anxiety | Switching vs Continuation | Migraine | Education_levelLess than a high school degree | 1.246 | 1.011 | 1.536 | 3.91e-02 | NA | 22696 | 11798 | 10898 |
| European | Recorded Anxiety | Switching vs Continuation | Migraine | Education_levelTwelve or GED | 1.405 | 1.273 | 1.550 | 1.18e-11 | NA | 22696 | 11798 | 10898 |
| European | Recorded Anxiety | Switching vs Continuation | Migraine | Migraine | 1.049 | 1.021 | 1.077 | 4.93e-04 | 3.45e-03 | 22696 | 11798 | 10898 |
| European | Recorded Anxiety | Discontinuation vs Continuation | Schizophrenia | (Intercept) | 0.323 | 0.270 | 0.386 | <2.2e-16 | NA | 15204 | 11798 | 3406 |
| European | Recorded Anxiety | Discontinuation vs Continuation | Schizophrenia | age_at_first_AD | 1.005 | 1.003 | 1.007 | 5.34e-05 | NA | 15204 | 11798 | 3406 |
| European | Recorded Anxiety | Discontinuation vs Continuation | Schizophrenia | sex_at_birthMale | 1.222 | 1.123 | 1.329 | 3.13e-06 | NA | 15204 | 11798 | 3406 |
| European | Recorded Anxiety | Discontinuation vs Continuation | Schizophrenia | Income_quantitative | 0.879 | 0.862 | 0.897 | <2.2e-16 | NA | 15204 | 11798 | 3406 |
| European | Recorded Anxiety | Discontinuation vs Continuation | Schizophrenia | Education_levelCollege Undergraduate | 1.098 | 0.997 | 1.209 | 5.88e-02 | NA | 15204 | 11798 | 3406 |
| European | Recorded Anxiety | Discontinuation vs Continuation | Schizophrenia | Education_levelLess than a high school degree | 1.750 | 1.349 | 2.271 | 2.55e-05 | NA | 15204 | 11798 | 3406 |
| European | Recorded Anxiety | Discontinuation vs Continuation | Schizophrenia | Education_levelTwelve or GED | 1.417 | 1.235 | 1.627 | 7.09e-07 | NA | 15204 | 11798 | 3406 |
| European | Recorded Anxiety | Discontinuation vs Continuation | Schizophrenia | Schizophrenia | 1.053 | 1.013 | 1.095 | 9.72e-03 | 6.81e-02 | 15204 | 11798 | 3406 |
| European | Recorded Anxiety | Switching vs Continuation | Schizophrenia | (Intercept) | 3.068 | 2.712 | 3.470 | <2.2e-16 | NA | 22696 | 11798 | 10898 |
| European | Recorded Anxiety | Switching vs Continuation | Schizophrenia | age_at_first_AD | 0.982 | 0.981 | 0.984 | <2.2e-16 | NA | 22696 | 11798 | 10898 |
| European | Recorded Anxiety | Switching vs Continuation | Schizophrenia | sex_at_birthMale | 1.160 | 1.092 | 1.232 | 1.31e-06 | NA | 22696 | 11798 | 10898 |
| European | Recorded Anxiety | Switching vs Continuation | Schizophrenia | Income_quantitative | 0.883 | 0.871 | 0.895 | <2.2e-16 | NA | 22696 | 11798 | 10898 |

| | | | | | | | | | | | | |
|---|---|---|---|---|---|---|---|---|---|---|---|---|
| European | Recorded Anxiety | Switching vs Continuation | Schizophrenia | Education_levelCollege Undergraduate | 1.113 | 1.043 | 1.188 | 1.24e-03 | NA | 22696 | 11798 | 10898 |
| European | Recorded Anxiety | Switching vs Continuation | Schizophrenia | Education_levelLess than a high school degree | 1.248 | 1.013 | 1.538 | 3.76e-02 | NA | 22696 | 11798 | 10898 |
| European | Recorded Anxiety | Switching vs Continuation | Schizophrenia | Education_levelTwelve or GED | 1.407 | 1.276 | 1.552 | 9.10e-12 | NA | 22696 | 11798 | 10898 |
| European | Recorded Anxiety | Switching vs Continuation | Schizophrenia | Schizophrenia | 1.006 | 0.979 | 1.033 | 6.90e-01 | 1.00e+00 | 22696 | 11798 | 10898 |
| European | CIDI-MDD | Discontinuation vs Continuation | ADHD | (Intercept) | 0.351 | 0.238 | 0.519 | 1.42e-07 | NA | 3093 | 2349 | 744 |
| European | CIDI-MDD | Discontinuation vs Continuation | ADHD | age_at_first_AD | 1.000 | 0.994 | 1.005 | 8.93e-01 | NA | 3093 | 2349 | 744 |
| European | CIDI-MDD | Discontinuation vs Continuation | ADHD | sex_at_birthMale | 1.047 | 0.864 | 1.269 | 6.41e-01 | NA | 3093 | 2349 | 744 |
| European | CIDI-MDD | Discontinuation vs Continuation | ADHD | Income_quantitative | 0.950 | 0.911 | 0.990 | 1.47e-02 | NA | 3093 | 2349 | 744 |
| European | CIDI-MDD | Discontinuation vs Continuation | ADHD | Education_levelCollege Undergraduate | 1.236 | 1.023 | 1.493 | 2.84e-02 | NA | 3093 | 2349 | 744 |
| European | CIDI-MDD | Discontinuation vs Continuation | ADHD | Education_levelLess than a high school degree | 0.948 | 0.339 | 2.651 | 9.19e-01 | NA | 3093 | 2349 | 744 |
| European | CIDI-MDD | Discontinuation vs Continuation | ADHD | Education_levelTwelve or GED | 1.109 | 0.760 | 1.618 | 5.91e-01 | NA | 3093 | 2349 | 744 |
| European | CIDI-MDD | Discontinuation vs Continuation | ADHD | ADHD | 1.052 | 0.966 | 1.146 | 2.41e-01 | 1.00e+00 | 3093 | 2349 | 744 |
| European | CIDI-MDD | Switching vs Continuation | ADHD | (Intercept) | 4.245 | 3.194 | 5.642 | <2.2e-16 | NA | 4499 | 2349 | 2150 |
| European | CIDI-MDD | Switching vs Continuation | ADHD | age_at_first_AD | 0.976 | 0.971 | 0.980 | <2.2e-16 | NA | 4499 | 2349 | 2150 |
| European | CIDI-MDD | Switching vs Continuation | ADHD | sex_at_birthMale | 1.103 | 0.958 | 1.269 | 1.73e-01 | NA | 4499 | 2349 | 2150 |
| European | CIDI-MDD | Switching vs Continuation | ADHD | Income_quantitative | 0.896 | 0.869 | 0.924 | 2.73e-12 | NA | 4499 | 2349 | 2150 |
| European | CIDI-MDD | Switching vs Continuation | ADHD | Education_levelCollege Undergraduate | 1.130 | 0.985 | 1.297 | 8.05e-02 | NA | 4499 | 2349 | 2150 |
| European | CIDI-MDD | Switching vs Continuation | ADHD | Education_levelLess than a high school degree | 0.541 | 0.249 | 1.178 | 1.22e-01 | NA | 4499 | 2349 | 2150 |
| European | CIDI-MDD | Switching vs Continuation | ADHD | Education_levelTwelve or GED | 1.313 | 1.012 | 1.704 | 4.06e-02 | NA | 4499 | 2349 | 2150 |
| European | CIDI-MDD | Switching vs Continuation | ADHD | ADHD | 1.047 | 0.983 | 1.115 | 1.55e-01 | 1.00e+00 | 4499 | 2349 | 2150 |
| European | CIDI-MDD | Discontinuation vs Continuation | BMI | (Intercept) | 0.353 | 0.240 | 0.522 | 1.61e-07 | NA | 3093 | 2349 | 744 |
| European | CIDI-MDD | Discontinuation vs Continuation | BMI | age_at_first_AD | 1.000 | 0.994 | 1.005 | 8.81e-01 | NA | 3093 | 2349 | 744 |
| European | CIDI-MDD | Discontinuation vs Continuation | BMI | sex_at_birthMale | 1.047 | 0.864 | 1.270 | 6.38e-01 | NA | 3093 | 2349 | 744 |
| European | CIDI-MDD | Discontinuation vs Continuation | BMI | Income_quantitative | 0.946 | 0.908 | 0.986 | 9.15e-03 | NA | 3093 | 2349 | 744 |
| European | CIDI-MDD | Discontinuation vs Continuation | BMI | Education_levelCollege Undergraduate | 1.253 | 1.038 | 1.513 | 1.91e-02 | NA | 3093 | 2349 | 744 |
| European | CIDI-MDD | Discontinuation vs Continuation | BMI | Education_levelLess than a high school degree | 0.997 | 0.357 | 2.787 | 9.96e-01 | NA | 3093 | 2349 | 744 |
| European | CIDI-MDD | Discontinuation vs Continuation | BMI | Education_levelTwelve or GED | 1.135 | 0.778 | 1.655 | 5.11e-01 | NA | 3093 | 2349 | 744 |
| European | CIDI-MDD | Discontinuation vs Continuation | BMI | BMI | 0.959 | 0.880 | 1.045 | 3.38e-01 | 1.00e+00 | 3093 | 2349 | 744 |
| European | CIDI-MDD | Switching vs Continuation | BMI | (Intercept) | 4.208 | 3.165 | 5.594 | <2.2e-16 | NA | 4499 | 2349 | 2150 |
| European | CIDI-MDD | Switching vs Continuation | BMI | age_at_first_AD | 0.976 | 0.972 | 0.980 | <2.2e-16 | NA | 4499 | 2349 | 2150 |
| European | CIDI-MDD | Switching vs Continuation | BMI | sex_at_birthMale | 1.100 | 0.955 | 1.266 | 1.86e-01 | NA | 4499 | 2349 | 2150 |
| European | CIDI-MDD | Switching vs Continuation | BMI | Income_quantitative | 0.897 | 0.870 | 0.925 | 3.88e-12 | NA | 4499 | 2349 | 2150 |
| European | CIDI-MDD | Switching vs Continuation | BMI | Education_levelCollege Undergraduate | 1.135 | 0.990 | 1.302 | 7.02e-02 | NA | 4499 | 2349 | 2150 |
| European | CIDI-MDD | Switching vs Continuation | BMI | Education_levelLess than a high school degree | 0.547 | 0.251 | 1.192 | 1.29e-01 | NA | 4499 | 2349 | 2150 |
| European | CIDI-MDD | Switching vs Continuation | BMI | Education_levelTwelve or GED | 1.313 | 1.012 | 1.704 | 4.03e-02 | NA | 4499 | 2349 | 2150 |
| European | CIDI-MDD | Switching vs Continuation | BMI | BMI | 1.066 | 1.002 | 1.134 | 4.14e-02 | 2.90e-01 | 4499 | 2349 | 2150 |
| European | CIDI-MDD | Discontinuation vs Continuation | Bipolar Disorder | (Intercept) | 0.357 | 0.241 | 0.526 | 2.15e-07 | NA | 3093 | 2349 | 744 |
| European | CIDI-MDD | Discontinuation vs Continuation | Bipolar Disorder | age_at_first_AD | 0.999 | 0.994 | 1.005 | 7.81e-01 | NA | 3093 | 2349 | 744 |
| European | CIDI-MDD | Discontinuation vs Continuation | Bipolar Disorder | sex_at_birthMale | 1.045 | 0.862 | 1.266 | 6.57e-01 | NA | 3093 | 2349 | 744 |
| European | CIDI-MDD | Discontinuation vs Continuation | Bipolar Disorder | Income_quantitative | 0.948 | 0.909 | 0.988 | 1.07e-02 | NA | 3093 | 2349 | 744 |
| European | CIDI-MDD | Discontinuation vs Continuation | Bipolar Disorder | Education_levelCollege Undergraduate | 1.256 | 1.040 | 1.517 | 1.77e-02 | NA | 3093 | 2349 | 744 |
| European | CIDI-MDD | Discontinuation vs Continuation | Bipolar Disorder | Education_levelLess than a high school degree | 1.023 | 0.366 | 2.862 | 9.65e-01 | NA | 3093 | 2349 | 744 |
| European | CIDI-MDD | Discontinuation vs Continuation | Bipolar Disorder | Education_levelTwelve or GED | 1.147 | 0.787 | 1.673 | 4.76e-01 | NA | 3093 | 2349 | 744 |
| European | CIDI-MDD | Discontinuation vs Continuation | Bipolar Disorder | Bipolar Disorder | 1.139 | 1.041 | 1.246 | 4.46e-03 | 3.12e-02 | 3093 | 2349 | 744 |
| European | CIDI-MDD | Switching vs Continuation | Bipolar Disorder | (Intercept) | 4.277 | 3.218 | 5.684 | <2.2e-16 | NA | 4499 | 2349 | 2150 |
| European | CIDI-MDD | Switching vs Continuation | Bipolar Disorder | age_at_first_AD | 0.975 | 0.971 | 0.980 | <2.2e-16 | NA | 4499 | 2349 | 2150 |
| European | CIDI-MDD | Switching vs Continuation | Bipolar Disorder | sex_at_birthMale | 1.102 | 0.957 | 1.269 | 1.76e-01 | NA | 4499 | 2349 | 2150 |
| European | CIDI-MDD | Switching vs Continuation | Bipolar Disorder | Income_quantitative | 0.894 | 0.867 | 0.922 | 1.01e-12 | NA | 4499 | 2349 | 2150 |
| European | CIDI-MDD | Switching vs Continuation | Bipolar Disorder | Education_levelCollege Undergraduate | 1.142 | 0.996 | 1.310 | 5.68e-02 | NA | 4499 | 2349 | 2150 |
| European | CIDI-MDD | Switching vs Continuation | Bipolar Disorder | Education_levelLess than a high school degree | 0.561 | 0.258 | 1.222 | 1.46e-01 | NA | 4499 | 2349 | 2150 |
| European | CIDI-MDD | Switching vs Continuation | Bipolar Disorder | Education_levelTwelve or GED | 1.337 | 1.031 | 1.735 | 2.85e-02 | NA | 4499 | 2349 | 2150 |
| European | CIDI-MDD | Switching vs Continuation | Bipolar Disorder | Bipolar Disorder | 1.047 | 0.982 | 1.117 | 1.57e-01 | 1.00e+00 | 4499 | 2349 | 2150 |
| European | CIDI-MDD | Discontinuation vs Continuation | Depression | (Intercept) | 0.352 | 0.238 | 0.519 | 1.52e-07 | NA | 3093 | 2349 | 744 |
| European | CIDI-MDD | Discontinuation vs Continuation | Depression | age_at_first_AD | 1.000 | 0.994 | 1.005 | 8.67e-01 | NA | 3093 | 2349 | 744 |
| European | CIDI-MDD | Discontinuation vs Continuation | Depression | sex_at_birthMale | 1.040 | 0.858 | 1.262 | 6.87e-01 | NA | 3093 | 2349 | 744 |
| European | CIDI-MDD | Discontinuation vs Continuation | Depression | Income_quantitative | 0.952 | 0.914 | 0.993 | 2.05e-02 | NA | 3093 | 2349 | 744 |
| European | CIDI-MDD | Discontinuation vs Continuation | Depression | Education_levelCollege Undergraduate | 1.219 | 1.009 | 1.472 | 4.02e-02 | NA | 3093 | 2349 | 744 |

| | | | | | | | | | | | | |
|---|---|---|---|---|---|---|---|---|---|---|---|---|
| European | CIDI-MDD | Discontinuation vs Continuation | Depression | Education_levelLess than a high school degree | 0.971 | 0.347 | 2.716 | 9.55e-01 | NA | 3093 | 2349 | 744 |
| European | CIDI-MDD | Discontinuation vs Continuation | Depression | Education_levelTwelve or GED | 1.102 | 0.756 | 1.609 | 6.13e-01 | NA | 3093 | 2349 | 744 |
| European | CIDI-MDD | Discontinuation vs Continuation | Depression | Depression | 1.201 | 1.100 | 1.311 | 4.12e-05 | 2.88e-04 | 3093 | 2349 | 744 |
| European | CIDI-MDD | Switching vs Continuation | Depression | (Intercept) | 4.271 | 3.213 | 5.679 | <2.2e-16 | NA | 4499 | 2349 | 2150 |
| European | CIDI-MDD | Switching vs Continuation | Depression | age_at_first_AD | 0.975 | 0.971 | 0.980 | <2.2e-16 | NA | 4499 | 2349 | 2150 |
| European | CIDI-MDD | Switching vs Continuation | Depression | sex_at_birthMale | 1.098 | 0.954 | 1.264 | 1.92e-01 | NA | 4499 | 2349 | 2150 |
| European | CIDI-MDD | Switching vs Continuation | Depression | Income_quantitative | 0.897 | 0.870 | 0.925 | 4.11e-12 | NA | 4499 | 2349 | 2150 |
| European | CIDI-MDD | Switching vs Continuation | Depression | Education_levelCollege Undergraduate | 1.124 | 0.980 | 1.290 | 9.43e-02 | NA | 4499 | 2349 | 2150 |
| European | CIDI-MDD | Switching vs Continuation | Depression | Education_levelLess than a high school degree | 0.546 | 0.250 | 1.189 | 1.27e-01 | NA | 4499 | 2349 | 2150 |
| European | CIDI-MDD | Switching vs Continuation | Depression | Education_levelTwelve or GED | 1.318 | 1.016 | 1.711 | 3.77e-02 | NA | 4499 | 2349 | 2150 |
| European | CIDI-MDD | Switching vs Continuation | Depression | Depression | 1.128 | 1.058 | 1.203 | 2.15e-04 | 1.50e-03 | 4499 | 2349 | 2150 |
| European | CIDI-MDD | Discontinuation vs Continuation | Insomnia | (Intercept) | 0.352 | 0.238 | 0.520 | 1.51e-07 | NA | 3093 | 2349 | 744 |
| European | CIDI-MDD | Discontinuation vs Continuation | Insomnia | age_at_first_AD | 1.000 | 0.994 | 1.005 | 8.48e-01 | NA | 3093 | 2349 | 744 |
| European | CIDI-MDD | Discontinuation vs Continuation | Insomnia | sex_at_birthMale | 1.052 | 0.867 | 1.275 | 6.09e-01 | NA | 3093 | 2349 | 744 |
| European | CIDI-MDD | Discontinuation vs Continuation | Insomnia | Income_quantitative | 0.951 | 0.912 | 0.991 | 1.65e-02 | NA | 3093 | 2349 | 744 |
| European | CIDI-MDD | Discontinuation vs Continuation | Insomnia | Education_levelCollege Undergraduate | 1.232 | 1.020 | 1.488 | 3.06e-02 | NA | 3093 | 2349 | 744 |
| European | CIDI-MDD | Discontinuation vs Continuation | Insomnia | Education_levelLess than a high school degree | 0.947 | 0.339 | 2.647 | 9.17e-01 | NA | 3093 | 2349 | 744 |
| European | CIDI-MDD | Discontinuation vs Continuation | Insomnia | Education_levelTwelve or GED | 1.113 | 0.763 | 1.622 | 5.79e-01 | NA | 3093 | 2349 | 744 |
| European | CIDI-MDD | Discontinuation vs Continuation | Insomnia | Insomnia | 1.119 | 1.025 | 1.221 | 1.19e-02 | 8.30e-02 | 3093 | 2349 | 744 |
| European | CIDI-MDD | Switching vs Continuation | Insomnia | (Intercept) | 4.260 | 3.205 | 5.661 | <2.2e-16 | NA | 4499 | 2349 | 2150 |
| European | CIDI-MDD | Switching vs Continuation | Insomnia | age_at_first_AD | 0.976 | 0.971 | 0.980 | <2.2e-16 | NA | 4499 | 2349 | 2150 |
| European | CIDI-MDD | Switching vs Continuation | Insomnia | sex_at_birthMale | 1.101 | 0.957 | 1.268 | 1.79e-01 | NA | 4499 | 2349 | 2150 |
| European | CIDI-MDD | Switching vs Continuation | Insomnia | Income_quantitative | 0.895 | 0.868 | 0.923 | 1.16e-12 | NA | 4499 | 2349 | 2150 |
| European | CIDI-MDD | Switching vs Continuation | Insomnia | Education_levelCollege Undergraduate | 1.140 | 0.994 | 1.307 | 6.15e-02 | NA | 4499 | 2349 | 2150 |
| European | CIDI-MDD | Switching vs Continuation | Insomnia | Education_levelLess than a high school degree | 0.551 | 0.253 | 1.199 | 1.33e-01 | NA | 4499 | 2349 | 2150 |
| European | CIDI-MDD | Switching vs Continuation | Insomnia | Education_levelTwelve or GED | 1.331 | 1.026 | 1.726 | 3.12e-02 | NA | 4499 | 2349 | 2150 |
| European | CIDI-MDD | Switching vs Continuation | Insomnia | Insomnia | 1.000 | 0.938 | 1.065 | 9.92e-01 | 1.00e+00 | 4499 | 2349 | 2150 |
| European | CIDI-MDD | Discontinuation vs Continuation | Migraine | (Intercept) | 0.352 | 0.238 | 0.519 | 1.44e-07 | NA | 3093 | 2349 | 744 |
| European | CIDI-MDD | Discontinuation vs Continuation | Migraine | age_at_first_AD | 1.000 | 0.994 | 1.005 | 8.92e-01 | NA | 3093 | 2349 | 744 |
| European | CIDI-MDD | Discontinuation vs Continuation | Migraine | sex_at_birthMale | 1.044 | 0.862 | 1.266 | 6.58e-01 | NA | 3093 | 2349 | 744 |
| European | CIDI-MDD | Discontinuation vs Continuation | Migraine | Income_quantitative | 0.948 | 0.910 | 0.988 | 1.12e-02 | NA | 3093 | 2349 | 744 |
| European | CIDI-MDD | Discontinuation vs Continuation | Migraine | Education_levelCollege Undergraduate | 1.249 | 1.035 | 1.508 | 2.07e-02 | NA | 3093 | 2349 | 744 |
| European | CIDI-MDD | Discontinuation vs Continuation | Migraine | Education_levelLess than a high school degree | 0.981 | 0.351 | 2.741 | 9.71e-01 | NA | 3093 | 2349 | 744 |
| European | CIDI-MDD | Discontinuation vs Continuation | Migraine | Education_levelTwelve or GED | 1.127 | 0.774 | 1.643 | 5.33e-01 | NA | 3093 | 2349 | 744 |
| European | CIDI-MDD | Discontinuation vs Continuation | Migraine | Migraine | 0.997 | 0.919 | 1.081 | 9.37e-01 | 1.00e+00 | 3093 | 2349 | 744 |
| European | CIDI-MDD | Switching vs Continuation | Migraine | (Intercept) | 4.258 | 3.204 | 5.659 | <2.2e-16 | NA | 4499 | 2349 | 2150 |
| European | CIDI-MDD | Switching vs Continuation | Migraine | age_at_first_AD | 0.976 | 0.971 | 0.980 | <2.2e-16 | NA | 4499 | 2349 | 2150 |
| European | CIDI-MDD | Switching vs Continuation | Migraine | sex_at_birthMale | 1.101 | 0.956 | 1.267 | 1.81e-01 | NA | 4499 | 2349 | 2150 |
| European | CIDI-MDD | Switching vs Continuation | Migraine | Income_quantitative | 0.895 | 0.868 | 0.923 | 1.22e-12 | NA | 4499 | 2349 | 2150 |
| European | CIDI-MDD | Switching vs Continuation | Migraine | Education_levelCollege Undergraduate | 1.139 | 0.994 | 1.306 | 6.19e-02 | NA | 4499 | 2349 | 2150 |
| European | CIDI-MDD | Switching vs Continuation | Migraine | Education_levelLess than a high school degree | 0.551 | 0.253 | 1.199 | 1.33e-01 | NA | 4499 | 2349 | 2150 |
| European | CIDI-MDD | Switching vs Continuation | Migraine | Education_levelTwelve or GED | 1.330 | 1.025 | 1.725 | 3.16e-02 | NA | 4499 | 2349 | 2150 |
| European | CIDI-MDD | Switching vs Continuation | Migraine | Migraine | 1.009 | 0.951 | 1.072 | 7.63e-01 | 1.00e+00 | 4499 | 2349 | 2150 |
| European | CIDI-MDD | Discontinuation vs Continuation | Schizophrenia | (Intercept) | 0.352 | 0.238 | 0.519 | 1.44e-07 | NA | 3093 | 2349 | 744 |
| European | CIDI-MDD | Discontinuation vs Continuation | Schizophrenia | age_at_first_AD | 1.000 | 0.994 | 1.005 | 8.83e-01 | NA | 3093 | 2349 | 744 |
| European | CIDI-MDD | Discontinuation vs Continuation | Schizophrenia | sex_at_birthMale | 1.047 | 0.863 | 1.269 | 6.43e-01 | NA | 3093 | 2349 | 744 |
| European | CIDI-MDD | Discontinuation vs Continuation | Schizophrenia | Income_quantitative | 0.948 | 0.910 | 0.988 | 1.08e-02 | NA | 3093 | 2349 | 744 |
| European | CIDI-MDD | Discontinuation vs Continuation | Schizophrenia | Education_levelCollege Undergraduate | 1.252 | 1.037 | 1.512 | 1.95e-02 | NA | 3093 | 2349 | 744 |
| European | CIDI-MDD | Discontinuation vs Continuation | Schizophrenia | Education_levelLess than a high school degree | 0.998 | 0.357 | 2.789 | 9.97e-01 | NA | 3093 | 2349 | 744 |
| European | CIDI-MDD | Discontinuation vs Continuation | Schizophrenia | Education_levelTwelve or GED | 1.136 | 0.779 | 1.657 | 5.07e-01 | NA | 3093 | 2349 | 744 |
| European | CIDI-MDD | Discontinuation vs Continuation | Schizophrenia | Schizophrenia | 1.082 | 0.994 | 1.179 | 6.88e-02 | 4.81e-01 | 3093 | 2349 | 744 |
| European | CIDI-MDD | Switching vs Continuation | Schizophrenia | (Intercept) | 4.257 | 3.203 | 5.658 | <2.2e-16 | NA | 4499 | 2349 | 2150 |
| European | CIDI-MDD | Switching vs Continuation | Schizophrenia | age_at_first_AD | 0.976 | 0.971 | 0.980 | <2.2e-16 | NA | 4499 | 2349 | 2150 |
| European | CIDI-MDD | Switching vs Continuation | Schizophrenia | sex_at_birthMale | 1.104 | 0.959 | 1.271 | 1.69e-01 | NA | 4499 | 2349 | 2150 |
| European | CIDI-MDD | Switching vs Continuation | Schizophrenia | Income_quantitative | 0.895 | 0.868 | 0.923 | 1.24e-12 | NA | 4499 | 2349 | 2150 |
| European | CIDI-MDD | Switching vs Continuation | Schizophrenia | Education_levelCollege Undergraduate | 1.140 | 0.994 | 1.307 | 6.14e-02 | NA | 4499 | 2349 | 2150 |
| European | CIDI-MDD | Switching vs Continuation | Schizophrenia | Education_levelLess than a high school degree | 0.555 | 0.255 | 1.209 | 1.38e-01 | NA | 4499 | 2349 | 2150 |

| | | | | | | | | | | | | |
|---|---|---|---|---|---|---|---|---|---|---|---|---|
| European | CIDI-MDD | Switching vs Continuation | Schizophrenia | Education_levelTwelve or GED | 1.333 | 1.028 | 1.730 | 3.01e-02 | NA | 4499 | 2349 | 2150 |
| European | CIDI-MDD | Switching vs Continuation | Schizophrenia | Schizophrenia | 1.039 | 0.977 | 1.106 | 2.24e-01 | 1.00e+00 | 4499 | 2349 | 2150 |
| European | CIDI-noMDD | Discontinuation vs Continuation | ADHD | (Intercept) | 0.107 | 0.062 | 0.184 | 8.67e-16 | NA | 2624 | 2019 | 605 |
| European | CIDI-noMDD | Discontinuation vs Continuation | ADHD | age_at_first_AD | 1.015 | 1.008 | 1.022 | 4.19e-05 | NA | 2624 | 2019 | 605 |
| European | CIDI-noMDD | Discontinuation vs Continuation | ADHD | sex_at_birthMale | 1.214 | 1.000 | 1.474 | 4.94e-02 | NA | 2624 | 2019 | 605 |
| European | CIDI-noMDD | Discontinuation vs Continuation | ADHD | Income_quantitative | 0.982 | 0.934 | 1.033 | 4.85e-01 | NA | 2624 | 2019 | 605 |
| European | CIDI-noMDD | Discontinuation vs Continuation | ADHD | Education_levelCollege Undergraduate | 1.378 | 1.117 | 1.701 | 2.79e-03 | NA | 2624 | 2019 | 605 |
| European | CIDI-noMDD | Discontinuation vs Continuation | ADHD | Education_levelLess than a high school degree | 0.527 | 0.065 | 4.291 | 5.49e-01 | NA | 2624 | 2019 | 605 |
| European | CIDI-noMDD | Discontinuation vs Continuation | ADHD | Education_levelTwelve or GED | 1.326 | 0.917 | 1.918 | 1.34e-01 | NA | 2624 | 2019 | 605 |
| European | CIDI-noMDD | Discontinuation vs Continuation | ADHD | ADHD | 0.982 | 0.894 | 1.077 | 6.96e-01 | 1.00e+00 | 2624 | 2019 | 605 |
| European | CIDI-noMDD | Switching vs Continuation | ADHD | (Intercept) | 2.254 | 1.395 | 3.643 | 9.07e-04 | NA | 2664 | 2019 | 645 |
| European | CIDI-noMDD | Switching vs Continuation | ADHD | age_at_first_AD | 0.971 | 0.965 | 0.977 | <2.2e-16 | NA | 2664 | 2019 | 645 |
| European | CIDI-noMDD | Switching vs Continuation | ADHD | sex_at_birthMale | 1.324 | 1.087 | 1.612 | 5.30e-03 | NA | 2664 | 2019 | 645 |
| European | CIDI-noMDD | Switching vs Continuation | ADHD | Income_quantitative | 0.893 | 0.850 | 0.937 | 5.52e-06 | NA | 2664 | 2019 | 645 |
| European | CIDI-noMDD | Switching vs Continuation | ADHD | Education_levelCollege Undergraduate | 1.109 | 0.898 | 1.370 | 3.36e-01 | NA | 2664 | 2019 | 645 |
| European | CIDI-noMDD | Switching vs Continuation | ADHD | Education_levelLess than a high school degree | 1.357 | 0.424 | 4.340 | 6.07e-01 | NA | 2664 | 2019 | 645 |
| European | CIDI-noMDD | Switching vs Continuation | ADHD | Education_levelTwelve or GED | 1.256 | 0.887 | 1.779 | 1.98e-01 | NA | 2664 | 2019 | 645 |
| European | CIDI-noMDD | Switching vs Continuation | ADHD | ADHD | 1.070 | 0.974 | 1.175 | 1.57e-01 | 1.00e+00 | 2664 | 2019 | 645 |
| European | CIDI-noMDD | Discontinuation vs Continuation | BMI | (Intercept) | 0.107 | 0.062 | 0.185 | 9.94e-16 | NA | 2624 | 2019 | 605 |
| European | CIDI-noMDD | Discontinuation vs Continuation | BMI | age_at_first_AD | 1.015 | 1.008 | 1.022 | 4.25e-05 | NA | 2624 | 2019 | 605 |
| European | CIDI-noMDD | Discontinuation vs Continuation | BMI | sex_at_birthMale | 1.215 | 1.001 | 1.475 | 4.86e-02 | NA | 2624 | 2019 | 605 |
| European | CIDI-noMDD | Discontinuation vs Continuation | BMI | Income_quantitative | 0.982 | 0.934 | 1.033 | 4.85e-01 | NA | 2624 | 2019 | 605 |
| European | CIDI-noMDD | Discontinuation vs Continuation | BMI | Education_levelCollege Undergraduate | 1.375 | 1.115 | 1.697 | 2.92e-03 | NA | 2624 | 2019 | 605 |
| European | CIDI-noMDD | Discontinuation vs Continuation | BMI | Education_levelLess than a high school degree | 0.523 | 0.064 | 4.255 | 5.44e-01 | NA | 2624 | 2019 | 605 |
| European | CIDI-noMDD | Discontinuation vs Continuation | BMI | Education_levelTwelve or GED | 1.322 | 0.914 | 1.912 | 1.38e-01 | NA | 2624 | 2019 | 605 |
| European | CIDI-noMDD | Discontinuation vs Continuation | BMI | BMI | 0.987 | 0.899 | 1.082 | 7.73e-01 | 1.00e+00 | 2624 | 2019 | 605 |
| European | CIDI-noMDD | Switching vs Continuation | BMI | (Intercept) | 2.265 | 1.401 | 3.662 | 8.49e-04 | NA | 2664 | 2019 | 645 |
| European | CIDI-noMDD | Switching vs Continuation | BMI | age_at_first_AD | 0.971 | 0.965 | 0.977 | <2.2e-16 | NA | 2664 | 2019 | 645 |
| European | CIDI-noMDD | Switching vs Continuation | BMI | sex_at_birthMale | 1.320 | 1.084 | 1.608 | 5.71e-03 | NA | 2664 | 2019 | 645 |
| European | CIDI-noMDD | Switching vs Continuation | BMI | Income_quantitative | 0.892 | 0.849 | 0.937 | 4.87e-06 | NA | 2664 | 2019 | 645 |
| European | CIDI-noMDD | Switching vs Continuation | BMI | Education_levelCollege Undergraduate | 1.120 | 0.908 | 1.382 | 2.91e-01 | NA | 2664 | 2019 | 645 |
| European | CIDI-noMDD | Switching vs Continuation | BMI | Education_levelLess than a high school degree | 1.419 | 0.445 | 4.525 | 5.54e-01 | NA | 2664 | 2019 | 645 |
| European | CIDI-noMDD | Switching vs Continuation | BMI | Education_levelTwelve or GED | 1.274 | 0.901 | 1.803 | 1.71e-01 | NA | 2664 | 2019 | 645 |
| European | CIDI-noMDD | Switching vs Continuation | BMI | BMI | 1.031 | 0.941 | 1.130 | 5.08e-01 | 1.00e+00 | 2664 | 2019 | 645 |
| European | CIDI-noMDD | Discontinuation vs Continuation | Bipolar Disorder | (Intercept) | 0.106 | 0.062 | 0.184 | 8.51e-16 | NA | 2624 | 2019 | 605 |
| European | CIDI-noMDD | Discontinuation vs Continuation | Bipolar Disorder | age_at_first_AD | 1.015 | 1.008 | 1.022 | 3.32e-05 | NA | 2624 | 2019 | 605 |
| European | CIDI-noMDD | Discontinuation vs Continuation | Bipolar Disorder | sex_at_birthMale | 1.218 | 1.003 | 1.478 | 4.64e-02 | NA | 2624 | 2019 | 605 |
| European | CIDI-noMDD | Discontinuation vs Continuation | Bipolar Disorder | Income_quantitative | 0.981 | 0.933 | 1.032 | 4.61e-01 | NA | 2624 | 2019 | 605 |
| European | CIDI-noMDD | Discontinuation vs Continuation | Bipolar Disorder | Education_levelCollege Undergraduate | 1.366 | 1.107 | 1.686 | 3.60e-03 | NA | 2624 | 2019 | 605 |
| European | CIDI-noMDD | Discontinuation vs Continuation | Bipolar Disorder | Education_levelLess than a high school degree | 0.536 | 0.066 | 4.363 | 5.60e-01 | NA | 2624 | 2019 | 605 |
| European | CIDI-noMDD | Discontinuation vs Continuation | Bipolar Disorder | Education_levelTwelve or GED | 1.292 | 0.892 | 1.871 | 1.75e-01 | NA | 2624 | 2019 | 605 |
| European | CIDI-noMDD | Discontinuation vs Continuation | Bipolar Disorder | Bipolar Disorder | 0.933 | 0.844 | 1.032 | 1.77e-01 | 1.00e+00 | 2624 | 2019 | 605 |
| European | CIDI-noMDD | Switching vs Continuation | Bipolar Disorder | (Intercept) | 2.279 | 1.410 | 3.683 | 7.67e-04 | NA | 2664 | 2019 | 645 |
| European | CIDI-noMDD | Switching vs Continuation | Bipolar Disorder | age_at_first_AD | 0.971 | 0.964 | 0.977 | <2.2e-16 | NA | 2664 | 2019 | 645 |
| European | CIDI-noMDD | Switching vs Continuation | Bipolar Disorder | sex_at_birthMale | 1.319 | 1.083 | 1.607 | 5.92e-03 | NA | 2664 | 2019 | 645 |
| European | CIDI-noMDD | Switching vs Continuation | Bipolar Disorder | Income_quantitative | 0.892 | 0.850 | 0.937 | 5.10e-06 | NA | 2664 | 2019 | 645 |
| European | CIDI-noMDD | Switching vs Continuation | Bipolar Disorder | Education_levelCollege Undergraduate | 1.124 | 0.911 | 1.388 | 2.75e-01 | NA | 2664 | 2019 | 645 |
| European | CIDI-noMDD | Switching vs Continuation | Bipolar Disorder | Education_levelLess than a high school degree | 1.398 | 0.436 | 4.479 | 5.73e-01 | NA | 2664 | 2019 | 645 |
| European | CIDI-noMDD | Switching vs Continuation | Bipolar Disorder | Education_levelTwelve or GED | 1.305 | 0.922 | 1.847 | 1.34e-01 | NA | 2664 | 2019 | 645 |
| European | CIDI-noMDD | Switching vs Continuation | Bipolar Disorder | Bipolar Disorder | 1.097 | 0.995 | 1.209 | 6.29e-02 | 4.40e-01 | 2664 | 2019 | 645 |
| European | CIDI-noMDD | Discontinuation vs Continuation | Depression | (Intercept) | 0.107 | 0.062 | 0.184 | 8.16e-16 | NA | 2624 | 2019 | 605 |
| European | CIDI-noMDD | Discontinuation vs Continuation | Depression | age_at_first_AD | 1.015 | 1.008 | 1.022 | 4.13e-05 | NA | 2624 | 2019 | 605 |
| European | CIDI-noMDD | Discontinuation vs Continuation | Depression | sex_at_birthMale | 1.217 | 1.003 | 1.477 | 4.69e-02 | NA | 2624 | 2019 | 605 |
| European | CIDI-noMDD | Discontinuation vs Continuation | Depression | Income_quantitative | 0.982 | 0.934 | 1.033 | 4.89e-01 | NA | 2624 | 2019 | 605 |
| European | CIDI-noMDD | Discontinuation vs Continuation | Depression | Education_levelCollege Undergraduate | 1.376 | 1.115 | 1.698 | 2.89e-03 | NA | 2624 | 2019 | 605 |
| European | CIDI-noMDD | Discontinuation vs Continuation | Depression | Education_levelLess than a high school degree | 0.525 | 0.064 | 4.273 | 5.47e-01 | NA | 2624 | 2019 | 605 |
| European | CIDI-noMDD | Discontinuation vs Continuation | Depression | Education_levelTwelve or GED | 1.326 | 0.917 | 1.919 | 1.34e-01 | NA | 2624 | 2019 | 605 |

| | | | | | | | | | | | | |
|---|---|---|---|---|---|---|---|---|---|---|---|---|
| European | CIDI-noMDD | Discontinuation vs Continuation | Depression | Depression | 0.983 | 0.891 | 1.084 | 7.31e-01 | 1.00e+00 | 2624 | 2019 | 605 |
| European | CIDI-noMDD | Switching vs Continuation | Depression | (Intercept) | 2.225 | 1.375 | 3.598 | 1.12e-03 | NA | 2664 | 2019 | 645 |
| European | CIDI-noMDD | Switching vs Continuation | Depression | age_at_first_AD | 0.971 | 0.965 | 0.978 | <2.2e-16 | NA | 2664 | 2019 | 645 |
| European | CIDI-noMDD | Switching vs Continuation | Depression | sex_at_birthMale | 1.302 | 1.068 | 1.586 | 8.88e-03 | NA | 2664 | 2019 | 645 |
| European | CIDI-noMDD | Switching vs Continuation | Depression | Income_quantitative | 0.894 | 0.851 | 0.939 | 7.57e-06 | NA | 2664 | 2019 | 645 |
| European | CIDI-noMDD | Switching vs Continuation | Depression | Education_levelCollege Undergraduate | 1.108 | 0.897 | 1.367 | 3.42e-01 | NA | 2664 | 2019 | 645 |
| European | CIDI-noMDD | Switching vs Continuation | Depression | Education_levelLess than a high school degree | 1.275 | 0.393 | 4.133 | 6.85e-01 | NA | 2664 | 2019 | 645 |
| European | CIDI-noMDD | Switching vs Continuation | Depression | Education_levelTwelve or GED | 1.235 | 0.872 | 1.749 | 2.36e-01 | NA | 2664 | 2019 | 645 |
| European | CIDI-noMDD | Switching vs Continuation | Depression | Depression | 1.152 | 1.045 | 1.270 | 4.44e-03 | 3.11e-02 | 2664 | 2019 | 645 |
| European | CIDI-noMDD | Discontinuation vs Continuation | Insomnia | (Intercept) | 0.106 | 0.062 | 0.184 | 7.85e-16 | NA | 2624 | 2019 | 605 |
| European | CIDI-noMDD | Discontinuation vs Continuation | Insomnia | age_at_first_AD | 1.015 | 1.008 | 1.022 | 4.02e-05 | NA | 2624 | 2019 | 605 |
| European | CIDI-noMDD | Discontinuation vs Continuation | Insomnia | sex_at_birthMale | 1.216 | 1.002 | 1.476 | 4.80e-02 | NA | 2624 | 2019 | 605 |
| European | CIDI-noMDD | Discontinuation vs Continuation | Insomnia | Income_quantitative | 0.983 | 0.934 | 1.034 | 5.04e-01 | NA | 2624 | 2019 | 605 |
| European | CIDI-noMDD | Discontinuation vs Continuation | Insomnia | Education_levelCollege Undergraduate | 1.374 | 1.113 | 1.695 | 3.05e-03 | NA | 2624 | 2019 | 605 |
| European | CIDI-noMDD | Discontinuation vs Continuation | Insomnia | Education_levelLess than a high school degree | 0.518 | 0.064 | 4.216 | 5.39e-01 | NA | 2624 | 2019 | 605 |
| European | CIDI-noMDD | Discontinuation vs Continuation | Insomnia | Education_levelTwelve or GED | 1.320 | 0.913 | 1.909 | 1.40e-01 | NA | 2624 | 2019 | 605 |
| European | CIDI-noMDD | Discontinuation vs Continuation | Insomnia | Insomnia | 1.010 | 0.917 | 1.112 | 8.42e-01 | 1.00e+00 | 2624 | 2019 | 605 |
| European | CIDI-noMDD | Switching vs Continuation | Insomnia | (Intercept) | 2.235 | 1.382 | 3.614 | 1.04e-03 | NA | 2664 | 2019 | 645 |
| European | CIDI-noMDD | Switching vs Continuation | Insomnia | age_at_first_AD | 0.971 | 0.965 | 0.977 | <2.2e-16 | NA | 2664 | 2019 | 645 |
| European | CIDI-noMDD | Switching vs Continuation | Insomnia | sex_at_birthMale | 1.320 | 1.084 | 1.608 | 5.79e-03 | NA | 2664 | 2019 | 645 |
| European | CIDI-noMDD | Switching vs Continuation | Insomnia | Income_quantitative | 0.894 | 0.851 | 0.939 | 8.00e-06 | NA | 2664 | 2019 | 645 |
| European | CIDI-noMDD | Switching vs Continuation | Insomnia | Education_levelCollege Undergraduate | 1.111 | 0.900 | 1.371 | 3.29e-01 | NA | 2664 | 2019 | 645 |
| European | CIDI-noMDD | Switching vs Continuation | Insomnia | Education_levelLess than a high school degree | 1.367 | 0.427 | 4.375 | 5.98e-01 | NA | 2664 | 2019 | 645 |
| European | CIDI-noMDD | Switching vs Continuation | Insomnia | Education_levelTwelve or GED | 1.261 | 0.891 | 1.785 | 1.91e-01 | NA | 2664 | 2019 | 645 |
| European | CIDI-noMDD | Switching vs Continuation | Insomnia | Insomnia | 1.112 | 1.010 | 1.225 | 3.10e-02 | 2.17e-01 | 2664 | 2019 | 645 |
| European | CIDI-noMDD | Discontinuation vs Continuation | Migraine | (Intercept) | 0.107 | 0.062 | 0.185 | 1.03e-15 | NA | 2624 | 2019 | 605 |
| European | CIDI-noMDD | Discontinuation vs Continuation | Migraine | age_at_first_AD | 1.015 | 1.008 | 1.022 | 4.51e-05 | NA | 2624 | 2019 | 605 |
| European | CIDI-noMDD | Discontinuation vs Continuation | Migraine | sex_at_birthMale | 1.220 | 1.005 | 1.481 | 4.44e-02 | NA | 2624 | 2019 | 605 |
| European | CIDI-noMDD | Discontinuation vs Continuation | Migraine | Income_quantitative | 0.981 | 0.933 | 1.032 | 4.66e-01 | NA | 2624 | 2019 | 605 |
| European | CIDI-noMDD | Discontinuation vs Continuation | Migraine | Education_levelCollege Undergraduate | 1.378 | 1.117 | 1.700 | 2.76e-03 | NA | 2624 | 2019 | 605 |
| European | CIDI-noMDD | Discontinuation vs Continuation | Migraine | Education_levelLess than a high school degree | 0.522 | 0.064 | 4.246 | 5.43e-01 | NA | 2624 | 2019 | 605 |
| European | CIDI-noMDD | Discontinuation vs Continuation | Migraine | Education_levelTwelve or GED | 1.325 | 0.916 | 1.916 | 1.35e-01 | NA | 2624 | 2019 | 605 |
| European | CIDI-noMDD | Discontinuation vs Continuation | Migraine | Migraine | 0.954 | 0.872 | 1.044 | 3.04e-01 | 1.00e+00 | 2624 | 2019 | 605 |
| European | CIDI-noMDD | Switching vs Continuation | Migraine | (Intercept) | 2.288 | 1.416 | 3.696 | 7.20e-04 | NA | 2664 | 2019 | 645 |
| European | CIDI-noMDD | Switching vs Continuation | Migraine | age_at_first_AD | 0.971 | 0.965 | 0.977 | <2.2e-16 | NA | 2664 | 2019 | 645 |
| European | CIDI-noMDD | Switching vs Continuation | Migraine | sex_at_birthMale | 1.321 | 1.084 | 1.608 | 5.68e-03 | NA | 2664 | 2019 | 645 |
| European | CIDI-noMDD | Switching vs Continuation | Migraine | Income_quantitative | 0.890 | 0.848 | 0.935 | 3.32e-06 | NA | 2664 | 2019 | 645 |
| European | CIDI-noMDD | Switching vs Continuation | Migraine | Education_levelCollege Undergraduate | 1.123 | 0.910 | 1.385 | 2.81e-01 | NA | 2664 | 2019 | 645 |
| European | CIDI-noMDD | Switching vs Continuation | Migraine | Education_levelLess than a high school degree | 1.434 | 0.450 | 4.572 | 5.43e-01 | NA | 2664 | 2019 | 645 |
| European | CIDI-noMDD | Switching vs Continuation | Migraine | Education_levelTwelve or GED | 1.279 | 0.904 | 1.809 | 1.64e-01 | NA | 2664 | 2019 | 645 |
| European | CIDI-noMDD | Switching vs Continuation | Migraine | Migraine | 0.995 | 0.910 | 1.088 | 9.09e-01 | 1.00e+00 | 2664 | 2019 | 645 |
| European | CIDI-noMDD | Discontinuation vs Continuation | Schizophrenia | (Intercept) | 0.106 | 0.062 | 0.183 | 7.47e-16 | NA | 2624 | 2019 | 605 |
| European | CIDI-noMDD | Discontinuation vs Continuation | Schizophrenia | age_at_first_AD | 1.015 | 1.008 | 1.022 | 3.99e-05 | NA | 2624 | 2019 | 605 |
| European | CIDI-noMDD | Discontinuation vs Continuation | Schizophrenia | sex_at_birthMale | 1.217 | 1.003 | 1.477 | 4.70e-02 | NA | 2624 | 2019 | 605 |
| European | CIDI-noMDD | Discontinuation vs Continuation | Schizophrenia | Income_quantitative | 0.983 | 0.934 | 1.034 | 5.04e-01 | NA | 2624 | 2019 | 605 |
| European | CIDI-noMDD | Discontinuation vs Continuation | Schizophrenia | Education_levelCollege Undergraduate | 1.375 | 1.115 | 1.696 | 2.95e-03 | NA | 2624 | 2019 | 605 |
| European | CIDI-noMDD | Discontinuation vs Continuation | Schizophrenia | Education_levelLess than a high school degree | 0.519 | 0.064 | 4.223 | 5.40e-01 | NA | 2624 | 2019 | 605 |
| European | CIDI-noMDD | Discontinuation vs Continuation | Schizophrenia | Education_levelTwelve or GED | 1.326 | 0.917 | 1.919 | 1.34e-01 | NA | 2624 | 2019 | 605 |
| European | CIDI-noMDD | Discontinuation vs Continuation | Schizophrenia | Schizophrenia | 1.022 | 0.931 | 1.123 | 6.44e-01 | 1.00e+00 | 2624 | 2019 | 605 |
| European | CIDI-noMDD | Switching vs Continuation | Schizophrenia | (Intercept) | 2.274 | 1.408 | 3.674 | 7.88e-04 | NA | 2664 | 2019 | 645 |
| European | CIDI-noMDD | Switching vs Continuation | Schizophrenia | age_at_first_AD | 0.971 | 0.965 | 0.977 | <2.2e-16 | NA | 2664 | 2019 | 645 |
| European | CIDI-noMDD | Switching vs Continuation | Schizophrenia | sex_at_birthMale | 1.324 | 1.087 | 1.612 | 5.32e-03 | NA | 2664 | 2019 | 645 |
| European | CIDI-noMDD | Switching vs Continuation | Schizophrenia | Income_quantitative | 0.891 | 0.848 | 0.935 | 3.46e-06 | NA | 2664 | 2019 | 645 |
| European | CIDI-noMDD | Switching vs Continuation | Schizophrenia | Education_levelCollege Undergraduate | 1.121 | 0.909 | 1.384 | 2.86e-01 | NA | 2664 | 2019 | 645 |
| European | CIDI-noMDD | Switching vs Continuation | Schizophrenia | Education_levelLess than a high school degree | 1.420 | 0.444 | 4.536 | 5.54e-01 | NA | 2664 | 2019 | 645 |
| European | CIDI-noMDD | Switching vs Continuation | Schizophrenia | Education_levelTwelve or GED | 1.281 | 0.905 | 1.811 | 1.62e-01 | NA | 2664 | 2019 | 645 |
| European | CIDI-noMDD | Switching vs Continuation | Schizophrenia | Schizophrenia | 1.056 | 0.961 | 1.160 | 2.57e-01 | 1.00e+00 | 2664 | 2019 | 645 |

| | | | | | | | | | | | | |
|---|---|---|---|---|---|---|---|---|---|---|---|---|
| African | Full Cohort | Discontinuation vs Continuation | ADHD | (Intercept) | 0.402 | 0.294 | 0.550 | 1.28e-08 | NA | 5972 | 4164 | 1808 |
| African | Full Cohort | Discontinuation vs Continuation | ADHD | age_at_first_AD | 0.999 | 0.995 | 1.003 | 7.18e-01 | NA | 5972 | 4164 | 1808 |
| African | Full Cohort | Discontinuation vs Continuation | ADHD | sex_at_birthMale | 1.215 | 1.073 | 1.376 | 2.07e-03 | NA | 5972 | 4164 | 1808 |
| African | Full Cohort | Discontinuation vs Continuation | ADHD | Income_quantitative | 0.982 | 0.945 | 1.020 | 3.40e-01 | NA | 5972 | 4164 | 1808 |
| African | Full Cohort | Discontinuation vs Continuation | ADHD | Education_levelCollege Undergraduate | 1.119 | 0.910 | 1.376 | 2.87e-01 | NA | 5972 | 4164 | 1808 |
| African | Full Cohort | Discontinuation vs Continuation | ADHD | Education_levelLess than a high school degree | 1.043 | 0.793 | 1.374 | 7.62e-01 | NA | 5972 | 4164 | 1808 |
| African | Full Cohort | Discontinuation vs Continuation | ADHD | Education_levelTwelve or GED | 1.138 | 0.902 | 1.435 | 2.75e-01 | NA | 5972 | 4164 | 1808 |
| African | Full Cohort | Discontinuation vs Continuation | ADHD | ADHD | 1.024 | 0.947 | 1.107 | 5.57e-01 | 1.00e+00 | 5972 | 4164 | 1808 |
| African | Full Cohort | Switching vs Continuation | ADHD | (Intercept) | 2.077 | 1.566 | 2.755 | 3.97e-07 | NA | 6693 | 4164 | 2529 |
| African | Full Cohort | Switching vs Continuation | ADHD | age_at_first_AD | 0.981 | 0.978 | 0.985 | <2.2e-16 | NA | 6693 | 4164 | 2529 |
| African | Full Cohort | Switching vs Continuation | ADHD | sex_at_birthMale | 0.936 | 0.832 | 1.053 | 2.70e-01 | NA | 6693 | 4164 | 2529 |
| African | Full Cohort | Switching vs Continuation | ADHD | Income_quantitative | 0.899 | 0.868 | 0.932 | 4.80e-09 | NA | 6693 | 4164 | 2529 |
| African | Full Cohort | Switching vs Continuation | ADHD | Education_levelCollege Undergraduate | 0.919 | 0.764 | 1.105 | 3.70e-01 | NA | 6693 | 4164 | 2529 |
| African | Full Cohort | Switching vs Continuation | ADHD | Education_levelLess than a high school degree | 0.918 | 0.720 | 1.171 | 4.91e-01 | NA | 6693 | 4164 | 2529 |
| African | Full Cohort | Switching vs Continuation | ADHD | Education_levelTwelve or GED | 0.848 | 0.689 | 1.043 | 1.18e-01 | NA | 6693 | 4164 | 2529 |
| African | Full Cohort | Switching vs Continuation | ADHD | ADHD | 1.038 | 0.967 | 1.115 | 3.04e-01 | 1.00e+00 | 6693 | 4164 | 2529 |
| African | Full Cohort | Discontinuation vs Continuation | BMI | (Intercept) | 0.403 | 0.295 | 0.552 | 1.44e-08 | NA | 5972 | 4164 | 1808 |
| African | Full Cohort | Discontinuation vs Continuation | BMI | age_at_first_AD | 0.999 | 0.995 | 1.003 | 6.88e-01 | NA | 5972 | 4164 | 1808 |
| African | Full Cohort | Discontinuation vs Continuation | BMI | sex_at_birthMale | 1.214 | 1.072 | 1.374 | 2.19e-03 | NA | 5972 | 4164 | 1808 |
| African | Full Cohort | Discontinuation vs Continuation | BMI | Income_quantitative | 0.981 | 0.944 | 1.019 | 3.16e-01 | NA | 5972 | 4164 | 1808 |
| African | Full Cohort | Discontinuation vs Continuation | BMI | Education_levelCollege Undergraduate | 1.122 | 0.913 | 1.381 | 2.74e-01 | NA | 5972 | 4164 | 1808 |
| African | Full Cohort | Discontinuation vs Continuation | BMI | Education_levelLess than a high school degree | 1.049 | 0.797 | 1.381 | 7.31e-01 | NA | 5972 | 4164 | 1808 |
| African | Full Cohort | Discontinuation vs Continuation | BMI | Education_levelTwelve or GED | 1.143 | 0.906 | 1.441 | 2.61e-01 | NA | 5972 | 4164 | 1808 |
| African | Full Cohort | Discontinuation vs Continuation | BMI | BMI | 0.978 | 0.909 | 1.053 | 5.54e-01 | 1.00e+00 | 5972 | 4164 | 1808 |
| African | Full Cohort | Switching vs Continuation | BMI | (Intercept) | 2.083 | 1.570 | 2.764 | 3.61e-07 | NA | 6693 | 4164 | 2529 |
| African | Full Cohort | Switching vs Continuation | BMI | age_at_first_AD | 0.981 | 0.977 | 0.985 | <2.2e-16 | NA | 6693 | 4164 | 2529 |
| African | Full Cohort | Switching vs Continuation | BMI | sex_at_birthMale | 0.933 | 0.829 | 1.049 | 2.47e-01 | NA | 6693 | 4164 | 2529 |
| African | Full Cohort | Switching vs Continuation | BMI | Income_quantitative | 0.898 | 0.867 | 0.931 | 3.35e-09 | NA | 6693 | 4164 | 2529 |
| African | Full Cohort | Switching vs Continuation | BMI | Education_levelCollege Undergraduate | 0.924 | 0.768 | 1.111 | 4.01e-01 | NA | 6693 | 4164 | 2529 |
| African | Full Cohort | Switching vs Continuation | BMI | Education_levelLess than a high school degree | 0.927 | 0.727 | 1.182 | 5.42e-01 | NA | 6693 | 4164 | 2529 |
| African | Full Cohort | Switching vs Continuation | BMI | Education_levelTwelve or GED | 0.854 | 0.695 | 1.050 | 1.35e-01 | NA | 6693 | 4164 | 2529 |
| African | Full Cohort | Switching vs Continuation | BMI | BMI | 0.964 | 0.902 | 1.029 | 2.71e-01 | 1.00e+00 | 6693 | 4164 | 2529 |
| African | Full Cohort | Discontinuation vs Continuation | Bipolar Disorder | (Intercept) | 0.401 | 0.293 | 0.549 | 1.18e-08 | NA | 5972 | 4164 | 1808 |
| African | Full Cohort | Discontinuation vs Continuation | Bipolar Disorder | age_at_first_AD | 0.999 | 0.995 | 1.003 | 7.20e-01 | NA | 5972 | 4164 | 1808 |
| African | Full Cohort | Discontinuation vs Continuation | Bipolar Disorder | sex_at_birthMale | 1.214 | 1.072 | 1.374 | 2.19e-03 | NA | 5972 | 4164 | 1808 |
| African | Full Cohort | Discontinuation vs Continuation | Bipolar Disorder | Income_quantitative | 0.981 | 0.945 | 1.019 | 3.29e-01 | NA | 5972 | 4164 | 1808 |
| African | Full Cohort | Discontinuation vs Continuation | Bipolar Disorder | Education_levelCollege Undergraduate | 1.122 | 0.912 | 1.380 | 2.76e-01 | NA | 5972 | 4164 | 1808 |
| African | Full Cohort | Discontinuation vs Continuation | Bipolar Disorder | Education_levelLess than a high school degree | 1.048 | 0.797 | 1.380 | 7.36e-01 | NA | 5972 | 4164 | 1808 |
| African | Full Cohort | Discontinuation vs Continuation | Bipolar Disorder | Education_levelTwelve or GED | 1.145 | 0.908 | 1.444 | 2.54e-01 | NA | 5972 | 4164 | 1808 |
| African | Full Cohort | Discontinuation vs Continuation | Bipolar Disorder | Bipolar Disorder | 1.064 | 0.982 | 1.153 | 1.31e-01 | 9.15e-01 | 5972 | 4164 | 1808 |
| African | Full Cohort | Switching vs Continuation | Bipolar Disorder | (Intercept) | 2.070 | 1.561 | 2.747 | 4.49e-07 | NA | 6693 | 4164 | 2529 |
| African | Full Cohort | Switching vs Continuation | Bipolar Disorder | age_at_first_AD | 0.981 | 0.977 | 0.985 | <2.2e-16 | NA | 6693 | 4164 | 2529 |
| African | Full Cohort | Switching vs Continuation | Bipolar Disorder | sex_at_birthMale | 0.935 | 0.831 | 1.051 | 2.59e-01 | NA | 6693 | 4164 | 2529 |
| African | Full Cohort | Switching vs Continuation | Bipolar Disorder | Income_quantitative | 0.900 | 0.869 | 0.932 | 5.72e-09 | NA | 6693 | 4164 | 2529 |
| African | Full Cohort | Switching vs Continuation | Bipolar Disorder | Education_levelCollege Undergraduate | 0.925 | 0.769 | 1.112 | 4.08e-01 | NA | 6693 | 4164 | 2529 |
| African | Full Cohort | Switching vs Continuation | Bipolar Disorder | Education_levelLess than a high school degree | 0.926 | 0.727 | 1.180 | 5.35e-01 | NA | 6693 | 4164 | 2529 |
| African | Full Cohort | Switching vs Continuation | Bipolar Disorder | Education_levelTwelve or GED | 0.857 | 0.697 | 1.053 | 1.42e-01 | NA | 6693 | 4164 | 2529 |
| African | Full Cohort | Switching vs Continuation | Bipolar Disorder | Bipolar Disorder | 1.085 | 1.009 | 1.167 | 2.86e-02 | 2.00e-01 | 6693 | 4164 | 2529 |
| African | Full Cohort | Discontinuation vs Continuation | Depression | (Intercept) | 0.403 | 0.294 | 0.551 | 1.38e-08 | NA | 5972 | 4164 | 1808 |
| African | Full Cohort | Discontinuation vs Continuation | Depression | age_at_first_AD | 0.999 | 0.995 | 1.003 | 7.03e-01 | NA | 5972 | 4164 | 1808 |
| African | Full Cohort | Discontinuation vs Continuation | Depression | sex_at_birthMale | 1.214 | 1.072 | 1.374 | 2.19e-03 | NA | 5972 | 4164 | 1808 |
| African | Full Cohort | Discontinuation vs Continuation | Depression | Income_quantitative | 0.981 | 0.944 | 1.019 | 3.13e-01 | NA | 5972 | 4164 | 1808 |
| African | Full Cohort | Discontinuation vs Continuation | Depression | Education_levelCollege Undergraduate | 1.121 | 0.911 | 1.378 | 2.80e-01 | NA | 5972 | 4164 | 1808 |
| African | Full Cohort | Discontinuation vs Continuation | Depression | Education_levelLess than a high school degree | 1.048 | 0.796 | 1.380 | 7.37e-01 | NA | 5972 | 4164 | 1808 |
| African | Full Cohort | Discontinuation vs Continuation | Depression | Education_levelTwelve or GED | 1.140 | 0.904 | 1.438 | 2.69e-01 | NA | 5972 | 4164 | 1808 |
| African | Full Cohort | Discontinuation vs Continuation | Depression | Depression | 0.965 | 0.893 | 1.043 | 3.65e-01 | 1.00e+00 | 5972 | 4164 | 1808 |
| African | Full Cohort | Switching vs Continuation | Depression | (Intercept) | 2.067 | 1.558 | 2.743 | 4.78e-07 | NA | 6693 | 4164 | 2529 |

| | | | | | | | | | | | | |
|---|---|---|---|---|---|---|---|---|---|---|---|---|
| African | Full Cohort | Switching vs Continuation | Depression | age_at_first_AD | 0.981 | 0.978 | 0.985 | <2.2e-16 | NA | 6693 | 4164 | 2529 |
| African | Full Cohort | Switching vs Continuation | Depression | sex_at_birthMale | 0.935 | 0.831 | 1.052 | 2.64e-01 | NA | 6693 | 4164 | 2529 |
| African | Full Cohort | Switching vs Continuation | Depression | Income_quantitative | 0.901 | 0.869 | 0.933 | 7.70e-09 | NA | 6693 | 4164 | 2529 |
| African | Full Cohort | Switching vs Continuation | Depression | Education_levelCollege Undergraduate | 0.920 | 0.765 | 1.107 | 3.77e-01 | NA | 6693 | 4164 | 2529 |
| African | Full Cohort | Switching vs Continuation | Depression | Education_levelLess than a high school degree | 0.921 | 0.722 | 1.174 | 5.05e-01 | NA | 6693 | 4164 | 2529 |
| African | Full Cohort | Switching vs Continuation | Depression | Education_levelTwelve or GED | 0.851 | 0.692 | 1.046 | 1.26e-01 | NA | 6693 | 4164 | 2529 |
| African | Full Cohort | Switching vs Continuation | Depression | Depression | 1.102 | 1.027 | 1.183 | 6.88e-03 | 4.82e-02 | 6693 | 4164 | 2529 |
| African | Full Cohort | Discontinuation vs Continuation | Insomnia | (Intercept) | 0.403 | 0.294 | 0.552 | 1.43e-08 | NA | 5972 | 4164 | 1808 |
| African | Full Cohort | Discontinuation vs Continuation | Insomnia | age_at_first_AD | 0.999 | 0.995 | 1.003 | 7.13e-01 | NA | 5972 | 4164 | 1808 |
| African | Full Cohort | Discontinuation vs Continuation | Insomnia | sex_at_birthMale | 1.214 | 1.072 | 1.374 | 2.22e-03 | NA | 5972 | 4164 | 1808 |
| African | Full Cohort | Discontinuation vs Continuation | Insomnia | Income_quantitative | 0.982 | 0.946 | 1.020 | 3.49e-01 | NA | 5972 | 4164 | 1808 |
| African | Full Cohort | Discontinuation vs Continuation | Insomnia | Education_levelCollege Undergraduate | 1.117 | 0.908 | 1.374 | 2.95e-01 | NA | 5972 | 4164 | 1808 |
| African | Full Cohort | Discontinuation vs Continuation | Insomnia | Education_levelLess than a high school degree | 1.036 | 0.787 | 1.365 | 7.99e-01 | NA | 5972 | 4164 | 1808 |
| African | Full Cohort | Discontinuation vs Continuation | Insomnia | Education_levelTwelve or GED | 1.137 | 0.902 | 1.434 | 2.77e-01 | NA | 5972 | 4164 | 1808 |
| African | Full Cohort | Discontinuation vs Continuation | Insomnia | Insomnia | 1.088 | 1.003 | 1.180 | 4.13e-02 | 2.89e-01 | 5972 | 4164 | 1808 |
| African | Full Cohort | Switching vs Continuation | Insomnia | (Intercept) | 2.090 | 1.575 | 2.774 | 3.24e-07 | NA | 6693 | 4164 | 2529 |
| African | Full Cohort | Switching vs Continuation | Insomnia | age_at_first_AD | 0.981 | 0.977 | 0.985 | <2.2e-16 | NA | 6693 | 4164 | 2529 |
| African | Full Cohort | Switching vs Continuation | Insomnia | sex_at_birthMale | 0.936 | 0.832 | 1.053 | 2.71e-01 | NA | 6693 | 4164 | 2529 |
| African | Full Cohort | Switching vs Continuation | Insomnia | Income_quantitative | 0.899 | 0.868 | 0.932 | 4.79e-09 | NA | 6693 | 4164 | 2529 |
| African | Full Cohort | Switching vs Continuation | Insomnia | Education_levelCollege Undergraduate | 0.916 | 0.762 | 1.102 | 3.54e-01 | NA | 6693 | 4164 | 2529 |
| African | Full Cohort | Switching vs Continuation | Insomnia | Education_levelLess than a high school degree | 0.913 | 0.716 | 1.164 | 4.61e-01 | NA | 6693 | 4164 | 2529 |
| African | Full Cohort | Switching vs Continuation | Insomnia | Education_levelTwelve or GED | 0.846 | 0.688 | 1.041 | 1.14e-01 | NA | 6693 | 4164 | 2529 |
| African | Full Cohort | Switching vs Continuation | Insomnia | Insomnia | 1.134 | 1.054 | 1.220 | 7.32e-04 | 5.12e-03 | 6693 | 4164 | 2529 |
| African | Full Cohort | Discontinuation vs Continuation | Migraine | (Intercept) | 0.403 | 0.294 | 0.551 | 1.34e-08 | NA | 5972 | 4164 | 1808 |
| African | Full Cohort | Discontinuation vs Continuation | Migraine | age_at_first_AD | 0.999 | 0.995 | 1.003 | 7.04e-01 | NA | 5972 | 4164 | 1808 |
| African | Full Cohort | Discontinuation vs Continuation | Migraine | sex_at_birthMale | 1.215 | 1.073 | 1.375 | 2.09e-03 | NA | 5972 | 4164 | 1808 |
| African | Full Cohort | Discontinuation vs Continuation | Migraine | Income_quantitative | 0.981 | 0.945 | 1.019 | 3.28e-01 | NA | 5972 | 4164 | 1808 |
| African | Full Cohort | Discontinuation vs Continuation | Migraine | Education_levelCollege Undergraduate | 1.121 | 0.911 | 1.378 | 2.80e-01 | NA | 5972 | 4164 | 1808 |
| African | Full Cohort | Discontinuation vs Continuation | Migraine | Education_levelLess than a high school degree | 1.046 | 0.795 | 1.377 | 7.46e-01 | NA | 5972 | 4164 | 1808 |
| African | Full Cohort | Discontinuation vs Continuation | Migraine | Education_levelTwelve or GED | 1.140 | 0.904 | 1.437 | 2.69e-01 | NA | 5972 | 4164 | 1808 |
| African | Full Cohort | Discontinuation vs Continuation | Migraine | Migraine | 1.001 | 0.936 | 1.071 | 9.74e-01 | 1.00e+00 | 5972 | 4164 | 1808 |
| African | Full Cohort | Switching vs Continuation | Migraine | (Intercept) | 2.077 | 1.566 | 2.756 | 3.95e-07 | NA | 6693 | 4164 | 2529 |
| African | Full Cohort | Switching vs Continuation | Migraine | age_at_first_AD | 0.981 | 0.978 | 0.985 | <2.2e-16 | NA | 6693 | 4164 | 2529 |
| African | Full Cohort | Switching vs Continuation | Migraine | sex_at_birthMale | 0.936 | 0.832 | 1.052 | 2.67e-01 | NA | 6693 | 4164 | 2529 |
| African | Full Cohort | Switching vs Continuation | Migraine | Income_quantitative | 0.899 | 0.868 | 0.931 | 4.02e-09 | NA | 6693 | 4164 | 2529 |
| African | Full Cohort | Switching vs Continuation | Migraine | Education_levelCollege Undergraduate | 0.921 | 0.766 | 1.108 | 3.84e-01 | NA | 6693 | 4164 | 2529 |
| African | Full Cohort | Switching vs Continuation | Migraine | Education_levelLess than a high school degree | 0.923 | 0.724 | 1.176 | 5.18e-01 | NA | 6693 | 4164 | 2529 |
| African | Full Cohort | Switching vs Continuation | Migraine | Education_levelTwelve or GED | 0.851 | 0.692 | 1.046 | 1.24e-01 | NA | 6693 | 4164 | 2529 |
| African | Full Cohort | Switching vs Continuation | Migraine | Migraine | 0.991 | 0.932 | 1.053 | 7.65e-01 | 1.00e+00 | 6693 | 4164 | 2529 |
| African | Full Cohort | Discontinuation vs Continuation | Schizophrenia | (Intercept) | 0.403 | 0.294 | 0.552 | 1.40e-08 | NA | 5972 | 4164 | 1808 |
| African | Full Cohort | Discontinuation vs Continuation | Schizophrenia | age_at_first_AD | 0.999 | 0.995 | 1.003 | 7.00e-01 | NA | 5972 | 4164 | 1808 |
| African | Full Cohort | Discontinuation vs Continuation | Schizophrenia | sex_at_birthMale | 1.215 | 1.073 | 1.375 | 2.10e-03 | NA | 5972 | 4164 | 1808 |
| African | Full Cohort | Discontinuation vs Continuation | Schizophrenia | Income_quantitative | 0.981 | 0.945 | 1.019 | 3.21e-01 | NA | 5972 | 4164 | 1808 |
| African | Full Cohort | Discontinuation vs Continuation | Schizophrenia | Education_levelCollege Undergraduate | 1.121 | 0.911 | 1.378 | 2.81e-01 | NA | 5972 | 4164 | 1808 |
| African | Full Cohort | Discontinuation vs Continuation | Schizophrenia | Education_levelLess than a high school degree | 1.047 | 0.796 | 1.378 | 7.42e-01 | NA | 5972 | 4164 | 1808 |
| African | Full Cohort | Discontinuation vs Continuation | Schizophrenia | Education_levelTwelve or GED | 1.140 | 0.904 | 1.437 | 2.69e-01 | NA | 5972 | 4164 | 1808 |
| African | Full Cohort | Discontinuation vs Continuation | Schizophrenia | Schizophrenia | 0.981 | 0.915 | 1.052 | 5.95e-01 | 1.00e+00 | 5972 | 4164 | 1808 |
| African | Full Cohort | Switching vs Continuation | Schizophrenia | (Intercept) | 2.081 | 1.569 | 2.761 | 3.72e-07 | NA | 6693 | 4164 | 2529 |
| African | Full Cohort | Switching vs Continuation | Schizophrenia | age_at_first_AD | 0.981 | 0.978 | 0.985 | <2.2e-16 | NA | 6693 | 4164 | 2529 |
| African | Full Cohort | Switching vs Continuation | Schizophrenia | sex_at_birthMale | 0.936 | 0.832 | 1.053 | 2.69e-01 | NA | 6693 | 4164 | 2529 |
| African | Full Cohort | Switching vs Continuation | Schizophrenia | Income_quantitative | 0.899 | 0.867 | 0.931 | 3.51e-09 | NA | 6693 | 4164 | 2529 |
| African | Full Cohort | Switching vs Continuation | Schizophrenia | Education_levelCollege Undergraduate | 0.921 | 0.766 | 1.107 | 3.79e-01 | NA | 6693 | 4164 | 2529 |
| African | Full Cohort | Switching vs Continuation | Schizophrenia | Education_levelLess than a high school degree | 0.923 | 0.724 | 1.176 | 5.17e-01 | NA | 6693 | 4164 | 2529 |
| African | Full Cohort | Switching vs Continuation | Schizophrenia | Education_levelTwelve or GED | 0.849 | 0.691 | 1.044 | 1.21e-01 | NA | 6693 | 4164 | 2529 |
| African | Full Cohort | Switching vs Continuation | Schizophrenia | Schizophrenia | 0.975 | 0.915 | 1.039 | 4.39e-01 | 1.00e+00 | 6693 | 4164 | 2529 |
| African | Recorded Depression | Discontinuation vs Continuation | ADHD | (Intercept) | 0.590 | 0.349 | 0.997 | 4.86e-02 | NA | 2367 | 1728 | 639 |
| African | Recorded Depression | Discontinuation vs Continuation | ADHD | age_at_first_AD | 0.993 | 0.987 | 1.000 | 4.76e-02 | NA | 2367 | 1728 | 639 |

| | | | | | | | | | | | | |
|---|---|---|---|---|---|---|---|---|---|---|---|---|
| African | Recorded Depression | Discontinuation vs Continuation | ADHD | sex_at_birthMale | 1.253 | 1.015 | 1.546 | 3.59e-02 | NA | 2367 | 1728 | 639 |
| African | Recorded Depression | Discontinuation vs Continuation | ADHD | Income_quantitative | 0.921 | 0.862 | 0.984 | 1.53e-02 | NA | 2367 | 1728 | 639 |
| African | Recorded Depression | Discontinuation vs Continuation | ADHD | Education_levelCollege Undergraduate | 0.944 | 0.661 | 1.348 | 7.52e-01 | NA | 2367 | 1728 | 639 |
| African | Recorded Depression | Discontinuation vs Continuation | ADHD | Education_levelLess than a high school degree | 0.914 | 0.573 | 1.459 | 7.06e-01 | NA | 2367 | 1728 | 639 |
| African | Recorded Depression | Discontinuation vs Continuation | ADHD | Education_levelTwelve or GED | 0.973 | 0.656 | 1.441 | 8.90e-01 | NA | 2367 | 1728 | 639 |
| African | Recorded Depression | Discontinuation vs Continuation | ADHD | ADHD | 1.024 | 0.900 | 1.165 | 7.24e-01 | 1.00e+00 | 2367 | 1728 | 639 |
| African | Recorded Depression | Switching vs Continuation | ADHD | (Intercept) | 2.710 | 1.863 | 3.941 | 1.83e-07 | NA | 3605 | 1728 | 1877 |
| African | Recorded Depression | Switching vs Continuation | ADHD | age_at_first_AD | 0.989 | 0.984 | 0.994 | 8.97e-06 | NA | 3605 | 1728 | 1877 |
| African | Recorded Depression | Switching vs Continuation | ADHD | sex_at_birthMale | 1.039 | 0.888 | 1.216 | 6.32e-01 | NA | 3605 | 1728 | 1877 |
| African | Recorded Depression | Switching vs Continuation | ADHD | Income_quantitative | 0.897 | 0.856 | 0.941 | 6.21e-06 | NA | 3605 | 1728 | 1877 |
| African | Recorded Depression | Switching vs Continuation | ADHD | Education_levelCollege Undergraduate | 0.867 | 0.678 | 1.108 | 2.55e-01 | NA | 3605 | 1728 | 1877 |
| African | Recorded Depression | Switching vs Continuation | ADHD | Education_levelLess than a high school degree | 0.839 | 0.604 | 1.165 | 2.95e-01 | NA | 3605 | 1728 | 1877 |
| African | Recorded Depression | Switching vs Continuation | ADHD | Education_levelTwelve or GED | 0.755 | 0.574 | 0.993 | 4.43e-02 | NA | 3605 | 1728 | 1877 |
| African | Recorded Depression | Switching vs Continuation | ADHD | ADHD | 1.016 | 0.925 | 1.117 | 7.40e-01 | 1.00e+00 | 3605 | 1728 | 1877 |
| African | Recorded Depression | Discontinuation vs Continuation | BMI | (Intercept) | 0.590 | 0.349 | 0.996 | 4.83e-02 | NA | 2367 | 1728 | 639 |
| African | Recorded Depression | Discontinuation vs Continuation | BMI | age_at_first_AD | 0.993 | 0.987 | 1.000 | 4.53e-02 | NA | 2367 | 1728 | 639 |
| African | Recorded Depression | Discontinuation vs Continuation | BMI | sex_at_birthMale | 1.250 | 1.012 | 1.543 | 3.80e-02 | NA | 2367 | 1728 | 639 |
| African | Recorded Depression | Discontinuation vs Continuation | BMI | Income_quantitative | 0.920 | 0.861 | 0.984 | 1.43e-02 | NA | 2367 | 1728 | 639 |
| African | Recorded Depression | Discontinuation vs Continuation | BMI | Education_levelCollege Undergraduate | 0.951 | 0.665 | 1.358 | 7.81e-01 | NA | 2367 | 1728 | 639 |
| African | Recorded Depression | Discontinuation vs Continuation | BMI | Education_levelLess than a high school degree | 0.921 | 0.577 | 1.470 | 7.29e-01 | NA | 2367 | 1728 | 639 |
| African | Recorded Depression | Discontinuation vs Continuation | BMI | Education_levelTwelve or GED | 0.981 | 0.662 | 1.454 | 9.24e-01 | NA | 2367 | 1728 | 639 |
| African | Recorded Depression | Discontinuation vs Continuation | BMI | BMI | 0.969 | 0.859 | 1.094 | 6.14e-01 | 1.00e+00 | 2367 | 1728 | 639 |
| African | Recorded Depression | Switching vs Continuation | BMI | (Intercept) | 2.723 | 1.872 | 3.962 | 1.61e-07 | NA | 3605 | 1728 | 1877 |
| African | Recorded Depression | Switching vs Continuation | BMI | age_at_first_AD | 0.988 | 0.983 | 0.993 | 6.04e-06 | NA | 3605 | 1728 | 1877 |
| African | Recorded Depression | Switching vs Continuation | BMI | sex_at_birthMale | 1.031 | 0.880 | 1.206 | 7.08e-01 | NA | 3605 | 1728 | 1877 |
| African | Recorded Depression | Switching vs Continuation | BMI | Income_quantitative | 0.896 | 0.855 | 0.940 | 5.09e-06 | NA | 3605 | 1728 | 1877 |
| African | Recorded Depression | Switching vs Continuation | BMI | Education_levelCollege Undergraduate | 0.876 | 0.685 | 1.120 | 2.90e-01 | NA | 3605 | 1728 | 1877 |
| African | Recorded Depression | Switching vs Continuation | BMI | Education_levelLess than a high school degree | 0.852 | 0.614 | 1.182 | 3.37e-01 | NA | 3605 | 1728 | 1877 |
| African | Recorded Depression | Switching vs Continuation | BMI | Education_levelTwelve or GED | 0.765 | 0.581 | 1.006 | 5.50e-02 | NA | 3605 | 1728 | 1877 |
| African | Recorded Depression | Switching vs Continuation | BMI | BMI | 0.933 | 0.855 | 1.017 | 1.15e-01 | 8.02e-01 | 3605 | 1728 | 1877 |
| African | Recorded Depression | Discontinuation vs Continuation | Bipolar Disorder | (Intercept) | 0.588 | 0.348 | 0.993 | 4.71e-02 | NA | 2367 | 1728 | 639 |
| African | Recorded Depression | Discontinuation vs Continuation | Bipolar Disorder | age_at_first_AD | 0.993 | 0.987 | 1.000 | 4.63e-02 | NA | 2367 | 1728 | 639 |
| African | Recorded Depression | Discontinuation vs Continuation | Bipolar Disorder | sex_at_birthMale | 1.249 | 1.012 | 1.541 | 3.87e-02 | NA | 2367 | 1728 | 639 |
| African | Recorded Depression | Discontinuation vs Continuation | Bipolar Disorder | Income_quantitative | 0.921 | 0.862 | 0.984 | 1.53e-02 | NA | 2367 | 1728 | 639 |
| African | Recorded Depression | Discontinuation vs Continuation | Bipolar Disorder | Education_levelCollege Undergraduate | 0.950 | 0.665 | 1.357 | 7.78e-01 | NA | 2367 | 1728 | 639 |
| African | Recorded Depression | Discontinuation vs Continuation | Bipolar Disorder | Education_levelLess than a high school degree | 0.920 | 0.577 | 1.470 | 7.28e-01 | NA | 2367 | 1728 | 639 |
| African | Recorded Depression | Discontinuation vs Continuation | Bipolar Disorder | Education_levelTwelve or GED | 0.983 | 0.663 | 1.456 | 9.31e-01 | NA | 2367 | 1728 | 639 |
| African | Recorded Depression | Discontinuation vs Continuation | Bipolar Disorder | Bipolar Disorder | 1.085 | 0.951 | 1.238 | 2.23e-01 | 1.00e+00 | 2367 | 1728 | 639 |
| African | Recorded Depression | Switching vs Continuation | Bipolar Disorder | (Intercept) | 2.694 | 1.852 | 3.918 | 2.15e-07 | NA | 3605 | 1728 | 1877 |
| African | Recorded Depression | Switching vs Continuation | Bipolar Disorder | age_at_first_AD | 0.989 | 0.984 | 0.994 | 8.03e-06 | NA | 3605 | 1728 | 1877 |
| African | Recorded Depression | Switching vs Continuation | Bipolar Disorder | sex_at_birthMale | 1.034 | 0.884 | 1.211 | 6.73e-01 | NA | 3605 | 1728 | 1877 |
| African | Recorded Depression | Switching vs Continuation | Bipolar Disorder | Income_quantitative | 0.898 | 0.857 | 0.941 | 7.20e-06 | NA | 3605 | 1728 | 1877 |
| African | Recorded Depression | Switching vs Continuation | Bipolar Disorder | Education_levelCollege Undergraduate | 0.875 | 0.684 | 1.119 | 2.87e-01 | NA | 3605 | 1728 | 1877 |
| African | Recorded Depression | Switching vs Continuation | Bipolar Disorder | Education_levelLess than a high school degree | 0.846 | 0.609 | 1.174 | 3.16e-01 | NA | 3605 | 1728 | 1877 |
| African | Recorded Depression | Switching vs Continuation | Bipolar Disorder | Education_levelTwelve or GED | 0.763 | 0.580 | 1.003 | 5.25e-02 | NA | 3605 | 1728 | 1877 |
| African | Recorded Depression | Switching vs Continuation | Bipolar Disorder | Bipolar Disorder | 1.087 | 0.986 | 1.198 | 9.34e-02 | 6.54e-01 | 3605 | 1728 | 1877 |
| African | Recorded Depression | Discontinuation vs Continuation | Depression | (Intercept) | 0.590 | 0.349 | 0.997 | 4.86e-02 | NA | 2367 | 1728 | 639 |
| African | Recorded Depression | Discontinuation vs Continuation | Depression | age_at_first_AD | 0.993 | 0.987 | 1.000 | 4.66e-02 | NA | 2367 | 1728 | 639 |
| African | Recorded Depression | Discontinuation vs Continuation | Depression | sex_at_birthMale | 1.253 | 1.015 | 1.546 | 3.57e-02 | NA | 2367 | 1728 | 639 |
| African | Recorded Depression | Discontinuation vs Continuation | Depression | Income_quantitative | 0.921 | 0.862 | 0.984 | 1.50e-02 | NA | 2367 | 1728 | 639 |
| African | Recorded Depression | Discontinuation vs Continuation | Depression | Education_levelCollege Undergraduate | 0.946 | 0.663 | 1.351 | 7.61e-01 | NA | 2367 | 1728 | 639 |
| African | Recorded Depression | Discontinuation vs Continuation | Depression | Education_levelLess than a high school degree | 0.915 | 0.573 | 1.461 | 7.10e-01 | NA | 2367 | 1728 | 639 |
| African | Recorded Depression | Discontinuation vs Continuation | Depression | Education_levelTwelve or GED | 0.975 | 0.659 | 1.445 | 9.01e-01 | NA | 2367 | 1728 | 639 |
| African | Recorded Depression | Discontinuation vs Continuation | Depression | Depression | 1.013 | 0.888 | 1.154 | 8.52e-01 | 1.00e+00 | 2367 | 1728 | 639 |
| African | Recorded Depression | Switching vs Continuation | Depression | (Intercept) | 2.701 | 1.857 | 3.929 | 2.00e-07 | NA | 3605 | 1728 | 1877 |
| African | Recorded Depression | Switching vs Continuation | Depression | age_at_first_AD | 0.989 | 0.984 | 0.994 | 9.88e-06 | NA | 3605 | 1728 | 1877 |
| African | Recorded Depression | Switching vs Continuation | Depression | sex_at_birthMale | 1.038 | 0.887 | 1.215 | 6.43e-01 | NA | 3605 | 1728 | 1877 |

| | | | | | | | | | | | | |
|---|---|---|---|---|---|---|---|---|---|---|---|---|
| African | Recorded Depression | Switching vs Continuation | Depression | Income_quantitative | 0.899 | 0.857 | 0.942 | 8.34e-06 | NA | 3605 | 1728 | 1877 |
| African | Recorded Depression | Switching vs Continuation | Depression | Education_levelCollege Undergraduate | 0.865 | 0.677 | 1.106 | 2.48e-01 | NA | 3605 | 1728 | 1877 |
| African | Recorded Depression | Switching vs Continuation | Depression | Education_levelLess than a high school degree | 0.834 | 0.601 | 1.158 | 2.79e-01 | NA | 3605 | 1728 | 1877 |
| African | Recorded Depression | Switching vs Continuation | Depression | Education_levelTwelve or GED | 0.754 | 0.573 | 0.991 | 4.29e-02 | NA | 3605 | 1728 | 1877 |
| African | Recorded Depression | Switching vs Continuation | Depression | Depression | 1.106 | 1.007 | 1.215 | 3.60e-02 | 2.52e-01 | 3605 | 1728 | 1877 |
| African | Recorded Depression | Discontinuation vs Continuation | Insomnia | (Intercept) | 0.590 | 0.349 | 0.997 | 4.87e-02 | NA | 2367 | 1728 | 639 |
| African | Recorded Depression | Discontinuation vs Continuation | Insomnia | age_at_first_AD | 0.994 | 0.987 | 1.000 | 5.18e-02 | NA | 2367 | 1728 | 639 |
| African | Recorded Depression | Discontinuation vs Continuation | Insomnia | sex_at_birthMale | 1.249 | 1.012 | 1.542 | 3.84e-02 | NA | 2367 | 1728 | 639 |
| African | Recorded Depression | Discontinuation vs Continuation | Insomnia | Income_quantitative | 0.920 | 0.861 | 0.984 | 1.46e-02 | NA | 2367 | 1728 | 639 |
| African | Recorded Depression | Discontinuation vs Continuation | Insomnia | Education_levelCollege Undergraduate | 0.945 | 0.661 | 1.350 | 7.56e-01 | NA | 2367 | 1728 | 639 |
| African | Recorded Depression | Discontinuation vs Continuation | Insomnia | Education_levelLess than a high school degree | 0.897 | 0.561 | 1.433 | 6.49e-01 | NA | 2367 | 1728 | 639 |
| African | Recorded Depression | Discontinuation vs Continuation | Insomnia | Education_levelTwelve or GED | 0.971 | 0.655 | 1.439 | 8.84e-01 | NA | 2367 | 1728 | 639 |
| African | Recorded Depression | Discontinuation vs Continuation | Insomnia | Insomnia | 1.147 | 1.002 | 1.314 | 4.70e-02 | 3.29e-01 | 2367 | 1728 | 639 |
| African | Recorded Depression | Switching vs Continuation | Insomnia | (Intercept) | 2.725 | 1.873 | 3.964 | 1.60e-07 | NA | 3605 | 1728 | 1877 |
| African | Recorded Depression | Switching vs Continuation | Insomnia | age_at_first_AD | 0.989 | 0.984 | 0.994 | 9.18e-06 | NA | 3605 | 1728 | 1877 |
| African | Recorded Depression | Switching vs Continuation | Insomnia | sex_at_birthMale | 1.042 | 0.890 | 1.219 | 6.11e-01 | NA | 3605 | 1728 | 1877 |
| African | Recorded Depression | Switching vs Continuation | Insomnia | Income_quantitative | 0.897 | 0.856 | 0.940 | 5.94e-06 | NA | 3605 | 1728 | 1877 |
| African | Recorded Depression | Switching vs Continuation | Insomnia | Education_levelCollege Undergraduate | 0.863 | 0.675 | 1.104 | 2.42e-01 | NA | 3605 | 1728 | 1877 |
| African | Recorded Depression | Switching vs Continuation | Insomnia | Education_levelLess than a high school degree | 0.830 | 0.598 | 1.153 | 2.67e-01 | NA | 3605 | 1728 | 1877 |
| African | Recorded Depression | Switching vs Continuation | Insomnia | Education_levelTwelve or GED | 0.750 | 0.570 | 0.986 | 3.94e-02 | NA | 3605 | 1728 | 1877 |
| African | Recorded Depression | Switching vs Continuation | Insomnia | Insomnia | 1.127 | 1.024 | 1.241 | 1.46e-02 | 1.02e-01 | 3605 | 1728 | 1877 |
| African | Recorded Depression | Discontinuation vs Continuation | Migraine | (Intercept) | 0.591 | 0.349 | 0.998 | 4.91e-02 | NA | 2367 | 1728 | 639 |
| African | Recorded Depression | Discontinuation vs Continuation | Migraine | age_at_first_AD | 0.993 | 0.987 | 1.000 | 4.64e-02 | NA | 2367 | 1728 | 639 |
| African | Recorded Depression | Discontinuation vs Continuation | Migraine | sex_at_birthMale | 1.252 | 1.014 | 1.545 | 3.66e-02 | NA | 2367 | 1728 | 639 |
| African | Recorded Depression | Discontinuation vs Continuation | Migraine | Income_quantitative | 0.921 | 0.862 | 0.984 | 1.51e-02 | NA | 2367 | 1728 | 639 |
| African | Recorded Depression | Discontinuation vs Continuation | Migraine | Education_levelCollege Undergraduate | 0.945 | 0.662 | 1.350 | 7.57e-01 | NA | 2367 | 1728 | 639 |
| African | Recorded Depression | Discontinuation vs Continuation | Migraine | Education_levelLess than a high school degree | 0.917 | 0.574 | 1.463 | 7.15e-01 | NA | 2367 | 1728 | 639 |
| African | Recorded Depression | Discontinuation vs Continuation | Migraine | Education_levelTwelve or GED | 0.974 | 0.658 | 1.443 | 8.96e-01 | NA | 2367 | 1728 | 639 |
| African | Recorded Depression | Discontinuation vs Continuation | Migraine | Migraine | 1.021 | 0.912 | 1.143 | 7.19e-01 | 1.00e+00 | 2367 | 1728 | 639 |
| African | Recorded Depression | Switching vs Continuation | Migraine | (Intercept) | 2.707 | 1.861 | 3.937 | 1.87e-07 | NA | 3605 | 1728 | 1877 |
| African | Recorded Depression | Switching vs Continuation | Migraine | age_at_first_AD | 0.989 | 0.984 | 0.994 | 8.97e-06 | NA | 3605 | 1728 | 1877 |
| African | Recorded Depression | Switching vs Continuation | Migraine | sex_at_birthMale | 1.039 | 0.888 | 1.216 | 6.29e-01 | NA | 3605 | 1728 | 1877 |
| African | Recorded Depression | Switching vs Continuation | Migraine | Income_quantitative | 0.897 | 0.856 | 0.940 | 6.13e-06 | NA | 3605 | 1728 | 1877 |
| African | Recorded Depression | Switching vs Continuation | Migraine | Education_levelCollege Undergraduate | 0.868 | 0.678 | 1.109 | 2.57e-01 | NA | 3605 | 1728 | 1877 |
| African | Recorded Depression | Switching vs Continuation | Migraine | Education_levelLess than a high school degree | 0.841 | 0.606 | 1.167 | 2.99e-01 | NA | 3605 | 1728 | 1877 |
| African | Recorded Depression | Switching vs Continuation | Migraine | Education_levelTwelve or GED | 0.756 | 0.575 | 0.994 | 4.54e-02 | NA | 3605 | 1728 | 1877 |
| African | Recorded Depression | Switching vs Continuation | Migraine | Migraine | 0.990 | 0.913 | 1.073 | 7.98e-01 | 1.00e+00 | 3605 | 1728 | 1877 |
| African | Recorded Depression | Discontinuation vs Continuation | Schizophrenia | (Intercept) | 0.589 | 0.348 | 0.996 | 4.81e-02 | NA | 2367 | 1728 | 639 |
| African | Recorded Depression | Discontinuation vs Continuation | Schizophrenia | age_at_first_AD | 0.993 | 0.987 | 1.000 | 4.72e-02 | NA | 2367 | 1728 | 639 |
| African | Recorded Depression | Discontinuation vs Continuation | Schizophrenia | sex_at_birthMale | 1.253 | 1.015 | 1.546 | 3.59e-02 | NA | 2367 | 1728 | 639 |
| African | Recorded Depression | Discontinuation vs Continuation | Schizophrenia | Income_quantitative | 0.921 | 0.862 | 0.984 | 1.51e-02 | NA | 2367 | 1728 | 639 |
| African | Recorded Depression | Discontinuation vs Continuation | Schizophrenia | Education_levelCollege Undergraduate | 0.947 | 0.663 | 1.352 | 7.64e-01 | NA | 2367 | 1728 | 639 |
| African | Recorded Depression | Discontinuation vs Continuation | Schizophrenia | Education_levelLess than a high school degree | 0.917 | 0.574 | 1.463 | 7.15e-01 | NA | 2367 | 1728 | 639 |
| African | Recorded Depression | Discontinuation vs Continuation | Schizophrenia | Education_levelTwelve or GED | 0.976 | 0.659 | 1.446 | 9.04e-01 | NA | 2367 | 1728 | 639 |
| African | Recorded Depression | Discontinuation vs Continuation | Schizophrenia | Schizophrenia | 1.008 | 0.901 | 1.129 | 8.86e-01 | 1.00e+00 | 2367 | 1728 | 639 |
| African | Recorded Depression | Switching vs Continuation | Schizophrenia | (Intercept) | 2.717 | 1.868 | 3.951 | 1.71e-07 | NA | 3605 | 1728 | 1877 |
| African | Recorded Depression | Switching vs Continuation | Schizophrenia | age_at_first_AD | 0.989 | 0.984 | 0.994 | 8.71e-06 | NA | 3605 | 1728 | 1877 |
| African | Recorded Depression | Switching vs Continuation | Schizophrenia | sex_at_birthMale | 1.040 | 0.889 | 1.218 | 6.22e-01 | NA | 3605 | 1728 | 1877 |
| African | Recorded Depression | Switching vs Continuation | Schizophrenia | Income_quantitative | 0.897 | 0.856 | 0.940 | 5.60e-06 | NA | 3605 | 1728 | 1877 |
| African | Recorded Depression | Switching vs Continuation | Schizophrenia | Education_levelCollege Undergraduate | 0.866 | 0.677 | 1.107 | 2.50e-01 | NA | 3605 | 1728 | 1877 |
| African | Recorded Depression | Switching vs Continuation | Schizophrenia | Education_levelLess than a high school degree | 0.840 | 0.605 | 1.165 | 2.97e-01 | NA | 3605 | 1728 | 1877 |
| African | Recorded Depression | Switching vs Continuation | Schizophrenia | Education_levelTwelve or GED | 0.754 | 0.573 | 0.991 | 4.31e-02 | NA | 3605 | 1728 | 1877 |
| African | Recorded Depression | Switching vs Continuation | Schizophrenia | Schizophrenia | 0.956 | 0.879 | 1.039 | 2.89e-01 | 1.00e+00 | 3605 | 1728 | 1877 |
| African | No Recorded Depression | Discontinuation vs Continuation | ADHD | (Intercept) | 0.360 | 0.242 | 0.535 | 4.48e-07 | NA | 3605 | 2436 | 1169 |
| African | No Recorded Depression | Discontinuation vs Continuation | ADHD | age_at_first_AD | 1.001 | 0.996 | 1.006 | 6.88e-01 | NA | 3605 | 2436 | 1169 |
| African | No Recorded Depression | Discontinuation vs Continuation | ADHD | sex_at_birthMale | 1.179 | 1.010 | 1.375 | 3.69e-02 | NA | 3605 | 2436 | 1169 |
| African | No Recorded Depression | Discontinuation vs Continuation | ADHD | Income_quantitative | 1.007 | 0.961 | 1.055 | 7.81e-01 | NA | 3605 | 2436 | 1169 |

| | | | | | | | | | | | | |
|---|---|---|---|---|---|---|---|---|---|---|---|---|
| African | No Recorded Depression | Discontinuation vs Continuation | ADHD | Education_levelCollege Undergraduate | 1.224 | 0.949 | 1.580 | 1.20e-01 | NA | 3605 | 2436 | 1169 |
| African | No Recorded Depression | Discontinuation vs Continuation | ADHD | Education_levelLess than a high school degree | 1.100 | 0.782 | 1.548 | 5.83e-01 | NA | 3605 | 2436 | 1169 |
| African | No Recorded Depression | Discontinuation vs Continuation | ADHD | Education_levelTwelve or GED | 1.230 | 0.922 | 1.643 | 1.60e-01 | NA | 3605 | 2436 | 1169 |
| African | No Recorded Depression | Discontinuation vs Continuation | ADHD | ADHD | 1.026 | 0.930 | 1.133 | 6.06e-01 | 1.00e+00 | 3605 | 2436 | 1169 |
| African | No Recorded Depression | Switching vs Continuation | ADHD | (Intercept) | 0.819 | 0.503 | 1.335 | 4.23e-01 | NA | 3088 | 2436 | 652 |
| African | No Recorded Depression | Switching vs Continuation | ADHD | age_at_first_AD | 0.980 | 0.973 | 0.986 | 7.88e-10 | NA | 3088 | 2436 | 652 |
| African | No Recorded Depression | Switching vs Continuation | ADHD | sex_at_birthMale | 0.887 | 0.724 | 1.087 | 2.48e-01 | NA | 3088 | 2436 | 652 |
| African | No Recorded Depression | Switching vs Continuation | ADHD | Income_quantitative | 0.952 | 0.896 | 1.012 | 1.17e-01 | NA | 3088 | 2436 | 652 |
| African | No Recorded Depression | Switching vs Continuation | ADHD | Education_levelCollege Undergraduate | 0.919 | 0.669 | 1.264 | 6.05e-01 | NA | 3088 | 2436 | 652 |
| African | No Recorded Depression | Switching vs Continuation | ADHD | Education_levelLess than a high school degree | 1.281 | 0.851 | 1.929 | 2.35e-01 | NA | 3088 | 2436 | 652 |
| African | No Recorded Depression | Switching vs Continuation | ADHD | Education_levelTwelve or GED | 1.074 | 0.751 | 1.536 | 6.95e-01 | NA | 3088 | 2436 | 652 |
| African | No Recorded Depression | Switching vs Continuation | ADHD | ADHD | 1.089 | 0.961 | 1.233 | 1.81e-01 | 1.00e+00 | 3088 | 2436 | 652 |
| African | No Recorded Depression | Discontinuation vs Continuation | BMI | (Intercept) | 0.361 | 0.243 | 0.537 | 4.95e-07 | NA | 3605 | 2436 | 1169 |
| African | No Recorded Depression | Discontinuation vs Continuation | BMI | age_at_first_AD | 1.001 | 0.996 | 1.006 | 7.07e-01 | NA | 3605 | 2436 | 1169 |
| African | No Recorded Depression | Discontinuation vs Continuation | BMI | sex_at_birthMale | 1.178 | 1.010 | 1.375 | 3.73e-02 | NA | 3605 | 2436 | 1169 |
| African | No Recorded Depression | Discontinuation vs Continuation | BMI | Income_quantitative | 1.006 | 0.960 | 1.054 | 8.09e-01 | NA | 3605 | 2436 | 1169 |
| African | No Recorded Depression | Discontinuation vs Continuation | BMI | Education_levelCollege Undergraduate | 1.226 | 0.950 | 1.582 | 1.17e-01 | NA | 3605 | 2436 | 1169 |
| African | No Recorded Depression | Discontinuation vs Continuation | BMI | Education_levelLess than a high school degree | 1.105 | 0.786 | 1.554 | 5.66e-01 | NA | 3605 | 2436 | 1169 |
| African | No Recorded Depression | Discontinuation vs Continuation | BMI | Education_levelTwelve or GED | 1.232 | 0.923 | 1.645 | 1.57e-01 | NA | 3605 | 2436 | 1169 |
| African | No Recorded Depression | Discontinuation vs Continuation | BMI | BMI | 0.990 | 0.903 | 1.087 | 8.37e-01 | 1.00e+00 | 3605 | 2436 | 1169 |
| African | No Recorded Depression | Switching vs Continuation | BMI | (Intercept) | 0.828 | 0.508 | 1.349 | 4.49e-01 | NA | 3088 | 2436 | 652 |
| African | No Recorded Depression | Switching vs Continuation | BMI | age_at_first_AD | 0.980 | 0.973 | 0.986 | 6.25e-10 | NA | 3088 | 2436 | 652 |
| African | No Recorded Depression | Switching vs Continuation | BMI | sex_at_birthMale | 0.885 | 0.722 | 1.084 | 2.37e-01 | NA | 3088 | 2436 | 652 |
| African | No Recorded Depression | Switching vs Continuation | BMI | Income_quantitative | 0.950 | 0.894 | 1.010 | 1.00e-01 | NA | 3088 | 2436 | 652 |
| African | No Recorded Depression | Switching vs Continuation | BMI | Education_levelCollege Undergraduate | 0.921 | 0.670 | 1.265 | 6.10e-01 | NA | 3088 | 2436 | 652 |
| African | No Recorded Depression | Switching vs Continuation | BMI | Education_levelLess than a high school degree | 1.288 | 0.856 | 1.939 | 2.25e-01 | NA | 3088 | 2436 | 652 |
| African | No Recorded Depression | Switching vs Continuation | BMI | Education_levelTwelve or GED | 1.074 | 0.751 | 1.536 | 6.95e-01 | NA | 3088 | 2436 | 652 |
| African | No Recorded Depression | Switching vs Continuation | BMI | BMI | 1.007 | 0.897 | 1.130 | 9.04e-01 | 1.00e+00 | 3088 | 2436 | 652 |
| African | No Recorded Depression | Discontinuation vs Continuation | Bipolar Disorder | (Intercept) | 0.359 | 0.242 | 0.534 | 4.23e-07 | NA | 3605 | 2436 | 1169 |
| African | No Recorded Depression | Discontinuation vs Continuation | Bipolar Disorder | age_at_first_AD | 1.001 | 0.996 | 1.006 | 6.79e-01 | NA | 3605 | 2436 | 1169 |
| African | No Recorded Depression | Discontinuation vs Continuation | Bipolar Disorder | sex_at_birthMale | 1.178 | 1.010 | 1.375 | 3.72e-02 | NA | 3605 | 2436 | 1169 |
| African | No Recorded Depression | Discontinuation vs Continuation | Bipolar Disorder | Income_quantitative | 1.006 | 0.960 | 1.054 | 8.01e-01 | NA | 3605 | 2436 | 1169 |
| African | No Recorded Depression | Discontinuation vs Continuation | Bipolar Disorder | Education_levelCollege Undergraduate | 1.226 | 0.950 | 1.582 | 1.18e-01 | NA | 3605 | 2436 | 1169 |
| African | No Recorded Depression | Discontinuation vs Continuation | Bipolar Disorder | Education_levelLess than a high school degree | 1.105 | 0.786 | 1.554 | 5.67e-01 | NA | 3605 | 2436 | 1169 |
| African | No Recorded Depression | Discontinuation vs Continuation | Bipolar Disorder | Education_levelTwelve or GED | 1.235 | 0.925 | 1.650 | 1.52e-01 | NA | 3605 | 2436 | 1169 |
| African | No Recorded Depression | Discontinuation vs Continuation | Bipolar Disorder | Bipolar Disorder | 1.059 | 0.956 | 1.174 | 2.70e-01 | 1.00e+00 | 3605 | 2436 | 1169 |
| African | No Recorded Depression | Switching vs Continuation | Bipolar Disorder | (Intercept) | 0.826 | 0.507 | 1.346 | 4.43e-01 | NA | 3088 | 2436 | 652 |
| African | No Recorded Depression | Switching vs Continuation | Bipolar Disorder | age_at_first_AD | 0.979 | 0.973 | 0.986 | 5.71e-10 | NA | 3088 | 2436 | 652 |
| African | No Recorded Depression | Switching vs Continuation | Bipolar Disorder | sex_at_birthMale | 0.887 | 0.724 | 1.087 | 2.46e-01 | NA | 3088 | 2436 | 652 |
| African | No Recorded Depression | Switching vs Continuation | Bipolar Disorder | Income_quantitative | 0.951 | 0.895 | 1.011 | 1.08e-01 | NA | 3088 | 2436 | 652 |
| African | No Recorded Depression | Switching vs Continuation | Bipolar Disorder | Education_levelCollege Undergraduate | 0.920 | 0.669 | 1.265 | 6.09e-01 | NA | 3088 | 2436 | 652 |
| African | No Recorded Depression | Switching vs Continuation | Bipolar Disorder | Education_levelLess than a high school degree | 1.290 | 0.857 | 1.941 | 2.22e-01 | NA | 3088 | 2436 | 652 |
| African | No Recorded Depression | Switching vs Continuation | Bipolar Disorder | Education_levelTwelve or GED | 1.081 | 0.756 | 1.546 | 6.69e-01 | NA | 3088 | 2436 | 652 |
| African | No Recorded Depression | Switching vs Continuation | Bipolar Disorder | Bipolar Disorder | 1.073 | 0.945 | 1.218 | 2.76e-01 | 1.00e+00 | 3088 | 2436 | 652 |
| African | No Recorded Depression | Discontinuation vs Continuation | Depression | (Intercept) | 0.362 | 0.243 | 0.538 | 5.04e-07 | NA | 3605 | 2436 | 1169 |
| African | No Recorded Depression | Discontinuation vs Continuation | Depression | age_at_first_AD | 1.001 | 0.996 | 1.006 | 7.00e-01 | NA | 3605 | 2436 | 1169 |
| African | No Recorded Depression | Discontinuation vs Continuation | Depression | sex_at_birthMale | 1.176 | 1.008 | 1.373 | 3.92e-02 | NA | 3605 | 2436 | 1169 |
| African | No Recorded Depression | Discontinuation vs Continuation | Depression | Income_quantitative | 1.005 | 0.959 | 1.053 | 8.45e-01 | NA | 3605 | 2436 | 1169 |
| African | No Recorded Depression | Discontinuation vs Continuation | Depression | Education_levelCollege Undergraduate | 1.227 | 0.950 | 1.583 | 1.17e-01 | NA | 3605 | 2436 | 1169 |
| African | No Recorded Depression | Discontinuation vs Continuation | Depression | Education_levelLess than a high school degree | 1.105 | 0.786 | 1.554 | 5.65e-01 | NA | 3605 | 2436 | 1169 |
| African | No Recorded Depression | Discontinuation vs Continuation | Depression | Education_levelTwelve or GED | 1.230 | 0.921 | 1.642 | 1.60e-01 | NA | 3605 | 2436 | 1169 |
| African | No Recorded Depression | Discontinuation vs Continuation | Depression | Depression | 0.946 | 0.858 | 1.042 | 2.57e-01 | 1.00e+00 | 3605 | 2436 | 1169 |
| African | No Recorded Depression | Switching vs Continuation | Depression | (Intercept) | 0.825 | 0.506 | 1.344 | 4.40e-01 | NA | 3088 | 2436 | 652 |
| African | No Recorded Depression | Switching vs Continuation | Depression | age_at_first_AD | 0.979 | 0.973 | 0.986 | 5.76e-10 | NA | 3088 | 2436 | 652 |
| African | No Recorded Depression | Switching vs Continuation | Depression | sex_at_birthMale | 0.885 | 0.722 | 1.084 | 2.37e-01 | NA | 3088 | 2436 | 652 |
| African | No Recorded Depression | Switching vs Continuation | Depression | Income_quantitative | 0.952 | 0.895 | 1.011 | 1.11e-01 | NA | 3088 | 2436 | 652 |
| African | No Recorded Depression | Switching vs Continuation | Depression | Education_levelCollege Undergraduate | 0.921 | 0.670 | 1.267 | 6.14e-01 | NA | 3088 | 2436 | 652 |

| | | | | | | | | | | | | |
|---|---|---|---|---|---|---|---|---|---|---|---|---|
| African | No Recorded Depression | Switching vs Continuation | Depression | Education_levelLess than a high school degree | 1.292 | 0.858 | 1.945 | 2.19e-01 | NA | 3088 | 2436 | 652 |
| African | No Recorded Depression | Switching vs Continuation | Depression | Education_levelTwelve or GED | 1.079 | 0.754 | 1.542 | 6.78e-01 | NA | 3088 | 2436 | 652 |
| African | No Recorded Depression | Switching vs Continuation | Depression | Depression | 1.069 | 0.946 | 1.208 | 2.85e-01 | 1.00e+00 | 3088 | 2436 | 652 |
| African | No Recorded Depression | Discontinuation vs Continuation | Insomnia | (Intercept) | 0.362 | 0.243 | 0.538 | 5.07e-07 | NA | 3605 | 2436 | 1169 |
| African | No Recorded Depression | Discontinuation vs Continuation | Insomnia | age_at_first_AD | 1.001 | 0.996 | 1.006 | 7.10e-01 | NA | 3605 | 2436 | 1169 |
| African | No Recorded Depression | Discontinuation vs Continuation | Insomnia | sex_at_birthMale | 1.178 | 1.009 | 1.374 | 3.79e-02 | NA | 3605 | 2436 | 1169 |
| African | No Recorded Depression | Discontinuation vs Continuation | Insomnia | Income_quantitative | 1.007 | 0.961 | 1.055 | 7.71e-01 | NA | 3605 | 2436 | 1169 |
| African | No Recorded Depression | Discontinuation vs Continuation | Insomnia | Education_levelCollege Undergraduate | 1.221 | 0.946 | 1.577 | 1.25e-01 | NA | 3605 | 2436 | 1169 |
| African | No Recorded Depression | Discontinuation vs Continuation | Insomnia | Education_levelLess than a high school degree | 1.098 | 0.781 | 1.545 | 5.91e-01 | NA | 3605 | 2436 | 1169 |
| African | No Recorded Depression | Discontinuation vs Continuation | Insomnia | Education_levelTwelve or GED | 1.230 | 0.921 | 1.642 | 1.61e-01 | NA | 3605 | 2436 | 1169 |
| African | No Recorded Depression | Discontinuation vs Continuation | Insomnia | Insomnia | 1.063 | 0.960 | 1.177 | 2.40e-01 | 1.00e+00 | 3605 | 2436 | 1169 |
| African | No Recorded Depression | Switching vs Continuation | Insomnia | (Intercept) | 0.832 | 0.511 | 1.355 | 4.60e-01 | NA | 3088 | 2436 | 652 |
| African | No Recorded Depression | Switching vs Continuation | Insomnia | age_at_first_AD | 0.979 | 0.973 | 0.986 | 5.39e-10 | NA | 3088 | 2436 | 652 |
| African | No Recorded Depression | Switching vs Continuation | Insomnia | sex_at_birthMale | 0.883 | 0.721 | 1.082 | 2.31e-01 | NA | 3088 | 2436 | 652 |
| African | No Recorded Depression | Switching vs Continuation | Insomnia | Income_quantitative | 0.951 | 0.895 | 1.011 | 1.06e-01 | NA | 3088 | 2436 | 652 |
| African | No Recorded Depression | Switching vs Continuation | Insomnia | Education_levelCollege Undergraduate | 0.918 | 0.668 | 1.262 | 5.99e-01 | NA | 3088 | 2436 | 652 |
| African | No Recorded Depression | Switching vs Continuation | Insomnia | Education_levelLess than a high school degree | 1.281 | 0.851 | 1.928 | 2.36e-01 | NA | 3088 | 2436 | 652 |
| African | No Recorded Depression | Switching vs Continuation | Insomnia | Education_levelTwelve or GED | 1.075 | 0.752 | 1.537 | 6.91e-01 | NA | 3088 | 2436 | 652 |
| African | No Recorded Depression | Switching vs Continuation | Insomnia | Insomnia | 1.098 | 0.967 | 1.247 | 1.50e-01 | 1.00e+00 | 3088 | 2436 | 652 |
| African | No Recorded Depression | Discontinuation vs Continuation | Migraine | (Intercept) | 0.361 | 0.243 | 0.536 | 4.65e-07 | NA | 3605 | 2436 | 1169 |
| African | No Recorded Depression | Discontinuation vs Continuation | Migraine | age_at_first_AD | 1.001 | 0.996 | 1.006 | 7.01e-01 | NA | 3605 | 2436 | 1169 |
| African | No Recorded Depression | Discontinuation vs Continuation | Migraine | sex_at_birthMale | 1.178 | 1.010 | 1.375 | 3.72e-02 | NA | 3605 | 2436 | 1169 |
| African | No Recorded Depression | Discontinuation vs Continuation | Migraine | Income_quantitative | 1.006 | 0.960 | 1.054 | 7.99e-01 | NA | 3605 | 2436 | 1169 |
| African | No Recorded Depression | Discontinuation vs Continuation | Migraine | Education_levelCollege Undergraduate | 1.227 | 0.951 | 1.583 | 1.16e-01 | NA | 3605 | 2436 | 1169 |
| African | No Recorded Depression | Discontinuation vs Continuation | Migraine | Education_levelLess than a high school degree | 1.105 | 0.786 | 1.554 | 5.65e-01 | NA | 3605 | 2436 | 1169 |
| African | No Recorded Depression | Discontinuation vs Continuation | Migraine | Education_levelTwelve or GED | 1.232 | 0.923 | 1.644 | 1.58e-01 | NA | 3605 | 2436 | 1169 |
| African | No Recorded Depression | Discontinuation vs Continuation | Migraine | Migraine | 0.990 | 0.909 | 1.077 | 8.08e-01 | 1.00e+00 | 3605 | 2436 | 1169 |
| African | No Recorded Depression | Switching vs Continuation | Migraine | (Intercept) | 0.828 | 0.509 | 1.349 | 4.49e-01 | NA | 3088 | 2436 | 652 |
| African | No Recorded Depression | Switching vs Continuation | Migraine | age_at_first_AD | 0.980 | 0.973 | 0.986 | 6.17e-10 | NA | 3088 | 2436 | 652 |
| African | No Recorded Depression | Switching vs Continuation | Migraine | sex_at_birthMale | 0.884 | 0.722 | 1.083 | 2.35e-01 | NA | 3088 | 2436 | 652 |
| African | No Recorded Depression | Switching vs Continuation | Migraine | Income_quantitative | 0.950 | 0.894 | 1.010 | 9.92e-02 | NA | 3088 | 2436 | 652 |
| African | No Recorded Depression | Switching vs Continuation | Migraine | Education_levelCollege Undergraduate | 0.921 | 0.670 | 1.266 | 6.11e-01 | NA | 3088 | 2436 | 652 |
| African | No Recorded Depression | Switching vs Continuation | Migraine | Education_levelLess than a high school degree | 1.289 | 0.856 | 1.940 | 2.24e-01 | NA | 3088 | 2436 | 652 |
| African | No Recorded Depression | Switching vs Continuation | Migraine | Education_levelTwelve or GED | 1.075 | 0.752 | 1.537 | 6.92e-01 | NA | 3088 | 2436 | 652 |
| African | No Recorded Depression | Switching vs Continuation | Migraine | Migraine | 0.998 | 0.897 | 1.110 | 9.63e-01 | 1.00e+00 | 3088 | 2436 | 652 |
| African | No Recorded Depression | Discontinuation vs Continuation | Schizophrenia | (Intercept) | 0.361 | 0.243 | 0.536 | 4.68e-07 | NA | 3605 | 2436 | 1169 |
| African | No Recorded Depression | Discontinuation vs Continuation | Schizophrenia | age_at_first_AD | 1.001 | 0.996 | 1.006 | 6.99e-01 | NA | 3605 | 2436 | 1169 |
| African | No Recorded Depression | Discontinuation vs Continuation | Schizophrenia | sex_at_birthMale | 1.178 | 1.009 | 1.374 | 3.79e-02 | NA | 3605 | 2436 | 1169 |
| African | No Recorded Depression | Discontinuation vs Continuation | Schizophrenia | Income_quantitative | 1.005 | 0.960 | 1.053 | 8.21e-01 | NA | 3605 | 2436 | 1169 |
| African | No Recorded Depression | Discontinuation vs Continuation | Schizophrenia | Education_levelCollege Undergraduate | 1.227 | 0.951 | 1.584 | 1.16e-01 | NA | 3605 | 2436 | 1169 |
| African | No Recorded Depression | Discontinuation vs Continuation | Schizophrenia | Education_levelLess than a high school degree | 1.107 | 0.787 | 1.557 | 5.58e-01 | NA | 3605 | 2436 | 1169 |
| African | No Recorded Depression | Discontinuation vs Continuation | Schizophrenia | Education_levelTwelve or GED | 1.232 | 0.923 | 1.646 | 1.56e-01 | NA | 3605 | 2436 | 1169 |
| African | No Recorded Depression | Discontinuation vs Continuation | Schizophrenia | Schizophrenia | 0.966 | 0.883 | 1.056 | 4.46e-01 | 1.00e+00 | 3605 | 2436 | 1169 |
| African | No Recorded Depression | Switching vs Continuation | Schizophrenia | (Intercept) | 0.827 | 0.507 | 1.347 | 4.44e-01 | NA | 3088 | 2436 | 652 |
| African | No Recorded Depression | Switching vs Continuation | Schizophrenia | age_at_first_AD | 0.979 | 0.973 | 0.986 | 5.91e-10 | NA | 3088 | 2436 | 652 |
| African | No Recorded Depression | Switching vs Continuation | Schizophrenia | sex_at_birthMale | 0.884 | 0.722 | 1.084 | 2.36e-01 | NA | 3088 | 2436 | 652 |
| African | No Recorded Depression | Switching vs Continuation | Schizophrenia | Income_quantitative | 0.951 | 0.895 | 1.011 | 1.07e-01 | NA | 3088 | 2436 | 652 |
| African | No Recorded Depression | Switching vs Continuation | Schizophrenia | Education_levelCollege Undergraduate | 0.921 | 0.670 | 1.267 | 6.14e-01 | NA | 3088 | 2436 | 652 |
| African | No Recorded Depression | Switching vs Continuation | Schizophrenia | Education_levelLess than a high school degree | 1.288 | 0.856 | 1.939 | 2.25e-01 | NA | 3088 | 2436 | 652 |
| African | No Recorded Depression | Switching vs Continuation | Schizophrenia | Education_levelTwelve or GED | 1.077 | 0.753 | 1.540 | 6.84e-01 | NA | 3088 | 2436 | 652 |
| African | No Recorded Depression | Switching vs Continuation | Schizophrenia | Schizophrenia | 1.037 | 0.927 | 1.160 | 5.28e-01 | 1.00e+00 | 3088 | 2436 | 652 |
| African | Recorded Anxiety | Discontinuation vs Continuation | ADHD | (Intercept) | 0.476 | 0.302 | 0.750 | 1.38e-03 | NA | 2793 | 2014 | 779 |
| African | Recorded Anxiety | Discontinuation vs Continuation | ADHD | age_at_first_AD | 0.996 | 0.990 | 1.002 | 2.38e-01 | NA | 2793 | 2014 | 779 |
| African | Recorded Anxiety | Discontinuation vs Continuation | ADHD | sex_at_birthMale | 1.218 | 1.001 | 1.483 | 4.91e-02 | NA | 2793 | 2014 | 779 |
| African | Recorded Anxiety | Discontinuation vs Continuation | ADHD | Income_quantitative | 0.922 | 0.870 | 0.976 | 5.57e-03 | NA | 2793 | 2014 | 779 |
| African | Recorded Anxiety | Discontinuation vs Continuation | ADHD | Education_levelCollege Undergraduate | 1.111 | 0.823 | 1.499 | 4.91e-01 | NA | 2793 | 2014 | 779 |
| African | Recorded Anxiety | Discontinuation vs Continuation | ADHD | Education_levelLess than a high school degree | 1.112 | 0.732 | 1.691 | 6.18e-01 | NA | 2793 | 2014 | 779 |

| | | | | | | | | | | | | |
|---|---|---|---|---|---|---|---|---|---|---|---|---|
| African | Recorded Anxiety | Discontinuation vs Continuation | ADHD | Education_levelTwelve or GED | 1.125 | 0.802 | 1.576 | 4.95e-01 | NA | 2793 | 2014 | 779 |
| African | Recorded Anxiety | Discontinuation vs Continuation | ADHD | ADHD | 1.059 | 0.942 | 1.191 | 3.39e-01 | 1.00e+00 | 2793 | 2014 | 779 |
| African | Recorded Anxiety | Switching vs Continuation | ADHD | (Intercept) | 2.314 | 1.632 | 3.281 | 2.49e-06 | NA | 3909 | 2014 | 1895 |
| African | Recorded Anxiety | Switching vs Continuation | ADHD | age_at_first_AD | 0.986 | 0.981 | 0.991 | 2.12e-08 | NA | 3909 | 2014 | 1895 |
| African | Recorded Anxiety | Switching vs Continuation | ADHD | sex_at_birthMale | 1.079 | 0.924 | 1.261 | 3.36e-01 | NA | 3909 | 2014 | 1895 |
| African | Recorded Anxiety | Switching vs Continuation | ADHD | Income_quantitative | 0.889 | 0.851 | 0.928 | 9.70e-08 | NA | 3909 | 2014 | 1895 |
| African | Recorded Anxiety | Switching vs Continuation | ADHD | Education_levelCollege Undergraduate | 0.996 | 0.797 | 1.244 | 9.71e-01 | NA | 3909 | 2014 | 1895 |
| African | Recorded Anxiety | Switching vs Continuation | ADHD | Education_levelLess than a high school degree | 1.083 | 0.792 | 1.480 | 6.20e-01 | NA | 3909 | 2014 | 1895 |
| African | Recorded Anxiety | Switching vs Continuation | ADHD | Education_levelTwelve or GED | 0.909 | 0.705 | 1.171 | 4.59e-01 | NA | 3909 | 2014 | 1895 |
| African | Recorded Anxiety | Switching vs Continuation | ADHD | ADHD | 1.024 | 0.936 | 1.120 | 6.02e-01 | 1.00e+00 | 3909 | 2014 | 1895 |
| African | Recorded Anxiety | Discontinuation vs Continuation | BMI | (Intercept) | 0.478 | 0.303 | 0.753 | 1.48e-03 | NA | 2793 | 2014 | 779 |
| African | Recorded Anxiety | Discontinuation vs Continuation | BMI | age_at_first_AD | 0.996 | 0.990 | 1.002 | 2.08e-01 | NA | 2793 | 2014 | 779 |
| African | Recorded Anxiety | Discontinuation vs Continuation | BMI | sex_at_birthMale | 1.215 | 0.998 | 1.480 | 5.22e-02 | NA | 2793 | 2014 | 779 |
| African | Recorded Anxiety | Discontinuation vs Continuation | BMI | Income_quantitative | 0.919 | 0.868 | 0.974 | 4.26e-03 | NA | 2793 | 2014 | 779 |
| African | Recorded Anxiety | Discontinuation vs Continuation | BMI | Education_levelCollege Undergraduate | 1.127 | 0.835 | 1.521 | 4.36e-01 | NA | 2793 | 2014 | 779 |
| African | Recorded Anxiety | Discontinuation vs Continuation | BMI | Education_levelLess than a high school degree | 1.128 | 0.742 | 1.714 | 5.74e-01 | NA | 2793 | 2014 | 779 |
| African | Recorded Anxiety | Discontinuation vs Continuation | BMI | Education_levelTwelve or GED | 1.146 | 0.817 | 1.607 | 4.30e-01 | NA | 2793 | 2014 | 779 |
| African | Recorded Anxiety | Discontinuation vs Continuation | BMI | BMI | 0.918 | 0.822 | 1.025 | 1.26e-01 | 8.85e-01 | 2793 | 2014 | 779 |
| African | Recorded Anxiety | Switching vs Continuation | BMI | (Intercept) | 2.324 | 1.639 | 3.296 | 2.23e-06 | NA | 3909 | 2014 | 1895 |
| African | Recorded Anxiety | Switching vs Continuation | BMI | age_at_first_AD | 0.986 | 0.981 | 0.991 | 1.42e-08 | NA | 3909 | 2014 | 1895 |
| African | Recorded Anxiety | Switching vs Continuation | BMI | sex_at_birthMale | 1.074 | 0.919 | 1.254 | 3.72e-01 | NA | 3909 | 2014 | 1895 |
| African | Recorded Anxiety | Switching vs Continuation | BMI | Income_quantitative | 0.887 | 0.850 | 0.927 | 7.23e-08 | NA | 3909 | 2014 | 1895 |
| African | Recorded Anxiety | Switching vs Continuation | BMI | Education_levelCollege Undergraduate | 1.004 | 0.804 | 1.254 | 9.72e-01 | NA | 3909 | 2014 | 1895 |
| African | Recorded Anxiety | Switching vs Continuation | BMI | Education_levelLess than a high school degree | 1.095 | 0.801 | 1.497 | 5.70e-01 | NA | 3909 | 2014 | 1895 |
| African | Recorded Anxiety | Switching vs Continuation | BMI | Education_levelTwelve or GED | 0.920 | 0.714 | 1.186 | 5.19e-01 | NA | 3909 | 2014 | 1895 |
| African | Recorded Anxiety | Switching vs Continuation | BMI | BMI | 0.940 | 0.865 | 1.022 | 1.47e-01 | 1.00e+00 | 3909 | 2014 | 1895 |
| African | Recorded Anxiety | Discontinuation vs Continuation | Bipolar Disorder | (Intercept) | 0.476 | 0.302 | 0.751 | 1.41e-03 | NA | 2793 | 2014 | 779 |
| African | Recorded Anxiety | Discontinuation vs Continuation | Bipolar Disorder | age_at_first_AD | 0.996 | 0.990 | 1.002 | 2.28e-01 | NA | 2793 | 2014 | 779 |
| African | Recorded Anxiety | Discontinuation vs Continuation | Bipolar Disorder | sex_at_birthMale | 1.220 | 1.002 | 1.485 | 4.80e-02 | NA | 2793 | 2014 | 779 |
| African | Recorded Anxiety | Discontinuation vs Continuation | Bipolar Disorder | Income_quantitative | 0.921 | 0.870 | 0.976 | 5.42e-03 | NA | 2793 | 2014 | 779 |
| African | Recorded Anxiety | Discontinuation vs Continuation | Bipolar Disorder | Education_levelCollege Undergraduate | 1.114 | 0.825 | 1.502 | 4.82e-01 | NA | 2793 | 2014 | 779 |
| African | Recorded Anxiety | Discontinuation vs Continuation | Bipolar Disorder | Education_levelLess than a high school degree | 1.116 | 0.734 | 1.696 | 6.07e-01 | NA | 2793 | 2014 | 779 |
| African | Recorded Anxiety | Discontinuation vs Continuation | Bipolar Disorder | Education_levelTwelve or GED | 1.130 | 0.806 | 1.583 | 4.79e-01 | NA | 2793 | 2014 | 779 |
| African | Recorded Anxiety | Discontinuation vs Continuation | Bipolar Disorder | Bipolar Disorder | 1.020 | 0.905 | 1.151 | 7.45e-01 | 1.00e+00 | 2793 | 2014 | 779 |
| African | Recorded Anxiety | Switching vs Continuation | Bipolar Disorder | (Intercept) | 2.312 | 1.630 | 3.278 | 2.55e-06 | NA | 3909 | 2014 | 1895 |
| African | Recorded Anxiety | Switching vs Continuation | Bipolar Disorder | age_at_first_AD | 0.986 | 0.981 | 0.991 | 1.72e-08 | NA | 3909 | 2014 | 1895 |
| African | Recorded Anxiety | Switching vs Continuation | Bipolar Disorder | sex_at_birthMale | 1.077 | 0.922 | 1.258 | 3.51e-01 | NA | 3909 | 2014 | 1895 |
| African | Recorded Anxiety | Switching vs Continuation | Bipolar Disorder | Income_quantitative | 0.889 | 0.852 | 0.929 | 1.21e-07 | NA | 3909 | 2014 | 1895 |
| African | Recorded Anxiety | Switching vs Continuation | Bipolar Disorder | Education_levelCollege Undergraduate | 0.999 | 0.800 | 1.248 | 9.95e-01 | NA | 3909 | 2014 | 1895 |
| African | Recorded Anxiety | Switching vs Continuation | Bipolar Disorder | Education_levelLess than a high school degree | 1.087 | 0.795 | 1.486 | 6.00e-01 | NA | 3909 | 2014 | 1895 |
| African | Recorded Anxiety | Switching vs Continuation | Bipolar Disorder | Education_levelTwelve or GED | 0.915 | 0.710 | 1.178 | 4.90e-01 | NA | 3909 | 2014 | 1895 |
| African | Recorded Anxiety | Switching vs Continuation | Bipolar Disorder | Bipolar Disorder | 1.058 | 0.964 | 1.162 | 2.32e-01 | 1.00e+00 | 3909 | 2014 | 1895 |
| African | Recorded Anxiety | Discontinuation vs Continuation | Depression | (Intercept) | 0.476 | 0.302 | 0.751 | 1.41e-03 | NA | 2793 | 2014 | 779 |
| African | Recorded Anxiety | Discontinuation vs Continuation | Depression | age_at_first_AD | 0.996 | 0.990 | 1.002 | 2.25e-01 | NA | 2793 | 2014 | 779 |
| African | Recorded Anxiety | Discontinuation vs Continuation | Depression | sex_at_birthMale | 1.220 | 1.002 | 1.485 | 4.74e-02 | NA | 2793 | 2014 | 779 |
| African | Recorded Anxiety | Discontinuation vs Continuation | Depression | Income_quantitative | 0.922 | 0.870 | 0.976 | 5.62e-03 | NA | 2793 | 2014 | 779 |
| African | Recorded Anxiety | Discontinuation vs Continuation | Depression | Education_levelCollege Undergraduate | 1.116 | 0.827 | 1.506 | 4.73e-01 | NA | 2793 | 2014 | 779 |
| African | Recorded Anxiety | Discontinuation vs Continuation | Depression | Education_levelLess than a high school degree | 1.115 | 0.734 | 1.695 | 6.09e-01 | NA | 2793 | 2014 | 779 |
| African | Recorded Anxiety | Discontinuation vs Continuation | Depression | Education_levelTwelve or GED | 1.129 | 0.806 | 1.583 | 4.80e-01 | NA | 2793 | 2014 | 779 |
| African | Recorded Anxiety | Discontinuation vs Continuation | Depression | Depression | 1.029 | 0.915 | 1.156 | 6.37e-01 | 1.00e+00 | 2793 | 2014 | 779 |
| African | Recorded Anxiety | Switching vs Continuation | Depression | (Intercept) | 2.304 | 1.625 | 3.267 | 2.81e-06 | NA | 3909 | 2014 | 1895 |
| African | Recorded Anxiety | Switching vs Continuation | Depression | age_at_first_AD | 0.986 | 0.981 | 0.991 | 2.10e-08 | NA | 3909 | 2014 | 1895 |
| African | Recorded Anxiety | Switching vs Continuation | Depression | sex_at_birthMale | 1.078 | 0.923 | 1.259 | 3.44e-01 | NA | 3909 | 2014 | 1895 |
| African | Recorded Anxiety | Switching vs Continuation | Depression | Income_quantitative | 0.890 | 0.852 | 0.930 | 1.56e-07 | NA | 3909 | 2014 | 1895 |
| African | Recorded Anxiety | Switching vs Continuation | Depression | Education_levelCollege Undergraduate | 0.996 | 0.798 | 1.245 | 9.74e-01 | NA | 3909 | 2014 | 1895 |
| African | Recorded Anxiety | Switching vs Continuation | Depression | Education_levelLess than a high school degree | 1.082 | 0.791 | 1.479 | 6.22e-01 | NA | 3909 | 2014 | 1895 |
| African | Recorded Anxiety | Switching vs Continuation | Depression | Education_levelTwelve or GED | 0.910 | 0.707 | 1.173 | 4.68e-01 | NA | 3909 | 2014 | 1895 |

| | | | | | | | | | | | | |
|---|---|---|---|---|---|---|---|---|---|---|---|---|
| African | Recorded Anxiety | Switching vs Continuation | Depression | Depression | 1.102 | 1.007 | 1.206 | 3.42e-02 | 2.39e-01 | 3909 | 2014 | 1895 |
| African | Recorded Anxiety | Discontinuation vs Continuation | Insomnia | (Intercept) | 0.480 | 0.304 | 0.757 | 1.60e-03 | NA | 2793 | 2014 | 779 |
| African | Recorded Anxiety | Discontinuation vs Continuation | Insomnia | age_at_first_AD | 0.996 | 0.990 | 1.002 | 2.26e-01 | NA | 2793 | 2014 | 779 |
| African | Recorded Anxiety | Discontinuation vs Continuation | Insomnia | sex_at_birthMale | 1.216 | 0.999 | 1.480 | 5.16e-02 | NA | 2793 | 2014 | 779 |
| African | Recorded Anxiety | Discontinuation vs Continuation | Insomnia | Income_quantitative | 0.921 | 0.870 | 0.976 | 5.45e-03 | NA | 2793 | 2014 | 779 |
| African | Recorded Anxiety | Discontinuation vs Continuation | Insomnia | Education_levelCollege Undergraduate | 1.109 | 0.822 | 1.497 | 4.97e-01 | NA | 2793 | 2014 | 779 |
| African | Recorded Anxiety | Discontinuation vs Continuation | Insomnia | Education_levelLess than a high school degree | 1.105 | 0.727 | 1.680 | 6.40e-01 | NA | 2793 | 2014 | 779 |
| African | Recorded Anxiety | Discontinuation vs Continuation | Insomnia | Education_levelTwelve or GED | 1.123 | 0.801 | 1.574 | 5.01e-01 | NA | 2793 | 2014 | 779 |
| African | Recorded Anxiety | Discontinuation vs Continuation | Insomnia | Insomnia | 1.108 | 0.982 | 1.250 | 9.75e-02 | 6.83e-01 | 2793 | 2014 | 779 |
| African | Recorded Anxiety | Switching vs Continuation | Insomnia | (Intercept) | 2.358 | 1.662 | 3.346 | 1.55e-06 | NA | 3909 | 2014 | 1895 |
| African | Recorded Anxiety | Switching vs Continuation | Insomnia | age_at_first_AD | 0.986 | 0.981 | 0.991 | 1.68e-08 | NA | 3909 | 2014 | 1895 |
| African | Recorded Anxiety | Switching vs Continuation | Insomnia | sex_at_birthMale | 1.082 | 0.926 | 1.264 | 3.23e-01 | NA | 3909 | 2014 | 1895 |
| African | Recorded Anxiety | Switching vs Continuation | Insomnia | Income_quantitative | 0.888 | 0.850 | 0.927 | 7.80e-08 | NA | 3909 | 2014 | 1895 |
| African | Recorded Anxiety | Switching vs Continuation | Insomnia | Education_levelCollege Undergraduate | 0.984 | 0.788 | 1.230 | 8.89e-01 | NA | 3909 | 2014 | 1895 |
| African | Recorded Anxiety | Switching vs Continuation | Insomnia | Education_levelLess than a high school degree | 1.069 | 0.782 | 1.463 | 6.75e-01 | NA | 3909 | 2014 | 1895 |
| African | Recorded Anxiety | Switching vs Continuation | Insomnia | Education_levelTwelve or GED | 0.895 | 0.695 | 1.154 | 3.92e-01 | NA | 3909 | 2014 | 1895 |
| African | Recorded Anxiety | Switching vs Continuation | Insomnia | Insomnia | 1.175 | 1.072 | 1.288 | 5.81e-04 | 4.06e-03 | 3909 | 2014 | 1895 |
| African | Recorded Anxiety | Discontinuation vs Continuation | Migraine | (Intercept) | 0.480 | 0.304 | 0.757 | 1.60e-03 | NA | 2793 | 2014 | 779 |
| African | Recorded Anxiety | Discontinuation vs Continuation | Migraine | age_at_first_AD | 0.996 | 0.990 | 1.002 | 2.24e-01 | NA | 2793 | 2014 | 779 |
| African | Recorded Anxiety | Discontinuation vs Continuation | Migraine | sex_at_birthMale | 1.217 | 1.000 | 1.482 | 5.02e-02 | NA | 2793 | 2014 | 779 |
| African | Recorded Anxiety | Discontinuation vs Continuation | Migraine | Income_quantitative | 0.921 | 0.869 | 0.975 | 4.93e-03 | NA | 2793 | 2014 | 779 |
| African | Recorded Anxiety | Discontinuation vs Continuation | Migraine | Education_levelCollege Undergraduate | 1.109 | 0.822 | 1.496 | 4.99e-01 | NA | 2793 | 2014 | 779 |
| African | Recorded Anxiety | Discontinuation vs Continuation | Migraine | Education_levelLess than a high school degree | 1.113 | 0.732 | 1.692 | 6.16e-01 | NA | 2793 | 2014 | 779 |
| African | Recorded Anxiety | Discontinuation vs Continuation | Migraine | Education_levelTwelve or GED | 1.122 | 0.801 | 1.573 | 5.04e-01 | NA | 2793 | 2014 | 779 |
| African | Recorded Anxiety | Discontinuation vs Continuation | Migraine | Migraine | 1.061 | 0.957 | 1.176 | 2.60e-01 | 1.00e+00 | 2793 | 2014 | 779 |
| African | Recorded Anxiety | Switching vs Continuation | Migraine | (Intercept) | 2.311 | 1.630 | 3.277 | 2.57e-06 | NA | 3909 | 2014 | 1895 |
| African | Recorded Anxiety | Switching vs Continuation | Migraine | age_at_first_AD | 0.986 | 0.981 | 0.991 | 2.03e-08 | NA | 3909 | 2014 | 1895 |
| African | Recorded Anxiety | Switching vs Continuation | Migraine | sex_at_birthMale | 1.080 | 0.924 | 1.261 | 3.35e-01 | NA | 3909 | 2014 | 1895 |
| African | Recorded Anxiety | Switching vs Continuation | Migraine | Income_quantitative | 0.889 | 0.851 | 0.928 | 9.61e-08 | NA | 3909 | 2014 | 1895 |
| African | Recorded Anxiety | Switching vs Continuation | Migraine | Education_levelCollege Undergraduate | 0.998 | 0.799 | 1.246 | 9.84e-01 | NA | 3909 | 2014 | 1895 |
| African | Recorded Anxiety | Switching vs Continuation | Migraine | Education_levelLess than a high school degree | 1.087 | 0.796 | 1.486 | 5.99e-01 | NA | 3909 | 2014 | 1895 |
| African | Recorded Anxiety | Switching vs Continuation | Migraine | Education_levelTwelve or GED | 0.911 | 0.707 | 1.174 | 4.71e-01 | NA | 3909 | 2014 | 1895 |
| African | Recorded Anxiety | Switching vs Continuation | Migraine | Migraine | 1.002 | 0.928 | 1.083 | 9.52e-01 | 1.00e+00 | 3909 | 2014 | 1895 |
| African | Recorded Anxiety | Discontinuation vs Continuation | Schizophrenia | (Intercept) | 0.479 | 0.303 | 0.755 | 1.51e-03 | NA | 2793 | 2014 | 779 |
| African | Recorded Anxiety | Discontinuation vs Continuation | Schizophrenia | age_at_first_AD | 0.996 | 0.990 | 1.002 | 2.23e-01 | NA | 2793 | 2014 | 779 |
| African | Recorded Anxiety | Discontinuation vs Continuation | Schizophrenia | sex_at_birthMale | 1.220 | 1.002 | 1.485 | 4.77e-02 | NA | 2793 | 2014 | 779 |
| African | Recorded Anxiety | Discontinuation vs Continuation | Schizophrenia | Income_quantitative | 0.920 | 0.869 | 0.975 | 4.73e-03 | NA | 2793 | 2014 | 779 |
| African | Recorded Anxiety | Discontinuation vs Continuation | Schizophrenia | Education_levelCollege Undergraduate | 1.114 | 0.825 | 1.502 | 4.82e-01 | NA | 2793 | 2014 | 779 |
| African | Recorded Anxiety | Discontinuation vs Continuation | Schizophrenia | Education_levelLess than a high school degree | 1.124 | 0.740 | 1.709 | 5.84e-01 | NA | 2793 | 2014 | 779 |
| African | Recorded Anxiety | Discontinuation vs Continuation | Schizophrenia | Education_levelTwelve or GED | 1.129 | 0.805 | 1.582 | 4.82e-01 | NA | 2793 | 2014 | 779 |
| African | Recorded Anxiety | Discontinuation vs Continuation | Schizophrenia | Schizophrenia | 0.923 | 0.831 | 1.025 | 1.32e-01 | 9.26e-01 | 2793 | 2014 | 779 |
| African | Recorded Anxiety | Switching vs Continuation | Schizophrenia | (Intercept) | 2.315 | 1.632 | 3.282 | 2.48e-06 | NA | 3909 | 2014 | 1895 |
| African | Recorded Anxiety | Switching vs Continuation | Schizophrenia | age_at_first_AD | 0.986 | 0.981 | 0.991 | 2.25e-08 | NA | 3909 | 2014 | 1895 |
| African | Recorded Anxiety | Switching vs Continuation | Schizophrenia | sex_at_birthMale | 1.081 | 0.925 | 1.263 | 3.25e-01 | NA | 3909 | 2014 | 1895 |
| African | Recorded Anxiety | Switching vs Continuation | Schizophrenia | Income_quantitative | 0.888 | 0.850 | 0.927 | 7.91e-08 | NA | 3909 | 2014 | 1895 |
| African | Recorded Anxiety | Switching vs Continuation | Schizophrenia | Education_levelCollege Undergraduate | 0.996 | 0.798 | 1.244 | 9.73e-01 | NA | 3909 | 2014 | 1895 |
| African | Recorded Anxiety | Switching vs Continuation | Schizophrenia | Education_levelLess than a high school degree | 1.088 | 0.796 | 1.488 | 5.95e-01 | NA | 3909 | 2014 | 1895 |
| African | Recorded Anxiety | Switching vs Continuation | Schizophrenia | Education_levelTwelve or GED | 0.909 | 0.705 | 1.170 | 4.58e-01 | NA | 3909 | 2014 | 1895 |
| African | Recorded Anxiety | Switching vs Continuation | Schizophrenia | Schizophrenia | 0.930 | 0.857 | 1.008 | 7.83e-02 | 5.48e-01 | 3909 | 2014 | 1895 |
| Latino/Mixed-American | Full Cohort | Discontinuation vs Continuation | ADHD | (Intercept) | 0.485 | 0.340 | 0.691 | 6.29e-05 | NA | 3862 | 2456 | 1406 |
| Latino/Mixed-American | Full Cohort | Discontinuation vs Continuation | ADHD | age_at_first_AD | 1.000 | 0.995 | 1.004 | 8.65e-01 | NA | 3862 | 2456 | 1406 |
| Latino/Mixed-American | Full Cohort | Discontinuation vs Continuation | ADHD | sex_at_birthMale | 1.224 | 1.058 | 1.416 | 6.57e-03 | NA | 3862 | 2456 | 1406 |
| Latino/Mixed-American | Full Cohort | Discontinuation vs Continuation | ADHD | Income_quantitative | 0.940 | 0.904 | 0.977 | 1.68e-03 | NA | 3862 | 2456 | 1406 |
| Latino/Mixed-American | Full Cohort | Discontinuation vs Continuation | ADHD | Education_levelCollege Undergraduate | 1.488 | 1.155 | 1.918 | 2.11e-03 | NA | 3862 | 2456 | 1406 |
| Latino/Mixed-American | Full Cohort | Discontinuation vs Continuation | ADHD | Education_levelLess than a high school degree | 1.106 | 0.814 | 1.503 | 5.21e-01 | NA | 3862 | 2456 | 1406 |
| Latino/Mixed-American | Full Cohort | Discontinuation vs Continuation | ADHD | Education_levelTwelve or GED | 1.395 | 1.043 | 1.864 | 2.47e-02 | NA | 3862 | 2456 | 1406 |
| Latino/Mixed-American | Full Cohort | Discontinuation vs Continuation | ADHD | ADHD | 1.038 | 0.964 | 1.119 | 3.22e-01 | 1.00e+00 | 3862 | 2456 | 1406 |

| | | | | | | | | | | | | |
|---|---|---|---|---|---|---|---|---|---|---|---|---|
| Latino/Mixed-American | Full Cohort | Switching vs Continuation | ADHD | (Intercept) | 1.246 | 0.889 | 1.745 | 2.02e-01 | NA | 3931 | 2456 | 1475 |
| Latino/Mixed-American | Full Cohort | Switching vs Continuation | ADHD | age_at_first_AD | 0.986 | 0.982 | 0.991 | 4.59e-09 | NA | 3931 | 2456 | 1475 |
| Latino/Mixed-American | Full Cohort | Switching vs Continuation | ADHD | sex_at_birthMale | 1.063 | 0.916 | 1.234 | 4.20e-01 | NA | 3931 | 2456 | 1475 |
| Latino/Mixed-American | Full Cohort | Switching vs Continuation | ADHD | Income_quantitative | 0.942 | 0.907 | 0.978 | 1.98e-03 | NA | 3931 | 2456 | 1475 |
| Latino/Mixed-American | Full Cohort | Switching vs Continuation | ADHD | Education_levelCollege Undergraduate | 1.076 | 0.854 | 1.355 | 5.35e-01 | NA | 3931 | 2456 | 1475 |
| Latino/Mixed-American | Full Cohort | Switching vs Continuation | ADHD | Education_levelLess than a high school degree | 0.986 | 0.741 | 1.311 | 9.21e-01 | NA | 3931 | 2456 | 1475 |
| Latino/Mixed-American | Full Cohort | Switching vs Continuation | ADHD | Education_levelTwelve or GED | 0.942 | 0.719 | 1.236 | 6.67e-01 | NA | 3931 | 2456 | 1475 |
| Latino/Mixed-American | Full Cohort | Switching vs Continuation | ADHD | ADHD | 1.046 | 0.973 | 1.124 | 2.23e-01 | 1.00e+00 | 3931 | 2456 | 1475 |
| Latino/Mixed-American | Full Cohort | Discontinuation vs Continuation | BMI | (Intercept) | 0.483 | 0.339 | 0.689 | 5.77e-05 | NA | 3862 | 2456 | 1406 |
| Latino/Mixed-American | Full Cohort | Discontinuation vs Continuation | BMI | age_at_first_AD | 1.000 | 0.995 | 1.004 | 8.41e-01 | NA | 3862 | 2456 | 1406 |
| Latino/Mixed-American | Full Cohort | Discontinuation vs Continuation | BMI | sex_at_birthMale | 1.225 | 1.059 | 1.418 | 6.33e-03 | NA | 3862 | 2456 | 1406 |
| Latino/Mixed-American | Full Cohort | Discontinuation vs Continuation | BMI | Income_quantitative | 0.943 | 0.907 | 0.980 | 2.81e-03 | NA | 3862 | 2456 | 1406 |
| Latino/Mixed-American | Full Cohort | Discontinuation vs Continuation | BMI | Education_levelCollege Undergraduate | 1.484 | 1.152 | 1.913 | 2.27e-03 | NA | 3862 | 2456 | 1406 |
| Latino/Mixed-American | Full Cohort | Discontinuation vs Continuation | BMI | Education_levelLess than a high school degree | 1.107 | 0.815 | 1.505 | 5.16e-01 | NA | 3862 | 2456 | 1406 |
| Latino/Mixed-American | Full Cohort | Discontinuation vs Continuation | BMI | Education_levelTwelve or GED | 1.394 | 1.043 | 1.863 | 2.47e-02 | NA | 3862 | 2456 | 1406 |
| Latino/Mixed-American | Full Cohort | Discontinuation vs Continuation | BMI | BMI | 1.091 | 1.019 | 1.169 | 1.28e-02 | 8.96e-02 | 3862 | 2456 | 1406 |
| Latino/Mixed-American | Full Cohort | Switching vs Continuation | BMI | (Intercept) | 1.247 | 0.890 | 1.747 | 2.00e-01 | NA | 3931 | 2456 | 1475 |
| Latino/Mixed-American | Full Cohort | Switching vs Continuation | BMI | age_at_first_AD | 0.986 | 0.982 | 0.991 | 3.56e-09 | NA | 3931 | 2456 | 1475 |
| Latino/Mixed-American | Full Cohort | Switching vs Continuation | BMI | sex_at_birthMale | 1.065 | 0.918 | 1.236 | 4.06e-01 | NA | 3931 | 2456 | 1475 |
| Latino/Mixed-American | Full Cohort | Switching vs Continuation | BMI | Income_quantitative | 0.943 | 0.908 | 0.979 | 2.41e-03 | NA | 3931 | 2456 | 1475 |
| Latino/Mixed-American | Full Cohort | Switching vs Continuation | BMI | Education_levelCollege Undergraduate | 1.074 | 0.852 | 1.353 | 5.46e-01 | NA | 3931 | 2456 | 1475 |
| Latino/Mixed-American | Full Cohort | Switching vs Continuation | BMI | Education_levelLess than a high school degree | 0.993 | 0.747 | 1.320 | 9.62e-01 | NA | 3931 | 2456 | 1475 |
| Latino/Mixed-American | Full Cohort | Switching vs Continuation | BMI | Education_levelTwelve or GED | 0.943 | 0.719 | 1.237 | 6.73e-01 | NA | 3931 | 2456 | 1475 |
| Latino/Mixed-American | Full Cohort | Switching vs Continuation | BMI | BMI | 1.059 | 0.990 | 1.134 | 9.75e-02 | 6.82e-01 | 3931 | 2456 | 1475 |
| Latino/Mixed-American | Full Cohort | Discontinuation vs Continuation | Bipolar Disorder | (Intercept) | 0.486 | 0.341 | 0.693 | 6.56e-05 | NA | 3862 | 2456 | 1406 |
| Latino/Mixed-American | Full Cohort | Discontinuation vs Continuation | Bipolar Disorder | age_at_first_AD | 1.000 | 0.995 | 1.004 | 8.13e-01 | NA | 3862 | 2456 | 1406 |
| Latino/Mixed-American | Full Cohort | Discontinuation vs Continuation | Bipolar Disorder | sex_at_birthMale | 1.226 | 1.059 | 1.418 | 6.23e-03 | NA | 3862 | 2456 | 1406 |
| Latino/Mixed-American | Full Cohort | Discontinuation vs Continuation | Bipolar Disorder | Income_quantitative | 0.939 | 0.903 | 0.976 | 1.36e-03 | NA | 3862 | 2456 | 1406 |
| Latino/Mixed-American | Full Cohort | Discontinuation vs Continuation | Bipolar Disorder | Education_levelCollege Undergraduate | 1.498 | 1.163 | 1.930 | 1.77e-03 | NA | 3862 | 2456 | 1406 |
| Latino/Mixed-American | Full Cohort | Discontinuation vs Continuation | Bipolar Disorder | Education_levelLess than a high school degree | 1.117 | 0.822 | 1.517 | 4.81e-01 | NA | 3862 | 2456 | 1406 |
| Latino/Mixed-American | Full Cohort | Discontinuation vs Continuation | Bipolar Disorder | Education_levelTwelve or GED | 1.409 | 1.054 | 1.883 | 2.04e-02 | NA | 3862 | 2456 | 1406 |
| Latino/Mixed-American | Full Cohort | Discontinuation vs Continuation | Bipolar Disorder | Bipolar Disorder | 1.050 | 0.976 | 1.130 | 1.91e-01 | 1.00e+00 | 3862 | 2456 | 1406 |
| Latino/Mixed-American | Full Cohort | Switching vs Continuation | Bipolar Disorder | (Intercept) | 1.240 | 0.885 | 1.737 | 2.12e-01 | NA | 3931 | 2456 | 1475 |
| Latino/Mixed-American | Full Cohort | Switching vs Continuation | Bipolar Disorder | age_at_first_AD | 0.986 | 0.982 | 0.991 | 3.24e-09 | NA | 3931 | 2456 | 1475 |
| Latino/Mixed-American | Full Cohort | Switching vs Continuation | Bipolar Disorder | sex_at_birthMale | 1.064 | 0.917 | 1.235 | 4.11e-01 | NA | 3931 | 2456 | 1475 |
| Latino/Mixed-American | Full Cohort | Switching vs Continuation | Bipolar Disorder | Income_quantitative | 0.942 | 0.907 | 0.978 | 1.97e-03 | NA | 3931 | 2456 | 1475 |
| Latino/Mixed-American | Full Cohort | Switching vs Continuation | Bipolar Disorder | Education_levelCollege Undergraduate | 1.086 | 0.862 | 1.368 | 4.85e-01 | NA | 3931 | 2456 | 1475 |
| Latino/Mixed-American | Full Cohort | Switching vs Continuation | Bipolar Disorder | Education_levelLess than a high school degree | 1.000 | 0.752 | 1.330 | 9.99e-01 | NA | 3931 | 2456 | 1475 |
| Latino/Mixed-American | Full Cohort | Switching vs Continuation | Bipolar Disorder | Education_levelTwelve or GED | 0.955 | 0.729 | 1.253 | 7.41e-01 | NA | 3931 | 2456 | 1475 |
| Latino/Mixed-American | Full Cohort | Switching vs Continuation | Bipolar Disorder | Bipolar Disorder | 1.041 | 0.968 | 1.119 | 2.76e-01 | 1.00e+00 | 3931 | 2456 | 1475 |
| Latino/Mixed-American | Full Cohort | Discontinuation vs Continuation | Depression | (Intercept) | 0.482 | 0.338 | 0.687 | 5.54e-05 | NA | 3862 | 2456 | 1406 |
| Latino/Mixed-American | Full Cohort | Discontinuation vs Continuation | Depression | age_at_first_AD | 1.000 | 0.995 | 1.004 | 9.40e-01 | NA | 3862 | 2456 | 1406 |
| Latino/Mixed-American | Full Cohort | Discontinuation vs Continuation | Depression | sex_at_birthMale | 1.222 | 1.056 | 1.414 | 7.22e-03 | NA | 3862 | 2456 | 1406 |
| Latino/Mixed-American | Full Cohort | Discontinuation vs Continuation | Depression | Income_quantitative | 0.941 | 0.905 | 0.978 | 2.02e-03 | NA | 3862 | 2456 | 1406 |
| Latino/Mixed-American | Full Cohort | Discontinuation vs Continuation | Depression | Education_levelCollege Undergraduate | 1.480 | 1.149 | 1.908 | 2.45e-03 | NA | 3862 | 2456 | 1406 |
| Latino/Mixed-American | Full Cohort | Discontinuation vs Continuation | Depression | Education_levelLess than a high school degree | 1.095 | 0.805 | 1.488 | 5.64e-01 | NA | 3862 | 2456 | 1406 |
| Latino/Mixed-American | Full Cohort | Discontinuation vs Continuation | Depression | Education_levelTwelve or GED | 1.390 | 1.040 | 1.858 | 2.60e-02 | NA | 3862 | 2456 | 1406 |
| Latino/Mixed-American | Full Cohort | Discontinuation vs Continuation | Depression | Depression | 1.148 | 1.065 | 1.238 | 3.13e-04 | 2.19e-03 | 3862 | 2456 | 1406 |
| Latino/Mixed-American | Full Cohort | Switching vs Continuation | Depression | (Intercept) | 1.233 | 0.880 | 1.728 | 2.24e-01 | NA | 3931 | 2456 | 1475 |
| Latino/Mixed-American | Full Cohort | Switching vs Continuation | Depression | age_at_first_AD | 0.986 | 0.982 | 0.991 | 4.95e-09 | NA | 3931 | 2456 | 1475 |
| Latino/Mixed-American | Full Cohort | Switching vs Continuation | Depression | sex_at_birthMale | 1.063 | 0.916 | 1.234 | 4.20e-01 | NA | 3931 | 2456 | 1475 |
| Latino/Mixed-American | Full Cohort | Switching vs Continuation | Depression | Income_quantitative | 0.944 | 0.909 | 0.981 | 3.11e-03 | NA | 3931 | 2456 | 1475 |
| Latino/Mixed-American | Full Cohort | Switching vs Continuation | Depression | Education_levelCollege Undergraduate | 1.079 | 0.856 | 1.359 | 5.20e-01 | NA | 3931 | 2456 | 1475 |
| Latino/Mixed-American | Full Cohort | Switching vs Continuation | Depression | Education_levelLess than a high school degree | 0.988 | 0.743 | 1.314 | 9.32e-01 | NA | 3931 | 2456 | 1475 |
| Latino/Mixed-American | Full Cohort | Switching vs Continuation | Depression | Education_levelTwelve or GED | 0.950 | 0.724 | 1.246 | 7.10e-01 | NA | 3931 | 2456 | 1475 |
| Latino/Mixed-American | Full Cohort | Switching vs Continuation | Depression | Depression | 1.148 | 1.066 | 1.236 | 2.41e-04 | 1.69e-03 | 3931 | 2456 | 1475 |
| Latino/Mixed-American | Full Cohort | Discontinuation vs Continuation | Insomnia | (Intercept) | 0.487 | 0.342 | 0.694 | 6.85e-05 | NA | 3862 | 2456 | 1406 |

| | | | | | | | | | | | | |
|---|---|---|---|---|---|---|---|---|---|---|---|---|
| Latino/Mixed-American | Full Cohort | Discontinuation vs Continuation | Insomnia | age_at_first_AD | 1.000 | 0.995 | 1.004 | 8.28e-01 | NA | 3862 | 2456 | 1406 |
| Latino/Mixed-American | Full Cohort | Discontinuation vs Continuation | Insomnia | sex_at_birthMale | 1.224 | 1.058 | 1.417 | 6.50e-03 | NA | 3862 | 2456 | 1406 |
| Latino/Mixed-American | Full Cohort | Discontinuation vs Continuation | Insomnia | Income_quantitative | 0.939 | 0.903 | 0.976 | 1.43e-03 | NA | 3862 | 2456 | 1406 |
| Latino/Mixed-American | Full Cohort | Discontinuation vs Continuation | Insomnia | Education_levelCollege Undergraduate | 1.491 | 1.157 | 1.922 | 2.00e-03 | NA | 3862 | 2456 | 1406 |
| Latino/Mixed-American | Full Cohort | Discontinuation vs Continuation | Insomnia | Education_levelLess than a high school degree | 1.111 | 0.818 | 1.510 | 5.01e-01 | NA | 3862 | 2456 | 1406 |
| Latino/Mixed-American | Full Cohort | Discontinuation vs Continuation | Insomnia | Education_levelTwelve or GED | 1.403 | 1.050 | 1.874 | 2.21e-02 | NA | 3862 | 2456 | 1406 |
| Latino/Mixed-American | Full Cohort | Discontinuation vs Continuation | Insomnia | Insomnia | 1.016 | 0.941 | 1.096 | 6.90e-01 | 1.00e+00 | 3862 | 2456 | 1406 |
| Latino/Mixed-American | Full Cohort | Switching vs Continuation | Insomnia | (Intercept) | 1.249 | 0.892 | 1.751 | 1.96e-01 | NA | 3931 | 2456 | 1475 |
| Latino/Mixed-American | Full Cohort | Switching vs Continuation | Insomnia | age_at_first_AD | 0.986 | 0.982 | 0.991 | 3.79e-09 | NA | 3931 | 2456 | 1475 |
| Latino/Mixed-American | Full Cohort | Switching vs Continuation | Insomnia | sex_at_birthMale | 1.065 | 0.918 | 1.236 | 4.07e-01 | NA | 3931 | 2456 | 1475 |
| Latino/Mixed-American | Full Cohort | Switching vs Continuation | Insomnia | Income_quantitative | 0.942 | 0.906 | 0.978 | 1.85e-03 | NA | 3931 | 2456 | 1475 |
| Latino/Mixed-American | Full Cohort | Switching vs Continuation | Insomnia | Education_levelCollege Undergraduate | 1.074 | 0.852 | 1.353 | 5.46e-01 | NA | 3931 | 2456 | 1475 |
| Latino/Mixed-American | Full Cohort | Switching vs Continuation | Insomnia | Education_levelLess than a high school degree | 0.988 | 0.743 | 1.314 | 9.36e-01 | NA | 3931 | 2456 | 1475 |
| Latino/Mixed-American | Full Cohort | Switching vs Continuation | Insomnia | Education_levelTwelve or GED | 0.944 | 0.720 | 1.237 | 6.75e-01 | NA | 3931 | 2456 | 1475 |
| Latino/Mixed-American | Full Cohort | Switching vs Continuation | Insomnia | Insomnia | 1.063 | 0.986 | 1.146 | 1.11e-01 | 7.74e-01 | 3931 | 2456 | 1475 |
| Latino/Mixed-American | Full Cohort | Discontinuation vs Continuation | Migraine | (Intercept) | 0.487 | 0.342 | 0.694 | 6.78e-05 | NA | 3862 | 2456 | 1406 |
| Latino/Mixed-American | Full Cohort | Discontinuation vs Continuation | Migraine | age_at_first_AD | 1.000 | 0.995 | 1.004 | 8.43e-01 | NA | 3862 | 2456 | 1406 |
| Latino/Mixed-American | Full Cohort | Discontinuation vs Continuation | Migraine | sex_at_birthMale | 1.225 | 1.059 | 1.418 | 6.33e-03 | NA | 3862 | 2456 | 1406 |
| Latino/Mixed-American | Full Cohort | Discontinuation vs Continuation | Migraine | Income_quantitative | 0.939 | 0.903 | 0.976 | 1.35e-03 | NA | 3862 | 2456 | 1406 |
| Latino/Mixed-American | Full Cohort | Discontinuation vs Continuation | Migraine | Education_levelCollege Undergraduate | 1.490 | 1.157 | 1.920 | 2.03e-03 | NA | 3862 | 2456 | 1406 |
| Latino/Mixed-American | Full Cohort | Discontinuation vs Continuation | Migraine | Education_levelLess than a high school degree | 1.109 | 0.816 | 1.508 | 5.07e-01 | NA | 3862 | 2456 | 1406 |
| Latino/Mixed-American | Full Cohort | Discontinuation vs Continuation | Migraine | Education_levelTwelve or GED | 1.399 | 1.047 | 1.870 | 2.31e-02 | NA | 3862 | 2456 | 1406 |
| Latino/Mixed-American | Full Cohort | Discontinuation vs Continuation | Migraine | Migraine | 1.042 | 0.973 | 1.117 | 2.39e-01 | 1.00e+00 | 3862 | 2456 | 1406 |
| Latino/Mixed-American | Full Cohort | Switching vs Continuation | Migraine | (Intercept) | 1.248 | 0.891 | 1.748 | 1.98e-01 | NA | 3931 | 2456 | 1475 |
| Latino/Mixed-American | Full Cohort | Switching vs Continuation | Migraine | age_at_first_AD | 0.986 | 0.982 | 0.991 | 3.75e-09 | NA | 3931 | 2456 | 1475 |
| Latino/Mixed-American | Full Cohort | Switching vs Continuation | Migraine | sex_at_birthMale | 1.064 | 0.917 | 1.234 | 4.17e-01 | NA | 3931 | 2456 | 1475 |
| Latino/Mixed-American | Full Cohort | Switching vs Continuation | Migraine | Income_quantitative | 0.941 | 0.906 | 0.977 | 1.65e-03 | NA | 3931 | 2456 | 1475 |
| Latino/Mixed-American | Full Cohort | Switching vs Continuation | Migraine | Education_levelCollege Undergraduate | 1.079 | 0.857 | 1.360 | 5.16e-01 | NA | 3931 | 2456 | 1475 |
| Latino/Mixed-American | Full Cohort | Switching vs Continuation | Migraine | Education_levelLess than a high school degree | 0.993 | 0.747 | 1.320 | 9.59e-01 | NA | 3931 | 2456 | 1475 |
| Latino/Mixed-American | Full Cohort | Switching vs Continuation | Migraine | Education_levelTwelve or GED | 0.947 | 0.722 | 1.242 | 6.94e-01 | NA | 3931 | 2456 | 1475 |
| Latino/Mixed-American | Full Cohort | Switching vs Continuation | Migraine | Migraine | 1.027 | 0.960 | 1.099 | 4.39e-01 | 1.00e+00 | 3931 | 2456 | 1475 |
| Latino/Mixed-American | Full Cohort | Discontinuation vs Continuation | Schizophrenia | (Intercept) | 0.485 | 0.340 | 0.691 | 6.21e-05 | NA | 3862 | 2456 | 1406 |
| Latino/Mixed-American | Full Cohort | Discontinuation vs Continuation | Schizophrenia | age_at_first_AD | 0.999 | 0.995 | 1.004 | 8.04e-01 | NA | 3862 | 2456 | 1406 |
| Latino/Mixed-American | Full Cohort | Discontinuation vs Continuation | Schizophrenia | sex_at_birthMale | 1.223 | 1.057 | 1.416 | 6.70e-03 | NA | 3862 | 2456 | 1406 |
| Latino/Mixed-American | Full Cohort | Discontinuation vs Continuation | Schizophrenia | Income_quantitative | 0.940 | 0.904 | 0.977 | 1.58e-03 | NA | 3862 | 2456 | 1406 |
| Latino/Mixed-American | Full Cohort | Discontinuation vs Continuation | Schizophrenia | Education_levelCollege Undergraduate | 1.502 | 1.166 | 1.936 | 1.66e-03 | NA | 3862 | 2456 | 1406 |
| Latino/Mixed-American | Full Cohort | Discontinuation vs Continuation | Schizophrenia | Education_levelLess than a high school degree | 1.120 | 0.824 | 1.522 | 4.68e-01 | NA | 3862 | 2456 | 1406 |
| Latino/Mixed-American | Full Cohort | Discontinuation vs Continuation | Schizophrenia | Education_levelTwelve or GED | 1.409 | 1.055 | 1.883 | 2.04e-02 | NA | 3862 | 2456 | 1406 |
| Latino/Mixed-American | Full Cohort | Discontinuation vs Continuation | Schizophrenia | Schizophrenia | 1.047 | 0.978 | 1.120 | 1.85e-01 | 1.00e+00 | 3862 | 2456 | 1406 |
| Latino/Mixed-American | Full Cohort | Switching vs Continuation | Schizophrenia | (Intercept) | 1.240 | 0.885 | 1.737 | 2.11e-01 | NA | 3931 | 2456 | 1475 |
| Latino/Mixed-American | Full Cohort | Switching vs Continuation | Schizophrenia | age_at_first_AD | 0.986 | 0.982 | 0.991 | 3.28e-09 | NA | 3931 | 2456 | 1475 |
| Latino/Mixed-American | Full Cohort | Switching vs Continuation | Schizophrenia | sex_at_birthMale | 1.064 | 0.917 | 1.235 | 4.12e-01 | NA | 3931 | 2456 | 1475 |
| Latino/Mixed-American | Full Cohort | Switching vs Continuation | Schizophrenia | Income_quantitative | 0.942 | 0.907 | 0.978 | 1.88e-03 | NA | 3931 | 2456 | 1475 |
| Latino/Mixed-American | Full Cohort | Switching vs Continuation | Schizophrenia | Education_levelCollege Undergraduate | 1.086 | 0.862 | 1.369 | 4.82e-01 | NA | 3931 | 2456 | 1475 |
| Latino/Mixed-American | Full Cohort | Switching vs Continuation | Schizophrenia | Education_levelLess than a high school degree | 0.999 | 0.752 | 1.328 | 9.95e-01 | NA | 3931 | 2456 | 1475 |
| Latino/Mixed-American | Full Cohort | Switching vs Continuation | Schizophrenia | Education_levelTwelve or GED | 0.956 | 0.729 | 1.253 | 7.43e-01 | NA | 3931 | 2456 | 1475 |
| Latino/Mixed-American | Full Cohort | Switching vs Continuation | Schizophrenia | Schizophrenia | 1.038 | 0.972 | 1.108 | 2.65e-01 | 1.00e+00 | 3931 | 2456 | 1475 |
| Latino/Mixed-American | Recorded Depression | Discontinuation vs Continuation | ADHD | (Intercept) | 0.440 | 0.248 | 0.780 | 4.94e-03 | NA | 1805 | 1179 | 626 |
| Latino/Mixed-American | Recorded Depression | Discontinuation vs Continuation | ADHD | age_at_first_AD | 0.998 | 0.991 | 1.005 | 5.29e-01 | NA | 1805 | 1179 | 626 |
| Latino/Mixed-American | Recorded Depression | Discontinuation vs Continuation | ADHD | sex_at_birthMale | 1.131 | 0.911 | 1.404 | 2.64e-01 | NA | 1805 | 1179 | 626 |
| Latino/Mixed-American | Recorded Depression | Discontinuation vs Continuation | ADHD | Income_quantitative | 0.948 | 0.891 | 1.008 | 8.80e-02 | NA | 1805 | 1179 | 626 |
| Latino/Mixed-American | Recorded Depression | Discontinuation vs Continuation | ADHD | Education_levelCollege Undergraduate | 1.677 | 1.092 | 2.576 | 1.81e-02 | NA | 1805 | 1179 | 626 |
| Latino/Mixed-American | Recorded Depression | Discontinuation vs Continuation | ADHD | Education_levelLess than a high school degree | 1.327 | 0.811 | 2.171 | 2.60e-01 | NA | 1805 | 1179 | 626 |
| Latino/Mixed-American | Recorded Depression | Discontinuation vs Continuation | ADHD | Education_levelTwelve or GED | 1.331 | 0.829 | 2.135 | 2.36e-01 | NA | 1805 | 1179 | 626 |
| Latino/Mixed-American | Recorded Depression | Discontinuation vs Continuation | ADHD | ADHD | 0.984 | 0.882 | 1.099 | 7.79e-01 | 1.00e+00 | 1805 | 1179 | 626 |
| Latino/Mixed-American | Recorded Depression | Switching vs Continuation | ADHD | (Intercept) | 1.888 | 1.213 | 2.937 | 4.83e-03 | NA | 2309 | 1179 | 1130 |
| Latino/Mixed-American | Recorded Depression | Switching vs Continuation | ADHD | age_at_first_AD | 0.989 | 0.983 | 0.995 | 3.83e-04 | NA | 2309 | 1179 | 1130 |

| | | | | | | | | | | | | |
|---|---|---|---|---|---|---|---|---|---|---|---|---|
| Latino/Mixed-American | Recorded Depression | Switching vs Continuation | ADHD | sex_at_birthMale | 0.965 | 0.799 | 1.165 | 7.08e-01 | NA | 2309 | 1179 | 1130 |
| Latino/Mixed-American | Recorded Depression | Switching vs Continuation | ADHD | Income_quantitative | 0.970 | 0.922 | 1.020 | 2.34e-01 | NA | 2309 | 1179 | 1130 |
| Latino/Mixed-American | Recorded Depression | Switching vs Continuation | ADHD | Education_levelCollege Undergraduate | 0.953 | 0.701 | 1.294 | 7.56e-01 | NA | 2309 | 1179 | 1130 |
| Latino/Mixed-American | Recorded Depression | Switching vs Continuation | ADHD | Education_levelLess than a high school degree | 0.858 | 0.595 | 1.237 | 4.12e-01 | NA | 2309 | 1179 | 1130 |
| Latino/Mixed-American | Recorded Depression | Switching vs Continuation | ADHD | Education_levelTwelve or GED | 0.724 | 0.512 | 1.025 | 6.84e-02 | NA | 2309 | 1179 | 1130 |
| Latino/Mixed-American | Recorded Depression | Switching vs Continuation | ADHD | ADHD | 0.987 | 0.900 | 1.083 | 7.87e-01 | 1.00e+00 | 2309 | 1179 | 1130 |
| Latino/Mixed-American | Recorded Depression | Discontinuation vs Continuation | BMI | (Intercept) | 0.436 | 0.246 | 0.774 | 4.58e-03 | NA | 1805 | 1179 | 626 |
| Latino/Mixed-American | Recorded Depression | Discontinuation vs Continuation | BMI | age_at_first_AD | 0.998 | 0.991 | 1.005 | 5.07e-01 | NA | 1805 | 1179 | 626 |
| Latino/Mixed-American | Recorded Depression | Discontinuation vs Continuation | BMI | sex_at_birthMale | 1.131 | 0.911 | 1.405 | 2.65e-01 | NA | 1805 | 1179 | 626 |
| Latino/Mixed-American | Recorded Depression | Discontinuation vs Continuation | BMI | Income_quantitative | 0.954 | 0.896 | 1.014 | 1.32e-01 | NA | 1805 | 1179 | 626 |
| Latino/Mixed-American | Recorded Depression | Discontinuation vs Continuation | BMI | Education_levelCollege Undergraduate | 1.670 | 1.087 | 2.567 | 1.93e-02 | NA | 1805 | 1179 | 626 |
| Latino/Mixed-American | Recorded Depression | Discontinuation vs Continuation | BMI | Education_levelLess than a high school degree | 1.312 | 0.801 | 2.148 | 2.80e-01 | NA | 1805 | 1179 | 626 |
| Latino/Mixed-American | Recorded Depression | Discontinuation vs Continuation | BMI | Education_levelTwelve or GED | 1.328 | 0.827 | 2.132 | 2.41e-01 | NA | 1805 | 1179 | 626 |
| Latino/Mixed-American | Recorded Depression | Discontinuation vs Continuation | BMI | BMI | 1.136 | 1.025 | 1.258 | 1.52e-02 | 1.06e-01 | 1805 | 1179 | 626 |
| Latino/Mixed-American | Recorded Depression | Switching vs Continuation | BMI | (Intercept) | 1.888 | 1.214 | 2.938 | 4.82e-03 | NA | 2309 | 1179 | 1130 |
| Latino/Mixed-American | Recorded Depression | Switching vs Continuation | BMI | age_at_first_AD | 0.989 | 0.984 | 0.995 | 3.91e-04 | NA | 2309 | 1179 | 1130 |
| Latino/Mixed-American | Recorded Depression | Switching vs Continuation | BMI | sex_at_birthMale | 0.965 | 0.799 | 1.165 | 7.08e-01 | NA | 2309 | 1179 | 1130 |
| Latino/Mixed-American | Recorded Depression | Switching vs Continuation | BMI | Income_quantitative | 0.970 | 0.923 | 1.020 | 2.41e-01 | NA | 2309 | 1179 | 1130 |
| Latino/Mixed-American | Recorded Depression | Switching vs Continuation | BMI | Education_levelCollege Undergraduate | 0.951 | 0.700 | 1.292 | 7.48e-01 | NA | 2309 | 1179 | 1130 |
| Latino/Mixed-American | Recorded Depression | Switching vs Continuation | BMI | Education_levelLess than a high school degree | 0.855 | 0.594 | 1.232 | 4.02e-01 | NA | 2309 | 1179 | 1130 |
| Latino/Mixed-American | Recorded Depression | Switching vs Continuation | BMI | Education_levelTwelve or GED | 0.722 | 0.510 | 1.022 | 6.61e-02 | NA | 2309 | 1179 | 1130 |
| Latino/Mixed-American | Recorded Depression | Switching vs Continuation | BMI | BMI | 1.007 | 0.924 | 1.098 | 8.72e-01 | 1.00e+00 | 2309 | 1179 | 1130 |
| Latino/Mixed-American | Recorded Depression | Discontinuation vs Continuation | Bipolar Disorder | (Intercept) | 0.436 | 0.246 | 0.773 | 4.49e-03 | NA | 1805 | 1179 | 626 |
| Latino/Mixed-American | Recorded Depression | Discontinuation vs Continuation | Bipolar Disorder | age_at_first_AD | 0.998 | 0.991 | 1.005 | 5.32e-01 | NA | 1805 | 1179 | 626 |
| Latino/Mixed-American | Recorded Depression | Discontinuation vs Continuation | Bipolar Disorder | sex_at_birthMale | 1.133 | 0.913 | 1.407 | 2.57e-01 | NA | 1805 | 1179 | 626 |
| Latino/Mixed-American | Recorded Depression | Discontinuation vs Continuation | Bipolar Disorder | Income_quantitative | 0.948 | 0.892 | 1.008 | 9.08e-02 | NA | 1805 | 1179 | 626 |
| Latino/Mixed-American | Recorded Depression | Discontinuation vs Continuation | Bipolar Disorder | Education_levelCollege Undergraduate | 1.688 | 1.099 | 2.594 | 1.68e-02 | NA | 1805 | 1179 | 626 |
| Latino/Mixed-American | Recorded Depression | Discontinuation vs Continuation | Bipolar Disorder | Education_levelLess than a high school degree | 1.337 | 0.817 | 2.188 | 2.48e-01 | NA | 1805 | 1179 | 626 |
| Latino/Mixed-American | Recorded Depression | Discontinuation vs Continuation | Bipolar Disorder | Education_levelTwelve or GED | 1.340 | 0.835 | 2.151 | 2.25e-01 | NA | 1805 | 1179 | 626 |
| Latino/Mixed-American | Recorded Depression | Discontinuation vs Continuation | Bipolar Disorder | Bipolar Disorder | 1.046 | 0.940 | 1.164 | 4.12e-01 | 1.00e+00 | 1805 | 1179 | 626 |
| Latino/Mixed-American | Recorded Depression | Switching vs Continuation | Bipolar Disorder | (Intercept) | 1.874 | 1.204 | 2.917 | 5.36e-03 | NA | 2309 | 1179 | 1130 |
| Latino/Mixed-American | Recorded Depression | Switching vs Continuation | Bipolar Disorder | age_at_first_AD | 0.989 | 0.983 | 0.995 | 3.71e-04 | NA | 2309 | 1179 | 1130 |
| Latino/Mixed-American | Recorded Depression | Switching vs Continuation | Bipolar Disorder | sex_at_birthMale | 0.963 | 0.797 | 1.163 | 6.94e-01 | NA | 2309 | 1179 | 1130 |
| Latino/Mixed-American | Recorded Depression | Switching vs Continuation | Bipolar Disorder | Income_quantitative | 0.971 | 0.923 | 1.021 | 2.47e-01 | NA | 2309 | 1179 | 1130 |
| Latino/Mixed-American | Recorded Depression | Switching vs Continuation | Bipolar Disorder | Education_levelCollege Undergraduate | 0.961 | 0.707 | 1.306 | 7.98e-01 | NA | 2309 | 1179 | 1130 |
| Latino/Mixed-American | Recorded Depression | Switching vs Continuation | Bipolar Disorder | Education_levelLess than a high school degree | 0.863 | 0.599 | 1.244 | 4.30e-01 | NA | 2309 | 1179 | 1130 |
| Latino/Mixed-American | Recorded Depression | Switching vs Continuation | Bipolar Disorder | Education_levelTwelve or GED | 0.729 | 0.515 | 1.032 | 7.46e-02 | NA | 2309 | 1179 | 1130 |
| Latino/Mixed-American | Recorded Depression | Switching vs Continuation | Bipolar Disorder | Bipolar Disorder | 1.058 | 0.966 | 1.160 | 2.25e-01 | 1.00e+00 | 2309 | 1179 | 1130 |
| Latino/Mixed-American | Recorded Depression | Discontinuation vs Continuation | Depression | (Intercept) | 0.437 | 0.247 | 0.776 | 4.67e-03 | NA | 1805 | 1179 | 626 |
| Latino/Mixed-American | Recorded Depression | Discontinuation vs Continuation | Depression | age_at_first_AD | 0.998 | 0.991 | 1.005 | 5.98e-01 | NA | 1805 | 1179 | 626 |
| Latino/Mixed-American | Recorded Depression | Discontinuation vs Continuation | Depression | sex_at_birthMale | 1.125 | 0.906 | 1.397 | 2.88e-01 | NA | 1805 | 1179 | 626 |
| Latino/Mixed-American | Recorded Depression | Discontinuation vs Continuation | Depression | Income_quantitative | 0.949 | 0.892 | 1.009 | 9.70e-02 | NA | 1805 | 1179 | 626 |
| Latino/Mixed-American | Recorded Depression | Discontinuation vs Continuation | Depression | Education_levelCollege Undergraduate | 1.663 | 1.082 | 2.555 | 2.04e-02 | NA | 1805 | 1179 | 626 |
| Latino/Mixed-American | Recorded Depression | Discontinuation vs Continuation | Depression | Education_levelLess than a high school degree | 1.303 | 0.796 | 2.133 | 2.93e-01 | NA | 1805 | 1179 | 626 |
| Latino/Mixed-American | Recorded Depression | Discontinuation vs Continuation | Depression | Education_levelTwelve or GED | 1.315 | 0.819 | 2.112 | 2.56e-01 | NA | 1805 | 1179 | 626 |
| Latino/Mixed-American | Recorded Depression | Discontinuation vs Continuation | Depression | Depression | 1.134 | 1.014 | 1.269 | 2.76e-02 | 1.94e-01 | 1805 | 1179 | 626 |
| Latino/Mixed-American | Recorded Depression | Switching vs Continuation | Depression | (Intercept) | 1.879 | 1.207 | 2.925 | 5.23e-03 | NA | 2309 | 1179 | 1130 |
| Latino/Mixed-American | Recorded Depression | Switching vs Continuation | Depression | age_at_first_AD | 0.989 | 0.984 | 0.995 | 4.65e-04 | NA | 2309 | 1179 | 1130 |
| Latino/Mixed-American | Recorded Depression | Switching vs Continuation | Depression | sex_at_birthMale | 0.964 | 0.798 | 1.164 | 7.03e-01 | NA | 2309 | 1179 | 1130 |
| Latino/Mixed-American | Recorded Depression | Switching vs Continuation | Depression | Income_quantitative | 0.971 | 0.923 | 1.021 | 2.53e-01 | NA | 2309 | 1179 | 1130 |
| Latino/Mixed-American | Recorded Depression | Switching vs Continuation | Depression | Education_levelCollege Undergraduate | 0.952 | 0.701 | 1.294 | 7.55e-01 | NA | 2309 | 1179 | 1130 |
| Latino/Mixed-American | Recorded Depression | Switching vs Continuation | Depression | Education_levelLess than a high school degree | 0.849 | 0.589 | 1.223 | 3.79e-01 | NA | 2309 | 1179 | 1130 |
| Latino/Mixed-American | Recorded Depression | Switching vs Continuation | Depression | Education_levelTwelve or GED | 0.722 | 0.510 | 1.023 | 6.68e-02 | NA | 2309 | 1179 | 1130 |
| Latino/Mixed-American | Recorded Depression | Switching vs Continuation | Depression | Depression | 1.117 | 1.017 | 1.225 | 2.00e-02 | 1.40e-01 | 2309 | 1179 | 1130 |
| Latino/Mixed-American | Recorded Depression | Discontinuation vs Continuation | Insomnia | (Intercept) | 0.438 | 0.247 | 0.777 | 4.71e-03 | NA | 1805 | 1179 | 626 |
| Latino/Mixed-American | Recorded Depression | Discontinuation vs Continuation | Insomnia | age_at_first_AD | 0.998 | 0.991 | 1.005 | 5.34e-01 | NA | 1805 | 1179 | 626 |
| Latino/Mixed-American | Recorded Depression | Discontinuation vs Continuation | Insomnia | sex_at_birthMale | 1.132 | 0.912 | 1.405 | 2.62e-01 | NA | 1805 | 1179 | 626 |

| | | | | | | | | | | | | |
|---|---|---|---|---|---|---|---|---|---|---|---|---|
| Latino/Mixed-American | Recorded Depression | Discontinuation vs Continuation | Insomnia | Income_quantitative | 0.948 | 0.891 | 1.008 | 8.90e-02 | NA | 1805 | 1179 | 626 |
| Latino/Mixed-American | Recorded Depression | Discontinuation vs Continuation | Insomnia | Education_levelCollege Undergraduate | 1.681 | 1.094 | 2.582 | 1.77e-02 | NA | 1805 | 1179 | 626 |
| Latino/Mixed-American | Recorded Depression | Discontinuation vs Continuation | Insomnia | Education_levelLess than a high school degree | 1.328 | 0.812 | 2.172 | 2.59e-01 | NA | 1805 | 1179 | 626 |
| Latino/Mixed-American | Recorded Depression | Discontinuation vs Continuation | Insomnia | Education_levelTwelve or GED | 1.332 | 0.830 | 2.138 | 2.34e-01 | NA | 1805 | 1179 | 626 |
| Latino/Mixed-American | Recorded Depression | Discontinuation vs Continuation | Insomnia | Insomnia | 0.976 | 0.872 | 1.093 | 6.73e-01 | 1.00e+00 | 1805 | 1179 | 626 |
| Latino/Mixed-American | Recorded Depression | Switching vs Continuation | Insomnia | (Intercept) | 1.890 | 1.215 | 2.940 | 4.77e-03 | NA | 2309 | 1179 | 1130 |
| Latino/Mixed-American | Recorded Depression | Switching vs Continuation | Insomnia | age_at_first_AD | 0.989 | 0.984 | 0.995 | 4.02e-04 | NA | 2309 | 1179 | 1130 |
| Latino/Mixed-American | Recorded Depression | Switching vs Continuation | Insomnia | sex_at_birthMale | 0.965 | 0.799 | 1.165 | 7.09e-01 | NA | 2309 | 1179 | 1130 |
| Latino/Mixed-American | Recorded Depression | Switching vs Continuation | Insomnia | Income_quantitative | 0.970 | 0.922 | 1.020 | 2.37e-01 | NA | 2309 | 1179 | 1130 |
| Latino/Mixed-American | Recorded Depression | Switching vs Continuation | Insomnia | Education_levelCollege Undergraduate | 0.949 | 0.699 | 1.289 | 7.39e-01 | NA | 2309 | 1179 | 1130 |
| Latino/Mixed-American | Recorded Depression | Switching vs Continuation | Insomnia | Education_levelLess than a high school degree | 0.853 | 0.592 | 1.229 | 3.95e-01 | NA | 2309 | 1179 | 1130 |
| Latino/Mixed-American | Recorded Depression | Switching vs Continuation | Insomnia | Education_levelTwelve or GED | 0.721 | 0.509 | 1.020 | 6.46e-02 | NA | 2309 | 1179 | 1130 |
| Latino/Mixed-American | Recorded Depression | Switching vs Continuation | Insomnia | Insomnia | 1.038 | 0.945 | 1.139 | 4.38e-01 | 1.00e+00 | 2309 | 1179 | 1130 |
| Latino/Mixed-American | Recorded Depression | Discontinuation vs Continuation | Migraine | (Intercept) | 0.441 | 0.249 | 0.783 | 5.13e-03 | NA | 1805 | 1179 | 626 |
| Latino/Mixed-American | Recorded Depression | Discontinuation vs Continuation | Migraine | age_at_first_AD | 0.998 | 0.991 | 1.005 | 5.39e-01 | NA | 1805 | 1179 | 626 |
| Latino/Mixed-American | Recorded Depression | Discontinuation vs Continuation | Migraine | sex_at_birthMale | 1.135 | 0.914 | 1.410 | 2.51e-01 | NA | 1805 | 1179 | 626 |
| Latino/Mixed-American | Recorded Depression | Discontinuation vs Continuation | Migraine | Income_quantitative | 0.948 | 0.891 | 1.008 | 8.91e-02 | NA | 1805 | 1179 | 626 |
| Latino/Mixed-American | Recorded Depression | Discontinuation vs Continuation | Migraine | Education_levelCollege Undergraduate | 1.673 | 1.089 | 2.571 | 1.88e-02 | NA | 1805 | 1179 | 626 |
| Latino/Mixed-American | Recorded Depression | Discontinuation vs Continuation | Migraine | Education_levelLess than a high school degree | 1.313 | 0.802 | 2.149 | 2.79e-01 | NA | 1805 | 1179 | 626 |
| Latino/Mixed-American | Recorded Depression | Discontinuation vs Continuation | Migraine | Education_levelTwelve or GED | 1.312 | 0.817 | 2.106 | 2.61e-01 | NA | 1805 | 1179 | 626 |
| Latino/Mixed-American | Recorded Depression | Discontinuation vs Continuation | Migraine | Migraine | 1.103 | 0.993 | 1.226 | 6.69e-02 | 4.68e-01 | 1805 | 1179 | 626 |
| Latino/Mixed-American | Recorded Depression | Switching vs Continuation | Migraine | (Intercept) | 1.907 | 1.225 | 2.969 | 4.24e-03 | NA | 2309 | 1179 | 1130 |
| Latino/Mixed-American | Recorded Depression | Switching vs Continuation | Migraine | age_at_first_AD | 0.989 | 0.984 | 0.995 | 4.15e-04 | NA | 2309 | 1179 | 1130 |
| Latino/Mixed-American | Recorded Depression | Switching vs Continuation | Migraine | sex_at_birthMale | 0.965 | 0.799 | 1.165 | 7.10e-01 | NA | 2309 | 1179 | 1130 |
| Latino/Mixed-American | Recorded Depression | Switching vs Continuation | Migraine | Income_quantitative | 0.969 | 0.921 | 1.019 | 2.17e-01 | NA | 2309 | 1179 | 1130 |
| Latino/Mixed-American | Recorded Depression | Switching vs Continuation | Migraine | Education_levelCollege Undergraduate | 0.949 | 0.698 | 1.289 | 7.37e-01 | NA | 2309 | 1179 | 1130 |
| Latino/Mixed-American | Recorded Depression | Switching vs Continuation | Migraine | Education_levelLess than a high school degree | 0.845 | 0.587 | 1.218 | 3.68e-01 | NA | 2309 | 1179 | 1130 |
| Latino/Mixed-American | Recorded Depression | Switching vs Continuation | Migraine | Education_levelTwelve or GED | 0.711 | 0.502 | 1.006 | 5.42e-02 | NA | 2309 | 1179 | 1130 |
| Latino/Mixed-American | Recorded Depression | Switching vs Continuation | Migraine | Migraine | 1.090 | 1.001 | 1.188 | 4.86e-02 | 3.40e-01 | 2309 | 1179 | 1130 |
| Latino/Mixed-American | Recorded Depression | Discontinuation vs Continuation | Schizophrenia | (Intercept) | 0.430 | 0.242 | 0.763 | 3.90e-03 | NA | 1805 | 1179 | 626 |
| Latino/Mixed-American | Recorded Depression | Discontinuation vs Continuation | Schizophrenia | age_at_first_AD | 0.998 | 0.991 | 1.005 | 5.42e-01 | NA | 1805 | 1179 | 626 |
| Latino/Mixed-American | Recorded Depression | Discontinuation vs Continuation | Schizophrenia | sex_at_birthMale | 1.135 | 0.914 | 1.410 | 2.52e-01 | NA | 1805 | 1179 | 626 |
| Latino/Mixed-American | Recorded Depression | Discontinuation vs Continuation | Schizophrenia | Income_quantitative | 0.948 | 0.892 | 1.008 | 9.05e-02 | NA | 1805 | 1179 | 626 |
| Latino/Mixed-American | Recorded Depression | Discontinuation vs Continuation | Schizophrenia | Education_levelCollege Undergraduate | 1.712 | 1.114 | 2.632 | 1.42e-02 | NA | 1805 | 1179 | 626 |
| Latino/Mixed-American | Recorded Depression | Discontinuation vs Continuation | Schizophrenia | Education_levelLess than a high school degree | 1.347 | 0.823 | 2.205 | 2.36e-01 | NA | 1805 | 1179 | 626 |
| Latino/Mixed-American | Recorded Depression | Discontinuation vs Continuation | Schizophrenia | Education_levelTwelve or GED | 1.348 | 0.840 | 2.165 | 2.16e-01 | NA | 1805 | 1179 | 626 |
| Latino/Mixed-American | Recorded Depression | Discontinuation vs Continuation | Schizophrenia | Schizophrenia | 1.112 | 1.005 | 1.229 | 3.94e-02 | 2.76e-01 | 1805 | 1179 | 626 |
| Latino/Mixed-American | Recorded Depression | Switching vs Continuation | Schizophrenia | (Intercept) | 1.881 | 1.209 | 2.926 | 5.10e-03 | NA | 2309 | 1179 | 1130 |
| Latino/Mixed-American | Recorded Depression | Switching vs Continuation | Schizophrenia | age_at_first_AD | 0.989 | 0.984 | 0.995 | 3.93e-04 | NA | 2309 | 1179 | 1130 |
| Latino/Mixed-American | Recorded Depression | Switching vs Continuation | Schizophrenia | sex_at_birthMale | 0.965 | 0.799 | 1.165 | 7.08e-01 | NA | 2309 | 1179 | 1130 |
| Latino/Mixed-American | Recorded Depression | Switching vs Continuation | Schizophrenia | Income_quantitative | 0.970 | 0.922 | 1.020 | 2.35e-01 | NA | 2309 | 1179 | 1130 |
| Latino/Mixed-American | Recorded Depression | Switching vs Continuation | Schizophrenia | Education_levelCollege Undergraduate | 0.957 | 0.704 | 1.301 | 7.79e-01 | NA | 2309 | 1179 | 1130 |
| Latino/Mixed-American | Recorded Depression | Switching vs Continuation | Schizophrenia | Education_levelLess than a high school degree | 0.858 | 0.596 | 1.236 | 4.11e-01 | NA | 2309 | 1179 | 1130 |
| Latino/Mixed-American | Recorded Depression | Switching vs Continuation | Schizophrenia | Education_levelTwelve or GED | 0.726 | 0.513 | 1.028 | 7.11e-02 | NA | 2309 | 1179 | 1130 |
| Latino/Mixed-American | Recorded Depression | Switching vs Continuation | Schizophrenia | Schizophrenia | 1.029 | 0.946 | 1.118 | 5.09e-01 | 1.00e+00 | 2309 | 1179 | 1130 |
| Latino/Mixed-American | No Recorded Depression | Discontinuation vs Continuation | ADHD | (Intercept) | 0.543 | 0.344 | 0.858 | 8.94e-03 | NA | 2057 | 1277 | 780 |
| Latino/Mixed-American | No Recorded Depression | Discontinuation vs Continuation | ADHD | age_at_first_AD | 1.000 | 0.994 | 1.006 | 9.47e-01 | NA | 2057 | 1277 | 780 |
| Latino/Mixed-American | No Recorded Depression | Discontinuation vs Continuation | ADHD | sex_at_birthMale | 1.310 | 1.073 | 1.598 | 7.85e-03 | NA | 2057 | 1277 | 780 |
| Latino/Mixed-American | No Recorded Depression | Discontinuation vs Continuation | ADHD | Income_quantitative | 0.927 | 0.881 | 0.975 | 3.27e-03 | NA | 2057 | 1277 | 780 |
| Latino/Mixed-American | No Recorded Depression | Discontinuation vs Continuation | ADHD | Education_levelCollege Undergraduate | 1.387 | 1.012 | 1.902 | 4.21e-02 | NA | 2057 | 1277 | 780 |
| Latino/Mixed-American | No Recorded Depression | Discontinuation vs Continuation | ADHD | Education_levelLess than a high school degree | 0.969 | 0.649 | 1.446 | 8.77e-01 | NA | 2057 | 1277 | 780 |
| Latino/Mixed-American | No Recorded Depression | Discontinuation vs Continuation | ADHD | Education_levelTwelve or GED | 1.556 | 1.069 | 2.264 | 2.09e-02 | NA | 2057 | 1277 | 780 |
| Latino/Mixed-American | No Recorded Depression | Discontinuation vs Continuation | ADHD | ADHD | 1.079 | 0.973 | 1.195 | 1.49e-01 | 1.00e+00 | 2057 | 1277 | 780 |
| Latino/Mixed-American | No Recorded Depression | Switching vs Continuation | ADHD | (Intercept) | 0.408 | 0.225 | 0.740 | 3.21e-03 | NA | 1622 | 1277 | 345 |
| Latino/Mixed-American | No Recorded Depression | Switching vs Continuation | ADHD | age_at_first_AD | 0.984 | 0.975 | 0.992 | 1.90e-04 | NA | 1622 | 1277 | 345 |
| Latino/Mixed-American | No Recorded Depression | Switching vs Continuation | ADHD | sex_at_birthMale | 1.245 | 0.950 | 1.632 | 1.12e-01 | NA | 1622 | 1277 | 345 |
| Latino/Mixed-American | No Recorded Depression | Switching vs Continuation | ADHD | Income_quantitative | 1.001 | 0.938 | 1.069 | 9.66e-01 | NA | 1622 | 1277 | 345 |

| | | | | | | | | | | | | |
|---|---|---|---|---|---|---|---|---|---|---|---|---|
| Latino/Mixed-American | No Recorded Depression | Switching vs Continuation | ADHD | Education_levelCollege Undergraduate | 1.360 | 0.908 | 2.038 | 1.36e-01 | NA | 1622 | 1277 | 345 |
| Latino/Mixed-American | No Recorded Depression | Switching vs Continuation | ADHD | Education_levelLess than a high school degree | 1.173 | 0.690 | 1.994 | 5.56e-01 | NA | 1622 | 1277 | 345 |
| Latino/Mixed-American | No Recorded Depression | Switching vs Continuation | ADHD | Education_levelTwelve or GED | 1.313 | 0.796 | 2.166 | 2.86e-01 | NA | 1622 | 1277 | 345 |
| Latino/Mixed-American | No Recorded Depression | Switching vs Continuation | ADHD | ADHD | 1.081 | 0.949 | 1.231 | 2.43e-01 | 1.00e+00 | 1622 | 1277 | 345 |
| Latino/Mixed-American | No Recorded Depression | Discontinuation vs Continuation | BMI | (Intercept) | 0.541 | 0.342 | 0.855 | 8.56e-03 | NA | 2057 | 1277 | 780 |
| Latino/Mixed-American | No Recorded Depression | Discontinuation vs Continuation | BMI | age_at_first_AD | 1.000 | 0.994 | 1.006 | 9.84e-01 | NA | 2057 | 1277 | 780 |
| Latino/Mixed-American | No Recorded Depression | Discontinuation vs Continuation | BMI | sex_at_birthMale | 1.310 | 1.074 | 1.598 | 7.76e-03 | NA | 2057 | 1277 | 780 |
| Latino/Mixed-American | No Recorded Depression | Discontinuation vs Continuation | BMI | Income_quantitative | 0.927 | 0.882 | 0.976 | 3.55e-03 | NA | 2057 | 1277 | 780 |
| Latino/Mixed-American | No Recorded Depression | Discontinuation vs Continuation | BMI | Education_levelCollege Undergraduate | 1.394 | 1.017 | 1.911 | 3.91e-02 | NA | 2057 | 1277 | 780 |
| Latino/Mixed-American | No Recorded Depression | Discontinuation vs Continuation | BMI | Education_levelLess than a high school degree | 0.986 | 0.661 | 1.471 | 9.47e-01 | NA | 2057 | 1277 | 780 |
| Latino/Mixed-American | No Recorded Depression | Discontinuation vs Continuation | BMI | Education_levelTwelve or GED | 1.580 | 1.087 | 2.296 | 1.66e-02 | NA | 2057 | 1277 | 780 |
| Latino/Mixed-American | No Recorded Depression | Discontinuation vs Continuation | BMI | BMI | 1.066 | 0.970 | 1.171 | 1.87e-01 | 1.00e+00 | 2057 | 1277 | 780 |
| Latino/Mixed-American | No Recorded Depression | Switching vs Continuation | BMI | (Intercept) | 0.407 | 0.224 | 0.739 | 3.16e-03 | NA | 1622 | 1277 | 345 |
| Latino/Mixed-American | No Recorded Depression | Switching vs Continuation | BMI | age_at_first_AD | 0.984 | 0.975 | 0.992 | 1.59e-04 | NA | 1622 | 1277 | 345 |
| Latino/Mixed-American | No Recorded Depression | Switching vs Continuation | BMI | sex_at_birthMale | 1.250 | 0.954 | 1.638 | 1.06e-01 | NA | 1622 | 1277 | 345 |
| Latino/Mixed-American | No Recorded Depression | Switching vs Continuation | BMI | Income_quantitative | 1.002 | 0.938 | 1.070 | 9.55e-01 | NA | 1622 | 1277 | 345 |
| Latino/Mixed-American | No Recorded Depression | Switching vs Continuation | BMI | Education_levelCollege Undergraduate | 1.367 | 0.912 | 2.047 | 1.30e-01 | NA | 1622 | 1277 | 345 |
| Latino/Mixed-American | No Recorded Depression | Switching vs Continuation | BMI | Education_levelLess than a high school degree | 1.199 | 0.706 | 2.037 | 5.03e-01 | NA | 1622 | 1277 | 345 |
| Latino/Mixed-American | No Recorded Depression | Switching vs Continuation | BMI | Education_levelTwelve or GED | 1.332 | 0.808 | 2.196 | 2.61e-01 | NA | 1622 | 1277 | 345 |
| Latino/Mixed-American | No Recorded Depression | Switching vs Continuation | BMI | BMI | 1.063 | 0.937 | 1.205 | 3.42e-01 | 1.00e+00 | 1622 | 1277 | 345 |
| Latino/Mixed-American | No Recorded Depression | Discontinuation vs Continuation | Bipolar Disorder | (Intercept) | 0.546 | 0.346 | 0.863 | 9.62e-03 | NA | 2057 | 1277 | 780 |
| Latino/Mixed-American | No Recorded Depression | Discontinuation vs Continuation | Bipolar Disorder | age_at_first_AD | 1.000 | 0.994 | 1.006 | 9.77e-01 | NA | 2057 | 1277 | 780 |
| Latino/Mixed-American | No Recorded Depression | Discontinuation vs Continuation | Bipolar Disorder | sex_at_birthMale | 1.309 | 1.073 | 1.597 | 7.88e-03 | NA | 2057 | 1277 | 780 |
| Latino/Mixed-American | No Recorded Depression | Discontinuation vs Continuation | Bipolar Disorder | Income_quantitative | 0.925 | 0.879 | 0.973 | 2.41e-03 | NA | 2057 | 1277 | 780 |
| Latino/Mixed-American | No Recorded Depression | Discontinuation vs Continuation | Bipolar Disorder | Education_levelCollege Undergraduate | 1.404 | 1.024 | 1.924 | 3.51e-02 | NA | 2057 | 1277 | 780 |
| Latino/Mixed-American | No Recorded Depression | Discontinuation vs Continuation | Bipolar Disorder | Education_levelLess than a high school degree | 0.986 | 0.661 | 1.471 | 9.46e-01 | NA | 2057 | 1277 | 780 |
| Latino/Mixed-American | No Recorded Depression | Discontinuation vs Continuation | Bipolar Disorder | Education_levelTwelve or GED | 1.595 | 1.097 | 2.318 | 1.44e-02 | NA | 2057 | 1277 | 780 |
| Latino/Mixed-American | No Recorded Depression | Discontinuation vs Continuation | Bipolar Disorder | Bipolar Disorder | 1.055 | 0.952 | 1.168 | 3.08e-01 | 1.00e+00 | 2057 | 1277 | 780 |
| Latino/Mixed-American | No Recorded Depression | Switching vs Continuation | Bipolar Disorder | (Intercept) | 0.411 | 0.226 | 0.746 | 3.49e-03 | NA | 1622 | 1277 | 345 |
| Latino/Mixed-American | No Recorded Depression | Switching vs Continuation | Bipolar Disorder | age_at_first_AD | 0.984 | 0.975 | 0.992 | 1.46e-04 | NA | 1622 | 1277 | 345 |
| Latino/Mixed-American | No Recorded Depression | Switching vs Continuation | Bipolar Disorder | sex_at_birthMale | 1.245 | 0.950 | 1.632 | 1.12e-01 | NA | 1622 | 1277 | 345 |
| Latino/Mixed-American | No Recorded Depression | Switching vs Continuation | Bipolar Disorder | Income_quantitative | 1.000 | 0.936 | 1.067 | 9.87e-01 | NA | 1622 | 1277 | 345 |
| Latino/Mixed-American | No Recorded Depression | Switching vs Continuation | Bipolar Disorder | Education_levelCollege Undergraduate | 1.371 | 0.915 | 2.054 | 1.26e-01 | NA | 1622 | 1277 | 345 |
| Latino/Mixed-American | No Recorded Depression | Switching vs Continuation | Bipolar Disorder | Education_levelLess than a high school degree | 1.195 | 0.704 | 2.031 | 5.09e-01 | NA | 1622 | 1277 | 345 |
| Latino/Mixed-American | No Recorded Depression | Switching vs Continuation | Bipolar Disorder | Education_levelTwelve or GED | 1.338 | 0.812 | 2.206 | 2.53e-01 | NA | 1622 | 1277 | 345 |
| Latino/Mixed-American | No Recorded Depression | Switching vs Continuation | Bipolar Disorder | Bipolar Disorder | 0.976 | 0.854 | 1.114 | 7.15e-01 | 1.00e+00 | 1622 | 1277 | 345 |
| Latino/Mixed-American | No Recorded Depression | Discontinuation vs Continuation | Depression | (Intercept) | 0.536 | 0.339 | 0.847 | 7.59e-03 | NA | 2057 | 1277 | 780 |
| Latino/Mixed-American | No Recorded Depression | Discontinuation vs Continuation | Depression | age_at_first_AD | 1.000 | 0.994 | 1.006 | 9.20e-01 | NA | 2057 | 1277 | 780 |
| Latino/Mixed-American | No Recorded Depression | Discontinuation vs Continuation | Depression | sex_at_birthMale | 1.311 | 1.074 | 1.600 | 7.73e-03 | NA | 2057 | 1277 | 780 |
| Latino/Mixed-American | No Recorded Depression | Discontinuation vs Continuation | Depression | Income_quantitative | 0.928 | 0.882 | 0.976 | 3.58e-03 | NA | 2057 | 1277 | 780 |
| Latino/Mixed-American | No Recorded Depression | Discontinuation vs Continuation | Depression | Education_levelCollege Undergraduate | 1.391 | 1.014 | 1.908 | 4.05e-02 | NA | 2057 | 1277 | 780 |
| Latino/Mixed-American | No Recorded Depression | Discontinuation vs Continuation | Depression | Education_levelLess than a high school degree | 0.972 | 0.651 | 1.451 | 8.90e-01 | NA | 2057 | 1277 | 780 |
| Latino/Mixed-American | No Recorded Depression | Discontinuation vs Continuation | Depression | Education_levelTwelve or GED | 1.582 | 1.088 | 2.300 | 1.63e-02 | NA | 2057 | 1277 | 780 |
| Latino/Mixed-American | No Recorded Depression | Discontinuation vs Continuation | Depression | Depression | 1.167 | 1.053 | 1.293 | 3.12e-03 | 2.18e-02 | 2057 | 1277 | 780 |
| Latino/Mixed-American | No Recorded Depression | Switching vs Continuation | Depression | (Intercept) | 0.405 | 0.223 | 0.735 | 2.98e-03 | NA | 1622 | 1277 | 345 |
| Latino/Mixed-American | No Recorded Depression | Switching vs Continuation | Depression | age_at_first_AD | 0.984 | 0.975 | 0.992 | 1.52e-04 | NA | 1622 | 1277 | 345 |
| Latino/Mixed-American | No Recorded Depression | Switching vs Continuation | Depression | sex_at_birthMale | 1.249 | 0.953 | 1.636 | 1.07e-01 | NA | 1622 | 1277 | 345 |
| Latino/Mixed-American | No Recorded Depression | Switching vs Continuation | Depression | Income_quantitative | 1.004 | 0.941 | 1.072 | 8.99e-01 | NA | 1622 | 1277 | 345 |
| Latino/Mixed-American | No Recorded Depression | Switching vs Continuation | Depression | Education_levelCollege Undergraduate | 1.368 | 0.914 | 2.049 | 1.28e-01 | NA | 1622 | 1277 | 345 |
| Latino/Mixed-American | No Recorded Depression | Switching vs Continuation | Depression | Education_levelLess than a high school degree | 1.193 | 0.702 | 2.026 | 5.14e-01 | NA | 1622 | 1277 | 345 |
| Latino/Mixed-American | No Recorded Depression | Switching vs Continuation | Depression | Education_levelTwelve or GED | 1.342 | 0.814 | 2.211 | 2.48e-01 | NA | 1622 | 1277 | 345 |
| Latino/Mixed-American | No Recorded Depression | Switching vs Continuation | Depression | Depression | 1.115 | 0.973 | 1.278 | 1.19e-01 | 8.30e-01 | 1622 | 1277 | 345 |
| Latino/Mixed-American | No Recorded Depression | Discontinuation vs Continuation | Insomnia | (Intercept) | 0.546 | 0.345 | 0.862 | 9.41e-03 | NA | 2057 | 1277 | 780 |
| Latino/Mixed-American | No Recorded Depression | Discontinuation vs Continuation | Insomnia | age_at_first_AD | 1.000 | 0.994 | 1.006 | 9.97e-01 | NA | 2057 | 1277 | 780 |
| Latino/Mixed-American | No Recorded Depression | Discontinuation vs Continuation | Insomnia | sex_at_birthMale | 1.312 | 1.076 | 1.601 | 7.41e-03 | NA | 2057 | 1277 | 780 |
| Latino/Mixed-American | No Recorded Depression | Discontinuation vs Continuation | Insomnia | Income_quantitative | 0.926 | 0.880 | 0.974 | 2.68e-03 | NA | 2057 | 1277 | 780 |
| Latino/Mixed-American | No Recorded Depression | Discontinuation vs Continuation | Insomnia | Education_levelCollege Undergraduate | 1.397 | 1.019 | 1.914 | 3.80e-02 | NA | 2057 | 1277 | 780 |

| | | | | | | | | | | | | |
|---|---|---|---|---|---|---|---|---|---|---|---|---|
| Latino/Mixed-American | No Recorded Depression | Discontinuation vs Continuation | Insomnia | Education_levelLess than a high school degree | 0.978 | 0.655 | 1.459 | 9.13e-01 | NA | 2057 | 1277 | 780 |
| Latino/Mixed-American | No Recorded Depression | Discontinuation vs Continuation | Insomnia | Education_levelTwelve or GED | 1.590 | 1.094 | 2.310 | 1.51e-02 | NA | 2057 | 1277 | 780 |
| Latino/Mixed-American | No Recorded Depression | Discontinuation vs Continuation | Insomnia | Insomnia | 1.053 | 0.948 | 1.169 | 3.36e-01 | 1.00e+00 | 2057 | 1277 | 780 |
| Latino/Mixed-American | No Recorded Depression | Switching vs Continuation | Insomnia | (Intercept) | 0.409 | 0.225 | 0.743 | 3.34e-03 | NA | 1622 | 1277 | 345 |
| Latino/Mixed-American | No Recorded Depression | Switching vs Continuation | Insomnia | age_at_first_AD | 0.984 | 0.975 | 0.992 | 1.46e-04 | NA | 1622 | 1277 | 345 |
| Latino/Mixed-American | No Recorded Depression | Switching vs Continuation | Insomnia | sex_at_birthMale | 1.249 | 0.953 | 1.636 | 1.07e-01 | NA | 1622 | 1277 | 345 |
| Latino/Mixed-American | No Recorded Depression | Switching vs Continuation | Insomnia | Income_quantitative | 1.000 | 0.937 | 1.068 | 9.95e-01 | NA | 1622 | 1277 | 345 |
| Latino/Mixed-American | No Recorded Depression | Switching vs Continuation | Insomnia | Education_levelCollege Undergraduate | 1.373 | 0.916 | 2.056 | 1.25e-01 | NA | 1622 | 1277 | 345 |
| Latino/Mixed-American | No Recorded Depression | Switching vs Continuation | Insomnia | Education_levelLess than a high school degree | 1.196 | 0.704 | 2.032 | 5.09e-01 | NA | 1622 | 1277 | 345 |
| Latino/Mixed-American | No Recorded Depression | Switching vs Continuation | Insomnia | Education_levelTwelve or GED | 1.340 | 0.813 | 2.210 | 2.51e-01 | NA | 1622 | 1277 | 345 |
| Latino/Mixed-American | No Recorded Depression | Switching vs Continuation | Insomnia | Insomnia | 1.006 | 0.873 | 1.160 | 9.30e-01 | 1.00e+00 | 1622 | 1277 | 345 |
| Latino/Mixed-American | No Recorded Depression | Discontinuation vs Continuation | Migraine | (Intercept) | 0.547 | 0.346 | 0.864 | 9.64e-03 | NA | 2057 | 1277 | 780 |
| Latino/Mixed-American | No Recorded Depression | Discontinuation vs Continuation | Migraine | age_at_first_AD | 1.000 | 0.994 | 1.006 | 9.78e-01 | NA | 2057 | 1277 | 780 |
| Latino/Mixed-American | No Recorded Depression | Discontinuation vs Continuation | Migraine | sex_at_birthMale | 1.308 | 1.073 | 1.596 | 8.01e-03 | NA | 2057 | 1277 | 780 |
| Latino/Mixed-American | No Recorded Depression | Discontinuation vs Continuation | Migraine | Income_quantitative | 0.925 | 0.880 | 0.973 | 2.53e-03 | NA | 2057 | 1277 | 780 |
| Latino/Mixed-American | No Recorded Depression | Discontinuation vs Continuation | Migraine | Education_levelCollege Undergraduate | 1.402 | 1.023 | 1.922 | 3.58e-02 | NA | 2057 | 1277 | 780 |
| Latino/Mixed-American | No Recorded Depression | Discontinuation vs Continuation | Migraine | Education_levelLess than a high school degree | 0.988 | 0.662 | 1.474 | 9.53e-01 | NA | 2057 | 1277 | 780 |
| Latino/Mixed-American | No Recorded Depression | Discontinuation vs Continuation | Migraine | Education_levelTwelve or GED | 1.592 | 1.095 | 2.313 | 1.48e-02 | NA | 2057 | 1277 | 780 |
| Latino/Mixed-American | No Recorded Depression | Discontinuation vs Continuation | Migraine | Migraine | 0.993 | 0.905 | 1.090 | 8.86e-01 | 1.00e+00 | 2057 | 1277 | 780 |
| Latino/Mixed-American | No Recorded Depression | Switching vs Continuation | Migraine | (Intercept) | 0.410 | 0.226 | 0.745 | 3.44e-03 | NA | 1622 | 1277 | 345 |
| Latino/Mixed-American | No Recorded Depression | Switching vs Continuation | Migraine | age_at_first_AD | 0.984 | 0.975 | 0.992 | 1.38e-04 | NA | 1622 | 1277 | 345 |
| Latino/Mixed-American | No Recorded Depression | Switching vs Continuation | Migraine | sex_at_birthMale | 1.249 | 0.954 | 1.637 | 1.06e-01 | NA | 1622 | 1277 | 345 |
| Latino/Mixed-American | No Recorded Depression | Switching vs Continuation | Migraine | Income_quantitative | 1.000 | 0.936 | 1.068 | 9.97e-01 | NA | 1622 | 1277 | 345 |
| Latino/Mixed-American | No Recorded Depression | Switching vs Continuation | Migraine | Education_levelCollege Undergraduate | 1.375 | 0.918 | 2.059 | 1.23e-01 | NA | 1622 | 1277 | 345 |
| Latino/Mixed-American | No Recorded Depression | Switching vs Continuation | Migraine | Education_levelLess than a high school degree | 1.200 | 0.706 | 2.039 | 5.01e-01 | NA | 1622 | 1277 | 345 |
| Latino/Mixed-American | No Recorded Depression | Switching vs Continuation | Migraine | Education_levelTwelve or GED | 1.343 | 0.815 | 2.213 | 2.47e-01 | NA | 1622 | 1277 | 345 |
| Latino/Mixed-American | No Recorded Depression | Switching vs Continuation | Migraine | Migraine | 0.971 | 0.856 | 1.103 | 6.53e-01 | 1.00e+00 | 1622 | 1277 | 345 |
| Latino/Mixed-American | No Recorded Depression | Discontinuation vs Continuation | Schizophrenia | (Intercept) | 0.546 | 0.346 | 0.863 | 9.61e-03 | NA | 2057 | 1277 | 780 |
| Latino/Mixed-American | No Recorded Depression | Discontinuation vs Continuation | Schizophrenia | age_at_first_AD | 1.000 | 0.994 | 1.006 | 9.87e-01 | NA | 2057 | 1277 | 780 |
| Latino/Mixed-American | No Recorded Depression | Discontinuation vs Continuation | Schizophrenia | sex_at_birthMale | 1.309 | 1.073 | 1.597 | 7.93e-03 | NA | 2057 | 1277 | 780 |
| Latino/Mixed-American | No Recorded Depression | Discontinuation vs Continuation | Schizophrenia | Income_quantitative | 0.925 | 0.879 | 0.973 | 2.49e-03 | NA | 2057 | 1277 | 780 |
| Latino/Mixed-American | No Recorded Depression | Discontinuation vs Continuation | Schizophrenia | Education_levelCollege Undergraduate | 1.400 | 1.021 | 1.919 | 3.65e-02 | NA | 2057 | 1277 | 780 |
| Latino/Mixed-American | No Recorded Depression | Discontinuation vs Continuation | Schizophrenia | Education_levelLess than a high school degree | 0.986 | 0.661 | 1.472 | 9.47e-01 | NA | 2057 | 1277 | 780 |
| Latino/Mixed-American | No Recorded Depression | Discontinuation vs Continuation | Schizophrenia | Education_levelTwelve or GED | 1.591 | 1.095 | 2.312 | 1.49e-02 | NA | 2057 | 1277 | 780 |
| Latino/Mixed-American | No Recorded Depression | Discontinuation vs Continuation | Schizophrenia | Schizophrenia | 0.992 | 0.905 | 1.087 | 8.62e-01 | 1.00e+00 | 2057 | 1277 | 780 |
| Latino/Mixed-American | No Recorded Depression | Switching vs Continuation | Schizophrenia | (Intercept) | 0.402 | 0.221 | 0.731 | 2.80e-03 | NA | 1622 | 1277 | 345 |
| Latino/Mixed-American | No Recorded Depression | Switching vs Continuation | Schizophrenia | age_at_first_AD | 0.984 | 0.975 | 0.992 | 1.37e-04 | NA | 1622 | 1277 | 345 |
| Latino/Mixed-American | No Recorded Depression | Switching vs Continuation | Schizophrenia | sex_at_birthMale | 1.251 | 0.955 | 1.639 | 1.04e-01 | NA | 1622 | 1277 | 345 |
| Latino/Mixed-American | No Recorded Depression | Switching vs Continuation | Schizophrenia | Income_quantitative | 1.003 | 0.940 | 1.071 | 9.25e-01 | NA | 1622 | 1277 | 345 |
| Latino/Mixed-American | No Recorded Depression | Switching vs Continuation | Schizophrenia | Education_levelCollege Undergraduate | 1.385 | 0.925 | 2.075 | 1.14e-01 | NA | 1622 | 1277 | 345 |
| Latino/Mixed-American | No Recorded Depression | Switching vs Continuation | Schizophrenia | Education_levelLess than a high school degree | 1.207 | 0.711 | 2.051 | 4.86e-01 | NA | 1622 | 1277 | 345 |
| Latino/Mixed-American | No Recorded Depression | Switching vs Continuation | Schizophrenia | Education_levelTwelve or GED | 1.356 | 0.823 | 2.235 | 2.32e-01 | NA | 1622 | 1277 | 345 |
| Latino/Mixed-American | No Recorded Depression | Switching vs Continuation | Schizophrenia | Schizophrenia | 1.084 | 0.962 | 1.222 | 1.87e-01 | 1.00e+00 | 1622 | 1277 | 345 |
| Latino/Mixed-American | Recorded Anxiety | Discontinuation vs Continuation | ADHD | (Intercept) | 0.464 | 0.287 | 0.752 | 1.81e-03 | NA | 2246 | 1462 | 784 |
| Latino/Mixed-American | Recorded Anxiety | Discontinuation vs Continuation | ADHD | age_at_first_AD | 1.002 | 0.996 | 1.008 | 5.54e-01 | NA | 2246 | 1462 | 784 |
| Latino/Mixed-American | Recorded Anxiety | Discontinuation vs Continuation | ADHD | sex_at_birthMale | 1.293 | 1.065 | 1.568 | 9.27e-03 | NA | 2246 | 1462 | 784 |
| Latino/Mixed-American | Recorded Anxiety | Discontinuation vs Continuation | ADHD | Income_quantitative | 0.931 | 0.883 | 0.982 | 8.26e-03 | NA | 2246 | 1462 | 784 |
| Latino/Mixed-American | Recorded Anxiety | Discontinuation vs Continuation | ADHD | Education_levelCollege Undergraduate | 1.302 | 0.921 | 1.841 | 1.35e-01 | NA | 2246 | 1462 | 784 |
| Latino/Mixed-American | Recorded Anxiety | Discontinuation vs Continuation | ADHD | Education_levelLess than a high school degree | 1.030 | 0.679 | 1.561 | 8.91e-01 | NA | 2246 | 1462 | 784 |
| Latino/Mixed-American | Recorded Anxiety | Discontinuation vs Continuation | ADHD | Education_levelTwelve or GED | 1.179 | 0.799 | 1.738 | 4.07e-01 | NA | 2246 | 1462 | 784 |
| Latino/Mixed-American | Recorded Anxiety | Discontinuation vs Continuation | ADHD | ADHD | 1.032 | 0.935 | 1.138 | 5.33e-01 | 1.00e+00 | 2246 | 1462 | 784 |
| Latino/Mixed-American | Recorded Anxiety | Switching vs Continuation | ADHD | (Intercept) | 1.306 | 0.876 | 1.948 | 1.90e-01 | NA | 2697 | 1462 | 1235 |
| Latino/Mixed-American | Recorded Anxiety | Switching vs Continuation | ADHD | age_at_first_AD | 0.993 | 0.987 | 0.998 | 1.13e-02 | NA | 2697 | 1462 | 1235 |
| Latino/Mixed-American | Recorded Anxiety | Switching vs Continuation | ADHD | sex_at_birthMale | 1.095 | 0.919 | 1.305 | 3.10e-01 | NA | 2697 | 1462 | 1235 |
| Latino/Mixed-American | Recorded Anxiety | Switching vs Continuation | ADHD | Income_quantitative | 0.967 | 0.924 | 1.011 | 1.42e-01 | NA | 2697 | 1462 | 1235 |
| Latino/Mixed-American | Recorded Anxiety | Switching vs Continuation | ADHD | Education_levelCollege Undergraduate | 0.986 | 0.751 | 1.294 | 9.18e-01 | NA | 2697 | 1462 | 1235 |
| Latino/Mixed-American | Recorded Anxiety | Switching vs Continuation | ADHD | Education_levelLess than a high school degree | 0.952 | 0.679 | 1.335 | 7.76e-01 | NA | 2697 | 1462 | 1235 |

| | | | | | | | | | | | | |
|---|---|---|---|---|---|---|---|---|---|---|---|---|
| Latino/Mixed-American | Recorded Anxiety | Switching vs Continuation | ADHD | Education_levelTwelve or GED | 0.791 | 0.576 | 1.085 | 1.45e-01 | NA | 2697 | 1462 | 1235 |
| Latino/Mixed-American | Recorded Anxiety | Switching vs Continuation | ADHD | ADHD | 1.063 | 0.977 | 1.157 | 1.55e-01 | 1.00e+00 | 2697 | 1462 | 1235 |
| Latino/Mixed-American | Recorded Anxiety | Discontinuation vs Continuation | BMI | (Intercept) | 0.463 | 0.286 | 0.751 | 1.77e-03 | NA | 2246 | 1462 | 784 |
| Latino/Mixed-American | Recorded Anxiety | Discontinuation vs Continuation | BMI | age_at_first_AD | 1.002 | 0.996 | 1.008 | 5.90e-01 | NA | 2246 | 1462 | 784 |
| Latino/Mixed-American | Recorded Anxiety | Discontinuation vs Continuation | BMI | sex_at_birthMale | 1.292 | 1.065 | 1.568 | 9.42e-03 | NA | 2246 | 1462 | 784 |
| Latino/Mixed-American | Recorded Anxiety | Discontinuation vs Continuation | BMI | Income_quantitative | 0.934 | 0.886 | 0.985 | 1.21e-02 | NA | 2246 | 1462 | 784 |
| Latino/Mixed-American | Recorded Anxiety | Discontinuation vs Continuation | BMI | Education_levelCollege Undergraduate | 1.301 | 0.920 | 1.839 | 1.37e-01 | NA | 2246 | 1462 | 784 |
| Latino/Mixed-American | Recorded Anxiety | Discontinuation vs Continuation | BMI | Education_levelLess than a high school degree | 1.023 | 0.675 | 1.551 | 9.14e-01 | NA | 2246 | 1462 | 784 |
| Latino/Mixed-American | Recorded Anxiety | Discontinuation vs Continuation | BMI | Education_levelTwelve or GED | 1.179 | 0.799 | 1.738 | 4.07e-01 | NA | 2246 | 1462 | 784 |
| Latino/Mixed-American | Recorded Anxiety | Discontinuation vs Continuation | BMI | BMI | 1.110 | 1.014 | 1.215 | 2.36e-02 | 1.65e-01 | 2246 | 1462 | 784 |
| Latino/Mixed-American | Recorded Anxiety | Switching vs Continuation | BMI | (Intercept) | 1.309 | 0.878 | 1.951 | 1.87e-01 | NA | 2697 | 1462 | 1235 |
| Latino/Mixed-American | Recorded Anxiety | Switching vs Continuation | BMI | age_at_first_AD | 0.993 | 0.987 | 0.998 | 9.58e-03 | NA | 2697 | 1462 | 1235 |
| Latino/Mixed-American | Recorded Anxiety | Switching vs Continuation | BMI | sex_at_birthMale | 1.098 | 0.921 | 1.308 | 2.97e-01 | NA | 2697 | 1462 | 1235 |
| Latino/Mixed-American | Recorded Anxiety | Switching vs Continuation | BMI | Income_quantitative | 0.967 | 0.924 | 1.011 | 1.41e-01 | NA | 2697 | 1462 | 1235 |
| Latino/Mixed-American | Recorded Anxiety | Switching vs Continuation | BMI | Education_levelCollege Undergraduate | 0.989 | 0.753 | 1.298 | 9.34e-01 | NA | 2697 | 1462 | 1235 |
| Latino/Mixed-American | Recorded Anxiety | Switching vs Continuation | BMI | Education_levelLess than a high school degree | 0.965 | 0.688 | 1.352 | 8.35e-01 | NA | 2697 | 1462 | 1235 |
| Latino/Mixed-American | Recorded Anxiety | Switching vs Continuation | BMI | Education_levelTwelve or GED | 0.796 | 0.580 | 1.092 | 1.57e-01 | NA | 2697 | 1462 | 1235 |
| Latino/Mixed-American | Recorded Anxiety | Switching vs Continuation | BMI | BMI | 1.033 | 0.955 | 1.118 | 4.13e-01 | 1.00e+00 | 2697 | 1462 | 1235 |
| Latino/Mixed-American | Recorded Anxiety | Discontinuation vs Continuation | Bipolar Disorder | (Intercept) | 0.466 | 0.288 | 0.755 | 1.90e-03 | NA | 2246 | 1462 | 784 |
| Latino/Mixed-American | Recorded Anxiety | Discontinuation vs Continuation | Bipolar Disorder | age_at_first_AD | 1.002 | 0.996 | 1.008 | 5.78e-01 | NA | 2246 | 1462 | 784 |
| Latino/Mixed-American | Recorded Anxiety | Discontinuation vs Continuation | Bipolar Disorder | sex_at_birthMale | 1.293 | 1.066 | 1.569 | 9.18e-03 | NA | 2246 | 1462 | 784 |
| Latino/Mixed-American | Recorded Anxiety | Discontinuation vs Continuation | Bipolar Disorder | Income_quantitative | 0.930 | 0.882 | 0.981 | 7.40e-03 | NA | 2246 | 1462 | 784 |
| Latino/Mixed-American | Recorded Anxiety | Discontinuation vs Continuation | Bipolar Disorder | Education_levelCollege Undergraduate | 1.306 | 0.924 | 1.846 | 1.31e-01 | NA | 2246 | 1462 | 784 |
| Latino/Mixed-American | Recorded Anxiety | Discontinuation vs Continuation | Bipolar Disorder | Education_levelLess than a high school degree | 1.035 | 0.683 | 1.569 | 8.71e-01 | NA | 2246 | 1462 | 784 |
| Latino/Mixed-American | Recorded Anxiety | Discontinuation vs Continuation | Bipolar Disorder | Education_levelTwelve or GED | 1.184 | 0.803 | 1.745 | 3.94e-01 | NA | 2246 | 1462 | 784 |
| Latino/Mixed-American | Recorded Anxiety | Discontinuation vs Continuation | Bipolar Disorder | Bipolar Disorder | 1.014 | 0.919 | 1.118 | 7.88e-01 | 1.00e+00 | 2246 | 1462 | 784 |
| Latino/Mixed-American | Recorded Anxiety | Switching vs Continuation | Bipolar Disorder | (Intercept) | 1.302 | 0.873 | 1.941 | 1.96e-01 | NA | 2697 | 1462 | 1235 |
| Latino/Mixed-American | Recorded Anxiety | Switching vs Continuation | Bipolar Disorder | age_at_first_AD | 0.993 | 0.987 | 0.998 | 9.09e-03 | NA | 2697 | 1462 | 1235 |
| Latino/Mixed-American | Recorded Anxiety | Switching vs Continuation | Bipolar Disorder | sex_at_birthMale | 1.098 | 0.922 | 1.308 | 2.96e-01 | NA | 2697 | 1462 | 1235 |
| Latino/Mixed-American | Recorded Anxiety | Switching vs Continuation | Bipolar Disorder | Income_quantitative | 0.967 | 0.924 | 1.011 | 1.41e-01 | NA | 2697 | 1462 | 1235 |
| Latino/Mixed-American | Recorded Anxiety | Switching vs Continuation | Bipolar Disorder | Education_levelCollege Undergraduate | 0.995 | 0.758 | 1.307 | 9.74e-01 | NA | 2697 | 1462 | 1235 |
| Latino/Mixed-American | Recorded Anxiety | Switching vs Continuation | Bipolar Disorder | Education_levelLess than a high school degree | 0.972 | 0.693 | 1.362 | 8.69e-01 | NA | 2697 | 1462 | 1235 |
| Latino/Mixed-American | Recorded Anxiety | Switching vs Continuation | Bipolar Disorder | Education_levelTwelve or GED | 0.803 | 0.585 | 1.101 | 1.73e-01 | NA | 2697 | 1462 | 1235 |
| Latino/Mixed-American | Recorded Anxiety | Switching vs Continuation | Bipolar Disorder | Bipolar Disorder | 1.048 | 0.962 | 1.141 | 2.86e-01 | 1.00e+00 | 2697 | 1462 | 1235 |
| Latino/Mixed-American | Recorded Anxiety | Discontinuation vs Continuation | Depression | (Intercept) | 0.460 | 0.284 | 0.746 | 1.62e-03 | NA | 2246 | 1462 | 784 |
| Latino/Mixed-American | Recorded Anxiety | Discontinuation vs Continuation | Depression | age_at_first_AD | 1.002 | 0.996 | 1.008 | 5.17e-01 | NA | 2246 | 1462 | 784 |
| Latino/Mixed-American | Recorded Anxiety | Discontinuation vs Continuation | Depression | sex_at_birthMale | 1.293 | 1.066 | 1.570 | 9.21e-03 | NA | 2246 | 1462 | 784 |
| Latino/Mixed-American | Recorded Anxiety | Discontinuation vs Continuation | Depression | Income_quantitative | 0.933 | 0.885 | 0.984 | 1.04e-02 | NA | 2246 | 1462 | 784 |
| Latino/Mixed-American | Recorded Anxiety | Discontinuation vs Continuation | Depression | Education_levelCollege Undergraduate | 1.298 | 0.918 | 1.835 | 1.41e-01 | NA | 2246 | 1462 | 784 |
| Latino/Mixed-American | Recorded Anxiety | Discontinuation vs Continuation | Depression | Education_levelLess than a high school degree | 1.025 | 0.676 | 1.554 | 9.09e-01 | NA | 2246 | 1462 | 784 |
| Latino/Mixed-American | Recorded Anxiety | Discontinuation vs Continuation | Depression | Education_levelTwelve or GED | 1.173 | 0.796 | 1.731 | 4.20e-01 | NA | 2246 | 1462 | 784 |
| Latino/Mixed-American | Recorded Anxiety | Discontinuation vs Continuation | Depression | Depression | 1.158 | 1.048 | 1.280 | 3.93e-03 | 2.75e-02 | 2246 | 1462 | 784 |
| Latino/Mixed-American | Recorded Anxiety | Switching vs Continuation | Depression | (Intercept) | 1.301 | 0.872 | 1.941 | 1.98e-01 | NA | 2697 | 1462 | 1235 |
| Latino/Mixed-American | Recorded Anxiety | Switching vs Continuation | Depression | age_at_first_AD | 0.993 | 0.987 | 0.998 | 9.31e-03 | NA | 2697 | 1462 | 1235 |
| Latino/Mixed-American | Recorded Anxiety | Switching vs Continuation | Depression | sex_at_birthMale | 1.102 | 0.925 | 1.313 | 2.78e-01 | NA | 2697 | 1462 | 1235 |
| Latino/Mixed-American | Recorded Anxiety | Switching vs Continuation | Depression | Income_quantitative | 0.969 | 0.925 | 1.014 | 1.67e-01 | NA | 2697 | 1462 | 1235 |
| Latino/Mixed-American | Recorded Anxiety | Switching vs Continuation | Depression | Education_levelCollege Undergraduate | 0.993 | 0.756 | 1.304 | 9.59e-01 | NA | 2697 | 1462 | 1235 |
| Latino/Mixed-American | Recorded Anxiety | Switching vs Continuation | Depression | Education_levelLess than a high school degree | 0.962 | 0.686 | 1.349 | 8.22e-01 | NA | 2697 | 1462 | 1235 |
| Latino/Mixed-American | Recorded Anxiety | Switching vs Continuation | Depression | Education_levelTwelve or GED | 0.798 | 0.582 | 1.095 | 1.62e-01 | NA | 2697 | 1462 | 1235 |
| Latino/Mixed-American | Recorded Anxiety | Switching vs Continuation | Depression | Depression | 1.141 | 1.047 | 1.244 | 2.74e-03 | 1.92e-02 | 2697 | 1462 | 1235 |
| Latino/Mixed-American | Recorded Anxiety | Discontinuation vs Continuation | Insomnia | (Intercept) | 0.466 | 0.288 | 0.755 | 1.90e-03 | NA | 2246 | 1462 | 784 |
| Latino/Mixed-American | Recorded Anxiety | Discontinuation vs Continuation | Insomnia | age_at_first_AD | 1.002 | 0.996 | 1.008 | 5.74e-01 | NA | 2246 | 1462 | 784 |
| Latino/Mixed-American | Recorded Anxiety | Discontinuation vs Continuation | Insomnia | sex_at_birthMale | 1.292 | 1.065 | 1.568 | 9.34e-03 | NA | 2246 | 1462 | 784 |
| Latino/Mixed-American | Recorded Anxiety | Discontinuation vs Continuation | Insomnia | Income_quantitative | 0.930 | 0.882 | 0.981 | 7.27e-03 | NA | 2246 | 1462 | 784 |
| Latino/Mixed-American | Recorded Anxiety | Discontinuation vs Continuation | Insomnia | Education_levelCollege Undergraduate | 1.306 | 0.924 | 1.846 | 1.31e-01 | NA | 2246 | 1462 | 784 |
| Latino/Mixed-American | Recorded Anxiety | Discontinuation vs Continuation | Insomnia | Education_levelLess than a high school degree | 1.034 | 0.682 | 1.567 | 8.75e-01 | NA | 2246 | 1462 | 784 |
| Latino/Mixed-American | Recorded Anxiety | Discontinuation vs Continuation | Insomnia | Education_levelTwelve or GED | 1.184 | 0.803 | 1.745 | 3.95e-01 | NA | 2246 | 1462 | 784 |

| | | | | | | | | | | | | |
|---|---|---|---|---|---|---|---|---|---|---|---|---|
| Latino/Mixed-American | Recorded Anxiety | Discontinuation vs Continuation | Insomnia | Insomnia | 1.004 | 0.908 | 1.110 | 9.42e-01 | 1.00e+00 | 2246 | 1462 | 784 |
| Latino/Mixed-American | Recorded Anxiety | Switching vs Continuation | Insomnia | (Intercept) | 1.310 | 0.879 | 1.954 | 1.85e-01 | NA | 2697 | 1462 | 1235 |
| Latino/Mixed-American | Recorded Anxiety | Switching vs Continuation | Insomnia | age_at_first_AD | 0.993 | 0.987 | 0.998 | 1.01e-02 | NA | 2697 | 1462 | 1235 |
| Latino/Mixed-American | Recorded Anxiety | Switching vs Continuation | Insomnia | sex_at_birthMale | 1.102 | 0.925 | 1.313 | 2.76e-01 | NA | 2697 | 1462 | 1235 |
| Latino/Mixed-American | Recorded Anxiety | Switching vs Continuation | Insomnia | Income_quantitative | 0.966 | 0.923 | 1.011 | 1.34e-01 | NA | 2697 | 1462 | 1235 |
| Latino/Mixed-American | Recorded Anxiety | Switching vs Continuation | Insomnia | Education_levelCollege Undergraduate | 0.985 | 0.750 | 1.293 | 9.10e-01 | NA | 2697 | 1462 | 1235 |
| Latino/Mixed-American | Recorded Anxiety | Switching vs Continuation | Insomnia | Education_levelLess than a high school degree | 0.959 | 0.684 | 1.344 | 8.06e-01 | NA | 2697 | 1462 | 1235 |
| Latino/Mixed-American | Recorded Anxiety | Switching vs Continuation | Insomnia | Education_levelTwelve or GED | 0.793 | 0.578 | 1.088 | 1.50e-01 | NA | 2697 | 1462 | 1235 |
| Latino/Mixed-American | Recorded Anxiety | Switching vs Continuation | Insomnia | Insomnia | 1.076 | 0.986 | 1.174 | 9.98e-02 | 6.99e-01 | 2697 | 1462 | 1235 |
| Latino/Mixed-American | Recorded Anxiety | Discontinuation vs Continuation | Migraine | (Intercept) | 0.465 | 0.287 | 0.753 | 1.83e-03 | NA | 2246 | 1462 | 784 |
| Latino/Mixed-American | Recorded Anxiety | Discontinuation vs Continuation | Migraine | age_at_first_AD | 1.002 | 0.996 | 1.008 | 5.72e-01 | NA | 2246 | 1462 | 784 |
| Latino/Mixed-American | Recorded Anxiety | Discontinuation vs Continuation | Migraine | sex_at_birthMale | 1.291 | 1.064 | 1.567 | 9.54e-03 | NA | 2246 | 1462 | 784 |
| Latino/Mixed-American | Recorded Anxiety | Discontinuation vs Continuation | Migraine | Income_quantitative | 0.930 | 0.882 | 0.981 | 7.23e-03 | NA | 2246 | 1462 | 784 |
| Latino/Mixed-American | Recorded Anxiety | Discontinuation vs Continuation | Migraine | Education_levelCollege Undergraduate | 1.308 | 0.926 | 1.849 | 1.28e-01 | NA | 2246 | 1462 | 784 |
| Latino/Mixed-American | Recorded Anxiety | Discontinuation vs Continuation | Migraine | Education_levelLess than a high school degree | 1.037 | 0.684 | 1.572 | 8.63e-01 | NA | 2246 | 1462 | 784 |
| Latino/Mixed-American | Recorded Anxiety | Discontinuation vs Continuation | Migraine | Education_levelTwelve or GED | 1.187 | 0.805 | 1.750 | 3.87e-01 | NA | 2246 | 1462 | 784 |
| Latino/Mixed-American | Recorded Anxiety | Discontinuation vs Continuation | Migraine | Migraine | 0.963 | 0.878 | 1.057 | 4.32e-01 | 1.00e+00 | 2246 | 1462 | 784 |
| Latino/Mixed-American | Recorded Anxiety | Switching vs Continuation | Migraine | (Intercept) | 1.309 | 0.878 | 1.952 | 1.86e-01 | NA | 2697 | 1462 | 1235 |
| Latino/Mixed-American | Recorded Anxiety | Switching vs Continuation | Migraine | age_at_first_AD | 0.993 | 0.987 | 0.998 | 9.75e-03 | NA | 2697 | 1462 | 1235 |
| Latino/Mixed-American | Recorded Anxiety | Switching vs Continuation | Migraine | sex_at_birthMale | 1.097 | 0.921 | 1.307 | 3.00e-01 | NA | 2697 | 1462 | 1235 |
| Latino/Mixed-American | Recorded Anxiety | Switching vs Continuation | Migraine | Income_quantitative | 0.965 | 0.922 | 1.010 | 1.24e-01 | NA | 2697 | 1462 | 1235 |
| Latino/Mixed-American | Recorded Anxiety | Switching vs Continuation | Migraine | Education_levelCollege Undergraduate | 0.992 | 0.756 | 1.302 | 9.54e-01 | NA | 2697 | 1462 | 1235 |
| Latino/Mixed-American | Recorded Anxiety | Switching vs Continuation | Migraine | Education_levelLess than a high school degree | 0.964 | 0.688 | 1.352 | 8.33e-01 | NA | 2697 | 1462 | 1235 |
| Latino/Mixed-American | Recorded Anxiety | Switching vs Continuation | Migraine | Education_levelTwelve or GED | 0.797 | 0.581 | 1.094 | 1.60e-01 | NA | 2697 | 1462 | 1235 |
| Latino/Mixed-American | Recorded Anxiety | Switching vs Continuation | Migraine | Migraine | 1.021 | 0.944 | 1.105 | 6.02e-01 | 1.00e+00 | 2697 | 1462 | 1235 |
| Latino/Mixed-American | Recorded Anxiety | Discontinuation vs Continuation | Schizophrenia | (Intercept) | 0.464 | 0.287 | 0.751 | 1.79e-03 | NA | 2246 | 1462 | 784 |
| Latino/Mixed-American | Recorded Anxiety | Discontinuation vs Continuation | Schizophrenia | age_at_first_AD | 1.002 | 0.996 | 1.008 | 6.15e-01 | NA | 2246 | 1462 | 784 |
| Latino/Mixed-American | Recorded Anxiety | Discontinuation vs Continuation | Schizophrenia | sex_at_birthMale | 1.294 | 1.066 | 1.570 | 9.07e-03 | NA | 2246 | 1462 | 784 |
| Latino/Mixed-American | Recorded Anxiety | Discontinuation vs Continuation | Schizophrenia | Income_quantitative | 0.931 | 0.883 | 0.982 | 8.69e-03 | NA | 2246 | 1462 | 784 |
| Latino/Mixed-American | Recorded Anxiety | Discontinuation vs Continuation | Schizophrenia | Education_levelCollege Undergraduate | 1.319 | 0.933 | 1.865 | 1.18e-01 | NA | 2246 | 1462 | 784 |
| Latino/Mixed-American | Recorded Anxiety | Discontinuation vs Continuation | Schizophrenia | Education_levelLess than a high school degree | 1.045 | 0.689 | 1.584 | 8.36e-01 | NA | 2246 | 1462 | 784 |
| Latino/Mixed-American | Recorded Anxiety | Discontinuation vs Continuation | Schizophrenia | Education_levelTwelve or GED | 1.190 | 0.807 | 1.755 | 3.79e-01 | NA | 2246 | 1462 | 784 |
| Latino/Mixed-American | Recorded Anxiety | Discontinuation vs Continuation | Schizophrenia | Schizophrenia | 1.071 | 0.980 | 1.170 | 1.31e-01 | 9.20e-01 | 2246 | 1462 | 784 |
| Latino/Mixed-American | Recorded Anxiety | Switching vs Continuation | Schizophrenia | (Intercept) | 1.294 | 0.868 | 1.931 | 2.06e-01 | NA | 2697 | 1462 | 1235 |
| Latino/Mixed-American | Recorded Anxiety | Switching vs Continuation | Schizophrenia | age_at_first_AD | 0.993 | 0.987 | 0.998 | 9.02e-03 | NA | 2697 | 1462 | 1235 |
| Latino/Mixed-American | Recorded Anxiety | Switching vs Continuation | Schizophrenia | sex_at_birthMale | 1.100 | 0.924 | 1.311 | 2.85e-01 | NA | 2697 | 1462 | 1235 |
| Latino/Mixed-American | Recorded Anxiety | Switching vs Continuation | Schizophrenia | Income_quantitative | 0.967 | 0.924 | 1.012 | 1.44e-01 | NA | 2697 | 1462 | 1235 |
| Latino/Mixed-American | Recorded Anxiety | Switching vs Continuation | Schizophrenia | Education_levelCollege Undergraduate | 1.003 | 0.764 | 1.317 | 9.85e-01 | NA | 2697 | 1462 | 1235 |
| Latino/Mixed-American | Recorded Anxiety | Switching vs Continuation | Schizophrenia | Education_levelLess than a high school degree | 0.973 | 0.694 | 1.364 | 8.75e-01 | NA | 2697 | 1462 | 1235 |
| Latino/Mixed-American | Recorded Anxiety | Switching vs Continuation | Schizophrenia | Education_levelTwelve or GED | 0.808 | 0.589 | 1.109 | 1.87e-01 | NA | 2697 | 1462 | 1235 |
| Latino/Mixed-American | Recorded Anxiety | Switching vs Continuation | Schizophrenia | Schizophrenia | 1.064 | 0.985 | 1.148 | 1.15e-01 | 8.02e-01 | 2697 | 1462 | 1235 |

**Supplementary Table 5. Multivariable logistic regression results for PGS associations with antidepressant treatment outcomes across clinical subgroups in the Pharmlines cohort (European ancestry participants).** Each model estimates the association between 7 PGS traits and one of two binary outcomes — discontinuation versus continuation, or switching versus continuation — fitted separately within each combination of clinical group (Full Cohort; and subgroups defined by recorded depression status), and PGS exposure. Within each model, one PGS exposure is modelled alongside a common covariate set (age at first antidepressant, genetic sex, income, and educational attainment). All PGS were residualised on the top 20 within-European-ancestry principal components and the residuals standardised to mean 0, SD 1; each odds ratio therefore reflects the change in odds per one-standard-deviation increase in the PC-adjusted PGS. Because PCs were regressed out at this stage, they are not included in the model covariate set.
Columns: Population = inferred genetic ancestry (all European); ClinicalGroup = cohort or clinical subgroup; Outcome = the binary contrast modelled; PGS = the PGS trait for that model; variable = the individual model term; odds_ratio = exponentiated coefficient; lower_95/upper_95 = 95% confidence interval bounds for the odds ratio; p_value = nominal (uncorrected) p-value; Bonf_P = Bonferroni-corrected p-value (capped at 1.00); N_Total = model sample size; N_Reference = number in the reference (continuation) group; N_Case = number in the comparison (discontinuation or switching) group. Odds ratios >1 indicate higher odds of the comparison outcome relative to continuation. Bonferroni correction was applied within each specification (ancestry × clinical group × outcome contrast) for the max 7 PGS traits tested. P-values reported as "<2.2e-16" reflect R's floating-point reporting limit.
Reference (omitted) categories: Sex at birth: Female.

| Population | ClinicalGroup | Outcome | PGS | variable | odds_ratio | lower_95 | upper_95 | p_value | Bonf_p | N_Total | N_Reference | N_Case |
|---|---|---|---|---|---|---|---|---|---|---|---|---|
| European | Full Cohort | Discontinuation vs Continuation | ADHD | (Intercept) | 0.241 | 0.171 | 0.339 | 3.25e-16 | 2.27e-15 | 5629 | 4208 | 1421 |
| European | Full Cohort | Discontinuation vs Continuation | ADHD | age_at_first_AD | 1.015 | 1.010 | 1.020 | 1.53e-09 | 1.07e-08 | 5629 | 4208 | 1421 |
| European | Full Cohort | Discontinuation vs Continuation | ADHD | sex_at_birth | 1.299 | 1.140 | 1.479 | 8.35e-05 | 5.85e-04 | 5629 | 4208 | 1421 |
| European | Full Cohort | Discontinuation vs Continuation | ADHD | Income_level | 1.000 | 1.000 | 1.000 | 2.98e-01 | 1.00e+00 | 5629 | 4208 | 1421 |
| European | Full Cohort | Discontinuation vs Continuation | ADHD | Education_level | 0.963 | 0.947 | 0.978 | 2.91e-06 | 2.04e-05 | 5629 | 4208 | 1421 |
| European | Full Cohort | Discontinuation vs Continuation | ADHD | ADHD | 1.082 | 1.018 | 1.151 | 1.16e-02 | 8.11e-02 | 5629 | 4208 | 1421 |
| European | Full Cohort | Switching vs Continuation | ADHD | (Intercept) | 0.842 | 0.594 | 1.194 | 3.35e-01 | 1.00e+00 | 5493 | 4208 | 1285 |
| European | Full Cohort | Switching vs Continuation | ADHD | age_at_first_AD | 0.985 | 0.980 | 0.990 | 1.08e-08 | 7.55e-08 | 5493 | 4208 | 1285 |
| European | Full Cohort | Switching vs Continuation | ADHD | sex_at_birth | 1.253 | 1.092 | 1.438 | 1.28e-03 | 8.93e-03 | 5493 | 4208 | 1285 |
| European | Full Cohort | Switching vs Continuation | ADHD | Income_level | 1.000 | 1.000 | 1.000 | 1.24e-02 | 8.66e-02 | 5493 | 4208 | 1285 |
| European | Full Cohort | Switching vs Continuation | ADHD | Education_level | 0.988 | 0.972 | 1.004 | 1.36e-01 | 9.53e-01 | 5493 | 4208 | 1285 |
| European | Full Cohort | Switching vs Continuation | ADHD | ADHD | 1.076 | 1.010 | 1.147 | 2.42e-02 | 1.69e-01 | 5493 | 4208 | 1285 |
| European | Full Cohort | Discontinuation vs Continuation | BMI | (Intercept) | 0.243 | 0.173 | 0.342 | 4.96e-16 | 3.47e-15 | 5629 | 4208 | 1421 |
| European | Full Cohort | Discontinuation vs Continuation | BMI | age_at_first_AD | 1.015 | 1.010 | 1.020 | 1.91e-09 | 1.34e-08 | 5629 | 4208 | 1421 |
| European | Full Cohort | Discontinuation vs Continuation | BMI | sex_at_birth | 1.303 | 1.144 | 1.485 | 6.68e-05 | 4.67e-04 | 5629 | 4208 | 1421 |
| European | Full Cohort | Discontinuation vs Continuation | BMI | Income_level | 1.000 | 1.000 | 1.000 | 3.02e-01 | 1.00e+00 | 5629 | 4208 | 1421 |
| European | Full Cohort | Discontinuation vs Continuation | BMI | Education_level | 0.962 | 0.947 | 0.978 | 2.29e-06 | 1.60e-05 | 5629 | 4208 | 1421 |
| European | Full Cohort | Discontinuation vs Continuation | BMI | BMI | 1.066 | 1.004 | 1.132 | 3.73e-02 | 2.61e-01 | 5629 | 4208 | 1421 |
| European | Full Cohort | Switching vs Continuation | BMI | (Intercept) | 0.869 | 0.613 | 1.232 | 4.31e-01 | 1.00e+00 | 5493 | 4208 | 1285 |
| European | Full Cohort | Switching vs Continuation | BMI | age_at_first_AD | 0.985 | 0.979 | 0.990 | 7.64e-09 | 5.35e-08 | 5493 | 4208 | 1285 |
| European | Full Cohort | Switching vs Continuation | BMI | sex_at_birth | 1.255 | 1.094 | 1.439 | 1.20e-03 | 8.37e-03 | 5493 | 4208 | 1285 |
| European | Full Cohort | Switching vs Continuation | BMI | Income_level | 1.000 | 1.000 | 1.000 | 9.98e-03 | 6.99e-02 | 5493 | 4208 | 1285 |
| European | Full Cohort | Switching vs Continuation | BMI | Education_level | 0.986 | 0.970 | 1.003 | 1.01e-01 | 7.04e-01 | 5493 | 4208 | 1285 |
| European | Full Cohort | Switching vs Continuation | BMI | BMI | 1.010 | 0.949 | 1.075 | 7.57e-01 | 1.00e+00 | 5493 | 4208 | 1285 |
| European | Full Cohort | Discontinuation vs Continuation | Bipolar Disorder | (Intercept) | 0.250 | 0.178 | 0.352 | 1.43e-15 | 1.00e-14 | 5629 | 4208 | 1421 |
| European | Full Cohort | Discontinuation vs Continuation | Bipolar Disorder | age_at_first_AD | 1.015 | 1.010 | 1.020 | 2.70e-09 | 1.89e-08 | 5629 | 4208 | 1421 |
| European | Full Cohort | Discontinuation vs Continuation | Bipolar Disorder | sex_at_birth | 1.300 | 1.141 | 1.480 | 7.96e-05 | 5.57e-04 | 5629 | 4208 | 1421 |
| European | Full Cohort | Discontinuation vs Continuation | Bipolar Disorder | Income_level | 1.000 | 1.000 | 1.000 | 3.16e-01 | 1.00e+00 | 5629 | 4208 | 1421 |
| European | Full Cohort | Discontinuation vs Continuation | Bipolar Disorder | Education_level | 0.961 | 0.946 | 0.976 | 9.98e-07 | 6.99e-06 | 5629 | 4208 | 1421 |
| European | Full Cohort | Discontinuation vs Continuation | Bipolar Disorder | Bipolar Disorder | 1.007 | 0.947 | 1.070 | 8.30e-01 | 1.00e+00 | 5629 | 4208 | 1421 |
| European | Full Cohort | Switching vs Continuation | Bipolar Disorder | (Intercept) | 0.868 | 0.613 | 1.230 | 4.27e-01 | 1.00e+00 | 5493 | 4208 | 1285 |
| European | Full Cohort | Switching vs Continuation | Bipolar Disorder | age_at_first_AD | 0.985 | 0.979 | 0.990 | 9.16e-09 | 6.41e-08 | 5493 | 4208 | 1285 |
| European | Full Cohort | Switching vs Continuation | Bipolar Disorder | sex_at_birth | 1.254 | 1.093 | 1.439 | 1.22e-03 | 8.53e-03 | 5493 | 4208 | 1285 |
| European | Full Cohort | Switching vs Continuation | Bipolar Disorder | Income_level | 1.000 | 1.000 | 1.000 | 9.95e-03 | 6.97e-02 | 5493 | 4208 | 1285 |
| European | Full Cohort | Switching vs Continuation | Bipolar Disorder | Education_level | 0.986 | 0.970 | 1.002 | 9.11e-02 | 6.38e-01 | 5493 | 4208 | 1285 |
| European | Full Cohort | Switching vs Continuation | Bipolar Disorder | Bipolar Disorder | 1.107 | 1.039 | 1.179 | 1.59e-03 | 1.11e-02 | 5493 | 4208 | 1285 |

| | | | | | | | | | | | | |
|---|---|---|---|---|---|---|---|---|---|---|---|---|
| European | Full Cohort | Discontinuation vs Continuation | Depression | (Intercept) | 0.253 | 0.180 | 0.356 | 2.88e-15 | 2.02e-14 | 5629 | 4208 | 1421 |
| European | Full Cohort | Discontinuation vs Continuation | Depression | age_at_first_AD | 1.014 | 1.010 | 1.019 | 4.31e-09 | 3.02e-08 | 5629 | 4208 | 1421 |
| European | Full Cohort | Discontinuation vs Continuation | Depression | sex_at_birth | 1.300 | 1.141 | 1.481 | 7.80e-05 | 5.46e-04 | 5629 | 4208 | 1421 |
| European | Full Cohort | Discontinuation vs Continuation | Depression | Income_level | 1.000 | 1.000 | 1.000 | 3.21e-01 | 1.00e+00 | 5629 | 4208 | 1421 |
| European | Full Cohort | Discontinuation vs Continuation | Depression | Education_level | 0.961 | 0.945 | 0.976 | 7.81e-07 | 5.47e-06 | 5629 | 4208 | 1421 |
| European | Full Cohort | Discontinuation vs Continuation | Depression | Depression | 0.965 | 0.908 | 1.025 | 2.43e-01 | 1.00e+00 | 5629 | 4208 | 1421 |
| European | Full Cohort | Switching vs Continuation | Depression | (Intercept) | 0.820 | 0.578 | 1.163 | 2.66e-01 | 1.00e+00 | 5493 | 4208 | 1285 |
| European | Full Cohort | Switching vs Continuation | Depression | age_at_first_AD | 0.985 | 0.980 | 0.991 | 3.88e-08 | 2.72e-07 | 5493 | 4208 | 1285 |
| European | Full Cohort | Switching vs Continuation | Depression | sex_at_birth | 1.247 | 1.087 | 1.431 | 1.64e-03 | 1.15e-02 | 5493 | 4208 | 1285 |
| European | Full Cohort | Switching vs Continuation | Depression | Income_level | 1.000 | 1.000 | 1.000 | 1.10e-02 | 7.71e-02 | 5493 | 4208 | 1285 |
| European | Full Cohort | Switching vs Continuation | Depression | Education_level | 0.988 | 0.971 | 1.004 | 1.35e-01 | 9.44e-01 | 5493 | 4208 | 1285 |
| European | Full Cohort | Switching vs Continuation | Depression | Depression | 1.163 | 1.092 | 1.239 | 2.82e-06 | 1.97e-05 | 5493 | 4208 | 1285 |
| European | Full Cohort | Discontinuation vs Continuation | Insomnia | (Intercept) | 0.250 | 0.177 | 0.351 | 1.38e-15 | 9.69e-15 | 5629 | 4208 | 1421 |
| European | Full Cohort | Discontinuation vs Continuation | Insomnia | age_at_first_AD | 1.015 | 1.010 | 1.020 | 2.61e-09 | 1.83e-08 | 5629 | 4208 | 1421 |
| European | Full Cohort | Discontinuation vs Continuation | Insomnia | sex_at_birth | 1.299 | 1.140 | 1.479 | 8.21e-05 | 5.74e-04 | 5629 | 4208 | 1421 |
| European | Full Cohort | Discontinuation vs Continuation | Insomnia | Income_level | 1.000 | 1.000 | 1.000 | 3.12e-01 | 1.00e+00 | 5629 | 4208 | 1421 |
| European | Full Cohort | Discontinuation vs Continuation | Insomnia | Education_level | 0.961 | 0.946 | 0.977 | 1.07e-06 | 7.47e-06 | 5629 | 4208 | 1421 |
| European | Full Cohort | Discontinuation vs Continuation | Insomnia | Insomnia | 1.011 | 0.951 | 1.075 | 7.32e-01 | 1.00e+00 | 5629 | 4208 | 1421 |
| European | Full Cohort | Switching vs Continuation | Insomnia | (Intercept) | 0.842 | 0.594 | 1.194 | 3.35e-01 | 1.00e+00 | 5493 | 4208 | 1285 |
| European | Full Cohort | Switching vs Continuation | Insomnia | age_at_first_AD | 0.985 | 0.980 | 0.990 | 1.31e-08 | 9.16e-08 | 5493 | 4208 | 1285 |
| European | Full Cohort | Switching vs Continuation | Insomnia | sex_at_birth | 1.253 | 1.092 | 1.437 | 1.30e-03 | 9.09e-03 | 5493 | 4208 | 1285 |
| European | Full Cohort | Switching vs Continuation | Insomnia | Income_level | 1.000 | 1.000 | 1.000 | 1.21e-02 | 8.45e-02 | 5493 | 4208 | 1285 |
| European | Full Cohort | Switching vs Continuation | Insomnia | Education_level | 0.987 | 0.971 | 1.004 | 1.25e-01 | 8.74e-01 | 5493 | 4208 | 1285 |
| European | Full Cohort | Switching vs Continuation | Insomnia | Insomnia | 1.095 | 1.027 | 1.166 | 5.16e-03 | 3.61e-02 | 5493 | 4208 | 1285 |
| European | Full Cohort | Discontinuation vs Continuation | Migraine | (Intercept) | 0.251 | 0.179 | 0.353 | 1.74e-15 | 1.22e-14 | 5629 | 4208 | 1421 |
| European | Full Cohort | Discontinuation vs Continuation | Migraine | age_at_first_AD | 1.015 | 1.010 | 1.019 | 2.75e-09 | 1.92e-08 | 5629 | 4208 | 1421 |
| European | Full Cohort | Discontinuation vs Continuation | Migraine | sex_at_birth | 1.300 | 1.141 | 1.480 | 7.87e-05 | 5.51e-04 | 5629 | 4208 | 1421 |
| European | Full Cohort | Discontinuation vs Continuation | Migraine | Income_level | 1.000 | 1.000 | 1.000 | 3.31e-01 | 1.00e+00 | 5629 | 4208 | 1421 |
| European | Full Cohort | Discontinuation vs Continuation | Migraine | Education_level | 0.961 | 0.946 | 0.976 | 9.44e-07 | 6.61e-06 | 5629 | 4208 | 1421 |
| European | Full Cohort | Discontinuation vs Continuation | Migraine | Migraine | 0.972 | 0.916 | 1.033 | 3.60e-01 | 1.00e+00 | 5629 | 4208 | 1421 |
| European | Full Cohort | Switching vs Continuation | Migraine | (Intercept) | 0.869 | 0.614 | 1.231 | 4.29e-01 | 1.00e+00 | 5493 | 4208 | 1285 |
| European | Full Cohort | Switching vs Continuation | Migraine | age_at_first_AD | 0.985 | 0.979 | 0.990 | 7.12e-09 | 4.99e-08 | 5493 | 4208 | 1285 |
| European | Full Cohort | Switching vs Continuation | Migraine | sex_at_birth | 1.253 | 1.092 | 1.437 | 1.30e-03 | 9.08e-03 | 5493 | 4208 | 1285 |
| European | Full Cohort | Switching vs Continuation | Migraine | Income_level | 1.000 | 1.000 | 1.000 | 1.03e-02 | 7.24e-02 | 5493 | 4208 | 1285 |
| European | Full Cohort | Switching vs Continuation | Migraine | Education_level | 0.986 | 0.970 | 1.003 | 9.95e-02 | 6.97e-01 | 5493 | 4208 | 1285 |
| European | Full Cohort | Switching vs Continuation | Migraine | Migraine | 1.032 | 0.970 | 1.099 | 3.13e-01 | 1.00e+00 | 5493 | 4208 | 1285 |
| European | Full Cohort | Discontinuation vs Continuation | Schizophrenia | (Intercept) | 0.250 | 0.178 | 0.352 | 1.47e-15 | 1.03e-14 | 5629 | 4208 | 1421 |
| European | Full Cohort | Discontinuation vs Continuation | Schizophrenia | age_at_first_AD | 1.015 | 1.010 | 1.019 | 2.85e-09 | 1.99e-08 | 5629 | 4208 | 1421 |
| European | Full Cohort | Discontinuation vs Continuation | Schizophrenia | sex_at_birth | 1.299 | 1.141 | 1.480 | 8.09e-05 | 5.67e-04 | 5629 | 4208 | 1421 |
| European | Full Cohort | Discontinuation vs Continuation | Schizophrenia | Income_level | 1.000 | 1.000 | 1.000 | 3.16e-01 | 1.00e+00 | 5629 | 4208 | 1421 |
| European | Full Cohort | Discontinuation vs Continuation | Schizophrenia | Education_level | 0.961 | 0.946 | 0.976 | 9.93e-07 | 6.95e-06 | 5629 | 4208 | 1421 |
| European | Full Cohort | Discontinuation vs Continuation | Schizophrenia | Schizophrenia | 0.999 | 0.941 | 1.062 | 9.83e-01 | 1.00e+00 | 5629 | 4208 | 1421 |
| European | Full Cohort | Switching vs Continuation | Schizophrenia | (Intercept) | 0.863 | 0.609 | 1.223 | 4.07e-01 | 1.00e+00 | 5493 | 4208 | 1285 |
| European | Full Cohort | Switching vs Continuation | Schizophrenia | age_at_first_AD | 0.985 | 0.980 | 0.990 | 1.26e-08 | 8.81e-08 | 5493 | 4208 | 1285 |
| European | Full Cohort | Switching vs Continuation | Schizophrenia | sex_at_birth | 1.252 | 1.092 | 1.436 | 1.33e-03 | 9.29e-03 | 5493 | 4208 | 1285 |
| European | Full Cohort | Switching vs Continuation | Schizophrenia | Income_level | 1.000 | 1.000 | 1.000 | 9.93e-03 | 6.95e-02 | 5493 | 4208 | 1285 |
| European | Full Cohort | Switching vs Continuation | Schizophrenia | Education_level | 0.986 | 0.970 | 1.002 | 9.41e-02 | 6.59e-01 | 5493 | 4208 | 1285 |
| European | Full Cohort | Switching vs Continuation | Schizophrenia | Schizophrenia | 1.068 | 1.004 | 1.137 | 3.80e-02 | 2.66e-01 | 5493 | 4208 | 1285 |

| | | | | | | | | | | | | |
|---|---|---|---|---|---|---|---|---|---|---|---|---|
| European | Recorded Depression | Discontinuation vs Continuation | ADHD | (Intercept) | 0.233 | 0.121 | 0.448 | 1.27e-05 | 8.91e-05 | 1958 | 1596 | 362 |
| European | Recorded Depression | Discontinuation vs Continuation | ADHD | age_at_first_AD | 1.012 | 1.001 | 1.022 | 2.57e-02 | 1.80e-01 | 1958 | 1596 | 362 |
| European | Recorded Depression | Discontinuation vs Continuation | ADHD | sex_at_birth | 1.297 | 1.008 | 1.669 | 4.34e-02 | 3.03e-01 | 1958 | 1596 | 362 |
| European | Recorded Depression | Discontinuation vs Continuation | ADHD | Income_level | 1.000 | 1.000 | 1.000 | 8.39e-01 | 1.00e+00 | 1958 | 1596 | 362 |
| European | Recorded Depression | Discontinuation vs Continuation | ADHD | Education_level | 0.960 | 0.932 | 0.989 | 7.03e-03 | 4.92e-02 | 1958 | 1596 | 362 |
| European | Recorded Depression | Discontinuation vs Continuation | ADHD | ADHD | 1.101 | 0.979 | 1.238 | 1.09e-01 | 7.63e-01 | 1958 | 1596 | 362 |
| European | Recorded Depression | Switching vs Continuation | ADHD | (Intercept) | 1.137 | 0.679 | 1.904 | 6.25e-01 | 1.00e+00 | 2251 | 1596 | 655 |
| European | Recorded Depression | Switching vs Continuation | ADHD | age_at_first_AD | 0.990 | 0.982 | 0.998 | 2.02e-02 | 1.42e-01 | 2251 | 1596 | 655 |
| European | Recorded Depression | Switching vs Continuation | ADHD | sex_at_birth | 1.264 | 1.029 | 1.553 | 2.54e-02 | 1.78e-01 | 2251 | 1596 | 655 |
| European | Recorded Depression | Switching vs Continuation | ADHD | Income_level | 1.000 | 1.000 | 1.000 | 6.26e-02 | 4.38e-01 | 2251 | 1596 | 655 |
| European | Recorded Depression | Switching vs Continuation | ADHD | Education_level | 0.972 | 0.950 | 0.996 | 2.01e-02 | 1.41e-01 | 2251 | 1596 | 655 |
| European | Recorded Depression | Switching vs Continuation | ADHD | ADHD | 1.068 | 0.972 | 1.174 | 1.69e-01 | 1.00e+00 | 2251 | 1596 | 655 |
| European | Recorded Depression | Discontinuation vs Continuation | BMI | (Intercept) | 0.234 | 0.122 | 0.451 | 1.37e-05 | 9.60e-05 | 1958 | 1596 | 362 |
| European | Recorded Depression | Discontinuation vs Continuation | BMI | age_at_first_AD | 1.012 | 1.001 | 1.022 | 2.59e-02 | 1.82e-01 | 1958 | 1596 | 362 |
| European | Recorded Depression | Discontinuation vs Continuation | BMI | sex_at_birth | 1.319 | 1.025 | 1.698 | 3.14e-02 | 2.20e-01 | 1958 | 1596 | 362 |
| European | Recorded Depression | Discontinuation vs Continuation | BMI | Income_level | 1.000 | 1.000 | 1.000 | 8.44e-01 | 1.00e+00 | 1958 | 1596 | 362 |
| European | Recorded Depression | Discontinuation vs Continuation | BMI | Education_level | 0.959 | 0.931 | 0.988 | 5.97e-03 | 4.18e-02 | 1958 | 1596 | 362 |
| European | Recorded Depression | Discontinuation vs Continuation | BMI | BMI | 1.097 | 0.981 | 1.227 | 1.06e-01 | 7.40e-01 | 1958 | 1596 | 362 |
| European | Recorded Depression | Switching vs Continuation | BMI | (Intercept) | 1.163 | 0.696 | 1.945 | 5.65e-01 | 1.00e+00 | 2251 | 1596 | 655 |
| European | Recorded Depression | Switching vs Continuation | BMI | age_at_first_AD | 0.990 | 0.982 | 0.998 | 1.92e-02 | 1.34e-01 | 2251 | 1596 | 655 |
| European | Recorded Depression | Switching vs Continuation | BMI | sex_at_birth | 1.269 | 1.033 | 1.559 | 2.31e-02 | 1.62e-01 | 2251 | 1596 | 655 |
| European | Recorded Depression | Switching vs Continuation | BMI | Income_level | 1.000 | 1.000 | 1.000 | 5.66e-02 | 3.96e-01 | 2251 | 1596 | 655 |
| European | Recorded Depression | Switching vs Continuation | BMI | Education_level | 0.971 | 0.949 | 0.995 | 1.62e-02 | 1.14e-01 | 2251 | 1596 | 655 |
| European | Recorded Depression | Switching vs Continuation | BMI | BMI | 1.026 | 0.937 | 1.123 | 5.85e-01 | 1.00e+00 | 2251 | 1596 | 655 |
| European | Recorded Depression | Discontinuation vs Continuation | Bipolar Disorder | (Intercept) | 0.243 | 0.127 | 0.466 | 2.05e-05 | 1.44e-04 | 1958 | 1596 | 362 |
| European | Recorded Depression | Discontinuation vs Continuation | Bipolar Disorder | age_at_first_AD | 1.012 | 1.001 | 1.022 | 2.82e-02 | 1.98e-01 | 1958 | 1596 | 362 |
| European | Recorded Depression | Discontinuation vs Continuation | Bipolar Disorder | sex_at_birth | 1.307 | 1.016 | 1.681 | 3.73e-02 | 2.61e-01 | 1958 | 1596 | 362 |
| European | Recorded Depression | Discontinuation vs Continuation | Bipolar Disorder | Income_level | 1.000 | 1.000 | 1.000 | 8.35e-01 | 1.00e+00 | 1958 | 1596 | 362 |
| European | Recorded Depression | Discontinuation vs Continuation | Bipolar Disorder | Education_level | 0.958 | 0.930 | 0.987 | 4.23e-03 | 2.96e-02 | 1958 | 1596 | 362 |
| European | Recorded Depression | Discontinuation vs Continuation | Bipolar Disorder | Bipolar Disorder | 1.020 | 0.908 | 1.146 | 7.37e-01 | 1.00e+00 | 1958 | 1596 | 362 |
| European | Recorded Depression | Switching vs Continuation | Bipolar Disorder | (Intercept) | 1.167 | 0.699 | 1.950 | 5.54e-01 | 1.00e+00 | 2251 | 1596 | 655 |
| European | Recorded Depression | Switching vs Continuation | Bipolar Disorder | age_at_first_AD | 0.990 | 0.982 | 0.998 | 1.78e-02 | 1.25e-01 | 2251 | 1596 | 655 |
| European | Recorded Depression | Switching vs Continuation | Bipolar Disorder | sex_at_birth | 1.259 | 1.025 | 1.547 | 2.84e-02 | 1.99e-01 | 2251 | 1596 | 655 |
| European | Recorded Depression | Switching vs Continuation | Bipolar Disorder | Income_level | 1.000 | 1.000 | 1.000 | 7.48e-02 | 5.24e-01 | 2251 | 1596 | 655 |
| European | Recorded Depression | Switching vs Continuation | Bipolar Disorder | Education_level | 0.970 | 0.948 | 0.993 | 1.22e-02 | 8.53e-02 | 2251 | 1596 | 655 |
| European | Recorded Depression | Switching vs Continuation | Bipolar Disorder | Bipolar Disorder | 1.114 | 1.014 | 1.223 | 2.39e-02 | 1.68e-01 | 2251 | 1596 | 655 |
| European | Recorded Depression | Discontinuation vs Continuation | Depression | (Intercept) | 0.252 | 0.132 | 0.484 | 3.32e-05 | 2.33e-04 | 1958 | 1596 | 362 |
| European | Recorded Depression | Discontinuation vs Continuation | Depression | age_at_first_AD | 1.011 | 1.001 | 1.022 | 3.56e-02 | 2.49e-01 | 1958 | 1596 | 362 |
| European | Recorded Depression | Discontinuation vs Continuation | Depression | sex_at_birth | 1.321 | 1.026 | 1.701 | 3.06e-02 | 2.14e-01 | 1958 | 1596 | 362 |
| European | Recorded Depression | Discontinuation vs Continuation | Depression | Income_level | 1.000 | 1.000 | 1.000 | 8.69e-01 | 1.00e+00 | 1958 | 1596 | 362 |
| European | Recorded Depression | Discontinuation vs Continuation | Depression | Education_level | 0.956 | 0.928 | 0.984 | 2.69e-03 | 1.88e-02 | 1958 | 1596 | 362 |
| European | Recorded Depression | Discontinuation vs Continuation | Depression | Depression | 0.899 | 0.800 | 1.011 | 7.46e-02 | 5.22e-01 | 1958 | 1596 | 362 |
| European | Recorded Depression | Switching vs Continuation | Depression | (Intercept) | 1.127 | 0.674 | 1.886 | 6.48e-01 | 1.00e+00 | 2251 | 1596 | 655 |
| European | Recorded Depression | Switching vs Continuation | Depression | age_at_first_AD | 0.990 | 0.982 | 0.999 | 2.43e-02 | 1.70e-01 | 2251 | 1596 | 655 |
| European | Recorded Depression | Switching vs Continuation | Depression | sex_at_birth | 1.254 | 1.021 | 1.541 | 3.12e-02 | 2.19e-01 | 2251 | 1596 | 655 |
| European | Recorded Depression | Switching vs Continuation | Depression | Income_level | 1.000 | 1.000 | 1.000 | 5.57e-02 | 3.90e-01 | 2251 | 1596 | 655 |
| European | Recorded Depression | Switching vs Continuation | Depression | Education_level | 0.973 | 0.950 | 0.996 | 2.08e-02 | 1.46e-01 | 2251 | 1596 | 655 |
| European | Recorded Depression | Switching vs Continuation | Depression | Depression | 1.113 | 1.013 | 1.223 | 2.58e-02 | 1.81e-01 | 2251 | 1596 | 655 |

| | | | | | | | | | | | | |
|---|---|---|---|---|---|---|---|---|---|---|---|---|
| European | Recorded Depression | Discontinuation vs Continuation | Insomnia | (Intercept) | 0.241 | 0.126 | 0.462 | 1.85e-05 | 1.30e-04 | 1958 | 1596 | 362 |
| European | Recorded Depression | Discontinuation vs Continuation | Insomnia | age_at_first_AD | 1.012 | 1.001 | 1.022 | 2.81e-02 | 1.97e-01 | 1958 | 1596 | 362 |
| European | Recorded Depression | Discontinuation vs Continuation | Insomnia | sex_at_birth | 1.309 | 1.018 | 1.685 | 3.60e-02 | 2.52e-01 | 1958 | 1596 | 362 |
| European | Recorded Depression | Discontinuation vs Continuation | Insomnia | Income_level | 1.000 | 1.000 | 1.000 | 8.43e-01 | 1.00e+00 | 1958 | 1596 | 362 |
| European | Recorded Depression | Discontinuation vs Continuation | Insomnia | Education_level | 0.958 | 0.930 | 0.987 | 4.68e-03 | 3.28e-02 | 1958 | 1596 | 362 |
| European | Recorded Depression | Discontinuation vs Continuation | Insomnia | Insomnia | 1.049 | 0.934 | 1.179 | 4.19e-01 | 1.00e+00 | 1958 | 1596 | 362 |
| European | Recorded Depression | Switching vs Continuation | Insomnia | (Intercept) | 1.148 | 0.687 | 1.919 | 5.99e-01 | 1.00e+00 | 2251 | 1596 | 655 |
| European | Recorded Depression | Switching vs Continuation | Insomnia | age_at_first_AD | 0.990 | 0.982 | 0.998 | 2.00e-02 | 1.40e-01 | 2251 | 1596 | 655 |
| European | Recorded Depression | Switching vs Continuation | Insomnia | sex_at_birth | 1.266 | 1.031 | 1.556 | 2.45e-02 | 1.71e-01 | 2251 | 1596 | 655 |
| European | Recorded Depression | Switching vs Continuation | Insomnia | Income_level | 1.000 | 1.000 | 1.000 | 5.89e-02 | 4.12e-01 | 2251 | 1596 | 655 |
| European | Recorded Depression | Switching vs Continuation | Insomnia | Education_level | 0.972 | 0.949 | 0.995 | 1.81e-02 | 1.27e-01 | 2251 | 1596 | 655 |
| European | Recorded Depression | Switching vs Continuation | Insomnia | Insomnia | 1.076 | 0.981 | 1.179 | 1.20e-01 | 8.37e-01 | 2251 | 1596 | 655 |
| European | Recorded Depression | Discontinuation vs Continuation | Migraine | (Intercept) | 0.241 | 0.126 | 0.461 | 1.75e-05 | 1.23e-04 | 1958 | 1596 | 362 |
| European | Recorded Depression | Discontinuation vs Continuation | Migraine | age_at_first_AD | 1.012 | 1.001 | 1.022 | 2.75e-02 | 1.93e-01 | 1958 | 1596 | 362 |
| European | Recorded Depression | Discontinuation vs Continuation | Migraine | sex_at_birth | 1.306 | 1.015 | 1.681 | 3.78e-02 | 2.65e-01 | 1958 | 1596 | 362 |
| European | Recorded Depression | Discontinuation vs Continuation | Migraine | Income_level | 1.000 | 1.000 | 1.000 | 8.23e-01 | 1.00e+00 | 1958 | 1596 | 362 |
| European | Recorded Depression | Discontinuation vs Continuation | Migraine | Education_level | 0.959 | 0.931 | 0.987 | 4.93e-03 | 3.45e-02 | 1958 | 1596 | 362 |
| European | Recorded Depression | Discontinuation vs Continuation | Migraine | Migraine | 1.090 | 0.972 | 1.221 | 1.39e-01 | 9.76e-01 | 1958 | 1596 | 362 |
| European | Recorded Depression | Switching vs Continuation | Migraine | (Intercept) | 1.168 | 0.699 | 1.950 | 5.54e-01 | 1.00e+00 | 2251 | 1596 | 655 |
| European | Recorded Depression | Switching vs Continuation | Migraine | age_at_first_AD | 0.990 | 0.982 | 0.998 | 1.82e-02 | 1.27e-01 | 2251 | 1596 | 655 |
| European | Recorded Depression | Switching vs Continuation | Migraine | sex_at_birth | 1.266 | 1.031 | 1.555 | 2.46e-02 | 1.72e-01 | 2251 | 1596 | 655 |
| European | Recorded Depression | Switching vs Continuation | Migraine | Income_level | 1.000 | 1.000 | 1.000 | 5.87e-02 | 4.11e-01 | 2251 | 1596 | 655 |
| European | Recorded Depression | Switching vs Continuation | Migraine | Education_level | 0.971 | 0.949 | 0.994 | 1.54e-02 | 1.08e-01 | 2251 | 1596 | 655 |
| European | Recorded Depression | Switching vs Continuation | Migraine | Migraine | 1.051 | 0.959 | 1.152 | 2.87e-01 | 1.00e+00 | 2251 | 1596 | 655 |
| European | Recorded Depression | Discontinuation vs Continuation | Schizophrenia | (Intercept) | 0.244 | 0.127 | 0.467 | 2.09e-05 | 1.46e-04 | 1958 | 1596 | 362 |
| European | Recorded Depression | Discontinuation vs Continuation | Schizophrenia | age_at_first_AD | 1.012 | 1.001 | 1.022 | 2.86e-02 | 2.00e-01 | 1958 | 1596 | 362 |
| European | Recorded Depression | Discontinuation vs Continuation | Schizophrenia | sex_at_birth | 1.307 | 1.016 | 1.681 | 3.75e-02 | 2.62e-01 | 1958 | 1596 | 362 |
| European | Recorded Depression | Discontinuation vs Continuation | Schizophrenia | Income_level | 1.000 | 1.000 | 1.000 | 8.31e-01 | 1.00e+00 | 1958 | 1596 | 362 |
| European | Recorded Depression | Discontinuation vs Continuation | Schizophrenia | Education_level | 0.958 | 0.930 | 0.987 | 4.25e-03 | 2.97e-02 | 1958 | 1596 | 362 |
| European | Recorded Depression | Discontinuation vs Continuation | Schizophrenia | Schizophrenia | 1.006 | 0.897 | 1.128 | 9.16e-01 | 1.00e+00 | 1958 | 1596 | 362 |
| European | Recorded Depression | Switching vs Continuation | Schizophrenia | (Intercept) | 1.173 | 0.702 | 1.959 | 5.42e-01 | 1.00e+00 | 2251 | 1596 | 655 |
| European | Recorded Depression | Switching vs Continuation | Schizophrenia | age_at_first_AD | 0.990 | 0.982 | 0.998 | 1.84e-02 | 1.29e-01 | 2251 | 1596 | 655 |
| European | Recorded Depression | Switching vs Continuation | Schizophrenia | sex_at_birth | 1.268 | 1.032 | 1.557 | 2.38e-02 | 1.66e-01 | 2251 | 1596 | 655 |
| European | Recorded Depression | Switching vs Continuation | Schizophrenia | Income_level | 1.000 | 1.000 | 1.000 | 5.80e-02 | 4.06e-01 | 2251 | 1596 | 655 |
| European | Recorded Depression | Switching vs Continuation | Schizophrenia | Education_level | 0.971 | 0.948 | 0.994 | 1.42e-02 | 9.95e-02 | 2251 | 1596 | 655 |
| European | Recorded Depression | Switching vs Continuation | Schizophrenia | Schizophrenia | 1.009 | 0.922 | 1.104 | 8.48e-01 | 1.00e+00 | 2251 | 1596 | 655 |
| European | No Recorded Depression | Discontinuation vs Continuation | ADHD | (Intercept) | 0.264 | 0.175 | 0.396 | 1.52e-10 | 1.06e-09 | 3619 | 2572 | 1047 |
| European | No Recorded Depression | Discontinuation vs Continuation | ADHD | age_at_first_AD | 1.014 | 1.008 | 1.019 | 1.23e-06 | 8.63e-06 | 3619 | 2572 | 1047 |
| European | No Recorded Depression | Discontinuation vs Continuation | ADHD | sex_at_birth | 1.274 | 1.092 | 1.486 | 2.11e-03 | 1.47e-02 | 3619 | 2572 | 1047 |
| European | No Recorded Depression | Discontinuation vs Continuation | ADHD | Income_level | 1.000 | 1.000 | 1.000 | 2.70e-01 | 1.00e+00 | 3619 | 2572 | 1047 |
| European | No Recorded Depression | Discontinuation vs Continuation | ADHD | Education_level | 0.970 | 0.951 | 0.989 | 1.94e-03 | 1.36e-02 | 3619 | 2572 | 1047 |
| European | No Recorded Depression | Discontinuation vs Continuation | ADHD | ADHD | 1.076 | 1.001 | 1.157 | 4.80e-02 | 3.36e-01 | 3619 | 2572 | 1047 |
| European | No Recorded Depression | Switching vs Continuation | ADHD | (Intercept) | 0.560 | 0.342 | 0.915 | 2.07e-02 | 1.45e-01 | 3182 | 2572 | 610 |
| European | No Recorded Depression | Switching vs Continuation | ADHD | age_at_first_AD | 0.986 | 0.979 | 0.993 | 6.10e-05 | 4.27e-04 | 3182 | 2572 | 610 |
| European | No Recorded Depression | Switching vs Continuation | ADHD | sex_at_birth | 1.345 | 1.114 | 1.625 | 2.09e-03 | 1.46e-02 | 3182 | 2572 | 610 |
| European | No Recorded Depression | Switching vs Continuation | ADHD | Income_level | 1.000 | 1.000 | 1.000 | 1.25e-01 | 8.77e-01 | 3182 | 2572 | 610 |
| European | No Recorded Depression | Switching vs Continuation | ADHD | Education_level | 0.993 | 0.970 | 1.016 | 5.35e-01 | 1.00e+00 | 3182 | 2572 | 610 |
| European | No Recorded Depression | Switching vs Continuation | ADHD | ADHD | 1.061 | 0.970 | 1.161 | 1.93e-01 | 1.00e+00 | 3182 | 2572 | 610 |

| | | | | | | | | | | | | |
|---|---|---|---|---|---|---|---|---|---|---|---|---|
| European | No Recorded Depression | Discontinuation vs Continuation | BMI | (Intercept) | 0.267 | 0.177 | 0.401 | 2.16e-10 | 1.51e-09 | 3619 | 2572 | 1047 |
| European | No Recorded Depression | Discontinuation vs Continuation | BMI | age_at_first_AD | 1.014 | 1.008 | 1.019 | 1.47e-06 | 1.03e-05 | 3619 | 2572 | 1047 |
| European | No Recorded Depression | Discontinuation vs Continuation | BMI | sex_at_birth | 1.273 | 1.091 | 1.485 | 2.16e-03 | 1.51e-02 | 3619 | 2572 | 1047 |
| European | No Recorded Depression | Discontinuation vs Continuation | BMI | Income_level | 1.000 | 1.000 | 1.000 | 2.78e-01 | 1.00e+00 | 3619 | 2572 | 1047 |
| European | No Recorded Depression | Discontinuation vs Continuation | BMI | Education_level | 0.970 | 0.951 | 0.989 | 1.76e-03 | 1.23e-02 | 3619 | 2572 | 1047 |
| European | No Recorded Depression | Discontinuation vs Continuation | BMI | BMI | 1.053 | 0.980 | 1.133 | 1.60e-01 | 1.00e+00 | 3619 | 2572 | 1047 |
| European | No Recorded Depression | Switching vs Continuation | BMI | (Intercept) | 0.581 | 0.356 | 0.950 | 3.06e-02 | 2.14e-01 | 3182 | 2572 | 610 |
| European | No Recorded Depression | Switching vs Continuation | BMI | age_at_first_AD | 0.985 | 0.978 | 0.992 | 4.85e-05 | 3.40e-04 | 3182 | 2572 | 610 |
| European | No Recorded Depression | Switching vs Continuation | BMI | sex_at_birth | 1.345 | 1.113 | 1.624 | 2.11e-03 | 1.48e-02 | 3182 | 2572 | 610 |
| European | No Recorded Depression | Switching vs Continuation | BMI | Income_level | 1.000 | 1.000 | 1.000 | 1.10e-01 | 7.71e-01 | 3182 | 2572 | 610 |
| European | No Recorded Depression | Switching vs Continuation | BMI | Education_level | 0.991 | 0.968 | 1.015 | 4.57e-01 | 1.00e+00 | 3182 | 2572 | 610 |
| European | No Recorded Depression | Switching vs Continuation | BMI | BMI | 0.979 | 0.896 | 1.070 | 6.43e-01 | 1.00e+00 | 3182 | 2572 | 610 |
| European | No Recorded Depression | Discontinuation vs Continuation | Bipolar Disorder | (Intercept) | 0.273 | 0.182 | 0.410 | 3.75e-10 | 2.63e-09 | 3619 | 2572 | 1047 |
| European | No Recorded Depression | Discontinuation vs Continuation | Bipolar Disorder | age_at_first_AD | 1.014 | 1.008 | 1.019 | 1.74e-06 | 1.21e-05 | 3619 | 2572 | 1047 |
| European | No Recorded Depression | Discontinuation vs Continuation | Bipolar Disorder | sex_at_birth | 1.273 | 1.091 | 1.485 | 2.18e-03 | 1.53e-02 | 3619 | 2572 | 1047 |
| European | No Recorded Depression | Discontinuation vs Continuation | Bipolar Disorder | Income_level | 1.000 | 1.000 | 1.000 | 2.89e-01 | 1.00e+00 | 3619 | 2572 | 1047 |
| European | No Recorded Depression | Discontinuation vs Continuation | Bipolar Disorder | Education_level | 0.969 | 0.950 | 0.987 | 1.15e-03 | 8.08e-03 | 3619 | 2572 | 1047 |
| European | No Recorded Depression | Discontinuation vs Continuation | Bipolar Disorder | Bipolar Disorder | 1.014 | 0.944 | 1.090 | 6.97e-01 | 1.00e+00 | 3619 | 2572 | 1047 |
| European | No Recorded Depression | Switching vs Continuation | Bipolar Disorder | (Intercept) | 0.574 | 0.351 | 0.937 | 2.65e-02 | 1.85e-01 | 3182 | 2572 | 610 |
| European | No Recorded Depression | Switching vs Continuation | Bipolar Disorder | age_at_first_AD | 0.986 | 0.979 | 0.993 | 6.12e-05 | 4.29e-04 | 3182 | 2572 | 610 |
| European | No Recorded Depression | Switching vs Continuation | Bipolar Disorder | sex_at_birth | 1.350 | 1.118 | 1.631 | 1.84e-03 | 1.29e-02 | 3182 | 2572 | 610 |
| European | No Recorded Depression | Switching vs Continuation | Bipolar Disorder | Income_level | 1.000 | 1.000 | 1.000 | 1.02e-01 | 7.14e-01 | 3182 | 2572 | 610 |
| European | No Recorded Depression | Switching vs Continuation | Bipolar Disorder | Education_level | 0.992 | 0.969 | 1.015 | 4.84e-01 | 1.00e+00 | 3182 | 2572 | 610 |
| European | No Recorded Depression | Switching vs Continuation | Bipolar Disorder | Bipolar Disorder | 1.080 | 0.989 | 1.179 | 8.70e-02 | 6.09e-01 | 3182 | 2572 | 610 |
| European | No Recorded Depression | Discontinuation vs Continuation | Depression | (Intercept) | 0.273 | 0.182 | 0.410 | 3.99e-10 | 2.79e-09 | 3619 | 2572 | 1047 |
| European | No Recorded Depression | Discontinuation vs Continuation | Depression | age_at_first_AD | 1.014 | 1.008 | 1.019 | 1.81e-06 | 1.27e-05 | 3619 | 2572 | 1047 |
| European | No Recorded Depression | Discontinuation vs Continuation | Depression | sex_at_birth | 1.272 | 1.090 | 1.483 | 2.26e-03 | 1.58e-02 | 3619 | 2572 | 1047 |
| European | No Recorded Depression | Discontinuation vs Continuation | Depression | Income_level | 1.000 | 1.000 | 1.000 | 2.85e-01 | 1.00e+00 | 3619 | 2572 | 1047 |
| European | No Recorded Depression | Discontinuation vs Continuation | Depression | Education_level | 0.968 | 0.950 | 0.987 | 1.15e-03 | 8.05e-03 | 3619 | 2572 | 1047 |
| European | No Recorded Depression | Discontinuation vs Continuation | Depression | Depression | 1.005 | 0.935 | 1.080 | 8.97e-01 | 1.00e+00 | 3619 | 2572 | 1047 |
| European | No Recorded Depression | Switching vs Continuation | Depression | (Intercept) | 0.542 | 0.331 | 0.887 | 1.47e-02 | 1.03e-01 | 3182 | 2572 | 610 |
| European | No Recorded Depression | Switching vs Continuation | Depression | age_at_first_AD | 0.986 | 0.979 | 0.993 | 1.23e-04 | 8.59e-04 | 3182 | 2572 | 610 |
| European | No Recorded Depression | Switching vs Continuation | Depression | sex_at_birth | 1.345 | 1.114 | 1.626 | 2.11e-03 | 1.47e-02 | 3182 | 2572 | 610 |
| European | No Recorded Depression | Switching vs Continuation | Depression | Income_level | 1.000 | 1.000 | 1.000 | 1.27e-01 | 8.87e-01 | 3182 | 2572 | 610 |
| European | No Recorded Depression | Switching vs Continuation | Depression | Education_level | 0.992 | 0.969 | 1.016 | 5.19e-01 | 1.00e+00 | 3182 | 2572 | 610 |
| European | No Recorded Depression | Switching vs Continuation | Depression | Depression | 1.172 | 1.073 | 1.280 | 4.40e-04 | 3.08e-03 | 3182 | 2572 | 610 |
| European | No Recorded Depression | Discontinuation vs Continuation | Insomnia | (Intercept) | 0.275 | 0.183 | 0.412 | 4.85e-10 | 3.40e-09 | 3619 | 2572 | 1047 |
| European | No Recorded Depression | Discontinuation vs Continuation | Insomnia | age_at_first_AD | 1.013 | 1.008 | 1.019 | 2.03e-06 | 1.42e-05 | 3619 | 2572 | 1047 |
| European | No Recorded Depression | Discontinuation vs Continuation | Insomnia | sex_at_birth | 1.272 | 1.090 | 1.484 | 2.21e-03 | 1.54e-02 | 3619 | 2572 | 1047 |
| European | No Recorded Depression | Discontinuation vs Continuation | Insomnia | Income_level | 1.000 | 1.000 | 1.000 | 2.91e-01 | 1.00e+00 | 3619 | 2572 | 1047 |
| European | No Recorded Depression | Discontinuation vs Continuation | Insomnia | Education_level | 0.968 | 0.950 | 0.987 | 1.10e-03 | 7.68e-03 | 3619 | 2572 | 1047 |
| European | No Recorded Depression | Discontinuation vs Continuation | Insomnia | Insomnia | 0.988 | 0.918 | 1.063 | 7.39e-01 | 1.00e+00 | 3619 | 2572 | 1047 |
| European | No Recorded Depression | Switching vs Continuation | Insomnia | (Intercept) | 0.555 | 0.339 | 0.907 | 1.87e-02 | 1.31e-01 | 3182 | 2572 | 610 |
| European | No Recorded Depression | Switching vs Continuation | Insomnia | age_at_first_AD | 0.986 | 0.979 | 0.993 | 7.24e-05 | 5.07e-04 | 3182 | 2572 | 610 |
| European | No Recorded Depression | Switching vs Continuation | Insomnia | sex_at_birth | 1.343 | 1.112 | 1.623 | 2.19e-03 | 1.53e-02 | 3182 | 2572 | 610 |
| European | No Recorded Depression | Switching vs Continuation | Insomnia | Income_level | 1.000 | 1.000 | 1.000 | 1.29e-01 | 9.00e-01 | 3182 | 2572 | 610 |
| European | No Recorded Depression | Switching vs Continuation | Insomnia | Education_level | 0.993 | 0.969 | 1.016 | 5.30e-01 | 1.00e+00 | 3182 | 2572 | 610 |
| European | No Recorded Depression | Switching vs Continuation | Insomnia | Insomnia | 1.090 | 0.996 | 1.193 | 6.07e-02 | 4.25e-01 | 3182 | 2572 | 610 |

| | | | | | | | | | | | | |
|---|---|---|---|---|---|---|---|---|---|---|---|---|
| European | No Recorded Depression | Discontinuation vs Continuation | Migraine | (Intercept) | 0.275 | 0.183 | 0.413 | 4.91e-10 | 3.43e-09 | 3619 | 2572 | 1047 |
| European | No Recorded Depression | Discontinuation vs Continuation | Migraine | age_at_first_AD | 1.014 | 1.008 | 1.019 | 1.64e-06 | 1.15e-05 | 3619 | 2572 | 1047 |
| European | No Recorded Depression | Discontinuation vs Continuation | Migraine | sex_at_birth | 1.274 | 1.092 | 1.486 | 2.12e-03 | 1.48e-02 | 3619 | 2572 | 1047 |
| European | No Recorded Depression | Discontinuation vs Continuation | Migraine | Income_level | 1.000 | 1.000 | 1.000 | 3.32e-01 | 1.00e+00 | 3619 | 2572 | 1047 |
| European | No Recorded Depression | Discontinuation vs Continuation | Migraine | Education_level | 0.968 | 0.950 | 0.987 | 1.10e-03 | 7.71e-03 | 3619 | 2572 | 1047 |
| European | No Recorded Depression | Discontinuation vs Continuation | Migraine | Migraine | 0.933 | 0.868 | 1.003 | 6.05e-02 | 4.23e-01 | 3619 | 2572 | 1047 |
| European | No Recorded Depression | Switching vs Continuation | Migraine | (Intercept) | 0.574 | 0.352 | 0.937 | 2.64e-02 | 1.85e-01 | 3182 | 2572 | 610 |
| European | No Recorded Depression | Switching vs Continuation | Migraine | age_at_first_AD | 0.985 | 0.978 | 0.993 | 5.15e-05 | 3.61e-04 | 3182 | 2572 | 610 |
| European | No Recorded Depression | Switching vs Continuation | Migraine | sex_at_birth | 1.344 | 1.113 | 1.623 | 2.14e-03 | 1.50e-02 | 3182 | 2572 | 610 |
| European | No Recorded Depression | Switching vs Continuation | Migraine | Income_level | 1.000 | 1.000 | 1.000 | 1.15e-01 | 8.03e-01 | 3182 | 2572 | 610 |
| European | No Recorded Depression | Switching vs Continuation | Migraine | Education_level | 0.992 | 0.969 | 1.015 | 4.83e-01 | 1.00e+00 | 3182 | 2572 | 610 |
| European | No Recorded Depression | Switching vs Continuation | Migraine | Migraine | 1.022 | 0.937 | 1.115 | 6.23e-01 | 1.00e+00 | 3182 | 2572 | 610 |
| European | No Recorded Depression | Discontinuation vs Continuation | Schizophrenia | (Intercept) | 0.273 | 0.182 | 0.410 | 3.87e-10 | 2.71e-09 | 3619 | 2572 | 1047 |
| European | No Recorded Depression | Discontinuation vs Continuation | Schizophrenia | age_at_first_AD | 1.014 | 1.008 | 1.019 | 1.76e-06 | 1.23e-05 | 3619 | 2572 | 1047 |
| European | No Recorded Depression | Discontinuation vs Continuation | Schizophrenia | sex_at_birth | 1.272 | 1.090 | 1.483 | 2.25e-03 | 1.58e-02 | 3619 | 2572 | 1047 |
| European | No Recorded Depression | Discontinuation vs Continuation | Schizophrenia | Income_level | 1.000 | 1.000 | 1.000 | 2.88e-01 | 1.00e+00 | 3619 | 2572 | 1047 |
| European | No Recorded Depression | Discontinuation vs Continuation | Schizophrenia | Education_level | 0.968 | 0.950 | 0.987 | 1.12e-03 | 7.87e-03 | 3619 | 2572 | 1047 |
| European | No Recorded Depression | Discontinuation vs Continuation | Schizophrenia | Schizophrenia | 1.010 | 0.939 | 1.086 | 7.83e-01 | 1.00e+00 | 3619 | 2572 | 1047 |
| European | No Recorded Depression | Switching vs Continuation | Schizophrenia | (Intercept) | 0.566 | 0.346 | 0.925 | 2.31e-02 | 1.61e-01 | 3182 | 2572 | 610 |
| European | No Recorded Depression | Switching vs Continuation | Schizophrenia | age_at_first_AD | 0.986 | 0.979 | 0.993 | 9.17e-05 | 6.42e-04 | 3182 | 2572 | 610 |
| European | No Recorded Depression | Switching vs Continuation | Schizophrenia | sex_at_birth | 1.344 | 1.113 | 1.624 | 2.16e-03 | 1.51e-02 | 3182 | 2572 | 610 |
| European | No Recorded Depression | Switching vs Continuation | Schizophrenia | Income_level | 1.000 | 1.000 | 1.000 | 1.06e-01 | 7.45e-01 | 3182 | 2572 | 610 |
| European | No Recorded Depression | Switching vs Continuation | Schizophrenia | Education_level | 0.991 | 0.968 | 1.015 | 4.65e-01 | 1.00e+00 | 3182 | 2572 | 610 |
| European | No Recorded Depression | Switching vs Continuation | Schizophrenia | Schizophrenia | 1.115 | 1.021 | 1.219 | 1.59e-02 | 1.11e-01 | 3182 | 2572 | 610 |

**Supplementary Table 6. Multivariable logistic regression results for PGS associations with antidepressant switching (sensitivity definition) across ancestry groups and clinical subgroups in All of Us.** Each model estimates the association between 7 PGS traits and the binary outcome switching versus continuation — where switching is operationalised using the sensitivity definition, restricting switching events to those occurring within six months of index antidepressant initiation (retaining the same 90-day post-discontinuation switching window and <28-day cross-taper exclusion as the primary definition used in Supplementary Table 4) — fitted separately within each combination of genetically inferred ancestry (European, African, Latino/Mixed-American), clinical group (Full Cohort; subgroups defined by recorded depression status, recorded anxiety, and CIDI-SF–derived MDD classification), and PGS exposure. Within each model, one PGS exposure is modelled alongside a common covariate set (age at first antidepressant, sex at birth, income, and educational attainment). All PGS were residualised on the top 16 global genetic ancestry principal components and the residuals standardised to mean 0, SD 1; each odds ratio therefore reflects the change in odds per one-standard-deviation increase in the PC-adjusted PGS. Because ancestry PCs were regressed out at this stage, they are not included in the model covariate set. Columns: Population = inferred genetic ancestry; ClinicalGroup = cohort or clinical subgroup; Outcome = the binary contrast modelled; PGS = the PGS trait for that model; variable = the individual model term; odds_ratio = exponentiated coefficient; lower_95/upper_95 = 95% confidence interval bounds for the odds ratio; p_value = nominal (uncorrected) p-value; Bonf_P = Bonferroni-corrected p-value (capped at 1.00); N_Total = model sample size; N_Reference = number in the reference (continuation) group; N_Case = number in the switching group. Odds ratios >1 indicate higher odds of switching relative to continuation. Bonferroni correction was applied within each specification (ancestry × clinical group) for the max 7 PGS traits tested. P-values reported as "<2.2e-16" reflect R's floating-point reporting limit.
Reference (omitted) categories: Sex at birth: Female; Income: lowest band (<$25k); Education level: lowest band (less than high school / fewer than twelve years).

| Population | ClinicalGroup | Outcome | PGS | variable | odds_ratio | lower_95 | upper_95 | p_value | Bonf_P | N_Total | N_Reference | N_Case |
|---|---|---|---|---|---|---|---|---|---|---|---|---|
| European | Full Cohort | Switching vs Continuation | ADHD | (Intercept) | 1.628 | 1.445 | 1.834 | 1.05e-15 | NA | 30916 | 22804 | 8112 |
| European | Full Cohort | Switching vs Continuation | ADHD | age_at_first_AD | 0.978 | 0.977 | 0.980 | <2.2e-16 | NA | 30916 | 22804 | 8112 |
| European | Full Cohort | Switching vs Continuation | ADHD | sex_at_birthMale | 1.080 | 1.019 | 1.144 | 9.04e-03 | NA | 30916 | 22804 | 8112 |
| European | Full Cohort | Switching vs Continuation | ADHD | Income_quantitative | 0.867 | 0.855 | 0.878 | <2.2e-16 | NA | 30916 | 22804 | 8112 |
| European | Full Cohort | Switching vs Continuation | ADHD | Education_levelCollege Undergraduate | 1.102 | 1.032 | 1.176 | 3.48e-03 | NA | 30916 | 22804 | 8112 |
| European | Full Cohort | Switching vs Continuation | ADHD | Education_levelLess than a high school degree | 1.341 | 1.099 | 1.637 | 3.85e-03 | NA | 30916 | 22804 | 8112 |
| European | Full Cohort | Switching vs Continuation | ADHD | Education_levelTwelve or GED | 1.336 | 1.215 | 1.468 | 1.85e-09 | NA | 30916 | 22804 | 8112 |
| European | Full Cohort | Switching vs Continuation | ADHD | ADHD | 1.081 | 1.052 | 1.110 | 1.41e-08 | 9.87e-08 | 30916 | 22804 | 8112 |
| European | Full Cohort | Switching vs Continuation | BMI | (Intercept) | 1.628 | 1.445 | 1.834 | 1.07e-15 | NA | 30916 | 22804 | 8112 |
| European | Full Cohort | Switching vs Continuation | BMI | age_at_first_AD | 0.978 | 0.977 | 0.980 | <2.2e-16 | NA | 30916 | 22804 | 8112 |
| European | Full Cohort | Switching vs Continuation | BMI | sex_at_birthMale | 1.077 | 1.017 | 1.141 | 1.16e-02 | NA | 30916 | 22804 | 8112 |
| European | Full Cohort | Switching vs Continuation | BMI | Income_quantitative | 0.866 | 0.854 | 0.877 | <2.2e-16 | NA | 30916 | 22804 | 8112 |
| European | Full Cohort | Switching vs Continuation | BMI | Education_levelCollege Undergraduate | 1.110 | 1.040 | 1.185 | 1.62e-03 | NA | 30916 | 22804 | 8112 |
| European | Full Cohort | Switching vs Continuation | BMI | Education_levelLess than a high school degree | 1.377 | 1.129 | 1.679 | 1.61e-03 | NA | 30916 | 22804 | 8112 |
| European | Full Cohort | Switching vs Continuation | BMI | Education_levelTwelve or GED | 1.351 | 1.230 | 1.485 | 3.98e-10 | NA | 30916 | 22804 | 8112 |
| European | Full Cohort | Switching vs Continuation | BMI | BMI | 1.049 | 1.021 | 1.077 | 4.13e-04 | 2.89e-03 | 30916 | 22804 | 8112 |
| European | Full Cohort | Switching vs Continuation | Bipolar Disorder | (Intercept) | 1.641 | 1.456 | 1.848 | 3.57e-16 | NA | 30916 | 22804 | 8112 |
| European | Full Cohort | Switching vs Continuation | Bipolar Disorder | age_at_first_AD | 0.978 | 0.977 | 0.980 | <2.2e-16 | NA | 30916 | 22804 | 8112 |
| European | Full Cohort | Switching vs Continuation | Bipolar Disorder | sex_at_birthMale | 1.075 | 1.015 | 1.139 | 1.38e-02 | NA | 30916 | 22804 | 8112 |
| European | Full Cohort | Switching vs Continuation | Bipolar Disorder | Income_quantitative | 0.864 | 0.853 | 0.876 | <2.2e-16 | NA | 30916 | 22804 | 8112 |
| European | Full Cohort | Switching vs Continuation | Bipolar Disorder | Education_levelCollege Undergraduate | 1.119 | 1.048 | 1.194 | 7.03e-04 | NA | 30916 | 22804 | 8112 |
| European | Full Cohort | Switching vs Continuation | Bipolar Disorder | Education_levelLess than a high school degree | 1.399 | 1.147 | 1.706 | 9.21e-04 | NA | 30916 | 22804 | 8112 |
| European | Full Cohort | Switching vs Continuation | Bipolar Disorder | Education_levelTwelve or GED | 1.372 | 1.249 | 1.508 | 4.41e-11 | NA | 30916 | 22804 | 8112 |
| European | Full Cohort | Switching vs Continuation | Bipolar Disorder | Bipolar Disorder | 1.058 | 1.030 | 1.088 | 5.41e-05 | 3.79e-04 | 30916 | 22804 | 8112 |
| European | Full Cohort | Switching vs Continuation | Depression | (Intercept) | 1.588 | 1.409 | 1.789 | 3.52e-14 | NA | 30916 | 22804 | 8112 |
| European | Full Cohort | Switching vs Continuation | Depression | age_at_first_AD | 0.979 | 0.977 | 0.980 | <2.2e-16 | NA | 30916 | 22804 | 8112 |
| European | Full Cohort | Switching vs Continuation | Depression | sex_at_birthMale | 1.068 | 1.008 | 1.131 | 2.56e-02 | NA | 30916 | 22804 | 8112 |
| European | Full Cohort | Switching vs Continuation | Depression | Income_quantitative | 0.870 | 0.858 | 0.881 | <2.2e-16 | NA | 30916 | 22804 | 8112 |
| European | Full Cohort | Switching vs Continuation | Depression | Education_levelCollege Undergraduate | 1.102 | 1.032 | 1.176 | 3.56e-03 | NA | 30916 | 22804 | 8112 |
| European | Full Cohort | Switching vs Continuation | Depression | Education_levelLess than a high school degree | 1.307 | 1.071 | 1.596 | 8.56e-03 | NA | 30916 | 22804 | 8112 |
| European | Full Cohort | Switching vs Continuation | Depression | Education_levelTwelve or GED | 1.344 | 1.223 | 1.477 | 8.49e-10 | NA | 30916 | 22804 | 8112 |
| European | Full Cohort | Switching vs Continuation | Depression | Depression | 1.213 | 1.180 | 1.247 | <2.2e-16 | <2.2e-16 | 30916 | 22804 | 8112 |
| European | Full Cohort | Switching vs Continuation | Insomnia | (Intercept) | 1.611 | 1.430 | 1.815 | 4.63e-15 | NA | 30916 | 22804 | 8112 |
| European | Full Cohort | Switching vs Continuation | Insomnia | age_at_first_AD | 0.979 | 0.977 | 0.980 | <2.2e-16 | NA | 30916 | 22804 | 8112 |
| European | Full Cohort | Switching vs Continuation | Insomnia | sex_at_birthMale | 1.075 | 1.015 | 1.139 | 1.35e-02 | NA | 30916 | 22804 | 8112 |
| European | Full Cohort | Switching vs Continuation | Insomnia | Income_quantitative | 0.867 | 0.856 | 0.879 | <2.2e-16 | NA | 30916 | 22804 | 8112 |
| European | Full Cohort | Switching vs Continuation | Insomnia | Education_levelCollege Undergraduate | 1.106 | 1.036 | 1.180 | 2.37e-03 | NA | 30916 | 22804 | 8112 |
| European | Full Cohort | Switching vs Continuation | Insomnia | Education_levelLess than a high school degree | 1.332 | 1.091 | 1.625 | 4.78e-03 | NA | 30916 | 22804 | 8112 |
| European | Full Cohort | Switching vs Continuation | Insomnia | Education_levelTwelve or GED | 1.343 | 1.222 | 1.475 | 9.23e-10 | NA | 30916 | 22804 | 8112 |
| European | Full Cohort | Switching vs Continuation | Insomnia | Insomnia | 1.129 | 1.099 | 1.161 | <2.2e-16 | <2.2e-16 | 30916 | 22804 | 8112 |
| European | Full Cohort | Switching vs Continuation | Migraine | (Intercept) | 1.638 | 1.455 | 1.845 | 4.23e-16 | NA | 30916 | 22804 | 8112 |
| European | Full Cohort | Switching vs Continuation | Migraine | age_at_first_AD | 0.978 | 0.977 | 0.980 | <2.2e-16 | NA | 30916 | 22804 | 8112 |
| European | Full Cohort | Switching vs Continuation | Migraine | sex_at_birthMale | 1.076 | 1.016 | 1.140 | 1.22e-02 | NA | 30916 | 22804 | 8112 |
| European | Full Cohort | Switching vs Continuation | Migraine | Income_quantitative | 0.864 | 0.853 | 0.876 | <2.2e-16 | NA | 30916 | 22804 | 8112 |
| European | Full Cohort | Switching vs Continuation | Migraine | Education_levelCollege Undergraduate | 1.115 | 1.045 | 1.189 | 1.03e-03 | NA | 30916 | 22804 | 8112 |
| European | Full Cohort | Switching vs Continuation | Migraine | Education_levelLess than a high school degree | 1.393 | 1.142 | 1.698 | 1.07e-03 | NA | 30916 | 22804 | 8112 |
| European | Full Cohort | Switching vs Continuation | Migraine | Education_levelTwelve or GED | 1.362 | 1.240 | 1.497 | 1.23e-10 | NA | 30916 | 22804 | 8112 |
| European | Full Cohort | Switching vs Continuation | Migraine | Migraine | 1.049 | 1.022 | 1.076 | 3.67e-04 | 2.57e-03 | 30916 | 22804 | 8112 |

| | | | | | | | | | | | | |
|---|---|---|---|---|---|---|---|---|---|---|---|---|
| European | Full Cohort | Switching vs Continuation | Schizophrenia | (Intercept) | 1.637 | 1.454 | 1.844 | 4.61e-16 | NA | 30916 | 22804 | 8112 |
| European | Full Cohort | Switching vs Continuation | Schizophrenia | age_at_first_AD | 0.978 | 0.977 | 0.980 | <2.2e-16 | NA | 30916 | 22804 | 8112 |
| European | Full Cohort | Switching vs Continuation | Schizophrenia | sex_at_birthMale | 1.076 | 1.016 | 1.140 | 1.27e-02 | NA | 30916 | 22804 | 8112 |
| European | Full Cohort | Switching vs Continuation | Schizophrenia | Income_quantitative | 0.864 | 0.853 | 0.876 | <2.2e-16 | NA | 30916 | 22804 | 8112 |
| European | Full Cohort | Switching vs Continuation | Schizophrenia | Education_levelCollege Undergraduate | 1.117 | 1.047 | 1.192 | 8.40e-04 | NA | 30916 | 22804 | 8112 |
| European | Full Cohort | Switching vs Continuation | Schizophrenia | Education_levelLess than a high school degree | 1.392 | 1.142 | 1.698 | 1.09e-03 | NA | 30916 | 22804 | 8112 |
| European | Full Cohort | Switching vs Continuation | Schizophrenia | Education_levelTwelve or GED | 1.366 | 1.244 | 1.501 | 8.06e-11 | NA | 30916 | 22804 | 8112 |
| European | Full Cohort | Switching vs Continuation | Schizophrenia | Schizophrenia | 1.026 | 0.999 | 1.054 | 5.61e-02 | 3.92e-01 | 30916 | 22804 | 8112 |
| European | Recorded Depression | Switching vs Continuation | ADHD | (Intercept) | 2.251 | 1.917 | 2.644 | <2.2e-16 | NA | 14441 | 8835 | 5606 |
| European | Recorded Depression | Switching vs Continuation | ADHD | age_at_first_AD | 0.979 | 0.977 | 0.981 | <2.2e-16 | NA | 14441 | 8835 | 5606 |
| European | Recorded Depression | Switching vs Continuation | ADHD | sex_at_birthMale | 1.162 | 1.077 | 1.253 | 9.99e-05 | NA | 14441 | 8835 | 5606 |
| European | Recorded Depression | Switching vs Continuation | ADHD | Income_quantitative | 0.892 | 0.876 | 0.908 | <2.2e-16 | NA | 14441 | 8835 | 5606 |
| European | Recorded Depression | Switching vs Continuation | ADHD | Education_levelCollege Undergraduate | 1.163 | 1.066 | 1.268 | 6.62e-04 | NA | 14441 | 8835 | 5606 |
| European | Recorded Depression | Switching vs Continuation | ADHD | Education_levelLess than a high school degree | 1.448 | 1.117 | 1.876 | 5.13e-03 | NA | 14441 | 8835 | 5606 |
| European | Recorded Depression | Switching vs Continuation | ADHD | Education_levelTwelve or GED | 1.438 | 1.269 | 1.629 | 1.26e-08 | NA | 14441 | 8835 | 5606 |
| European | Recorded Depression | Switching vs Continuation | ADHD | ADHD | 1.054 | 1.018 | 1.092 | 3.41e-03 | 2.39e-02 | 14441 | 8835 | 5606 |
| European | Recorded Depression | Switching vs Continuation | BMI | (Intercept) | 2.257 | 1.922 | 2.651 | <2.2e-16 | NA | 14441 | 8835 | 5606 |
| European | Recorded Depression | Switching vs Continuation | BMI | age_at_first_AD | 0.979 | 0.977 | 0.981 | <2.2e-16 | NA | 14441 | 8835 | 5606 |
| European | Recorded Depression | Switching vs Continuation | BMI | sex_at_birthMale | 1.159 | 1.075 | 1.250 | 1.25e-04 | NA | 14441 | 8835 | 5606 |
| European | Recorded Depression | Switching vs Continuation | BMI | Income_quantitative | 0.890 | 0.874 | 0.906 | <2.2e-16 | NA | 14441 | 8835 | 5606 |
| European | Recorded Depression | Switching vs Continuation | BMI | Education_levelCollege Undergraduate | 1.174 | 1.077 | 1.281 | 2.68e-04 | NA | 14441 | 8835 | 5606 |
| European | Recorded Depression | Switching vs Continuation | BMI | Education_levelLess than a high school degree | 1.486 | 1.148 | 1.925 | 2.68e-03 | NA | 14441 | 8835 | 5606 |
| European | Recorded Depression | Switching vs Continuation | BMI | Education_levelTwelve or GED | 1.461 | 1.289 | 1.655 | 2.67e-09 | NA | 14441 | 8835 | 5606 |
| European | Recorded Depression | Switching vs Continuation | BMI | BMI | 0.998 | 0.964 | 1.034 | 9.19e-01 | 1.00e+00 | 14441 | 8835 | 5606 |
| European | Recorded Depression | Switching vs Continuation | Bipolar Disorder | (Intercept) | 2.259 | 1.924 | 2.653 | <2.2e-16 | NA | 14441 | 8835 | 5606 |
| European | Recorded Depression | Switching vs Continuation | Bipolar Disorder | age_at_first_AD | 0.979 | 0.977 | 0.981 | <2.2e-16 | NA | 14441 | 8835 | 5606 |
| European | Recorded Depression | Switching vs Continuation | Bipolar Disorder | sex_at_birthMale | 1.159 | 1.075 | 1.250 | 1.27e-04 | NA | 14441 | 8835 | 5606 |
| European | Recorded Depression | Switching vs Continuation | Bipolar Disorder | Income_quantitative | 0.890 | 0.875 | 0.906 | <2.2e-16 | NA | 14441 | 8835 | 5606 |
| European | Recorded Depression | Switching vs Continuation | Bipolar Disorder | Education_levelCollege Undergraduate | 1.175 | 1.078 | 1.281 | 2.58e-04 | NA | 14441 | 8835 | 5606 |
| European | Recorded Depression | Switching vs Continuation | Bipolar Disorder | Education_levelLess than a high school degree | 1.486 | 1.148 | 1.924 | 2.66e-03 | NA | 14441 | 8835 | 5606 |
| European | Recorded Depression | Switching vs Continuation | Bipolar Disorder | Education_levelTwelve or GED | 1.463 | 1.291 | 1.657 | 2.26e-09 | NA | 14441 | 8835 | 5606 |
| European | Recorded Depression | Switching vs Continuation | Bipolar Disorder | Bipolar Disorder | 1.021 | 0.985 | 1.059 | 2.55e-01 | 1.00e+00 | 14441 | 8835 | 5606 |
| European | Recorded Depression | Switching vs Continuation | Depression | (Intercept) | 2.235 | 1.902 | 2.625 | <2.2e-16 | NA | 14441 | 8835 | 5606 |
| European | Recorded Depression | Switching vs Continuation | Depression | age_at_first_AD | 0.979 | 0.977 | 0.981 | <2.2e-16 | NA | 14441 | 8835 | 5606 |
| European | Recorded Depression | Switching vs Continuation | Depression | sex_at_birthMale | 1.153 | 1.069 | 1.244 | 2.27e-04 | NA | 14441 | 8835 | 5606 |
| European | Recorded Depression | Switching vs Continuation | Depression | Income_quantitative | 0.894 | 0.879 | 0.910 | <2.2e-16 | NA | 14441 | 8835 | 5606 |
| European | Recorded Depression | Switching vs Continuation | Depression | Education_levelCollege Undergraduate | 1.164 | 1.067 | 1.269 | 6.01e-04 | NA | 14441 | 8835 | 5606 |
| European | Recorded Depression | Switching vs Continuation | Depression | Education_levelLess than a high school degree | 1.414 | 1.091 | 1.832 | 8.86e-03 | NA | 14441 | 8835 | 5606 |
| European | Recorded Depression | Switching vs Continuation | Depression | Education_levelTwelve or GED | 1.444 | 1.275 | 1.637 | 8.03e-09 | NA | 14441 | 8835 | 5606 |
| European | Recorded Depression | Switching vs Continuation | Depression | Depression | 1.153 | 1.112 | 1.196 | 1.23e-14 | 8.63e-14 | 14441 | 8835 | 5606 |
| European | Recorded Depression | Switching vs Continuation | Insomnia | (Intercept) | 2.227 | 1.896 | 2.615 | <2.2e-16 | NA | 14441 | 8835 | 5606 |
| European | Recorded Depression | Switching vs Continuation | Insomnia | age_at_first_AD | 0.979 | 0.977 | 0.981 | <2.2e-16 | NA | 14441 | 8835 | 5606 |
| European | Recorded Depression | Switching vs Continuation | Insomnia | sex_at_birthMale | 1.157 | 1.073 | 1.248 | 1.60e-04 | NA | 14441 | 8835 | 5606 |
| European | Recorded Depression | Switching vs Continuation | Insomnia | Income_quantitative | 0.893 | 0.877 | 0.909 | <2.2e-16 | NA | 14441 | 8835 | 5606 |
| European | Recorded Depression | Switching vs Continuation | Insomnia | Education_levelCollege Undergraduate | 1.167 | 1.070 | 1.272 | 4.65e-04 | NA | 14441 | 8835 | 5606 |
| European | Recorded Depression | Switching vs Continuation | Insomnia | Education_levelLess than a high school degree | 1.432 | 1.105 | 1.855 | 6.61e-03 | NA | 14441 | 8835 | 5606 |
| European | Recorded Depression | Switching vs Continuation | Insomnia | Education_levelTwelve or GED | 1.442 | 1.273 | 1.634 | 8.98e-09 | NA | 14441 | 8835 | 5606 |
| European | Recorded Depression | Switching vs Continuation | Insomnia | Insomnia | 1.092 | 1.053 | 1.133 | 2.32e-06 | 1.62e-05 | 14441 | 8835 | 5606 |
| European | Recorded Depression | Switching vs Continuation | Migraine | (Intercept) | 2.255 | 1.920 | 2.648 | <2.2e-16 | NA | 14441 | 8835 | 5606 |
| European | Recorded Depression | Switching vs Continuation | Migraine | age_at_first_AD | 0.979 | 0.977 | 0.981 | <2.2e-16 | NA | 14441 | 8835 | 5606 |
| European | Recorded Depression | Switching vs Continuation | Migraine | sex_at_birthMale | 1.160 | 1.076 | 1.251 | 1.17e-04 | NA | 14441 | 8835 | 5606 |
| European | Recorded Depression | Switching vs Continuation | Migraine | Income_quantitative | 0.891 | 0.875 | 0.907 | <2.2e-16 | NA | 14441 | 8835 | 5606 |
| European | Recorded Depression | Switching vs Continuation | Migraine | Education_levelCollege Undergraduate | 1.173 | 1.076 | 1.279 | 3.00e-04 | NA | 14441 | 8835 | 5606 |
| European | Recorded Depression | Switching vs Continuation | Migraine | Education_levelLess than a high school degree | 1.488 | 1.149 | 1.927 | 2.58e-03 | NA | 14441 | 8835 | 5606 |
| European | Recorded Depression | Switching vs Continuation | Migraine | Education_levelTwelve or GED | 1.460 | 1.289 | 1.653 | 2.76e-09 | NA | 14441 | 8835 | 5606 |
| European | Recorded Depression | Switching vs Continuation | Migraine | Migraine | 1.064 | 1.028 | 1.101 | 4.35e-04 | 3.05e-03 | 14441 | 8835 | 5606 |
| European | Recorded Depression | Switching vs Continuation | Schizophrenia | (Intercept) | 2.254 | 1.920 | 2.647 | <2.2e-16 | NA | 14441 | 8835 | 5606 |
| European | Recorded Depression | Switching vs Continuation | Schizophrenia | age_at_first_AD | 0.979 | 0.977 | 0.981 | <2.2e-16 | NA | 14441 | 8835 | 5606 |
| European | Recorded Depression | Switching vs Continuation | Schizophrenia | sex_at_birthMale | 1.159 | 1.075 | 1.250 | 1.29e-04 | NA | 14441 | 8835 | 5606 |
| European | Recorded Depression | Switching vs Continuation | Schizophrenia | Income_quantitative | 0.890 | 0.875 | 0.906 | <2.2e-16 | NA | 14441 | 8835 | 5606 |

| | | | | | | | | | | | | |
|---|---|---|---|---|---|---|---|---|---|---|---|---|
| European | Recorded Depression | Switching vs Continuation | Schizophrenia | Education_levelCollege Undergraduate | 1.174 | 1.077 | 1.280 | 2.68e-04 | NA | 14441 | 8835 | 5606 |
| European | Recorded Depression | Switching vs Continuation | Schizophrenia | Education_levelLess than a high school degree | 1.481 | 1.144 | 1.918 | 2.89e-03 | NA | 14441 | 8835 | 5606 |
| European | Recorded Depression | Switching vs Continuation | Schizophrenia | Education_levelTwelve or GED | 1.461 | 1.290 | 1.655 | 2.46e-09 | NA | 14441 | 8835 | 5606 |
| European | Recorded Depression | Switching vs Continuation | Schizophrenia | Schizophrenia | 1.022 | 0.987 | 1.059 | 2.18e-01 | 1.00e+00 | 14441 | 8835 | 5606 |
| European | No Recorded Depression | Switching vs Continuation | ADHD | (Intercept) | 0.724 | 0.596 | 0.879 | 1.09e-03 | NA | 16475 | 13969 | 2506 |
| European | No Recorded Depression | Switching vs Continuation | ADHD | age_at_first_AD | 0.981 | 0.978 | 0.983 | <2.2e-16 | NA | 16475 | 13969 | 2506 |
| European | No Recorded Depression | Switching vs Continuation | ADHD | sex_at_birthMale | 0.925 | 0.839 | 1.020 | 1.17e-01 | NA | 16475 | 13969 | 2506 |
| European | No Recorded Depression | Switching vs Continuation | ADHD | Income_quantitative | 0.898 | 0.879 | 0.918 | <2.2e-16 | NA | 16475 | 13969 | 2506 |
| European | No Recorded Depression | Switching vs Continuation | ADHD | Education_levelCollege Undergraduate | 1.039 | 0.935 | 1.154 | 4.77e-01 | NA | 16475 | 13969 | 2506 |
| European | No Recorded Depression | Switching vs Continuation | ADHD | Education_levelLess than a high school degree | 1.263 | 0.893 | 1.787 | 1.87e-01 | NA | 16475 | 13969 | 2506 |
| European | No Recorded Depression | Switching vs Continuation | ADHD | Education_levelTwelve or GED | 1.257 | 1.075 | 1.471 | 4.21e-03 | NA | 16475 | 13969 | 2506 |
| European | No Recorded Depression | Switching vs Continuation | ADHD | ADHD | 1.089 | 1.042 | 1.138 | 1.72e-04 | 1.20e-03 | 16475 | 13969 | 2506 |
| European | No Recorded Depression | Switching vs Continuation | BMI | (Intercept) | 0.723 | 0.596 | 0.878 | 1.07e-03 | NA | 16475 | 13969 | 2506 |
| European | No Recorded Depression | Switching vs Continuation | BMI | age_at_first_AD | 0.981 | 0.978 | 0.983 | <2.2e-16 | NA | 16475 | 13969 | 2506 |
| European | No Recorded Depression | Switching vs Continuation | BMI | sex_at_birthMale | 0.922 | 0.837 | 1.017 | 1.04e-01 | NA | 16475 | 13969 | 2506 |
| European | No Recorded Depression | Switching vs Continuation | BMI | Income_quantitative | 0.898 | 0.879 | 0.918 | <2.2e-16 | NA | 16475 | 13969 | 2506 |
| European | No Recorded Depression | Switching vs Continuation | BMI | Education_levelCollege Undergraduate | 1.040 | 0.937 | 1.155 | 4.60e-01 | NA | 16475 | 13969 | 2506 |
| European | No Recorded Depression | Switching vs Continuation | BMI | Education_levelLess than a high school degree | 1.298 | 0.918 | 1.835 | 1.39e-01 | NA | 16475 | 13969 | 2506 |
| European | No Recorded Depression | Switching vs Continuation | BMI | Education_levelTwelve or GED | 1.263 | 1.080 | 1.477 | 3.53e-03 | NA | 16475 | 13969 | 2506 |
| European | No Recorded Depression | Switching vs Continuation | BMI | BMI | 1.084 | 1.038 | 1.133 | 2.73e-04 | 1.91e-03 | 16475 | 13969 | 2506 |
| European | No Recorded Depression | Switching vs Continuation | Bipolar Disorder | (Intercept) | 0.729 | 0.601 | 0.885 | 1.40e-03 | NA | 16475 | 13969 | 2506 |
| European | No Recorded Depression | Switching vs Continuation | Bipolar Disorder | age_at_first_AD | 0.981 | 0.978 | 0.983 | <2.2e-16 | NA | 16475 | 13969 | 2506 |
| European | No Recorded Depression | Switching vs Continuation | Bipolar Disorder | sex_at_birthMale | 0.919 | 0.833 | 1.013 | 8.83e-02 | NA | 16475 | 13969 | 2506 |
| European | No Recorded Depression | Switching vs Continuation | Bipolar Disorder | Income_quantitative | 0.897 | 0.877 | 0.916 | <2.2e-16 | NA | 16475 | 13969 | 2506 |
| European | No Recorded Depression | Switching vs Continuation | Bipolar Disorder | Education_levelCollege Undergraduate | 1.056 | 0.951 | 1.172 | 3.11e-01 | NA | 16475 | 13969 | 2506 |
| European | No Recorded Depression | Switching vs Continuation | Bipolar Disorder | Education_levelLess than a high school degree | 1.329 | 0.940 | 1.879 | 1.07e-01 | NA | 16475 | 13969 | 2506 |
| European | No Recorded Depression | Switching vs Continuation | Bipolar Disorder | Education_levelTwelve or GED | 1.299 | 1.111 | 1.519 | 1.07e-03 | NA | 16475 | 13969 | 2506 |
| European | No Recorded Depression | Switching vs Continuation | Bipolar Disorder | Bipolar Disorder | 1.084 | 1.035 | 1.134 | 5.58e-04 | 3.91e-03 | 16475 | 13969 | 2506 |
| European | No Recorded Depression | Switching vs Continuation | Depression | (Intercept) | 0.704 | 0.580 | 0.856 | 4.21e-04 | NA | 16475 | 13969 | 2506 |
| European | No Recorded Depression | Switching vs Continuation | Depression | age_at_first_AD | 0.981 | 0.978 | 0.984 | <2.2e-16 | NA | 16475 | 13969 | 2506 |
| European | No Recorded Depression | Switching vs Continuation | Depression | sex_at_birthMale | 0.914 | 0.829 | 1.008 | 7.14e-02 | NA | 16475 | 13969 | 2506 |
| European | No Recorded Depression | Switching vs Continuation | Depression | Income_quantitative | 0.901 | 0.881 | 0.921 | <2.2e-16 | NA | 16475 | 13969 | 2506 |
| European | No Recorded Depression | Switching vs Continuation | Depression | Education_levelCollege Undergraduate | 1.037 | 0.934 | 1.152 | 4.93e-01 | NA | 16475 | 13969 | 2506 |
| European | No Recorded Depression | Switching vs Continuation | Depression | Education_levelLess than a high school degree | 1.242 | 0.877 | 1.758 | 2.22e-01 | NA | 16475 | 13969 | 2506 |
| European | No Recorded Depression | Switching vs Continuation | Depression | Education_levelTwelve or GED | 1.266 | 1.082 | 1.481 | 3.21e-03 | NA | 16475 | 13969 | 2506 |
| European | No Recorded Depression | Switching vs Continuation | Depression | Depression | 1.205 | 1.151 | 1.261 | 1.02e-15 | 7.16e-15 | 16475 | 13969 | 2506 |
| European | No Recorded Depression | Switching vs Continuation | Insomnia | (Intercept) | 0.717 | 0.590 | 0.871 | 7.85e-04 | NA | 16475 | 13969 | 2506 |
| European | No Recorded Depression | Switching vs Continuation | Insomnia | age_at_first_AD | 0.981 | 0.978 | 0.983 | <2.2e-16 | NA | 16475 | 13969 | 2506 |
| European | No Recorded Depression | Switching vs Continuation | Insomnia | sex_at_birthMale | 0.923 | 0.837 | 1.017 | 1.06e-01 | NA | 16475 | 13969 | 2506 |
| European | No Recorded Depression | Switching vs Continuation | Insomnia | Income_quantitative | 0.899 | 0.880 | 0.919 | <2.2e-16 | NA | 16475 | 13969 | 2506 |
| European | No Recorded Depression | Switching vs Continuation | Insomnia | Education_levelCollege Undergraduate | 1.039 | 0.936 | 1.154 | 4.74e-01 | NA | 16475 | 13969 | 2506 |
| European | No Recorded Depression | Switching vs Continuation | Insomnia | Education_levelLess than a high school degree | 1.257 | 0.889 | 1.779 | 1.96e-01 | NA | 16475 | 13969 | 2506 |
| European | No Recorded Depression | Switching vs Continuation | Insomnia | Education_levelTwelve or GED | 1.262 | 1.079 | 1.476 | 3.58e-03 | NA | 16475 | 13969 | 2506 |
| European | No Recorded Depression | Switching vs Continuation | Insomnia | Insomnia | 1.162 | 1.110 | 1.216 | 1.03e-10 | 7.23e-10 | 16475 | 13969 | 2506 |
| European | No Recorded Depression | Switching vs Continuation | Migraine | (Intercept) | 0.730 | 0.601 | 0.886 | 1.42e-03 | NA | 16475 | 13969 | 2506 |
| European | No Recorded Depression | Switching vs Continuation | Migraine | age_at_first_AD | 0.981 | 0.978 | 0.983 | <2.2e-16 | NA | 16475 | 13969 | 2506 |
| European | No Recorded Depression | Switching vs Continuation | Migraine | sex_at_birthMale | 0.921 | 0.836 | 1.016 | 9.91e-02 | NA | 16475 | 13969 | 2506 |
| European | No Recorded Depression | Switching vs Continuation | Migraine | Income_quantitative | 0.896 | 0.877 | 0.916 | <2.2e-16 | NA | 16475 | 13969 | 2506 |
| European | No Recorded Depression | Switching vs Continuation | Migraine | Education_levelCollege Undergraduate | 1.051 | 0.946 | 1.166 | 3.56e-01 | NA | 16475 | 13969 | 2506 |
| European | No Recorded Depression | Switching vs Continuation | Migraine | Education_levelLess than a high school degree | 1.318 | 0.933 | 1.862 | 1.18e-01 | NA | 16475 | 13969 | 2506 |
| European | No Recorded Depression | Switching vs Continuation | Migraine | Education_levelTwelve or GED | 1.282 | 1.097 | 1.499 | 1.84e-03 | NA | 16475 | 13969 | 2506 |
| European | No Recorded Depression | Switching vs Continuation | Migraine | Migraine | 1.039 | 0.995 | 1.085 | 8.13e-02 | 5.69e-01 | 16475 | 13969 | 2506 |
| European | No Recorded Depression | Switching vs Continuation | Schizophrenia | (Intercept) | 0.729 | 0.601 | 0.885 | 1.39e-03 | NA | 16475 | 13969 | 2506 |
| European | No Recorded Depression | Switching vs Continuation | Schizophrenia | age_at_first_AD | 0.981 | 0.978 | 0.983 | <2.2e-16 | NA | 16475 | 13969 | 2506 |
| European | No Recorded Depression | Switching vs Continuation | Schizophrenia | sex_at_birthMale | 0.921 | 0.836 | 1.015 | 9.87e-02 | NA | 16475 | 13969 | 2506 |
| European | No Recorded Depression | Switching vs Continuation | Schizophrenia | Income_quantitative | 0.896 | 0.877 | 0.916 | <2.2e-16 | NA | 16475 | 13969 | 2506 |
| European | No Recorded Depression | Switching vs Continuation | Schizophrenia | Education_levelCollege Undergraduate | 1.053 | 0.948 | 1.169 | 3.36e-01 | NA | 16475 | 13969 | 2506 |
| European | No Recorded Depression | Switching vs Continuation | Schizophrenia | Education_levelLess than a high school degree | 1.323 | 0.936 | 1.869 | 1.13e-01 | NA | 16475 | 13969 | 2506 |
| European | No Recorded Depression | Switching vs Continuation | Schizophrenia | Education_levelTwelve or GED | 1.287 | 1.101 | 1.505 | 1.55e-03 | NA | 16475 | 13969 | 2506 |
| European | No Recorded Depression | Switching vs Continuation | Schizophrenia | Schizophrenia | 1.025 | 0.981 | 1.071 | 2.68e-01 | 1.00e+00 | 16475 | 13969 | 2506 |

| | | | | | | | | | | | | |
|---|---|---|---|---|---|---|---|---|---|---|---|---|
| European | Recorded Anxiety | Switching vs Continuation | ADHD | (Intercept) | 1.775 | 1.543 | 2.044 | 1.23e-15 | NA | 19319 | 12943 | 6376 |
| European | Recorded Anxiety | Switching vs Continuation | ADHD | age_at_first_AD | 0.982 | 0.980 | 0.984 | <2.2e-16 | NA | 19319 | 12943 | 6376 |
| European | Recorded Anxiety | Switching vs Continuation | ADHD | sex_at_birthMale | 1.203 | 1.123 | 1.289 | 1.61e-07 | NA | 19319 | 12943 | 6376 |
| European | Recorded Anxiety | Switching vs Continuation | ADHD | Income_quantitative | 0.863 | 0.849 | 0.877 | <2.2e-16 | NA | 19319 | 12943 | 6376 |
| European | Recorded Anxiety | Switching vs Continuation | ADHD | Education_levelCollege Undergraduate | 1.114 | 1.031 | 1.204 | 6.23e-03 | NA | 19319 | 12943 | 6376 |
| European | Recorded Anxiety | Switching vs Continuation | ADHD | Education_levelLess than a high school degree | 1.234 | 0.978 | 1.557 | 7.62e-02 | NA | 19319 | 12943 | 6376 |
| European | Recorded Anxiety | Switching vs Continuation | ADHD | Education_levelTwelve or GED | 1.448 | 1.295 | 1.619 | 8.33e-11 | NA | 19319 | 12943 | 6376 |
| European | Recorded Anxiety | Switching vs Continuation | ADHD | ADHD | 1.065 | 1.032 | 1.100 | 1.06e-04 | 7.43e-04 | 19319 | 12943 | 6376 |
| European | Recorded Anxiety | Switching vs Continuation | BMI | (Intercept) | 1.772 | 1.540 | 2.040 | 1.51e-15 | NA | 19319 | 12943 | 6376 |
| European | Recorded Anxiety | Switching vs Continuation | BMI | age_at_first_AD | 0.982 | 0.980 | 0.984 | <2.2e-16 | NA | 19319 | 12943 | 6376 |
| European | Recorded Anxiety | Switching vs Continuation | BMI | sex_at_birthMale | 1.201 | 1.121 | 1.286 | 2.08e-07 | NA | 19319 | 12943 | 6376 |
| European | Recorded Anxiety | Switching vs Continuation | BMI | Income_quantitative | 0.862 | 0.849 | 0.876 | <2.2e-16 | NA | 19319 | 12943 | 6376 |
| European | Recorded Anxiety | Switching vs Continuation | BMI | Education_levelCollege Undergraduate | 1.119 | 1.036 | 1.209 | 4.39e-03 | NA | 19319 | 12943 | 6376 |
| European | Recorded Anxiety | Switching vs Continuation | BMI | Education_levelLess than a high school degree | 1.256 | 0.996 | 1.583 | 5.43e-02 | NA | 19319 | 12943 | 6376 |
| European | Recorded Anxiety | Switching vs Continuation | BMI | Education_levelTwelve or GED | 1.456 | 1.302 | 1.628 | 4.21e-11 | NA | 19319 | 12943 | 6376 |
| European | Recorded Anxiety | Switching vs Continuation | BMI | BMI | 1.049 | 1.016 | 1.082 | 2.85e-03 | 1.99e-02 | 19319 | 12943 | 6376 |
| European | Recorded Anxiety | Switching vs Continuation | Bipolar Disorder | (Intercept) | 1.783 | 1.549 | 2.052 | 7.29e-16 | NA | 19319 | 12943 | 6376 |
| European | Recorded Anxiety | Switching vs Continuation | Bipolar Disorder | age_at_first_AD | 0.982 | 0.980 | 0.984 | <2.2e-16 | NA | 19319 | 12943 | 6376 |
| European | Recorded Anxiety | Switching vs Continuation | Bipolar Disorder | sex_at_birthMale | 1.198 | 1.118 | 1.284 | 2.79e-07 | NA | 19319 | 12943 | 6376 |
| European | Recorded Anxiety | Switching vs Continuation | Bipolar Disorder | Income_quantitative | 0.861 | 0.848 | 0.875 | <2.2e-16 | NA | 19319 | 12943 | 6376 |
| European | Recorded Anxiety | Switching vs Continuation | Bipolar Disorder | Education_levelCollege Undergraduate | 1.128 | 1.044 | 1.219 | 2.22e-03 | NA | 19319 | 12943 | 6376 |
| European | Recorded Anxiety | Switching vs Continuation | Bipolar Disorder | Education_levelLess than a high school degree | 1.276 | 1.012 | 1.609 | 3.93e-02 | NA | 19319 | 12943 | 6376 |
| European | Recorded Anxiety | Switching vs Continuation | Bipolar Disorder | Education_levelTwelve or GED | 1.478 | 1.322 | 1.652 | 6.47e-12 | NA | 19319 | 12943 | 6376 |
| European | Recorded Anxiety | Switching vs Continuation | Bipolar Disorder | Bipolar Disorder | 1.049 | 1.015 | 1.083 | 4.29e-03 | 3.01e-02 | 19319 | 12943 | 6376 |
| European | Recorded Anxiety | Switching vs Continuation | Depression | (Intercept) | 1.746 | 1.517 | 2.011 | 9.60e-15 | NA | 19319 | 12943 | 6376 |
| European | Recorded Anxiety | Switching vs Continuation | Depression | age_at_first_AD | 0.982 | 0.980 | 0.984 | <2.2e-16 | NA | 19319 | 12943 | 6376 |
| European | Recorded Anxiety | Switching vs Continuation | Depression | sex_at_birthMale | 1.191 | 1.111 | 1.276 | 7.38e-07 | NA | 19319 | 12943 | 6376 |
| European | Recorded Anxiety | Switching vs Continuation | Depression | Income_quantitative | 0.866 | 0.853 | 0.880 | <2.2e-16 | NA | 19319 | 12943 | 6376 |
| European | Recorded Anxiety | Switching vs Continuation | Depression | Education_levelCollege Undergraduate | 1.111 | 1.028 | 1.201 | 7.79e-03 | NA | 19319 | 12943 | 6376 |
| European | Recorded Anxiety | Switching vs Continuation | Depression | Education_levelLess than a high school degree | 1.205 | 0.955 | 1.520 | 1.17e-01 | NA | 19319 | 12943 | 6376 |
| European | Recorded Anxiety | Switching vs Continuation | Depression | Education_levelTwelve or GED | 1.447 | 1.294 | 1.619 | 9.08e-11 | NA | 19319 | 12943 | 6376 |
| European | Recorded Anxiety | Switching vs Continuation | Depression | Depression | 1.200 | 1.161 | 1.240 | <2.2e-16 | <2.2e-16 | 19319 | 12943 | 6376 |
| European | Recorded Anxiety | Switching vs Continuation | Insomnia | (Intercept) | 1.757 | 1.527 | 2.023 | 4.25e-15 | NA | 19319 | 12943 | 6376 |
| European | Recorded Anxiety | Switching vs Continuation | Insomnia | age_at_first_AD | 0.982 | 0.980 | 0.984 | <2.2e-16 | NA | 19319 | 12943 | 6376 |
| European | Recorded Anxiety | Switching vs Continuation | Insomnia | sex_at_birthMale | 1.199 | 1.119 | 1.285 | 2.55e-07 | NA | 19319 | 12943 | 6376 |
| European | Recorded Anxiety | Switching vs Continuation | Insomnia | Income_quantitative | 0.864 | 0.850 | 0.878 | <2.2e-16 | NA | 19319 | 12943 | 6376 |
| European | Recorded Anxiety | Switching vs Continuation | Insomnia | Education_levelCollege Undergraduate | 1.114 | 1.031 | 1.204 | 6.18e-03 | NA | 19319 | 12943 | 6376 |
| European | Recorded Anxiety | Switching vs Continuation | Insomnia | Education_levelLess than a high school degree | 1.217 | 0.965 | 1.535 | 9.75e-02 | NA | 19319 | 12943 | 6376 |
| European | Recorded Anxiety | Switching vs Continuation | Insomnia | Education_levelTwelve or GED | 1.446 | 1.293 | 1.617 | 9.70e-11 | NA | 19319 | 12943 | 6376 |
| European | Recorded Anxiety | Switching vs Continuation | Insomnia | Insomnia | 1.130 | 1.094 | 1.168 | 2.52e-13 | 1.76e-12 | 19319 | 12943 | 6376 |
| European | Recorded Anxiety | Switching vs Continuation | Migraine | (Intercept) | 1.781 | 1.548 | 2.050 | 8.35e-16 | NA | 19319 | 12943 | 6376 |
| European | Recorded Anxiety | Switching vs Continuation | Migraine | age_at_first_AD | 0.982 | 0.980 | 0.984 | <2.2e-16 | NA | 19319 | 12943 | 6376 |
| European | Recorded Anxiety | Switching vs Continuation | Migraine | sex_at_birthMale | 1.200 | 1.120 | 1.286 | 2.31e-07 | NA | 19319 | 12943 | 6376 |
| European | Recorded Anxiety | Switching vs Continuation | Migraine | Income_quantitative | 0.861 | 0.848 | 0.875 | <2.2e-16 | NA | 19319 | 12943 | 6376 |
| European | Recorded Anxiety | Switching vs Continuation | Migraine | Education_levelCollege Undergraduate | 1.125 | 1.041 | 1.215 | 2.93e-03 | NA | 19319 | 12943 | 6376 |
| European | Recorded Anxiety | Switching vs Continuation | Migraine | Education_levelLess than a high school degree | 1.272 | 1.009 | 1.603 | 4.20e-02 | NA | 19319 | 12943 | 6376 |
| European | Recorded Anxiety | Switching vs Continuation | Migraine | Education_levelTwelve or GED | 1.470 | 1.315 | 1.643 | 1.22e-11 | NA | 19319 | 12943 | 6376 |
| European | Recorded Anxiety | Switching vs Continuation | Migraine | Migraine | 1.049 | 1.017 | 1.082 | 2.69e-03 | 1.88e-02 | 19319 | 12943 | 6376 |
| European | Recorded Anxiety | Switching vs Continuation | Schizophrenia | (Intercept) | 1.781 | 1.548 | 2.050 | 8.30e-16 | NA | 19319 | 12943 | 6376 |
| European | Recorded Anxiety | Switching vs Continuation | Schizophrenia | age_at_first_AD | 0.982 | 0.980 | 0.984 | <2.2e-16 | NA | 19319 | 12943 | 6376 |
| European | Recorded Anxiety | Switching vs Continuation | Schizophrenia | sex_at_birthMale | 1.200 | 1.120 | 1.285 | 2.34e-07 | NA | 19319 | 12943 | 6376 |
| European | Recorded Anxiety | Switching vs Continuation | Schizophrenia | Income_quantitative | 0.861 | 0.848 | 0.875 | <2.2e-16 | NA | 19319 | 12943 | 6376 |
| European | Recorded Anxiety | Switching vs Continuation | Schizophrenia | Education_levelCollege Undergraduate | 1.127 | 1.043 | 1.217 | 2.53e-03 | NA | 19319 | 12943 | 6376 |
| European | Recorded Anxiety | Switching vs Continuation | Schizophrenia | Education_levelLess than a high school degree | 1.271 | 1.009 | 1.603 | 4.22e-02 | NA | 19319 | 12943 | 6376 |
| European | Recorded Anxiety | Switching vs Continuation | Schizophrenia | Education_levelTwelve or GED | 1.473 | 1.318 | 1.647 | 9.31e-12 | NA | 19319 | 12943 | 6376 |
| European | Recorded Anxiety | Switching vs Continuation | Schizophrenia | Schizophrenia | 1.019 | 0.988 | 1.052 | 2.30e-01 | 1.00e+00 | 19319 | 12943 | 6376 |
| European | CIDI-MDD | Switching vs Continuation | ADHD | (Intercept) | 2.286 | 1.664 | 3.141 | 3.34e-07 | NA | 3857 | 2568 | 1289 |
| European | CIDI-MDD | Switching vs Continuation | ADHD | age_at_first_AD | 0.977 | 0.972 | 0.981 | <2.2e-16 | NA | 3857 | 2568 | 1289 |
| European | CIDI-MDD | Switching vs Continuation | ADHD | sex_at_birthMale | 1.097 | 0.933 | 1.290 | 2.61e-01 | NA | 3857 | 2568 | 1289 |
| European | CIDI-MDD | Switching vs Continuation | ADHD | Income_quantitative | 0.880 | 0.850 | 0.912 | 9.53e-13 | NA | 3857 | 2568 | 1289 |

| | | | | | | | | | | | | |
|---|---|---|---|---|---|---|---|---|---|---|---|---|
| European | CIDI-MDD | Switching vs Continuation | ADHD | Education_levelCollege Undergraduate | 1.154 | 0.983 | 1.355 | 8.08e-02 | NA | 3857 | 2568 | 1289 |
| European | CIDI-MDD | Switching vs Continuation | ADHD | Education_levelLess than a high school degree | 0.833 | 0.366 | 1.899 | 6.64e-01 | NA | 3857 | 2568 | 1289 |
| European | CIDI-MDD | Switching vs Continuation | ADHD | Education_levelTwelve or GED | 1.475 | 1.104 | 1.970 | 8.55e-03 | NA | 3857 | 2568 | 1289 |
| European | CIDI-MDD | Switching vs Continuation | ADHD | ADHD | 1.065 | 0.991 | 1.144 | 8.47e-02 | 5.93e-01 | 3857 | 2568 | 1289 |
| European | CIDI-MDD | Switching vs Continuation | BMI | (Intercept) | 2.291 | 1.667 | 3.147 | 3.16e-07 | NA | 3857 | 2568 | 1289 |
| European | CIDI-MDD | Switching vs Continuation | BMI | age_at_first_AD | 0.977 | 0.972 | 0.981 | <2.2e-16 | NA | 3857 | 2568 | 1289 |
| European | CIDI-MDD | Switching vs Continuation | BMI | sex_at_birthMale | 1.095 | 0.931 | 1.287 | 2.72e-01 | NA | 3857 | 2568 | 1289 |
| European | CIDI-MDD | Switching vs Continuation | BMI | Income_quantitative | 0.878 | 0.848 | 0.910 | 4.52e-13 | NA | 3857 | 2568 | 1289 |
| European | CIDI-MDD | Switching vs Continuation | BMI | Education_levelCollege Undergraduate | 1.166 | 0.994 | 1.369 | 5.93e-02 | NA | 3857 | 2568 | 1289 |
| European | CIDI-MDD | Switching vs Continuation | BMI | Education_levelLess than a high school degree | 0.852 | 0.374 | 1.943 | 7.04e-01 | NA | 3857 | 2568 | 1289 |
| European | CIDI-MDD | Switching vs Continuation | BMI | Education_levelTwelve or GED | 1.501 | 1.124 | 2.004 | 5.85e-03 | NA | 3857 | 2568 | 1289 |
| European | CIDI-MDD | Switching vs Continuation | BMI | BMI | 1.014 | 0.944 | 1.088 | 7.04e-01 | 1.00e+00 | 3857 | 2568 | 1289 |
| European | CIDI-MDD | Switching vs Continuation | Bipolar Disorder | (Intercept) | 2.307 | 1.679 | 3.169 | 2.49e-07 | NA | 3857 | 2568 | 1289 |
| European | CIDI-MDD | Switching vs Continuation | Bipolar Disorder | age_at_first_AD | 0.976 | 0.972 | 0.981 | <2.2e-16 | NA | 3857 | 2568 | 1289 |
| European | CIDI-MDD | Switching vs Continuation | Bipolar Disorder | sex_at_birthMale | 1.096 | 0.933 | 1.288 | 2.66e-01 | NA | 3857 | 2568 | 1289 |
| European | CIDI-MDD | Switching vs Continuation | Bipolar Disorder | Income_quantitative | 0.878 | 0.848 | 0.909 | 2.91e-13 | NA | 3857 | 2568 | 1289 |
| European | CIDI-MDD | Switching vs Continuation | Bipolar Disorder | Education_levelCollege Undergraduate | 1.171 | 0.998 | 1.375 | 5.27e-02 | NA | 3857 | 2568 | 1289 |
| European | CIDI-MDD | Switching vs Continuation | Bipolar Disorder | Education_levelLess than a high school degree | 0.886 | 0.388 | 2.021 | 7.74e-01 | NA | 3857 | 2568 | 1289 |
| European | CIDI-MDD | Switching vs Continuation | Bipolar Disorder | Education_levelTwelve or GED | 1.519 | 1.138 | 2.028 | 4.55e-03 | NA | 3857 | 2568 | 1289 |
| European | CIDI-MDD | Switching vs Continuation | Bipolar Disorder | Bipolar Disorder | 1.076 | 1.000 | 1.159 | 5.07e-02 | 3.55e-01 | 3857 | 2568 | 1289 |
| European | CIDI-MDD | Switching vs Continuation | Depression | (Intercept) | 2.302 | 1.675 | 3.164 | 2.73e-07 | NA | 3857 | 2568 | 1289 |
| European | CIDI-MDD | Switching vs Continuation | Depression | age_at_first_AD | 0.977 | 0.972 | 0.981 | <2.2e-16 | NA | 3857 | 2568 | 1289 |
| European | CIDI-MDD | Switching vs Continuation | Depression | sex_at_birthMale | 1.094 | 0.930 | 1.286 | 2.78e-01 | NA | 3857 | 2568 | 1289 |
| European | CIDI-MDD | Switching vs Continuation | Depression | Income_quantitative | 0.880 | 0.850 | 0.912 | 9.41e-13 | NA | 3857 | 2568 | 1289 |
| European | CIDI-MDD | Switching vs Continuation | Depression | Education_levelCollege Undergraduate | 1.150 | 0.979 | 1.350 | 8.86e-02 | NA | 3857 | 2568 | 1289 |
| European | CIDI-MDD | Switching vs Continuation | Depression | Education_levelLess than a high school degree | 0.845 | 0.370 | 1.927 | 6.88e-01 | NA | 3857 | 2568 | 1289 |
| European | CIDI-MDD | Switching vs Continuation | Depression | Education_levelTwelve or GED | 1.489 | 1.115 | 1.988 | 7.00e-03 | NA | 3857 | 2568 | 1289 |
| European | CIDI-MDD | Switching vs Continuation | Depression | Depression | 1.117 | 1.039 | 1.202 | 2.94e-03 | 2.06e-02 | 3857 | 2568 | 1289 |
| European | CIDI-MDD | Switching vs Continuation | Insomnia | (Intercept) | 2.297 | 1.672 | 3.155 | 2.81e-07 | NA | 3857 | 2568 | 1289 |
| European | CIDI-MDD | Switching vs Continuation | Insomnia | age_at_first_AD | 0.977 | 0.972 | 0.981 | <2.2e-16 | NA | 3857 | 2568 | 1289 |
| European | CIDI-MDD | Switching vs Continuation | Insomnia | sex_at_birthMale | 1.095 | 0.932 | 1.287 | 2.70e-01 | NA | 3857 | 2568 | 1289 |
| European | CIDI-MDD | Switching vs Continuation | Insomnia | Income_quantitative | 0.878 | 0.848 | 0.909 | 3.00e-13 | NA | 3857 | 2568 | 1289 |
| European | CIDI-MDD | Switching vs Continuation | Insomnia | Education_levelCollege Undergraduate | 1.169 | 0.996 | 1.372 | 5.64e-02 | NA | 3857 | 2568 | 1289 |
| European | CIDI-MDD | Switching vs Continuation | Insomnia | Education_levelLess than a high school degree | 0.857 | 0.376 | 1.953 | 7.13e-01 | NA | 3857 | 2568 | 1289 |
| European | CIDI-MDD | Switching vs Continuation | Insomnia | Education_levelTwelve or GED | 1.507 | 1.129 | 2.011 | 5.37e-03 | NA | 3857 | 2568 | 1289 |
| European | CIDI-MDD | Switching vs Continuation | Insomnia | Insomnia | 0.992 | 0.922 | 1.067 | 8.23e-01 | 1.00e+00 | 3857 | 2568 | 1289 |
| European | CIDI-MDD | Switching vs Continuation | Migraine | (Intercept) | 2.296 | 1.671 | 3.153 | 2.87e-07 | NA | 3857 | 2568 | 1289 |
| European | CIDI-MDD | Switching vs Continuation | Migraine | age_at_first_AD | 0.977 | 0.972 | 0.981 | <2.2e-16 | NA | 3857 | 2568 | 1289 |
| European | CIDI-MDD | Switching vs Continuation | Migraine | sex_at_birthMale | 1.095 | 0.932 | 1.287 | 2.70e-01 | NA | 3857 | 2568 | 1289 |
| European | CIDI-MDD | Switching vs Continuation | Migraine | Income_quantitative | 0.878 | 0.848 | 0.909 | 3.26e-13 | NA | 3857 | 2568 | 1289 |
| European | CIDI-MDD | Switching vs Continuation | Migraine | Education_levelCollege Undergraduate | 1.167 | 0.995 | 1.370 | 5.81e-02 | NA | 3857 | 2568 | 1289 |
| European | CIDI-MDD | Switching vs Continuation | Migraine | Education_levelLess than a high school degree | 0.854 | 0.375 | 1.947 | 7.08e-01 | NA | 3857 | 2568 | 1289 |
| European | CIDI-MDD | Switching vs Continuation | Migraine | Education_levelTwelve or GED | 1.504 | 1.127 | 2.007 | 5.57e-03 | NA | 3857 | 2568 | 1289 |
| European | CIDI-MDD | Switching vs Continuation | Migraine | Migraine | 1.008 | 0.941 | 1.080 | 8.19e-01 | 1.00e+00 | 3857 | 2568 | 1289 |
| European | CIDI-MDD | Switching vs Continuation | Schizophrenia | (Intercept) | 2.294 | 1.670 | 3.152 | 2.96e-07 | NA | 3857 | 2568 | 1289 |
| European | CIDI-MDD | Switching vs Continuation | Schizophrenia | age_at_first_AD | 0.977 | 0.972 | 0.981 | <2.2e-16 | NA | 3857 | 2568 | 1289 |
| European | CIDI-MDD | Switching vs Continuation | Schizophrenia | sex_at_birthMale | 1.101 | 0.937 | 1.294 | 2.44e-01 | NA | 3857 | 2568 | 1289 |
| European | CIDI-MDD | Switching vs Continuation | Schizophrenia | Income_quantitative | 0.878 | 0.848 | 0.910 | 3.56e-13 | NA | 3857 | 2568 | 1289 |
| European | CIDI-MDD | Switching vs Continuation | Schizophrenia | Education_levelCollege Undergraduate | 1.167 | 0.995 | 1.370 | 5.82e-02 | NA | 3857 | 2568 | 1289 |
| European | CIDI-MDD | Switching vs Continuation | Schizophrenia | Education_levelLess than a high school degree | 0.869 | 0.381 | 1.981 | 7.38e-01 | NA | 3857 | 2568 | 1289 |
| European | CIDI-MDD | Switching vs Continuation | Schizophrenia | Education_levelTwelve or GED | 1.513 | 1.133 | 2.019 | 4.96e-03 | NA | 3857 | 2568 | 1289 |
| European | CIDI-MDD | Switching vs Continuation | Schizophrenia | Schizophrenia | 1.070 | 0.997 | 1.149 | 6.24e-02 | 4.37e-01 | 3857 | 2568 | 1289 |
| European | CIDI-noMDD | Switching vs Continuation | ADHD | (Intercept) | 1.096 | 0.609 | 1.973 | 7.60e-01 | NA | 2467 | 2114 | 353 |
| European | CIDI-noMDD | Switching vs Continuation | ADHD | age_at_first_AD | 0.971 | 0.963 | 0.979 | 1.65e-12 | NA | 2467 | 2114 | 353 |
| European | CIDI-noMDD | Switching vs Continuation | ADHD | sex_at_birthMale | 1.316 | 1.025 | 1.689 | 3.13e-02 | NA | 2467 | 2114 | 353 |
| European | CIDI-noMDD | Switching vs Continuation | ADHD | Income_quantitative | 0.895 | 0.841 | 0.952 | 4.05e-04 | NA | 2467 | 2114 | 353 |
| European | CIDI-noMDD | Switching vs Continuation | ADHD | Education_levelCollege Undergraduate | 1.166 | 0.889 | 1.528 | 2.66e-01 | NA | 2467 | 2114 | 353 |
| European | CIDI-noMDD | Switching vs Continuation | ADHD | Education_levelLess than a high school degree | 2.190 | 0.623 | 7.696 | 2.22e-01 | NA | 2467 | 2114 | 353 |
| European | CIDI-noMDD | Switching vs Continuation | ADHD | Education_levelTwelve or GED | 1.344 | 0.872 | 2.071 | 1.80e-01 | NA | 2467 | 2114 | 353 |
| European | CIDI-noMDD | Switching vs Continuation | ADHD | ADHD | 1.127 | 1.001 | 1.269 | 4.78e-02 | 3.35e-01 | 2467 | 2114 | 353 |

| | | | | | | | | | | | | |
|---|---|---|---|---|---|---|---|---|---|---|---|---|
| European | CIDI-noMDD | Switching vs Continuation | BMI | (Intercept) | 1.093 | 0.607 | 1.967 | 7.67e-01 | NA | 2467 | 2114 | 353 |
| European | CIDI-noMDD | Switching vs Continuation | BMI | age_at_first_AD | 0.971 | 0.963 | 0.979 | 1.50e-12 | NA | 2467 | 2114 | 353 |
| European | CIDI-noMDD | Switching vs Continuation | BMI | sex_at_birthMale | 1.311 | 1.021 | 1.683 | 3.36e-02 | NA | 2467 | 2114 | 353 |
| European | CIDI-noMDD | Switching vs Continuation | BMI | Income_quantitative | 0.895 | 0.841 | 0.952 | 4.12e-04 | NA | 2467 | 2114 | 353 |
| European | CIDI-noMDD | Switching vs Continuation | BMI | Education_levelCollege Undergraduate | 1.182 | 0.902 | 1.548 | 2.25e-01 | NA | 2467 | 2114 | 353 |
| European | CIDI-noMDD | Switching vs Continuation | BMI | Education_levelLess than a high school degree | 2.317 | 0.661 | 8.122 | 1.89e-01 | NA | 2467 | 2114 | 353 |
| European | CIDI-noMDD | Switching vs Continuation | BMI | Education_levelTwelve or GED | 1.370 | 0.890 | 2.108 | 1.53e-01 | NA | 2467 | 2114 | 353 |
| European | CIDI-noMDD | Switching vs Continuation | BMI | BMI | 1.103 | 0.982 | 1.238 | 9.77e-02 | 6.84e-01 | 2467 | 2114 | 353 |
| European | CIDI-noMDD | Switching vs Continuation | Bipolar Disorder | (Intercept) | 1.106 | 0.615 | 1.991 | 7.36e-01 | NA | 2467 | 2114 | 353 |
| European | CIDI-noMDD | Switching vs Continuation | Bipolar Disorder | age_at_first_AD | 0.971 | 0.963 | 0.979 | 7.07e-13 | NA | 2467 | 2114 | 353 |
| European | CIDI-noMDD | Switching vs Continuation | Bipolar Disorder | sex_at_birthMale | 1.303 | 1.015 | 1.672 | 3.82e-02 | NA | 2467 | 2114 | 353 |
| European | CIDI-noMDD | Switching vs Continuation | Bipolar Disorder | Income_quantitative | 0.895 | 0.841 | 0.952 | 4.03e-04 | NA | 2467 | 2114 | 353 |
| European | CIDI-noMDD | Switching vs Continuation | Bipolar Disorder | Education_levelCollege Undergraduate | 1.193 | 0.911 | 1.563 | 2.00e-01 | NA | 2467 | 2114 | 353 |
| European | CIDI-noMDD | Switching vs Continuation | Bipolar Disorder | Education_levelLess than a high school degree | 2.315 | 0.656 | 8.172 | 1.92e-01 | NA | 2467 | 2114 | 353 |
| European | CIDI-noMDD | Switching vs Continuation | Bipolar Disorder | Education_levelTwelve or GED | 1.426 | 0.925 | 2.198 | 1.08e-01 | NA | 2467 | 2114 | 353 |
| European | CIDI-noMDD | Switching vs Continuation | Bipolar Disorder | Bipolar Disorder | 1.166 | 1.031 | 1.319 | 1.46e-02 | 1.02e-01 | 2467 | 2114 | 353 |
| European | CIDI-noMDD | Switching vs Continuation | Depression | (Intercept) | 1.093 | 0.607 | 1.969 | 7.67e-01 | NA | 2467 | 2114 | 353 |
| European | CIDI-noMDD | Switching vs Continuation | Depression | age_at_first_AD | 0.971 | 0.963 | 0.979 | 2.08e-12 | NA | 2467 | 2114 | 353 |
| European | CIDI-noMDD | Switching vs Continuation | Depression | sex_at_birthMale | 1.290 | 1.005 | 1.657 | 4.56e-02 | NA | 2467 | 2114 | 353 |
| European | CIDI-noMDD | Switching vs Continuation | Depression | Income_quantitative | 0.895 | 0.841 | 0.952 | 3.97e-04 | NA | 2467 | 2114 | 353 |
| European | CIDI-noMDD | Switching vs Continuation | Depression | Education_levelCollege Undergraduate | 1.171 | 0.894 | 1.535 | 2.52e-01 | NA | 2467 | 2114 | 353 |
| European | CIDI-noMDD | Switching vs Continuation | Depression | Education_levelLess than a high school degree | 2.127 | 0.597 | 7.575 | 2.44e-01 | NA | 2467 | 2114 | 353 |
| European | CIDI-noMDD | Switching vs Continuation | Depression | Education_levelTwelve or GED | 1.329 | 0.862 | 2.049 | 1.97e-01 | NA | 2467 | 2114 | 353 |
| European | CIDI-noMDD | Switching vs Continuation | Depression | Depression | 1.151 | 1.019 | 1.301 | 2.38e-02 | 1.66e-01 | 2467 | 2114 | 353 |
| European | CIDI-noMDD | Switching vs Continuation | Insomnia | (Intercept) | 1.071 | 0.594 | 1.933 | 8.19e-01 | NA | 2467 | 2114 | 353 |
| European | CIDI-noMDD | Switching vs Continuation | Insomnia | age_at_first_AD | 0.971 | 0.963 | 0.979 | 1.76e-12 | NA | 2467 | 2114 | 353 |
| European | CIDI-noMDD | Switching vs Continuation | Insomnia | sex_at_birthMale | 1.314 | 1.023 | 1.687 | 3.25e-02 | NA | 2467 | 2114 | 353 |
| European | CIDI-noMDD | Switching vs Continuation | Insomnia | Income_quantitative | 0.898 | 0.844 | 0.955 | 6.31e-04 | NA | 2467 | 2114 | 353 |
| European | CIDI-noMDD | Switching vs Continuation | Insomnia | Education_levelCollege Undergraduate | 1.169 | 0.892 | 1.533 | 2.59e-01 | NA | 2467 | 2114 | 353 |
| European | CIDI-noMDD | Switching vs Continuation | Insomnia | Education_levelLess than a high school degree | 2.266 | 0.645 | 7.960 | 2.02e-01 | NA | 2467 | 2114 | 353 |
| European | CIDI-noMDD | Switching vs Continuation | Insomnia | Education_levelTwelve or GED | 1.353 | 0.877 | 2.085 | 1.72e-01 | NA | 2467 | 2114 | 353 |
| European | CIDI-noMDD | Switching vs Continuation | Insomnia | Insomnia | 1.208 | 1.069 | 1.365 | 2.40e-03 | 1.68e-02 | 2467 | 2114 | 353 |
| European | CIDI-noMDD | Switching vs Continuation | Migraine | (Intercept) | 1.129 | 0.628 | 2.030 | 6.84e-01 | NA | 2467 | 2114 | 353 |
| European | CIDI-noMDD | Switching vs Continuation | Migraine | age_at_first_AD | 0.971 | 0.963 | 0.979 | 1.02e-12 | NA | 2467 | 2114 | 353 |
| European | CIDI-noMDD | Switching vs Continuation | Migraine | sex_at_birthMale | 1.307 | 1.018 | 1.677 | 3.55e-02 | NA | 2467 | 2114 | 353 |
| European | CIDI-noMDD | Switching vs Continuation | Migraine | Income_quantitative | 0.890 | 0.837 | 0.947 | 2.16e-04 | NA | 2467 | 2114 | 353 |
| European | CIDI-noMDD | Switching vs Continuation | Migraine | Education_levelCollege Undergraduate | 1.191 | 0.909 | 1.560 | 2.05e-01 | NA | 2467 | 2114 | 353 |
| European | CIDI-noMDD | Switching vs Continuation | Migraine | Education_levelLess than a high school degree | 2.391 | 0.682 | 8.385 | 1.73e-01 | NA | 2467 | 2114 | 353 |
| European | CIDI-noMDD | Switching vs Continuation | Migraine | Education_levelTwelve or GED | 1.382 | 0.898 | 2.127 | 1.41e-01 | NA | 2467 | 2114 | 353 |
| European | CIDI-noMDD | Switching vs Continuation | Migraine | Migraine | 0.973 | 0.870 | 1.088 | 6.29e-01 | 1.00e+00 | 2467 | 2114 | 353 |
| European | CIDI-noMDD | Switching vs Continuation | Schizophrenia | (Intercept) | 1.113 | 0.619 | 2.003 | 7.20e-01 | NA | 2467 | 2114 | 353 |
| European | CIDI-noMDD | Switching vs Continuation | Schizophrenia | age_at_first_AD | 0.971 | 0.963 | 0.979 | 1.29e-12 | NA | 2467 | 2114 | 353 |
| European | CIDI-noMDD | Switching vs Continuation | Schizophrenia | sex_at_birthMale | 1.310 | 1.021 | 1.682 | 3.39e-02 | NA | 2467 | 2114 | 353 |
| European | CIDI-noMDD | Switching vs Continuation | Schizophrenia | Income_quantitative | 0.891 | 0.838 | 0.948 | 2.49e-04 | NA | 2467 | 2114 | 353 |
| European | CIDI-noMDD | Switching vs Continuation | Schizophrenia | Education_levelCollege Undergraduate | 1.188 | 0.907 | 1.556 | 2.11e-01 | NA | 2467 | 2114 | 353 |
| European | CIDI-noMDD | Switching vs Continuation | Schizophrenia | Education_levelLess than a high school degree | 2.356 | 0.671 | 8.278 | 1.81e-01 | NA | 2467 | 2114 | 353 |
| European | CIDI-noMDD | Switching vs Continuation | Schizophrenia | Education_levelTwelve or GED | 1.382 | 0.898 | 2.127 | 1.41e-01 | NA | 2467 | 2114 | 353 |
| European | CIDI-noMDD | Switching vs Continuation | Schizophrenia | Schizophrenia | 1.093 | 0.971 | 1.231 | 1.40e-01 | 9.81e-01 | 2467 | 2114 | 353 |
| African | Full Cohort | Switching vs Continuation | ADHD | (Intercept) | 1.018 | 0.735 | 1.412 | 9.13e-01 | NA | 6074 | 4493 | 1581 |
| African | Full Cohort | Switching vs Continuation | ADHD | age_at_first_AD | 0.983 | 0.979 | 0.987 | 1.26e-14 | NA | 6074 | 4493 | 1581 |
| African | Full Cohort | Switching vs Continuation | ADHD | sex_at_birthMale | 1.001 | 0.874 | 1.146 | 9.90e-01 | NA | 6074 | 4493 | 1581 |
| African | Full Cohort | Switching vs Continuation | ADHD | Income_quantitative | 0.907 | 0.870 | 0.945 | 3.60e-06 | NA | 6074 | 4493 | 1581 |
| African | Full Cohort | Switching vs Continuation | ADHD | Education_levelCollege Undergraduate | 0.968 | 0.779 | 1.204 | 7.71e-01 | NA | 6074 | 4493 | 1581 |
| African | Full Cohort | Switching vs Continuation | ADHD | Education_levelLess than a high school degree | 0.991 | 0.746 | 1.316 | 9.50e-01 | NA | 6074 | 4493 | 1581 |
| African | Full Cohort | Switching vs Continuation | ADHD | Education_levelTwelve or GED | 0.873 | 0.684 | 1.114 | 2.74e-01 | NA | 6074 | 4493 | 1581 |
| African | Full Cohort | Switching vs Continuation | ADHD | ADHD | 1.024 | 0.944 | 1.112 | 5.67e-01 | 1.00e+00 | 6074 | 4493 | 1581 |
| African | Full Cohort | Switching vs Continuation | BMI | (Intercept) | 1.019 | 0.735 | 1.412 | 9.11e-01 | NA | 6074 | 4493 | 1581 |
| African | Full Cohort | Switching vs Continuation | BMI | age_at_first_AD | 0.983 | 0.979 | 0.987 | 1.16e-14 | NA | 6074 | 4493 | 1581 |
| African | Full Cohort | Switching vs Continuation | BMI | sex_at_birthMale | 1.000 | 0.873 | 1.146 | 9.96e-01 | NA | 6074 | 4493 | 1581 |
| African | Full Cohort | Switching vs Continuation | BMI | Income_quantitative | 0.906 | 0.870 | 0.945 | 3.29e-06 | NA | 6074 | 4493 | 1581 |

| | | | | | | | | | | | | |
|---|---|---|---|---|---|---|---|---|---|---|---|---|
| African | Full Cohort | Switching vs Continuation | BMI | Education_levelCollege Undergraduate | 0.970 | 0.780 | 1.206 | 7.85e-01 | NA | 6074 | 4493 | 1581 |
| African | Full Cohort | Switching vs Continuation | BMI | Education_levelLess than a high school degree | 0.995 | 0.750 | 1.322 | 9.74e-01 | NA | 6074 | 4493 | 1581 |
| African | Full Cohort | Switching vs Continuation | BMI | Education_levelTwelve or GED | 0.875 | 0.686 | 1.116 | 2.83e-01 | NA | 6074 | 4493 | 1581 |
| African | Full Cohort | Switching vs Continuation | BMI | BMI | 0.996 | 0.923 | 1.075 | 9.17e-01 | 1.00e+00 | 6074 | 4493 | 1581 |
| African | Full Cohort | Switching vs Continuation | Bipolar Disorder | (Intercept) | 1.015 | 0.732 | 1.408 | 9.28e-01 | NA | 6074 | 4493 | 1581 |
| African | Full Cohort | Switching vs Continuation | Bipolar Disorder | age_at_first_AD | 0.983 | 0.979 | 0.987 | 8.88e-15 | NA | 6074 | 4493 | 1581 |
| African | Full Cohort | Switching vs Continuation | Bipolar Disorder | sex_at_birthMale | 0.999 | 0.872 | 1.144 | 9.86e-01 | NA | 6074 | 4493 | 1581 |
| African | Full Cohort | Switching vs Continuation | Bipolar Disorder | Income_quantitative | 0.907 | 0.870 | 0.946 | 4.06e-06 | NA | 6074 | 4493 | 1581 |
| African | Full Cohort | Switching vs Continuation | Bipolar Disorder | Education_levelCollege Undergraduate | 0.975 | 0.784 | 1.212 | 8.16e-01 | NA | 6074 | 4493 | 1581 |
| African | Full Cohort | Switching vs Continuation | Bipolar Disorder | Education_levelLess than a high school degree | 1.000 | 0.753 | 1.327 | 9.97e-01 | NA | 6074 | 4493 | 1581 |
| African | Full Cohort | Switching vs Continuation | Bipolar Disorder | Education_levelTwelve or GED | 0.882 | 0.692 | 1.126 | 3.14e-01 | NA | 6074 | 4493 | 1581 |
| African | Full Cohort | Switching vs Continuation | Bipolar Disorder | Bipolar Disorder | 1.101 | 1.012 | 1.199 | 2.58e-02 | 1.80e-01 | 6074 | 4493 | 1581 |
| African | Full Cohort | Switching vs Continuation | Depression | (Intercept) | 1.010 | 0.728 | 1.400 | 9.53e-01 | NA | 6074 | 4493 | 1581 |
| African | Full Cohort | Switching vs Continuation | Depression | age_at_first_AD | 0.983 | 0.979 | 0.987 | 1.07e-14 | NA | 6074 | 4493 | 1581 |
| African | Full Cohort | Switching vs Continuation | Depression | sex_at_birthMale | 1.000 | 0.873 | 1.146 | 9.98e-01 | NA | 6074 | 4493 | 1581 |
| African | Full Cohort | Switching vs Continuation | Depression | Income_quantitative | 0.909 | 0.872 | 0.947 | 5.99e-06 | NA | 6074 | 4493 | 1581 |
| African | Full Cohort | Switching vs Continuation | Depression | Education_levelCollege Undergraduate | 0.971 | 0.781 | 1.207 | 7.90e-01 | NA | 6074 | 4493 | 1581 |
| African | Full Cohort | Switching vs Continuation | Depression | Education_levelLess than a high school degree | 0.997 | 0.751 | 1.324 | 9.83e-01 | NA | 6074 | 4493 | 1581 |
| African | Full Cohort | Switching vs Continuation | Depression | Education_levelTwelve or GED | 0.879 | 0.689 | 1.121 | 2.97e-01 | NA | 6074 | 4493 | 1581 |
| African | Full Cohort | Switching vs Continuation | Depression | Depression | 1.133 | 1.044 | 1.229 | 2.71e-03 | 1.90e-02 | 6074 | 4493 | 1581 |
| African | Full Cohort | Switching vs Continuation | Insomnia | (Intercept) | 1.025 | 0.739 | 1.421 | 8.83e-01 | NA | 6074 | 4493 | 1581 |
| African | Full Cohort | Switching vs Continuation | Insomnia | age_at_first_AD | 0.983 | 0.979 | 0.987 | 9.73e-15 | NA | 6074 | 4493 | 1581 |
| African | Full Cohort | Switching vs Continuation | Insomnia | sex_at_birthMale | 1.000 | 0.873 | 1.145 | 9.99e-01 | NA | 6074 | 4493 | 1581 |
| African | Full Cohort | Switching vs Continuation | Insomnia | Income_quantitative | 0.907 | 0.870 | 0.945 | 3.63e-06 | NA | 6074 | 4493 | 1581 |
| African | Full Cohort | Switching vs Continuation | Insomnia | Education_levelCollege Undergraduate | 0.965 | 0.776 | 1.200 | 7.48e-01 | NA | 6074 | 4493 | 1581 |
| African | Full Cohort | Switching vs Continuation | Insomnia | Education_levelLess than a high school degree | 0.983 | 0.740 | 1.305 | 9.03e-01 | NA | 6074 | 4493 | 1581 |
| African | Full Cohort | Switching vs Continuation | Insomnia | Education_levelTwelve or GED | 0.872 | 0.683 | 1.112 | 2.69e-01 | NA | 6074 | 4493 | 1581 |
| African | Full Cohort | Switching vs Continuation | Insomnia | Insomnia | 1.140 | 1.047 | 1.240 | 2.38e-03 | 1.66e-02 | 6074 | 4493 | 1581 |
| African | Full Cohort | Switching vs Continuation | Migraine | (Intercept) | 1.018 | 0.735 | 1.412 | 9.13e-01 | NA | 6074 | 4493 | 1581 |
| African | Full Cohort | Switching vs Continuation | Migraine | age_at_first_AD | 0.983 | 0.979 | 0.987 | 1.22e-14 | NA | 6074 | 4493 | 1581 |
| African | Full Cohort | Switching vs Continuation | Migraine | sex_at_birthMale | 1.001 | 0.874 | 1.146 | 9.90e-01 | NA | 6074 | 4493 | 1581 |
| African | Full Cohort | Switching vs Continuation | Migraine | Income_quantitative | 0.906 | 0.870 | 0.945 | 3.22e-06 | NA | 6074 | 4493 | 1581 |
| African | Full Cohort | Switching vs Continuation | Migraine | Education_levelCollege Undergraduate | 0.969 | 0.780 | 1.205 | 7.77e-01 | NA | 6074 | 4493 | 1581 |
| African | Full Cohort | Switching vs Continuation | Migraine | Education_levelLess than a high school degree | 0.994 | 0.749 | 1.319 | 9.66e-01 | NA | 6074 | 4493 | 1581 |
| African | Full Cohort | Switching vs Continuation | Migraine | Education_levelTwelve or GED | 0.874 | 0.686 | 1.115 | 2.79e-01 | NA | 6074 | 4493 | 1581 |
| African | Full Cohort | Switching vs Continuation | Migraine | Migraine | 0.964 | 0.898 | 1.034 | 3.04e-01 | 1.00e+00 | 6074 | 4493 | 1581 |
| African | Full Cohort | Switching vs Continuation | Schizophrenia | (Intercept) | 1.019 | 0.735 | 1.413 | 9.08e-01 | NA | 6074 | 4493 | 1581 |
| African | Full Cohort | Switching vs Continuation | Schizophrenia | age_at_first_AD | 0.983 | 0.979 | 0.987 | 1.19e-14 | NA | 6074 | 4493 | 1581 |
| African | Full Cohort | Switching vs Continuation | Schizophrenia | sex_at_birthMale | 1.001 | 0.874 | 1.146 | 9.90e-01 | NA | 6074 | 4493 | 1581 |
| African | Full Cohort | Switching vs Continuation | Schizophrenia | Income_quantitative | 0.906 | 0.869 | 0.944 | 3.08e-06 | NA | 6074 | 4493 | 1581 |
| African | Full Cohort | Switching vs Continuation | Schizophrenia | Education_levelCollege Undergraduate | 0.969 | 0.779 | 1.205 | 7.78e-01 | NA | 6074 | 4493 | 1581 |
| African | Full Cohort | Switching vs Continuation | Schizophrenia | Education_levelLess than a high school degree | 0.995 | 0.749 | 1.321 | 9.70e-01 | NA | 6074 | 4493 | 1581 |
| African | Full Cohort | Switching vs Continuation | Schizophrenia | Education_levelTwelve or GED | 0.874 | 0.685 | 1.115 | 2.78e-01 | NA | 6074 | 4493 | 1581 |
| African | Full Cohort | Switching vs Continuation | Schizophrenia | Schizophrenia | 0.980 | 0.910 | 1.055 | 5.92e-01 | 1.00e+00 | 6074 | 4493 | 1581 |
| African | Recorded Depression | Switching vs Continuation | ADHD | (Intercept) | 1.310 | 0.864 | 1.988 | 2.03e-01 | NA | 3124 | 1930 | 1194 |
| African | Recorded Depression | Switching vs Continuation | ADHD | age_at_first_AD | 0.989 | 0.983 | 0.994 | 6.67e-05 | NA | 3124 | 1930 | 1194 |
| African | Recorded Depression | Switching vs Continuation | ADHD | sex_at_birthMale | 1.060 | 0.891 | 1.260 | 5.13e-01 | NA | 3124 | 1930 | 1194 |
| African | Recorded Depression | Switching vs Continuation | ADHD | Income_quantitative | 0.911 | 0.864 | 0.960 | 4.68e-04 | NA | 3124 | 1930 | 1194 |
| African | Recorded Depression | Switching vs Continuation | ADHD | Education_levelCollege Undergraduate | 0.975 | 0.736 | 1.291 | 8.61e-01 | NA | 3124 | 1930 | 1194 |
| African | Recorded Depression | Switching vs Continuation | ADHD | Education_levelLess than a high school degree | 0.970 | 0.670 | 1.403 | 8.70e-01 | NA | 3124 | 1930 | 1194 |
| African | Recorded Depression | Switching vs Continuation | ADHD | Education_levelTwelve or GED | 0.864 | 0.633 | 1.179 | 3.56e-01 | NA | 3124 | 1930 | 1194 |
| African | Recorded Depression | Switching vs Continuation | ADHD | ADHD | 1.032 | 0.930 | 1.145 | 5.52e-01 | 1.00e+00 | 3124 | 1930 | 1194 |
| African | Recorded Depression | Switching vs Continuation | BMI | (Intercept) | 1.310 | 0.864 | 1.987 | 2.04e-01 | NA | 3124 | 1930 | 1194 |
| African | Recorded Depression | Switching vs Continuation | BMI | age_at_first_AD | 0.989 | 0.983 | 0.994 | 5.77e-05 | NA | 3124 | 1930 | 1194 |
| African | Recorded Depression | Switching vs Continuation | BMI | sex_at_birthMale | 1.057 | 0.888 | 1.257 | 5.33e-01 | NA | 3124 | 1930 | 1194 |
| African | Recorded Depression | Switching vs Continuation | BMI | Income_quantitative | 0.910 | 0.864 | 0.959 | 4.42e-04 | NA | 3124 | 1930 | 1194 |
| African | Recorded Depression | Switching vs Continuation | BMI | Education_levelCollege Undergraduate | 0.982 | 0.742 | 1.301 | 9.01e-01 | NA | 3124 | 1930 | 1194 |
| African | Recorded Depression | Switching vs Continuation | BMI | Education_levelLess than a high school degree | 0.981 | 0.679 | 1.418 | 9.19e-01 | NA | 3124 | 1930 | 1194 |
| African | Recorded Depression | Switching vs Continuation | BMI | Education_levelTwelve or GED | 0.872 | 0.639 | 1.189 | 3.86e-01 | NA | 3124 | 1930 | 1194 |
| African | Recorded Depression | Switching vs Continuation | BMI | BMI | 0.971 | 0.882 | 1.068 | 5.44e-01 | 1.00e+00 | 3124 | 1930 | 1194 |

| | | | | | | | | | | | | |
|---|---|---|---|---|---|---|---|---|---|---|---|---|
| African | Recorded Depression | Switching vs Continuation | Bipolar Disorder | (Intercept) | 1.300 | 0.857 | 1.972 | 2.17e-01 | NA | 3124 | 1930 | 1194 |
| African | Recorded Depression | Switching vs Continuation | Bipolar Disorder | age_at_first_AD | 0.989 | 0.983 | 0.994 | 5.67e-05 | NA | 3124 | 1930 | 1194 |
| African | Recorded Depression | Switching vs Continuation | Bipolar Disorder | sex_at_birthMale | 1.053 | 0.885 | 1.252 | 5.63e-01 | NA | 3124 | 1930 | 1194 |
| African | Recorded Depression | Switching vs Continuation | Bipolar Disorder | Income_quantitative | 0.911 | 0.865 | 0.960 | 5.17e-04 | NA | 3124 | 1930 | 1194 |
| African | Recorded Depression | Switching vs Continuation | Bipolar Disorder | Education_levelCollege Undergraduate | 0.989 | 0.747 | 1.309 | 9.37e-01 | NA | 3124 | 1930 | 1194 |
| African | Recorded Depression | Switching vs Continuation | Bipolar Disorder | Education_levelLess than a high school degree | 0.985 | 0.681 | 1.423 | 9.34e-01 | NA | 3124 | 1930 | 1194 |
| African | Recorded Depression | Switching vs Continuation | Bipolar Disorder | Education_levelTwelve or GED | 0.878 | 0.643 | 1.198 | 4.10e-01 | NA | 3124 | 1930 | 1194 |
| African | Recorded Depression | Switching vs Continuation | Bipolar Disorder | Bipolar Disorder | 1.107 | 0.994 | 1.233 | 6.49e-02 | 4.54e-01 | 3124 | 1930 | 1194 |
| African | Recorded Depression | Switching vs Continuation | Depression | (Intercept) | 1.300 | 0.857 | 1.972 | 2.17e-01 | NA | 3124 | 1930 | 1194 |
| African | Recorded Depression | Switching vs Continuation | Depression | age_at_first_AD | 0.989 | 0.983 | 0.994 | 6.65e-05 | NA | 3124 | 1930 | 1194 |
| African | Recorded Depression | Switching vs Continuation | Depression | sex_at_birthMale | 1.056 | 0.888 | 1.257 | 5.35e-01 | NA | 3124 | 1930 | 1194 |
| African | Recorded Depression | Switching vs Continuation | Depression | Income_quantitative | 0.913 | 0.866 | 0.962 | 6.63e-04 | NA | 3124 | 1930 | 1194 |
| African | Recorded Depression | Switching vs Continuation | Depression | Education_levelCollege Undergraduate | 0.977 | 0.738 | 1.294 | 8.73e-01 | NA | 3124 | 1930 | 1194 |
| African | Recorded Depression | Switching vs Continuation | Depression | Education_levelLess than a high school degree | 0.972 | 0.672 | 1.406 | 8.80e-01 | NA | 3124 | 1930 | 1194 |
| African | Recorded Depression | Switching vs Continuation | Depression | Education_levelTwelve or GED | 0.867 | 0.635 | 1.183 | 3.68e-01 | NA | 3124 | 1930 | 1194 |
| African | Recorded Depression | Switching vs Continuation | Depression | Depression | 1.155 | 1.041 | 1.282 | 6.44e-03 | 4.51e-02 | 3124 | 1930 | 1194 |
| African | Recorded Depression | Switching vs Continuation | Insomnia | (Intercept) | 1.314 | 0.866 | 1.993 | 1.99e-01 | NA | 3124 | 1930 | 1194 |
| African | Recorded Depression | Switching vs Continuation | Insomnia | age_at_first_AD | 0.989 | 0.983 | 0.994 | 6.76e-05 | NA | 3124 | 1930 | 1194 |
| African | Recorded Depression | Switching vs Continuation | Insomnia | sex_at_birthMale | 1.060 | 0.891 | 1.261 | 5.08e-01 | NA | 3124 | 1930 | 1194 |
| African | Recorded Depression | Switching vs Continuation | Insomnia | Income_quantitative | 0.911 | 0.864 | 0.959 | 4.48e-04 | NA | 3124 | 1930 | 1194 |
| African | Recorded Depression | Switching vs Continuation | Insomnia | Education_levelCollege Undergraduate | 0.974 | 0.736 | 1.289 | 8.53e-01 | NA | 3124 | 1930 | 1194 |
| African | Recorded Depression | Switching vs Continuation | Insomnia | Education_levelLess than a high school degree | 0.964 | 0.667 | 1.394 | 8.45e-01 | NA | 3124 | 1930 | 1194 |
| African | Recorded Depression | Switching vs Continuation | Insomnia | Education_levelTwelve or GED | 0.862 | 0.632 | 1.176 | 3.48e-01 | NA | 3124 | 1930 | 1194 |
| African | Recorded Depression | Switching vs Continuation | Insomnia | Insomnia | 1.108 | 0.997 | 1.231 | 5.77e-02 | 4.04e-01 | 3124 | 1930 | 1194 |
| African | Recorded Depression | Switching vs Continuation | Migraine | (Intercept) | 1.307 | 0.862 | 1.982 | 2.08e-01 | NA | 3124 | 1930 | 1194 |
| African | Recorded Depression | Switching vs Continuation | Migraine | age_at_first_AD | 0.989 | 0.983 | 0.994 | 6.73e-05 | NA | 3124 | 1930 | 1194 |
| African | Recorded Depression | Switching vs Continuation | Migraine | sex_at_birthMale | 1.060 | 0.892 | 1.261 | 5.07e-01 | NA | 3124 | 1930 | 1194 |
| African | Recorded Depression | Switching vs Continuation | Migraine | Income_quantitative | 0.911 | 0.864 | 0.960 | 4.56e-04 | NA | 3124 | 1930 | 1194 |
| African | Recorded Depression | Switching vs Continuation | Migraine | Education_levelCollege Undergraduate | 0.977 | 0.738 | 1.294 | 8.73e-01 | NA | 3124 | 1930 | 1194 |
| African | Recorded Depression | Switching vs Continuation | Migraine | Education_levelLess than a high school degree | 0.974 | 0.674 | 1.409 | 8.90e-01 | NA | 3124 | 1930 | 1194 |
| African | Recorded Depression | Switching vs Continuation | Migraine | Education_levelTwelve or GED | 0.867 | 0.636 | 1.183 | 3.67e-01 | NA | 3124 | 1930 | 1194 |
| African | Recorded Depression | Switching vs Continuation | Migraine | Migraine | 0.980 | 0.897 | 1.072 | 6.61e-01 | 1.00e+00 | 3124 | 1930 | 1194 |
| African | Recorded Depression | Switching vs Continuation | Schizophrenia | (Intercept) | 1.307 | 0.862 | 1.982 | 2.08e-01 | NA | 3124 | 1930 | 1194 |
| African | Recorded Depression | Switching vs Continuation | Schizophrenia | age_at_first_AD | 0.989 | 0.983 | 0.994 | 6.57e-05 | NA | 3124 | 1930 | 1194 |
| African | Recorded Depression | Switching vs Continuation | Schizophrenia | sex_at_birthMale | 1.061 | 0.892 | 1.261 | 5.06e-01 | NA | 3124 | 1930 | 1194 |
| African | Recorded Depression | Switching vs Continuation | Schizophrenia | Income_quantitative | 0.911 | 0.864 | 0.960 | 4.55e-04 | NA | 3124 | 1930 | 1194 |
| African | Recorded Depression | Switching vs Continuation | Schizophrenia | Education_levelCollege Undergraduate | 0.978 | 0.739 | 1.295 | 8.76e-01 | NA | 3124 | 1930 | 1194 |
| African | Recorded Depression | Switching vs Continuation | Schizophrenia | Education_levelLess than a high school degree | 0.976 | 0.675 | 1.411 | 8.96e-01 | NA | 3124 | 1930 | 1194 |
| African | Recorded Depression | Switching vs Continuation | Schizophrenia | Education_levelTwelve or GED | 0.867 | 0.636 | 1.183 | 3.67e-01 | NA | 3124 | 1930 | 1194 |
| African | Recorded Depression | Switching vs Continuation | Schizophrenia | Schizophrenia | 0.974 | 0.888 | 1.067 | 5.70e-01 | 1.00e+00 | 3124 | 1930 | 1194 |
| African | No Recorded Depression | Switching vs Continuation | ADHD | (Intercept) | 0.442 | 0.245 | 0.798 | 6.76e-03 | NA | 2950 | 2563 | 387 |
| African | No Recorded Depression | Switching vs Continuation | ADHD | age_at_first_AD | 0.984 | 0.976 | 0.992 | 6.64e-05 | NA | 2950 | 2563 | 387 |
| African | No Recorded Depression | Switching vs Continuation | ADHD | sex_at_birthMale | 1.042 | 0.816 | 1.331 | 7.43e-01 | NA | 2950 | 2563 | 387 |
| African | No Recorded Depression | Switching vs Continuation | ADHD | Income_quantitative | 0.938 | 0.869 | 1.011 | 9.53e-02 | NA | 2950 | 2563 | 387 |
| African | No Recorded Depression | Switching vs Continuation | ADHD | Education_levelCollege Undergraduate | 0.785 | 0.537 | 1.148 | 2.12e-01 | NA | 2950 | 2563 | 387 |
| African | No Recorded Depression | Switching vs Continuation | ADHD | Education_levelLess than a high school degree | 1.121 | 0.689 | 1.823 | 6.46e-01 | NA | 2950 | 2563 | 387 |
| African | No Recorded Depression | Switching vs Continuation | ADHD | Education_levelTwelve or GED | 0.821 | 0.534 | 1.263 | 3.70e-01 | NA | 2950 | 2563 | 387 |
| African | No Recorded Depression | Switching vs Continuation | ADHD | ADHD | 1.001 | 0.860 | 1.166 | 9.86e-01 | 1.00e+00 | 2950 | 2563 | 387 |
| African | No Recorded Depression | Switching vs Continuation | BMI | (Intercept) | 0.442 | 0.245 | 0.797 | 6.67e-03 | NA | 2950 | 2563 | 387 |
| African | No Recorded Depression | Switching vs Continuation | BMI | age_at_first_AD | 0.984 | 0.976 | 0.992 | 6.70e-05 | NA | 2950 | 2563 | 387 |
| African | No Recorded Depression | Switching vs Continuation | BMI | sex_at_birthMale | 1.042 | 0.816 | 1.331 | 7.39e-01 | NA | 2950 | 2563 | 387 |
| African | No Recorded Depression | Switching vs Continuation | BMI | Income_quantitative | 0.938 | 0.870 | 1.012 | 9.64e-02 | NA | 2950 | 2563 | 387 |
| African | No Recorded Depression | Switching vs Continuation | BMI | Education_levelCollege Undergraduate | 0.785 | 0.537 | 1.147 | 2.11e-01 | NA | 2950 | 2563 | 387 |
| African | No Recorded Depression | Switching vs Continuation | BMI | Education_levelLess than a high school degree | 1.118 | 0.688 | 1.819 | 6.52e-01 | NA | 2950 | 2563 | 387 |
| African | No Recorded Depression | Switching vs Continuation | BMI | Education_levelTwelve or GED | 0.820 | 0.533 | 1.261 | 3.66e-01 | NA | 2950 | 2563 | 387 |
| African | No Recorded Depression | Switching vs Continuation | BMI | BMI | 1.025 | 0.888 | 1.182 | 7.37e-01 | 1.00e+00 | 2950 | 2563 | 387 |
| African | No Recorded Depression | Switching vs Continuation | Bipolar Disorder | (Intercept) | 0.441 | 0.244 | 0.796 | 6.54e-03 | NA | 2950 | 2563 | 387 |
| African | No Recorded Depression | Switching vs Continuation | Bipolar Disorder | age_at_first_AD | 0.984 | 0.976 | 0.992 | 6.30e-05 | NA | 2950 | 2563 | 387 |
| African | No Recorded Depression | Switching vs Continuation | Bipolar Disorder | sex_at_birthMale | 1.045 | 0.818 | 1.335 | 7.25e-01 | NA | 2950 | 2563 | 387 |
| African | No Recorded Depression | Switching vs Continuation | Bipolar Disorder | Income_quantitative | 0.939 | 0.870 | 1.012 | 1.00e-01 | NA | 2950 | 2563 | 387 |

| | | | | | | | | | | | | |
|---|---|---|---|---|---|---|---|---|---|---|---|---|
| African | No Recorded Depression | Switching vs Continuation | Bipolar Disorder | Education_levelCollege Undergraduate | 0.785 | 0.537 | 1.148 | 2.12e-01 | NA | 2950 | 2563 | 387 |
| African | No Recorded Depression | Switching vs Continuation | Bipolar Disorder | Education_levelLess than a high school degree | 1.122 | 0.690 | 1.825 | 6.42e-01 | NA | 2950 | 2563 | 387 |
| African | No Recorded Depression | Switching vs Continuation | Bipolar Disorder | Education_levelTwelve or GED | 0.827 | 0.537 | 1.271 | 3.86e-01 | NA | 2950 | 2563 | 387 |
| African | No Recorded Depression | Switching vs Continuation | Bipolar Disorder | Bipolar Disorder | 1.086 | 0.929 | 1.270 | 3.02e-01 | 1.00e+00 | 2950 | 2563 | 387 |
| African | No Recorded Depression | Switching vs Continuation | Depression | (Intercept) | 0.441 | 0.244 | 0.796 | 6.58e-03 | NA | 2950 | 2563 | 387 |
| African | No Recorded Depression | Switching vs Continuation | Depression | age_at_first_AD | 0.984 | 0.976 | 0.992 | 6.40e-05 | NA | 2950 | 2563 | 387 |
| African | No Recorded Depression | Switching vs Continuation | Depression | sex_at_birthMale | 1.042 | 0.816 | 1.331 | 7.40e-01 | NA | 2950 | 2563 | 387 |
| African | No Recorded Depression | Switching vs Continuation | Depression | Income_quantitative | 0.938 | 0.870 | 1.012 | 9.98e-02 | NA | 2950 | 2563 | 387 |
| African | No Recorded Depression | Switching vs Continuation | Depression | Education_levelCollege Undergraduate | 0.786 | 0.537 | 1.149 | 2.14e-01 | NA | 2950 | 2563 | 387 |
| African | No Recorded Depression | Switching vs Continuation | Depression | Education_levelLess than a high school degree | 1.124 | 0.691 | 1.828 | 6.39e-01 | NA | 2950 | 2563 | 387 |
| African | No Recorded Depression | Switching vs Continuation | Depression | Education_levelTwelve or GED | 0.824 | 0.535 | 1.267 | 3.77e-01 | NA | 2950 | 2563 | 387 |
| African | No Recorded Depression | Switching vs Continuation | Depression | Depression | 1.040 | 0.896 | 1.208 | 6.05e-01 | 1.00e+00 | 2950 | 2563 | 387 |
| African | No Recorded Depression | Switching vs Continuation | Insomnia | (Intercept) | 0.444 | 0.246 | 0.802 | 7.07e-03 | NA | 2950 | 2563 | 387 |
| African | No Recorded Depression | Switching vs Continuation | Insomnia | age_at_first_AD | 0.984 | 0.976 | 0.992 | 5.76e-05 | NA | 2950 | 2563 | 387 |
| African | No Recorded Depression | Switching vs Continuation | Insomnia | sex_at_birthMale | 1.040 | 0.814 | 1.328 | 7.55e-01 | NA | 2950 | 2563 | 387 |
| African | No Recorded Depression | Switching vs Continuation | Insomnia | Income_quantitative | 0.939 | 0.870 | 1.012 | 1.01e-01 | NA | 2950 | 2563 | 387 |
| African | No Recorded Depression | Switching vs Continuation | Insomnia | Education_levelCollege Undergraduate | 0.784 | 0.536 | 1.146 | 2.09e-01 | NA | 2950 | 2563 | 387 |
| African | No Recorded Depression | Switching vs Continuation | Insomnia | Education_levelLess than a high school degree | 1.113 | 0.684 | 1.810 | 6.67e-01 | NA | 2950 | 2563 | 387 |
| African | No Recorded Depression | Switching vs Continuation | Insomnia | Education_levelTwelve or GED | 0.824 | 0.535 | 1.267 | 3.77e-01 | NA | 2950 | 2563 | 387 |
| African | No Recorded Depression | Switching vs Continuation | Insomnia | Insomnia | 1.135 | 0.970 | 1.328 | 1.13e-01 | 7.92e-01 | 2950 | 2563 | 387 |
| African | No Recorded Depression | Switching vs Continuation | Migraine | (Intercept) | 0.440 | 0.244 | 0.794 | 6.42e-03 | NA | 2950 | 2563 | 387 |
| African | No Recorded Depression | Switching vs Continuation | Migraine | age_at_first_AD | 0.984 | 0.976 | 0.992 | 6.68e-05 | NA | 2950 | 2563 | 387 |
| African | No Recorded Depression | Switching vs Continuation | Migraine | sex_at_birthMale | 1.041 | 0.815 | 1.329 | 7.50e-01 | NA | 2950 | 2563 | 387 |
| African | No Recorded Depression | Switching vs Continuation | Migraine | Income_quantitative | 0.938 | 0.870 | 1.012 | 9.70e-02 | NA | 2950 | 2563 | 387 |
| African | No Recorded Depression | Switching vs Continuation | Migraine | Education_levelCollege Undergraduate | 0.788 | 0.539 | 1.152 | 2.18e-01 | NA | 2950 | 2563 | 387 |
| African | No Recorded Depression | Switching vs Continuation | Migraine | Education_levelLess than a high school degree | 1.126 | 0.692 | 1.832 | 6.32e-01 | NA | 2950 | 2563 | 387 |
| African | No Recorded Depression | Switching vs Continuation | Migraine | Education_levelTwelve or GED | 0.824 | 0.535 | 1.267 | 3.77e-01 | NA | 2950 | 2563 | 387 |
| African | No Recorded Depression | Switching vs Continuation | Migraine | Migraine | 0.943 | 0.827 | 1.076 | 3.86e-01 | 1.00e+00 | 2950 | 2563 | 387 |
| African | No Recorded Depression | Switching vs Continuation | Schizophrenia | (Intercept) | 0.442 | 0.245 | 0.797 | 6.70e-03 | NA | 2950 | 2563 | 387 |
| African | No Recorded Depression | Switching vs Continuation | Schizophrenia | age_at_first_AD | 0.984 | 0.976 | 0.992 | 6.47e-05 | NA | 2950 | 2563 | 387 |
| African | No Recorded Depression | Switching vs Continuation | Schizophrenia | sex_at_birthMale | 1.042 | 0.816 | 1.331 | 7.43e-01 | NA | 2950 | 2563 | 387 |
| African | No Recorded Depression | Switching vs Continuation | Schizophrenia | Income_quantitative | 0.938 | 0.870 | 1.012 | 9.74e-02 | NA | 2950 | 2563 | 387 |
| African | No Recorded Depression | Switching vs Continuation | Schizophrenia | Education_levelCollege Undergraduate | 0.786 | 0.537 | 1.148 | 2.13e-01 | NA | 2950 | 2563 | 387 |
| African | No Recorded Depression | Switching vs Continuation | Schizophrenia | Education_levelLess than a high school degree | 1.121 | 0.689 | 1.823 | 6.46e-01 | NA | 2950 | 2563 | 387 |
| African | No Recorded Depression | Switching vs Continuation | Schizophrenia | Education_levelTwelve or GED | 0.822 | 0.534 | 1.264 | 3.72e-01 | NA | 2950 | 2563 | 387 |
| African | No Recorded Depression | Switching vs Continuation | Schizophrenia | Schizophrenia | 1.019 | 0.886 | 1.172 | 7.92e-01 | 1.00e+00 | 2950 | 2563 | 387 |
| African | Recorded Anxiety | Switching vs Continuation | ADHD | (Intercept) | 1.022 | 0.689 | 1.517 | 9.14e-01 | NA | 3429 | 2223 | 1206 |
| African | Recorded Anxiety | Switching vs Continuation | ADHD | age_at_first_AD | 0.990 | 0.984 | 0.995 | 1.31e-04 | NA | 3429 | 2223 | 1206 |
| African | Recorded Anxiety | Switching vs Continuation | ADHD | sex_at_birthMale | 1.127 | 0.948 | 1.340 | 1.76e-01 | NA | 3429 | 2223 | 1206 |
| African | Recorded Anxiety | Switching vs Continuation | ADHD | Income_quantitative | 0.889 | 0.846 | 0.934 | 3.35e-06 | NA | 3429 | 2223 | 1206 |
| African | Recorded Anxiety | Switching vs Continuation | ADHD | Education_levelCollege Undergraduate | 1.106 | 0.853 | 1.435 | 4.47e-01 | NA | 3429 | 2223 | 1206 |
| African | Recorded Anxiety | Switching vs Continuation | ADHD | Education_levelLess than a high school degree | 1.132 | 0.793 | 1.616 | 4.95e-01 | NA | 3429 | 2223 | 1206 |
| African | Recorded Anxiety | Switching vs Continuation | ADHD | Education_levelTwelve or GED | 1.028 | 0.768 | 1.378 | 8.51e-01 | NA | 3429 | 2223 | 1206 |
| African | Recorded Anxiety | Switching vs Continuation | ADHD | ADHD | 1.035 | 0.937 | 1.144 | 4.95e-01 | 1.00e+00 | 3429 | 2223 | 1206 |
| African | Recorded Anxiety | Switching vs Continuation | BMI | (Intercept) | 1.026 | 0.691 | 1.524 | 8.97e-01 | NA | 3429 | 2223 | 1206 |
| African | Recorded Anxiety | Switching vs Continuation | BMI | age_at_first_AD | 0.989 | 0.984 | 0.995 | 1.00e-04 | NA | 3429 | 2223 | 1206 |
| African | Recorded Anxiety | Switching vs Continuation | BMI | sex_at_birthMale | 1.122 | 0.944 | 1.335 | 1.91e-01 | NA | 3429 | 2223 | 1206 |
| African | Recorded Anxiety | Switching vs Continuation | BMI | Income_quantitative | 0.888 | 0.845 | 0.933 | 2.74e-06 | NA | 3429 | 2223 | 1206 |
| African | Recorded Anxiety | Switching vs Continuation | BMI | Education_levelCollege Undergraduate | 1.114 | 0.859 | 1.445 | 4.15e-01 | NA | 3429 | 2223 | 1206 |
| African | Recorded Anxiety | Switching vs Continuation | BMI | Education_levelLess than a high school degree | 1.147 | 0.803 | 1.636 | 4.51e-01 | NA | 3429 | 2223 | 1206 |
| African | Recorded Anxiety | Switching vs Continuation | BMI | Education_levelTwelve or GED | 1.040 | 0.776 | 1.394 | 7.91e-01 | NA | 3429 | 2223 | 1206 |
| African | Recorded Anxiety | Switching vs Continuation | BMI | BMI | 0.948 | 0.863 | 1.040 | 2.56e-01 | 1.00e+00 | 3429 | 2223 | 1206 |
| African | Recorded Anxiety | Switching vs Continuation | Bipolar Disorder | (Intercept) | 1.023 | 0.689 | 1.518 | 9.12e-01 | NA | 3429 | 2223 | 1206 |
| African | Recorded Anxiety | Switching vs Continuation | Bipolar Disorder | age_at_first_AD | 0.989 | 0.984 | 0.995 | 1.03e-04 | NA | 3429 | 2223 | 1206 |
| African | Recorded Anxiety | Switching vs Continuation | Bipolar Disorder | sex_at_birthMale | 1.121 | 0.942 | 1.333 | 1.98e-01 | NA | 3429 | 2223 | 1206 |
| African | Recorded Anxiety | Switching vs Continuation | Bipolar Disorder | Income_quantitative | 0.890 | 0.847 | 0.935 | 4.05e-06 | NA | 3429 | 2223 | 1206 |
| African | Recorded Anxiety | Switching vs Continuation | Bipolar Disorder | Education_levelCollege Undergraduate | 1.112 | 0.857 | 1.442 | 4.24e-01 | NA | 3429 | 2223 | 1206 |
| African | Recorded Anxiety | Switching vs Continuation | Bipolar Disorder | Education_levelLess than a high school degree | 1.139 | 0.798 | 1.625 | 4.73e-01 | NA | 3429 | 2223 | 1206 |
| African | Recorded Anxiety | Switching vs Continuation | Bipolar Disorder | Education_levelTwelve or GED | 1.039 | 0.776 | 1.392 | 7.99e-01 | NA | 3429 | 2223 | 1206 |
| African | Recorded Anxiety | Switching vs Continuation | Bipolar Disorder | Bipolar Disorder | 1.095 | 0.987 | 1.216 | 8.74e-02 | 6.12e-01 | 3429 | 2223 | 1206 |

| | | | | | | | | | | | | |
|---|---|---|---|---|---|---|---|---|---|---|---|---|
| African | Recorded Anxiety | Switching vs Continuation | Depression | (Intercept) | 1.012 | 0.681 | 1.503 | 9.53e-01 | NA | 3429 | 2223 | 1206 |
| African | Recorded Anxiety | Switching vs Continuation | Depression | age_at_first_AD | 0.990 | 0.984 | 0.995 | 1.12e-04 | NA | 3429 | 2223 | 1206 |
| African | Recorded Anxiety | Switching vs Continuation | Depression | sex_at_birthMale | 1.123 | 0.944 | 1.335 | 1.90e-01 | NA | 3429 | 2223 | 1206 |
| African | Recorded Anxiety | Switching vs Continuation | Depression | Income_quantitative | 0.892 | 0.849 | 0.938 | 6.56e-06 | NA | 3429 | 2223 | 1206 |
| African | Recorded Anxiety | Switching vs Continuation | Depression | Education_levelCollege Undergraduate | 1.113 | 0.858 | 1.444 | 4.20e-01 | NA | 3429 | 2223 | 1206 |
| African | Recorded Anxiety | Switching vs Continuation | Depression | Education_levelLess than a high school degree | 1.142 | 0.800 | 1.630 | 4.65e-01 | NA | 3429 | 2223 | 1206 |
| African | Recorded Anxiety | Switching vs Continuation | Depression | Education_levelTwelve or GED | 1.038 | 0.774 | 1.390 | 8.05e-01 | NA | 3429 | 2223 | 1206 |
| African | Recorded Anxiety | Switching vs Continuation | Depression | Depression | 1.177 | 1.064 | 1.302 | 1.54e-03 | 1.08e-02 | 3429 | 2223 | 1206 |
| African | Recorded Anxiety | Switching vs Continuation | Insomnia | (Intercept) | 1.044 | 0.703 | 1.552 | 8.30e-01 | NA | 3429 | 2223 | 1206 |
| African | Recorded Anxiety | Switching vs Continuation | Insomnia | age_at_first_AD | 0.989 | 0.984 | 0.995 | 1.03e-04 | NA | 3429 | 2223 | 1206 |
| African | Recorded Anxiety | Switching vs Continuation | Insomnia | sex_at_birthMale | 1.128 | 0.948 | 1.341 | 1.74e-01 | NA | 3429 | 2223 | 1206 |
| African | Recorded Anxiety | Switching vs Continuation | Insomnia | Income_quantitative | 0.888 | 0.845 | 0.933 | 2.52e-06 | NA | 3429 | 2223 | 1206 |
| African | Recorded Anxiety | Switching vs Continuation | Insomnia | Education_levelCollege Undergraduate | 1.093 | 0.842 | 1.418 | 5.04e-01 | NA | 3429 | 2223 | 1206 |
| African | Recorded Anxiety | Switching vs Continuation | Insomnia | Education_levelLess than a high school degree | 1.119 | 0.784 | 1.599 | 5.35e-01 | NA | 3429 | 2223 | 1206 |
| African | Recorded Anxiety | Switching vs Continuation | Insomnia | Education_levelTwelve or GED | 1.014 | 0.756 | 1.358 | 9.28e-01 | NA | 3429 | 2223 | 1206 |
| African | Recorded Anxiety | Switching vs Continuation | Insomnia | Insomnia | 1.192 | 1.076 | 1.321 | 7.89e-04 | 5.52e-03 | 3429 | 2223 | 1206 |
| African | Recorded Anxiety | Switching vs Continuation | Migraine | (Intercept) | 1.020 | 0.687 | 1.514 | 9.23e-01 | NA | 3429 | 2223 | 1206 |
| African | Recorded Anxiety | Switching vs Continuation | Migraine | age_at_first_AD | 0.990 | 0.984 | 0.995 | 1.28e-04 | NA | 3429 | 2223 | 1206 |
| African | Recorded Anxiety | Switching vs Continuation | Migraine | sex_at_birthMale | 1.127 | 0.948 | 1.340 | 1.75e-01 | NA | 3429 | 2223 | 1206 |
| African | Recorded Anxiety | Switching vs Continuation | Migraine | Income_quantitative | 0.889 | 0.846 | 0.934 | 3.33e-06 | NA | 3429 | 2223 | 1206 |
| African | Recorded Anxiety | Switching vs Continuation | Migraine | Education_levelCollege Undergraduate | 1.109 | 0.855 | 1.438 | 4.34e-01 | NA | 3429 | 2223 | 1206 |
| African | Recorded Anxiety | Switching vs Continuation | Migraine | Education_levelLess than a high school degree | 1.139 | 0.799 | 1.626 | 4.72e-01 | NA | 3429 | 2223 | 1206 |
| African | Recorded Anxiety | Switching vs Continuation | Migraine | Education_levelTwelve or GED | 1.032 | 0.771 | 1.383 | 8.30e-01 | NA | 3429 | 2223 | 1206 |
| African | Recorded Anxiety | Switching vs Continuation | Migraine | Migraine | 0.993 | 0.911 | 1.083 | 8.76e-01 | 1.00e+00 | 3429 | 2223 | 1206 |
| African | Recorded Anxiety | Switching vs Continuation | Schizophrenia | (Intercept) | 1.019 | 0.686 | 1.513 | 9.27e-01 | NA | 3429 | 2223 | 1206 |
| African | Recorded Anxiety | Switching vs Continuation | Schizophrenia | age_at_first_AD | 0.990 | 0.984 | 0.995 | 1.42e-04 | NA | 3429 | 2223 | 1206 |
| African | Recorded Anxiety | Switching vs Continuation | Schizophrenia | sex_at_birthMale | 1.129 | 0.950 | 1.343 | 1.69e-01 | NA | 3429 | 2223 | 1206 |
| African | Recorded Anxiety | Switching vs Continuation | Schizophrenia | Income_quantitative | 0.888 | 0.846 | 0.934 | 2.89e-06 | NA | 3429 | 2223 | 1206 |
| African | Recorded Anxiety | Switching vs Continuation | Schizophrenia | Education_levelCollege Undergraduate | 1.108 | 0.854 | 1.437 | 4.41e-01 | NA | 3429 | 2223 | 1206 |
| African | Recorded Anxiety | Switching vs Continuation | Schizophrenia | Education_levelLess than a high school degree | 1.141 | 0.800 | 1.628 | 4.67e-01 | NA | 3429 | 2223 | 1206 |
| African | Recorded Anxiety | Switching vs Continuation | Schizophrenia | Education_levelTwelve or GED | 1.030 | 0.769 | 1.380 | 8.41e-01 | NA | 3429 | 2223 | 1206 |
| African | Recorded Anxiety | Switching vs Continuation | Schizophrenia | Schizophrenia | 0.926 | 0.846 | 1.014 | 9.76e-02 | 6.83e-01 | 3429 | 2223 | 1206 |
| Latino/Mixed-American | Full Cohort | Switching vs Continuation | ADHD | (Intercept) | 0.677 | 0.457 | 1.002 | 5.12e-02 | NA | 3553 | 2629 | 924 |
| Latino/Mixed-American | Full Cohort | Switching vs Continuation | ADHD | age_at_first_AD | 0.987 | 0.981 | 0.992 | 7.05e-07 | NA | 3553 | 2629 | 924 |
| Latino/Mixed-American | Full Cohort | Switching vs Continuation | ADHD | sex_at_birthMale | 1.170 | 0.986 | 1.388 | 7.21e-02 | NA | 3553 | 2629 | 924 |
| Latino/Mixed-American | Full Cohort | Switching vs Continuation | ADHD | Income_quantitative | 0.941 | 0.900 | 0.983 | 6.52e-03 | NA | 3553 | 2629 | 924 |
| Latino/Mixed-American | Full Cohort | Switching vs Continuation | ADHD | Education_levelCollege Undergraduate | 1.156 | 0.879 | 1.520 | 3.00e-01 | NA | 3553 | 2629 | 924 |
| Latino/Mixed-American | Full Cohort | Switching vs Continuation | ADHD | Education_levelLess than a high school degree | 0.845 | 0.601 | 1.187 | 3.31e-01 | NA | 3553 | 2629 | 924 |
| Latino/Mixed-American | Full Cohort | Switching vs Continuation | ADHD | Education_levelTwelve or GED | 1.054 | 0.767 | 1.450 | 7.45e-01 | NA | 3553 | 2629 | 924 |
| Latino/Mixed-American | Full Cohort | Switching vs Continuation | ADHD | ADHD | 1.041 | 0.958 | 1.132 | 3.42e-01 | 1.00e+00 | 3553 | 2629 | 924 |
| Latino/Mixed-American | Full Cohort | Switching vs Continuation | BMI | (Intercept) | 0.678 | 0.458 | 1.003 | 5.19e-02 | NA | 3553 | 2629 | 924 |
| Latino/Mixed-American | Full Cohort | Switching vs Continuation | BMI | age_at_first_AD | 0.986 | 0.981 | 0.992 | 5.55e-07 | NA | 3553 | 2629 | 924 |
| Latino/Mixed-American | Full Cohort | Switching vs Continuation | BMI | sex_at_birthMale | 1.173 | 0.988 | 1.391 | 6.81e-02 | NA | 3553 | 2629 | 924 |
| Latino/Mixed-American | Full Cohort | Switching vs Continuation | BMI | Income_quantitative | 0.942 | 0.901 | 0.984 | 7.70e-03 | NA | 3553 | 2629 | 924 |
| Latino/Mixed-American | Full Cohort | Switching vs Continuation | BMI | Education_levelCollege Undergraduate | 1.155 | 0.878 | 1.519 | 3.02e-01 | NA | 3553 | 2629 | 924 |
| Latino/Mixed-American | Full Cohort | Switching vs Continuation | BMI | Education_levelLess than a high school degree | 0.850 | 0.605 | 1.195 | 3.50e-01 | NA | 3553 | 2629 | 924 |
| Latino/Mixed-American | Full Cohort | Switching vs Continuation | BMI | Education_levelTwelve or GED | 1.057 | 0.769 | 1.453 | 7.32e-01 | NA | 3553 | 2629 | 924 |
| Latino/Mixed-American | Full Cohort | Switching vs Continuation | BMI | BMI | 1.054 | 0.974 | 1.140 | 1.91e-01 | 1.00e+00 | 3553 | 2629 | 924 |
| Latino/Mixed-American | Full Cohort | Switching vs Continuation | Bipolar Disorder | (Intercept) | 0.675 | 0.456 | 1.000 | 4.99e-02 | NA | 3553 | 2629 | 924 |
| Latino/Mixed-American | Full Cohort | Switching vs Continuation | Bipolar Disorder | age_at_first_AD | 0.986 | 0.981 | 0.992 | 5.51e-07 | NA | 3553 | 2629 | 924 |
| Latino/Mixed-American | Full Cohort | Switching vs Continuation | Bipolar Disorder | sex_at_birthMale | 1.171 | 0.987 | 1.390 | 6.99e-02 | NA | 3553 | 2629 | 924 |
| Latino/Mixed-American | Full Cohort | Switching vs Continuation | Bipolar Disorder | Income_quantitative | 0.941 | 0.900 | 0.983 | 6.31e-03 | NA | 3553 | 2629 | 924 |
| Latino/Mixed-American | Full Cohort | Switching vs Continuation | Bipolar Disorder | Education_levelCollege Undergraduate | 1.164 | 0.885 | 1.531 | 2.77e-01 | NA | 3553 | 2629 | 924 |
| Latino/Mixed-American | Full Cohort | Switching vs Continuation | Bipolar Disorder | Education_levelLess than a high school degree | 0.855 | 0.608 | 1.202 | 3.67e-01 | NA | 3553 | 2629 | 924 |
| Latino/Mixed-American | Full Cohort | Switching vs Continuation | Bipolar Disorder | Education_levelTwelve or GED | 1.066 | 0.776 | 1.465 | 6.94e-01 | NA | 3553 | 2629 | 924 |
| Latino/Mixed-American | Full Cohort | Switching vs Continuation | Bipolar Disorder | Bipolar Disorder | 1.025 | 0.942 | 1.115 | 5.64e-01 | 1.00e+00 | 3553 | 2629 | 924 |
| Latino/Mixed-American | Full Cohort | Switching vs Continuation | Depression | (Intercept) | 0.672 | 0.454 | 0.995 | 4.70e-02 | NA | 3553 | 2629 | 924 |
| Latino/Mixed-American | Full Cohort | Switching vs Continuation | Depression | age_at_first_AD | 0.987 | 0.981 | 0.992 | 7.38e-07 | NA | 3553 | 2629 | 924 |
| Latino/Mixed-American | Full Cohort | Switching vs Continuation | Depression | sex_at_birthMale | 1.168 | 0.984 | 1.386 | 7.61e-02 | NA | 3553 | 2629 | 924 |
| Latino/Mixed-American | Full Cohort | Switching vs Continuation | Depression | Income_quantitative | 0.944 | 0.903 | 0.986 | 9.99e-03 | NA | 3553 | 2629 | 924 |

| | | | | | | | | | | | | |
|---|---|---|---|---|---|---|---|---|---|---|---|---|
| Latino/Mixed-American | Full Cohort | Switching vs Continuation | Depression | Education_levelCollege Undergraduate | 1.153 | 0.876 | 1.516 | 3.10e-01 | NA | 3553 | 2629 | 924 |
| Latino/Mixed-American | Full Cohort | Switching vs Continuation | Depression | Education_levelLess than a high school degree | 0.842 | 0.599 | 1.184 | 3.23e-01 | NA | 3553 | 2629 | 924 |
| Latino/Mixed-American | Full Cohort | Switching vs Continuation | Depression | Education_levelTwelve or GED | 1.057 | 0.769 | 1.454 | 7.31e-01 | NA | 3553 | 2629 | 924 |
| Latino/Mixed-American | Full Cohort | Switching vs Continuation | Depression | Depression | 1.156 | 1.060 | 1.260 | 9.91e-04 | 6.94e-03 | 3553 | 2629 | 924 |
| Latino/Mixed-American | Full Cohort | Switching vs Continuation | Insomnia | (Intercept) | 0.680 | 0.460 | 1.007 | 5.43e-02 | NA | 3553 | 2629 | 924 |
| Latino/Mixed-American | Full Cohort | Switching vs Continuation | Insomnia | age_at_first_AD | 0.986 | 0.981 | 0.992 | 6.02e-07 | NA | 3553 | 2629 | 924 |
| Latino/Mixed-American | Full Cohort | Switching vs Continuation | Insomnia | sex_at_birthMale | 1.174 | 0.989 | 1.393 | 6.64e-02 | NA | 3553 | 2629 | 924 |
| Latino/Mixed-American | Full Cohort | Switching vs Continuation | Insomnia | Income_quantitative | 0.941 | 0.900 | 0.983 | 6.52e-03 | NA | 3553 | 2629 | 924 |
| Latino/Mixed-American | Full Cohort | Switching vs Continuation | Insomnia | Education_levelCollege Undergraduate | 1.150 | 0.874 | 1.512 | 3.19e-01 | NA | 3553 | 2629 | 924 |
| Latino/Mixed-American | Full Cohort | Switching vs Continuation | Insomnia | Education_levelLess than a high school degree | 0.841 | 0.598 | 1.182 | 3.19e-01 | NA | 3553 | 2629 | 924 |
| Latino/Mixed-American | Full Cohort | Switching vs Continuation | Insomnia | Education_levelTwelve or GED | 1.051 | 0.765 | 1.445 | 7.58e-01 | NA | 3553 | 2629 | 924 |
| Latino/Mixed-American | Full Cohort | Switching vs Continuation | Insomnia | Insomnia | 1.098 | 1.006 | 1.198 | 3.70e-02 | 2.59e-01 | 3553 | 2629 | 924 |
| Latino/Mixed-American | Full Cohort | Switching vs Continuation | Migraine | (Intercept) | 0.677 | 0.458 | 1.002 | 5.15e-02 | NA | 3553 | 2629 | 924 |
| Latino/Mixed-American | Full Cohort | Switching vs Continuation | Migraine | age_at_first_AD | 0.986 | 0.981 | 0.992 | 5.90e-07 | NA | 3553 | 2629 | 924 |
| Latino/Mixed-American | Full Cohort | Switching vs Continuation | Migraine | sex_at_birthMale | 1.170 | 0.986 | 1.388 | 7.15e-02 | NA | 3553 | 2629 | 924 |
| Latino/Mixed-American | Full Cohort | Switching vs Continuation | Migraine | Income_quantitative | 0.940 | 0.900 | 0.982 | 5.85e-03 | NA | 3553 | 2629 | 924 |
| Latino/Mixed-American | Full Cohort | Switching vs Continuation | Migraine | Education_levelCollege Undergraduate | 1.161 | 0.883 | 1.527 | 2.85e-01 | NA | 3553 | 2629 | 924 |
| Latino/Mixed-American | Full Cohort | Switching vs Continuation | Migraine | Education_levelLess than a high school degree | 0.852 | 0.606 | 1.197 | 3.55e-01 | NA | 3553 | 2629 | 924 |
| Latino/Mixed-American | Full Cohort | Switching vs Continuation | Migraine | Education_levelTwelve or GED | 1.062 | 0.773 | 1.459 | 7.12e-01 | NA | 3553 | 2629 | 924 |
| Latino/Mixed-American | Full Cohort | Switching vs Continuation | Migraine | Migraine | 1.012 | 0.935 | 1.096 | 7.59e-01 | 1.00e+00 | 3553 | 2629 | 924 |
| Latino/Mixed-American | Full Cohort | Switching vs Continuation | Schizophrenia | (Intercept) | 0.674 | 0.456 | 0.998 | 4.89e-02 | NA | 3553 | 2629 | 924 |
| Latino/Mixed-American | Full Cohort | Switching vs Continuation | Schizophrenia | age_at_first_AD | 0.986 | 0.981 | 0.992 | 5.43e-07 | NA | 3553 | 2629 | 924 |
| Latino/Mixed-American | Full Cohort | Switching vs Continuation | Schizophrenia | sex_at_birthMale | 1.171 | 0.987 | 1.389 | 7.08e-02 | NA | 3553 | 2629 | 924 |
| Latino/Mixed-American | Full Cohort | Switching vs Continuation | Schizophrenia | Income_quantitative | 0.941 | 0.900 | 0.983 | 6.45e-03 | NA | 3553 | 2629 | 924 |
| Latino/Mixed-American | Full Cohort | Switching vs Continuation | Schizophrenia | Education_levelCollege Undergraduate | 1.166 | 0.887 | 1.534 | 2.72e-01 | NA | 3553 | 2629 | 924 |
| Latino/Mixed-American | Full Cohort | Switching vs Continuation | Schizophrenia | Education_levelLess than a high school degree | 0.855 | 0.609 | 1.202 | 3.68e-01 | NA | 3553 | 2629 | 924 |
| Latino/Mixed-American | Full Cohort | Switching vs Continuation | Schizophrenia | Education_levelTwelve or GED | 1.068 | 0.777 | 1.469 | 6.83e-01 | NA | 3553 | 2629 | 924 |
| Latino/Mixed-American | Full Cohort | Switching vs Continuation | Schizophrenia | Schizophrenia | 1.034 | 0.958 | 1.115 | 3.90e-01 | 1.00e+00 | 3553 | 2629 | 924 |
| Latino/Mixed-American | Recorded Depression | Switching vs Continuation | ADHD | (Intercept) | 0.972 | 0.590 | 1.602 | 9.12e-01 | NA | 1996 | 1301 | 695 |
| Latino/Mixed-American | Recorded Depression | Switching vs Continuation | ADHD | age_at_first_AD | 0.991 | 0.985 | 0.998 | 8.09e-03 | NA | 1996 | 1301 | 695 |
| Latino/Mixed-American | Recorded Depression | Switching vs Continuation | ADHD | sex_at_birthMale | 1.093 | 0.885 | 1.350 | 4.09e-01 | NA | 1996 | 1301 | 695 |
| Latino/Mixed-American | Recorded Depression | Switching vs Continuation | ADHD | Income_quantitative | 0.952 | 0.899 | 1.008 | 9.05e-02 | NA | 1996 | 1301 | 695 |
| Latino/Mixed-American | Recorded Depression | Switching vs Continuation | ADHD | Education_levelCollege Undergraduate | 1.004 | 0.706 | 1.428 | 9.82e-01 | NA | 1996 | 1301 | 695 |
| Latino/Mixed-American | Recorded Depression | Switching vs Continuation | ADHD | Education_levelLess than a high school degree | 0.702 | 0.460 | 1.071 | 1.00e-01 | NA | 1996 | 1301 | 695 |
| Latino/Mixed-American | Recorded Depression | Switching vs Continuation | ADHD | Education_levelTwelve or GED | 0.801 | 0.538 | 1.193 | 2.75e-01 | NA | 1996 | 1301 | 695 |
| Latino/Mixed-American | Recorded Depression | Switching vs Continuation | ADHD | ADHD | 0.980 | 0.882 | 1.088 | 7.04e-01 | 1.00e+00 | 1996 | 1301 | 695 |
| Latino/Mixed-American | Recorded Depression | Switching vs Continuation | BMI | (Intercept) | 0.973 | 0.591 | 1.604 | 9.16e-01 | NA | 1996 | 1301 | 695 |
| Latino/Mixed-American | Recorded Depression | Switching vs Continuation | BMI | age_at_first_AD | 0.991 | 0.985 | 0.998 | 8.22e-03 | NA | 1996 | 1301 | 695 |
| Latino/Mixed-American | Recorded Depression | Switching vs Continuation | BMI | sex_at_birthMale | 1.093 | 0.885 | 1.351 | 4.08e-01 | NA | 1996 | 1301 | 695 |
| Latino/Mixed-American | Recorded Depression | Switching vs Continuation | BMI | Income_quantitative | 0.953 | 0.900 | 1.009 | 9.66e-02 | NA | 1996 | 1301 | 695 |
| Latino/Mixed-American | Recorded Depression | Switching vs Continuation | BMI | Education_levelCollege Undergraduate | 1.001 | 0.704 | 1.424 | 9.97e-01 | NA | 1996 | 1301 | 695 |
| Latino/Mixed-American | Recorded Depression | Switching vs Continuation | BMI | Education_levelLess than a high school degree | 0.698 | 0.458 | 1.065 | 9.54e-02 | NA | 1996 | 1301 | 695 |
| Latino/Mixed-American | Recorded Depression | Switching vs Continuation | BMI | Education_levelTwelve or GED | 0.798 | 0.536 | 1.188 | 2.66e-01 | NA | 1996 | 1301 | 695 |
| Latino/Mixed-American | Recorded Depression | Switching vs Continuation | BMI | BMI | 1.013 | 0.920 | 1.116 | 7.89e-01 | 1.00e+00 | 1996 | 1301 | 695 |
| Latino/Mixed-American | Recorded Depression | Switching vs Continuation | Bipolar Disorder | (Intercept) | 0.967 | 0.587 | 1.594 | 8.96e-01 | NA | 1996 | 1301 | 695 |
| Latino/Mixed-American | Recorded Depression | Switching vs Continuation | Bipolar Disorder | age_at_first_AD | 0.991 | 0.985 | 0.998 | 8.05e-03 | NA | 1996 | 1301 | 695 |
| Latino/Mixed-American | Recorded Depression | Switching vs Continuation | Bipolar Disorder | sex_at_birthMale | 1.093 | 0.885 | 1.350 | 4.10e-01 | NA | 1996 | 1301 | 695 |
| Latino/Mixed-American | Recorded Depression | Switching vs Continuation | Bipolar Disorder | Income_quantitative | 0.953 | 0.900 | 1.009 | 9.58e-02 | NA | 1996 | 1301 | 695 |
| Latino/Mixed-American | Recorded Depression | Switching vs Continuation | Bipolar Disorder | Education_levelCollege Undergraduate | 1.009 | 0.709 | 1.435 | 9.62e-01 | NA | 1996 | 1301 | 695 |
| Latino/Mixed-American | Recorded Depression | Switching vs Continuation | Bipolar Disorder | Education_levelLess than a high school degree | 0.705 | 0.462 | 1.076 | 1.05e-01 | NA | 1996 | 1301 | 695 |
| Latino/Mixed-American | Recorded Depression | Switching vs Continuation | Bipolar Disorder | Education_levelTwelve or GED | 0.803 | 0.540 | 1.196 | 2.81e-01 | NA | 1996 | 1301 | 695 |
| Latino/Mixed-American | Recorded Depression | Switching vs Continuation | Bipolar Disorder | Bipolar Disorder | 1.043 | 0.941 | 1.157 | 4.21e-01 | 1.00e+00 | 1996 | 1301 | 695 |
| Latino/Mixed-American | Recorded Depression | Switching vs Continuation | Depression | (Intercept) | 0.975 | 0.591 | 1.608 | 9.22e-01 | NA | 1996 | 1301 | 695 |
| Latino/Mixed-American | Recorded Depression | Switching vs Continuation | Depression | age_at_first_AD | 0.991 | 0.985 | 0.998 | 8.85e-03 | NA | 1996 | 1301 | 695 |
| Latino/Mixed-American | Recorded Depression | Switching vs Continuation | Depression | sex_at_birthMale | 1.090 | 0.882 | 1.347 | 4.25e-01 | NA | 1996 | 1301 | 695 |
| Latino/Mixed-American | Recorded Depression | Switching vs Continuation | Depression | Income_quantitative | 0.954 | 0.901 | 1.010 | 1.03e-01 | NA | 1996 | 1301 | 695 |
| Latino/Mixed-American | Recorded Depression | Switching vs Continuation | Depression | Education_levelCollege Undergraduate | 0.996 | 0.700 | 1.417 | 9.81e-01 | NA | 1996 | 1301 | 695 |
| Latino/Mixed-American | Recorded Depression | Switching vs Continuation | Depression | Education_levelLess than a high school degree | 0.690 | 0.452 | 1.052 | 8.47e-02 | NA | 1996 | 1301 | 695 |
| Latino/Mixed-American | Recorded Depression | Switching vs Continuation | Depression | Education_levelTwelve or GED | 0.792 | 0.532 | 1.180 | 2.52e-01 | NA | 1996 | 1301 | 695 |
| Latino/Mixed-American | Recorded Depression | Switching vs Continuation | Depression | Depression | 1.138 | 1.023 | 1.266 | 1.71e-02 | 1.20e-01 | 1996 | 1301 | 695 |

| | | | | | | | | | | | | |
|---|---|---|---|---|---|---|---|---|---|---|---|---|
| Latino/Mixed-American | Recorded Depression | Switching vs Continuation | Insomnia | (Intercept) | 0.978 | 0.593 | 1.612 | 9.31e-01 | NA | 1996 | 1301 | 695 |
| Latino/Mixed-American | Recorded Depression | Switching vs Continuation | Insomnia | age_at_first_AD | 0.991 | 0.985 | 0.998 | 8.30e-03 | NA | 1996 | 1301 | 695 |
| Latino/Mixed-American | Recorded Depression | Switching vs Continuation | Insomnia | sex_at_birthMale | 1.094 | 0.886 | 1.352 | 4.03e-01 | NA | 1996 | 1301 | 695 |
| Latino/Mixed-American | Recorded Depression | Switching vs Continuation | Insomnia | Income_quantitative | 0.952 | 0.900 | 1.008 | 9.32e-02 | NA | 1996 | 1301 | 695 |
| Latino/Mixed-American | Recorded Depression | Switching vs Continuation | Insomnia | Education_levelCollege Undergraduate | 0.995 | 0.699 | 1.415 | 9.77e-01 | NA | 1996 | 1301 | 695 |
| Latino/Mixed-American | Recorded Depression | Switching vs Continuation | Insomnia | Education_levelLess than a high school degree | 0.693 | 0.454 | 1.056 | 8.81e-02 | NA | 1996 | 1301 | 695 |
| Latino/Mixed-American | Recorded Depression | Switching vs Continuation | Insomnia | Education_levelTwelve or GED | 0.793 | 0.533 | 1.180 | 2.53e-01 | NA | 1996 | 1301 | 695 |
| Latino/Mixed-American | Recorded Depression | Switching vs Continuation | Insomnia | Insomnia | 1.083 | 0.974 | 1.204 | 1.40e-01 | 9.79e-01 | 1996 | 1301 | 695 |
| Latino/Mixed-American | Recorded Depression | Switching vs Continuation | Migraine | (Intercept) | 0.978 | 0.593 | 1.611 | 9.29e-01 | NA | 1996 | 1301 | 695 |
| Latino/Mixed-American | Recorded Depression | Switching vs Continuation | Migraine | age_at_first_AD | 0.991 | 0.985 | 0.998 | 8.67e-03 | NA | 1996 | 1301 | 695 |
| Latino/Mixed-American | Recorded Depression | Switching vs Continuation | Migraine | sex_at_birthMale | 1.093 | 0.885 | 1.350 | 4.09e-01 | NA | 1996 | 1301 | 695 |
| Latino/Mixed-American | Recorded Depression | Switching vs Continuation | Migraine | Income_quantitative | 0.951 | 0.899 | 1.007 | 8.70e-02 | NA | 1996 | 1301 | 695 |
| Latino/Mixed-American | Recorded Depression | Switching vs Continuation | Migraine | Education_levelCollege Undergraduate | 1.002 | 0.704 | 1.425 | 9.93e-01 | NA | 1996 | 1301 | 695 |
| Latino/Mixed-American | Recorded Depression | Switching vs Continuation | Migraine | Education_levelLess than a high school degree | 0.694 | 0.455 | 1.058 | 8.98e-02 | NA | 1996 | 1301 | 695 |
| Latino/Mixed-American | Recorded Depression | Switching vs Continuation | Migraine | Education_levelTwelve or GED | 0.789 | 0.529 | 1.174 | 2.42e-01 | NA | 1996 | 1301 | 695 |
| Latino/Mixed-American | Recorded Depression | Switching vs Continuation | Migraine | Migraine | 1.070 | 0.971 | 1.180 | 1.73e-01 | 1.00e+00 | 1996 | 1301 | 695 |
| Latino/Mixed-American | Recorded Depression | Switching vs Continuation | Schizophrenia | (Intercept) | 0.967 | 0.587 | 1.594 | 8.95e-01 | NA | 1996 | 1301 | 695 |
| Latino/Mixed-American | Recorded Depression | Switching vs Continuation | Schizophrenia | age_at_first_AD | 0.991 | 0.985 | 0.998 | 8.29e-03 | NA | 1996 | 1301 | 695 |
| Latino/Mixed-American | Recorded Depression | Switching vs Continuation | Schizophrenia | sex_at_birthMale | 1.093 | 0.885 | 1.350 | 4.09e-01 | NA | 1996 | 1301 | 695 |
| Latino/Mixed-American | Recorded Depression | Switching vs Continuation | Schizophrenia | Income_quantitative | 0.952 | 0.900 | 1.008 | 9.31e-02 | NA | 1996 | 1301 | 695 |
| Latino/Mixed-American | Recorded Depression | Switching vs Continuation | Schizophrenia | Education_levelCollege Undergraduate | 1.010 | 0.710 | 1.436 | 9.58e-01 | NA | 1996 | 1301 | 695 |
| Latino/Mixed-American | Recorded Depression | Switching vs Continuation | Schizophrenia | Education_levelLess than a high school degree | 0.703 | 0.461 | 1.072 | 1.01e-01 | NA | 1996 | 1301 | 695 |
| Latino/Mixed-American | Recorded Depression | Switching vs Continuation | Schizophrenia | Education_levelTwelve or GED | 0.805 | 0.540 | 1.198 | 2.84e-01 | NA | 1996 | 1301 | 695 |
| Latino/Mixed-American | Recorded Depression | Switching vs Continuation | Schizophrenia | Schizophrenia | 1.048 | 0.954 | 1.151 | 3.27e-01 | 1.00e+00 | 1996 | 1301 | 695 |
| Latino/Mixed-American | No Recorded Depression | Switching vs Continuation | ADHD | (Intercept) | 0.262 | 0.130 | 0.529 | 1.86e-04 | NA | 1557 | 1328 | 229 |
| Latino/Mixed-American | No Recorded Depression | Switching vs Continuation | ADHD | age_at_first_AD | 0.980 | 0.970 | 0.990 | 1.29e-04 | NA | 1557 | 1328 | 229 |
| Latino/Mixed-American | No Recorded Depression | Switching vs Continuation | ADHD | sex_at_birthMale | 1.317 | 0.959 | 1.809 | 8.86e-02 | NA | 1557 | 1328 | 229 |
| Latino/Mixed-American | No Recorded Depression | Switching vs Continuation | ADHD | Income_quantitative | 1.024 | 0.950 | 1.105 | 5.34e-01 | NA | 1557 | 1328 | 229 |
| Latino/Mixed-American | No Recorded Depression | Switching vs Continuation | ADHD | Education_levelCollege Undergraduate | 1.467 | 0.908 | 2.368 | 1.17e-01 | NA | 1557 | 1328 | 229 |
| Latino/Mixed-American | No Recorded Depression | Switching vs Continuation | ADHD | Education_levelLess than a high school degree | 1.074 | 0.562 | 2.052 | 8.29e-01 | NA | 1557 | 1328 | 229 |
| Latino/Mixed-American | No Recorded Depression | Switching vs Continuation | ADHD | Education_levelTwelve or GED | 1.532 | 0.851 | 2.758 | 1.55e-01 | NA | 1557 | 1328 | 229 |
| Latino/Mixed-American | No Recorded Depression | Switching vs Continuation | ADHD | ADHD | 1.082 | 0.928 | 1.260 | 3.14e-01 | 1.00e+00 | 1557 | 1328 | 229 |
| Latino/Mixed-American | No Recorded Depression | Switching vs Continuation | BMI | (Intercept) | 0.261 | 0.129 | 0.527 | 1.79e-04 | NA | 1557 | 1328 | 229 |
| Latino/Mixed-American | No Recorded Depression | Switching vs Continuation | BMI | age_at_first_AD | 0.980 | 0.970 | 0.990 | 1.06e-04 | NA | 1557 | 1328 | 229 |
| Latino/Mixed-American | No Recorded Depression | Switching vs Continuation | BMI | sex_at_birthMale | 1.323 | 0.964 | 1.817 | 8.33e-02 | NA | 1557 | 1328 | 229 |
| Latino/Mixed-American | No Recorded Depression | Switching vs Continuation | BMI | Income_quantitative | 1.025 | 0.950 | 1.106 | 5.23e-01 | NA | 1557 | 1328 | 229 |
| Latino/Mixed-American | No Recorded Depression | Switching vs Continuation | BMI | Education_levelCollege Undergraduate | 1.480 | 0.917 | 2.389 | 1.08e-01 | NA | 1557 | 1328 | 229 |
| Latino/Mixed-American | No Recorded Depression | Switching vs Continuation | BMI | Education_levelLess than a high school degree | 1.103 | 0.578 | 2.106 | 7.66e-01 | NA | 1557 | 1328 | 229 |
| Latino/Mixed-American | No Recorded Depression | Switching vs Continuation | BMI | Education_levelTwelve or GED | 1.563 | 0.869 | 2.811 | 1.36e-01 | NA | 1557 | 1328 | 229 |
| Latino/Mixed-American | No Recorded Depression | Switching vs Continuation | BMI | BMI | 1.055 | 0.911 | 1.222 | 4.77e-01 | 1.00e+00 | 1557 | 1328 | 229 |
| Latino/Mixed-American | No Recorded Depression | Switching vs Continuation | Bipolar Disorder | (Intercept) | 0.264 | 0.131 | 0.532 | 2.01e-04 | NA | 1557 | 1328 | 229 |
| Latino/Mixed-American | No Recorded Depression | Switching vs Continuation | Bipolar Disorder | age_at_first_AD | 0.980 | 0.970 | 0.990 | 1.00e-04 | NA | 1557 | 1328 | 229 |
| Latino/Mixed-American | No Recorded Depression | Switching vs Continuation | Bipolar Disorder | sex_at_birthMale | 1.315 | 0.957 | 1.806 | 9.09e-02 | NA | 1557 | 1328 | 229 |
| Latino/Mixed-American | No Recorded Depression | Switching vs Continuation | Bipolar Disorder | Income_quantitative | 1.022 | 0.948 | 1.103 | 5.67e-01 | NA | 1557 | 1328 | 229 |
| Latino/Mixed-American | No Recorded Depression | Switching vs Continuation | Bipolar Disorder | Education_levelCollege Undergraduate | 1.483 | 0.919 | 2.393 | 1.07e-01 | NA | 1557 | 1328 | 229 |
| Latino/Mixed-American | No Recorded Depression | Switching vs Continuation | Bipolar Disorder | Education_levelLess than a high school degree | 1.102 | 0.577 | 2.104 | 7.69e-01 | NA | 1557 | 1328 | 229 |
| Latino/Mixed-American | No Recorded Depression | Switching vs Continuation | Bipolar Disorder | Education_levelTwelve or GED | 1.567 | 0.871 | 2.817 | 1.34e-01 | NA | 1557 | 1328 | 229 |
| Latino/Mixed-American | No Recorded Depression | Switching vs Continuation | Bipolar Disorder | Bipolar Disorder | 0.955 | 0.816 | 1.117 | 5.63e-01 | 1.00e+00 | 1557 | 1328 | 229 |
| Latino/Mixed-American | No Recorded Depression | Switching vs Continuation | Depression | (Intercept) | 0.260 | 0.129 | 0.525 | 1.70e-04 | NA | 1557 | 1328 | 229 |
| Latino/Mixed-American | No Recorded Depression | Switching vs Continuation | Depression | age_at_first_AD | 0.980 | 0.970 | 0.990 | 1.07e-04 | NA | 1557 | 1328 | 229 |
| Latino/Mixed-American | No Recorded Depression | Switching vs Continuation | Depression | sex_at_birthMale | 1.320 | 0.961 | 1.813 | 8.60e-02 | NA | 1557 | 1328 | 229 |
| Latino/Mixed-American | No Recorded Depression | Switching vs Continuation | Depression | Income_quantitative | 1.027 | 0.952 | 1.108 | 4.90e-01 | NA | 1557 | 1328 | 229 |
| Latino/Mixed-American | No Recorded Depression | Switching vs Continuation | Depression | Education_levelCollege Undergraduate | 1.476 | 0.915 | 2.382 | 1.11e-01 | NA | 1557 | 1328 | 229 |
| Latino/Mixed-American | No Recorded Depression | Switching vs Continuation | Depression | Education_levelLess than a high school degree | 1.094 | 0.573 | 2.087 | 7.86e-01 | NA | 1557 | 1328 | 229 |
| Latino/Mixed-American | No Recorded Depression | Switching vs Continuation | Depression | Education_levelTwelve or GED | 1.569 | 0.873 | 2.821 | 1.32e-01 | NA | 1557 | 1328 | 229 |
| Latino/Mixed-American | No Recorded Depression | Switching vs Continuation | Depression | Depression | 1.101 | 0.937 | 1.294 | 2.43e-01 | 1.00e+00 | 1557 | 1328 | 229 |
| Latino/Mixed-American | No Recorded Depression | Switching vs Continuation | Insomnia | (Intercept) | 0.263 | 0.130 | 0.530 | 1.89e-04 | NA | 1557 | 1328 | 229 |
| Latino/Mixed-American | No Recorded Depression | Switching vs Continuation | Insomnia | age_at_first_AD | 0.980 | 0.970 | 0.990 | 1.00e-04 | NA | 1557 | 1328 | 229 |
| Latino/Mixed-American | No Recorded Depression | Switching vs Continuation | Insomnia | sex_at_birthMale | 1.323 | 0.964 | 1.818 | 8.34e-02 | NA | 1557 | 1328 | 229 |
| Latino/Mixed-American | No Recorded Depression | Switching vs Continuation | Insomnia | Income_quantitative | 1.024 | 0.949 | 1.105 | 5.44e-01 | NA | 1557 | 1328 | 229 |

| | | | | | | | | | | | | |
|---|---|---|---|---|---|---|---|---|---|---|---|---|
| Latino/Mixed-American | No Recorded Depression | Switching vs Continuation | Insomnia | Education_levelCollege Undergraduate | 1.481 | 0.918 | 2.391 | 1.08e-01 | NA | 1557 | 1328 | 229 |
| Latino/Mixed-American | No Recorded Depression | Switching vs Continuation | Insomnia | Education_levelLess than a high school degree | 1.096 | 0.574 | 2.093 | 7.81e-01 | NA | 1557 | 1328 | 229 |
| Latino/Mixed-American | No Recorded Depression | Switching vs Continuation | Insomnia | Education_levelTwelve or GED | 1.565 | 0.870 | 2.816 | 1.35e-01 | NA | 1557 | 1328 | 229 |
| Latino/Mixed-American | No Recorded Depression | Switching vs Continuation | Insomnia | Insomnia | 1.028 | 0.870 | 1.216 | 7.46e-01 | 1.00e+00 | 1557 | 1328 | 229 |
| Latino/Mixed-American | No Recorded Depression | Switching vs Continuation | Migraine | (Intercept) | 0.264 | 0.130 | 0.533 | 2.02e-04 | NA | 1557 | 1328 | 229 |
| Latino/Mixed-American | No Recorded Depression | Switching vs Continuation | Migraine | age_at_first_AD | 0.980 | 0.970 | 0.990 | 9.16e-05 | NA | 1557 | 1328 | 229 |
| Latino/Mixed-American | No Recorded Depression | Switching vs Continuation | Migraine | sex_at_birthMale | 1.323 | 0.964 | 1.817 | 8.34e-02 | NA | 1557 | 1328 | 229 |
| Latino/Mixed-American | No Recorded Depression | Switching vs Continuation | Migraine | Income_quantitative | 1.023 | 0.948 | 1.104 | 5.59e-01 | NA | 1557 | 1328 | 229 |
| Latino/Mixed-American | No Recorded Depression | Switching vs Continuation | Migraine | Education_levelCollege Undergraduate | 1.486 | 0.921 | 2.399 | 1.05e-01 | NA | 1557 | 1328 | 229 |
| Latino/Mixed-American | No Recorded Depression | Switching vs Continuation | Migraine | Education_levelLess than a high school degree | 1.104 | 0.578 | 2.108 | 7.64e-01 | NA | 1557 | 1328 | 229 |
| Latino/Mixed-American | No Recorded Depression | Switching vs Continuation | Migraine | Education_levelTwelve or GED | 1.574 | 0.875 | 2.831 | 1.30e-01 | NA | 1557 | 1328 | 229 |
| Latino/Mixed-American | No Recorded Depression | Switching vs Continuation | Migraine | Migraine | 0.955 | 0.822 | 1.109 | 5.46e-01 | 1.00e+00 | 1557 | 1328 | 229 |
| Latino/Mixed-American | No Recorded Depression | Switching vs Continuation | Schizophrenia | (Intercept) | 0.261 | 0.129 | 0.527 | 1.81e-04 | NA | 1557 | 1328 | 229 |
| Latino/Mixed-American | No Recorded Depression | Switching vs Continuation | Schizophrenia | age_at_first_AD | 0.980 | 0.970 | 0.990 | 9.76e-05 | NA | 1557 | 1328 | 229 |
| Latino/Mixed-American | No Recorded Depression | Switching vs Continuation | Schizophrenia | sex_at_birthMale | 1.321 | 0.962 | 1.814 | 8.48e-02 | NA | 1557 | 1328 | 229 |
| Latino/Mixed-American | No Recorded Depression | Switching vs Continuation | Schizophrenia | Income_quantitative | 1.024 | 0.949 | 1.106 | 5.35e-01 | NA | 1557 | 1328 | 229 |
| Latino/Mixed-American | No Recorded Depression | Switching vs Continuation | Schizophrenia | Education_levelCollege Undergraduate | 1.489 | 0.922 | 2.403 | 1.03e-01 | NA | 1557 | 1328 | 229 |
| Latino/Mixed-American | No Recorded Depression | Switching vs Continuation | Schizophrenia | Education_levelLess than a high school degree | 1.104 | 0.578 | 2.106 | 7.65e-01 | NA | 1557 | 1328 | 229 |
| Latino/Mixed-American | No Recorded Depression | Switching vs Continuation | Schizophrenia | Education_levelTwelve or GED | 1.576 | 0.876 | 2.834 | 1.29e-01 | NA | 1557 | 1328 | 229 |
| Latino/Mixed-American | No Recorded Depression | Switching vs Continuation | Schizophrenia | Schizophrenia | 1.023 | 0.889 | 1.176 | 7.55e-01 | 1.00e+00 | 1557 | 1328 | 229 |
| Latino/Mixed-American | Recorded Anxiety | Switching vs Continuation | ADHD | (Intercept) | 0.638 | 0.405 | 1.008 | 5.40e-02 | NA | 2360 | 1590 | 770 |
| Latino/Mixed-American | Recorded Anxiety | Switching vs Continuation | ADHD | age_at_first_AD | 0.994 | 0.988 | 1.001 | 7.38e-02 | NA | 2360 | 1590 | 770 |
| Latino/Mixed-American | Recorded Anxiety | Switching vs Continuation | ADHD | sex_at_birthMale | 1.218 | 1.001 | 1.483 | 4.89e-02 | NA | 2360 | 1590 | 770 |
| Latino/Mixed-American | Recorded Anxiety | Switching vs Continuation | ADHD | Income_quantitative | 0.961 | 0.913 | 1.012 | 1.30e-01 | NA | 2360 | 1590 | 770 |
| Latino/Mixed-American | Recorded Anxiety | Switching vs Continuation | ADHD | Education_levelCollege Undergraduate | 1.114 | 0.811 | 1.530 | 5.06e-01 | NA | 2360 | 1590 | 770 |
| Latino/Mixed-American | Recorded Anxiety | Switching vs Continuation | ADHD | Education_levelLess than a high school degree | 0.860 | 0.580 | 1.276 | 4.55e-01 | NA | 2360 | 1590 | 770 |
| Latino/Mixed-American | Recorded Anxiety | Switching vs Continuation | ADHD | Education_levelTwelve or GED | 0.942 | 0.654 | 1.358 | 7.49e-01 | NA | 2360 | 1590 | 770 |
| Latino/Mixed-American | Recorded Anxiety | Switching vs Continuation | ADHD | ADHD | 1.063 | 0.966 | 1.170 | 2.10e-01 | 1.00e+00 | 2360 | 1590 | 770 |
| Latino/Mixed-American | Recorded Anxiety | Switching vs Continuation | BMI | (Intercept) | 0.639 | 0.405 | 1.008 | 5.43e-02 | NA | 2360 | 1590 | 770 |
| Latino/Mixed-American | Recorded Anxiety | Switching vs Continuation | BMI | age_at_first_AD | 0.994 | 0.988 | 1.000 | 6.56e-02 | NA | 2360 | 1590 | 770 |
| Latino/Mixed-American | Recorded Anxiety | Switching vs Continuation | BMI | sex_at_birthMale | 1.221 | 1.003 | 1.486 | 4.65e-02 | NA | 2360 | 1590 | 770 |
| Latino/Mixed-American | Recorded Anxiety | Switching vs Continuation | BMI | Income_quantitative | 0.961 | 0.912 | 1.011 | 1.26e-01 | NA | 2360 | 1590 | 770 |
| Latino/Mixed-American | Recorded Anxiety | Switching vs Continuation | BMI | Education_levelCollege Undergraduate | 1.120 | 0.816 | 1.539 | 4.83e-01 | NA | 2360 | 1590 | 770 |
| Latino/Mixed-American | Recorded Anxiety | Switching vs Continuation | BMI | Education_levelLess than a high school degree | 0.873 | 0.589 | 1.294 | 4.99e-01 | NA | 2360 | 1590 | 770 |
| Latino/Mixed-American | Recorded Anxiety | Switching vs Continuation | BMI | Education_levelTwelve or GED | 0.952 | 0.661 | 1.372 | 7.93e-01 | NA | 2360 | 1590 | 770 |
| Latino/Mixed-American | Recorded Anxiety | Switching vs Continuation | BMI | BMI | 1.019 | 0.933 | 1.113 | 6.83e-01 | 1.00e+00 | 2360 | 1590 | 770 |
| Latino/Mixed-American | Recorded Anxiety | Switching vs Continuation | Bipolar Disorder | (Intercept) | 0.638 | 0.404 | 1.006 | 5.33e-02 | NA | 2360 | 1590 | 770 |
| Latino/Mixed-American | Recorded Anxiety | Switching vs Continuation | Bipolar Disorder | age_at_first_AD | 0.994 | 0.988 | 1.000 | 6.57e-02 | NA | 2360 | 1590 | 770 |
| Latino/Mixed-American | Recorded Anxiety | Switching vs Continuation | Bipolar Disorder | sex_at_birthMale | 1.221 | 1.003 | 1.486 | 4.64e-02 | NA | 2360 | 1590 | 770 |
| Latino/Mixed-American | Recorded Anxiety | Switching vs Continuation | Bipolar Disorder | Income_quantitative | 0.960 | 0.912 | 1.011 | 1.22e-01 | NA | 2360 | 1590 | 770 |
| Latino/Mixed-American | Recorded Anxiety | Switching vs Continuation | Bipolar Disorder | Education_levelCollege Undergraduate | 1.123 | 0.818 | 1.542 | 4.74e-01 | NA | 2360 | 1590 | 770 |
| Latino/Mixed-American | Recorded Anxiety | Switching vs Continuation | Bipolar Disorder | Education_levelLess than a high school degree | 0.876 | 0.591 | 1.298 | 5.08e-01 | NA | 2360 | 1590 | 770 |
| Latino/Mixed-American | Recorded Anxiety | Switching vs Continuation | Bipolar Disorder | Education_levelTwelve or GED | 0.954 | 0.663 | 1.375 | 8.02e-01 | NA | 2360 | 1590 | 770 |
| Latino/Mixed-American | Recorded Anxiety | Switching vs Continuation | Bipolar Disorder | Bipolar Disorder | 1.013 | 0.919 | 1.117 | 7.93e-01 | 1.00e+00 | 2360 | 1590 | 770 |
| Latino/Mixed-American | Recorded Anxiety | Switching vs Continuation | Depression | (Intercept) | 0.639 | 0.405 | 1.009 | 5.45e-02 | NA | 2360 | 1590 | 770 |
| Latino/Mixed-American | Recorded Anxiety | Switching vs Continuation | Depression | age_at_first_AD | 0.994 | 0.988 | 1.000 | 6.28e-02 | NA | 2360 | 1590 | 770 |
| Latino/Mixed-American | Recorded Anxiety | Switching vs Continuation | Depression | sex_at_birthMale | 1.224 | 1.005 | 1.490 | 4.44e-02 | NA | 2360 | 1590 | 770 |
| Latino/Mixed-American | Recorded Anxiety | Switching vs Continuation | Depression | Income_quantitative | 0.963 | 0.915 | 1.014 | 1.55e-01 | NA | 2360 | 1590 | 770 |
| Latino/Mixed-American | Recorded Anxiety | Switching vs Continuation | Depression | Education_levelCollege Undergraduate | 1.115 | 0.812 | 1.531 | 5.02e-01 | NA | 2360 | 1590 | 770 |
| Latino/Mixed-American | Recorded Anxiety | Switching vs Continuation | Depression | Education_levelLess than a high school degree | 0.866 | 0.584 | 1.283 | 4.73e-01 | NA | 2360 | 1590 | 770 |
| Latino/Mixed-American | Recorded Anxiety | Switching vs Continuation | Depression | Education_levelTwelve or GED | 0.945 | 0.656 | 1.362 | 7.63e-01 | NA | 2360 | 1590 | 770 |
| Latino/Mixed-American | Recorded Anxiety | Switching vs Continuation | Depression | Depression | 1.144 | 1.037 | 1.263 | 7.55e-03 | 5.28e-02 | 2360 | 1590 | 770 |
| Latino/Mixed-American | Recorded Anxiety | Switching vs Continuation | Insomnia | (Intercept) | 0.643 | 0.407 | 1.014 | 5.75e-02 | NA | 2360 | 1590 | 770 |
| Latino/Mixed-American | Recorded Anxiety | Switching vs Continuation | Insomnia | age_at_first_AD | 0.994 | 0.988 | 1.000 | 6.63e-02 | NA | 2360 | 1590 | 770 |
| Latino/Mixed-American | Recorded Anxiety | Switching vs Continuation | Insomnia | sex_at_birthMale | 1.230 | 1.010 | 1.498 | 3.90e-02 | NA | 2360 | 1590 | 770 |
| Latino/Mixed-American | Recorded Anxiety | Switching vs Continuation | Insomnia | Income_quantitative | 0.961 | 0.913 | 1.011 | 1.26e-01 | NA | 2360 | 1590 | 770 |
| Latino/Mixed-American | Recorded Anxiety | Switching vs Continuation | Insomnia | Education_levelCollege Undergraduate | 1.108 | 0.807 | 1.523 | 5.26e-01 | NA | 2360 | 1590 | 770 |
| Latino/Mixed-American | Recorded Anxiety | Switching vs Continuation | Insomnia | Education_levelLess than a high school degree | 0.860 | 0.580 | 1.276 | 4.54e-01 | NA | 2360 | 1590 | 770 |
| Latino/Mixed-American | Recorded Anxiety | Switching vs Continuation | Insomnia | Education_levelTwelve or GED | 0.941 | 0.653 | 1.357 | 7.45e-01 | NA | 2360 | 1590 | 770 |
| Latino/Mixed-American | Recorded Anxiety | Switching vs Continuation | Insomnia | Insomnia | 1.103 | 0.998 | 1.218 | 5.43e-02 | 3.80e-01 | 2360 | 1590 | 770 |

| | | | | | | | | | | | | |
|---|---|---|---|---|---|---|---|---|---|---|---|---|
| Latino/Mixed-American | Recorded Anxiety | Switching vs Continuation | Migraine | (Intercept) | 0.639 | 0.405 | 1.008 | 5.40e-02 | NA | 2360 | 1590 | 770 |
| Latino/Mixed-American | Recorded Anxiety | Switching vs Continuation | Migraine | age_at_first_AD | 0.994 | 0.988 | 1.000 | 6.68e-02 | NA | 2360 | 1590 | 770 |
| Latino/Mixed-American | Recorded Anxiety | Switching vs Continuation | Migraine | sex_at_birthMale | 1.220 | 1.002 | 1.485 | 4.74e-02 | NA | 2360 | 1590 | 770 |
| Latino/Mixed-American | Recorded Anxiety | Switching vs Continuation | Migraine | Income_quantitative | 0.960 | 0.912 | 1.010 | 1.17e-01 | NA | 2360 | 1590 | 770 |
| Latino/Mixed-American | Recorded Anxiety | Switching vs Continuation | Migraine | Education_levelCollege Undergraduate | 1.123 | 0.818 | 1.543 | 4.72e-01 | NA | 2360 | 1590 | 770 |
| Latino/Mixed-American | Recorded Anxiety | Switching vs Continuation | Migraine | Education_levelLess than a high school degree | 0.873 | 0.589 | 1.294 | 4.98e-01 | NA | 2360 | 1590 | 770 |
| Latino/Mixed-American | Recorded Anxiety | Switching vs Continuation | Migraine | Education_levelTwelve or GED | 0.952 | 0.661 | 1.371 | 7.91e-01 | NA | 2360 | 1590 | 770 |
| Latino/Mixed-American | Recorded Anxiety | Switching vs Continuation | Migraine | Migraine | 1.022 | 0.934 | 1.118 | 6.40e-01 | 1.00e+00 | 2360 | 1590 | 770 |
| Latino/Mixed-American | Recorded Anxiety | Switching vs Continuation | Schizophrenia | (Intercept) | 0.634 | 0.402 | 1.001 | 5.03e-02 | NA | 2360 | 1590 | 770 |
| Latino/Mixed-American | Recorded Anxiety | Switching vs Continuation | Schizophrenia | age_at_first_AD | 0.994 | 0.988 | 1.000 | 6.33e-02 | NA | 2360 | 1590 | 770 |
| Latino/Mixed-American | Recorded Anxiety | Switching vs Continuation | Schizophrenia | sex_at_birthMale | 1.223 | 1.005 | 1.488 | 4.49e-02 | NA | 2360 | 1590 | 770 |
| Latino/Mixed-American | Recorded Anxiety | Switching vs Continuation | Schizophrenia | Income_quantitative | 0.961 | 0.913 | 1.012 | 1.32e-01 | NA | 2360 | 1590 | 770 |
| Latino/Mixed-American | Recorded Anxiety | Switching vs Continuation | Schizophrenia | Education_levelCollege Undergraduate | 1.129 | 0.822 | 1.551 | 4.53e-01 | NA | 2360 | 1590 | 770 |
| Latino/Mixed-American | Recorded Anxiety | Switching vs Continuation | Schizophrenia | Education_levelLess than a high school degree | 0.879 | 0.593 | 1.303 | 5.21e-01 | NA | 2360 | 1590 | 770 |
| Latino/Mixed-American | Recorded Anxiety | Switching vs Continuation | Schizophrenia | Education_levelTwelve or GED | 0.961 | 0.667 | 1.384 | 8.30e-01 | NA | 2360 | 1590 | 770 |
| Latino/Mixed-American | Recorded Anxiety | Switching vs Continuation | Schizophrenia | Schizophrenia | 1.048 | 0.961 | 1.142 | 2.91e-01 | 1.00e+00 | 2360 | 1590 | 770 |

**Supplementary Table 7. Multivariable logistic regression results for PGS associations with antidepressant switching (sensitivity definition) across clinical subgroups in the Pharmlines cohort (European ancestry participants)**. Each model estimates the association between 7 PGS traits and the binary outcome switching versus continuation — where switching is operationalised using the sensitivity definition, restricting switching events to those occurring within six months of index antidepressant initiation (retaining the same 90-day post-discontinuation switching window and <28-day cross-taper exclusion as the primary definition used in Supplementary Table 5 — fitted separately within each combination of clinical group (Full Cohort; and subgroups defined by recorded depression status), and PGS exposure. Within each model, one PGS exposure is modelled alongside a common covariate set (age at first antidepressant, genetic sex, income, and educational attainment). All PGS were residualised on the top 20 within-European-ancestry principal components and the residuals standardised to mean 0, SD 1; each odds ratio therefore reflects the change in odds per one-standard-deviation increase in the PC-adjusted PGS. Because PCs were regressed out at this stage, they are not included in the model covariate set.
Columns: Population = inferred genetic ancestry (all European); ClinicalGroup = cohort or clinical subgroup; Outcome = the binary contrast modelled; PGS = the PGS trait for that model; variable = the individual model term; odds_ratio = exponentiated coefficient; lower_95/upper_95 = 95% confidence interval bounds for the odds ratio; p_value = nominal (uncorrected) p-value; Bonf_P = Bonferroni-corrected p-value (capped at 1.00); N_Total = model sample size; N_Reference = number in the reference (continuation) group; N_Case = number in the comparison (discontinuation or switching) group. Odds ratios >1 indicate higher odds of the comparison outcome relative to continuation. Bonferroni correction was applied within each specification (ancestry × clinical group × outcome contrast) for the max 7 PGS traits tested. P-values reported as "<2.2e-16" reflect R's floating-point reporting limit.
Reference (omitted) categories: Sex at birth: Female.

| Population | ClinicalGroup | Outcome | PGS | variable | odds_ratio | lower_95 | upper_95 | p_value | Bonf_p | N_Total | N_Reference | N_Case |
|---|---|---|---|---|---|---|---|---|---|---|---|---|
| European | Full Cohort | Switching vs Continuation | ADHD | (Intercept) | 0.446 | 0.287 | 0.694 | 3.37e-04 | 2.36e-03 | 5153 | 4445 | 708 |
| European | Full Cohort | Switching vs Continuation | ADHD | age_at_first_AD | 0.986 | 0.979 | 0.992 | 2.84e-05 | 1.99e-04 | 5153 | 4445 | 708 |
| European | Full Cohort | Switching vs Continuation | ADHD | sex_at_birth | 1.374 | 1.158 | 1.632 | 2.80e-04 | 1.96e-03 | 5153 | 4445 | 708 |
| European | Full Cohort | Switching vs Continuation | ADHD | Income_level | 1.000 | 1.000 | 1.000 | 5.91e-03 | 4.14e-02 | 5153 | 4445 | 708 |
| European | Full Cohort | Switching vs Continuation | ADHD | Education_level | 0.988 | 0.967 | 1.008 | 2.42e-01 | 1.00e+00 | 5153 | 4445 | 708 |
| European | Full Cohort | Switching vs Continuation | ADHD | ADHD | 1.087 | 1.002 | 1.178 | 4.45e-02 | 3.11e-01 | 5153 | 4445 | 708 |
| European | Full Cohort | Switching vs Continuation | BMI | (Intercept) | 0.468 | 0.301 | 0.727 | 7.29e-04 | 5.10e-03 | 5153 | 4445 | 708 |
| European | Full Cohort | Switching vs Continuation | BMI | age_at_first_AD | 0.986 | 0.979 | 0.992 | 2.06e-05 | 1.44e-04 | 5153 | 4445 | 708 |
| European | Full Cohort | Switching vs Continuation | BMI | sex_at_birth | 1.376 | 1.159 | 1.634 | 2.62e-04 | 1.83e-03 | 5153 | 4445 | 708 |
| European | Full Cohort | Switching vs Continuation | BMI | Income_level | 1.000 | 1.000 | 1.000 | 4.80e-03 | 3.36e-02 | 5153 | 4445 | 708 |
| European | Full Cohort | Switching vs Continuation | BMI | Education_level | 0.986 | 0.965 | 1.006 | 1.74e-01 | 1.00e+00 | 5153 | 4445 | 708 |
| European | Full Cohort | Switching vs Continuation | BMI | BMI | 0.983 | 0.908 | 1.063 | 6.66e-01 | 1.00e+00 | 5153 | 4445 | 708 |
| European | Full Cohort | Switching vs Continuation | Bipolar Disorder | (Intercept) | 0.460 | 0.296 | 0.715 | 5.54e-04 | 3.88e-03 | 5153 | 4445 | 708 |
| European | Full Cohort | Switching vs Continuation | Bipolar Disorder | age_at_first_AD | 0.986 | 0.979 | 0.992 | 2.63e-05 | 1.84e-04 | 5153 | 4445 | 708 |
| European | Full Cohort | Switching vs Continuation | Bipolar Disorder | sex_at_birth | 1.379 | 1.161 | 1.637 | 2.46e-04 | 1.72e-03 | 5153 | 4445 | 708 |
| European | Full Cohort | Switching vs Continuation | Bipolar Disorder | Income_level | 1.000 | 1.000 | 1.000 | 5.01e-03 | 3.51e-02 | 5153 | 4445 | 708 |
| European | Full Cohort | Switching vs Continuation | Bipolar Disorder | Education_level | 0.986 | 0.965 | 1.006 | 1.76e-01 | 1.00e+00 | 5153 | 4445 | 708 |
| European | Full Cohort | Switching vs Continuation | Bipolar Disorder | Bipolar Disorder | 1.131 | 1.044 | 1.225 | 2.47e-03 | 1.73e-02 | 5153 | 4445 | 708 |
| European | Full Cohort | Switching vs Continuation | Depression | (Intercept) | 0.438 | 0.281 | 0.681 | 2.45e-04 | 1.71e-03 | 5153 | 4445 | 708 |
| European | Full Cohort | Switching vs Continuation | Depression | age_at_first_AD | 0.986 | 0.980 | 0.993 | 5.71e-05 | 3.99e-04 | 5153 | 4445 | 708 |
| European | Full Cohort | Switching vs Continuation | Depression | sex_at_birth | 1.370 | 1.154 | 1.627 | 3.23e-04 | 2.26e-03 | 5153 | 4445 | 708 |
| European | Full Cohort | Switching vs Continuation | Depression | Income_level | 1.000 | 1.000 | 1.000 | 5.16e-03 | 3.61e-02 | 5153 | 4445 | 708 |
| European | Full Cohort | Switching vs Continuation | Depression | Education_level | 0.987 | 0.967 | 1.008 | 2.28e-01 | 1.00e+00 | 5153 | 4445 | 708 |
| European | Full Cohort | Switching vs Continuation | Depression | Depression | 1.152 | 1.063 | 1.249 | 5.52e-04 | 3.86e-03 | 5153 | 4445 | 708 |
| European | Full Cohort | Switching vs Continuation | Insomnia | (Intercept) | 0.443 | 0.285 | 0.688 | 2.91e-04 | 2.04e-03 | 5153 | 4445 | 708 |
| European | Full Cohort | Switching vs Continuation | Insomnia | age_at_first_AD | 0.986 | 0.979 | 0.993 | 3.43e-05 | 2.40e-04 | 5153 | 4445 | 708 |
| European | Full Cohort | Switching vs Continuation | Insomnia | sex_at_birth | 1.374 | 1.157 | 1.631 | 2.88e-04 | 2.02e-03 | 5153 | 4445 | 708 |
| European | Full Cohort | Switching vs Continuation | Insomnia | Income_level | 1.000 | 1.000 | 1.000 | 5.88e-03 | 4.12e-02 | 5153 | 4445 | 708 |
| European | Full Cohort | Switching vs Continuation | Insomnia | Education_level | 0.987 | 0.967 | 1.008 | 2.33e-01 | 1.00e+00 | 5153 | 4445 | 708 |
| European | Full Cohort | Switching vs Continuation | Insomnia | Insomnia | 1.130 | 1.043 | 1.225 | 2.73e-03 | 1.91e-02 | 5153 | 4445 | 708 |
| European | Full Cohort | Switching vs Continuation | Migraine | (Intercept) | 0.463 | 0.298 | 0.719 | 6.00e-04 | 4.20e-03 | 5153 | 4445 | 708 |
| European | Full Cohort | Switching vs Continuation | Migraine | age_at_first_AD | 0.986 | 0.979 | 0.992 | 2.13e-05 | 1.49e-04 | 5153 | 4445 | 708 |
| European | Full Cohort | Switching vs Continuation | Migraine | sex_at_birth | 1.376 | 1.159 | 1.633 | 2.68e-04 | 1.87e-03 | 5153 | 4445 | 708 |
| European | Full Cohort | Switching vs Continuation | Migraine | Income_level | 1.000 | 1.000 | 1.000 | 5.07e-03 | 3.55e-02 | 5153 | 4445 | 708 |
| European | Full Cohort | Switching vs Continuation | Migraine | Education_level | 0.986 | 0.966 | 1.007 | 1.88e-01 | 1.00e+00 | 5153 | 4445 | 708 |
| European | Full Cohort | Switching vs Continuation | Migraine | Migraine | 1.027 | 0.949 | 1.111 | 5.08e-01 | 1.00e+00 | 5153 | 4445 | 708 |
| European | Full Cohort | Switching vs Continuation | Schizophrenia | (Intercept) | 0.455 | 0.293 | 0.707 | 4.54e-04 | 3.18e-03 | 5153 | 4445 | 708 |

| | | | | | | | | | | | | |
|---|---|---|---|---|---|---|---|---|---|---|---|---|
| European | Full Cohort | Switching vs Continuation | Schizophrenia | age_at_first_AD | 0.986 | 0.979 | 0.993 | 3.88e-05 | 2.72e-04 | 5153 | 4445 | 708 |
| European | Full Cohort | Switching vs Continuation | Schizophrenia | sex_at_birth | 1.372 | 1.156 | 1.629 | 3.05e-04 | 2.13e-03 | 5153 | 4445 | 708 |
| European | Full Cohort | Switching vs Continuation | Schizophrenia | Income_level | 1.000 | 1.000 | 1.000 | 4.79e-03 | 3.35e-02 | 5153 | 4445 | 708 |
| European | Full Cohort | Switching vs Continuation | Schizophrenia | Education_level | 0.986 | 0.965 | 1.007 | 1.80e-01 | 1.00e+00 | 5153 | 4445 | 708 |
| European | Full Cohort | Switching vs Continuation | Schizophrenia | Schizophrenia | 1.120 | 1.035 | 1.212 | 4.83e-03 | 3.38e-02 | 5153 | 4445 | 708 |
| European | Recorded Depression | Switching vs Continuation | ADHD | (Intercept) | 0.695 | 0.373 | 1.298 | 2.54e-01 | 1.00e+00 | 2073 | 1692 | 381 |
| European | Recorded Depression | Switching vs Continuation | ADHD | age_at_first_AD | 0.989 | 0.979 | 0.999 | 3.06e-02 | 2.14e-01 | 2073 | 1692 | 381 |
| European | Recorded Depression | Switching vs Continuation | ADHD | sex_at_birth | 1.276 | 0.994 | 1.639 | 5.58e-02 | 3.91e-01 | 2073 | 1692 | 381 |
| European | Recorded Depression | Switching vs Continuation | ADHD | Income_level | 1.000 | 1.000 | 1.000 | 5.22e-02 | 3.66e-01 | 2073 | 1692 | 381 |
| European | Recorded Depression | Switching vs Continuation | ADHD | Education_level | 0.973 | 0.945 | 1.002 | 6.83e-02 | 4.78e-01 | 2073 | 1692 | 381 |
| European | Recorded Depression | Switching vs Continuation | ADHD | ADHD | 0.980 | 0.874 | 1.100 | 7.32e-01 | 1.00e+00 | 2073 | 1692 | 381 |
| European | Recorded Depression | Switching vs Continuation | BMI | (Intercept) | 0.684 | 0.367 | 1.277 | 2.33e-01 | 1.00e+00 | 2073 | 1692 | 381 |
| European | Recorded Depression | Switching vs Continuation | BMI | age_at_first_AD | 0.989 | 0.979 | 0.999 | 3.18e-02 | 2.23e-01 | 2073 | 1692 | 381 |
| European | Recorded Depression | Switching vs Continuation | BMI | sex_at_birth | 1.275 | 0.993 | 1.637 | 5.67e-02 | 3.97e-01 | 2073 | 1692 | 381 |
| European | Recorded Depression | Switching vs Continuation | BMI | Income_level | 1.000 | 1.000 | 1.000 | 5.29e-02 | 3.70e-01 | 2073 | 1692 | 381 |
| European | Recorded Depression | Switching vs Continuation | BMI | Education_level | 0.974 | 0.946 | 1.003 | 7.75e-02 | 5.43e-01 | 2073 | 1692 | 381 |
| European | Recorded Depression | Switching vs Continuation | BMI | BMI | 1.021 | 0.915 | 1.139 | 7.14e-01 | 1.00e+00 | 2073 | 1692 | 381 |
| European | Recorded Depression | Switching vs Continuation | Bipolar Disorder | (Intercept) | 0.679 | 0.364 | 1.267 | 2.24e-01 | 1.00e+00 | 2073 | 1692 | 381 |
| European | Recorded Depression | Switching vs Continuation | Bipolar Disorder | age_at_first_AD | 0.989 | 0.979 | 0.999 | 3.24e-02 | 2.27e-01 | 2073 | 1692 | 381 |
| European | Recorded Depression | Switching vs Continuation | Bipolar Disorder | sex_at_birth | 1.265 | 0.985 | 1.625 | 6.54e-02 | 4.58e-01 | 2073 | 1692 | 381 |
| European | Recorded Depression | Switching vs Continuation | Bipolar Disorder | Income_level | 1.000 | 1.000 | 1.000 | 6.94e-02 | 4.86e-01 | 2073 | 1692 | 381 |
| European | Recorded Depression | Switching vs Continuation | Bipolar Disorder | Education_level | 0.973 | 0.945 | 1.001 | 6.25e-02 | 4.37e-01 | 2073 | 1692 | 381 |
| European | Recorded Depression | Switching vs Continuation | Bipolar Disorder | Bipolar Disorder | 1.150 | 1.025 | 1.289 | 1.69e-02 | 1.18e-01 | 2073 | 1692 | 381 |
| European | Recorded Depression | Switching vs Continuation | Depression | (Intercept) | 0.664 | 0.356 | 1.240 | 1.99e-01 | 1.00e+00 | 2073 | 1692 | 381 |
| European | Recorded Depression | Switching vs Continuation | Depression | age_at_first_AD | 0.989 | 0.979 | 0.999 | 3.77e-02 | 2.64e-01 | 2073 | 1692 | 381 |
| European | Recorded Depression | Switching vs Continuation | Depression | sex_at_birth | 1.262 | 0.982 | 1.621 | 6.90e-02 | 4.83e-01 | 2073 | 1692 | 381 |
| European | Recorded Depression | Switching vs Continuation | Depression | Income_level | 1.000 | 1.000 | 1.000 | 5.14e-02 | 3.60e-01 | 2073 | 1692 | 381 |
| European | Recorded Depression | Switching vs Continuation | Depression | Education_level | 0.975 | 0.947 | 1.004 | 9.10e-02 | 6.37e-01 | 2073 | 1692 | 381 |
| European | Recorded Depression | Switching vs Continuation | Depression | Depression | 1.095 | 0.976 | 1.229 | 1.22e-01 | 8.56e-01 | 2073 | 1692 | 381 |
| European | Recorded Depression | Switching vs Continuation | Insomnia | (Intercept) | 0.685 | 0.367 | 1.277 | 2.33e-01 | 1.00e+00 | 2073 | 1692 | 381 |
| European | Recorded Depression | Switching vs Continuation | Insomnia | age_at_first_AD | 0.989 | 0.979 | 0.999 | 3.16e-02 | 2.21e-01 | 2073 | 1692 | 381 |
| European | Recorded Depression | Switching vs Continuation | Insomnia | sex_at_birth | 1.274 | 0.992 | 1.636 | 5.74e-02 | 4.02e-01 | 2073 | 1692 | 381 |
| European | Recorded Depression | Switching vs Continuation | Insomnia | Income_level | 1.000 | 1.000 | 1.000 | 5.32e-02 | 3.72e-01 | 2073 | 1692 | 381 |
| European | Recorded Depression | Switching vs Continuation | Insomnia | Education_level | 0.974 | 0.946 | 1.003 | 7.63e-02 | 5.34e-01 | 2073 | 1692 | 381 |
| European | Recorded Depression | Switching vs Continuation | Insomnia | Insomnia | 1.023 | 0.915 | 1.145 | 6.86e-01 | 1.00e+00 | 2073 | 1692 | 381 |
| European | Recorded Depression | Switching vs Continuation | Migraine | (Intercept) | 0.689 | 0.370 | 1.283 | 2.40e-01 | 1.00e+00 | 2073 | 1692 | 381 |
| European | Recorded Depression | Switching vs Continuation | Migraine | age_at_first_AD | 0.989 | 0.979 | 0.999 | 3.10e-02 | 2.17e-01 | 2073 | 1692 | 381 |
| European | Recorded Depression | Switching vs Continuation | Migraine | sex_at_birth | 1.275 | 0.993 | 1.637 | 5.71e-02 | 3.99e-01 | 2073 | 1692 | 381 |
| European | Recorded Depression | Switching vs Continuation | Migraine | Income_level | 1.000 | 1.000 | 1.000 | 5.34e-02 | 3.74e-01 | 2073 | 1692 | 381 |
| European | Recorded Depression | Switching vs Continuation | Migraine | Education_level | 0.974 | 0.946 | 1.002 | 7.29e-02 | 5.10e-01 | 2073 | 1692 | 381 |
| European | Recorded Depression | Switching vs Continuation | Migraine | Migraine | 1.011 | 0.904 | 1.131 | 8.48e-01 | 1.00e+00 | 2073 | 1692 | 381 |
| European | Recorded Depression | Switching vs Continuation | Schizophrenia | (Intercept) | 0.683 | 0.366 | 1.272 | 2.29e-01 | 1.00e+00 | 2073 | 1692 | 381 |
| European | Recorded Depression | Switching vs Continuation | Schizophrenia | age_at_first_AD | 0.989 | 0.979 | 0.999 | 3.44e-02 | 2.41e-01 | 2073 | 1692 | 381 |
| European | Recorded Depression | Switching vs Continuation | Schizophrenia | sex_at_birth | 1.267 | 0.987 | 1.628 | 6.34e-02 | 4.44e-01 | 2073 | 1692 | 381 |
| European | Recorded Depression | Switching vs Continuation | Schizophrenia | Income_level | 1.000 | 1.000 | 1.000 | 5.55e-02 | 3.89e-01 | 2073 | 1692 | 381 |
| European | Recorded Depression | Switching vs Continuation | Schizophrenia | Education_level | 0.974 | 0.946 | 1.002 | 6.99e-02 | 4.89e-01 | 2073 | 1692 | 381 |
| European | Recorded Depression | Switching vs Continuation | Schizophrenia | Schizophrenia | 1.074 | 0.962 | 1.198 | 2.04e-01 | 1.00e+00 | 2073 | 1692 | 381 |
| European | No Recorded Depression | Switching vs Continuation | ADHD | (Intercept) | 0.227 | 0.118 | 0.436 | 8.41e-06 | 5.89e-05 | 3024 | 2713 | 311 |
| European | No Recorded Depression | Switching vs Continuation | ADHD | age_at_first_AD | 0.990 | 0.981 | 1.000 | 4.02e-02 | 2.81e-01 | 3024 | 2713 | 311 |

| | | | | | | | | | | | | |
|---|---|---|---|---|---|---|---|---|---|---|---|---|
| European | No Recorded Depression | Switching vs Continuation | ADHD | sex_at_birth | 1.686 | 1.320 | 2.152 | 2.79e-05 | 1.95e-04 | 3024 | 2713 | 311 |
| European | No Recorded Depression | Switching vs Continuation | ADHD | Income_level | 1.000 | 1.000 | 1.000 | 9.93e-02 | 6.95e-01 | 3024 | 2713 | 311 |
| European | No Recorded Depression | Switching vs Continuation | ADHD | Education_level | 0.990 | 0.959 | 1.022 | 5.34e-01 | 1.00e+00 | 3024 | 2713 | 311 |
| European | No Recorded Depression | Switching vs Continuation | ADHD | ADHD | 1.199 | 1.064 | 1.352 | 2.98e-03 | 2.09e-02 | 3024 | 2713 | 311 |
| European | No Recorded Depression | Switching vs Continuation | BMI | (Intercept) | 0.258 | 0.135 | 0.495 | 4.58e-05 | 3.20e-04 | 3024 | 2713 | 311 |
| European | No Recorded Depression | Switching vs Continuation | BMI | age_at_first_AD | 0.990 | 0.980 | 0.999 | 2.85e-02 | 2.00e-01 | 3024 | 2713 | 311 |
| European | No Recorded Depression | Switching vs Continuation | BMI | sex_at_birth | 1.682 | 1.318 | 2.146 | 2.95e-05 | 2.06e-04 | 3024 | 2713 | 311 |
| European | No Recorded Depression | Switching vs Continuation | BMI | Income_level | 1.000 | 1.000 | 1.000 | 7.25e-02 | 5.08e-01 | 3024 | 2713 | 311 |
| European | No Recorded Depression | Switching vs Continuation | BMI | Education_level | 0.985 | 0.955 | 1.017 | 3.58e-01 | 1.00e+00 | 3024 | 2713 | 311 |
| European | No Recorded Depression | Switching vs Continuation | BMI | BMI | 0.921 | 0.819 | 1.035 | 1.65e-01 | 1.00e+00 | 3024 | 2713 | 311 |
| European | No Recorded Depression | Switching vs Continuation | Bipolar Disorder | (Intercept) | 0.248 | 0.129 | 0.475 | 2.64e-05 | 1.85e-04 | 3024 | 2713 | 311 |
| European | No Recorded Depression | Switching vs Continuation | Bipolar Disorder | age_at_first_AD | 0.990 | 0.981 | 0.999 | 3.39e-02 | 2.37e-01 | 3024 | 2713 | 311 |
| European | No Recorded Depression | Switching vs Continuation | Bipolar Disorder | sex_at_birth | 1.691 | 1.325 | 2.158 | 2.43e-05 | 1.70e-04 | 3024 | 2713 | 311 |
| European | No Recorded Depression | Switching vs Continuation | Bipolar Disorder | Income_level | 1.000 | 1.000 | 1.000 | 7.38e-02 | 5.16e-01 | 3024 | 2713 | 311 |
| European | No Recorded Depression | Switching vs Continuation | Bipolar Disorder | Education_level | 0.987 | 0.957 | 1.019 | 4.18e-01 | 1.00e+00 | 3024 | 2713 | 311 |
| European | No Recorded Depression | Switching vs Continuation | Bipolar Disorder | Bipolar Disorder | 1.088 | 0.969 | 1.223 | 1.55e-01 | 1.00e+00 | 3024 | 2713 | 311 |
| European | No Recorded Depression | Switching vs Continuation | Depression | (Intercept) | 0.234 | 0.122 | 0.449 | 1.28e-05 | 8.94e-05 | 3024 | 2713 | 311 |
| European | No Recorded Depression | Switching vs Continuation | Depression | age_at_first_AD | 0.991 | 0.981 | 1.000 | 4.74e-02 | 3.32e-01 | 3024 | 2713 | 311 |
| European | No Recorded Depression | Switching vs Continuation | Depression | sex_at_birth | 1.688 | 1.322 | 2.155 | 2.63e-05 | 1.84e-04 | 3024 | 2713 | 311 |
| European | No Recorded Depression | Switching vs Continuation | Depression | Income_level | 1.000 | 1.000 | 1.000 | 8.72e-02 | 6.10e-01 | 3024 | 2713 | 311 |
| European | No Recorded Depression | Switching vs Continuation | Depression | Education_level | 0.988 | 0.957 | 1.019 | 4.31e-01 | 1.00e+00 | 3024 | 2713 | 311 |
| European | No Recorded Depression | Switching vs Continuation | Depression | Depression | 1.177 | 1.046 | 1.325 | 6.84e-03 | 4.79e-02 | 3024 | 2713 | 311 |
| European | No Recorded Depression | Switching vs Continuation | Insomnia | (Intercept) | 0.227 | 0.118 | 0.435 | 7.88e-06 | 5.52e-05 | 3024 | 2713 | 311 |
| European | No Recorded Depression | Switching vs Continuation | Insomnia | age_at_first_AD | 0.990 | 0.981 | 1.000 | 4.52e-02 | 3.16e-01 | 3024 | 2713 | 311 |
| European | No Recorded Depression | Switching vs Continuation | Insomnia | sex_at_birth | 1.674 | 1.311 | 2.138 | 3.60e-05 | 2.52e-04 | 3024 | 2713 | 311 |
| European | No Recorded Depression | Switching vs Continuation | Insomnia | Income_level | 1.000 | 1.000 | 1.000 | 9.93e-02 | 6.95e-01 | 3024 | 2713 | 311 |
| European | No Recorded Depression | Switching vs Continuation | Insomnia | Education_level | 0.989 | 0.958 | 1.021 | 4.89e-01 | 1.00e+00 | 3024 | 2713 | 311 |
| European | No Recorded Depression | Switching vs Continuation | Insomnia | Insomnia | 1.231 | 1.093 | 1.387 | 6.42e-04 | 4.50e-03 | 3024 | 2713 | 311 |
| European | No Recorded Depression | Switching vs Continuation | Migraine | (Intercept) | 0.248 | 0.129 | 0.475 | 2.57e-05 | 1.80e-04 | 3024 | 2713 | 311 |
| European | No Recorded Depression | Switching vs Continuation | Migraine | age_at_first_AD | 0.990 | 0.980 | 0.999 | 3.14e-02 | 2.20e-01 | 3024 | 2713 | 311 |
| European | No Recorded Depression | Switching vs Continuation | Migraine | sex_at_birth | 1.681 | 1.317 | 2.146 | 2.99e-05 | 2.09e-04 | 3024 | 2713 | 311 |
| European | No Recorded Depression | Switching vs Continuation | Migraine | Income_level | 1.000 | 1.000 | 1.000 | 8.16e-02 | 5.71e-01 | 3024 | 2713 | 311 |
| European | No Recorded Depression | Switching vs Continuation | Migraine | Education_level | 0.987 | 0.957 | 1.019 | 4.20e-01 | 1.00e+00 | 3024 | 2713 | 311 |
| European | No Recorded Depression | Switching vs Continuation | Migraine | Migraine | 1.042 | 0.928 | 1.170 | 4.86e-01 | 1.00e+00 | 3024 | 2713 | 311 |
| European | No Recorded Depression | Switching vs Continuation | Schizophrenia | (Intercept) | 0.242 | 0.126 | 0.465 | 1.97e-05 | 1.38e-04 | 3024 | 2713 | 311 |
| European | No Recorded Depression | Switching vs Continuation | Schizophrenia | age_at_first_AD | 0.990 | 0.981 | 1.000 | 4.35e-02 | 3.04e-01 | 3024 | 2713 | 311 |
| European | No Recorded Depression | Switching vs Continuation | Schizophrenia | sex_at_birth | 1.683 | 1.319 | 2.148 | 2.89e-05 | 2.02e-04 | 3024 | 2713 | 311 |
| European | No Recorded Depression | Switching vs Continuation | Schizophrenia | Income_level | 1.000 | 1.000 | 1.000 | 7.60e-02 | 5.32e-01 | 3024 | 2713 | 311 |
| European | No Recorded Depression | Switching vs Continuation | Schizophrenia | Education_level | 0.987 | 0.956 | 1.018 | 4.04e-01 | 1.00e+00 | 3024 | 2713 | 311 |
| European | No Recorded Depression | Switching vs Continuation | Schizophrenia | Schizophrenia | 1.155 | 1.027 | 1.299 | 1.58e-02 | 1.11e-01 | 3024 | 2713 | 311 |

**Supplementary Table S8. Multivariable logistic regression results for PGS associations with antidepressant discontinuation (sensitivity definition) across ancestry groups and clinical subgroups in All of Us.** Each model estimates the association between 7 PGS traits and the binary outcome discontinuation versus continuation, fitted separately within each combination of genetically inferred ancestry (European, African, Latino/Mixed-American), clinical group (Full Cohort; subgroups defined by recorded depression status, recorded anxiety, and CIDI-SF–derived MDD classification), and PGS exposure. Individuals who met augmentation or combination criteria at any point during follow-up were excluded from this analysis, so that the discontinuation-versus-continuation contrast is estimated among those who did not escalate treatment via augmentation or combination, and only had short antidepresant monotherapy episodes throughout their full prescribing history. Within each model, one PGS exposure is modelled alongside a common covariate set (age at first antidepressant, sex at birth, income, and educational attainment). All PGS were residualised on the top 16 global genetic ancestry principal components and the residuals standardised to mean 0, SD 1; each odds ratio therefore reflects the change in odds per one-standard-deviation increase in the PC-adjusted PGS. Because ancestry PCs were regressed out at this stage, they are not included in the model covariate set. Columns: Population = inferred genetic ancestry; ClinicalGroup = cohort or clinical subgroup; Outcome = the binary contrast modelled; PGS = the PGS trait for that model; variable = the individual model term; odds_ratio = exponentiated coefficient; lower_95/upper_95 = 95% confidence interval bounds for the odds ratio; p_value = nominal (uncorrected) p-value; Bonf_P = Bonferroni-corrected p-value (capped at 1.00); N_Total = model sample size; N_Reference = number in the reference (continuation) group; N_Case = number in the discontinuation group. Odds ratios >1 indicate higher odds of discontinuation relative to continuation. Bonferroni correction was applied within each specification (ancestry × clinical group) for the max 7 PGS traits tested. P-values reported as "<2.2e-16" reflect R's floating-point reporting limit.
Reference (omitted) categories: Sex at birth: Female; Income: lowest band (<$25k); Education level: lowest band (less than high school / fewer than twelve years).

| Population | ClinicalGroup | Outcome | PGS | variable | odds_ratio | lower_95 | upper_95 | p_value | Bonf_P | N_Total | N_Reference | N_Case |
|---|---|---|---|---|---|---|---|---|---|---|---|---|
| European | Full Cohort | Discontinuation vs Continuation | ADHD | (Intercept) | 0.247 | 0.216 | 0.283 | <2.2e-16 | NA | 27749 | 21256 | 6493 |
| European | Full Cohort | Discontinuation vs Continuation | ADHD | age_at_first_AD | 1.007 | 1.005 | 1.009 | 1.40e-14 | NA | 27749 | 21256 | 6493 |
| European | Full Cohort | Discontinuation vs Continuation | ADHD | sex_at_birthMale | 1.142 | 1.076 | 1.213 | 1.37e-05 | NA | 27749 | 21256 | 6493 |
| European | Full Cohort | Discontinuation vs Continuation | ADHD | Income_quantitative | 0.934 | 0.921 | 0.947 | <2.2e-16 | NA | 27749 | 21256 | 6493 |
| European | Full Cohort | Discontinuation vs Continuation | ADHD | Education_levelCollege Undergraduate | 1.108 | 1.035 | 1.186 | 3.11e-03 | NA | 27749 | 21256 | 6493 |
| European | Full Cohort | Discontinuation vs Continuation | ADHD | Education_levelLess than a high school degree | 1.658 | 1.340 | 2.052 | 3.24e-06 | NA | 27749 | 21256 | 6493 |
| European | Full Cohort | Discontinuation vs Continuation | ADHD | Education_levelTwelve or GED | 1.327 | 1.199 | 1.469 | 4.48e-08 | NA | 27749 | 21256 | 6493 |
| European | Full Cohort | Discontinuation vs Continuation | ADHD | ADHD | 1.040 | 1.010 | 1.070 | 7.59e-03 | 5.31e-02 | 27749 | 21256 | 6493 |
| European | Full Cohort | Discontinuation vs Continuation | BMI | (Intercept) | 0.249 | 0.218 | 0.285 | <2.2e-16 | NA | 27749 | 21256 | 6493 |
| European | Full Cohort | Discontinuation vs Continuation | BMI | age_at_first_AD | 1.007 | 1.005 | 1.009 | 3.17e-14 | NA | 27749 | 21256 | 6493 |
| European | Full Cohort | Discontinuation vs Continuation | BMI | sex_at_birthMale | 1.140 | 1.074 | 1.211 | 1.74e-05 | NA | 27749 | 21256 | 6493 |
| European | Full Cohort | Discontinuation vs Continuation | BMI | Income_quantitative | 0.932 | 0.919 | 0.946 | <2.2e-16 | NA | 27749 | 21256 | 6493 |
| European | Full Cohort | Discontinuation vs Continuation | BMI | Education_levelCollege Undergraduate | 1.115 | 1.042 | 1.193 | 1.66e-03 | NA | 27749 | 21256 | 6493 |
| European | Full Cohort | Discontinuation vs Continuation | BMI | Education_levelLess than a high school degree | 1.697 | 1.372 | 2.099 | 1.08e-06 | NA | 27749 | 21256 | 6493 |
| European | Full Cohort | Discontinuation vs Continuation | BMI | Education_levelTwelve or GED | 1.343 | 1.213 | 1.486 | 1.21e-08 | NA | 27749 | 21256 | 6493 |
| European | Full Cohort | Discontinuation vs Continuation | BMI | BMI | 0.995 | 0.968 | 1.024 | 7.47e-01 | 1.00e+00 | 27749 | 21256 | 6493 |
| European | Full Cohort | Discontinuation vs Continuation | Bipolar Disorder | (Intercept) | 0.248 | 0.217 | 0.284 | <2.2e-16 | NA | 27749 | 21256 | 6493 |
| European | Full Cohort | Discontinuation vs Continuation | Bipolar Disorder | age_at_first_AD | 1.007 | 1.005 | 1.009 | 3.44e-14 | NA | 27749 | 21256 | 6493 |
| European | Full Cohort | Discontinuation vs Continuation | Bipolar Disorder | sex_at_birthMale | 1.139 | 1.073 | 1.210 | 1.96e-05 | NA | 27749 | 21256 | 6493 |
| European | Full Cohort | Discontinuation vs Continuation | Bipolar Disorder | Income_quantitative | 0.933 | 0.920 | 0.946 | <2.2e-16 | NA | 27749 | 21256 | 6493 |
| European | Full Cohort | Discontinuation vs Continuation | Bipolar Disorder | Education_levelCollege Undergraduate | 1.117 | 1.044 | 1.195 | 1.41e-03 | NA | 27749 | 21256 | 6493 |
| European | Full Cohort | Discontinuation vs Continuation | Bipolar Disorder | Education_levelLess than a high school degree | 1.699 | 1.374 | 2.102 | 9.90e-07 | NA | 27749 | 21256 | 6493 |
| European | Full Cohort | Discontinuation vs Continuation | Bipolar Disorder | Education_levelTwelve or GED | 1.347 | 1.218 | 1.491 | 7.84e-09 | NA | 27749 | 21256 | 6493 |
| European | Full Cohort | Discontinuation vs Continuation | Bipolar Disorder | Bipolar Disorder | 1.039 | 1.008 | 1.070 | 1.24e-02 | 8.65e-02 | 27749 | 21256 | 6493 |
| European | Full Cohort | Discontinuation vs Continuation | Depression | (Intercept) | 0.246 | 0.215 | 0.281 | <2.2e-16 | NA | 27749 | 21256 | 6493 |
| European | Full Cohort | Discontinuation vs Continuation | Depression | age_at_first_AD | 1.007 | 1.005 | 1.009 | 8.21e-15 | NA | 27749 | 21256 | 6493 |
| European | Full Cohort | Discontinuation vs Continuation | Depression | sex_at_birthMale | 1.138 | 1.072 | 1.208 | 2.38e-05 | NA | 27749 | 21256 | 6493 |
| European | Full Cohort | Discontinuation vs Continuation | Depression | Income_quantitative | 0.935 | 0.922 | 0.948 | <2.2e-16 | NA | 27749 | 21256 | 6493 |
| European | Full Cohort | Discontinuation vs Continuation | Depression | Education_levelCollege Undergraduate | 1.110 | 1.037 | 1.188 | 2.57e-03 | NA | 27749 | 21256 | 6493 |
| European | Full Cohort | Discontinuation vs Continuation | Depression | Education_levelLess than a high school degree | 1.654 | 1.337 | 2.046 | 3.48e-06 | NA | 27749 | 21256 | 6493 |
| European | Full Cohort | Discontinuation vs Continuation | Depression | Education_levelTwelve or GED | 1.333 | 1.205 | 1.475 | 2.56e-08 | NA | 27749 | 21256 | 6493 |
| European | Full Cohort | Discontinuation vs Continuation | Depression | Depression | 1.072 | 1.041 | 1.103 | 2.98e-06 | 2.09e-05 | 27749 | 21256 | 6493 |
| European | Full Cohort | Discontinuation vs Continuation | Insomnia | (Intercept) | 0.246 | 0.215 | 0.281 | <2.2e-16 | NA | 27749 | 21256 | 6493 |
| European | Full Cohort | Discontinuation vs Continuation | Insomnia | age_at_first_AD | 1.007 | 1.005 | 1.009 | 9.16e-15 | NA | 27749 | 21256 | 6493 |
| European | Full Cohort | Discontinuation vs Continuation | Insomnia | sex_at_birthMale | 1.140 | 1.074 | 1.211 | 1.76e-05 | NA | 27749 | 21256 | 6493 |
| European | Full Cohort | Discontinuation vs Continuation | Insomnia | Income_quantitative | 0.935 | 0.922 | 0.948 | <2.2e-16 | NA | 27749 | 21256 | 6493 |
| European | Full Cohort | Discontinuation vs Continuation | Insomnia | Education_levelCollege Undergraduate | 1.108 | 1.035 | 1.186 | 3.02e-03 | NA | 27749 | 21256 | 6493 |
| European | Full Cohort | Discontinuation vs Continuation | Insomnia | Education_levelLess than a high school degree | 1.648 | 1.333 | 2.039 | 4.13e-06 | NA | 27749 | 21256 | 6493 |
| European | Full Cohort | Discontinuation vs Continuation | Insomnia | Education_levelTwelve or GED | 1.329 | 1.201 | 1.470 | 3.89e-08 | NA | 27749 | 21256 | 6493 |
| European | Full Cohort | Discontinuation vs Continuation | Insomnia | Insomnia | 1.075 | 1.044 | 1.107 | 1.53e-06 | 1.07e-05 | 27749 | 21256 | 6493 |
| European | Full Cohort | Discontinuation vs Continuation | Migraine | (Intercept) | 0.249 | 0.218 | 0.284 | <2.2e-16 | NA | 27749 | 21256 | 6493 |
| European | Full Cohort | Discontinuation vs Continuation | Migraine | age_at_first_AD | 1.007 | 1.005 | 1.009 | 2.67e-14 | NA | 27749 | 21256 | 6493 |
| European | Full Cohort | Discontinuation vs Continuation | Migraine | sex_at_birthMale | 1.140 | 1.074 | 1.211 | 1.74e-05 | NA | 27749 | 21256 | 6493 |
| European | Full Cohort | Discontinuation vs Continuation | Migraine | Income_quantitative | 0.933 | 0.920 | 0.946 | <2.2e-16 | NA | 27749 | 21256 | 6493 |

| | | | | | | | | | | | | |
|---|---|---|---|---|---|---|---|---|---|---|---|---|
| European | Full Cohort | Discontinuation vs Continuation | Migraine | Education_levelCollege Undergraduate | 1.114 | 1.041 | 1.192 | 1.77e-03 | NA | 27749 | 21256 | 6493 |
| European | Full Cohort | Discontinuation vs Continuation | Migraine | Education_levelLess than a high school degree | 1.694 | 1.370 | 2.095 | 1.15e-06 | NA | 27749 | 21256 | 6493 |
| European | Full Cohort | Discontinuation vs Continuation | Migraine | Education_levelTwelve or GED | 1.341 | 1.212 | 1.484 | 1.34e-08 | NA | 27749 | 21256 | 6493 |
| European | Full Cohort | Discontinuation vs Continuation | Migraine | Migraine | 1.006 | 0.978 | 1.034 | 6.88e-01 | 1.00e+00 | 27749 | 21256 | 6493 |
| European | Full Cohort | Discontinuation vs Continuation | Schizophrenia | (Intercept) | 0.248 | 0.217 | 0.284 | <2.2e-16 | NA | 27749 | 21256 | 6493 |
| European | Full Cohort | Discontinuation vs Continuation | Schizophrenia | age_at_first_AD | 1.007 | 1.005 | 1.009 | 2.39e-14 | NA | 27749 | 21256 | 6493 |
| European | Full Cohort | Discontinuation vs Continuation | Schizophrenia | sex_at_birthMale | 1.140 | 1.074 | 1.210 | 1.79e-05 | NA | 27749 | 21256 | 6493 |
| European | Full Cohort | Discontinuation vs Continuation | Schizophrenia | Income_quantitative | 0.933 | 0.920 | 0.946 | <2.2e-16 | NA | 27749 | 21256 | 6493 |
| European | Full Cohort | Discontinuation vs Continuation | Schizophrenia | Education_levelCollege Undergraduate | 1.115 | 1.042 | 1.193 | 1.59e-03 | NA | 27749 | 21256 | 6493 |
| European | Full Cohort | Discontinuation vs Continuation | Schizophrenia | Education_levelLess than a high school degree | 1.689 | 1.366 | 2.089 | 1.31e-06 | NA | 27749 | 21256 | 6493 |
| European | Full Cohort | Discontinuation vs Continuation | Schizophrenia | Education_levelTwelve or GED | 1.344 | 1.215 | 1.487 | 1.04e-08 | NA | 27749 | 21256 | 6493 |
| European | Full Cohort | Discontinuation vs Continuation | Schizophrenia | Schizophrenia | 1.028 | 0.999 | 1.058 | 5.57e-02 | 3.90e-01 | 27749 | 21256 | 6493 |
| European | Recorded Depression | Discontinuation vs Continuation | ADHD | (Intercept) | 0.181 | 0.140 | 0.234 | <2.2e-16 | NA | 9641 | 7969 | 1672 |
| European | Recorded Depression | Discontinuation vs Continuation | ADHD | age_at_first_AD | 1.007 | 1.004 | 1.011 | 3.50e-05 | NA | 9641 | 7969 | 1672 |
| European | Recorded Depression | Discontinuation vs Continuation | ADHD | sex_at_birthMale | 1.214 | 1.084 | 1.361 | 8.34e-04 | NA | 9641 | 7969 | 1672 |
| European | Recorded Depression | Discontinuation vs Continuation | ADHD | Income_quantitative | 0.876 | 0.851 | 0.900 | <2.2e-16 | NA | 9641 | 7969 | 1672 |
| European | Recorded Depression | Discontinuation vs Continuation | ADHD | Education_levelCollege Undergraduate | 1.270 | 1.104 | 1.461 | 8.22e-04 | NA | 9641 | 7969 | 1672 |
| European | Recorded Depression | Discontinuation vs Continuation | ADHD | Education_levelLess than a high school degree | 2.236 | 1.591 | 3.143 | 3.57e-06 | NA | 9641 | 7969 | 1672 |
| European | Recorded Depression | Discontinuation vs Continuation | ADHD | Education_levelTwelve or GED | 1.585 | 1.309 | 1.918 | 2.31e-06 | NA | 9641 | 7969 | 1672 |
| European | Recorded Depression | Discontinuation vs Continuation | ADHD | ADHD | 1.079 | 1.021 | 1.140 | 7.07e-03 | 4.95e-02 | 9641 | 7969 | 1672 |
| European | Recorded Depression | Discontinuation vs Continuation | BMI | (Intercept) | 0.182 | 0.141 | 0.236 | <2.2e-16 | NA | 9641 | 7969 | 1672 |
| European | Recorded Depression | Discontinuation vs Continuation | BMI | age_at_first_AD | 1.007 | 1.004 | 1.011 | 4.10e-05 | NA | 9641 | 7969 | 1672 |
| European | Recorded Depression | Discontinuation vs Continuation | BMI | sex_at_birthMale | 1.208 | 1.078 | 1.353 | 1.14e-03 | NA | 9641 | 7969 | 1672 |
| European | Recorded Depression | Discontinuation vs Continuation | BMI | Income_quantitative | 0.873 | 0.849 | 0.898 | <2.2e-16 | NA | 9641 | 7969 | 1672 |
| European | Recorded Depression | Discontinuation vs Continuation | BMI | Education_levelCollege Undergraduate | 1.283 | 1.115 | 1.476 | 4.81e-04 | NA | 9641 | 7969 | 1672 |
| European | Recorded Depression | Discontinuation vs Continuation | BMI | Education_levelLess than a high school degree | 2.314 | 1.648 | 3.249 | 1.27e-06 | NA | 9641 | 7969 | 1672 |
| European | Recorded Depression | Discontinuation vs Continuation | BMI | Education_levelTwelve or GED | 1.610 | 1.331 | 1.949 | 9.82e-07 | NA | 9641 | 7969 | 1672 |
| European | Recorded Depression | Discontinuation vs Continuation | BMI | BMI | 1.022 | 0.968 | 1.078 | 4.41e-01 | 1.00e+00 | 9641 | 7969 | 1672 |
| European | Recorded Depression | Discontinuation vs Continuation | Bipolar Disorder | (Intercept) | 0.183 | 0.141 | 0.237 | <2.2e-16 | NA | 9641 | 7969 | 1672 |
| European | Recorded Depression | Discontinuation vs Continuation | Bipolar Disorder | age_at_first_AD | 1.007 | 1.004 | 1.010 | 5.31e-05 | NA | 9641 | 7969 | 1672 |
| European | Recorded Depression | Discontinuation vs Continuation | Bipolar Disorder | sex_at_birthMale | 1.207 | 1.077 | 1.353 | 1.19e-03 | NA | 9641 | 7969 | 1672 |
| European | Recorded Depression | Discontinuation vs Continuation | Bipolar Disorder | Income_quantitative | 0.872 | 0.848 | 0.897 | <2.2e-16 | NA | 9641 | 7969 | 1672 |
| European | Recorded Depression | Discontinuation vs Continuation | Bipolar Disorder | Education_levelCollege Undergraduate | 1.287 | 1.119 | 1.481 | 4.02e-04 | NA | 9641 | 7969 | 1672 |
| European | Recorded Depression | Discontinuation vs Continuation | Bipolar Disorder | Education_levelLess than a high school degree | 2.336 | 1.664 | 3.279 | 9.58e-07 | NA | 9641 | 7969 | 1672 |
| European | Recorded Depression | Discontinuation vs Continuation | Bipolar Disorder | Education_levelTwelve or GED | 1.626 | 1.344 | 1.967 | 5.69e-07 | NA | 9641 | 7969 | 1672 |
| European | Recorded Depression | Discontinuation vs Continuation | Bipolar Disorder | Bipolar Disorder | 1.068 | 1.010 | 1.130 | 2.18e-02 | 1.52e-01 | 9641 | 7969 | 1672 |
| European | Recorded Depression | Discontinuation vs Continuation | Depression | (Intercept) | 0.181 | 0.139 | 0.234 | <2.2e-16 | NA | 9641 | 7969 | 1672 |
| European | Recorded Depression | Discontinuation vs Continuation | Depression | age_at_first_AD | 1.007 | 1.004 | 1.011 | 3.19e-05 | NA | 9641 | 7969 | 1672 |
| European | Recorded Depression | Discontinuation vs Continuation | Depression | sex_at_birthMale | 1.206 | 1.076 | 1.351 | 1.28e-03 | NA | 9641 | 7969 | 1672 |
| European | Recorded Depression | Discontinuation vs Continuation | Depression | Income_quantitative | 0.876 | 0.852 | 0.901 | <2.2e-16 | NA | 9641 | 7969 | 1672 |
| European | Recorded Depression | Discontinuation vs Continuation | Depression | Education_levelCollege Undergraduate | 1.275 | 1.108 | 1.466 | 6.87e-04 | NA | 9641 | 7969 | 1672 |
| European | Recorded Depression | Discontinuation vs Continuation | Depression | Education_levelLess than a high school degree | 2.229 | 1.587 | 3.131 | 3.80e-06 | NA | 9641 | 7969 | 1672 |
| European | Recorded Depression | Discontinuation vs Continuation | Depression | Education_levelTwelve or GED | 1.588 | 1.312 | 1.922 | 2.04e-06 | NA | 9641 | 7969 | 1672 |
| European | Recorded Depression | Discontinuation vs Continuation | Depression | Depression | 1.144 | 1.082 | 1.209 | 2.19e-06 | 1.53e-05 | 9641 | 7969 | 1672 |
| European | Recorded Depression | Discontinuation vs Continuation | Insomnia | (Intercept) | 0.181 | 0.139 | 0.234 | <2.2e-16 | NA | 9641 | 7969 | 1672 |
| European | Recorded Depression | Discontinuation vs Continuation | Insomnia | age_at_first_AD | 1.007 | 1.004 | 1.011 | 3.26e-05 | NA | 9641 | 7969 | 1672 |
| European | Recorded Depression | Discontinuation vs Continuation | Insomnia | sex_at_birthMale | 1.208 | 1.078 | 1.354 | 1.14e-03 | NA | 9641 | 7969 | 1672 |
| European | Recorded Depression | Discontinuation vs Continuation | Insomnia | Income_quantitative | 0.875 | 0.851 | 0.900 | <2.2e-16 | NA | 9641 | 7969 | 1672 |
| European | Recorded Depression | Discontinuation vs Continuation | Insomnia | Education_levelCollege Undergraduate | 1.277 | 1.110 | 1.469 | 6.13e-04 | NA | 9641 | 7969 | 1672 |
| European | Recorded Depression | Discontinuation vs Continuation | Insomnia | Education_levelLess than a high school degree | 2.227 | 1.585 | 3.130 | 4.00e-06 | NA | 9641 | 7969 | 1672 |
| European | Recorded Depression | Discontinuation vs Continuation | Insomnia | Education_levelTwelve or GED | 1.593 | 1.316 | 1.928 | 1.73e-06 | NA | 9641 | 7969 | 1672 |
| European | Recorded Depression | Discontinuation vs Continuation | Insomnia | Insomnia | 1.100 | 1.040 | 1.164 | 9.40e-04 | 6.58e-03 | 9641 | 7969 | 1672 |
| European | Recorded Depression | Discontinuation vs Continuation | Migraine | (Intercept) | 0.183 | 0.141 | 0.237 | <2.2e-16 | NA | 9641 | 7969 | 1672 |
| European | Recorded Depression | Discontinuation vs Continuation | Migraine | age_at_first_AD | 1.007 | 1.004 | 1.010 | 4.72e-05 | NA | 9641 | 7969 | 1672 |
| European | Recorded Depression | Discontinuation vs Continuation | Migraine | sex_at_birthMale | 1.208 | 1.078 | 1.354 | 1.11e-03 | NA | 9641 | 7969 | 1672 |
| European | Recorded Depression | Discontinuation vs Continuation | Migraine | Income_quantitative | 0.873 | 0.848 | 0.897 | <2.2e-16 | NA | 9641 | 7969 | 1672 |
| European | Recorded Depression | Discontinuation vs Continuation | Migraine | Education_levelCollege Undergraduate | 1.285 | 1.117 | 1.478 | 4.47e-04 | NA | 9641 | 7969 | 1672 |

| | | | | | | | | | | | | |
|---|---|---|---|---|---|---|---|---|---|---|---|---|
| European | Recorded Depression | Discontinuation vs Continuation | Migraine | Education_levelLess than a high school degree | 2.325 | 1.656 | 3.263 | 1.08e-06 | NA | 9641 | 7969 | 1672 |
| European | Recorded Depression | Discontinuation vs Continuation | Migraine | Education_levelTwelve or GED | 1.617 | 1.337 | 1.956 | 7.58e-07 | NA | 9641 | 7969 | 1672 |
| European | Recorded Depression | Discontinuation vs Continuation | Migraine | Migraine | 1.011 | 0.959 | 1.067 | 6.82e-01 | 1.00e+00 | 9641 | 7969 | 1672 |
| European | Recorded Depression | Discontinuation vs Continuation | Schizophrenia | (Intercept) | 0.183 | 0.141 | 0.236 | <2.2e-16 | NA | 9641 | 7969 | 1672 |
| European | Recorded Depression | Discontinuation vs Continuation | Schizophrenia | age_at_first_AD | 1.007 | 1.004 | 1.011 | 4.60e-05 | NA | 9641 | 7969 | 1672 |
| European | Recorded Depression | Discontinuation vs Continuation | Schizophrenia | sex_at_birthMale | 1.209 | 1.079 | 1.354 | 1.10e-03 | NA | 9641 | 7969 | 1672 |
| European | Recorded Depression | Discontinuation vs Continuation | Schizophrenia | Income_quantitative | 0.873 | 0.849 | 0.897 | <2.2e-16 | NA | 9641 | 7969 | 1672 |
| European | Recorded Depression | Discontinuation vs Continuation | Schizophrenia | Education_levelCollege Undergraduate | 1.286 | 1.118 | 1.479 | 4.27e-04 | NA | 9641 | 7969 | 1672 |
| European | Recorded Depression | Discontinuation vs Continuation | Schizophrenia | Education_levelLess than a high school degree | 2.311 | 1.646 | 3.244 | 1.30e-06 | NA | 9641 | 7969 | 1672 |
| European | Recorded Depression | Discontinuation vs Continuation | Schizophrenia | Education_levelTwelve or GED | 1.620 | 1.339 | 1.960 | 6.86e-07 | NA | 9641 | 7969 | 1672 |
| European | Recorded Depression | Discontinuation vs Continuation | Schizophrenia | Schizophrenia | 1.044 | 0.989 | 1.102 | 1.22e-01 | 8.51e-01 | 9641 | 7969 | 1672 |
| European | No Recorded Depression | Discontinuation vs Continuation | ADHD | (Intercept) | 0.327 | 0.279 | 0.384 | <2.2e-16 | NA | 18108 | 13287 | 4821 |
| European | No Recorded Depression | Discontinuation vs Continuation | ADHD | age_at_first_AD | 1.006 | 1.004 | 1.008 | 1.10e-08 | NA | 18108 | 13287 | 4821 |
| European | No Recorded Depression | Discontinuation vs Continuation | ADHD | sex_at_birthMale | 1.117 | 1.040 | 1.199 | 2.27e-03 | NA | 18108 | 13287 | 4821 |
| European | No Recorded Depression | Discontinuation vs Continuation | ADHD | Income_quantitative | 0.934 | 0.919 | 0.950 | 1.77e-15 | NA | 18108 | 13287 | 4821 |
| European | No Recorded Depression | Discontinuation vs Continuation | ADHD | Education_levelCollege Undergraduate | 1.051 | 0.972 | 1.137 | 2.11e-01 | NA | 18108 | 13287 | 4821 |
| European | No Recorded Depression | Discontinuation vs Continuation | ADHD | Education_levelLess than a high school degree | 1.385 | 1.049 | 1.828 | 2.15e-02 | NA | 18108 | 13287 | 4821 |
| European | No Recorded Depression | Discontinuation vs Continuation | ADHD | Education_levelTwelve or GED | 1.215 | 1.075 | 1.372 | 1.74e-03 | NA | 18108 | 13287 | 4821 |
| European | No Recorded Depression | Discontinuation vs Continuation | ADHD | ADHD | 1.034 | 0.999 | 1.070 | 5.61e-02 | 3.93e-01 | 18108 | 13287 | 4821 |
| European | No Recorded Depression | Discontinuation vs Continuation | BMI | (Intercept) | 0.329 | 0.281 | 0.386 | <2.2e-16 | NA | 18108 | 13287 | 4821 |
| European | No Recorded Depression | Discontinuation vs Continuation | BMI | age_at_first_AD | 1.006 | 1.004 | 1.008 | 1.71e-08 | NA | 18108 | 13287 | 4821 |
| European | No Recorded Depression | Discontinuation vs Continuation | BMI | sex_at_birthMale | 1.116 | 1.040 | 1.198 | 2.45e-03 | NA | 18108 | 13287 | 4821 |
| European | No Recorded Depression | Discontinuation vs Continuation | BMI | Income_quantitative | 0.933 | 0.918 | 0.949 | 5.58e-16 | NA | 18108 | 13287 | 4821 |
| European | No Recorded Depression | Discontinuation vs Continuation | BMI | Education_levelCollege Undergraduate | 1.057 | 0.977 | 1.143 | 1.67e-01 | NA | 18108 | 13287 | 4821 |
| European | No Recorded Depression | Discontinuation vs Continuation | BMI | Education_levelLess than a high school degree | 1.412 | 1.070 | 1.864 | 1.47e-02 | NA | 18108 | 13287 | 4821 |
| European | No Recorded Depression | Discontinuation vs Continuation | BMI | Education_levelTwelve or GED | 1.226 | 1.085 | 1.384 | 1.03e-03 | NA | 18108 | 13287 | 4821 |
| European | No Recorded Depression | Discontinuation vs Continuation | BMI | BMI | 0.998 | 0.965 | 1.033 | 9.28e-01 | 1.00e+00 | 18108 | 13287 | 4821 |
| European | No Recorded Depression | Discontinuation vs Continuation | Bipolar Disorder | (Intercept) | 0.328 | 0.280 | 0.385 | <2.2e-16 | NA | 18108 | 13287 | 4821 |
| European | No Recorded Depression | Discontinuation vs Continuation | Bipolar Disorder | age_at_first_AD | 1.006 | 1.004 | 1.008 | 1.91e-08 | NA | 18108 | 13287 | 4821 |
| European | No Recorded Depression | Discontinuation vs Continuation | Bipolar Disorder | sex_at_birthMale | 1.115 | 1.038 | 1.197 | 2.72e-03 | NA | 18108 | 13287 | 4821 |
| European | No Recorded Depression | Discontinuation vs Continuation | Bipolar Disorder | Income_quantitative | 0.934 | 0.918 | 0.949 | 7.31e-16 | NA | 18108 | 13287 | 4821 |
| European | No Recorded Depression | Discontinuation vs Continuation | Bipolar Disorder | Education_levelCollege Undergraduate | 1.059 | 0.980 | 1.146 | 1.49e-01 | NA | 18108 | 13287 | 4821 |
| European | No Recorded Depression | Discontinuation vs Continuation | Bipolar Disorder | Education_levelLess than a high school degree | 1.416 | 1.073 | 1.869 | 1.38e-02 | NA | 18108 | 13287 | 4821 |
| European | No Recorded Depression | Discontinuation vs Continuation | Bipolar Disorder | Education_levelTwelve or GED | 1.232 | 1.091 | 1.392 | 7.57e-04 | NA | 18108 | 13287 | 4821 |
| European | No Recorded Depression | Discontinuation vs Continuation | Bipolar Disorder | Bipolar Disorder | 1.040 | 1.004 | 1.077 | 2.92e-02 | 2.04e-01 | 18108 | 13287 | 4821 |
| European | No Recorded Depression | Discontinuation vs Continuation | Depression | (Intercept) | 0.325 | 0.277 | 0.381 | <2.2e-16 | NA | 18108 | 13287 | 4821 |
| European | No Recorded Depression | Discontinuation vs Continuation | Depression | age_at_first_AD | 1.006 | 1.004 | 1.008 | 6.65e-09 | NA | 18108 | 13287 | 4821 |
| European | No Recorded Depression | Discontinuation vs Continuation | Depression | sex_at_birthMale | 1.113 | 1.037 | 1.195 | 3.16e-03 | NA | 18108 | 13287 | 4821 |
| European | No Recorded Depression | Discontinuation vs Continuation | Depression | Income_quantitative | 0.935 | 0.920 | 0.951 | 4.06e-15 | NA | 18108 | 13287 | 4821 |
| European | No Recorded Depression | Discontinuation vs Continuation | Depression | Education_levelCollege Undergraduate | 1.052 | 0.973 | 1.138 | 2.03e-01 | NA | 18108 | 13287 | 4821 |
| European | No Recorded Depression | Discontinuation vs Continuation | Depression | Education_levelLess than a high school degree | 1.377 | 1.043 | 1.817 | 2.39e-02 | NA | 18108 | 13287 | 4821 |
| European | No Recorded Depression | Discontinuation vs Continuation | Depression | Education_levelTwelve or GED | 1.220 | 1.081 | 1.378 | 1.33e-03 | NA | 18108 | 13287 | 4821 |
| European | No Recorded Depression | Discontinuation vs Continuation | Depression | Depression | 1.072 | 1.036 | 1.110 | 7.76e-05 | 5.44e-04 | 18108 | 13287 | 4821 |
| European | No Recorded Depression | Discontinuation vs Continuation | Insomnia | (Intercept) | 0.325 | 0.277 | 0.381 | <2.2e-16 | NA | 18108 | 13287 | 4821 |
| European | No Recorded Depression | Discontinuation vs Continuation | Insomnia | age_at_first_AD | 1.006 | 1.004 | 1.008 | 8.14e-09 | NA | 18108 | 13287 | 4821 |
| European | No Recorded Depression | Discontinuation vs Continuation | Insomnia | sex_at_birthMale | 1.116 | 1.039 | 1.198 | 2.47e-03 | NA | 18108 | 13287 | 4821 |
| European | No Recorded Depression | Discontinuation vs Continuation | Insomnia | Income_quantitative | 0.935 | 0.920 | 0.951 | 4.37e-15 | NA | 18108 | 13287 | 4821 |
| European | No Recorded Depression | Discontinuation vs Continuation | Insomnia | Education_levelCollege Undergraduate | 1.050 | 0.971 | 1.136 | 2.21e-01 | NA | 18108 | 13287 | 4821 |
| European | No Recorded Depression | Discontinuation vs Continuation | Insomnia | Education_levelLess than a high school degree | 1.380 | 1.046 | 1.822 | 2.28e-02 | NA | 18108 | 13287 | 4821 |
| European | No Recorded Depression | Discontinuation vs Continuation | Insomnia | Education_levelTwelve or GED | 1.215 | 1.076 | 1.372 | 1.67e-03 | NA | 18108 | 13287 | 4821 |
| European | No Recorded Depression | Discontinuation vs Continuation | Insomnia | Insomnia | 1.073 | 1.036 | 1.111 | 6.94e-05 | 4.86e-04 | 18108 | 13287 | 4821 |
| European | No Recorded Depression | Discontinuation vs Continuation | Migraine | (Intercept) | 0.329 | 0.281 | 0.386 | <2.2e-16 | NA | 18108 | 13287 | 4821 |
| European | No Recorded Depression | Discontinuation vs Continuation | Migraine | age_at_first_AD | 1.006 | 1.004 | 1.008 | 1.63e-08 | NA | 18108 | 13287 | 4821 |
| European | No Recorded Depression | Discontinuation vs Continuation | Migraine | sex_at_birthMale | 1.116 | 1.039 | 1.198 | 2.46e-03 | NA | 18108 | 13287 | 4821 |
| European | No Recorded Depression | Discontinuation vs Continuation | Migraine | Income_quantitative | 0.933 | 0.918 | 0.949 | 4.92e-16 | NA | 18108 | 13287 | 4821 |
| European | No Recorded Depression | Discontinuation vs Continuation | Migraine | Education_levelCollege Undergraduate | 1.057 | 0.977 | 1.143 | 1.69e-01 | NA | 18108 | 13287 | 4821 |
| European | No Recorded Depression | Discontinuation vs Continuation | Migraine | Education_levelLess than a high school degree | 1.411 | 1.070 | 1.862 | 1.48e-02 | NA | 18108 | 13287 | 4821 |

| | | | | | | | | | | | | |
|---|---|---|---|---|---|---|---|---|---|---|---|---|
| European | No Recorded Depression | Discontinuation vs Continuation | Migraine | Education_levelTwelve or GED | 1.225 | 1.085 | 1.383 | 1.04e-03 | NA | 18108 | 13287 | 4821 |
| European | No Recorded Depression | Discontinuation vs Continuation | Migraine | Migraine | 1.002 | 0.969 | 1.036 | 8.97e-01 | 1.00e+00 | 18108 | 13287 | 4821 |
| European | No Recorded Depression | Discontinuation vs Continuation | Schizophrenia | (Intercept) | 0.328 | 0.280 | 0.384 | <2.2e-16 | NA | 18108 | 13287 | 4821 |
| European | No Recorded Depression | Discontinuation vs Continuation | Schizophrenia | age_at_first_AD | 1.006 | 1.004 | 1.008 | 1.49e-08 | NA | 18108 | 13287 | 4821 |
| European | No Recorded Depression | Discontinuation vs Continuation | Schizophrenia | sex_at_birthMale | 1.116 | 1.039 | 1.198 | 2.56e-03 | NA | 18108 | 13287 | 4821 |
| European | No Recorded Depression | Discontinuation vs Continuation | Schizophrenia | Income_quantitative | 0.934 | 0.918 | 0.950 | 8.41e-16 | NA | 18108 | 13287 | 4821 |
| European | No Recorded Depression | Discontinuation vs Continuation | Schizophrenia | Education_levelCollege Undergraduate | 1.058 | 0.978 | 1.144 | 1.60e-01 | NA | 18108 | 13287 | 4821 |
| European | No Recorded Depression | Discontinuation vs Continuation | Schizophrenia | Education_levelLess than a high school degree | 1.409 | 1.068 | 1.858 | 1.54e-02 | NA | 18108 | 13287 | 4821 |
| European | No Recorded Depression | Discontinuation vs Continuation | Schizophrenia | Education_levelTwelve or GED | 1.228 | 1.088 | 1.387 | 9.17e-04 | NA | 18108 | 13287 | 4821 |
| European | No Recorded Depression | Discontinuation vs Continuation | Schizophrenia | Schizophrenia | 1.025 | 0.991 | 1.060 | 1.48e-01 | 1.00e+00 | 18108 | 13287 | 4821 |
| European | Recorded Anxiety | Discontinuation vs Continuation | ADHD | (Intercept) | 0.232 | 0.191 | 0.282 | <2.2e-16 | NA | 14583 | 11798 | 2785 |
| European | Recorded Anxiety | Discontinuation vs Continuation | ADHD | age_at_first_AD | 1.007 | 1.004 | 1.009 | 4.55e-07 | NA | 14583 | 11798 | 2785 |
| European | Recorded Anxiety | Discontinuation vs Continuation | ADHD | sex_at_birthMale | 1.238 | 1.131 | 1.356 | 3.78e-06 | NA | 14583 | 11798 | 2785 |
| European | Recorded Anxiety | Discontinuation vs Continuation | ADHD | Income_quantitative | 0.894 | 0.875 | 0.913 | <2.2e-16 | NA | 14583 | 11798 | 2785 |
| European | Recorded Anxiety | Discontinuation vs Continuation | ADHD | Education_levelCollege Undergraduate | 1.074 | 0.968 | 1.191 | 1.80e-01 | NA | 14583 | 11798 | 2785 |
| European | Recorded Anxiety | Discontinuation vs Continuation | ADHD | Education_levelLess than a high school degree | 1.573 | 1.180 | 2.096 | 2.03e-03 | NA | 14583 | 11798 | 2785 |
| European | Recorded Anxiety | Discontinuation vs Continuation | ADHD | Education_levelTwelve or GED | 1.328 | 1.143 | 1.542 | 2.02e-04 | NA | 14583 | 11798 | 2785 |
| European | Recorded Anxiety | Discontinuation vs Continuation | ADHD | ADHD | 1.044 | 1.000 | 1.090 | 4.88e-02 | 3.41e-01 | 14583 | 11798 | 2785 |
| European | Recorded Anxiety | Discontinuation vs Continuation | BMI | (Intercept) | 0.234 | 0.193 | 0.284 | <2.2e-16 | NA | 14583 | 11798 | 2785 |
| European | Recorded Anxiety | Discontinuation vs Continuation | BMI | age_at_first_AD | 1.007 | 1.004 | 1.009 | 7.02e-07 | NA | 14583 | 11798 | 2785 |
| European | Recorded Anxiety | Discontinuation vs Continuation | BMI | sex_at_birthMale | 1.236 | 1.129 | 1.353 | 4.60e-06 | NA | 14583 | 11798 | 2785 |
| European | Recorded Anxiety | Discontinuation vs Continuation | BMI | Income_quantitative | 0.892 | 0.873 | 0.911 | <2.2e-16 | NA | 14583 | 11798 | 2785 |
| European | Recorded Anxiety | Discontinuation vs Continuation | BMI | Education_levelCollege Undergraduate | 1.085 | 0.978 | 1.203 | 1.25e-01 | NA | 14583 | 11798 | 2785 |
| European | Recorded Anxiety | Discontinuation vs Continuation | BMI | Education_levelLess than a high school degree | 1.618 | 1.215 | 2.156 | 1.01e-03 | NA | 14583 | 11798 | 2785 |
| European | Recorded Anxiety | Discontinuation vs Continuation | BMI | Education_levelTwelve or GED | 1.351 | 1.163 | 1.568 | 8.17e-05 | NA | 14583 | 11798 | 2785 |
| European | Recorded Anxiety | Discontinuation vs Continuation | BMI | BMI | 0.978 | 0.937 | 1.019 | 2.89e-01 | 1.00e+00 | 14583 | 11798 | 2785 |
| European | Recorded Anxiety | Discontinuation vs Continuation | Bipolar Disorder | (Intercept) | 0.233 | 0.192 | 0.283 | <2.2e-16 | NA | 14583 | 11798 | 2785 |
| European | Recorded Anxiety | Discontinuation vs Continuation | Bipolar Disorder | age_at_first_AD | 1.007 | 1.004 | 1.009 | 7.68e-07 | NA | 14583 | 11798 | 2785 |
| European | Recorded Anxiety | Discontinuation vs Continuation | Bipolar Disorder | sex_at_birthMale | 1.232 | 1.126 | 1.349 | 6.17e-06 | NA | 14583 | 11798 | 2785 |
| European | Recorded Anxiety | Discontinuation vs Continuation | Bipolar Disorder | Income_quantitative | 0.893 | 0.874 | 0.912 | <2.2e-16 | NA | 14583 | 11798 | 2785 |
| European | Recorded Anxiety | Discontinuation vs Continuation | Bipolar Disorder | Education_levelCollege Undergraduate | 1.086 | 0.979 | 1.204 | 1.21e-01 | NA | 14583 | 11798 | 2785 |
| European | Recorded Anxiety | Discontinuation vs Continuation | Bipolar Disorder | Education_levelLess than a high school degree | 1.621 | 1.216 | 2.159 | 9.72e-04 | NA | 14583 | 11798 | 2785 |
| European | Recorded Anxiety | Discontinuation vs Continuation | Bipolar Disorder | Education_levelTwelve or GED | 1.353 | 1.165 | 1.571 | 7.26e-05 | NA | 14583 | 11798 | 2785 |
| European | Recorded Anxiety | Discontinuation vs Continuation | Bipolar Disorder | Bipolar Disorder | 1.072 | 1.025 | 1.120 | 2.18e-03 | 1.52e-02 | 14583 | 11798 | 2785 |
| European | Recorded Anxiety | Discontinuation vs Continuation | Depression | (Intercept) | 0.231 | 0.190 | 0.281 | <2.2e-16 | NA | 14583 | 11798 | 2785 |
| European | Recorded Anxiety | Discontinuation vs Continuation | Depression | age_at_first_AD | 1.007 | 1.004 | 1.009 | 3.57e-07 | NA | 14583 | 11798 | 2785 |
| European | Recorded Anxiety | Discontinuation vs Continuation | Depression | sex_at_birthMale | 1.232 | 1.125 | 1.349 | 6.60e-06 | NA | 14583 | 11798 | 2785 |
| European | Recorded Anxiety | Discontinuation vs Continuation | Depression | Income_quantitative | 0.896 | 0.877 | 0.915 | <2.2e-16 | NA | 14583 | 11798 | 2785 |
| European | Recorded Anxiety | Discontinuation vs Continuation | Depression | Education_levelCollege Undergraduate | 1.072 | 0.966 | 1.189 | 1.88e-01 | NA | 14583 | 11798 | 2785 |
| European | Recorded Anxiety | Discontinuation vs Continuation | Depression | Education_levelLess than a high school degree | 1.559 | 1.170 | 2.078 | 2.44e-03 | NA | 14583 | 11798 | 2785 |
| European | Recorded Anxiety | Discontinuation vs Continuation | Depression | Education_levelTwelve or GED | 1.325 | 1.141 | 1.539 | 2.22e-04 | NA | 14583 | 11798 | 2785 |
| European | Recorded Anxiety | Discontinuation vs Continuation | Depression | Depression | 1.111 | 1.064 | 1.160 | 2.24e-06 | 1.56e-05 | 14583 | 11798 | 2785 |
| European | Recorded Anxiety | Discontinuation vs Continuation | Insomnia | (Intercept) | 0.231 | 0.190 | 0.281 | <2.2e-16 | NA | 14583 | 11798 | 2785 |
| European | Recorded Anxiety | Discontinuation vs Continuation | Insomnia | age_at_first_AD | 1.007 | 1.004 | 1.009 | 3.29e-07 | NA | 14583 | 11798 | 2785 |
| European | Recorded Anxiety | Discontinuation vs Continuation | Insomnia | sex_at_birthMale | 1.235 | 1.128 | 1.352 | 4.98e-06 | NA | 14583 | 11798 | 2785 |
| European | Recorded Anxiety | Discontinuation vs Continuation | Insomnia | Income_quantitative | 0.895 | 0.876 | 0.914 | <2.2e-16 | NA | 14583 | 11798 | 2785 |
| European | Recorded Anxiety | Discontinuation vs Continuation | Insomnia | Education_levelCollege Undergraduate | 1.070 | 0.965 | 1.187 | 2.01e-01 | NA | 14583 | 11798 | 2785 |
| European | Recorded Anxiety | Discontinuation vs Continuation | Insomnia | Education_levelLess than a high school degree | 1.541 | 1.156 | 2.054 | 3.20e-03 | NA | 14583 | 11798 | 2785 |
| European | Recorded Anxiety | Discontinuation vs Continuation | Insomnia | Education_levelTwelve or GED | 1.319 | 1.136 | 1.532 | 2.84e-04 | NA | 14583 | 11798 | 2785 |
| European | Recorded Anxiety | Discontinuation vs Continuation | Insomnia | Insomnia | 1.109 | 1.061 | 1.159 | 4.44e-06 | 3.11e-05 | 14583 | 11798 | 2785 |
| European | Recorded Anxiety | Discontinuation vs Continuation | Migraine | (Intercept) | 0.234 | 0.192 | 0.284 | <2.2e-16 | NA | 14583 | 11798 | 2785 |
| European | Recorded Anxiety | Discontinuation vs Continuation | Migraine | age_at_first_AD | 1.007 | 1.004 | 1.009 | 5.63e-07 | NA | 14583 | 11798 | 2785 |
| European | Recorded Anxiety | Discontinuation vs Continuation | Migraine | sex_at_birthMale | 1.235 | 1.128 | 1.352 | 4.79e-06 | NA | 14583 | 11798 | 2785 |
| European | Recorded Anxiety | Discontinuation vs Continuation | Migraine | Income_quantitative | 0.893 | 0.874 | 0.912 | <2.2e-16 | NA | 14583 | 11798 | 2785 |
| European | Recorded Anxiety | Discontinuation vs Continuation | Migraine | Education_levelCollege Undergraduate | 1.080 | 0.973 | 1.198 | 1.47e-01 | NA | 14583 | 11798 | 2785 |
| European | Recorded Anxiety | Discontinuation vs Continuation | Migraine | Education_levelLess than a high school degree | 1.602 | 1.202 | 2.134 | 1.29e-03 | NA | 14583 | 11798 | 2785 |
| European | Recorded Anxiety | Discontinuation vs Continuation | Migraine | Education_levelTwelve or GED | 1.341 | 1.155 | 1.557 | 1.14e-04 | NA | 14583 | 11798 | 2785 |

| | | | | | | | | | | | | |
|---|---|---|---|---|---|---|---|---|---|---|---|---|
| European | Recorded Anxiety | Discontinuation vs Continuation | Migraine | Migraine | 1.023 | 0.982 | 1.067 | 2.77e-01 | 1.00e+00 | 14583 | 11798 | 2785 |
| European | Recorded Anxiety | Discontinuation vs Continuation | Schizophrenia | (Intercept) | 0.232 | 0.191 | 0.282 | <2.2e-16 | NA | 14583 | 11798 | 2785 |
| European | Recorded Anxiety | Discontinuation vs Continuation | Schizophrenia | age_at_first_AD | 1.007 | 1.004 | 1.009 | 5.95e-07 | NA | 14583 | 11798 | 2785 |
| European | Recorded Anxiety | Discontinuation vs Continuation | Schizophrenia | sex_at_birthMale | 1.234 | 1.127 | 1.351 | 5.30e-06 | NA | 14583 | 11798 | 2785 |
| European | Recorded Anxiety | Discontinuation vs Continuation | Schizophrenia | Income_quantitative | 0.893 | 0.875 | 0.912 | <2.2e-16 | NA | 14583 | 11798 | 2785 |
| European | Recorded Anxiety | Discontinuation vs Continuation | Schizophrenia | Education_levelCollege Undergraduate | 1.083 | 0.976 | 1.201 | 1.34e-01 | NA | 14583 | 11798 | 2785 |
| European | Recorded Anxiety | Discontinuation vs Continuation | Schizophrenia | Education_levelLess than a high school degree | 1.600 | 1.201 | 2.131 | 1.32e-03 | NA | 14583 | 11798 | 2785 |
| European | Recorded Anxiety | Discontinuation vs Continuation | Schizophrenia | Education_levelTwelve or GED | 1.347 | 1.160 | 1.564 | 9.15e-05 | NA | 14583 | 11798 | 2785 |
| European | Recorded Anxiety | Discontinuation vs Continuation | Schizophrenia | Schizophrenia | 1.047 | 1.004 | 1.093 | 3.31e-02 | 2.32e-01 | 14583 | 11798 | 2785 |
| European | CIDI-MDD | Discontinuation vs Continuation | ADHD | (Intercept) | 0.254 | 0.167 | 0.389 | 2.46e-10 | NA | 2956 | 2349 | 607 |
| European | CIDI-MDD | Discontinuation vs Continuation | ADHD | age_at_first_AD | 1.002 | 0.996 | 1.008 | 6.00e-01 | NA | 2956 | 2349 | 607 |
| European | CIDI-MDD | Discontinuation vs Continuation | ADHD | sex_at_birthMale | 1.077 | 0.876 | 1.324 | 4.83e-01 | NA | 2956 | 2349 | 607 |
| European | CIDI-MDD | Discontinuation vs Continuation | ADHD | Income_quantitative | 0.958 | 0.916 | 1.002 | 5.94e-02 | NA | 2956 | 2349 | 607 |
| European | CIDI-MDD | Discontinuation vs Continuation | ADHD | Education_levelCollege Undergraduate | 1.190 | 0.971 | 1.459 | 9.32e-02 | NA | 2956 | 2349 | 607 |
| European | CIDI-MDD | Discontinuation vs Continuation | ADHD | Education_levelLess than a high school degree | 0.696 | 0.198 | 2.445 | 5.72e-01 | NA | 2956 | 2349 | 607 |
| European | CIDI-MDD | Discontinuation vs Continuation | ADHD | Education_levelTwelve or GED | 1.135 | 0.759 | 1.697 | 5.37e-01 | NA | 2956 | 2349 | 607 |
| European | CIDI-MDD | Discontinuation vs Continuation | ADHD | ADHD | 1.071 | 0.977 | 1.176 | 1.45e-01 | 1.00e+00 | 2956 | 2349 | 607 |
| European | CIDI-MDD | Discontinuation vs Continuation | BMI | (Intercept) | 0.257 | 0.168 | 0.392 | 2.98e-10 | NA | 2956 | 2349 | 607 |
| European | CIDI-MDD | Discontinuation vs Continuation | BMI | age_at_first_AD | 1.002 | 0.996 | 1.008 | 6.12e-01 | NA | 2956 | 2349 | 607 |
| European | CIDI-MDD | Discontinuation vs Continuation | BMI | sex_at_birthMale | 1.075 | 0.874 | 1.321 | 4.93e-01 | NA | 2956 | 2349 | 607 |
| European | CIDI-MDD | Discontinuation vs Continuation | BMI | Income_quantitative | 0.954 | 0.912 | 0.998 | 3.91e-02 | NA | 2956 | 2349 | 607 |
| European | CIDI-MDD | Discontinuation vs Continuation | BMI | Education_levelCollege Undergraduate | 1.209 | 0.987 | 1.481 | 6.65e-02 | NA | 2956 | 2349 | 607 |
| European | CIDI-MDD | Discontinuation vs Continuation | BMI | Education_levelLess than a high school degree | 0.739 | 0.211 | 2.591 | 6.36e-01 | NA | 2956 | 2349 | 607 |
| European | CIDI-MDD | Discontinuation vs Continuation | BMI | Education_levelTwelve or GED | 1.167 | 0.781 | 1.744 | 4.51e-01 | NA | 2956 | 2349 | 607 |
| European | CIDI-MDD | Discontinuation vs Continuation | BMI | BMI | 0.967 | 0.881 | 1.061 | 4.76e-01 | 1.00e+00 | 2956 | 2349 | 607 |
| European | CIDI-MDD | Discontinuation vs Continuation | Bipolar Disorder | (Intercept) | 0.259 | 0.170 | 0.396 | 4.43e-10 | NA | 2956 | 2349 | 607 |
| European | CIDI-MDD | Discontinuation vs Continuation | Bipolar Disorder | age_at_first_AD | 1.001 | 0.995 | 1.007 | 7.09e-01 | NA | 2956 | 2349 | 607 |
| European | CIDI-MDD | Discontinuation vs Continuation | Bipolar Disorder | sex_at_birthMale | 1.073 | 0.873 | 1.319 | 5.05e-01 | NA | 2956 | 2349 | 607 |
| European | CIDI-MDD | Discontinuation vs Continuation | Bipolar Disorder | Income_quantitative | 0.954 | 0.913 | 0.998 | 4.10e-02 | NA | 2956 | 2349 | 607 |
| European | CIDI-MDD | Discontinuation vs Continuation | Bipolar Disorder | Education_levelCollege Undergraduate | 1.214 | 0.991 | 1.487 | 6.09e-02 | NA | 2956 | 2349 | 607 |
| European | CIDI-MDD | Discontinuation vs Continuation | Bipolar Disorder | Education_levelLess than a high school degree | 0.774 | 0.220 | 2.720 | 6.90e-01 | NA | 2956 | 2349 | 607 |
| European | CIDI-MDD | Discontinuation vs Continuation | Bipolar Disorder | Education_levelTwelve or GED | 1.184 | 0.792 | 1.770 | 4.09e-01 | NA | 2956 | 2349 | 607 |
| European | CIDI-MDD | Discontinuation vs Continuation | Bipolar Disorder | Bipolar Disorder | 1.152 | 1.045 | 1.269 | 4.30e-03 | 3.01e-02 | 2956 | 2349 | 607 |
| European | CIDI-MDD | Discontinuation vs Continuation | Depression | (Intercept) | 0.256 | 0.168 | 0.392 | 3.19e-10 | NA | 2956 | 2349 | 607 |
| European | CIDI-MDD | Discontinuation vs Continuation | Depression | age_at_first_AD | 1.001 | 0.995 | 1.007 | 6.40e-01 | NA | 2956 | 2349 | 607 |
| European | CIDI-MDD | Discontinuation vs Continuation | Depression | sex_at_birthMale | 1.068 | 0.868 | 1.313 | 5.34e-01 | NA | 2956 | 2349 | 607 |
| European | CIDI-MDD | Discontinuation vs Continuation | Depression | Income_quantitative | 0.960 | 0.918 | 1.004 | 7.09e-02 | NA | 2956 | 2349 | 607 |
| European | CIDI-MDD | Discontinuation vs Continuation | Depression | Education_levelCollege Undergraduate | 1.174 | 0.958 | 1.439 | 1.21e-01 | NA | 2956 | 2349 | 607 |
| European | CIDI-MDD | Discontinuation vs Continuation | Depression | Education_levelLess than a high school degree | 0.718 | 0.205 | 2.522 | 6.06e-01 | NA | 2956 | 2349 | 607 |
| European | CIDI-MDD | Discontinuation vs Continuation | Depression | Education_levelTwelve or GED | 1.133 | 0.758 | 1.694 | 5.43e-01 | NA | 2956 | 2349 | 607 |
| European | CIDI-MDD | Discontinuation vs Continuation | Depression | Depression | 1.215 | 1.104 | 1.336 | 6.04e-05 | 4.23e-04 | 2956 | 2349 | 607 |
| European | CIDI-MDD | Discontinuation vs Continuation | Insomnia | (Intercept) | 0.255 | 0.167 | 0.390 | 2.76e-10 | NA | 2956 | 2349 | 607 |
| European | CIDI-MDD | Discontinuation vs Continuation | Insomnia | age_at_first_AD | 1.001 | 0.995 | 1.007 | 6.56e-01 | NA | 2956 | 2349 | 607 |
| European | CIDI-MDD | Discontinuation vs Continuation | Insomnia | sex_at_birthMale | 1.081 | 0.879 | 1.329 | 4.59e-01 | NA | 2956 | 2349 | 607 |
| European | CIDI-MDD | Discontinuation vs Continuation | Insomnia | Income_quantitative | 0.959 | 0.917 | 1.003 | 6.43e-02 | NA | 2956 | 2349 | 607 |
| European | CIDI-MDD | Discontinuation vs Continuation | Insomnia | Education_levelCollege Undergraduate | 1.189 | 0.971 | 1.457 | 9.44e-02 | NA | 2956 | 2349 | 607 |
| European | CIDI-MDD | Discontinuation vs Continuation | Insomnia | Education_levelLess than a high school degree | 0.702 | 0.200 | 2.462 | 5.80e-01 | NA | 2956 | 2349 | 607 |
| European | CIDI-MDD | Discontinuation vs Continuation | Insomnia | Education_levelTwelve or GED | 1.145 | 0.766 | 1.711 | 5.08e-01 | NA | 2956 | 2349 | 607 |
| European | CIDI-MDD | Discontinuation vs Continuation | Insomnia | Insomnia | 1.142 | 1.038 | 1.256 | 6.25e-03 | 4.38e-02 | 2956 | 2349 | 607 |
| European | CIDI-MDD | Discontinuation vs Continuation | Migraine | (Intercept) | 0.256 | 0.167 | 0.390 | 2.63e-10 | NA | 2956 | 2349 | 607 |
| European | CIDI-MDD | Discontinuation vs Continuation | Migraine | age_at_first_AD | 1.002 | 0.996 | 1.008 | 6.04e-01 | NA | 2956 | 2349 | 607 |
| European | CIDI-MDD | Discontinuation vs Continuation | Migraine | sex_at_birthMale | 1.072 | 0.872 | 1.318 | 5.08e-01 | NA | 2956 | 2349 | 607 |
| European | CIDI-MDD | Discontinuation vs Continuation | Migraine | Income_quantitative | 0.955 | 0.914 | 0.999 | 4.53e-02 | NA | 2956 | 2349 | 607 |
| European | CIDI-MDD | Discontinuation vs Continuation | Migraine | Education_levelCollege Undergraduate | 1.206 | 0.985 | 1.477 | 7.04e-02 | NA | 2956 | 2349 | 607 |
| European | CIDI-MDD | Discontinuation vs Continuation | Migraine | Education_levelLess than a high school degree | 0.728 | 0.208 | 2.553 | 6.20e-01 | NA | 2956 | 2349 | 607 |
| European | CIDI-MDD | Discontinuation vs Continuation | Migraine | Education_levelTwelve or GED | 1.159 | 0.776 | 1.731 | 4.71e-01 | NA | 2956 | 2349 | 607 |
| European | CIDI-MDD | Discontinuation vs Continuation | Migraine | Migraine | 1.012 | 0.926 | 1.105 | 7.95e-01 | 1.00e+00 | 2956 | 2349 | 607 |

| | | | | | | | | | | | | |
|---|---|---|---|---|---|---|---|---|---|---|---|---|
| European | CIDI-MDD | Discontinuation vs Continuation | Schizophrenia | (Intercept) | 0.254 | 0.166 | 0.389 | 2.45e-10 | NA | 2956 | 2349 | 607 |
| European | CIDI-MDD | Discontinuation vs Continuation | Schizophrenia | age_at_first_AD | 1.002 | 0.996 | 1.008 | 6.09e-01 | NA | 2956 | 2349 | 607 |
| European | CIDI-MDD | Discontinuation vs Continuation | Schizophrenia | sex_at_birthMale | 1.075 | 0.874 | 1.322 | 4.93e-01 | NA | 2956 | 2349 | 607 |
| European | CIDI-MDD | Discontinuation vs Continuation | Schizophrenia | Income_quantitative | 0.955 | 0.913 | 0.999 | 4.33e-02 | NA | 2956 | 2349 | 607 |
| European | CIDI-MDD | Discontinuation vs Continuation | Schizophrenia | Education_levelCollege Undergraduate | 1.211 | 0.989 | 1.483 | 6.46e-02 | NA | 2956 | 2349 | 607 |
| European | CIDI-MDD | Discontinuation vs Continuation | Schizophrenia | Education_levelLess than a high school degree | 0.760 | 0.216 | 2.666 | 6.68e-01 | NA | 2956 | 2349 | 607 |
| European | CIDI-MDD | Discontinuation vs Continuation | Schizophrenia | Education_levelTwelve or GED | 1.175 | 0.786 | 1.755 | 4.33e-01 | NA | 2956 | 2349 | 607 |
| European | CIDI-MDD | Discontinuation vs Continuation | Schizophrenia | Schizophrenia | 1.130 | 1.030 | 1.239 | 9.56e-03 | 6.69e-02 | 2956 | 2349 | 607 |
| European | CIDI-noMDD | Discontinuation vs Continuation | ADHD | (Intercept) | 0.091 | 0.052 | 0.159 | <2.2e-16 | NA | 2586 | 2019 | 567 |
| European | CIDI-noMDD | Discontinuation vs Continuation | ADHD | age_at_first_AD | 1.016 | 1.009 | 1.024 | 1.32e-05 | NA | 2586 | 2019 | 567 |
| European | CIDI-noMDD | Discontinuation vs Continuation | ADHD | sex_at_birthMale | 1.194 | 0.979 | 1.456 | 8.08e-02 | NA | 2586 | 2019 | 567 |
| European | CIDI-noMDD | Discontinuation vs Continuation | ADHD | Income_quantitative | 0.982 | 0.932 | 1.034 | 4.95e-01 | NA | 2586 | 2019 | 567 |
| European | CIDI-noMDD | Discontinuation vs Continuation | ADHD | Education_levelCollege Undergraduate | 1.422 | 1.145 | 1.766 | 1.45e-03 | NA | 2586 | 2019 | 567 |
| European | CIDI-noMDD | Discontinuation vs Continuation | ADHD | Education_levelLess than a high school degree | 0.582 | 0.071 | 4.748 | 6.14e-01 | NA | 2586 | 2019 | 567 |
| European | CIDI-noMDD | Discontinuation vs Continuation | ADHD | Education_levelTwelve or GED | 1.361 | 0.932 | 1.988 | 1.10e-01 | NA | 2586 | 2019 | 567 |
| European | CIDI-noMDD | Discontinuation vs Continuation | ADHD | ADHD | 0.983 | 0.894 | 1.082 | 7.30e-01 | 1.00e+00 | 2586 | 2019 | 567 |
| European | CIDI-noMDD | Discontinuation vs Continuation | BMI | (Intercept) | 0.091 | 0.052 | 0.160 | <2.2e-16 | NA | 2586 | 2019 | 567 |
| European | CIDI-noMDD | Discontinuation vs Continuation | BMI | age_at_first_AD | 1.016 | 1.009 | 1.024 | 1.35e-05 | NA | 2586 | 2019 | 567 |
| European | CIDI-noMDD | Discontinuation vs Continuation | BMI | sex_at_birthMale | 1.194 | 0.979 | 1.457 | 8.02e-02 | NA | 2586 | 2019 | 567 |
| European | CIDI-noMDD | Discontinuation vs Continuation | BMI | Income_quantitative | 0.982 | 0.932 | 1.034 | 4.91e-01 | NA | 2586 | 2019 | 567 |
| European | CIDI-noMDD | Discontinuation vs Continuation | BMI | Education_levelCollege Undergraduate | 1.420 | 1.144 | 1.763 | 1.49e-03 | NA | 2586 | 2019 | 567 |
| European | CIDI-noMDD | Discontinuation vs Continuation | BMI | Education_levelLess than a high school degree | 0.580 | 0.071 | 4.724 | 6.11e-01 | NA | 2586 | 2019 | 567 |
| European | CIDI-noMDD | Discontinuation vs Continuation | BMI | Education_levelTwelve or GED | 1.359 | 0.931 | 1.983 | 1.12e-01 | NA | 2586 | 2019 | 567 |
| European | CIDI-noMDD | Discontinuation vs Continuation | BMI | BMI | 0.983 | 0.894 | 1.080 | 7.15e-01 | 1.00e+00 | 2586 | 2019 | 567 |
| European | CIDI-noMDD | Discontinuation vs Continuation | Bipolar Disorder | (Intercept) | 0.090 | 0.051 | 0.159 | <2.2e-16 | NA | 2586 | 2019 | 567 |
| European | CIDI-noMDD | Discontinuation vs Continuation | Bipolar Disorder | age_at_first_AD | 1.016 | 1.009 | 1.024 | 1.03e-05 | NA | 2586 | 2019 | 567 |
| European | CIDI-noMDD | Discontinuation vs Continuation | Bipolar Disorder | sex_at_birthMale | 1.197 | 0.981 | 1.460 | 7.62e-02 | NA | 2586 | 2019 | 567 |
| European | CIDI-noMDD | Discontinuation vs Continuation | Bipolar Disorder | Income_quantitative | 0.981 | 0.931 | 1.033 | 4.67e-01 | NA | 2586 | 2019 | 567 |
| European | CIDI-noMDD | Discontinuation vs Continuation | Bipolar Disorder | Education_levelCollege Undergraduate | 1.409 | 1.135 | 1.749 | 1.91e-03 | NA | 2586 | 2019 | 567 |
| European | CIDI-noMDD | Discontinuation vs Continuation | Bipolar Disorder | Education_levelLess than a high school degree | 0.595 | 0.073 | 4.843 | 6.27e-01 | NA | 2586 | 2019 | 567 |
| European | CIDI-noMDD | Discontinuation vs Continuation | Bipolar Disorder | Education_levelTwelve or GED | 1.325 | 0.906 | 1.937 | 1.47e-01 | NA | 2586 | 2019 | 567 |
| European | CIDI-noMDD | Discontinuation vs Continuation | Bipolar Disorder | Bipolar Disorder | 0.929 | 0.838 | 1.029 | 1.58e-01 | 1.00e+00 | 2586 | 2019 | 567 |
| European | CIDI-noMDD | Discontinuation vs Continuation | Depression | (Intercept) | 0.091 | 0.052 | 0.159 | <2.2e-16 | NA | 2586 | 2019 | 567 |
| European | CIDI-noMDD | Discontinuation vs Continuation | Depression | age_at_first_AD | 1.016 | 1.009 | 1.024 | 1.34e-05 | NA | 2586 | 2019 | 567 |
| European | CIDI-noMDD | Discontinuation vs Continuation | Depression | sex_at_birthMale | 1.198 | 0.982 | 1.462 | 7.50e-02 | NA | 2586 | 2019 | 567 |
| European | CIDI-noMDD | Discontinuation vs Continuation | Depression | Income_quantitative | 0.982 | 0.932 | 1.034 | 4.87e-01 | NA | 2586 | 2019 | 567 |
| European | CIDI-noMDD | Discontinuation vs Continuation | Depression | Education_levelCollege Undergraduate | 1.421 | 1.145 | 1.764 | 1.44e-03 | NA | 2586 | 2019 | 567 |
| European | CIDI-noMDD | Discontinuation vs Continuation | Depression | Education_levelLess than a high school degree | 0.587 | 0.072 | 4.782 | 6.18e-01 | NA | 2586 | 2019 | 567 |
| European | CIDI-noMDD | Discontinuation vs Continuation | Depression | Education_levelTwelve or GED | 1.368 | 0.937 | 1.998 | 1.05e-01 | NA | 2586 | 2019 | 567 |
| European | CIDI-noMDD | Discontinuation vs Continuation | Depression | Depression | 0.965 | 0.873 | 1.068 | 4.93e-01 | 1.00e+00 | 2586 | 2019 | 567 |
| European | CIDI-noMDD | Discontinuation vs Continuation | Insomnia | (Intercept) | 0.090 | 0.051 | 0.159 | <2.2e-16 | NA | 2586 | 2019 | 567 |
| European | CIDI-noMDD | Discontinuation vs Continuation | Insomnia | age_at_first_AD | 1.016 | 1.009 | 1.024 | 1.28e-05 | NA | 2586 | 2019 | 567 |
| European | CIDI-noMDD | Discontinuation vs Continuation | Insomnia | sex_at_birthMale | 1.195 | 0.979 | 1.457 | 7.94e-02 | NA | 2586 | 2019 | 567 |
| European | CIDI-noMDD | Discontinuation vs Continuation | Insomnia | Income_quantitative | 0.983 | 0.933 | 1.035 | 5.06e-01 | NA | 2586 | 2019 | 567 |
| European | CIDI-noMDD | Discontinuation vs Continuation | Insomnia | Education_levelCollege Undergraduate | 1.419 | 1.143 | 1.761 | 1.53e-03 | NA | 2586 | 2019 | 567 |
| European | CIDI-noMDD | Discontinuation vs Continuation | Insomnia | Education_levelLess than a high school degree | 0.576 | 0.071 | 4.689 | 6.06e-01 | NA | 2586 | 2019 | 567 |
| European | CIDI-noMDD | Discontinuation vs Continuation | Insomnia | Education_levelTwelve or GED | 1.358 | 0.930 | 1.983 | 1.13e-01 | NA | 2586 | 2019 | 567 |
| European | CIDI-noMDD | Discontinuation vs Continuation | Insomnia | Insomnia | 0.997 | 0.903 | 1.100 | 9.53e-01 | 1.00e+00 | 2586 | 2019 | 567 |
| European | CIDI-noMDD | Discontinuation vs Continuation | Migraine | (Intercept) | 0.091 | 0.052 | 0.161 | <2.2e-16 | NA | 2586 | 2019 | 567 |
| European | CIDI-noMDD | Discontinuation vs Continuation | Migraine | age_at_first_AD | 1.016 | 1.009 | 1.024 | 1.48e-05 | NA | 2586 | 2019 | 567 |
| European | CIDI-noMDD | Discontinuation vs Continuation | Migraine | sex_at_birthMale | 1.201 | 0.984 | 1.465 | 7.15e-02 | NA | 2586 | 2019 | 567 |
| European | CIDI-noMDD | Discontinuation vs Continuation | Migraine | Income_quantitative | 0.981 | 0.931 | 1.033 | 4.64e-01 | NA | 2586 | 2019 | 567 |
| European | CIDI-noMDD | Discontinuation vs Continuation | Migraine | Education_levelCollege Undergraduate | 1.424 | 1.147 | 1.767 | 1.37e-03 | NA | 2586 | 2019 | 567 |
| European | CIDI-noMDD | Discontinuation vs Continuation | Migraine | Education_levelLess than a high school degree | 0.579 | 0.071 | 4.714 | 6.09e-01 | NA | 2586 | 2019 | 567 |
| European | CIDI-noMDD | Discontinuation vs Continuation | Migraine | Education_levelTwelve or GED | 1.362 | 0.933 | 1.988 | 1.09e-01 | NA | 2586 | 2019 | 567 |
| European | CIDI-noMDD | Discontinuation vs Continuation | Migraine | Migraine | 0.939 | 0.856 | 1.029 | 1.79e-01 | 1.00e+00 | 2586 | 2019 | 567 |
| European | CIDI-noMDD | Discontinuation vs Continuation | Schizophrenia | (Intercept) | 0.090 | 0.051 | 0.158 | <2.2e-16 | NA | 2586 | 2019 | 567 |

| | | | | | | | | | | | | |
|---|---|---|---|---|---|---|---|---|---|---|---|---|
| European | CIDI-noMDD | Discontinuation vs Continuation | Schizophrenia | age_at_first_AD | 1.016 | 1.009 | 1.024 | 1.25e-05 | NA | 2586 | 2019 | 567 |
| European | CIDI-noMDD | Discontinuation vs Continuation | Schizophrenia | sex_at_birthMale | 1.196 | 0.980 | 1.459 | 7.78e-02 | NA | 2586 | 2019 | 567 |
| European | CIDI-noMDD | Discontinuation vs Continuation | Schizophrenia | Income_quantitative | 0.983 | 0.933 | 1.035 | 5.12e-01 | NA | 2586 | 2019 | 567 |
| European | CIDI-noMDD | Discontinuation vs Continuation | Schizophrenia | Education_levelCollege Undergraduate | 1.419 | 1.143 | 1.761 | 1.52e-03 | NA | 2586 | 2019 | 567 |
| European | CIDI-noMDD | Discontinuation vs Continuation | Schizophrenia | Education_levelLess than a high school degree | 0.575 | 0.071 | 4.681 | 6.05e-01 | NA | 2586 | 2019 | 567 |
| European | CIDI-noMDD | Discontinuation vs Continuation | Schizophrenia | Education_levelTwelve or GED | 1.362 | 0.933 | 1.988 | 1.10e-01 | NA | 2586 | 2019 | 567 |
| European | CIDI-noMDD | Discontinuation vs Continuation | Schizophrenia | Schizophrenia | 1.016 | 0.923 | 1.119 | 7.41e-01 | 1.00e+00 | 2586 | 2019 | 567 |
| African | Full Cohort | Discontinuation vs Continuation | ADHD | (Intercept) | 0.302 | 0.218 | 0.419 | 7.86e-13 | NA | 5743 | 4164 | 1579 |
| African | Full Cohort | Discontinuation vs Continuation | ADHD | age_at_first_AD | 1.001 | 0.997 | 1.006 | 5.41e-01 | NA | 5743 | 4164 | 1579 |
| African | Full Cohort | Discontinuation vs Continuation | ADHD | sex_at_birthMale | 1.166 | 1.023 | 1.328 | 2.15e-02 | NA | 5743 | 4164 | 1579 |
| African | Full Cohort | Discontinuation vs Continuation | ADHD | Income_quantitative | 1.009 | 0.970 | 1.049 | 6.60e-01 | NA | 5743 | 4164 | 1579 |
| African | Full Cohort | Discontinuation vs Continuation | ADHD | Education_levelCollege Undergraduate | 1.113 | 0.899 | 1.378 | 3.26e-01 | NA | 5743 | 4164 | 1579 |
| African | Full Cohort | Discontinuation vs Continuation | ADHD | Education_levelLess than a high school degree | 1.050 | 0.788 | 1.400 | 7.37e-01 | NA | 5743 | 4164 | 1579 |
| African | Full Cohort | Discontinuation vs Continuation | ADHD | Education_levelTwelve or GED | 1.158 | 0.910 | 1.473 | 2.33e-01 | NA | 5743 | 4164 | 1579 |
| African | Full Cohort | Discontinuation vs Continuation | ADHD | ADHD | 1.025 | 0.944 | 1.113 | 5.57e-01 | 1.00e+00 | 5743 | 4164 | 1579 |
| African | Full Cohort | Discontinuation vs Continuation | BMI | (Intercept) | 0.303 | 0.219 | 0.421 | 9.78e-13 | NA | 5743 | 4164 | 1579 |
| African | Full Cohort | Discontinuation vs Continuation | BMI | age_at_first_AD | 1.001 | 0.997 | 1.005 | 5.77e-01 | NA | 5743 | 4164 | 1579 |
| African | Full Cohort | Discontinuation vs Continuation | BMI | sex_at_birthMale | 1.164 | 1.022 | 1.327 | 2.25e-02 | NA | 5743 | 4164 | 1579 |
| African | Full Cohort | Discontinuation vs Continuation | BMI | Income_quantitative | 1.008 | 0.969 | 1.048 | 7.03e-01 | NA | 5743 | 4164 | 1579 |
| African | Full Cohort | Discontinuation vs Continuation | BMI | Education_levelCollege Undergraduate | 1.116 | 0.902 | 1.383 | 3.12e-01 | NA | 5743 | 4164 | 1579 |
| African | Full Cohort | Discontinuation vs Continuation | BMI | Education_levelLess than a high school degree | 1.057 | 0.793 | 1.408 | 7.07e-01 | NA | 5743 | 4164 | 1579 |
| African | Full Cohort | Discontinuation vs Continuation | BMI | Education_levelTwelve or GED | 1.163 | 0.914 | 1.480 | 2.20e-01 | NA | 5743 | 4164 | 1579 |
| African | Full Cohort | Discontinuation vs Continuation | BMI | BMI | 0.971 | 0.899 | 1.049 | 4.57e-01 | 1.00e+00 | 5743 | 4164 | 1579 |
| African | Full Cohort | Discontinuation vs Continuation | Bipolar Disorder | (Intercept) | 0.301 | 0.217 | 0.418 | 7.18e-13 | NA | 5743 | 4164 | 1579 |
| African | Full Cohort | Discontinuation vs Continuation | Bipolar Disorder | age_at_first_AD | 1.001 | 0.997 | 1.006 | 5.42e-01 | NA | 5743 | 4164 | 1579 |
| African | Full Cohort | Discontinuation vs Continuation | Bipolar Disorder | sex_at_birthMale | 1.164 | 1.022 | 1.327 | 2.26e-02 | NA | 5743 | 4164 | 1579 |
| African | Full Cohort | Discontinuation vs Continuation | Bipolar Disorder | Income_quantitative | 1.008 | 0.970 | 1.049 | 6.76e-01 | NA | 5743 | 4164 | 1579 |
| African | Full Cohort | Discontinuation vs Continuation | Bipolar Disorder | Education_levelCollege Undergraduate | 1.117 | 0.902 | 1.383 | 3.11e-01 | NA | 5743 | 4164 | 1579 |
| African | Full Cohort | Discontinuation vs Continuation | Bipolar Disorder | Education_levelLess than a high school degree | 1.055 | 0.792 | 1.406 | 7.15e-01 | NA | 5743 | 4164 | 1579 |
| African | Full Cohort | Discontinuation vs Continuation | Bipolar Disorder | Education_levelTwelve or GED | 1.165 | 0.916 | 1.483 | 2.13e-01 | NA | 5743 | 4164 | 1579 |
| African | Full Cohort | Discontinuation vs Continuation | Bipolar Disorder | Bipolar Disorder | 1.069 | 0.982 | 1.163 | 1.22e-01 | 8.51e-01 | 5743 | 4164 | 1579 |
| African | Full Cohort | Discontinuation vs Continuation | Depression | (Intercept) | 0.303 | 0.218 | 0.421 | 9.10e-13 | NA | 5743 | 4164 | 1579 |
| African | Full Cohort | Discontinuation vs Continuation | Depression | age_at_first_AD | 1.001 | 0.997 | 1.006 | 5.59e-01 | NA | 5743 | 4164 | 1579 |
| African | Full Cohort | Discontinuation vs Continuation | Depression | sex_at_birthMale | 1.164 | 1.021 | 1.327 | 2.27e-02 | NA | 5743 | 4164 | 1579 |
| African | Full Cohort | Discontinuation vs Continuation | Depression | Income_quantitative | 1.007 | 0.969 | 1.048 | 7.14e-01 | NA | 5743 | 4164 | 1579 |
| African | Full Cohort | Discontinuation vs Continuation | Depression | Education_levelCollege Undergraduate | 1.114 | 0.900 | 1.380 | 3.20e-01 | NA | 5743 | 4164 | 1579 |
| African | Full Cohort | Discontinuation vs Continuation | Depression | Education_levelLess than a high school degree | 1.055 | 0.792 | 1.406 | 7.13e-01 | NA | 5743 | 4164 | 1579 |
| African | Full Cohort | Discontinuation vs Continuation | Depression | Education_levelTwelve or GED | 1.159 | 0.911 | 1.475 | 2.28e-01 | NA | 5743 | 4164 | 1579 |
| African | Full Cohort | Discontinuation vs Continuation | Depression | Depression | 0.949 | 0.875 | 1.029 | 2.03e-01 | 1.00e+00 | 5743 | 4164 | 1579 |
| African | Full Cohort | Discontinuation vs Continuation | Insomnia | (Intercept) | 0.303 | 0.218 | 0.420 | 8.79e-13 | NA | 5743 | 4164 | 1579 |
| African | Full Cohort | Discontinuation vs Continuation | Insomnia | age_at_first_AD | 1.001 | 0.997 | 1.006 | 5.53e-01 | NA | 5743 | 4164 | 1579 |
| African | Full Cohort | Discontinuation vs Continuation | Insomnia | sex_at_birthMale | 1.165 | 1.022 | 1.328 | 2.20e-02 | NA | 5743 | 4164 | 1579 |
| African | Full Cohort | Discontinuation vs Continuation | Insomnia | Income_quantitative | 1.009 | 0.970 | 1.049 | 6.64e-01 | NA | 5743 | 4164 | 1579 |
| African | Full Cohort | Discontinuation vs Continuation | Insomnia | Education_levelCollege Undergraduate | 1.113 | 0.899 | 1.379 | 3.25e-01 | NA | 5743 | 4164 | 1579 |
| African | Full Cohort | Discontinuation vs Continuation | Insomnia | Education_levelLess than a high school degree | 1.049 | 0.787 | 1.398 | 7.46e-01 | NA | 5743 | 4164 | 1579 |
| African | Full Cohort | Discontinuation vs Continuation | Insomnia | Education_levelTwelve or GED | 1.159 | 0.911 | 1.474 | 2.30e-01 | NA | 5743 | 4164 | 1579 |
| African | Full Cohort | Discontinuation vs Continuation | Insomnia | Insomnia | 1.040 | 0.956 | 1.133 | 3.59e-01 | 1.00e+00 | 5743 | 4164 | 1579 |
| African | Full Cohort | Discontinuation vs Continuation | Migraine | (Intercept) | 0.302 | 0.218 | 0.420 | 8.36e-13 | NA | 5743 | 4164 | 1579 |
| African | Full Cohort | Discontinuation vs Continuation | Migraine | age_at_first_AD | 1.001 | 0.997 | 1.006 | 5.56e-01 | NA | 5743 | 4164 | 1579 |
| African | Full Cohort | Discontinuation vs Continuation | Migraine | sex_at_birthMale | 1.166 | 1.023 | 1.328 | 2.16e-02 | NA | 5743 | 4164 | 1579 |
| African | Full Cohort | Discontinuation vs Continuation | Migraine | Income_quantitative | 1.008 | 0.970 | 1.049 | 6.78e-01 | NA | 5743 | 4164 | 1579 |
| African | Full Cohort | Discontinuation vs Continuation | Migraine | Education_levelCollege Undergraduate | 1.115 | 0.901 | 1.381 | 3.18e-01 | NA | 5743 | 4164 | 1579 |
| African | Full Cohort | Discontinuation vs Continuation | Migraine | Education_levelLess than a high school degree | 1.054 | 0.791 | 1.404 | 7.21e-01 | NA | 5743 | 4164 | 1579 |
| African | Full Cohort | Discontinuation vs Continuation | Migraine | Education_levelTwelve or GED | 1.160 | 0.912 | 1.476 | 2.27e-01 | NA | 5743 | 4164 | 1579 |
| African | Full Cohort | Discontinuation vs Continuation | Migraine | Migraine | 0.995 | 0.927 | 1.068 | 8.81e-01 | 1.00e+00 | 5743 | 4164 | 1579 |
| African | Full Cohort | Discontinuation vs Continuation | Schizophrenia | (Intercept) | 0.303 | 0.218 | 0.420 | 9.08e-13 | NA | 5743 | 4164 | 1579 |
| African | Full Cohort | Discontinuation vs Continuation | Schizophrenia | age_at_first_AD | 1.001 | 0.997 | 1.006 | 5.59e-01 | NA | 5743 | 4164 | 1579 |

| | | | | | | | | | | | | |
|---|---|---|---|---|---|---|---|---|---|---|---|---|
| African | Full Cohort | Discontinuation vs Continuation | Schizophrenia | sex_at_birthMale | 1.165 | 1.022 | 1.328 | 2.19e-02 | NA | 5743 | 4164 | 1579 |
| African | Full Cohort | Discontinuation vs Continuation | Schizophrenia | Income_quantitative | 1.008 | 0.969 | 1.048 | 6.91e-01 | NA | 5743 | 4164 | 1579 |
| African | Full Cohort | Discontinuation vs Continuation | Schizophrenia | Education_levelCollege Undergraduate | 1.114 | 0.900 | 1.380 | 3.21e-01 | NA | 5743 | 4164 | 1579 |
| African | Full Cohort | Discontinuation vs Continuation | Schizophrenia | Education_levelLess than a high school degree | 1.054 | 0.791 | 1.405 | 7.18e-01 | NA | 5743 | 4164 | 1579 |
| African | Full Cohort | Discontinuation vs Continuation | Schizophrenia | Education_levelTwelve or GED | 1.159 | 0.911 | 1.475 | 2.29e-01 | NA | 5743 | 4164 | 1579 |
| African | Full Cohort | Discontinuation vs Continuation | Schizophrenia | Schizophrenia | 0.978 | 0.909 | 1.053 | 5.56e-01 | 1.00e+00 | 5743 | 4164 | 1579 |
| African | Recorded Depression | Discontinuation vs Continuation | ADHD | (Intercept) | 0.402 | 0.228 | 0.708 | 1.63e-03 | NA | 2233 | 1728 | 505 |
| African | Recorded Depression | Discontinuation vs Continuation | ADHD | age_at_first_AD | 0.995 | 0.988 | 1.002 | 1.71e-01 | NA | 2233 | 1728 | 505 |
| African | Recorded Depression | Discontinuation vs Continuation | ADHD | sex_at_birthMale | 1.154 | 0.915 | 1.455 | 2.28e-01 | NA | 2233 | 1728 | 505 |
| African | Recorded Depression | Discontinuation vs Continuation | ADHD | Income_quantitative | 0.958 | 0.893 | 1.029 | 2.40e-01 | NA | 2233 | 1728 | 505 |
| African | Recorded Depression | Discontinuation vs Continuation | ADHD | Education_levelCollege Undergraduate | 0.939 | 0.641 | 1.375 | 7.45e-01 | NA | 2233 | 1728 | 505 |
| African | Recorded Depression | Discontinuation vs Continuation | ADHD | Education_levelLess than a high school degree | 0.964 | 0.582 | 1.597 | 8.86e-01 | NA | 2233 | 1728 | 505 |
| African | Recorded Depression | Discontinuation vs Continuation | ADHD | Education_levelTwelve or GED | 1.016 | 0.666 | 1.549 | 9.42e-01 | NA | 2233 | 1728 | 505 |
| African | Recorded Depression | Discontinuation vs Continuation | ADHD | ADHD | 1.066 | 0.926 | 1.227 | 3.74e-01 | 1.00e+00 | 2233 | 1728 | 505 |
| African | Recorded Depression | Discontinuation vs Continuation | BMI | (Intercept) | 0.403 | 0.229 | 0.711 | 1.68e-03 | NA | 2233 | 1728 | 505 |
| African | Recorded Depression | Discontinuation vs Continuation | BMI | age_at_first_AD | 0.995 | 0.988 | 1.002 | 1.59e-01 | NA | 2233 | 1728 | 505 |
| African | Recorded Depression | Discontinuation vs Continuation | BMI | sex_at_birthMale | 1.154 | 0.915 | 1.455 | 2.27e-01 | NA | 2233 | 1728 | 505 |
| African | Recorded Depression | Discontinuation vs Continuation | BMI | Income_quantitative | 0.957 | 0.892 | 1.028 | 2.29e-01 | NA | 2233 | 1728 | 505 |
| African | Recorded Depression | Discontinuation vs Continuation | BMI | Education_levelCollege Undergraduate | 0.945 | 0.645 | 1.385 | 7.73e-01 | NA | 2233 | 1728 | 505 |
| African | Recorded Depression | Discontinuation vs Continuation | BMI | Education_levelLess than a high school degree | 0.972 | 0.586 | 1.610 | 9.11e-01 | NA | 2233 | 1728 | 505 |
| African | Recorded Depression | Discontinuation vs Continuation | BMI | Education_levelTwelve or GED | 1.025 | 0.672 | 1.564 | 9.08e-01 | NA | 2233 | 1728 | 505 |
| African | Recorded Depression | Discontinuation vs Continuation | BMI | BMI | 0.989 | 0.866 | 1.129 | 8.68e-01 | 1.00e+00 | 2233 | 1728 | 505 |
| African | Recorded Depression | Discontinuation vs Continuation | Bipolar Disorder | (Intercept) | 0.400 | 0.227 | 0.705 | 1.53e-03 | NA | 2233 | 1728 | 505 |
| African | Recorded Depression | Discontinuation vs Continuation | Bipolar Disorder | age_at_first_AD | 0.995 | 0.988 | 1.002 | 1.59e-01 | NA | 2233 | 1728 | 505 |
| African | Recorded Depression | Discontinuation vs Continuation | Bipolar Disorder | sex_at_birthMale | 1.148 | 0.910 | 1.448 | 2.44e-01 | NA | 2233 | 1728 | 505 |
| African | Recorded Depression | Discontinuation vs Continuation | Bipolar Disorder | Income_quantitative | 0.958 | 0.893 | 1.029 | 2.40e-01 | NA | 2233 | 1728 | 505 |
| African | Recorded Depression | Discontinuation vs Continuation | Bipolar Disorder | Education_levelCollege Undergraduate | 0.952 | 0.650 | 1.395 | 8.00e-01 | NA | 2233 | 1728 | 505 |
| African | Recorded Depression | Discontinuation vs Continuation | Bipolar Disorder | Education_levelLess than a high school degree | 0.978 | 0.590 | 1.620 | 9.30e-01 | NA | 2233 | 1728 | 505 |
| African | Recorded Depression | Discontinuation vs Continuation | Bipolar Disorder | Education_levelTwelve or GED | 1.036 | 0.679 | 1.580 | 8.70e-01 | NA | 2233 | 1728 | 505 |
| African | Recorded Depression | Discontinuation vs Continuation | Bipolar Disorder | Bipolar Disorder | 1.129 | 0.978 | 1.303 | 9.80e-02 | 6.86e-01 | 2233 | 1728 | 505 |
| African | Recorded Depression | Discontinuation vs Continuation | Depression | (Intercept) | 0.403 | 0.228 | 0.710 | 1.68e-03 | NA | 2233 | 1728 | 505 |
| African | Recorded Depression | Discontinuation vs Continuation | Depression | age_at_first_AD | 0.995 | 0.988 | 1.002 | 1.60e-01 | NA | 2233 | 1728 | 505 |
| African | Recorded Depression | Discontinuation vs Continuation | Depression | sex_at_birthMale | 1.155 | 0.916 | 1.456 | 2.24e-01 | NA | 2233 | 1728 | 505 |
| African | Recorded Depression | Discontinuation vs Continuation | Depression | Income_quantitative | 0.958 | 0.892 | 1.028 | 2.30e-01 | NA | 2233 | 1728 | 505 |
| African | Recorded Depression | Discontinuation vs Continuation | Depression | Education_levelCollege Undergraduate | 0.944 | 0.644 | 1.382 | 7.66e-01 | NA | 2233 | 1728 | 505 |
| African | Recorded Depression | Discontinuation vs Continuation | Depression | Education_levelLess than a high school degree | 0.971 | 0.586 | 1.609 | 9.09e-01 | NA | 2233 | 1728 | 505 |
| African | Recorded Depression | Discontinuation vs Continuation | Depression | Education_levelTwelve or GED | 1.024 | 0.671 | 1.561 | 9.14e-01 | NA | 2233 | 1728 | 505 |
| African | Recorded Depression | Discontinuation vs Continuation | Depression | Depression | 0.991 | 0.860 | 1.143 | 9.02e-01 | 1.00e+00 | 2233 | 1728 | 505 |
| African | Recorded Depression | Discontinuation vs Continuation | Insomnia | (Intercept) | 0.402 | 0.228 | 0.710 | 1.67e-03 | NA | 2233 | 1728 | 505 |
| African | Recorded Depression | Discontinuation vs Continuation | Insomnia | age_at_first_AD | 0.995 | 0.988 | 1.002 | 1.69e-01 | NA | 2233 | 1728 | 505 |
| African | Recorded Depression | Discontinuation vs Continuation | Insomnia | sex_at_birthMale | 1.153 | 0.914 | 1.454 | 2.29e-01 | NA | 2233 | 1728 | 505 |
| African | Recorded Depression | Discontinuation vs Continuation | Insomnia | Income_quantitative | 0.958 | 0.892 | 1.028 | 2.30e-01 | NA | 2233 | 1728 | 505 |
| African | Recorded Depression | Discontinuation vs Continuation | Insomnia | Education_levelCollege Undergraduate | 0.944 | 0.644 | 1.382 | 7.65e-01 | NA | 2233 | 1728 | 505 |
| African | Recorded Depression | Discontinuation vs Continuation | Insomnia | Education_levelLess than a high school degree | 0.958 | 0.578 | 1.589 | 8.68e-01 | NA | 2233 | 1728 | 505 |
| African | Recorded Depression | Discontinuation vs Continuation | Insomnia | Education_levelTwelve or GED | 1.022 | 0.670 | 1.558 | 9.21e-01 | NA | 2233 | 1728 | 505 |
| African | Recorded Depression | Discontinuation vs Continuation | Insomnia | Insomnia | 1.088 | 0.939 | 1.261 | 2.62e-01 | 1.00e+00 | 2233 | 1728 | 505 |
| African | Recorded Depression | Discontinuation vs Continuation | Migraine | (Intercept) | 0.403 | 0.228 | 0.710 | 1.67e-03 | NA | 2233 | 1728 | 505 |
| African | Recorded Depression | Discontinuation vs Continuation | Migraine | age_at_first_AD | 0.995 | 0.988 | 1.002 | 1.60e-01 | NA | 2233 | 1728 | 505 |
| African | Recorded Depression | Discontinuation vs Continuation | Migraine | sex_at_birthMale | 1.155 | 0.916 | 1.457 | 2.23e-01 | NA | 2233 | 1728 | 505 |
| African | Recorded Depression | Discontinuation vs Continuation | Migraine | Income_quantitative | 0.958 | 0.892 | 1.028 | 2.30e-01 | NA | 2233 | 1728 | 505 |
| African | Recorded Depression | Discontinuation vs Continuation | Migraine | Education_levelCollege Undergraduate | 0.944 | 0.645 | 1.383 | 7.68e-01 | NA | 2233 | 1728 | 505 |
| African | Recorded Depression | Discontinuation vs Continuation | Migraine | Education_levelLess than a high school degree | 0.970 | 0.585 | 1.607 | 9.06e-01 | NA | 2233 | 1728 | 505 |
| African | Recorded Depression | Discontinuation vs Continuation | Migraine | Education_levelTwelve or GED | 1.024 | 0.671 | 1.561 | 9.13e-01 | NA | 2233 | 1728 | 505 |
| African | Recorded Depression | Discontinuation vs Continuation | Migraine | Migraine | 0.996 | 0.881 | 1.126 | 9.44e-01 | 1.00e+00 | 2233 | 1728 | 505 |
| African | Recorded Depression | Discontinuation vs Continuation | Schizophrenia | (Intercept) | 0.402 | 0.228 | 0.709 | 1.64e-03 | NA | 2233 | 1728 | 505 |
| African | Recorded Depression | Discontinuation vs Continuation | Schizophrenia | age_at_first_AD | 0.995 | 0.988 | 1.002 | 1.62e-01 | NA | 2233 | 1728 | 505 |
| African | Recorded Depression | Discontinuation vs Continuation | Schizophrenia | sex_at_birthMale | 1.155 | 0.916 | 1.456 | 2.24e-01 | NA | 2233 | 1728 | 505 |

| | | | | | | | | | | | | |
|---|---|---|---|---|---|---|---|---|---|---|---|---|
| African | Recorded Depression | Discontinuation vs Continuation | Schizophrenia | Income_quantitative | 0.958 | 0.892 | 1.028 | 2.33e-01 | NA | 2233 | 1728 | 505 |
| African | Recorded Depression | Discontinuation vs Continuation | Schizophrenia | Education_levelCollege Undergraduate | 0.945 | 0.645 | 1.385 | 7.73e-01 | NA | 2233 | 1728 | 505 |
| African | Recorded Depression | Discontinuation vs Continuation | Schizophrenia | Education_levelLess than a high school degree | 0.971 | 0.586 | 1.609 | 9.09e-01 | NA | 2233 | 1728 | 505 |
| African | Recorded Depression | Discontinuation vs Continuation | Schizophrenia | Education_levelTwelve or GED | 1.025 | 0.672 | 1.563 | 9.10e-01 | NA | 2233 | 1728 | 505 |
| African | Recorded Depression | Discontinuation vs Continuation | Schizophrenia | Schizophrenia | 1.012 | 0.894 | 1.145 | 8.53e-01 | 1.00e+00 | 2233 | 1728 | 505 |
| African | No Recorded Depression | Discontinuation vs Continuation | ADHD | (Intercept) | 0.305 | 0.203 | 0.458 | 1.02e-08 | NA | 3510 | 2436 | 1074 |
| African | No Recorded Depression | Discontinuation vs Continuation | ADHD | age_at_first_AD | 1.002 | 0.997 | 1.008 | 4.04e-01 | NA | 3510 | 2436 | 1074 |
| African | No Recorded Depression | Discontinuation vs Continuation | ADHD | sex_at_birthMale | 1.143 | 0.975 | 1.341 | 1.01e-01 | NA | 3510 | 2436 | 1074 |
| African | No Recorded Depression | Discontinuation vs Continuation | ADHD | Income_quantitative | 1.023 | 0.976 | 1.073 | 3.39e-01 | NA | 3510 | 2436 | 1074 |
| African | No Recorded Depression | Discontinuation vs Continuation | ADHD | Education_levelCollege Undergraduate | 1.211 | 0.935 | 1.570 | 1.48e-01 | NA | 3510 | 2436 | 1074 |
| African | No Recorded Depression | Discontinuation vs Continuation | ADHD | Education_levelLess than a high school degree | 1.074 | 0.756 | 1.525 | 6.91e-01 | NA | 3510 | 2436 | 1074 |
| African | No Recorded Depression | Discontinuation vs Continuation | ADHD | Education_levelTwelve or GED | 1.228 | 0.914 | 1.650 | 1.72e-01 | NA | 3510 | 2436 | 1074 |
| African | No Recorded Depression | Discontinuation vs Continuation | ADHD | ADHD | 1.009 | 0.911 | 1.117 | 8.68e-01 | 1.00e+00 | 3510 | 2436 | 1074 |
| African | No Recorded Depression | Discontinuation vs Continuation | BMI | (Intercept) | 0.307 | 0.204 | 0.460 | 1.17e-08 | NA | 3510 | 2436 | 1074 |
| African | No Recorded Depression | Discontinuation vs Continuation | BMI | age_at_first_AD | 1.002 | 0.997 | 1.008 | 4.21e-01 | NA | 3510 | 2436 | 1074 |
| African | No Recorded Depression | Discontinuation vs Continuation | BMI | sex_at_birthMale | 1.142 | 0.974 | 1.340 | 1.02e-01 | NA | 3510 | 2436 | 1074 |
| African | No Recorded Depression | Discontinuation vs Continuation | BMI | Income_quantitative | 1.023 | 0.975 | 1.072 | 3.57e-01 | NA | 3510 | 2436 | 1074 |
| African | No Recorded Depression | Discontinuation vs Continuation | BMI | Education_levelCollege Undergraduate | 1.212 | 0.935 | 1.571 | 1.46e-01 | NA | 3510 | 2436 | 1074 |
| African | No Recorded Depression | Discontinuation vs Continuation | BMI | Education_levelLess than a high school degree | 1.078 | 0.759 | 1.531 | 6.76e-01 | NA | 3510 | 2436 | 1074 |
| African | No Recorded Depression | Discontinuation vs Continuation | BMI | Education_levelTwelve or GED | 1.231 | 0.916 | 1.654 | 1.68e-01 | NA | 3510 | 2436 | 1074 |
| African | No Recorded Depression | Discontinuation vs Continuation | BMI | BMI | 0.972 | 0.884 | 1.069 | 5.60e-01 | 1.00e+00 | 3510 | 2436 | 1074 |
| African | No Recorded Depression | Discontinuation vs Continuation | Bipolar Disorder | (Intercept) | 0.304 | 0.203 | 0.457 | 9.46e-09 | NA | 3510 | 2436 | 1074 |
| African | No Recorded Depression | Discontinuation vs Continuation | Bipolar Disorder | age_at_first_AD | 1.002 | 0.997 | 1.008 | 3.95e-01 | NA | 3510 | 2436 | 1074 |
| African | No Recorded Depression | Discontinuation vs Continuation | Bipolar Disorder | sex_at_birthMale | 1.143 | 0.974 | 1.340 | 1.01e-01 | NA | 3510 | 2436 | 1074 |
| African | No Recorded Depression | Discontinuation vs Continuation | Bipolar Disorder | Income_quantitative | 1.023 | 0.976 | 1.073 | 3.45e-01 | NA | 3510 | 2436 | 1074 |
| African | No Recorded Depression | Discontinuation vs Continuation | Bipolar Disorder | Education_levelCollege Undergraduate | 1.212 | 0.935 | 1.571 | 1.46e-01 | NA | 3510 | 2436 | 1074 |
| African | No Recorded Depression | Discontinuation vs Continuation | Bipolar Disorder | Education_levelLess than a high school degree | 1.075 | 0.757 | 1.526 | 6.87e-01 | NA | 3510 | 2436 | 1074 |
| African | No Recorded Depression | Discontinuation vs Continuation | Bipolar Disorder | Education_levelTwelve or GED | 1.232 | 0.917 | 1.656 | 1.66e-01 | NA | 3510 | 2436 | 1074 |
| African | No Recorded Depression | Discontinuation vs Continuation | Bipolar Disorder | Bipolar Disorder | 1.050 | 0.945 | 1.167 | 3.61e-01 | 1.00e+00 | 3510 | 2436 | 1074 |
| African | No Recorded Depression | Discontinuation vs Continuation | Depression | (Intercept) | 0.307 | 0.204 | 0.460 | 1.14e-08 | NA | 3510 | 2436 | 1074 |
| African | No Recorded Depression | Discontinuation vs Continuation | Depression | age_at_first_AD | 1.002 | 0.997 | 1.008 | 4.06e-01 | NA | 3510 | 2436 | 1074 |
| African | No Recorded Depression | Discontinuation vs Continuation | Depression | sex_at_birthMale | 1.140 | 0.972 | 1.337 | 1.07e-01 | NA | 3510 | 2436 | 1074 |
| African | No Recorded Depression | Discontinuation vs Continuation | Depression | Income_quantitative | 1.022 | 0.974 | 1.071 | 3.79e-01 | NA | 3510 | 2436 | 1074 |
| African | No Recorded Depression | Discontinuation vs Continuation | Depression | Education_levelCollege Undergraduate | 1.212 | 0.935 | 1.571 | 1.46e-01 | NA | 3510 | 2436 | 1074 |
| African | No Recorded Depression | Discontinuation vs Continuation | Depression | Education_levelLess than a high school degree | 1.075 | 0.757 | 1.527 | 6.86e-01 | NA | 3510 | 2436 | 1074 |
| African | No Recorded Depression | Discontinuation vs Continuation | Depression | Education_levelTwelve or GED | 1.227 | 0.913 | 1.648 | 1.75e-01 | NA | 3510 | 2436 | 1074 |
| African | No Recorded Depression | Discontinuation vs Continuation | Depression | Depression | 0.934 | 0.846 | 1.032 | 1.80e-01 | 1.00e+00 | 3510 | 2436 | 1074 |
| African | No Recorded Depression | Discontinuation vs Continuation | Insomnia | (Intercept) | 0.306 | 0.204 | 0.459 | 1.07e-08 | NA | 3510 | 2436 | 1074 |
| African | No Recorded Depression | Discontinuation vs Continuation | Insomnia | age_at_first_AD | 1.002 | 0.997 | 1.008 | 4.10e-01 | NA | 3510 | 2436 | 1074 |
| African | No Recorded Depression | Discontinuation vs Continuation | Insomnia | sex_at_birthMale | 1.143 | 0.974 | 1.340 | 1.01e-01 | NA | 3510 | 2436 | 1074 |
| African | No Recorded Depression | Discontinuation vs Continuation | Insomnia | Income_quantitative | 1.024 | 0.976 | 1.073 | 3.36e-01 | NA | 3510 | 2436 | 1074 |
| African | No Recorded Depression | Discontinuation vs Continuation | Insomnia | Education_levelCollege Undergraduate | 1.210 | 0.934 | 1.569 | 1.50e-01 | NA | 3510 | 2436 | 1074 |
| African | No Recorded Depression | Discontinuation vs Continuation | Insomnia | Education_levelLess than a high school degree | 1.073 | 0.755 | 1.523 | 6.96e-01 | NA | 3510 | 2436 | 1074 |
| African | No Recorded Depression | Discontinuation vs Continuation | Insomnia | Education_levelTwelve or GED | 1.228 | 0.914 | 1.650 | 1.72e-01 | NA | 3510 | 2436 | 1074 |
| African | No Recorded Depression | Discontinuation vs Continuation | Insomnia | Insomnia | 1.024 | 0.923 | 1.137 | 6.53e-01 | 1.00e+00 | 3510 | 2436 | 1074 |
| African | No Recorded Depression | Discontinuation vs Continuation | Migraine | (Intercept) | 0.305 | 0.203 | 0.458 | 1.03e-08 | NA | 3510 | 2436 | 1074 |
| African | No Recorded Depression | Discontinuation vs Continuation | Migraine | age_at_first_AD | 1.002 | 0.997 | 1.008 | 4.07e-01 | NA | 3510 | 2436 | 1074 |
| African | No Recorded Depression | Discontinuation vs Continuation | Migraine | sex_at_birthMale | 1.143 | 0.974 | 1.340 | 1.01e-01 | NA | 3510 | 2436 | 1074 |
| African | No Recorded Depression | Discontinuation vs Continuation | Migraine | Income_quantitative | 1.023 | 0.976 | 1.073 | 3.42e-01 | NA | 3510 | 2436 | 1074 |
| African | No Recorded Depression | Discontinuation vs Continuation | Migraine | Education_levelCollege Undergraduate | 1.212 | 0.935 | 1.571 | 1.46e-01 | NA | 3510 | 2436 | 1074 |
| African | No Recorded Depression | Discontinuation vs Continuation | Migraine | Education_levelLess than a high school degree | 1.075 | 0.757 | 1.528 | 6.85e-01 | NA | 3510 | 2436 | 1074 |
| African | No Recorded Depression | Discontinuation vs Continuation | Migraine | Education_levelTwelve or GED | 1.229 | 0.915 | 1.651 | 1.71e-01 | NA | 3510 | 2436 | 1074 |
| African | No Recorded Depression | Discontinuation vs Continuation | Migraine | Migraine | 0.993 | 0.910 | 1.084 | 8.72e-01 | 1.00e+00 | 3510 | 2436 | 1074 |
| African | No Recorded Depression | Discontinuation vs Continuation | Schizophrenia | (Intercept) | 0.305 | 0.203 | 0.458 | 1.02e-08 | NA | 3510 | 2436 | 1074 |
| African | No Recorded Depression | Discontinuation vs Continuation | Schizophrenia | age_at_first_AD | 1.002 | 0.997 | 1.008 | 4.05e-01 | NA | 3510 | 2436 | 1074 |
| African | No Recorded Depression | Discontinuation vs Continuation | Schizophrenia | sex_at_birthMale | 1.142 | 0.973 | 1.339 | 1.03e-01 | NA | 3510 | 2436 | 1074 |
| African | No Recorded Depression | Discontinuation vs Continuation | Schizophrenia | Income_quantitative | 1.023 | 0.975 | 1.072 | 3.56e-01 | NA | 3510 | 2436 | 1074 |

| | | | | | | | | | | | | |
|---|---|---|---|---|---|---|---|---|---|---|---|---|
| African | No Recorded Depression | Discontinuation vs Continuation | Schizophrenia | Education_levelCollege Undergraduate | 1.213 | 0.936 | 1.572 | 1.45e-01 | NA | 3510 | 2436 | 1074 |
| African | No Recorded Depression | Discontinuation vs Continuation | Schizophrenia | Education_levelLess than a high school degree | 1.079 | 0.760 | 1.532 | 6.72e-01 | NA | 3510 | 2436 | 1074 |
| African | No Recorded Depression | Discontinuation vs Continuation | Schizophrenia | Education_levelTwelve or GED | 1.230 | 0.915 | 1.652 | 1.70e-01 | NA | 3510 | 2436 | 1074 |
| African | No Recorded Depression | Discontinuation vs Continuation | Schizophrenia | Schizophrenia | 0.963 | 0.878 | 1.056 | 4.17e-01 | 1.00e+00 | 3510 | 2436 | 1074 |
| African | Recorded Anxiety | Discontinuation vs Continuation | ADHD | (Intercept) | 0.335 | 0.207 | 0.544 | 9.35e-06 | NA | 2663 | 2014 | 649 |
| African | Recorded Anxiety | Discontinuation vs Continuation | ADHD | age_at_first_AD | 0.999 | 0.993 | 1.005 | 7.32e-01 | NA | 2663 | 2014 | 649 |
| African | Recorded Anxiety | Discontinuation vs Continuation | ADHD | sex_at_birthMale | 1.132 | 0.916 | 1.400 | 2.51e-01 | NA | 2663 | 2014 | 649 |
| African | Recorded Anxiety | Discontinuation vs Continuation | ADHD | Income_quantitative | 0.956 | 0.899 | 1.015 | 1.41e-01 | NA | 2663 | 2014 | 649 |
| African | Recorded Anxiety | Discontinuation vs Continuation | ADHD | Education_levelCollege Undergraduate | 1.072 | 0.783 | 1.466 | 6.66e-01 | NA | 2663 | 2014 | 649 |
| African | Recorded Anxiety | Discontinuation vs Continuation | ADHD | Education_levelLess than a high school degree | 1.113 | 0.713 | 1.736 | 6.38e-01 | NA | 2663 | 2014 | 649 |
| African | Recorded Anxiety | Discontinuation vs Continuation | ADHD | Education_levelTwelve or GED | 1.162 | 0.815 | 1.656 | 4.08e-01 | NA | 2663 | 2014 | 649 |
| African | Recorded Anxiety | Discontinuation vs Continuation | ADHD | ADHD | 1.100 | 0.970 | 1.247 | 1.36e-01 | 9.51e-01 | 2663 | 2014 | 649 |
| African | Recorded Anxiety | Discontinuation vs Continuation | BMI | (Intercept) | 0.340 | 0.209 | 0.550 | 1.17e-05 | NA | 2663 | 2014 | 649 |
| African | Recorded Anxiety | Discontinuation vs Continuation | BMI | age_at_first_AD | 0.999 | 0.992 | 1.005 | 6.42e-01 | NA | 2663 | 2014 | 649 |
| African | Recorded Anxiety | Discontinuation vs Continuation | BMI | sex_at_birthMale | 1.132 | 0.916 | 1.400 | 2.52e-01 | NA | 2663 | 2014 | 649 |
| African | Recorded Anxiety | Discontinuation vs Continuation | BMI | Income_quantitative | 0.952 | 0.896 | 1.012 | 1.15e-01 | NA | 2663 | 2014 | 649 |
| African | Recorded Anxiety | Discontinuation vs Continuation | BMI | Education_levelCollege Undergraduate | 1.088 | 0.795 | 1.489 | 6.00e-01 | NA | 2663 | 2014 | 649 |
| African | Recorded Anxiety | Discontinuation vs Continuation | BMI | Education_levelLess than a high school degree | 1.129 | 0.724 | 1.763 | 5.92e-01 | NA | 2663 | 2014 | 649 |
| African | Recorded Anxiety | Discontinuation vs Continuation | BMI | Education_levelTwelve or GED | 1.185 | 0.831 | 1.691 | 3.48e-01 | NA | 2663 | 2014 | 649 |
| African | Recorded Anxiety | Discontinuation vs Continuation | BMI | BMI | 0.914 | 0.812 | 1.028 | 1.33e-01 | 9.30e-01 | 2663 | 2014 | 649 |
| African | Recorded Anxiety | Discontinuation vs Continuation | Bipolar Disorder | (Intercept) | 0.338 | 0.208 | 0.547 | 1.05e-05 | NA | 2663 | 2014 | 649 |
| African | Recorded Anxiety | Discontinuation vs Continuation | Bipolar Disorder | age_at_first_AD | 0.999 | 0.992 | 1.005 | 6.87e-01 | NA | 2663 | 2014 | 649 |
| African | Recorded Anxiety | Discontinuation vs Continuation | Bipolar Disorder | sex_at_birthMale | 1.136 | 0.919 | 1.405 | 2.39e-01 | NA | 2663 | 2014 | 649 |
| African | Recorded Anxiety | Discontinuation vs Continuation | Bipolar Disorder | Income_quantitative | 0.955 | 0.899 | 1.015 | 1.36e-01 | NA | 2663 | 2014 | 649 |
| African | Recorded Anxiety | Discontinuation vs Continuation | Bipolar Disorder | Education_levelCollege Undergraduate | 1.076 | 0.787 | 1.472 | 6.46e-01 | NA | 2663 | 2014 | 649 |
| African | Recorded Anxiety | Discontinuation vs Continuation | Bipolar Disorder | Education_levelLess than a high school degree | 1.118 | 0.717 | 1.744 | 6.23e-01 | NA | 2663 | 2014 | 649 |
| African | Recorded Anxiety | Discontinuation vs Continuation | Bipolar Disorder | Education_levelTwelve or GED | 1.169 | 0.820 | 1.667 | 3.88e-01 | NA | 2663 | 2014 | 649 |
| African | Recorded Anxiety | Discontinuation vs Continuation | Bipolar Disorder | Bipolar Disorder | 1.028 | 0.904 | 1.169 | 6.73e-01 | 1.00e+00 | 2663 | 2014 | 649 |
| African | Recorded Anxiety | Discontinuation vs Continuation | Depression | (Intercept) | 0.337 | 0.208 | 0.547 | 1.04e-05 | NA | 2663 | 2014 | 649 |
| African | Recorded Anxiety | Discontinuation vs Continuation | Depression | age_at_first_AD | 0.999 | 0.992 | 1.005 | 6.89e-01 | NA | 2663 | 2014 | 649 |
| African | Recorded Anxiety | Discontinuation vs Continuation | Depression | sex_at_birthMale | 1.137 | 0.919 | 1.406 | 2.37e-01 | NA | 2663 | 2014 | 649 |
| African | Recorded Anxiety | Discontinuation vs Continuation | Depression | Income_quantitative | 0.955 | 0.899 | 1.014 | 1.35e-01 | NA | 2663 | 2014 | 649 |
| African | Recorded Anxiety | Discontinuation vs Continuation | Depression | Education_levelCollege Undergraduate | 1.078 | 0.788 | 1.474 | 6.41e-01 | NA | 2663 | 2014 | 649 |
| African | Recorded Anxiety | Discontinuation vs Continuation | Depression | Education_levelLess than a high school degree | 1.118 | 0.717 | 1.745 | 6.22e-01 | NA | 2663 | 2014 | 649 |
| African | Recorded Anxiety | Discontinuation vs Continuation | Depression | Education_levelTwelve or GED | 1.168 | 0.819 | 1.664 | 3.92e-01 | NA | 2663 | 2014 | 649 |
| African | Recorded Anxiety | Discontinuation vs Continuation | Depression | Depression | 1.014 | 0.895 | 1.148 | 8.32e-01 | 1.00e+00 | 2663 | 2014 | 649 |
| African | Recorded Anxiety | Discontinuation vs Continuation | Insomnia | (Intercept) | 0.339 | 0.209 | 0.549 | 1.14e-05 | NA | 2663 | 2014 | 649 |
| African | Recorded Anxiety | Discontinuation vs Continuation | Insomnia | age_at_first_AD | 0.999 | 0.992 | 1.005 | 6.89e-01 | NA | 2663 | 2014 | 649 |
| African | Recorded Anxiety | Discontinuation vs Continuation | Insomnia | sex_at_birthMale | 1.134 | 0.917 | 1.403 | 2.45e-01 | NA | 2663 | 2014 | 649 |
| African | Recorded Anxiety | Discontinuation vs Continuation | Insomnia | Income_quantitative | 0.955 | 0.899 | 1.014 | 1.35e-01 | NA | 2663 | 2014 | 649 |
| African | Recorded Anxiety | Discontinuation vs Continuation | Insomnia | Education_levelCollege Undergraduate | 1.074 | 0.785 | 1.470 | 6.54e-01 | NA | 2663 | 2014 | 649 |
| African | Recorded Anxiety | Discontinuation vs Continuation | Insomnia | Education_levelLess than a high school degree | 1.113 | 0.713 | 1.736 | 6.38e-01 | NA | 2663 | 2014 | 649 |
| African | Recorded Anxiety | Discontinuation vs Continuation | Insomnia | Education_levelTwelve or GED | 1.164 | 0.817 | 1.660 | 4.00e-01 | NA | 2663 | 2014 | 649 |
| African | Recorded Anxiety | Discontinuation vs Continuation | Insomnia | Insomnia | 1.057 | 0.929 | 1.203 | 3.98e-01 | 1.00e+00 | 2663 | 2014 | 649 |
| African | Recorded Anxiety | Discontinuation vs Continuation | Migraine | (Intercept) | 0.340 | 0.209 | 0.551 | 1.19e-05 | NA | 2663 | 2014 | 649 |
| African | Recorded Anxiety | Discontinuation vs Continuation | Migraine | age_at_first_AD | 0.999 | 0.992 | 1.005 | 6.85e-01 | NA | 2663 | 2014 | 649 |
| African | Recorded Anxiety | Discontinuation vs Continuation | Migraine | sex_at_birthMale | 1.135 | 0.918 | 1.403 | 2.44e-01 | NA | 2663 | 2014 | 649 |
| African | Recorded Anxiety | Discontinuation vs Continuation | Migraine | Income_quantitative | 0.954 | 0.898 | 1.014 | 1.27e-01 | NA | 2663 | 2014 | 649 |
| African | Recorded Anxiety | Discontinuation vs Continuation | Migraine | Education_levelCollege Undergraduate | 1.073 | 0.784 | 1.468 | 6.60e-01 | NA | 2663 | 2014 | 649 |
| African | Recorded Anxiety | Discontinuation vs Continuation | Migraine | Education_levelLess than a high school degree | 1.116 | 0.715 | 1.741 | 6.29e-01 | NA | 2663 | 2014 | 649 |
| African | Recorded Anxiety | Discontinuation vs Continuation | Migraine | Education_levelTwelve or GED | 1.162 | 0.815 | 1.657 | 4.08e-01 | NA | 2663 | 2014 | 649 |
| African | Recorded Anxiety | Discontinuation vs Continuation | Migraine | Migraine | 1.049 | 0.940 | 1.170 | 3.94e-01 | 1.00e+00 | 2663 | 2014 | 649 |
| African | Recorded Anxiety | Discontinuation vs Continuation | Schizophrenia | (Intercept) | 0.339 | 0.209 | 0.550 | 1.17e-05 | NA | 2663 | 2014 | 649 |
| African | Recorded Anxiety | Discontinuation vs Continuation | Schizophrenia | age_at_first_AD | 0.999 | 0.992 | 1.005 | 6.83e-01 | NA | 2663 | 2014 | 649 |
| African | Recorded Anxiety | Discontinuation vs Continuation | Schizophrenia | sex_at_birthMale | 1.136 | 0.919 | 1.404 | 2.40e-01 | NA | 2663 | 2014 | 649 |
| African | Recorded Anxiety | Discontinuation vs Continuation | Schizophrenia | Income_quantitative | 0.953 | 0.897 | 1.013 | 1.23e-01 | NA | 2663 | 2014 | 649 |
| African | Recorded Anxiety | Discontinuation vs Continuation | Schizophrenia | Education_levelCollege Undergraduate | 1.075 | 0.786 | 1.471 | 6.51e-01 | NA | 2663 | 2014 | 649 |

| | | | | | | | | | | | | |
|---|---|---|---|---|---|---|---|---|---|---|---|---|
| African | Recorded Anxiety | Discontinuation vs Continuation | Schizophrenia | Education_levelLess than a high school degree | 1.128 | 0.723 | 1.760 | 5.97e-01 | NA | 2663 | 2014 | 649 |
| African | Recorded Anxiety | Discontinuation vs Continuation | Schizophrenia | Education_levelTwelve or GED | 1.165 | 0.817 | 1.661 | 3.98e-01 | NA | 2663 | 2014 | 649 |
| African | Recorded Anxiety | Discontinuation vs Continuation | Schizophrenia | Schizophrenia | 0.919 | 0.822 | 1.029 | 1.44e-01 | 1.00e+00 | 2663 | 2014 | 649 |
| Latino/Mixed-American | Full Cohort | Discontinuation vs Continuation | ADHD | (Intercept) | 0.408 | 0.283 | 0.589 | 1.71e-06 | NA | 3688 | 2456 | 1232 |
| Latino/Mixed-American | Full Cohort | Discontinuation vs Continuation | ADHD | age_at_first_AD | 1.000 | 0.995 | 1.004 | 8.46e-01 | NA | 3688 | 2456 | 1232 |
| Latino/Mixed-American | Full Cohort | Discontinuation vs Continuation | ADHD | sex_at_birthMale | 1.207 | 1.037 | 1.405 | 1.54e-02 | NA | 3688 | 2456 | 1232 |
| Latino/Mixed-American | Full Cohort | Discontinuation vs Continuation | ADHD | Income_quantitative | 0.959 | 0.922 | 0.999 | 4.19e-02 | NA | 3688 | 2456 | 1232 |
| Latino/Mixed-American | Full Cohort | Discontinuation vs Continuation | ADHD | Education_levelCollege Undergraduate | 1.465 | 1.128 | 1.902 | 4.24e-03 | NA | 3688 | 2456 | 1232 |
| Latino/Mixed-American | Full Cohort | Discontinuation vs Continuation | ADHD | Education_levelLess than a high school degree | 1.108 | 0.806 | 1.525 | 5.28e-01 | NA | 3688 | 2456 | 1232 |
| Latino/Mixed-American | Full Cohort | Discontinuation vs Continuation | ADHD | Education_levelTwelve or GED | 1.415 | 1.048 | 1.911 | 2.32e-02 | NA | 3688 | 2456 | 1232 |
| Latino/Mixed-American | Full Cohort | Discontinuation vs Continuation | ADHD | ADHD | 1.032 | 0.954 | 1.116 | 4.31e-01 | 1.00e+00 | 3688 | 2456 | 1232 |
| Latino/Mixed-American | Full Cohort | Discontinuation vs Continuation | BMI | (Intercept) | 0.406 | 0.281 | 0.587 | 1.54e-06 | NA | 3688 | 2456 | 1232 |
| Latino/Mixed-American | Full Cohort | Discontinuation vs Continuation | BMI | age_at_first_AD | 1.000 | 0.995 | 1.004 | 8.30e-01 | NA | 3688 | 2456 | 1232 |
| Latino/Mixed-American | Full Cohort | Discontinuation vs Continuation | BMI | sex_at_birthMale | 1.209 | 1.038 | 1.407 | 1.47e-02 | NA | 3688 | 2456 | 1232 |
| Latino/Mixed-American | Full Cohort | Discontinuation vs Continuation | BMI | Income_quantitative | 0.962 | 0.924 | 1.001 | 5.92e-02 | NA | 3688 | 2456 | 1232 |
| Latino/Mixed-American | Full Cohort | Discontinuation vs Continuation | BMI | Education_levelCollege Undergraduate | 1.461 | 1.125 | 1.898 | 4.51e-03 | NA | 3688 | 2456 | 1232 |
| Latino/Mixed-American | Full Cohort | Discontinuation vs Continuation | BMI | Education_levelLess than a high school degree | 1.107 | 0.805 | 1.523 | 5.32e-01 | NA | 3688 | 2456 | 1232 |
| Latino/Mixed-American | Full Cohort | Discontinuation vs Continuation | BMI | Education_levelTwelve or GED | 1.413 | 1.047 | 1.907 | 2.39e-02 | NA | 3688 | 2456 | 1232 |
| Latino/Mixed-American | Full Cohort | Discontinuation vs Continuation | BMI | BMI | 1.087 | 1.012 | 1.168 | 2.25e-02 | 1.57e-01 | 3688 | 2456 | 1232 |
| Latino/Mixed-American | Full Cohort | Discontinuation vs Continuation | Bipolar Disorder | (Intercept) | 0.409 | 0.283 | 0.590 | 1.77e-06 | NA | 3688 | 2456 | 1232 |
| Latino/Mixed-American | Full Cohort | Discontinuation vs Continuation | Bipolar Disorder | age_at_first_AD | 0.999 | 0.995 | 1.004 | 8.03e-01 | NA | 3688 | 2456 | 1232 |
| Latino/Mixed-American | Full Cohort | Discontinuation vs Continuation | Bipolar Disorder | sex_at_birthMale | 1.209 | 1.039 | 1.408 | 1.44e-02 | NA | 3688 | 2456 | 1232 |
| Latino/Mixed-American | Full Cohort | Discontinuation vs Continuation | Bipolar Disorder | Income_quantitative | 0.958 | 0.921 | 0.998 | 3.75e-02 | NA | 3688 | 2456 | 1232 |
| Latino/Mixed-American | Full Cohort | Discontinuation vs Continuation | Bipolar Disorder | Education_levelCollege Undergraduate | 1.473 | 1.134 | 1.913 | 3.71e-03 | NA | 3688 | 2456 | 1232 |
| Latino/Mixed-American | Full Cohort | Discontinuation vs Continuation | Bipolar Disorder | Education_levelLess than a high school degree | 1.118 | 0.813 | 1.537 | 4.93e-01 | NA | 3688 | 2456 | 1232 |
| Latino/Mixed-American | Full Cohort | Discontinuation vs Continuation | Bipolar Disorder | Education_levelTwelve or GED | 1.428 | 1.058 | 1.928 | 1.98e-02 | NA | 3688 | 2456 | 1232 |
| Latino/Mixed-American | Full Cohort | Discontinuation vs Continuation | Bipolar Disorder | Bipolar Disorder | 1.044 | 0.967 | 1.127 | 2.74e-01 | 1.00e+00 | 3688 | 2456 | 1232 |
| Latino/Mixed-American | Full Cohort | Discontinuation vs Continuation | Depression | (Intercept) | 0.407 | 0.282 | 0.588 | 1.62e-06 | NA | 3688 | 2456 | 1232 |
| Latino/Mixed-American | Full Cohort | Discontinuation vs Continuation | Depression | age_at_first_AD | 1.000 | 0.995 | 1.004 | 9.06e-01 | NA | 3688 | 2456 | 1232 |
| Latino/Mixed-American | Full Cohort | Discontinuation vs Continuation | Depression | sex_at_birthMale | 1.205 | 1.035 | 1.404 | 1.62e-02 | NA | 3688 | 2456 | 1232 |
| Latino/Mixed-American | Full Cohort | Discontinuation vs Continuation | Depression | Income_quantitative | 0.960 | 0.922 | 0.999 | 4.64e-02 | NA | 3688 | 2456 | 1232 |
| Latino/Mixed-American | Full Cohort | Discontinuation vs Continuation | Depression | Education_levelCollege Undergraduate | 1.454 | 1.119 | 1.889 | 5.03e-03 | NA | 3688 | 2456 | 1232 |
| Latino/Mixed-American | Full Cohort | Discontinuation vs Continuation | Depression | Education_levelLess than a high school degree | 1.096 | 0.797 | 1.508 | 5.71e-01 | NA | 3688 | 2456 | 1232 |
| Latino/Mixed-American | Full Cohort | Discontinuation vs Continuation | Depression | Education_levelTwelve or GED | 1.408 | 1.043 | 1.901 | 2.53e-02 | NA | 3688 | 2456 | 1232 |
| Latino/Mixed-American | Full Cohort | Discontinuation vs Continuation | Depression | Depression | 1.122 | 1.038 | 1.213 | 3.81e-03 | 2.67e-02 | 3688 | 2456 | 1232 |
| Latino/Mixed-American | Full Cohort | Discontinuation vs Continuation | Insomnia | (Intercept) | 0.409 | 0.284 | 0.591 | 1.85e-06 | NA | 3688 | 2456 | 1232 |
| Latino/Mixed-American | Full Cohort | Discontinuation vs Continuation | Insomnia | age_at_first_AD | 0.999 | 0.995 | 1.004 | 8.14e-01 | NA | 3688 | 2456 | 1232 |
| Latino/Mixed-American | Full Cohort | Discontinuation vs Continuation | Insomnia | sex_at_birthMale | 1.207 | 1.037 | 1.405 | 1.53e-02 | NA | 3688 | 2456 | 1232 |
| Latino/Mixed-American | Full Cohort | Discontinuation vs Continuation | Insomnia | Income_quantitative | 0.959 | 0.921 | 0.998 | 3.81e-02 | NA | 3688 | 2456 | 1232 |
| Latino/Mixed-American | Full Cohort | Discontinuation vs Continuation | Insomnia | Education_levelCollege Undergraduate | 1.468 | 1.130 | 1.907 | 3.99e-03 | NA | 3688 | 2456 | 1232 |
| Latino/Mixed-American | Full Cohort | Discontinuation vs Continuation | Insomnia | Education_levelLess than a high school degree | 1.114 | 0.810 | 1.532 | 5.08e-01 | NA | 3688 | 2456 | 1232 |
| Latino/Mixed-American | Full Cohort | Discontinuation vs Continuation | Insomnia | Education_levelTwelve or GED | 1.423 | 1.055 | 1.921 | 2.10e-02 | NA | 3688 | 2456 | 1232 |
| Latino/Mixed-American | Full Cohort | Discontinuation vs Continuation | Insomnia | Insomnia | 1.006 | 0.929 | 1.089 | 8.84e-01 | 1.00e+00 | 3688 | 2456 | 1232 |
| Latino/Mixed-American | Full Cohort | Discontinuation vs Continuation | Migraine | (Intercept) | 0.409 | 0.284 | 0.591 | 1.86e-06 | NA | 3688 | 2456 | 1232 |
| Latino/Mixed-American | Full Cohort | Discontinuation vs Continuation | Migraine | age_at_first_AD | 1.000 | 0.995 | 1.004 | 8.26e-01 | NA | 3688 | 2456 | 1232 |
| Latino/Mixed-American | Full Cohort | Discontinuation vs Continuation | Migraine | sex_at_birthMale | 1.208 | 1.037 | 1.406 | 1.49e-02 | NA | 3688 | 2456 | 1232 |
| Latino/Mixed-American | Full Cohort | Discontinuation vs Continuation | Migraine | Income_quantitative | 0.958 | 0.921 | 0.998 | 3.77e-02 | NA | 3688 | 2456 | 1232 |
| Latino/Mixed-American | Full Cohort | Discontinuation vs Continuation | Migraine | Education_levelCollege Undergraduate | 1.465 | 1.128 | 1.903 | 4.18e-03 | NA | 3688 | 2456 | 1232 |
| Latino/Mixed-American | Full Cohort | Discontinuation vs Continuation | Migraine | Education_levelLess than a high school degree | 1.111 | 0.808 | 1.528 | 5.17e-01 | NA | 3688 | 2456 | 1232 |
| Latino/Mixed-American | Full Cohort | Discontinuation vs Continuation | Migraine | Education_levelTwelve or GED | 1.419 | 1.051 | 1.915 | 2.22e-02 | NA | 3688 | 2456 | 1232 |
| Latino/Mixed-American | Full Cohort | Discontinuation vs Continuation | Migraine | Migraine | 1.044 | 0.971 | 1.122 | 2.48e-01 | 1.00e+00 | 3688 | 2456 | 1232 |
| Latino/Mixed-American | Full Cohort | Discontinuation vs Continuation | Schizophrenia | (Intercept) | 0.408 | 0.283 | 0.589 | 1.73e-06 | NA | 3688 | 2456 | 1232 |
| Latino/Mixed-American | Full Cohort | Discontinuation vs Continuation | Schizophrenia | age_at_first_AD | 0.999 | 0.995 | 1.004 | 7.91e-01 | NA | 3688 | 2456 | 1232 |
| Latino/Mixed-American | Full Cohort | Discontinuation vs Continuation | Schizophrenia | sex_at_birthMale | 1.206 | 1.036 | 1.405 | 1.57e-02 | NA | 3688 | 2456 | 1232 |
| Latino/Mixed-American | Full Cohort | Discontinuation vs Continuation | Schizophrenia | Income_quantitative | 0.959 | 0.921 | 0.998 | 4.04e-02 | NA | 3688 | 2456 | 1232 |
| Latino/Mixed-American | Full Cohort | Discontinuation vs Continuation | Schizophrenia | Education_levelCollege Undergraduate | 1.476 | 1.136 | 1.917 | 3.55e-03 | NA | 3688 | 2456 | 1232 |
| Latino/Mixed-American | Full Cohort | Discontinuation vs Continuation | Schizophrenia | Education_levelLess than a high school degree | 1.120 | 0.815 | 1.541 | 4.84e-01 | NA | 3688 | 2456 | 1232 |

| | | | | | | | | | | | | |
|---|---|---|---|---|---|---|---|---|---|---|---|---|
| Latino/Mixed-American | Full Cohort | Discontinuation vs Continuation | Schizophrenia | Education_levelTwelve or GED | 1.428 | 1.058 | 1.927 | 1.99e-02 | NA | 3688 | 2456 | 1232 |
| Latino/Mixed-American | Full Cohort | Discontinuation vs Continuation | Schizophrenia | Schizophrenia | 1.039 | 0.969 | 1.115 | 2.83e-01 | 1.00e+00 | 3688 | 2456 | 1232 |
| Latino/Mixed-American | Recorded Depression | Discontinuation vs Continuation | ADHD | (Intercept) | 0.367 | 0.201 | 0.668 | 1.05e-03 | NA | 1698 | 1179 | 519 |
| Latino/Mixed-American | Recorded Depression | Discontinuation vs Continuation | ADHD | age_at_first_AD | 0.997 | 0.990 | 1.004 | 4.14e-01 | NA | 1698 | 1179 | 519 |
| Latino/Mixed-American | Recorded Depression | Discontinuation vs Continuation | ADHD | sex_at_birthMale | 1.117 | 0.887 | 1.407 | 3.46e-01 | NA | 1698 | 1179 | 519 |
| Latino/Mixed-American | Recorded Depression | Discontinuation vs Continuation | ADHD | Income_quantitative | 0.974 | 0.913 | 1.038 | 4.14e-01 | NA | 1698 | 1179 | 519 |
| Latino/Mixed-American | Recorded Depression | Discontinuation vs Continuation | ADHD | Education_levelCollege Undergraduate | 1.606 | 1.028 | 2.508 | 3.75e-02 | NA | 1698 | 1179 | 519 |
| Latino/Mixed-American | Recorded Depression | Discontinuation vs Continuation | ADHD | Education_levelLess than a high school degree | 1.300 | 0.776 | 2.178 | 3.18e-01 | NA | 1698 | 1179 | 519 |
| Latino/Mixed-American | Recorded Depression | Discontinuation vs Continuation | ADHD | Education_levelTwelve or GED | 1.318 | 0.805 | 2.158 | 2.73e-01 | NA | 1698 | 1179 | 519 |
| Latino/Mixed-American | Recorded Depression | Discontinuation vs Continuation | ADHD | ADHD | 0.955 | 0.849 | 1.074 | 4.37e-01 | 1.00e+00 | 1698 | 1179 | 519 |
| Latino/Mixed-American | Recorded Depression | Discontinuation vs Continuation | BMI | (Intercept) | 0.362 | 0.198 | 0.660 | 9.23e-04 | NA | 1698 | 1179 | 519 |
| Latino/Mixed-American | Recorded Depression | Discontinuation vs Continuation | BMI | age_at_first_AD | 0.997 | 0.990 | 1.004 | 4.08e-01 | NA | 1698 | 1179 | 519 |
| Latino/Mixed-American | Recorded Depression | Discontinuation vs Continuation | BMI | sex_at_birthMale | 1.118 | 0.888 | 1.408 | 3.44e-01 | NA | 1698 | 1179 | 519 |
| Latino/Mixed-American | Recorded Depression | Discontinuation vs Continuation | BMI | Income_quantitative | 0.980 | 0.919 | 1.046 | 5.43e-01 | NA | 1698 | 1179 | 519 |
| Latino/Mixed-American | Recorded Depression | Discontinuation vs Continuation | BMI | Education_levelCollege Undergraduate | 1.601 | 1.024 | 2.502 | 3.89e-02 | NA | 1698 | 1179 | 519 |
| Latino/Mixed-American | Recorded Depression | Discontinuation vs Continuation | BMI | Education_levelLess than a high school degree | 1.280 | 0.764 | 2.145 | 3.49e-01 | NA | 1698 | 1179 | 519 |
| Latino/Mixed-American | Recorded Depression | Discontinuation vs Continuation | BMI | Education_levelTwelve or GED | 1.311 | 0.800 | 2.149 | 2.82e-01 | NA | 1698 | 1179 | 519 |
| Latino/Mixed-American | Recorded Depression | Discontinuation vs Continuation | BMI | BMI | 1.140 | 1.022 | 1.271 | 1.87e-02 | 1.31e-01 | 1698 | 1179 | 519 |
| Latino/Mixed-American | Recorded Depression | Discontinuation vs Continuation | Bipolar Disorder | (Intercept) | 0.362 | 0.199 | 0.659 | 8.95e-04 | NA | 1698 | 1179 | 519 |
| Latino/Mixed-American | Recorded Depression | Discontinuation vs Continuation | Bipolar Disorder | age_at_first_AD | 0.997 | 0.990 | 1.004 | 4.37e-01 | NA | 1698 | 1179 | 519 |
| Latino/Mixed-American | Recorded Depression | Discontinuation vs Continuation | Bipolar Disorder | sex_at_birthMale | 1.120 | 0.889 | 1.410 | 3.37e-01 | NA | 1698 | 1179 | 519 |
| Latino/Mixed-American | Recorded Depression | Discontinuation vs Continuation | Bipolar Disorder | Income_quantitative | 0.975 | 0.914 | 1.039 | 4.31e-01 | NA | 1698 | 1179 | 519 |
| Latino/Mixed-American | Recorded Depression | Discontinuation vs Continuation | Bipolar Disorder | Education_levelCollege Undergraduate | 1.614 | 1.033 | 2.523 | 3.54e-02 | NA | 1698 | 1179 | 519 |
| Latino/Mixed-American | Recorded Depression | Discontinuation vs Continuation | Bipolar Disorder | Education_levelLess than a high school degree | 1.305 | 0.779 | 2.187 | 3.12e-01 | NA | 1698 | 1179 | 519 |
| Latino/Mixed-American | Recorded Depression | Discontinuation vs Continuation | Bipolar Disorder | Education_levelTwelve or GED | 1.325 | 0.809 | 2.170 | 2.64e-01 | NA | 1698 | 1179 | 519 |
| Latino/Mixed-American | Recorded Depression | Discontinuation vs Continuation | Bipolar Disorder | Bipolar Disorder | 1.035 | 0.924 | 1.160 | 5.50e-01 | 1.00e+00 | 1698 | 1179 | 519 |
| Latino/Mixed-American | Recorded Depression | Discontinuation vs Continuation | Depression | (Intercept) | 0.364 | 0.200 | 0.664 | 9.69e-04 | NA | 1698 | 1179 | 519 |
| Latino/Mixed-American | Recorded Depression | Discontinuation vs Continuation | Depression | age_at_first_AD | 0.997 | 0.990 | 1.005 | 4.68e-01 | NA | 1698 | 1179 | 519 |
| Latino/Mixed-American | Recorded Depression | Discontinuation vs Continuation | Depression | sex_at_birthMale | 1.114 | 0.885 | 1.403 | 3.58e-01 | NA | 1698 | 1179 | 519 |
| Latino/Mixed-American | Recorded Depression | Discontinuation vs Continuation | Depression | Income_quantitative | 0.975 | 0.914 | 1.040 | 4.41e-01 | NA | 1698 | 1179 | 519 |
| Latino/Mixed-American | Recorded Depression | Discontinuation vs Continuation | Depression | Education_levelCollege Undergraduate | 1.594 | 1.020 | 2.491 | 4.05e-02 | NA | 1698 | 1179 | 519 |
| Latino/Mixed-American | Recorded Depression | Discontinuation vs Continuation | Depression | Education_levelLess than a high school degree | 1.280 | 0.764 | 2.144 | 3.49e-01 | NA | 1698 | 1179 | 519 |
| Latino/Mixed-American | Recorded Depression | Discontinuation vs Continuation | Depression | Education_levelTwelve or GED | 1.305 | 0.797 | 2.137 | 2.91e-01 | NA | 1698 | 1179 | 519 |
| Latino/Mixed-American | Recorded Depression | Discontinuation vs Continuation | Depression | Depression | 1.088 | 0.967 | 1.225 | 1.62e-01 | 1.00e+00 | 1698 | 1179 | 519 |
| Latino/Mixed-American | Recorded Depression | Discontinuation vs Continuation | Insomnia | (Intercept) | 0.363 | 0.199 | 0.661 | 9.29e-04 | NA | 1698 | 1179 | 519 |
| Latino/Mixed-American | Recorded Depression | Discontinuation vs Continuation | Insomnia | age_at_first_AD | 0.997 | 0.990 | 1.004 | 4.34e-01 | NA | 1698 | 1179 | 519 |
| Latino/Mixed-American | Recorded Depression | Discontinuation vs Continuation | Insomnia | sex_at_birthMale | 1.119 | 0.889 | 1.409 | 3.37e-01 | NA | 1698 | 1179 | 519 |
| Latino/Mixed-American | Recorded Depression | Discontinuation vs Continuation | Insomnia | Income_quantitative | 0.974 | 0.914 | 1.039 | 4.26e-01 | NA | 1698 | 1179 | 519 |
| Latino/Mixed-American | Recorded Depression | Discontinuation vs Continuation | Insomnia | Education_levelCollege Undergraduate | 1.611 | 1.031 | 2.517 | 3.62e-02 | NA | 1698 | 1179 | 519 |
| Latino/Mixed-American | Recorded Depression | Discontinuation vs Continuation | Insomnia | Education_levelLess than a high school degree | 1.299 | 0.776 | 2.176 | 3.20e-01 | NA | 1698 | 1179 | 519 |
| Latino/Mixed-American | Recorded Depression | Discontinuation vs Continuation | Insomnia | Education_levelTwelve or GED | 1.318 | 0.805 | 2.158 | 2.72e-01 | NA | 1698 | 1179 | 519 |
| Latino/Mixed-American | Recorded Depression | Discontinuation vs Continuation | Insomnia | Insomnia | 0.951 | 0.843 | 1.073 | 4.15e-01 | 1.00e+00 | 1698 | 1179 | 519 |
| Latino/Mixed-American | Recorded Depression | Discontinuation vs Continuation | Migraine | (Intercept) | 0.367 | 0.201 | 0.668 | 1.05e-03 | NA | 1698 | 1179 | 519 |
| Latino/Mixed-American | Recorded Depression | Discontinuation vs Continuation | Migraine | age_at_first_AD | 0.997 | 0.990 | 1.004 | 4.35e-01 | NA | 1698 | 1179 | 519 |
| Latino/Mixed-American | Recorded Depression | Discontinuation vs Continuation | Migraine | sex_at_birthMale | 1.120 | 0.889 | 1.410 | 3.36e-01 | NA | 1698 | 1179 | 519 |
| Latino/Mixed-American | Recorded Depression | Discontinuation vs Continuation | Migraine | Income_quantitative | 0.975 | 0.914 | 1.039 | 4.33e-01 | NA | 1698 | 1179 | 519 |
| Latino/Mixed-American | Recorded Depression | Discontinuation vs Continuation | Migraine | Education_levelCollege Undergraduate | 1.599 | 1.023 | 2.498 | 3.94e-02 | NA | 1698 | 1179 | 519 |
| Latino/Mixed-American | Recorded Depression | Discontinuation vs Continuation | Migraine | Education_levelLess than a high school degree | 1.284 | 0.766 | 2.151 | 3.43e-01 | NA | 1698 | 1179 | 519 |
| Latino/Mixed-American | Recorded Depression | Discontinuation vs Continuation | Migraine | Education_levelTwelve or GED | 1.296 | 0.791 | 2.124 | 3.03e-01 | NA | 1698 | 1179 | 519 |
| Latino/Mixed-American | Recorded Depression | Discontinuation vs Continuation | Migraine | Migraine | 1.097 | 0.980 | 1.228 | 1.06e-01 | 7.43e-01 | 1698 | 1179 | 519 |
| Latino/Mixed-American | Recorded Depression | Discontinuation vs Continuation | Schizophrenia | (Intercept) | 0.357 | 0.196 | 0.652 | 7.90e-04 | NA | 1698 | 1179 | 519 |
| Latino/Mixed-American | Recorded Depression | Discontinuation vs Continuation | Schizophrenia | age_at_first_AD | 0.997 | 0.990 | 1.004 | 4.37e-01 | NA | 1698 | 1179 | 519 |
| Latino/Mixed-American | Recorded Depression | Discontinuation vs Continuation | Schizophrenia | sex_at_birthMale | 1.120 | 0.889 | 1.411 | 3.35e-01 | NA | 1698 | 1179 | 519 |
| Latino/Mixed-American | Recorded Depression | Discontinuation vs Continuation | Schizophrenia | Income_quantitative | 0.974 | 0.913 | 1.039 | 4.19e-01 | NA | 1698 | 1179 | 519 |
| Latino/Mixed-American | Recorded Depression | Discontinuation vs Continuation | Schizophrenia | Education_levelCollege Undergraduate | 1.639 | 1.048 | 2.562 | 3.02e-02 | NA | 1698 | 1179 | 519 |
| Latino/Mixed-American | Recorded Depression | Discontinuation vs Continuation | Schizophrenia | Education_levelLess than a high school degree | 1.316 | 0.786 | 2.205 | 2.96e-01 | NA | 1698 | 1179 | 519 |
| Latino/Mixed-American | Recorded Depression | Discontinuation vs Continuation | Schizophrenia | Education_levelTwelve or GED | 1.337 | 0.816 | 2.190 | 2.49e-01 | NA | 1698 | 1179 | 519 |

| | | | | | | | | | | | | |
|---|---|---|---|---|---|---|---|---|---|---|---|---|
| Latino/Mixed-American | Recorded Depression | Discontinuation vs Continuation | Schizophrenia | Schizophrenia | 1.116 | 1.002 | 1.243 | 4.60e-02 | 3.22e-01 | 1698 | 1179 | 519 |
| Latino/Mixed-American | No Recorded Depression | Discontinuation vs Continuation | ADHD | (Intercept) | 0.475 | 0.297 | 0.760 | 1.92e-03 | NA | 1990 | 1277 | 713 |
| Latino/Mixed-American | No Recorded Depression | Discontinuation vs Continuation | ADHD | age_at_first_AD | 1.000 | 0.994 | 1.007 | 9.26e-01 | NA | 1990 | 1277 | 713 |
| Latino/Mixed-American | No Recorded Depression | Discontinuation vs Continuation | ADHD | sex_at_birthMale | 1.286 | 1.048 | 1.578 | 1.60e-02 | NA | 1990 | 1277 | 713 |
| Latino/Mixed-American | No Recorded Depression | Discontinuation vs Continuation | ADHD | Income_quantitative | 0.940 | 0.892 | 0.990 | 1.87e-02 | NA | 1990 | 1277 | 713 |
| Latino/Mixed-American | No Recorded Depression | Discontinuation vs Continuation | ADHD | Education_levelCollege Undergraduate | 1.384 | 1.001 | 1.915 | 4.95e-02 | NA | 1990 | 1277 | 713 |
| Latino/Mixed-American | No Recorded Depression | Discontinuation vs Continuation | ADHD | Education_levelLess than a high school degree | 0.991 | 0.656 | 1.498 | 9.66e-01 | NA | 1990 | 1277 | 713 |
| Latino/Mixed-American | No Recorded Depression | Discontinuation vs Continuation | ADHD | Education_levelTwelve or GED | 1.595 | 1.085 | 2.346 | 1.76e-02 | NA | 1990 | 1277 | 713 |
| Latino/Mixed-American | No Recorded Depression | Discontinuation vs Continuation | ADHD | ADHD | 1.089 | 0.980 | 1.210 | 1.15e-01 | 8.03e-01 | 1990 | 1277 | 713 |
| Latino/Mixed-American | No Recorded Depression | Discontinuation vs Continuation | BMI | (Intercept) | 0.474 | 0.296 | 0.758 | 1.85e-03 | NA | 1990 | 1277 | 713 |
| Latino/Mixed-American | No Recorded Depression | Discontinuation vs Continuation | BMI | age_at_first_AD | 1.000 | 0.994 | 1.006 | 9.68e-01 | NA | 1990 | 1277 | 713 |
| Latino/Mixed-American | No Recorded Depression | Discontinuation vs Continuation | BMI | sex_at_birthMale | 1.287 | 1.049 | 1.580 | 1.55e-02 | NA | 1990 | 1277 | 713 |
| Latino/Mixed-American | No Recorded Depression | Discontinuation vs Continuation | BMI | Income_quantitative | 0.940 | 0.892 | 0.990 | 1.93e-02 | NA | 1990 | 1277 | 713 |
| Latino/Mixed-American | No Recorded Depression | Discontinuation vs Continuation | BMI | Education_levelCollege Undergraduate | 1.393 | 1.007 | 1.926 | 4.52e-02 | NA | 1990 | 1277 | 713 |
| Latino/Mixed-American | No Recorded Depression | Discontinuation vs Continuation | BMI | Education_levelLess than a high school degree | 1.010 | 0.669 | 1.526 | 9.62e-01 | NA | 1990 | 1277 | 713 |
| Latino/Mixed-American | No Recorded Depression | Discontinuation vs Continuation | BMI | Education_levelTwelve or GED | 1.623 | 1.105 | 2.384 | 1.36e-02 | NA | 1990 | 1277 | 713 |
| Latino/Mixed-American | No Recorded Depression | Discontinuation vs Continuation | BMI | BMI | 1.065 | 0.966 | 1.174 | 2.03e-01 | 1.00e+00 | 1990 | 1277 | 713 |
| Latino/Mixed-American | No Recorded Depression | Discontinuation vs Continuation | Bipolar Disorder | (Intercept) | 0.479 | 0.299 | 0.766 | 2.13e-03 | NA | 1990 | 1277 | 713 |
| Latino/Mixed-American | No Recorded Depression | Discontinuation vs Continuation | Bipolar Disorder | age_at_first_AD | 1.000 | 0.994 | 1.006 | 9.87e-01 | NA | 1990 | 1277 | 713 |
| Latino/Mixed-American | No Recorded Depression | Discontinuation vs Continuation | Bipolar Disorder | sex_at_birthMale | 1.287 | 1.049 | 1.579 | 1.57e-02 | NA | 1990 | 1277 | 713 |
| Latino/Mixed-American | No Recorded Depression | Discontinuation vs Continuation | Bipolar Disorder | Income_quantitative | 0.937 | 0.890 | 0.987 | 1.43e-02 | NA | 1990 | 1277 | 713 |
| Latino/Mixed-American | No Recorded Depression | Discontinuation vs Continuation | Bipolar Disorder | Education_levelCollege Undergraduate | 1.401 | 1.013 | 1.937 | 4.12e-02 | NA | 1990 | 1277 | 713 |
| Latino/Mixed-American | No Recorded Depression | Discontinuation vs Continuation | Bipolar Disorder | Education_levelLess than a high school degree | 1.012 | 0.670 | 1.528 | 9.55e-01 | NA | 1990 | 1277 | 713 |
| Latino/Mixed-American | No Recorded Depression | Discontinuation vs Continuation | Bipolar Disorder | Education_levelTwelve or GED | 1.638 | 1.116 | 2.406 | 1.18e-02 | NA | 1990 | 1277 | 713 |
| Latino/Mixed-American | No Recorded Depression | Discontinuation vs Continuation | Bipolar Disorder | Bipolar Disorder | 1.049 | 0.945 | 1.165 | 3.67e-01 | 1.00e+00 | 1990 | 1277 | 713 |
| Latino/Mixed-American | No Recorded Depression | Discontinuation vs Continuation | Depression | (Intercept) | 0.470 | 0.294 | 0.752 | 1.66e-03 | NA | 1990 | 1277 | 713 |
| Latino/Mixed-American | No Recorded Depression | Discontinuation vs Continuation | Depression | age_at_first_AD | 1.000 | 0.994 | 1.007 | 9.01e-01 | NA | 1990 | 1277 | 713 |
| Latino/Mixed-American | No Recorded Depression | Discontinuation vs Continuation | Depression | sex_at_birthMale | 1.286 | 1.048 | 1.579 | 1.61e-02 | NA | 1990 | 1277 | 713 |
| Latino/Mixed-American | No Recorded Depression | Discontinuation vs Continuation | Depression | Income_quantitative | 0.940 | 0.892 | 0.990 | 1.96e-02 | NA | 1990 | 1277 | 713 |
| Latino/Mixed-American | No Recorded Depression | Discontinuation vs Continuation | Depression | Education_levelCollege Undergraduate | 1.384 | 1.001 | 1.914 | 4.95e-02 | NA | 1990 | 1277 | 713 |
| Latino/Mixed-American | No Recorded Depression | Discontinuation vs Continuation | Depression | Education_levelLess than a high school degree | 0.995 | 0.658 | 1.503 | 9.79e-01 | NA | 1990 | 1277 | 713 |
| Latino/Mixed-American | No Recorded Depression | Discontinuation vs Continuation | Depression | Education_levelTwelve or GED | 1.620 | 1.103 | 2.380 | 1.39e-02 | NA | 1990 | 1277 | 713 |
| Latino/Mixed-American | No Recorded Depression | Discontinuation vs Continuation | Depression | Depression | 1.158 | 1.043 | 1.287 | 6.13e-03 | 4.29e-02 | 1990 | 1277 | 713 |
| Latino/Mixed-American | No Recorded Depression | Discontinuation vs Continuation | Insomnia | (Intercept) | 0.478 | 0.299 | 0.765 | 2.07e-03 | NA | 1990 | 1277 | 713 |
| Latino/Mixed-American | No Recorded Depression | Discontinuation vs Continuation | Insomnia | age_at_first_AD | 1.000 | 0.994 | 1.006 | 9.89e-01 | NA | 1990 | 1277 | 713 |
| Latino/Mixed-American | No Recorded Depression | Discontinuation vs Continuation | Insomnia | sex_at_birthMale | 1.289 | 1.051 | 1.582 | 1.49e-02 | NA | 1990 | 1277 | 713 |
| Latino/Mixed-American | No Recorded Depression | Discontinuation vs Continuation | Insomnia | Income_quantitative | 0.938 | 0.891 | 0.988 | 1.56e-02 | NA | 1990 | 1277 | 713 |
| Latino/Mixed-American | No Recorded Depression | Discontinuation vs Continuation | Insomnia | Education_levelCollege Undergraduate | 1.394 | 1.008 | 1.928 | 4.45e-02 | NA | 1990 | 1277 | 713 |
| Latino/Mixed-American | No Recorded Depression | Discontinuation vs Continuation | Insomnia | Education_levelLess than a high school degree | 1.002 | 0.663 | 1.514 | 9.94e-01 | NA | 1990 | 1277 | 713 |
| Latino/Mixed-American | No Recorded Depression | Discontinuation vs Continuation | Insomnia | Education_levelTwelve or GED | 1.631 | 1.111 | 2.396 | 1.25e-02 | NA | 1990 | 1277 | 713 |
| Latino/Mixed-American | No Recorded Depression | Discontinuation vs Continuation | Insomnia | Insomnia | 1.058 | 0.950 | 1.178 | 3.06e-01 | 1.00e+00 | 1990 | 1277 | 713 |
| Latino/Mixed-American | No Recorded Depression | Discontinuation vs Continuation | Migraine | (Intercept) | 0.478 | 0.299 | 0.765 | 2.09e-03 | NA | 1990 | 1277 | 713 |
| Latino/Mixed-American | No Recorded Depression | Discontinuation vs Continuation | Migraine | age_at_first_AD | 1.000 | 0.994 | 1.006 | 9.96e-01 | NA | 1990 | 1277 | 713 |
| Latino/Mixed-American | No Recorded Depression | Discontinuation vs Continuation | Migraine | sex_at_birthMale | 1.286 | 1.048 | 1.577 | 1.60e-02 | NA | 1990 | 1277 | 713 |
| Latino/Mixed-American | No Recorded Depression | Discontinuation vs Continuation | Migraine | Income_quantitative | 0.937 | 0.890 | 0.987 | 1.47e-02 | NA | 1990 | 1277 | 713 |
| Latino/Mixed-American | No Recorded Depression | Discontinuation vs Continuation | Migraine | Education_levelCollege Undergraduate | 1.401 | 1.013 | 1.936 | 4.15e-02 | NA | 1990 | 1277 | 713 |
| Latino/Mixed-American | No Recorded Depression | Discontinuation vs Continuation | Migraine | Education_levelLess than a high school degree | 1.013 | 0.671 | 1.530 | 9.50e-01 | NA | 1990 | 1277 | 713 |
| Latino/Mixed-American | No Recorded Depression | Discontinuation vs Continuation | Migraine | Education_levelTwelve or GED | 1.637 | 1.115 | 2.403 | 1.20e-02 | NA | 1990 | 1277 | 713 |
| Latino/Mixed-American | No Recorded Depression | Discontinuation vs Continuation | Migraine | Migraine | 1.001 | 0.910 | 1.102 | 9.80e-01 | 1.00e+00 | 1990 | 1277 | 713 |
| Latino/Mixed-American | No Recorded Depression | Discontinuation vs Continuation | Schizophrenia | (Intercept) | 0.479 | 0.299 | 0.766 | 2.11e-03 | NA | 1990 | 1277 | 713 |
| Latino/Mixed-American | No Recorded Depression | Discontinuation vs Continuation | Schizophrenia | age_at_first_AD | 1.000 | 0.994 | 1.006 | 9.91e-01 | NA | 1990 | 1277 | 713 |
| Latino/Mixed-American | No Recorded Depression | Discontinuation vs Continuation | Schizophrenia | sex_at_birthMale | 1.287 | 1.049 | 1.579 | 1.56e-02 | NA | 1990 | 1277 | 713 |
| Latino/Mixed-American | No Recorded Depression | Discontinuation vs Continuation | Schizophrenia | Income_quantitative | 0.937 | 0.889 | 0.987 | 1.39e-02 | NA | 1990 | 1277 | 713 |
| Latino/Mixed-American | No Recorded Depression | Discontinuation vs Continuation | Schizophrenia | Education_levelCollege Undergraduate | 1.398 | 1.011 | 1.933 | 4.26e-02 | NA | 1990 | 1277 | 713 |
| Latino/Mixed-American | No Recorded Depression | Discontinuation vs Continuation | Schizophrenia | Education_levelLess than a high school degree | 1.010 | 0.669 | 1.526 | 9.62e-01 | NA | 1990 | 1277 | 713 |
| Latino/Mixed-American | No Recorded Depression | Discontinuation vs Continuation | Schizophrenia | Education_levelTwelve or GED | 1.635 | 1.113 | 2.401 | 1.22e-02 | NA | 1990 | 1277 | 713 |
| Latino/Mixed-American | No Recorded Depression | Discontinuation vs Continuation | Schizophrenia | Schizophrenia | 0.979 | 0.891 | 1.076 | 6.59e-01 | 1.00e+00 | 1990 | 1277 | 713 |

| | | | | | | | | | | | | |
|---|---|---|---|---|---|---|---|---|---|---|---|---|
| Latino/Mixed-American | Recorded Anxiety | Discontinuation vs Continuation | ADHD | (Intercept) | 0.397 | 0.241 | 0.654 | 2.86e-04 | NA | 2138 | 1462 | 676 |
| Latino/Mixed-American | Recorded Anxiety | Discontinuation vs Continuation | ADHD | age_at_first_AD | 1.001 | 0.995 | 1.008 | 6.44e-01 | NA | 2138 | 1462 | 676 |
| Latino/Mixed-American | Recorded Anxiety | Discontinuation vs Continuation | ADHD | sex_at_birthMale | 1.312 | 1.072 | 1.605 | 8.46e-03 | NA | 2138 | 1462 | 676 |
| Latino/Mixed-American | Recorded Anxiety | Discontinuation vs Continuation | ADHD | Income_quantitative | 0.949 | 0.898 | 1.003 | 6.42e-02 | NA | 2138 | 1462 | 676 |
| Latino/Mixed-American | Recorded Anxiety | Discontinuation vs Continuation | ADHD | Education_levelCollege Undergraduate | 1.232 | 0.862 | 1.760 | 2.53e-01 | NA | 2138 | 1462 | 676 |
| Latino/Mixed-American | Recorded Anxiety | Discontinuation vs Continuation | ADHD | Education_levelLess than a high school degree | 1.021 | 0.663 | 1.573 | 9.24e-01 | NA | 2138 | 1462 | 676 |
| Latino/Mixed-American | Recorded Anxiety | Discontinuation vs Continuation | ADHD | Education_levelTwelve or GED | 1.184 | 0.793 | 1.769 | 4.08e-01 | NA | 2138 | 1462 | 676 |
| Latino/Mixed-American | Recorded Anxiety | Discontinuation vs Continuation | ADHD | ADHD | 1.016 | 0.916 | 1.127 | 7.64e-01 | 1.00e+00 | 2138 | 1462 | 676 |
| Latino/Mixed-American | Recorded Anxiety | Discontinuation vs Continuation | BMI | (Intercept) | 0.395 | 0.240 | 0.652 | 2.73e-04 | NA | 2138 | 1462 | 676 |
| Latino/Mixed-American | Recorded Anxiety | Discontinuation vs Continuation | BMI | age_at_first_AD | 1.001 | 0.995 | 1.008 | 6.67e-01 | NA | 2138 | 1462 | 676 |
| Latino/Mixed-American | Recorded Anxiety | Discontinuation vs Continuation | BMI | sex_at_birthMale | 1.312 | 1.072 | 1.606 | 8.40e-03 | NA | 2138 | 1462 | 676 |
| Latino/Mixed-American | Recorded Anxiety | Discontinuation vs Continuation | BMI | Income_quantitative | 0.952 | 0.901 | 1.007 | 8.49e-02 | NA | 2138 | 1462 | 676 |
| Latino/Mixed-American | Recorded Anxiety | Discontinuation vs Continuation | BMI | Education_levelCollege Undergraduate | 1.230 | 0.861 | 1.758 | 2.56e-01 | NA | 2138 | 1462 | 676 |
| Latino/Mixed-American | Recorded Anxiety | Discontinuation vs Continuation | BMI | Education_levelLess than a high school degree | 1.014 | 0.658 | 1.561 | 9.51e-01 | NA | 2138 | 1462 | 676 |
| Latino/Mixed-American | Recorded Anxiety | Discontinuation vs Continuation | BMI | Education_levelTwelve or GED | 1.181 | 0.791 | 1.764 | 4.16e-01 | NA | 2138 | 1462 | 676 |
| Latino/Mixed-American | Recorded Anxiety | Discontinuation vs Continuation | BMI | BMI | 1.093 | 0.994 | 1.201 | 6.70e-02 | 4.69e-01 | 2138 | 1462 | 676 |
| Latino/Mixed-American | Recorded Anxiety | Discontinuation vs Continuation | Bipolar Disorder | (Intercept) | 0.398 | 0.242 | 0.655 | 2.94e-04 | NA | 2138 | 1462 | 676 |
| Latino/Mixed-American | Recorded Anxiety | Discontinuation vs Continuation | Bipolar Disorder | age_at_first_AD | 1.001 | 0.995 | 1.008 | 6.56e-01 | NA | 2138 | 1462 | 676 |
| Latino/Mixed-American | Recorded Anxiety | Discontinuation vs Continuation | Bipolar Disorder | sex_at_birthMale | 1.311 | 1.071 | 1.605 | 8.61e-03 | NA | 2138 | 1462 | 676 |
| Latino/Mixed-American | Recorded Anxiety | Discontinuation vs Continuation | Bipolar Disorder | Income_quantitative | 0.948 | 0.897 | 1.002 | 6.07e-02 | NA | 2138 | 1462 | 676 |
| Latino/Mixed-American | Recorded Anxiety | Discontinuation vs Continuation | Bipolar Disorder | Education_levelCollege Undergraduate | 1.234 | 0.863 | 1.762 | 2.49e-01 | NA | 2138 | 1462 | 676 |
| Latino/Mixed-American | Recorded Anxiety | Discontinuation vs Continuation | Bipolar Disorder | Education_levelLess than a high school degree | 1.023 | 0.665 | 1.575 | 9.16e-01 | NA | 2138 | 1462 | 676 |
| Latino/Mixed-American | Recorded Anxiety | Discontinuation vs Continuation | Bipolar Disorder | Education_levelTwelve or GED | 1.187 | 0.795 | 1.773 | 4.02e-01 | NA | 2138 | 1462 | 676 |
| Latino/Mixed-American | Recorded Anxiety | Discontinuation vs Continuation | Bipolar Disorder | Bipolar Disorder | 0.996 | 0.899 | 1.104 | 9.39e-01 | 1.00e+00 | 2138 | 1462 | 676 |
| Latino/Mixed-American | Recorded Anxiety | Discontinuation vs Continuation | Depression | (Intercept) | 0.395 | 0.239 | 0.650 | 2.63e-04 | NA | 2138 | 1462 | 676 |
| Latino/Mixed-American | Recorded Anxiety | Discontinuation vs Continuation | Depression | age_at_first_AD | 1.002 | 0.995 | 1.008 | 6.08e-01 | NA | 2138 | 1462 | 676 |
| Latino/Mixed-American | Recorded Anxiety | Discontinuation vs Continuation | Depression | sex_at_birthMale | 1.315 | 1.074 | 1.609 | 8.00e-03 | NA | 2138 | 1462 | 676 |
| Latino/Mixed-American | Recorded Anxiety | Discontinuation vs Continuation | Depression | Income_quantitative | 0.951 | 0.900 | 1.005 | 7.64e-02 | NA | 2138 | 1462 | 676 |
| Latino/Mixed-American | Recorded Anxiety | Discontinuation vs Continuation | Depression | Education_levelCollege Undergraduate | 1.222 | 0.855 | 1.747 | 2.70e-01 | NA | 2138 | 1462 | 676 |
| Latino/Mixed-American | Recorded Anxiety | Discontinuation vs Continuation | Depression | Education_levelLess than a high school degree | 1.014 | 0.659 | 1.561 | 9.49e-01 | NA | 2138 | 1462 | 676 |
| Latino/Mixed-American | Recorded Anxiety | Discontinuation vs Continuation | Depression | Education_levelTwelve or GED | 1.175 | 0.787 | 1.755 | 4.30e-01 | NA | 2138 | 1462 | 676 |
| Latino/Mixed-American | Recorded Anxiety | Discontinuation vs Continuation | Depression | Depression | 1.129 | 1.017 | 1.254 | 2.28e-02 | 1.60e-01 | 2138 | 1462 | 676 |
| Latino/Mixed-American | Recorded Anxiety | Discontinuation vs Continuation | Insomnia | (Intercept) | 0.398 | 0.242 | 0.656 | 2.97e-04 | NA | 2138 | 1462 | 676 |
| Latino/Mixed-American | Recorded Anxiety | Discontinuation vs Continuation | Insomnia | age_at_first_AD | 1.001 | 0.995 | 1.008 | 6.63e-01 | NA | 2138 | 1462 | 676 |
| Latino/Mixed-American | Recorded Anxiety | Discontinuation vs Continuation | Insomnia | sex_at_birthMale | 1.311 | 1.071 | 1.604 | 8.62e-03 | NA | 2138 | 1462 | 676 |
| Latino/Mixed-American | Recorded Anxiety | Discontinuation vs Continuation | Insomnia | Income_quantitative | 0.948 | 0.897 | 1.002 | 5.96e-02 | NA | 2138 | 1462 | 676 |
| Latino/Mixed-American | Recorded Anxiety | Discontinuation vs Continuation | Insomnia | Education_levelCollege Undergraduate | 1.235 | 0.865 | 1.765 | 2.46e-01 | NA | 2138 | 1462 | 676 |
| Latino/Mixed-American | Recorded Anxiety | Discontinuation vs Continuation | Insomnia | Education_levelLess than a high school degree | 1.025 | 0.666 | 1.579 | 9.09e-01 | NA | 2138 | 1462 | 676 |
| Latino/Mixed-American | Recorded Anxiety | Discontinuation vs Continuation | Insomnia | Education_levelTwelve or GED | 1.188 | 0.795 | 1.773 | 4.00e-01 | NA | 2138 | 1462 | 676 |
| Latino/Mixed-American | Recorded Anxiety | Discontinuation vs Continuation | Insomnia | Insomnia | 0.982 | 0.884 | 1.092 | 7.41e-01 | 1.00e+00 | 2138 | 1462 | 676 |
| Latino/Mixed-American | Recorded Anxiety | Discontinuation vs Continuation | Migraine | (Intercept) | 0.396 | 0.241 | 0.653 | 2.79e-04 | NA | 2138 | 1462 | 676 |
| Latino/Mixed-American | Recorded Anxiety | Discontinuation vs Continuation | Migraine | age_at_first_AD | 1.001 | 0.995 | 1.008 | 6.51e-01 | NA | 2138 | 1462 | 676 |
| Latino/Mixed-American | Recorded Anxiety | Discontinuation vs Continuation | Migraine | sex_at_birthMale | 1.311 | 1.071 | 1.604 | 8.67e-03 | NA | 2138 | 1462 | 676 |
| Latino/Mixed-American | Recorded Anxiety | Discontinuation vs Continuation | Migraine | Income_quantitative | 0.948 | 0.897 | 1.002 | 6.05e-02 | NA | 2138 | 1462 | 676 |
| Latino/Mixed-American | Recorded Anxiety | Discontinuation vs Continuation | Migraine | Education_levelCollege Undergraduate | 1.236 | 0.865 | 1.766 | 2.44e-01 | NA | 2138 | 1462 | 676 |
| Latino/Mixed-American | Recorded Anxiety | Discontinuation vs Continuation | Migraine | Education_levelLess than a high school degree | 1.027 | 0.667 | 1.581 | 9.04e-01 | NA | 2138 | 1462 | 676 |
| Latino/Mixed-American | Recorded Anxiety | Discontinuation vs Continuation | Migraine | Education_levelTwelve or GED | 1.191 | 0.798 | 1.779 | 3.92e-01 | NA | 2138 | 1462 | 676 |
| Latino/Mixed-American | Recorded Anxiety | Discontinuation vs Continuation | Migraine | Migraine | 0.960 | 0.871 | 1.059 | 4.16e-01 | 1.00e+00 | 2138 | 1462 | 676 |
| Latino/Mixed-American | Recorded Anxiety | Discontinuation vs Continuation | Schizophrenia | (Intercept) | 0.396 | 0.240 | 0.652 | 2.76e-04 | NA | 2138 | 1462 | 676 |
| Latino/Mixed-American | Recorded Anxiety | Discontinuation vs Continuation | Schizophrenia | age_at_first_AD | 1.001 | 0.995 | 1.008 | 7.00e-01 | NA | 2138 | 1462 | 676 |
| Latino/Mixed-American | Recorded Anxiety | Discontinuation vs Continuation | Schizophrenia | sex_at_birthMale | 1.313 | 1.072 | 1.607 | 8.34e-03 | NA | 2138 | 1462 | 676 |
| Latino/Mixed-American | Recorded Anxiety | Discontinuation vs Continuation | Schizophrenia | Income_quantitative | 0.950 | 0.899 | 1.004 | 7.00e-02 | NA | 2138 | 1462 | 676 |
| Latino/Mixed-American | Recorded Anxiety | Discontinuation vs Continuation | Schizophrenia | Education_levelCollege Undergraduate | 1.245 | 0.871 | 1.779 | 2.30e-01 | NA | 2138 | 1462 | 676 |
| Latino/Mixed-American | Recorded Anxiety | Discontinuation vs Continuation | Schizophrenia | Education_levelLess than a high school degree | 1.035 | 0.672 | 1.594 | 8.76e-01 | NA | 2138 | 1462 | 676 |
| Latino/Mixed-American | Recorded Anxiety | Discontinuation vs Continuation | Schizophrenia | Education_levelTwelve or GED | 1.195 | 0.800 | 1.785 | 3.84e-01 | NA | 2138 | 1462 | 676 |
| Latino/Mixed-American | Recorded Anxiety | Discontinuation vs Continuation | Schizophrenia | Schizophrenia | 1.071 | 0.976 | 1.176 | 1.47e-01 | 1.00e+00 | 2138 | 1462 | 676 |

**Supplementary Table 9. Multivariable logistic regression results for PGS associations with antidepressant discontinuation (sensitivity definition) across clinical subgroups in the Pharmlines cohort (European ancestry participants).**
Each model estimates the association between 7 PGS traits and the binary outcome discontinuation versus continuation, fitted separately within each combination of clinical group (Full Cohort; and subgroups defined by recorded depression status), and PGS exposure. Individuals who met augmentation or combination criteria at any point during follow-up were excluded from this analysis, so that the discontinuation-versus-continuation contrast is estimated among those who did not escalate treatment via augmentation or combination, and only had short antidepresant monotherapy episodes throughout their full prescribing history. Within each model, one PGS exposure is modelled alongside a common covariate set (age at first antidepressant, genetic sex, income, and educational attainment). All PGS were residualised on the top 20 within-European-ancestry principal components and the residuals standardised to mean 0, SD 1; each odds ratio therefore reflects the change in odds per one-standard-deviation increase in the PC-adjusted PGS. Because PCs were regressed out at this stage, they are not included in the model covariate set.
Columns: Population = inferred genetic ancestry (all European); ClinicalGroup = cohort or clinical subgroup; Outcome = the binary contrast modelled; PGS = the PGS trait for that model; variable = the individual model term; odds_ratio = exponentiated coefficient; lower_95/upper_95 = 95% confidence interval bounds for the odds ratio; p_value = nominal (uncorrected) p-value; Bonf_P = Bonferroni-corrected p-value (capped at 1.00); N_Total = model sample size; N_Reference = number in the reference (continuation) group; N_Case = number in the comparison (discontinuation or switching) group. Odds ratios >1 indicate higher odds of the comparison outcome relative to continuation. Bonferroni correction was applied within each specification (ancestry × clinical group × outcome contrast) for the max 7 PGS traits tested. P-values reported as "<2.2e-16" reflect R's floating-point reporting limit.
Reference (omitted) categories: Sex at birth: Female.

| Population | ClinicalGroup | Outcome | PGS | variable | odds_ratio | lower_95 | upper_95 | p_value | Bonf_p | N_Total | N_Reference | N_Case |
|---|---|---|---|---|---|---|---|---|---|---|---|---|
| European | Full Cohort | Discontinuation vs Continuation | ADHD | (Intercept) | 0.204 | 0.145 | 0.287 | <2.2e-16 | <2.2e-16 | 5824 | 4445 | 1379 |
| European | Full Cohort | Discontinuation vs Continuation | ADHD | age_at_first_AD | 1.016 | 1.011 | 1.021 | 1.50e-10 | 1.05e-09 | 5824 | 4445 | 1379 |
| European | Full Cohort | Discontinuation vs Continuation | ADHD | sex_at_birth | 1.279 | 1.121 | 1.458 | 2.41e-04 | 1.69e-03 | 5824 | 4445 | 1379 |
| European | Full Cohort | Discontinuation vs Continuation | ADHD | Income_level | 1.000 | 1.000 | 1.000 | 1.96e-01 | 1.00e+00 | 5824 | 4445 | 1379 |
| European | Full Cohort | Discontinuation vs Continuation | ADHD | Education_level | 0.964 | 0.949 | 0.980 | 8.02e-06 | 5.61e-05 | 5824 | 4445 | 1379 |
| European | Full Cohort | Discontinuation vs Continuation | ADHD | ADHD | 1.080 | 1.015 | 1.149 | 1.46e-02 | 1.02e-01 | 5824 | 4445 | 1379 |
| European | Full Cohort | Discontinuation vs Continuation | BMI | (Intercept) | 0.206 | 0.146 | 0.291 | <2.2e-16 | <2.2e-16 | 5824 | 4445 | 1379 |
| European | Full Cohort | Discontinuation vs Continuation | BMI | age_at_first_AD | 1.016 | 1.011 | 1.021 | 1.96e-10 | 1.38e-09 | 5824 | 4445 | 1379 |
| European | Full Cohort | Discontinuation vs Continuation | BMI | sex_at_birth | 1.282 | 1.125 | 1.462 | 2.03e-04 | 1.42e-03 | 5824 | 4445 | 1379 |
| European | Full Cohort | Discontinuation vs Continuation | BMI | Income_level | 1.000 | 1.000 | 1.000 | 1.97e-01 | 1.00e+00 | 5824 | 4445 | 1379 |
| European | Full Cohort | Discontinuation vs Continuation | BMI | Education_level | 0.964 | 0.948 | 0.979 | 6.02e-06 | 4.21e-05 | 5824 | 4445 | 1379 |
| European | Full Cohort | Discontinuation vs Continuation | BMI | BMI | 1.063 | 1.001 | 1.129 | 4.61e-02 | 3.23e-01 | 5824 | 4445 | 1379 |
| European | Full Cohort | Discontinuation vs Continuation | Bipolar Disorder | (Intercept) | 0.212 | 0.151 | 0.298 | <2.2e-16 | <2.2e-16 | 5824 | 4445 | 1379 |
| European | Full Cohort | Discontinuation vs Continuation | Bipolar Disorder | age_at_first_AD | 1.016 | 1.011 | 1.020 | 2.69e-10 | 1.89e-09 | 5824 | 4445 | 1379 |
| European | Full Cohort | Discontinuation vs Continuation | Bipolar Disorder | sex_at_birth | 1.279 | 1.122 | 1.458 | 2.35e-04 | 1.64e-03 | 5824 | 4445 | 1379 |
| European | Full Cohort | Discontinuation vs Continuation | Bipolar Disorder | Income_level | 1.000 | 1.000 | 1.000 | 2.07e-01 | 1.00e+00 | 5824 | 4445 | 1379 |
| European | Full Cohort | Discontinuation vs Continuation | Bipolar Disorder | Education_level | 0.962 | 0.947 | 0.978 | 2.84e-06 | 1.99e-05 | 5824 | 4445 | 1379 |
| European | Full Cohort | Discontinuation vs Continuation | Bipolar Disorder | Bipolar Disorder | 1.006 | 0.946 | 1.069 | 8.51e-01 | 1.00e+00 | 5824 | 4445 | 1379 |
| European | Full Cohort | Discontinuation vs Continuation | Depression | (Intercept) | 0.216 | 0.153 | 0.304 | <2.2e-16 | <2.2e-16 | 5824 | 4445 | 1379 |
| European | Full Cohort | Discontinuation vs Continuation | Depression | age_at_first_AD | 1.015 | 1.010 | 1.020 | 5.35e-10 | 3.74e-09 | 5824 | 4445 | 1379 |
| European | Full Cohort | Discontinuation vs Continuation | Depression | sex_at_birth | 1.280 | 1.122 | 1.459 | 2.28e-04 | 1.60e-03 | 5824 | 4445 | 1379 |
| European | Full Cohort | Discontinuation vs Continuation | Depression | Income_level | 1.000 | 1.000 | 1.000 | 2.10e-01 | 1.00e+00 | 5824 | 4445 | 1379 |
| European | Full Cohort | Discontinuation vs Continuation | Depression | Education_level | 0.962 | 0.947 | 0.977 | 2.04e-06 | 1.43e-05 | 5824 | 4445 | 1379 |
| European | Full Cohort | Discontinuation vs Continuation | Depression | Depression | 0.954 | 0.897 | 1.013 | 1.25e-01 | 8.75e-01 | 5824 | 4445 | 1379 |
| European | Full Cohort | Discontinuation vs Continuation | Insomnia | (Intercept) | 0.212 | 0.151 | 0.299 | <2.2e-16 | <2.2e-16 | 5824 | 4445 | 1379 |
| European | Full Cohort | Discontinuation vs Continuation | Insomnia | age_at_first_AD | 1.016 | 1.011 | 1.020 | 2.90e-10 | 2.03e-09 | 5824 | 4445 | 1379 |
| European | Full Cohort | Discontinuation vs Continuation | Insomnia | sex_at_birth | 1.279 | 1.122 | 1.458 | 2.37e-04 | 1.66e-03 | 5824 | 4445 | 1379 |
| European | Full Cohort | Discontinuation vs Continuation | Insomnia | Income_level | 1.000 | 1.000 | 1.000 | 2.07e-01 | 1.00e+00 | 5824 | 4445 | 1379 |
| European | Full Cohort | Discontinuation vs Continuation | Insomnia | Education_level | 0.962 | 0.947 | 0.978 | 2.83e-06 | 1.98e-05 | 5824 | 4445 | 1379 |
| European | Full Cohort | Discontinuation vs Continuation | Insomnia | Insomnia | 0.998 | 0.938 | 1.061 | 9.48e-01 | 1.00e+00 | 5824 | 4445 | 1379 |
| European | Full Cohort | Discontinuation vs Continuation | Migraine | (Intercept) | 0.213 | 0.151 | 0.299 | <2.2e-16 | <2.2e-16 | 5824 | 4445 | 1379 |
| European | Full Cohort | Discontinuation vs Continuation | Migraine | age_at_first_AD | 1.016 | 1.011 | 1.020 | 2.71e-10 | 1.90e-09 | 5824 | 4445 | 1379 |
| European | Full Cohort | Discontinuation vs Continuation | Migraine | sex_at_birth | 1.279 | 1.122 | 1.459 | 2.31e-04 | 1.62e-03 | 5824 | 4445 | 1379 |
| European | Full Cohort | Discontinuation vs Continuation | Migraine | Income_level | 1.000 | 1.000 | 1.000 | 2.14e-01 | 1.00e+00 | 5824 | 4445 | 1379 |

| | | | | | | | | | | | | |
|---|---|---|---|---|---|---|---|---|---|---|---|---|
| European | Full Cohort | Discontinuation vs Continuation | Migraine | Education_level | 0.962 | 0.947 | 0.978 | 2.66e-06 | 1.86e-05 | 5824 | 4445 | 1379 |
| European | Full Cohort | Discontinuation vs Continuation | Migraine | Migraine | 0.974 | 0.917 | 1.034 | 3.84e-01 | 1.00e+00 | 5824 | 4445 | 1379 |
| European | Full Cohort | Discontinuation vs Continuation | Schizophrenia | (Intercept) | 0.212 | 0.151 | 0.298 | <2.2e-16 | <2.2e-16 | 5824 | 4445 | 1379 |
| European | Full Cohort | Discontinuation vs Continuation | Schizophrenia | age_at_first_AD | 1.016 | 1.011 | 1.020 | 3.02e-10 | 2.11e-09 | 5824 | 4445 | 1379 |
| European | Full Cohort | Discontinuation vs Continuation | Schizophrenia | sex_at_birth | 1.279 | 1.122 | 1.458 | 2.37e-04 | 1.66e-03 | 5824 | 4445 | 1379 |
| European | Full Cohort | Discontinuation vs Continuation | Schizophrenia | Income_level | 1.000 | 1.000 | 1.000 | 2.06e-01 | 1.00e+00 | 5824 | 4445 | 1379 |
| European | Full Cohort | Discontinuation vs Continuation | Schizophrenia | Education_level | 0.962 | 0.947 | 0.978 | 2.84e-06 | 1.99e-05 | 5824 | 4445 | 1379 |
| European | Full Cohort | Discontinuation vs Continuation | Schizophrenia | Schizophrenia | 0.994 | 0.935 | 1.056 | 8.40e-01 | 1.00e+00 | 5824 | 4445 | 1379 |
| European | Recorded Depression | Discontinuation vs Continuation | ADHD | (Intercept) | 0.172 | 0.089 | 0.333 | 1.66e-07 | 1.16e-06 | 2040 | 1692 | 348 |
| European | Recorded Depression | Discontinuation vs Continuation | ADHD | age_at_first_AD | 1.014 | 1.003 | 1.024 | 1.14e-02 | 7.99e-02 | 2040 | 1692 | 348 |
| European | Recorded Depression | Discontinuation vs Continuation | ADHD | sex_at_birth | 1.255 | 0.972 | 1.621 | 8.20e-02 | 5.74e-01 | 2040 | 1692 | 348 |
| European | Recorded Depression | Discontinuation vs Continuation | ADHD | Income_level | 1.000 | 1.000 | 1.000 | 9.98e-01 | 1.00e+00 | 2040 | 1692 | 348 |
| European | Recorded Depression | Discontinuation vs Continuation | ADHD | Education_level | 0.967 | 0.939 | 0.996 | 2.82e-02 | 1.98e-01 | 2040 | 1692 | 348 |
| European | Recorded Depression | Discontinuation vs Continuation | ADHD | ADHD | 1.101 | 0.977 | 1.241 | 1.13e-01 | 7.91e-01 | 2040 | 1692 | 348 |
| European | Recorded Depression | Discontinuation vs Continuation | BMI | (Intercept) | 0.174 | 0.090 | 0.336 | 1.99e-07 | 1.39e-06 | 2040 | 1692 | 348 |
| European | Recorded Depression | Discontinuation vs Continuation | BMI | age_at_first_AD | 1.014 | 1.003 | 1.024 | 1.16e-02 | 8.12e-02 | 2040 | 1692 | 348 |
| European | Recorded Depression | Discontinuation vs Continuation | BMI | sex_at_birth | 1.274 | 0.986 | 1.646 | 6.39e-02 | 4.47e-01 | 2040 | 1692 | 348 |
| European | Recorded Depression | Discontinuation vs Continuation | BMI | Income_level | 1.000 | 1.000 | 1.000 | 9.86e-01 | 1.00e+00 | 2040 | 1692 | 348 |
| European | Recorded Depression | Discontinuation vs Continuation | BMI | Education_level | 0.966 | 0.938 | 0.995 | 2.32e-02 | 1.62e-01 | 2040 | 1692 | 348 |
| European | Recorded Depression | Discontinuation vs Continuation | BMI | BMI | 1.091 | 0.975 | 1.221 | 1.29e-01 | 9.04e-01 | 2040 | 1692 | 348 |
| European | Recorded Depression | Discontinuation vs Continuation | Bipolar Disorder | (Intercept) | 0.180 | 0.093 | 0.347 | 2.89e-07 | 2.02e-06 | 2040 | 1692 | 348 |
| European | Recorded Depression | Discontinuation vs Continuation | Bipolar Disorder | age_at_first_AD | 1.013 | 1.003 | 1.024 | 1.24e-02 | 8.68e-02 | 2040 | 1692 | 348 |
| European | Recorded Depression | Discontinuation vs Continuation | Bipolar Disorder | sex_at_birth | 1.264 | 0.979 | 1.633 | 7.25e-02 | 5.08e-01 | 2040 | 1692 | 348 |
| European | Recorded Depression | Discontinuation vs Continuation | Bipolar Disorder | Income_level | 1.000 | 1.000 | 1.000 | 9.95e-01 | 1.00e+00 | 2040 | 1692 | 348 |
| European | Recorded Depression | Discontinuation vs Continuation | Bipolar Disorder | Education_level | 0.965 | 0.936 | 0.994 | 1.78e-02 | 1.24e-01 | 2040 | 1692 | 348 |
| European | Recorded Depression | Discontinuation vs Continuation | Bipolar Disorder | Bipolar Disorder | 1.022 | 0.909 | 1.149 | 7.19e-01 | 1.00e+00 | 2040 | 1692 | 348 |
| European | Recorded Depression | Discontinuation vs Continuation | Depression | (Intercept) | 0.188 | 0.098 | 0.362 | 5.81e-07 | 4.07e-06 | 2040 | 1692 | 348 |
| European | Recorded Depression | Discontinuation vs Continuation | Depression | age_at_first_AD | 1.013 | 1.002 | 1.023 | 1.67e-02 | 1.17e-01 | 2040 | 1692 | 348 |
| European | Recorded Depression | Discontinuation vs Continuation | Depression | sex_at_birth | 1.281 | 0.991 | 1.656 | 5.84e-02 | 4.09e-01 | 2040 | 1692 | 348 |
| European | Recorded Depression | Discontinuation vs Continuation | Depression | Income_level | 1.000 | 1.000 | 1.000 | 9.62e-01 | 1.00e+00 | 2040 | 1692 | 348 |
| European | Recorded Depression | Discontinuation vs Continuation | Depression | Education_level | 0.962 | 0.934 | 0.991 | 1.14e-02 | 7.97e-02 | 2040 | 1692 | 348 |
| European | Recorded Depression | Discontinuation vs Continuation | Depression | Depression | 0.887 | 0.788 | 0.998 | 4.61e-02 | 3.23e-01 | 2040 | 1692 | 348 |
| European | Recorded Depression | Discontinuation vs Continuation | Insomnia | (Intercept) | 0.179 | 0.093 | 0.346 | 2.85e-07 | 1.99e-06 | 2040 | 1692 | 348 |
| European | Recorded Depression | Discontinuation vs Continuation | Insomnia | age_at_first_AD | 1.013 | 1.003 | 1.024 | 1.26e-02 | 8.84e-02 | 2040 | 1692 | 348 |
| European | Recorded Depression | Discontinuation vs Continuation | Insomnia | sex_at_birth | 1.266 | 0.980 | 1.635 | 7.10e-02 | 4.97e-01 | 2040 | 1692 | 348 |
| European | Recorded Depression | Discontinuation vs Continuation | Insomnia | Income_level | 1.000 | 1.000 | 1.000 | 9.96e-01 | 1.00e+00 | 2040 | 1692 | 348 |
| European | Recorded Depression | Discontinuation vs Continuation | Insomnia | Education_level | 0.965 | 0.937 | 0.994 | 1.89e-02 | 1.32e-01 | 2040 | 1692 | 348 |
| European | Recorded Depression | Discontinuation vs Continuation | Insomnia | Insomnia | 1.023 | 0.910 | 1.150 | 7.07e-01 | 1.00e+00 | 2040 | 1692 | 348 |
| European | Recorded Depression | Discontinuation vs Continuation | Migraine | (Intercept) | 0.178 | 0.092 | 0.343 | 2.39e-07 | 1.67e-06 | 2040 | 1692 | 348 |
| European | Recorded Depression | Discontinuation vs Continuation | Migraine | age_at_first_AD | 1.013 | 1.003 | 1.024 | 1.20e-02 | 8.42e-02 | 2040 | 1692 | 348 |
| European | Recorded Depression | Discontinuation vs Continuation | Migraine | sex_at_birth | 1.264 | 0.979 | 1.633 | 7.26e-02 | 5.08e-01 | 2040 | 1692 | 348 |
| European | Recorded Depression | Discontinuation vs Continuation | Migraine | Income_level | 1.000 | 1.000 | 1.000 | 9.86e-01 | 1.00e+00 | 2040 | 1692 | 348 |
| European | Recorded Depression | Discontinuation vs Continuation | Migraine | Education_level | 0.965 | 0.937 | 0.995 | 2.02e-02 | 1.42e-01 | 2040 | 1692 | 348 |
| European | Recorded Depression | Discontinuation vs Continuation | Migraine | Migraine | 1.093 | 0.973 | 1.227 | 1.34e-01 | 9.39e-01 | 2040 | 1692 | 348 |
| European | Recorded Depression | Discontinuation vs Continuation | Schizophrenia | (Intercept) | 0.181 | 0.094 | 0.348 | 3.06e-07 | 2.14e-06 | 2040 | 1692 | 348 |
| European | Recorded Depression | Discontinuation vs Continuation | Schizophrenia | age_at_first_AD | 1.013 | 1.003 | 1.024 | 1.30e-02 | 9.11e-02 | 2040 | 1692 | 348 |
| European | Recorded Depression | Discontinuation vs Continuation | Schizophrenia | sex_at_birth | 1.264 | 0.979 | 1.633 | 7.23e-02 | 5.06e-01 | 2040 | 1692 | 348 |
| European | Recorded Depression | Discontinuation vs Continuation | Schizophrenia | Income_level | 1.000 | 1.000 | 1.000 | 9.97e-01 | 1.00e+00 | 2040 | 1692 | 348 |

| | | | | | | | | | | | | |
|---|---|---|---|---|---|---|---|---|---|---|---|---|
| European | Recorded Depression | Discontinuation vs Continuation | Schizophrenia | Education_level | 0.965 | 0.936 | 0.994 | 1.80e-02 | 1.26e-01 | 2040 | 1692 | 348 |
| European | Recorded Depression | Discontinuation vs Continuation | Schizophrenia | Schizophrenia | 0.995 | 0.887 | 1.116 | 9.27e-01 | 1.00e+00 | 2040 | 1692 | 348 |
| European | No Recorded Depression | Discontinuation vs Continuation | ADHD | (Intercept) | 0.235 | 0.156 | 0.353 | 3.72e-12 | 2.61e-11 | 3732 | 2713 | 1019 |
| European | No Recorded Depression | Discontinuation vs Continuation | ADHD | age_at_first_AD | 1.014 | 1.009 | 1.020 | 4.20e-07 | 2.94e-06 | 3732 | 2713 | 1019 |
| European | No Recorded Depression | Discontinuation vs Continuation | ADHD | sex_at_birth | 1.261 | 1.080 | 1.472 | 3.40e-03 | 2.38e-02 | 3732 | 2713 | 1019 |
| European | No Recorded Depression | Discontinuation vs Continuation | ADHD | Income_level | 1.000 | 1.000 | 1.000 | 1.91e-01 | 1.00e+00 | 3732 | 2713 | 1019 |
| European | No Recorded Depression | Discontinuation vs Continuation | ADHD | Education_level | 0.969 | 0.950 | 0.988 | 1.60e-03 | 1.12e-02 | 3732 | 2713 | 1019 |
| European | No Recorded Depression | Discontinuation vs Continuation | ADHD | ADHD | 1.076 | 1.001 | 1.158 | 4.83e-02 | 3.38e-01 | 3732 | 2713 | 1019 |
| European | No Recorded Depression | Discontinuation vs Continuation | BMI | (Intercept) | 0.238 | 0.158 | 0.358 | 5.50e-12 | 3.85e-11 | 3732 | 2713 | 1019 |
| European | No Recorded Depression | Discontinuation vs Continuation | BMI | age_at_first_AD | 1.014 | 1.009 | 1.020 | 5.10e-07 | 3.57e-06 | 3732 | 2713 | 1019 |
| European | No Recorded Depression | Discontinuation vs Continuation | BMI | sex_at_birth | 1.260 | 1.079 | 1.471 | 3.51e-03 | 2.46e-02 | 3732 | 2713 | 1019 |
| European | No Recorded Depression | Discontinuation vs Continuation | BMI | Income_level | 1.000 | 1.000 | 1.000 | 1.97e-01 | 1.00e+00 | 3732 | 2713 | 1019 |
| European | No Recorded Depression | Discontinuation vs Continuation | BMI | Education_level | 0.969 | 0.950 | 0.988 | 1.42e-03 | 9.91e-03 | 3732 | 2713 | 1019 |
| European | No Recorded Depression | Discontinuation vs Continuation | BMI | BMI | 1.052 | 0.979 | 1.132 | 1.69e-01 | 1.00e+00 | 3732 | 2713 | 1019 |
| European | No Recorded Depression | Discontinuation vs Continuation | Bipolar Disorder | (Intercept) | 0.243 | 0.162 | 0.365 | 9.75e-12 | 6.82e-11 | 3732 | 2713 | 1019 |
| European | No Recorded Depression | Discontinuation vs Continuation | Bipolar Disorder | age_at_first_AD | 1.014 | 1.009 | 1.020 | 6.02e-07 | 4.21e-06 | 3732 | 2713 | 1019 |
| European | No Recorded Depression | Discontinuation vs Continuation | Bipolar Disorder | sex_at_birth | 1.259 | 1.078 | 1.470 | 3.55e-03 | 2.49e-02 | 3732 | 2713 | 1019 |
| European | No Recorded Depression | Discontinuation vs Continuation | Bipolar Disorder | Income_level | 1.000 | 1.000 | 1.000 | 2.05e-01 | 1.00e+00 | 3732 | 2713 | 1019 |
| European | No Recorded Depression | Discontinuation vs Continuation | Bipolar Disorder | Education_level | 0.968 | 0.949 | 0.987 | 9.35e-04 | 6.54e-03 | 3732 | 2713 | 1019 |
| European | No Recorded Depression | Discontinuation vs Continuation | Bipolar Disorder | Bipolar Disorder | 1.013 | 0.943 | 1.089 | 7.24e-01 | 1.00e+00 | 3732 | 2713 | 1019 |
| European | No Recorded Depression | Discontinuation vs Continuation | Depression | (Intercept) | 0.244 | 0.162 | 0.366 | 1.16e-11 | 8.15e-11 | 3732 | 2713 | 1019 |
| European | No Recorded Depression | Discontinuation vs Continuation | Depression | age_at_first_AD | 1.014 | 1.008 | 1.020 | 6.74e-07 | 4.72e-06 | 3732 | 2713 | 1019 |
| European | No Recorded Depression | Discontinuation vs Continuation | Depression | sex_at_birth | 1.258 | 1.077 | 1.469 | 3.69e-03 | 2.58e-02 | 3732 | 2713 | 1019 |
| European | No Recorded Depression | Discontinuation vs Continuation | Depression | Income_level | 1.000 | 1.000 | 1.000 | 2.03e-01 | 1.00e+00 | 3732 | 2713 | 1019 |
| European | No Recorded Depression | Discontinuation vs Continuation | Depression | Education_level | 0.968 | 0.949 | 0.987 | 9.10e-04 | 6.37e-03 | 3732 | 2713 | 1019 |
| European | No Recorded Depression | Discontinuation vs Continuation | Depression | Depression | 0.996 | 0.926 | 1.070 | 9.08e-01 | 1.00e+00 | 3732 | 2713 | 1019 |
| European | No Recorded Depression | Discontinuation vs Continuation | Insomnia | (Intercept) | 0.245 | 0.163 | 0.368 | 1.36e-11 | 9.50e-11 | 3732 | 2713 | 1019 |
| European | No Recorded Depression | Discontinuation vs Continuation | Insomnia | age_at_first_AD | 1.014 | 1.008 | 1.020 | 7.34e-07 | 5.14e-06 | 3732 | 2713 | 1019 |
| European | No Recorded Depression | Discontinuation vs Continuation | Insomnia | sex_at_birth | 1.259 | 1.078 | 1.470 | 3.57e-03 | 2.50e-02 | 3732 | 2713 | 1019 |
| European | No Recorded Depression | Discontinuation vs Continuation | Insomnia | Income_level | 1.000 | 1.000 | 1.000 | 2.06e-01 | 1.00e+00 | 3732 | 2713 | 1019 |
| European | No Recorded Depression | Discontinuation vs Continuation | Insomnia | Education_level | 0.968 | 0.949 | 0.987 | 8.70e-04 | 6.09e-03 | 3732 | 2713 | 1019 |
| European | No Recorded Depression | Discontinuation vs Continuation | Insomnia | Insomnia | 0.983 | 0.913 | 1.058 | 6.38e-01 | 1.00e+00 | 3732 | 2713 | 1019 |
| European | No Recorded Depression | Discontinuation vs Continuation | Migraine | (Intercept) | 0.245 | 0.163 | 0.367 | 1.20e-11 | 8.41e-11 | 3732 | 2713 | 1019 |
| European | No Recorded Depression | Discontinuation vs Continuation | Migraine | age_at_first_AD | 1.014 | 1.009 | 1.020 | 5.49e-07 | 3.84e-06 | 3732 | 2713 | 1019 |
| European | No Recorded Depression | Discontinuation vs Continuation | Migraine | sex_at_birth | 1.260 | 1.079 | 1.472 | 3.44e-03 | 2.41e-02 | 3732 | 2713 | 1019 |
| European | No Recorded Depression | Discontinuation vs Continuation | Migraine | Income_level | 1.000 | 1.000 | 1.000 | 2.28e-01 | 1.00e+00 | 3732 | 2713 | 1019 |
| European | No Recorded Depression | Discontinuation vs Continuation | Migraine | Education_level | 0.968 | 0.949 | 0.987 | 8.59e-04 | 6.01e-03 | 3732 | 2713 | 1019 |
| European | No Recorded Depression | Discontinuation vs Continuation | Migraine | Migraine | 0.937 | 0.872 | 1.006 | 7.36e-02 | 5.15e-01 | 3732 | 2713 | 1019 |
| European | No Recorded Depression | Discontinuation vs Continuation | Schizophrenia | (Intercept) | 0.243 | 0.162 | 0.365 | 9.99e-12 | 6.99e-11 | 3732 | 2713 | 1019 |
| European | No Recorded Depression | Discontinuation vs Continuation | Schizophrenia | age_at_first_AD | 1.014 | 1.009 | 1.020 | 6.17e-07 | 4.32e-06 | 3732 | 2713 | 1019 |
| European | No Recorded Depression | Discontinuation vs Continuation | Schizophrenia | sex_at_birth | 1.258 | 1.078 | 1.469 | 3.66e-03 | 2.56e-02 | 3732 | 2713 | 1019 |
| European | No Recorded Depression | Discontinuation vs Continuation | Schizophrenia | Income_level | 1.000 | 1.000 | 1.000 | 2.03e-01 | 1.00e+00 | 3732 | 2713 | 1019 |
| European | No Recorded Depression | Discontinuation vs Continuation | Schizophrenia | Education_level | 0.968 | 0.949 | 0.987 | 9.15e-04 | 6.40e-03 | 3732 | 2713 | 1019 |
| European | No Recorded Depression | Discontinuation vs Continuation | Schizophrenia | Schizophrenia | 1.006 | 0.936 | 1.082 | 8.68e-01 | 1.00e+00 | 3732 | 2713 | 1019 |

**Supplementary Table 10. Multivariable logistic regression results for PGS associations with antidepressant augmentation across ancestry groups and clinical subgroups in All of Us.** Each model estimates the association between 7 PGS traits and the binary outcome augmentation versus continuation — a higher-intensity treatment outcome defined at the level of the individual's overall prescribing history, in which an antidepressant episode overlapped with an atypical antipsychotic or lithium episode by at least 28 days at any point — fitted separately within each combination of genetically inferred ancestry (European, African, Latino/Mixed-American), clinical group (Full Cohort; subgroups defined by recorded depression status, recorded anxiety, and CIDI-SF–derived MDD classification), and PGS exposure. Within each model, one PGS exposure is modelled alongside a common covariate set (age at first antidepressant, sex at birth, income, and educational attainment). All PGS were residualised on the top 16 global genetic ancestry principal components and the residuals standardised to mean 0, SD 1; each odds ratio therefore reflects the change in odds per one-standard-deviation increase in the PC-adjusted PGS. Because ancestry PCs were regressed out at this stage, they are not included in the model covariate set. Columns: Population = inferred genetic ancestry; ClinicalGroup = cohort or clinical subgroup; Outcome = the binary contrast modelled; PGS = the PGS trait for that model; variable = the individual model term; odds_ratio = exponentiated coefficient; lower_95/upper_95 = 95% confidence interval bounds for the odds ratio; p_value = nominal (uncorrected) p-value; Bonf_P = Bonferroni-corrected p-value (capped at 1.00); N_Total = model sample size; N_Reference = number in the reference (continuation) group; N_Case = number in the augmentation group. Odds ratios >1 indicate higher odds of augmentation relative to continuation. Bonferroni correction was applied within each specification (ancestry × clinical group) for the max 7 PGS traits tested. P-values reported as "<2.2e-16" reflect R's floating-point reporting limit.
Reference (omitted) categories: Sex at birth: Female; Income: lowest band (<$25k); Education level: lowest band (less than high school / fewer than twelve years).

| Population | ClinicalGroup | Outcome | PGS | variable | odds_ratio | lower_95 | upper_95 | p_value | Bonf_P | N_Total | N_Cases | N_Controls |
|---|---|---|---|---|---|---|---|---|---|---|---|---|
| European | Full Cohort | Augmentation | ADHD | (Intercept) | 1.502 | 1.282 | 1.759 | 4.66e-07 | 3.26e-06 | 25145 | 3889 | 21256 |
| European | Full Cohort | Augmentation | ADHD | age_at_first_AD | 0.974 | 0.972 | 0.976 | <2.2e-16 | <2.2e-16 | 25145 | 3889 | 21256 |
| European | Full Cohort | Augmentation | ADHD | sex_at_birthMale | 1.341 | 1.242 | 1.448 | 6.15e-14 | 4.31e-13 | 25145 | 3889 | 21256 |
| European | Full Cohort | Augmentation | ADHD | Income_quantitative | 0.780 | 0.765 | 0.794 | <2.2e-16 | <2.2e-16 | 25145 | 3889 | 21256 |
| European | Full Cohort | Augmentation | ADHD | Education_levelCollege Undergraduate | 0.958 | 0.874 | 1.051 | 3.63e-01 | 1.00e+00 | 25145 | 3889 | 21256 |
| European | Full Cohort | Augmentation | ADHD | Education_levelLess than a high school degree | 1.789 | 1.435 | 2.227 | 2.06e-07 | 1.44e-06 | 25145 | 3889 | 21256 |
| European | Full Cohort | Augmentation | ADHD | Education_levelTwelve or GED | 1.264 | 1.115 | 1.433 | 2.41e-04 | 1.69e-03 | 25145 | 3889 | 21256 |
| European | Full Cohort | Augmentation | ADHD | ADHD | 1.098 | 1.059 | 1.139 | 5.14e-07 | 3.60e-06 | 25145 | 3889 | 21256 |
| European | Full Cohort | Augmentation | Migraine | (Intercept) | 1.519 | 1.297 | 1.779 | 2.09e-07 | 1.46e-06 | 25145 | 3889 | 21256 |
| European | Full Cohort | Augmentation | Migraine | age_at_first_AD | 0.974 | 0.972 | 0.976 | <2.2e-16 | <2.2e-16 | 25145 | 3889 | 21256 |
| European | Full Cohort | Augmentation | Migraine | sex_at_birthMale | 1.334 | 1.236 | 1.440 | 1.51e-13 | 1.06e-12 | 25145 | 3889 | 21256 |
| European | Full Cohort | Augmentation | Migraine | Income_quantitative | 0.777 | 0.763 | 0.791 | <2.2e-16 | <2.2e-16 | 25145 | 3889 | 21256 |
| European | Full Cohort | Augmentation | Migraine | Education_levelCollege Undergraduate | 0.972 | 0.886 | 1.065 | 5.38e-01 | 1.00e+00 | 25145 | 3889 | 21256 |
| European | Full Cohort | Augmentation | Migraine | Education_levelLess than a high school degree | 1.876 | 1.506 | 2.332 | 1.70e-08 | 1.19e-07 | 25145 | 3889 | 21256 |
| European | Full Cohort | Augmentation | Migraine | Education_levelTwelve or GED | 1.296 | 1.144 | 1.468 | 4.57e-05 | 3.20e-04 | 25145 | 3889 | 21256 |
| European | Full Cohort | Augmentation | Migraine | Migraine | 1.038 | 1.002 | 1.076 | 4.05e-02 | 2.84e-01 | 25145 | 3889 | 21256 |
| European | Full Cohort | Augmentation | Insomnia | (Intercept) | 1.489 | 1.271 | 1.744 | 8.40e-07 | 5.88e-06 | 25145 | 3889 | 21256 |
| European | Full Cohort | Augmentation | Insomnia | age_at_first_AD | 0.974 | 0.972 | 0.976 | <2.2e-16 | <2.2e-16 | 25145 | 3889 | 21256 |
| European | Full Cohort | Augmentation | Insomnia | sex_at_birthMale | 1.339 | 1.240 | 1.445 | 9.00e-14 | 6.30e-13 | 25145 | 3889 | 21256 |
| European | Full Cohort | Augmentation | Insomnia | Income_quantitative | 0.780 | 0.766 | 0.795 | <2.2e-16 | <2.2e-16 | 25145 | 3889 | 21256 |
| European | Full Cohort | Augmentation | Insomnia | Education_levelCollege Undergraduate | 0.960 | 0.876 | 1.053 | 3.85e-01 | 1.00e+00 | 25145 | 3889 | 21256 |
| European | Full Cohort | Augmentation | Insomnia | Education_levelLess than a high school degree | 1.759 | 1.410 | 2.190 | 4.76e-07 | 3.33e-06 | 25145 | 3889 | 21256 |
| European | Full Cohort | Augmentation | Insomnia | Education_levelTwelve or GED | 1.269 | 1.119 | 1.438 | 1.91e-04 | 1.34e-03 | 25145 | 3889 | 21256 |
| European | Full Cohort | Augmentation | Insomnia | Insomnia | 1.163 | 1.120 | 1.207 | 4.07e-15 | 2.85e-14 | 25145 | 3889 | 21256 |
| European | Full Cohort | Augmentation | BMI | (Intercept) | 1.512 | 1.291 | 1.770 | 2.96e-07 | 2.07e-06 | 25145 | 3889 | 21256 |
| European | Full Cohort | Augmentation | BMI | age_at_first_AD | 0.974 | 0.972 | 0.976 | <2.2e-16 | <2.2e-16 | 25145 | 3889 | 21256 |
| European | Full Cohort | Augmentation | BMI | sex_at_birthMale | 1.335 | 1.237 | 1.441 | 1.33e-13 | 9.34e-13 | 25145 | 3889 | 21256 |
| European | Full Cohort | Augmentation | BMI | Income_quantitative | 0.778 | 0.763 | 0.792 | <2.2e-16 | <2.2e-16 | 25145 | 3889 | 21256 |
| European | Full Cohort | Augmentation | BMI | Education_levelCollege Undergraduate | 0.968 | 0.883 | 1.061 | 4.83e-01 | 1.00e+00 | 25145 | 3889 | 21256 |
| European | Full Cohort | Augmentation | BMI | Education_levelLess than a high school degree | 1.858 | 1.491 | 2.310 | 2.83e-08 | 1.98e-07 | 25145 | 3889 | 21256 |
| European | Full Cohort | Augmentation | BMI | Education_levelTwelve or GED | 1.285 | 1.134 | 1.456 | 8.39e-05 | 5.87e-04 | 25145 | 3889 | 21256 |
| European | Full Cohort | Augmentation | BMI | BMI | 1.048 | 1.011 | 1.086 | 1.05e-02 | 7.37e-02 | 25145 | 3889 | 21256 |
| European | Full Cohort | Augmentation | Bipolar Disorder | (Intercept) | 1.510 | 1.289 | 1.768 | 3.30e-07 | 2.31e-06 | 25145 | 3889 | 21256 |
| European | Full Cohort | Augmentation | Bipolar Disorder | age_at_first_AD | 0.974 | 0.972 | 0.976 | <2.2e-16 | <2.2e-16 | 25145 | 3889 | 21256 |
| European | Full Cohort | Augmentation | Bipolar Disorder | sex_at_birthMale | 1.331 | 1.233 | 1.437 | 2.48e-13 | 1.73e-12 | 25145 | 3889 | 21256 |
| European | Full Cohort | Augmentation | Bipolar Disorder | Income_quantitative | 0.777 | 0.763 | 0.792 | <2.2e-16 | <2.2e-16 | 25145 | 3889 | 21256 |
| European | Full Cohort | Augmentation | Bipolar Disorder | Education_levelCollege Undergraduate | 0.977 | 0.891 | 1.071 | 6.14e-01 | 1.00e+00 | 25145 | 3889 | 21256 |
| European | Full Cohort | Augmentation | Bipolar Disorder | Education_levelLess than a high school degree | 1.888 | 1.515 | 2.348 | 1.28e-08 | 8.96e-08 | 25145 | 3889 | 21256 |
| European | Full Cohort | Augmentation | Bipolar Disorder | Education_levelTwelve or GED | 1.310 | 1.156 | 1.484 | 2.23e-05 | 1.56e-04 | 25145 | 3889 | 21256 |
| European | Full Cohort | Augmentation | Bipolar Disorder | Bipolar Disorder | 1.117 | 1.076 | 1.160 | 7.24e-09 | 5.07e-08 | 25145 | 3889 | 21256 |
| European | Full Cohort | Augmentation | Depression | (Intercept) | 1.419 | 1.209 | 1.664 | 1.74e-05 | 1.22e-04 | 25145 | 3889 | 21256 |
| European | Full Cohort | Augmentation | Depression | age_at_first_AD | 0.974 | 0.972 | 0.976 | <2.2e-16 | <2.2e-16 | 25145 | 3889 | 21256 |
| European | Full Cohort | Augmentation | Depression | sex_at_birthMale | 1.330 | 1.231 | 1.436 | 4.28e-13 | 3.00e-12 | 25145 | 3889 | 21256 |

| | | | | | | | | | | | | |
|---|---|---|---|---|---|---|---|---|---|---|---|---|
| European | Full Cohort | Augmentation | Depression | Income_quantitative | 0.785 | 0.770 | 0.800 | <2.2e-16 | <2.2e-16 | 25145 | 3889 | 21256 |
| European | Full Cohort | Augmentation | Depression | Education_levelCollege Undergraduate | 0.949 | 0.866 | 1.042 | 2.72e-01 | 1.00e+00 | 25145 | 3889 | 21256 |
| European | Full Cohort | Augmentation | Depression | Education_levelLess than a high school degree | 1.697 | 1.358 | 2.115 | 2.88e-06 | 2.02e-05 | 25145 | 3889 | 21256 |
| European | Full Cohort | Augmentation | Depression | Education_levelTwelve or GED | 1.254 | 1.106 | 1.422 | 4.21e-04 | 2.94e-03 | 25145 | 3889 | 21256 |
| European | Full Cohort | Augmentation | Depression | Depression | 1.373 | 1.322 | 1.426 | <2.2e-16 | <2.2e-16 | 25145 | 3889 | 21256 |
| European | Full Cohort | Augmentation | Schizophrenia | (Intercept) | 1.498 | 1.279 | 1.754 | 5.44e-07 | 3.81e-06 | 25145 | 3889 | 21256 |
| European | Full Cohort | Augmentation | Schizophrenia | age_at_first_AD | 0.974 | 0.972 | 0.976 | <2.2e-16 | <2.2e-16 | 25145 | 3889 | 21256 |
| European | Full Cohort | Augmentation | Schizophrenia | sex_at_birthMale | 1.330 | 1.231 | 1.435 | 3.16e-13 | 2.21e-12 | 25145 | 3889 | 21256 |
| European | Full Cohort | Augmentation | Schizophrenia | Income_quantitative | 0.778 | 0.764 | 0.793 | <2.2e-16 | <2.2e-16 | 25145 | 3889 | 21256 |
| European | Full Cohort | Augmentation | Schizophrenia | Education_levelCollege Undergraduate | 0.975 | 0.889 | 1.069 | 5.86e-01 | 1.00e+00 | 25145 | 3889 | 21256 |
| European | Full Cohort | Augmentation | Schizophrenia | Education_levelLess than a high school degree | 1.865 | 1.497 | 2.319 | 2.33e-08 | 1.63e-07 | 25145 | 3889 | 21256 |
| European | Full Cohort | Augmentation | Schizophrenia | Education_levelTwelve or GED | 1.304 | 1.150 | 1.477 | 3.15e-05 | 2.21e-04 | 25145 | 3889 | 21256 |
| European | Full Cohort | Augmentation | Schizophrenia | Schizophrenia | 1.109 | 1.070 | 1.150 | 1.59e-08 | 1.11e-07 | 25145 | 3889 | 21256 |
| European | Recorded Depression | Augmentation | ADHD | (Intercept) | 2.356 | 1.928 | 2.879 | <2.2e-16 | 3.74e-16 | 10915 | 2946 | 7969 |
| European | Recorded Depression | Augmentation | ADHD | age_at_first_AD | 0.973 | 0.970 | 0.976 | <2.2e-16 | <2.2e-16 | 10915 | 2946 | 7969 |
| European | Recorded Depression | Augmentation | ADHD | sex_at_birthMale | 1.386 | 1.259 | 1.525 | 2.14e-11 | 1.50e-10 | 10915 | 2946 | 7969 |
| European | Recorded Depression | Augmentation | ADHD | Income_quantitative | 0.822 | 0.803 | 0.841 | <2.2e-16 | <2.2e-16 | 10915 | 2946 | 7969 |
| European | Recorded Depression | Augmentation | ADHD | Education_levelCollege Undergraduate | 0.985 | 0.880 | 1.104 | 7.94e-01 | 1.00e+00 | 10915 | 2946 | 7969 |
| European | Recorded Depression | Augmentation | ADHD | Education_levelLess than a high school degree | 1.840 | 1.391 | 2.435 | 1.97e-05 | 1.38e-04 | 10915 | 2946 | 7969 |
| European | Recorded Depression | Augmentation | ADHD | Education_levelTwelve or GED | 1.330 | 1.137 | 1.555 | 3.58e-04 | 2.50e-03 | 10915 | 2946 | 7969 |
| European | Recorded Depression | Augmentation | ADHD | ADHD | 1.065 | 1.017 | 1.114 | 6.90e-03 | 4.83e-02 | 10915 | 2946 | 7969 |
| European | Recorded Depression | Augmentation | Migraine | (Intercept) | 2.367 | 1.937 | 2.893 | <2.2e-16 | 2.47e-16 | 10915 | 2946 | 7969 |
| European | Recorded Depression | Augmentation | Migraine | age_at_first_AD | 0.973 | 0.970 | 0.976 | <2.2e-16 | <2.2e-16 | 10915 | 2946 | 7969 |
| European | Recorded Depression | Augmentation | Migraine | sex_at_birthMale | 1.381 | 1.255 | 1.519 | 3.26e-11 | 2.29e-10 | 10915 | 2946 | 7969 |
| European | Recorded Depression | Augmentation | Migraine | Income_quantitative | 0.820 | 0.801 | 0.839 | <2.2e-16 | <2.2e-16 | 10915 | 2946 | 7969 |
| European | Recorded Depression | Augmentation | Migraine | Education_levelCollege Undergraduate | 0.995 | 0.888 | 1.114 | 9.24e-01 | 1.00e+00 | 10915 | 2946 | 7969 |
| European | Recorded Depression | Augmentation | Migraine | Education_levelLess than a high school degree | 1.902 | 1.439 | 2.516 | 6.24e-06 | 4.37e-05 | 10915 | 2946 | 7969 |
| European | Recorded Depression | Augmentation | Migraine | Education_levelTwelve or GED | 1.354 | 1.159 | 1.583 | 1.38e-04 | 9.63e-04 | 10915 | 2946 | 7969 |
| European | Recorded Depression | Augmentation | Migraine | Migraine | 1.058 | 1.013 | 1.106 | 1.17e-02 | 8.18e-02 | 10915 | 2946 | 7969 |
| European | Recorded Depression | Augmentation | Insomnia | (Intercept) | 2.333 | 1.909 | 2.852 | <2.2e-16 | 9.08e-16 | 10915 | 2946 | 7969 |
| European | Recorded Depression | Augmentation | Insomnia | age_at_first_AD | 0.973 | 0.971 | 0.976 | <2.2e-16 | <2.2e-16 | 10915 | 2946 | 7969 |
| European | Recorded Depression | Augmentation | Insomnia | sex_at_birthMale | 1.383 | 1.257 | 1.522 | 2.87e-11 | 2.01e-10 | 10915 | 2946 | 7969 |
| European | Recorded Depression | Augmentation | Insomnia | Income_quantitative | 0.823 | 0.804 | 0.842 | <2.2e-16 | <2.2e-16 | 10915 | 2946 | 7969 |
| European | Recorded Depression | Augmentation | Insomnia | Education_levelCollege Undergraduate | 0.986 | 0.881 | 1.105 | 8.10e-01 | 1.00e+00 | 10915 | 2946 | 7969 |
| European | Recorded Depression | Augmentation | Insomnia | Education_levelLess than a high school degree | 1.792 | 1.354 | 2.372 | 4.51e-05 | 3.16e-04 | 10915 | 2946 | 7969 |
| European | Recorded Depression | Augmentation | Insomnia | Education_levelTwelve or GED | 1.332 | 1.139 | 1.557 | 3.29e-04 | 2.30e-03 | 10915 | 2946 | 7969 |
| European | Recorded Depression | Augmentation | Insomnia | Insomnia | 1.128 | 1.077 | 1.183 | 4.65e-07 | 3.25e-06 | 10915 | 2946 | 7969 |
| European | Recorded Depression | Augmentation | BMI | (Intercept) | 2.371 | 1.940 | 2.897 | <2.2e-16 | 2.22e-16 | 10915 | 2946 | 7969 |
| European | Recorded Depression | Augmentation | BMI | age_at_first_AD | 0.973 | 0.970 | 0.976 | <2.2e-16 | <2.2e-16 | 10915 | 2946 | 7969 |
| European | Recorded Depression | Augmentation | BMI | sex_at_birthMale | 1.380 | 1.254 | 1.518 | 3.86e-11 | 2.70e-10 | 10915 | 2946 | 7969 |
| European | Recorded Depression | Augmentation | BMI | Income_quantitative | 0.820 | 0.801 | 0.839 | <2.2e-16 | <2.2e-16 | 10915 | 2946 | 7969 |
| European | Recorded Depression | Augmentation | BMI | Education_levelCollege Undergraduate | 0.996 | 0.890 | 1.116 | 9.43e-01 | 1.00e+00 | 10915 | 2946 | 7969 |
| European | Recorded Depression | Augmentation | BMI | Education_levelLess than a high school degree | 1.899 | 1.437 | 2.512 | 6.70e-06 | 4.69e-05 | 10915 | 2946 | 7969 |
| European | Recorded Depression | Augmentation | BMI | Education_levelTwelve or GED | 1.355 | 1.159 | 1.584 | 1.39e-04 | 9.74e-04 | 10915 | 2946 | 7969 |
| European | Recorded Depression | Augmentation | BMI | BMI | 1.001 | 0.957 | 1.047 | 9.71e-01 | 1.00e+00 | 10915 | 2946 | 7969 |
| European | Recorded Depression | Augmentation | Bipolar Disorder | (Intercept) | 2.379 | 1.947 | 2.908 | <2.2e-16 | <2.2e-16 | 10915 | 2946 | 7969 |
| European | Recorded Depression | Augmentation | Bipolar Disorder | age_at_first_AD | 0.973 | 0.970 | 0.976 | <2.2e-16 | <2.2e-16 | 10915 | 2946 | 7969 |
| European | Recorded Depression | Augmentation | Bipolar Disorder | sex_at_birthMale | 1.381 | 1.255 | 1.519 | 3.50e-11 | 2.45e-10 | 10915 | 2946 | 7969 |
| European | Recorded Depression | Augmentation | Bipolar Disorder | Income_quantitative | 0.819 | 0.801 | 0.838 | <2.2e-16 | <2.2e-16 | 10915 | 2946 | 7969 |
| European | Recorded Depression | Augmentation | Bipolar Disorder | Education_levelCollege Undergraduate | 0.997 | 0.891 | 1.117 | 9.60e-01 | 1.00e+00 | 10915 | 2946 | 7969 |
| European | Recorded Depression | Augmentation | Bipolar Disorder | Education_levelLess than a high school degree | 1.904 | 1.440 | 2.518 | 6.15e-06 | 4.30e-05 | 10915 | 2946 | 7969 |
| European | Recorded Depression | Augmentation | Bipolar Disorder | Education_levelTwelve or GED | 1.361 | 1.164 | 1.590 | 1.09e-04 | 7.66e-04 | 10915 | 2946 | 7969 |
| European | Recorded Depression | Augmentation | Bipolar Disorder | Bipolar Disorder | 1.078 | 1.029 | 1.129 | 1.57e-03 | 1.10e-02 | 10915 | 2946 | 7969 |
| European | Recorded Depression | Augmentation | Depression | (Intercept) | 2.324 | 1.900 | 2.843 | 2.45e-16 | 1.71e-15 | 10915 | 2946 | 7969 |
| European | Recorded Depression | Augmentation | Depression | age_at_first_AD | 0.973 | 0.970 | 0.976 | <2.2e-16 | <2.2e-16 | 10915 | 2946 | 7969 |

| | | | | | | | | | | | | |
|---|---|---|---|---|---|---|---|---|---|---|---|---|
| European | Recorded Depression | Augmentation | Depression | sex_at_birthMale | 1.383 | 1.256 | 1.522 | 3.54e-11 | 2.48e-10 | 10915 | 2946 | 7969 |
| European | Recorded Depression | Augmentation | Depression | Income_quantitative | 0.826 | 0.807 | 0.845 | <2.2e-16 | <2.2e-16 | 10915 | 2946 | 7969 |
| European | Recorded Depression | Augmentation | Depression | Education_levelCollege Undergraduate | 0.975 | 0.870 | 1.093 | 6.62e-01 | 1.00e+00 | 10915 | 2946 | 7969 |
| European | Recorded Depression | Augmentation | Depression | Education_levelLess than a high school degree | 1.757 | 1.327 | 2.328 | 8.35e-05 | 5.85e-04 | 10915 | 2946 | 7969 |
| European | Recorded Depression | Augmentation | Depression | Education_levelTwelve or GED | 1.315 | 1.124 | 1.539 | 6.26e-04 | 4.38e-03 | 10915 | 2946 | 7969 |
| European | Recorded Depression | Augmentation | Depression | Depression | 1.292 | 1.234 | 1.354 | <2.2e-16 | <2.2e-16 | 10915 | 2946 | 7969 |
| European | Recorded Depression | Augmentation | Schizophrenia | (Intercept) | 2.362 | 1.933 | 2.886 | <2.2e-16 | 3.07e-16 | 10915 | 2946 | 7969 |
| European | Recorded Depression | Augmentation | Schizophrenia | age_at_first_AD | 0.973 | 0.970 | 0.976 | <2.2e-16 | <2.2e-16 | 10915 | 2946 | 7969 |
| European | Recorded Depression | Augmentation | Schizophrenia | sex_at_birthMale | 1.379 | 1.253 | 1.517 | 4.41e-11 | 3.08e-10 | 10915 | 2946 | 7969 |
| European | Recorded Depression | Augmentation | Schizophrenia | Income_quantitative | 0.820 | 0.801 | 0.839 | <2.2e-16 | <2.2e-16 | 10915 | 2946 | 7969 |
| European | Recorded Depression | Augmentation | Schizophrenia | Education_levelCollege Undergraduate | 0.996 | 0.890 | 1.116 | 9.45e-01 | 1.00e+00 | 10915 | 2946 | 7969 |
| European | Recorded Depression | Augmentation | Schizophrenia | Education_levelLess than a high school degree | 1.870 | 1.415 | 2.474 | 1.10e-05 | 7.72e-05 | 10915 | 2946 | 7969 |
| European | Recorded Depression | Augmentation | Schizophrenia | Education_levelTwelve or GED | 1.358 | 1.161 | 1.587 | 1.22e-04 | 8.57e-04 | 10915 | 2946 | 7969 |
| European | Recorded Depression | Augmentation | Schizophrenia | Schizophrenia | 1.093 | 1.046 | 1.143 | 8.74e-05 | 6.12e-04 | 10915 | 2946 | 7969 |
| European | No Recorded Depression | Augmentation | ADHD | (Intercept) | 0.491 | 0.364 | 0.659 | 2.56e-06 | 1.79e-05 | 14230 | 943 | 13287 |
| European | No Recorded Depression | Augmentation | ADHD | age_at_first_AD | 0.979 | 0.975 | 0.983 | <2.2e-16 | <2.2e-16 | 14230 | 943 | 13287 |
| European | No Recorded Depression | Augmentation | ADHD | sex_at_birthMale | 1.269 | 1.097 | 1.465 | 1.23e-03 | 8.61e-03 | 14230 | 943 | 13287 |
| European | No Recorded Depression | Augmentation | ADHD | Income_quantitative | 0.776 | 0.749 | 0.803 | <2.2e-16 | <2.2e-16 | 14230 | 943 | 13287 |
| European | No Recorded Depression | Augmentation | ADHD | Education_levelCollege Undergraduate | 0.980 | 0.822 | 1.171 | 8.20e-01 | 1.00e+00 | 14230 | 943 | 13287 |
| European | No Recorded Depression | Augmentation | ADHD | Education_levelLess than a high school degree | 1.875 | 1.241 | 2.783 | 2.23e-03 | 1.56e-02 | 14230 | 943 | 13287 |
| European | No Recorded Depression | Augmentation | ADHD | Education_levelTwelve or GED | 1.312 | 1.036 | 1.659 | 2.39e-02 | 1.67e-01 | 14230 | 943 | 13287 |
| European | No Recorded Depression | Augmentation | ADHD | ADHD | 1.132 | 1.057 | 1.213 | 4.16e-04 | 2.91e-03 | 14230 | 943 | 13287 |
| European | No Recorded Depression | Augmentation | Migraine | (Intercept) | 0.499 | 0.371 | 0.670 | 4.08e-06 | 2.85e-05 | 14230 | 943 | 13287 |
| European | No Recorded Depression | Augmentation | Migraine | age_at_first_AD | 0.979 | 0.975 | 0.983 | <2.2e-16 | <2.2e-16 | 14230 | 943 | 13287 |
| European | No Recorded Depression | Augmentation | Migraine | sex_at_birthMale | 1.263 | 1.092 | 1.458 | 1.53e-03 | 1.07e-02 | 14230 | 943 | 13287 |
| European | No Recorded Depression | Augmentation | Migraine | Income_quantitative | 0.773 | 0.746 | 0.800 | <2.2e-16 | <2.2e-16 | 14230 | 943 | 13287 |
| European | No Recorded Depression | Augmentation | Migraine | Education_levelCollege Undergraduate | 0.999 | 0.839 | 1.194 | 9.94e-01 | 1.00e+00 | 14230 | 943 | 13287 |
| European | No Recorded Depression | Augmentation | Migraine | Education_levelLess than a high school degree | 2.011 | 1.334 | 2.978 | 6.32e-04 | 4.42e-03 | 14230 | 943 | 13287 |
| European | No Recorded Depression | Augmentation | Migraine | Education_levelTwelve or GED | 1.357 | 1.072 | 1.714 | 1.08e-02 | 7.58e-02 | 14230 | 943 | 13287 |
| European | No Recorded Depression | Augmentation | Migraine | Migraine | 1.005 | 0.939 | 1.075 | 8.92e-01 | 1.00e+00 | 14230 | 943 | 13287 |
| European | No Recorded Depression | Augmentation | Insomnia | (Intercept) | 0.490 | 0.363 | 0.658 | 2.45e-06 | 1.72e-05 | 14230 | 943 | 13287 |
| European | No Recorded Depression | Augmentation | Insomnia | age_at_first_AD | 0.979 | 0.975 | 0.983 | <2.2e-16 | <2.2e-16 | 14230 | 943 | 13287 |
| European | No Recorded Depression | Augmentation | Insomnia | sex_at_birthMale | 1.268 | 1.096 | 1.464 | 1.29e-03 | 9.06e-03 | 14230 | 943 | 13287 |
| European | No Recorded Depression | Augmentation | Insomnia | Income_quantitative | 0.776 | 0.749 | 0.803 | <2.2e-16 | <2.2e-16 | 14230 | 943 | 13287 |
| European | No Recorded Depression | Augmentation | Insomnia | Education_levelCollege Undergraduate | 0.983 | 0.825 | 1.175 | 8.51e-01 | 1.00e+00 | 14230 | 943 | 13287 |
| European | No Recorded Depression | Augmentation | Insomnia | Education_levelLess than a high school degree | 1.885 | 1.248 | 2.797 | 2.04e-03 | 1.43e-02 | 14230 | 943 | 13287 |
| European | No Recorded Depression | Augmentation | Insomnia | Education_levelTwelve or GED | 1.322 | 1.044 | 1.671 | 2.01e-02 | 1.41e-01 | 14230 | 943 | 13287 |
| European | No Recorded Depression | Augmentation | Insomnia | Insomnia | 1.181 | 1.101 | 1.268 | 3.83e-06 | 2.68e-05 | 14230 | 943 | 13287 |
| European | No Recorded Depression | Augmentation | BMI | (Intercept) | 0.498 | 0.370 | 0.668 | 3.89e-06 | 2.72e-05 | 14230 | 943 | 13287 |
| European | No Recorded Depression | Augmentation | BMI | age_at_first_AD | 0.979 | 0.975 | 0.983 | <2.2e-16 | <2.2e-16 | 14230 | 943 | 13287 |
| European | No Recorded Depression | Augmentation | BMI | sex_at_birthMale | 1.264 | 1.093 | 1.460 | 1.46e-03 | 1.02e-02 | 14230 | 943 | 13287 |
| European | No Recorded Depression | Augmentation | BMI | Income_quantitative | 0.774 | 0.747 | 0.801 | <2.2e-16 | <2.2e-16 | 14230 | 943 | 13287 |
| European | No Recorded Depression | Augmentation | BMI | Education_levelCollege Undergraduate | 0.991 | 0.832 | 1.184 | 9.19e-01 | 1.00e+00 | 14230 | 943 | 13287 |
| European | No Recorded Depression | Augmentation | BMI | Education_levelLess than a high school degree | 1.980 | 1.313 | 2.933 | 8.41e-04 | 5.89e-03 | 14230 | 943 | 13287 |
| European | No Recorded Depression | Augmentation | BMI | Education_levelTwelve or GED | 1.339 | 1.058 | 1.693 | 1.49e-02 | 1.04e-01 | 14230 | 943 | 13287 |
| European | No Recorded Depression | Augmentation | BMI | BMI | 1.060 | 0.991 | 1.135 | 9.01e-02 | 6.31e-01 | 14230 | 943 | 13287 |
| European | No Recorded Depression | Augmentation | Bipolar Disorder | (Intercept) | 0.485 | 0.360 | 0.652 | 1.79e-06 | 1.25e-05 | 14230 | 943 | 13287 |
| European | No Recorded Depression | Augmentation | Bipolar Disorder | age_at_first_AD | 0.979 | 0.975 | 0.983 | <2.2e-16 | <2.2e-16 | 14230 | 943 | 13287 |
| European | No Recorded Depression | Augmentation | Bipolar Disorder | sex_at_birthMale | 1.254 | 1.084 | 1.448 | 2.14e-03 | 1.50e-02 | 14230 | 943 | 13287 |
| European | No Recorded Depression | Augmentation | Bipolar Disorder | Income_quantitative | 0.775 | 0.748 | 0.802 | <2.2e-16 | <2.2e-16 | 14230 | 943 | 13287 |
| European | No Recorded Depression | Augmentation | Bipolar Disorder | Education_levelCollege Undergraduate | 1.007 | 0.845 | 1.203 | 9.39e-01 | 1.00e+00 | 14230 | 943 | 13287 |
| European | No Recorded Depression | Augmentation | Bipolar Disorder | Education_levelLess than a high school degree | 2.015 | 1.336 | 2.988 | 6.26e-04 | 4.38e-03 | 14230 | 943 | 13287 |
| European | No Recorded Depression | Augmentation | Bipolar Disorder | Education_levelTwelve or GED | 1.380 | 1.090 | 1.744 | 7.24e-03 | 5.07e-02 | 14230 | 943 | 13287 |
| European | No Recorded Depression | Augmentation | Bipolar Disorder | Bipolar Disorder | 1.175 | 1.094 | 1.262 | 9.97e-06 | 6.98e-05 | 14230 | 943 | 13287 |
| European | No Recorded Depression | Augmentation | Depression | (Intercept) | 0.451 | 0.333 | 0.607 | 1.83e-07 | 1.28e-06 | 14230 | 943 | 13287 |

| | | | | | | | | | | | | |
|---|---|---|---|---|---|---|---|---|---|---|---|---|
| European | No Recorded Depression | Augmentation | Depression | age_at_first_AD | 0.980 | 0.976 | 0.984 | <2.2e-16 | <2.2e-16 | 14230 | 943 | 13287 |
| European | No Recorded Depression | Augmentation | Depression | sex_at_birthMale | 1.249 | 1.079 | 1.442 | 2.67e-03 | 1.87e-02 | 14230 | 943 | 13287 |
| European | No Recorded Depression | Augmentation | Depression | Income_quantitative | 0.780 | 0.754 | 0.808 | <2.2e-16 | <2.2e-16 | 14230 | 943 | 13287 |
| European | No Recorded Depression | Augmentation | Depression | Education_levelCollege Undergraduate | 0.976 | 0.819 | 1.167 | 7.89e-01 | 1.00e+00 | 14230 | 943 | 13287 |
| European | No Recorded Depression | Augmentation | Depression | Education_levelLess than a high school degree | 1.767 | 1.166 | 2.631 | 5.99e-03 | 4.20e-02 | 14230 | 943 | 13287 |
| European | No Recorded Depression | Augmentation | Depression | Education_levelTwelve or GED | 1.308 | 1.032 | 1.655 | 2.57e-02 | 1.80e-01 | 14230 | 943 | 13287 |
| European | No Recorded Depression | Augmentation | Depression | Depression | 1.400 | 1.304 | 1.504 | <2.2e-16 | <2.2e-16 | 14230 | 943 | 13287 |
| European | No Recorded Depression | Augmentation | Schizophrenia | (Intercept) | 0.480 | 0.356 | 0.644 | 1.22e-06 | 8.56e-06 | 14230 | 943 | 13287 |
| European | No Recorded Depression | Augmentation | Schizophrenia | age_at_first_AD | 0.979 | 0.975 | 0.983 | <2.2e-16 | <2.2e-16 | 14230 | 943 | 13287 |
| European | No Recorded Depression | Augmentation | Schizophrenia | sex_at_birthMale | 1.254 | 1.084 | 1.448 | 2.15e-03 | 1.50e-02 | 14230 | 943 | 13287 |
| European | No Recorded Depression | Augmentation | Schizophrenia | Income_quantitative | 0.776 | 0.749 | 0.803 | <2.2e-16 | <2.2e-16 | 14230 | 943 | 13287 |
| European | No Recorded Depression | Augmentation | Schizophrenia | Education_levelCollege Undergraduate | 1.004 | 0.843 | 1.200 | 9.63e-01 | 1.00e+00 | 14230 | 943 | 13287 |
| European | No Recorded Depression | Augmentation | Schizophrenia | Education_levelLess than a high school degree | 2.023 | 1.342 | 2.998 | 5.71e-04 | 4.00e-03 | 14230 | 943 | 13287 |
| European | No Recorded Depression | Augmentation | Schizophrenia | Education_levelTwelve or GED | 1.367 | 1.080 | 1.728 | 9.04e-03 | 6.33e-02 | 14230 | 943 | 13287 |
| European | No Recorded Depression | Augmentation | Schizophrenia | Schizophrenia | 1.165 | 1.088 | 1.247 | 1.25e-05 | 8.77e-05 | 14230 | 943 | 13287 |
| European | Recorded Anxiety | Augmentation | ADHD | (Intercept) | 1.630 | 1.356 | 1.958 | 1.86e-07 | 1.30e-06 | 14887 | 3089 | 11798 |
| European | Recorded Anxiety | Augmentation | ADHD | age_at_first_AD | 0.978 | 0.976 | 0.981 | <2.2e-16 | <2.2e-16 | 14887 | 3089 | 11798 |
| European | Recorded Anxiety | Augmentation | ADHD | sex_at_birthMale | 1.497 | 1.368 | 1.638 | <2.2e-16 | <2.2e-16 | 14887 | 3089 | 11798 |
| European | Recorded Anxiety | Augmentation | ADHD | Income_quantitative | 0.775 | 0.758 | 0.792 | <2.2e-16 | <2.2e-16 | 14887 | 3089 | 11798 |
| European | Recorded Anxiety | Augmentation | ADHD | Education_levelCollege Undergraduate | 0.962 | 0.864 | 1.072 | 4.82e-01 | 1.00e+00 | 14887 | 3089 | 11798 |
| European | Recorded Anxiety | Augmentation | ADHD | Education_levelLess than a high school degree | 1.513 | 1.165 | 1.962 | 1.80e-03 | 1.26e-02 | 14887 | 3089 | 11798 |
| European | Recorded Anxiety | Augmentation | ADHD | Education_levelTwelve or GED | 1.328 | 1.147 | 1.537 | 1.43e-04 | 9.98e-04 | 14887 | 3089 | 11798 |
| European | Recorded Anxiety | Augmentation | ADHD | ADHD | 1.068 | 1.024 | 1.115 | 2.51e-03 | 1.76e-02 | 14887 | 3089 | 11798 |
| European | Recorded Anxiety | Augmentation | Migraine | (Intercept) | 1.638 | 1.363 | 1.968 | 1.36e-07 | 9.53e-07 | 14887 | 3089 | 11798 |
| European | Recorded Anxiety | Augmentation | Migraine | age_at_first_AD | 0.978 | 0.976 | 0.981 | <2.2e-16 | <2.2e-16 | 14887 | 3089 | 11798 |
| European | Recorded Anxiety | Augmentation | Migraine | sex_at_birthMale | 1.492 | 1.363 | 1.632 | <2.2e-16 | <2.2e-16 | 14887 | 3089 | 11798 |
| European | Recorded Anxiety | Augmentation | Migraine | Income_quantitative | 0.773 | 0.757 | 0.790 | <2.2e-16 | <2.2e-16 | 14887 | 3089 | 11798 |
| European | Recorded Anxiety | Augmentation | Migraine | Education_levelCollege Undergraduate | 0.971 | 0.872 | 1.081 | 5.85e-01 | 1.00e+00 | 14887 | 3089 | 11798 |
| European | Recorded Anxiety | Augmentation | Migraine | Education_levelLess than a high school degree | 1.560 | 1.203 | 2.021 | 7.72e-04 | 5.41e-03 | 14887 | 3089 | 11798 |
| European | Recorded Anxiety | Augmentation | Migraine | Education_levelTwelve or GED | 1.350 | 1.167 | 1.561 | 5.50e-05 | 3.85e-04 | 14887 | 3089 | 11798 |
| European | Recorded Anxiety | Augmentation | Migraine | Migraine | 1.058 | 1.015 | 1.103 | 7.93e-03 | 5.55e-02 | 14887 | 3089 | 11798 |
| European | Recorded Anxiety | Augmentation | Insomnia | (Intercept) | 1.618 | 1.346 | 1.945 | 2.94e-07 | 2.06e-06 | 14887 | 3089 | 11798 |
| European | Recorded Anxiety | Augmentation | Insomnia | age_at_first_AD | 0.978 | 0.976 | 0.981 | <2.2e-16 | <2.2e-16 | 14887 | 3089 | 11798 |
| European | Recorded Anxiety | Augmentation | Insomnia | sex_at_birthMale | 1.499 | 1.369 | 1.640 | <2.2e-16 | <2.2e-16 | 14887 | 3089 | 11798 |
| European | Recorded Anxiety | Augmentation | Insomnia | Income_quantitative | 0.776 | 0.759 | 0.793 | <2.2e-16 | <2.2e-16 | 14887 | 3089 | 11798 |
| European | Recorded Anxiety | Augmentation | Insomnia | Education_levelCollege Undergraduate | 0.957 | 0.859 | 1.066 | 4.18e-01 | 1.00e+00 | 14887 | 3089 | 11798 |
| European | Recorded Anxiety | Augmentation | Insomnia | Education_levelLess than a high school degree | 1.459 | 1.123 | 1.892 | 4.52e-03 | 3.17e-02 | 14887 | 3089 | 11798 |
| European | Recorded Anxiety | Augmentation | Insomnia | Education_levelTwelve or GED | 1.318 | 1.139 | 1.525 | 2.14e-04 | 1.50e-03 | 14887 | 3089 | 11798 |
| European | Recorded Anxiety | Augmentation | Insomnia | Insomnia | 1.161 | 1.111 | 1.213 | 3.50e-11 | 2.45e-10 | 14887 | 3089 | 11798 |
| European | Recorded Anxiety | Augmentation | BMI | (Intercept) | 1.634 | 1.360 | 1.964 | 1.57e-07 | 1.10e-06 | 14887 | 3089 | 11798 |
| European | Recorded Anxiety | Augmentation | BMI | age_at_first_AD | 0.978 | 0.976 | 0.981 | <2.2e-16 | <2.2e-16 | 14887 | 3089 | 11798 |
| European | Recorded Anxiety | Augmentation | BMI | sex_at_birthMale | 1.494 | 1.365 | 1.634 | <2.2e-16 | <2.2e-16 | 14887 | 3089 | 11798 |
| European | Recorded Anxiety | Augmentation | BMI | Income_quantitative | 0.774 | 0.757 | 0.791 | <2.2e-16 | <2.2e-16 | 14887 | 3089 | 11798 |
| European | Recorded Anxiety | Augmentation | BMI | Education_levelCollege Undergraduate | 0.967 | 0.869 | 1.077 | 5.37e-01 | 1.00e+00 | 14887 | 3089 | 11798 |
| European | Recorded Anxiety | Augmentation | BMI | Education_levelLess than a high school degree | 1.545 | 1.191 | 2.001 | 1.01e-03 | 7.07e-03 | 14887 | 3089 | 11798 |
| European | Recorded Anxiety | Augmentation | BMI | Education_levelTwelve or GED | 1.338 | 1.156 | 1.548 | 9.45e-05 | 6.61e-04 | 14887 | 3089 | 11798 |
| European | Recorded Anxiety | Augmentation | BMI | BMI | 1.049 | 1.007 | 1.094 | 2.27e-02 | 1.59e-01 | 14887 | 3089 | 11798 |
| European | Recorded Anxiety | Augmentation | Bipolar Disorder | (Intercept) | 1.633 | 1.359 | 1.962 | 1.68e-07 | 1.18e-06 | 14887 | 3089 | 11798 |
| European | Recorded Anxiety | Augmentation | Bipolar Disorder | age_at_first_AD | 0.978 | 0.976 | 0.981 | <2.2e-16 | <2.2e-16 | 14887 | 3089 | 11798 |
| European | Recorded Anxiety | Augmentation | Bipolar Disorder | sex_at_birthMale | 1.489 | 1.361 | 1.629 | <2.2e-16 | <2.2e-16 | 14887 | 3089 | 11798 |
| European | Recorded Anxiety | Augmentation | Bipolar Disorder | Income_quantitative | 0.774 | 0.757 | 0.790 | <2.2e-16 | <2.2e-16 | 14887 | 3089 | 11798 |
| European | Recorded Anxiety | Augmentation | Bipolar Disorder | Education_levelCollege Undergraduate | 0.977 | 0.877 | 1.087 | 6.63e-01 | 1.00e+00 | 14887 | 3089 | 11798 |
| European | Recorded Anxiety | Augmentation | Bipolar Disorder | Education_levelLess than a high school degree | 1.568 | 1.208 | 2.031 | 6.84e-04 | 4.79e-03 | 14887 | 3089 | 11798 |
| European | Recorded Anxiety | Augmentation | Bipolar Disorder | Education_levelTwelve or GED | 1.361 | 1.176 | 1.574 | 3.47e-05 | 2.43e-04 | 14887 | 3089 | 11798 |
| European | Recorded Anxiety | Augmentation | Bipolar Disorder | Bipolar Disorder | 1.108 | 1.060 | 1.157 | 4.59e-06 | 3.21e-05 | 14887 | 3089 | 11798 |

| | | | | | | | | | | | | |
|---|---|---|---|---|---|---|---|---|---|---|---|---|
| European | Recorded Anxiety | Augmentation | Depression | (Intercept) | 1.574 | 1.308 | 1.894 | 1.56e-06 | 1.10e-05 | 14887 | 3089 | 11798 |
| European | Recorded Anxiety | Augmentation | Depression | age_at_first_AD | 0.978 | 0.976 | 0.981 | <2.2e-16 | <2.2e-16 | 14887 | 3089 | 11798 |
| European | Recorded Anxiety | Augmentation | Depression | sex_at_birthMale | 1.491 | 1.362 | 1.632 | <2.2e-16 | <2.2e-16 | 14887 | 3089 | 11798 |
| European | Recorded Anxiety | Augmentation | Depression | Income_quantitative | 0.780 | 0.763 | 0.797 | <2.2e-16 | <2.2e-16 | 14887 | 3089 | 11798 |
| European | Recorded Anxiety | Augmentation | Depression | Education_levelCollege Undergraduate | 0.945 | 0.848 | 1.053 | 3.02e-01 | 1.00e+00 | 14887 | 3089 | 11798 |
| European | Recorded Anxiety | Augmentation | Depression | Education_levelLess than a high school degree | 1.431 | 1.100 | 1.858 | 7.35e-03 | 5.14e-02 | 14887 | 3089 | 11798 |
| European | Recorded Anxiety | Augmentation | Depression | Education_levelTwelve or GED | 1.303 | 1.125 | 1.509 | 4.16e-04 | 2.91e-03 | 14887 | 3089 | 11798 |
| European | Recorded Anxiety | Augmentation | Depression | Depression | 1.337 | 1.280 | 1.397 | <2.2e-16 | <2.2e-16 | 14887 | 3089 | 11798 |
| European | Recorded Anxiety | Augmentation | Schizophrenia | (Intercept) | 1.629 | 1.356 | 1.957 | 1.92e-07 | 1.34e-06 | 14887 | 3089 | 11798 |
| European | Recorded Anxiety | Augmentation | Schizophrenia | age_at_first_AD | 0.978 | 0.976 | 0.981 | <2.2e-16 | <2.2e-16 | 14887 | 3089 | 11798 |
| European | Recorded Anxiety | Augmentation | Schizophrenia | sex_at_birthMale | 1.487 | 1.359 | 1.627 | <2.2e-16 | <2.2e-16 | 14887 | 3089 | 11798 |
| European | Recorded Anxiety | Augmentation | Schizophrenia | Income_quantitative | 0.774 | 0.758 | 0.791 | <2.2e-16 | <2.2e-16 | 14887 | 3089 | 11798 |
| European | Recorded Anxiety | Augmentation | Schizophrenia | Education_levelCollege Undergraduate | 0.973 | 0.875 | 1.084 | 6.22e-01 | 1.00e+00 | 14887 | 3089 | 11798 |
| European | Recorded Anxiety | Augmentation | Schizophrenia | Education_levelLess than a high school degree | 1.553 | 1.197 | 2.011 | 8.83e-04 | 6.18e-03 | 14887 | 3089 | 11798 |
| European | Recorded Anxiety | Augmentation | Schizophrenia | Education_levelTwelve or GED | 1.355 | 1.171 | 1.568 | 4.43e-05 | 3.10e-04 | 14887 | 3089 | 11798 |
| European | Recorded Anxiety | Augmentation | Schizophrenia | Schizophrenia | 1.086 | 1.042 | 1.133 | 1.04e-04 | 7.29e-04 | 14887 | 3089 | 11798 |
| European | CIDI-MDD | Augmentation | ADHD | (Intercept) | 2.298 | 1.541 | 3.429 | 4.53e-05 | 3.17e-04 | 3022 | 673 | 2349 |
| European | CIDI-MDD | Augmentation | ADHD | age_at_first_AD | 0.971 | 0.965 | 0.977 | <2.2e-16 | <2.2e-16 | 3022 | 673 | 2349 |
| European | CIDI-MDD | Augmentation | ADHD | sex_at_birthMale | 1.437 | 1.172 | 1.759 | 4.66e-04 | 3.26e-03 | 3022 | 673 | 2349 |
| European | CIDI-MDD | Augmentation | ADHD | Income_quantitative | 0.814 | 0.778 | 0.852 | <2.2e-16 | <2.2e-16 | 3022 | 673 | 2349 |
| European | CIDI-MDD | Augmentation | ADHD | Education_levelCollege Undergraduate | 1.027 | 0.832 | 1.271 | 8.05e-01 | 1.00e+00 | 3022 | 673 | 2349 |
| European | CIDI-MDD | Augmentation | ADHD | Education_levelLess than a high school degree | 0.470 | 0.130 | 1.346 | 1.93e-01 | 1.00e+00 | 3022 | 673 | 2349 |
| European | CIDI-MDD | Augmentation | ADHD | Education_levelTwelve or GED | 1.484 | 1.038 | 2.111 | 2.91e-02 | 2.04e-01 | 3022 | 673 | 2349 |
| European | CIDI-MDD | Augmentation | ADHD | ADHD | 1.121 | 1.022 | 1.230 | 1.54e-02 | 1.08e-01 | 3022 | 673 | 2349 |
| European | CIDI-MDD | Augmentation | Migraine | (Intercept) | 2.312 | 1.552 | 3.449 | 3.87e-05 | 2.71e-04 | 3022 | 673 | 2349 |
| European | CIDI-MDD | Augmentation | Migraine | age_at_first_AD | 0.971 | 0.965 | 0.977 | <2.2e-16 | <2.2e-16 | 3022 | 673 | 2349 |
| European | CIDI-MDD | Augmentation | Migraine | sex_at_birthMale | 1.422 | 1.160 | 1.741 | 6.59e-04 | 4.61e-03 | 3022 | 673 | 2349 |
| European | CIDI-MDD | Augmentation | Migraine | Income_quantitative | 0.811 | 0.776 | 0.849 | <2.2e-16 | <2.2e-16 | 3022 | 673 | 2349 |
| European | CIDI-MDD | Augmentation | Migraine | Education_levelCollege Undergraduate | 1.047 | 0.848 | 1.294 | 6.71e-01 | 1.00e+00 | 3022 | 673 | 2349 |
| European | CIDI-MDD | Augmentation | Migraine | Education_levelLess than a high school degree | 0.485 | 0.134 | 1.392 | 2.14e-01 | 1.00e+00 | 3022 | 673 | 2349 |
| European | CIDI-MDD | Augmentation | Migraine | Education_levelTwelve or GED | 1.540 | 1.079 | 2.186 | 1.65e-02 | 1.16e-01 | 3022 | 673 | 2349 |
| European | CIDI-MDD | Augmentation | Migraine | Migraine | 1.039 | 0.951 | 1.135 | 3.97e-01 | 1.00e+00 | 3022 | 673 | 2349 |
| European | CIDI-MDD | Augmentation | Insomnia | (Intercept) | 2.317 | 1.554 | 3.458 | 3.77e-05 | 2.64e-04 | 3022 | 673 | 2349 |
| European | CIDI-MDD | Augmentation | Insomnia | age_at_first_AD | 0.971 | 0.965 | 0.977 | <2.2e-16 | <2.2e-16 | 3022 | 673 | 2349 |
| European | CIDI-MDD | Augmentation | Insomnia | sex_at_birthMale | 1.440 | 1.174 | 1.763 | 4.30e-04 | 3.01e-03 | 3022 | 673 | 2349 |
| European | CIDI-MDD | Augmentation | Insomnia | Income_quantitative | 0.813 | 0.777 | 0.850 | <2.2e-16 | <2.2e-16 | 3022 | 673 | 2349 |
| European | CIDI-MDD | Augmentation | Insomnia | Education_levelCollege Undergraduate | 1.038 | 0.841 | 1.284 | 7.31e-01 | 1.00e+00 | 3022 | 673 | 2349 |
| European | CIDI-MDD | Augmentation | Insomnia | Education_levelLess than a high school degree | 0.468 | 0.129 | 1.348 | 1.93e-01 | 1.00e+00 | 3022 | 673 | 2349 |
| European | CIDI-MDD | Augmentation | Insomnia | Education_levelTwelve or GED | 1.517 | 1.062 | 2.155 | 2.09e-02 | 1.46e-01 | 3022 | 673 | 2349 |
| European | CIDI-MDD | Augmentation | Insomnia | Insomnia | 1.133 | 1.031 | 1.247 | 9.80e-03 | 6.86e-02 | 3022 | 673 | 2349 |
| European | CIDI-MDD | Augmentation | BMI | (Intercept) | 2.317 | 1.555 | 3.457 | 3.69e-05 | 2.58e-04 | 3022 | 673 | 2349 |
| European | CIDI-MDD | Augmentation | BMI | age_at_first_AD | 0.971 | 0.965 | 0.977 | <2.2e-16 | <2.2e-16 | 3022 | 673 | 2349 |
| European | CIDI-MDD | Augmentation | BMI | sex_at_birthMale | 1.424 | 1.161 | 1.742 | 6.34e-04 | 4.44e-03 | 3022 | 673 | 2349 |
| European | CIDI-MDD | Augmentation | BMI | Income_quantitative | 0.811 | 0.775 | 0.848 | <2.2e-16 | <2.2e-16 | 3022 | 673 | 2349 |
| European | CIDI-MDD | Augmentation | BMI | Education_levelCollege Undergraduate | 1.048 | 0.849 | 1.295 | 6.66e-01 | 1.00e+00 | 3022 | 673 | 2349 |
| European | CIDI-MDD | Augmentation | BMI | Education_levelLess than a high school degree | 0.489 | 0.135 | 1.401 | 2.18e-01 | 1.00e+00 | 3022 | 673 | 2349 |
| European | CIDI-MDD | Augmentation | BMI | Education_levelTwelve or GED | 1.541 | 1.079 | 2.189 | 1.65e-02 | 1.16e-01 | 3022 | 673 | 2349 |
| European | CIDI-MDD | Augmentation | BMI | BMI | 1.003 | 0.916 | 1.099 | 9.41e-01 | 1.00e+00 | 3022 | 673 | 2349 |
| European | CIDI-MDD | Augmentation | Bipolar Disorder | (Intercept) | 2.337 | 1.567 | 3.489 | 3.20e-05 | 2.24e-04 | 3022 | 673 | 2349 |
| European | CIDI-MDD | Augmentation | Bipolar Disorder | age_at_first_AD | 0.970 | 0.964 | 0.976 | <2.2e-16 | <2.2e-16 | 3022 | 673 | 2349 |
| European | CIDI-MDD | Augmentation | Bipolar Disorder | sex_at_birthMale | 1.427 | 1.163 | 1.747 | 6.10e-04 | 4.27e-03 | 3022 | 673 | 2349 |
| European | CIDI-MDD | Augmentation | Bipolar Disorder | Income_quantitative | 0.811 | 0.775 | 0.848 | <2.2e-16 | <2.2e-16 | 3022 | 673 | 2349 |
| European | CIDI-MDD | Augmentation | Bipolar Disorder | Education_levelCollege Undergraduate | 1.060 | 0.859 | 1.312 | 5.86e-01 | 1.00e+00 | 3022 | 673 | 2349 |
| European | CIDI-MDD | Augmentation | Bipolar Disorder | Education_levelLess than a high school degree | 0.535 | 0.148 | 1.539 | 2.83e-01 | 1.00e+00 | 3022 | 673 | 2349 |
| European | CIDI-MDD | Augmentation | Bipolar Disorder | Education_levelTwelve or GED | 1.590 | 1.112 | 2.261 | 1.04e-02 | 7.25e-02 | 3022 | 673 | 2349 |

| | | | | | | | | | | | | |
|---|---|---|---|---|---|---|---|---|---|---|---|---|
| European | CIDI-MDD | Augmentation | Bipolar Disorder | Bipolar Disorder | 1.181 | 1.072 | 1.301 | 7.47e-04 | 5.23e-03 | 3022 | 673 | 2349 |
| European | CIDI-MDD | Augmentation | Depression | (Intercept) | 2.311 | 1.546 | 3.457 | 4.51e-05 | 3.15e-04 | 3022 | 673 | 2349 |
| European | CIDI-MDD | Augmentation | Depression | age_at_first_AD | 0.970 | 0.964 | 0.976 | <2.2e-16 | <2.2e-16 | 3022 | 673 | 2349 |
| European | CIDI-MDD | Augmentation | Depression | sex_at_birthMale | 1.432 | 1.166 | 1.755 | 5.66e-04 | 3.96e-03 | 3022 | 673 | 2349 |
| European | CIDI-MDD | Augmentation | Depression | Income_quantitative | 0.815 | 0.779 | 0.853 | <2.2e-16 | <2.2e-16 | 3022 | 673 | 2349 |
| European | CIDI-MDD | Augmentation | Depression | Education_levelCollege Undergraduate | 1.020 | 0.826 | 1.263 | 8.53e-01 | 1.00e+00 | 3022 | 673 | 2349 |
| European | CIDI-MDD | Augmentation | Depression | Education_levelLess than a high school degree | 0.474 | 0.131 | 1.372 | 2.02e-01 | 1.00e+00 | 3022 | 673 | 2349 |
| European | CIDI-MDD | Augmentation | Depression | Education_levelTwelve or GED | 1.525 | 1.065 | 2.172 | 2.02e-02 | 1.42e-01 | 3022 | 673 | 2349 |
| European | CIDI-MDD | Augmentation | Depression | Depression | 1.341 | 1.218 | 1.477 | 2.37e-09 | 1.66e-08 | 3022 | 673 | 2349 |
| European | CIDI-MDD | Augmentation | Schizophrenia | (Intercept) | 2.310 | 1.548 | 3.449 | 4.14e-05 | 2.90e-04 | 3022 | 673 | 2349 |
| European | CIDI-MDD | Augmentation | Schizophrenia | age_at_first_AD | 0.971 | 0.965 | 0.977 | <2.2e-16 | <2.2e-16 | 3022 | 673 | 2349 |
| European | CIDI-MDD | Augmentation | Schizophrenia | sex_at_birthMale | 1.428 | 1.164 | 1.749 | 5.94e-04 | 4.16e-03 | 3022 | 673 | 2349 |
| European | CIDI-MDD | Augmentation | Schizophrenia | Income_quantitative | 0.811 | 0.775 | 0.849 | <2.2e-16 | <2.2e-16 | 3022 | 673 | 2349 |
| European | CIDI-MDD | Augmentation | Schizophrenia | Education_levelCollege Undergraduate | 1.052 | 0.852 | 1.301 | 6.42e-01 | 1.00e+00 | 3022 | 673 | 2349 |
| European | CIDI-MDD | Augmentation | Schizophrenia | Education_levelLess than a high school degree | 0.511 | 0.142 | 1.469 | 2.49e-01 | 1.00e+00 | 3022 | 673 | 2349 |
| European | CIDI-MDD | Augmentation | Schizophrenia | Education_levelTwelve or GED | 1.563 | 1.093 | 2.222 | 1.35e-02 | 9.47e-02 | 3022 | 673 | 2349 |
| European | CIDI-MDD | Augmentation | Schizophrenia | Schizophrenia | 1.186 | 1.082 | 1.301 | 2.69e-04 | 1.88e-03 | 3022 | 673 | 2349 |
| European | CIDI-noMDD | Augmentation | ADHD | (Intercept) | 0.529 | 0.158 | 1.669 | 2.88e-01 | 1.00e+00 | 2087 | 68 | 2019 |
| European | CIDI-noMDD | Augmentation | ADHD | age_at_first_AD | 0.962 | 0.946 | 0.979 | 8.29e-06 | 5.81e-05 | 2087 | 68 | 2019 |
| European | CIDI-noMDD | Augmentation | ADHD | sex_at_birthMale | 2.105 | 1.253 | 3.512 | 4.45e-03 | 3.11e-02 | 2087 | 68 | 2019 |
| European | CIDI-noMDD | Augmentation | ADHD | Income_quantitative | 0.783 | 0.685 | 0.893 | 2.90e-04 | 2.03e-03 | 2087 | 68 | 2019 |
| European | CIDI-noMDD | Augmentation | ADHD | Education_levelCollege Undergraduate | 1.104 | 0.608 | 2.065 | 7.50e-01 | 1.00e+00 | 2087 | 68 | 2019 |
| European | CIDI-noMDD | Augmentation | ADHD | Education_levelLess than a high school degree | 1.682 | 0.084 | 11.240 | 6.48e-01 | 1.00e+00 | 2087 | 68 | 2019 |
| European | CIDI-noMDD | Augmentation | ADHD | Education_levelTwelve or GED | 1.649 | 0.692 | 3.786 | 2.45e-01 | 1.00e+00 | 2087 | 68 | 2019 |
| European | CIDI-noMDD | Augmentation | ADHD | ADHD | 1.043 | 0.807 | 1.349 | 7.46e-01 | 1.00e+00 | 2087 | 68 | 2019 |
| European | CIDI-noMDD | Augmentation | Migraine | (Intercept) | 0.539 | 0.162 | 1.702 | 3.03e-01 | 1.00e+00 | 2087 | 68 | 2019 |
| European | CIDI-noMDD | Augmentation | Migraine | age_at_first_AD | 0.962 | 0.946 | 0.978 | 7.14e-06 | 5.00e-05 | 2087 | 68 | 2019 |
| European | CIDI-noMDD | Augmentation | Migraine | sex_at_birthMale | 2.111 | 1.257 | 3.520 | 4.28e-03 | 2.99e-02 | 2087 | 68 | 2019 |
| European | CIDI-noMDD | Augmentation | Migraine | Income_quantitative | 0.779 | 0.682 | 0.889 | 2.20e-04 | 1.54e-03 | 2087 | 68 | 2019 |
| European | CIDI-noMDD | Augmentation | Migraine | Education_levelCollege Undergraduate | 1.124 | 0.619 | 2.103 | 7.05e-01 | 1.00e+00 | 2087 | 68 | 2019 |
| European | CIDI-noMDD | Augmentation | Migraine | Education_levelLess than a high school degree | 1.740 | 0.087 | 11.610 | 6.26e-01 | 1.00e+00 | 2087 | 68 | 2019 |
| European | CIDI-noMDD | Augmentation | Migraine | Education_levelTwelve or GED | 1.680 | 0.706 | 3.854 | 2.27e-01 | 1.00e+00 | 2087 | 68 | 2019 |
| European | CIDI-noMDD | Augmentation | Migraine | Migraine | 0.905 | 0.711 | 1.150 | 4.14e-01 | 1.00e+00 | 2087 | 68 | 2019 |
| European | CIDI-noMDD | Augmentation | Insomnia | (Intercept) | 0.529 | 0.158 | 1.671 | 2.88e-01 | 1.00e+00 | 2087 | 68 | 2019 |
| European | CIDI-noMDD | Augmentation | Insomnia | age_at_first_AD | 0.962 | 0.946 | 0.979 | 8.21e-06 | 5.75e-05 | 2087 | 68 | 2019 |
| European | CIDI-noMDD | Augmentation | Insomnia | sex_at_birthMale | 2.107 | 1.254 | 3.516 | 4.42e-03 | 3.09e-02 | 2087 | 68 | 2019 |
| European | CIDI-noMDD | Augmentation | Insomnia | Income_quantitative | 0.783 | 0.685 | 0.893 | 2.90e-04 | 2.03e-03 | 2087 | 68 | 2019 |
| European | CIDI-noMDD | Augmentation | Insomnia | Education_levelCollege Undergraduate | 1.104 | 0.608 | 2.066 | 7.50e-01 | 1.00e+00 | 2087 | 68 | 2019 |
| European | CIDI-noMDD | Augmentation | Insomnia | Education_levelLess than a high school degree | 1.703 | 0.085 | 11.310 | 6.39e-01 | 1.00e+00 | 2087 | 68 | 2019 |
| European | CIDI-noMDD | Augmentation | Insomnia | Education_levelTwelve or GED | 1.652 | 0.693 | 3.792 | 2.43e-01 | 1.00e+00 | 2087 | 68 | 2019 |
| European | CIDI-noMDD | Augmentation | Insomnia | Insomnia | 1.047 | 0.804 | 1.362 | 7.31e-01 | 1.00e+00 | 2087 | 68 | 2019 |
| European | CIDI-noMDD | Augmentation | BMI | (Intercept) | 0.530 | 0.158 | 1.674 | 2.90e-01 | 1.00e+00 | 2087 | 68 | 2019 |
| European | CIDI-noMDD | Augmentation | BMI | age_at_first_AD | 0.962 | 0.946 | 0.979 | 8.10e-06 | 5.67e-05 | 2087 | 68 | 2019 |
| European | CIDI-noMDD | Augmentation | BMI | sex_at_birthMale | 2.101 | 1.251 | 3.504 | 4.52e-03 | 3.17e-02 | 2087 | 68 | 2019 |
| European | CIDI-noMDD | Augmentation | BMI | Income_quantitative | 0.783 | 0.685 | 0.893 | 2.93e-04 | 2.05e-03 | 2087 | 68 | 2019 |
| European | CIDI-noMDD | Augmentation | BMI | Education_levelCollege Undergraduate | 1.108 | 0.611 | 2.072 | 7.40e-01 | 1.00e+00 | 2087 | 68 | 2019 |
| European | CIDI-noMDD | Augmentation | BMI | Education_levelLess than a high school degree | 1.722 | 0.086 | 11.390 | 6.32e-01 | 1.00e+00 | 2087 | 68 | 2019 |
| European | CIDI-noMDD | Augmentation | BMI | Education_levelTwelve or GED | 1.665 | 0.700 | 3.815 | 2.35e-01 | 1.00e+00 | 2087 | 68 | 2019 |
| European | CIDI-noMDD | Augmentation | BMI | BMI | 1.026 | 0.804 | 1.307 | 8.35e-01 | 1.00e+00 | 2087 | 68 | 2019 |
| European | CIDI-noMDD | Augmentation | Bipolar Disorder | (Intercept) | 0.532 | 0.159 | 1.683 | 2.93e-01 | 1.00e+00 | 2087 | 68 | 2019 |
| European | CIDI-noMDD | Augmentation | Bipolar Disorder | age_at_first_AD | 0.962 | 0.946 | 0.979 | 8.14e-06 | 5.70e-05 | 2087 | 68 | 2019 |
| European | CIDI-noMDD | Augmentation | Bipolar Disorder | sex_at_birthMale | 2.097 | 1.249 | 3.496 | 4.61e-03 | 3.23e-02 | 2087 | 68 | 2019 |
| European | CIDI-noMDD | Augmentation | Bipolar Disorder | Income_quantitative | 0.782 | 0.684 | 0.892 | 2.75e-04 | 1.92e-03 | 2087 | 68 | 2019 |
| European | CIDI-noMDD | Augmentation | Bipolar Disorder | Education_levelCollege Undergraduate | 1.110 | 0.611 | 2.075 | 7.37e-01 | 1.00e+00 | 2087 | 68 | 2019 |
| European | CIDI-noMDD | Augmentation | Bipolar Disorder | Education_levelLess than a high school degree | 1.731 | 0.086 | 11.470 | 6.29e-01 | 1.00e+00 | 2087 | 68 | 2019 |

| | | | | | | | | | | | | |
|---|---|---|---|---|---|---|---|---|---|---|---|---|
| European | CIDI-noMDD | Augmentation | Bipolar Disorder | Education_levelTwelve or GED | 1.668 | 0.700 | 3.828 | 2.34e-01 | 1.00e+00 | 2087 | 68 | 2019 |
| European | CIDI-noMDD | Augmentation | Bipolar Disorder | Bipolar Disorder | 1.009 | 0.775 | 1.313 | 9.46e-01 | 1.00e+00 | 2087 | 68 | 2019 |
| European | CIDI-noMDD | Augmentation | Depression | (Intercept) | 0.498 | 0.148 | 1.586 | 2.49e-01 | 1.00e+00 | 2087 | 68 | 2019 |
| European | CIDI-noMDD | Augmentation | Depression | age_at_first_AD | 0.963 | 0.946 | 0.979 | 1.14e-05 | 8.00e-05 | 2087 | 68 | 2019 |
| European | CIDI-noMDD | Augmentation | Depression | sex_at_birthMale | 2.068 | 1.230 | 3.451 | 5.58e-03 | 3.91e-02 | 2087 | 68 | 2019 |
| European | CIDI-noMDD | Augmentation | Depression | Income_quantitative | 0.788 | 0.690 | 0.899 | 4.25e-04 | 2.98e-03 | 2087 | 68 | 2019 |
| European | CIDI-noMDD | Augmentation | Depression | Education_levelCollege Undergraduate | 1.082 | 0.596 | 2.025 | 7.99e-01 | 1.00e+00 | 2087 | 68 | 2019 |
| European | CIDI-noMDD | Augmentation | Depression | Education_levelLess than a high school degree | 1.331 | 0.065 | 9.354 | 8.05e-01 | 1.00e+00 | 2087 | 68 | 2019 |
| European | CIDI-noMDD | Augmentation | Depression | Education_levelTwelve or GED | 1.547 | 0.646 | 3.563 | 3.13e-01 | 1.00e+00 | 2087 | 68 | 2019 |
| European | CIDI-noMDD | Augmentation | Depression | Depression | 1.273 | 0.981 | 1.651 | 6.89e-02 | 4.82e-01 | 2087 | 68 | 2019 |
| European | CIDI-noMDD | Augmentation | Schizophrenia | (Intercept) | 0.527 | 0.157 | 1.664 | 2.85e-01 | 1.00e+00 | 2087 | 68 | 2019 |
| European | CIDI-noMDD | Augmentation | Schizophrenia | age_at_first_AD | 0.962 | 0.946 | 0.979 | 8.84e-06 | 6.19e-05 | 2087 | 68 | 2019 |
| European | CIDI-noMDD | Augmentation | Schizophrenia | sex_at_birthMale | 2.106 | 1.254 | 3.512 | 4.41e-03 | 3.09e-02 | 2087 | 68 | 2019 |
| European | CIDI-noMDD | Augmentation | Schizophrenia | Income_quantitative | 0.782 | 0.684 | 0.892 | 2.60e-04 | 1.82e-03 | 2087 | 68 | 2019 |
| European | CIDI-noMDD | Augmentation | Schizophrenia | Education_levelCollege Undergraduate | 1.107 | 0.610 | 2.070 | 7.43e-01 | 1.00e+00 | 2087 | 68 | 2019 |
| European | CIDI-noMDD | Augmentation | Schizophrenia | Education_levelLess than a high school degree | 1.731 | 0.086 | 11.500 | 6.29e-01 | 1.00e+00 | 2087 | 68 | 2019 |
| European | CIDI-noMDD | Augmentation | Schizophrenia | Education_levelTwelve or GED | 1.668 | 0.701 | 3.821 | 2.33e-01 | 1.00e+00 | 2087 | 68 | 2019 |
| European | CIDI-noMDD | Augmentation | Schizophrenia | Schizophrenia | 1.065 | 0.828 | 1.372 | 6.24e-01 | 1.00e+00 | 2087 | 68 | 2019 |
| African | Full Cohort | Augmentation | ADHD | (Intercept) | 1.213 | 0.792 | 1.851 | 3.72e-01 | 1.00e+00 | 5138 | 974 | 4164 |
| African | Full Cohort | Augmentation | ADHD | age_at_first_AD | 0.975 | 0.970 | 0.980 | <2.2e-16 | <2.2e-16 | 5138 | 974 | 4164 |
| African | Full Cohort | Augmentation | ADHD | sex_at_birthMale | 1.485 | 1.270 | 1.734 | 6.58e-07 | 4.60e-06 | 5138 | 974 | 4164 |
| African | Full Cohort | Augmentation | ADHD | Income_quantitative | 0.780 | 0.734 | 0.828 | 1.18e-15 | 8.26e-15 | 5138 | 974 | 4164 |
| African | Full Cohort | Augmentation | ADHD | Education_levelCollege Undergraduate | 0.812 | 0.605 | 1.100 | 1.71e-01 | 1.00e+00 | 5138 | 974 | 4164 |
| African | Full Cohort | Augmentation | ADHD | Education_levelLess than a high school degree | 1.141 | 0.803 | 1.634 | 4.65e-01 | 1.00e+00 | 5138 | 974 | 4164 |
| African | Full Cohort | Augmentation | ADHD | Education_levelTwelve or GED | 0.899 | 0.655 | 1.246 | 5.16e-01 | 1.00e+00 | 5138 | 974 | 4164 |
| African | Full Cohort | Augmentation | ADHD | ADHD | 1.061 | 0.958 | 1.176 | 2.53e-01 | 1.00e+00 | 5138 | 974 | 4164 |
| African | Full Cohort | Augmentation | Migraine | (Intercept) | 1.214 | 0.792 | 1.852 | 3.71e-01 | 1.00e+00 | 5138 | 974 | 4164 |
| African | Full Cohort | Augmentation | Migraine | age_at_first_AD | 0.975 | 0.970 | 0.980 | <2.2e-16 | <2.2e-16 | 5138 | 974 | 4164 |
| African | Full Cohort | Augmentation | Migraine | sex_at_birthMale | 1.487 | 1.271 | 1.736 | 6.05e-07 | 4.24e-06 | 5138 | 974 | 4164 |
| African | Full Cohort | Augmentation | Migraine | Income_quantitative | 0.779 | 0.733 | 0.827 | 8.92e-16 | 6.24e-15 | 5138 | 974 | 4164 |
| African | Full Cohort | Augmentation | Migraine | Education_levelCollege Undergraduate | 0.815 | 0.607 | 1.105 | 1.79e-01 | 1.00e+00 | 5138 | 974 | 4164 |
| African | Full Cohort | Augmentation | Migraine | Education_levelLess than a high school degree | 1.151 | 0.810 | 1.647 | 4.36e-01 | 1.00e+00 | 5138 | 974 | 4164 |
| African | Full Cohort | Augmentation | Migraine | Education_levelTwelve or GED | 0.904 | 0.658 | 1.252 | 5.37e-01 | 1.00e+00 | 5138 | 974 | 4164 |
| African | Full Cohort | Augmentation | Migraine | Migraine | 0.966 | 0.885 | 1.055 | 4.45e-01 | 1.00e+00 | 5138 | 974 | 4164 |
| African | Full Cohort | Augmentation | Insomnia | (Intercept) | 1.221 | 0.796 | 1.864 | 3.58e-01 | 1.00e+00 | 5138 | 974 | 4164 |
| African | Full Cohort | Augmentation | Insomnia | age_at_first_AD | 0.975 | 0.970 | 0.980 | <2.2e-16 | <2.2e-16 | 5138 | 974 | 4164 |
| African | Full Cohort | Augmentation | Insomnia | sex_at_birthMale | 1.483 | 1.268 | 1.732 | 7.42e-07 | 5.20e-06 | 5138 | 974 | 4164 |
| African | Full Cohort | Augmentation | Insomnia | Income_quantitative | 0.780 | 0.733 | 0.828 | 1.13e-15 | 7.92e-15 | 5138 | 974 | 4164 |
| African | Full Cohort | Augmentation | Insomnia | Education_levelCollege Undergraduate | 0.809 | 0.603 | 1.097 | 1.65e-01 | 1.00e+00 | 5138 | 974 | 4164 |
| African | Full Cohort | Augmentation | Insomnia | Education_levelLess than a high school degree | 1.133 | 0.796 | 1.622 | 4.92e-01 | 1.00e+00 | 5138 | 974 | 4164 |
| African | Full Cohort | Augmentation | Insomnia | Education_levelTwelve or GED | 0.899 | 0.655 | 1.246 | 5.18e-01 | 1.00e+00 | 5138 | 974 | 4164 |
| African | Full Cohort | Augmentation | Insomnia | Insomnia | 1.170 | 1.052 | 1.300 | 3.66e-03 | 2.56e-02 | 5138 | 974 | 4164 |
| African | Full Cohort | Augmentation | BMI | (Intercept) | 1.213 | 0.792 | 1.851 | 3.72e-01 | 1.00e+00 | 5138 | 974 | 4164 |
| African | Full Cohort | Augmentation | BMI | age_at_first_AD | 0.975 | 0.970 | 0.980 | <2.2e-16 | <2.2e-16 | 5138 | 974 | 4164 |
| African | Full Cohort | Augmentation | BMI | sex_at_birthMale | 1.488 | 1.272 | 1.738 | 6.00e-07 | 4.20e-06 | 5138 | 974 | 4164 |
| African | Full Cohort | Augmentation | BMI | Income_quantitative | 0.780 | 0.733 | 0.828 | 9.75e-16 | 6.83e-15 | 5138 | 974 | 4164 |
| African | Full Cohort | Augmentation | BMI | Education_levelCollege Undergraduate | 0.814 | 0.607 | 1.104 | 1.78e-01 | 1.00e+00 | 5138 | 974 | 4164 |
| African | Full Cohort | Augmentation | BMI | Education_levelLess than a high school degree | 1.149 | 0.808 | 1.644 | 4.44e-01 | 1.00e+00 | 5138 | 974 | 4164 |
| African | Full Cohort | Augmentation | BMI | Education_levelTwelve or GED | 0.902 | 0.657 | 1.249 | 5.28e-01 | 1.00e+00 | 5138 | 974 | 4164 |
| African | Full Cohort | Augmentation | BMI | BMI | 1.018 | 0.926 | 1.119 | 7.12e-01 | 1.00e+00 | 5138 | 974 | 4164 |
| African | Full Cohort | Augmentation | Bipolar Disorder | (Intercept) | 1.207 | 0.788 | 1.842 | 3.85e-01 | 1.00e+00 | 5138 | 974 | 4164 |
| African | Full Cohort | Augmentation | Bipolar Disorder | age_at_first_AD | 0.975 | 0.970 | 0.980 | <2.2e-16 | <2.2e-16 | 5138 | 974 | 4164 |
| African | Full Cohort | Augmentation | Bipolar Disorder | sex_at_birthMale | 1.480 | 1.266 | 1.729 | 8.27e-07 | 5.79e-06 | 5138 | 974 | 4164 |
| African | Full Cohort | Augmentation | Bipolar Disorder | Income_quantitative | 0.780 | 0.733 | 0.828 | 1.25e-15 | 8.78e-15 | 5138 | 974 | 4164 |
| African | Full Cohort | Augmentation | Bipolar Disorder | Education_levelCollege Undergraduate | 0.815 | 0.608 | 1.106 | 1.81e-01 | 1.00e+00 | 5138 | 974 | 4164 |

| | | | | | | | | | | | | |
|---|---|---|---|---|---|---|---|---|---|---|---|---|
| African | Full Cohort | Augmentation | Bipolar Disorder | Education_levelLess than a high school degree | 1.153 | 0.811 | 1.651 | 4.31e-01 | 1.00e+00 | 5138 | 974 | 4164 |
| African | Full Cohort | Augmentation | Bipolar Disorder | Education_levelTwelve or GED | 0.911 | 0.663 | 1.261 | 5.67e-01 | 1.00e+00 | 5138 | 974 | 4164 |
| African | Full Cohort | Augmentation | Bipolar Disorder | Bipolar Disorder | 1.176 | 1.059 | 1.306 | 2.37e-03 | 1.66e-02 | 5138 | 974 | 4164 |
| African | Full Cohort | Augmentation | Depression | (Intercept) | 1.210 | 0.789 | 1.848 | 3.80e-01 | 1.00e+00 | 5138 | 974 | 4164 |
| African | Full Cohort | Augmentation | Depression | age_at_first_AD | 0.975 | 0.970 | 0.980 | <2.2e-16 | <2.2e-16 | 5138 | 974 | 4164 |
| African | Full Cohort | Augmentation | Depression | sex_at_birthMale | 1.489 | 1.273 | 1.739 | 5.71e-07 | 4.00e-06 | 5138 | 974 | 4164 |
| African | Full Cohort | Augmentation | Depression | Income_quantitative | 0.782 | 0.736 | 0.831 | 2.30e-15 | 1.61e-14 | 5138 | 974 | 4164 |
| African | Full Cohort | Augmentation | Depression | Education_levelCollege Undergraduate | 0.815 | 0.607 | 1.106 | 1.81e-01 | 1.00e+00 | 5138 | 974 | 4164 |
| African | Full Cohort | Augmentation | Depression | Education_levelLess than a high school degree | 1.146 | 0.806 | 1.641 | 4.51e-01 | 1.00e+00 | 5138 | 974 | 4164 |
| African | Full Cohort | Augmentation | Depression | Education_levelTwelve or GED | 0.905 | 0.659 | 1.254 | 5.42e-01 | 1.00e+00 | 5138 | 974 | 4164 |
| African | Full Cohort | Augmentation | Depression | Depression | 1.222 | 1.104 | 1.353 | 1.13e-04 | 7.94e-04 | 5138 | 974 | 4164 |
| African | Full Cohort | Augmentation | Schizophrenia | (Intercept) | 1.210 | 0.789 | 1.846 | 3.79e-01 | 1.00e+00 | 5138 | 974 | 4164 |
| African | Full Cohort | Augmentation | Schizophrenia | age_at_first_AD | 0.975 | 0.970 | 0.980 | <2.2e-16 | <2.2e-16 | 5138 | 974 | 4164 |
| African | Full Cohort | Augmentation | Schizophrenia | sex_at_birthMale | 1.486 | 1.271 | 1.735 | 6.34e-07 | 4.44e-06 | 5138 | 974 | 4164 |
| African | Full Cohort | Augmentation | Schizophrenia | Income_quantitative | 0.780 | 0.733 | 0.828 | 1.18e-15 | 8.26e-15 | 5138 | 974 | 4164 |
| African | Full Cohort | Augmentation | Schizophrenia | Education_levelCollege Undergraduate | 0.816 | 0.608 | 1.106 | 1.82e-01 | 1.00e+00 | 5138 | 974 | 4164 |
| African | Full Cohort | Augmentation | Schizophrenia | Education_levelLess than a high school degree | 1.151 | 0.810 | 1.647 | 4.36e-01 | 1.00e+00 | 5138 | 974 | 4164 |
| African | Full Cohort | Augmentation | Schizophrenia | Education_levelTwelve or GED | 0.905 | 0.659 | 1.253 | 5.41e-01 | 1.00e+00 | 5138 | 974 | 4164 |
| African | Full Cohort | Augmentation | Schizophrenia | Schizophrenia | 1.033 | 0.943 | 1.132 | 4.83e-01 | 1.00e+00 | 5138 | 974 | 4164 |
| African | Recorded Depression | Augmentation | ADHD | (Intercept) | 1.514 | 0.889 | 2.574 | 1.26e-01 | 8.81e-01 | 2402 | 674 | 1728 |
| African | Recorded Depression | Augmentation | ADHD | age_at_first_AD | 0.983 | 0.976 | 0.990 | 9.50e-07 | 6.65e-06 | 2402 | 674 | 1728 |
| African | Recorded Depression | Augmentation | ADHD | sex_at_birthMale | 1.505 | 1.225 | 1.846 | 9.17e-05 | 6.42e-04 | 2402 | 674 | 1728 |
| African | Recorded Depression | Augmentation | ADHD | Income_quantitative | 0.810 | 0.751 | 0.870 | 1.77e-08 | 1.24e-07 | 2402 | 674 | 1728 |
| African | Recorded Depression | Augmentation | ADHD | Education_levelCollege Undergraduate | 0.683 | 0.477 | 0.987 | 3.96e-02 | 2.77e-01 | 2402 | 674 | 1728 |
| African | Recorded Depression | Augmentation | ADHD | Education_levelLess than a high school degree | 1.000 | 0.642 | 1.566 | 1.00e+00 | 1.00e+00 | 2402 | 674 | 1728 |
| African | Recorded Depression | Augmentation | ADHD | Education_levelTwelve or GED | 0.740 | 0.500 | 1.101 | 1.33e-01 | 9.33e-01 | 2402 | 674 | 1728 |
| African | Recorded Depression | Augmentation | ADHD | ADHD | 1.023 | 0.898 | 1.166 | 7.30e-01 | 1.00e+00 | 2402 | 674 | 1728 |
| African | Recorded Depression | Augmentation | Migraine | (Intercept) | 1.510 | 0.887 | 2.566 | 1.28e-01 | 8.98e-01 | 2402 | 674 | 1728 |
| African | Recorded Depression | Augmentation | Migraine | age_at_first_AD | 0.983 | 0.976 | 0.990 | 9.38e-07 | 6.56e-06 | 2402 | 674 | 1728 |
| African | Recorded Depression | Augmentation | Migraine | sex_at_birthMale | 1.506 | 1.226 | 1.848 | 8.93e-05 | 6.25e-04 | 2402 | 674 | 1728 |
| African | Recorded Depression | Augmentation | Migraine | Income_quantitative | 0.810 | 0.751 | 0.870 | 1.81e-08 | 1.26e-07 | 2402 | 674 | 1728 |
| African | Recorded Depression | Augmentation | Migraine | Education_levelCollege Undergraduate | 0.685 | 0.479 | 0.990 | 4.09e-02 | 2.87e-01 | 2402 | 674 | 1728 |
| African | Recorded Depression | Augmentation | Migraine | Education_levelLess than a high school degree | 1.003 | 0.644 | 1.571 | 9.88e-01 | 1.00e+00 | 2402 | 674 | 1728 |
| African | Recorded Depression | Augmentation | Migraine | Education_levelTwelve or GED | 0.742 | 0.503 | 1.104 | 1.37e-01 | 9.62e-01 | 2402 | 674 | 1728 |
| African | Recorded Depression | Augmentation | Migraine | Migraine | 0.997 | 0.891 | 1.117 | 9.63e-01 | 1.00e+00 | 2402 | 674 | 1728 |
| African | Recorded Depression | Augmentation | Insomnia | (Intercept) | 1.523 | 0.894 | 2.590 | 1.21e-01 | 8.46e-01 | 2402 | 674 | 1728 |
| African | Recorded Depression | Augmentation | Insomnia | age_at_first_AD | 0.983 | 0.976 | 0.990 | 8.43e-07 | 5.90e-06 | 2402 | 674 | 1728 |
| African | Recorded Depression | Augmentation | Insomnia | sex_at_birthMale | 1.504 | 1.224 | 1.845 | 9.46e-05 | 6.62e-04 | 2402 | 674 | 1728 |
| African | Recorded Depression | Augmentation | Insomnia | Income_quantitative | 0.810 | 0.751 | 0.870 | 1.73e-08 | 1.21e-07 | 2402 | 674 | 1728 |
| African | Recorded Depression | Augmentation | Insomnia | Education_levelCollege Undergraduate | 0.683 | 0.477 | 0.986 | 3.88e-02 | 2.72e-01 | 2402 | 674 | 1728 |
| African | Recorded Depression | Augmentation | Insomnia | Education_levelLess than a high school degree | 0.993 | 0.637 | 1.555 | 9.76e-01 | 1.00e+00 | 2402 | 674 | 1728 |
| African | Recorded Depression | Augmentation | Insomnia | Education_levelTwelve or GED | 0.739 | 0.500 | 1.099 | 1.31e-01 | 9.19e-01 | 2402 | 674 | 1728 |
| African | Recorded Depression | Augmentation | Insomnia | Insomnia | 1.099 | 0.961 | 1.258 | 1.69e-01 | 1.00e+00 | 2402 | 674 | 1728 |
| African | Recorded Depression | Augmentation | BMI | (Intercept) | 1.508 | 0.886 | 2.563 | 1.29e-01 | 9.05e-01 | 2402 | 674 | 1728 |
| African | Recorded Depression | Augmentation | BMI | age_at_first_AD | 0.983 | 0.976 | 0.990 | 8.53e-07 | 5.97e-06 | 2402 | 674 | 1728 |
| African | Recorded Depression | Augmentation | BMI | sex_at_birthMale | 1.493 | 1.215 | 1.832 | 1.31e-04 | 9.15e-04 | 2402 | 674 | 1728 |
| African | Recorded Depression | Augmentation | BMI | Income_quantitative | 0.809 | 0.751 | 0.870 | 1.56e-08 | 1.09e-07 | 2402 | 674 | 1728 |
| African | Recorded Depression | Augmentation | BMI | Education_levelCollege Undergraduate | 0.691 | 0.483 | 0.998 | 4.59e-02 | 3.21e-01 | 2402 | 674 | 1728 |
| African | Recorded Depression | Augmentation | BMI | Education_levelLess than a high school degree | 1.012 | 0.649 | 1.585 | 9.57e-01 | 1.00e+00 | 2402 | 674 | 1728 |
| African | Recorded Depression | Augmentation | BMI | Education_levelTwelve or GED | 0.750 | 0.507 | 1.116 | 1.52e-01 | 1.00e+00 | 2402 | 674 | 1728 |
| African | Recorded Depression | Augmentation | BMI | BMI | 0.929 | 0.823 | 1.048 | 2.31e-01 | 1.00e+00 | 2402 | 674 | 1728 |
| African | Recorded Depression | Augmentation | Bipolar Disorder | (Intercept) | 1.511 | 0.887 | 2.569 | 1.28e-01 | 8.93e-01 | 2402 | 674 | 1728 |
| African | Recorded Depression | Augmentation | Bipolar Disorder | age_at_first_AD | 0.983 | 0.976 | 0.990 | 7.83e-07 | 5.48e-06 | 2402 | 674 | 1728 |
| African | Recorded Depression | Augmentation | Bipolar Disorder | sex_at_birthMale | 1.487 | 1.210 | 1.825 | 1.51e-04 | 1.06e-03 | 2402 | 674 | 1728 |
| African | Recorded Depression | Augmentation | Bipolar Disorder | Income_quantitative | 0.810 | 0.752 | 0.871 | 2.08e-08 | 1.45e-07 | 2402 | 674 | 1728 |

| | | | | | | | | | | | | |
|---|---|---|---|---|---|---|---|---|---|---|---|---|
| African | Recorded Depression | Augmentation | Bipolar Disorder | Education_levelCollege Undergraduate | 0.689 | 0.482 | 0.995 | 4.41e-02 | 3.09e-01 | 2402 | 674 | 1728 |
| African | Recorded Depression | Augmentation | Bipolar Disorder | Education_levelLess than a high school degree | 1.005 | 0.645 | 1.574 | 9.83e-01 | 1.00e+00 | 2402 | 674 | 1728 |
| African | Recorded Depression | Augmentation | Bipolar Disorder | Education_levelTwelve or GED | 0.750 | 0.507 | 1.116 | 1.51e-01 | 1.00e+00 | 2402 | 674 | 1728 |
| African | Recorded Depression | Augmentation | Bipolar Disorder | Bipolar Disorder | 1.180 | 1.031 | 1.350 | 1.60e-02 | 1.12e-01 | 2402 | 674 | 1728 |
| African | Recorded Depression | Augmentation | Depression | (Intercept) | 1.522 | 0.894 | 2.589 | 1.21e-01 | 8.47e-01 | 2402 | 674 | 1728 |
| African | Recorded Depression | Augmentation | Depression | age_at_first_AD | 0.983 | 0.976 | 0.990 | 6.68e-07 | 4.68e-06 | 2402 | 674 | 1728 |
| African | Recorded Depression | Augmentation | Depression | sex_at_birthMale | 1.505 | 1.225 | 1.847 | 9.31e-05 | 6.52e-04 | 2402 | 674 | 1728 |
| African | Recorded Depression | Augmentation | Depression | Income_quantitative | 0.811 | 0.753 | 0.872 | 2.35e-08 | 1.65e-07 | 2402 | 674 | 1728 |
| African | Recorded Depression | Augmentation | Depression | Education_levelCollege Undergraduate | 0.685 | 0.478 | 0.989 | 4.07e-02 | 2.85e-01 | 2402 | 674 | 1728 |
| African | Recorded Depression | Augmentation | Depression | Education_levelLess than a high school degree | 0.995 | 0.638 | 1.558 | 9.82e-01 | 1.00e+00 | 2402 | 674 | 1728 |
| African | Recorded Depression | Augmentation | Depression | Education_levelTwelve or GED | 0.739 | 0.500 | 1.099 | 1.31e-01 | 9.19e-01 | 2402 | 674 | 1728 |
| African | Recorded Depression | Augmentation | Depression | Depression | 1.184 | 1.038 | 1.353 | 1.23e-02 | 8.61e-02 | 2402 | 674 | 1728 |
| African | Recorded Depression | Augmentation | Schizophrenia | (Intercept) | 1.501 | 0.882 | 2.552 | 1.34e-01 | 9.35e-01 | 2402 | 674 | 1728 |
| African | Recorded Depression | Augmentation | Schizophrenia | age_at_first_AD | 0.983 | 0.976 | 0.990 | 9.57e-07 | 6.70e-06 | 2402 | 674 | 1728 |
| African | Recorded Depression | Augmentation | Schizophrenia | sex_at_birthMale | 1.507 | 1.227 | 1.848 | 8.77e-05 | 6.14e-04 | 2402 | 674 | 1728 |
| African | Recorded Depression | Augmentation | Schizophrenia | Income_quantitative | 0.810 | 0.752 | 0.871 | 2.07e-08 | 1.45e-07 | 2402 | 674 | 1728 |
| African | Recorded Depression | Augmentation | Schizophrenia | Education_levelCollege Undergraduate | 0.688 | 0.481 | 0.993 | 4.29e-02 | 3.01e-01 | 2402 | 674 | 1728 |
| African | Recorded Depression | Augmentation | Schizophrenia | Education_levelLess than a high school degree | 1.006 | 0.646 | 1.575 | 9.78e-01 | 1.00e+00 | 2402 | 674 | 1728 |
| African | Recorded Depression | Augmentation | Schizophrenia | Education_levelTwelve or GED | 0.744 | 0.504 | 1.107 | 1.41e-01 | 9.87e-01 | 2402 | 674 | 1728 |
| African | Recorded Depression | Augmentation | Schizophrenia | Schizophrenia | 1.039 | 0.927 | 1.165 | 5.11e-01 | 1.00e+00 | 2402 | 674 | 1728 |
| African | No Recorded Depression | Augmentation | ADHD | (Intercept) | 0.598 | 0.264 | 1.303 | 2.06e-01 | 1.00e+00 | 2736 | 300 | 2436 |
| African | No Recorded Depression | Augmentation | ADHD | age_at_first_AD | 0.971 | 0.961 | 0.980 | 2.56e-10 | 1.79e-09 | 2736 | 300 | 2436 |
| African | No Recorded Depression | Augmentation | ADHD | sex_at_birthMale | 1.719 | 1.327 | 2.223 | 3.77e-05 | 2.64e-04 | 2736 | 300 | 2436 |
| African | No Recorded Depression | Augmentation | ADHD | Income_quantitative | 0.731 | 0.647 | 0.820 | 2.23e-07 | 1.56e-06 | 2736 | 300 | 2436 |
| African | No Recorded Depression | Augmentation | ADHD | Education_levelCollege Undergraduate | 1.124 | 0.633 | 2.137 | 7.04e-01 | 1.00e+00 | 2736 | 300 | 2436 |
| African | No Recorded Depression | Augmentation | ADHD | Education_levelLess than a high school degree | 1.710 | 0.891 | 3.461 | 1.19e-01 | 8.35e-01 | 2736 | 300 | 2436 |
| African | No Recorded Depression | Augmentation | ADHD | Education_levelTwelve or GED | 1.354 | 0.739 | 2.637 | 3.48e-01 | 1.00e+00 | 2736 | 300 | 2436 |
| African | No Recorded Depression | Augmentation | ADHD | ADHD | 1.160 | 0.972 | 1.384 | 9.93e-02 | 6.95e-01 | 2736 | 300 | 2436 |
| African | No Recorded Depression | Augmentation | Migraine | (Intercept) | 0.606 | 0.267 | 1.321 | 2.18e-01 | 1.00e+00 | 2736 | 300 | 2436 |
| African | No Recorded Depression | Augmentation | Migraine | age_at_first_AD | 0.970 | 0.961 | 0.979 | 1.79e-10 | 1.25e-09 | 2736 | 300 | 2436 |
| African | No Recorded Depression | Augmentation | Migraine | sex_at_birthMale | 1.718 | 1.326 | 2.220 | 3.89e-05 | 2.72e-04 | 2736 | 300 | 2436 |
| African | No Recorded Depression | Augmentation | Migraine | Income_quantitative | 0.728 | 0.644 | 0.816 | 1.58e-07 | 1.10e-06 | 2736 | 300 | 2436 |
| African | No Recorded Depression | Augmentation | Migraine | Education_levelCollege Undergraduate | 1.134 | 0.638 | 2.155 | 6.83e-01 | 1.00e+00 | 2736 | 300 | 2436 |
| African | No Recorded Depression | Augmentation | Migraine | Education_levelLess than a high school degree | 1.757 | 0.915 | 3.555 | 1.02e-01 | 7.11e-01 | 2736 | 300 | 2436 |
| African | No Recorded Depression | Augmentation | Migraine | Education_levelTwelve or GED | 1.362 | 0.744 | 2.652 | 3.38e-01 | 1.00e+00 | 2736 | 300 | 2436 |
| African | No Recorded Depression | Augmentation | Migraine | Migraine | 0.901 | 0.774 | 1.049 | 1.82e-01 | 1.00e+00 | 2736 | 300 | 2436 |
| African | No Recorded Depression | Augmentation | Insomnia | (Intercept) | 0.601 | 0.265 | 1.308 | 2.09e-01 | 1.00e+00 | 2736 | 300 | 2436 |
| African | No Recorded Depression | Augmentation | Insomnia | age_at_first_AD | 0.970 | 0.961 | 0.979 | 2.33e-10 | 1.63e-09 | 2736 | 300 | 2436 |
| African | No Recorded Depression | Augmentation | Insomnia | sex_at_birthMale | 1.709 | 1.319 | 2.210 | 4.62e-05 | 3.24e-04 | 2736 | 300 | 2436 |
| African | No Recorded Depression | Augmentation | Insomnia | Income_quantitative | 0.730 | 0.646 | 0.818 | 1.88e-07 | 1.31e-06 | 2736 | 300 | 2436 |
| African | No Recorded Depression | Augmentation | Insomnia | Education_levelCollege Undergraduate | 1.122 | 0.631 | 2.132 | 7.09e-01 | 1.00e+00 | 2736 | 300 | 2436 |
| African | No Recorded Depression | Augmentation | Insomnia | Education_levelLess than a high school degree | 1.701 | 0.886 | 3.443 | 1.23e-01 | 8.59e-01 | 2736 | 300 | 2436 |
| African | No Recorded Depression | Augmentation | Insomnia | Education_levelTwelve or GED | 1.361 | 0.744 | 2.651 | 3.39e-01 | 1.00e+00 | 2736 | 300 | 2436 |
| African | No Recorded Depression | Augmentation | Insomnia | Insomnia | 1.275 | 1.063 | 1.531 | 9.13e-03 | 6.39e-02 | 2736 | 300 | 2436 |
| African | No Recorded Depression | Augmentation | BMI | (Intercept) | 0.605 | 0.267 | 1.317 | 2.15e-01 | 1.00e+00 | 2736 | 300 | 2436 |
| African | No Recorded Depression | Augmentation | BMI | age_at_first_AD | 0.970 | 0.961 | 0.979 | 1.98e-10 | 1.39e-09 | 2736 | 300 | 2436 |
| African | No Recorded Depression | Augmentation | BMI | sex_at_birthMale | 1.741 | 1.343 | 2.252 | 2.61e-05 | 1.83e-04 | 2736 | 300 | 2436 |
| African | No Recorded Depression | Augmentation | BMI | Income_quantitative | 0.729 | 0.645 | 0.817 | 1.71e-07 | 1.20e-06 | 2736 | 300 | 2436 |
| African | No Recorded Depression | Augmentation | BMI | Education_levelCollege Undergraduate | 1.130 | 0.636 | 2.146 | 6.92e-01 | 1.00e+00 | 2736 | 300 | 2436 |
| African | No Recorded Depression | Augmentation | BMI | Education_levelLess than a high school degree | 1.704 | 0.888 | 3.448 | 1.21e-01 | 8.48e-01 | 2736 | 300 | 2436 |
| African | No Recorded Depression | Augmentation | BMI | Education_levelTwelve or GED | 1.332 | 0.728 | 2.594 | 3.73e-01 | 1.00e+00 | 2736 | 300 | 2436 |
| African | No Recorded Depression | Augmentation | BMI | BMI | 1.217 | 1.033 | 1.435 | 1.92e-02 | 1.34e-01 | 2736 | 300 | 2436 |
| African | No Recorded Depression | Augmentation | Bipolar Disorder | (Intercept) | 0.603 | 0.266 | 1.313 | 2.13e-01 | 1.00e+00 | 2736 | 300 | 2436 |
| African | No Recorded Depression | Augmentation | Bipolar Disorder | age_at_first_AD | 0.970 | 0.961 | 0.979 | 2.23e-10 | 1.56e-09 | 2736 | 300 | 2436 |
| African | No Recorded Depression | Augmentation | Bipolar Disorder | sex_at_birthMale | 1.723 | 1.329 | 2.227 | 3.55e-05 | 2.48e-04 | 2736 | 300 | 2436 |

| | | | | | | | | | | | | |
|---|---|---|---|---|---|---|---|---|---|---|---|---|
| African | No Recorded Depression | Augmentation | Bipolar Disorder | Income_quantitative | 0.728 | 0.644 | 0.816 | 1.54e-07 | 1.08e-06 | 2736 | 300 | 2436 |
| African | No Recorded Depression | Augmentation | Bipolar Disorder | Education_levelCollege Undergraduate | 1.124 | 0.633 | 2.136 | 7.04e-01 | 1.00e+00 | 2736 | 300 | 2436 |
| African | No Recorded Depression | Augmentation | Bipolar Disorder | Education_levelLess than a high school degree | 1.740 | 0.907 | 3.520 | 1.07e-01 | 7.50e-01 | 2736 | 300 | 2436 |
| African | No Recorded Depression | Augmentation | Bipolar Disorder | Education_levelTwelve or GED | 1.365 | 0.746 | 2.658 | 3.34e-01 | 1.00e+00 | 2736 | 300 | 2436 |
| African | No Recorded Depression | Augmentation | Bipolar Disorder | Bipolar Disorder | 1.150 | 0.961 | 1.377 | 1.28e-01 | 8.95e-01 | 2736 | 300 | 2436 |
| African | No Recorded Depression | Augmentation | Depression | (Intercept) | 0.597 | 0.263 | 1.303 | 2.05e-01 | 1.00e+00 | 2736 | 300 | 2436 |
| African | No Recorded Depression | Augmentation | Depression | age_at_first_AD | 0.970 | 0.961 | 0.979 | 1.61e-10 | 1.12e-09 | 2736 | 300 | 2436 |
| African | No Recorded Depression | Augmentation | Depression | sex_at_birthMale | 1.727 | 1.332 | 2.233 | 3.33e-05 | 2.33e-04 | 2736 | 300 | 2436 |
| African | No Recorded Depression | Augmentation | Depression | Income_quantitative | 0.733 | 0.648 | 0.821 | 2.49e-07 | 1.74e-06 | 2736 | 300 | 2436 |
| African | No Recorded Depression | Augmentation | Depression | Education_levelCollege Undergraduate | 1.133 | 0.637 | 2.154 | 6.86e-01 | 1.00e+00 | 2736 | 300 | 2436 |
| African | No Recorded Depression | Augmentation | Depression | Education_levelLess than a high school degree | 1.748 | 0.910 | 3.539 | 1.05e-01 | 7.36e-01 | 2736 | 300 | 2436 |
| African | No Recorded Depression | Augmentation | Depression | Education_levelTwelve or GED | 1.373 | 0.749 | 2.677 | 3.26e-01 | 1.00e+00 | 2736 | 300 | 2436 |
| African | No Recorded Depression | Augmentation | Depression | Depression | 1.289 | 1.084 | 1.534 | 4.10e-03 | 2.87e-02 | 2736 | 300 | 2436 |
| African | No Recorded Depression | Augmentation | Schizophrenia | (Intercept) | 0.609 | 0.269 | 1.325 | 2.21e-01 | 1.00e+00 | 2736 | 300 | 2436 |
| African | No Recorded Depression | Augmentation | Schizophrenia | age_at_first_AD | 0.970 | 0.961 | 0.979 | 1.93e-10 | 1.35e-09 | 2736 | 300 | 2436 |
| African | No Recorded Depression | Augmentation | Schizophrenia | sex_at_birthMale | 1.715 | 1.323 | 2.216 | 4.09e-05 | 2.86e-04 | 2736 | 300 | 2436 |
| African | No Recorded Depression | Augmentation | Schizophrenia | Income_quantitative | 0.728 | 0.644 | 0.816 | 1.53e-07 | 1.07e-06 | 2736 | 300 | 2436 |
| African | No Recorded Depression | Augmentation | Schizophrenia | Education_levelCollege Undergraduate | 1.130 | 0.636 | 2.146 | 6.92e-01 | 1.00e+00 | 2736 | 300 | 2436 |
| African | No Recorded Depression | Augmentation | Schizophrenia | Education_levelLess than a high school degree | 1.737 | 0.906 | 3.514 | 1.08e-01 | 7.58e-01 | 2736 | 300 | 2436 |
| African | No Recorded Depression | Augmentation | Schizophrenia | Education_levelTwelve or GED | 1.358 | 0.742 | 2.644 | 3.42e-01 | 1.00e+00 | 2736 | 300 | 2436 |
| African | No Recorded Depression | Augmentation | Schizophrenia | Schizophrenia | 1.025 | 0.874 | 1.201 | 7.65e-01 | 1.00e+00 | 2736 | 300 | 2436 |
| African | Recorded Anxiety | Augmentation | ADHD | (Intercept) | 1.208 | 0.720 | 2.017 | 4.71e-01 | 1.00e+00 | 2677 | 663 | 2014 |
| African | Recorded Anxiety | Augmentation | ADHD | age_at_first_AD | 0.981 | 0.974 | 0.987 | 1.70e-08 | 1.19e-07 | 2677 | 663 | 2014 |
| African | Recorded Anxiety | Augmentation | ADHD | sex_at_birthMale | 1.580 | 1.285 | 1.940 | 1.33e-05 | 9.29e-05 | 2677 | 663 | 2014 |
| African | Recorded Anxiety | Augmentation | ADHD | Income_quantitative | 0.772 | 0.717 | 0.828 | 1.46e-12 | 1.02e-11 | 2677 | 663 | 2014 |
| African | Recorded Anxiety | Augmentation | ADHD | Education_levelCollege Undergraduate | 0.949 | 0.670 | 1.362 | 7.71e-01 | 1.00e+00 | 2677 | 663 | 2014 |
| African | Recorded Anxiety | Augmentation | ADHD | Education_levelLess than a high school degree | 1.306 | 0.839 | 2.045 | 2.40e-01 | 1.00e+00 | 2677 | 663 | 2014 |
| African | Recorded Anxiety | Augmentation | ADHD | Education_levelTwelve or GED | 0.942 | 0.642 | 1.396 | 7.62e-01 | 1.00e+00 | 2677 | 663 | 2014 |
| African | Recorded Anxiety | Augmentation | ADHD | ADHD | 1.014 | 0.893 | 1.152 | 8.31e-01 | 1.00e+00 | 2677 | 663 | 2014 |
| African | Recorded Anxiety | Augmentation | Migraine | (Intercept) | 1.205 | 0.719 | 2.013 | 4.77e-01 | 1.00e+00 | 2677 | 663 | 2014 |
| African | Recorded Anxiety | Augmentation | Migraine | age_at_first_AD | 0.981 | 0.974 | 0.987 | 1.72e-08 | 1.21e-07 | 2677 | 663 | 2014 |
| African | Recorded Anxiety | Augmentation | Migraine | sex_at_birthMale | 1.582 | 1.287 | 1.943 | 1.26e-05 | 8.82e-05 | 2677 | 663 | 2014 |
| African | Recorded Anxiety | Augmentation | Migraine | Income_quantitative | 0.772 | 0.717 | 0.828 | 1.50e-12 | 1.05e-11 | 2677 | 663 | 2014 |
| African | Recorded Anxiety | Augmentation | Migraine | Education_levelCollege Undergraduate | 0.950 | 0.671 | 1.364 | 7.76e-01 | 1.00e+00 | 2677 | 663 | 2014 |
| African | Recorded Anxiety | Augmentation | Migraine | Education_levelLess than a high school degree | 1.308 | 0.841 | 2.048 | 2.36e-01 | 1.00e+00 | 2677 | 663 | 2014 |
| African | Recorded Anxiety | Augmentation | Migraine | Education_levelTwelve or GED | 0.944 | 0.644 | 1.399 | 7.71e-01 | 1.00e+00 | 2677 | 663 | 2014 |
| African | Recorded Anxiety | Augmentation | Migraine | Migraine | 0.981 | 0.877 | 1.098 | 7.41e-01 | 1.00e+00 | 2677 | 663 | 2014 |
| African | Recorded Anxiety | Augmentation | Insomnia | (Intercept) | 1.224 | 0.729 | 2.045 | 4.43e-01 | 1.00e+00 | 2677 | 663 | 2014 |
| African | Recorded Anxiety | Augmentation | Insomnia | age_at_first_AD | 0.981 | 0.974 | 0.987 | 1.46e-08 | 1.02e-07 | 2677 | 663 | 2014 |
| African | Recorded Anxiety | Augmentation | Insomnia | sex_at_birthMale | 1.575 | 1.280 | 1.934 | 1.57e-05 | 1.10e-04 | 2677 | 663 | 2014 |
| African | Recorded Anxiety | Augmentation | Insomnia | Income_quantitative | 0.771 | 0.717 | 0.827 | 1.32e-12 | 9.23e-12 | 2677 | 663 | 2014 |
| African | Recorded Anxiety | Augmentation | Insomnia | Education_levelCollege Undergraduate | 0.941 | 0.664 | 1.352 | 7.38e-01 | 1.00e+00 | 2677 | 663 | 2014 |
| African | Recorded Anxiety | Augmentation | Insomnia | Education_levelLess than a high school degree | 1.295 | 0.831 | 2.029 | 2.56e-01 | 1.00e+00 | 2677 | 663 | 2014 |
| African | Recorded Anxiety | Augmentation | Insomnia | Education_levelTwelve or GED | 0.935 | 0.637 | 1.385 | 7.32e-01 | 1.00e+00 | 2677 | 663 | 2014 |
| African | Recorded Anxiety | Augmentation | Insomnia | Insomnia | 1.148 | 1.006 | 1.311 | 4.12e-02 | 2.89e-01 | 2677 | 663 | 2014 |
| African | Recorded Anxiety | Augmentation | BMI | (Intercept) | 1.207 | 0.720 | 2.015 | 4.74e-01 | 1.00e+00 | 2677 | 663 | 2014 |
| African | Recorded Anxiety | Augmentation | BMI | age_at_first_AD | 0.981 | 0.974 | 0.987 | 1.65e-08 | 1.16e-07 | 2677 | 663 | 2014 |
| African | Recorded Anxiety | Augmentation | BMI | sex_at_birthMale | 1.579 | 1.283 | 1.939 | 1.43e-05 | 1.00e-04 | 2677 | 663 | 2014 |
| African | Recorded Anxiety | Augmentation | BMI | Income_quantitative | 0.771 | 0.717 | 0.828 | 1.42e-12 | 9.95e-12 | 2677 | 663 | 2014 |
| African | Recorded Anxiety | Augmentation | BMI | Education_levelCollege Undergraduate | 0.951 | 0.671 | 1.366 | 7.82e-01 | 1.00e+00 | 2677 | 663 | 2014 |
| African | Recorded Anxiety | Augmentation | BMI | Education_levelLess than a high school degree | 1.310 | 0.842 | 2.052 | 2.34e-01 | 1.00e+00 | 2677 | 663 | 2014 |
| African | Recorded Anxiety | Augmentation | BMI | Education_levelTwelve or GED | 0.946 | 0.645 | 1.402 | 7.78e-01 | 1.00e+00 | 2677 | 663 | 2014 |
| African | Recorded Anxiety | Augmentation | BMI | BMI | 0.984 | 0.873 | 1.108 | 7.85e-01 | 1.00e+00 | 2677 | 663 | 2014 |
| African | Recorded Anxiety | Augmentation | Bipolar Disorder | (Intercept) | 1.222 | 0.728 | 2.041 | 4.46e-01 | 1.00e+00 | 2677 | 663 | 2014 |
| African | Recorded Anxiety | Augmentation | Bipolar Disorder | age_at_first_AD | 0.981 | 0.974 | 0.987 | 1.21e-08 | 8.45e-08 | 2677 | 663 | 2014 |

| | | | | | | | | | | | | |
|---|---|---|---|---|---|---|---|---|---|---|---|---|
| African | Recorded Anxiety | Augmentation | Bipolar Disorder | sex_at_birthMale | 1.570 | 1.276 | 1.928 | 1.81e-05 | 1.27e-04 | 2677 | 663 | 2014 |
| African | Recorded Anxiety | Augmentation | Bipolar Disorder | Income_quantitative | 0.772 | 0.717 | 0.829 | 1.77e-12 | 1.24e-11 | 2677 | 663 | 2014 |
| African | Recorded Anxiety | Augmentation | Bipolar Disorder | Education_levelCollege Undergraduate | 0.945 | 0.667 | 1.356 | 7.54e-01 | 1.00e+00 | 2677 | 663 | 2014 |
| African | Recorded Anxiety | Augmentation | Bipolar Disorder | Education_levelLess than a high school degree | 1.303 | 0.837 | 2.040 | 2.44e-01 | 1.00e+00 | 2677 | 663 | 2014 |
| African | Recorded Anxiety | Augmentation | Bipolar Disorder | Education_levelTwelve or GED | 0.945 | 0.644 | 1.400 | 7.73e-01 | 1.00e+00 | 2677 | 663 | 2014 |
| African | Recorded Anxiety | Augmentation | Bipolar Disorder | Bipolar Disorder | 1.124 | 0.985 | 1.283 | 8.21e-02 | 5.75e-01 | 2677 | 663 | 2014 |
| African | Recorded Anxiety | Augmentation | Depression | (Intercept) | 1.216 | 0.724 | 2.033 | 4.58e-01 | 1.00e+00 | 2677 | 663 | 2014 |
| African | Recorded Anxiety | Augmentation | Depression | age_at_first_AD | 0.980 | 0.974 | 0.987 | 8.15e-09 | 5.70e-08 | 2677 | 663 | 2014 |
| African | Recorded Anxiety | Augmentation | Depression | sex_at_birthMale | 1.577 | 1.282 | 1.937 | 1.52e-05 | 1.06e-04 | 2677 | 663 | 2014 |
| African | Recorded Anxiety | Augmentation | Depression | Income_quantitative | 0.774 | 0.720 | 0.831 | 2.98e-12 | 2.09e-11 | 2677 | 663 | 2014 |
| African | Recorded Anxiety | Augmentation | Depression | Education_levelCollege Undergraduate | 0.955 | 0.673 | 1.371 | 7.97e-01 | 1.00e+00 | 2677 | 663 | 2014 |
| African | Recorded Anxiety | Augmentation | Depression | Education_levelLess than a high school degree | 1.312 | 0.842 | 2.055 | 2.33e-01 | 1.00e+00 | 2677 | 663 | 2014 |
| African | Recorded Anxiety | Augmentation | Depression | Education_levelTwelve or GED | 0.948 | 0.646 | 1.405 | 7.88e-01 | 1.00e+00 | 2677 | 663 | 2014 |
| African | Recorded Anxiety | Augmentation | Depression | Depression | 1.214 | 1.066 | 1.384 | 3.56e-03 | 2.50e-02 | 2677 | 663 | 2014 |
| African | Recorded Anxiety | Augmentation | Schizophrenia | (Intercept) | 1.208 | 0.720 | 2.017 | 4.72e-01 | 1.00e+00 | 2677 | 663 | 2014 |
| African | Recorded Anxiety | Augmentation | Schizophrenia | age_at_first_AD | 0.981 | 0.974 | 0.988 | 1.78e-08 | 1.25e-07 | 2677 | 663 | 2014 |
| African | Recorded Anxiety | Augmentation | Schizophrenia | sex_at_birthMale | 1.580 | 1.285 | 1.940 | 1.34e-05 | 9.40e-05 | 2677 | 663 | 2014 |
| African | Recorded Anxiety | Augmentation | Schizophrenia | Income_quantitative | 0.771 | 0.716 | 0.827 | 1.30e-12 | 9.10e-12 | 2677 | 663 | 2014 |
| African | Recorded Anxiety | Augmentation | Schizophrenia | Education_levelCollege Undergraduate | 0.949 | 0.670 | 1.362 | 7.72e-01 | 1.00e+00 | 2677 | 663 | 2014 |
| African | Recorded Anxiety | Augmentation | Schizophrenia | Education_levelLess than a high school degree | 1.309 | 0.841 | 2.049 | 2.36e-01 | 1.00e+00 | 2677 | 663 | 2014 |
| African | Recorded Anxiety | Augmentation | Schizophrenia | Education_levelTwelve or GED | 0.943 | 0.643 | 1.397 | 7.65e-01 | 1.00e+00 | 2677 | 663 | 2014 |
| African | Recorded Anxiety | Augmentation | Schizophrenia | Schizophrenia | 0.959 | 0.854 | 1.075 | 4.71e-01 | 1.00e+00 | 2677 | 663 | 2014 |
| Latino/Mixed-American | Full Cohort | Augmentation | ADHD | (Intercept) | 0.569 | 0.338 | 0.949 | 3.21e-02 | 2.25e-01 | 2971 | 515 | 2456 |
| Latino/Mixed-American | Full Cohort | Augmentation | ADHD | age_at_first_AD | 0.989 | 0.982 | 0.996 | 1.29e-03 | 9.01e-03 | 2971 | 515 | 2456 |
| Latino/Mixed-American | Full Cohort | Augmentation | ADHD | sex_at_birthMale | 1.597 | 1.297 | 1.963 | 9.45e-06 | 6.61e-05 | 2971 | 515 | 2456 |
| Latino/Mixed-American | Full Cohort | Augmentation | ADHD | Income_quantitative | 0.804 | 0.753 | 0.857 | 4.47e-11 | 3.13e-10 | 2971 | 515 | 2456 |
| Latino/Mixed-American | Full Cohort | Augmentation | ADHD | Education_levelCollege Undergraduate | 0.875 | 0.608 | 1.282 | 4.83e-01 | 1.00e+00 | 2971 | 515 | 2456 |
| Latino/Mixed-American | Full Cohort | Augmentation | ADHD | Education_levelLess than a high school degree | 0.915 | 0.599 | 1.416 | 6.86e-01 | 1.00e+00 | 2971 | 515 | 2456 |
| Latino/Mixed-American | Full Cohort | Augmentation | ADHD | Education_levelTwelve or GED | 0.785 | 0.518 | 1.204 | 2.60e-01 | 1.00e+00 | 2971 | 515 | 2456 |
| Latino/Mixed-American | Full Cohort | Augmentation | ADHD | ADHD | 1.084 | 0.973 | 1.209 | 1.44e-01 | 1.00e+00 | 2971 | 515 | 2456 |
| Latino/Mixed-American | Full Cohort | Augmentation | Migraine | (Intercept) | 0.574 | 0.341 | 0.956 | 3.44e-02 | 2.41e-01 | 2971 | 515 | 2456 |
| Latino/Mixed-American | Full Cohort | Augmentation | Migraine | age_at_first_AD | 0.989 | 0.982 | 0.996 | 1.03e-03 | 7.22e-03 | 2971 | 515 | 2456 |
| Latino/Mixed-American | Full Cohort | Augmentation | Migraine | sex_at_birthMale | 1.596 | 1.296 | 1.961 | 9.79e-06 | 6.85e-05 | 2971 | 515 | 2456 |
| Latino/Mixed-American | Full Cohort | Augmentation | Migraine | Income_quantitative | 0.803 | 0.752 | 0.856 | 3.32e-11 | 2.33e-10 | 2971 | 515 | 2456 |
| Latino/Mixed-American | Full Cohort | Augmentation | Migraine | Education_levelCollege Undergraduate | 0.880 | 0.611 | 1.289 | 5.01e-01 | 1.00e+00 | 2971 | 515 | 2456 |
| Latino/Mixed-American | Full Cohort | Augmentation | Migraine | Education_levelLess than a high school degree | 0.925 | 0.605 | 1.430 | 7.21e-01 | 1.00e+00 | 2971 | 515 | 2456 |
| Latino/Mixed-American | Full Cohort | Augmentation | Migraine | Education_levelTwelve or GED | 0.793 | 0.523 | 1.216 | 2.79e-01 | 1.00e+00 | 2971 | 515 | 2456 |
| Latino/Mixed-American | Full Cohort | Augmentation | Migraine | Migraine | 1.035 | 0.935 | 1.146 | 5.06e-01 | 1.00e+00 | 2971 | 515 | 2456 |
| Latino/Mixed-American | Full Cohort | Augmentation | Insomnia | (Intercept) | 0.572 | 0.340 | 0.954 | 3.36e-02 | 2.35e-01 | 2971 | 515 | 2456 |
| Latino/Mixed-American | Full Cohort | Augmentation | Insomnia | age_at_first_AD | 0.989 | 0.982 | 0.996 | 1.13e-03 | 7.88e-03 | 2971 | 515 | 2456 |
| Latino/Mixed-American | Full Cohort | Augmentation | Insomnia | sex_at_birthMale | 1.601 | 1.300 | 1.968 | 8.67e-06 | 6.07e-05 | 2971 | 515 | 2456 |
| Latino/Mixed-American | Full Cohort | Augmentation | Insomnia | Income_quantitative | 0.804 | 0.753 | 0.857 | 4.11e-11 | 2.88e-10 | 2971 | 515 | 2456 |
| Latino/Mixed-American | Full Cohort | Augmentation | Insomnia | Education_levelCollege Undergraduate | 0.872 | 0.605 | 1.277 | 4.71e-01 | 1.00e+00 | 2971 | 515 | 2456 |
| Latino/Mixed-American | Full Cohort | Augmentation | Insomnia | Education_levelLess than a high school degree | 0.913 | 0.597 | 1.412 | 6.76e-01 | 1.00e+00 | 2971 | 515 | 2456 |
| Latino/Mixed-American | Full Cohort | Augmentation | Insomnia | Education_levelTwelve or GED | 0.789 | 0.521 | 1.210 | 2.71e-01 | 1.00e+00 | 2971 | 515 | 2456 |
| Latino/Mixed-American | Full Cohort | Augmentation | Insomnia | Insomnia | 1.139 | 1.019 | 1.275 | 2.24e-02 | 1.57e-01 | 2971 | 515 | 2456 |
| Latino/Mixed-American | Full Cohort | Augmentation | BMI | (Intercept) | 0.575 | 0.342 | 0.958 | 3.51e-02 | 2.46e-01 | 2971 | 515 | 2456 |
| Latino/Mixed-American | Full Cohort | Augmentation | BMI | age_at_first_AD | 0.989 | 0.982 | 0.995 | 9.51e-04 | 6.66e-03 | 2971 | 515 | 2456 |
| Latino/Mixed-American | Full Cohort | Augmentation | BMI | sex_at_birthMale | 1.595 | 1.295 | 1.960 | 9.90e-06 | 6.93e-05 | 2971 | 515 | 2456 |
| Latino/Mixed-American | Full Cohort | Augmentation | BMI | Income_quantitative | 0.802 | 0.751 | 0.855 | 2.84e-11 | 1.98e-10 | 2971 | 515 | 2456 |
| Latino/Mixed-American | Full Cohort | Augmentation | BMI | Education_levelCollege Undergraduate | 0.884 | 0.614 | 1.294 | 5.14e-01 | 1.00e+00 | 2971 | 515 | 2456 |
| Latino/Mixed-American | Full Cohort | Augmentation | BMI | Education_levelLess than a high school degree | 0.928 | 0.608 | 1.436 | 7.34e-01 | 1.00e+00 | 2971 | 515 | 2456 |
| Latino/Mixed-American | Full Cohort | Augmentation | BMI | Education_levelTwelve or GED | 0.798 | 0.527 | 1.224 | 2.94e-01 | 1.00e+00 | 2971 | 515 | 2456 |
| Latino/Mixed-American | Full Cohort | Augmentation | BMI | BMI | 0.972 | 0.879 | 1.074 | 5.74e-01 | 1.00e+00 | 2971 | 515 | 2456 |
| Latino/Mixed-American | Full Cohort | Augmentation | Bipolar Disorder | (Intercept) | 0.557 | 0.331 | 0.929 | 2.62e-02 | 1.83e-01 | 2971 | 515 | 2456 |

| | | | | | | | | | | | | |
|---|---|---|---|---|---|---|---|---|---|---|---|---|
| Latino/Mixed-American | Full Cohort | Augmentation | Bipolar Disorder | age_at_first_AD | 0.989 | 0.982 | 0.996 | 1.03e-03 | 7.21e-03 | 2971 | 515 | 2456 |
| Latino/Mixed-American | Full Cohort | Augmentation | Bipolar Disorder | sex_at_birthMale | 1.615 | 1.311 | 1.987 | 6.05e-06 | 4.23e-05 | 2971 | 515 | 2456 |
| Latino/Mixed-American | Full Cohort | Augmentation | Bipolar Disorder | Income_quantitative | 0.804 | 0.753 | 0.857 | 4.05e-11 | 2.84e-10 | 2971 | 515 | 2456 |
| Latino/Mixed-American | Full Cohort | Augmentation | Bipolar Disorder | Education_levelCollege Undergraduate | 0.894 | 0.620 | 1.309 | 5.54e-01 | 1.00e+00 | 2971 | 515 | 2456 |
| Latino/Mixed-American | Full Cohort | Augmentation | Bipolar Disorder | Education_levelLess than a high school degree | 0.937 | 0.614 | 1.450 | 7.68e-01 | 1.00e+00 | 2971 | 515 | 2456 |
| Latino/Mixed-American | Full Cohort | Augmentation | Bipolar Disorder | Education_levelTwelve or GED | 0.814 | 0.537 | 1.248 | 3.38e-01 | 1.00e+00 | 2971 | 515 | 2456 |
| Latino/Mixed-American | Full Cohort | Augmentation | Bipolar Disorder | Bipolar Disorder | 1.195 | 1.073 | 1.331 | 1.22e-03 | 8.55e-03 | 2971 | 515 | 2456 |
| Latino/Mixed-American | Full Cohort | Augmentation | Depression | (Intercept) | 0.553 | 0.328 | 0.923 | 2.47e-02 | 1.73e-01 | 2971 | 515 | 2456 |
| Latino/Mixed-American | Full Cohort | Augmentation | Depression | age_at_first_AD | 0.989 | 0.983 | 0.996 | 1.76e-03 | 1.23e-02 | 2971 | 515 | 2456 |
| Latino/Mixed-American | Full Cohort | Augmentation | Depression | sex_at_birthMale | 1.597 | 1.295 | 1.964 | 1.06e-05 | 7.45e-05 | 2971 | 515 | 2456 |
| Latino/Mixed-American | Full Cohort | Augmentation | Depression | Income_quantitative | 0.807 | 0.755 | 0.860 | 8.53e-11 | 5.97e-10 | 2971 | 515 | 2456 |
| Latino/Mixed-American | Full Cohort | Augmentation | Depression | Education_levelCollege Undergraduate | 0.861 | 0.597 | 1.262 | 4.31e-01 | 1.00e+00 | 2971 | 515 | 2456 |
| Latino/Mixed-American | Full Cohort | Augmentation | Depression | Education_levelLess than a high school degree | 0.902 | 0.590 | 1.395 | 6.37e-01 | 1.00e+00 | 2971 | 515 | 2456 |
| Latino/Mixed-American | Full Cohort | Augmentation | Depression | Education_levelTwelve or GED | 0.789 | 0.521 | 1.211 | 2.71e-01 | 1.00e+00 | 2971 | 515 | 2456 |
| Latino/Mixed-American | Full Cohort | Augmentation | Depression | Depression | 1.337 | 1.196 | 1.496 | 3.87e-07 | 2.71e-06 | 2971 | 515 | 2456 |
| Latino/Mixed-American | Full Cohort | Augmentation | Schizophrenia | (Intercept) | 0.564 | 0.335 | 0.941 | 2.94e-02 | 2.06e-01 | 2971 | 515 | 2456 |
| Latino/Mixed-American | Full Cohort | Augmentation | Schizophrenia | age_at_first_AD | 0.989 | 0.982 | 0.995 | 8.86e-04 | 6.20e-03 | 2971 | 515 | 2456 |
| Latino/Mixed-American | Full Cohort | Augmentation | Schizophrenia | sex_at_birthMale | 1.600 | 1.299 | 1.967 | 8.76e-06 | 6.13e-05 | 2971 | 515 | 2456 |
| Latino/Mixed-American | Full Cohort | Augmentation | Schizophrenia | Income_quantitative | 0.805 | 0.754 | 0.858 | 4.95e-11 | 3.46e-10 | 2971 | 515 | 2456 |
| Latino/Mixed-American | Full Cohort | Augmentation | Schizophrenia | Education_levelCollege Undergraduate | 0.892 | 0.620 | 1.307 | 5.49e-01 | 1.00e+00 | 2971 | 515 | 2456 |
| Latino/Mixed-American | Full Cohort | Augmentation | Schizophrenia | Education_levelLess than a high school degree | 0.939 | 0.615 | 1.451 | 7.73e-01 | 1.00e+00 | 2971 | 515 | 2456 |
| Latino/Mixed-American | Full Cohort | Augmentation | Schizophrenia | Education_levelTwelve or GED | 0.806 | 0.532 | 1.236 | 3.15e-01 | 1.00e+00 | 2971 | 515 | 2456 |
| Latino/Mixed-American | Full Cohort | Augmentation | Schizophrenia | Schizophrenia | 1.106 | 1.003 | 1.220 | 4.30e-02 | 3.01e-01 | 2971 | 515 | 2456 |
| Latino/Mixed-American | Recorded Depression | Augmentation | ADHD | (Intercept) | 0.633 | 0.328 | 1.208 | 1.69e-01 | 1.00e+00 | 1565 | 386 | 1179 |
| Latino/Mixed-American | Recorded Depression | Augmentation | ADHD | age_at_first_AD | 0.991 | 0.983 | 0.999 | 2.84e-02 | 1.99e-01 | 1565 | 386 | 1179 |
| Latino/Mixed-American | Recorded Depression | Augmentation | ADHD | sex_at_birthMale | 1.386 | 1.072 | 1.788 | 1.23e-02 | 8.58e-02 | 1565 | 386 | 1179 |
| Latino/Mixed-American | Recorded Depression | Augmentation | ADHD | Income_quantitative | 0.876 | 0.809 | 0.947 | 9.36e-04 | 6.55e-03 | 1565 | 386 | 1179 |
| Latino/Mixed-American | Recorded Depression | Augmentation | ADHD | Education_levelCollege Undergraduate | 0.954 | 0.599 | 1.554 | 8.44e-01 | 1.00e+00 | 1565 | 386 | 1179 |
| Latino/Mixed-American | Recorded Depression | Augmentation | ADHD | Education_levelLess than a high school degree | 1.124 | 0.661 | 1.948 | 6.70e-01 | 1.00e+00 | 1565 | 386 | 1179 |
| Latino/Mixed-American | Recorded Depression | Augmentation | ADHD | Education_levelTwelve or GED | 0.769 | 0.456 | 1.319 | 3.31e-01 | 1.00e+00 | 1565 | 386 | 1179 |
| Latino/Mixed-American | Recorded Depression | Augmentation | ADHD | ADHD | 1.035 | 0.905 | 1.183 | 6.19e-01 | 1.00e+00 | 1565 | 386 | 1179 |
| Latino/Mixed-American | Recorded Depression | Augmentation | Migraine | (Intercept) | 0.648 | 0.335 | 1.238 | 1.93e-01 | 1.00e+00 | 1565 | 386 | 1179 |
| Latino/Mixed-American | Recorded Depression | Augmentation | Migraine | age_at_first_AD | 0.991 | 0.983 | 0.999 | 2.71e-02 | 1.90e-01 | 1565 | 386 | 1179 |
| Latino/Mixed-American | Recorded Depression | Augmentation | Migraine | sex_at_birthMale | 1.385 | 1.070 | 1.787 | 1.27e-02 | 8.89e-02 | 1565 | 386 | 1179 |
| Latino/Mixed-American | Recorded Depression | Augmentation | Migraine | Income_quantitative | 0.875 | 0.808 | 0.945 | 8.48e-04 | 5.94e-03 | 1565 | 386 | 1179 |
| Latino/Mixed-American | Recorded Depression | Augmentation | Migraine | Education_levelCollege Undergraduate | 0.938 | 0.589 | 1.530 | 7.92e-01 | 1.00e+00 | 1565 | 386 | 1179 |
| Latino/Mixed-American | Recorded Depression | Augmentation | Migraine | Education_levelLess than a high school degree | 1.099 | 0.645 | 1.908 | 7.32e-01 | 1.00e+00 | 1565 | 386 | 1179 |
| Latino/Mixed-American | Recorded Depression | Augmentation | Migraine | Education_levelTwelve or GED | 0.742 | 0.439 | 1.275 | 2.71e-01 | 1.00e+00 | 1565 | 386 | 1179 |
| Latino/Mixed-American | Recorded Depression | Augmentation | Migraine | Migraine | 1.155 | 1.020 | 1.308 | 2.34e-02 | 1.64e-01 | 1565 | 386 | 1179 |
| Latino/Mixed-American | Recorded Depression | Augmentation | Insomnia | (Intercept) | 0.635 | 0.329 | 1.212 | 1.72e-01 | 1.00e+00 | 1565 | 386 | 1179 |
| Latino/Mixed-American | Recorded Depression | Augmentation | Insomnia | age_at_first_AD | 0.991 | 0.983 | 0.999 | 2.92e-02 | 2.05e-01 | 1565 | 386 | 1179 |
| Latino/Mixed-American | Recorded Depression | Augmentation | Insomnia | sex_at_birthMale | 1.384 | 1.070 | 1.785 | 1.27e-02 | 8.90e-02 | 1565 | 386 | 1179 |
| Latino/Mixed-American | Recorded Depression | Augmentation | Insomnia | Income_quantitative | 0.875 | 0.808 | 0.945 | 8.48e-04 | 5.94e-03 | 1565 | 386 | 1179 |
| Latino/Mixed-American | Recorded Depression | Augmentation | Insomnia | Education_levelCollege Undergraduate | 0.948 | 0.595 | 1.545 | 8.25e-01 | 1.00e+00 | 1565 | 386 | 1179 |
| Latino/Mixed-American | Recorded Depression | Augmentation | Insomnia | Education_levelLess than a high school degree | 1.116 | 0.656 | 1.934 | 6.89e-01 | 1.00e+00 | 1565 | 386 | 1179 |
| Latino/Mixed-American | Recorded Depression | Augmentation | Insomnia | Education_levelTwelve or GED | 0.767 | 0.455 | 1.316 | 3.27e-01 | 1.00e+00 | 1565 | 386 | 1179 |
| Latino/Mixed-American | Recorded Depression | Augmentation | Insomnia | Insomnia | 1.119 | 0.979 | 1.280 | 1.01e-01 | 7.06e-01 | 1565 | 386 | 1179 |
| Latino/Mixed-American | Recorded Depression | Augmentation | BMI | (Intercept) | 0.634 | 0.328 | 1.209 | 1.70e-01 | 1.00e+00 | 1565 | 386 | 1179 |
| Latino/Mixed-American | Recorded Depression | Augmentation | BMI | age_at_first_AD | 0.991 | 0.983 | 0.999 | 2.63e-02 | 1.84e-01 | 1565 | 386 | 1179 |
| Latino/Mixed-American | Recorded Depression | Augmentation | BMI | sex_at_birthMale | 1.385 | 1.071 | 1.786 | 1.26e-02 | 8.82e-02 | 1565 | 386 | 1179 |
| Latino/Mixed-American | Recorded Depression | Augmentation | BMI | Income_quantitative | 0.873 | 0.807 | 0.944 | 7.10e-04 | 4.97e-03 | 1565 | 386 | 1179 |
| Latino/Mixed-American | Recorded Depression | Augmentation | BMI | Education_levelCollege Undergraduate | 0.963 | 0.605 | 1.570 | 8.77e-01 | 1.00e+00 | 1565 | 386 | 1179 |
| Latino/Mixed-American | Recorded Depression | Augmentation | BMI | Education_levelLess than a high school degree | 1.136 | 0.668 | 1.969 | 6.43e-01 | 1.00e+00 | 1565 | 386 | 1179 |
| Latino/Mixed-American | Recorded Depression | Augmentation | BMI | Education_levelTwelve or GED | 0.777 | 0.461 | 1.333 | 3.51e-01 | 1.00e+00 | 1565 | 386 | 1179 |
| Latino/Mixed-American | Recorded Depression | Augmentation | BMI | BMI | 0.927 | 0.821 | 1.046 | 2.18e-01 | 1.00e+00 | 1565 | 386 | 1179 |

| | | | | | | | | | | | | |
|---|---|---|---|---|---|---|---|---|---|---|---|---|
| Latino/Mixed-American | Recorded Depression | Augmentation | Bipolar Disorder | (Intercept) | 0.609 | 0.315 | 1.165 | 1.37e-01 | 9.61e-01 | 1565 | 386 | 1179 |
| Latino/Mixed-American | Recorded Depression | Augmentation | Bipolar Disorder | age_at_first_AD | 0.991 | 0.983 | 0.999 | 2.62e-02 | 1.83e-01 | 1565 | 386 | 1179 |
| Latino/Mixed-American | Recorded Depression | Augmentation | Bipolar Disorder | sex_at_birthMale | 1.399 | 1.081 | 1.806 | 1.03e-02 | 7.24e-02 | 1565 | 386 | 1179 |
| Latino/Mixed-American | Recorded Depression | Augmentation | Bipolar Disorder | Income_quantitative | 0.877 | 0.810 | 0.947 | 9.68e-04 | 6.78e-03 | 1565 | 386 | 1179 |
| Latino/Mixed-American | Recorded Depression | Augmentation | Bipolar Disorder | Education_levelCollege Undergraduate | 0.982 | 0.616 | 1.603 | 9.40e-01 | 1.00e+00 | 1565 | 386 | 1179 |
| Latino/Mixed-American | Recorded Depression | Augmentation | Bipolar Disorder | Education_levelLess than a high school degree | 1.163 | 0.683 | 2.018 | 5.84e-01 | 1.00e+00 | 1565 | 386 | 1179 |
| Latino/Mixed-American | Recorded Depression | Augmentation | Bipolar Disorder | Education_levelTwelve or GED | 0.803 | 0.475 | 1.379 | 4.17e-01 | 1.00e+00 | 1565 | 386 | 1179 |
| Latino/Mixed-American | Recorded Depression | Augmentation | Bipolar Disorder | Bipolar Disorder | 1.259 | 1.106 | 1.435 | 5.15e-04 | 3.60e-03 | 1565 | 386 | 1179 |
| Latino/Mixed-American | Recorded Depression | Augmentation | Depression | (Intercept) | 0.645 | 0.334 | 1.233 | 1.88e-01 | 1.00e+00 | 1565 | 386 | 1179 |
| Latino/Mixed-American | Recorded Depression | Augmentation | Depression | age_at_first_AD | 0.991 | 0.983 | 0.999 | 3.36e-02 | 2.35e-01 | 1565 | 386 | 1179 |
| Latino/Mixed-American | Recorded Depression | Augmentation | Depression | sex_at_birthMale | 1.378 | 1.064 | 1.779 | 1.44e-02 | 1.01e-01 | 1565 | 386 | 1179 |
| Latino/Mixed-American | Recorded Depression | Augmentation | Depression | Income_quantitative | 0.875 | 0.808 | 0.945 | 8.17e-04 | 5.72e-03 | 1565 | 386 | 1179 |
| Latino/Mixed-American | Recorded Depression | Augmentation | Depression | Education_levelCollege Undergraduate | 0.919 | 0.576 | 1.499 | 7.27e-01 | 1.00e+00 | 1565 | 386 | 1179 |
| Latino/Mixed-American | Recorded Depression | Augmentation | Depression | Education_levelLess than a high school degree | 1.066 | 0.625 | 1.850 | 8.17e-01 | 1.00e+00 | 1565 | 386 | 1179 |
| Latino/Mixed-American | Recorded Depression | Augmentation | Depression | Education_levelTwelve or GED | 0.747 | 0.442 | 1.282 | 2.81e-01 | 1.00e+00 | 1565 | 386 | 1179 |
| Latino/Mixed-American | Recorded Depression | Augmentation | Depression | Depression | 1.289 | 1.124 | 1.478 | 2.78e-04 | 1.95e-03 | 1565 | 386 | 1179 |
| Latino/Mixed-American | Recorded Depression | Augmentation | Schizophrenia | (Intercept) | 0.621 | 0.321 | 1.185 | 1.51e-01 | 1.00e+00 | 1565 | 386 | 1179 |
| Latino/Mixed-American | Recorded Depression | Augmentation | Schizophrenia | age_at_first_AD | 0.991 | 0.983 | 0.999 | 2.74e-02 | 1.92e-01 | 1565 | 386 | 1179 |
| Latino/Mixed-American | Recorded Depression | Augmentation | Schizophrenia | sex_at_birthMale | 1.392 | 1.076 | 1.796 | 1.13e-02 | 7.91e-02 | 1565 | 386 | 1179 |
| Latino/Mixed-American | Recorded Depression | Augmentation | Schizophrenia | Income_quantitative | 0.877 | 0.810 | 0.947 | 9.92e-04 | 6.95e-03 | 1565 | 386 | 1179 |
| Latino/Mixed-American | Recorded Depression | Augmentation | Schizophrenia | Education_levelCollege Undergraduate | 0.971 | 0.610 | 1.583 | 9.04e-01 | 1.00e+00 | 1565 | 386 | 1179 |
| Latino/Mixed-American | Recorded Depression | Augmentation | Schizophrenia | Education_levelLess than a high school degree | 1.140 | 0.670 | 1.975 | 6.34e-01 | 1.00e+00 | 1565 | 386 | 1179 |
| Latino/Mixed-American | Recorded Depression | Augmentation | Schizophrenia | Education_levelTwelve or GED | 0.781 | 0.463 | 1.339 | 3.60e-01 | 1.00e+00 | 1565 | 386 | 1179 |
| Latino/Mixed-American | Recorded Depression | Augmentation | Schizophrenia | Schizophrenia | 1.129 | 1.003 | 1.271 | 4.57e-02 | 3.20e-01 | 1565 | 386 | 1179 |
| Latino/Mixed-American | No Recorded Depression | Augmentation | ADHD | (Intercept) | 0.440 | 0.174 | 1.075 | 7.57e-02 | 5.30e-01 | 1406 | 129 | 1277 |
| Latino/Mixed-American | No Recorded Depression | Augmentation | ADHD | age_at_first_AD | 0.990 | 0.977 | 1.002 | 1.01e-01 | 7.10e-01 | 1406 | 129 | 1277 |
| Latino/Mixed-American | No Recorded Depression | Augmentation | ADHD | sex_at_birthMale | 2.252 | 1.535 | 3.292 | 2.92e-05 | 2.05e-04 | 1406 | 129 | 1277 |
| Latino/Mixed-American | No Recorded Depression | Augmentation | ADHD | Income_quantitative | 0.720 | 0.632 | 0.813 | 2.79e-07 | 1.95e-06 | 1406 | 129 | 1277 |
| Latino/Mixed-American | No Recorded Depression | Augmentation | ADHD | Education_levelCollege Undergraduate | 0.668 | 0.362 | 1.292 | 2.10e-01 | 1.00e+00 | 1406 | 129 | 1277 |
| Latino/Mixed-American | No Recorded Depression | Augmentation | ADHD | Education_levelLess than a high school degree | 0.429 | 0.200 | 0.942 | 3.17e-02 | 2.22e-01 | 1406 | 129 | 1277 |
| Latino/Mixed-American | No Recorded Depression | Augmentation | ADHD | Education_levelTwelve or GED | 0.622 | 0.302 | 1.316 | 2.03e-01 | 1.00e+00 | 1406 | 129 | 1277 |
| Latino/Mixed-American | No Recorded Depression | Augmentation | ADHD | ADHD | 1.109 | 0.906 | 1.359 | 3.15e-01 | 1.00e+00 | 1406 | 129 | 1277 |
| Latino/Mixed-American | No Recorded Depression | Augmentation | Migraine | (Intercept) | 0.457 | 0.181 | 1.122 | 9.20e-02 | 6.44e-01 | 1406 | 129 | 1277 |
| Latino/Mixed-American | No Recorded Depression | Augmentation | Migraine | age_at_first_AD | 0.989 | 0.976 | 1.001 | 8.01e-02 | 5.61e-01 | 1406 | 129 | 1277 |
| Latino/Mixed-American | No Recorded Depression | Augmentation | Migraine | sex_at_birthMale | 2.247 | 1.531 | 3.286 | 3.11e-05 | 2.18e-04 | 1406 | 129 | 1277 |
| Latino/Mixed-American | No Recorded Depression | Augmentation | Migraine | Income_quantitative | 0.716 | 0.628 | 0.809 | 2.05e-07 | 1.44e-06 | 1406 | 129 | 1277 |
| Latino/Mixed-American | No Recorded Depression | Augmentation | Migraine | Education_levelCollege Undergraduate | 0.672 | 0.364 | 1.301 | 2.18e-01 | 1.00e+00 | 1406 | 129 | 1277 |
| Latino/Mixed-American | No Recorded Depression | Augmentation | Migraine | Education_levelLess than a high school degree | 0.436 | 0.203 | 0.958 | 3.52e-02 | 2.47e-01 | 1406 | 129 | 1277 |
| Latino/Mixed-American | No Recorded Depression | Augmentation | Migraine | Education_levelTwelve or GED | 0.634 | 0.308 | 1.342 | 2.22e-01 | 1.00e+00 | 1406 | 129 | 1277 |
| Latino/Mixed-American | No Recorded Depression | Augmentation | Migraine | Migraine | 0.873 | 0.717 | 1.062 | 1.76e-01 | 1.00e+00 | 1406 | 129 | 1277 |
| Latino/Mixed-American | No Recorded Depression | Augmentation | Insomnia | (Intercept) | 0.442 | 0.175 | 1.078 | 7.69e-02 | 5.39e-01 | 1406 | 129 | 1277 |
| Latino/Mixed-American | No Recorded Depression | Augmentation | Insomnia | age_at_first_AD | 0.989 | 0.977 | 1.002 | 9.16e-02 | 6.41e-01 | 1406 | 129 | 1277 |
| Latino/Mixed-American | No Recorded Depression | Augmentation | Insomnia | sex_at_birthMale | 2.267 | 1.545 | 3.317 | 2.57e-05 | 1.80e-04 | 1406 | 129 | 1277 |
| Latino/Mixed-American | No Recorded Depression | Augmentation | Insomnia | Income_quantitative | 0.720 | 0.632 | 0.813 | 2.96e-07 | 2.07e-06 | 1406 | 129 | 1277 |
| Latino/Mixed-American | No Recorded Depression | Augmentation | Insomnia | Education_levelCollege Undergraduate | 0.671 | 0.364 | 1.298 | 2.16e-01 | 1.00e+00 | 1406 | 129 | 1277 |
| Latino/Mixed-American | No Recorded Depression | Augmentation | Insomnia | Education_levelLess than a high school degree | 0.433 | 0.202 | 0.950 | 3.35e-02 | 2.34e-01 | 1406 | 129 | 1277 |
| Latino/Mixed-American | No Recorded Depression | Augmentation | Insomnia | Education_levelTwelve or GED | 0.635 | 0.309 | 1.343 | 2.24e-01 | 1.00e+00 | 1406 | 129 | 1277 |
| Latino/Mixed-American | No Recorded Depression | Augmentation | Insomnia | Insomnia | 1.107 | 0.892 | 1.375 | 3.55e-01 | 1.00e+00 | 1406 | 129 | 1277 |
| Latino/Mixed-American | No Recorded Depression | Augmentation | BMI | (Intercept) | 0.449 | 0.178 | 1.097 | 8.34e-02 | 5.84e-01 | 1406 | 129 | 1277 |
| Latino/Mixed-American | No Recorded Depression | Augmentation | BMI | age_at_first_AD | 0.989 | 0.977 | 1.002 | 8.73e-02 | 6.11e-01 | 1406 | 129 | 1277 |
| Latino/Mixed-American | No Recorded Depression | Augmentation | BMI | sex_at_birthMale | 2.248 | 1.533 | 3.286 | 3.02e-05 | 2.11e-04 | 1406 | 129 | 1277 |
| Latino/Mixed-American | No Recorded Depression | Augmentation | BMI | Income_quantitative | 0.717 | 0.630 | 0.810 | 2.30e-07 | 1.61e-06 | 1406 | 129 | 1277 |
| Latino/Mixed-American | No Recorded Depression | Augmentation | BMI | Education_levelCollege Undergraduate | 0.674 | 0.366 | 1.304 | 2.21e-01 | 1.00e+00 | 1406 | 129 | 1277 |
| Latino/Mixed-American | No Recorded Depression | Augmentation | BMI | Education_levelLess than a high school degree | 0.438 | 0.204 | 0.961 | 3.59e-02 | 2.52e-01 | 1406 | 129 | 1277 |
| Latino/Mixed-American | No Recorded Depression | Augmentation | BMI | Education_levelTwelve or GED | 0.639 | 0.310 | 1.353 | 2.31e-01 | 1.00e+00 | 1406 | 129 | 1277 |

| | | | | | | | | | | | | |
|---|---|---|---|---|---|---|---|---|---|---|---|---|
| Latino/Mixed-American | No Recorded Depression | Augmentation | BMI | BMI | 0.945 | 0.780 | 1.145 | 5.66e-01 | 1.00e+00 | 1406 | 129 | 1277 |
| Latino/Mixed-American | No Recorded Depression | Augmentation | Bipolar Disorder | (Intercept) | 0.446 | 0.177 | 1.090 | 8.08e-02 | 5.65e-01 | 1406 | 129 | 1277 |
| Latino/Mixed-American | No Recorded Depression | Augmentation | Bipolar Disorder | age_at_first_AD | 0.989 | 0.977 | 1.002 | 9.19e-02 | 6.43e-01 | 1406 | 129 | 1277 |
| Latino/Mixed-American | No Recorded Depression | Augmentation | Bipolar Disorder | sex_at_birthMale | 2.241 | 1.527 | 3.277 | 3.34e-05 | 2.34e-04 | 1406 | 129 | 1277 |
| Latino/Mixed-American | No Recorded Depression | Augmentation | Bipolar Disorder | Income_quantitative | 0.718 | 0.630 | 0.811 | 2.46e-07 | 1.72e-06 | 1406 | 129 | 1277 |
| Latino/Mixed-American | No Recorded Depression | Augmentation | Bipolar Disorder | Education_levelCollege Undergraduate | 0.674 | 0.365 | 1.303 | 2.20e-01 | 1.00e+00 | 1406 | 129 | 1277 |
| Latino/Mixed-American | No Recorded Depression | Augmentation | Bipolar Disorder | Education_levelLess than a high school degree | 0.439 | 0.205 | 0.963 | 3.64e-02 | 2.55e-01 | 1406 | 129 | 1277 |
| Latino/Mixed-American | No Recorded Depression | Augmentation | Bipolar Disorder | Education_levelTwelve or GED | 0.635 | 0.308 | 1.343 | 2.24e-01 | 1.00e+00 | 1406 | 129 | 1277 |
| Latino/Mixed-American | No Recorded Depression | Augmentation | Bipolar Disorder | Bipolar Disorder | 0.975 | 0.791 | 1.200 | 8.08e-01 | 1.00e+00 | 1406 | 129 | 1277 |
| Latino/Mixed-American | No Recorded Depression | Augmentation | Depression | (Intercept) | 0.410 | 0.162 | 1.005 | 5.47e-02 | 3.83e-01 | 1406 | 129 | 1277 |
| Latino/Mixed-American | No Recorded Depression | Augmentation | Depression | age_at_first_AD | 0.990 | 0.977 | 1.002 | 1.05e-01 | 7.35e-01 | 1406 | 129 | 1277 |
| Latino/Mixed-American | No Recorded Depression | Augmentation | Depression | sex_at_birthMale | 2.268 | 1.544 | 3.321 | 2.67e-05 | 1.87e-04 | 1406 | 129 | 1277 |
| Latino/Mixed-American | No Recorded Depression | Augmentation | Depression | Income_quantitative | 0.726 | 0.637 | 0.820 | 5.62e-07 | 3.93e-06 | 1406 | 129 | 1277 |
| Latino/Mixed-American | No Recorded Depression | Augmentation | Depression | Education_levelCollege Undergraduate | 0.671 | 0.364 | 1.298 | 2.16e-01 | 1.00e+00 | 1406 | 129 | 1277 |
| Latino/Mixed-American | No Recorded Depression | Augmentation | Depression | Education_levelLess than a high school degree | 0.444 | 0.207 | 0.973 | 3.88e-02 | 2.72e-01 | 1406 | 129 | 1277 |
| Latino/Mixed-American | No Recorded Depression | Augmentation | Depression | Education_levelTwelve or GED | 0.655 | 0.318 | 1.384 | 2.56e-01 | 1.00e+00 | 1406 | 129 | 1277 |
| Latino/Mixed-American | No Recorded Depression | Augmentation | Depression | Depression | 1.342 | 1.086 | 1.663 | 6.67e-03 | 4.67e-02 | 1406 | 129 | 1277 |
| Latino/Mixed-American | No Recorded Depression | Augmentation | Schizophrenia | (Intercept) | 0.439 | 0.175 | 1.073 | 7.52e-02 | 5.26e-01 | 1406 | 129 | 1277 |
| Latino/Mixed-American | No Recorded Depression | Augmentation | Schizophrenia | age_at_first_AD | 0.989 | 0.977 | 1.002 | 8.64e-02 | 6.05e-01 | 1406 | 129 | 1277 |
| Latino/Mixed-American | No Recorded Depression | Augmentation | Schizophrenia | sex_at_birthMale | 2.258 | 1.539 | 3.301 | 2.76e-05 | 1.93e-04 | 1406 | 129 | 1277 |
| Latino/Mixed-American | No Recorded Depression | Augmentation | Schizophrenia | Income_quantitative | 0.720 | 0.632 | 0.813 | 2.93e-07 | 2.05e-06 | 1406 | 129 | 1277 |
| Latino/Mixed-American | No Recorded Depression | Augmentation | Schizophrenia | Education_levelCollege Undergraduate | 0.681 | 0.370 | 1.316 | 2.33e-01 | 1.00e+00 | 1406 | 129 | 1277 |
| Latino/Mixed-American | No Recorded Depression | Augmentation | Schizophrenia | Education_levelLess than a high school degree | 0.445 | 0.207 | 0.975 | 3.93e-02 | 2.75e-01 | 1406 | 129 | 1277 |
| Latino/Mixed-American | No Recorded Depression | Augmentation | Schizophrenia | Education_levelTwelve or GED | 0.645 | 0.313 | 1.364 | 2.40e-01 | 1.00e+00 | 1406 | 129 | 1277 |
| Latino/Mixed-American | No Recorded Depression | Augmentation | Schizophrenia | Schizophrenia | 1.075 | 0.894 | 1.295 | 4.42e-01 | 1.00e+00 | 1406 | 129 | 1277 |
| Latino/Mixed-American | Recorded Anxiety | Augmentation | ADHD | (Intercept) | 0.388 | 0.203 | 0.729 | 3.65e-03 | 2.55e-02 | 1861 | 399 | 1462 |
| Latino/Mixed-American | Recorded Anxiety | Augmentation | ADHD | age_at_first_AD | 0.998 | 0.990 | 1.006 | 5.60e-01 | 1.00e+00 | 1861 | 399 | 1462 |
| Latino/Mixed-American | Recorded Anxiety | Augmentation | ADHD | sex_at_birthMale | 1.586 | 1.241 | 2.023 | 2.12e-04 | 1.48e-03 | 1861 | 399 | 1462 |
| Latino/Mixed-American | Recorded Anxiety | Augmentation | ADHD | Income_quantitative | 0.833 | 0.771 | 0.897 | 2.37e-06 | 1.66e-05 | 1861 | 399 | 1462 |
| Latino/Mixed-American | Recorded Anxiety | Augmentation | ADHD | Education_levelCollege Undergraduate | 1.069 | 0.676 | 1.741 | 7.82e-01 | 1.00e+00 | 1861 | 399 | 1462 |
| Latino/Mixed-American | Recorded Anxiety | Augmentation | ADHD | Education_levelLess than a high school degree | 1.227 | 0.724 | 2.126 | 4.56e-01 | 1.00e+00 | 1861 | 399 | 1462 |
| Latino/Mixed-American | Recorded Anxiety | Augmentation | ADHD | Education_levelTwelve or GED | 0.887 | 0.530 | 1.517 | 6.54e-01 | 1.00e+00 | 1861 | 399 | 1462 |
| Latino/Mixed-American | Recorded Anxiety | Augmentation | ADHD | ADHD | 1.091 | 0.962 | 1.238 | 1.75e-01 | 1.00e+00 | 1861 | 399 | 1462 |
| Latino/Mixed-American | Recorded Anxiety | Augmentation | Migraine | (Intercept) | 0.392 | 0.205 | 0.738 | 4.12e-03 | 2.89e-02 | 1861 | 399 | 1462 |
| Latino/Mixed-American | Recorded Anxiety | Augmentation | Migraine | age_at_first_AD | 0.997 | 0.989 | 1.005 | 5.08e-01 | 1.00e+00 | 1861 | 399 | 1462 |
| Latino/Mixed-American | Recorded Anxiety | Augmentation | Migraine | sex_at_birthMale | 1.585 | 1.240 | 2.022 | 2.14e-04 | 1.50e-03 | 1861 | 399 | 1462 |
| Latino/Mixed-American | Recorded Anxiety | Augmentation | Migraine | Income_quantitative | 0.830 | 0.769 | 0.895 | 1.67e-06 | 1.17e-05 | 1861 | 399 | 1462 |
| Latino/Mixed-American | Recorded Anxiety | Augmentation | Migraine | Education_levelCollege Undergraduate | 1.080 | 0.683 | 1.758 | 7.49e-01 | 1.00e+00 | 1861 | 399 | 1462 |
| Latino/Mixed-American | Recorded Anxiety | Augmentation | Migraine | Education_levelLess than a high school degree | 1.241 | 0.732 | 2.150 | 4.31e-01 | 1.00e+00 | 1861 | 399 | 1462 |
| Latino/Mixed-American | Recorded Anxiety | Augmentation | Migraine | Education_levelTwelve or GED | 0.897 | 0.536 | 1.533 | 6.84e-01 | 1.00e+00 | 1861 | 399 | 1462 |
| Latino/Mixed-American | Recorded Anxiety | Augmentation | Migraine | Migraine | 1.023 | 0.909 | 1.152 | 7.02e-01 | 1.00e+00 | 1861 | 399 | 1462 |
| Latino/Mixed-American | Recorded Anxiety | Augmentation | Insomnia | (Intercept) | 0.387 | 0.202 | 0.728 | 3.59e-03 | 2.52e-02 | 1861 | 399 | 1462 |
| Latino/Mixed-American | Recorded Anxiety | Augmentation | Insomnia | age_at_first_AD | 0.998 | 0.990 | 1.006 | 5.49e-01 | 1.00e+00 | 1861 | 399 | 1462 |
| Latino/Mixed-American | Recorded Anxiety | Augmentation | Insomnia | sex_at_birthMale | 1.597 | 1.249 | 2.038 | 1.74e-04 | 1.22e-03 | 1861 | 399 | 1462 |
| Latino/Mixed-American | Recorded Anxiety | Augmentation | Insomnia | Income_quantitative | 0.832 | 0.770 | 0.896 | 2.03e-06 | 1.42e-05 | 1861 | 399 | 1462 |
| Latino/Mixed-American | Recorded Anxiety | Augmentation | Insomnia | Education_levelCollege Undergraduate | 1.070 | 0.677 | 1.744 | 7.78e-01 | 1.00e+00 | 1861 | 399 | 1462 |
| Latino/Mixed-American | Recorded Anxiety | Augmentation | Insomnia | Education_levelLess than a high school degree | 1.227 | 0.724 | 2.127 | 4.55e-01 | 1.00e+00 | 1861 | 399 | 1462 |
| Latino/Mixed-American | Recorded Anxiety | Augmentation | Insomnia | Education_levelTwelve or GED | 0.895 | 0.535 | 1.531 | 6.79e-01 | 1.00e+00 | 1861 | 399 | 1462 |
| Latino/Mixed-American | Recorded Anxiety | Augmentation | Insomnia | Insomnia | 1.154 | 1.013 | 1.315 | 3.10e-02 | 2.17e-01 | 1861 | 399 | 1462 |
| Latino/Mixed-American | Recorded Anxiety | Augmentation | BMI | (Intercept) | 0.392 | 0.205 | 0.737 | 4.05e-03 | 2.84e-02 | 1861 | 399 | 1462 |
| Latino/Mixed-American | Recorded Anxiety | Augmentation | BMI | age_at_first_AD | 0.997 | 0.989 | 1.005 | 5.06e-01 | 1.00e+00 | 1861 | 399 | 1462 |
| Latino/Mixed-American | Recorded Anxiety | Augmentation | BMI | sex_at_birthMale | 1.588 | 1.242 | 2.025 | 2.05e-04 | 1.43e-03 | 1861 | 399 | 1462 |
| Latino/Mixed-American | Recorded Anxiety | Augmentation | BMI | Income_quantitative | 0.829 | 0.767 | 0.893 | 1.38e-06 | 9.63e-06 | 1861 | 399 | 1462 |
| Latino/Mixed-American | Recorded Anxiety | Augmentation | BMI | Education_levelCollege Undergraduate | 1.086 | 0.687 | 1.769 | 7.30e-01 | 1.00e+00 | 1861 | 399 | 1462 |
| Latino/Mixed-American | Recorded Anxiety | Augmentation | BMI | Education_levelLess than a high school degree | 1.247 | 0.736 | 2.161 | 4.20e-01 | 1.00e+00 | 1861 | 399 | 1462 |

| | | | | | | | | | | | |
|---|---|---|---|---|---|---|---|---|---|---|---|
| Latino/Mixed-American | Recorded Anxiety | Augmentation | BMI | Education_levelTwelve or GED | 0.904 | 0.541 | 1.545 | 7.05e-01 | 1.00e+00 | 1861 | 399 | 1462 |
| Latino/Mixed-American | Recorded Anxiety | Augmentation | BMI | BMI | 0.956 | 0.852 | 1.072 | 4.38e-01 | 1.00e+00 | 1861 | 399 | 1462 |
| Latino/Mixed-American | Recorded Anxiety | Augmentation | Bipolar Disorder | (Intercept) | 0.388 | 0.203 | 0.730 | 3.71e-03 | 2.60e-02 | 1861 | 399 | 1462 |
| Latino/Mixed-American | Recorded Anxiety | Augmentation | Bipolar Disorder | age_at_first_AD | 0.997 | 0.989 | 1.005 | 4.83e-01 | 1.00e+00 | 1861 | 399 | 1462 |
| Latino/Mixed-American | Recorded Anxiety | Augmentation | Bipolar Disorder | sex_at_birthMale | 1.607 | 1.257 | 2.051 | 1.44e-04 | 1.01e-03 | 1861 | 399 | 1462 |
| Latino/Mixed-American | Recorded Anxiety | Augmentation | Bipolar Disorder | Income_quantitative | 0.833 | 0.771 | 0.897 | 2.39e-06 | 1.67e-05 | 1861 | 399 | 1462 |
| Latino/Mixed-American | Recorded Anxiety | Augmentation | Bipolar Disorder | Education_levelCollege Undergraduate | 1.074 | 0.680 | 1.750 | 7.65e-01 | 1.00e+00 | 1861 | 399 | 1462 |
| Latino/Mixed-American | Recorded Anxiety | Augmentation | Bipolar Disorder | Education_levelLess than a high school degree | 1.250 | 0.737 | 2.165 | 4.16e-01 | 1.00e+00 | 1861 | 399 | 1462 |
| Latino/Mixed-American | Recorded Anxiety | Augmentation | Bipolar Disorder | Education_levelTwelve or GED | 0.906 | 0.541 | 1.548 | 7.10e-01 | 1.00e+00 | 1861 | 399 | 1462 |
| Latino/Mixed-American | Recorded Anxiety | Augmentation | Bipolar Disorder | Bipolar Disorder | 1.184 | 1.042 | 1.345 | 9.52e-03 | 6.67e-02 | 1861 | 399 | 1462 |
| Latino/Mixed-American | Recorded Anxiety | Augmentation | Depression | (Intercept) | 0.383 | 0.200 | 0.722 | 3.34e-03 | 2.34e-02 | 1861 | 399 | 1462 |
| Latino/Mixed-American | Recorded Anxiety | Augmentation | Depression | age_at_first_AD | 0.998 | 0.990 | 1.005 | 5.38e-01 | 1.00e+00 | 1861 | 399 | 1462 |
| Latino/Mixed-American | Recorded Anxiety | Augmentation | Depression | sex_at_birthMale | 1.600 | 1.250 | 2.043 | 1.73e-04 | 1.21e-03 | 1861 | 399 | 1462 |
| Latino/Mixed-American | Recorded Anxiety | Augmentation | Depression | Income_quantitative | 0.834 | 0.772 | 0.899 | 3.08e-06 | 2.16e-05 | 1861 | 399 | 1462 |
| Latino/Mixed-American | Recorded Anxiety | Augmentation | Depression | Education_levelCollege Undergraduate | 1.058 | 0.669 | 1.725 | 8.14e-01 | 1.00e+00 | 1861 | 399 | 1462 |
| Latino/Mixed-American | Recorded Anxiety | Augmentation | Depression | Education_levelLess than a high school degree | 1.227 | 0.724 | 2.128 | 4.55e-01 | 1.00e+00 | 1861 | 399 | 1462 |
| Latino/Mixed-American | Recorded Anxiety | Augmentation | Depression | Education_levelTwelve or GED | 0.888 | 0.531 | 1.519 | 6.57e-01 | 1.00e+00 | 1861 | 399 | 1462 |
| Latino/Mixed-American | Recorded Anxiety | Augmentation | Depression | Depression | 1.307 | 1.147 | 1.491 | 6.20e-05 | 4.34e-04 | 1861 | 399 | 1462 |
| Latino/Mixed-American | Recorded Anxiety | Augmentation | Schizophrenia | (Intercept) | 0.388 | 0.203 | 0.730 | 3.69e-03 | 2.58e-02 | 1861 | 399 | 1462 |
| Latino/Mixed-American | Recorded Anxiety | Augmentation | Schizophrenia | age_at_first_AD | 0.997 | 0.989 | 1.005 | 4.76e-01 | 1.00e+00 | 1861 | 399 | 1462 |
| Latino/Mixed-American | Recorded Anxiety | Augmentation | Schizophrenia | sex_at_birthMale | 1.593 | 1.246 | 2.031 | 1.88e-04 | 1.32e-03 | 1861 | 399 | 1462 |
| Latino/Mixed-American | Recorded Anxiety | Augmentation | Schizophrenia | Income_quantitative | 0.833 | 0.771 | 0.897 | 2.39e-06 | 1.67e-05 | 1861 | 399 | 1462 |
| Latino/Mixed-American | Recorded Anxiety | Augmentation | Schizophrenia | Education_levelCollege Undergraduate | 1.090 | 0.689 | 1.774 | 7.21e-01 | 1.00e+00 | 1861 | 399 | 1462 |
| Latino/Mixed-American | Recorded Anxiety | Augmentation | Schizophrenia | Education_levelLess than a high school degree | 1.254 | 0.741 | 2.172 | 4.08e-01 | 1.00e+00 | 1861 | 399 | 1462 |
| Latino/Mixed-American | Recorded Anxiety | Augmentation | Schizophrenia | Education_levelTwelve or GED | 0.907 | 0.543 | 1.550 | 7.15e-01 | 1.00e+00 | 1861 | 399 | 1462 |
| Latino/Mixed-American | Recorded Anxiety | Augmentation | Schizophrenia | Schizophrenia | 1.092 | 0.975 | 1.224 | 1.27e-01 | 8.90e-01 | 1861 | 399 | 1462 |

**Supplementary Table 11. Multivariable logistic regression results for PGS associations with antidepressant augmentation across clinical subgroups in the Pharmlines cohort (European ancestry participants).** Each model estimates the association between 7 PGS traits and the binary outcome augmentation versus continuation — a higher-intensity treatment outcome defined at the level of the individual's overall prescribing history, in which an antidepressant episode overlapped with an atypical antipsychotic or lithium episode by at least 28 days at any point — fitted separately within each combination of clinical group (Full Cohort and subgroups defined by recorded depression status), and PGS exposure. Within each model, one PGS exposure is modelled alongside a common covariate set (age at first antidepressant, genetic sex, income, and educational attainment). All PGS were residualised on the top 20 within-European-ancestry principal components and the residuals standardised to mean 0, SD 1; each odds ratio therefore reflects the change in odds per one-standard-deviation increase in the PC-adjusted PGS. Because PCs were regressed out at this stage, they are not included in the model covariate set. Columns: Population = inferred genetic ancestry (all European); ClinicalGroup = cohort or clinical subgroup; Outcome = the binary contrast modelled; PGS = the PGS trait for that model; variable = the individual model term; odds_ratio = exponentiated coefficient; lower_95/upper_95 = 95% confidence interval bounds for the odds ratio; p_value = nominal (uncorrected) p-value; Bonf_P = Bonferroni-corrected p-value (capped at 1.00); N_Total = model sample size; N_Reference = number in the reference (continuation) group; N_Case = number in the augmentation group. Odds ratios >1 indicate higher odds of augmentation relative to continuation. Bonferroni correction was applied within each specification (clinical group) for the max 7 PGS traits tested. P-values reported as "<2.2e-16" reflect R's floating-point reporting limit.
Reference (omitted) categories: Sex at birth: Female.

| Population | ClinicalGroup | Outcome | PGS | variable | odds_ratio | lower_95 | upper_95 | p_value | Bonf_p | N_Total | N_Cases | N_Controls |
|---|---|---|---|---|---|---|---|---|---|---|---|---|
| European | Full Cohort | Augmentation | ADHD | (Intercept) | 0.3977 | 0.2172 | 0.7269 | 2.76e-03 | 1.93e-02 | 4565 | 357 | 4208 |
| European | Full Cohort | Augmentation | ADHD | age_at_first_AD | 0.9769 | 0.9677 | 0.986 | 9.37e-07 | 6.56e-06 | 4565 | 357 | 4208 |
| European | Full Cohort | Augmentation | ADHD | sex_at_birth | 1.53 | 1.213 | 1.921 | 2.87e-04 | 2.01e-03 | 4565 | 357 | 4208 |
| European | Full Cohort | Augmentation | ADHD | Income_level | 0.9997 | 0.9995 | 0.9999 | 5.54e-03 | 3.88e-02 | 4565 | 357 | 4208 |
| European | Full Cohort | Augmentation | ADHD | Education_level | 0.9842 | 0.9562 | 1.013 | 2.77e-01 | 1.00e+00 | 4565 | 357 | 4208 |
| European | Full Cohort | Augmentation | ADHD | ADHD | 1.002 | 0.8983 | 1.118 | 9.68e-01 | 1.00e+00 | 4565 | 357 | 4208 |
| European | Full Cohort | Augmentation | Bipolar Disorder | (Intercept) | 0.3905 | 0.2134 | 0.7133 | 2.25e-03 | 1.57e-02 | 4565 | 357 | 4208 |
| European | Full Cohort | Augmentation | Bipolar Disorder | age_at_first_AD | 0.9772 | 0.968 | 0.9863 | 1.33e-06 | 9.33e-06 | 4565 | 357 | 4208 |
| European | Full Cohort | Augmentation | Bipolar Disorder | sex_at_birth | 1.533 | 1.216 | 1.926 | 2.68e-04 | 1.88e-03 | 4565 | 357 | 4208 |
| European | Full Cohort | Augmentation | Bipolar Disorder | Income_level | 0.9997 | 0.9995 | 0.9999 | 5.58e-03 | 3.90e-02 | 4565 | 357 | 4208 |
| European | Full Cohort | Augmentation | Bipolar Disorder | Education_level | 0.9838 | 0.9558 | 1.012 | 2.62e-01 | 1.00e+00 | 4565 | 357 | 4208 |
| European | Full Cohort | Augmentation | Bipolar Disorder | Bipolar Disorder | 1.173 | 1.052 | 1.309 | 4.00e-03 | 2.80e-02 | 4565 | 357 | 4208 |
| European | Full Cohort | Augmentation | Migraine | (Intercept) | 0.3962 | 0.2169 | 0.7229 | 2.57e-03 | 1.80e-02 | 4565 | 357 | 4208 |
| European | Full Cohort | Augmentation | Migraine | age_at_first_AD | 0.9769 | 0.9677 | 0.986 | 9.29e-07 | 6.51e-06 | 4565 | 357 | 4208 |
| European | Full Cohort | Augmentation | Migraine | sex_at_birth | 1.529 | 1.212 | 1.92 | 2.94e-04 | 2.05e-03 | 4565 | 357 | 4208 |
| European | Full Cohort | Augmentation | Migraine | Income_level | 0.9997 | 0.9995 | 0.9999 | 5.80e-03 | 4.06e-02 | 4565 | 357 | 4208 |
| European | Full Cohort | Augmentation | Migraine | Education_level | 0.9843 | 0.9563 | 1.013 | 2.77e-01 | 1.00e+00 | 4565 | 357 | 4208 |
| European | Full Cohort | Augmentation | Migraine | Migraine | 1.029 | 0.9238 | 1.147 | 6.02e-01 | 1.00e+00 | 4565 | 357 | 4208 |
| European | Full Cohort | Augmentation | Schizophrenia | (Intercept) | 0.3911 | 0.2138 | 0.7144 | 2.28e-03 | 1.60e-02 | 4565 | 357 | 4208 |
| European | Full Cohort | Augmentation | Schizophrenia | age_at_first_AD | 0.9773 | 0.9681 | 0.9864 | 1.53e-06 | 1.07e-05 | 4565 | 357 | 4208 |
| European | Full Cohort | Augmentation | Schizophrenia | sex_at_birth | 1.523 | 1.207 | 1.913 | 3.37e-04 | 2.36e-03 | 4565 | 357 | 4208 |
| European | Full Cohort | Augmentation | Schizophrenia | Income_level | 0.9997 | 0.9995 | 0.9999 | 5.38e-03 | 3.77e-02 | 4565 | 357 | 4208 |
| European | Full Cohort | Augmentation | Schizophrenia | Education_level | 0.9837 | 0.9558 | 1.012 | 2.61e-01 | 1.00e+00 | 4565 | 357 | 4208 |
| European | Full Cohort | Augmentation | Schizophrenia | Schizophrenia | 1.145 | 1.028 | 1.276 | 1.41e-02 | 9.88e-02 | 4565 | 357 | 4208 |
| European | Full Cohort | Augmentation | BMI | (Intercept) | 0.41 | 0.2243 | 0.7482 | 3.71e-03 | 2.60e-02 | 4565 | 357 | 4208 |
| European | Full Cohort | Augmentation | BMI | age_at_first_AD | 0.9767 | 0.9675 | 0.9858 | 7.57e-07 | 5.30e-06 | 4565 | 357 | 4208 |
| European | Full Cohort | Augmentation | BMI | sex_at_birth | 1.524 | 1.208 | 1.914 | 3.29e-04 | 2.30e-03 | 4565 | 357 | 4208 |
| European | Full Cohort | Augmentation | BMI | Income_level | 0.9997 | 0.9995 | 0.9999 | 5.37e-03 | 3.76e-02 | 4565 | 357 | 4208 |
| European | Full Cohort | Augmentation | BMI | Education_level | 0.9827 | 0.9548 | 1.011 | 2.33e-01 | 1.00e+00 | 4565 | 357 | 4208 |
| European | Full Cohort | Augmentation | BMI | BMI | 0.9255 | 0.8306 | 1.031 | 1.61e-01 | 1.00e+00 | 4565 | 357 | 4208 |
| European | Full Cohort | Augmentation | Depression | (Intercept) | 0.3918 | 0.2141 | 0.7156 | 2.32e-03 | 1.63e-02 | 4565 | 357 | 4208 |

| | | | | | | | | | | | | |
|---|---|---|---|---|---|---|---|---|---|---|---|---|
| European | Full Cohort | Augmentation | Depression | age_at_first_AD | 0.9771 | 0.9679 | 0.9862 | 1.20e-06 | 8.43e-06 | 4565 | 357 | 4208 |
| European | Full Cohort | Augmentation | Depression | sex_at_birth | 1.527 | 1.211 | 1.918 | 3.03e-04 | 2.12e-03 | 4565 | 357 | 4208 |
| European | Full Cohort | Augmentation | Depression | Income_level | 0.9997 | 0.9995 | 0.9999 | 5.69e-03 | 3.98e-02 | 4565 | 357 | 4208 |
| European | Full Cohort | Augmentation | Depression | Education_level | 0.9845 | 0.9565 | 1.013 | 2.84e-01 | 1.00e+00 | 4565 | 357 | 4208 |
| European | Full Cohort | Augmentation | Depression | Depression | 1.048 | 0.9401 | 1.169 | 3.95e-01 | 1.00e+00 | 4565 | 357 | 4208 |
| European | Full Cohort | Augmentation | Insomnia | (Intercept) | 0.4007 | 0.219 | 0.7321 | 2.96e-03 | 2.07e-02 | 4565 | 357 | 4208 |
| European | Full Cohort | Augmentation | Insomnia | age_at_first_AD | 0.9768 | 0.9677 | 0.9859 | 8.84e-07 | 6.18e-06 | 4565 | 357 | 4208 |
| European | Full Cohort | Augmentation | Insomnia | sex_at_birth | 1.53 | 1.214 | 1.922 | 2.82e-04 | 1.98e-03 | 4565 | 357 | 4208 |
| European | Full Cohort | Augmentation | Insomnia | Income_level | 0.9997 | 0.9995 | 0.9999 | 5.35e-03 | 3.75e-02 | 4565 | 357 | 4208 |
| European | Full Cohort | Augmentation | Insomnia | Education_level | 0.984 | 0.956 | 1.012 | 2.69e-01 | 1.00e+00 | 4565 | 357 | 4208 |
| European | Full Cohort | Augmentation | Insomnia | Insomnia | 0.9811 | 0.8789 | 1.095 | 7.34e-01 | 1.00e+00 | 4565 | 357 | 4208 |
| European | Recorded Depression | Augmentation | ADHD | (Intercept) | 0.7116 | 0.3104 | 1.629 | 4.21e-01 | 1.00e+00 | 1796 | 200 | 1596 |
| European | Recorded Depression | Augmentation | ADHD | age_at_first_AD | 0.9861 | 0.9726 | 0.9997 | 4.62e-02 | 3.24e-01 | 1796 | 200 | 1596 |
| European | Recorded Depression | Augmentation | ADHD | sex_at_birth | 1.447 | 1.041 | 1.994 | 2.55e-02 | 1.79e-01 | 1796 | 200 | 1596 |
| European | Recorded Depression | Augmentation | ADHD | Income_level | 0.9996 | 0.9993 | 0.9999 | 1.24e-02 | 8.66e-02 | 1796 | 200 | 1596 |
| European | Recorded Depression | Augmentation | ADHD | Education_level | 0.9524 | 0.9147 | 0.9906 | 1.62e-02 | 1.13e-01 | 1796 | 200 | 1596 |
| European | Recorded Depression | Augmentation | ADHD | ADHD | 0.9562 | 0.8223 | 1.112 | 5.60e-01 | 1.00e+00 | 1796 | 200 | 1596 |
| European | Recorded Depression | Augmentation | Bipolar Disorder | (Intercept) | 0.6922 | 0.3024 | 1.582 | 3.83e-01 | 1.00e+00 | 1796 | 200 | 1596 |
| European | Recorded Depression | Augmentation | Bipolar Disorder | age_at_first_AD | 0.9863 | 0.9728 | 1 | 4.97e-02 | 3.48e-01 | 1796 | 200 | 1596 |
| European | Recorded Depression | Augmentation | Bipolar Disorder | sex_at_birth | 1.442 | 1.037 | 1.987 | 2.71e-02 | 1.90e-01 | 1796 | 200 | 1596 |
| European | Recorded Depression | Augmentation | Bipolar Disorder | Income_level | 0.9996 | 0.9993 | 0.9999 | 1.45e-02 | 1.02e-01 | 1796 | 200 | 1596 |
| European | Recorded Depression | Augmentation | Bipolar Disorder | Education_level | 0.9524 | 0.915 | 0.9905 | 1.59e-02 | 1.11e-01 | 1796 | 200 | 1596 |
| European | Recorded Depression | Augmentation | Bipolar Disorder | Bipolar Disorder | 1.125 | 0.9661 | 1.31 | 1.30e-01 | 9.07e-01 | 1796 | 200 | 1596 |
| European | Recorded Depression | Augmentation | Migraine | (Intercept) | 0.6986 | 0.3052 | 1.596 | 3.95e-01 | 1.00e+00 | 1796 | 200 | 1596 |
| European | Recorded Depression | Augmentation | Migraine | age_at_first_AD | 0.9862 | 0.9727 | 0.9998 | 4.75e-02 | 3.33e-01 | 1796 | 200 | 1596 |
| European | Recorded Depression | Augmentation | Migraine | sex_at_birth | 1.442 | 1.038 | 1.987 | 2.68e-02 | 1.88e-01 | 1796 | 200 | 1596 |
| European | Recorded Depression | Augmentation | Migraine | Income_level | 0.9996 | 0.9993 | 0.9999 | 1.30e-02 | 9.07e-02 | 1796 | 200 | 1596 |
| European | Recorded Depression | Augmentation | Migraine | Education_level | 0.9532 | 0.9156 | 0.9913 | 1.78e-02 | 1.25e-01 | 1796 | 200 | 1596 |
| European | Recorded Depression | Augmentation | Migraine | Migraine | 1.01 | 0.8705 | 1.173 | 8.94e-01 | 1.00e+00 | 1796 | 200 | 1596 |
| European | Recorded Depression | Augmentation | Schizophrenia | (Intercept) | 0.6973 | 0.3045 | 1.594 | 3.93e-01 | 1.00e+00 | 1796 | 200 | 1596 |
| European | Recorded Depression | Augmentation | Schizophrenia | age_at_first_AD | 0.9863 | 0.9728 | 1 | 4.98e-02 | 3.49e-01 | 1796 | 200 | 1596 |
| European | Recorded Depression | Augmentation | Schizophrenia | sex_at_birth | 1.44 | 1.036 | 1.984 | 2.75e-02 | 1.93e-01 | 1796 | 200 | 1596 |
| European | Recorded Depression | Augmentation | Schizophrenia | Income_level | 0.9996 | 0.9993 | 0.9999 | 1.29e-02 | 9.00e-02 | 1796 | 200 | 1596 |
| European | Recorded Depression | Augmentation | Schizophrenia | Education_level | 0.953 | 0.9154 | 0.9911 | 1.73e-02 | 1.21e-01 | 1796 | 200 | 1596 |
| European | Recorded Depression | Augmentation | Schizophrenia | Schizophrenia | 1.053 | 0.9086 | 1.22 | 4.96e-01 | 1.00e+00 | 1796 | 200 | 1596 |
| European | Recorded Depression | Augmentation | BMI | (Intercept) | 0.7204 | 0.3153 | 1.644 | 4.36e-01 | 1.00e+00 | 1796 | 200 | 1596 |
| European | Recorded Depression | Augmentation | BMI | age_at_first_AD | 0.9859 | 0.9724 | 0.9995 | 4.32e-02 | 3.02e-01 | 1796 | 200 | 1596 |
| European | Recorded Depression | Augmentation | BMI | sex_at_birth | 1.433 | 1.031 | 1.975 | 2.99e-02 | 2.09e-01 | 1796 | 200 | 1596 |
| European | Recorded Depression | Augmentation | BMI | Income_level | 0.9996 | 0.9993 | 0.9999 | 1.42e-02 | 9.94e-02 | 1796 | 200 | 1596 |
| European | Recorded Depression | Augmentation | BMI | Education_level | 0.9513 | 0.9138 | 0.9895 | 1.39e-02 | 9.73e-02 | 1796 | 200 | 1596 |
| European | Recorded Depression | Augmentation | BMI | BMI | 0.8997 | 0.7758 | 1.043 | 1.62e-01 | 1.00e+00 | 1796 | 200 | 1596 |
| European | Recorded Depression | Augmentation | Depression | (Intercept) | 0.6987 | 0.3049 | 1.598 | 3.96e-01 | 1.00e+00 | 1796 | 200 | 1596 |
| European | Recorded Depression | Augmentation | Depression | age_at_first_AD | 0.9862 | 0.9726 | 0.9998 | 4.79e-02 | 3.35e-01 | 1796 | 200 | 1596 |
| European | Recorded Depression | Augmentation | Depression | sex_at_birth | 1.442 | 1.037 | 1.987 | 2.72e-02 | 1.90e-01 | 1796 | 200 | 1596 |

| | | | | | | | | | | | | |
|---|---|---|---|---|---|---|---|---|---|---|---|---|
| European | Recorded Depression | Augmentation | Depression | Income_level | 0.9996 | 0.9993 | 0.9999 | 1.29e-02 | 9.00e-02 | 1796 | 200 | 1596 |
| European | Recorded Depression | Augmentation | Depression | Education_level | 0.9532 | 0.9156 | 0.9914 | 1.81e-02 | 1.27e-01 | 1796 | 200 | 1596 |
| European | Recorded Depression | Augmentation | Depression | Depression | 1.005 | 0.8627 | 1.171 | 9.49e-01 | 1.00e+00 | 1796 | 200 | 1596 |
| European | Recorded Depression | Augmentation | Insomnia | (Intercept) | 0.708 | 0.309 | 1.62 | 4.14e-01 | 1.00e+00 | 1796 | 200 | 1596 |
| European | Recorded Depression | Augmentation | Insomnia | age_at_first_AD | 0.9861 | 0.9725 | 0.9997 | 4.56e-02 | 3.19e-01 | 1796 | 200 | 1596 |
| European | Recorded Depression | Augmentation | Insomnia | sex_at_birth | 1.444 | 1.039 | 1.99 | 2.62e-02 | 1.83e-01 | 1796 | 200 | 1596 |
| European | Recorded Depression | Augmentation | Insomnia | Income_level | 0.9996 | 0.9993 | 0.9999 | 1.27e-02 | 8.87e-02 | 1796 | 200 | 1596 |
| European | Recorded Depression | Augmentation | Insomnia | Education_level | 0.9527 | 0.9151 | 0.9909 | 1.68e-02 | 1.18e-01 | 1796 | 200 | 1596 |
| European | Recorded Depression | Augmentation | Insomnia | Insomnia | 0.9597 | 0.8256 | 1.115 | 5.92e-01 | 1.00e+00 | 1796 | 200 | 1596 |
| European | No Recorded Depression | Augmentation | ADHD | (Intercept) | 0.1674 | 0.06605 | 0.4223 | 1.58e-04 | 1.11e-03 | 2719 | 147 | 2572 |
| European | No Recorded Depression | Augmentation | ADHD | age_at_first_AD | 0.9762 | 0.9627 | 0.9897 | 6.44e-04 | 4.51e-03 | 2719 | 147 | 2572 |
| European | No Recorded Depression | Augmentation | ADHD | sex_at_birth | 1.86 | 1.318 | 2.613 | 3.66e-04 | 2.56e-03 | 2719 | 147 | 2572 |
| European | No Recorded Depression | Augmentation | ADHD | Income_level | 0.9998 | 0.9995 | 1 | 2.22e-01 | 1.00e+00 | 2719 | 147 | 2572 |
| European | No Recorded Depression | Augmentation | ADHD | Education_level | 1.005 | 0.9612 | 1.049 | 8.37e-01 | 1.00e+00 | 2719 | 147 | 2572 |
| European | No Recorded Depression | Augmentation | ADHD | ADHD | 1.039 | 0.878 | 1.23 | 6.57e-01 | 1.00e+00 | 2719 | 147 | 2572 |
| European | No Recorded Depression | Augmentation | Bipolar Disorder | (Intercept) | 0.1637 | 0.06444 | 0.4137 | 1.35e-04 | 9.47e-04 | 2719 | 147 | 2572 |
| European | No Recorded Depression | Augmentation | Bipolar Disorder | age_at_first_AD | 0.9766 | 0.963 | 0.9902 | 8.46e-04 | 5.92e-03 | 2719 | 147 | 2572 |
| European | No Recorded Depression | Augmentation | Bipolar Disorder | sex_at_birth | 1.873 | 1.326 | 2.631 | 3.20e-04 | 2.24e-03 | 2719 | 147 | 2572 |
| European | No Recorded Depression | Augmentation | Bipolar Disorder | Income_level | 0.9998 | 0.9995 | 1 | 2.08e-01 | 1.00e+00 | 2719 | 147 | 2572 |
| European | No Recorded Depression | Augmentation | Bipolar Disorder | Education_level | 1.004 | 0.9609 | 1.049 | 8.51e-01 | 1.00e+00 | 2719 | 147 | 2572 |
| European | No Recorded Depression | Augmentation | Bipolar Disorder | Bipolar Disorder | 1.218 | 1.032 | 1.438 | 1.97e-02 | 1.38e-01 | 2719 | 147 | 2572 |
| European | No Recorded Depression | Augmentation | Migraine | (Intercept) | 0.1681 | 0.06658 | 0.4225 | 1.54e-04 | 1.08e-03 | 2719 | 147 | 2572 |
| European | No Recorded Depression | Augmentation | Migraine | age_at_first_AD | 0.9761 | 0.9625 | 0.9895 | 5.97e-04 | 4.18e-03 | 2719 | 147 | 2572 |
| European | No Recorded Depression | Augmentation | Migraine | sex_at_birth | 1.857 | 1.316 | 2.609 | 3.82e-04 | 2.67e-03 | 2719 | 147 | 2572 |
| European | No Recorded Depression | Augmentation | Migraine | Income_level | 0.9998 | 0.9995 | 1 | 2.32e-01 | 1.00e+00 | 2719 | 147 | 2572 |
| European | No Recorded Depression | Augmentation | Migraine | Education_level | 1.004 | 0.9608 | 1.048 | 8.58e-01 | 1.00e+00 | 2719 | 147 | 2572 |
| European | No Recorded Depression | Augmentation | Migraine | Migraine | 1.095 | 0.9279 | 1.292 | 2.84e-01 | 1.00e+00 | 2719 | 147 | 2572 |
| European | No Recorded Depression | Augmentation | Schizophrenia | (Intercept) | 0.1644 | 0.06473 | 0.4151 | 1.38e-04 | 9.69e-04 | 2719 | 147 | 2572 |
| European | No Recorded Depression | Augmentation | Schizophrenia | age_at_first_AD | 0.9769 | 0.9633 | 0.9904 | 9.42e-04 | 6.60e-03 | 2719 | 147 | 2572 |
| European | No Recorded Depression | Augmentation | Schizophrenia | sex_at_birth | 1.842 | 1.305 | 2.589 | 4.60e-04 | 3.22e-03 | 2719 | 147 | 2572 |
| European | No Recorded Depression | Augmentation | Schizophrenia | Income_level | 0.9998 | 0.9995 | 1 | 2.16e-01 | 1.00e+00 | 2719 | 147 | 2572 |
| European | No Recorded Depression | Augmentation | Schizophrenia | Education_level | 1.003 | 0.9593 | 1.047 | 9.08e-01 | 1.00e+00 | 2719 | 147 | 2572 |
| European | No Recorded Depression | Augmentation | Schizophrenia | Schizophrenia | 1.258 | 1.065 | 1.488 | 6.99e-03 | 4.89e-02 | 2719 | 147 | 2572 |
| European | No Recorded Depression | Augmentation | BMI | (Intercept) | 0.1757 | 0.06935 | 0.4427 | 2.34e-04 | 1.64e-03 | 2719 | 147 | 2572 |
| European | No Recorded Depression | Augmentation | BMI | age_at_first_AD | 0.976 | 0.9624 | 0.9894 | 5.62e-04 | 3.94e-03 | 2719 | 147 | 2572 |
| European | No Recorded Depression | Augmentation | BMI | sex_at_birth | 1.855 | 1.314 | 2.605 | 3.91e-04 | 2.74e-03 | 2719 | 147 | 2572 |
| European | No Recorded Depression | Augmentation | BMI | Income_level | 0.9998 | 0.9995 | 1 | 2.09e-01 | 1.00e+00 | 2719 | 147 | 2572 |
| European | No Recorded Depression | Augmentation | BMI | Education_level | 1.003 | 0.9593 | 1.047 | 9.07e-01 | 1.00e+00 | 2719 | 147 | 2572 |
| European | No Recorded Depression | Augmentation | BMI | BMI | 0.9417 | 0.7978 | 1.113 | 4.79e-01 | 1.00e+00 | 2719 | 147 | 2572 |
| European | No Recorded Depression | Augmentation | Depression | (Intercept) | 0.1689 | 0.0668 | 0.4251 | 1.64e-04 | 1.15e-03 | 2719 | 147 | 2572 |
| European | No Recorded Depression | Augmentation | Depression | age_at_first_AD | 0.9762 | 0.9627 | 0.9897 | 6.62e-04 | 4.64e-03 | 2719 | 147 | 2572 |
| European | No Recorded Depression | Augmentation | Depression | sex_at_birth | 1.86 | 1.318 | 2.612 | 3.67e-04 | 2.57e-03 | 2719 | 147 | 2572 |
| European | No Recorded Depression | Augmentation | Depression | Income_level | 0.9998 | 0.9995 | 1 | 2.21e-01 | 1.00e+00 | 2719 | 147 | 2572 |
| European | No Recorded Depression | Augmentation | Depression | Education_level | 1.004 | 0.9606 | 1.048 | 8.64e-01 | 1.00e+00 | 2719 | 147 | 2572 |

| | | | | | | | | | | | | |
|---|---|---|---|---|---|---|---|---|---|---|---|---|
| European | No Recorded Depression | Augmentation | Depression | Depression | 1.039 | 0.8798 | 1.227 | 6.50e-01 | 1.00e+00 | 2719 | 147 | 2572 |
| European | No Recorded Depression | Augmentation | Insomnia | (Intercept) | 0.1712 | 0.06766 | 0.4316 | 1.88e-04 | 1.31e-03 | 2719 | 147 | 2572 |
| European | No Recorded Depression | Augmentation | Insomnia | age_at_first_AD | 0.9761 | 0.9625 | 0.9896 | 6.04e-04 | 4.23e-03 | 2719 | 147 | 2572 |
| European | No Recorded Depression | Augmentation | Insomnia | sex_at_birth | 1.861 | 1.318 | 2.613 | 3.65e-04 | 2.56e-03 | 2719 | 147 | 2572 |
| European | No Recorded Depression | Augmentation | Insomnia | Income_level | 0.9998 | 0.9995 | 1 | 2.14e-01 | 1.00e+00 | 2719 | 147 | 2572 |
| European | No Recorded Depression | Augmentation | Insomnia | Education_level | 1.004 | 0.9606 | 1.048 | 8.64e-01 | 1.00e+00 | 2719 | 147 | 2572 |
| European | No Recorded Depression | Augmentation | Insomnia | Insomnia | 0.9903 | 0.836 | 1.173 | 9.10e-01 | 1.00e+00 | 2719 | 147 | 2572 |

**Supplementary Table 12. Ordinal (proportional-odds) logistic regression results for PGS associations with antidepressant treatment-escalation severity across ancestry groups and clinical subgroups in All of Us.** Each model estimates the association between 7 PGS traits and a single ordered three-level outcome reflecting increasing treatment-escalation intensity — continuation < switching < augmentation — fitted separately within each combination of genetically inferred ancestry (European, African, Latino/Mixed-American), clinical group (Full Cohort; subgroups defined by recorded depression status, recorded anxiety, and CIDI-SF–derived MDD classification), and PGS exposure. The proportional-odds model estimates a single odds ratio per predictor for the odds of falling in a higher (more escalated) versus lower outcome category, assumed constant across both category thresholds (continuation vs. switching-or-augmentation, and continuation-or-switching vs. augmentation). Within each model, one PGS exposure is modelled alongside a common covariate set (age at first antidepressant, sex at birth, income, and educational attainment). All PGS were residualised on the top 16 global genetic ancestry principal components and the residuals standardised to mean 0, SD 1; each odds ratio therefore reflects the change in odds of a more escalated outcome per one-standard-deviation increase in the PC-adjusted PGS. Because ancestry PCs were regressed out at this stage, they are not included in the model covariate set. Columns: Population = inferred genetic ancestry; ClinicalGroup = cohort or clinical subgroup; PGS = the PGS trait for that model; variable = the individual model term; odds_ratio = exponentiated coefficient (proportional-odds ratio); lower_95/upper_95 = 95% confidence interval bounds for the odds ratio; p_value = nominal (uncorrected) p-value; Bonf_P = Bonferroni-corrected p-value (capped at 1.00); N_Total = model sample size; N_Continuation = number in the continuation category; N_Switching = number in the switching category; N_Augmentation = number in the augmentation category. Odds ratios >1 indicate higher odds of a more escalated treatment outcome (i.e. movement up the continuation → switching → augmentation gradient) per one-SD increase in PGS. Bonferroni correction was applied within each specification (ancestry × clinical group) for the max 7 PGS traits tested. P-values reported as "<2.2e-16" reflect R's floating-point reporting limit.
Reference (omitted) categories: Sex at birth: Female; Income: lowest band (<$25k); Education level: lowest band (less than high school / fewer than twelve years).

| Population | ClinicalGroup | PGS | variable | odds_ratio | lower_95 | upper_95 | p_value | Bonf_P | N_Total | N_Continuation | N_Switching | N_Augmentation |
|---|---|---|---|---|---|---|---|---|---|---|---|---|
| European | Full Cohort | ADHD | age_at_first_AD | 0.9785 | 0.9772 | 0.9798 | <2.2e-16 | <2.2e-16 | 36790 | 21256 | 11645 | 3889 |
| European | Full Cohort | ADHD | sex_at_birthMale | 1.129 | 1.078 | 1.181 | 1.88e-07 | 1.32e-06 | 36790 | 21256 | 11645 | 3889 |
| European | Full Cohort | ADHD | Income_quantitative | 0.8614 | 0.8524 | 0.8705 | <2.2e-16 | <2.2e-16 | 36790 | 21256 | 11645 | 3889 |
| European | Full Cohort | ADHD | Education_levelCollege Undergraduate | 1.046 | 0.994 | 1.1 | 8.40e-02 | 5.88e-01 | 36790 | 21256 | 11645 | 3889 |
| European | Full Cohort | ADHD | Education_levelLess than a high school degree | 1.629 | 1.392 | 1.907 | 1.26e-09 | 8.84e-09 | 36790 | 21256 | 11645 | 3889 |
| European | Full Cohort | ADHD | Education_levelTwelve or GED | 1.261 | 1.169 | 1.359 | 1.57e-09 | 1.10e-08 | 36790 | 21256 | 11645 | 3889 |
| European | Full Cohort | ADHD | ADHD | 1.069 | 1.046 | 1.092 | 8.70e-10 | 6.09e-09 | 36790 | 21256 | 11645 | 3889 |
| European | Full Cohort | BMI | age_at_first_AD | 0.9785 | 0.9772 | 0.9798 | <2.2e-16 | <2.2e-16 | 36790 | 21256 | 11645 | 3889 |
| European | Full Cohort | BMI | sex_at_birthMale | 1.126 | 1.076 | 1.179 | 2.99e-07 | 2.09e-06 | 36790 | 21256 | 11645 | 3889 |
| European | Full Cohort | BMI | Income_quantitative | 0.8605 | 0.8515 | 0.8696 | <2.2e-16 | <2.2e-16 | 36790 | 21256 | 11645 | 3889 |
| European | Full Cohort | BMI | Education_levelCollege Undergraduate | 1.051 | 0.9993 | 1.106 | 5.30e-02 | 3.71e-01 | 36790 | 21256 | 11645 | 3889 |
| European | Full Cohort | BMI | Education_levelLess than a high school degree | 1.66 | 1.419 | 1.943 | 2.63e-10 | 1.84e-09 | 36790 | 21256 | 11645 | 3889 |
| European | Full Cohort | BMI | Education_levelTwelve or GED | 1.272 | 1.18 | 1.371 | 3.51e-10 | 2.46e-09 | 36790 | 21256 | 11645 | 3889 |
| European | Full Cohort | BMI | BMI | 1.045 | 1.023 | 1.067 | 3.74e-05 | 2.62e-04 | 36790 | 21256 | 11645 | 3889 |
| European | Full Cohort | Bipolar Disorder | age_at_first_AD | 0.9783 | 0.977 | 0.9796 | <2.2e-16 | <2.2e-16 | 36790 | 21256 | 11645 | 3889 |
| European | Full Cohort | Bipolar Disorder | sex_at_birthMale | 1.125 | 1.075 | 1.177 | 3.90e-07 | 2.73e-06 | 36790 | 21256 | 11645 | 3889 |
| European | Full Cohort | Bipolar Disorder | Income_quantitative | 0.8592 | 0.8503 | 0.8683 | <2.2e-16 | <2.2e-16 | 36790 | 21256 | 11645 | 3889 |
| European | Full Cohort | Bipolar Disorder | Education_levelCollege Undergraduate | 1.059 | 1.007 | 1.114 | 2.57e-02 | 1.80e-01 | 36790 | 21256 | 11645 | 3889 |
| European | Full Cohort | Bipolar Disorder | Education_levelLess than a high school degree | 1.684 | 1.439 | 1.97 | 8.13e-11 | 5.69e-10 | 36790 | 21256 | 11645 | 3889 |
| European | Full Cohort | Bipolar Disorder | Education_levelTwelve or GED | 1.292 | 1.199 | 1.393 | 2.05e-11 | 1.43e-10 | 36790 | 21256 | 11645 | 3889 |
| European | Full Cohort | Bipolar Disorder | Bipolar Disorder | 1.064 | 1.041 | 1.087 | 2.15e-08 | 1.51e-07 | 36790 | 21256 | 11645 | 3889 |
| European | Full Cohort | Depression | age_at_first_AD | 0.9787 | 0.9774 | 0.98 | <2.2e-16 | <2.2e-16 | 36790 | 21256 | 11645 | 3889 |
| European | Full Cohort | Depression | sex_at_birthMale | 1.116 | 1.066 | 1.168 | 2.47e-06 | 1.73e-05 | 36790 | 21256 | 11645 | 3889 |
| European | Full Cohort | Depression | Income_quantitative | 0.8651 | 0.856 | 0.8743 | <2.2e-16 | <2.2e-16 | 36790 | 21256 | 11645 | 3889 |
| European | Full Cohort | Depression | Education_levelCollege Undergraduate | 1.045 | 0.9932 | 1.099 | 8.97e-02 | 6.28e-01 | 36790 | 21256 | 11645 | 3889 |
| European | Full Cohort | Depression | Education_levelLess than a high school degree | 1.568 | 1.339 | 1.836 | 2.25e-08 | 1.58e-07 | 36790 | 21256 | 11645 | 3889 |
| European | Full Cohort | Depression | Education_levelTwelve or GED | 1.264 | 1.173 | 1.363 | 9.64e-10 | 6.75e-09 | 36790 | 21256 | 11645 | 3889 |
| European | Full Cohort | Depression | Depression | 1.234 | 1.207 | 1.261 | <2.2e-16 | <2.2e-16 | 36790 | 21256 | 11645 | 3889 |
| European | Full Cohort | Insomnia | age_at_first_AD | 0.9786 | 0.9772 | 0.9799 | <2.2e-16 | <2.2e-16 | 36790 | 21256 | 11645 | 3889 |
| European | Full Cohort | Insomnia | sex_at_birthMale | 1.123 | 1.073 | 1.176 | 5.41e-07 | 3.79e-06 | 36790 | 21256 | 11645 | 3889 |
| European | Full Cohort | Insomnia | Income_quantitative | 0.8619 | 0.8529 | 0.871 | <2.2e-16 | <2.2e-16 | 36790 | 21256 | 11645 | 3889 |
| European | Full Cohort | Insomnia | Education_levelCollege Undergraduate | 1.049 | 0.9969 | 1.103 | 6.60e-02 | 4.62e-01 | 36790 | 21256 | 11645 | 3889 |
| European | Full Cohort | Insomnia | Education_levelLess than a high school degree | 1.62 | 1.384 | 1.896 | 1.91e-09 | 1.34e-08 | 36790 | 21256 | 11645 | 3889 |
| European | Full Cohort | Insomnia | Education_levelTwelve or GED | 1.266 | 1.175 | 1.365 | 6.88e-10 | 4.81e-09 | 36790 | 21256 | 11645 | 3889 |
| European | Full Cohort | Insomnia | Insomnia | 1.111 | 1.087 | 1.135 | <2.2e-16 | <2.2e-16 | 36790 | 21256 | 11645 | 3889 |
| European | Full Cohort | Migraine | age_at_first_AD | 0.9784 | 0.9771 | 0.9797 | <2.2e-16 | <2.2e-16 | 36790 | 21256 | 11645 | 3889 |
| European | Full Cohort | Migraine | sex_at_birthMale | 1.126 | 1.075 | 1.178 | 3.50e-07 | 2.45e-06 | 36790 | 21256 | 11645 | 3889 |
| European | Full Cohort | Migraine | Income_quantitative | 0.8593 | 0.8503 | 0.8684 | <2.2e-16 | <2.2e-16 | 36790 | 21256 | 11645 | 3889 |
| European | Full Cohort | Migraine | Education_levelCollege Undergraduate | 1.056 | 1.004 | 1.11 | 3.58e-02 | 2.50e-01 | 36790 | 21256 | 11645 | 3889 |
| European | Full Cohort | Migraine | Education_levelLess than a high school degree | 1.678 | 1.434 | 1.963 | 1.09e-10 | 7.61e-10 | 36790 | 21256 | 11645 | 3889 |
| European | Full Cohort | Migraine | Education_levelTwelve or GED | 1.282 | 1.19 | 1.382 | 7.78e-11 | 5.45e-10 | 36790 | 21256 | 11645 | 3889 |
| European | Full Cohort | Migraine | Migraine | 1.038 | 1.017 | 1.06 | 3.99e-04 | 2.79e-03 | 36790 | 21256 | 11645 | 3889 |
| European | Full Cohort | Schizophrenia | age_at_first_AD | 0.9784 | 0.9771 | 0.9797 | <2.2e-16 | <2.2e-16 | 36790 | 21256 | 11645 | 3889 |
| European | Full Cohort | Schizophrenia | sex_at_birthMale | 1.126 | 1.076 | 1.178 | 3.46e-07 | 2.43e-06 | 36790 | 21256 | 11645 | 3889 |

| | | | | | | | | | | | | |
|---|---|---|---|---|---|---|---|---|---|---|---|---|
| European | Full Cohort | Schizophrenia | Income_quantitative | 0.8596 | 0.8506 | 0.8687 | <2.2e-16 | <2.2e-16 | 36790 | 21256 | 11645 | 3889 |
| European | Full Cohort | Schizophrenia | Education_levelCollege Undergraduate | 1.058 | 1.006 | 1.112 | 2.97e-02 | 2.08e-01 | 36790 | 21256 | 11645 | 3889 |
| European | Full Cohort | Schizophrenia | Education_levelLess than a high school degree | 1.675 | 1.431 | 1.96 | 1.25e-10 | 8.74e-10 | 36790 | 21256 | 11645 | 3889 |
| European | Full Cohort | Schizophrenia | Education_levelTwelve or GED | 1.287 | 1.194 | 1.387 | 4.21e-11 | 2.95e-10 | 36790 | 21256 | 11645 | 3889 |
| European | Full Cohort | Schizophrenia | Schizophrenia | 1.043 | 1.022 | 1.065 | 7.06e-05 | 4.94e-04 | 36790 | 21256 | 11645 | 3889 |
| European | Recorded Depression | ADHD | age_at_first_AD | 0.9793 | 0.9775 | 0.9811 | <2.2e-16 | <2.2e-16 | 18692 | 7969 | 7777 | 2946 |
| European | Recorded Depression | ADHD | sex_at_birthMale | 1.21 | 1.139 | 1.285 | 6.25e-10 | 4.38e-09 | 18692 | 7969 | 7777 | 2946 |
| European | Recorded Depression | ADHD | Income_quantitative | 0.8886 | 0.8761 | 0.9013 | <2.2e-16 | <2.2e-16 | 18692 | 7969 | 7777 | 2946 |
| European | Recorded Depression | ADHD | Education_levelCollege Undergraduate | 1.077 | 1.005 | 1.153 | 3.46e-02 | 2.43e-01 | 18692 | 7969 | 7777 | 2946 |
| European | Recorded Depression | ADHD | Education_levelLess than a high school degree | 1.726 | 1.413 | 2.108 | 8.73e-08 | 6.11e-07 | 18692 | 7969 | 7777 | 2946 |
| European | Recorded Depression | ADHD | Education_levelTwelve or GED | 1.329 | 1.203 | 1.468 | 2.22e-08 | 1.55e-07 | 18692 | 7969 | 7777 | 2946 |
| European | Recorded Depression | ADHD | ADHD | 1.05 | 1.021 | 1.08 | 7.09e-04 | 4.96e-03 | 18692 | 7969 | 7777 | 2946 |
| European | Recorded Depression | BMI | age_at_first_AD | 0.9793 | 0.9775 | 0.9811 | <2.2e-16 | <2.2e-16 | 18692 | 7969 | 7777 | 2946 |
| European | Recorded Depression | BMI | sex_at_birthMale | 1.208 | 1.138 | 1.284 | 8.03e-10 | 5.62e-09 | 18692 | 7969 | 7777 | 2946 |
| European | Recorded Depression | BMI | Income_quantitative | 0.887 | 0.8745 | 0.8997 | <2.2e-16 | <2.2e-16 | 18692 | 7969 | 7777 | 2946 |
| European | Recorded Depression | BMI | Education_levelCollege Undergraduate | 1.085 | 1.013 | 1.162 | 1.95e-02 | 1.37e-01 | 18692 | 7969 | 7777 | 2946 |
| European | Recorded Depression | BMI | Education_levelLess than a high school degree | 1.757 | 1.439 | 2.146 | 3.14e-08 | 2.20e-07 | 18692 | 7969 | 7777 | 2946 |
| European | Recorded Depression | BMI | Education_levelTwelve or GED | 1.346 | 1.218 | 1.486 | 4.79e-09 | 3.35e-08 | 18692 | 7969 | 7777 | 2946 |
| European | Recorded Depression | BMI | BMI | 1.007 | 0.9793 | 1.035 | 6.32e-01 | 1.00e+00 | 18692 | 7969 | 7777 | 2946 |
| European | Recorded Depression | Bipolar Disorder | age_at_first_AD | 0.9792 | 0.9774 | 0.981 | <2.2e-16 | <2.2e-16 | 18692 | 7969 | 7777 | 2946 |
| European | Recorded Depression | Bipolar Disorder | sex_at_birthMale | 1.208 | 1.138 | 1.284 | 8.00e-10 | 5.60e-09 | 18692 | 7969 | 7777 | 2946 |
| European | Recorded Depression | Bipolar Disorder | Income_quantitative | 0.8866 | 0.8742 | 0.8993 | <2.2e-16 | <2.2e-16 | 18692 | 7969 | 7777 | 2946 |
| European | Recorded Depression | Bipolar Disorder | Education_levelCollege Undergraduate | 1.087 | 1.015 | 1.164 | 1.71e-02 | 1.20e-01 | 18692 | 7969 | 7777 | 2946 |
| European | Recorded Depression | Bipolar Disorder | Education_levelLess than a high school degree | 1.76 | 1.442 | 2.149 | 2.76e-08 | 1.93e-07 | 18692 | 7969 | 7777 | 2946 |
| European | Recorded Depression | Bipolar Disorder | Education_levelTwelve or GED | 1.351 | 1.223 | 1.492 | 2.85e-09 | 2.00e-08 | 18692 | 7969 | 7777 | 2946 |
| European | Recorded Depression | Bipolar Disorder | Bipolar Disorder | 1.034 | 1.005 | 1.065 | 2.09e-02 | 1.46e-01 | 18692 | 7969 | 7777 | 2946 |
| European | Recorded Depression | Depression | age_at_first_AD | 0.9793 | 0.9775 | 0.9811 | <2.2e-16 | <2.2e-16 | 18692 | 7969 | 7777 | 2946 |
| European | Recorded Depression | Depression | sex_at_birthMale | 1.199 | 1.128 | 1.273 | 4.29e-09 | 3.00e-08 | 18692 | 7969 | 7777 | 2946 |
| European | Recorded Depression | Depression | Income_quantitative | 0.8914 | 0.8788 | 0.9041 | <2.2e-16 | <2.2e-16 | 18692 | 7969 | 7777 | 2946 |
| European | Recorded Depression | Depression | Education_levelCollege Undergraduate | 1.077 | 1.006 | 1.153 | 3.37e-02 | 2.36e-01 | 18692 | 7969 | 7777 | 2946 |
| European | Recorded Depression | Depression | Education_levelLess than a high school degree | 1.67 | 1.367 | 2.039 | 5.15e-07 | 3.61e-06 | 18692 | 7969 | 7777 | 2946 |
| European | Recorded Depression | Depression | Education_levelTwelve or GED | 1.333 | 1.207 | 1.473 | 1.41e-08 | 9.88e-08 | 18692 | 7969 | 7777 | 2946 |
| European | Recorded Depression | Depression | Depression | 1.178 | 1.145 | 1.213 | <2.2e-16 | <2.2e-16 | 18692 | 7969 | 7777 | 2946 |
| European | Recorded Depression | Insomnia | age_at_first_AD | 0.9794 | 0.9776 | 0.9812 | <2.2e-16 | <2.2e-16 | 18692 | 7969 | 7777 | 2946 |
| European | Recorded Depression | Insomnia | sex_at_birthMale | 1.205 | 1.134 | 1.28 | 1.54e-09 | 1.08e-08 | 18692 | 7969 | 7777 | 2946 |
| European | Recorded Depression | Insomnia | Income_quantitative | 0.8891 | 0.8766 | 0.9018 | <2.2e-16 | <2.2e-16 | 18692 | 7969 | 7777 | 2946 |
| European | Recorded Depression | Insomnia | Education_levelCollege Undergraduate | 1.08 | 1.008 | 1.156 | 2.81e-02 | 1.97e-01 | 18692 | 7969 | 7777 | 2946 |
| European | Recorded Depression | Insomnia | Education_levelLess than a high school degree | 1.712 | 1.402 | 2.091 | 1.36e-07 | 9.52e-07 | 18692 | 7969 | 7777 | 2946 |
| European | Recorded Depression | Insomnia | Education_levelTwelve or GED | 1.333 | 1.207 | 1.472 | 1.44e-08 | 1.00e-07 | 18692 | 7969 | 7777 | 2946 |
| European | Recorded Depression | Insomnia | Insomnia | 1.082 | 1.051 | 1.114 | 9.80e-08 | 6.86e-07 | 18692 | 7969 | 7777 | 2946 |
| European | Recorded Depression | Migraine | age_at_first_AD | 0.9793 | 0.9775 | 0.9811 | <2.2e-16 | <2.2e-16 | 18692 | 7969 | 7777 | 2946 |
| European | Recorded Depression | Migraine | sex_at_birthMale | 1.208 | 1.137 | 1.283 | 8.41e-10 | 5.88e-09 | 18692 | 7969 | 7777 | 2946 |
| European | Recorded Depression | Migraine | Income_quantitative | 0.8872 | 0.8747 | 0.8998 | <2.2e-16 | <2.2e-16 | 18692 | 7969 | 7777 | 2946 |
| European | Recorded Depression | Migraine | Education_levelCollege Undergraduate | 1.084 | 1.013 | 1.161 | 2.01e-02 | 1.41e-01 | 18692 | 7969 | 7777 | 2946 |
| European | Recorded Depression | Migraine | Education_levelLess than a high school degree | 1.763 | 1.444 | 2.153 | 2.54e-08 | 1.78e-07 | 18692 | 7969 | 7777 | 2946 |
| European | Recorded Depression | Migraine | Education_levelTwelve or GED | 1.347 | 1.22 | 1.487 | 4.09e-09 | 2.87e-08 | 18692 | 7969 | 7777 | 2946 |
| European | Recorded Depression | Migraine | Migraine | 1.049 | 1.02 | 1.078 | 7.07e-04 | 4.95e-03 | 18692 | 7969 | 7777 | 2946 |
| European | Recorded Depression | Schizophrenia | age_at_first_AD | 0.9793 | 0.9775 | 0.9811 | <2.2e-16 | <2.2e-16 | 18692 | 7969 | 7777 | 2946 |
| European | Recorded Depression | Schizophrenia | sex_at_birthMale | 1.208 | 1.137 | 1.283 | 8.67e-10 | 6.07e-09 | 18692 | 7969 | 7777 | 2946 |
| European | Recorded Depression | Schizophrenia | Income_quantitative | 0.8871 | 0.8747 | 0.8998 | <2.2e-16 | <2.2e-16 | 18692 | 7969 | 7777 | 2946 |
| European | Recorded Depression | Schizophrenia | Education_levelCollege Undergraduate | 1.087 | 1.015 | 1.164 | 1.74e-02 | 1.22e-01 | 18692 | 7969 | 7777 | 2946 |
| European | Recorded Depression | Schizophrenia | Education_levelLess than a high school degree | 1.754 | 1.437 | 2.141 | 3.42e-08 | 2.40e-07 | 18692 | 7969 | 7777 | 2946 |
| European | Recorded Depression | Schizophrenia | Education_levelTwelve or GED | 1.349 | 1.221 | 1.49 | 3.45e-09 | 2.42e-08 | 18692 | 7969 | 7777 | 2946 |
| European | Recorded Depression | Schizophrenia | Schizophrenia | 1.04 | 1.012 | 1.07 | 5.58e-03 | 3.90e-02 | 18692 | 7969 | 7777 | 2946 |
| European | No Recorded Depression | ADHD | age_at_first_AD | 0.9809 | 0.9788 | 0.983 | <2.2e-16 | <2.2e-16 | 18098 | 13287 | 3868 | 943 |
| European | No Recorded Depression | ADHD | sex_at_birthMale | 0.981 | 0.9104 | 1.057 | 6.16e-01 | 1.00e+00 | 18098 | 13287 | 3868 | 943 |
| European | No Recorded Depression | ADHD | Income_quantitative | 0.8898 | 0.8748 | 0.9049 | <2.2e-16 | <2.2e-16 | 18098 | 13287 | 3868 | 943 |

| | | | | | | | | | | | | |
|---|---|---|---|---|---|---|---|---|---|---|---|---|
| European | No Recorded Depression | ADHD | Education_levelCollege Undergraduate | 1.01 | 0.932 | 1.093 | 8.17e-01 | 1.00e+00 | 18098 | 13287 | 3868 | 943 |
| European | No Recorded Depression | ADHD | Education_levelLess than a high school degree | 1.515 | 1.155 | 1.987 | 2.67e-03 | 1.87e-02 | 18098 | 13287 | 3868 | 943 |
| European | No Recorded Depression | ADHD | Education_levelTwelve or GED | 1.195 | 1.057 | 1.352 | 4.38e-03 | 3.06e-02 | 18098 | 13287 | 3868 | 943 |
| European | No Recorded Depression | ADHD | ADHD | 1.065 | 1.029 | 1.102 | 3.46e-04 | 2.42e-03 | 18098 | 13287 | 3868 | 943 |
| European | No Recorded Depression | BMI | age_at_first_AD | 0.9809 | 0.9788 | 0.983 | <2.2e-16 | <2.2e-16 | 18098 | 13287 | 3868 | 943 |
| European | No Recorded Depression | BMI | sex_at_birthMale | 0.9785 | 0.9081 | 1.054 | 5.69e-01 | 1.00e+00 | 18098 | 13287 | 3868 | 943 |
| European | No Recorded Depression | BMI | Income_quantitative | 0.8894 | 0.8745 | 0.9046 | <2.2e-16 | <2.2e-16 | 18098 | 13287 | 3868 | 943 |
| European | No Recorded Depression | BMI | Education_levelCollege Undergraduate | 1.012 | 0.9339 | 1.096 | 7.78e-01 | 1.00e+00 | 18098 | 13287 | 3868 | 943 |
| European | No Recorded Depression | BMI | Education_levelLess than a high school degree | 1.55 | 1.183 | 2.032 | 1.48e-03 | 1.04e-02 | 18098 | 13287 | 3868 | 943 |
| European | No Recorded Depression | BMI | Education_levelTwelve or GED | 1.201 | 1.063 | 1.358 | 3.39e-03 | 2.38e-02 | 18098 | 13287 | 3868 | 943 |
| European | No Recorded Depression | BMI | BMI | 1.053 | 1.018 | 1.089 | 2.59e-03 | 1.82e-02 | 18098 | 13287 | 3868 | 943 |
| European | No Recorded Depression | Bipolar Disorder | age_at_first_AD | 0.9807 | 0.9787 | 0.9828 | <2.2e-16 | <2.2e-16 | 18098 | 13287 | 3868 | 943 |
| European | No Recorded Depression | Bipolar Disorder | sex_at_birthMale | 0.9758 | 0.9056 | 1.051 | 5.20e-01 | 1.00e+00 | 18098 | 13287 | 3868 | 943 |
| European | No Recorded Depression | Bipolar Disorder | Income_quantitative | 0.8884 | 0.8736 | 0.9035 | <2.2e-16 | <2.2e-16 | 18098 | 13287 | 3868 | 943 |
| European | No Recorded Depression | Bipolar Disorder | Education_levelCollege Undergraduate | 1.022 | 0.9436 | 1.107 | 5.94e-01 | 1.00e+00 | 18098 | 13287 | 3868 | 943 |
| European | No Recorded Depression | Bipolar Disorder | Education_levelLess than a high school degree | 1.578 | 1.204 | 2.068 | 9.57e-04 | 6.70e-03 | 18098 | 13287 | 3868 | 943 |
| European | No Recorded Depression | Bipolar Disorder | Education_levelTwelve or GED | 1.228 | 1.086 | 1.388 | 1.04e-03 | 7.26e-03 | 18098 | 13287 | 3868 | 943 |
| European | No Recorded Depression | Bipolar Disorder | Bipolar Disorder | 1.075 | 1.038 | 1.114 | 6.18e-05 | 4.33e-04 | 18098 | 13287 | 3868 | 943 |
| European | No Recorded Depression | Depression | age_at_first_AD | 0.9812 | 0.9791 | 0.9833 | <2.2e-16 | <2.2e-16 | 18098 | 13287 | 3868 | 943 |
| European | No Recorded Depression | Depression | sex_at_birthMale | 0.9708 | 0.9008 | 1.046 | 4.38e-01 | 1.00e+00 | 18098 | 13287 | 3868 | 943 |
| European | No Recorded Depression | Depression | Income_quantitative | 0.8928 | 0.8778 | 0.908 | <2.2e-16 | <2.2e-16 | 18098 | 13287 | 3868 | 943 |
| European | No Recorded Depression | Depression | Education_levelCollege Undergraduate | 1.007 | 0.9293 | 1.091 | 8.70e-01 | 1.00e+00 | 18098 | 13287 | 3868 | 943 |
| European | No Recorded Depression | Depression | Education_levelLess than a high school degree | 1.47 | 1.121 | 1.929 | 5.38e-03 | 3.77e-02 | 18098 | 13287 | 3868 | 943 |
| European | No Recorded Depression | Depression | Education_levelTwelve or GED | 1.196 | 1.058 | 1.353 | 4.20e-03 | 2.94e-02 | 18098 | 13287 | 3868 | 943 |
| European | No Recorded Depression | Depression | Depression | 1.218 | 1.176 | 1.262 | <2.2e-16 | <2.2e-16 | 18098 | 13287 | 3868 | 943 |
| European | No Recorded Depression | Insomnia | age_at_first_AD | 0.981 | 0.9789 | 0.9831 | <2.2e-16 | <2.2e-16 | 18098 | 13287 | 3868 | 943 |
| European | No Recorded Depression | Insomnia | sex_at_birthMale | 0.978 | 0.9076 | 1.054 | 5.60e-01 | 1.00e+00 | 18098 | 13287 | 3868 | 943 |
| European | No Recorded Depression | Insomnia | Income_quantitative | 0.8906 | 0.8757 | 0.9058 | <2.2e-16 | <2.2e-16 | 18098 | 13287 | 3868 | 943 |
| European | No Recorded Depression | Insomnia | Education_levelCollege Undergraduate | 1.009 | 0.9318 | 1.093 | 8.20e-01 | 1.00e+00 | 18098 | 13287 | 3868 | 943 |
| European | No Recorded Depression | Insomnia | Education_levelLess than a high school degree | 1.509 | 1.151 | 1.979 | 2.90e-03 | 2.03e-02 | 18098 | 13287 | 3868 | 943 |
| European | No Recorded Depression | Insomnia | Education_levelTwelve or GED | 1.199 | 1.06 | 1.355 | 3.78e-03 | 2.65e-02 | 18098 | 13287 | 3868 | 943 |
| European | No Recorded Depression | Insomnia | Insomnia | 1.127 | 1.088 | 1.167 | 2.39e-11 | 1.68e-10 | 18098 | 13287 | 3868 | 943 |
| European | No Recorded Depression | Migraine | age_at_first_AD | 0.9809 | 0.9788 | 0.9829 | <2.2e-16 | <2.2e-16 | 18098 | 13287 | 3868 | 943 |
| European | No Recorded Depression | Migraine | sex_at_birthMale | 0.9777 | 0.9074 | 1.054 | 5.55e-01 | 1.00e+00 | 18098 | 13287 | 3868 | 943 |
| European | No Recorded Depression | Migraine | Income_quantitative | 0.8879 | 0.8731 | 0.903 | <2.2e-16 | <2.2e-16 | 18098 | 13287 | 3868 | 943 |
| European | No Recorded Depression | Migraine | Education_levelCollege Undergraduate | 1.018 | 0.9401 | 1.103 | 6.58e-01 | 1.00e+00 | 18098 | 13287 | 3868 | 943 |
| European | No Recorded Depression | Migraine | Education_levelLess than a high school degree | 1.565 | 1.194 | 2.051 | 1.17e-03 | 8.16e-03 | 18098 | 13287 | 3868 | 943 |
| European | No Recorded Depression | Migraine | Education_levelTwelve or GED | 1.214 | 1.074 | 1.372 | 1.92e-03 | 1.34e-02 | 18098 | 13287 | 3868 | 943 |
| European | No Recorded Depression | Migraine | Migraine | 1.024 | 0.9905 | 1.059 | 1.62e-01 | 1.00e+00 | 18098 | 13287 | 3868 | 943 |
| European | No Recorded Depression | Schizophrenia | age_at_first_AD | 0.9808 | 0.9788 | 0.9829 | <2.2e-16 | <2.2e-16 | 18098 | 13287 | 3868 | 943 |
| European | No Recorded Depression | Schizophrenia | sex_at_birthMale | 0.9777 | 0.9073 | 1.053 | 5.53e-01 | 1.00e+00 | 18098 | 13287 | 3868 | 943 |
| European | No Recorded Depression | Schizophrenia | Income_quantitative | 0.8885 | 0.8737 | 0.9036 | <2.2e-16 | <2.2e-16 | 18098 | 13287 | 3868 | 943 |
| European | No Recorded Depression | Schizophrenia | Education_levelCollege Undergraduate | 1.02 | 0.9414 | 1.104 | 6.34e-01 | 1.00e+00 | 18098 | 13287 | 3868 | 943 |
| European | No Recorded Depression | Schizophrenia | Education_levelLess than a high school degree | 1.567 | 1.196 | 2.054 | 1.12e-03 | 7.85e-03 | 18098 | 13287 | 3868 | 943 |
| European | No Recorded Depression | Schizophrenia | Education_levelTwelve or GED | 1.22 | 1.079 | 1.379 | 1.46e-03 | 1.02e-02 | 18098 | 13287 | 3868 | 943 |
| European | No Recorded Depression | Schizophrenia | Schizophrenia | 1.043 | 1.008 | 1.079 | 1.43e-02 | 1.00e-01 | 18098 | 13287 | 3868 | 943 |
| European | Recorded Anxiety | ADHD | age_at_first_AD | 0.9826 | 0.981 | 0.9842 | <2.2e-16 | <2.2e-16 | 23907 | 11798 | 9020 | 3089 |
| European | Recorded Anxiety | ADHD | sex_at_birthMale | 1.254 | 1.186 | 1.325 | 1.14e-15 | 7.98e-15 | 23907 | 11798 | 9020 | 3089 |
| European | Recorded Anxiety | ADHD | Income_quantitative | 0.8579 | 0.847 | 0.8688 | <2.2e-16 | <2.2e-16 | 23907 | 11798 | 9020 | 3089 |
| European | Recorded Anxiety | ADHD | Education_levelCollege Undergraduate | 1.045 | 0.9829 | 1.111 | 1.59e-01 | 1.00e+00 | 23907 | 11798 | 9020 | 3089 |
| European | Recorded Anxiety | ADHD | Education_levelLess than a high school degree | 1.446 | 1.201 | 1.74 | 9.55e-05 | 6.69e-04 | 23907 | 11798 | 9020 | 3089 |
| European | Recorded Anxiety | ADHD | Education_levelTwelve or GED | 1.327 | 1.213 | 1.452 | 6.43e-10 | 4.50e-09 | 23907 | 11798 | 9020 | 3089 |
| European | Recorded Anxiety | ADHD | ADHD | 1.049 | 1.023 | 1.076 | 2.23e-04 | 1.56e-03 | 23907 | 11798 | 9020 | 3089 |
| European | Recorded Anxiety | BMI | age_at_first_AD | 0.9826 | 0.981 | 0.9842 | <2.2e-16 | <2.2e-16 | 23907 | 11798 | 9020 | 3089 |
| European | Recorded Anxiety | BMI | sex_at_birthMale | 1.252 | 1.185 | 1.324 | 1.54e-15 | 1.08e-14 | 23907 | 11798 | 9020 | 3089 |
| European | Recorded Anxiety | BMI | Income_quantitative | 0.8574 | 0.8466 | 0.8684 | <2.2e-16 | <2.2e-16 | 23907 | 11798 | 9020 | 3089 |
| European | Recorded Anxiety | BMI | Education_levelCollege Undergraduate | 1.047 | 0.9851 | 1.113 | 1.39e-01 | 9.76e-01 | 23907 | 11798 | 9020 | 3089 |

| | | | | | | | | | | | | |
|---|---|---|---|---|---|---|---|---|---|---|---|---|
| European | Recorded Anxiety | BMI | Education_levelLess than a high school degree | 1.462 | 1.215 | 1.759 | 5.73e-05 | 4.01e-04 | 23907 | 11798 | 9020 | 3089 |
| European | Recorded Anxiety | BMI | Education_levelTwelve or GED | 1.331 | 1.217 | 1.456 | 4.19e-10 | 2.93e-09 | 23907 | 11798 | 9020 | 3089 |
| European | Recorded Anxiety | BMI | BMI | 1.042 | 1.016 | 1.068 | 1.26e-03 | 8.85e-03 | 23907 | 11798 | 9020 | 3089 |
| European | Recorded Anxiety | Bipolar Disorder | age_at_first_AD | 0.9824 | 0.9808 | 0.984 | <2.2e-16 | <2.2e-16 | 23907 | 11798 | 9020 | 3089 |
| European | Recorded Anxiety | Bipolar Disorder | sex_at_birthMale | 1.251 | 1.183 | 1.322 | 2.28e-15 | 1.59e-14 | 23907 | 11798 | 9020 | 3089 |
| European | Recorded Anxiety | Bipolar Disorder | Income_quantitative | 0.8563 | 0.8455 | 0.8672 | <2.2e-16 | <2.2e-16 | 23907 | 11798 | 9020 | 3089 |
| European | Recorded Anxiety | Bipolar Disorder | Education_levelCollege Undergraduate | 1.055 | 0.9927 | 1.122 | 8.47e-02 | 5.93e-01 | 23907 | 11798 | 9020 | 3089 |
| European | Recorded Anxiety | Bipolar Disorder | Education_levelLess than a high school degree | 1.48 | 1.23 | 1.78 | 3.26e-05 | 2.28e-04 | 23907 | 11798 | 9020 | 3089 |
| European | Recorded Anxiety | Bipolar Disorder | Education_levelTwelve or GED | 1.35 | 1.235 | 1.477 | 4.72e-11 | 3.30e-10 | 23907 | 11798 | 9020 | 3089 |
| European | Recorded Anxiety | Bipolar Disorder | Bipolar Disorder | 1.057 | 1.03 | 1.085 | 2.63e-05 | 1.84e-04 | 23907 | 11798 | 9020 | 3089 |
| European | Recorded Anxiety | Depression | age_at_first_AD | 0.9826 | 0.981 | 0.9842 | <2.2e-16 | <2.2e-16 | 23907 | 11798 | 9020 | 3089 |
| European | Recorded Anxiety | Depression | sex_at_birthMale | 1.239 | 1.172 | 1.31 | 3.45e-14 | 2.41e-13 | 23907 | 11798 | 9020 | 3089 |
| European | Recorded Anxiety | Depression | Income_quantitative | 0.8617 | 0.8509 | 0.8727 | <2.2e-16 | <2.2e-16 | 23907 | 11798 | 9020 | 3089 |
| European | Recorded Anxiety | Depression | Education_levelCollege Undergraduate | 1.041 | 0.9795 | 1.107 | 1.95e-01 | 1.00e+00 | 23907 | 11798 | 9020 | 3089 |
| European | Recorded Anxiety | Depression | Education_levelLess than a high school degree | 1.39 | 1.155 | 1.672 | 5.02e-04 | 3.52e-03 | 23907 | 11798 | 9020 | 3089 |
| European | Recorded Anxiety | Depression | Education_levelTwelve or GED | 1.322 | 1.209 | 1.446 | 1.02e-09 | 7.12e-09 | 23907 | 11798 | 9020 | 3089 |
| European | Recorded Anxiety | Depression | Depression | 1.214 | 1.182 | 1.246 | <2.2e-16 | <2.2e-16 | 23907 | 11798 | 9020 | 3089 |
| European | Recorded Anxiety | Insomnia | age_at_first_AD | 0.9826 | 0.981 | 0.9842 | <2.2e-16 | <2.2e-16 | 23907 | 11798 | 9020 | 3089 |
| European | Recorded Anxiety | Insomnia | sex_at_birthMale | 1.248 | 1.181 | 1.319 | 4.05e-15 | 2.83e-14 | 23907 | 11798 | 9020 | 3089 |
| European | Recorded Anxiety | Insomnia | Income_quantitative | 0.859 | 0.8482 | 0.87 | <2.2e-16 | <2.2e-16 | 23907 | 11798 | 9020 | 3089 |
| European | Recorded Anxiety | Insomnia | Education_levelCollege Undergraduate | 1.043 | 0.9813 | 1.109 | 1.76e-01 | 1.00e+00 | 23907 | 11798 | 9020 | 3089 |
| European | Recorded Anxiety | Insomnia | Education_levelLess than a high school degree | 1.426 | 1.185 | 1.716 | 1.73e-04 | 1.21e-03 | 23907 | 11798 | 9020 | 3089 |
| European | Recorded Anxiety | Insomnia | Education_levelTwelve or GED | 1.323 | 1.209 | 1.447 | 9.31e-10 | 6.52e-09 | 23907 | 11798 | 9020 | 3089 |
| European | Recorded Anxiety | Insomnia | Insomnia | 1.112 | 1.083 | 1.141 | 1.78e-15 | 1.25e-14 | 23907 | 11798 | 9020 | 3089 |
| European | Recorded Anxiety | Migraine | age_at_first_AD | 0.9825 | 0.9809 | 0.9841 | <2.2e-16 | <2.2e-16 | 23907 | 11798 | 9020 | 3089 |
| European | Recorded Anxiety | Migraine | sex_at_birthMale | 1.251 | 1.184 | 1.322 | 2.10e-15 | 1.47e-14 | 23907 | 11798 | 9020 | 3089 |
| European | Recorded Anxiety | Migraine | Income_quantitative | 0.8564 | 0.8457 | 0.8673 | <2.2e-16 | <2.2e-16 | 23907 | 11798 | 9020 | 3089 |
| European | Recorded Anxiety | Migraine | Education_levelCollege Undergraduate | 1.051 | 0.9892 | 1.118 | 1.07e-01 | 7.52e-01 | 23907 | 11798 | 9020 | 3089 |
| European | Recorded Anxiety | Migraine | Education_levelLess than a high school degree | 1.478 | 1.228 | 1.778 | 3.49e-05 | 2.45e-04 | 23907 | 11798 | 9020 | 3089 |
| European | Recorded Anxiety | Migraine | Education_levelTwelve or GED | 1.342 | 1.227 | 1.468 | 1.16e-10 | 8.13e-10 | 23907 | 11798 | 9020 | 3089 |
| European | Recorded Anxiety | Migraine | Migraine | 1.044 | 1.019 | 1.071 | 5.59e-04 | 3.91e-03 | 23907 | 11798 | 9020 | 3089 |
| European | Recorded Anxiety | Schizophrenia | age_at_first_AD | 0.9825 | 0.9809 | 0.9841 | <2.2e-16 | <2.2e-16 | 23907 | 11798 | 9020 | 3089 |
| European | Recorded Anxiety | Schizophrenia | sex_at_birthMale | 1.252 | 1.184 | 1.323 | 1.87e-15 | 1.31e-14 | 23907 | 11798 | 9020 | 3089 |
| European | Recorded Anxiety | Schizophrenia | Income_quantitative | 0.8566 | 0.8458 | 0.8675 | <2.2e-16 | <2.2e-16 | 23907 | 11798 | 9020 | 3089 |
| European | Recorded Anxiety | Schizophrenia | Education_levelCollege Undergraduate | 1.054 | 0.9912 | 1.12 | 9.39e-02 | 6.58e-01 | 23907 | 11798 | 9020 | 3089 |
| European | Recorded Anxiety | Schizophrenia | Education_levelLess than a high school degree | 1.474 | 1.226 | 1.774 | 3.84e-05 | 2.69e-04 | 23907 | 11798 | 9020 | 3089 |
| European | Recorded Anxiety | Schizophrenia | Education_levelTwelve or GED | 1.346 | 1.231 | 1.472 | 7.27e-11 | 5.09e-10 | 23907 | 11798 | 9020 | 3089 |
| European | Recorded Anxiety | Schizophrenia | Schizophrenia | 1.034 | 1.008 | 1.06 | 8.98e-03 | 6.29e-02 | 23907 | 11798 | 9020 | 3089 |
| European | CIDI-MDD | ADHD | age_at_first_AD | 0.9767 | 0.9729 | 0.9805 | <2.2e-16 | <2.2e-16 | 4758 | 2349 | 1736 | 673 |
| European | CIDI-MDD | ADHD | sex_at_birthMale | 1.193 | 1.049 | 1.358 | 7.27e-03 | 5.09e-02 | 4758 | 2349 | 1736 | 673 |
| European | CIDI-MDD | ADHD | Income_quantitative | 0.8758 | 0.8514 | 0.901 | <2.2e-16 | <2.2e-16 | 4758 | 2349 | 1736 | 673 |
| European | CIDI-MDD | ADHD | Education_levelCollege Undergraduate | 1.09 | 0.96 | 1.238 | 1.83e-01 | 1.00e+00 | 4758 | 2349 | 1736 | 673 |
| European | CIDI-MDD | ADHD | Education_levelLess than a high school degree | 0.504 | 0.2412 | 1.053 | 6.84e-02 | 4.79e-01 | 4758 | 2349 | 1736 | 673 |
| European | CIDI-MDD | ADHD | Education_levelTwelve or GED | 1.327 | 1.05 | 1.677 | 1.77e-02 | 1.24e-01 | 4758 | 2349 | 1736 | 673 |
| European | CIDI-MDD | ADHD | ADHD | 1.066 | 1.006 | 1.13 | 2.93e-02 | 2.05e-01 | 4758 | 2349 | 1736 | 673 |
| European | CIDI-MDD | BMI | age_at_first_AD | 0.9767 | 0.9729 | 0.9804 | <2.2e-16 | <2.2e-16 | 4758 | 2349 | 1736 | 673 |
| European | CIDI-MDD | BMI | sex_at_birthMale | 1.192 | 1.047 | 1.356 | 7.75e-03 | 5.42e-02 | 4758 | 2349 | 1736 | 673 |
| European | CIDI-MDD | BMI | Income_quantitative | 0.8748 | 0.8505 | 0.8999 | <2.2e-16 | <2.2e-16 | 4758 | 2349 | 1736 | 673 |
| European | CIDI-MDD | BMI | Education_levelCollege Undergraduate | 1.1 | 0.9691 | 1.249 | 1.40e-01 | 9.82e-01 | 4758 | 2349 | 1736 | 673 |
| European | CIDI-MDD | BMI | Education_levelLess than a high school degree | 0.513 | 0.2456 | 1.072 | 7.58e-02 | 5.31e-01 | 4758 | 2349 | 1736 | 673 |
| European | CIDI-MDD | BMI | Education_levelTwelve or GED | 1.343 | 1.064 | 1.697 | 1.32e-02 | 9.25e-02 | 4758 | 2349 | 1736 | 673 |
| European | CIDI-MDD | BMI | BMI | 1.034 | 0.9777 | 1.094 | 2.40e-01 | 1.00e+00 | 4758 | 2349 | 1736 | 673 |
| European | CIDI-MDD | Bipolar Disorder | age_at_first_AD | 0.9765 | 0.9727 | 0.9802 | <2.2e-16 | <2.2e-16 | 4758 | 2349 | 1736 | 673 |
| European | CIDI-MDD | Bipolar Disorder | sex_at_birthMale | 1.196 | 1.051 | 1.36 | 6.75e-03 | 4.73e-02 | 4758 | 2349 | 1736 | 673 |
| European | CIDI-MDD | Bipolar Disorder | Income_quantitative | 0.8734 | 0.8491 | 0.8984 | <2.2e-16 | <2.2e-16 | 4758 | 2349 | 1736 | 673 |
| European | CIDI-MDD | Bipolar Disorder | Education_levelCollege Undergraduate | 1.107 | 0.9754 | 1.257 | 1.15e-01 | 8.07e-01 | 4758 | 2349 | 1736 | 673 |
| European | CIDI-MDD | Bipolar Disorder | Education_levelLess than a high school degree | 0.5333 | 0.2554 | 1.113 | 9.41e-02 | 6.59e-01 | 4758 | 2349 | 1736 | 673 |

| | | | | | | | | | | | | |
|---|---|---|---|---|---|---|---|---|---|---|---|---|
| European | CIDI-MDD | Bipolar Disorder | Education_levelTwelve or GED | 1.364 | 1.08 | 1.723 | 9.10e-03 | 6.37e-02 | 4758 | 2349 | 1736 | 673 |
| European | CIDI-MDD | Bipolar Disorder | Bipolar Disorder | 1.084 | 1.022 | 1.15 | 7.00e-03 | 4.90e-02 | 4758 | 2349 | 1736 | 673 |
| European | CIDI-MDD | Depression | age_at_first_AD | 0.9766 | 0.9728 | 0.9803 | <2.2e-16 | <2.2e-16 | 4758 | 2349 | 1736 | 673 |
| European | CIDI-MDD | Depression | sex_at_birthMale | 1.186 | 1.042 | 1.349 | 9.81e-03 | 6.87e-02 | 4758 | 2349 | 1736 | 673 |
| European | CIDI-MDD | Depression | Income_quantitative | 0.877 | 0.8525 | 0.9021 | <2.2e-16 | <2.2e-16 | 4758 | 2349 | 1736 | 673 |
| European | CIDI-MDD | Depression | Education_levelCollege Undergraduate | 1.08 | 0.951 | 1.227 | 2.36e-01 | 1.00e+00 | 4758 | 2349 | 1736 | 673 |
| European | CIDI-MDD | Depression | Education_levelLess than a high school degree | 0.5153 | 0.2467 | 1.076 | 7.77e-02 | 5.44e-01 | 4758 | 2349 | 1736 | 673 |
| European | CIDI-MDD | Depression | Education_levelTwelve or GED | 1.333 | 1.055 | 1.684 | 1.60e-02 | 1.12e-01 | 4758 | 2349 | 1736 | 673 |
| European | CIDI-MDD | Depression | Depression | 1.175 | 1.108 | 1.246 | 6.70e-08 | 4.69e-07 | 4758 | 2349 | 1736 | 673 |
| European | CIDI-MDD | Insomnia | age_at_first_AD | 0.9766 | 0.9729 | 0.9804 | <2.2e-16 | <2.2e-16 | 4758 | 2349 | 1736 | 673 |
| European | CIDI-MDD | Insomnia | sex_at_birthMale | 1.193 | 1.049 | 1.358 | 7.28e-03 | 5.09e-02 | 4758 | 2349 | 1736 | 673 |
| European | CIDI-MDD | Insomnia | Income_quantitative | 0.8743 | 0.85 | 0.8993 | <2.2e-16 | <2.2e-16 | 4758 | 2349 | 1736 | 673 |
| European | CIDI-MDD | Insomnia | Education_levelCollege Undergraduate | 1.099 | 0.9683 | 1.248 | 1.43e-01 | 1.00e+00 | 4758 | 2349 | 1736 | 673 |
| European | CIDI-MDD | Insomnia | Education_levelLess than a high school degree | 0.5109 | 0.2445 | 1.067 | 7.40e-02 | 5.18e-01 | 4758 | 2349 | 1736 | 673 |
| European | CIDI-MDD | Insomnia | Education_levelTwelve or GED | 1.347 | 1.066 | 1.701 | 1.25e-02 | 8.72e-02 | 4758 | 2349 | 1736 | 673 |
| European | CIDI-MDD | Insomnia | Insomnia | 1.032 | 0.9738 | 1.094 | 2.87e-01 | 1.00e+00 | 4758 | 2349 | 1736 | 673 |
| European | CIDI-MDD | Migraine | age_at_first_AD | 0.9766 | 0.9729 | 0.9804 | <2.2e-16 | <2.2e-16 | 4758 | 2349 | 1736 | 673 |
| European | CIDI-MDD | Migraine | sex_at_birthMale | 1.192 | 1.048 | 1.357 | 7.64e-03 | 5.35e-02 | 4758 | 2349 | 1736 | 673 |
| European | CIDI-MDD | Migraine | Income_quantitative | 0.874 | 0.8496 | 0.8989 | <2.2e-16 | <2.2e-16 | 4758 | 2349 | 1736 | 673 |
| European | CIDI-MDD | Migraine | Education_levelCollege Undergraduate | 1.103 | 0.9717 | 1.252 | 1.29e-01 | 9.06e-01 | 4758 | 2349 | 1736 | 673 |
| European | CIDI-MDD | Migraine | Education_levelLess than a high school degree | 0.5178 | 0.2479 | 1.081 | 7.98e-02 | 5.59e-01 | 4758 | 2349 | 1736 | 673 |
| European | CIDI-MDD | Migraine | Education_levelTwelve or GED | 1.352 | 1.071 | 1.707 | 1.13e-02 | 7.93e-02 | 4758 | 2349 | 1736 | 673 |
| European | CIDI-MDD | Migraine | Migraine | 1.014 | 0.9597 | 1.071 | 6.24e-01 | 1.00e+00 | 4758 | 2349 | 1736 | 673 |
| European | CIDI-MDD | Schizophrenia | age_at_first_AD | 0.9766 | 0.9728 | 0.9803 | <2.2e-16 | <2.2e-16 | 4758 | 2349 | 1736 | 673 |
| European | CIDI-MDD | Schizophrenia | sex_at_birthMale | 1.2 | 1.055 | 1.366 | 5.66e-03 | 3.96e-02 | 4758 | 2349 | 1736 | 673 |
| European | CIDI-MDD | Schizophrenia | Income_quantitative | 0.8743 | 0.85 | 0.8993 | <2.2e-16 | <2.2e-16 | 4758 | 2349 | 1736 | 673 |
| European | CIDI-MDD | Schizophrenia | Education_levelCollege Undergraduate | 1.103 | 0.9719 | 1.253 | 1.29e-01 | 9.00e-01 | 4758 | 2349 | 1736 | 673 |
| European | CIDI-MDD | Schizophrenia | Education_levelLess than a high school degree | 0.5257 | 0.252 | 1.097 | 8.66e-02 | 6.06e-01 | 4758 | 2349 | 1736 | 673 |
| European | CIDI-MDD | Schizophrenia | Education_levelTwelve or GED | 1.357 | 1.075 | 1.714 | 1.03e-02 | 7.21e-02 | 4758 | 2349 | 1736 | 673 |
| European | CIDI-MDD | Schizophrenia | Schizophrenia | 1.082 | 1.022 | 1.145 | 6.43e-03 | 4.50e-02 | 4758 | 2349 | 1736 | 673 |
| European | CIDI-noMDD | ADHD | age_at_first_AD | 0.971 | 0.9647 | 0.9772 | <2.2e-16 | <2.2e-16 | 2698 | 2019 | 611 | 68 |
| European | CIDI-noMDD | ADHD | sex_at_birthMale | 1.379 | 1.137 | 1.671 | 1.06e-03 | 7.45e-03 | 2698 | 2019 | 611 | 68 |
| European | CIDI-noMDD | ADHD | Income_quantitative | 0.8882 | 0.8466 | 0.9318 | 1.25e-06 | 8.77e-06 | 2698 | 2019 | 611 | 68 |
| European | CIDI-noMDD | ADHD | Education_levelCollege Undergraduate | 1.132 | 0.921 | 1.392 | 2.38e-01 | 1.00e+00 | 2698 | 2019 | 611 | 68 |
| European | CIDI-noMDD | ADHD | Education_levelLess than a high school degree | 1.385 | 0.4336 | 4.424 | 5.83e-01 | 1.00e+00 | 2698 | 2019 | 611 | 68 |
| European | CIDI-noMDD | ADHD | Education_levelTwelve or GED | 1.307 | 0.9307 | 1.835 | 1.22e-01 | 8.56e-01 | 2698 | 2019 | 611 | 68 |
| European | CIDI-noMDD | ADHD | ADHD | 1.059 | 0.9666 | 1.16 | 2.19e-01 | 1.00e+00 | 2698 | 2019 | 611 | 68 |
| European | CIDI-noMDD | BMI | age_at_first_AD | 0.9709 | 0.9646 | 0.9771 | <2.2e-16 | <2.2e-16 | 2698 | 2019 | 611 | 68 |
| European | CIDI-noMDD | BMI | sex_at_birthMale | 1.376 | 1.135 | 1.667 | 1.15e-03 | 8.02e-03 | 2698 | 2019 | 611 | 68 |
| European | CIDI-noMDD | BMI | Income_quantitative | 0.8872 | 0.8457 | 0.9309 | 1.04e-06 | 7.28e-06 | 2698 | 2019 | 611 | 68 |
| European | CIDI-noMDD | BMI | Education_levelCollege Undergraduate | 1.143 | 0.9303 | 1.404 | 2.03e-01 | 1.00e+00 | 2698 | 2019 | 611 | 68 |
| European | CIDI-noMDD | BMI | Education_levelLess than a high school degree | 1.447 | 0.4548 | 4.606 | 5.31e-01 | 1.00e+00 | 2698 | 2019 | 611 | 68 |
| European | CIDI-noMDD | BMI | Education_levelTwelve or GED | 1.324 | 0.9434 | 1.858 | 1.05e-01 | 7.32e-01 | 2698 | 2019 | 611 | 68 |
| European | CIDI-noMDD | BMI | BMI | 1.02 | 0.9332 | 1.116 | 6.57e-01 | 1.00e+00 | 2698 | 2019 | 611 | 68 |
| European | CIDI-noMDD | Bipolar Disorder | age_at_first_AD | 0.9706 | 0.9644 | 0.9769 | <2.2e-16 | <2.2e-16 | 2698 | 2019 | 611 | 68 |
| European | CIDI-noMDD | Bipolar Disorder | sex_at_birthMale | 1.376 | 1.135 | 1.667 | 1.16e-03 | 8.09e-03 | 2698 | 2019 | 611 | 68 |
| European | CIDI-noMDD | Bipolar Disorder | Income_quantitative | 0.8878 | 0.8463 | 0.9314 | 1.11e-06 | 7.80e-06 | 2698 | 2019 | 611 | 68 |
| European | CIDI-noMDD | Bipolar Disorder | Education_levelCollege Undergraduate | 1.146 | 0.9325 | 1.407 | 1.95e-01 | 1.00e+00 | 2698 | 2019 | 611 | 68 |
| European | CIDI-noMDD | Bipolar Disorder | Education_levelLess than a high school degree | 1.417 | 0.443 | 4.535 | 5.57e-01 | 1.00e+00 | 2698 | 2019 | 611 | 68 |
| European | CIDI-noMDD | Bipolar Disorder | Education_levelTwelve or GED | 1.352 | 0.9625 | 1.898 | 8.19e-02 | 5.74e-01 | 2698 | 2019 | 611 | 68 |
| European | CIDI-noMDD | Bipolar Disorder | Bipolar Disorder | 1.079 | 0.9812 | 1.187 | 1.16e-01 | 8.14e-01 | 2698 | 2019 | 611 | 68 |
| European | CIDI-noMDD | Depression | age_at_first_AD | 0.9711 | 0.9649 | 0.9774 | <2.2e-16 | <2.2e-16 | 2698 | 2019 | 611 | 68 |
| European | CIDI-noMDD | Depression | sex_at_birthMale | 1.358 | 1.12 | 1.646 | 1.85e-03 | 1.30e-02 | 2698 | 2019 | 611 | 68 |
| European | CIDI-noMDD | Depression | Income_quantitative | 0.8899 | 0.8483 | 0.9336 | 1.86e-06 | 1.30e-05 | 2698 | 2019 | 611 | 68 |
| European | CIDI-noMDD | Depression | Education_levelCollege Undergraduate | 1.128 | 0.9179 | 1.386 | 2.52e-01 | 1.00e+00 | 2698 | 2019 | 611 | 68 |
| European | CIDI-noMDD | Depression | Education_levelLess than a high school degree | 1.294 | 0.4005 | 4.182 | 6.67e-01 | 1.00e+00 | 2698 | 2019 | 611 | 68 |
| European | CIDI-noMDD | Depression | Education_levelTwelve or GED | 1.283 | 0.9131 | 1.802 | 1.51e-01 | 1.00e+00 | 2698 | 2019 | 611 | 68 |

| | | | | | | | | | | | | |
|---|---|---|---|---|---|---|---|---|---|---|---|---|
| European | CIDI-noMDD | Depression | Depression | 1.152 | 1.047 | 1.267 | 3.58e-03 | 2.51e-02 | 2698 | 2019 | 611 | 68 |
| European | CIDI-noMDD | Insomnia | age_at_first_AD | 0.9709 | 0.9647 | 0.9772 | <2.2e-16 | <2.2e-16 | 2698 | 2019 | 611 | 68 |
| European | CIDI-noMDD | Insomnia | sex_at_birthMale | 1.373 | 1.133 | 1.665 | 1.22e-03 | 8.55e-03 | 2698 | 2019 | 611 | 68 |
| European | CIDI-noMDD | Insomnia | Income_quantitative | 0.89 | 0.8483 | 0.9338 | 1.98e-06 | 1.39e-05 | 2698 | 2019 | 611 | 68 |
| European | CIDI-noMDD | Insomnia | Education_levelCollege Undergraduate | 1.133 | 0.9219 | 1.392 | 2.35e-01 | 1.00e+00 | 2698 | 2019 | 611 | 68 |
| European | CIDI-noMDD | Insomnia | Education_levelLess than a high school degree | 1.387 | 0.4347 | 4.427 | 5.80e-01 | 1.00e+00 | 2698 | 2019 | 611 | 68 |
| European | CIDI-noMDD | Insomnia | Education_levelTwelve or GED | 1.309 | 0.9323 | 1.838 | 1.20e-01 | 8.39e-01 | 2698 | 2019 | 611 | 68 |
| European | CIDI-noMDD | Insomnia | Insomnia | 1.105 | 1.006 | 1.214 | 3.78e-02 | 2.64e-01 | 2698 | 2019 | 611 | 68 |
| European | CIDI-noMDD | Migraine | age_at_first_AD | 0.9708 | 0.9646 | 0.9771 | <2.2e-16 | <2.2e-16 | 2698 | 2019 | 611 | 68 |
| European | CIDI-noMDD | Migraine | sex_at_birthMale | 1.376 | 1.135 | 1.668 | 1.14e-03 | 7.96e-03 | 2698 | 2019 | 611 | 68 |
| European | CIDI-noMDD | Migraine | Income_quantitative | 0.8863 | 0.8449 | 0.9297 | 7.71e-07 | 5.40e-06 | 2698 | 2019 | 611 | 68 |
| European | CIDI-noMDD | Migraine | Education_levelCollege Undergraduate | 1.145 | 0.9318 | 1.406 | 1.98e-01 | 1.00e+00 | 2698 | 2019 | 611 | 68 |
| European | CIDI-noMDD | Migraine | Education_levelLess than a high school degree | 1.457 | 0.4576 | 4.638 | 5.24e-01 | 1.00e+00 | 2698 | 2019 | 611 | 68 |
| European | CIDI-noMDD | Migraine | Education_levelTwelve or GED | 1.327 | 0.9461 | 1.862 | 1.01e-01 | 7.08e-01 | 2698 | 2019 | 611 | 68 |
| European | CIDI-noMDD | Migraine | Migraine | 0.9946 | 0.9116 | 1.085 | 9.04e-01 | 1.00e+00 | 2698 | 2019 | 611 | 68 |
| European | CIDI-noMDD | Schizophrenia | age_at_first_AD | 0.9709 | 0.9646 | 0.9771 | <2.2e-16 | <2.2e-16 | 2698 | 2019 | 611 | 68 |
| European | CIDI-noMDD | Schizophrenia | sex_at_birthMale | 1.379 | 1.138 | 1.671 | 1.06e-03 | 7.44e-03 | 2698 | 2019 | 611 | 68 |
| European | CIDI-noMDD | Schizophrenia | Income_quantitative | 0.8864 | 0.845 | 0.9298 | 7.79e-07 | 5.45e-06 | 2698 | 2019 | 611 | 68 |
| European | CIDI-noMDD | Schizophrenia | Education_levelCollege Undergraduate | 1.143 | 0.9307 | 1.404 | 2.02e-01 | 1.00e+00 | 2698 | 2019 | 611 | 68 |
| European | CIDI-noMDD | Schizophrenia | Education_levelLess than a high school degree | 1.434 | 0.4497 | 4.573 | 5.42e-01 | 1.00e+00 | 2698 | 2019 | 611 | 68 |
| European | CIDI-noMDD | Schizophrenia | Education_levelTwelve or GED | 1.329 | 0.9475 | 1.865 | 9.94e-02 | 6.96e-01 | 2698 | 2019 | 611 | 68 |
| European | CIDI-noMDD | Schizophrenia | Schizophrenia | 1.046 | 0.9545 | 1.146 | 3.35e-01 | 1.00e+00 | 2698 | 2019 | 611 | 68 |
| African | Full Cohort | ADHD | age_at_first_AD | 0.9801 | 0.9767 | 0.9835 | <2.2e-16 | <2.2e-16 | 7216 | 4164 | 2078 | 974 |
| African | Full Cohort | ADHD | sex_at_birthMale | 1.137 | 1.022 | 1.264 | 1.80e-02 | 1.26e-01 | 7216 | 4164 | 2078 | 974 |
| African | Full Cohort | ADHD | Income_quantitative | 0.8666 | 0.8382 | 0.8959 | <2.2e-16 | 2.49e-16 | 7216 | 4164 | 2078 | 974 |
| African | Full Cohort | ADHD | Education_levelCollege Undergraduate | 0.8951 | 0.7535 | 1.063 | 2.08e-01 | 1.00e+00 | 7216 | 4164 | 2078 | 974 |
| African | Full Cohort | ADHD | Education_levelLess than a high school degree | 1.036 | 0.8288 | 1.294 | 7.58e-01 | 1.00e+00 | 7216 | 4164 | 2078 | 974 |
| African | Full Cohort | ADHD | Education_levelTwelve or GED | 0.8952 | 0.7392 | 1.084 | 2.57e-01 | 1.00e+00 | 7216 | 4164 | 2078 | 974 |
| African | Full Cohort | ADHD | ADHD | 1.045 | 0.9792 | 1.116 | 1.83e-01 | 1.00e+00 | 7216 | 4164 | 2078 | 974 |
| African | Full Cohort | BMI | age_at_first_AD | 0.98 | 0.9766 | 0.9835 | <2.2e-16 | <2.2e-16 | 7216 | 4164 | 2078 | 974 |
| African | Full Cohort | BMI | sex_at_birthMale | 1.134 | 1.02 | 1.261 | 2.03e-02 | 1.42e-01 | 7216 | 4164 | 2078 | 974 |
| African | Full Cohort | BMI | Income_quantitative | 0.8658 | 0.8374 | 0.8951 | <2.2e-16 | <2.2e-16 | 7216 | 4164 | 2078 | 974 |
| African | Full Cohort | BMI | Education_levelCollege Undergraduate | 0.8991 | 0.7568 | 1.068 | 2.26e-01 | 1.00e+00 | 7216 | 4164 | 2078 | 974 |
| African | Full Cohort | BMI | Education_levelLess than a high school degree | 1.046 | 0.8371 | 1.306 | 6.94e-01 | 1.00e+00 | 7216 | 4164 | 2078 | 974 |
| African | Full Cohort | BMI | Education_levelTwelve or GED | 0.9007 | 0.7437 | 1.091 | 2.84e-01 | 1.00e+00 | 7216 | 4164 | 2078 | 974 |
| African | Full Cohort | BMI | BMI | 0.9819 | 0.9247 | 1.043 | 5.52e-01 | 1.00e+00 | 7216 | 4164 | 2078 | 974 |
| African | Full Cohort | Bipolar Disorder | age_at_first_AD | 0.98 | 0.9766 | 0.9834 | <2.2e-16 | <2.2e-16 | 7216 | 4164 | 2078 | 974 |
| African | Full Cohort | Bipolar Disorder | sex_at_birthMale | 1.135 | 1.021 | 1.262 | 1.95e-02 | 1.37e-01 | 7216 | 4164 | 2078 | 974 |
| African | Full Cohort | Bipolar Disorder | Income_quantitative | 0.8669 | 0.8385 | 0.8963 | <2.2e-16 | 3.08e-16 | 7216 | 4164 | 2078 | 974 |
| African | Full Cohort | Bipolar Disorder | Education_levelCollege Undergraduate | 0.9019 | 0.7592 | 1.071 | 2.40e-01 | 1.00e+00 | 7216 | 4164 | 2078 | 974 |
| African | Full Cohort | Bipolar Disorder | Education_levelLess than a high school degree | 1.047 | 0.8383 | 1.308 | 6.84e-01 | 1.00e+00 | 7216 | 4164 | 2078 | 974 |
| African | Full Cohort | Bipolar Disorder | Education_levelTwelve or GED | 0.9067 | 0.7487 | 1.098 | 3.16e-01 | 1.00e+00 | 7216 | 4164 | 2078 | 974 |
| African | Full Cohort | Bipolar Disorder | Bipolar Disorder | 1.105 | 1.034 | 1.182 | 3.33e-03 | 2.33e-02 | 7216 | 4164 | 2078 | 974 |
| African | Full Cohort | Depression | age_at_first_AD | 0.9801 | 0.9767 | 0.9835 | <2.2e-16 | <2.2e-16 | 7216 | 4164 | 2078 | 974 |
| African | Full Cohort | Depression | sex_at_birthMale | 1.136 | 1.022 | 1.264 | 1.86e-02 | 1.30e-01 | 7216 | 4164 | 2078 | 974 |
| African | Full Cohort | Depression | Income_quantitative | 0.8679 | 0.8395 | 0.8973 | <2.2e-16 | 5.37e-16 | 7216 | 4164 | 2078 | 974 |
| African | Full Cohort | Depression | Education_levelCollege Undergraduate | 0.8947 | 0.7531 | 1.063 | 2.06e-01 | 1.00e+00 | 7216 | 4164 | 2078 | 974 |
| African | Full Cohort | Depression | Education_levelLess than a high school degree | 1.037 | 0.8298 | 1.295 | 7.51e-01 | 1.00e+00 | 7216 | 4164 | 2078 | 974 |
| African | Full Cohort | Depression | Education_levelTwelve or GED | 0.8986 | 0.7419 | 1.088 | 2.74e-01 | 1.00e+00 | 7216 | 4164 | 2078 | 974 |
| African | Full Cohort | Depression | Depression | 1.134 | 1.063 | 1.21 | 1.42e-04 | 9.95e-04 | 7216 | 4164 | 2078 | 974 |
| African | Full Cohort | Insomnia | age_at_first_AD | 0.98 | 0.9766 | 0.9835 | <2.2e-16 | <2.2e-16 | 7216 | 4164 | 2078 | 974 |
| African | Full Cohort | Insomnia | sex_at_birthMale | 1.138 | 1.024 | 1.266 | 1.69e-02 | 1.18e-01 | 7216 | 4164 | 2078 | 974 |
| African | Full Cohort | Insomnia | Income_quantitative | 0.8664 | 0.838 | 0.8957 | <2.2e-16 | <2.2e-16 | 7216 | 4164 | 2078 | 974 |
| African | Full Cohort | Insomnia | Education_levelCollege Undergraduate | 0.8918 | 0.7505 | 1.06 | 1.93e-01 | 1.00e+00 | 7216 | 4164 | 2078 | 974 |
| African | Full Cohort | Insomnia | Education_levelLess than a high school degree | 1.03 | 0.8247 | 1.287 | 7.92e-01 | 1.00e+00 | 7216 | 4164 | 2078 | 974 |
| African | Full Cohort | Insomnia | Education_levelTwelve or GED | 0.8934 | 0.7377 | 1.082 | 2.49e-01 | 1.00e+00 | 7216 | 4164 | 2078 | 974 |
| African | Full Cohort | Insomnia | Insomnia | 1.144 | 1.071 | 1.223 | 7.28e-05 | 5.10e-04 | 7216 | 4164 | 2078 | 974 |

| | | | | | | | | | | | | |
|---|---|---|---|---|---|---|---|---|---|---|---|---|
| African | Full Cohort | Migraine | age_at_first_AD | 0.9801 | 0.9766 | 0.9835 | <2.2e-16 | <2.2e-16 | 7216 | 4164 | 2078 | 974 |
| African | Full Cohort | Migraine | sex_at_birthMale | 1.136 | 1.021 | 1.263 | 1.87e-02 | 1.31e-01 | 7216 | 4164 | 2078 | 974 |
| African | Full Cohort | Migraine | Income_quantitative | 0.866 | 0.8377 | 0.8953 | <2.2e-16 | <2.2e-16 | 7216 | 4164 | 2078 | 974 |
| African | Full Cohort | Migraine | Education_levelCollege Undergraduate | 0.8976 | 0.7556 | 1.066 | 2.19e-01 | 1.00e+00 | 7216 | 4164 | 2078 | 974 |
| African | Full Cohort | Migraine | Education_levelLess than a high school degree | 1.043 | 0.8349 | 1.303 | 7.11e-01 | 1.00e+00 | 7216 | 4164 | 2078 | 974 |
| African | Full Cohort | Migraine | Education_levelTwelve or GED | 0.8987 | 0.7422 | 1.088 | 2.74e-01 | 1.00e+00 | 7216 | 4164 | 2078 | 974 |
| African | Full Cohort | Migraine | Migraine | 0.9884 | 0.9347 | 1.045 | 6.83e-01 | 1.00e+00 | 7216 | 4164 | 2078 | 974 |
| African | Full Cohort | Schizophrenia | age_at_first_AD | 0.9801 | 0.9766 | 0.9835 | <2.2e-16 | <2.2e-16 | 7216 | 4164 | 2078 | 974 |
| African | Full Cohort | Schizophrenia | sex_at_birthMale | 1.136 | 1.021 | 1.263 | 1.88e-02 | 1.31e-01 | 7216 | 4164 | 2078 | 974 |
| African | Full Cohort | Schizophrenia | Income_quantitative | 0.866 | 0.8376 | 0.8954 | <2.2e-16 | <2.2e-16 | 7216 | 4164 | 2078 | 974 |
| African | Full Cohort | Schizophrenia | Education_levelCollege Undergraduate | 0.8978 | 0.7558 | 1.067 | 2.20e-01 | 1.00e+00 | 7216 | 4164 | 2078 | 974 |
| African | Full Cohort | Schizophrenia | Education_levelLess than a high school degree | 1.043 | 0.8353 | 1.303 | 7.08e-01 | 1.00e+00 | 7216 | 4164 | 2078 | 974 |
| African | Full Cohort | Schizophrenia | Education_levelTwelve or GED | 0.8987 | 0.7422 | 1.088 | 2.74e-01 | 1.00e+00 | 7216 | 4164 | 2078 | 974 |
| African | Full Cohort | Schizophrenia | Schizophrenia | 0.9966 | 0.9402 | 1.056 | 9.09e-01 | 1.00e+00 | 7216 | 4164 | 2078 | 974 |
| African | Recorded Depression | ADHD | age_at_first_AD | 0.9875 | 0.983 | 0.992 | 7.69e-08 | 5.38e-07 | 3912 | 1728 | 1510 | 674 |
| African | Recorded Depression | ADHD | sex_at_birthMale | 1.228 | 1.067 | 1.414 | 4.22e-03 | 2.96e-02 | 3912 | 1728 | 1510 | 674 |
| African | Recorded Depression | ADHD | Income_quantitative | 0.8767 | 0.8396 | 0.9155 | 2.52e-09 | 1.76e-08 | 3912 | 1728 | 1510 | 674 |
| African | Recorded Depression | ADHD | Education_levelCollege Undergraduate | 0.8308 | 0.6643 | 1.039 | 1.04e-01 | 7.29e-01 | 3912 | 1728 | 1510 | 674 |
| African | Recorded Depression | ADHD | Education_levelLess than a high school degree | 1.01 | 0.7518 | 1.356 | 9.48e-01 | 1.00e+00 | 3912 | 1728 | 1510 | 674 |
| African | Recorded Depression | ADHD | Education_levelTwelve or GED | 0.8212 | 0.6408 | 1.052 | 1.19e-01 | 8.36e-01 | 3912 | 1728 | 1510 | 674 |
| African | Recorded Depression | ADHD | ADHD | 1.009 | 0.9266 | 1.098 | 8.38e-01 | 1.00e+00 | 3912 | 1728 | 1510 | 674 |
| African | Recorded Depression | BMI | age_at_first_AD | 0.9873 | 0.9827 | 0.9918 | 4.67e-08 | 3.27e-07 | 3912 | 1728 | 1510 | 674 |
| African | Recorded Depression | BMI | sex_at_birthMale | 1.218 | 1.057 | 1.402 | 6.24e-03 | 4.37e-02 | 3912 | 1728 | 1510 | 674 |
| African | Recorded Depression | BMI | Income_quantitative | 0.8758 | 0.8387 | 0.9146 | 1.97e-09 | 1.38e-08 | 3912 | 1728 | 1510 | 674 |
| African | Recorded Depression | BMI | Education_levelCollege Undergraduate | 0.8384 | 0.6704 | 1.049 | 1.22e-01 | 8.57e-01 | 3912 | 1728 | 1510 | 674 |
| African | Recorded Depression | BMI | Education_levelLess than a high school degree | 1.027 | 0.7644 | 1.379 | 8.62e-01 | 1.00e+00 | 3912 | 1728 | 1510 | 674 |
| African | Recorded Depression | BMI | Education_levelTwelve or GED | 0.8314 | 0.6488 | 1.065 | 1.44e-01 | 1.00e+00 | 3912 | 1728 | 1510 | 674 |
| African | Recorded Depression | BMI | BMI | 0.9305 | 0.8609 | 1.006 | 6.96e-02 | 4.87e-01 | 3912 | 1728 | 1510 | 674 |
| African | Recorded Depression | Bipolar Disorder | age_at_first_AD | 0.9874 | 0.9829 | 0.992 | 6.77e-08 | 4.74e-07 | 3912 | 1728 | 1510 | 674 |
| African | Recorded Depression | Bipolar Disorder | sex_at_birthMale | 1.224 | 1.063 | 1.409 | 5.03e-03 | 3.52e-02 | 3912 | 1728 | 1510 | 674 |
| African | Recorded Depression | Bipolar Disorder | Income_quantitative | 0.8772 | 0.84 | 0.916 | 2.97e-09 | 2.08e-08 | 3912 | 1728 | 1510 | 674 |
| African | Recorded Depression | Bipolar Disorder | Education_levelCollege Undergraduate | 0.8385 | 0.6704 | 1.049 | 1.23e-01 | 8.58e-01 | 3912 | 1728 | 1510 | 674 |
| African | Recorded Depression | Bipolar Disorder | Education_levelLess than a high school degree | 1.017 | 0.7575 | 1.365 | 9.11e-01 | 1.00e+00 | 3912 | 1728 | 1510 | 674 |
| African | Recorded Depression | Bipolar Disorder | Education_levelTwelve or GED | 0.8287 | 0.6468 | 1.062 | 1.37e-01 | 9.60e-01 | 3912 | 1728 | 1510 | 674 |
| African | Recorded Depression | Bipolar Disorder | Bipolar Disorder | 1.104 | 1.012 | 1.205 | 2.66e-02 | 1.86e-01 | 3912 | 1728 | 1510 | 674 |
| African | Recorded Depression | Depression | age_at_first_AD | 0.9875 | 0.983 | 0.9921 | 9.05e-08 | 6.33e-07 | 3912 | 1728 | 1510 | 674 |
| African | Recorded Depression | Depression | sex_at_birthMale | 1.227 | 1.066 | 1.413 | 4.40e-03 | 3.08e-02 | 3912 | 1728 | 1510 | 674 |
| African | Recorded Depression | Depression | Income_quantitative | 0.8779 | 0.8407 | 0.9167 | 3.63e-09 | 2.54e-08 | 3912 | 1728 | 1510 | 674 |
| African | Recorded Depression | Depression | Education_levelCollege Undergraduate | 0.8265 | 0.6608 | 1.034 | 9.49e-02 | 6.64e-01 | 3912 | 1728 | 1510 | 674 |
| African | Recorded Depression | Depression | Education_levelLess than a high school degree | 0.9992 | 0.7441 | 1.342 | 9.96e-01 | 1.00e+00 | 3912 | 1728 | 1510 | 674 |
| African | Recorded Depression | Depression | Education_levelTwelve or GED | 0.8176 | 0.6381 | 1.048 | 1.11e-01 | 7.78e-01 | 3912 | 1728 | 1510 | 674 |
| African | Recorded Depression | Depression | Depression | 1.117 | 1.026 | 1.216 | 1.05e-02 | 7.38e-02 | 3912 | 1728 | 1510 | 674 |
| African | Recorded Depression | Insomnia | age_at_first_AD | 0.9875 | 0.983 | 0.992 | 7.78e-08 | 5.44e-07 | 3912 | 1728 | 1510 | 674 |
| African | Recorded Depression | Insomnia | sex_at_birthMale | 1.233 | 1.071 | 1.42 | 3.58e-03 | 2.51e-02 | 3912 | 1728 | 1510 | 674 |
| African | Recorded Depression | Insomnia | Income_quantitative | 0.8764 | 0.8393 | 0.9152 | 2.30e-09 | 1.61e-08 | 3912 | 1728 | 1510 | 674 |
| African | Recorded Depression | Insomnia | Education_levelCollege Undergraduate | 0.8272 | 0.6615 | 1.034 | 9.63e-02 | 6.74e-01 | 3912 | 1728 | 1510 | 674 |
| African | Recorded Depression | Insomnia | Education_levelLess than a high school degree | 1.001 | 0.7457 | 1.344 | 9.93e-01 | 1.00e+00 | 3912 | 1728 | 1510 | 674 |
| African | Recorded Depression | Insomnia | Education_levelTwelve or GED | 0.8158 | 0.6368 | 1.045 | 1.07e-01 | 7.52e-01 | 3912 | 1728 | 1510 | 674 |
| African | Recorded Depression | Insomnia | Insomnia | 1.1 | 1.009 | 1.199 | 2.96e-02 | 2.07e-01 | 3912 | 1728 | 1510 | 674 |
| African | Recorded Depression | Migraine | age_at_first_AD | 0.9875 | 0.9829 | 0.992 | 7.60e-08 | 5.32e-07 | 3912 | 1728 | 1510 | 674 |
| African | Recorded Depression | Migraine | sex_at_birthMale | 1.228 | 1.067 | 1.414 | 4.23e-03 | 2.96e-02 | 3912 | 1728 | 1510 | 674 |
| African | Recorded Depression | Migraine | Income_quantitative | 0.8767 | 0.8396 | 0.9155 | 2.57e-09 | 1.80e-08 | 3912 | 1728 | 1510 | 674 |
| African | Recorded Depression | Migraine | Education_levelCollege Undergraduate | 0.8317 | 0.6651 | 1.04 | 1.06e-01 | 7.42e-01 | 3912 | 1728 | 1510 | 674 |
| African | Recorded Depression | Migraine | Education_levelLess than a high school degree | 1.012 | 0.7538 | 1.359 | 9.37e-01 | 1.00e+00 | 3912 | 1728 | 1510 | 674 |
| African | Recorded Depression | Migraine | Education_levelTwelve or GED | 0.8224 | 0.6419 | 1.054 | 1.22e-01 | 8.52e-01 | 3912 | 1728 | 1510 | 674 |
| African | Recorded Depression | Migraine | Migraine | 1.001 | 0.9307 | 1.076 | 9.81e-01 | 1.00e+00 | 3912 | 1728 | 1510 | 674 |
| African | Recorded Depression | Schizophrenia | age_at_first_AD | 0.9875 | 0.9829 | 0.992 | 7.58e-08 | 5.31e-07 | 3912 | 1728 | 1510 | 674 |

| | | | | | | | | | | | | |
|---|---|---|---|---|---|---|---|---|---|---|---|---|
| African | Recorded Depression | Schizophrenia | sex_at_birthMale | 1.228 | 1.067 | 1.414 | 4.21e-03 | 2.95e-02 | 3912 | 1728 | 1510 | 674 |
| African | Recorded Depression | Schizophrenia | Income_quantitative | 0.8767 | 0.8395 | 0.9155 | 2.52e-09 | 1.76e-08 | 3912 | 1728 | 1510 | 674 |
| African | Recorded Depression | Schizophrenia | Education_levelCollege Undergraduate | 0.8314 | 0.6649 | 1.04 | 1.05e-01 | 7.37e-01 | 3912 | 1728 | 1510 | 674 |
| African | Recorded Depression | Schizophrenia | Education_levelLess than a high school degree | 1.012 | 0.7538 | 1.358 | 9.38e-01 | 1.00e+00 | 3912 | 1728 | 1510 | 674 |
| African | Recorded Depression | Schizophrenia | Education_levelTwelve or GED | 0.822 | 0.6416 | 1.053 | 1.21e-01 | 8.47e-01 | 3912 | 1728 | 1510 | 674 |
| African | Recorded Depression | Schizophrenia | Schizophrenia | 0.9956 | 0.9233 | 1.074 | 9.09e-01 | 1.00e+00 | 3912 | 1728 | 1510 | 674 |
| African | No Recorded Depression | ADHD | age_at_first_AD | 0.9769 | 0.9711 | 0.9826 | 8.44e-15 | 5.91e-14 | 3304 | 2436 | 568 | 300 |
| African | No Recorded Depression | ADHD | sex_at_birthMale | 1.167 | 0.9797 | 1.39 | 8.36e-02 | 5.85e-01 | 3304 | 2436 | 568 | 300 |
| African | No Recorded Depression | ADHD | Income_quantitative | 0.8867 | 0.8377 | 0.9386 | 3.38e-05 | 2.37e-04 | 3304 | 2436 | 568 | 300 |
| African | No Recorded Depression | ADHD | Education_levelCollege Undergraduate | 0.9303 | 0.6919 | 1.251 | 6.32e-01 | 1.00e+00 | 3304 | 2436 | 568 | 300 |
| African | No Recorded Depression | ADHD | Education_levelLess than a high school degree | 1.306 | 0.9019 | 1.89 | 1.58e-01 | 1.00e+00 | 3304 | 2436 | 568 | 300 |
| African | No Recorded Depression | ADHD | Education_levelTwelve or GED | 1.061 | 0.7641 | 1.473 | 7.24e-01 | 1.00e+00 | 3304 | 2436 | 568 | 300 |
| African | No Recorded Depression | ADHD | ADHD | 1.136 | 1.016 | 1.269 | 2.51e-02 | 1.76e-01 | 3304 | 2436 | 568 | 300 |
| African | No Recorded Depression | BMI | age_at_first_AD | 0.9767 | 0.9709 | 0.9825 | 5.52e-15 | 3.86e-14 | 3304 | 2436 | 568 | 300 |
| African | No Recorded Depression | BMI | sex_at_birthMale | 1.165 | 0.9776 | 1.387 | 8.79e-02 | 6.15e-01 | 3304 | 2436 | 568 | 300 |
| African | No Recorded Depression | BMI | Income_quantitative | 0.8847 | 0.8359 | 0.9363 | 2.32e-05 | 1.63e-04 | 3304 | 2436 | 568 | 300 |
| African | No Recorded Depression | BMI | Education_levelCollege Undergraduate | 0.9321 | 0.6934 | 1.253 | 6.41e-01 | 1.00e+00 | 3304 | 2436 | 568 | 300 |
| African | No Recorded Depression | BMI | Education_levelLess than a high school degree | 1.315 | 0.9085 | 1.903 | 1.47e-01 | 1.00e+00 | 3304 | 2436 | 568 | 300 |
| African | No Recorded Depression | BMI | Education_levelTwelve or GED | 1.058 | 0.7626 | 1.469 | 7.34e-01 | 1.00e+00 | 3304 | 2436 | 568 | 300 |
| African | No Recorded Depression | BMI | BMI | 1.081 | 0.9756 | 1.197 | 1.37e-01 | 9.59e-01 | 3304 | 2436 | 568 | 300 |
| African | No Recorded Depression | Bipolar Disorder | age_at_first_AD | 0.9766 | 0.9708 | 0.9824 | 4.13e-15 | 2.89e-14 | 3304 | 2436 | 568 | 300 |
| African | No Recorded Depression | Bipolar Disorder | sex_at_birthMale | 1.165 | 0.9784 | 1.388 | 8.62e-02 | 6.03e-01 | 3304 | 2436 | 568 | 300 |
| African | No Recorded Depression | Bipolar Disorder | Income_quantitative | 0.8846 | 0.8358 | 0.9363 | 2.33e-05 | 1.63e-04 | 3304 | 2436 | 568 | 300 |
| African | No Recorded Depression | Bipolar Disorder | Education_levelCollege Undergraduate | 0.9319 | 0.6932 | 1.253 | 6.40e-01 | 1.00e+00 | 3304 | 2436 | 568 | 300 |
| African | No Recorded Depression | Bipolar Disorder | Education_levelLess than a high school degree | 1.321 | 0.9128 | 1.912 | 1.40e-01 | 9.80e-01 | 3304 | 2436 | 568 | 300 |
| African | No Recorded Depression | Bipolar Disorder | Education_levelTwelve or GED | 1.073 | 0.7726 | 1.489 | 6.76e-01 | 1.00e+00 | 3304 | 2436 | 568 | 300 |
| African | No Recorded Depression | Bipolar Disorder | Bipolar Disorder | 1.108 | 0.9898 | 1.241 | 7.46e-02 | 5.22e-01 | 3304 | 2436 | 568 | 300 |
| African | No Recorded Depression | Depression | age_at_first_AD | 0.9766 | 0.9708 | 0.9824 | 4.17e-15 | 2.92e-14 | 3304 | 2436 | 568 | 300 |
| African | No Recorded Depression | Depression | sex_at_birthMale | 1.163 | 0.9761 | 1.385 | 9.13e-02 | 6.39e-01 | 3304 | 2436 | 568 | 300 |
| African | No Recorded Depression | Depression | Income_quantitative | 0.8858 | 0.8369 | 0.9375 | 2.82e-05 | 1.97e-04 | 3304 | 2436 | 568 | 300 |
| African | No Recorded Depression | Depression | Education_levelCollege Undergraduate | 0.9324 | 0.6935 | 1.254 | 6.43e-01 | 1.00e+00 | 3304 | 2436 | 568 | 300 |
| African | No Recorded Depression | Depression | Education_levelLess than a high school degree | 1.322 | 0.9134 | 1.913 | 1.39e-01 | 9.73e-01 | 3304 | 2436 | 568 | 300 |
| African | No Recorded Depression | Depression | Education_levelTwelve or GED | 1.069 | 0.7698 | 1.484 | 6.91e-01 | 1.00e+00 | 3304 | 2436 | 568 | 300 |
| African | No Recorded Depression | Depression | Depression | 1.139 | 1.02 | 1.271 | 2.05e-02 | 1.43e-01 | 3304 | 2436 | 568 | 300 |
| African | No Recorded Depression | Insomnia | age_at_first_AD | 0.9765 | 0.9708 | 0.9823 | 3.68e-15 | 2.57e-14 | 3304 | 2436 | 568 | 300 |
| African | No Recorded Depression | Insomnia | sex_at_birthMale | 1.157 | 0.9715 | 1.379 | 1.02e-01 | 7.12e-01 | 3304 | 2436 | 568 | 300 |
| African | No Recorded Depression | Insomnia | Income_quantitative | 0.8848 | 0.836 | 0.9364 | 2.35e-05 | 1.64e-04 | 3304 | 2436 | 568 | 300 |
| African | No Recorded Depression | Insomnia | Education_levelCollege Undergraduate | 0.9276 | 0.6898 | 1.247 | 6.19e-01 | 1.00e+00 | 3304 | 2436 | 568 | 300 |
| African | No Recorded Depression | Insomnia | Education_levelLess than a high school degree | 1.306 | 0.9024 | 1.892 | 1.57e-01 | 1.00e+00 | 3304 | 2436 | 568 | 300 |
| African | No Recorded Depression | Insomnia | Education_levelTwelve or GED | 1.063 | 0.7656 | 1.476 | 7.15e-01 | 1.00e+00 | 3304 | 2436 | 568 | 300 |
| African | No Recorded Depression | Insomnia | Insomnia | 1.176 | 1.049 | 1.317 | 5.25e-03 | 3.67e-02 | 3304 | 2436 | 568 | 300 |
| African | No Recorded Depression | Migraine | age_at_first_AD | 0.9767 | 0.9709 | 0.9825 | 4.90e-15 | 3.43e-14 | 3304 | 2436 | 568 | 300 |
| African | No Recorded Depression | Migraine | sex_at_birthMale | 1.16 | 0.9742 | 1.382 | 9.54e-02 | 6.68e-01 | 3304 | 2436 | 568 | 300 |
| African | No Recorded Depression | Migraine | Income_quantitative | 0.8835 | 0.8348 | 0.9351 | 1.88e-05 | 1.32e-04 | 3304 | 2436 | 568 | 300 |
| African | No Recorded Depression | Migraine | Education_levelCollege Undergraduate | 0.9344 | 0.6951 | 1.256 | 6.53e-01 | 1.00e+00 | 3304 | 2436 | 568 | 300 |
| African | No Recorded Depression | Migraine | Education_levelLess than a high school degree | 1.322 | 0.9138 | 1.914 | 1.38e-01 | 9.69e-01 | 3304 | 2436 | 568 | 300 |
| African | No Recorded Depression | Migraine | Education_levelTwelve or GED | 1.064 | 0.7666 | 1.477 | 7.11e-01 | 1.00e+00 | 3304 | 2436 | 568 | 300 |
| African | No Recorded Depression | Migraine | Migraine | 0.9699 | 0.882 | 1.067 | 5.29e-01 | 1.00e+00 | 3304 | 2436 | 568 | 300 |
| African | No Recorded Depression | Schizophrenia | age_at_first_AD | 0.9767 | 0.9709 | 0.9824 | 4.64e-15 | 3.25e-14 | 3304 | 2436 | 568 | 300 |
| African | No Recorded Depression | Schizophrenia | sex_at_birthMale | 1.161 | 0.9746 | 1.383 | 9.46e-02 | 6.62e-01 | 3304 | 2436 | 568 | 300 |
| African | No Recorded Depression | Schizophrenia | Income_quantitative | 0.8841 | 0.8353 | 0.9358 | 2.14e-05 | 1.50e-04 | 3304 | 2436 | 568 | 300 |
| African | No Recorded Depression | Schizophrenia | Education_levelCollege Undergraduate | 0.9339 | 0.6948 | 1.255 | 6.51e-01 | 1.00e+00 | 3304 | 2436 | 568 | 300 |
| African | No Recorded Depression | Schizophrenia | Education_levelLess than a high school degree | 1.319 | 0.9113 | 1.908 | 1.42e-01 | 9.96e-01 | 3304 | 2436 | 568 | 300 |
| African | No Recorded Depression | Schizophrenia | Education_levelTwelve or GED | 1.065 | 0.7671 | 1.478 | 7.08e-01 | 1.00e+00 | 3304 | 2436 | 568 | 300 |
| African | No Recorded Depression | Schizophrenia | Schizophrenia | 1.035 | 0.9364 | 1.143 | 5.03e-01 | 1.00e+00 | 3304 | 2436 | 568 | 300 |
| African | Recorded Anxiety | ADHD | age_at_first_AD | 0.9851 | 0.9807 | 0.9895 | 4.11e-11 | 2.88e-10 | 4213 | 2014 | 1536 | 663 |
| African | Recorded Anxiety | ADHD | sex_at_birthMale | 1.265 | 1.099 | 1.457 | 1.08e-03 | 7.53e-03 | 4213 | 2014 | 1536 | 663 |

| | | | | | | | | | | | | |
|---|---|---|---|---|---|---|---|---|---|---|---|---|
| African | Recorded Anxiety | ADHD | Income_quantitative | 0.8584 | 0.8243 | 0.894 | 1.57e-13 | 1.10e-12 | 4213 | 2014 | 1536 | 663 |
| African | Recorded Anxiety | ADHD | Education_levelCollege Undergraduate | 0.978 | 0.795 | 1.203 | 8.33e-01 | 1.00e+00 | 4213 | 2014 | 1536 | 663 |
| African | Recorded Anxiety | ADHD | Education_levelLess than a high school degree | 1.189 | 0.8939 | 1.582 | 2.34e-01 | 1.00e+00 | 4213 | 2014 | 1536 | 663 |
| African | Recorded Anxiety | ADHD | Education_levelTwelve or GED | 0.9456 | 0.7483 | 1.195 | 6.39e-01 | 1.00e+00 | 4213 | 2014 | 1536 | 663 |
| African | Recorded Anxiety | ADHD | ADHD | 1.009 | 0.9294 | 1.095 | 8.34e-01 | 1.00e+00 | 4213 | 2014 | 1536 | 663 |
| African | Recorded Anxiety | BMI | age_at_first_AD | 0.985 | 0.9806 | 0.9894 | 3.16e-11 | 2.22e-10 | 4213 | 2014 | 1536 | 663 |
| African | Recorded Anxiety | BMI | sex_at_birthMale | 1.26 | 1.094 | 1.451 | 1.32e-03 | 9.25e-03 | 4213 | 2014 | 1536 | 663 |
| African | Recorded Anxiety | BMI | Income_quantitative | 0.8578 | 0.8237 | 0.8933 | 1.26e-13 | 8.83e-13 | 4213 | 2014 | 1536 | 663 |
| African | Recorded Anxiety | BMI | Education_levelCollege Undergraduate | 0.9826 | 0.7988 | 1.209 | 8.68e-01 | 1.00e+00 | 4213 | 2014 | 1536 | 663 |
| African | Recorded Anxiety | BMI | Education_levelLess than a high school degree | 1.197 | 0.9002 | 1.592 | 2.16e-01 | 1.00e+00 | 4213 | 2014 | 1536 | 663 |
| African | Recorded Anxiety | BMI | Education_levelTwelve or GED | 0.9522 | 0.7535 | 1.203 | 6.82e-01 | 1.00e+00 | 4213 | 2014 | 1536 | 663 |
| African | Recorded Anxiety | BMI | BMI | 0.9611 | 0.8914 | 1.036 | 3.00e-01 | 1.00e+00 | 4213 | 2014 | 1536 | 663 |
| African | Recorded Anxiety | Bipolar Disorder | age_at_first_AD | 0.985 | 0.9806 | 0.9894 | 3.12e-11 | 2.19e-10 | 4213 | 2014 | 1536 | 663 |
| African | Recorded Anxiety | Bipolar Disorder | sex_at_birthMale | 1.262 | 1.096 | 1.454 | 1.20e-03 | 8.38e-03 | 4213 | 2014 | 1536 | 663 |
| African | Recorded Anxiety | Bipolar Disorder | Income_quantitative | 0.8591 | 0.825 | 0.8947 | 2.16e-13 | 1.51e-12 | 4213 | 2014 | 1536 | 663 |
| African | Recorded Anxiety | Bipolar Disorder | Education_levelCollege Undergraduate | 0.9803 | 0.797 | 1.206 | 8.51e-01 | 1.00e+00 | 4213 | 2014 | 1536 | 663 |
| African | Recorded Anxiety | Bipolar Disorder | Education_levelLess than a high school degree | 1.192 | 0.8962 | 1.585 | 2.28e-01 | 1.00e+00 | 4213 | 2014 | 1536 | 663 |
| African | Recorded Anxiety | Bipolar Disorder | Education_levelTwelve or GED | 0.9506 | 0.7523 | 1.201 | 6.71e-01 | 1.00e+00 | 4213 | 2014 | 1536 | 663 |
| African | Recorded Anxiety | Bipolar Disorder | Bipolar Disorder | 1.067 | 0.9803 | 1.161 | 1.34e-01 | 9.40e-01 | 4213 | 2014 | 1536 | 663 |
| African | Recorded Anxiety | Depression | age_at_first_AD | 0.9851 | 0.9808 | 0.9895 | 4.59e-11 | 3.21e-10 | 4213 | 2014 | 1536 | 663 |
| African | Recorded Anxiety | Depression | sex_at_birthMale | 1.264 | 1.097 | 1.455 | 1.14e-03 | 8.01e-03 | 4213 | 2014 | 1536 | 663 |
| African | Recorded Anxiety | Depression | Income_quantitative | 0.86 | 0.8259 | 0.8956 | 3.13e-13 | 2.19e-12 | 4213 | 2014 | 1536 | 663 |
| African | Recorded Anxiety | Depression | Education_levelCollege Undergraduate | 0.9747 | 0.7923 | 1.199 | 8.09e-01 | 1.00e+00 | 4213 | 2014 | 1536 | 663 |
| African | Recorded Anxiety | Depression | Education_levelLess than a high school degree | 1.18 | 0.8874 | 1.57 | 2.55e-01 | 1.00e+00 | 4213 | 2014 | 1536 | 663 |
| African | Recorded Anxiety | Depression | Education_levelTwelve or GED | 0.9437 | 0.7468 | 1.193 | 6.28e-01 | 1.00e+00 | 4213 | 2014 | 1536 | 663 |
| African | Recorded Anxiety | Depression | Depression | 1.121 | 1.033 | 1.217 | 6.41e-03 | 4.49e-02 | 4213 | 2014 | 1536 | 663 |
| African | Recorded Anxiety | Insomnia | age_at_first_AD | 0.985 | 0.9806 | 0.9894 | 3.08e-11 | 2.16e-10 | 4213 | 2014 | 1536 | 663 |
| African | Recorded Anxiety | Insomnia | sex_at_birthMale | 1.271 | 1.104 | 1.464 | 8.55e-04 | 5.98e-03 | 4213 | 2014 | 1536 | 663 |
| African | Recorded Anxiety | Insomnia | Income_quantitative | 0.8575 | 0.8234 | 0.893 | 1.07e-13 | 7.52e-13 | 4213 | 2014 | 1536 | 663 |
| African | Recorded Anxiety | Insomnia | Education_levelCollege Undergraduate | 0.9657 | 0.7849 | 1.188 | 7.42e-01 | 1.00e+00 | 4213 | 2014 | 1536 | 663 |
| African | Recorded Anxiety | Insomnia | Education_levelLess than a high school degree | 1.172 | 0.8813 | 1.559 | 2.75e-01 | 1.00e+00 | 4213 | 2014 | 1536 | 663 |
| African | Recorded Anxiety | Insomnia | Education_levelTwelve or GED | 0.9312 | 0.7368 | 1.177 | 5.51e-01 | 1.00e+00 | 4213 | 2014 | 1536 | 663 |
| African | Recorded Anxiety | Insomnia | Insomnia | 1.149 | 1.058 | 1.249 | 1.04e-03 | 7.29e-03 | 4213 | 2014 | 1536 | 663 |
| African | Recorded Anxiety | Migraine | age_at_first_AD | 0.9851 | 0.9807 | 0.9895 | 4.00e-11 | 2.80e-10 | 4213 | 2014 | 1536 | 663 |
| African | Recorded Anxiety | Migraine | sex_at_birthMale | 1.265 | 1.099 | 1.456 | 1.08e-03 | 7.56e-03 | 4213 | 2014 | 1536 | 663 |
| African | Recorded Anxiety | Migraine | Income_quantitative | 0.8584 | 0.8243 | 0.8939 | 1.55e-13 | 1.08e-12 | 4213 | 2014 | 1536 | 663 |
| African | Recorded Anxiety | Migraine | Education_levelCollege Undergraduate | 0.9786 | 0.7956 | 1.204 | 8.38e-01 | 1.00e+00 | 4213 | 2014 | 1536 | 663 |
| African | Recorded Anxiety | Migraine | Education_levelLess than a high school degree | 1.191 | 0.896 | 1.584 | 2.28e-01 | 1.00e+00 | 4213 | 2014 | 1536 | 663 |
| African | Recorded Anxiety | Migraine | Education_levelTwelve or GED | 0.9465 | 0.7492 | 1.196 | 6.45e-01 | 1.00e+00 | 4213 | 2014 | 1536 | 663 |
| African | Recorded Anxiety | Migraine | Migraine | 1.003 | 0.9351 | 1.076 | 9.30e-01 | 1.00e+00 | 4213 | 2014 | 1536 | 663 |
| African | Recorded Anxiety | Schizophrenia | age_at_first_AD | 0.9852 | 0.9808 | 0.9896 | 4.69e-11 | 3.28e-10 | 4213 | 2014 | 1536 | 663 |
| African | Recorded Anxiety | Schizophrenia | sex_at_birthMale | 1.266 | 1.099 | 1.457 | 1.04e-03 | 7.30e-03 | 4213 | 2014 | 1536 | 663 |
| African | Recorded Anxiety | Schizophrenia | Income_quantitative | 0.858 | 0.8239 | 0.8935 | 1.34e-13 | 9.38e-13 | 4213 | 2014 | 1536 | 663 |
| African | Recorded Anxiety | Schizophrenia | Education_levelCollege Undergraduate | 0.978 | 0.7951 | 1.203 | 8.33e-01 | 1.00e+00 | 4213 | 2014 | 1536 | 663 |
| African | Recorded Anxiety | Schizophrenia | Education_levelLess than a high school degree | 1.194 | 0.8981 | 1.588 | 2.22e-01 | 1.00e+00 | 4213 | 2014 | 1536 | 663 |
| African | Recorded Anxiety | Schizophrenia | Education_levelTwelve or GED | 0.9448 | 0.7478 | 1.194 | 6.34e-01 | 1.00e+00 | 4213 | 2014 | 1536 | 663 |
| African | Recorded Anxiety | Schizophrenia | Schizophrenia | 0.9502 | 0.8825 | 1.023 | 1.75e-01 | 1.00e+00 | 4213 | 2014 | 1536 | 663 |
| Latino/Mixed-American | Full Cohort | ADHD | age_at_first_AD | 0.9883 | 0.984 | 0.9925 | 7.70e-08 | 5.39e-07 | 4199 | 2456 | 1228 | 515 |
| Latino/Mixed-American | Full Cohort | ADHD | sex_at_birthMale | 1.231 | 1.074 | 1.411 | 2.90e-03 | 2.03e-02 | 4199 | 2456 | 1228 | 515 |
| Latino/Mixed-American | Full Cohort | ADHD | Income_quantitative | 0.9002 | 0.8687 | 0.9328 | 6.88e-09 | 4.82e-08 | 4199 | 2456 | 1228 | 515 |
| Latino/Mixed-American | Full Cohort | ADHD | Education_levelCollege Undergraduate | 1.006 | 0.8117 | 1.248 | 9.54e-01 | 1.00e+00 | 4199 | 2456 | 1228 | 515 |
| Latino/Mixed-American | Full Cohort | ADHD | Education_levelLess than a high school degree | 0.9812 | 0.7546 | 1.276 | 8.87e-01 | 1.00e+00 | 4199 | 2456 | 1228 | 515 |
| Latino/Mixed-American | Full Cohort | ADHD | Education_levelTwelve or GED | 0.9052 | 0.7044 | 1.163 | 4.36e-01 | 1.00e+00 | 4199 | 2456 | 1228 | 515 |
| Latino/Mixed-American | Full Cohort | ADHD | ADHD | 1.051 | 0.9841 | 1.123 | 1.38e-01 | 9.63e-01 | 4199 | 2456 | 1228 | 515 |
| Latino/Mixed-American | Full Cohort | BMI | age_at_first_AD | 0.9882 | 0.9839 | 0.9924 | 6.07e-08 | 4.25e-07 | 4199 | 2456 | 1228 | 515 |
| Latino/Mixed-American | Full Cohort | BMI | sex_at_birthMale | 1.232 | 1.075 | 1.413 | 2.71e-03 | 1.90e-02 | 4199 | 2456 | 1228 | 515 |
| Latino/Mixed-American | Full Cohort | BMI | Income_quantitative | 0.9 | 0.8685 | 0.9327 | 6.79e-09 | 4.75e-08 | 4199 | 2456 | 1228 | 515 |

| | | | | | | | | | | | | |
|---|---|---|---|---|---|---|---|---|---|---|---|---|
| Latino/Mixed-American | Full Cohort | BMI | Education_levelCollege Undergraduate | 1.008 | 0.8134 | 1.25 | 9.39e-01 | 1.00e+00 | 4199 | 2456 | 1228 | 515 |
| Latino/Mixed-American | Full Cohort | BMI | Education_levelLess than a high school degree | 0.9908 | 0.7623 | 1.288 | 9.45e-01 | 1.00e+00 | 4199 | 2456 | 1228 | 515 |
| Latino/Mixed-American | Full Cohort | BMI | Education_levelTwelve or GED | 0.9103 | 0.7085 | 1.17 | 4.63e-01 | 1.00e+00 | 4199 | 2456 | 1228 | 515 |
| Latino/Mixed-American | Full Cohort | BMI | BMI | 1.022 | 0.9599 | 1.089 | 4.93e-01 | 1.00e+00 | 4199 | 2456 | 1228 | 515 |
| Latino/Mixed-American | Full Cohort | Bipolar Disorder | age_at_first_AD | 0.9881 | 0.9839 | 0.9924 | 5.30e-08 | 3.71e-07 | 4199 | 2456 | 1228 | 515 |
| Latino/Mixed-American | Full Cohort | Bipolar Disorder | sex_at_birthMale | 1.233 | 1.076 | 1.414 | 2.63e-03 | 1.84e-02 | 4199 | 2456 | 1228 | 515 |
| Latino/Mixed-American | Full Cohort | Bipolar Disorder | Income_quantitative | 0.9008 | 0.8693 | 0.9334 | 8.58e-09 | 6.01e-08 | 4199 | 2456 | 1228 | 515 |
| Latino/Mixed-American | Full Cohort | Bipolar Disorder | Education_levelCollege Undergraduate | 1.021 | 0.8236 | 1.266 | 8.47e-01 | 1.00e+00 | 4199 | 2456 | 1228 | 515 |
| Latino/Mixed-American | Full Cohort | Bipolar Disorder | Education_levelLess than a high school degree | 1.002 | 0.7706 | 1.302 | 9.89e-01 | 1.00e+00 | 4199 | 2456 | 1228 | 515 |
| Latino/Mixed-American | Full Cohort | Bipolar Disorder | Education_levelTwelve or GED | 0.9227 | 0.718 | 1.186 | 5.30e-01 | 1.00e+00 | 4199 | 2456 | 1228 | 515 |
| Latino/Mixed-American | Full Cohort | Bipolar Disorder | Bipolar Disorder | 1.068 | 0.9992 | 1.142 | 5.27e-02 | 3.69e-01 | 4199 | 2456 | 1228 | 515 |
| Latino/Mixed-American | Full Cohort | Depression | age_at_first_AD | 0.9882 | 0.984 | 0.9925 | 7.33e-08 | 5.13e-07 | 4199 | 2456 | 1228 | 515 |
| Latino/Mixed-American | Full Cohort | Depression | sex_at_birthMale | 1.234 | 1.076 | 1.415 | 2.61e-03 | 1.83e-02 | 4199 | 2456 | 1228 | 515 |
| Latino/Mixed-American | Full Cohort | Depression | Income_quantitative | 0.9028 | 0.8712 | 0.9355 | 1.85e-08 | 1.30e-07 | 4199 | 2456 | 1228 | 515 |
| Latino/Mixed-American | Full Cohort | Depression | Education_levelCollege Undergraduate | 1.013 | 0.8172 | 1.256 | 9.04e-01 | 1.00e+00 | 4199 | 2456 | 1228 | 515 |
| Latino/Mixed-American | Full Cohort | Depression | Education_levelLess than a high school degree | 0.9893 | 0.7609 | 1.286 | 9.36e-01 | 1.00e+00 | 4199 | 2456 | 1228 | 515 |
| Latino/Mixed-American | Full Cohort | Depression | Education_levelTwelve or GED | 0.9179 | 0.7142 | 1.18 | 5.03e-01 | 1.00e+00 | 4199 | 2456 | 1228 | 515 |
| Latino/Mixed-American | Full Cohort | Depression | Depression | 1.188 | 1.11 | 1.271 | 6.79e-07 | 4.75e-06 | 4199 | 2456 | 1228 | 515 |
| Latino/Mixed-American | Full Cohort | Insomnia | age_at_first_AD | 0.9882 | 0.984 | 0.9925 | 6.55e-08 | 4.58e-07 | 4199 | 2456 | 1228 | 515 |
| Latino/Mixed-American | Full Cohort | Insomnia | sex_at_birthMale | 1.234 | 1.077 | 1.415 | 2.54e-03 | 1.78e-02 | 4199 | 2456 | 1228 | 515 |
| Latino/Mixed-American | Full Cohort | Insomnia | Income_quantitative | 0.8998 | 0.8684 | 0.9324 | 6.00e-09 | 4.20e-08 | 4199 | 2456 | 1228 | 515 |
| Latino/Mixed-American | Full Cohort | Insomnia | Education_levelCollege Undergraduate | 1.004 | 0.8101 | 1.245 | 9.69e-01 | 1.00e+00 | 4199 | 2456 | 1228 | 515 |
| Latino/Mixed-American | Full Cohort | Insomnia | Education_levelLess than a high school degree | 0.9846 | 0.7575 | 1.28 | 9.08e-01 | 1.00e+00 | 4199 | 2456 | 1228 | 515 |
| Latino/Mixed-American | Full Cohort | Insomnia | Education_levelTwelve or GED | 0.9057 | 0.7049 | 1.164 | 4.39e-01 | 1.00e+00 | 4199 | 2456 | 1228 | 515 |
| Latino/Mixed-American | Full Cohort | Insomnia | Insomnia | 1.073 | 1.001 | 1.149 | 4.68e-02 | 3.27e-01 | 4199 | 2456 | 1228 | 515 |
| Latino/Mixed-American | Full Cohort | Migraine | age_at_first_AD | 0.9882 | 0.984 | 0.9925 | 6.89e-08 | 4.82e-07 | 4199 | 2456 | 1228 | 515 |
| Latino/Mixed-American | Full Cohort | Migraine | sex_at_birthMale | 1.232 | 1.074 | 1.412 | 2.80e-03 | 1.96e-02 | 4199 | 2456 | 1228 | 515 |
| Latino/Mixed-American | Full Cohort | Migraine | Income_quantitative | 0.8992 | 0.8677 | 0.9317 | 4.69e-09 | 3.28e-08 | 4199 | 2456 | 1228 | 515 |
| Latino/Mixed-American | Full Cohort | Migraine | Education_levelCollege Undergraduate | 1.01 | 0.8149 | 1.252 | 9.26e-01 | 1.00e+00 | 4199 | 2456 | 1228 | 515 |
| Latino/Mixed-American | Full Cohort | Migraine | Education_levelLess than a high school degree | 0.988 | 0.76 | 1.284 | 9.28e-01 | 1.00e+00 | 4199 | 2456 | 1228 | 515 |
| Latino/Mixed-American | Full Cohort | Migraine | Education_levelTwelve or GED | 0.9095 | 0.7078 | 1.169 | 4.58e-01 | 1.00e+00 | 4199 | 2456 | 1228 | 515 |
| Latino/Mixed-American | Full Cohort | Migraine | Migraine | 1.032 | 0.9698 | 1.099 | 3.18e-01 | 1.00e+00 | 4199 | 2456 | 1228 | 515 |
| Latino/Mixed-American | Full Cohort | Schizophrenia | age_at_first_AD | 0.9882 | 0.9839 | 0.9924 | 5.73e-08 | 4.01e-07 | 4199 | 2456 | 1228 | 515 |
| Latino/Mixed-American | Full Cohort | Schizophrenia | sex_at_birthMale | 1.233 | 1.075 | 1.413 | 2.68e-03 | 1.87e-02 | 4199 | 2456 | 1228 | 515 |
| Latino/Mixed-American | Full Cohort | Schizophrenia | Income_quantitative | 0.9003 | 0.8689 | 0.9329 | 7.25e-09 | 5.07e-08 | 4199 | 2456 | 1228 | 515 |
| Latino/Mixed-American | Full Cohort | Schizophrenia | Education_levelCollege Undergraduate | 1.022 | 0.8239 | 1.267 | 8.45e-01 | 1.00e+00 | 4199 | 2456 | 1228 | 515 |
| Latino/Mixed-American | Full Cohort | Schizophrenia | Education_levelLess than a high school degree | 1 | 0.7693 | 1.3 | 1.00e+00 | 1.00e+00 | 4199 | 2456 | 1228 | 515 |
| Latino/Mixed-American | Full Cohort | Schizophrenia | Education_levelTwelve or GED | 0.9232 | 0.7184 | 1.186 | 5.32e-01 | 1.00e+00 | 4199 | 2456 | 1228 | 515 |
| Latino/Mixed-American | Full Cohort | Schizophrenia | Schizophrenia | 1.065 | 1.002 | 1.131 | 4.16e-02 | 2.91e-01 | 4199 | 2456 | 1228 | 515 |
| Latino/Mixed-American | Recorded Depression | ADHD | age_at_first_AD | 0.9914 | 0.9861 | 0.9968 | 1.86e-03 | 1.30e-02 | 2480 | 1179 | 915 | 386 |
| Latino/Mixed-American | Recorded Depression | ADHD | sex_at_birthMale | 1.11 | 0.9339 | 1.319 | 2.36e-01 | 1.00e+00 | 2480 | 1179 | 915 | 386 |
| Latino/Mixed-American | Recorded Depression | ADHD | Income_quantitative | 0.9379 | 0.8955 | 0.9823 | 6.55e-03 | 4.58e-02 | 2480 | 1179 | 915 | 386 |
| Latino/Mixed-American | Recorded Depression | ADHD | Education_levelCollege Undergraduate | 0.9626 | 0.7284 | 1.272 | 7.89e-01 | 1.00e+00 | 2480 | 1179 | 915 | 386 |
| Latino/Mixed-American | Recorded Depression | ADHD | Education_levelLess than a high school degree | 0.9949 | 0.7141 | 1.386 | 9.76e-01 | 1.00e+00 | 2480 | 1179 | 915 | 386 |
| Latino/Mixed-American | Recorded Depression | ADHD | Education_levelTwelve or GED | 0.7763 | 0.5655 | 1.066 | 1.17e-01 | 8.21e-01 | 2480 | 1179 | 915 | 386 |
| Latino/Mixed-American | Recorded Depression | ADHD | ADHD | 1.003 | 0.9222 | 1.091 | 9.45e-01 | 1.00e+00 | 2480 | 1179 | 915 | 386 |
| Latino/Mixed-American | Recorded Depression | BMI | age_at_first_AD | 0.9914 | 0.9861 | 0.9968 | 1.90e-03 | 1.33e-02 | 2480 | 1179 | 915 | 386 |
| Latino/Mixed-American | Recorded Depression | BMI | sex_at_birthMale | 1.109 | 0.933 | 1.318 | 2.41e-01 | 1.00e+00 | 2480 | 1179 | 915 | 386 |
| Latino/Mixed-American | Recorded Depression | BMI | Income_quantitative | 0.937 | 0.8947 | 0.9814 | 5.90e-03 | 4.13e-02 | 2480 | 1179 | 915 | 386 |
| Latino/Mixed-American | Recorded Depression | BMI | Education_levelCollege Undergraduate | 0.9664 | 0.7313 | 1.277 | 8.10e-01 | 1.00e+00 | 2480 | 1179 | 915 | 386 |
| Latino/Mixed-American | Recorded Depression | BMI | Education_levelLess than a high school degree | 0.998 | 0.7167 | 1.39 | 9.91e-01 | 1.00e+00 | 2480 | 1179 | 915 | 386 |
| Latino/Mixed-American | Recorded Depression | BMI | Education_levelTwelve or GED | 0.7799 | 0.5681 | 1.071 | 1.24e-01 | 8.68e-01 | 2480 | 1179 | 915 | 386 |
| Latino/Mixed-American | Recorded Depression | BMI | BMI | 0.9728 | 0.8991 | 1.052 | 4.92e-01 | 1.00e+00 | 2480 | 1179 | 915 | 386 |
| Latino/Mixed-American | Recorded Depression | Bipolar Disorder | age_at_first_AD | 0.9913 | 0.9859 | 0.9967 | 1.55e-03 | 1.08e-02 | 2480 | 1179 | 915 | 386 |
| Latino/Mixed-American | Recorded Depression | Bipolar Disorder | sex_at_birthMale | 1.107 | 0.9316 | 1.316 | 2.48e-01 | 1.00e+00 | 2480 | 1179 | 915 | 386 |
| Latino/Mixed-American | Recorded Depression | Bipolar Disorder | Income_quantitative | 0.9396 | 0.8971 | 0.9841 | 8.28e-03 | 5.79e-02 | 2480 | 1179 | 915 | 386 |
| Latino/Mixed-American | Recorded Depression | Bipolar Disorder | Education_levelCollege Undergraduate | 0.9808 | 0.7418 | 1.297 | 8.92e-01 | 1.00e+00 | 2480 | 1179 | 915 | 386 |

| | | | | | | | | | | | | |
|---|---|---|---|---|---|---|---|---|---|---|---|---|
| Latino/Mixed-American | Recorded Depression | Bipolar Disorder | Education_levelLess than a high school degree | 1.012 | 0.7268 | 1.41 | 9.42e-01 | 1.00e+00 | 2480 | 1179 | 915 | 386 |
| Latino/Mixed-American | Recorded Depression | Bipolar Disorder | Education_levelTwelve or GED | 0.7898 | 0.5752 | 1.085 | 1.45e-01 | 1.00e+00 | 2480 | 1179 | 915 | 386 |
| Latino/Mixed-American | Recorded Depression | Bipolar Disorder | Bipolar Disorder | 1.114 | 1.025 | 1.211 | 1.14e-02 | 7.95e-02 | 2480 | 1179 | 915 | 386 |
| Latino/Mixed-American | Recorded Depression | Depression | age_at_first_AD | 0.9915 | 0.9861 | 0.9969 | 2.12e-03 | 1.48e-02 | 2480 | 1179 | 915 | 386 |
| Latino/Mixed-American | Recorded Depression | Depression | sex_at_birthMale | 1.112 | 0.9352 | 1.322 | 2.30e-01 | 1.00e+00 | 2480 | 1179 | 915 | 386 |
| Latino/Mixed-American | Recorded Depression | Depression | Income_quantitative | 0.9386 | 0.8961 | 0.983 | 7.26e-03 | 5.08e-02 | 2480 | 1179 | 915 | 386 |
| Latino/Mixed-American | Recorded Depression | Depression | Education_levelCollege Undergraduate | 0.9684 | 0.7327 | 1.28 | 8.21e-01 | 1.00e+00 | 2480 | 1179 | 915 | 386 |
| Latino/Mixed-American | Recorded Depression | Depression | Education_levelLess than a high school degree | 0.9914 | 0.7118 | 1.381 | 9.59e-01 | 1.00e+00 | 2480 | 1179 | 915 | 386 |
| Latino/Mixed-American | Recorded Depression | Depression | Education_levelTwelve or GED | 0.7812 | 0.569 | 1.073 | 1.27e-01 | 8.87e-01 | 2480 | 1179 | 915 | 386 |
| Latino/Mixed-American | Recorded Depression | Depression | Depression | 1.158 | 1.064 | 1.26 | 6.60e-04 | 4.62e-03 | 2480 | 1179 | 915 | 386 |
| Latino/Mixed-American | Recorded Depression | Insomnia | age_at_first_AD | 0.9914 | 0.9861 | 0.9968 | 1.88e-03 | 1.31e-02 | 2480 | 1179 | 915 | 386 |
| Latino/Mixed-American | Recorded Depression | Insomnia | sex_at_birthMale | 1.112 | 0.9355 | 1.322 | 2.29e-01 | 1.00e+00 | 2480 | 1179 | 915 | 386 |
| Latino/Mixed-American | Recorded Depression | Insomnia | Income_quantitative | 0.9378 | 0.8954 | 0.9821 | 6.41e-03 | 4.49e-02 | 2480 | 1179 | 915 | 386 |
| Latino/Mixed-American | Recorded Depression | Insomnia | Education_levelCollege Undergraduate | 0.9595 | 0.7261 | 1.268 | 7.71e-01 | 1.00e+00 | 2480 | 1179 | 915 | 386 |
| Latino/Mixed-American | Recorded Depression | Insomnia | Education_levelLess than a high school degree | 0.9916 | 0.7121 | 1.381 | 9.60e-01 | 1.00e+00 | 2480 | 1179 | 915 | 386 |
| Latino/Mixed-American | Recorded Depression | Insomnia | Education_levelTwelve or GED | 0.7731 | 0.5632 | 1.061 | 1.11e-01 | 7.79e-01 | 2480 | 1179 | 915 | 386 |
| Latino/Mixed-American | Recorded Depression | Insomnia | Insomnia | 1.051 | 0.9653 | 1.143 | 2.53e-01 | 1.00e+00 | 2480 | 1179 | 915 | 386 |
| Latino/Mixed-American | Recorded Depression | Migraine | age_at_first_AD | 0.9915 | 0.9861 | 0.9969 | 2.00e-03 | 1.40e-02 | 2480 | 1179 | 915 | 386 |
| Latino/Mixed-American | Recorded Depression | Migraine | sex_at_birthMale | 1.111 | 0.9345 | 1.32 | 2.33e-01 | 1.00e+00 | 2480 | 1179 | 915 | 386 |
| Latino/Mixed-American | Recorded Depression | Migraine | Income_quantitative | 0.9359 | 0.8936 | 0.9803 | 5.04e-03 | 3.53e-02 | 2480 | 1179 | 915 | 386 |
| Latino/Mixed-American | Recorded Depression | Migraine | Education_levelCollege Undergraduate | 0.9614 | 0.7274 | 1.271 | 7.82e-01 | 1.00e+00 | 2480 | 1179 | 915 | 386 |
| Latino/Mixed-American | Recorded Depression | Migraine | Education_levelLess than a high school degree | 0.9813 | 0.7044 | 1.367 | 9.11e-01 | 1.00e+00 | 2480 | 1179 | 915 | 386 |
| Latino/Mixed-American | Recorded Depression | Migraine | Education_levelTwelve or GED | 0.7624 | 0.5551 | 1.047 | 9.38e-02 | 6.57e-01 | 2480 | 1179 | 915 | 386 |
| Latino/Mixed-American | Recorded Depression | Migraine | Migraine | 1.107 | 1.024 | 1.197 | 1.02e-02 | 7.11e-02 | 2480 | 1179 | 915 | 386 |
| Latino/Mixed-American | Recorded Depression | Schizophrenia | age_at_first_AD | 0.9914 | 0.986 | 0.9968 | 1.80e-03 | 1.26e-02 | 2480 | 1179 | 915 | 386 |
| Latino/Mixed-American | Recorded Depression | Schizophrenia | sex_at_birthMale | 1.11 | 0.9342 | 1.32 | 2.35e-01 | 1.00e+00 | 2480 | 1179 | 915 | 386 |
| Latino/Mixed-American | Recorded Depression | Schizophrenia | Income_quantitative | 0.9378 | 0.8954 | 0.9821 | 6.42e-03 | 4.50e-02 | 2480 | 1179 | 915 | 386 |
| Latino/Mixed-American | Recorded Depression | Schizophrenia | Education_levelCollege Undergraduate | 0.9751 | 0.7376 | 1.289 | 8.59e-01 | 1.00e+00 | 2480 | 1179 | 915 | 386 |
| Latino/Mixed-American | Recorded Depression | Schizophrenia | Education_levelLess than a high school degree | 1.003 | 0.7204 | 1.397 | 9.84e-01 | 1.00e+00 | 2480 | 1179 | 915 | 386 |
| Latino/Mixed-American | Recorded Depression | Schizophrenia | Education_levelTwelve or GED | 0.7856 | 0.5722 | 1.079 | 1.36e-01 | 9.50e-01 | 2480 | 1179 | 915 | 386 |
| Latino/Mixed-American | Recorded Depression | Schizophrenia | Schizophrenia | 1.068 | 0.9896 | 1.152 | 9.08e-02 | 6.36e-01 | 2480 | 1179 | 915 | 386 |
| Latino/Mixed-American | No Recorded Depression | ADHD | age_at_first_AD | 0.9863 | 0.9788 | 0.994 | 4.74e-04 | 3.32e-03 | 1719 | 1277 | 313 | 129 |
| Latino/Mixed-American | No Recorded Depression | ADHD | sex_at_birthMale | 1.535 | 1.21 | 1.948 | 4.19e-04 | 2.93e-03 | 1719 | 1277 | 313 | 129 |
| Latino/Mixed-American | No Recorded Depression | ADHD | Income_quantitative | 0.917 | 0.8631 | 0.9742 | 5.03e-03 | 3.52e-02 | 1719 | 1277 | 313 | 129 |
| Latino/Mixed-American | No Recorded Depression | ADHD | Education_levelCollege Undergraduate | 1.103 | 0.7672 | 1.587 | 5.96e-01 | 1.00e+00 | 1719 | 1277 | 313 | 129 |
| Latino/Mixed-American | No Recorded Depression | ADHD | Education_levelLess than a high school degree | 0.8889 | 0.5556 | 1.422 | 6.23e-01 | 1.00e+00 | 1719 | 1277 | 313 | 129 |
| Latino/Mixed-American | No Recorded Depression | ADHD | Education_levelTwelve or GED | 1.08 | 0.6926 | 1.684 | 7.34e-01 | 1.00e+00 | 1719 | 1277 | 313 | 129 |
| Latino/Mixed-American | No Recorded Depression | ADHD | ADHD | 1.083 | 0.9643 | 1.217 | 1.78e-01 | 1.00e+00 | 1719 | 1277 | 313 | 129 |
| Latino/Mixed-American | No Recorded Depression | BMI | age_at_first_AD | 0.9861 | 0.9785 | 0.9937 | 3.77e-04 | 2.64e-03 | 1719 | 1277 | 313 | 129 |
| Latino/Mixed-American | No Recorded Depression | BMI | sex_at_birthMale | 1.541 | 1.215 | 1.955 | 3.71e-04 | 2.60e-03 | 1719 | 1277 | 313 | 129 |
| Latino/Mixed-American | No Recorded Depression | BMI | Income_quantitative | 0.9164 | 0.8625 | 0.9737 | 4.80e-03 | 3.36e-02 | 1719 | 1277 | 313 | 129 |
| Latino/Mixed-American | No Recorded Depression | BMI | Education_levelCollege Undergraduate | 1.109 | 0.7708 | 1.595 | 5.78e-01 | 1.00e+00 | 1719 | 1277 | 313 | 129 |
| Latino/Mixed-American | No Recorded Depression | BMI | Education_levelLess than a high school degree | 0.9046 | 0.5657 | 1.447 | 6.75e-01 | 1.00e+00 | 1719 | 1277 | 313 | 129 |
| Latino/Mixed-American | No Recorded Depression | BMI | Education_levelTwelve or GED | 1.095 | 0.7028 | 1.708 | 6.87e-01 | 1.00e+00 | 1719 | 1277 | 313 | 129 |
| Latino/Mixed-American | No Recorded Depression | BMI | BMI | 1.027 | 0.9172 | 1.15 | 6.42e-01 | 1.00e+00 | 1719 | 1277 | 313 | 129 |
| Latino/Mixed-American | No Recorded Depression | Bipolar Disorder | age_at_first_AD | 0.9861 | 0.9785 | 0.9937 | 3.66e-04 | 2.56e-03 | 1719 | 1277 | 313 | 129 |
| Latino/Mixed-American | No Recorded Depression | Bipolar Disorder | sex_at_birthMale | 1.538 | 1.212 | 1.952 | 3.99e-04 | 2.79e-03 | 1719 | 1277 | 313 | 129 |
| Latino/Mixed-American | No Recorded Depression | Bipolar Disorder | Income_quantitative | 0.9153 | 0.8614 | 0.9726 | 4.26e-03 | 2.98e-02 | 1719 | 1277 | 313 | 129 |
| Latino/Mixed-American | No Recorded Depression | Bipolar Disorder | Education_levelCollege Undergraduate | 1.108 | 0.7702 | 1.594 | 5.80e-01 | 1.00e+00 | 1719 | 1277 | 313 | 129 |
| Latino/Mixed-American | No Recorded Depression | Bipolar Disorder | Education_levelLess than a high school degree | 0.9015 | 0.5636 | 1.442 | 6.65e-01 | 1.00e+00 | 1719 | 1277 | 313 | 129 |
| Latino/Mixed-American | No Recorded Depression | Bipolar Disorder | Education_levelTwelve or GED | 1.096 | 0.7029 | 1.708 | 6.86e-01 | 1.00e+00 | 1719 | 1277 | 313 | 129 |
| Latino/Mixed-American | No Recorded Depression | Bipolar Disorder | Bipolar Disorder | 0.9831 | 0.8716 | 1.109 | 7.81e-01 | 1.00e+00 | 1719 | 1277 | 313 | 129 |
| Latino/Mixed-American | No Recorded Depression | Depression | age_at_first_AD | 0.986 | 0.9784 | 0.9936 | 3.34e-04 | 2.34e-03 | 1719 | 1277 | 313 | 129 |
| Latino/Mixed-American | No Recorded Depression | Depression | sex_at_birthMale | 1.545 | 1.217 | 1.961 | 3.50e-04 | 2.45e-03 | 1719 | 1277 | 313 | 129 |
| Latino/Mixed-American | No Recorded Depression | Depression | Income_quantitative | 0.9214 | 0.8671 | 0.9791 | 8.27e-03 | 5.79e-02 | 1719 | 1277 | 313 | 129 |
| Latino/Mixed-American | No Recorded Depression | Depression | Education_levelCollege Undergraduate | 1.111 | 0.7724 | 1.598 | 5.70e-01 | 1.00e+00 | 1719 | 1277 | 313 | 129 |
| Latino/Mixed-American | No Recorded Depression | Depression | Education_levelLess than a high school degree | 0.908 | 0.5678 | 1.452 | 6.87e-01 | 1.00e+00 | 1719 | 1277 | 313 | 129 |

| | | | | | | | | | | | | |
|---|---|---|---|---|---|---|---|---|---|---|---|---|
| Latino/Mixed-American | No Recorded Depression | Depression | Education_levelTwelve or GED | 1.108 | 0.7105 | 1.727 | 6.52e-01 | 1.00e+00 | 1719 | 1277 | 313 | 129 |
| Latino/Mixed-American | No Recorded Depression | Depression | Depression | 1.169 | 1.033 | 1.323 | 1.35e-02 | 9.48e-02 | 1719 | 1277 | 313 | 129 |
| Latino/Mixed-American | No Recorded Depression | Insomnia | age_at_first_AD | 0.9861 | 0.9785 | 0.9937 | 3.69e-04 | 2.58e-03 | 1719 | 1277 | 313 | 129 |
| Latino/Mixed-American | No Recorded Depression | Insomnia | sex_at_birthMale | 1.543 | 1.216 | 1.958 | 3.59e-04 | 2.51e-03 | 1719 | 1277 | 313 | 129 |
| Latino/Mixed-American | No Recorded Depression | Insomnia | Income_quantitative | 0.9163 | 0.8625 | 0.9736 | 4.71e-03 | 3.30e-02 | 1719 | 1277 | 313 | 129 |
| Latino/Mixed-American | No Recorded Depression | Insomnia | Education_levelCollege Undergraduate | 1.107 | 0.7694 | 1.592 | 5.85e-01 | 1.00e+00 | 1719 | 1277 | 313 | 129 |
| Latino/Mixed-American | No Recorded Depression | Insomnia | Education_levelLess than a high school degree | 0.8991 | 0.5622 | 1.438 | 6.57e-01 | 1.00e+00 | 1719 | 1277 | 313 | 129 |
| Latino/Mixed-American | No Recorded Depression | Insomnia | Education_levelTwelve or GED | 1.094 | 0.7018 | 1.705 | 6.92e-01 | 1.00e+00 | 1719 | 1277 | 313 | 129 |
| Latino/Mixed-American | No Recorded Depression | Insomnia | Insomnia | 1.044 | 0.9184 | 1.188 | 5.08e-01 | 1.00e+00 | 1719 | 1277 | 313 | 129 |
| Latino/Mixed-American | No Recorded Depression | Migraine | age_at_first_AD | 0.9859 | 0.9783 | 0.9935 | 3.12e-04 | 2.19e-03 | 1719 | 1277 | 313 | 129 |
| Latino/Mixed-American | No Recorded Depression | Migraine | sex_at_birthMale | 1.541 | 1.214 | 1.955 | 3.72e-04 | 2.61e-03 | 1719 | 1277 | 313 | 129 |
| Latino/Mixed-American | No Recorded Depression | Migraine | Income_quantitative | 0.9151 | 0.8612 | 0.9723 | 4.15e-03 | 2.91e-02 | 1719 | 1277 | 313 | 129 |
| Latino/Mixed-American | No Recorded Depression | Migraine | Education_levelCollege Undergraduate | 1.112 | 0.7731 | 1.599 | 5.67e-01 | 1.00e+00 | 1719 | 1277 | 313 | 129 |
| Latino/Mixed-American | No Recorded Depression | Migraine | Education_levelLess than a high school degree | 0.9078 | 0.5676 | 1.452 | 6.87e-01 | 1.00e+00 | 1719 | 1277 | 313 | 129 |
| Latino/Mixed-American | No Recorded Depression | Migraine | Education_levelTwelve or GED | 1.1 | 0.706 | 1.715 | 6.73e-01 | 1.00e+00 | 1719 | 1277 | 313 | 129 |
| Latino/Mixed-American | No Recorded Depression | Migraine | Migraine | 0.9397 | 0.8377 | 1.054 | 2.89e-01 | 1.00e+00 | 1719 | 1277 | 313 | 129 |
| Latino/Mixed-American | No Recorded Depression | Schizophrenia | age_at_first_AD | 0.9861 | 0.9785 | 0.9937 | 3.48e-04 | 2.44e-03 | 1719 | 1277 | 313 | 129 |
| Latino/Mixed-American | No Recorded Depression | Schizophrenia | sex_at_birthMale | 1.546 | 1.218 | 1.962 | 3.37e-04 | 2.36e-03 | 1719 | 1277 | 313 | 129 |
| Latino/Mixed-American | No Recorded Depression | Schizophrenia | Income_quantitative | 0.9186 | 0.8645 | 0.976 | 6.09e-03 | 4.26e-02 | 1719 | 1277 | 313 | 129 |
| Latino/Mixed-American | No Recorded Depression | Schizophrenia | Education_levelCollege Undergraduate | 1.125 | 0.782 | 1.619 | 5.25e-01 | 1.00e+00 | 1719 | 1277 | 313 | 129 |
| Latino/Mixed-American | No Recorded Depression | Schizophrenia | Education_levelLess than a high school degree | 0.9168 | 0.5732 | 1.466 | 7.17e-01 | 1.00e+00 | 1719 | 1277 | 313 | 129 |
| Latino/Mixed-American | No Recorded Depression | Schizophrenia | Education_levelTwelve or GED | 1.116 | 0.7159 | 1.74 | 6.28e-01 | 1.00e+00 | 1719 | 1277 | 313 | 129 |
| Latino/Mixed-American | No Recorded Depression | Schizophrenia | Schizophrenia | 1.088 | 0.9768 | 1.212 | 1.25e-01 | 8.75e-01 | 1719 | 1277 | 313 | 129 |
| Latino/Mixed-American | Recorded Anxiety | ADHD | age_at_first_AD | 0.9957 | 0.9906 | 1.001 | 1.00e-01 | 7.01e-01 | 2878 | 1462 | 1017 | 399 |
| Latino/Mixed-American | Recorded Anxiety | ADHD | sex_at_birthMale | 1.246 | 1.06 | 1.465 | 7.54e-03 | 5.28e-02 | 2878 | 1462 | 1017 | 399 |
| Latino/Mixed-American | Recorded Anxiety | ADHD | Income_quantitative | 0.9234 | 0.8853 | 0.9632 | 2.15e-04 | 1.50e-03 | 2878 | 1462 | 1017 | 399 |
| Latino/Mixed-American | Recorded Anxiety | ADHD | Education_levelCollege Undergraduate | 1.025 | 0.7965 | 1.32 | 8.45e-01 | 1.00e+00 | 2878 | 1462 | 1017 | 399 |
| Latino/Mixed-American | Recorded Anxiety | ADHD | Education_levelLess than a high school degree | 1.061 | 0.7769 | 1.449 | 7.09e-01 | 1.00e+00 | 2878 | 1462 | 1017 | 399 |
| Latino/Mixed-American | Recorded Anxiety | ADHD | Education_levelTwelve or GED | 0.8407 | 0.627 | 1.127 | 2.46e-01 | 1.00e+00 | 2878 | 1462 | 1017 | 399 |
| Latino/Mixed-American | Recorded Anxiety | ADHD | ADHD | 1.062 | 0.9832 | 1.148 | 1.25e-01 | 8.78e-01 | 2878 | 1462 | 1017 | 399 |
| Latino/Mixed-American | Recorded Anxiety | BMI | age_at_first_AD | 0.9956 | 0.9905 | 1.001 | 9.28e-02 | 6.50e-01 | 2878 | 1462 | 1017 | 399 |
| Latino/Mixed-American | Recorded Anxiety | BMI | sex_at_birthMale | 1.249 | 1.063 | 1.468 | 6.94e-03 | 4.86e-02 | 2878 | 1462 | 1017 | 399 |
| Latino/Mixed-American | Recorded Anxiety | BMI | Income_quantitative | 0.9223 | 0.8842 | 0.9621 | 1.74e-04 | 1.22e-03 | 2878 | 1462 | 1017 | 399 |
| Latino/Mixed-American | Recorded Anxiety | BMI | Education_levelCollege Undergraduate | 1.032 | 0.8014 | 1.329 | 8.08e-01 | 1.00e+00 | 2878 | 1462 | 1017 | 399 |
| Latino/Mixed-American | Recorded Anxiety | BMI | Education_levelLess than a high school degree | 1.077 | 0.7892 | 1.471 | 6.39e-01 | 1.00e+00 | 2878 | 1462 | 1017 | 399 |
| Latino/Mixed-American | Recorded Anxiety | BMI | Education_levelTwelve or GED | 0.8493 | 0.6334 | 1.139 | 2.75e-01 | 1.00e+00 | 2878 | 1462 | 1017 | 399 |
| Latino/Mixed-American | Recorded Anxiety | BMI | BMI | 0.9951 | 0.9253 | 1.07 | 8.95e-01 | 1.00e+00 | 2878 | 1462 | 1017 | 399 |
| Latino/Mixed-American | Recorded Anxiety | Bipolar Disorder | age_at_first_AD | 0.9955 | 0.9904 | 1.001 | 8.35e-02 | 5.84e-01 | 2878 | 1462 | 1017 | 399 |
| Latino/Mixed-American | Recorded Anxiety | Bipolar Disorder | sex_at_birthMale | 1.25 | 1.063 | 1.469 | 6.79e-03 | 4.76e-02 | 2878 | 1462 | 1017 | 399 |
| Latino/Mixed-American | Recorded Anxiety | Bipolar Disorder | Income_quantitative | 0.9242 | 0.886 | 0.964 | 2.51e-04 | 1.76e-03 | 2878 | 1462 | 1017 | 399 |
| Latino/Mixed-American | Recorded Anxiety | Bipolar Disorder | Education_levelCollege Undergraduate | 1.038 | 0.8062 | 1.337 | 7.72e-01 | 1.00e+00 | 2878 | 1462 | 1017 | 399 |
| Latino/Mixed-American | Recorded Anxiety | Bipolar Disorder | Education_levelLess than a high school degree | 1.087 | 0.7962 | 1.484 | 5.99e-01 | 1.00e+00 | 2878 | 1462 | 1017 | 399 |
| Latino/Mixed-American | Recorded Anxiety | Bipolar Disorder | Education_levelTwelve or GED | 0.8549 | 0.6376 | 1.146 | 2.95e-01 | 1.00e+00 | 2878 | 1462 | 1017 | 399 |
| Latino/Mixed-American | Recorded Anxiety | Bipolar Disorder | Bipolar Disorder | 1.067 | 0.9865 | 1.155 | 1.05e-01 | 7.33e-01 | 2878 | 1462 | 1017 | 399 |
| Latino/Mixed-American | Recorded Anxiety | Depression | age_at_first_AD | 0.9955 | 0.9904 | 1.001 | 8.24e-02 | 5.77e-01 | 2878 | 1462 | 1017 | 399 |
| Latino/Mixed-American | Recorded Anxiety | Depression | sex_at_birthMale | 1.256 | 1.068 | 1.476 | 5.82e-03 | 4.07e-02 | 2878 | 1462 | 1017 | 399 |
| Latino/Mixed-American | Recorded Anxiety | Depression | Income_quantitative | 0.9252 | 0.887 | 0.9652 | 3.11e-04 | 2.18e-03 | 2878 | 1462 | 1017 | 399 |
| Latino/Mixed-American | Recorded Anxiety | Depression | Education_levelCollege Undergraduate | 1.037 | 0.8049 | 1.335 | 7.81e-01 | 1.00e+00 | 2878 | 1462 | 1017 | 399 |
| Latino/Mixed-American | Recorded Anxiety | Depression | Education_levelLess than a high school degree | 1.077 | 0.7884 | 1.47 | 6.43e-01 | 1.00e+00 | 2878 | 1462 | 1017 | 399 |
| Latino/Mixed-American | Recorded Anxiety | Depression | Education_levelTwelve or GED | 0.8512 | 0.6348 | 1.142 | 2.82e-01 | 1.00e+00 | 2878 | 1462 | 1017 | 399 |
| Latino/Mixed-American | Recorded Anxiety | Depression | Depression | 1.175 | 1.085 | 1.272 | 6.82e-05 | 4.78e-04 | 2878 | 1462 | 1017 | 399 |
| Latino/Mixed-American | Recorded Anxiety | Insomnia | age_at_first_AD | 0.9956 | 0.9905 | 1.001 | 9.23e-02 | 6.46e-01 | 2878 | 1462 | 1017 | 399 |
| Latino/Mixed-American | Recorded Anxiety | Insomnia | sex_at_birthMale | 1.257 | 1.069 | 1.477 | 5.58e-03 | 3.90e-02 | 2878 | 1462 | 1017 | 399 |
| Latino/Mixed-American | Recorded Anxiety | Insomnia | Income_quantitative | 0.9228 | 0.8847 | 0.9625 | 1.88e-04 | 1.31e-03 | 2878 | 1462 | 1017 | 399 |
| Latino/Mixed-American | Recorded Anxiety | Insomnia | Education_levelCollege Undergraduate | 1.023 | 0.7947 | 1.317 | 8.59e-01 | 1.00e+00 | 2878 | 1462 | 1017 | 399 |
| Latino/Mixed-American | Recorded Anxiety | Insomnia | Education_levelLess than a high school degree | 1.068 | 0.7824 | 1.458 | 6.78e-01 | 1.00e+00 | 2878 | 1462 | 1017 | 399 |
| Latino/Mixed-American | Recorded Anxiety | Insomnia | Education_levelTwelve or GED | 0.8404 | 0.6268 | 1.127 | 2.45e-01 | 1.00e+00 | 2878 | 1462 | 1017 | 399 |

| | | | | | | | | | | | | |
|---|---|---|---|---|---|---|---|---|---|---|---|---|
| Latino/Mixed-American | Recorded Anxiety | Insomnia | Insomnia | 1.088 | 1.004 | 1.179 | 4.06e-02 | 2.84e-01 | 2878 | 1462 | 1017 | 399 |
| Latino/Mixed-American | Recorded Anxiety | Migraine | age_at_first_AD | 0.9956 | 0.9905 | 1.001 | 9.33e-02 | 6.53e-01 | 2878 | 1462 | 1017 | 399 |
| Latino/Mixed-American | Recorded Anxiety | Migraine | sex_at_birthMale | 1.249 | 1.063 | 1.468 | 6.93e-03 | 4.85e-02 | 2878 | 1462 | 1017 | 399 |
| Latino/Mixed-American | Recorded Anxiety | Migraine | Income_quantitative | 0.9219 | 0.8839 | 0.9617 | 1.60e-04 | 1.12e-03 | 2878 | 1462 | 1017 | 399 |
| Latino/Mixed-American | Recorded Anxiety | Migraine | Education_levelCollege Undergraduate | 1.032 | 0.8015 | 1.328 | 8.08e-01 | 1.00e+00 | 2878 | 1462 | 1017 | 399 |
| Latino/Mixed-American | Recorded Anxiety | Migraine | Education_levelLess than a high school degree | 1.073 | 0.7859 | 1.465 | 6.57e-01 | 1.00e+00 | 2878 | 1462 | 1017 | 399 |
| Latino/Mixed-American | Recorded Anxiety | Migraine | Education_levelTwelve or GED | 0.8457 | 0.6308 | 1.134 | 2.63e-01 | 1.00e+00 | 2878 | 1462 | 1017 | 399 |
| Latino/Mixed-American | Recorded Anxiety | Migraine | Migraine | 1.029 | 0.9567 | 1.106 | 4.44e-01 | 1.00e+00 | 2878 | 1462 | 1017 | 399 |
| Latino/Mixed-American | Recorded Anxiety | Schizophrenia | age_at_first_AD | 0.9955 | 0.9904 | 1.001 | 8.50e-02 | 5.95e-01 | 2878 | 1462 | 1017 | 399 |
| Latino/Mixed-American | Recorded Anxiety | Schizophrenia | sex_at_birthMale | 1.254 | 1.067 | 1.474 | 6.10e-03 | 4.27e-02 | 2878 | 1462 | 1017 | 399 |
| Latino/Mixed-American | Recorded Anxiety | Schizophrenia | Income_quantitative | 0.9239 | 0.8858 | 0.9638 | 2.38e-04 | 1.67e-03 | 2878 | 1462 | 1017 | 399 |
| Latino/Mixed-American | Recorded Anxiety | Schizophrenia | Education_levelCollege Undergraduate | 1.045 | 0.8112 | 1.345 | 7.35e-01 | 1.00e+00 | 2878 | 1462 | 1017 | 399 |
| Latino/Mixed-American | Recorded Anxiety | Schizophrenia | Education_levelLess than a high school degree | 1.087 | 0.7961 | 1.484 | 5.99e-01 | 1.00e+00 | 2878 | 1462 | 1017 | 399 |
| Latino/Mixed-American | Recorded Anxiety | Schizophrenia | Education_levelTwelve or GED | 0.8602 | 0.6414 | 1.154 | 3.14e-01 | 1.00e+00 | 2878 | 1462 | 1017 | 399 |
| Latino/Mixed-American | Recorded Anxiety | Schizophrenia | Schizophrenia | 1.075 | 1.002 | 1.154 | 4.47e-02 | 3.13e-01 | 2878 | 1462 | 1017 | 399 |

**Supplementary Table 13. Multivariable logistic regression results for PGS × drug-class interactions on antidepressant switching versus continuation in All of Us.** Each model estimates the association between one PGS trait and switching (versus continuation), together with its interaction with antidepressant drug class, fitted separately within each combination of genetically inferred ancestry (European, African, Latino/Mixed-American) and clinical group (Full Cohort; subgroups defined by recorded depression status, recorded anxiety, and CIDI-SF–derived MDD classification). The CIDI-SF–derived MDD subgroups (CIDI-MDD, CIDI-noMDD) were analysed within European-ancestry participants only. The outcome is modelled as switch versus continue ~ PGS × DrugClass + covariates, with SSRI as the reference drug class. Because a participant can contribute switching/continuation observations for multiple drug classes, standard errors are cluster-robust, clustered on individual. Within each model, one PGS exposure is modelled alongside a common covariate set (age at first antidepressant, sex at birth, income, and educational attainment). All PGS were residualised on the top 16 global genetic ancestry principal components and the residuals standardised to mean 0, SD 1; each odds ratio therefore reflects the change in odds per one-standard-deviation increase in the PC-adjusted PGS. Because ancestry PCs were regressed out at this stage, they are not included in the model covariate set. For each PGS × clinical group model, three types of estimate are reported (type column): Within-class slope = the PGS–switching association within each drug class (the simple slope, i.e. the PGS effect on switching for that class); Interaction (vs SSRI) = the difference in the PGS slope for each class relative to the SSRI reference class; and Joint_Interaction_Test = an omnibus test of whether the PGS slope differs across drug classes (all interaction terms jointly zero), for which the statistic column reports the test statistic and no odds ratio is defined. Columns: Population = inferred genetic ancestry; ClinicalGroup = cohort or clinical subgroup; PGS = the PGS trait for that model; type = estimate type (within-class slope, interaction vs SSRI, or joint interaction test); DrugClass = the antidepressant class for that term; estimate = model coefficient on the log-odds scale; std.error = cluster-robust standard error; statistic = z-statistic (or joint-test statistic for Joint_Interaction_Test rows); odds_ratio = exponentiated coefficient; lower_95/upper_95 = 95% confidence interval bounds for the odds ratio; p_value = nominal (uncorrected) p-value; Bonf_P = Bonferroni-corrected p-value (capped at 1.00); N_Total = number of person–drug-class observations in the model; N_Persons = number of unique individuals contributing observations. Odds ratios >1 (within-class slopes) indicate higher odds of switching per one-SD increase in PGS; for interaction terms, odds ratios >1 indicate a steeper PGS–switching slope relative to SSRI. Bonferroni correction was applied within each specification (ancestry × clinical group) for the max 7 PGS traits tested.

| Population | ClinicalGroup | PGS | type | DrugClass | estimate | std.error | statistic | odds_ratio | lower_95 | upper_95 | p_value | Bonf_P | N_Total | N_Persons |
|---|---|---|---|---|---|---|---|---|---|---|---|---|---|---|
| European | Full Cohort | ADHD | Within-class slope | SSRI | 0.081 | 0.015 | 5.381 | 1.084 | 1.053 | 1.117 | 7.40e-08 | 5.18e-07 | 43921 | 34993 |
| European | Full Cohort | ADHD | Within-class slope | SNRI | 0.043 | 0.024 | 1.807 | 1.044 | 0.996 | 1.094 | 7.08e-02 | 4.95e-01 | 43921 | 34993 |
| European | Full Cohort | ADHD | Within-class slope | TCA | 0.030 | 0.031 | 0.947 | 1.030 | 0.969 | 1.095 | 3.43e-01 | 1.00e+00 | 43921 | 34993 |
| European | Full Cohort | ADHD | Within-class slope | NaSSA | 0.057 | 0.062 | 0.920 | 1.059 | 0.937 | 1.196 | 3.58e-01 | 1.00e+00 | 43921 | 34993 |
| European | Full Cohort | ADHD | Within-class slope | SARI | 0.105 | 0.034 | 3.099 | 1.110 | 1.039 | 1.186 | 1.94e-03 | 1.36e-02 | 43921 | 34993 |
| European | Full Cohort | ADHD | Within-class slope | NDRI | 0.010 | 0.029 | 0.349 | 1.010 | 0.955 | 1.068 | 7.27e-01 | 1.00e+00 | 43921 | 34993 |
| European | Full Cohort | ADHD | Interaction (vs SSRI) | SNRI | -0.038 | 0.027 | -1.424 | 0.963 | 0.914 | 1.014 | 1.54e-01 | 1.00e+00 | 43921 | 34993 |
| European | Full Cohort | ADHD | Interaction (vs SSRI) | TCA | -0.051 | 0.033 | -1.540 | 0.950 | 0.890 | 1.014 | 1.23e-01 | 8.64e-01 | 43921 | 34993 |
| European | Full Cohort | ADHD | Interaction (vs SSRI) | NaSSA | -0.024 | 0.063 | -0.377 | 0.976 | 0.862 | 1.105 | 7.06e-01 | 1.00e+00 | 43921 | 34993 |
| European | Full Cohort | ADHD | Interaction (vs SSRI) | SARI | 0.024 | 0.036 | 0.661 | 1.024 | 0.955 | 1.098 | 5.09e-01 | 1.00e+00 | 43921 | 34993 |
| European | Full Cohort | ADHD | Interaction (vs SSRI) | NDRI | -0.071 | 0.030 | -2.332 | 0.931 | 0.877 | 0.989 | 1.97e-02 | 1.38e-01 | 43921 | 34993 |
| European | Full Cohort | ADHD | Joint_Interaction_Test | | | | 9.132 | | | | 1.04e-01 | 7.28e-01 | 43921 | 34993 |
| European | Full Cohort | Migraine | Within-class slope | SSRI | 0.056 | 0.015 | 3.811 | 1.057 | 1.027 | 1.088 | 1.38e-04 | 9.69e-04 | 43921 | 34993 |
| European | Full Cohort | Migraine | Within-class slope | SNRI | 0.028 | 0.023 | 1.229 | 1.029 | 0.983 | 1.077 | 2.19e-01 | 1.00e+00 | 43921 | 34993 |
| European | Full Cohort | Migraine | Within-class slope | TCA | 0.018 | 0.031 | 0.596 | 1.018 | 0.959 | 1.082 | 5.51e-01 | 1.00e+00 | 43921 | 34993 |
| European | Full Cohort | Migraine | Within-class slope | NaSSA | 0.049 | 0.062 | 0.795 | 1.050 | 0.931 | 1.185 | 4.27e-01 | 1.00e+00 | 43921 | 34993 |
| European | Full Cohort | Migraine | Within-class slope | SARI | 0.007 | 0.033 | 0.221 | 1.007 | 0.945 | 1.074 | 8.25e-01 | 1.00e+00 | 43921 | 34993 |
| European | Full Cohort | Migraine | Within-class slope | NDRI | 0.095 | 0.028 | 3.367 | 1.100 | 1.041 | 1.163 | 7.60e-04 | 5.32e-03 | 43921 | 34993 |
| European | Full Cohort | Migraine | Interaction (vs SSRI) | SNRI | -0.027 | 0.026 | -1.058 | 0.973 | 0.925 | 1.024 | 2.90e-01 | 1.00e+00 | 43921 | 34993 |
| European | Full Cohort | Migraine | Interaction (vs SSRI) | TCA | -0.038 | 0.033 | -1.136 | 0.963 | 0.903 | 1.028 | 2.56e-01 | 1.00e+00 | 43921 | 34993 |
| European | Full Cohort | Migraine | Interaction (vs SSRI) | NaSSA | -0.007 | 0.063 | -0.109 | 0.993 | 0.878 | 1.123 | 9.13e-01 | 1.00e+00 | 43921 | 34993 |
| European | Full Cohort | Migraine | Interaction (vs SSRI) | SARI | -0.049 | 0.035 | -1.401 | 0.953 | 0.890 | 1.020 | 1.61e-01 | 1.00e+00 | 43921 | 34993 |
| European | Full Cohort | Migraine | Interaction (vs SSRI) | NDRI | 0.040 | 0.030 | 1.309 | 1.040 | 0.981 | 1.104 | 1.91e-01 | 1.00e+00 | 43921 | 34993 |
| European | Full Cohort | Migraine | Joint_Interaction_Test | | | | 6.721 | | | | 2.42e-01 | 1.00e+00 | 43921 | 34993 |
| European | Full Cohort | Insomnia | Within-class slope | SSRI | 0.111 | 0.016 | 7.130 | 1.117 | 1.084 | 1.152 | 1.00e-12 | 7.01e-12 | 43921 | 34993 |
| European | Full Cohort | Insomnia | Within-class slope | SNRI | 0.076 | 0.025 | 3.088 | 1.079 | 1.028 | 1.133 | 2.02e-03 | 1.41e-02 | 43921 | 34993 |
| European | Full Cohort | Insomnia | Within-class slope | TCA | 0.129 | 0.032 | 4.085 | 1.138 | 1.069 | 1.211 | 4.41e-05 | 3.09e-04 | 43921 | 34993 |
| European | Full Cohort | Insomnia | Within-class slope | NaSSA | 0.042 | 0.064 | 0.659 | 1.043 | 0.920 | 1.182 | 5.10e-01 | 1.00e+00 | 43921 | 34993 |
| European | Full Cohort | Insomnia | Within-class slope | SARI | 0.082 | 0.034 | 2.409 | 1.086 | 1.015 | 1.161 | 1.60e-02 | 1.12e-01 | 43921 | 34993 |
| European | Full Cohort | Insomnia | Within-class slope | NDRI | 0.074 | 0.029 | 2.542 | 1.077 | 1.017 | 1.140 | 1.10e-02 | 7.72e-02 | 43921 | 34993 |
| European | Full Cohort | Insomnia | Interaction (vs SSRI) | SNRI | -0.035 | 0.027 | -1.261 | 0.966 | 0.915 | 1.019 | 2.07e-01 | 1.00e+00 | 43921 | 34993 |
| European | Full Cohort | Insomnia | Interaction (vs SSRI) | TCA | 0.018 | 0.034 | 0.537 | 1.018 | 0.953 | 1.089 | 5.91e-01 | 1.00e+00 | 43921 | 34993 |
| European | Full Cohort | Insomnia | Interaction (vs SSRI) | NaSSA | -0.069 | 0.065 | -1.061 | 0.933 | 0.822 | 1.060 | 2.89e-01 | 1.00e+00 | 43921 | 34993 |
| European | Full Cohort | Insomnia | Interaction (vs SSRI) | SARI | -0.028 | 0.036 | -0.786 | 0.972 | 0.905 | 1.043 | 4.32e-01 | 1.00e+00 | 43921 | 34993 |
| European | Full Cohort | Insomnia | Interaction (vs SSRI) | NDRI | -0.037 | 0.031 | -1.174 | 0.964 | 0.906 | 1.025 | 2.41e-01 | 1.00e+00 | 43921 | 34993 |
| European | Full Cohort | Insomnia | Joint_Interaction_Test | | | | 4.447 | | | | 4.87e-01 | 1.00e+00 | 43921 | 34993 |
| European | Full Cohort | Depression | Within-class slope | SSRI | 0.178 | 0.016 | 11.419 | 1.195 | 1.159 | 1.232 | <2.2e-16 | <2.2e-16 | 43921 | 34993 |
| European | Full Cohort | Depression | Within-class slope | SNRI | 0.191 | 0.025 | 7.748 | 1.211 | 1.153 | 1.271 | 9.34e-15 | 6.54e-14 | 43921 | 34993 |

| | | | | | | | | | | | | | | |
|---|---|---|---|---|---|---|---|---|---|---|---|---|---|---|
| European | Full Cohort | Depression | Within-class slope | TCA | 0.193 | 0.032 | 6.070 | 1.213 | 1.139 | 1.290 | 1.28e-09 | 8.96e-09 | 43921 | 34993 |
| European | Full Cohort | Depression | Within-class slope | NaSSA | 0.137 | 0.063 | 2.173 | 1.147 | 1.014 | 1.298 | 2.98e-02 | 2.08e-01 | 43921 | 34993 |
| European | Full Cohort | Depression | Within-class slope | SARI | 0.229 | 0.034 | 6.717 | 1.258 | 1.176 | 1.345 | 1.85e-11 | 1.30e-10 | 43921 | 34993 |
| European | Full Cohort | Depression | Within-class slope | NDRI | 0.203 | 0.028 | 7.144 | 1.225 | 1.159 | 1.295 | 9.09e-13 | 6.36e-12 | 43921 | 34993 |
| European | Full Cohort | Depression | Interaction (vs SSRI) | SNRI | 0.013 | 0.028 | 0.485 | 1.013 | 0.960 | 1.070 | 6.27e-01 | 1.00e+00 | 43921 | 34993 |
| European | Full Cohort | Depression | Interaction (vs SSRI) | TCA | 0.015 | 0.034 | 0.438 | 1.015 | 0.949 | 1.085 | 6.62e-01 | 1.00e+00 | 43921 | 34993 |
| European | Full Cohort | Depression | Interaction (vs SSRI) | NaSSA | -0.041 | 0.064 | -0.634 | 0.960 | 0.846 | 1.089 | 5.26e-01 | 1.00e+00 | 43921 | 34993 |
| European | Full Cohort | Depression | Interaction (vs SSRI) | SARI | 0.051 | 0.036 | 1.419 | 1.053 | 0.981 | 1.130 | 1.56e-01 | 1.00e+00 | 43921 | 34993 |
| European | Full Cohort | Depression | Interaction (vs SSRI) | NDRI | 0.025 | 0.031 | 0.824 | 1.026 | 0.966 | 1.089 | 4.10e-01 | 1.00e+00 | 43921 | 34993 |
| European | Full Cohort | Depression | Joint_Interaction_Test | | | | 3.105 | | | | 6.84e-01 | 1.00e+00 | 43921 | 34993 |
| European | Full Cohort | Schizophrenia | Within-class slope | SSRI | 0.012 | 0.015 | 0.789 | 1.012 | 0.983 | 1.042 | 4.30e-01 | 1.00e+00 | 43921 | 34993 |
| European | Full Cohort | Schizophrenia | Within-class slope | SNRI | 0.036 | 0.024 | 1.481 | 1.037 | 0.988 | 1.087 | 1.39e-01 | 9.71e-01 | 43921 | 34993 |
| European | Full Cohort | Schizophrenia | Within-class slope | TCA | -0.009 | 0.031 | -0.285 | 0.991 | 0.932 | 1.054 | 7.76e-01 | 1.00e+00 | 43921 | 34993 |
| European | Full Cohort | Schizophrenia | Within-class slope | NaSSA | 0.013 | 0.063 | 0.200 | 1.013 | 0.895 | 1.146 | 8.42e-01 | 1.00e+00 | 43921 | 34993 |
| European | Full Cohort | Schizophrenia | Within-class slope | SARI | 0.010 | 0.033 | 0.320 | 1.011 | 0.948 | 1.077 | 7.49e-01 | 1.00e+00 | 43921 | 34993 |
| European | Full Cohort | Schizophrenia | Within-class slope | NDRI | 0.027 | 0.028 | 0.956 | 1.027 | 0.972 | 1.085 | 3.39e-01 | 1.00e+00 | 43921 | 34993 |
| European | Full Cohort | Schizophrenia | Interaction (vs SSRI) | SNRI | 0.024 | 0.027 | 0.895 | 1.024 | 0.972 | 1.080 | 3.71e-01 | 1.00e+00 | 43921 | 34993 |
| European | Full Cohort | Schizophrenia | Interaction (vs SSRI) | TCA | -0.021 | 0.034 | -0.617 | 0.980 | 0.917 | 1.046 | 5.37e-01 | 1.00e+00 | 43921 | 34993 |
| European | Full Cohort | Schizophrenia | Interaction (vs SSRI) | NaSSA | 0.001 | 0.064 | 0.013 | 1.001 | 0.883 | 1.135 | 9.89e-01 | 1.00e+00 | 43921 | 34993 |
| European | Full Cohort | Schizophrenia | Interaction (vs SSRI) | SARI | -0.001 | 0.035 | -0.036 | 0.999 | 0.933 | 1.069 | 9.71e-01 | 1.00e+00 | 43921 | 34993 |
| European | Full Cohort | Schizophrenia | Interaction (vs SSRI) | NDRI | 0.015 | 0.030 | 0.501 | 1.015 | 0.957 | 1.077 | 6.16e-01 | 1.00e+00 | 43921 | 34993 |
| European | Full Cohort | Schizophrenia | Joint_Interaction_Test | | | | 1.702 | | | | 8.89e-01 | 1.00e+00 | 43921 | 34993 |
| European | Full Cohort | Bipolar Disorder | Within-class slope | SSRI | 0.037 | 0.015 | 2.411 | 1.038 | 1.007 | 1.070 | 1.59e-02 | 1.11e-01 | 43921 | 34993 |
| European | Full Cohort | Bipolar Disorder | Within-class slope | SNRI | 0.036 | 0.025 | 1.445 | 1.036 | 0.987 | 1.088 | 1.48e-01 | 1.00e+00 | 43921 | 34993 |
| European | Full Cohort | Bipolar Disorder | Within-class slope | TCA | 0.048 | 0.032 | 1.497 | 1.050 | 0.985 | 1.118 | 1.34e-01 | 9.41e-01 | 43921 | 34993 |
| European | Full Cohort | Bipolar Disorder | Within-class slope | NaSSA | -0.036 | 0.065 | -0.553 | 0.965 | 0.849 | 1.096 | 5.80e-01 | 1.00e+00 | 43921 | 34993 |
| European | Full Cohort | Bipolar Disorder | Within-class slope | SARI | 0.088 | 0.034 | 2.558 | 1.092 | 1.021 | 1.168 | 1.05e-02 | 7.37e-02 | 43921 | 34993 |
| European | Full Cohort | Bipolar Disorder | Within-class slope | NDRI | 0.035 | 0.029 | 1.195 | 1.035 | 0.978 | 1.096 | 2.32e-01 | 1.00e+00 | 43921 | 34993 |
| European | Full Cohort | Bipolar Disorder | Interaction (vs SSRI) | SNRI | -0.001 | 0.028 | -0.049 | 0.999 | 0.946 | 1.054 | 9.61e-01 | 1.00e+00 | 43921 | 34993 |
| European | Full Cohort | Bipolar Disorder | Interaction (vs SSRI) | TCA | 0.011 | 0.035 | 0.324 | 1.011 | 0.945 | 1.082 | 7.46e-01 | 1.00e+00 | 43921 | 34993 |
| European | Full Cohort | Bipolar Disorder | Interaction (vs SSRI) | NaSSA | -0.073 | 0.066 | -1.107 | 0.929 | 0.816 | 1.058 | 2.68e-01 | 1.00e+00 | 43921 | 34993 |
| European | Full Cohort | Bipolar Disorder | Interaction (vs SSRI) | SARI | 0.051 | 0.037 | 1.394 | 1.052 | 0.980 | 1.130 | 1.63e-01 | 1.00e+00 | 43921 | 34993 |
| European | Full Cohort | Bipolar Disorder | Interaction (vs SSRI) | NDRI | -0.002 | 0.031 | -0.078 | 0.998 | 0.938 | 1.061 | 9.38e-01 | 1.00e+00 | 43921 | 34993 |
| European | Full Cohort | Bipolar Disorder | Joint_Interaction_Test | | | | 3.686 | | | | 5.95e-01 | 1.00e+00 | 43921 | 34993 |
| European | Full Cohort | BMI | Within-class slope | SSRI | 0.054 | 0.015 | 3.679 | 1.056 | 1.026 | 1.087 | 2.34e-04 | 1.64e-03 | 43921 | 34993 |
| European | Full Cohort | BMI | Within-class slope | SNRI | 0.020 | 0.024 | 0.848 | 1.020 | 0.974 | 1.068 | 3.97e-01 | 1.00e+00 | 43921 | 34993 |
| European | Full Cohort | BMI | Within-class slope | TCA | 0.063 | 0.031 | 2.029 | 1.065 | 1.002 | 1.133 | 4.24e-02 | 2.97e-01 | 43921 | 34993 |
| European | Full Cohort | BMI | Within-class slope | NaSSA | 0.042 | 0.062 | 0.680 | 1.043 | 0.924 | 1.177 | 4.96e-01 | 1.00e+00 | 43921 | 34993 |
| European | Full Cohort | BMI | Within-class slope | SARI | 0.052 | 0.033 | 1.594 | 1.053 | 0.988 | 1.123 | 1.11e-01 | 7.77e-01 | 43921 | 34993 |
| European | Full Cohort | BMI | Within-class slope | NDRI | -0.032 | 0.028 | -1.123 | 0.969 | 0.917 | 1.024 | 2.62e-01 | 1.00e+00 | 43921 | 34993 |
| European | Full Cohort | BMI | Interaction (vs SSRI) | SNRI | -0.034 | 0.026 | -1.310 | 0.966 | 0.918 | 1.017 | 1.90e-01 | 1.00e+00 | 43921 | 34993 |
| European | Full Cohort | BMI | Interaction (vs SSRI) | TCA | 0.009 | 0.033 | 0.271 | 1.009 | 0.945 | 1.077 | 7.86e-01 | 1.00e+00 | 43921 | 34993 |
| European | Full Cohort | BMI | Interaction (vs SSRI) | NaSSA | -0.012 | 0.063 | -0.194 | 0.988 | 0.873 | 1.118 | 8.46e-01 | 1.00e+00 | 43921 | 34993 |
| European | Full Cohort | BMI | Interaction (vs SSRI) | SARI | -0.002 | 0.034 | -0.068 | 0.998 | 0.933 | 1.067 | 9.46e-01 | 1.00e+00 | 43921 | 34993 |
| European | Full Cohort | BMI | Interaction (vs SSRI) | NDRI | -0.086 | 0.030 | -2.854 | 0.918 | 0.865 | 0.973 | 4.32e-03 | 3.02e-02 | 43921 | 34993 |
| European | Full Cohort | BMI | Joint_Interaction_Test | | | | 9.639 | | | | 8.61e-02 | 6.03e-01 | 43921 | 34993 |
| European | Recorded Depression | ADHD | Within-class slope | SSRI | 0.071 | 0.020 | 3.617 | 1.073 | 1.033 | 1.115 | 2.98e-04 | 2.09e-03 | 23614 | 17516 |
| European | Recorded Depression | ADHD | Within-class slope | SNRI | 0.008 | 0.032 | 0.247 | 1.008 | 0.946 | 1.074 | 8.05e-01 | 1.00e+00 | 23614 | 17516 |
| European | Recorded Depression | ADHD | Within-class slope | TCA | -0.052 | 0.052 | -0.990 | 0.950 | 0.858 | 1.052 | 3.22e-01 | 1.00e+00 | 23614 | 17516 |
| European | Recorded Depression | ADHD | Within-class slope | NaSSA | 0.019 | 0.090 | 0.211 | 1.019 | 0.854 | 1.216 | 8.33e-01 | 1.00e+00 | 23614 | 17516 |
| European | Recorded Depression | ADHD | Within-class slope | SARI | 0.024 | 0.057 | 0.423 | 1.024 | 0.916 | 1.145 | 6.72e-01 | 1.00e+00 | 23614 | 17516 |
| European | Recorded Depression | ADHD | Within-class slope | NDRI | -0.004 | 0.037 | -0.101 | 0.996 | 0.927 | 1.071 | 9.19e-01 | 1.00e+00 | 23614 | 17516 |
| European | Recorded Depression | ADHD | Interaction (vs SSRI) | SNRI | -0.063 | 0.036 | -1.760 | 0.939 | 0.876 | 1.007 | 7.85e-02 | 5.49e-01 | 23614 | 17516 |
| European | Recorded Depression | ADHD | Interaction (vs SSRI) | TCA | -0.122 | 0.054 | -2.261 | 0.885 | 0.796 | 0.984 | 2.38e-02 | 1.66e-01 | 23614 | 17516 |

| | | | | | | | | | | | | | | |
|---|---|---|---|---|---|---|---|---|---|---|---|---|---|---|
| European | Recorded Depression | ADHD | Interaction (vs SSRI) | NaSSA | -0.052 | 0.092 | -0.564 | 0.950 | 0.794 | 1.136 | 5.73e-01 | 1.00e+00 | 23614 | 17516 |
| European | Recorded Depression | ADHD | Interaction (vs SSRI) | SARI | -0.047 | 0.058 | -0.799 | 0.954 | 0.851 | 1.070 | 4.24e-01 | 1.00e+00 | 23614 | 17516 |
| European | Recorded Depression | ADHD | Interaction (vs SSRI) | NDRI | -0.074 | 0.039 | -1.891 | 0.928 | 0.859 | 1.003 | 5.86e-02 | 4.10e-01 | 23614 | 17516 |
| European | Recorded Depression | ADHD | Joint_Interaction_Test | | | | 8.859 | | | | 1.15e-01 | 8.04e-01 | 23614 | 17516 |
| European | Recorded Depression | Migraine | Within-class slope | SSRI | 0.067 | 0.019 | 3.509 | 1.069 | 1.030 | 1.110 | 4.50e-04 | 3.15e-03 | 23614 | 17516 |
| European | Recorded Depression | Migraine | Within-class slope | SNRI | 0.035 | 0.032 | 1.118 | 1.036 | 0.974 | 1.102 | 2.63e-01 | 1.00e+00 | 23614 | 17516 |
| European | Recorded Depression | Migraine | Within-class slope | TCA | 0.020 | 0.053 | 0.381 | 1.020 | 0.920 | 1.131 | 7.03e-01 | 1.00e+00 | 23614 | 17516 |
| European | Recorded Depression | Migraine | Within-class slope | NaSSA | 0.046 | 0.087 | 0.531 | 1.048 | 0.883 | 1.243 | 5.95e-01 | 1.00e+00 | 23614 | 17516 |
| European | Recorded Depression | Migraine | Within-class slope | SARI | 0.003 | 0.054 | 0.062 | 1.003 | 0.902 | 1.116 | 9.51e-01 | 1.00e+00 | 23614 | 17516 |
| European | Recorded Depression | Migraine | Within-class slope | NDRI | 0.127 | 0.036 | 3.525 | 1.135 | 1.058 | 1.219 | 4.24e-04 | 2.97e-03 | 23614 | 17516 |
| European | Recorded Depression | Migraine | Interaction (vs SSRI) | SNRI | -0.032 | 0.035 | -0.908 | 0.969 | 0.905 | 1.037 | 3.64e-01 | 1.00e+00 | 23614 | 17516 |
| European | Recorded Depression | Migraine | Interaction (vs SSRI) | TCA | -0.047 | 0.055 | -0.861 | 0.954 | 0.857 | 1.062 | 3.89e-01 | 1.00e+00 | 23614 | 17516 |
| European | Recorded Depression | Migraine | Interaction (vs SSRI) | NaSSA | -0.021 | 0.089 | -0.232 | 0.980 | 0.824 | 1.165 | 8.16e-01 | 1.00e+00 | 23614 | 17516 |
| European | Recorded Depression | Migraine | Interaction (vs SSRI) | SARI | -0.064 | 0.056 | -1.135 | 0.938 | 0.841 | 1.047 | 2.56e-01 | 1.00e+00 | 23614 | 17516 |
| European | Recorded Depression | Migraine | Interaction (vs SSRI) | NDRI | 0.060 | 0.038 | 1.559 | 1.062 | 0.985 | 1.145 | 1.19e-01 | 8.34e-01 | 23614 | 17516 |
| European | Recorded Depression | Migraine | Joint_Interaction_Test | | | | 6.212 | | | | 2.86e-01 | 1.00e+00 | 23614 | 17516 |
| European | Recorded Depression | Insomnia | Within-class slope | SSRI | 0.075 | 0.020 | 3.722 | 1.078 | 1.036 | 1.121 | 1.98e-04 | 1.39e-03 | 23614 | 17516 |
| European | Recorded Depression | Insomnia | Within-class slope | SNRI | 0.053 | 0.033 | 1.577 | 1.054 | 0.987 | 1.126 | 1.15e-01 | 8.04e-01 | 23614 | 17516 |
| European | Recorded Depression | Insomnia | Within-class slope | TCA | 0.066 | 0.055 | 1.201 | 1.068 | 0.959 | 1.190 | 2.30e-01 | 1.00e+00 | 23614 | 17516 |
| European | Recorded Depression | Insomnia | Within-class slope | NaSSA | -0.022 | 0.087 | -0.256 | 0.978 | 0.824 | 1.160 | 7.98e-01 | 1.00e+00 | 23614 | 17516 |
| European | Recorded Depression | Insomnia | Within-class slope | SARI | -0.009 | 0.058 | -0.149 | 0.991 | 0.885 | 1.111 | 8.82e-01 | 1.00e+00 | 23614 | 17516 |
| European | Recorded Depression | Insomnia | Within-class slope | NDRI | 0.095 | 0.038 | 2.528 | 1.100 | 1.022 | 1.184 | 1.15e-02 | 8.02e-02 | 23614 | 17516 |
| European | Recorded Depression | Insomnia | Interaction (vs SSRI) | SNRI | -0.022 | 0.037 | -0.605 | 0.978 | 0.910 | 1.051 | 5.45e-01 | 1.00e+00 | 23614 | 17516 |
| European | Recorded Depression | Insomnia | Interaction (vs SSRI) | TCA | -0.009 | 0.057 | -0.156 | 0.991 | 0.886 | 1.109 | 8.76e-01 | 1.00e+00 | 23614 | 17516 |
| European | Recorded Depression | Insomnia | Interaction (vs SSRI) | NaSSA | -0.097 | 0.089 | -1.098 | 0.907 | 0.763 | 1.079 | 2.72e-01 | 1.00e+00 | 23614 | 17516 |
| European | Recorded Depression | Insomnia | Interaction (vs SSRI) | SARI | -0.084 | 0.060 | -1.401 | 0.920 | 0.818 | 1.034 | 1.61e-01 | 1.00e+00 | 23614 | 17516 |
| European | Recorded Depression | Insomnia | Interaction (vs SSRI) | NDRI | 0.020 | 0.040 | 0.500 | 1.020 | 0.943 | 1.105 | 6.17e-01 | 1.00e+00 | 23614 | 17516 |
| European | Recorded Depression | Insomnia | Joint_Interaction_Test | | | | 3.834 | | | | 5.74e-01 | 1.00e+00 | 23614 | 17516 |
| European | Recorded Depression | Depression | Within-class slope | SSRI | 0.149 | 0.020 | 7.392 | 1.160 | 1.116 | 1.207 | 1.45e-13 | 1.02e-12 | 23614 | 17516 |
| European | Recorded Depression | Depression | Within-class slope | SNRI | 0.104 | 0.033 | 3.148 | 1.109 | 1.040 | 1.183 | 1.64e-03 | 1.15e-02 | 23614 | 17516 |
| European | Recorded Depression | Depression | Within-class slope | TCA | 0.070 | 0.054 | 1.292 | 1.073 | 0.964 | 1.194 | 1.96e-01 | 1.00e+00 | 23614 | 17516 |
| European | Recorded Depression | Depression | Within-class slope | NaSSA | 0.156 | 0.092 | 1.693 | 1.168 | 0.976 | 1.399 | 9.05e-02 | 6.33e-01 | 23614 | 17516 |
| European | Recorded Depression | Depression | Within-class slope | SARI | 0.082 | 0.058 | 1.417 | 1.085 | 0.969 | 1.215 | 1.57e-01 | 1.00e+00 | 23614 | 17516 |
| European | Recorded Depression | Depression | Within-class slope | NDRI | 0.166 | 0.037 | 4.535 | 1.181 | 1.099 | 1.269 | 5.77e-06 | 4.04e-05 | 23614 | 17516 |
| European | Recorded Depression | Depression | Interaction (vs SSRI) | SNRI | -0.045 | 0.036 | -1.248 | 0.956 | 0.890 | 1.026 | 2.12e-01 | 1.00e+00 | 23614 | 17516 |
| European | Recorded Depression | Depression | Interaction (vs SSRI) | TCA | -0.078 | 0.057 | -1.389 | 0.925 | 0.828 | 1.033 | 1.65e-01 | 1.00e+00 | 23614 | 17516 |
| European | Recorded Depression | Depression | Interaction (vs SSRI) | NaSSA | 0.007 | 0.094 | 0.073 | 1.007 | 0.838 | 1.210 | 9.42e-01 | 1.00e+00 | 23614 | 17516 |
| European | Recorded Depression | Depression | Interaction (vs SSRI) | SARI | -0.067 | 0.059 | -1.132 | 0.935 | 0.832 | 1.050 | 2.58e-01 | 1.00e+00 | 23614 | 17516 |
| European | Recorded Depression | Depression | Interaction (vs SSRI) | NDRI | 0.017 | 0.040 | 0.440 | 1.018 | 0.942 | 1.100 | 6.60e-01 | 1.00e+00 | 23614 | 17516 |
| European | Recorded Depression | Depression | Joint_Interaction_Test | | | | 4.637 | | | | 4.62e-01 | 1.00e+00 | 23614 | 17516 |
| European | Recorded Depression | Schizophrenia | Within-class slope | SSRI | 0.005 | 0.019 | 0.243 | 1.005 | 0.967 | 1.044 | 8.08e-01 | 1.00e+00 | 23614 | 17516 |
| European | Recorded Depression | Schizophrenia | Within-class slope | SNRI | 0.016 | 0.033 | 0.489 | 1.016 | 0.953 | 1.083 | 6.25e-01 | 1.00e+00 | 23614 | 17516 |
| European | Recorded Depression | Schizophrenia | Within-class slope | TCA | -0.048 | 0.055 | -0.882 | 0.953 | 0.856 | 1.061 | 3.78e-01 | 1.00e+00 | 23614 | 17516 |
| European | Recorded Depression | Schizophrenia | Within-class slope | NaSSA | 0.054 | 0.087 | 0.618 | 1.055 | 0.890 | 1.250 | 5.36e-01 | 1.00e+00 | 23614 | 17516 |
| European | Recorded Depression | Schizophrenia | Within-class slope | SARI | -0.094 | 0.056 | -1.683 | 0.911 | 0.817 | 1.016 | 9.24e-02 | 6.47e-01 | 23614 | 17516 |
| European | Recorded Depression | Schizophrenia | Within-class slope | NDRI | 0.033 | 0.037 | 0.903 | 1.034 | 0.962 | 1.110 | 3.66e-01 | 1.00e+00 | 23614 | 17516 |
| European | Recorded Depression | Schizophrenia | Interaction (vs SSRI) | SNRI | 0.011 | 0.036 | 0.313 | 1.011 | 0.942 | 1.085 | 7.54e-01 | 1.00e+00 | 23614 | 17516 |
| European | Recorded Depression | Schizophrenia | Interaction (vs SSRI) | TCA | -0.053 | 0.057 | -0.939 | 0.948 | 0.849 | 1.059 | 3.48e-01 | 1.00e+00 | 23614 | 17516 |
| European | Recorded Depression | Schizophrenia | Interaction (vs SSRI) | NaSSA | 0.049 | 0.088 | 0.556 | 1.050 | 0.884 | 1.248 | 5.78e-01 | 1.00e+00 | 23614 | 17516 |
| European | Recorded Depression | Schizophrenia | Interaction (vs SSRI) | SARI | -0.098 | 0.057 | -1.710 | 0.906 | 0.810 | 1.014 | 8.73e-02 | 6.11e-01 | 23614 | 17516 |
| European | Recorded Depression | Schizophrenia | Interaction (vs SSRI) | NDRI | 0.028 | 0.039 | 0.724 | 1.029 | 0.953 | 1.111 | 4.69e-01 | 1.00e+00 | 23614 | 17516 |
| European | Recorded Depression | Schizophrenia | Joint_Interaction_Test | | | | 5.354 | | | | 3.74e-01 | 1.00e+00 | 23614 | 17516 |
| European | Recorded Depression | Bipolar Disorder | Within-class slope | SSRI | 0.010 | 0.020 | 0.495 | 1.010 | 0.971 | 1.050 | 6.20e-01 | 1.00e+00 | 23614 | 17516 |
| European | Recorded Depression | Bipolar Disorder | Within-class slope | SNRI | 0.024 | 0.033 | 0.731 | 1.025 | 0.960 | 1.094 | 4.65e-01 | 1.00e+00 | 23614 | 17516 |

| | | | | | | | | | | | | | | |
|---|---|---|---|---|---|---|---|---|---|---|---|---|---|---|
| European | Recorded Depression | Bipolar Disorder | Within-class slope | TCA | -0.021 | 0.054 | -0.384 | 0.980 | 0.882 | 1.088 | 7.01e-01 | 1.00e+00 | 23614 | 17516 |
| European | Recorded Depression | Bipolar Disorder | Within-class slope | NaSSA | -0.060 | 0.089 | -0.674 | 0.942 | 0.791 | 1.122 | 5.00e-01 | 1.00e+00 | 23614 | 17516 |
| European | Recorded Depression | Bipolar Disorder | Within-class slope | SARI | -0.015 | 0.058 | -0.263 | 0.985 | 0.880 | 1.103 | 7.93e-01 | 1.00e+00 | 23614 | 17516 |
| European | Recorded Depression | Bipolar Disorder | Within-class slope | NDRI | -0.020 | 0.038 | -0.516 | 0.981 | 0.911 | 1.056 | 6.06e-01 | 1.00e+00 | 23614 | 17516 |
| European | Recorded Depression | Bipolar Disorder | Interaction (vs SSRI) | SNRI | 0.014 | 0.037 | 0.395 | 1.015 | 0.944 | 1.090 | 6.93e-01 | 1.00e+00 | 23614 | 17516 |
| European | Recorded Depression | Bipolar Disorder | Interaction (vs SSRI) | TCA | -0.030 | 0.056 | -0.547 | 0.970 | 0.870 | 1.082 | 5.84e-01 | 1.00e+00 | 23614 | 17516 |
| European | Recorded Depression | Bipolar Disorder | Interaction (vs SSRI) | NaSSA | -0.070 | 0.091 | -0.773 | 0.932 | 0.781 | 1.114 | 4.40e-01 | 1.00e+00 | 23614 | 17516 |
| European | Recorded Depression | Bipolar Disorder | Interaction (vs SSRI) | SARI | -0.025 | 0.059 | -0.421 | 0.975 | 0.868 | 1.096 | 6.74e-01 | 1.00e+00 | 23614 | 17516 |
| European | Recorded Depression | Bipolar Disorder | Interaction (vs SSRI) | NDRI | -0.029 | 0.040 | -0.727 | 0.971 | 0.897 | 1.051 | 4.67e-01 | 1.00e+00 | 23614 | 17516 |
| European | Recorded Depression | Bipolar Disorder | Joint_Interaction_Test | | | | 1.778 | | | | 8.79e-01 | 1.00e+00 | 23614 | 17516 |
| European | Recorded Depression | BMI | Within-class slope | SSRI | 0.007 | 0.019 | 0.383 | 1.007 | 0.970 | 1.046 | 7.02e-01 | 1.00e+00 | 23614 | 17516 |
| European | Recorded Depression | BMI | Within-class slope | SNRI | -0.010 | 0.031 | -0.334 | 0.990 | 0.931 | 1.052 | 7.38e-01 | 1.00e+00 | 23614 | 17516 |
| European | Recorded Depression | BMI | Within-class slope | TCA | -0.051 | 0.052 | -0.999 | 0.950 | 0.859 | 1.051 | 3.18e-01 | 1.00e+00 | 23614 | 17516 |
| European | Recorded Depression | BMI | Within-class slope | NaSSA | 0.064 | 0.082 | 0.788 | 1.066 | 0.909 | 1.252 | 4.31e-01 | 1.00e+00 | 23614 | 17516 |
| European | Recorded Depression | BMI | Within-class slope | SARI | -0.070 | 0.053 | -1.328 | 0.933 | 0.841 | 1.034 | 1.84e-01 | 1.00e+00 | 23614 | 17516 |
| European | Recorded Depression | BMI | Within-class slope | NDRI | -0.011 | 0.036 | -0.294 | 0.989 | 0.922 | 1.062 | 7.69e-01 | 1.00e+00 | 23614 | 17516 |
| European | Recorded Depression | BMI | Interaction (vs SSRI) | SNRI | -0.018 | 0.035 | -0.514 | 0.982 | 0.918 | 1.051 | 6.07e-01 | 1.00e+00 | 23614 | 17516 |
| European | Recorded Depression | BMI | Interaction (vs SSRI) | TCA | -0.059 | 0.053 | -1.098 | 0.943 | 0.849 | 1.047 | 2.72e-01 | 1.00e+00 | 23614 | 17516 |
| European | Recorded Depression | BMI | Interaction (vs SSRI) | NaSSA | 0.057 | 0.083 | 0.687 | 1.059 | 0.900 | 1.246 | 4.92e-01 | 1.00e+00 | 23614 | 17516 |
| European | Recorded Depression | BMI | Interaction (vs SSRI) | SARI | -0.077 | 0.054 | -1.418 | 0.926 | 0.832 | 1.030 | 1.56e-01 | 1.00e+00 | 23614 | 17516 |
| European | Recorded Depression | BMI | Interaction (vs SSRI) | NDRI | -0.018 | 0.039 | -0.463 | 0.982 | 0.911 | 1.060 | 6.43e-01 | 1.00e+00 | 23614 | 17516 |
| European | Recorded Depression | BMI | Joint_Interaction_Test | | | | 3.680 | | | | 5.96e-01 | 1.00e+00 | 23614 | 17516 |
| European | No Recorded Depression | ADHD | Within-class slope | SSRI | 0.074 | 0.025 | 2.962 | 1.077 | 1.025 | 1.131 | 3.06e-03 | 2.14e-02 | 20307 | 17477 |
| European | No Recorded Depression | ADHD | Within-class slope | SNRI | 0.077 | 0.040 | 1.923 | 1.080 | 0.999 | 1.168 | 5.44e-02 | 3.81e-01 | 20307 | 17477 |
| European | No Recorded Depression | ADHD | Within-class slope | TCA | 0.036 | 0.044 | 0.824 | 1.037 | 0.952 | 1.129 | 4.10e-01 | 1.00e+00 | 20307 | 17477 |
| European | No Recorded Depression | ADHD | Within-class slope | NaSSA | 0.044 | 0.100 | 0.439 | 1.045 | 0.859 | 1.271 | 6.61e-01 | 1.00e+00 | 20307 | 17477 |
| European | No Recorded Depression | ADHD | Within-class slope | SARI | 0.048 | 0.051 | 0.940 | 1.049 | 0.949 | 1.159 | 3.47e-01 | 1.00e+00 | 20307 | 17477 |
| European | No Recorded Depression | ADHD | Within-class slope | NDRI | 0.037 | 0.050 | 0.741 | 1.038 | 0.941 | 1.146 | 4.58e-01 | 1.00e+00 | 20307 | 17477 |
| European | No Recorded Depression | ADHD | Interaction (vs SSRI) | SNRI | 0.003 | 0.045 | 0.057 | 1.003 | 0.917 | 1.096 | 9.54e-01 | 1.00e+00 | 20307 | 17477 |
| European | No Recorded Depression | ADHD | Interaction (vs SSRI) | TCA | -0.038 | 0.049 | -0.788 | 0.962 | 0.875 | 1.059 | 4.31e-01 | 1.00e+00 | 20307 | 17477 |
| European | No Recorded Depression | ADHD | Interaction (vs SSRI) | NaSSA | -0.030 | 0.102 | -0.298 | 0.970 | 0.794 | 1.184 | 7.65e-01 | 1.00e+00 | 20307 | 17477 |
| European | No Recorded Depression | ADHD | Interaction (vs SSRI) | SARI | -0.026 | 0.055 | -0.482 | 0.974 | 0.874 | 1.085 | 6.30e-01 | 1.00e+00 | 20307 | 17477 |
| European | No Recorded Depression | ADHD | Interaction (vs SSRI) | NDRI | -0.037 | 0.054 | -0.678 | 0.964 | 0.866 | 1.072 | 4.98e-01 | 1.00e+00 | 20307 | 17477 |
| European | No Recorded Depression | ADHD | Joint_Interaction_Test | | | | 1.177 | | | | 9.47e-01 | 1.00e+00 | 20307 | 17477 |
| European | No Recorded Depression | Migraine | Within-class slope | SSRI | 0.043 | 0.025 | 1.742 | 1.044 | 0.995 | 1.095 | 8.15e-02 | 5.71e-01 | 20307 | 17477 |
| European | No Recorded Depression | Migraine | Within-class slope | SNRI | -0.001 | 0.040 | -0.031 | 0.999 | 0.923 | 1.081 | 9.76e-01 | 1.00e+00 | 20307 | 17477 |
| European | No Recorded Depression | Migraine | Within-class slope | TCA | 0.025 | 0.043 | 0.586 | 1.026 | 0.942 | 1.117 | 5.58e-01 | 1.00e+00 | 20307 | 17477 |
| European | No Recorded Depression | Migraine | Within-class slope | NaSSA | 0.044 | 0.098 | 0.455 | 1.045 | 0.863 | 1.266 | 6.49e-01 | 1.00e+00 | 20307 | 17477 |
| European | No Recorded Depression | Migraine | Within-class slope | SARI | -0.017 | 0.050 | -0.337 | 0.983 | 0.892 | 1.084 | 7.36e-01 | 1.00e+00 | 20307 | 17477 |
| European | No Recorded Depression | Migraine | Within-class slope | NDRI | 0.059 | 0.051 | 1.138 | 1.060 | 0.959 | 1.173 | 2.55e-01 | 1.00e+00 | 20307 | 17477 |
| European | No Recorded Depression | Migraine | Interaction (vs SSRI) | SNRI | -0.044 | 0.045 | -0.977 | 0.957 | 0.876 | 1.045 | 3.29e-01 | 1.00e+00 | 20307 | 17477 |
| European | No Recorded Depression | Migraine | Interaction (vs SSRI) | TCA | -0.017 | 0.048 | -0.358 | 0.983 | 0.894 | 1.081 | 7.21e-01 | 1.00e+00 | 20307 | 17477 |
| European | No Recorded Depression | Migraine | Interaction (vs SSRI) | NaSSA | 0.002 | 0.100 | 0.016 | 1.002 | 0.824 | 1.218 | 9.87e-01 | 1.00e+00 | 20307 | 17477 |
| European | No Recorded Depression | Migraine | Interaction (vs SSRI) | SARI | -0.060 | 0.054 | -1.101 | 0.942 | 0.847 | 1.048 | 2.71e-01 | 1.00e+00 | 20307 | 17477 |
| European | No Recorded Depression | Migraine | Interaction (vs SSRI) | NDRI | 0.016 | 0.055 | 0.288 | 1.016 | 0.912 | 1.132 | 7.73e-01 | 1.00e+00 | 20307 | 17477 |
| European | No Recorded Depression | Migraine | Joint_Interaction_Test | | | | 2.231 | | | | 8.16e-01 | 1.00e+00 | 20307 | 17477 |
| European | No Recorded Depression | Insomnia | Within-class slope | SSRI | 0.145 | 0.026 | 5.520 | 1.156 | 1.098 | 1.217 | 3.39e-08 | 2.37e-07 | 20307 | 17477 |
| European | No Recorded Depression | Insomnia | Within-class slope | SNRI | 0.099 | 0.043 | 2.329 | 1.104 | 1.016 | 1.201 | 1.98e-02 | 1.39e-01 | 20307 | 17477 |
| European | No Recorded Depression | Insomnia | Within-class slope | TCA | 0.126 | 0.045 | 2.804 | 1.134 | 1.039 | 1.238 | 5.04e-03 | 3.53e-02 | 20307 | 17477 |
| European | No Recorded Depression | Insomnia | Within-class slope | NaSSA | 0.063 | 0.103 | 0.614 | 1.065 | 0.871 | 1.302 | 5.39e-01 | 1.00e+00 | 20307 | 17477 |
| European | No Recorded Depression | Insomnia | Within-class slope | SARI | 0.120 | 0.051 | 2.354 | 1.127 | 1.020 | 1.245 | 1.86e-02 | 1.30e-01 | 20307 | 17477 |
| European | No Recorded Depression | Insomnia | Within-class slope | NDRI | 0.070 | 0.052 | 1.360 | 1.073 | 0.969 | 1.188 | 1.74e-01 | 1.00e+00 | 20307 | 17477 |
| European | No Recorded Depression | Insomnia | Interaction (vs SSRI) | SNRI | -0.045 | 0.048 | -0.946 | 0.956 | 0.870 | 1.050 | 3.44e-01 | 1.00e+00 | 20307 | 17477 |
| European | No Recorded Depression | Insomnia | Interaction (vs SSRI) | TCA | -0.019 | 0.050 | -0.380 | 0.981 | 0.889 | 1.082 | 7.04e-01 | 1.00e+00 | 20307 | 17477 |

| | | | | | | | | | | | | | | |
|---|---|---|---|---|---|---|---|---|---|---|---|---|---|---|
| European | No Recorded Depression | Insomnia | Interaction (vs SSRI) | NaSSA | -0.082 | 0.105 | -0.781 | 0.921 | 0.750 | 1.132 | 4.35e-01 | 1.00e+00 | 20307 | 17477 |
| European | No Recorded Depression | Insomnia | Interaction (vs SSRI) | SARI | -0.025 | 0.055 | -0.454 | 0.975 | 0.875 | 1.087 | 6.50e-01 | 1.00e+00 | 20307 | 17477 |
| European | No Recorded Depression | Insomnia | Interaction (vs SSRI) | NDRI | -0.074 | 0.056 | -1.333 | 0.928 | 0.832 | 1.036 | 1.82e-01 | 1.00e+00 | 20307 | 17477 |
| European | No Recorded Depression | Insomnia | Joint_Interaction_Test | | | | 2.556 | | | | 7.68e-01 | 1.00e+00 | 20307 | 17477 |
| European | No Recorded Depression | Depression | Within-class slope | SSRI | 0.161 | 0.026 | 6.163 | 1.175 | 1.116 | 1.236 | 7.13e-10 | 4.99e-09 | 20307 | 17477 |
| European | No Recorded Depression | Depression | Within-class slope | SNRI | 0.242 | 0.043 | 5.588 | 1.273 | 1.170 | 1.386 | 2.29e-08 | 1.61e-07 | 20307 | 17477 |
| European | No Recorded Depression | Depression | Within-class slope | TCA | 0.177 | 0.045 | 3.916 | 1.193 | 1.092 | 1.304 | 9.01e-05 | 6.31e-04 | 20307 | 17477 |
| European | No Recorded Depression | Depression | Within-class slope | NaSSA | -0.036 | 0.097 | -0.376 | 0.964 | 0.798 | 1.166 | 7.07e-01 | 1.00e+00 | 20307 | 17477 |
| European | No Recorded Depression | Depression | Within-class slope | SARI | 0.164 | 0.051 | 3.234 | 1.178 | 1.067 | 1.301 | 1.22e-03 | 8.55e-03 | 20307 | 17477 |
| European | No Recorded Depression | Depression | Within-class slope | NDRI | 0.200 | 0.050 | 4.005 | 1.221 | 1.107 | 1.347 | 6.20e-05 | 4.34e-04 | 20307 | 17477 |
| European | No Recorded Depression | Depression | Interaction (vs SSRI) | SNRI | 0.081 | 0.048 | 1.665 | 1.084 | 0.986 | 1.192 | 9.58e-02 | 6.71e-01 | 20307 | 17477 |
| European | No Recorded Depression | Depression | Interaction (vs SSRI) | TCA | 0.016 | 0.051 | 0.311 | 1.016 | 0.920 | 1.122 | 7.56e-01 | 1.00e+00 | 20307 | 17477 |
| European | No Recorded Depression | Depression | Interaction (vs SSRI) | NaSSA | -0.197 | 0.099 | -1.988 | 0.821 | 0.676 | 0.997 | 4.68e-02 | 3.27e-01 | 20307 | 17477 |
| European | No Recorded Depression | Depression | Interaction (vs SSRI) | SARI | 0.003 | 0.056 | 0.053 | 1.003 | 0.899 | 1.118 | 9.58e-01 | 1.00e+00 | 20307 | 17477 |
| European | No Recorded Depression | Depression | Interaction (vs SSRI) | NDRI | 0.039 | 0.054 | 0.718 | 1.040 | 0.935 | 1.156 | 4.73e-01 | 1.00e+00 | 20307 | 17477 |
| European | No Recorded Depression | Depression | Joint_Interaction_Test | | | | 8.100 | | | | 1.51e-01 | 1.00e+00 | 20307 | 17477 |
| European | No Recorded Depression | Schizophrenia | Within-class slope | SSRI | 0.014 | 0.025 | 0.570 | 1.014 | 0.966 | 1.065 | 5.69e-01 | 1.00e+00 | 20307 | 17477 |
| European | No Recorded Depression | Schizophrenia | Within-class slope | SNRI | 0.068 | 0.041 | 1.653 | 1.071 | 0.987 | 1.161 | 9.84e-02 | 6.89e-01 | 20307 | 17477 |
| European | No Recorded Depression | Schizophrenia | Within-class slope | TCA | -0.023 | 0.044 | -0.515 | 0.978 | 0.897 | 1.066 | 6.07e-01 | 1.00e+00 | 20307 | 17477 |
| European | No Recorded Depression | Schizophrenia | Within-class slope | NaSSA | -0.051 | 0.100 | -0.509 | 0.951 | 0.782 | 1.156 | 6.11e-01 | 1.00e+00 | 20307 | 17477 |
| European | No Recorded Depression | Schizophrenia | Within-class slope | SARI | 0.096 | 0.051 | 1.864 | 1.100 | 0.995 | 1.217 | 6.23e-02 | 4.36e-01 | 20307 | 17477 |
| European | No Recorded Depression | Schizophrenia | Within-class slope | NDRI | 0.017 | 0.049 | 0.358 | 1.018 | 0.925 | 1.120 | 7.20e-01 | 1.00e+00 | 20307 | 17477 |
| European | No Recorded Depression | Schizophrenia | Interaction (vs SSRI) | SNRI | 0.054 | 0.047 | 1.159 | 1.056 | 0.963 | 1.157 | 2.46e-01 | 1.00e+00 | 20307 | 17477 |
| European | No Recorded Depression | Schizophrenia | Interaction (vs SSRI) | TCA | -0.037 | 0.049 | -0.756 | 0.964 | 0.876 | 1.060 | 4.50e-01 | 1.00e+00 | 20307 | 17477 |
| European | No Recorded Depression | Schizophrenia | Interaction (vs SSRI) | NaSSA | -0.065 | 0.102 | -0.635 | 0.937 | 0.767 | 1.145 | 5.25e-01 | 1.00e+00 | 20307 | 17477 |
| European | No Recorded Depression | Schizophrenia | Interaction (vs SSRI) | SARI | 0.082 | 0.056 | 1.466 | 1.085 | 0.973 | 1.210 | 1.43e-01 | 9.98e-01 | 20307 | 17477 |
| European | No Recorded Depression | Schizophrenia | Interaction (vs SSRI) | NDRI | 0.003 | 0.053 | 0.062 | 1.003 | 0.905 | 1.113 | 9.50e-01 | 1.00e+00 | 20307 | 17477 |
| European | No Recorded Depression | Schizophrenia | Joint_Interaction_Test | | | | 5.151 | | | | 3.98e-01 | 1.00e+00 | 20307 | 17477 |
| European | No Recorded Depression | Bipolar Disorder | Within-class slope | SSRI | 0.057 | 0.026 | 2.205 | 1.059 | 1.006 | 1.114 | 2.74e-02 | 1.92e-01 | 20307 | 17477 |
| European | No Recorded Depression | Bipolar Disorder | Within-class slope | SNRI | 0.031 | 0.043 | 0.738 | 1.032 | 0.949 | 1.122 | 4.61e-01 | 1.00e+00 | 20307 | 17477 |
| European | No Recorded Depression | Bipolar Disorder | Within-class slope | TCA | 0.063 | 0.046 | 1.369 | 1.065 | 0.973 | 1.164 | 1.71e-01 | 1.00e+00 | 20307 | 17477 |
| European | No Recorded Depression | Bipolar Disorder | Within-class slope | NaSSA | -0.028 | 0.111 | -0.252 | 0.972 | 0.783 | 1.208 | 8.01e-01 | 1.00e+00 | 20307 | 17477 |
| European | No Recorded Depression | Bipolar Disorder | Within-class slope | SARI | 0.143 | 0.053 | 2.685 | 1.154 | 1.039 | 1.281 | 7.25e-03 | 5.08e-02 | 20307 | 17477 |
| European | No Recorded Depression | Bipolar Disorder | Within-class slope | NDRI | 0.094 | 0.050 | 1.856 | 1.098 | 0.995 | 1.212 | 6.35e-02 | 4.45e-01 | 20307 | 17477 |
| European | No Recorded Depression | Bipolar Disorder | Interaction (vs SSRI) | SNRI | -0.026 | 0.048 | -0.533 | 0.975 | 0.887 | 1.071 | 5.94e-01 | 1.00e+00 | 20307 | 17477 |
| European | No Recorded Depression | Bipolar Disorder | Interaction (vs SSRI) | TCA | 0.005 | 0.051 | 0.106 | 1.005 | 0.910 | 1.111 | 9.16e-01 | 1.00e+00 | 20307 | 17477 |
| European | No Recorded Depression | Bipolar Disorder | Interaction (vs SSRI) | NaSSA | -0.085 | 0.113 | -0.753 | 0.918 | 0.736 | 1.146 | 4.52e-01 | 1.00e+00 | 20307 | 17477 |
| European | No Recorded Depression | Bipolar Disorder | Interaction (vs SSRI) | SARI | 0.086 | 0.058 | 1.486 | 1.090 | 0.973 | 1.221 | 1.37e-01 | 9.61e-01 | 20307 | 17477 |
| European | No Recorded Depression | Bipolar Disorder | Interaction (vs SSRI) | NDRI | 0.037 | 0.055 | 0.664 | 1.037 | 0.931 | 1.155 | 5.06e-01 | 1.00e+00 | 20307 | 17477 |
| European | No Recorded Depression | Bipolar Disorder | Joint_Interaction_Test | | | | 4.265 | | | | 5.12e-01 | 1.00e+00 | 20307 | 17477 |
| European | No Recorded Depression | BMI | Within-class slope | SSRI | 0.095 | 0.025 | 3.795 | 1.099 | 1.047 | 1.155 | 1.48e-04 | 1.03e-03 | 20307 | 17477 |
| European | No Recorded Depression | BMI | Within-class slope | SNRI | 0.044 | 0.040 | 1.095 | 1.045 | 0.966 | 1.131 | 2.74e-01 | 1.00e+00 | 20307 | 17477 |
| European | No Recorded Depression | BMI | Within-class slope | TCA | 0.066 | 0.045 | 1.473 | 1.068 | 0.978 | 1.166 | 1.41e-01 | 9.84e-01 | 20307 | 17477 |
| European | No Recorded Depression | BMI | Within-class slope | NaSSA | -0.004 | 0.098 | -0.037 | 0.996 | 0.822 | 1.208 | 9.71e-01 | 1.00e+00 | 20307 | 17477 |
| European | No Recorded Depression | BMI | Within-class slope | SARI | 0.042 | 0.051 | 0.821 | 1.043 | 0.944 | 1.152 | 4.12e-01 | 1.00e+00 | 20307 | 17477 |
| European | No Recorded Depression | BMI | Within-class slope | NDRI | -0.018 | 0.051 | -0.354 | 0.982 | 0.889 | 1.085 | 7.23e-01 | 1.00e+00 | 20307 | 17477 |
| European | No Recorded Depression | BMI | Interaction (vs SSRI) | SNRI | -0.051 | 0.046 | -1.113 | 0.951 | 0.869 | 1.039 | 2.66e-01 | 1.00e+00 | 20307 | 17477 |
| European | No Recorded Depression | BMI | Interaction (vs SSRI) | TCA | -0.029 | 0.050 | -0.582 | 0.972 | 0.881 | 1.071 | 5.61e-01 | 1.00e+00 | 20307 | 17477 |
| European | No Recorded Depression | BMI | Interaction (vs SSRI) | NaSSA | -0.098 | 0.101 | -0.976 | 0.906 | 0.744 | 1.104 | 3.29e-01 | 1.00e+00 | 20307 | 17477 |
| European | No Recorded Depression | BMI | Interaction (vs SSRI) | SARI | -0.053 | 0.055 | -0.968 | 0.948 | 0.852 | 1.056 | 3.33e-01 | 1.00e+00 | 20307 | 17477 |
| European | No Recorded Depression | BMI | Interaction (vs SSRI) | NDRI | -0.113 | 0.054 | -2.088 | 0.893 | 0.804 | 0.993 | 3.68e-02 | 2.58e-01 | 20307 | 17477 |
| European | No Recorded Depression | BMI | Joint_Interaction_Test | | | | 5.478 | | | | 3.60e-01 | 1.00e+00 | 20307 | 17477 |
| European | Recorded Anxiety | ADHD | Within-class slope | SSRI | 0.070 | 0.017 | 4.014 | 1.072 | 1.036 | 1.109 | 5.97e-05 | 4.18e-04 | 29610 | 22635 |
| European | Recorded Anxiety | ADHD | Within-class slope | SNRI | 0.032 | 0.030 | 1.084 | 1.033 | 0.974 | 1.094 | 2.78e-01 | 1.00e+00 | 29610 | 22635 |

| | | | | | | | | | | | | | | |
|---|---|---|---|---|---|---|---|---|---|---|---|---|---|---|
| European | Recorded Anxiety | ADHD | Within-class slope | TCA | 0.020 | 0.041 | 0.490 | 1.020 | 0.942 | 1.105 | 6.24e-01 | 1.00e+00 | 29610 | 22635 |
| European | Recorded Anxiety | ADHD | Within-class slope | NaSSA | 0.026 | 0.073 | 0.349 | 1.026 | 0.888 | 1.185 | 7.27e-01 | 1.00e+00 | 29610 | 22635 |
| European | Recorded Anxiety | ADHD | Within-class slope | SARI | 0.026 | 0.044 | 0.590 | 1.026 | 0.941 | 1.119 | 5.55e-01 | 1.00e+00 | 29610 | 22635 |
| European | Recorded Anxiety | ADHD | Within-class slope | NDRI | -0.011 | 0.036 | -0.319 | 0.989 | 0.922 | 1.060 | 7.49e-01 | 1.00e+00 | 29610 | 22635 |
| European | Recorded Anxiety | ADHD | Interaction (vs SSRI) | SNRI | -0.037 | 0.033 | -1.148 | 0.963 | 0.904 | 1.027 | 2.51e-01 | 1.00e+00 | 29610 | 22635 |
| European | Recorded Anxiety | ADHD | Interaction (vs SSRI) | TCA | -0.050 | 0.043 | -1.153 | 0.952 | 0.875 | 1.035 | 2.49e-01 | 1.00e+00 | 29610 | 22635 |
| European | Recorded Anxiety | ADHD | Interaction (vs SSRI) | NaSSA | -0.044 | 0.075 | -0.588 | 0.957 | 0.827 | 1.108 | 5.57e-01 | 1.00e+00 | 29610 | 22635 |
| European | Recorded Anxiety | ADHD | Interaction (vs SSRI) | SARI | -0.044 | 0.046 | -0.948 | 0.957 | 0.875 | 1.048 | 3.43e-01 | 1.00e+00 | 29610 | 22635 |
| European | Recorded Anxiety | ADHD | Interaction (vs SSRI) | NDRI | -0.081 | 0.038 | -2.151 | 0.922 | 0.857 | 0.993 | 3.15e-02 | 2.20e-01 | 29610 | 22635 |
| European | Recorded Anxiety | ADHD | Joint_Interaction_Test | | | | 6.005 | | | | 3.06e-01 | 1.00e+00 | 29610 | 22635 |
| European | Recorded Anxiety | Migraine | Within-class slope | SSRI | 0.059 | 0.017 | 3.515 | 1.061 | 1.026 | 1.096 | 4.40e-04 | 3.08e-03 | 29610 | 22635 |
| European | Recorded Anxiety | Migraine | Within-class slope | SNRI | 0.023 | 0.029 | 0.809 | 1.024 | 0.967 | 1.083 | 4.18e-01 | 1.00e+00 | 29610 | 22635 |
| European | Recorded Anxiety | Migraine | Within-class slope | TCA | 0.017 | 0.040 | 0.423 | 1.017 | 0.940 | 1.101 | 6.72e-01 | 1.00e+00 | 29610 | 22635 |
| European | Recorded Anxiety | Migraine | Within-class slope | NaSSA | 0.060 | 0.075 | 0.806 | 1.062 | 0.917 | 1.230 | 4.20e-01 | 1.00e+00 | 29610 | 22635 |
| European | Recorded Anxiety | Migraine | Within-class slope | SARI | 0.000 | 0.043 | 0.011 | 1.000 | 0.920 | 1.088 | 9.91e-01 | 1.00e+00 | 29610 | 22635 |
| European | Recorded Anxiety | Migraine | Within-class slope | NDRI | 0.108 | 0.035 | 3.059 | 1.114 | 1.040 | 1.194 | 2.22e-03 | 1.55e-02 | 29610 | 22635 |
| European | Recorded Anxiety | Migraine | Interaction (vs SSRI) | SNRI | -0.036 | 0.032 | -1.123 | 0.965 | 0.907 | 1.027 | 2.61e-01 | 1.00e+00 | 29610 | 22635 |
| European | Recorded Anxiety | Migraine | Interaction (vs SSRI) | TCA | -0.042 | 0.042 | -0.986 | 0.959 | 0.882 | 1.042 | 3.24e-01 | 1.00e+00 | 29610 | 22635 |
| European | Recorded Anxiety | Migraine | Interaction (vs SSRI) | NaSSA | 0.001 | 0.076 | 0.018 | 1.001 | 0.863 | 1.162 | 9.86e-01 | 1.00e+00 | 29610 | 22635 |
| European | Recorded Anxiety | Migraine | Interaction (vs SSRI) | SARI | -0.058 | 0.045 | -1.306 | 0.943 | 0.864 | 1.030 | 1.91e-01 | 1.00e+00 | 29610 | 22635 |
| European | Recorded Anxiety | Migraine | Interaction (vs SSRI) | NDRI | 0.049 | 0.037 | 1.321 | 1.050 | 0.977 | 1.129 | 1.86e-01 | 1.00e+00 | 29610 | 22635 |
| European | Recorded Anxiety | Migraine | Joint_Interaction_Test | | | | 6.243 | | | | 2.83e-01 | 1.00e+00 | 29610 | 22635 |
| European | Recorded Anxiety | Insomnia | Within-class slope | SSRI | 0.119 | 0.018 | 6.691 | 1.127 | 1.088 | 1.167 | 2.22e-11 | 1.56e-10 | 29610 | 22635 |
| European | Recorded Anxiety | Insomnia | Within-class slope | SNRI | 0.075 | 0.031 | 2.436 | 1.078 | 1.015 | 1.145 | 1.48e-02 | 1.04e-01 | 29610 | 22635 |
| European | Recorded Anxiety | Insomnia | Within-class slope | TCA | 0.088 | 0.042 | 2.120 | 1.093 | 1.007 | 1.186 | 3.40e-02 | 2.38e-01 | 29610 | 22635 |
| European | Recorded Anxiety | Insomnia | Within-class slope | NaSSA | 0.022 | 0.078 | 0.280 | 1.022 | 0.877 | 1.191 | 7.79e-01 | 1.00e+00 | 29610 | 22635 |
| European | Recorded Anxiety | Insomnia | Within-class slope | SARI | 0.014 | 0.045 | 0.311 | 1.014 | 0.928 | 1.108 | 7.56e-01 | 1.00e+00 | 29610 | 22635 |
| European | Recorded Anxiety | Insomnia | Within-class slope | NDRI | 0.091 | 0.036 | 2.505 | 1.095 | 1.020 | 1.176 | 1.22e-02 | 8.56e-02 | 29610 | 22635 |
| European | Recorded Anxiety | Insomnia | Interaction (vs SSRI) | SNRI | -0.044 | 0.034 | -1.321 | 0.957 | 0.896 | 1.022 | 1.86e-01 | 1.00e+00 | 29610 | 22635 |
| European | Recorded Anxiety | Insomnia | Interaction (vs SSRI) | TCA | -0.031 | 0.044 | -0.704 | 0.970 | 0.890 | 1.057 | 4.81e-01 | 1.00e+00 | 29610 | 22635 |
| European | Recorded Anxiety | Insomnia | Interaction (vs SSRI) | NaSSA | -0.098 | 0.079 | -1.232 | 0.907 | 0.777 | 1.059 | 2.18e-01 | 1.00e+00 | 29610 | 22635 |
| European | Recorded Anxiety | Insomnia | Interaction (vs SSRI) | SARI | -0.105 | 0.047 | -2.244 | 0.900 | 0.821 | 0.987 | 2.48e-02 | 1.74e-01 | 29610 | 22635 |
| European | Recorded Anxiety | Insomnia | Interaction (vs SSRI) | NDRI | -0.029 | 0.038 | -0.743 | 0.972 | 0.901 | 1.048 | 4.57e-01 | 1.00e+00 | 29610 | 22635 |
| European | Recorded Anxiety | Insomnia | Joint_Interaction_Test | | | | 7.019 | | | | 2.19e-01 | 1.00e+00 | 29610 | 22635 |
| European | Recorded Anxiety | Depression | Within-class slope | SSRI | 0.187 | 0.018 | 10.464 | 1.206 | 1.164 | 1.249 | <2.2e-16 | <2.2e-16 | 29610 | 22635 |
| European | Recorded Anxiety | Depression | Within-class slope | SNRI | 0.143 | 0.030 | 4.719 | 1.154 | 1.087 | 1.224 | 2.36e-06 | 1.65e-05 | 29610 | 22635 |
| European | Recorded Anxiety | Depression | Within-class slope | TCA | 0.133 | 0.041 | 3.255 | 1.142 | 1.054 | 1.237 | 1.13e-03 | 7.94e-03 | 29610 | 22635 |
| European | Recorded Anxiety | Depression | Within-class slope | NaSSA | 0.087 | 0.076 | 1.144 | 1.091 | 0.940 | 1.266 | 2.52e-01 | 1.00e+00 | 29610 | 22635 |
| European | Recorded Anxiety | Depression | Within-class slope | SARI | 0.180 | 0.045 | 4.029 | 1.198 | 1.097 | 1.307 | 5.59e-05 | 3.92e-04 | 29610 | 22635 |
| European | Recorded Anxiety | Depression | Within-class slope | NDRI | 0.183 | 0.035 | 5.154 | 1.201 | 1.120 | 1.287 | 2.55e-07 | 1.79e-06 | 29610 | 22635 |
| European | Recorded Anxiety | Depression | Interaction (vs SSRI) | SNRI | -0.044 | 0.033 | -1.324 | 0.957 | 0.896 | 1.021 | 1.85e-01 | 1.00e+00 | 29610 | 22635 |
| European | Recorded Anxiety | Depression | Interaction (vs SSRI) | TCA | -0.054 | 0.043 | -1.249 | 0.947 | 0.870 | 1.031 | 2.12e-01 | 1.00e+00 | 29610 | 22635 |
| European | Recorded Anxiety | Depression | Interaction (vs SSRI) | NaSSA | -0.100 | 0.078 | -1.289 | 0.905 | 0.777 | 1.053 | 1.97e-01 | 1.00e+00 | 29610 | 22635 |
| European | Recorded Anxiety | Depression | Interaction (vs SSRI) | SARI | -0.007 | 0.047 | -0.145 | 0.993 | 0.906 | 1.089 | 8.85e-01 | 1.00e+00 | 29610 | 22635 |
| European | Recorded Anxiety | Depression | Interaction (vs SSRI) | NDRI | -0.004 | 0.038 | -0.110 | 0.996 | 0.925 | 1.072 | 9.13e-01 | 1.00e+00 | 29610 | 22635 |
| European | Recorded Anxiety | Depression | Joint_Interaction_Test | | | | 4.120 | | | | 5.32e-01 | 1.00e+00 | 29610 | 22635 |
| European | Recorded Anxiety | Schizophrenia | Within-class slope | SSRI | 0.010 | 0.017 | 0.612 | 1.010 | 0.977 | 1.045 | 5.40e-01 | 1.00e+00 | 29610 | 22635 |
| European | Recorded Anxiety | Schizophrenia | Within-class slope | SNRI | 0.036 | 0.030 | 1.207 | 1.037 | 0.978 | 1.099 | 2.27e-01 | 1.00e+00 | 29610 | 22635 |
| European | Recorded Anxiety | Schizophrenia | Within-class slope | TCA | -0.036 | 0.041 | -0.870 | 0.965 | 0.889 | 1.046 | 3.84e-01 | 1.00e+00 | 29610 | 22635 |
| European | Recorded Anxiety | Schizophrenia | Within-class slope | NaSSA | 0.035 | 0.079 | 0.440 | 1.035 | 0.887 | 1.207 | 6.60e-01 | 1.00e+00 | 29610 | 22635 |
| European | Recorded Anxiety | Schizophrenia | Within-class slope | SARI | 0.009 | 0.044 | 0.206 | 1.009 | 0.926 | 1.099 | 8.36e-01 | 1.00e+00 | 29610 | 22635 |
| European | Recorded Anxiety | Schizophrenia | Within-class slope | NDRI | -0.002 | 0.035 | -0.055 | 0.998 | 0.931 | 1.070 | 9.56e-01 | 1.00e+00 | 29610 | 22635 |
| European | Recorded Anxiety | Schizophrenia | Interaction (vs SSRI) | SNRI | 0.026 | 0.033 | 0.783 | 1.026 | 0.962 | 1.094 | 4.34e-01 | 1.00e+00 | 29610 | 22635 |
| European | Recorded Anxiety | Schizophrenia | Interaction (vs SSRI) | TCA | -0.046 | 0.043 | -1.073 | 0.955 | 0.877 | 1.039 | 2.83e-01 | 1.00e+00 | 29610 | 22635 |

| | | | | | | | | | | | | | | |
|---|---|---|---|---|---|---|---|---|---|---|---|---|---|---|
| European | Recorded Anxiety | Schizophrenia | Interaction (vs SSRI) | NaSSA | 0.024 | 0.080 | 0.302 | 1.024 | 0.876 | 1.198 | 7.62e-01 | 1.00e+00 | 29610 | 22635 |
| European | Recorded Anxiety | Schizophrenia | Interaction (vs SSRI) | SARI | -0.001 | 0.046 | -0.031 | 0.999 | 0.913 | 1.092 | 9.75e-01 | 1.00e+00 | 29610 | 22635 |
| European | Recorded Anxiety | Schizophrenia | Interaction (vs SSRI) | NDRI | -0.012 | 0.037 | -0.330 | 0.988 | 0.918 | 1.063 | 7.41e-01 | 1.00e+00 | 29610 | 22635 |
| European | Recorded Anxiety | Schizophrenia | Joint_Interaction_Test | | | | 2.439 | | | | 7.86e-01 | 1.00e+00 | 29610 | 22635 |
| European | Recorded Anxiety | Bipolar Disorder | Within-class slope | SSRI | 0.035 | 0.018 | 1.976 | 1.036 | 1.000 | 1.072 | 4.82e-02 | 3.37e-01 | 29610 | 22635 |
| European | Recorded Anxiety | Bipolar Disorder | Within-class slope | SNRI | 0.031 | 0.030 | 1.025 | 1.032 | 0.972 | 1.095 | 3.05e-01 | 1.00e+00 | 29610 | 22635 |
| European | Recorded Anxiety | Bipolar Disorder | Within-class slope | TCA | 0.051 | 0.042 | 1.211 | 1.052 | 0.969 | 1.143 | 2.26e-01 | 1.00e+00 | 29610 | 22635 |
| European | Recorded Anxiety | Bipolar Disorder | Within-class slope | NaSSA | -0.055 | 0.079 | -0.705 | 0.946 | 0.811 | 1.104 | 4.81e-01 | 1.00e+00 | 29610 | 22635 |
| European | Recorded Anxiety | Bipolar Disorder | Within-class slope | SARI | 0.081 | 0.045 | 1.788 | 1.085 | 0.992 | 1.186 | 7.38e-02 | 5.16e-01 | 29610 | 22635 |
| European | Recorded Anxiety | Bipolar Disorder | Within-class slope | NDRI | -0.017 | 0.037 | -0.458 | 0.983 | 0.915 | 1.056 | 6.47e-01 | 1.00e+00 | 29610 | 22635 |
| European | Recorded Anxiety | Bipolar Disorder | Interaction (vs SSRI) | SNRI | -0.004 | 0.033 | -0.112 | 0.996 | 0.933 | 1.064 | 9.11e-01 | 1.00e+00 | 29610 | 22635 |
| European | Recorded Anxiety | Bipolar Disorder | Interaction (vs SSRI) | TCA | 0.016 | 0.044 | 0.362 | 1.016 | 0.932 | 1.108 | 7.17e-01 | 1.00e+00 | 29610 | 22635 |
| European | Recorded Anxiety | Bipolar Disorder | Interaction (vs SSRI) | NaSSA | -0.090 | 0.080 | -1.134 | 0.914 | 0.782 | 1.068 | 2.57e-01 | 1.00e+00 | 29610 | 22635 |
| European | Recorded Anxiety | Bipolar Disorder | Interaction (vs SSRI) | SARI | 0.046 | 0.048 | 0.974 | 1.047 | 0.954 | 1.150 | 3.30e-01 | 1.00e+00 | 29610 | 22635 |
| European | Recorded Anxiety | Bipolar Disorder | Interaction (vs SSRI) | NDRI | -0.052 | 0.039 | -1.338 | 0.950 | 0.880 | 1.024 | 1.81e-01 | 1.00e+00 | 29610 | 22635 |
| European | Recorded Anxiety | Bipolar Disorder | Joint_Interaction_Test | | | | 4.665 | | | | 4.58e-01 | 1.00e+00 | 29610 | 22635 |
| European | Recorded Anxiety | BMI | Within-class slope | SSRI | 0.051 | 0.017 | 3.043 | 1.052 | 1.018 | 1.088 | 2.34e-03 | 1.64e-02 | 29610 | 22635 |
| European | Recorded Anxiety | BMI | Within-class slope | SNRI | 0.011 | 0.029 | 0.369 | 1.011 | 0.955 | 1.070 | 7.12e-01 | 1.00e+00 | 29610 | 22635 |
| European | Recorded Anxiety | BMI | Within-class slope | TCA | 0.036 | 0.040 | 0.881 | 1.036 | 0.957 | 1.121 | 3.78e-01 | 1.00e+00 | 29610 | 22635 |
| European | Recorded Anxiety | BMI | Within-class slope | NaSSA | 0.021 | 0.075 | 0.278 | 1.021 | 0.882 | 1.182 | 7.81e-01 | 1.00e+00 | 29610 | 22635 |
| European | Recorded Anxiety | BMI | Within-class slope | SARI | 0.034 | 0.042 | 0.801 | 1.034 | 0.952 | 1.124 | 4.23e-01 | 1.00e+00 | 29610 | 22635 |
| European | Recorded Anxiety | BMI | Within-class slope | NDRI | -0.018 | 0.035 | -0.500 | 0.983 | 0.917 | 1.053 | 6.17e-01 | 1.00e+00 | 29610 | 22635 |
| European | Recorded Anxiety | BMI | Interaction (vs SSRI) | SNRI | -0.040 | 0.032 | -1.269 | 0.960 | 0.902 | 1.022 | 2.05e-01 | 1.00e+00 | 29610 | 22635 |
| European | Recorded Anxiety | BMI | Interaction (vs SSRI) | TCA | -0.016 | 0.042 | -0.368 | 0.985 | 0.906 | 1.070 | 7.13e-01 | 1.00e+00 | 29610 | 22635 |
| European | Recorded Anxiety | BMI | Interaction (vs SSRI) | NaSSA | -0.030 | 0.076 | -0.400 | 0.970 | 0.836 | 1.126 | 6.89e-01 | 1.00e+00 | 29610 | 22635 |
| European | Recorded Anxiety | BMI | Interaction (vs SSRI) | SARI | -0.017 | 0.044 | -0.391 | 0.983 | 0.902 | 1.072 | 6.96e-01 | 1.00e+00 | 29610 | 22635 |
| European | Recorded Anxiety | BMI | Interaction (vs SSRI) | NDRI | -0.069 | 0.037 | -1.857 | 0.934 | 0.868 | 1.004 | 6.33e-02 | 4.43e-01 | 29610 | 22635 |
| European | Recorded Anxiety | BMI | Joint_Interaction_Test | | | | 4.330 | | | | 5.03e-01 | 1.00e+00 | 29610 | 22635 |
| European | CIDI-MDD | ADHD | Within-class slope | SSRI | 0.056 | 0.041 | 1.363 | 1.058 | 0.976 | 1.147 | 1.73e-01 | 1.00e+00 | 5843 | 4475 |
| European | CIDI-MDD | ADHD | Within-class slope | SNRI | -0.016 | 0.062 | -0.260 | 0.984 | 0.871 | 1.112 | 7.95e-01 | 1.00e+00 | 5843 | 4475 |
| European | CIDI-MDD | ADHD | Within-class slope | TCA | -0.091 | 0.092 | -0.996 | 0.913 | 0.763 | 1.092 | 3.19e-01 | 1.00e+00 | 5843 | 4475 |
| European | CIDI-MDD | ADHD | Within-class slope | NaSSA | -0.350 | 0.205 | -1.703 | 0.705 | 0.471 | 1.054 | 8.86e-02 | 6.20e-01 | 5843 | 4475 |
| European | CIDI-MDD | ADHD | Within-class slope | SARI | 0.197 | 0.104 | 1.902 | 1.218 | 0.994 | 1.493 | 5.72e-02 | 4.00e-01 | 5843 | 4475 |
| European | CIDI-MDD | ADHD | Within-class slope | NDRI | 0.070 | 0.074 | 0.940 | 1.072 | 0.927 | 1.240 | 3.47e-01 | 1.00e+00 | 5843 | 4475 |
| European | CIDI-MDD | ADHD | Interaction (vs SSRI) | SNRI | -0.072 | 0.070 | -1.030 | 0.930 | 0.811 | 1.067 | 3.03e-01 | 1.00e+00 | 5843 | 4475 |
| European | CIDI-MDD | ADHD | Interaction (vs SSRI) | TCA | -0.147 | 0.096 | -1.535 | 0.863 | 0.715 | 1.042 | 1.25e-01 | 8.74e-01 | 5843 | 4475 |
| European | CIDI-MDD | ADHD | Interaction (vs SSRI) | NaSSA | -0.406 | 0.210 | -1.937 | 0.666 | 0.442 | 1.005 | 5.27e-02 | 3.69e-01 | 5843 | 4475 |
| European | CIDI-MDD | ADHD | Interaction (vs SSRI) | SARI | 0.141 | 0.108 | 1.305 | 1.152 | 0.932 | 1.424 | 1.92e-01 | 1.00e+00 | 5843 | 4475 |
| European | CIDI-MDD | ADHD | Interaction (vs SSRI) | NDRI | 0.013 | 0.081 | 0.167 | 1.014 | 0.865 | 1.187 | 8.67e-01 | 1.00e+00 | 5843 | 4475 |
| European | CIDI-MDD | ADHD | Joint_Interaction_Test | | | | 9.440 | | | | 9.27e-02 | 6.49e-01 | 5843 | 4475 |
| European | CIDI-MDD | Migraine | Within-class slope | SSRI | 0.030 | 0.039 | 0.762 | 1.030 | 0.954 | 1.113 | 4.46e-01 | 1.00e+00 | 5843 | 4475 |
| European | CIDI-MDD | Migraine | Within-class slope | SNRI | -0.074 | 0.059 | -1.252 | 0.928 | 0.826 | 1.043 | 2.10e-01 | 1.00e+00 | 5843 | 4475 |
| European | CIDI-MDD | Migraine | Within-class slope | TCA | 0.136 | 0.092 | 1.475 | 1.145 | 0.956 | 1.371 | 1.40e-01 | 9.81e-01 | 5843 | 4475 |
| European | CIDI-MDD | Migraine | Within-class slope | NaSSA | -0.091 | 0.207 | -0.441 | 0.913 | 0.608 | 1.370 | 6.59e-01 | 1.00e+00 | 5843 | 4475 |
| European | CIDI-MDD | Migraine | Within-class slope | SARI | -0.027 | 0.097 | -0.280 | 0.973 | 0.804 | 1.177 | 7.79e-01 | 1.00e+00 | 5843 | 4475 |
| European | CIDI-MDD | Migraine | Within-class slope | NDRI | 0.052 | 0.074 | 0.697 | 1.053 | 0.911 | 1.217 | 4.86e-01 | 1.00e+00 | 5843 | 4475 |
| European | CIDI-MDD | Migraine | Interaction (vs SSRI) | SNRI | -0.104 | 0.068 | -1.542 | 0.901 | 0.789 | 1.029 | 1.23e-01 | 8.61e-01 | 5843 | 4475 |
| European | CIDI-MDD | Migraine | Interaction (vs SSRI) | TCA | 0.106 | 0.097 | 1.089 | 1.111 | 0.919 | 1.344 | 2.76e-01 | 1.00e+00 | 5843 | 4475 |
| European | CIDI-MDD | Migraine | Interaction (vs SSRI) | NaSSA | -0.121 | 0.210 | -0.576 | 0.886 | 0.586 | 1.338 | 5.65e-01 | 1.00e+00 | 5843 | 4475 |
| European | CIDI-MDD | Migraine | Interaction (vs SSRI) | SARI | -0.057 | 0.102 | -0.563 | 0.944 | 0.774 | 1.153 | 5.74e-01 | 1.00e+00 | 5843 | 4475 |
| European | CIDI-MDD | Migraine | Interaction (vs SSRI) | NDRI | 0.022 | 0.079 | 0.273 | 1.022 | 0.875 | 1.193 | 7.85e-01 | 1.00e+00 | 5843 | 4475 |
| European | CIDI-MDD | Migraine | Joint_Interaction_Test | | | | 5.189 | | | | 3.93e-01 | 1.00e+00 | 5843 | 4475 |
| European | CIDI-MDD | Insomnia | Within-class slope | SSRI | 0.000 | 0.043 | 0.007 | 1.000 | 0.919 | 1.089 | 9.94e-01 | 1.00e+00 | 5843 | 4475 |
| European | CIDI-MDD | Insomnia | Within-class slope | SNRI | -0.049 | 0.062 | -0.792 | 0.952 | 0.843 | 1.075 | 4.28e-01 | 1.00e+00 | 5843 | 4475 |

| | | | | | | | | | | | | | | |
|---|---|---|---|---|---|---|---|---|---|---|---|---|---|---|
| European | CIDI-MDD | Insomnia | Within-class slope | TCA | 0.047 | 0.097 | 0.481 | 1.048 | 0.866 | 1.268 | 6.31e-01 | 1.00e+00 | 5843 | 4475 |
| European | CIDI-MDD | Insomnia | Within-class slope | NaSSA | 0.082 | 0.230 | 0.358 | 1.086 | 0.692 | 1.703 | 7.20e-01 | 1.00e+00 | 5843 | 4475 |
| European | CIDI-MDD | Insomnia | Within-class slope | SARI | -0.097 | 0.101 | -0.961 | 0.907 | 0.744 | 1.107 | 3.37e-01 | 1.00e+00 | 5843 | 4475 |
| European | CIDI-MDD | Insomnia | Within-class slope | NDRI | -0.052 | 0.075 | -0.693 | 0.949 | 0.820 | 1.100 | 4.89e-01 | 1.00e+00 | 5843 | 4475 |
| European | CIDI-MDD | Insomnia | Interaction (vs SSRI) | SNRI | -0.049 | 0.071 | -0.690 | 0.952 | 0.828 | 1.095 | 4.90e-01 | 1.00e+00 | 5843 | 4475 |
| European | CIDI-MDD | Insomnia | Interaction (vs SSRI) | TCA | 0.047 | 0.102 | 0.457 | 1.048 | 0.858 | 1.279 | 6.48e-01 | 1.00e+00 | 5843 | 4475 |
| European | CIDI-MDD | Insomnia | Interaction (vs SSRI) | NaSSA | 0.082 | 0.231 | 0.354 | 1.085 | 0.690 | 1.707 | 7.23e-01 | 1.00e+00 | 5843 | 4475 |
| European | CIDI-MDD | Insomnia | Interaction (vs SSRI) | SARI | -0.098 | 0.107 | -0.916 | 0.907 | 0.736 | 1.118 | 3.60e-01 | 1.00e+00 | 5843 | 4475 |
| European | CIDI-MDD | Insomnia | Interaction (vs SSRI) | NDRI | -0.052 | 0.083 | -0.628 | 0.949 | 0.806 | 1.117 | 5.30e-01 | 1.00e+00 | 5843 | 4475 |
| European | CIDI-MDD | Insomnia | Joint_Interaction_Test | | | | 1.942 | | | | 8.57e-01 | 1.00e+00 | 5843 | 4475 |
| European | CIDI-MDD | Depression | Within-class slope | SSRI | 0.096 | 0.043 | 2.270 | 1.101 | 1.013 | 1.197 | 2.32e-02 | 1.63e-01 | 5843 | 4475 |
| European | CIDI-MDD | Depression | Within-class slope | SNRI | 0.098 | 0.063 | 1.561 | 1.103 | 0.975 | 1.248 | 1.19e-01 | 8.30e-01 | 5843 | 4475 |
| European | CIDI-MDD | Depression | Within-class slope | TCA | 0.027 | 0.095 | 0.284 | 1.027 | 0.853 | 1.238 | 7.76e-01 | 1.00e+00 | 5843 | 4475 |
| European | CIDI-MDD | Depression | Within-class slope | NaSSA | 0.013 | 0.232 | 0.055 | 1.013 | 0.643 | 1.596 | 9.56e-01 | 1.00e+00 | 5843 | 4475 |
| European | CIDI-MDD | Depression | Within-class slope | SARI | 0.019 | 0.104 | 0.186 | 1.020 | 0.831 | 1.251 | 8.53e-01 | 1.00e+00 | 5843 | 4475 |
| European | CIDI-MDD | Depression | Within-class slope | NDRI | 0.120 | 0.074 | 1.629 | 1.128 | 0.976 | 1.303 | 1.03e-01 | 7.24e-01 | 5843 | 4475 |
| European | CIDI-MDD | Depression | Interaction (vs SSRI) | SNRI | 0.002 | 0.072 | 0.025 | 1.002 | 0.870 | 1.154 | 9.80e-01 | 1.00e+00 | 5843 | 4475 |
| European | CIDI-MDD | Depression | Interaction (vs SSRI) | TCA | -0.069 | 0.102 | -0.681 | 0.933 | 0.764 | 1.140 | 4.96e-01 | 1.00e+00 | 5843 | 4475 |
| European | CIDI-MDD | Depression | Interaction (vs SSRI) | NaSSA | -0.084 | 0.237 | -0.354 | 0.920 | 0.579 | 1.462 | 7.23e-01 | 1.00e+00 | 5843 | 4475 |
| European | CIDI-MDD | Depression | Interaction (vs SSRI) | SARI | -0.077 | 0.110 | -0.703 | 0.926 | 0.747 | 1.148 | 4.82e-01 | 1.00e+00 | 5843 | 4475 |
| European | CIDI-MDD | Depression | Interaction (vs SSRI) | NDRI | 0.024 | 0.080 | 0.294 | 1.024 | 0.875 | 1.198 | 7.68e-01 | 1.00e+00 | 5843 | 4475 |
| European | CIDI-MDD | Depression | Joint_Interaction_Test | | | | 1.215 | | | | 9.43e-01 | 1.00e+00 | 5843 | 4475 |
| European | CIDI-MDD | Schizophrenia | Within-class slope | SSRI | 0.030 | 0.042 | 0.714 | 1.030 | 0.950 | 1.117 | 4.75e-01 | 1.00e+00 | 5843 | 4475 |
| European | CIDI-MDD | Schizophrenia | Within-class slope | SNRI | 0.086 | 0.064 | 1.341 | 1.090 | 0.961 | 1.235 | 1.80e-01 | 1.00e+00 | 5843 | 4475 |
| European | CIDI-MDD | Schizophrenia | Within-class slope | TCA | 0.125 | 0.101 | 1.239 | 1.133 | 0.930 | 1.382 | 2.15e-01 | 1.00e+00 | 5843 | 4475 |
| European | CIDI-MDD | Schizophrenia | Within-class slope | NaSSA | 0.024 | 0.214 | 0.113 | 1.024 | 0.673 | 1.560 | 9.10e-01 | 1.00e+00 | 5843 | 4475 |
| European | CIDI-MDD | Schizophrenia | Within-class slope | SARI | 0.129 | 0.101 | 1.276 | 1.137 | 0.933 | 1.386 | 2.02e-01 | 1.00e+00 | 5843 | 4475 |
| European | CIDI-MDD | Schizophrenia | Within-class slope | NDRI | 0.034 | 0.073 | 0.461 | 1.034 | 0.896 | 1.193 | 6.45e-01 | 1.00e+00 | 5843 | 4475 |
| European | CIDI-MDD | Schizophrenia | Interaction (vs SSRI) | SNRI | 0.056 | 0.073 | 0.766 | 1.058 | 0.916 | 1.222 | 4.44e-01 | 1.00e+00 | 5843 | 4475 |
| European | CIDI-MDD | Schizophrenia | Interaction (vs SSRI) | TCA | 0.096 | 0.106 | 0.903 | 1.100 | 0.894 | 1.354 | 3.67e-01 | 1.00e+00 | 5843 | 4475 |
| European | CIDI-MDD | Schizophrenia | Interaction (vs SSRI) | NaSSA | -0.005 | 0.218 | -0.025 | 0.995 | 0.649 | 1.523 | 9.80e-01 | 1.00e+00 | 5843 | 4475 |
| European | CIDI-MDD | Schizophrenia | Interaction (vs SSRI) | SARI | 0.099 | 0.107 | 0.930 | 1.104 | 0.896 | 1.361 | 3.53e-01 | 1.00e+00 | 5843 | 4475 |
| European | CIDI-MDD | Schizophrenia | Interaction (vs SSRI) | NDRI | 0.004 | 0.079 | 0.051 | 1.004 | 0.860 | 1.173 | 9.59e-01 | 1.00e+00 | 5843 | 4475 |
| European | CIDI-MDD | Schizophrenia | Joint_Interaction_Test | | | | 1.816 | | | | 8.74e-01 | 1.00e+00 | 5843 | 4475 |
| European | CIDI-MDD | Bipolar Disorder | Within-class slope | SSRI | 0.033 | 0.042 | 0.782 | 1.034 | 0.952 | 1.123 | 4.34e-01 | 1.00e+00 | 5843 | 4475 |
| European | CIDI-MDD | Bipolar Disorder | Within-class slope | SNRI | 0.064 | 0.064 | 0.998 | 1.066 | 0.941 | 1.207 | 3.18e-01 | 1.00e+00 | 5843 | 4475 |
| European | CIDI-MDD | Bipolar Disorder | Within-class slope | TCA | -0.043 | 0.106 | -0.408 | 0.958 | 0.778 | 1.179 | 6.84e-01 | 1.00e+00 | 5843 | 4475 |
| European | CIDI-MDD | Bipolar Disorder | Within-class slope | NaSSA | 0.091 | 0.200 | 0.453 | 1.095 | 0.740 | 1.620 | 6.50e-01 | 1.00e+00 | 5843 | 4475 |
| European | CIDI-MDD | Bipolar Disorder | Within-class slope | SARI | 0.044 | 0.108 | 0.403 | 1.045 | 0.845 | 1.292 | 6.87e-01 | 1.00e+00 | 5843 | 4475 |
| European | CIDI-MDD | Bipolar Disorder | Within-class slope | NDRI | 0.014 | 0.074 | 0.191 | 1.014 | 0.877 | 1.172 | 8.49e-01 | 1.00e+00 | 5843 | 4475 |
| European | CIDI-MDD | Bipolar Disorder | Interaction (vs SSRI) | SNRI | 0.031 | 0.073 | 0.420 | 1.031 | 0.894 | 1.189 | 6.74e-01 | 1.00e+00 | 5843 | 4475 |
| European | CIDI-MDD | Bipolar Disorder | Interaction (vs SSRI) | TCA | -0.076 | 0.111 | -0.686 | 0.927 | 0.745 | 1.152 | 4.93e-01 | 1.00e+00 | 5843 | 4475 |
| European | CIDI-MDD | Bipolar Disorder | Interaction (vs SSRI) | NaSSA | 0.058 | 0.203 | 0.284 | 1.059 | 0.712 | 1.577 | 7.76e-01 | 1.00e+00 | 5843 | 4475 |
| European | CIDI-MDD | Bipolar Disorder | Interaction (vs SSRI) | SARI | 0.011 | 0.114 | 0.095 | 1.011 | 0.809 | 1.264 | 9.24e-01 | 1.00e+00 | 5843 | 4475 |
| European | CIDI-MDD | Bipolar Disorder | Interaction (vs SSRI) | NDRI | -0.019 | 0.081 | -0.232 | 0.981 | 0.837 | 1.151 | 8.17e-01 | 1.00e+00 | 5843 | 4475 |
| European | CIDI-MDD | Bipolar Disorder | Joint_Interaction_Test | | | | 0.970 | | | | 9.65e-01 | 1.00e+00 | 5843 | 4475 |
| European | CIDI-MDD | BMI | Within-class slope | SSRI | 0.093 | 0.041 | 2.249 | 1.097 | 1.012 | 1.189 | 2.45e-02 | 1.72e-01 | 5843 | 4475 |
| European | CIDI-MDD | BMI | Within-class slope | SNRI | 0.000 | 0.062 | -0.006 | 1.000 | 0.885 | 1.129 | 9.95e-01 | 1.00e+00 | 5843 | 4475 |
| European | CIDI-MDD | BMI | Within-class slope | TCA | 0.191 | 0.100 | 1.905 | 1.211 | 0.995 | 1.475 | 5.67e-02 | 3.97e-01 | 5843 | 4475 |
| European | CIDI-MDD | BMI | Within-class slope | NaSSA | 0.056 | 0.234 | 0.239 | 1.057 | 0.668 | 1.673 | 8.11e-01 | 1.00e+00 | 5843 | 4475 |
| European | CIDI-MDD | BMI | Within-class slope | SARI | 0.100 | 0.101 | 0.995 | 1.106 | 0.907 | 1.347 | 3.20e-01 | 1.00e+00 | 5843 | 4475 |
| European | CIDI-MDD | BMI | Within-class slope | NDRI | -0.102 | 0.071 | -1.426 | 0.903 | 0.785 | 1.039 | 1.54e-01 | 1.00e+00 | 5843 | 4475 |
| European | CIDI-MDD | BMI | Interaction (vs SSRI) | SNRI | -0.093 | 0.070 | -1.322 | 0.911 | 0.794 | 1.046 | 1.86e-01 | 1.00e+00 | 5843 | 4475 |
| European | CIDI-MDD | BMI | Interaction (vs SSRI) | TCA | 0.099 | 0.107 | 0.929 | 1.104 | 0.896 | 1.360 | 3.53e-01 | 1.00e+00 | 5843 | 4475 |

| | | | | | | | | | | | | | | |
|---|---|---|---|---|---|---|---|---|---|---|---|---|---|---|
| European | CIDI-MDD | BMI | Interaction (vs SSRI) | NaSSA | -0.037 | 0.236 | -0.155 | 0.964 | 0.607 | 1.531 | 8.77e-01 | 1.00e+00 | 5843 | 4475 |
| European | CIDI-MDD | BMI | Interaction (vs SSRI) | SARI | 0.008 | 0.106 | 0.074 | 1.008 | 0.819 | 1.240 | 9.41e-01 | 1.00e+00 | 5843 | 4475 |
| European | CIDI-MDD | BMI | Interaction (vs SSRI) | NDRI | -0.194 | 0.077 | -2.515 | 0.823 | 0.707 | 0.958 | 1.19e-02 | 8.33e-02 | 5843 | 4475 |
| European | CIDI-MDD | BMI | Joint_Interaction_Test | | | | 9.410 | | | | 9.38e-02 | 6.56e-01 | 5843 | 4475 |
| European | CIDI-noMDD | ADHD | Within-class slope | SSRI | 0.000 | 0.066 | 0.004 | 1.000 | 0.879 | 1.138 | 9.97e-01 | 1.00e+00 | 3146 | 2660 |
| European | CIDI-noMDD | ADHD | Within-class slope | SNRI | 0.038 | 0.107 | 0.357 | 1.039 | 0.842 | 1.281 | 7.21e-01 | 1.00e+00 | 3146 | 2660 |
| European | CIDI-noMDD | ADHD | Within-class slope | TCA | 0.157 | 0.098 | 1.604 | 1.170 | 0.966 | 1.417 | 1.09e-01 | 7.61e-01 | 3146 | 2660 |
| European | CIDI-noMDD | ADHD | Within-class slope | NaSSA | 0.108 | 0.260 | 0.416 | 1.114 | 0.669 | 1.857 | 6.78e-01 | 1.00e+00 | 3146 | 2660 |
| European | CIDI-noMDD | ADHD | Within-class slope | SARI | 0.062 | 0.120 | 0.513 | 1.064 | 0.841 | 1.345 | 6.08e-01 | 1.00e+00 | 3146 | 2660 |
| European | CIDI-noMDD | ADHD | Within-class slope | NDRI | 0.124 | 0.136 | 0.911 | 1.132 | 0.867 | 1.478 | 3.63e-01 | 1.00e+00 | 3146 | 2660 |
| European | CIDI-noMDD | ADHD | Interaction (vs SSRI) | SNRI | 0.038 | 0.120 | 0.317 | 1.039 | 0.821 | 1.314 | 7.51e-01 | 1.00e+00 | 3146 | 2660 |
| European | CIDI-noMDD | ADHD | Interaction (vs SSRI) | TCA | 0.157 | 0.113 | 1.381 | 1.170 | 0.936 | 1.461 | 1.67e-01 | 1.00e+00 | 3146 | 2660 |
| European | CIDI-noMDD | ADHD | Interaction (vs SSRI) | NaSSA | 0.108 | 0.266 | 0.405 | 1.114 | 0.661 | 1.878 | 6.85e-01 | 1.00e+00 | 3146 | 2660 |
| European | CIDI-noMDD | ADHD | Interaction (vs SSRI) | SARI | 0.061 | 0.132 | 0.464 | 1.063 | 0.821 | 1.378 | 6.43e-01 | 1.00e+00 | 3146 | 2660 |
| European | CIDI-noMDD | ADHD | Interaction (vs SSRI) | NDRI | 0.124 | 0.139 | 0.892 | 1.132 | 0.862 | 1.485 | 3.73e-01 | 1.00e+00 | 3146 | 2660 |
| European | CIDI-noMDD | ADHD | Joint_Interaction_Test | | | | 2.445 | | | | 7.85e-01 | 1.00e+00 | 3146 | 2660 |
| European | CIDI-noMDD | Migraine | Within-class slope | SSRI | 0.003 | 0.061 | 0.047 | 1.003 | 0.891 | 1.129 | 9.62e-01 | 1.00e+00 | 3146 | 2660 |
| European | CIDI-noMDD | Migraine | Within-class slope | SNRI | -0.094 | 0.095 | -0.999 | 0.910 | 0.756 | 1.095 | 3.18e-01 | 1.00e+00 | 3146 | 2660 |
| European | CIDI-noMDD | Migraine | Within-class slope | TCA | -0.335 | 0.100 | -3.362 | 0.715 | 0.588 | 0.870 | 7.74e-04 | 5.41e-03 | 3146 | 2660 |
| European | CIDI-noMDD | Migraine | Within-class slope | NaSSA | 0.254 | 0.207 | 1.230 | 1.290 | 0.860 | 1.934 | 2.19e-01 | 1.00e+00 | 3146 | 2660 |
| European | CIDI-noMDD | Migraine | Within-class slope | SARI | 0.195 | 0.128 | 1.526 | 1.215 | 0.946 | 1.561 | 1.27e-01 | 8.88e-01 | 3146 | 2660 |
| European | CIDI-noMDD | Migraine | Within-class slope | NDRI | 0.050 | 0.137 | 0.364 | 1.051 | 0.804 | 1.375 | 7.16e-01 | 1.00e+00 | 3146 | 2660 |
| European | CIDI-noMDD | Migraine | Interaction (vs SSRI) | SNRI | -0.097 | 0.107 | -0.906 | 0.907 | 0.735 | 1.120 | 3.65e-01 | 1.00e+00 | 3146 | 2660 |
| European | CIDI-noMDD | Migraine | Interaction (vs SSRI) | TCA | -0.338 | 0.113 | -2.985 | 0.713 | 0.571 | 0.890 | 2.84e-03 | 1.98e-02 | 3146 | 2660 |
| European | CIDI-noMDD | Migraine | Interaction (vs SSRI) | NaSSA | 0.251 | 0.214 | 1.173 | 1.286 | 0.845 | 1.957 | 2.41e-01 | 1.00e+00 | 3146 | 2660 |
| European | CIDI-noMDD | Migraine | Interaction (vs SSRI) | SARI | 0.192 | 0.139 | 1.384 | 1.212 | 0.923 | 1.590 | 1.66e-01 | 1.00e+00 | 3146 | 2660 |
| European | CIDI-noMDD | Migraine | Interaction (vs SSRI) | NDRI | 0.047 | 0.141 | 0.334 | 1.048 | 0.795 | 1.382 | 7.38e-01 | 1.00e+00 | 3146 | 2660 |
| European | CIDI-noMDD | Migraine | Joint_Interaction_Test | | | | 16.214 | | | | 6.26e-03 | 4.38e-02 | 3146 | 2660 |
| European | CIDI-noMDD | Insomnia | Within-class slope | SSRI | 0.106 | 0.066 | 1.625 | 1.112 | 0.978 | 1.265 | 1.04e-01 | 7.30e-01 | 3146 | 2660 |
| European | CIDI-noMDD | Insomnia | Within-class slope | SNRI | 0.049 | 0.106 | 0.463 | 1.050 | 0.853 | 1.292 | 6.43e-01 | 1.00e+00 | 3146 | 2660 |
| European | CIDI-noMDD | Insomnia | Within-class slope | TCA | 0.156 | 0.121 | 1.284 | 1.168 | 0.921 | 1.482 | 1.99e-01 | 1.00e+00 | 3146 | 2660 |
| European | CIDI-noMDD | Insomnia | Within-class slope | NaSSA | 0.110 | 0.227 | 0.483 | 1.116 | 0.715 | 1.740 | 6.29e-01 | 1.00e+00 | 3146 | 2660 |
| European | CIDI-noMDD | Insomnia | Within-class slope | SARI | 0.197 | 0.126 | 1.571 | 1.218 | 0.952 | 1.559 | 1.16e-01 | 8.13e-01 | 3146 | 2660 |
| European | CIDI-noMDD | Insomnia | Within-class slope | NDRI | 0.222 | 0.131 | 1.691 | 1.249 | 0.965 | 1.616 | 9.08e-02 | 6.36e-01 | 3146 | 2660 |
| European | CIDI-noMDD | Insomnia | Interaction (vs SSRI) | SNRI | -0.057 | 0.119 | -0.481 | 0.944 | 0.747 | 1.193 | 6.30e-01 | 1.00e+00 | 3146 | 2660 |
| European | CIDI-noMDD | Insomnia | Interaction (vs SSRI) | TCA | 0.049 | 0.134 | 0.367 | 1.050 | 0.808 | 1.365 | 7.13e-01 | 1.00e+00 | 3146 | 2660 |
| European | CIDI-noMDD | Insomnia | Interaction (vs SSRI) | NaSSA | 0.003 | 0.233 | 0.013 | 1.003 | 0.635 | 1.584 | 9.90e-01 | 1.00e+00 | 3146 | 2660 |
| European | CIDI-noMDD | Insomnia | Interaction (vs SSRI) | SARI | 0.091 | 0.136 | 0.670 | 1.095 | 0.839 | 1.429 | 5.03e-01 | 1.00e+00 | 3146 | 2660 |
| European | CIDI-noMDD | Insomnia | Interaction (vs SSRI) | NDRI | 0.116 | 0.142 | 0.815 | 1.123 | 0.850 | 1.483 | 4.15e-01 | 1.00e+00 | 3146 | 2660 |
| European | CIDI-noMDD | Insomnia | Joint_Interaction_Test | | | | 1.677 | | | | 8.92e-01 | 1.00e+00 | 3146 | 2660 |
| European | CIDI-noMDD | Depression | Within-class slope | SSRI | 0.092 | 0.069 | 1.344 | 1.096 | 0.959 | 1.254 | 1.79e-01 | 1.00e+00 | 3146 | 2660 |
| European | CIDI-noMDD | Depression | Within-class slope | SNRI | 0.079 | 0.111 | 0.711 | 1.082 | 0.871 | 1.345 | 4.77e-01 | 1.00e+00 | 3146 | 2660 |
| European | CIDI-noMDD | Depression | Within-class slope | TCA | 0.253 | 0.112 | 2.257 | 1.288 | 1.034 | 1.605 | 2.40e-02 | 1.68e-01 | 3146 | 2660 |
| European | CIDI-noMDD | Depression | Within-class slope | NaSSA | 0.002 | 0.269 | 0.006 | 1.002 | 0.591 | 1.697 | 9.95e-01 | 1.00e+00 | 3146 | 2660 |
| European | CIDI-noMDD | Depression | Within-class slope | SARI | 0.101 | 0.127 | 0.794 | 1.106 | 0.862 | 1.419 | 4.27e-01 | 1.00e+00 | 3146 | 2660 |
| European | CIDI-noMDD | Depression | Within-class slope | NDRI | 0.198 | 0.139 | 1.425 | 1.219 | 0.929 | 1.599 | 1.54e-01 | 1.00e+00 | 3146 | 2660 |
| European | CIDI-noMDD | Depression | Interaction (vs SSRI) | SNRI | -0.013 | 0.123 | -0.108 | 0.987 | 0.775 | 1.256 | 9.14e-01 | 1.00e+00 | 3146 | 2660 |
| European | CIDI-noMDD | Depression | Interaction (vs SSRI) | TCA | 0.161 | 0.127 | 1.269 | 1.175 | 0.916 | 1.507 | 2.05e-01 | 1.00e+00 | 3146 | 2660 |
| European | CIDI-noMDD | Depression | Interaction (vs SSRI) | NaSSA | -0.090 | 0.275 | -0.329 | 0.914 | 0.533 | 1.565 | 7.42e-01 | 1.00e+00 | 3146 | 2660 |
| European | CIDI-noMDD | Depression | Interaction (vs SSRI) | SARI | 0.009 | 0.142 | 0.062 | 1.009 | 0.764 | 1.331 | 9.50e-01 | 1.00e+00 | 3146 | 2660 |
| European | CIDI-noMDD | Depression | Interaction (vs SSRI) | NDRI | 0.106 | 0.146 | 0.725 | 1.111 | 0.835 | 1.479 | 4.69e-01 | 1.00e+00 | 3146 | 2660 |
| European | CIDI-noMDD | Depression | Joint_Interaction_Test | | | | 2.456 | | | | 7.83e-01 | 1.00e+00 | 3146 | 2660 |
| European | CIDI-noMDD | Schizophrenia | Within-class slope | SSRI | 0.018 | 0.063 | 0.282 | 1.018 | 0.900 | 1.151 | 7.78e-01 | 1.00e+00 | 3146 | 2660 |
| European | CIDI-noMDD | Schizophrenia | Within-class slope | SNRI | 0.027 | 0.108 | 0.248 | 1.027 | 0.832 | 1.268 | 8.04e-01 | 1.00e+00 | 3146 | 2660 |

| | | | | | | | | | | | | | | |
|---|---|---|---|---|---|---|---|---|---|---|---|---|---|---|
| European | CIDI-noMDD | Schizophrenia | Within-class slope | TCA | 0.132 | 0.111 | 1.186 | 1.141 | 0.918 | 1.419 | 2.35e-01 | 1.00e+00 | 3146 | 2660 |
| European | CIDI-noMDD | Schizophrenia | Within-class slope | NaSSA | 0.149 | 0.281 | 0.530 | 1.161 | 0.669 | 2.015 | 5.96e-01 | 1.00e+00 | 3146 | 2660 |
| European | CIDI-noMDD | Schizophrenia | Within-class slope | SARI | 0.126 | 0.136 | 0.923 | 1.134 | 0.868 | 1.481 | 3.56e-01 | 1.00e+00 | 3146 | 2660 |
| European | CIDI-noMDD | Schizophrenia | Within-class slope | NDRI | 0.070 | 0.132 | 0.529 | 1.072 | 0.828 | 1.389 | 5.97e-01 | 1.00e+00 | 3146 | 2660 |
| European | CIDI-noMDD | Schizophrenia | Interaction (vs SSRI) | SNRI | 0.009 | 0.118 | 0.077 | 1.009 | 0.801 | 1.271 | 9.39e-01 | 1.00e+00 | 3146 | 2660 |
| European | CIDI-noMDD | Schizophrenia | Interaction (vs SSRI) | TCA | 0.114 | 0.124 | 0.924 | 1.121 | 0.880 | 1.428 | 3.55e-01 | 1.00e+00 | 3146 | 2660 |
| European | CIDI-noMDD | Schizophrenia | Interaction (vs SSRI) | NaSSA | 0.131 | 0.284 | 0.463 | 1.140 | 0.653 | 1.991 | 6.44e-01 | 1.00e+00 | 3146 | 2660 |
| European | CIDI-noMDD | Schizophrenia | Interaction (vs SSRI) | SARI | 0.108 | 0.147 | 0.737 | 1.114 | 0.836 | 1.485 | 4.61e-01 | 1.00e+00 | 3146 | 2660 |
| European | CIDI-noMDD | Schizophrenia | Interaction (vs SSRI) | NDRI | 0.052 | 0.136 | 0.382 | 1.053 | 0.806 | 1.377 | 7.03e-01 | 1.00e+00 | 3146 | 2660 |
| European | CIDI-noMDD | Schizophrenia | Joint_Interaction_Test | | | | 1.468 | | | | 9.17e-01 | 1.00e+00 | 3146 | 2660 |
| European | CIDI-noMDD | Bipolar Disorder | Within-class slope | SSRI | 0.039 | 0.067 | 0.572 | 1.039 | 0.911 | 1.186 | 5.67e-01 | 1.00e+00 | 3146 | 2660 |
| European | CIDI-noMDD | Bipolar Disorder | Within-class slope | SNRI | 0.035 | 0.114 | 0.306 | 1.035 | 0.829 | 1.294 | 7.59e-01 | 1.00e+00 | 3146 | 2660 |
| European | CIDI-noMDD | Bipolar Disorder | Within-class slope | TCA | 0.241 | 0.123 | 1.959 | 1.272 | 1.000 | 1.619 | 5.01e-02 | 3.50e-01 | 3146 | 2660 |
| European | CIDI-noMDD | Bipolar Disorder | Within-class slope | NaSSA | 0.145 | 0.260 | 0.558 | 1.156 | 0.694 | 1.926 | 5.77e-01 | 1.00e+00 | 3146 | 2660 |
| European | CIDI-noMDD | Bipolar Disorder | Within-class slope | SARI | 0.013 | 0.135 | 0.095 | 1.013 | 0.777 | 1.320 | 9.24e-01 | 1.00e+00 | 3146 | 2660 |
| European | CIDI-noMDD | Bipolar Disorder | Within-class slope | NDRI | 0.209 | 0.136 | 1.540 | 1.233 | 0.945 | 1.608 | 1.24e-01 | 8.65e-01 | 3146 | 2660 |
| European | CIDI-noMDD | Bipolar Disorder | Interaction (vs SSRI) | SNRI | -0.004 | 0.125 | -0.030 | 0.996 | 0.780 | 1.272 | 9.76e-01 | 1.00e+00 | 3146 | 2660 |
| European | CIDI-noMDD | Bipolar Disorder | Interaction (vs SSRI) | TCA | 0.202 | 0.134 | 1.504 | 1.224 | 0.941 | 1.593 | 1.33e-01 | 9.28e-01 | 3146 | 2660 |
| European | CIDI-noMDD | Bipolar Disorder | Interaction (vs SSRI) | NaSSA | 0.107 | 0.264 | 0.405 | 1.113 | 0.664 | 1.865 | 6.86e-01 | 1.00e+00 | 3146 | 2660 |
| European | CIDI-noMDD | Bipolar Disorder | Interaction (vs SSRI) | SARI | -0.026 | 0.148 | -0.174 | 0.975 | 0.730 | 1.302 | 8.62e-01 | 1.00e+00 | 3146 | 2660 |
| European | CIDI-noMDD | Bipolar Disorder | Interaction (vs SSRI) | NDRI | 0.170 | 0.142 | 1.201 | 1.186 | 0.898 | 1.566 | 2.30e-01 | 1.00e+00 | 3146 | 2660 |
| European | CIDI-noMDD | Bipolar Disorder | Joint_Interaction_Test | | | | 4.037 | | | | 5.44e-01 | 1.00e+00 | 3146 | 2660 |
| European | CIDI-noMDD | BMI | Within-class slope | SSRI | 0.024 | 0.063 | 0.381 | 1.024 | 0.905 | 1.159 | 7.03e-01 | 1.00e+00 | 3146 | 2660 |
| European | CIDI-noMDD | BMI | Within-class slope | SNRI | -0.109 | 0.107 | -1.023 | 0.896 | 0.727 | 1.105 | 3.06e-01 | 1.00e+00 | 3146 | 2660 |
| European | CIDI-noMDD | BMI | Within-class slope | TCA | 0.071 | 0.111 | 0.637 | 1.073 | 0.863 | 1.334 | 5.24e-01 | 1.00e+00 | 3146 | 2660 |
| European | CIDI-noMDD | BMI | Within-class slope | NaSSA | 0.070 | 0.232 | 0.301 | 1.072 | 0.681 | 1.688 | 7.63e-01 | 1.00e+00 | 3146 | 2660 |
| European | CIDI-noMDD | BMI | Within-class slope | SARI | 0.138 | 0.145 | 0.951 | 1.148 | 0.864 | 1.526 | 3.42e-01 | 1.00e+00 | 3146 | 2660 |
| European | CIDI-noMDD | BMI | Within-class slope | NDRI | 0.150 | 0.134 | 1.114 | 1.161 | 0.893 | 1.511 | 2.65e-01 | 1.00e+00 | 3146 | 2660 |
| European | CIDI-noMDD | BMI | Interaction (vs SSRI) | SNRI | -0.134 | 0.118 | -1.135 | 0.875 | 0.695 | 1.102 | 2.56e-01 | 1.00e+00 | 3146 | 2660 |
| European | CIDI-noMDD | BMI | Interaction (vs SSRI) | TCA | 0.047 | 0.125 | 0.374 | 1.048 | 0.821 | 1.338 | 7.09e-01 | 1.00e+00 | 3146 | 2660 |
| European | CIDI-noMDD | BMI | Interaction (vs SSRI) | NaSSA | 0.046 | 0.240 | 0.190 | 1.047 | 0.654 | 1.676 | 8.49e-01 | 1.00e+00 | 3146 | 2660 |
| European | CIDI-noMDD | BMI | Interaction (vs SSRI) | SARI | 0.114 | 0.151 | 0.756 | 1.121 | 0.834 | 1.506 | 4.49e-01 | 1.00e+00 | 3146 | 2660 |
| European | CIDI-noMDD | BMI | Interaction (vs SSRI) | NDRI | 0.126 | 0.140 | 0.897 | 1.134 | 0.862 | 1.492 | 3.70e-01 | 1.00e+00 | 3146 | 2660 |
| European | CIDI-noMDD | BMI | Joint_Interaction_Test | | | | 3.597 | | | | 6.09e-01 | 1.00e+00 | 3146 | 2660 |
| African | Full Cohort | ADHD | Within-class slope | SSRI | 0.030 | 0.051 | 0.586 | 1.030 | 0.932 | 1.139 | 5.58e-01 | 1.00e+00 | 8442 | 6685 |
| African | Full Cohort | ADHD | Within-class slope | SNRI | 0.021 | 0.074 | 0.284 | 1.021 | 0.883 | 1.181 | 7.77e-01 | 1.00e+00 | 8442 | 6685 |
| African | Full Cohort | ADHD | Within-class slope | TCA | 0.008 | 0.082 | 0.097 | 1.008 | 0.859 | 1.183 | 9.23e-01 | 1.00e+00 | 8442 | 6685 |
| African | Full Cohort | ADHD | Within-class slope | NaSSA | 0.067 | 0.152 | 0.443 | 1.070 | 0.794 | 1.441 | 6.58e-01 | 1.00e+00 | 8442 | 6685 |
| African | Full Cohort | ADHD | Within-class slope | SARI | -0.034 | 0.094 | -0.361 | 0.967 | 0.804 | 1.162 | 7.18e-01 | 1.00e+00 | 8442 | 6685 |
| African | Full Cohort | ADHD | Within-class slope | NDRI | 0.006 | 0.094 | 0.060 | 1.006 | 0.836 | 1.210 | 9.52e-01 | 1.00e+00 | 8442 | 6685 |
| African | Full Cohort | ADHD | Interaction (vs SSRI) | SNRI | -0.009 | 0.085 | -0.106 | 0.991 | 0.839 | 1.171 | 9.16e-01 | 1.00e+00 | 8442 | 6685 |
| African | Full Cohort | ADHD | Interaction (vs SSRI) | TCA | -0.022 | 0.093 | -0.236 | 0.978 | 0.814 | 1.175 | 8.13e-01 | 1.00e+00 | 8442 | 6685 |
| African | Full Cohort | ADHD | Interaction (vs SSRI) | NaSSA | 0.037 | 0.158 | 0.236 | 1.038 | 0.762 | 1.414 | 8.13e-01 | 1.00e+00 | 8442 | 6685 |
| African | Full Cohort | ADHD | Interaction (vs SSRI) | SARI | -0.064 | 0.103 | -0.618 | 0.938 | 0.766 | 1.149 | 5.37e-01 | 1.00e+00 | 8442 | 6685 |
| African | Full Cohort | ADHD | Interaction (vs SSRI) | NDRI | -0.024 | 0.102 | -0.238 | 0.976 | 0.798 | 1.193 | 8.12e-01 | 1.00e+00 | 8442 | 6685 |
| African | Full Cohort | ADHD | Joint_Interaction_Test | | | | 0.522 | | | | 9.91e-01 | 1.00e+00 | 8442 | 6685 |
| African | Full Cohort | Migraine | Within-class slope | SSRI | -0.029 | 0.043 | -0.675 | 0.971 | 0.892 | 1.057 | 5.00e-01 | 1.00e+00 | 8442 | 6685 |
| African | Full Cohort | Migraine | Within-class slope | SNRI | 0.017 | 0.066 | 0.257 | 1.017 | 0.894 | 1.157 | 7.98e-01 | 1.00e+00 | 8442 | 6685 |
| African | Full Cohort | Migraine | Within-class slope | TCA | 0.002 | 0.075 | 0.032 | 1.002 | 0.866 | 1.160 | 9.75e-01 | 1.00e+00 | 8442 | 6685 |
| African | Full Cohort | Migraine | Within-class slope | NaSSA | -0.114 | 0.129 | -0.881 | 0.892 | 0.693 | 1.150 | 3.78e-01 | 1.00e+00 | 8442 | 6685 |
| African | Full Cohort | Migraine | Within-class slope | SARI | 0.063 | 0.082 | 0.773 | 1.065 | 0.907 | 1.251 | 4.40e-01 | 1.00e+00 | 8442 | 6685 |
| African | Full Cohort | Migraine | Within-class slope | NDRI | 0.073 | 0.084 | 0.864 | 1.075 | 0.912 | 1.268 | 3.88e-01 | 1.00e+00 | 8442 | 6685 |
| African | Full Cohort | Migraine | Interaction (vs SSRI) | SNRI | 0.046 | 0.076 | 0.608 | 1.047 | 0.903 | 1.215 | 5.43e-01 | 1.00e+00 | 8442 | 6685 |
| African | Full Cohort | Migraine | Interaction (vs SSRI) | TCA | 0.032 | 0.084 | 0.374 | 1.032 | 0.875 | 1.217 | 7.08e-01 | 1.00e+00 | 8442 | 6685 |

| | | | | | | | | | | | | | | |
|---|---|---|---|---|---|---|---|---|---|---|---|---|---|---|
| African | Full Cohort | Migraine | Interaction (vs SSRI) | NaSSA | -0.085 | 0.134 | -0.632 | 0.919 | 0.706 | 1.195 | 5.28e-01 | 1.00e+00 | 8442 | 6685 |
| African | Full Cohort | Migraine | Interaction (vs SSRI) | SARI | 0.093 | 0.090 | 1.027 | 1.097 | 0.919 | 1.309 | 3.04e-01 | 1.00e+00 | 8442 | 6685 |
| African | Full Cohort | Migraine | Interaction (vs SSRI) | NDRI | 0.102 | 0.090 | 1.133 | 1.107 | 0.928 | 1.321 | 2.57e-01 | 1.00e+00 | 8442 | 6685 |
| African | Full Cohort | Migraine | Joint_Interaction_Test | | | | 2.866 | | | | 7.21e-01 | 1.00e+00 | 8442 | 6685 |
| African | Full Cohort | Insomnia | Within-class slope | SSRI | 0.090 | 0.052 | 1.722 | 1.094 | 0.988 | 1.213 | 8.51e-02 | 5.95e-01 | 8442 | 6685 |
| African | Full Cohort | Insomnia | Within-class slope | SNRI | 0.039 | 0.078 | 0.497 | 1.040 | 0.892 | 1.211 | 6.19e-01 | 1.00e+00 | 8442 | 6685 |
| African | Full Cohort | Insomnia | Within-class slope | TCA | 0.102 | 0.083 | 1.229 | 1.108 | 0.941 | 1.304 | 2.19e-01 | 1.00e+00 | 8442 | 6685 |
| African | Full Cohort | Insomnia | Within-class slope | NaSSA | 0.098 | 0.156 | 0.626 | 1.102 | 0.812 | 1.496 | 5.31e-01 | 1.00e+00 | 8442 | 6685 |
| African | Full Cohort | Insomnia | Within-class slope | SARI | 0.391 | 0.097 | 4.018 | 1.479 | 1.222 | 1.790 | 5.87e-05 | 4.11e-04 | 8442 | 6685 |
| African | Full Cohort | Insomnia | Within-class slope | NDRI | 0.044 | 0.103 | 0.427 | 1.045 | 0.854 | 1.278 | 6.70e-01 | 1.00e+00 | 8442 | 6685 |
| African | Full Cohort | Insomnia | Interaction (vs SSRI) | SNRI | -0.051 | 0.090 | -0.569 | 0.950 | 0.796 | 1.134 | 5.69e-01 | 1.00e+00 | 8442 | 6685 |
| African | Full Cohort | Insomnia | Interaction (vs SSRI) | TCA | 0.012 | 0.096 | 0.127 | 1.012 | 0.838 | 1.222 | 8.99e-01 | 1.00e+00 | 8442 | 6685 |
| African | Full Cohort | Insomnia | Interaction (vs SSRI) | NaSSA | 0.007 | 0.162 | 0.045 | 1.007 | 0.733 | 1.384 | 9.64e-01 | 1.00e+00 | 8442 | 6685 |
| African | Full Cohort | Insomnia | Interaction (vs SSRI) | SARI | 0.301 | 0.105 | 2.858 | 1.351 | 1.099 | 1.661 | 4.27e-03 | 2.99e-02 | 8442 | 6685 |
| African | Full Cohort | Insomnia | Interaction (vs SSRI) | NDRI | -0.046 | 0.109 | -0.425 | 0.955 | 0.771 | 1.182 | 6.71e-01 | 1.00e+00 | 8442 | 6685 |
| African | Full Cohort | Insomnia | Joint_Interaction_Test | | | | 10.236 | | | | 6.88e-02 | 4.82e-01 | 8442 | 6685 |
| African | Full Cohort | Depression | Within-class slope | SSRI | 0.091 | 0.051 | 1.786 | 1.096 | 0.991 | 1.211 | 7.40e-02 | 5.18e-01 | 8442 | 6685 |
| African | Full Cohort | Depression | Within-class slope | SNRI | 0.079 | 0.077 | 1.032 | 1.083 | 0.931 | 1.259 | 3.02e-01 | 1.00e+00 | 8442 | 6685 |
| African | Full Cohort | Depression | Within-class slope | TCA | 0.022 | 0.078 | 0.278 | 1.022 | 0.877 | 1.191 | 7.81e-01 | 1.00e+00 | 8442 | 6685 |
| African | Full Cohort | Depression | Within-class slope | NaSSA | -0.209 | 0.154 | -1.360 | 0.811 | 0.600 | 1.097 | 1.74e-01 | 1.00e+00 | 8442 | 6685 |
| African | Full Cohort | Depression | Within-class slope | SARI | 0.121 | 0.095 | 1.267 | 1.128 | 0.936 | 1.360 | 2.05e-01 | 1.00e+00 | 8442 | 6685 |
| African | Full Cohort | Depression | Within-class slope | NDRI | 0.284 | 0.098 | 2.900 | 1.328 | 1.096 | 1.608 | 3.74e-03 | 2.62e-02 | 8442 | 6685 |
| African | Full Cohort | Depression | Interaction (vs SSRI) | SNRI | -0.012 | 0.089 | -0.134 | 0.988 | 0.830 | 1.176 | 8.93e-01 | 1.00e+00 | 8442 | 6685 |
| African | Full Cohort | Depression | Interaction (vs SSRI) | TCA | -0.070 | 0.090 | -0.773 | 0.933 | 0.782 | 1.113 | 4.40e-01 | 1.00e+00 | 8442 | 6685 |
| African | Full Cohort | Depression | Interaction (vs SSRI) | NaSSA | -0.301 | 0.160 | -1.880 | 0.740 | 0.541 | 1.013 | 6.02e-02 | 4.21e-01 | 8442 | 6685 |
| African | Full Cohort | Depression | Interaction (vs SSRI) | SARI | 0.029 | 0.104 | 0.282 | 1.030 | 0.840 | 1.263 | 7.78e-01 | 1.00e+00 | 8442 | 6685 |
| African | Full Cohort | Depression | Interaction (vs SSRI) | NDRI | 0.192 | 0.105 | 1.840 | 1.212 | 0.988 | 1.488 | 6.58e-02 | 4.61e-01 | 8442 | 6685 |
| African | Full Cohort | Depression | Joint_Interaction_Test | | | | 8.999 | | | | 1.09e-01 | 7.64e-01 | 8442 | 6685 |
| African | Full Cohort | Schizophrenia | Within-class slope | SSRI | -0.028 | 0.046 | -0.616 | 0.972 | 0.889 | 1.063 | 5.38e-01 | 1.00e+00 | 8442 | 6685 |
| African | Full Cohort | Schizophrenia | Within-class slope | SNRI | 0.028 | 0.065 | 0.425 | 1.028 | 0.904 | 1.169 | 6.71e-01 | 1.00e+00 | 8442 | 6685 |
| African | Full Cohort | Schizophrenia | Within-class slope | TCA | -0.050 | 0.074 | -0.677 | 0.951 | 0.823 | 1.099 | 4.98e-01 | 1.00e+00 | 8442 | 6685 |
| African | Full Cohort | Schizophrenia | Within-class slope | NaSSA | 0.135 | 0.133 | 1.016 | 1.145 | 0.882 | 1.486 | 3.10e-01 | 1.00e+00 | 8442 | 6685 |
| African | Full Cohort | Schizophrenia | Within-class slope | SARI | 0.059 | 0.087 | 0.685 | 1.061 | 0.895 | 1.258 | 4.93e-01 | 1.00e+00 | 8442 | 6685 |
| African | Full Cohort | Schizophrenia | Within-class slope | NDRI | -0.035 | 0.087 | -0.406 | 0.965 | 0.815 | 1.144 | 6.85e-01 | 1.00e+00 | 8442 | 6685 |
| African | Full Cohort | Schizophrenia | Interaction (vs SSRI) | SNRI | 0.056 | 0.076 | 0.734 | 1.058 | 0.911 | 1.228 | 4.63e-01 | 1.00e+00 | 8442 | 6685 |
| African | Full Cohort | Schizophrenia | Interaction (vs SSRI) | TCA | -0.022 | 0.084 | -0.259 | 0.978 | 0.829 | 1.154 | 7.95e-01 | 1.00e+00 | 8442 | 6685 |
| African | Full Cohort | Schizophrenia | Interaction (vs SSRI) | NaSSA | 0.163 | 0.138 | 1.180 | 1.177 | 0.898 | 1.544 | 2.38e-01 | 1.00e+00 | 8442 | 6685 |
| African | Full Cohort | Schizophrenia | Interaction (vs SSRI) | SARI | 0.088 | 0.095 | 0.927 | 1.092 | 0.907 | 1.314 | 3.54e-01 | 1.00e+00 | 8442 | 6685 |
| African | Full Cohort | Schizophrenia | Interaction (vs SSRI) | NDRI | -0.007 | 0.094 | -0.075 | 0.993 | 0.826 | 1.194 | 9.40e-01 | 1.00e+00 | 8442 | 6685 |
| African | Full Cohort | Schizophrenia | Joint_Interaction_Test | | | | 2.875 | | | | 7.19e-01 | 1.00e+00 | 8442 | 6685 |
| African | Full Cohort | Bipolar Disorder | Within-class slope | SSRI | 0.081 | 0.053 | 1.509 | 1.084 | 0.976 | 1.204 | 1.31e-01 | 9.20e-01 | 8442 | 6685 |
| African | Full Cohort | Bipolar Disorder | Within-class slope | SNRI | 0.153 | 0.078 | 1.953 | 1.165 | 0.999 | 1.358 | 5.09e-02 | 3.56e-01 | 8442 | 6685 |
| African | Full Cohort | Bipolar Disorder | Within-class slope | TCA | -0.048 | 0.083 | -0.571 | 0.954 | 0.810 | 1.123 | 5.68e-01 | 1.00e+00 | 8442 | 6685 |
| African | Full Cohort | Bipolar Disorder | Within-class slope | NaSSA | -0.121 | 0.142 | -0.852 | 0.886 | 0.670 | 1.171 | 3.94e-01 | 1.00e+00 | 8442 | 6685 |
| African | Full Cohort | Bipolar Disorder | Within-class slope | SARI | 0.081 | 0.100 | 0.813 | 1.084 | 0.892 | 1.318 | 4.16e-01 | 1.00e+00 | 8442 | 6685 |
| African | Full Cohort | Bipolar Disorder | Within-class slope | NDRI | 0.072 | 0.104 | 0.697 | 1.075 | 0.877 | 1.318 | 4.86e-01 | 1.00e+00 | 8442 | 6685 |
| African | Full Cohort | Bipolar Disorder | Interaction (vs SSRI) | SNRI | 0.072 | 0.092 | 0.784 | 1.075 | 0.898 | 1.286 | 4.33e-01 | 1.00e+00 | 8442 | 6685 |
| African | Full Cohort | Bipolar Disorder | Interaction (vs SSRI) | TCA | -0.128 | 0.097 | -1.322 | 0.880 | 0.727 | 1.064 | 1.86e-01 | 1.00e+00 | 8442 | 6685 |
| African | Full Cohort | Bipolar Disorder | Interaction (vs SSRI) | NaSSA | -0.202 | 0.149 | -1.352 | 0.817 | 0.610 | 1.095 | 1.76e-01 | 1.00e+00 | 8442 | 6685 |
| African | Full Cohort | Bipolar Disorder | Interaction (vs SSRI) | SARI | 0.000 | 0.109 | 0.003 | 1.000 | 0.808 | 1.238 | 9.98e-01 | 1.00e+00 | 8442 | 6685 |
| African | Full Cohort | Bipolar Disorder | Interaction (vs SSRI) | NDRI | -0.008 | 0.113 | -0.074 | 0.992 | 0.795 | 1.236 | 9.41e-01 | 1.00e+00 | 8442 | 6685 |
| African | Full Cohort | Bipolar Disorder | Joint_Interaction_Test | | | | 5.154 | | | | 3.97e-01 | 1.00e+00 | 8442 | 6685 |
| African | Full Cohort | BMI | Within-class slope | SSRI | -0.058 | 0.047 | -1.247 | 0.943 | 0.860 | 1.034 | 2.12e-01 | 1.00e+00 | 8442 | 6685 |
| African | Full Cohort | BMI | Within-class slope | SNRI | -0.108 | 0.070 | -1.532 | 0.898 | 0.782 | 1.031 | 1.25e-01 | 8.78e-01 | 8442 | 6685 |

| | | | | | | | | | | | | | | |
|---|---|---|---|---|---|---|---|---|---|---|---|---|---|---|
| African | Full Cohort | BMI | Within-class slope | TCA | 0.011 | 0.073 | 0.150 | 1.011 | 0.875 | 1.168 | 8.81e-01 | 1.00e+00 | 8442 | 6685 |
| African | Full Cohort | BMI | Within-class slope | NaSSA | -0.031 | 0.140 | -0.224 | 0.969 | 0.737 | 1.275 | 8.23e-01 | 1.00e+00 | 8442 | 6685 |
| African | Full Cohort | BMI | Within-class slope | SARI | 0.033 | 0.088 | 0.378 | 1.034 | 0.870 | 1.229 | 7.05e-01 | 1.00e+00 | 8442 | 6685 |
| African | Full Cohort | BMI | Within-class slope | NDRI | -0.162 | 0.091 | -1.769 | 0.851 | 0.711 | 1.018 | 7.69e-02 | 5.38e-01 | 8442 | 6685 |
| African | Full Cohort | BMI | Interaction (vs SSRI) | SNRI | -0.049 | 0.081 | -0.610 | 0.952 | 0.812 | 1.115 | 5.42e-01 | 1.00e+00 | 8442 | 6685 |
| African | Full Cohort | BMI | Interaction (vs SSRI) | TCA | 0.069 | 0.085 | 0.821 | 1.072 | 0.908 | 1.265 | 4.12e-01 | 1.00e+00 | 8442 | 6685 |
| African | Full Cohort | BMI | Interaction (vs SSRI) | NaSSA | 0.027 | 0.145 | 0.187 | 1.028 | 0.773 | 1.366 | 8.52e-01 | 1.00e+00 | 8442 | 6685 |
| African | Full Cohort | BMI | Interaction (vs SSRI) | SARI | 0.092 | 0.096 | 0.958 | 1.096 | 0.908 | 1.323 | 3.38e-01 | 1.00e+00 | 8442 | 6685 |
| African | Full Cohort | BMI | Interaction (vs SSRI) | NDRI | -0.103 | 0.098 | -1.052 | 0.902 | 0.744 | 1.093 | 2.93e-01 | 1.00e+00 | 8442 | 6685 |
| African | Full Cohort | BMI | Joint_Interaction_Test | | | | 4.056 | | | | 5.41e-01 | 1.00e+00 | 8442 | 6685 |
| African | Recorded Depression | ADHD | Within-class slope | SSRI | -0.028 | 0.062 | -0.454 | 0.972 | 0.861 | 1.098 | 6.50e-01 | 1.00e+00 | 4849 | 3600 |
| African | Recorded Depression | ADHD | Within-class slope | SNRI | -0.033 | 0.098 | -0.338 | 0.967 | 0.798 | 1.172 | 7.36e-01 | 1.00e+00 | 4849 | 3600 |
| African | Recorded Depression | ADHD | Within-class slope | TCA | -0.031 | 0.120 | -0.258 | 0.970 | 0.767 | 1.226 | 7.96e-01 | 1.00e+00 | 4849 | 3600 |
| African | Recorded Depression | ADHD | Within-class slope | NaSSA | -0.038 | 0.192 | -0.196 | 0.963 | 0.661 | 1.404 | 8.45e-01 | 1.00e+00 | 4849 | 3600 |
| African | Recorded Depression | ADHD | Within-class slope | SARI | -0.017 | 0.144 | -0.116 | 0.983 | 0.741 | 1.304 | 9.08e-01 | 1.00e+00 | 4849 | 3600 |
| African | Recorded Depression | ADHD | Within-class slope | NDRI | -0.035 | 0.121 | -0.287 | 0.966 | 0.761 | 1.225 | 7.74e-01 | 1.00e+00 | 4849 | 3600 |
| African | Recorded Depression | ADHD | Interaction (vs SSRI) | SNRI | -0.005 | 0.110 | -0.046 | 0.995 | 0.802 | 1.235 | 9.63e-01 | 1.00e+00 | 4849 | 3600 |
| African | Recorded Depression | ADHD | Interaction (vs SSRI) | TCA | -0.003 | 0.129 | -0.022 | 0.997 | 0.774 | 1.285 | 9.82e-01 | 1.00e+00 | 4849 | 3600 |
| African | Recorded Depression | ADHD | Interaction (vs SSRI) | NaSSA | -0.010 | 0.199 | -0.048 | 0.990 | 0.670 | 1.464 | 9.61e-01 | 1.00e+00 | 4849 | 3600 |
| African | Recorded Depression | ADHD | Interaction (vs SSRI) | SARI | 0.011 | 0.153 | 0.074 | 1.011 | 0.749 | 1.366 | 9.41e-01 | 1.00e+00 | 4849 | 3600 |
| African | Recorded Depression | ADHD | Interaction (vs SSRI) | NDRI | -0.007 | 0.130 | -0.052 | 0.993 | 0.769 | 1.282 | 9.59e-01 | 1.00e+00 | 4849 | 3600 |
| African | Recorded Depression | ADHD | Joint_Interaction_Test | | | | 0.014 | | | | 1.00e+00 | 1.00e+00 | 4849 | 3600 |
| African | Recorded Depression | Migraine | Within-class slope | SSRI | -0.017 | 0.053 | -0.318 | 0.983 | 0.887 | 1.091 | 7.50e-01 | 1.00e+00 | 4849 | 3600 |
| African | Recorded Depression | Migraine | Within-class slope | SNRI | -0.038 | 0.086 | -0.439 | 0.963 | 0.813 | 1.140 | 6.60e-01 | 1.00e+00 | 4849 | 3600 |
| African | Recorded Depression | Migraine | Within-class slope | TCA | 0.074 | 0.106 | 0.691 | 1.076 | 0.874 | 1.326 | 4.89e-01 | 1.00e+00 | 4849 | 3600 |
| African | Recorded Depression | Migraine | Within-class slope | NaSSA | -0.158 | 0.167 | -0.945 | 0.854 | 0.615 | 1.185 | 3.45e-01 | 1.00e+00 | 4849 | 3600 |
| African | Recorded Depression | Migraine | Within-class slope | SARI | 0.206 | 0.119 | 1.731 | 1.228 | 0.973 | 1.551 | 8.34e-02 | 5.84e-01 | 4849 | 3600 |
| African | Recorded Depression | Migraine | Within-class slope | NDRI | 0.013 | 0.107 | 0.124 | 1.013 | 0.821 | 1.251 | 9.01e-01 | 1.00e+00 | 4849 | 3600 |
| African | Recorded Depression | Migraine | Interaction (vs SSRI) | SNRI | -0.021 | 0.097 | -0.216 | 0.979 | 0.809 | 1.185 | 8.29e-01 | 1.00e+00 | 4849 | 3600 |
| African | Recorded Depression | Migraine | Interaction (vs SSRI) | TCA | 0.090 | 0.116 | 0.782 | 1.095 | 0.873 | 1.373 | 4.34e-01 | 1.00e+00 | 4849 | 3600 |
| African | Recorded Depression | Migraine | Interaction (vs SSRI) | NaSSA | -0.141 | 0.173 | -0.817 | 0.868 | 0.619 | 1.218 | 4.14e-01 | 1.00e+00 | 4849 | 3600 |
| African | Recorded Depression | Migraine | Interaction (vs SSRI) | SARI | 0.223 | 0.128 | 1.741 | 1.249 | 0.972 | 1.605 | 8.17e-02 | 5.72e-01 | 4849 | 3600 |
| African | Recorded Depression | Migraine | Interaction (vs SSRI) | NDRI | 0.030 | 0.115 | 0.263 | 1.031 | 0.823 | 1.290 | 7.93e-01 | 1.00e+00 | 4849 | 3600 |
| African | Recorded Depression | Migraine | Joint_Interaction_Test | | | | 4.829 | | | | 4.37e-01 | 1.00e+00 | 4849 | 3600 |
| African | Recorded Depression | Insomnia | Within-class slope | SSRI | 0.025 | 0.063 | 0.394 | 1.025 | 0.906 | 1.160 | 6.93e-01 | 1.00e+00 | 4849 | 3600 |
| African | Recorded Depression | Insomnia | Within-class slope | SNRI | 0.024 | 0.103 | 0.233 | 1.024 | 0.838 | 1.252 | 8.16e-01 | 1.00e+00 | 4849 | 3600 |
| African | Recorded Depression | Insomnia | Within-class slope | TCA | 0.269 | 0.125 | 2.151 | 1.308 | 1.024 | 1.671 | 3.15e-02 | 2.20e-01 | 4849 | 3600 |
| African | Recorded Depression | Insomnia | Within-class slope | NaSSA | -0.110 | 0.194 | -0.567 | 0.896 | 0.612 | 1.311 | 5.71e-01 | 1.00e+00 | 4849 | 3600 |
| African | Recorded Depression | Insomnia | Within-class slope | SARI | 0.362 | 0.139 | 2.601 | 1.436 | 1.093 | 1.887 | 9.31e-03 | 6.51e-02 | 4849 | 3600 |
| African | Recorded Depression | Insomnia | Within-class slope | NDRI | -0.021 | 0.126 | -0.164 | 0.980 | 0.765 | 1.254 | 8.70e-01 | 1.00e+00 | 4849 | 3600 |
| African | Recorded Depression | Insomnia | Interaction (vs SSRI) | SNRI | -0.001 | 0.116 | -0.008 | 0.999 | 0.796 | 1.254 | 9.94e-01 | 1.00e+00 | 4849 | 3600 |
| African | Recorded Depression | Insomnia | Interaction (vs SSRI) | TCA | 0.244 | 0.136 | 1.795 | 1.276 | 0.978 | 1.665 | 7.26e-02 | 5.08e-01 | 4849 | 3600 |
| African | Recorded Depression | Insomnia | Interaction (vs SSRI) | NaSSA | -0.135 | 0.201 | -0.671 | 0.874 | 0.589 | 1.296 | 5.02e-01 | 1.00e+00 | 4849 | 3600 |
| African | Recorded Depression | Insomnia | Interaction (vs SSRI) | SARI | 0.337 | 0.148 | 2.283 | 1.401 | 1.049 | 1.872 | 2.24e-02 | 1.57e-01 | 4849 | 3600 |
| African | Recorded Depression | Insomnia | Interaction (vs SSRI) | NDRI | -0.045 | 0.134 | -0.339 | 0.956 | 0.735 | 1.243 | 7.35e-01 | 1.00e+00 | 4849 | 3600 |
| African | Recorded Depression | Insomnia | Joint_Interaction_Test | | | | 9.442 | | | | 9.27e-02 | 6.49e-01 | 4849 | 3600 |
| African | Recorded Depression | Depression | Within-class slope | SSRI | 0.041 | 0.062 | 0.671 | 1.042 | 0.924 | 1.176 | 5.02e-01 | 1.00e+00 | 4849 | 3600 |
| African | Recorded Depression | Depression | Within-class slope | SNRI | 0.163 | 0.103 | 1.588 | 1.177 | 0.963 | 1.439 | 1.12e-01 | 7.85e-01 | 4849 | 3600 |
| African | Recorded Depression | Depression | Within-class slope | TCA | 0.036 | 0.117 | 0.304 | 1.036 | 0.824 | 1.303 | 7.61e-01 | 1.00e+00 | 4849 | 3600 |
| African | Recorded Depression | Depression | Within-class slope | NaSSA | -0.265 | 0.202 | -1.313 | 0.767 | 0.516 | 1.139 | 1.89e-01 | 1.00e+00 | 4849 | 3600 |
| African | Recorded Depression | Depression | Within-class slope | SARI | 0.198 | 0.143 | 1.388 | 1.219 | 0.922 | 1.613 | 1.65e-01 | 1.00e+00 | 4849 | 3600 |
| African | Recorded Depression | Depression | Within-class slope | NDRI | 0.171 | 0.118 | 1.447 | 1.187 | 0.941 | 1.497 | 1.48e-01 | 1.00e+00 | 4849 | 3600 |
| African | Recorded Depression | Depression | Interaction (vs SSRI) | SNRI | 0.122 | 0.116 | 1.051 | 1.129 | 0.900 | 1.417 | 2.93e-01 | 1.00e+00 | 4849 | 3600 |
| African | Recorded Depression | Depression | Interaction (vs SSRI) | TCA | -0.006 | 0.127 | -0.046 | 0.994 | 0.774 | 1.276 | 9.64e-01 | 1.00e+00 | 4849 | 3600 |

| | | | | | | | | | | | | | | |
|---|---|---|---|---|---|---|---|---|---|---|---|---|---|---|
| African | Recorded Depression | Depression | Interaction (vs SSRI) | NaSSA | -0.306 | 0.209 | -1.468 | 0.736 | 0.489 | 1.108 | 1.42e-01 | 9.95e-01 | 4849 | 3600 |
| African | Recorded Depression | Depression | Interaction (vs SSRI) | SARI | 0.157 | 0.151 | 1.037 | 1.170 | 0.870 | 1.574 | 3.00e-01 | 1.00e+00 | 4849 | 3600 |
| African | Recorded Depression | Depression | Interaction (vs SSRI) | NDRI | 0.130 | 0.128 | 1.018 | 1.139 | 0.887 | 1.462 | 3.09e-01 | 1.00e+00 | 4849 | 3600 |
| African | Recorded Depression | Depression | Joint_Interaction_Test | | | | 5.922 | | | | 3.14e-01 | 1.00e+00 | 4849 | 3600 |
| African | Recorded Depression | Schizophrenia | Within-class slope | SSRI | -0.031 | 0.055 | -0.575 | 0.969 | 0.871 | 1.078 | 5.65e-01 | 1.00e+00 | 4849 | 3600 |
| African | Recorded Depression | Schizophrenia | Within-class slope | SNRI | 0.059 | 0.086 | 0.685 | 1.061 | 0.896 | 1.256 | 4.93e-01 | 1.00e+00 | 4849 | 3600 |
| African | Recorded Depression | Schizophrenia | Within-class slope | TCA | -0.071 | 0.109 | -0.650 | 0.932 | 0.753 | 1.153 | 5.16e-01 | 1.00e+00 | 4849 | 3600 |
| African | Recorded Depression | Schizophrenia | Within-class slope | NaSSA | 0.043 | 0.171 | 0.249 | 1.044 | 0.746 | 1.459 | 8.03e-01 | 1.00e+00 | 4849 | 3600 |
| African | Recorded Depression | Schizophrenia | Within-class slope | SARI | 0.036 | 0.124 | 0.294 | 1.037 | 0.813 | 1.323 | 7.69e-01 | 1.00e+00 | 4849 | 3600 |
| African | Recorded Depression | Schizophrenia | Within-class slope | NDRI | -0.101 | 0.111 | -0.905 | 0.904 | 0.727 | 1.125 | 3.66e-01 | 1.00e+00 | 4849 | 3600 |
| African | Recorded Depression | Schizophrenia | Interaction (vs SSRI) | SNRI | 0.090 | 0.097 | 0.929 | 1.095 | 0.904 | 1.325 | 3.53e-01 | 1.00e+00 | 4849 | 3600 |
| African | Recorded Depression | Schizophrenia | Interaction (vs SSRI) | TCA | -0.039 | 0.118 | -0.334 | 0.961 | 0.763 | 1.211 | 7.38e-01 | 1.00e+00 | 4849 | 3600 |
| African | Recorded Depression | Schizophrenia | Interaction (vs SSRI) | NaSSA | 0.074 | 0.177 | 0.418 | 1.077 | 0.761 | 1.524 | 6.76e-01 | 1.00e+00 | 4849 | 3600 |
| African | Recorded Depression | Schizophrenia | Interaction (vs SSRI) | SARI | 0.068 | 0.132 | 0.514 | 1.070 | 0.826 | 1.386 | 6.07e-01 | 1.00e+00 | 4849 | 3600 |
| African | Recorded Depression | Schizophrenia | Interaction (vs SSRI) | NDRI | -0.069 | 0.120 | -0.576 | 0.933 | 0.737 | 1.182 | 5.65e-01 | 1.00e+00 | 4849 | 3600 |
| African | Recorded Depression | Schizophrenia | Joint_Interaction_Test | | | | 2.100 | | | | 8.35e-01 | 1.00e+00 | 4849 | 3600 |
| African | Recorded Depression | Bipolar Disorder | Within-class slope | SSRI | 0.074 | 0.065 | 1.140 | 1.077 | 0.948 | 1.222 | 2.54e-01 | 1.00e+00 | 4849 | 3600 |
| African | Recorded Depression | Bipolar Disorder | Within-class slope | SNRI | 0.089 | 0.103 | 0.861 | 1.093 | 0.893 | 1.337 | 3.89e-01 | 1.00e+00 | 4849 | 3600 |
| African | Recorded Depression | Bipolar Disorder | Within-class slope | TCA | -0.153 | 0.128 | -1.194 | 0.858 | 0.667 | 1.103 | 2.32e-01 | 1.00e+00 | 4849 | 3600 |
| African | Recorded Depression | Bipolar Disorder | Within-class slope | NaSSA | -0.075 | 0.187 | -0.403 | 0.928 | 0.644 | 1.337 | 6.87e-01 | 1.00e+00 | 4849 | 3600 |
| African | Recorded Depression | Bipolar Disorder | Within-class slope | SARI | 0.036 | 0.147 | 0.245 | 1.037 | 0.777 | 1.384 | 8.06e-01 | 1.00e+00 | 4849 | 3600 |
| African | Recorded Depression | Bipolar Disorder | Within-class slope | NDRI | 0.057 | 0.129 | 0.441 | 1.059 | 0.822 | 1.364 | 6.59e-01 | 1.00e+00 | 4849 | 3600 |
| African | Recorded Depression | Bipolar Disorder | Interaction (vs SSRI) | SNRI | 0.015 | 0.118 | 0.126 | 1.015 | 0.805 | 1.279 | 8.99e-01 | 1.00e+00 | 4849 | 3600 |
| African | Recorded Depression | Bipolar Disorder | Interaction (vs SSRI) | TCA | -0.227 | 0.141 | -1.611 | 0.797 | 0.605 | 1.050 | 1.07e-01 | 7.50e-01 | 4849 | 3600 |
| African | Recorded Depression | Bipolar Disorder | Interaction (vs SSRI) | NaSSA | -0.149 | 0.195 | -0.763 | 0.862 | 0.588 | 1.263 | 4.45e-01 | 1.00e+00 | 4849 | 3600 |
| African | Recorded Depression | Bipolar Disorder | Interaction (vs SSRI) | SARI | -0.038 | 0.158 | -0.239 | 0.963 | 0.707 | 1.312 | 8.11e-01 | 1.00e+00 | 4849 | 3600 |
| African | Recorded Depression | Bipolar Disorder | Interaction (vs SSRI) | NDRI | -0.017 | 0.140 | -0.120 | 0.983 | 0.747 | 1.294 | 9.05e-01 | 1.00e+00 | 4849 | 3600 |
| African | Recorded Depression | Bipolar Disorder | Joint_Interaction_Test | | | | 3.300 | | | | 6.54e-01 | 1.00e+00 | 4849 | 3600 |
| African | Recorded Depression | BMI | Within-class slope | SSRI | -0.105 | 0.055 | -1.901 | 0.901 | 0.808 | 1.003 | 5.73e-02 | 4.01e-01 | 4849 | 3600 |
| African | Recorded Depression | BMI | Within-class slope | SNRI | -0.184 | 0.094 | -1.957 | 0.832 | 0.692 | 1.000 | 5.03e-02 | 3.52e-01 | 4849 | 3600 |
| African | Recorded Depression | BMI | Within-class slope | TCA | 0.073 | 0.111 | 0.659 | 1.076 | 0.865 | 1.338 | 5.10e-01 | 1.00e+00 | 4849 | 3600 |
| African | Recorded Depression | BMI | Within-class slope | NaSSA | 0.135 | 0.175 | 0.769 | 1.144 | 0.811 | 1.614 | 4.42e-01 | 1.00e+00 | 4849 | 3600 |
| African | Recorded Depression | BMI | Within-class slope | SARI | -0.177 | 0.121 | -1.467 | 0.837 | 0.661 | 1.061 | 1.42e-01 | 9.96e-01 | 4849 | 3600 |
| African | Recorded Depression | BMI | Within-class slope | NDRI | -0.192 | 0.114 | -1.677 | 0.826 | 0.660 | 1.033 | 9.35e-02 | 6.54e-01 | 4849 | 3600 |
| African | Recorded Depression | BMI | Interaction (vs SSRI) | SNRI | -0.079 | 0.104 | -0.761 | 0.924 | 0.753 | 1.133 | 4.47e-01 | 1.00e+00 | 4849 | 3600 |
| African | Recorded Depression | BMI | Interaction (vs SSRI) | TCA | 0.178 | 0.120 | 1.486 | 1.195 | 0.945 | 1.511 | 1.37e-01 | 9.62e-01 | 4849 | 3600 |
| African | Recorded Depression | BMI | Interaction (vs SSRI) | NaSSA | 0.240 | 0.181 | 1.323 | 1.271 | 0.891 | 1.813 | 1.86e-01 | 1.00e+00 | 4849 | 3600 |
| African | Recorded Depression | BMI | Interaction (vs SSRI) | SARI | -0.073 | 0.128 | -0.565 | 0.930 | 0.723 | 1.196 | 5.72e-01 | 1.00e+00 | 4849 | 3600 |
| African | Recorded Depression | BMI | Interaction (vs SSRI) | NDRI | -0.087 | 0.121 | -0.716 | 0.917 | 0.723 | 1.163 | 4.74e-01 | 1.00e+00 | 4849 | 3600 |
| African | Recorded Depression | BMI | Joint_Interaction_Test | | | | 6.639 | | | | 2.49e-01 | 1.00e+00 | 4849 | 3600 |
| African | No Recorded Depression | ADHD | Within-class slope | SSRI | 0.225 | 0.100 | 2.250 | 1.252 | 1.029 | 1.523 | 2.44e-02 | 1.71e-01 | 3593 | 3085 |
| African | No Recorded Depression | ADHD | Within-class slope | SNRI | 0.171 | 0.128 | 1.332 | 1.186 | 0.923 | 1.525 | 1.83e-01 | 1.00e+00 | 3593 | 3085 |
| African | No Recorded Depression | ADHD | Within-class slope | TCA | -0.069 | 0.124 | -0.554 | 0.933 | 0.731 | 1.191 | 5.80e-01 | 1.00e+00 | 3593 | 3085 |
| African | No Recorded Depression | ADHD | Within-class slope | NaSSA | 0.196 | 0.271 | 0.722 | 1.216 | 0.715 | 2.070 | 4.70e-01 | 1.00e+00 | 3593 | 3085 |
| African | No Recorded Depression | ADHD | Within-class slope | SARI | -0.148 | 0.145 | -1.017 | 0.863 | 0.649 | 1.147 | 3.09e-01 | 1.00e+00 | 3593 | 3085 |
| African | No Recorded Depression | ADHD | Within-class slope | NDRI | 0.389 | 0.195 | 1.993 | 1.475 | 1.006 | 2.162 | 4.63e-02 | 3.24e-01 | 3593 | 3085 |
| African | No Recorded Depression | ADHD | Interaction (vs SSRI) | SNRI | -0.054 | 0.158 | -0.342 | 0.947 | 0.694 | 1.292 | 7.32e-01 | 1.00e+00 | 3593 | 3085 |
| African | No Recorded Depression | ADHD | Interaction (vs SSRI) | TCA | -0.294 | 0.160 | -1.836 | 0.745 | 0.545 | 1.020 | 6.64e-02 | 4.65e-01 | 3593 | 3085 |
| African | No Recorded Depression | ADHD | Interaction (vs SSRI) | NaSSA | -0.029 | 0.282 | -0.104 | 0.971 | 0.559 | 1.687 | 9.17e-01 | 1.00e+00 | 3593 | 3085 |
| African | No Recorded Depression | ADHD | Interaction (vs SSRI) | SARI | -0.373 | 0.171 | -2.180 | 0.689 | 0.493 | 0.963 | 2.92e-02 | 2.05e-01 | 3593 | 3085 |
| African | No Recorded Depression | ADHD | Interaction (vs SSRI) | NDRI | 0.164 | 0.215 | 0.761 | 1.178 | 0.773 | 1.795 | 4.47e-01 | 1.00e+00 | 3593 | 3085 |
| African | No Recorded Depression | ADHD | Joint_Interaction_Test | | | | 9.222 | | | | 1.01e-01 | 7.04e-01 | 3593 | 3085 |
| African | No Recorded Depression | Migraine | Within-class slope | SSRI | -0.077 | 0.080 | -0.967 | 0.926 | 0.792 | 1.082 | 3.33e-01 | 1.00e+00 | 3593 | 3085 |
| African | No Recorded Depression | Migraine | Within-class slope | SNRI | 0.113 | 0.115 | 0.979 | 1.119 | 0.893 | 1.403 | 3.28e-01 | 1.00e+00 | 3593 | 3085 |

| | | | | | | | | | | | | | | |
|---|---|---|---|---|---|---|---|---|---|---|---|---|---|---|
| African | No Recorded Depression | Migraine | Within-class slope | TCA | -0.033 | 0.116 | -0.287 | 0.967 | 0.771 | 1.213 | 7.74e-01 | 1.00e+00 | 3593 | 3085 |
| African | No Recorded Depression | Migraine | Within-class slope | NaSSA | -0.146 | 0.229 | -0.639 | 0.864 | 0.552 | 1.353 | 5.23e-01 | 1.00e+00 | 3593 | 3085 |
| African | No Recorded Depression | Migraine | Within-class slope | SARI | 0.047 | 0.139 | 0.338 | 1.048 | 0.798 | 1.376 | 7.35e-01 | 1.00e+00 | 3593 | 3085 |
| African | No Recorded Depression | Migraine | Within-class slope | NDRI | 0.202 | 0.156 | 1.294 | 1.224 | 0.901 | 1.663 | 1.96e-01 | 1.00e+00 | 3593 | 3085 |
| African | No Recorded Depression | Migraine | Interaction (vs SSRI) | SNRI | 0.190 | 0.138 | 1.373 | 1.209 | 0.922 | 1.586 | 1.70e-01 | 1.00e+00 | 3593 | 3085 |
| African | No Recorded Depression | Migraine | Interaction (vs SSRI) | TCA | 0.044 | 0.140 | 0.313 | 1.045 | 0.794 | 1.376 | 7.54e-01 | 1.00e+00 | 3593 | 3085 |
| African | No Recorded Depression | Migraine | Interaction (vs SSRI) | NaSSA | -0.069 | 0.236 | -0.293 | 0.933 | 0.588 | 1.482 | 7.70e-01 | 1.00e+00 | 3593 | 3085 |
| African | No Recorded Depression | Migraine | Interaction (vs SSRI) | SARI | 0.124 | 0.158 | 0.785 | 1.132 | 0.830 | 1.544 | 4.33e-01 | 1.00e+00 | 3593 | 3085 |
| African | No Recorded Depression | Migraine | Interaction (vs SSRI) | NDRI | 0.279 | 0.166 | 1.679 | 1.322 | 0.954 | 1.832 | 9.32e-02 | 6.52e-01 | 3593 | 3085 |
| African | No Recorded Depression | Migraine | Joint_Interaction_Test | | | | 4.585 | | | | 4.69e-01 | 1.00e+00 | 3593 | 3085 |
| African | No Recorded Depression | Insomnia | Within-class slope | SSRI | 0.120 | 0.107 | 1.122 | 1.128 | 0.914 | 1.391 | 2.62e-01 | 1.00e+00 | 3593 | 3085 |
| African | No Recorded Depression | Insomnia | Within-class slope | SNRI | 0.100 | 0.136 | 0.735 | 1.105 | 0.847 | 1.441 | 4.62e-01 | 1.00e+00 | 3593 | 3085 |
| African | No Recorded Depression | Insomnia | Within-class slope | TCA | -0.077 | 0.130 | -0.596 | 0.926 | 0.718 | 1.193 | 5.51e-01 | 1.00e+00 | 3593 | 3085 |
| African | No Recorded Depression | Insomnia | Within-class slope | NaSSA | 0.545 | 0.278 | 1.958 | 1.724 | 0.999 | 2.975 | 5.03e-02 | 3.52e-01 | 3593 | 3085 |
| African | No Recorded Depression | Insomnia | Within-class slope | SARI | 0.406 | 0.168 | 2.420 | 1.501 | 1.080 | 2.085 | 1.55e-02 | 1.09e-01 | 3593 | 3085 |
| African | No Recorded Depression | Insomnia | Within-class slope | NDRI | 0.266 | 0.218 | 1.216 | 1.304 | 0.850 | 2.001 | 2.24e-01 | 1.00e+00 | 3593 | 3085 |
| African | No Recorded Depression | Insomnia | Interaction (vs SSRI) | SNRI | -0.021 | 0.170 | -0.121 | 0.980 | 0.702 | 1.367 | 9.04e-01 | 1.00e+00 | 3593 | 3085 |
| African | No Recorded Depression | Insomnia | Interaction (vs SSRI) | TCA | -0.197 | 0.166 | -1.186 | 0.821 | 0.592 | 1.137 | 2.36e-01 | 1.00e+00 | 3593 | 3085 |
| African | No Recorded Depression | Insomnia | Interaction (vs SSRI) | NaSSA | 0.425 | 0.294 | 1.443 | 1.529 | 0.859 | 2.722 | 1.49e-01 | 1.00e+00 | 3593 | 3085 |
| African | No Recorded Depression | Insomnia | Interaction (vs SSRI) | SARI | 0.286 | 0.188 | 1.518 | 1.331 | 0.920 | 1.925 | 1.29e-01 | 9.04e-01 | 3593 | 3085 |
| African | No Recorded Depression | Insomnia | Interaction (vs SSRI) | NDRI | 0.145 | 0.229 | 0.636 | 1.157 | 0.739 | 1.810 | 5.25e-01 | 1.00e+00 | 3593 | 3085 |
| African | No Recorded Depression | Insomnia | Joint_Interaction_Test | | | | 7.959 | | | | 1.59e-01 | 1.00e+00 | 3593 | 3085 |
| African | No Recorded Depression | Depression | Within-class slope | SSRI | 0.212 | 0.098 | 2.161 | 1.236 | 1.020 | 1.499 | 3.07e-02 | 2.15e-01 | 3593 | 3085 |
| African | No Recorded Depression | Depression | Within-class slope | SNRI | -0.065 | 0.125 | -0.524 | 0.937 | 0.733 | 1.196 | 6.00e-01 | 1.00e+00 | 3593 | 3085 |
| African | No Recorded Depression | Depression | Within-class slope | TCA | -0.149 | 0.116 | -1.283 | 0.862 | 0.686 | 1.082 | 2.00e-01 | 1.00e+00 | 3593 | 3085 |
| African | No Recorded Depression | Depression | Within-class slope | NaSSA | -0.217 | 0.250 | -0.866 | 0.805 | 0.493 | 1.315 | 3.87e-01 | 1.00e+00 | 3593 | 3085 |
| African | No Recorded Depression | Depression | Within-class slope | SARI | 0.041 | 0.155 | 0.264 | 1.042 | 0.769 | 1.410 | 7.92e-01 | 1.00e+00 | 3593 | 3085 |
| African | No Recorded Depression | Depression | Within-class slope | NDRI | 0.510 | 0.189 | 2.702 | 1.666 | 1.150 | 2.412 | 6.88e-03 | 4.82e-02 | 3593 | 3085 |
| African | No Recorded Depression | Depression | Interaction (vs SSRI) | SNRI | -0.278 | 0.157 | -1.765 | 0.758 | 0.557 | 1.031 | 7.75e-02 | 5.43e-01 | 3593 | 3085 |
| African | No Recorded Depression | Depression | Interaction (vs SSRI) | TCA | -0.361 | 0.151 | -2.390 | 0.697 | 0.518 | 0.937 | 1.69e-02 | 1.18e-01 | 3593 | 3085 |
| African | No Recorded Depression | Depression | Interaction (vs SSRI) | NaSSA | -0.429 | 0.260 | -1.651 | 0.651 | 0.391 | 1.084 | 9.87e-02 | 6.91e-01 | 3593 | 3085 |
| African | No Recorded Depression | Depression | Interaction (vs SSRI) | SARI | -0.171 | 0.178 | -0.962 | 0.842 | 0.594 | 1.195 | 3.36e-01 | 1.00e+00 | 3593 | 3085 |
| African | No Recorded Depression | Depression | Interaction (vs SSRI) | NDRI | 0.298 | 0.204 | 1.459 | 1.347 | 0.903 | 2.010 | 1.45e-01 | 1.00e+00 | 3593 | 3085 |
| African | No Recorded Depression | Depression | Joint_Interaction_Test | | | | 13.228 | | | | 2.13e-02 | 1.49e-01 | 3593 | 3085 |
| African | No Recorded Depression | Schizophrenia | Within-class slope | SSRI | 0.076 | 0.095 | 0.803 | 1.079 | 0.896 | 1.301 | 4.22e-01 | 1.00e+00 | 3593 | 3085 |
| African | No Recorded Depression | Schizophrenia | Within-class slope | SNRI | -0.057 | 0.122 | -0.468 | 0.945 | 0.744 | 1.199 | 6.40e-01 | 1.00e+00 | 3593 | 3085 |
| African | No Recorded Depression | Schizophrenia | Within-class slope | TCA | -0.047 | 0.122 | -0.383 | 0.954 | 0.752 | 1.212 | 7.01e-01 | 1.00e+00 | 3593 | 3085 |
| African | No Recorded Depression | Schizophrenia | Within-class slope | NaSSA | 0.265 | 0.227 | 1.168 | 1.303 | 0.836 | 2.033 | 2.43e-01 | 1.00e+00 | 3593 | 3085 |
| African | No Recorded Depression | Schizophrenia | Within-class slope | SARI | 0.130 | 0.151 | 0.859 | 1.139 | 0.847 | 1.531 | 3.90e-01 | 1.00e+00 | 3593 | 3085 |
| African | No Recorded Depression | Schizophrenia | Within-class slope | NDRI | 0.213 | 0.180 | 1.179 | 1.237 | 0.869 | 1.762 | 2.38e-01 | 1.00e+00 | 3593 | 3085 |
| African | No Recorded Depression | Schizophrenia | Interaction (vs SSRI) | SNRI | -0.133 | 0.154 | -0.868 | 0.875 | 0.648 | 1.182 | 3.85e-01 | 1.00e+00 | 3593 | 3085 |
| African | No Recorded Depression | Schizophrenia | Interaction (vs SSRI) | TCA | -0.123 | 0.152 | -0.812 | 0.884 | 0.657 | 1.190 | 4.17e-01 | 1.00e+00 | 3593 | 3085 |
| African | No Recorded Depression | Schizophrenia | Interaction (vs SSRI) | NaSSA | 0.188 | 0.238 | 0.793 | 1.207 | 0.758 | 1.924 | 4.28e-01 | 1.00e+00 | 3593 | 3085 |
| African | No Recorded Depression | Schizophrenia | Interaction (vs SSRI) | SARI | 0.053 | 0.171 | 0.313 | 1.055 | 0.755 | 1.474 | 7.55e-01 | 1.00e+00 | 3593 | 3085 |
| African | No Recorded Depression | Schizophrenia | Interaction (vs SSRI) | NDRI | 0.136 | 0.194 | 0.701 | 1.146 | 0.783 | 1.677 | 4.83e-01 | 1.00e+00 | 3593 | 3085 |
| African | No Recorded Depression | Schizophrenia | Joint_Interaction_Test | | | | 3.740 | | | | 5.87e-01 | 1.00e+00 | 3593 | 3085 |
| African | No Recorded Depression | Bipolar Disorder | Within-class slope | SSRI | 0.110 | 0.108 | 1.017 | 1.116 | 0.903 | 1.381 | 3.09e-01 | 1.00e+00 | 3593 | 3085 |
| African | No Recorded Depression | Bipolar Disorder | Within-class slope | SNRI | 0.232 | 0.135 | 1.718 | 1.261 | 0.968 | 1.644 | 8.58e-02 | 6.01e-01 | 3593 | 3085 |
| African | No Recorded Depression | Bipolar Disorder | Within-class slope | TCA | 0.011 | 0.124 | 0.085 | 1.011 | 0.793 | 1.288 | 9.32e-01 | 1.00e+00 | 3593 | 3085 |
| African | No Recorded Depression | Bipolar Disorder | Within-class slope | NaSSA | -0.240 | 0.239 | -1.005 | 0.787 | 0.493 | 1.256 | 3.15e-01 | 1.00e+00 | 3593 | 3085 |
| African | No Recorded Depression | Bipolar Disorder | Within-class slope | SARI | 0.076 | 0.149 | 0.508 | 1.079 | 0.805 | 1.445 | 6.11e-01 | 1.00e+00 | 3593 | 3085 |
| African | No Recorded Depression | Bipolar Disorder | Within-class slope | NDRI | 0.141 | 0.218 | 0.650 | 1.152 | 0.752 | 1.765 | 5.16e-01 | 1.00e+00 | 3593 | 3085 |
| African | No Recorded Depression | Bipolar Disorder | Interaction (vs SSRI) | SNRI | 0.122 | 0.171 | 0.712 | 1.130 | 0.807 | 1.581 | 4.76e-01 | 1.00e+00 | 3593 | 3085 |
| African | No Recorded Depression | Bipolar Disorder | Interaction (vs SSRI) | TCA | -0.100 | 0.163 | -0.611 | 0.905 | 0.658 | 1.246 | 5.41e-01 | 1.00e+00 | 3593 | 3085 |

| | | | | | | | | | | | | | | |
|---|---|---|---|---|---|---|---|---|---|---|---|---|---|---|
| African | No Recorded Depression | Bipolar Disorder | Interaction (vs SSRI) | NaSSA | -0.350 | 0.251 | -1.395 | 0.705 | 0.431 | 1.152 | 1.63e-01 | 1.00e+00 | 3593 | 3085 |
| African | No Recorded Depression | Bipolar Disorder | Interaction (vs SSRI) | SARI | -0.034 | 0.174 | -0.198 | 0.966 | 0.688 | 1.358 | 8.43e-01 | 1.00e+00 | 3593 | 3085 |
| African | No Recorded Depression | Bipolar Disorder | Interaction (vs SSRI) | NDRI | 0.031 | 0.236 | 0.133 | 1.032 | 0.650 | 1.637 | 8.94e-01 | 1.00e+00 | 3593 | 3085 |
| African | No Recorded Depression | Bipolar Disorder | Joint_Interaction_Test | | | | 3.580 | | | | 6.11e-01 | 1.00e+00 | 3593 | 3085 |
| African | No Recorded Depression | BMI | Within-class slope | SSRI | 0.079 | 0.099 | 0.801 | 1.082 | 0.892 | 1.313 | 4.23e-01 | 1.00e+00 | 3593 | 3085 |
| African | No Recorded Depression | BMI | Within-class slope | SNRI | 0.055 | 0.122 | 0.451 | 1.057 | 0.832 | 1.342 | 6.52e-01 | 1.00e+00 | 3593 | 3085 |
| African | No Recorded Depression | BMI | Within-class slope | TCA | -0.173 | 0.112 | -1.543 | 0.841 | 0.676 | 1.048 | 1.23e-01 | 8.60e-01 | 3593 | 3085 |
| African | No Recorded Depression | BMI | Within-class slope | NaSSA | -0.322 | 0.247 | -1.303 | 0.725 | 0.446 | 1.176 | 1.93e-01 | 1.00e+00 | 3593 | 3085 |
| African | No Recorded Depression | BMI | Within-class slope | SARI | 0.236 | 0.150 | 1.573 | 1.267 | 0.943 | 1.701 | 1.16e-01 | 8.10e-01 | 3593 | 3085 |
| African | No Recorded Depression | BMI | Within-class slope | NDRI | -0.105 | 0.176 | -0.598 | 0.900 | 0.637 | 1.272 | 5.50e-01 | 1.00e+00 | 3593 | 3085 |
| African | No Recorded Depression | BMI | Interaction (vs SSRI) | SNRI | -0.024 | 0.157 | -0.152 | 0.976 | 0.718 | 1.328 | 8.79e-01 | 1.00e+00 | 3593 | 3085 |
| African | No Recorded Depression | BMI | Interaction (vs SSRI) | TCA | -0.252 | 0.148 | -1.699 | 0.778 | 0.582 | 1.039 | 8.93e-02 | 6.25e-01 | 3593 | 3085 |
| African | No Recorded Depression | BMI | Interaction (vs SSRI) | NaSSA | -0.401 | 0.258 | -1.555 | 0.670 | 0.404 | 1.110 | 1.20e-01 | 8.39e-01 | 3593 | 3085 |
| African | No Recorded Depression | BMI | Interaction (vs SSRI) | SARI | 0.158 | 0.175 | 0.901 | 1.171 | 0.831 | 1.649 | 3.68e-01 | 1.00e+00 | 3593 | 3085 |
| African | No Recorded Depression | BMI | Interaction (vs SSRI) | NDRI | -0.184 | 0.196 | -0.942 | 0.832 | 0.567 | 1.220 | 3.46e-01 | 1.00e+00 | 3593 | 3085 |
| African | No Recorded Depression | BMI | Joint_Interaction_Test | | | | 8.436 | | | | 1.34e-01 | 9.36e-01 | 3593 | 3085 |
| African | Recorded Anxiety | ADHD | Within-class slope | SSRI | 0.026 | 0.060 | 0.428 | 1.026 | 0.912 | 1.154 | 6.68e-01 | 1.00e+00 | 5189 | 3903 |
| African | Recorded Anxiety | ADHD | Within-class slope | SNRI | -0.055 | 0.093 | -0.589 | 0.946 | 0.788 | 1.136 | 5.56e-01 | 1.00e+00 | 5189 | 3903 |
| African | Recorded Anxiety | ADHD | Within-class slope | TCA | -0.053 | 0.109 | -0.488 | 0.948 | 0.766 | 1.174 | 6.25e-01 | 1.00e+00 | 5189 | 3903 |
| African | Recorded Anxiety | ADHD | Within-class slope | NaSSA | 0.025 | 0.197 | 0.127 | 1.025 | 0.697 | 1.509 | 8.99e-01 | 1.00e+00 | 5189 | 3903 |
| African | Recorded Anxiety | ADHD | Within-class slope | SARI | -0.201 | 0.132 | -1.515 | 0.818 | 0.631 | 1.061 | 1.30e-01 | 9.08e-01 | 5189 | 3903 |
| African | Recorded Anxiety | ADHD | Within-class slope | NDRI | 0.016 | 0.122 | 0.134 | 1.016 | 0.801 | 1.290 | 8.94e-01 | 1.00e+00 | 5189 | 3903 |
| African | Recorded Anxiety | ADHD | Interaction (vs SSRI) | SNRI | -0.081 | 0.106 | -0.759 | 0.923 | 0.749 | 1.136 | 4.48e-01 | 1.00e+00 | 5189 | 3903 |
| African | Recorded Anxiety | ADHD | Interaction (vs SSRI) | TCA | -0.079 | 0.120 | -0.660 | 0.924 | 0.731 | 1.168 | 5.10e-01 | 1.00e+00 | 5189 | 3903 |
| African | Recorded Anxiety | ADHD | Interaction (vs SSRI) | NaSSA | -0.001 | 0.203 | -0.003 | 0.999 | 0.671 | 1.489 | 9.98e-01 | 1.00e+00 | 5189 | 3903 |
| African | Recorded Anxiety | ADHD | Interaction (vs SSRI) | SARI | -0.226 | 0.141 | -1.605 | 0.798 | 0.605 | 1.051 | 1.08e-01 | 7.59e-01 | 5189 | 3903 |
| African | Recorded Anxiety | ADHD | Interaction (vs SSRI) | NDRI | -0.009 | 0.129 | -0.073 | 0.991 | 0.770 | 1.275 | 9.42e-01 | 1.00e+00 | 5189 | 3903 |
| African | Recorded Anxiety | ADHD | Joint_Interaction_Test | | | | 2.977 | | | | 7.03e-01 | 1.00e+00 | 5189 | 3903 |
| African | Recorded Anxiety | Migraine | Within-class slope | SSRI | -0.009 | 0.052 | -0.171 | 0.991 | 0.896 | 1.097 | 8.64e-01 | 1.00e+00 | 5189 | 3903 |
| African | Recorded Anxiety | Migraine | Within-class slope | SNRI | 0.048 | 0.081 | 0.593 | 1.049 | 0.895 | 1.231 | 5.53e-01 | 1.00e+00 | 5189 | 3903 |
| African | Recorded Anxiety | Migraine | Within-class slope | TCA | 0.063 | 0.098 | 0.645 | 1.065 | 0.879 | 1.291 | 5.19e-01 | 1.00e+00 | 5189 | 3903 |
| African | Recorded Anxiety | Migraine | Within-class slope | NaSSA | -0.232 | 0.168 | -1.376 | 0.793 | 0.570 | 1.103 | 1.69e-01 | 1.00e+00 | 5189 | 3903 |
| African | Recorded Anxiety | Migraine | Within-class slope | SARI | 0.212 | 0.114 | 1.859 | 1.236 | 0.989 | 1.547 | 6.30e-02 | 4.41e-01 | 5189 | 3903 |
| African | Recorded Anxiety | Migraine | Within-class slope | NDRI | -0.061 | 0.109 | -0.556 | 0.941 | 0.760 | 1.166 | 5.79e-01 | 1.00e+00 | 5189 | 3903 |
| African | Recorded Anxiety | Migraine | Interaction (vs SSRI) | SNRI | 0.057 | 0.094 | 0.609 | 1.059 | 0.881 | 1.272 | 5.42e-01 | 1.00e+00 | 5189 | 3903 |
| African | Recorded Anxiety | Migraine | Interaction (vs SSRI) | TCA | 0.072 | 0.108 | 0.668 | 1.075 | 0.870 | 1.327 | 5.04e-01 | 1.00e+00 | 5189 | 3903 |
| African | Recorded Anxiety | Migraine | Interaction (vs SSRI) | NaSSA | -0.223 | 0.174 | -1.281 | 0.800 | 0.569 | 1.125 | 2.00e-01 | 1.00e+00 | 5189 | 3903 |
| African | Recorded Anxiety | Migraine | Interaction (vs SSRI) | SARI | 0.221 | 0.123 | 1.799 | 1.247 | 0.980 | 1.587 | 7.20e-02 | 5.04e-01 | 5189 | 3903 |
| African | Recorded Anxiety | Migraine | Interaction (vs SSRI) | NDRI | -0.052 | 0.115 | -0.452 | 0.949 | 0.758 | 1.189 | 6.52e-01 | 1.00e+00 | 5189 | 3903 |
| African | Recorded Anxiety | Migraine | Joint_Interaction_Test | | | | 6.334 | | | | 2.75e-01 | 1.00e+00 | 5189 | 3903 |
| African | Recorded Anxiety | Insomnia | Within-class slope | SSRI | 0.137 | 0.061 | 2.243 | 1.146 | 1.017 | 1.292 | 2.49e-02 | 1.74e-01 | 5189 | 3903 |
| African | Recorded Anxiety | Insomnia | Within-class slope | SNRI | 0.007 | 0.097 | 0.073 | 1.007 | 0.833 | 1.217 | 9.41e-01 | 1.00e+00 | 5189 | 3903 |
| African | Recorded Anxiety | Insomnia | Within-class slope | TCA | 0.211 | 0.112 | 1.875 | 1.234 | 0.991 | 1.538 | 6.07e-02 | 4.25e-01 | 5189 | 3903 |
| African | Recorded Anxiety | Insomnia | Within-class slope | NaSSA | -0.157 | 0.195 | -0.802 | 0.855 | 0.583 | 1.254 | 4.22e-01 | 1.00e+00 | 5189 | 3903 |
| African | Recorded Anxiety | Insomnia | Within-class slope | SARI | 0.504 | 0.129 | 3.896 | 1.655 | 1.284 | 2.132 | 9.77e-05 | 6.84e-04 | 5189 | 3903 |
| African | Recorded Anxiety | Insomnia | Within-class slope | NDRI | 0.105 | 0.127 | 0.829 | 1.111 | 0.866 | 1.425 | 4.07e-01 | 1.00e+00 | 5189 | 3903 |
| African | Recorded Anxiety | Insomnia | Interaction (vs SSRI) | SNRI | -0.129 | 0.110 | -1.175 | 0.879 | 0.708 | 1.090 | 2.40e-01 | 1.00e+00 | 5189 | 3903 |
| African | Recorded Anxiety | Insomnia | Interaction (vs SSRI) | TCA | 0.074 | 0.124 | 0.598 | 1.077 | 0.845 | 1.372 | 5.50e-01 | 1.00e+00 | 5189 | 3903 |
| African | Recorded Anxiety | Insomnia | Interaction (vs SSRI) | NaSSA | -0.293 | 0.202 | -1.452 | 0.746 | 0.502 | 1.108 | 1.47e-01 | 1.00e+00 | 5189 | 3903 |
| African | Recorded Anxiety | Insomnia | Interaction (vs SSRI) | SARI | 0.367 | 0.136 | 2.694 | 1.443 | 1.105 | 1.885 | 7.05e-03 | 4.93e-02 | 5189 | 3903 |
| African | Recorded Anxiety | Insomnia | Interaction (vs SSRI) | NDRI | -0.031 | 0.133 | -0.236 | 0.969 | 0.747 | 1.257 | 8.14e-01 | 1.00e+00 | 5189 | 3903 |
| African | Recorded Anxiety | Insomnia | Joint_Interaction_Test | | | | 13.093 | | | | 2.25e-02 | 1.58e-01 | 5189 | 3903 |
| African | Recorded Anxiety | Depression | Within-class slope | SSRI | 0.113 | 0.060 | 1.885 | 1.119 | 0.995 | 1.259 | 5.95e-02 | 4.16e-01 | 5189 | 3903 |
| African | Recorded Anxiety | Depression | Within-class slope | SNRI | 0.136 | 0.099 | 1.374 | 1.145 | 0.944 | 1.389 | 1.69e-01 | 1.00e+00 | 5189 | 3903 |

| | | | | | | | | | | | | | |
|---|---|---|---|---|---|---|---|---|---|---|---|---|---|
| African | Recorded Anxiety | Depression | Within-class slope | TCA | -0.047 | 0.108 | -0.429 | 0.955 | 0.772 | 1.180 | 6.68e-01 | 1.00e+00 | 5189 | 3903 |
| African | Recorded Anxiety | Depression | Within-class slope | NaSSA | -0.427 | 0.207 | -2.058 | 0.653 | 0.435 | 0.980 | 3.96e-02 | 2.77e-01 | 5189 | 3903 |
| African | Recorded Anxiety | Depression | Within-class slope | SARI | 0.130 | 0.130 | 0.996 | 1.138 | 0.882 | 1.469 | 3.19e-01 | 1.00e+00 | 5189 | 3903 |
| African | Recorded Anxiety | Depression | Within-class slope | NDRI | 0.350 | 0.127 | 2.756 | 1.418 | 1.106 | 1.819 | 5.85e-03 | 4.09e-02 | 5189 | 3903 |
| African | Recorded Anxiety | Depression | Interaction (vs SSRI) | SNRI | 0.023 | 0.112 | 0.203 | 1.023 | 0.821 | 1.274 | 8.39e-01 | 1.00e+00 | 5189 | 3903 |
| African | Recorded Anxiety | Depression | Interaction (vs SSRI) | TCA | -0.159 | 0.119 | -1.341 | 0.853 | 0.676 | 1.076 | 1.80e-01 | 1.00e+00 | 5189 | 3903 |
| African | Recorded Anxiety | Depression | Interaction (vs SSRI) | NaSSA | -0.540 | 0.214 | -2.520 | 0.583 | 0.383 | 0.887 | 1.17e-02 | 8.20e-02 | 5189 | 3903 |
| African | Recorded Anxiety | Depression | Interaction (vs SSRI) | SARI | 0.017 | 0.139 | 0.121 | 1.017 | 0.775 | 1.334 | 9.03e-01 | 1.00e+00 | 5189 | 3903 |
| African | Recorded Anxiety | Depression | Interaction (vs SSRI) | NDRI | 0.237 | 0.133 | 1.784 | 1.267 | 0.977 | 1.644 | 7.44e-02 | 5.21e-01 | 5189 | 3903 |
| African | Recorded Anxiety | Depression | Joint_Interaction_Test | | | | 13.023 | | | | 2.32e-02 | 1.62e-01 | 5189 | 3903 |
| African | Recorded Anxiety | Schizophrenia | Within-class slope | SSRI | -0.054 | 0.055 | -0.987 | 0.948 | 0.851 | 1.055 | 3.24e-01 | 1.00e+00 | 5189 | 3903 |
| African | Recorded Anxiety | Schizophrenia | Within-class slope | SNRI | 0.027 | 0.083 | 0.323 | 1.027 | 0.873 | 1.208 | 7.47e-01 | 1.00e+00 | 5189 | 3903 |
| African | Recorded Anxiety | Schizophrenia | Within-class slope | TCA | -0.127 | 0.100 | -1.272 | 0.880 | 0.723 | 1.071 | 2.03e-01 | 1.00e+00 | 5189 | 3903 |
| African | Recorded Anxiety | Schizophrenia | Within-class slope | NaSSA | -0.262 | 0.186 | -1.410 | 0.770 | 0.535 | 1.107 | 1.59e-01 | 1.00e+00 | 5189 | 3903 |
| African | Recorded Anxiety | Schizophrenia | Within-class slope | SARI | 0.030 | 0.114 | 0.264 | 1.030 | 0.825 | 1.288 | 7.92e-01 | 1.00e+00 | 5189 | 3903 |
| African | Recorded Anxiety | Schizophrenia | Within-class slope | NDRI | -0.005 | 0.117 | -0.046 | 0.995 | 0.791 | 1.251 | 9.63e-01 | 1.00e+00 | 5189 | 3903 |
| African | Recorded Anxiety | Schizophrenia | Interaction (vs SSRI) | SNRI | 0.081 | 0.095 | 0.851 | 1.084 | 0.900 | 1.305 | 3.95e-01 | 1.00e+00 | 5189 | 3903 |
| African | Recorded Anxiety | Schizophrenia | Interaction (vs SSRI) | TCA | -0.074 | 0.110 | -0.669 | 0.929 | 0.749 | 1.152 | 5.03e-01 | 1.00e+00 | 5189 | 3903 |
| African | Recorded Anxiety | Schizophrenia | Interaction (vs SSRI) | NaSSA | -0.208 | 0.192 | -1.085 | 0.812 | 0.558 | 1.182 | 2.78e-01 | 1.00e+00 | 5189 | 3903 |
| African | Recorded Anxiety | Schizophrenia | Interaction (vs SSRI) | SARI | 0.084 | 0.122 | 0.686 | 1.087 | 0.856 | 1.382 | 4.93e-01 | 1.00e+00 | 5189 | 3903 |
| African | Recorded Anxiety | Schizophrenia | Interaction (vs SSRI) | NDRI | 0.049 | 0.124 | 0.391 | 1.050 | 0.823 | 1.339 | 6.96e-01 | 1.00e+00 | 5189 | 3903 |
| African | Recorded Anxiety | Schizophrenia | Joint_Interaction_Test | | | | 3.628 | | | | 6.04e-01 | 1.00e+00 | 5189 | 3903 |
| African | Recorded Anxiety | Bipolar Disorder | Within-class slope | SSRI | 0.061 | 0.063 | 0.968 | 1.063 | 0.939 | 1.203 | 3.33e-01 | 1.00e+00 | 5189 | 3903 |
| African | Recorded Anxiety | Bipolar Disorder | Within-class slope | SNRI | 0.126 | 0.099 | 1.273 | 1.134 | 0.934 | 1.376 | 2.03e-01 | 1.00e+00 | 5189 | 3903 |
| African | Recorded Anxiety | Bipolar Disorder | Within-class slope | TCA | -0.206 | 0.112 | -1.838 | 0.814 | 0.653 | 1.014 | 6.60e-02 | 4.62e-01 | 5189 | 3903 |
| African | Recorded Anxiety | Bipolar Disorder | Within-class slope | NaSSA | -0.251 | 0.179 | -1.399 | 0.778 | 0.548 | 1.106 | 1.62e-01 | 1.00e+00 | 5189 | 3903 |
| African | Recorded Anxiety | Bipolar Disorder | Within-class slope | SARI | 0.067 | 0.143 | 0.466 | 1.069 | 0.808 | 1.415 | 6.41e-01 | 1.00e+00 | 5189 | 3903 |
| African | Recorded Anxiety | Bipolar Disorder | Within-class slope | NDRI | 0.172 | 0.135 | 1.269 | 1.187 | 0.911 | 1.548 | 2.04e-01 | 1.00e+00 | 5189 | 3903 |
| African | Recorded Anxiety | Bipolar Disorder | Interaction (vs SSRI) | SNRI | 0.065 | 0.114 | 0.570 | 1.067 | 0.854 | 1.333 | 5.69e-01 | 1.00e+00 | 5189 | 3903 |
| African | Recorded Anxiety | Bipolar Disorder | Interaction (vs SSRI) | TCA | -0.267 | 0.126 | -2.122 | 0.766 | 0.598 | 0.980 | 3.39e-02 | 2.37e-01 | 5189 | 3903 |
| African | Recorded Anxiety | Bipolar Disorder | Interaction (vs SSRI) | NaSSA | -0.312 | 0.188 | -1.659 | 0.732 | 0.507 | 1.058 | 9.71e-02 | 6.79e-01 | 5189 | 3903 |
| African | Recorded Anxiety | Bipolar Disorder | Interaction (vs SSRI) | SARI | 0.006 | 0.151 | 0.037 | 1.006 | 0.747 | 1.353 | 9.70e-01 | 1.00e+00 | 5189 | 3903 |
| African | Recorded Anxiety | Bipolar Disorder | Interaction (vs SSRI) | NDRI | 0.111 | 0.143 | 0.775 | 1.117 | 0.844 | 1.477 | 4.38e-01 | 1.00e+00 | 5189 | 3903 |
| African | Recorded Anxiety | Bipolar Disorder | Joint_Interaction_Test | | | | 9.559 | | | | 8.87e-02 | 6.21e-01 | 5189 | 3903 |
| African | Recorded Anxiety | BMI | Within-class slope | SSRI | -0.076 | 0.054 | -1.406 | 0.927 | 0.833 | 1.030 | 1.60e-01 | 1.00e+00 | 5189 | 3903 |
| African | Recorded Anxiety | BMI | Within-class slope | SNRI | -0.186 | 0.090 | -2.066 | 0.831 | 0.696 | 0.990 | 3.88e-02 | 2.72e-01 | 5189 | 3903 |
| African | Recorded Anxiety | BMI | Within-class slope | TCA | 0.072 | 0.101 | 0.711 | 1.074 | 0.882 | 1.309 | 4.77e-01 | 1.00e+00 | 5189 | 3903 |
| African | Recorded Anxiety | BMI | Within-class slope | NaSSA | 0.099 | 0.183 | 0.543 | 1.104 | 0.772 | 1.580 | 5.87e-01 | 1.00e+00 | 5189 | 3903 |
| African | Recorded Anxiety | BMI | Within-class slope | SARI | -0.174 | 0.114 | -1.526 | 0.840 | 0.672 | 1.051 | 1.27e-01 | 8.89e-01 | 5189 | 3903 |
| African | Recorded Anxiety | BMI | Within-class slope | NDRI | -0.087 | 0.117 | -0.742 | 0.917 | 0.729 | 1.153 | 4.58e-01 | 1.00e+00 | 5189 | 3903 |
| African | Recorded Anxiety | BMI | Interaction (vs SSRI) | SNRI | -0.109 | 0.101 | -1.086 | 0.896 | 0.736 | 1.092 | 2.78e-01 | 1.00e+00 | 5189 | 3903 |
| African | Recorded Anxiety | BMI | Interaction (vs SSRI) | TCA | 0.148 | 0.111 | 1.330 | 1.160 | 0.932 | 1.442 | 1.83e-01 | 1.00e+00 | 5189 | 3903 |
| African | Recorded Anxiety | BMI | Interaction (vs SSRI) | NaSSA | 0.176 | 0.188 | 0.933 | 1.192 | 0.824 | 1.723 | 3.51e-01 | 1.00e+00 | 5189 | 3903 |
| African | Recorded Anxiety | BMI | Interaction (vs SSRI) | SARI | -0.098 | 0.122 | -0.806 | 0.907 | 0.714 | 1.151 | 4.20e-01 | 1.00e+00 | 5189 | 3903 |
| African | Recorded Anxiety | BMI | Interaction (vs SSRI) | NDRI | -0.011 | 0.123 | -0.087 | 0.989 | 0.777 | 1.260 | 9.31e-01 | 1.00e+00 | 5189 | 3903 |
| African | Recorded Anxiety | BMI | Joint_Interaction_Test | | | | 6.030 | | | | 3.03e-01 | 1.00e+00 | 5189 | 3903 |
| African | CIDI-MDD | ADHD | Within-class slope | SSRI | 0.472 | 0.249 | 1.899 | 1.604 | 0.985 | 2.611 | 5.76e-02 | 4.03e-01 | 226 | 185 |
| African | CIDI-MDD | ADHD | Within-class slope | SNRI | 0.314 | 0.453 | 0.693 | 1.369 | 0.563 | 3.329 | 4.88e-01 | 1.00e+00 | 226 | 185 |
| African | CIDI-MDD | ADHD | Within-class slope | NDRI | 1.190 | 0.430 | 2.766 | 3.286 | 1.414 | 7.636 | 5.68e-03 | 3.98e-02 | 226 | 185 |
| African | CIDI-MDD | ADHD | Interaction (vs SSRI) | SNRI | -0.158 | 0.484 | -0.326 | 0.854 | 0.330 | 2.206 | 7.44e-01 | 1.00e+00 | 226 | 185 |
| African | CIDI-MDD | ADHD | Interaction (vs SSRI) | NDRI | 0.717 | 0.454 | 1.581 | 2.049 | 0.842 | 4.988 | 1.14e-01 | 7.98e-01 | 226 | 185 |
| African | CIDI-MDD | ADHD | Joint_Interaction_Test | | | | 2.905 | | | | 2.34e-01 | 1.00e+00 | 226 | 185 |
| African | CIDI-MDD | Migraine | Within-class slope | SSRI | -0.219 | 0.240 | -0.912 | 0.804 | 0.502 | 1.285 | 3.62e-01 | 1.00e+00 | 226 | 185 |
| African | CIDI-MDD | Migraine | Within-class slope | SNRI | -0.288 | 0.336 | -0.859 | 0.750 | 0.388 | 1.447 | 3.90e-01 | 1.00e+00 | 226 | 185 |

| | | | | | | | | | | | | | |
|---|---|---|---|---|---|---|---|---|---|---|---|---|---|
| African | CIDI-MDD | Migraine | Within-class slope | NDRI | -0.288 | 0.460 | -0.627 | 0.750 | 0.304 | 1.846 | 5.31e-01 | 1.00e+00 | 226 | 185 |
| African | CIDI-MDD | Migraine | Interaction (vs SSRI) | SNRI | -0.070 | 0.416 | -0.167 | 0.933 | 0.412 | 2.110 | 8.67e-01 | 1.00e+00 | 226 | 185 |
| African | CIDI-MDD | Migraine | Interaction (vs SSRI) | NDRI | -0.069 | 0.475 | -0.146 | 0.933 | 0.368 | 2.365 | 8.84e-01 | 1.00e+00 | 226 | 185 |
| African | CIDI-MDD | Migraine | Joint_Interaction_Test | | | | 0.038 | | | | 9.81e-01 | 1.00e+00 | 226 | 185 |
| African | CIDI-MDD | Insomnia | Within-class slope | SSRI | 0.587 | 0.310 | 1.894 | 1.799 | 0.980 | 3.302 | 5.82e-02 | 4.07e-01 | 226 | 185 |
| African | CIDI-MDD | Insomnia | Within-class slope | SNRI | -0.394 | 0.500 | -0.788 | 0.674 | 0.253 | 1.796 | 4.30e-01 | 1.00e+00 | 226 | 185 |
| African | CIDI-MDD | Insomnia | Within-class slope | NDRI | 1.190 | 0.632 | 1.883 | 3.286 | 0.953 | 11.334 | 5.97e-02 | 4.18e-01 | 226 | 185 |
| African | CIDI-MDD | Insomnia | Interaction (vs SSRI) | SNRI | -0.981 | 0.566 | -1.733 | 0.375 | 0.124 | 1.137 | 8.31e-02 | 5.82e-01 | 226 | 185 |
| African | CIDI-MDD | Insomnia | Interaction (vs SSRI) | NDRI | 0.603 | 0.679 | 0.888 | 1.827 | 0.483 | 6.909 | 3.75e-01 | 1.00e+00 | 226 | 185 |
| African | CIDI-MDD | Insomnia | Joint_Interaction_Test | | | | 4.981 | | | | 8.29e-02 | 5.80e-01 | 226 | 185 |
| African | CIDI-MDD | Depression | Within-class slope | SSRI | 0.443 | 0.302 | 1.468 | 1.557 | 0.862 | 2.813 | 1.42e-01 | 9.94e-01 | 226 | 185 |
| African | CIDI-MDD | Depression | Within-class slope | SNRI | 1.109 | 0.504 | 2.200 | 3.032 | 1.129 | 8.144 | 2.78e-02 | 1.95e-01 | 226 | 185 |
| African | CIDI-MDD | Depression | Within-class slope | NDRI | 1.425 | 0.495 | 2.881 | 4.158 | 1.577 | 10.965 | 3.97e-03 | 2.78e-02 | 226 | 185 |
| African | CIDI-MDD | Depression | Interaction (vs SSRI) | SNRI | 0.666 | 0.582 | 1.144 | 1.947 | 0.622 | 6.096 | 2.53e-01 | 1.00e+00 | 226 | 185 |
| African | CIDI-MDD | Depression | Interaction (vs SSRI) | NDRI | 0.982 | 0.558 | 1.760 | 2.670 | 0.895 | 7.970 | 7.84e-02 | 5.49e-01 | 226 | 185 |
| African | CIDI-MDD | Depression | Joint_Interaction_Test | | | | 3.462 | | | | 1.77e-01 | 1.00e+00 | 226 | 185 |
| African | CIDI-MDD | Schizophrenia | Within-class slope | SSRI | -0.099 | 0.222 | -0.447 | 0.906 | 0.587 | 1.398 | 6.55e-01 | 1.00e+00 | 226 | 185 |
| African | CIDI-MDD | Schizophrenia | Within-class slope | SNRI | 0.307 | 0.440 | 0.699 | 1.360 | 0.574 | 3.220 | 4.85e-01 | 1.00e+00 | 226 | 185 |
| African | CIDI-MDD | Schizophrenia | Within-class slope | NDRI | -0.212 | 0.482 | -0.439 | 0.809 | 0.315 | 2.081 | 6.61e-01 | 1.00e+00 | 226 | 185 |
| African | CIDI-MDD | Schizophrenia | Interaction (vs SSRI) | SNRI | 0.406 | 0.479 | 0.848 | 1.501 | 0.587 | 3.843 | 3.97e-01 | 1.00e+00 | 226 | 185 |
| African | CIDI-MDD | Schizophrenia | Interaction (vs SSRI) | NDRI | -0.113 | 0.500 | -0.225 | 0.894 | 0.335 | 2.383 | 8.22e-01 | 1.00e+00 | 226 | 185 |
| African | CIDI-MDD | Schizophrenia | Joint_Interaction_Test | | | | 0.897 | | | | 6.38e-01 | 1.00e+00 | 226 | 185 |
| African | CIDI-MDD | Bipolar Disorder | Within-class slope | SSRI | 0.394 | 0.286 | 1.377 | 1.483 | 0.846 | 2.601 | 1.69e-01 | 1.00e+00 | 226 | 185 |
| African | CIDI-MDD | Bipolar Disorder | Within-class slope | SNRI | -0.003 | 0.339 | -0.009 | 0.997 | 0.513 | 1.937 | 9.93e-01 | 1.00e+00 | 226 | 185 |
| African | CIDI-MDD | Bipolar Disorder | Within-class slope | NDRI | 0.164 | 0.448 | 0.366 | 1.178 | 0.489 | 2.837 | 7.15e-01 | 1.00e+00 | 226 | 185 |
| African | CIDI-MDD | Bipolar Disorder | Interaction (vs SSRI) | SNRI | -0.397 | 0.445 | -0.892 | 0.672 | 0.281 | 1.609 | 3.72e-01 | 1.00e+00 | 226 | 185 |
| African | CIDI-MDD | Bipolar Disorder | Interaction (vs SSRI) | NDRI | -0.230 | 0.533 | -0.432 | 0.794 | 0.279 | 2.259 | 6.66e-01 | 1.00e+00 | 226 | 185 |
| African | CIDI-MDD | Bipolar Disorder | Joint_Interaction_Test | | | | 0.802 | | | | 6.70e-01 | 1.00e+00 | 226 | 185 |
| African | CIDI-MDD | BMI | Within-class slope | SSRI | 0.012 | 0.238 | 0.053 | 1.013 | 0.635 | 1.614 | 9.58e-01 | 1.00e+00 | 226 | 185 |
| African | CIDI-MDD | BMI | Within-class slope | SNRI | 0.068 | 0.414 | 0.165 | 1.071 | 0.476 | 2.409 | 8.69e-01 | 1.00e+00 | 226 | 185 |
| African | CIDI-MDD | BMI | Within-class slope | NDRI | -0.593 | 0.425 | -1.395 | 0.553 | 0.240 | 1.272 | 1.63e-01 | 1.00e+00 | 226 | 185 |
| African | CIDI-MDD | BMI | Interaction (vs SSRI) | SNRI | 0.056 | 0.475 | 0.117 | 1.057 | 0.416 | 2.685 | 9.06e-01 | 1.00e+00 | 226 | 185 |
| African | CIDI-MDD | BMI | Interaction (vs SSRI) | NDRI | -0.605 | 0.461 | -1.314 | 0.546 | 0.221 | 1.347 | 1.89e-01 | 1.00e+00 | 226 | 185 |
| African | CIDI-MDD | BMI | Joint_Interaction_Test | | | | 2.025 | | | | 3.63e-01 | 1.00e+00 | 226 | 185 |
| Latino/Mixed-American | Full Cohort | ADHD | Within-class slope | SSRI | 0.102 | 0.048 | 2.129 | 1.108 | 1.008 | 1.217 | 3.33e-02 | 2.33e-01 | 4917 | 3922 |
| Latino/Mixed-American | Full Cohort | ADHD | Within-class slope | SNRI | 0.002 | 0.078 | 0.025 | 1.002 | 0.859 | 1.169 | 9.80e-01 | 1.00e+00 | 4917 | 3922 |
| Latino/Mixed-American | Full Cohort | ADHD | Within-class slope | TCA | -0.030 | 0.086 | -0.351 | 0.970 | 0.820 | 1.148 | 7.26e-01 | 1.00e+00 | 4917 | 3922 |
| Latino/Mixed-American | Full Cohort | ADHD | Within-class slope | NaSSA | 0.209 | 0.164 | 1.275 | 1.232 | 0.894 | 1.700 | 2.02e-01 | 1.00e+00 | 4917 | 3922 |
| Latino/Mixed-American | Full Cohort | ADHD | Within-class slope | SARI | 0.014 | 0.111 | 0.126 | 1.014 | 0.815 | 1.261 | 9.00e-01 | 1.00e+00 | 4917 | 3922 |
| Latino/Mixed-American | Full Cohort | ADHD | Within-class slope | NDRI | -0.107 | 0.098 | -1.100 | 0.898 | 0.742 | 1.088 | 2.71e-01 | 1.00e+00 | 4917 | 3922 |
| Latino/Mixed-American | Full Cohort | ADHD | Interaction (vs SSRI) | SNRI | -0.100 | 0.087 | -1.153 | 0.904 | 0.762 | 1.073 | 2.49e-01 | 1.00e+00 | 4917 | 3922 |
| Latino/Mixed-American | Full Cohort | ADHD | Interaction (vs SSRI) | TCA | -0.132 | 0.095 | -1.395 | 0.876 | 0.727 | 1.055 | 1.63e-01 | 1.00e+00 | 4917 | 3922 |
| Latino/Mixed-American | Full Cohort | ADHD | Interaction (vs SSRI) | NaSSA | 0.107 | 0.171 | 0.625 | 1.113 | 0.796 | 1.554 | 5.32e-01 | 1.00e+00 | 4917 | 3922 |
| Latino/Mixed-American | Full Cohort | ADHD | Interaction (vs SSRI) | SARI | -0.088 | 0.117 | -0.757 | 0.915 | 0.728 | 1.151 | 4.49e-01 | 1.00e+00 | 4917 | 3922 |
| Latino/Mixed-American | Full Cohort | ADHD | Interaction (vs SSRI) | NDRI | -0.210 | 0.103 | -2.032 | 0.811 | 0.662 | 0.993 | 4.21e-02 | 2.95e-01 | 4917 | 3922 |
| Latino/Mixed-American | Full Cohort | ADHD | Joint_Interaction_Test | | | | 6.913 | | | | 2.27e-01 | 1.00e+00 | 4917 | 3922 |
| Latino/Mixed-American | Full Cohort | Migraine | Within-class slope | SSRI | 0.056 | 0.046 | 1.224 | 1.058 | 0.967 | 1.158 | 2.21e-01 | 1.00e+00 | 4917 | 3922 |
| Latino/Mixed-American | Full Cohort | Migraine | Within-class slope | SNRI | 0.105 | 0.073 | 1.423 | 1.110 | 0.961 | 1.282 | 1.55e-01 | 1.00e+00 | 4917 | 3922 |
| Latino/Mixed-American | Full Cohort | Migraine | Within-class slope | TCA | 0.053 | 0.079 | 0.680 | 1.055 | 0.904 | 1.230 | 4.97e-01 | 1.00e+00 | 4917 | 3922 |
| Latino/Mixed-American | Full Cohort | Migraine | Within-class slope | NaSSA | -0.059 | 0.164 | -0.359 | 0.943 | 0.683 | 1.301 | 7.19e-01 | 1.00e+00 | 4917 | 3922 |
| Latino/Mixed-American | Full Cohort | Migraine | Within-class slope | SARI | 0.025 | 0.103 | 0.239 | 1.025 | 0.838 | 1.254 | 8.11e-01 | 1.00e+00 | 4917 | 3922 |
| Latino/Mixed-American | Full Cohort | Migraine | Within-class slope | NDRI | -0.019 | 0.098 | -0.193 | 0.981 | 0.809 | 1.190 | 8.47e-01 | 1.00e+00 | 4917 | 3922 |
| Latino/Mixed-American | Full Cohort | Migraine | Interaction (vs SSRI) | SNRI | 0.048 | 0.083 | 0.583 | 1.049 | 0.892 | 1.234 | 5.60e-01 | 1.00e+00 | 4917 | 3922 |
| Latino/Mixed-American | Full Cohort | Migraine | Interaction (vs SSRI) | TCA | -0.003 | 0.088 | -0.035 | 0.997 | 0.839 | 1.184 | 9.72e-01 | 1.00e+00 | 4917 | 3922 |

| | | | | | | | | | | | | | | |
|---|---|---|---|---|---|---|---|---|---|---|---|---|---|---|
| Latino/Mixed-American | Full Cohort | Migraine | Interaction (vs SSRI) | NaSSA | -0.115 | 0.168 | -0.686 | 0.891 | 0.641 | 1.239 | 4.92e-01 | 1.00e+00 | 4917 | 3922 |
| Latino/Mixed-American | Full Cohort | Migraine | Interaction (vs SSRI) | SARI | -0.032 | 0.107 | -0.297 | 0.969 | 0.785 | 1.196 | 7.67e-01 | 1.00e+00 | 4917 | 3922 |
| Latino/Mixed-American | Full Cohort | Migraine | Interaction (vs SSRI) | NDRI | -0.075 | 0.100 | -0.753 | 0.927 | 0.762 | 1.129 | 4.51e-01 | 1.00e+00 | 4917 | 3922 |
| Latino/Mixed-American | Full Cohort | Migraine | Joint_Interaction_Test | | | | 1.671 | | | | 8.93e-01 | 1.00e+00 | 4917 | 3922 |
| Latino/Mixed-American | Full Cohort | Insomnia | Within-class slope | SSRI | 0.078 | 0.050 | 1.554 | 1.081 | 0.980 | 1.193 | 1.20e-01 | 8.41e-01 | 4917 | 3922 |
| Latino/Mixed-American | Full Cohort | Insomnia | Within-class slope | SNRI | -0.033 | 0.082 | -0.405 | 0.967 | 0.823 | 1.137 | 6.86e-01 | 1.00e+00 | 4917 | 3922 |
| Latino/Mixed-American | Full Cohort | Insomnia | Within-class slope | TCA | 0.083 | 0.087 | 0.952 | 1.087 | 0.916 | 1.289 | 3.41e-01 | 1.00e+00 | 4917 | 3922 |
| Latino/Mixed-American | Full Cohort | Insomnia | Within-class slope | NaSSA | 0.199 | 0.191 | 1.045 | 1.220 | 0.840 | 1.773 | 2.96e-01 | 1.00e+00 | 4917 | 3922 |
| Latino/Mixed-American | Full Cohort | Insomnia | Within-class slope | SARI | 0.099 | 0.113 | 0.876 | 1.104 | 0.885 | 1.377 | 3.81e-01 | 1.00e+00 | 4917 | 3922 |
| Latino/Mixed-American | Full Cohort | Insomnia | Within-class slope | NDRI | 0.077 | 0.108 | 0.716 | 1.081 | 0.874 | 1.336 | 4.74e-01 | 1.00e+00 | 4917 | 3922 |
| Latino/Mixed-American | Full Cohort | Insomnia | Interaction (vs SSRI) | SNRI | -0.112 | 0.094 | -1.183 | 0.894 | 0.744 | 1.076 | 2.37e-01 | 1.00e+00 | 4917 | 3922 |
| Latino/Mixed-American | Full Cohort | Insomnia | Interaction (vs SSRI) | TCA | 0.005 | 0.096 | 0.050 | 1.005 | 0.833 | 1.213 | 9.60e-01 | 1.00e+00 | 4917 | 3922 |
| Latino/Mixed-American | Full Cohort | Insomnia | Interaction (vs SSRI) | NaSSA | 0.121 | 0.196 | 0.618 | 1.129 | 0.769 | 1.656 | 5.36e-01 | 1.00e+00 | 4917 | 3922 |
| Latino/Mixed-American | Full Cohort | Insomnia | Interaction (vs SSRI) | SARI | 0.021 | 0.119 | 0.173 | 1.021 | 0.808 | 1.289 | 8.63e-01 | 1.00e+00 | 4917 | 3922 |
| Latino/Mixed-American | Full Cohort | Insomnia | Interaction (vs SSRI) | NDRI | -0.001 | 0.114 | -0.006 | 0.999 | 0.799 | 1.250 | 9.95e-01 | 1.00e+00 | 4917 | 3922 |
| Latino/Mixed-American | Full Cohort | Insomnia | Joint_Interaction_Test | | | | 2.253 | | | | 8.13e-01 | 1.00e+00 | 4917 | 3922 |
| Latino/Mixed-American | Full Cohort | Depression | Within-class slope | SSRI | 0.185 | 0.050 | 3.712 | 1.204 | 1.091 | 1.328 | 2.06e-04 | 1.44e-03 | 4917 | 3922 |
| Latino/Mixed-American | Full Cohort | Depression | Within-class slope | SNRI | 0.183 | 0.081 | 2.255 | 1.201 | 1.024 | 1.407 | 2.41e-02 | 1.69e-01 | 4917 | 3922 |
| Latino/Mixed-American | Full Cohort | Depression | Within-class slope | TCA | 0.175 | 0.087 | 2.005 | 1.191 | 1.004 | 1.414 | 4.50e-02 | 3.15e-01 | 4917 | 3922 |
| Latino/Mixed-American | Full Cohort | Depression | Within-class slope | NaSSA | 0.121 | 0.166 | 0.727 | 1.128 | 0.815 | 1.562 | 4.67e-01 | 1.00e+00 | 4917 | 3922 |
| Latino/Mixed-American | Full Cohort | Depression | Within-class slope | SARI | 0.126 | 0.111 | 1.133 | 1.134 | 0.912 | 1.409 | 2.57e-01 | 1.00e+00 | 4917 | 3922 |
| Latino/Mixed-American | Full Cohort | Depression | Within-class slope | NDRI | 0.025 | 0.106 | 0.233 | 1.025 | 0.832 | 1.263 | 8.16e-01 | 1.00e+00 | 4917 | 3922 |
| Latino/Mixed-American | Full Cohort | Depression | Interaction (vs SSRI) | SNRI | -0.003 | 0.093 | -0.029 | 0.997 | 0.831 | 1.197 | 9.77e-01 | 1.00e+00 | 4917 | 3922 |
| Latino/Mixed-American | Full Cohort | Depression | Interaction (vs SSRI) | TCA | -0.010 | 0.098 | -0.107 | 0.990 | 0.817 | 1.199 | 9.15e-01 | 1.00e+00 | 4917 | 3922 |
| Latino/Mixed-American | Full Cohort | Depression | Interaction (vs SSRI) | NaSSA | -0.065 | 0.170 | -0.380 | 0.937 | 0.671 | 1.309 | 7.04e-01 | 1.00e+00 | 4917 | 3922 |
| Latino/Mixed-American | Full Cohort | Depression | Interaction (vs SSRI) | SARI | -0.060 | 0.117 | -0.510 | 0.942 | 0.749 | 1.185 | 6.10e-01 | 1.00e+00 | 4917 | 3922 |
| Latino/Mixed-American | Full Cohort | Depression | Interaction (vs SSRI) | NDRI | -0.161 | 0.112 | -1.432 | 0.852 | 0.683 | 1.061 | 1.52e-01 | 1.00e+00 | 4917 | 3922 |
| Latino/Mixed-American | Full Cohort | Depression | Joint_Interaction_Test | | | | 2.297 | | | | 8.07e-01 | 1.00e+00 | 4917 | 3922 |
| Latino/Mixed-American | Full Cohort | Schizophrenia | Within-class slope | SSRI | 0.079 | 0.044 | 1.815 | 1.083 | 0.994 | 1.179 | 6.95e-02 | 4.86e-01 | 4917 | 3922 |
| Latino/Mixed-American | Full Cohort | Schizophrenia | Within-class slope | SNRI | 0.071 | 0.074 | 0.956 | 1.073 | 0.928 | 1.241 | 3.39e-01 | 1.00e+00 | 4917 | 3922 |
| Latino/Mixed-American | Full Cohort | Schizophrenia | Within-class slope | TCA | -0.040 | 0.077 | -0.516 | 0.961 | 0.826 | 1.118 | 6.06e-01 | 1.00e+00 | 4917 | 3922 |
| Latino/Mixed-American | Full Cohort | Schizophrenia | Within-class slope | NaSSA | 0.143 | 0.168 | 0.852 | 1.154 | 0.830 | 1.603 | 3.94e-01 | 1.00e+00 | 4917 | 3922 |
| Latino/Mixed-American | Full Cohort | Schizophrenia | Within-class slope | SARI | 0.014 | 0.104 | 0.137 | 1.014 | 0.827 | 1.245 | 8.91e-01 | 1.00e+00 | 4917 | 3922 |
| Latino/Mixed-American | Full Cohort | Schizophrenia | Within-class slope | NDRI | -0.108 | 0.095 | -1.134 | 0.897 | 0.744 | 1.082 | 2.57e-01 | 1.00e+00 | 4917 | 3922 |
| Latino/Mixed-American | Full Cohort | Schizophrenia | Interaction (vs SSRI) | SNRI | -0.009 | 0.084 | -0.102 | 0.992 | 0.841 | 1.169 | 9.19e-01 | 1.00e+00 | 4917 | 3922 |
| Latino/Mixed-American | Full Cohort | Schizophrenia | Interaction (vs SSRI) | TCA | -0.119 | 0.086 | -1.387 | 0.888 | 0.750 | 1.051 | 1.66e-01 | 1.00e+00 | 4917 | 3922 |
| Latino/Mixed-American | Full Cohort | Schizophrenia | Interaction (vs SSRI) | NaSSA | 0.064 | 0.171 | 0.373 | 1.066 | 0.762 | 1.490 | 7.10e-01 | 1.00e+00 | 4917 | 3922 |
| Latino/Mixed-American | Full Cohort | Schizophrenia | Interaction (vs SSRI) | SARI | -0.065 | 0.109 | -0.594 | 0.937 | 0.756 | 1.161 | 5.53e-01 | 1.00e+00 | 4917 | 3922 |
| Latino/Mixed-American | Full Cohort | Schizophrenia | Interaction (vs SSRI) | NDRI | -0.188 | 0.101 | -1.862 | 0.829 | 0.680 | 1.010 | 6.26e-02 | 4.38e-01 | 4917 | 3922 |
| Latino/Mixed-American | Full Cohort | Schizophrenia | Joint_Interaction_Test | | | | 5.484 | | | | 3.60e-01 | 1.00e+00 | 4917 | 3922 |
| Latino/Mixed-American | Full Cohort | Bipolar Disorder | Within-class slope | SSRI | 0.062 | 0.047 | 1.314 | 1.064 | 0.970 | 1.167 | 1.89e-01 | 1.00e+00 | 4917 | 3922 |
| Latino/Mixed-American | Full Cohort | Bipolar Disorder | Within-class slope | SNRI | 0.150 | 0.081 | 1.853 | 1.162 | 0.991 | 1.361 | 6.39e-02 | 4.47e-01 | 4917 | 3922 |
| Latino/Mixed-American | Full Cohort | Bipolar Disorder | Within-class slope | TCA | 0.065 | 0.085 | 0.768 | 1.067 | 0.904 | 1.260 | 4.42e-01 | 1.00e+00 | 4917 | 3922 |
| Latino/Mixed-American | Full Cohort | Bipolar Disorder | Within-class slope | NaSSA | 0.287 | 0.180 | 1.595 | 1.332 | 0.937 | 1.895 | 1.11e-01 | 7.75e-01 | 4917 | 3922 |
| Latino/Mixed-American | Full Cohort | Bipolar Disorder | Within-class slope | SARI | -0.097 | 0.114 | -0.851 | 0.907 | 0.725 | 1.135 | 3.95e-01 | 1.00e+00 | 4917 | 3922 |
| Latino/Mixed-American | Full Cohort | Bipolar Disorder | Within-class slope | NDRI | -0.079 | 0.110 | -0.721 | 0.924 | 0.745 | 1.146 | 4.71e-01 | 1.00e+00 | 4917 | 3922 |
| Latino/Mixed-American | Full Cohort | Bipolar Disorder | Interaction (vs SSRI) | SNRI | 0.088 | 0.090 | 0.976 | 1.092 | 0.915 | 1.303 | 3.29e-01 | 1.00e+00 | 4917 | 3922 |
| Latino/Mixed-American | Full Cohort | Bipolar Disorder | Interaction (vs SSRI) | TCA | 0.003 | 0.094 | 0.035 | 1.003 | 0.835 | 1.205 | 9.72e-01 | 1.00e+00 | 4917 | 3922 |
| Latino/Mixed-American | Full Cohort | Bipolar Disorder | Interaction (vs SSRI) | NaSSA | 0.225 | 0.182 | 1.235 | 1.252 | 0.876 | 1.789 | 2.17e-01 | 1.00e+00 | 4917 | 3922 |
| Latino/Mixed-American | Full Cohort | Bipolar Disorder | Interaction (vs SSRI) | SARI | -0.159 | 0.117 | -1.356 | 0.853 | 0.677 | 1.073 | 1.75e-01 | 1.00e+00 | 4917 | 3922 |
| Latino/Mixed-American | Full Cohort | Bipolar Disorder | Interaction (vs SSRI) | NDRI | -0.141 | 0.116 | -1.219 | 0.868 | 0.692 | 1.090 | 2.23e-01 | 1.00e+00 | 4917 | 3922 |
| Latino/Mixed-American | Full Cohort | Bipolar Disorder | Joint_Interaction_Test | | | | 6.761 | | | | 2.39e-01 | 1.00e+00 | 4917 | 3922 |
| Latino/Mixed-American | Full Cohort | BMI | Within-class slope | SSRI | 0.046 | 0.045 | 1.018 | 1.047 | 0.958 | 1.145 | 3.09e-01 | 1.00e+00 | 4917 | 3922 |
| Latino/Mixed-American | Full Cohort | BMI | Within-class slope | SNRI | -0.062 | 0.074 | -0.844 | 0.940 | 0.813 | 1.086 | 3.99e-01 | 1.00e+00 | 4917 | 3922 |

| | | | | | | | | | | | | | | |
|---|---|---|---|---|---|---|---|---|---|---|---|---|---|---|
| Latino/Mixed-American | Full Cohort | BMI | Within-class slope | TCA | 0.062 | 0.082 | 0.759 | 1.064 | 0.906 | 1.250 | 4.48e-01 | 1.00e+00 | 4917 | 3922 |
| Latino/Mixed-American | Full Cohort | BMI | Within-class slope | NaSSA | 0.107 | 0.157 | 0.679 | 1.113 | 0.818 | 1.514 | 4.97e-01 | 1.00e+00 | 4917 | 3922 |
| Latino/Mixed-American | Full Cohort | BMI | Within-class slope | SARI | -0.010 | 0.102 | -0.102 | 0.990 | 0.810 | 1.209 | 9.19e-01 | 1.00e+00 | 4917 | 3922 |
| Latino/Mixed-American | Full Cohort | BMI | Within-class slope | NDRI | 0.107 | 0.099 | 1.078 | 1.113 | 0.916 | 1.351 | 2.81e-01 | 1.00e+00 | 4917 | 3922 |
| Latino/Mixed-American | Full Cohort | BMI | Interaction (vs SSRI) | SNRI | -0.109 | 0.084 | -1.290 | 0.897 | 0.761 | 1.058 | 1.97e-01 | 1.00e+00 | 4917 | 3922 |
| Latino/Mixed-American | Full Cohort | BMI | Interaction (vs SSRI) | TCA | 0.016 | 0.090 | 0.177 | 1.016 | 0.852 | 1.211 | 8.59e-01 | 1.00e+00 | 4917 | 3922 |
| Latino/Mixed-American | Full Cohort | BMI | Interaction (vs SSRI) | NaSSA | 0.061 | 0.159 | 0.380 | 1.062 | 0.777 | 1.452 | 7.04e-01 | 1.00e+00 | 4917 | 3922 |
| Latino/Mixed-American | Full Cohort | BMI | Interaction (vs SSRI) | SARI | -0.057 | 0.107 | -0.532 | 0.945 | 0.767 | 1.164 | 5.94e-01 | 1.00e+00 | 4917 | 3922 |
| Latino/Mixed-American | Full Cohort | BMI | Interaction (vs SSRI) | NDRI | 0.061 | 0.105 | 0.579 | 1.062 | 0.866 | 1.304 | 5.63e-01 | 1.00e+00 | 4917 | 3922 |
| Latino/Mixed-American | Full Cohort | BMI | Joint_Interaction_Test | | | | 3.046 | | | | 6.93e-01 | 1.00e+00 | 4917 | 3922 |
| Latino/Mixed-American | Recorded Depression | ADHD | Within-class slope | SSRI | 0.024 | 0.057 | 0.415 | 1.024 | 0.915 | 1.145 | 6.78e-01 | 1.00e+00 | 3065 | 2304 |
| Latino/Mixed-American | Recorded Depression | ADHD | Within-class slope | SNRI | 0.055 | 0.098 | 0.559 | 1.056 | 0.872 | 1.279 | 5.76e-01 | 1.00e+00 | 3065 | 2304 |
| Latino/Mixed-American | Recorded Depression | ADHD | Within-class slope | TCA | -0.024 | 0.118 | -0.203 | 0.976 | 0.775 | 1.230 | 8.39e-01 | 1.00e+00 | 3065 | 2304 |
| Latino/Mixed-American | Recorded Depression | ADHD | Within-class slope | NaSSA | 0.216 | 0.212 | 1.017 | 1.241 | 0.819 | 1.881 | 3.09e-01 | 1.00e+00 | 3065 | 2304 |
| Latino/Mixed-American | Recorded Depression | ADHD | Within-class slope | SARI | -0.209 | 0.160 | -1.302 | 0.812 | 0.593 | 1.111 | 1.93e-01 | 1.00e+00 | 3065 | 2304 |
| Latino/Mixed-American | Recorded Depression | ADHD | Within-class slope | NDRI | -0.068 | 0.119 | -0.567 | 0.935 | 0.740 | 1.181 | 5.70e-01 | 1.00e+00 | 3065 | 2304 |
| Latino/Mixed-American | Recorded Depression | ADHD | Interaction (vs SSRI) | SNRI | 0.031 | 0.108 | 0.286 | 1.031 | 0.835 | 1.273 | 7.75e-01 | 1.00e+00 | 3065 | 2304 |
| Latino/Mixed-American | Recorded Depression | ADHD | Interaction (vs SSRI) | TCA | -0.048 | 0.126 | -0.377 | 0.953 | 0.744 | 1.222 | 7.06e-01 | 1.00e+00 | 3065 | 2304 |
| Latino/Mixed-American | Recorded Depression | ADHD | Interaction (vs SSRI) | NaSSA | 0.192 | 0.219 | 0.876 | 1.212 | 0.788 | 1.862 | 3.81e-01 | 1.00e+00 | 3065 | 2304 |
| Latino/Mixed-American | Recorded Depression | ADHD | Interaction (vs SSRI) | SARI | -0.232 | 0.164 | -1.417 | 0.793 | 0.575 | 1.093 | 1.56e-01 | 1.00e+00 | 3065 | 2304 |
| Latino/Mixed-American | Recorded Depression | ADHD | Interaction (vs SSRI) | NDRI | -0.091 | 0.126 | -0.727 | 0.913 | 0.713 | 1.168 | 4.67e-01 | 1.00e+00 | 3065 | 2304 |
| Latino/Mixed-American | Recorded Depression | ADHD | Joint_Interaction_Test | | | | 3.893 | | | | 5.65e-01 | 1.00e+00 | 3065 | 2304 |
| Latino/Mixed-American | Recorded Depression | Migraine | Within-class slope | SSRI | 0.084 | 0.054 | 1.549 | 1.088 | 0.978 | 1.209 | 1.21e-01 | 8.50e-01 | 3065 | 2304 |
| Latino/Mixed-American | Recorded Depression | Migraine | Within-class slope | SNRI | 0.184 | 0.090 | 2.047 | 1.203 | 1.008 | 1.435 | 4.06e-02 | 2.84e-01 | 3065 | 2304 |
| Latino/Mixed-American | Recorded Depression | Migraine | Within-class slope | TCA | 0.183 | 0.113 | 1.623 | 1.201 | 0.963 | 1.497 | 1.05e-01 | 7.32e-01 | 3065 | 2304 |
| Latino/Mixed-American | Recorded Depression | Migraine | Within-class slope | NaSSA | -0.073 | 0.199 | -0.367 | 0.930 | 0.630 | 1.372 | 7.14e-01 | 1.00e+00 | 3065 | 2304 |
| Latino/Mixed-American | Recorded Depression | Migraine | Within-class slope | SARI | -0.009 | 0.142 | -0.061 | 0.991 | 0.750 | 1.311 | 9.51e-01 | 1.00e+00 | 3065 | 2304 |
| Latino/Mixed-American | Recorded Depression | Migraine | Within-class slope | NDRI | 0.045 | 0.114 | 0.395 | 1.046 | 0.837 | 1.307 | 6.93e-01 | 1.00e+00 | 3065 | 2304 |
| Latino/Mixed-American | Recorded Depression | Migraine | Interaction (vs SSRI) | SNRI | 0.100 | 0.101 | 0.999 | 1.106 | 0.908 | 1.347 | 3.18e-01 | 1.00e+00 | 3065 | 2304 |
| Latino/Mixed-American | Recorded Depression | Migraine | Interaction (vs SSRI) | TCA | 0.099 | 0.121 | 0.816 | 1.104 | 0.871 | 1.400 | 4.14e-01 | 1.00e+00 | 3065 | 2304 |
| Latino/Mixed-American | Recorded Depression | Migraine | Interaction (vs SSRI) | NaSSA | -0.157 | 0.203 | -0.772 | 0.855 | 0.574 | 1.273 | 4.40e-01 | 1.00e+00 | 3065 | 2304 |
| Latino/Mixed-American | Recorded Depression | Migraine | Interaction (vs SSRI) | SARI | -0.093 | 0.145 | -0.639 | 0.911 | 0.686 | 1.211 | 5.23e-01 | 1.00e+00 | 3065 | 2304 |
| Latino/Mixed-American | Recorded Depression | Migraine | Interaction (vs SSRI) | NDRI | -0.039 | 0.116 | -0.337 | 0.962 | 0.766 | 1.207 | 7.36e-01 | 1.00e+00 | 3065 | 2304 |
| Latino/Mixed-American | Recorded Depression | Migraine | Joint_Interaction_Test | | | | 3.165 | | | | 6.75e-01 | 1.00e+00 | 3065 | 2304 |
| Latino/Mixed-American | Recorded Depression | Insomnia | Within-class slope | SSRI | 0.007 | 0.058 | 0.124 | 1.007 | 0.899 | 1.129 | 9.02e-01 | 1.00e+00 | 3065 | 2304 |
| Latino/Mixed-American | Recorded Depression | Insomnia | Within-class slope | SNRI | 0.084 | 0.104 | 0.807 | 1.088 | 0.887 | 1.335 | 4.19e-01 | 1.00e+00 | 3065 | 2304 |
| Latino/Mixed-American | Recorded Depression | Insomnia | Within-class slope | TCA | 0.004 | 0.116 | 0.036 | 1.004 | 0.800 | 1.260 | 9.71e-01 | 1.00e+00 | 3065 | 2304 |
| Latino/Mixed-American | Recorded Depression | Insomnia | Within-class slope | NaSSA | 0.098 | 0.232 | 0.423 | 1.103 | 0.700 | 1.738 | 6.72e-01 | 1.00e+00 | 3065 | 2304 |
| Latino/Mixed-American | Recorded Depression | Insomnia | Within-class slope | SARI | 0.063 | 0.147 | 0.428 | 1.065 | 0.798 | 1.422 | 6.68e-01 | 1.00e+00 | 3065 | 2304 |
| Latino/Mixed-American | Recorded Depression | Insomnia | Within-class slope | NDRI | 0.108 | 0.134 | 0.802 | 1.114 | 0.856 | 1.449 | 4.23e-01 | 1.00e+00 | 3065 | 2304 |
| Latino/Mixed-American | Recorded Depression | Insomnia | Interaction (vs SSRI) | SNRI | 0.077 | 0.118 | 0.655 | 1.080 | 0.858 | 1.361 | 5.13e-01 | 1.00e+00 | 3065 | 2304 |
| Latino/Mixed-American | Recorded Depression | Insomnia | Interaction (vs SSRI) | TCA | -0.003 | 0.124 | -0.024 | 0.997 | 0.781 | 1.272 | 9.81e-01 | 1.00e+00 | 3065 | 2304 |
| Latino/Mixed-American | Recorded Depression | Insomnia | Interaction (vs SSRI) | NaSSA | 0.091 | 0.238 | 0.382 | 1.095 | 0.687 | 1.746 | 7.02e-01 | 1.00e+00 | 3065 | 2304 |
| Latino/Mixed-American | Recorded Depression | Insomnia | Interaction (vs SSRI) | SARI | 0.056 | 0.152 | 0.367 | 1.058 | 0.785 | 1.425 | 7.13e-01 | 1.00e+00 | 3065 | 2304 |
| Latino/Mixed-American | Recorded Depression | Insomnia | Interaction (vs SSRI) | NDRI | 0.101 | 0.141 | 0.712 | 1.106 | 0.839 | 1.458 | 4.76e-01 | 1.00e+00 | 3065 | 2304 |
| Latino/Mixed-American | Recorded Depression | Insomnia | Joint_Interaction_Test | | | | 0.958 | | | | 9.66e-01 | 1.00e+00 | 3065 | 2304 |
| Latino/Mixed-American | Recorded Depression | Depression | Within-class slope | SSRI | 0.160 | 0.059 | 2.712 | 1.174 | 1.045 | 1.318 | 6.69e-03 | 4.68e-02 | 3065 | 2304 |
| Latino/Mixed-American | Recorded Depression | Depression | Within-class slope | SNRI | 0.299 | 0.102 | 2.924 | 1.348 | 1.104 | 1.647 | 3.45e-03 | 2.42e-02 | 3065 | 2304 |
| Latino/Mixed-American | Recorded Depression | Depression | Within-class slope | TCA | 0.142 | 0.122 | 1.164 | 1.152 | 0.908 | 1.462 | 2.44e-01 | 1.00e+00 | 3065 | 2304 |
| Latino/Mixed-American | Recorded Depression | Depression | Within-class slope | NaSSA | 0.005 | 0.214 | 0.025 | 1.005 | 0.660 | 1.531 | 9.80e-01 | 1.00e+00 | 3065 | 2304 |
| Latino/Mixed-American | Recorded Depression | Depression | Within-class slope | SARI | 0.074 | 0.170 | 0.432 | 1.076 | 0.771 | 1.503 | 6.65e-01 | 1.00e+00 | 3065 | 2304 |
| Latino/Mixed-American | Recorded Depression | Depression | Within-class slope | NDRI | 0.040 | 0.126 | 0.317 | 1.041 | 0.813 | 1.333 | 7.51e-01 | 1.00e+00 | 3065 | 2304 |
| Latino/Mixed-American | Recorded Depression | Depression | Interaction (vs SSRI) | SNRI | 0.138 | 0.116 | 1.195 | 1.149 | 0.915 | 1.441 | 2.32e-01 | 1.00e+00 | 3065 | 2304 |
| Latino/Mixed-American | Recorded Depression | Depression | Interaction (vs SSRI) | TCA | -0.019 | 0.132 | -0.142 | 0.981 | 0.758 | 1.271 | 8.87e-01 | 1.00e+00 | 3065 | 2304 |

| | | | | | | | | | | | | | | |
|---|---|---|---|---|---|---|---|---|---|---|---|---|---|---|
| Latino/Mixed-American | Recorded Depression | Depression | Interaction (vs SSRI) | NaSSA | -0.155 | 0.219 | -0.707 | 0.856 | 0.557 | 1.316 | 4.80e-01 | 1.00e+00 | 3065 | 2304 |
| Latino/Mixed-American | Recorded Depression | Depression | Interaction (vs SSRI) | SARI | -0.087 | 0.175 | -0.495 | 0.917 | 0.651 | 1.292 | 6.21e-01 | 1.00e+00 | 3065 | 2304 |
| Latino/Mixed-American | Recorded Depression | Depression | Interaction (vs SSRI) | NDRI | -0.120 | 0.134 | -0.896 | 0.887 | 0.682 | 1.154 | 3.70e-01 | 1.00e+00 | 3065 | 2304 |
| Latino/Mixed-American | Recorded Depression | Depression | Joint_Interaction_Test | | | | 3.670 | | | | 5.98e-01 | 1.00e+00 | 3065 | 2304 |
| Latino/Mixed-American | Recorded Depression | Schizophrenia | Within-class slope | SSRI | 0.097 | 0.052 | 1.867 | 1.102 | 0.995 | 1.220 | 6.19e-02 | 4.33e-01 | 3065 | 2304 |
| Latino/Mixed-American | Recorded Depression | Schizophrenia | Within-class slope | SNRI | 0.062 | 0.096 | 0.643 | 1.064 | 0.881 | 1.285 | 5.20e-01 | 1.00e+00 | 3065 | 2304 |
| Latino/Mixed-American | Recorded Depression | Schizophrenia | Within-class slope | TCA | 0.063 | 0.110 | 0.573 | 1.065 | 0.858 | 1.323 | 5.67e-01 | 1.00e+00 | 3065 | 2304 |
| Latino/Mixed-American | Recorded Depression | Schizophrenia | Within-class slope | NaSSA | 0.084 | 0.226 | 0.372 | 1.088 | 0.698 | 1.695 | 7.10e-01 | 1.00e+00 | 3065 | 2304 |
| Latino/Mixed-American | Recorded Depression | Schizophrenia | Within-class slope | SARI | 0.001 | 0.147 | 0.007 | 1.001 | 0.751 | 1.334 | 9.95e-01 | 1.00e+00 | 3065 | 2304 |
| Latino/Mixed-American | Recorded Depression | Schizophrenia | Within-class slope | NDRI | -0.173 | 0.118 | -1.467 | 0.841 | 0.668 | 1.060 | 1.42e-01 | 9.96e-01 | 3065 | 2304 |
| Latino/Mixed-American | Recorded Depression | Schizophrenia | Interaction (vs SSRI) | SNRI | -0.035 | 0.107 | -0.329 | 0.966 | 0.783 | 1.190 | 7.43e-01 | 1.00e+00 | 3065 | 2304 |
| Latino/Mixed-American | Recorded Depression | Schizophrenia | Interaction (vs SSRI) | TCA | -0.034 | 0.118 | -0.285 | 0.967 | 0.767 | 1.219 | 7.76e-01 | 1.00e+00 | 3065 | 2304 |
| Latino/Mixed-American | Recorded Depression | Schizophrenia | Interaction (vs SSRI) | NaSSA | -0.013 | 0.230 | -0.056 | 0.987 | 0.629 | 1.549 | 9.55e-01 | 1.00e+00 | 3065 | 2304 |
| Latino/Mixed-American | Recorded Depression | Schizophrenia | Interaction (vs SSRI) | SARI | -0.096 | 0.150 | -0.638 | 0.908 | 0.677 | 1.220 | 5.23e-01 | 1.00e+00 | 3065 | 2304 |
| Latino/Mixed-American | Recorded Depression | Schizophrenia | Interaction (vs SSRI) | NDRI | -0.270 | 0.124 | -2.177 | 0.763 | 0.599 | 0.973 | 2.95e-02 | 2.06e-01 | 3065 | 2304 |
| Latino/Mixed-American | Recorded Depression | Schizophrenia | Joint_Interaction_Test | | | | 4.977 | | | | 4.19e-01 | 1.00e+00 | 3065 | 2304 |
| Latino/Mixed-American | Recorded Depression | Bipolar Disorder | Within-class slope | SSRI | 0.074 | 0.056 | 1.329 | 1.077 | 0.965 | 1.202 | 1.84e-01 | 1.00e+00 | 3065 | 2304 |
| Latino/Mixed-American | Recorded Depression | Bipolar Disorder | Within-class slope | SNRI | 0.152 | 0.099 | 1.544 | 1.164 | 0.960 | 1.412 | 1.23e-01 | 8.58e-01 | 3065 | 2304 |
| Latino/Mixed-American | Recorded Depression | Bipolar Disorder | Within-class slope | TCA | 0.206 | 0.125 | 1.647 | 1.228 | 0.962 | 1.569 | 9.96e-02 | 6.97e-01 | 3065 | 2304 |
| Latino/Mixed-American | Recorded Depression | Bipolar Disorder | Within-class slope | NaSSA | 0.413 | 0.238 | 1.739 | 1.512 | 0.949 | 2.408 | 8.20e-02 | 5.74e-01 | 3065 | 2304 |
| Latino/Mixed-American | Recorded Depression | Bipolar Disorder | Within-class slope | SARI | -0.093 | 0.169 | -0.547 | 0.911 | 0.654 | 1.271 | 5.84e-01 | 1.00e+00 | 3065 | 2304 |
| Latino/Mixed-American | Recorded Depression | Bipolar Disorder | Within-class slope | NDRI | -0.003 | 0.139 | -0.023 | 0.997 | 0.760 | 1.308 | 9.82e-01 | 1.00e+00 | 3065 | 2304 |
| Latino/Mixed-American | Recorded Depression | Bipolar Disorder | Interaction (vs SSRI) | SNRI | 0.078 | 0.109 | 0.711 | 1.081 | 0.872 | 1.339 | 4.77e-01 | 1.00e+00 | 3065 | 2304 |
| Latino/Mixed-American | Recorded Depression | Bipolar Disorder | Interaction (vs SSRI) | TCA | 0.131 | 0.132 | 0.997 | 1.140 | 0.881 | 1.476 | 3.19e-01 | 1.00e+00 | 3065 | 2304 |
| Latino/Mixed-American | Recorded Depression | Bipolar Disorder | Interaction (vs SSRI) | NaSSA | 0.339 | 0.239 | 1.415 | 1.403 | 0.878 | 2.244 | 1.57e-01 | 1.00e+00 | 3065 | 2304 |
| Latino/Mixed-American | Recorded Depression | Bipolar Disorder | Interaction (vs SSRI) | SARI | -0.167 | 0.171 | -0.975 | 0.846 | 0.605 | 1.184 | 3.30e-01 | 1.00e+00 | 3065 | 2304 |
| Latino/Mixed-American | Recorded Depression | Bipolar Disorder | Interaction (vs SSRI) | NDRI | -0.078 | 0.145 | -0.536 | 0.925 | 0.697 | 1.229 | 5.92e-01 | 1.00e+00 | 3065 | 2304 |
| Latino/Mixed-American | Recorded Depression | Bipolar Disorder | Joint_Interaction_Test | | | | 5.294 | | | | 3.81e-01 | 1.00e+00 | 3065 | 2304 |
| Latino/Mixed-American | Recorded Depression | BMI | Within-class slope | SSRI | -0.003 | 0.055 | -0.056 | 0.997 | 0.895 | 1.111 | 9.55e-01 | 1.00e+00 | 3065 | 2304 |
| Latino/Mixed-American | Recorded Depression | BMI | Within-class slope | SNRI | -0.147 | 0.093 | -1.569 | 0.864 | 0.719 | 1.037 | 1.17e-01 | 8.16e-01 | 3065 | 2304 |
| Latino/Mixed-American | Recorded Depression | BMI | Within-class slope | TCA | 0.164 | 0.109 | 1.501 | 1.178 | 0.951 | 1.460 | 1.33e-01 | 9.34e-01 | 3065 | 2304 |
| Latino/Mixed-American | Recorded Depression | BMI | Within-class slope | NaSSA | 0.119 | 0.205 | 0.579 | 1.126 | 0.753 | 1.683 | 5.63e-01 | 1.00e+00 | 3065 | 2304 |
| Latino/Mixed-American | Recorded Depression | BMI | Within-class slope | SARI | -0.064 | 0.139 | -0.464 | 0.938 | 0.714 | 1.231 | 6.42e-01 | 1.00e+00 | 3065 | 2304 |
| Latino/Mixed-American | Recorded Depression | BMI | Within-class slope | NDRI | 0.004 | 0.127 | 0.030 | 1.004 | 0.782 | 1.288 | 9.76e-01 | 1.00e+00 | 3065 | 2304 |
| Latino/Mixed-American | Recorded Depression | BMI | Interaction (vs SSRI) | SNRI | -0.144 | 0.106 | -1.353 | 0.866 | 0.704 | 1.066 | 1.76e-01 | 1.00e+00 | 3065 | 2304 |
| Latino/Mixed-American | Recorded Depression | BMI | Interaction (vs SSRI) | TCA | 0.167 | 0.118 | 1.420 | 1.182 | 0.938 | 1.488 | 1.56e-01 | 1.00e+00 | 3065 | 2304 |
| Latino/Mixed-American | Recorded Depression | BMI | Interaction (vs SSRI) | NaSSA | 0.122 | 0.209 | 0.584 | 1.130 | 0.751 | 1.700 | 5.59e-01 | 1.00e+00 | 3065 | 2304 |
| Latino/Mixed-American | Recorded Depression | BMI | Interaction (vs SSRI) | SARI | -0.061 | 0.141 | -0.433 | 0.941 | 0.713 | 1.241 | 6.65e-01 | 1.00e+00 | 3065 | 2304 |
| Latino/Mixed-American | Recorded Depression | BMI | Interaction (vs SSRI) | NDRI | 0.007 | 0.134 | 0.052 | 1.007 | 0.774 | 1.309 | 9.59e-01 | 1.00e+00 | 3065 | 2304 |
| Latino/Mixed-American | Recorded Depression | BMI | Joint_Interaction_Test | | | | 5.881 | | | | 3.18e-01 | 1.00e+00 | 3065 | 2304 |
| Latino/Mixed-American | No Recorded Depression | ADHD | Within-class slope | SSRI | 0.237 | 0.102 | 2.326 | 1.268 | 1.038 | 1.549 | 2.00e-02 | 1.40e-01 | 1791 | 1581 |
| Latino/Mixed-American | No Recorded Depression | ADHD | Within-class slope | SNRI | -0.212 | 0.147 | -1.448 | 0.809 | 0.607 | 1.078 | 1.48e-01 | 1.00e+00 | 1791 | 1581 |
| Latino/Mixed-American | No Recorded Depression | ADHD | Within-class slope | TCA | -0.075 | 0.155 | -0.481 | 0.928 | 0.685 | 1.258 | 6.31e-01 | 1.00e+00 | 1791 | 1581 |
| Latino/Mixed-American | No Recorded Depression | ADHD | Within-class slope | SARI | 0.224 | 0.199 | 1.126 | 1.251 | 0.847 | 1.848 | 2.60e-01 | 1.00e+00 | 1791 | 1581 |
| Latino/Mixed-American | No Recorded Depression | ADHD | Within-class slope | NDRI | -0.425 | 0.229 | -1.856 | 0.654 | 0.418 | 1.024 | 6.34e-02 | 4.44e-01 | 1791 | 1581 |
| Latino/Mixed-American | No Recorded Depression | ADHD | Interaction (vs SSRI) | SNRI | -0.450 | 0.171 | -2.624 | 0.638 | 0.456 | 0.892 | 8.70e-03 | 6.09e-02 | 1791 | 1581 |
| Latino/Mixed-American | No Recorded Depression | ADHD | Interaction (vs SSRI) | TCA | -0.312 | 0.183 | -1.708 | 0.732 | 0.512 | 1.047 | 8.77e-02 | 6.14e-01 | 1791 | 1581 |
| Latino/Mixed-American | No Recorded Depression | ADHD | Interaction (vs SSRI) | SARI | -0.013 | 0.217 | -0.061 | 0.987 | 0.645 | 1.509 | 9.51e-01 | 1.00e+00 | 1791 | 1581 |
| Latino/Mixed-American | No Recorded Depression | ADHD | Interaction (vs SSRI) | NDRI | -0.662 | 0.239 | -2.772 | 0.516 | 0.323 | 0.824 | 5.57e-03 | 3.90e-02 | 1791 | 1581 |
| Latino/Mixed-American | No Recorded Depression | ADHD | Joint_Interaction_Test | | | | 13.744 | | | | 8.16e-03 | 5.71e-02 | 1791 | 1581 |
| Latino/Mixed-American | No Recorded Depression | Migraine | Within-class slope | SSRI | 0.033 | 0.094 | 0.349 | 1.033 | 0.860 | 1.242 | 7.27e-01 | 1.00e+00 | 1791 | 1581 |
| Latino/Mixed-American | No Recorded Depression | Migraine | Within-class slope | SNRI | -0.115 | 0.137 | -0.842 | 0.891 | 0.681 | 1.165 | 4.00e-01 | 1.00e+00 | 1791 | 1581 |
| Latino/Mixed-American | No Recorded Depression | Migraine | Within-class slope | TCA | -0.019 | 0.128 | -0.145 | 0.982 | 0.764 | 1.261 | 8.84e-01 | 1.00e+00 | 1791 | 1581 |
| Latino/Mixed-American | No Recorded Depression | Migraine | Within-class slope | SARI | -0.027 | 0.187 | -0.145 | 0.973 | 0.674 | 1.405 | 8.85e-01 | 1.00e+00 | 1791 | 1581 |

| | | | | | | | | | | | | | | |
|---|---|---|---|---|---|---|---|---|---|---|---|---|---|---|
| Latino/Mixed-American | No Recorded Depression | Migraine | Within-class slope | NDRI | 0.011 | 0.220 | 0.048 | 1.011 | 0.657 | 1.556 | 9.61e-01 | 1.00e+00 | 1791 | 1581 |
| Latino/Mixed-American | No Recorded Depression | Migraine | Interaction (vs SSRI) | SNRI | -0.148 | 0.162 | -0.913 | 0.862 | 0.627 | 1.185 | 3.61e-01 | 1.00e+00 | 1791 | 1581 |
| Latino/Mixed-American | No Recorded Depression | Migraine | Interaction (vs SSRI) | TCA | -0.051 | 0.155 | -0.332 | 0.950 | 0.701 | 1.287 | 7.40e-01 | 1.00e+00 | 1791 | 1581 |
| Latino/Mixed-American | No Recorded Depression | Migraine | Interaction (vs SSRI) | SARI | -0.060 | 0.207 | -0.290 | 0.942 | 0.628 | 1.413 | 7.72e-01 | 1.00e+00 | 1791 | 1581 |
| Latino/Mixed-American | No Recorded Depression | Migraine | Interaction (vs SSRI) | NDRI | -0.022 | 0.230 | -0.096 | 0.978 | 0.623 | 1.536 | 9.23e-01 | 1.00e+00 | 1791 | 1581 |
| Latino/Mixed-American | No Recorded Depression | Migraine | Joint_Interaction_Test | | | | 0.836 | | | | 9.34e-01 | 1.00e+00 | 1791 | 1581 |
| Latino/Mixed-American | No Recorded Depression | Insomnia | Within-class slope | SSRI | 0.169 | 0.107 | 1.579 | 1.184 | 0.960 | 1.460 | 1.14e-01 | 8.00e-01 | 1791 | 1581 |
| Latino/Mixed-American | No Recorded Depression | Insomnia | Within-class slope | SNRI | -0.437 | 0.163 | -2.681 | 0.646 | 0.469 | 0.889 | 7.34e-03 | 5.14e-02 | 1791 | 1581 |
| Latino/Mixed-American | No Recorded Depression | Insomnia | Within-class slope | TCA | 0.013 | 0.142 | 0.093 | 1.013 | 0.767 | 1.338 | 9.26e-01 | 1.00e+00 | 1791 | 1581 |
| Latino/Mixed-American | No Recorded Depression | Insomnia | Within-class slope | SARI | -0.032 | 0.183 | -0.174 | 0.969 | 0.677 | 1.387 | 8.62e-01 | 1.00e+00 | 1791 | 1581 |
| Latino/Mixed-American | No Recorded Depression | Insomnia | Within-class slope | NDRI | 0.023 | 0.220 | 0.106 | 1.024 | 0.665 | 1.577 | 9.15e-01 | 1.00e+00 | 1791 | 1581 |
| Latino/Mixed-American | No Recorded Depression | Insomnia | Interaction (vs SSRI) | SNRI | -0.606 | 0.192 | -3.158 | 0.545 | 0.374 | 0.795 | 1.59e-03 | 1.11e-02 | 1791 | 1581 |
| Latino/Mixed-American | No Recorded Depression | Insomnia | Interaction (vs SSRI) | TCA | -0.156 | 0.174 | -0.892 | 0.856 | 0.608 | 1.205 | 3.73e-01 | 1.00e+00 | 1791 | 1581 |
| Latino/Mixed-American | No Recorded Depression | Insomnia | Interaction (vs SSRI) | SARI | -0.201 | 0.212 | -0.944 | 0.818 | 0.540 | 1.241 | 3.45e-01 | 1.00e+00 | 1791 | 1581 |
| Latino/Mixed-American | No Recorded Depression | Insomnia | Interaction (vs SSRI) | NDRI | -0.145 | 0.228 | -0.637 | 0.865 | 0.553 | 1.352 | 5.24e-01 | 1.00e+00 | 1791 | 1581 |
| Latino/Mixed-American | No Recorded Depression | Insomnia | Joint_Interaction_Test | | | | 10.049 | | | | 3.96e-02 | 2.77e-01 | 1791 | 1581 |
| Latino/Mixed-American | No Recorded Depression | Depression | Within-class slope | SSRI | 0.155 | 0.097 | 1.598 | 1.168 | 0.965 | 1.413 | 1.10e-01 | 7.70e-01 | 1791 | 1581 |
| Latino/Mixed-American | No Recorded Depression | Depression | Within-class slope | SNRI | -0.122 | 0.162 | -0.755 | 0.885 | 0.644 | 1.216 | 4.50e-01 | 1.00e+00 | 1791 | 1581 |
| Latino/Mixed-American | No Recorded Depression | Depression | Within-class slope | TCA | 0.149 | 0.143 | 1.045 | 1.161 | 0.878 | 1.535 | 2.96e-01 | 1.00e+00 | 1791 | 1581 |
| Latino/Mixed-American | No Recorded Depression | Depression | Within-class slope | SARI | 0.166 | 0.200 | 0.830 | 1.181 | 0.798 | 1.748 | 4.07e-01 | 1.00e+00 | 1791 | 1581 |
| Latino/Mixed-American | No Recorded Depression | Depression | Within-class slope | NDRI | -0.011 | 0.264 | -0.043 | 0.989 | 0.589 | 1.660 | 9.66e-01 | 1.00e+00 | 1791 | 1581 |
| Latino/Mixed-American | No Recorded Depression | Depression | Interaction (vs SSRI) | SNRI | -0.278 | 0.184 | -1.507 | 0.757 | 0.528 | 1.087 | 1.32e-01 | 9.23e-01 | 1791 | 1581 |
| Latino/Mixed-American | No Recorded Depression | Depression | Interaction (vs SSRI) | TCA | -0.006 | 0.171 | -0.038 | 0.994 | 0.710 | 1.390 | 9.70e-01 | 1.00e+00 | 1791 | 1581 |
| Latino/Mixed-American | No Recorded Depression | Depression | Interaction (vs SSRI) | SARI | 0.011 | 0.217 | 0.049 | 1.011 | 0.661 | 1.547 | 9.61e-01 | 1.00e+00 | 1791 | 1581 |
| Latino/Mixed-American | No Recorded Depression | Depression | Interaction (vs SSRI) | NDRI | -0.167 | 0.270 | -0.617 | 0.846 | 0.498 | 1.438 | 5.38e-01 | 1.00e+00 | 1791 | 1581 |
| Latino/Mixed-American | No Recorded Depression | Depression | Joint_Interaction_Test | | | | 2.903 | | | | 5.74e-01 | 1.00e+00 | 1791 | 1581 |
| Latino/Mixed-American | No Recorded Depression | Schizophrenia | Within-class slope | SSRI | 0.036 | 0.088 | 0.410 | 1.037 | 0.872 | 1.233 | 6.82e-01 | 1.00e+00 | 1791 | 1581 |
| Latino/Mixed-American | No Recorded Depression | Schizophrenia | Within-class slope | SNRI | 0.100 | 0.143 | 0.701 | 1.106 | 0.835 | 1.464 | 4.83e-01 | 1.00e+00 | 1791 | 1581 |
| Latino/Mixed-American | No Recorded Depression | Schizophrenia | Within-class slope | TCA | 0.016 | 0.124 | 0.131 | 1.016 | 0.797 | 1.296 | 8.95e-01 | 1.00e+00 | 1791 | 1581 |
| Latino/Mixed-American | No Recorded Depression | Schizophrenia | Within-class slope | SARI | 0.128 | 0.197 | 0.651 | 1.137 | 0.773 | 1.671 | 5.15e-01 | 1.00e+00 | 1791 | 1581 |
| Latino/Mixed-American | No Recorded Depression | Schizophrenia | Within-class slope | NDRI | -0.103 | 0.225 | -0.460 | 0.902 | 0.580 | 1.402 | 6.46e-01 | 1.00e+00 | 1791 | 1581 |
| Latino/Mixed-American | No Recorded Depression | Schizophrenia | Interaction (vs SSRI) | SNRI | 0.064 | 0.167 | 0.383 | 1.066 | 0.768 | 1.479 | 7.01e-01 | 1.00e+00 | 1791 | 1581 |
| Latino/Mixed-American | No Recorded Depression | Schizophrenia | Interaction (vs SSRI) | TCA | -0.020 | 0.149 | -0.134 | 0.980 | 0.732 | 1.312 | 8.93e-01 | 1.00e+00 | 1791 | 1581 |
| Latino/Mixed-American | No Recorded Depression | Schizophrenia | Interaction (vs SSRI) | SARI | 0.092 | 0.206 | 0.445 | 1.096 | 0.732 | 1.642 | 6.57e-01 | 1.00e+00 | 1791 | 1581 |
| Latino/Mixed-American | No Recorded Depression | Schizophrenia | Interaction (vs SSRI) | NDRI | -0.140 | 0.234 | -0.597 | 0.870 | 0.550 | 1.375 | 5.50e-01 | 1.00e+00 | 1791 | 1581 |
| Latino/Mixed-American | No Recorded Depression | Schizophrenia | Joint_Interaction_Test | | | | 0.871 | | | | 9.29e-01 | 1.00e+00 | 1791 | 1581 |
| Latino/Mixed-American | No Recorded Depression | Bipolar Disorder | Within-class slope | SSRI | -0.020 | 0.101 | -0.195 | 0.981 | 0.805 | 1.194 | 8.45e-01 | 1.00e+00 | 1791 | 1581 |
| Latino/Mixed-American | No Recorded Depression | Bipolar Disorder | Within-class slope | SNRI | 0.159 | 0.161 | 0.989 | 1.173 | 0.855 | 1.609 | 3.23e-01 | 1.00e+00 | 1791 | 1581 |
| Latino/Mixed-American | No Recorded Depression | Bipolar Disorder | Within-class slope | TCA | -0.027 | 0.134 | -0.202 | 0.973 | 0.748 | 1.267 | 8.40e-01 | 1.00e+00 | 1791 | 1581 |
| Latino/Mixed-American | No Recorded Depression | Bipolar Disorder | Within-class slope | SARI | -0.251 | 0.231 | -1.087 | 0.778 | 0.495 | 1.223 | 2.77e-01 | 1.00e+00 | 1791 | 1581 |
| Latino/Mixed-American | No Recorded Depression | Bipolar Disorder | Within-class slope | NDRI | -0.150 | 0.206 | -0.730 | 0.860 | 0.575 | 1.288 | 4.65e-01 | 1.00e+00 | 1791 | 1581 |
| Latino/Mixed-American | No Recorded Depression | Bipolar Disorder | Interaction (vs SSRI) | SNRI | 0.179 | 0.184 | 0.971 | 1.196 | 0.833 | 1.717 | 3.31e-01 | 1.00e+00 | 1791 | 1581 |
| Latino/Mixed-American | No Recorded Depression | Bipolar Disorder | Interaction (vs SSRI) | TCA | -0.008 | 0.162 | -0.047 | 0.992 | 0.723 | 1.363 | 9.63e-01 | 1.00e+00 | 1791 | 1581 |
| Latino/Mixed-American | No Recorded Depression | Bipolar Disorder | Interaction (vs SSRI) | SARI | -0.231 | 0.234 | -0.987 | 0.793 | 0.501 | 1.256 | 3.24e-01 | 1.00e+00 | 1791 | 1581 |
| Latino/Mixed-American | No Recorded Depression | Bipolar Disorder | Interaction (vs SSRI) | NDRI | -0.131 | 0.226 | -0.580 | 0.877 | 0.564 | 1.365 | 5.62e-01 | 1.00e+00 | 1791 | 1581 |
| Latino/Mixed-American | No Recorded Depression | Bipolar Disorder | Joint_Interaction_Test | | | | 2.792 | | | | 5.93e-01 | 1.00e+00 | 1791 | 1581 |
| Latino/Mixed-American | No Recorded Depression | BMI | Within-class slope | SSRI | 0.098 | 0.091 | 1.071 | 1.103 | 0.922 | 1.319 | 2.84e-01 | 1.00e+00 | 1791 | 1581 |
| Latino/Mixed-American | No Recorded Depression | BMI | Within-class slope | SNRI | 0.027 | 0.134 | 0.201 | 1.027 | 0.790 | 1.336 | 8.41e-01 | 1.00e+00 | 1791 | 1581 |
| Latino/Mixed-American | No Recorded Depression | BMI | Within-class slope | TCA | -0.192 | 0.143 | -1.350 | 0.825 | 0.624 | 1.091 | 1.77e-01 | 1.00e+00 | 1791 | 1581 |
| Latino/Mixed-American | No Recorded Depression | BMI | Within-class slope | SARI | -0.038 | 0.186 | -0.204 | 0.963 | 0.669 | 1.386 | 8.38e-01 | 1.00e+00 | 1791 | 1581 |
| Latino/Mixed-American | No Recorded Depression | BMI | Within-class slope | NDRI | 0.139 | 0.207 | 0.672 | 1.149 | 0.766 | 1.725 | 5.02e-01 | 1.00e+00 | 1791 | 1581 |
| Latino/Mixed-American | No Recorded Depression | BMI | Interaction (vs SSRI) | SNRI | -0.071 | 0.157 | -0.452 | 0.932 | 0.685 | 1.267 | 6.51e-01 | 1.00e+00 | 1791 | 1581 |
| Latino/Mixed-American | No Recorded Depression | BMI | Interaction (vs SSRI) | TCA | -0.290 | 0.162 | -1.791 | 0.748 | 0.544 | 1.028 | 7.33e-02 | 5.13e-01 | 1791 | 1581 |
| Latino/Mixed-American | No Recorded Depression | BMI | Interaction (vs SSRI) | SARI | -0.136 | 0.199 | -0.683 | 0.873 | 0.592 | 1.289 | 4.94e-01 | 1.00e+00 | 1791 | 1581 |

| | | | | | | | | | | | | | | |
|---|---|---|---|---|---|---|---|---|---|---|---|---|---|---|
| Latino/Mixed-American | No Recorded Depression | BMI | Interaction (vs SSRI) | NDRI | 0.041 | 0.217 | 0.190 | 1.042 | 0.681 | 1.596 | 8.49e-01 | 1.00e+00 | 1791 | 1581 |
| Latino/Mixed-American | No Recorded Depression | BMI | Joint_Interaction_Test | | | | 3.603 | | | | 4.62e-01 | 1.00e+00 | 1791 | 1581 |
| Latino/Mixed-American | Recorded Anxiety | ADHD | Within-class slope | SSRI | 0.105 | 0.053 | 1.957 | 1.110 | 1.000 | 1.233 | 5.04e-02 | 3.53e-01 | 3514 | 2692 |
| Latino/Mixed-American | Recorded Anxiety | ADHD | Within-class slope | SNRI | 0.034 | 0.091 | 0.378 | 1.035 | 0.866 | 1.237 | 7.05e-01 | 1.00e+00 | 3514 | 2692 |
| Latino/Mixed-American | Recorded Anxiety | ADHD | Within-class slope | TCA | -0.031 | 0.106 | -0.289 | 0.970 | 0.789 | 1.193 | 7.72e-01 | 1.00e+00 | 3514 | 2692 |
| Latino/Mixed-American | Recorded Anxiety | ADHD | Within-class slope | NaSSA | 0.275 | 0.212 | 1.301 | 1.317 | 0.870 | 1.995 | 1.93e-01 | 1.00e+00 | 3514 | 2692 |
| Latino/Mixed-American | Recorded Anxiety | ADHD | Within-class slope | SARI | -0.046 | 0.131 | -0.350 | 0.955 | 0.738 | 1.236 | 7.27e-01 | 1.00e+00 | 3514 | 2692 |
| Latino/Mixed-American | Recorded Anxiety | ADHD | Within-class slope | NDRI | 0.060 | 0.115 | 0.526 | 1.062 | 0.848 | 1.329 | 5.99e-01 | 1.00e+00 | 3514 | 2692 |
| Latino/Mixed-American | Recorded Anxiety | ADHD | Interaction (vs SSRI) | SNRI | -0.070 | 0.101 | -0.693 | 0.932 | 0.764 | 1.137 | 4.88e-01 | 1.00e+00 | 3514 | 2692 |
| Latino/Mixed-American | Recorded Anxiety | ADHD | Interaction (vs SSRI) | TCA | -0.135 | 0.114 | -1.184 | 0.874 | 0.699 | 1.093 | 2.36e-01 | 1.00e+00 | 3514 | 2692 |
| Latino/Mixed-American | Recorded Anxiety | ADHD | Interaction (vs SSRI) | NaSSA | 0.171 | 0.218 | 0.786 | 1.186 | 0.775 | 1.817 | 4.32e-01 | 1.00e+00 | 3514 | 2692 |
| Latino/Mixed-American | Recorded Anxiety | ADHD | Interaction (vs SSRI) | SARI | -0.151 | 0.137 | -1.097 | 0.860 | 0.657 | 1.126 | 2.73e-01 | 1.00e+00 | 3514 | 2692 |
| Latino/Mixed-American | Recorded Anxiety | ADHD | Interaction (vs SSRI) | NDRI | -0.044 | 0.120 | -0.369 | 0.957 | 0.756 | 1.211 | 7.12e-01 | 1.00e+00 | 3514 | 2692 |
| Latino/Mixed-American | Recorded Anxiety | ADHD | Joint_Interaction_Test | | | | 3.313 | | | | 6.52e-01 | 1.00e+00 | 3514 | 2692 |
| Latino/Mixed-American | Recorded Anxiety | Migraine | Within-class slope | SSRI | 0.040 | 0.050 | 0.788 | 1.041 | 0.943 | 1.149 | 4.31e-01 | 1.00e+00 | 3514 | 2692 |
| Latino/Mixed-American | Recorded Anxiety | Migraine | Within-class slope | SNRI | 0.104 | 0.087 | 1.204 | 1.110 | 0.937 | 1.316 | 2.29e-01 | 1.00e+00 | 3514 | 2692 |
| Latino/Mixed-American | Recorded Anxiety | Migraine | Within-class slope | TCA | 0.057 | 0.099 | 0.577 | 1.059 | 0.872 | 1.286 | 5.64e-01 | 1.00e+00 | 3514 | 2692 |
| Latino/Mixed-American | Recorded Anxiety | Migraine | Within-class slope | NaSSA | -0.124 | 0.189 | -0.657 | 0.883 | 0.609 | 1.280 | 5.11e-01 | 1.00e+00 | 3514 | 2692 |
| Latino/Mixed-American | Recorded Anxiety | Migraine | Within-class slope | SARI | 0.039 | 0.128 | 0.308 | 1.040 | 0.810 | 1.336 | 7.58e-01 | 1.00e+00 | 3514 | 2692 |
| Latino/Mixed-American | Recorded Anxiety | Migraine | Within-class slope | NDRI | -0.053 | 0.111 | -0.479 | 0.948 | 0.762 | 1.179 | 6.32e-01 | 1.00e+00 | 3514 | 2692 |
| Latino/Mixed-American | Recorded Anxiety | Migraine | Interaction (vs SSRI) | SNRI | 0.065 | 0.096 | 0.675 | 1.067 | 0.884 | 1.287 | 5.00e-01 | 1.00e+00 | 3514 | 2692 |
| Latino/Mixed-American | Recorded Anxiety | Migraine | Interaction (vs SSRI) | TCA | 0.017 | 0.107 | 0.163 | 1.018 | 0.825 | 1.255 | 8.71e-01 | 1.00e+00 | 3514 | 2692 |
| Latino/Mixed-American | Recorded Anxiety | Migraine | Interaction (vs SSRI) | NaSSA | -0.164 | 0.194 | -0.848 | 0.849 | 0.581 | 1.240 | 3.97e-01 | 1.00e+00 | 3514 | 2692 |
| Latino/Mixed-American | Recorded Anxiety | Migraine | Interaction (vs SSRI) | SARI | 0.000 | 0.131 | -0.004 | 1.000 | 0.773 | 1.293 | 9.97e-01 | 1.00e+00 | 3514 | 2692 |
| Latino/Mixed-American | Recorded Anxiety | Migraine | Interaction (vs SSRI) | NDRI | -0.093 | 0.113 | -0.822 | 0.911 | 0.730 | 1.137 | 4.11e-01 | 1.00e+00 | 3514 | 2692 |
| Latino/Mixed-American | Recorded Anxiety | Migraine | Joint_Interaction_Test | | | | 2.147 | | | | 8.28e-01 | 1.00e+00 | 3514 | 2692 |
| Latino/Mixed-American | Recorded Anxiety | Insomnia | Within-class slope | SSRI | 0.063 | 0.056 | 1.129 | 1.065 | 0.955 | 1.188 | 2.59e-01 | 1.00e+00 | 3514 | 2692 |
| Latino/Mixed-American | Recorded Anxiety | Insomnia | Within-class slope | SNRI | 0.023 | 0.096 | 0.241 | 1.023 | 0.847 | 1.237 | 8.10e-01 | 1.00e+00 | 3514 | 2692 |
| Latino/Mixed-American | Recorded Anxiety | Insomnia | Within-class slope | TCA | 0.092 | 0.106 | 0.864 | 1.096 | 0.890 | 1.350 | 3.87e-01 | 1.00e+00 | 3514 | 2692 |
| Latino/Mixed-American | Recorded Anxiety | Insomnia | Within-class slope | NaSSA | 0.294 | 0.232 | 1.268 | 1.342 | 0.852 | 2.115 | 2.05e-01 | 1.00e+00 | 3514 | 2692 |
| Latino/Mixed-American | Recorded Anxiety | Insomnia | Within-class slope | SARI | 0.066 | 0.133 | 0.494 | 1.068 | 0.823 | 1.387 | 6.21e-01 | 1.00e+00 | 3514 | 2692 |
| Latino/Mixed-American | Recorded Anxiety | Insomnia | Within-class slope | NDRI | 0.061 | 0.130 | 0.468 | 1.063 | 0.824 | 1.370 | 6.40e-01 | 1.00e+00 | 3514 | 2692 |
| Latino/Mixed-American | Recorded Anxiety | Insomnia | Interaction (vs SSRI) | SNRI | -0.040 | 0.109 | -0.366 | 0.961 | 0.776 | 1.189 | 7.15e-01 | 1.00e+00 | 3514 | 2692 |
| Latino/Mixed-American | Recorded Anxiety | Insomnia | Interaction (vs SSRI) | TCA | 0.029 | 0.114 | 0.252 | 1.029 | 0.822 | 1.288 | 8.01e-01 | 1.00e+00 | 3514 | 2692 |
| Latino/Mixed-American | Recorded Anxiety | Insomnia | Interaction (vs SSRI) | NaSSA | 0.231 | 0.237 | 0.976 | 1.260 | 0.792 | 2.006 | 3.29e-01 | 1.00e+00 | 3514 | 2692 |
| Latino/Mixed-American | Recorded Anxiety | Insomnia | Interaction (vs SSRI) | SARI | 0.003 | 0.139 | 0.020 | 1.003 | 0.763 | 1.318 | 9.84e-01 | 1.00e+00 | 3514 | 2692 |
| Latino/Mixed-American | Recorded Anxiety | Insomnia | Interaction (vs SSRI) | NDRI | -0.002 | 0.135 | -0.017 | 0.998 | 0.765 | 1.301 | 9.86e-01 | 1.00e+00 | 3514 | 2692 |
| Latino/Mixed-American | Recorded Anxiety | Insomnia | Joint_Interaction_Test | | | | 1.252 | | | | 9.40e-01 | 1.00e+00 | 3514 | 2692 |
| Latino/Mixed-American | Recorded Anxiety | Depression | Within-class slope | SSRI | 0.175 | 0.055 | 3.149 | 1.191 | 1.068 | 1.327 | 1.64e-03 | 1.15e-02 | 3514 | 2692 |
| Latino/Mixed-American | Recorded Anxiety | Depression | Within-class slope | SNRI | 0.280 | 0.095 | 2.936 | 1.323 | 1.097 | 1.594 | 3.33e-03 | 2.33e-02 | 3514 | 2692 |
| Latino/Mixed-American | Recorded Anxiety | Depression | Within-class slope | TCA | 0.159 | 0.111 | 1.435 | 1.173 | 0.943 | 1.457 | 1.51e-01 | 1.00e+00 | 3514 | 2692 |
| Latino/Mixed-American | Recorded Anxiety | Depression | Within-class slope | NaSSA | 0.145 | 0.210 | 0.691 | 1.156 | 0.766 | 1.744 | 4.90e-01 | 1.00e+00 | 3514 | 2692 |
| Latino/Mixed-American | Recorded Anxiety | Depression | Within-class slope | SARI | 0.101 | 0.143 | 0.701 | 1.106 | 0.835 | 1.464 | 4.83e-01 | 1.00e+00 | 3514 | 2692 |
| Latino/Mixed-American | Recorded Anxiety | Depression | Within-class slope | NDRI | 0.005 | 0.122 | 0.040 | 1.005 | 0.791 | 1.277 | 9.68e-01 | 1.00e+00 | 3514 | 2692 |
| Latino/Mixed-American | Recorded Anxiety | Depression | Interaction (vs SSRI) | SNRI | 0.105 | 0.108 | 0.970 | 1.111 | 0.898 | 1.374 | 3.32e-01 | 1.00e+00 | 3514 | 2692 |
| Latino/Mixed-American | Recorded Anxiety | Depression | Interaction (vs SSRI) | TCA | -0.015 | 0.121 | -0.128 | 0.985 | 0.778 | 1.247 | 8.98e-01 | 1.00e+00 | 3514 | 2692 |
| Latino/Mixed-American | Recorded Anxiety | Depression | Interaction (vs SSRI) | NaSSA | -0.030 | 0.214 | -0.139 | 0.971 | 0.639 | 1.475 | 8.90e-01 | 1.00e+00 | 3514 | 2692 |
| Latino/Mixed-American | Recorded Anxiety | Depression | Interaction (vs SSRI) | SARI | -0.074 | 0.148 | -0.499 | 0.929 | 0.694 | 1.242 | 6.18e-01 | 1.00e+00 | 3514 | 2692 |
| Latino/Mixed-American | Recorded Anxiety | Depression | Interaction (vs SSRI) | NDRI | -0.170 | 0.128 | -1.323 | 0.844 | 0.656 | 1.085 | 1.86e-01 | 1.00e+00 | 3514 | 2692 |
| Latino/Mixed-American | Recorded Anxiety | Depression | Joint_Interaction_Test | | | | 3.660 | | | | 5.99e-01 | 1.00e+00 | 3514 | 2692 |
| Latino/Mixed-American | Recorded Anxiety | Schizophrenia | Within-class slope | SSRI | 0.118 | 0.048 | 2.456 | 1.125 | 1.024 | 1.236 | 1.40e-02 | 9.83e-02 | 3514 | 2692 |
| Latino/Mixed-American | Recorded Anxiety | Schizophrenia | Within-class slope | SNRI | 0.100 | 0.091 | 1.103 | 1.105 | 0.925 | 1.320 | 2.70e-01 | 1.00e+00 | 3514 | 2692 |
| Latino/Mixed-American | Recorded Anxiety | Schizophrenia | Within-class slope | TCA | 0.006 | 0.099 | 0.058 | 1.006 | 0.828 | 1.221 | 9.54e-01 | 1.00e+00 | 3514 | 2692 |
| Latino/Mixed-American | Recorded Anxiety | Schizophrenia | Within-class slope | NaSSA | 0.156 | 0.223 | 0.699 | 1.169 | 0.755 | 1.811 | 4.84e-01 | 1.00e+00 | 3514 | 2692 |

| | | | | | | | | | | | | | | |
|---|---|---|---|---|---|---|---|---|---|---|---|---|---|---|
| Latino/Mixed-American | Recorded Anxiety | Schizophrenia | Within-class slope | SARI | -0.022 | 0.122 | -0.179 | 0.979 | 0.771 | 1.242 | 8.58e-01 | 1.00e+00 | 3514 | 2692 |
| Latino/Mixed-American | Recorded Anxiety | Schizophrenia | Within-class slope | NDRI | -0.191 | 0.115 | -1.658 | 0.826 | 0.659 | 1.036 | 9.74e-02 | 6.82e-01 | 3514 | 2692 |
| Latino/Mixed-American | Recorded Anxiety | Schizophrenia | Interaction (vs SSRI) | SNRI | -0.018 | 0.101 | -0.177 | 0.982 | 0.807 | 1.196 | 8.59e-01 | 1.00e+00 | 3514 | 2692 |
| Latino/Mixed-American | Recorded Anxiety | Schizophrenia | Interaction (vs SSRI) | TCA | -0.112 | 0.107 | -1.052 | 0.894 | 0.726 | 1.102 | 2.93e-01 | 1.00e+00 | 3514 | 2692 |
| Latino/Mixed-American | Recorded Anxiety | Schizophrenia | Interaction (vs SSRI) | NaSSA | 0.038 | 0.226 | 0.170 | 1.039 | 0.668 | 1.617 | 8.65e-01 | 1.00e+00 | 3514 | 2692 |
| Latino/Mixed-American | Recorded Anxiety | Schizophrenia | Interaction (vs SSRI) | SARI | -0.140 | 0.127 | -1.099 | 0.870 | 0.678 | 1.115 | 2.72e-01 | 1.00e+00 | 3514 | 2692 |
| Latino/Mixed-American | Recorded Anxiety | Schizophrenia | Interaction (vs SSRI) | NDRI | -0.309 | 0.120 | -2.583 | 0.734 | 0.581 | 0.928 | 9.81e-03 | 6.86e-02 | 3514 | 2692 |
| Latino/Mixed-American | Recorded Anxiety | Schizophrenia | Joint_Interaction_Test | | | | 8.268 | | | | 1.42e-01 | 9.95e-01 | 3514 | 2692 |
| Latino/Mixed-American | Recorded Anxiety | Bipolar Disorder | Within-class slope | SSRI | 0.053 | 0.053 | 1.010 | 1.055 | 0.951 | 1.170 | 3.13e-01 | 1.00e+00 | 3514 | 2692 |
| Latino/Mixed-American | Recorded Anxiety | Bipolar Disorder | Within-class slope | SNRI | 0.119 | 0.097 | 1.230 | 1.127 | 0.932 | 1.363 | 2.19e-01 | 1.00e+00 | 3514 | 2692 |
| Latino/Mixed-American | Recorded Anxiety | Bipolar Disorder | Within-class slope | TCA | 0.199 | 0.112 | 1.771 | 1.220 | 0.979 | 1.521 | 7.66e-02 | 5.36e-01 | 3514 | 2692 |
| Latino/Mixed-American | Recorded Anxiety | Bipolar Disorder | Within-class slope | NaSSA | 0.399 | 0.221 | 1.805 | 1.490 | 0.966 | 2.298 | 7.11e-02 | 4.98e-01 | 3514 | 2692 |
| Latino/Mixed-American | Recorded Anxiety | Bipolar Disorder | Within-class slope | SARI | -0.232 | 0.136 | -1.703 | 0.793 | 0.607 | 1.036 | 8.86e-02 | 6.20e-01 | 3514 | 2692 |
| Latino/Mixed-American | Recorded Anxiety | Bipolar Disorder | Within-class slope | NDRI | -0.044 | 0.132 | -0.335 | 0.957 | 0.738 | 1.240 | 7.38e-01 | 1.00e+00 | 3514 | 2692 |
| Latino/Mixed-American | Recorded Anxiety | Bipolar Disorder | Interaction (vs SSRI) | SNRI | 0.066 | 0.107 | 0.616 | 1.068 | 0.866 | 1.318 | 5.38e-01 | 1.00e+00 | 3514 | 2692 |
| Latino/Mixed-American | Recorded Anxiety | Bipolar Disorder | Interaction (vs SSRI) | TCA | 0.146 | 0.119 | 1.219 | 1.157 | 0.915 | 1.462 | 2.23e-01 | 1.00e+00 | 3514 | 2692 |
| Latino/Mixed-American | Recorded Anxiety | Bipolar Disorder | Interaction (vs SSRI) | NaSSA | 0.345 | 0.222 | 1.552 | 1.412 | 0.913 | 2.184 | 1.21e-01 | 8.44e-01 | 3514 | 2692 |
| Latino/Mixed-American | Recorded Anxiety | Bipolar Disorder | Interaction (vs SSRI) | SARI | -0.285 | 0.140 | -2.043 | 0.752 | 0.572 | 0.988 | 4.10e-02 | 2.87e-01 | 3514 | 2692 |
| Latino/Mixed-American | Recorded Anxiety | Bipolar Disorder | Interaction (vs SSRI) | NDRI | -0.098 | 0.137 | -0.712 | 0.907 | 0.693 | 1.187 | 4.77e-01 | 1.00e+00 | 3514 | 2692 |
| Latino/Mixed-American | Recorded Anxiety | Bipolar Disorder | Joint_Interaction_Test | | | | 10.381 | | | | 6.51e-02 | 4.56e-01 | 3514 | 2692 |
| Latino/Mixed-American | Recorded Anxiety | BMI | Within-class slope | SSRI | 0.044 | 0.050 | 0.891 | 1.045 | 0.948 | 1.153 | 3.73e-01 | 1.00e+00 | 3514 | 2692 |
| Latino/Mixed-American | Recorded Anxiety | BMI | Within-class slope | SNRI | -0.112 | 0.088 | -1.274 | 0.894 | 0.752 | 1.062 | 2.03e-01 | 1.00e+00 | 3514 | 2692 |
| Latino/Mixed-American | Recorded Anxiety | BMI | Within-class slope | TCA | 0.016 | 0.103 | 0.158 | 1.016 | 0.830 | 1.244 | 8.74e-01 | 1.00e+00 | 3514 | 2692 |
| Latino/Mixed-American | Recorded Anxiety | BMI | Within-class slope | NaSSA | 0.028 | 0.195 | 0.144 | 1.029 | 0.702 | 1.507 | 8.85e-01 | 1.00e+00 | 3514 | 2692 |
| Latino/Mixed-American | Recorded Anxiety | BMI | Within-class slope | SARI | -0.023 | 0.117 | -0.196 | 0.977 | 0.776 | 1.230 | 8.44e-01 | 1.00e+00 | 3514 | 2692 |
| Latino/Mixed-American | Recorded Anxiety | BMI | Within-class slope | NDRI | 0.091 | 0.118 | 0.771 | 1.095 | 0.869 | 1.381 | 4.40e-01 | 1.00e+00 | 3514 | 2692 |
| Latino/Mixed-American | Recorded Anxiety | BMI | Interaction (vs SSRI) | SNRI | -0.157 | 0.099 | -1.576 | 0.855 | 0.704 | 1.039 | 1.15e-01 | 8.06e-01 | 3514 | 2692 |
| Latino/Mixed-American | Recorded Anxiety | BMI | Interaction (vs SSRI) | TCA | -0.028 | 0.110 | -0.254 | 0.972 | 0.783 | 1.207 | 7.99e-01 | 1.00e+00 | 3514 | 2692 |
| Latino/Mixed-American | Recorded Anxiety | BMI | Interaction (vs SSRI) | NaSSA | -0.016 | 0.196 | -0.083 | 0.984 | 0.669 | 1.446 | 9.34e-01 | 1.00e+00 | 3514 | 2692 |
| Latino/Mixed-American | Recorded Anxiety | BMI | Interaction (vs SSRI) | SARI | -0.067 | 0.122 | -0.551 | 0.935 | 0.736 | 1.188 | 5.81e-01 | 1.00e+00 | 3514 | 2692 |
| Latino/Mixed-American | Recorded Anxiety | BMI | Interaction (vs SSRI) | NDRI | 0.047 | 0.123 | 0.378 | 1.048 | 0.823 | 1.334 | 7.05e-01 | 1.00e+00 | 3514 | 2692 |
| Latino/Mixed-American | Recorded Anxiety | BMI | Joint_Interaction_Test | | | | 3.071 | | | | 6.89e-01 | 1.00e+00 | 3514 | 2692 |
| Latino/Mixed-American | CIDI-MDD | ADHD | Within-class slope | SSRI | 0.431 | 0.218 | 1.983 | 1.540 | 1.005 | 2.358 | 4.74e-02 | 3.32e-01 | 201 | 187 |
| Latino/Mixed-American | CIDI-MDD | ADHD | Within-class slope | SNRI | -1.106 | 0.345 | -3.202 | 0.331 | 0.168 | 0.651 | 1.37e-03 | 9.56e-03 | 201 | 187 |
| Latino/Mixed-American | CIDI-MDD | ADHD | Interaction (vs SSRI) | SNRI | -1.537 | 0.406 | -3.786 | 0.215 | 0.097 | 0.476 | 1.53e-04 | 1.07e-03 | 201 | 187 |
| Latino/Mixed-American | CIDI-MDD | ADHD | Joint_Interaction_Test | | | | 14.337 | | | | 1.53e-04 | 1.07e-03 | 201 | 187 |
| Latino/Mixed-American | CIDI-MDD | Migraine | Within-class slope | SSRI | 0.152 | 0.172 | 0.882 | 1.164 | 0.831 | 1.631 | 3.78e-01 | 1.00e+00 | 201 | 187 |
| Latino/Mixed-American | CIDI-MDD | Migraine | Within-class slope | SNRI | -0.252 | 0.277 | -0.908 | 0.777 | 0.451 | 1.339 | 3.64e-01 | 1.00e+00 | 201 | 187 |
| Latino/Mixed-American | CIDI-MDD | Migraine | Interaction (vs SSRI) | SNRI | -0.404 | 0.327 | -1.236 | 0.668 | 0.352 | 1.267 | 2.17e-01 | 1.00e+00 | 201 | 187 |
| Latino/Mixed-American | CIDI-MDD | Migraine | Joint_Interaction_Test | | | | 1.527 | | | | 2.17e-01 | 1.00e+00 | 201 | 187 |
| Latino/Mixed-American | CIDI-MDD | Insomnia | Within-class slope | SSRI | 0.317 | 0.234 | 1.356 | 1.373 | 0.868 | 2.171 | 1.75e-01 | 1.00e+00 | 201 | 187 |
| Latino/Mixed-American | CIDI-MDD | Insomnia | Within-class slope | SNRI | -0.386 | 0.384 | -1.007 | 0.680 | 0.320 | 1.442 | 3.14e-01 | 1.00e+00 | 201 | 187 |
| Latino/Mixed-American | CIDI-MDD | Insomnia | Interaction (vs SSRI) | SNRI | -0.703 | 0.470 | -1.495 | 0.495 | 0.197 | 1.244 | 1.35e-01 | 9.44e-01 | 201 | 187 |
| Latino/Mixed-American | CIDI-MDD | Insomnia | Joint_Interaction_Test | | | | 2.236 | | | | 1.35e-01 | 9.44e-01 | 201 | 187 |
| Latino/Mixed-American | CIDI-MDD | Depression | Within-class slope | SSRI | 0.499 | 0.224 | 2.227 | 1.647 | 1.062 | 2.555 | 2.59e-02 | 1.81e-01 | 201 | 187 |
| Latino/Mixed-American | CIDI-MDD | Depression | Within-class slope | SNRI | 0.026 | 0.394 | 0.066 | 1.026 | 0.474 | 2.221 | 9.47e-01 | 1.00e+00 | 201 | 187 |
| Latino/Mixed-American | CIDI-MDD | Depression | Interaction (vs SSRI) | SNRI | -0.473 | 0.471 | -1.004 | 0.623 | 0.248 | 1.569 | 3.15e-01 | 1.00e+00 | 201 | 187 |
| Latino/Mixed-American | CIDI-MDD | Depression | Joint_Interaction_Test | | | | 1.008 | | | | 3.15e-01 | 1.00e+00 | 201 | 187 |
| Latino/Mixed-American | CIDI-MDD | Schizophrenia | Within-class slope | SSRI | 0.270 | 0.202 | 1.339 | 1.310 | 0.882 | 1.946 | 1.81e-01 | 1.00e+00 | 201 | 187 |
| Latino/Mixed-American | CIDI-MDD | Schizophrenia | Within-class slope | SNRI | -0.789 | 0.321 | -2.459 | 0.454 | 0.242 | 0.852 | 1.39e-02 | 9.76e-02 | 201 | 187 |
| Latino/Mixed-American | CIDI-MDD | Schizophrenia | Interaction (vs SSRI) | SNRI | -1.060 | 0.395 | -2.683 | 0.347 | 0.160 | 0.752 | 7.31e-03 | 5.11e-02 | 201 | 187 |
| Latino/Mixed-American | CIDI-MDD | Schizophrenia | Joint_Interaction_Test | | | | 7.196 | | | | 7.31e-03 | 5.11e-02 | 201 | 187 |
| Latino/Mixed-American | CIDI-MDD | Bipolar Disorder | Within-class slope | SSRI | 0.242 | 0.199 | 1.215 | 1.274 | 0.862 | 1.884 | 2.24e-01 | 1.00e+00 | 201 | 187 |
| Latino/Mixed-American | CIDI-MDD | Bipolar Disorder | Within-class slope | SNRI | -0.148 | 0.309 | -0.479 | 0.862 | 0.471 | 1.581 | 6.32e-01 | 1.00e+00 | 201 | 187 |

| | | | | | | | | | | | | | | |
|---|---|---|---|---|---|---|---|---|---|---|---|---|---|---|
| Latino/Mixed-American | CIDI-MDD | Bipolar Disorder | Interaction (vs SSRI) | SNRI | -0.390 | 0.385 | -1.015 | 0.677 | 0.319 | 1.438 | 3.10e-01 | 1.00e+00 | 201 | 187 |
| Latino/Mixed-American | CIDI-MDD | Bipolar Disorder | Joint_Interaction_Test | | | | 1.030 | | | | 3.10e-01 | 1.00e+00 | 201 | 187 |
| Latino/Mixed-American | CIDI-MDD | BMI | Within-class slope | SSRI | 0.233 | 0.214 | 1.087 | 1.262 | 0.830 | 1.920 | 2.77e-01 | 1.00e+00 | 201 | 187 |
| Latino/Mixed-American | CIDI-MDD | BMI | Within-class slope | SNRI | 0.197 | 0.260 | 0.758 | 1.217 | 0.732 | 2.025 | 4.49e-01 | 1.00e+00 | 201 | 187 |
| Latino/Mixed-American | CIDI-MDD | BMI | Interaction (vs SSRI) | SNRI | -0.036 | 0.335 | -0.107 | 0.965 | 0.500 | 1.861 | 9.15e-01 | 1.00e+00 | 201 | 187 |
| Latino/Mixed-American | CIDI-MDD | BMI | Joint_Interaction_Test | | | | 0.012 | | | | 9.15e-01 | 1.00e+00 | 201 | 187 |

**Supplementary Table 14. Multivariable logistic regression results for PGS × drug-class interactions on antidepressant switching versus continuation in the Pharmlines cohort (European ancestry participants).** Each model estimates the association between one PGS trait and switching (versus continuation), together with its interaction with antidepressant drug class, fitted separately within each clinical group (Full Cohort, and subgroups defined by recorded depression status). The outcome is modelled as switch versus continue ~ PGS × DrugClass + covariates, with SSRI as the reference drug class. Because a participant can contribute switching/continuation observations for multiple drug classes, standard errors are cluster-robust, clustered on individual. Within each model, one PGS exposure is modelled alongside a common covariate set (age at first antidepressant, genetic sex, income, and educational attainment). All PGS were residualised on the top 20 within-European-ancestry principal components and the residuals standardised to mean 0, SD 1; each odds ratio therefore reflects the change in odds per one-standard-deviation increase in the PC-adjusted PGS. Because PCs were regressed out at this stage, they are not included in the model covariate set. For each PGS × clinical group model, three types of estimate are reported (type column): Within-class slope = the PGS–switching association within each drug class (the simple slope, i.e. the PGS effect on switching for that class); Interaction (vs SSRI) = the difference in the PGS slope for each class relative to the SSRI reference class; and Joint_Interaction_Test = an omnibus test of whether the PGS slope differs across drug classes (all interaction terms jointly zero), for which the statistic column reports the test statistic and no odds ratio is defined. Columns: Population = inferred genetic ancestry (all European); ClinicalGroup = cohort or clinical subgroup; PGS = the PGS trait for that model; type = estimate type (within-class slope, interaction vs SSRI, or joint interaction test); DrugClass = the antidepressant class for that term; estimate = model coefficient on the log-odds scale; std.error = cluster-robust standard error; statistic = z-statistic (or joint-test statistic for Joint_Interaction_Test rows); odds_ratio = exponentiated coefficient; lower_95/upper_95 = 95% confidence interval bounds for the odds ratio; p_value = nominal (uncorrected) p-value; Bonf_P = Bonferroni-corrected p-value (capped at 1.00); N_Total = number of person–drug-class observations in the model; N_Persons = number of unique individuals contributing observations. Odds ratios >1 (within-class slopes) indicate higher odds of switching per one-SD increase in PGS; for interaction terms, odds ratios >1 indicate a steeper PGS–switching slope relative to SSRI. Bonferroni correction was applied within each specification (clinical group) for the max 7 PGS traits tested.

| Population | ClinicalGroup | PGS | type | DrugClass | estimate | std.error | statistic | odds_ratio | lower_95 | upper_95 | N_Total | N_Persons | p_value | Bonf_P |
|---|---|---|---|---|---|---|---|---|---|---|---|---|---|---|
| European | Full Cohort | ADHD | Within-class slope | SSRI | 0.079 | 0.041 | 1.918 | 1.082 | 0.998 | 1.173 | 6383 | 5439 | 5.51e-02 | 3.86e-01 |
| European | Full Cohort | ADHD | Within-class slope | SNRI | 0.057 | 0.076 | 0.754 | 1.059 | 0.912 | 1.230 | 6383 | 5439 | 4.51e-01 | 1.00e+00 |
| European | Full Cohort | ADHD | Within-class slope | TCA | 0.000 | 0.062 | -0.001 | 1.000 | 0.885 | 1.130 | 6383 | 5439 | 9.99e-01 | 1.00e+00 |
| European | Full Cohort | ADHD | Within-class slope | NaSSA | -0.009 | 0.094 | -0.092 | 0.991 | 0.824 | 1.193 | 6383 | 5439 | 9.27e-01 | 1.00e+00 |
| European | Full Cohort | ADHD | Within-class slope | NDRI | -0.058 | 0.165 | -0.352 | 0.944 | 0.683 | 1.304 | 6383 | 5439 | 7.25e-01 | 1.00e+00 |
| European | Full Cohort | ADHD | Interaction (vs SSRI) | SNRI | -0.021 | 0.081 | -0.266 | 0.979 | 0.836 | 1.146 | 6383 | 5439 | 7.90e-01 | 1.00e+00 |
| European | Full Cohort | ADHD | Interaction (vs SSRI) | TCA | -0.079 | 0.068 | -1.158 | 0.924 | 0.808 | 1.056 | 6383 | 5439 | 2.47e-01 | 1.00e+00 |
| European | Full Cohort | ADHD | Interaction (vs SSRI) | NaSSA | -0.088 | 0.101 | -0.869 | 0.916 | 0.752 | 1.116 | 6383 | 5439 | 3.85e-01 | 1.00e+00 |
| European | Full Cohort | ADHD | Interaction (vs SSRI) | NDRI | -0.137 | 0.167 | -0.822 | 0.872 | 0.629 | 1.209 | 6383 | 5439 | 4.11e-01 | 1.00e+00 |
| European | Full Cohort | ADHD | Joint_Interaction_Test | | | | 2.172 | | | | 6383 | 5439 | 7.04e-01 | 1.00e+00 |
| European | Full Cohort | Bipolar Disorder | Within-class slope | SSRI | 0.082 | 0.041 | 2.005 | 1.086 | 1.002 | 1.176 | 6383 | 5439 | 4.50e-02 | 3.15e-01 |
| European | Full Cohort | Bipolar Disorder | Within-class slope | SNRI | 0.157 | 0.074 | 2.123 | 1.170 | 1.012 | 1.352 | 6383 | 5439 | 3.38e-02 | 2.37e-01 |
| European | Full Cohort | Bipolar Disorder | Within-class slope | TCA | 0.196 | 0.066 | 2.980 | 1.216 | 1.069 | 1.383 | 6383 | 5439 | 2.88e-03 | 2.02e-02 |
| European | Full Cohort | Bipolar Disorder | Within-class slope | NaSSA | 0.013 | 0.095 | 0.138 | 1.013 | 0.841 | 1.221 | 6383 | 5439 | 8.90e-01 | 1.00e+00 |
| European | Full Cohort | Bipolar Disorder | Within-class slope | NDRI | -0.212 | 0.168 | -1.265 | 0.809 | 0.582 | 1.123 | 6383 | 5439 | 2.06e-01 | 1.00e+00 |
| European | Full Cohort | Bipolar Disorder | Interaction (vs SSRI) | SNRI | 0.075 | 0.078 | 0.953 | 1.078 | 0.924 | 1.256 | 6383 | 5439 | 3.41e-01 | 1.00e+00 |
| European | Full Cohort | Bipolar Disorder | Interaction (vs SSRI) | TCA | 0.114 | 0.073 | 1.563 | 1.120 | 0.972 | 1.292 | 6383 | 5439 | 1.18e-01 | 8.27e-01 |
| European | Full Cohort | Bipolar Disorder | Interaction (vs SSRI) | NaSSA | -0.069 | 0.100 | -0.690 | 0.933 | 0.767 | 1.135 | 6383 | 5439 | 4.90e-01 | 1.00e+00 |
| European | Full Cohort | Bipolar Disorder | Interaction (vs SSRI) | NDRI | -0.294 | 0.170 | -1.732 | 0.745 | 0.534 | 1.040 | 6383 | 5439 | 8.33e-02 | 5.83e-01 |
| European | Full Cohort | Bipolar Disorder | Joint_Interaction_Test | | | | 7.772 | | | | 6383 | 5439 | 1.00e-01 | 7.02e-01 |
| European | Full Cohort | Migraine | Within-class slope | SSRI | 0.022 | 0.041 | 0.533 | 1.022 | 0.943 | 1.107 | 6383 | 5439 | 5.94e-01 | 1.00e+00 |
| European | Full Cohort | Migraine | Within-class slope | SNRI | 0.007 | 0.070 | 0.097 | 1.007 | 0.878 | 1.155 | 6383 | 5439 | 9.22e-01 | 1.00e+00 |
| European | Full Cohort | Migraine | Within-class slope | TCA | 0.079 | 0.064 | 1.230 | 1.082 | 0.954 | 1.228 | 6383 | 5439 | 2.19e-01 | 1.00e+00 |
| European | Full Cohort | Migraine | Within-class slope | NaSSA | 0.013 | 0.095 | 0.141 | 1.014 | 0.841 | 1.222 | 6383 | 5439 | 8.88e-01 | 1.00e+00 |
| European | Full Cohort | Migraine | Within-class slope | NDRI | -0.322 | 0.177 | -1.815 | 0.725 | 0.512 | 1.026 | 6383 | 5439 | 6.95e-02 | 4.87e-01 |
| European | Full Cohort | Migraine | Interaction (vs SSRI) | SNRI | -0.015 | 0.077 | -0.194 | 0.985 | 0.847 | 1.145 | 6383 | 5439 | 8.46e-01 | 1.00e+00 |
| European | Full Cohort | Migraine | Interaction (vs SSRI) | TCA | 0.057 | 0.071 | 0.811 | 1.059 | 0.922 | 1.216 | 6383 | 5439 | 4.17e-01 | 1.00e+00 |
| European | Full Cohort | Migraine | Interaction (vs SSRI) | NaSSA | -0.008 | 0.101 | -0.083 | 0.992 | 0.814 | 1.208 | 6383 | 5439 | 9.34e-01 | 1.00e+00 |
| European | Full Cohort | Migraine | Interaction (vs SSRI) | NDRI | -0.344 | 0.180 | -1.911 | 0.709 | 0.499 | 1.009 | 6383 | 5439 | 5.60e-02 | 3.92e-01 |
| European | Full Cohort | Migraine | Joint_Interaction_Test | | | | 4.799 | | | | 6383 | 5439 | 3.09e-01 | 1.00e+00 |
| European | Full Cohort | Schizophrenia | Within-class slope | SSRI | 0.065 | 0.040 | 1.621 | 1.067 | 0.987 | 1.154 | 6383 | 5439 | 1.05e-01 | 7.35e-01 |
| European | Full Cohort | Schizophrenia | Within-class slope | SNRI | 0.041 | 0.076 | 0.545 | 1.042 | 0.899 | 1.208 | 6383 | 5439 | 5.86e-01 | 1.00e+00 |
| European | Full Cohort | Schizophrenia | Within-class slope | TCA | 0.076 | 0.063 | 1.207 | 1.079 | 0.954 | 1.220 | 6383 | 5439 | 2.27e-01 | 1.00e+00 |
| European | Full Cohort | Schizophrenia | Within-class slope | NaSSA | 0.072 | 0.096 | 0.749 | 1.075 | 0.890 | 1.298 | 6383 | 5439 | 4.54e-01 | 1.00e+00 |
| European | Full Cohort | Schizophrenia | Within-class slope | NDRI | 0.000 | 0.187 | 0.001 | 1.000 | 0.693 | 1.444 | 6383 | 5439 | 9.99e-01 | 1.00e+00 |
| European | Full Cohort | Schizophrenia | Interaction (vs SSRI) | SNRI | -0.024 | 0.077 | -0.306 | 0.977 | 0.839 | 1.136 | 6383 | 5439 | 7.59e-01 | 1.00e+00 |

| | | | | | | | | | | | | | | |
|---|---|---|---|---|---|---|---|---|---|---|---|---|---|---|
| European | Full Cohort | Schizophrenia | Interaction (vs SSRI) | TCA | 0.011 | 0.070 | 0.154 | 1.011 | 0.881 | 1.160 | 6383 | 5439 | 8.78e-01 | 1.00e+00 |
| European | Full Cohort | Schizophrenia | Interaction (vs SSRI) | NaSSA | 0.007 | 0.100 | 0.072 | 1.007 | 0.827 | 1.226 | 6383 | 5439 | 9.42e-01 | 1.00e+00 |
| European | Full Cohort | Schizophrenia | Interaction (vs SSRI) | NDRI | -0.065 | 0.189 | -0.341 | 0.937 | 0.647 | 1.359 | 6383 | 5439 | 7.33e-01 | 1.00e+00 |
| European | Full Cohort | Schizophrenia | Joint_Interaction_Test | | | | 0.269 | | | | 6383 | 5439 | 9.92e-01 | 1.00e+00 |
| European | Full Cohort | BMI | Within-class slope | SSRI | 0.045 | 0.040 | 1.143 | 1.046 | 0.968 | 1.131 | 6383 | 5439 | 2.53e-01 | 1.00e+00 |
| European | Full Cohort | BMI | Within-class slope | SNRI | -0.017 | 0.073 | -0.239 | 0.983 | 0.852 | 1.134 | 6383 | 5439 | 8.11e-01 | 1.00e+00 |
| European | Full Cohort | BMI | Within-class slope | TCA | -0.129 | 0.061 | -2.132 | 0.879 | 0.780 | 0.990 | 6383 | 5439 | 3.30e-02 | 2.31e-01 |
| European | Full Cohort | BMI | Within-class slope | NaSSA | -0.001 | 0.088 | -0.015 | 0.999 | 0.841 | 1.185 | 6383 | 5439 | 9.88e-01 | 1.00e+00 |
| European | Full Cohort | BMI | Within-class slope | NDRI | -0.267 | 0.184 | -1.451 | 0.766 | 0.534 | 1.098 | 6383 | 5439 | 1.47e-01 | 1.00e+00 |
| European | Full Cohort | BMI | Interaction (vs SSRI) | SNRI | -0.063 | 0.078 | -0.807 | 0.939 | 0.806 | 1.094 | 6383 | 5439 | 4.20e-01 | 1.00e+00 |
| European | Full Cohort | BMI | Interaction (vs SSRI) | TCA | -0.175 | 0.068 | -2.554 | 0.840 | 0.734 | 0.960 | 6383 | 5439 | 1.07e-02 | 7.46e-02 |
| European | Full Cohort | BMI | Interaction (vs SSRI) | NaSSA | -0.047 | 0.094 | -0.498 | 0.954 | 0.794 | 1.147 | 6383 | 5439 | 6.18e-01 | 1.00e+00 |
| European | Full Cohort | BMI | Interaction (vs SSRI) | NDRI | -0.312 | 0.187 | -1.675 | 0.732 | 0.508 | 1.055 | 6383 | 5439 | 9.39e-02 | 6.57e-01 |
| European | Full Cohort | BMI | Joint_Interaction_Test | | | | 8.609 | | | | 6383 | 5439 | 7.17e-02 | 5.02e-01 |
| European | Full Cohort | Depression | Within-class slope | SSRI | 0.118 | 0.041 | 2.874 | 1.125 | 1.038 | 1.219 | 6383 | 5439 | 4.06e-03 | 2.84e-02 |
| European | Full Cohort | Depression | Within-class slope | SNRI | 0.146 | 0.074 | 1.959 | 1.157 | 1.000 | 1.339 | 6383 | 5439 | 5.01e-02 | 3.51e-01 |
| European | Full Cohort | Depression | Within-class slope | TCA | 0.177 | 0.064 | 2.779 | 1.193 | 1.054 | 1.352 | 6383 | 5439 | 5.45e-03 | 3.81e-02 |
| European | Full Cohort | Depression | Within-class slope | NaSSA | 0.064 | 0.091 | 0.702 | 1.066 | 0.891 | 1.275 | 6383 | 5439 | 4.82e-01 | 1.00e+00 |
| European | Full Cohort | Depression | Within-class slope | NDRI | -0.135 | 0.185 | -0.731 | 0.874 | 0.608 | 1.255 | 6383 | 5439 | 4.65e-01 | 1.00e+00 |
| European | Full Cohort | Depression | Interaction (vs SSRI) | SNRI | 0.028 | 0.079 | 0.356 | 1.028 | 0.881 | 1.200 | 6383 | 5439 | 7.22e-01 | 1.00e+00 |
| European | Full Cohort | Depression | Interaction (vs SSRI) | TCA | 0.059 | 0.071 | 0.827 | 1.061 | 0.922 | 1.220 | 6383 | 5439 | 4.08e-01 | 1.00e+00 |
| European | Full Cohort | Depression | Interaction (vs SSRI) | NaSSA | -0.054 | 0.098 | -0.550 | 0.948 | 0.783 | 1.148 | 6383 | 5439 | 5.82e-01 | 1.00e+00 |
| European | Full Cohort | Depression | Interaction (vs SSRI) | NDRI | -0.253 | 0.187 | -1.351 | 0.776 | 0.538 | 1.121 | 6383 | 5439 | 1.77e-01 | 1.00e+00 |
| European | Full Cohort | Depression | Joint_Interaction_Test | | | | 3.561 | | | | 6383 | 5439 | 4.69e-01 | 1.00e+00 |
| European | Full Cohort | Insomnia | Within-class slope | SSRI | 0.096 | 0.041 | 2.328 | 1.101 | 1.015 | 1.193 | 6383 | 5439 | 1.99e-02 | 1.39e-01 |
| European | Full Cohort | Insomnia | Within-class slope | SNRI | 0.031 | 0.079 | 0.387 | 1.031 | 0.883 | 1.204 | 6383 | 5439 | 6.99e-01 | 1.00e+00 |
| European | Full Cohort | Insomnia | Within-class slope | TCA | 0.157 | 0.064 | 2.458 | 1.170 | 1.032 | 1.326 | 6383 | 5439 | 1.40e-02 | 9.78e-02 |
| European | Full Cohort | Insomnia | Within-class slope | NaSSA | 0.085 | 0.095 | 0.899 | 1.089 | 0.904 | 1.311 | 6383 | 5439 | 3.68e-01 | 1.00e+00 |
| European | Full Cohort | Insomnia | Within-class slope | NDRI | 0.030 | 0.179 | 0.166 | 1.030 | 0.725 | 1.463 | 6383 | 5439 | 8.69e-01 | 1.00e+00 |
| European | Full Cohort | Insomnia | Interaction (vs SSRI) | SNRI | -0.065 | 0.083 | -0.784 | 0.937 | 0.796 | 1.103 | 6383 | 5439 | 4.33e-01 | 1.00e+00 |
| European | Full Cohort | Insomnia | Interaction (vs SSRI) | TCA | 0.061 | 0.071 | 0.864 | 1.063 | 0.925 | 1.221 | 6383 | 5439 | 3.88e-01 | 1.00e+00 |
| European | Full Cohort | Insomnia | Interaction (vs SSRI) | NaSSA | -0.011 | 0.099 | -0.108 | 0.989 | 0.815 | 1.202 | 6383 | 5439 | 9.14e-01 | 1.00e+00 |
| European | Full Cohort | Insomnia | Interaction (vs SSRI) | NDRI | -0.066 | 0.180 | -0.369 | 0.936 | 0.658 | 1.331 | 6383 | 5439 | 7.12e-01 | 1.00e+00 |
| European | Full Cohort | Insomnia | Joint_Interaction_Test | | | | 1.967 | | | | 6383 | 5439 | 7.42e-01 | 1.00e+00 |
| European | Recorded Depression | ADHD | Within-class slope | SSRI | 0.150 | 0.059 | 2.567 | 1.162 | 1.036 | 1.304 | 2737 | 2229 | 1.03e-02 | 7.19e-02 |
| European | Recorded Depression | ADHD | Within-class slope | SNRI | -0.002 | 0.106 | -0.017 | 0.998 | 0.810 | 1.229 | 2737 | 2229 | 9.86e-01 | 1.00e+00 |
| European | Recorded Depression | ADHD | Within-class slope | TCA | -0.169 | 0.102 | -1.655 | 0.845 | 0.692 | 1.032 | 2737 | 2229 | 9.78e-02 | 6.85e-01 |
| European | Recorded Depression | ADHD | Within-class slope | NaSSA | -0.035 | 0.143 | -0.245 | 0.966 | 0.730 | 1.277 | 2737 | 2229 | 8.07e-01 | 1.00e+00 |
| European | Recorded Depression | ADHD | Within-class slope | NDRI | -0.468 | 0.271 | -1.727 | 0.626 | 0.369 | 1.065 | 2737 | 2229 | 8.41e-02 | 5.89e-01 |
| European | Recorded Depression | ADHD | Interaction (vs SSRI) | SNRI | -0.152 | 0.114 | -1.338 | 0.859 | 0.687 | 1.073 | 2737 | 2229 | 1.81e-01 | 1.00e+00 |
| European | Recorded Depression | ADHD | Interaction (vs SSRI) | TCA | -0.319 | 0.108 | -2.963 | 0.727 | 0.588 | 0.898 | 2737 | 2229 | 3.04e-03 | 2.13e-02 |
| European | Recorded Depression | ADHD | Interaction (vs SSRI) | NaSSA | -0.185 | 0.152 | -1.223 | 0.831 | 0.617 | 1.118 | 2737 | 2229 | 2.21e-01 | 1.00e+00 |
| European | Recorded Depression | ADHD | Interaction (vs SSRI) | NDRI | -0.618 | 0.274 | -2.253 | 0.539 | 0.315 | 0.923 | 2737 | 2229 | 2.43e-02 | 1.70e-01 |
| European | Recorded Depression | ADHD | Joint_Interaction_Test | | | | 13.079 | | | | 2737 | 2229 | 1.09e-02 | 7.63e-02 |
| European | Recorded Depression | Bipolar Disorder | Within-class slope | SSRI | 0.085 | 0.059 | 1.436 | 1.089 | 0.969 | 1.223 | 2737 | 2229 | 1.51e-01 | 1.00e+00 |
| European | Recorded Depression | Bipolar Disorder | Within-class slope | SNRI | 0.123 | 0.104 | 1.179 | 1.130 | 0.922 | 1.386 | 2737 | 2229 | 2.39e-01 | 1.00e+00 |
| European | Recorded Depression | Bipolar Disorder | Within-class slope | TCA | 0.271 | 0.105 | 2.582 | 1.312 | 1.068 | 1.612 | 2737 | 2229 | 9.81e-03 | 6.87e-02 |
| European | Recorded Depression | Bipolar Disorder | Within-class slope | NaSSA | -0.142 | 0.145 | -0.978 | 0.867 | 0.652 | 1.153 | 2737 | 2229 | 3.28e-01 | 1.00e+00 |
| European | Recorded Depression | Bipolar Disorder | Within-class slope | NDRI | -0.176 | 0.250 | -0.703 | 0.838 | 0.513 | 1.370 | 2737 | 2229 | 4.82e-01 | 1.00e+00 |
| European | Recorded Depression | Bipolar Disorder | Interaction (vs SSRI) | SNRI | 0.038 | 0.110 | 0.341 | 1.038 | 0.836 | 1.289 | 2737 | 2229 | 7.33e-01 | 1.00e+00 |

| | | | | | | | | | | | | | | |
|---|---|---|---|---|---|---|---|---|---|---|---|---|---|---|
| European | Recorded Depression | Bipolar Disorder | Interaction (vs SSRI) | TCA | 0.187 | 0.116 | 1.613 | 1.205 | 0.961 | 1.511 | 2737 | 2229 | 1.07e-01 | 7.46e-01 |
| European | Recorded Depression | Bipolar Disorder | Interaction (vs SSRI) | NaSSA | -0.227 | 0.153 | -1.481 | 0.797 | 0.590 | 1.076 | 2737 | 2229 | 1.39e-01 | 9.70e-01 |
| European | Recorded Depression | Bipolar Disorder | Interaction (vs SSRI) | NDRI | -0.261 | 0.256 | -1.021 | 0.770 | 0.467 | 1.272 | 2737 | 2229 | 3.07e-01 | 1.00e+00 |
| European | Recorded Depression | Bipolar Disorder | Joint_Interaction_Test | | | | 7.124 | | | | 2737 | 2229 | 1.29e-01 | 9.06e-01 |
| European | Recorded Depression | Migraine | Within-class slope | SSRI | 0.043 | 0.057 | 0.763 | 1.044 | 0.934 | 1.167 | 2737 | 2229 | 4.46e-01 | 1.00e+00 |
| European | Recorded Depression | Migraine | Within-class slope | SNRI | -0.079 | 0.106 | -0.744 | 0.924 | 0.750 | 1.138 | 2737 | 2229 | 4.57e-01 | 1.00e+00 |
| European | Recorded Depression | Migraine | Within-class slope | TCA | 0.096 | 0.101 | 0.948 | 1.101 | 0.902 | 1.343 | 2737 | 2229 | 3.43e-01 | 1.00e+00 |
| European | Recorded Depression | Migraine | Within-class slope | NaSSA | -0.078 | 0.136 | -0.578 | 0.925 | 0.708 | 1.206 | 2737 | 2229 | 5.63e-01 | 1.00e+00 |
| European | Recorded Depression | Migraine | Within-class slope | NDRI | -0.140 | 0.264 | -0.532 | 0.869 | 0.518 | 1.458 | 2737 | 2229 | 5.95e-01 | 1.00e+00 |
| European | Recorded Depression | Migraine | Interaction (vs SSRI) | SNRI | -0.122 | 0.114 | -1.071 | 0.885 | 0.707 | 1.107 | 2737 | 2229 | 2.84e-01 | 1.00e+00 |
| European | Recorded Depression | Migraine | Interaction (vs SSRI) | TCA | 0.053 | 0.105 | 0.503 | 1.054 | 0.858 | 1.296 | 2737 | 2229 | 6.15e-01 | 1.00e+00 |
| European | Recorded Depression | Migraine | Interaction (vs SSRI) | NaSSA | -0.122 | 0.144 | -0.846 | 0.885 | 0.668 | 1.174 | 2737 | 2229 | 3.98e-01 | 1.00e+00 |
| European | Recorded Depression | Migraine | Interaction (vs SSRI) | NDRI | -0.184 | 0.268 | -0.685 | 0.832 | 0.492 | 1.407 | 2737 | 2229 | 4.93e-01 | 1.00e+00 |
| European | Recorded Depression | Migraine | Joint_Interaction_Test | | | | 2.603 | | | | 2737 | 2229 | 6.26e-01 | 1.00e+00 |
| European | Recorded Depression | Schizophrenia | Within-class slope | SSRI | 0.027 | 0.056 | 0.473 | 1.027 | 0.920 | 1.147 | 2737 | 2229 | 6.36e-01 | 1.00e+00 |
| European | Recorded Depression | Schizophrenia | Within-class slope | SNRI | 0.034 | 0.106 | 0.323 | 1.035 | 0.841 | 1.272 | 2737 | 2229 | 7.47e-01 | 1.00e+00 |
| European | Recorded Depression | Schizophrenia | Within-class slope | TCA | 0.012 | 0.097 | 0.125 | 1.012 | 0.837 | 1.224 | 2737 | 2229 | 9.01e-01 | 1.00e+00 |
| European | Recorded Depression | Schizophrenia | Within-class slope | NaSSA | 0.068 | 0.135 | 0.507 | 1.071 | 0.822 | 1.395 | 2737 | 2229 | 6.12e-01 | 1.00e+00 |
| European | Recorded Depression | Schizophrenia | Within-class slope | NDRI | -0.248 | 0.300 | -0.827 | 0.780 | 0.433 | 1.405 | 2737 | 2229 | 4.08e-01 | 1.00e+00 |
| European | Recorded Depression | Schizophrenia | Interaction (vs SSRI) | SNRI | 0.007 | 0.108 | 0.069 | 1.007 | 0.815 | 1.246 | 2737 | 2229 | 9.45e-01 | 1.00e+00 |
| European | Recorded Depression | Schizophrenia | Interaction (vs SSRI) | TCA | -0.014 | 0.107 | -0.135 | 0.986 | 0.799 | 1.216 | 2737 | 2229 | 8.92e-01 | 1.00e+00 |
| European | Recorded Depression | Schizophrenia | Interaction (vs SSRI) | NaSSA | 0.042 | 0.140 | 0.298 | 1.043 | 0.792 | 1.373 | 2737 | 2229 | 7.66e-01 | 1.00e+00 |
| European | Recorded Depression | Schizophrenia | Interaction (vs SSRI) | NDRI | -0.275 | 0.303 | -0.907 | 0.760 | 0.419 | 1.376 | 2737 | 2229 | 3.64e-01 | 1.00e+00 |
| European | Recorded Depression | Schizophrenia | Joint_Interaction_Test | | | | 0.933 | | | | 2737 | 2229 | 9.20e-01 | 1.00e+00 |
| European | Recorded Depression | BMI | Within-class slope | SSRI | 0.087 | 0.056 | 1.554 | 1.091 | 0.978 | 1.217 | 2737 | 2229 | 1.20e-01 | 8.42e-01 |
| European | Recorded Depression | BMI | Within-class slope | SNRI | -0.076 | 0.107 | -0.711 | 0.927 | 0.752 | 1.143 | 2737 | 2229 | 4.77e-01 | 1.00e+00 |
| European | Recorded Depression | BMI | Within-class slope | TCA | -0.080 | 0.089 | -0.901 | 0.923 | 0.776 | 1.099 | 2737 | 2229 | 3.68e-01 | 1.00e+00 |
| European | Recorded Depression | BMI | Within-class slope | NaSSA | -0.011 | 0.128 | -0.087 | 0.989 | 0.770 | 1.270 | 2737 | 2229 | 9.31e-01 | 1.00e+00 |
| European | Recorded Depression | BMI | Within-class slope | NDRI | -0.122 | 0.265 | -0.462 | 0.885 | 0.526 | 1.488 | 2737 | 2229 | 6.44e-01 | 1.00e+00 |
| European | Recorded Depression | BMI | Interaction (vs SSRI) | SNRI | -0.163 | 0.115 | -1.423 | 0.850 | 0.679 | 1.063 | 2737 | 2229 | 1.55e-01 | 1.00e+00 |
| European | Recorded Depression | BMI | Interaction (vs SSRI) | TCA | -0.167 | 0.101 | -1.651 | 0.846 | 0.694 | 1.032 | 2737 | 2229 | 9.87e-02 | 6.91e-01 |
| European | Recorded Depression | BMI | Interaction (vs SSRI) | NaSSA | -0.098 | 0.139 | -0.708 | 0.907 | 0.691 | 1.189 | 2737 | 2229 | 4.79e-01 | 1.00e+00 |
| European | Recorded Depression | BMI | Interaction (vs SSRI) | NDRI | -0.209 | 0.271 | -0.773 | 0.811 | 0.477 | 1.380 | 2737 | 2229 | 4.40e-01 | 1.00e+00 |
| European | Recorded Depression | BMI | Joint_Interaction_Test | | | | 4.181 | | | | 2737 | 2229 | 3.82e-01 | 1.00e+00 |
| European | Recorded Depression | Depression | Within-class slope | SSRI | 0.138 | 0.059 | 2.358 | 1.148 | 1.024 | 1.287 | 2737 | 2229 | 1.84e-02 | 1.28e-01 |
| European | Recorded Depression | Depression | Within-class slope | SNRI | 0.065 | 0.106 | 0.612 | 1.067 | 0.866 | 1.315 | 2737 | 2229 | 5.40e-01 | 1.00e+00 |
| European | Recorded Depression | Depression | Within-class slope | TCA | 0.084 | 0.097 | 0.865 | 1.087 | 0.900 | 1.314 | 2737 | 2229 | 3.87e-01 | 1.00e+00 |
| European | Recorded Depression | Depression | Within-class slope | NaSSA | -0.011 | 0.139 | -0.080 | 0.989 | 0.753 | 1.299 | 2737 | 2229 | 9.37e-01 | 1.00e+00 |
| European | Recorded Depression | Depression | Within-class slope | NDRI | -0.382 | 0.304 | -1.257 | 0.683 | 0.377 | 1.238 | 2737 | 2229 | 2.09e-01 | 1.00e+00 |
| European | Recorded Depression | Depression | Interaction (vs SSRI) | SNRI | -0.073 | 0.112 | -0.647 | 0.930 | 0.746 | 1.159 | 2737 | 2229 | 5.17e-01 | 1.00e+00 |
| European | Recorded Depression | Depression | Interaction (vs SSRI) | TCA | -0.054 | 0.106 | -0.514 | 0.947 | 0.770 | 1.165 | 2737 | 2229 | 6.07e-01 | 1.00e+00 |
| European | Recorded Depression | Depression | Interaction (vs SSRI) | NaSSA | -0.149 | 0.149 | -1.000 | 0.862 | 0.643 | 1.154 | 2737 | 2229 | 3.17e-01 | 1.00e+00 |
| European | Recorded Depression | Depression | Interaction (vs SSRI) | NDRI | -0.520 | 0.308 | -1.685 | 0.595 | 0.325 | 1.089 | 2737 | 2229 | 9.21e-02 | 6.45e-01 |
| European | Recorded Depression | Depression | Joint_Interaction_Test | | | | 3.767 | | | | 2737 | 2229 | 4.38e-01 | 1.00e+00 |
| European | Recorded Depression | Insomnia | Within-class slope | SSRI | 0.129 | 0.058 | 2.225 | 1.138 | 1.016 | 1.276 | 2737 | 2229 | 2.61e-02 | 1.83e-01 |
| European | Recorded Depression | Insomnia | Within-class slope | SNRI | -0.100 | 0.109 | -0.917 | 0.905 | 0.731 | 1.120 | 2737 | 2229 | 3.59e-01 | 1.00e+00 |
| European | Recorded Depression | Insomnia | Within-class slope | TCA | 0.105 | 0.104 | 1.009 | 1.111 | 0.906 | 1.363 | 2737 | 2229 | 3.13e-01 | 1.00e+00 |
| European | Recorded Depression | Insomnia | Within-class slope | NaSSA | 0.115 | 0.131 | 0.875 | 1.122 | 0.867 | 1.451 | 2737 | 2229 | 3.81e-01 | 1.00e+00 |
| European | Recorded Depression | Insomnia | Within-class slope | NDRI | -0.147 | 0.255 | -0.575 | 0.863 | 0.524 | 1.424 | 2737 | 2229 | 5.65e-01 | 1.00e+00 |
| European | Recorded Depression | Insomnia | Interaction (vs SSRI) | SNRI | -0.229 | 0.116 | -1.970 | 0.795 | 0.633 | 0.999 | 2737 | 2229 | 4.89e-02 | 3.42e-01 |

| | | | | | | | | | | | | | |
|---|---|---|---|---|---|---|---|---|---|---|---|---|---|
| European | Recorded Depression | Insomnia | Interaction (vs SSRI) | TCA | -0.024 | 0.113 | -0.216 | 0.976 | 0.783 | 1.217 | 2737 | 2229 | 8.29e-01 | 1.00e+00 |
| European | Recorded Depression | Insomnia | Interaction (vs SSRI) | NaSSA | -0.015 | 0.138 | -0.105 | 0.986 | 0.752 | 1.292 | 2737 | 2229 | 9.16e-01 | 1.00e+00 |
| European | Recorded Depression | Insomnia | Interaction (vs SSRI) | NDRI | -0.276 | 0.258 | -1.072 | 0.759 | 0.458 | 1.257 | 2737 | 2229 | 2.84e-01 | 1.00e+00 |
| European | Recorded Depression | Insomnia | Joint_Interaction_Test | | | | 4.926 | | | | 2737 | 2229 | 2.95e-01 | 1.00e+00 |
| European | No Recorded Depression | ADHD | Within-class slope | SSRI | -0.013 | 0.059 | -0.216 | 0.987 | 0.879 | 1.109 | 3572 | 3150 | 8.29e-01 | 1.00e+00 |
| European | No Recorded Depression | ADHD | Within-class slope | SNRI | 0.118 | 0.113 | 1.045 | 1.126 | 0.902 | 1.405 | 3572 | 3150 | 2.96e-01 | 1.00e+00 |
| European | No Recorded Depression | ADHD | Within-class slope | TCA | 0.082 | 0.083 | 0.984 | 1.085 | 0.922 | 1.277 | 3572 | 3150 | 3.25e-01 | 1.00e+00 |
| European | No Recorded Depression | ADHD | Within-class slope | NaSSA | -0.102 | 0.138 | -0.740 | 0.903 | 0.690 | 1.183 | 3572 | 3150 | 4.60e-01 | 1.00e+00 |
| European | No Recorded Depression | ADHD | Within-class slope | NDRI | 0.283 | 0.288 | 0.984 | 1.327 | 0.755 | 2.333 | 3572 | 3150 | 3.25e-01 | 1.00e+00 |
| European | No Recorded Depression | ADHD | Interaction (vs SSRI) | SNRI | 0.131 | 0.119 | 1.100 | 1.140 | 0.903 | 1.440 | 3572 | 3150 | 2.71e-01 | 1.00e+00 |
| European | No Recorded Depression | ADHD | Interaction (vs SSRI) | TCA | 0.095 | 0.094 | 1.009 | 1.099 | 0.915 | 1.321 | 3572 | 3150 | 3.13e-01 | 1.00e+00 |
| European | No Recorded Depression | ADHD | Interaction (vs SSRI) | NaSSA | -0.089 | 0.147 | -0.606 | 0.915 | 0.686 | 1.220 | 3572 | 3150 | 5.45e-01 | 1.00e+00 |
| European | No Recorded Depression | ADHD | Interaction (vs SSRI) | NDRI | 0.296 | 0.285 | 1.038 | 1.344 | 0.769 | 2.351 | 3572 | 3150 | 2.99e-01 | 1.00e+00 |
| European | No Recorded Depression | ADHD | Joint_Interaction_Test | | | | 3.746 | | | | 3572 | 3150 | 4.42e-01 | 1.00e+00 |
| European | No Recorded Depression | Bipolar Disorder | Within-class slope | SSRI | 0.054 | 0.059 | 0.915 | 1.055 | 0.940 | 1.184 | 3572 | 3150 | 3.60e-01 | 1.00e+00 |
| European | No Recorded Depression | Bipolar Disorder | Within-class slope | SNRI | 0.192 | 0.106 | 1.819 | 1.212 | 0.985 | 1.491 | 3572 | 3150 | 6.89e-02 | 4.82e-01 |
| European | No Recorded Depression | Bipolar Disorder | Within-class slope | TCA | 0.138 | 0.088 | 1.556 | 1.147 | 0.965 | 1.364 | 3572 | 3150 | 1.20e-01 | 8.38e-01 |
| European | No Recorded Depression | Bipolar Disorder | Within-class slope | NaSSA | 0.040 | 0.133 | 0.298 | 1.040 | 0.802 | 1.349 | 3572 | 3150 | 7.65e-01 | 1.00e+00 |
| European | No Recorded Depression | Bipolar Disorder | Within-class slope | NDRI | -0.180 | 0.232 | -0.773 | 0.836 | 0.530 | 1.318 | 3572 | 3150 | 4.40e-01 | 1.00e+00 |
| European | No Recorded Depression | Bipolar Disorder | Interaction (vs SSRI) | SNRI | 0.139 | 0.114 | 1.218 | 1.149 | 0.919 | 1.436 | 3572 | 3150 | 2.23e-01 | 1.00e+00 |
| European | No Recorded Depression | Bipolar Disorder | Interaction (vs SSRI) | TCA | 0.084 | 0.099 | 0.848 | 1.087 | 0.896 | 1.320 | 3572 | 3150 | 3.96e-01 | 1.00e+00 |
| European | No Recorded Depression | Bipolar Disorder | Interaction (vs SSRI) | NaSSA | -0.014 | 0.139 | -0.102 | 0.986 | 0.751 | 1.294 | 3572 | 3150 | 9.19e-01 | 1.00e+00 |
| European | No Recorded Depression | Bipolar Disorder | Interaction (vs SSRI) | NDRI | -0.233 | 0.234 | -0.998 | 0.792 | 0.501 | 1.252 | 3572 | 3150 | 3.18e-01 | 1.00e+00 |
| European | No Recorded Depression | Bipolar Disorder | Joint_Interaction_Test | | | | 3.317 | | | | 3572 | 3150 | 5.06e-01 | 1.00e+00 |
| European | No Recorded Depression | Migraine | Within-class slope | SSRI | 0.015 | 0.060 | 0.249 | 1.015 | 0.902 | 1.143 | 3572 | 3150 | 8.03e-01 | 1.00e+00 |
| European | No Recorded Depression | Migraine | Within-class slope | SNRI | 0.086 | 0.096 | 0.889 | 1.089 | 0.902 | 1.316 | 3572 | 3150 | 3.74e-01 | 1.00e+00 |
| European | No Recorded Depression | Migraine | Within-class slope | TCA | 0.064 | 0.087 | 0.740 | 1.066 | 0.899 | 1.264 | 3572 | 3150 | 4.59e-01 | 1.00e+00 |
| European | No Recorded Depression | Migraine | Within-class slope | NaSSA | 0.069 | 0.143 | 0.481 | 1.071 | 0.809 | 1.418 | 3572 | 3150 | 6.30e-01 | 1.00e+00 |
| European | No Recorded Depression | Migraine | Within-class slope | NDRI | -0.416 | 0.246 | -1.694 | 0.660 | 0.408 | 1.068 | 3572 | 3150 | 9.03e-02 | 6.32e-01 |
| European | No Recorded Depression | Migraine | Interaction (vs SSRI) | SNRI | 0.071 | 0.108 | 0.651 | 1.073 | 0.868 | 1.327 | 3572 | 3150 | 5.15e-01 | 1.00e+00 |
| European | No Recorded Depression | Migraine | Interaction (vs SSRI) | TCA | 0.049 | 0.102 | 0.484 | 1.050 | 0.861 | 1.282 | 3572 | 3150 | 6.28e-01 | 1.00e+00 |
| European | No Recorded Depression | Migraine | Interaction (vs SSRI) | NaSSA | 0.054 | 0.150 | 0.359 | 1.055 | 0.787 | 1.415 | 3572 | 3150 | 7.20e-01 | 1.00e+00 |
| European | No Recorded Depression | Migraine | Interaction (vs SSRI) | NDRI | -0.431 | 0.249 | -1.733 | 0.650 | 0.399 | 1.058 | 3572 | 3150 | 8.30e-02 | 5.81e-01 |
| European | No Recorded Depression | Migraine | Joint_Interaction_Test | | | | 4.093 | | | | 3572 | 3150 | 3.94e-01 | 1.00e+00 |
| European | No Recorded Depression | Schizophrenia | Within-class slope | SSRI | 0.086 | 0.058 | 1.493 | 1.090 | 0.973 | 1.220 | 3572 | 3150 | 1.36e-01 | 9.49e-01 |
| European | No Recorded Depression | Schizophrenia | Within-class slope | SNRI | 0.071 | 0.111 | 0.642 | 1.074 | 0.864 | 1.335 | 3572 | 3150 | 5.21e-01 | 1.00e+00 |
| European | No Recorded Depression | Schizophrenia | Within-class slope | TCA | 0.102 | 0.086 | 1.185 | 1.108 | 0.935 | 1.312 | 3572 | 3150 | 2.36e-01 | 1.00e+00 |
| European | No Recorded Depression | Schizophrenia | Within-class slope | NaSSA | 0.046 | 0.142 | 0.326 | 1.047 | 0.793 | 1.382 | 3572 | 3150 | 7.45e-01 | 1.00e+00 |
| European | No Recorded Depression | Schizophrenia | Within-class slope | NDRI | 0.309 | 0.265 | 1.165 | 1.362 | 0.810 | 2.291 | 3572 | 3150 | 2.44e-01 | 1.00e+00 |
| European | No Recorded Depression | Schizophrenia | Interaction (vs SSRI) | SNRI | -0.015 | 0.114 | -0.128 | 0.985 | 0.788 | 1.233 | 3572 | 3150 | 8.98e-01 | 1.00e+00 |
| European | No Recorded Depression | Schizophrenia | Interaction (vs SSRI) | TCA | 0.016 | 0.098 | 0.167 | 1.016 | 0.839 | 1.231 | 3572 | 3150 | 8.67e-01 | 1.00e+00 |
| European | No Recorded Depression | Schizophrenia | Interaction (vs SSRI) | NaSSA | -0.040 | 0.148 | -0.269 | 0.961 | 0.719 | 1.284 | 3572 | 3150 | 7.88e-01 | 1.00e+00 |
| European | No Recorded Depression | Schizophrenia | Interaction (vs SSRI) | NDRI | 0.223 | 0.268 | 0.834 | 1.250 | 0.740 | 2.112 | 3572 | 3150 | 4.04e-01 | 1.00e+00 |
| European | No Recorded Depression | Schizophrenia | Joint_Interaction_Test | | | | 0.829 | | | | 3572 | 3150 | 9.34e-01 | 1.00e+00 |
| European | No Recorded Depression | BMI | Within-class slope | SSRI | -0.008 | 0.058 | -0.140 | 0.992 | 0.885 | 1.112 | 3572 | 3150 | 8.89e-01 | 1.00e+00 |
| European | No Recorded Depression | BMI | Within-class slope | SNRI | 0.008 | 0.107 | 0.072 | 1.008 | 0.817 | 1.243 | 3572 | 3150 | 9.43e-01 | 1.00e+00 |
| European | No Recorded Depression | BMI | Within-class slope | TCA | -0.201 | 0.085 | -2.376 | 0.818 | 0.692 | 0.965 | 3572 | 3150 | 1.75e-02 | 1.22e-01 |
| European | No Recorded Depression | BMI | Within-class slope | NaSSA | -0.106 | 0.133 | -0.798 | 0.900 | 0.694 | 1.167 | 3572 | 3150 | 4.25e-01 | 1.00e+00 |
| European | No Recorded Depression | BMI | Within-class slope | NDRI | -0.393 | 0.261 | -1.505 | 0.675 | 0.405 | 1.126 | 3572 | 3150 | 1.32e-01 | 9.27e-01 |
| European | No Recorded Depression | BMI | Interaction (vs SSRI) | SNRI | 0.016 | 0.113 | 0.140 | 1.016 | 0.814 | 1.268 | 3572 | 3150 | 8.88e-01 | 1.00e+00 |

| | | | | | | | | | | | | | | |
|---|---|---|---|---|---|---|---|---|---|---|---|---|---|---|
| European | No Recorded Depression | BMI | Interaction (vs SSRI) | TCA | -0.193 | 0.096 | -2.023 | 0.824 | 0.683 | 0.994 | 3572 | 3150 | 4.31e-02 | 3.01e-01 |
| European | No Recorded Depression | BMI | Interaction (vs SSRI) | NaSSA | -0.098 | 0.140 | -0.697 | 0.907 | 0.689 | 1.194 | 3572 | 3150 | 4.86e-01 | 1.00e+00 |
| European | No Recorded Depression | BMI | Interaction (vs SSRI) | NDRI | -0.385 | 0.261 | -1.474 | 0.681 | 0.408 | 1.135 | 3572 | 3150 | 1.40e-01 | 9.83e-01 |
| European | No Recorded Depression | BMI | Joint_Interaction_Test | | | | 6.536 | | | | 3572 | 3150 | 1.63e-01 | 1.00e+00 |
| European | No Recorded Depression | Depression | Within-class slope | SSRI | 0.092 | 0.060 | 1.543 | 1.096 | 0.975 | 1.232 | 3572 | 3150 | 1.23e-01 | 8.60e-01 |
| European | No Recorded Depression | Depression | Within-class slope | SNRI | 0.174 | 0.109 | 1.590 | 1.190 | 0.960 | 1.473 | 3572 | 3150 | 1.12e-01 | 7.82e-01 |
| European | No Recorded Depression | Depression | Within-class slope | TCA | 0.180 | 0.090 | 1.997 | 1.197 | 1.003 | 1.427 | 3572 | 3150 | 4.59e-02 | 3.21e-01 |
| European | No Recorded Depression | Depression | Within-class slope | NaSSA | 0.074 | 0.136 | 0.544 | 1.077 | 0.825 | 1.407 | 3572 | 3150 | 5.86e-01 | 1.00e+00 |
| European | No Recorded Depression | Depression | Within-class slope | NDRI | -0.070 | 0.280 | -0.250 | 0.932 | 0.538 | 1.614 | 3572 | 3150 | 8.03e-01 | 1.00e+00 |
| European | No Recorded Depression | Depression | Interaction (vs SSRI) | SNRI | 0.082 | 0.117 | 0.701 | 1.085 | 0.863 | 1.364 | 3572 | 3150 | 4.83e-01 | 1.00e+00 |
| European | No Recorded Depression | Depression | Interaction (vs SSRI) | TCA | 0.088 | 0.102 | 0.861 | 1.092 | 0.894 | 1.333 | 3572 | 3150 | 3.89e-01 | 1.00e+00 |
| European | No Recorded Depression | Depression | Interaction (vs SSRI) | NaSSA | -0.018 | 0.143 | -0.124 | 0.982 | 0.743 | 1.300 | 3572 | 3150 | 9.01e-01 | 1.00e+00 |
| European | No Recorded Depression | Depression | Interaction (vs SSRI) | NDRI | -0.162 | 0.281 | -0.575 | 0.851 | 0.490 | 1.476 | 3572 | 3150 | 5.65e-01 | 1.00e+00 |
| European | No Recorded Depression | Depression | Joint_Interaction_Test | | | | 1.603 | | | | 3572 | 3150 | 8.08e-01 | 1.00e+00 |
| European | No Recorded Depression | Insomnia | Within-class slope | SSRI | 0.061 | 0.060 | 1.029 | 1.063 | 0.946 | 1.195 | 3572 | 3150 | 3.04e-01 | 1.00e+00 |
| European | No Recorded Depression | Insomnia | Within-class slope | SNRI | 0.118 | 0.118 | 1.002 | 1.125 | 0.893 | 1.417 | 3572 | 3150 | 3.16e-01 | 1.00e+00 |
| European | No Recorded Depression | Insomnia | Within-class slope | TCA | 0.142 | 0.087 | 1.629 | 1.152 | 0.972 | 1.366 | 3572 | 3150 | 1.03e-01 | 7.23e-01 |
| European | No Recorded Depression | Insomnia | Within-class slope | NaSSA | -0.078 | 0.142 | -0.550 | 0.925 | 0.701 | 1.221 | 3572 | 3150 | 5.82e-01 | 1.00e+00 |
| European | No Recorded Depression | Insomnia | Within-class slope | NDRI | 0.138 | 0.266 | 0.520 | 1.148 | 0.682 | 1.933 | 3572 | 3150 | 6.03e-01 | 1.00e+00 |
| European | No Recorded Depression | Insomnia | Interaction (vs SSRI) | SNRI | 0.057 | 0.123 | 0.460 | 1.058 | 0.832 | 1.347 | 3572 | 3150 | 6.46e-01 | 1.00e+00 |
| European | No Recorded Depression | Insomnia | Interaction (vs SSRI) | TCA | 0.080 | 0.098 | 0.825 | 1.084 | 0.895 | 1.312 | 3572 | 3150 | 4.09e-01 | 1.00e+00 |
| European | No Recorded Depression | Insomnia | Interaction (vs SSRI) | NaSSA | -0.139 | 0.147 | -0.944 | 0.870 | 0.652 | 1.162 | 3572 | 3150 | 3.45e-01 | 1.00e+00 |
| European | No Recorded Depression | Insomnia | Interaction (vs SSRI) | NDRI | 0.077 | 0.264 | 0.292 | 1.080 | 0.644 | 1.812 | 3572 | 3150 | 7.71e-01 | 1.00e+00 |
| European | No Recorded Depression | Insomnia | Joint_Interaction_Test | | | | 2.287 | | | | 3572 | 3150 | 6.83e-01 | 1.00e+00 |

**Supplementary Table 15. Multivariable logistic regression results for PGS × drug interactions on antidepressant switching versus continuation in All of Us, modelled at the individual-antidepressant level.** Each model estimates the association between one PGS trait and switching (versus continuation), together with its interaction with the specific antidepressant, fitted separately within each combination of genetically inferred ancestry (European, African, Latino/Mixed-American) and clinical group (Full Cohort; subgroups defined by recorded depression status, recorded anxiety, and CIDI-SF–derived MDD classification). The CIDI-SF–derived MDD subgroups (CIDI-MDD, CIDI-noMDD) were analysed within European-ancestry participants only. The outcome is modelled as switch versus continue ~ PGS × Drug + covariates, with sertraline (SSRI) as the reference drug. Because a participant can contribute switching/continuation observations for multiple antidepressants, standard errors are cluster-robust, clustered on individual. Within each model, one PGS exposure is modelled alongside a common covariate set (age at first antidepressant, sex at birth, income, and educational attainment). All PGS were residualised on the top 16 global genetic ancestry principal components and the residuals standardised to mean 0, SD 1; each odds ratio therefore reflects the change in odds per one-standard-deviation increase in the PC-adjusted PGS. Because ancestry PCs were regressed out at this stage, they are not included in the model covariate set. For each PGS × clinical group model, three types of estimate are reported (type column): Within-class slope = the PGS–switching association for each individual antidepressant (the simple slope, i.e. the PGS effect on switching from that drug); Interaction (vs SSRI:sertraline) = the difference in the PGS slope for each drug relative to the sertraline reference; and Joint_Interaction_Test = an omnibus test of whether the PGS slope differs across drugs (all interaction terms jointly zero), for which the statistic column reports the test statistic and no odds ratio is defined. Columns: Population = inferred genetic ancestry; ClinicalGroup = cohort or clinical subgroup; Drug = the antidepressant (class:drug) for that term; PGS = the PGS trait for that model; type = estimate type (within-drug slope, interaction vs sertraline, or joint interaction test); estimate = model coefficient on the log-odds scale; std.error = cluster-robust standard error; statistic = z-statistic (or joint-test statistic for Joint_Interaction_Test rows); odds_ratio = exponentiated coefficient; lower_95/upper_95 = 95% confidence interval bounds for the odds ratio; p_value = nominal (uncorrected) p-value; Bonf_P = Bonferroni-corrected p-value (capped at 1.00); N_Total = number of person–antidepressant observations in the model; N_Persons = number of unique individuals contributing observations. Odds ratios >1 (within-drug slopes) indicate higher odds of switching per one-SD increase in PGS; for interaction terms, odds ratios >1 indicate a steeper PGS–switching slope relative to sertraline. Bonferroni correction was applied within each specification (ancestry × clinical group) for the max 7 PGS traits tested.

| Population | ClinicalGroup | Drug | PGS | type | estimate | std.error | statistic | odds_ratio | lower_95 | upper_95 | p_value | Bonf_P | N_Total | N_Persons |
|---|---|---|---|---|---|---|---|---|---|---|---|---|---|---|
| European | Full Cohort | SSRI:sertraline | ADHD | Within-class slope | 0.085 | 0.025 | 3.349 | 1.089 | 1.036 | 1.145 | 8.12e-04 | 5.69e-03 | 47198 | 34667 |
| European | Full Cohort | NaSSA:mirtazapine | ADHD | Within-class slope | 0.057 | 0.062 | 0.913 | 1.058 | 0.937 | 1.196 | 3.61e-01 | 1.00e+00 | 47198 | 34667 |
| European | Full Cohort | NDRI:bupropion | ADHD | Within-class slope | 0.010 | 0.029 | 0.340 | 1.010 | 0.955 | 1.068 | 7.34e-01 | 1.00e+00 | 47198 | 34667 |
| European | Full Cohort | SARI:trazodone | ADHD | Within-class slope | 0.101 | 0.034 | 2.971 | 1.107 | 1.035 | 1.183 | 2.96e-03 | 2.07e-02 | 47198 | 34667 |
| European | Full Cohort | SNRI:duloxetine | ADHD | Within-class slope | 0.045 | 0.032 | 1.378 | 1.046 | 0.981 | 1.114 | 1.68e-01 | 1.00e+00 | 47198 | 34667 |
| European | Full Cohort | SNRI:venlafaxine | ADHD | Within-class slope | 0.033 | 0.035 | 0.957 | 1.034 | 0.966 | 1.106 | 3.38e-01 | 1.00e+00 | 47198 | 34667 |
| European | Full Cohort | SSRI:citalopram | ADHD | Within-class slope | 0.036 | 0.031 | 1.185 | 1.037 | 0.976 | 1.101 | 2.36e-01 | 1.00e+00 | 47198 | 34667 |
| European | Full Cohort | SSRI:escitalopram | ADHD | Within-class slope | 0.117 | 0.030 | 3.968 | 1.125 | 1.061 | 1.192 | 7.26e-05 | 5.08e-04 | 47198 | 34667 |
| European | Full Cohort | SSRI:fluoxetine | ADHD | Within-class slope | 0.027 | 0.031 | 0.866 | 1.027 | 0.966 | 1.092 | 3.86e-01 | 1.00e+00 | 47198 | 34667 |
| European | Full Cohort | SSRI:paroxetine | ADHD | Within-class slope | 0.121 | 0.051 | 2.355 | 1.128 | 1.020 | 1.247 | 1.85e-02 | 1.30e-01 | 47198 | 34667 |
| European | Full Cohort | TCA:amitriptyline | ADHD | Within-class slope | 0.068 | 0.043 | 1.594 | 1.071 | 0.984 | 1.165 | 1.11e-01 | 7.76e-01 | 47198 | 34667 |
| European | Full Cohort | TCA:doxepin | ADHD | Within-class slope | 0.050 | 0.099 | 0.504 | 1.051 | 0.866 | 1.275 | 6.15e-01 | 1.00e+00 | 47198 | 34667 |
| European | Full Cohort | TCA:nortriptyline | ADHD | Within-class slope | 0.003 | 0.053 | 0.051 | 1.003 | 0.904 | 1.112 | 9.59e-01 | 1.00e+00 | 47198 | 34667 |
| European | Full Cohort | NaSSA:mirtazapine | ADHD | Interaction (vs SSRI:sertraline) | -0.028 | 0.067 | -0.428 | 0.972 | 0.853 | 1.107 | 6.69e-01 | 1.00e+00 | 47198 | 34667 |
| European | Full Cohort | NDRI:bupropion | ADHD | Interaction (vs SSRI:sertraline) | -0.076 | 0.037 | -2.068 | 0.927 | 0.863 | 0.996 | 3.86e-02 | 2.71e-01 | 47198 | 34667 |
| European | Full Cohort | SARI:trazodone | ADHD | Interaction (vs SSRI:sertraline) | 0.016 | 0.041 | 0.385 | 1.016 | 0.937 | 1.102 | 7.00e-01 | 1.00e+00 | 47198 | 34667 |
| European | Full Cohort | SNRI:duloxetine | ADHD | Interaction (vs SSRI:sertraline) | -0.041 | 0.040 | -1.019 | 0.960 | 0.888 | 1.038 | 3.08e-01 | 1.00e+00 | 47198 | 34667 |
| European | Full Cohort | SNRI:venlafaxine | ADHD | Interaction (vs SSRI:sertraline) | -0.052 | 0.042 | -1.250 | 0.949 | 0.874 | 1.030 | 2.11e-01 | 1.00e+00 | 47198 | 34667 |
| European | Full Cohort | SSRI:citalopram | ADHD | Interaction (vs SSRI:sertraline) | -0.049 | 0.039 | -1.266 | 0.952 | 0.883 | 1.027 | 2.05e-01 | 1.00e+00 | 47198 | 34667 |
| European | Full Cohort | SSRI:escitalopram | ADHD | Interaction (vs SSRI:sertraline) | 0.032 | 0.038 | 0.848 | 1.033 | 0.959 | 1.112 | 3.96e-01 | 1.00e+00 | 47198 | 34667 |
| European | Full Cohort | SSRI:fluoxetine | ADHD | Interaction (vs SSRI:sertraline) | -0.058 | 0.039 | -1.485 | 0.943 | 0.874 | 1.019 | 1.37e-01 | 9.62e-01 | 47198 | 34667 |
| European | Full Cohort | SSRI:paroxetine | ADHD | Interaction (vs SSRI:sertraline) | 0.035 | 0.056 | 0.625 | 1.036 | 0.928 | 1.156 | 5.32e-01 | 1.00e+00 | 47198 | 34667 |
| European | Full Cohort | TCA:amitriptyline | ADHD | Interaction (vs SSRI:sertraline) | -0.017 | 0.049 | -0.346 | 0.983 | 0.893 | 1.082 | 7.29e-01 | 1.00e+00 | 47198 | 34667 |
| European | Full Cohort | TCA:doxepin | ADHD | Interaction (vs SSRI:sertraline) | -0.036 | 0.102 | -0.351 | 0.965 | 0.791 | 1.178 | 7.26e-01 | 1.00e+00 | 47198 | 34667 |
| European | Full Cohort | TCA:nortriptyline | ADHD | Interaction (vs SSRI:sertraline) | -0.083 | 0.057 | -1.446 | 0.921 | 0.823 | 1.030 | 1.48e-01 | 1.00e+00 | 47198 | 34667 |
| European | Full Cohort | | ADHD | Joint_Interaction_Test | | | 15.328 | | | | 2.24e-01 | 1.00e+00 | 47198 | 34667 |
| European | Full Cohort | SSRI:sertraline | Migraine | Within-class slope | 0.077 | 0.025 | 3.099 | 1.080 | 1.029 | 1.134 | 1.94e-03 | 1.36e-02 | 47198 | 34667 |
| European | Full Cohort | NaSSA:mirtazapine | Migraine | Within-class slope | 0.049 | 0.062 | 0.790 | 1.050 | 0.930 | 1.185 | 4.30e-01 | 1.00e+00 | 47198 | 34667 |
| European | Full Cohort | NDRI:bupropion | Migraine | Within-class slope | 0.096 | 0.028 | 3.372 | 1.100 | 1.041 | 1.163 | 7.45e-04 | 5.22e-03 | 47198 | 34667 |
| European | Full Cohort | SARI:trazodone | Migraine | Within-class slope | 0.006 | 0.033 | 0.168 | 1.006 | 0.942 | 1.073 | 8.67e-01 | 1.00e+00 | 47198 | 34667 |
| European | Full Cohort | SNRI:duloxetine | Migraine | Within-class slope | -0.016 | 0.031 | -0.505 | 0.984 | 0.926 | 1.046 | 6.14e-01 | 1.00e+00 | 47198 | 34667 |
| European | Full Cohort | SNRI:venlafaxine | Migraine | Within-class slope | 0.078 | 0.034 | 2.290 | 1.081 | 1.011 | 1.156 | 2.20e-02 | 1.54e-01 | 47198 | 34667 |
| European | Full Cohort | SSRI:citalopram | Migraine | Within-class slope | 0.033 | 0.030 | 1.090 | 1.034 | 0.974 | 1.097 | 2.76e-01 | 1.00e+00 | 47198 | 34667 |
| European | Full Cohort | SSRI:escitalopram | Migraine | Within-class slope | 0.043 | 0.029 | 1.480 | 1.044 | 0.986 | 1.104 | 1.39e-01 | 9.72e-01 | 47198 | 34667 |
| European | Full Cohort | SSRI:fluoxetine | Migraine | Within-class slope | 0.066 | 0.031 | 2.127 | 1.068 | 1.005 | 1.135 | 3.34e-02 | 2.34e-01 | 47198 | 34667 |
| European | Full Cohort | SSRI:paroxetine | Migraine | Within-class slope | 0.067 | 0.048 | 1.401 | 1.070 | 0.974 | 1.175 | 1.61e-01 | 1.00e+00 | 47198 | 34667 |
| European | Full Cohort | TCA:amitriptyline | Migraine | Within-class slope | 0.045 | 0.042 | 1.084 | 1.046 | 0.964 | 1.135 | 2.78e-01 | 1.00e+00 | 47198 | 34667 |
| European | Full Cohort | TCA:doxepin | Migraine | Within-class slope | 0.153 | 0.093 | 1.642 | 1.165 | 0.971 | 1.398 | 1.01e-01 | 7.04e-01 | 47198 | 34667 |
| European | Full Cohort | TCA:nortriptyline | Migraine | Within-class slope | -0.028 | 0.053 | -0.517 | 0.973 | 0.876 | 1.080 | 6.05e-01 | 1.00e+00 | 47198 | 34667 |
| European | Full Cohort | NaSSA:mirtazapine | Migraine | Interaction (vs SSRI:sertraline) | -0.028 | 0.066 | -0.425 | 0.972 | 0.855 | 1.106 | 6.71e-01 | 1.00e+00 | 47198 | 34667 |
| European | Full Cohort | NDRI:bupropion | Migraine | Interaction (vs SSRI:sertraline) | 0.019 | 0.036 | 0.524 | 1.019 | 0.950 | 1.094 | 6.00e-01 | 1.00e+00 | 47198 | 34667 |
| European | Full Cohort | SARI:trazodone | Migraine | Interaction (vs SSRI:sertraline) | -0.071 | 0.040 | -1.759 | 0.931 | 0.860 | 1.008 | 7.85e-02 | 5.50e-01 | 47198 | 34667 |
| European | Full Cohort | SNRI:duloxetine | Migraine | Interaction (vs SSRI:sertraline) | -0.092 | 0.039 | -2.394 | 0.912 | 0.845 | 0.983 | 1.67e-02 | 1.17e-01 | 47198 | 34667 |
| European | Full Cohort | SNRI:venlafaxine | Migraine | Interaction (vs SSRI:sertraline) | 0.001 | 0.041 | 0.028 | 1.001 | 0.924 | 1.085 | 9.78e-01 | 1.00e+00 | 47198 | 34667 |
| European | Full Cohort | SSRI:citalopram | Migraine | Interaction (vs SSRI:sertraline) | -0.044 | 0.038 | -1.148 | 0.957 | 0.888 | 1.031 | 2.51e-01 | 1.00e+00 | 47198 | 34667 |

| | | | | | | | | | | | | | | |
|---|---|---|---|---|---|---|---|---|---|---|---|---|---|---|
| European | Full Cohort | SSRI:escitalopram | Migraine | Interaction (vs SSRI:sertraline) | -0.034 | 0.037 | -0.918 | 0.966 | 0.898 | 1.040 | 3.59e-01 | 1.00e+00 | 47198 | 34667 |
| European | Full Cohort | SSRI:fluoxetine | Migraine | Interaction (vs SSRI:sertraline) | -0.011 | 0.038 | -0.286 | 0.989 | 0.918 | 1.066 | 7.75e-01 | 1.00e+00 | 47198 | 34667 |
| European | Full Cohort | SSRI:paroxetine | Migraine | Interaction (vs SSRI:sertraline) | -0.009 | 0.053 | -0.178 | 0.991 | 0.893 | 1.099 | 8.59e-01 | 1.00e+00 | 47198 | 34667 |
| European | Full Cohort | TCA:amitriptyline | Migraine | Interaction (vs SSRI:sertraline) | -0.032 | 0.048 | -0.661 | 0.969 | 0.882 | 1.064 | 5.09e-01 | 1.00e+00 | 47198 | 34667 |
| European | Full Cohort | TCA:doxepin | Migraine | Interaction (vs SSRI:sertraline) | 0.076 | 0.096 | 0.791 | 1.079 | 0.894 | 1.302 | 4.29e-01 | 1.00e+00 | 47198 | 34667 |
| European | Full Cohort | TCA:nortriptyline | Migraine | Interaction (vs SSRI:sertraline) | -0.104 | 0.058 | -1.795 | 0.901 | 0.804 | 1.010 | 7.26e-02 | 5.08e-01 | 47198 | 34667 |
| European | Full Cohort | | Migraine | Joint_Interaction_Test | | | 15.520 | | | | 2.14e-01 | 1.00e+00 | 47198 | 34667 |
| European | Full Cohort | SSRI:sertraline | Insomnia | Within-class slope | 0.105 | 0.026 | 4.025 | 1.111 | 1.055 | 1.169 | 5.69e-05 | 3.98e-04 | 47198 | 34667 |
| European | Full Cohort | NaSSA:mirtazapine | Insomnia | Within-class slope | 0.042 | 0.064 | 0.656 | 1.043 | 0.920 | 1.182 | 5.12e-01 | 1.00e+00 | 47198 | 34667 |
| European | Full Cohort | NDRI:bupropion | Insomnia | Within-class slope | 0.074 | 0.029 | 2.542 | 1.077 | 1.017 | 1.140 | 1.10e-02 | 7.71e-02 | 47198 | 34667 |
| European | Full Cohort | SARI:trazodone | Insomnia | Within-class slope | 0.085 | 0.035 | 2.457 | 1.088 | 1.017 | 1.165 | 1.40e-02 | 9.80e-02 | 47198 | 34667 |
| European | Full Cohort | SNRI:duloxetine | Insomnia | Within-class slope | 0.107 | 0.033 | 3.201 | 1.113 | 1.042 | 1.188 | 1.37e-03 | 9.58e-03 | 47198 | 34667 |
| European | Full Cohort | SNRI:venlafaxine | Insomnia | Within-class slope | 0.088 | 0.037 | 2.384 | 1.091 | 1.016 | 1.173 | 1.71e-02 | 1.20e-01 | 47198 | 34667 |
| European | Full Cohort | SSRI:citalopram | Insomnia | Within-class slope | 0.088 | 0.032 | 2.785 | 1.092 | 1.026 | 1.162 | 5.36e-03 | 3.75e-02 | 47198 | 34667 |
| European | Full Cohort | SSRI:escitalopram | Insomnia | Within-class slope | 0.128 | 0.030 | 4.230 | 1.136 | 1.071 | 1.205 | 2.33e-05 | 1.63e-04 | 47198 | 34667 |
| European | Full Cohort | SSRI:fluoxetine | Insomnia | Within-class slope | 0.057 | 0.033 | 1.735 | 1.058 | 0.993 | 1.129 | 8.27e-02 | 5.79e-01 | 47198 | 34667 |
| European | Full Cohort | SSRI:paroxetine | Insomnia | Within-class slope | 0.144 | 0.053 | 2.715 | 1.155 | 1.041 | 1.281 | 6.63e-03 | 4.64e-02 | 47198 | 34667 |
| European | Full Cohort | TCA:amitriptyline | Insomnia | Within-class slope | 0.129 | 0.042 | 3.079 | 1.138 | 1.048 | 1.235 | 2.08e-03 | 1.45e-02 | 47198 | 34667 |
| European | Full Cohort | TCA:doxepin | Insomnia | Within-class slope | 0.249 | 0.100 | 2.494 | 1.283 | 1.055 | 1.561 | 1.26e-02 | 8.85e-02 | 47198 | 34667 |
| European | Full Cohort | TCA:nortriptyline | Insomnia | Within-class slope | 0.134 | 0.057 | 2.360 | 1.143 | 1.023 | 1.277 | 1.83e-02 | 1.28e-01 | 47198 | 34667 |
| European | Full Cohort | NaSSA:mirtazapine | Insomnia | Interaction (vs SSRI:sertraline) | -0.063 | 0.068 | -0.930 | 0.939 | 0.821 | 1.073 | 3.52e-01 | 1.00e+00 | 47198 | 34667 |
| European | Full Cohort | NDRI:bupropion | Insomnia | Interaction (vs SSRI:sertraline) | -0.031 | 0.038 | -0.824 | 0.969 | 0.900 | 1.044 | 4.10e-01 | 1.00e+00 | 47198 | 34667 |
| European | Full Cohort | SARI:trazodone | Insomnia | Interaction (vs SSRI:sertraline) | -0.020 | 0.042 | -0.486 | 0.980 | 0.902 | 1.064 | 6.27e-01 | 1.00e+00 | 47198 | 34667 |
| European | Full Cohort | SNRI:duloxetine | Insomnia | Interaction (vs SSRI:sertraline) | 0.001 | 0.041 | 0.034 | 1.001 | 0.924 | 1.086 | 9.73e-01 | 1.00e+00 | 47198 | 34667 |
| European | Full Cohort | SNRI:venlafaxine | Insomnia | Interaction (vs SSRI:sertraline) | -0.018 | 0.044 | -0.406 | 0.982 | 0.902 | 1.070 | 6.85e-01 | 1.00e+00 | 47198 | 34667 |
| European | Full Cohort | SSRI:citalopram | Insomnia | Interaction (vs SSRI:sertraline) | -0.017 | 0.040 | -0.425 | 0.983 | 0.909 | 1.063 | 6.71e-01 | 1.00e+00 | 47198 | 34667 |
| European | Full Cohort | SSRI:escitalopram | Insomnia | Interaction (vs SSRI:sertraline) | 0.022 | 0.039 | 0.578 | 1.023 | 0.948 | 1.103 | 5.63e-01 | 1.00e+00 | 47198 | 34667 |
| European | Full Cohort | SSRI:fluoxetine | Insomnia | Interaction (vs SSRI:sertraline) | -0.048 | 0.041 | -1.185 | 0.953 | 0.879 | 1.032 | 2.36e-01 | 1.00e+00 | 47198 | 34667 |
| European | Full Cohort | SSRI:paroxetine | Insomnia | Interaction (vs SSRI:sertraline) | 0.039 | 0.058 | 0.662 | 1.039 | 0.927 | 1.165 | 5.08e-01 | 1.00e+00 | 47198 | 34667 |
| European | Full Cohort | TCA:amitriptyline | Insomnia | Interaction (vs SSRI:sertraline) | 0.024 | 0.048 | 0.490 | 1.024 | 0.932 | 1.125 | 6.24e-01 | 1.00e+00 | 47198 | 34667 |
| European | Full Cohort | TCA:doxepin | Insomnia | Interaction (vs SSRI:sertraline) | 0.144 | 0.103 | 1.400 | 1.155 | 0.944 | 1.413 | 1.61e-01 | 1.00e+00 | 47198 | 34667 |
| European | Full Cohort | TCA:nortriptyline | Insomnia | Interaction (vs SSRI:sertraline) | 0.028 | 0.062 | 0.459 | 1.029 | 0.912 | 1.161 | 6.46e-01 | 1.00e+00 | 47198 | 34667 |
| European | Full Cohort | | Insomnia | Joint_Interaction_Test | | | 8.805 | | | | 7.19e-01 | 1.00e+00 | 47198 | 34667 |
| European | Full Cohort | SSRI:sertraline | Depression | Within-class slope | 0.202 | 0.026 | 7.751 | 1.223 | 1.162 | 1.287 | 9.12e-15 | 6.39e-14 | 47198 | 34667 |
| European | Full Cohort | NaSSA:mirtazapine | Depression | Within-class slope | 0.137 | 0.063 | 2.164 | 1.146 | 1.013 | 1.297 | 3.05e-02 | 2.13e-01 | 47198 | 34667 |
| European | Full Cohort | NDRI:bupropion | Depression | Within-class slope | 0.203 | 0.028 | 7.133 | 1.225 | 1.159 | 1.295 | 9.83e-13 | 6.88e-12 | 47198 | 34667 |
| European | Full Cohort | SARI:trazodone | Depression | Within-class slope | 0.228 | 0.034 | 6.655 | 1.257 | 1.175 | 1.344 | 2.83e-11 | 1.98e-10 | 47198 | 34667 |
| European | Full Cohort | SNRI:duloxetine | Depression | Within-class slope | 0.211 | 0.033 | 6.305 | 1.235 | 1.156 | 1.318 | 2.88e-10 | 2.02e-09 | 47198 | 34667 |
| European | Full Cohort | SNRI:venlafaxine | Depression | Within-class slope | 0.213 | 0.037 | 5.832 | 1.237 | 1.152 | 1.329 | 5.48e-09 | 3.84e-08 | 47198 | 34667 |
| European | Full Cohort | SSRI:citalopram | Depression | Within-class slope | 0.124 | 0.032 | 3.853 | 1.132 | 1.063 | 1.205 | 1.17e-04 | 8.17e-04 | 47198 | 34667 |
| European | Full Cohort | SSRI:escitalopram | Depression | Within-class slope | 0.183 | 0.031 | 5.960 | 1.200 | 1.130 | 1.275 | 2.52e-09 | 1.77e-08 | 47198 | 34667 |
| European | Full Cohort | SSRI:fluoxetine | Depression | Within-class slope | 0.118 | 0.032 | 3.661 | 1.126 | 1.057 | 1.199 | 2.52e-04 | 1.76e-03 | 47198 | 34667 |
| European | Full Cohort | SSRI:paroxetine | Depression | Within-class slope | 0.241 | 0.054 | 4.447 | 1.272 | 1.144 | 1.415 | 8.71e-06 | 6.09e-05 | 47198 | 34667 |
| European | Full Cohort | TCA:amitriptyline | Depression | Within-class slope | 0.224 | 0.043 | 5.236 | 1.251 | 1.150 | 1.360 | 1.64e-07 | 1.15e-06 | 47198 | 34667 |
| European | Full Cohort | TCA:doxepin | Depression | Within-class slope | 0.308 | 0.101 | 3.057 | 1.361 | 1.117 | 1.659 | 2.24e-03 | 1.57e-02 | 47198 | 34667 |
| European | Full Cohort | TCA:nortriptyline | Depression | Within-class slope | 0.129 | 0.055 | 2.340 | 1.138 | 1.021 | 1.268 | 1.93e-02 | 1.35e-01 | 47198 | 34667 |
| European | Full Cohort | NaSSA:mirtazapine | Depression | Interaction (vs SSRI:sertraline) | -0.065 | 0.068 | -0.960 | 0.937 | 0.821 | 1.070 | 3.37e-01 | 1.00e+00 | 47198 | 34667 |
| European | Full Cohort | NDRI:bupropion | Depression | Interaction (vs SSRI:sertraline) | 0.001 | 0.037 | 0.039 | 1.001 | 0.931 | 1.077 | 9.69e-01 | 1.00e+00 | 47198 | 34667 |
| European | Full Cohort | SARI:trazodone | Depression | Interaction (vs SSRI:sertraline) | 0.027 | 0.042 | 0.641 | 1.027 | 0.946 | 1.116 | 5.22e-01 | 1.00e+00 | 47198 | 34667 |
| European | Full Cohort | SNRI:duloxetine | Depression | Interaction (vs SSRI:sertraline) | 0.009 | 0.041 | 0.225 | 1.009 | 0.931 | 1.094 | 8.22e-01 | 1.00e+00 | 47198 | 34667 |
| European | Full Cohort | SNRI:venlafaxine | Depression | Interaction (vs SSRI:sertraline) | 0.012 | 0.043 | 0.267 | 1.012 | 0.929 | 1.101 | 7.89e-01 | 1.00e+00 | 47198 | 34667 |
| European | Full Cohort | SSRI:citalopram | Depression | Interaction (vs SSRI:sertraline) | -0.078 | 0.040 | -1.944 | 0.925 | 0.855 | 1.001 | 5.19e-02 | 3.63e-01 | 47198 | 34667 |
| European | Full Cohort | SSRI:escitalopram | Depression | Interaction (vs SSRI:sertraline) | -0.019 | 0.039 | -0.481 | 0.981 | 0.909 | 1.060 | 6.30e-01 | 1.00e+00 | 47198 | 34667 |
| European | Full Cohort | SSRI:fluoxetine | Depression | Interaction (vs SSRI:sertraline) | -0.083 | 0.040 | -2.065 | 0.920 | 0.850 | 0.996 | 3.90e-02 | 2.73e-01 | 47198 | 34667 |
| European | Full Cohort | SSRI:paroxetine | Depression | Interaction (vs SSRI:sertraline) | 0.039 | 0.059 | 0.662 | 1.040 | 0.926 | 1.168 | 5.08e-01 | 1.00e+00 | 47198 | 34667 |
| European | Full Cohort | TCA:amitriptyline | Depression | Interaction (vs SSRI:sertraline) | 0.022 | 0.049 | 0.453 | 1.022 | 0.929 | 1.126 | 6.50e-01 | 1.00e+00 | 47198 | 34667 |
| European | Full Cohort | TCA:doxepin | Depression | Interaction (vs SSRI:sertraline) | 0.107 | 0.104 | 1.032 | 1.113 | 0.908 | 1.363 | 3.02e-01 | 1.00e+00 | 47198 | 34667 |
| European | Full Cohort | TCA:nortriptyline | Depression | Interaction (vs SSRI:sertraline) | -0.072 | 0.060 | -1.202 | 0.930 | 0.827 | 1.047 | 2.29e-01 | 1.00e+00 | 47198 | 34667 |
| European | Full Cohort | | Depression | Joint_Interaction_Test | | | 17.281 | | | | 1.39e-01 | 9.75e-01 | 47198 | 34667 |

| | | | | | | | | | | | | | | |
|---|---|---|---|---|---|---|---|---|---|---|---|---|---|---|
| European | Full Cohort | SSRI:sertraline | Schizophrenia | Within-class slope | 0.044 | 0.025 | 1.768 | 1.045 | 0.995 | 1.098 | 7.70e-02 | 5.39e-01 | 47198 | 34667 |
| European | Full Cohort | NaSSA:mirtazapine | Schizophrenia | Within-class slope | 0.012 | 0.063 | 0.192 | 1.012 | 0.895 | 1.145 | 8.47e-01 | 1.00e+00 | 47198 | 34667 |
| European | Full Cohort | NDRI:bupropion | Schizophrenia | Within-class slope | 0.027 | 0.028 | 0.960 | 1.027 | 0.972 | 1.086 | 3.37e-01 | 1.00e+00 | 47198 | 34667 |
| European | Full Cohort | SARI:trazodone | Schizophrenia | Within-class slope | 0.010 | 0.033 | 0.292 | 1.010 | 0.946 | 1.077 | 7.70e-01 | 1.00e+00 | 47198 | 34667 |
| European | Full Cohort | SNRI:duloxetine | Schizophrenia | Within-class slope | 0.051 | 0.033 | 1.530 | 1.052 | 0.986 | 1.122 | 1.26e-01 | 8.83e-01 | 47198 | 34667 |
| European | Full Cohort | SNRI:venlafaxine | Schizophrenia | Within-class slope | 0.029 | 0.035 | 0.814 | 1.029 | 0.961 | 1.102 | 4.16e-01 | 1.00e+00 | 47198 | 34667 |
| European | Full Cohort | SSRI:citalopram | Schizophrenia | Within-class slope | -0.020 | 0.030 | -0.663 | 0.980 | 0.923 | 1.040 | 5.08e-01 | 1.00e+00 | 47198 | 34667 |
| European | Full Cohort | SSRI:escitalopram | Schizophrenia | Within-class slope | 0.038 | 0.029 | 1.290 | 1.038 | 0.981 | 1.100 | 1.97e-01 | 1.00e+00 | 47198 | 34667 |
| European | Full Cohort | SSRI:fluoxetine | Schizophrenia | Within-class slope | -0.004 | 0.032 | -0.120 | 0.996 | 0.936 | 1.060 | 9.04e-01 | 1.00e+00 | 47198 | 34667 |
| European | Full Cohort | SSRI:paroxetine | Schizophrenia | Within-class slope | 0.009 | 0.051 | 0.173 | 1.009 | 0.914 | 1.114 | 8.62e-01 | 1.00e+00 | 47198 | 34667 |
| European | Full Cohort | TCA:amitriptyline | Schizophrenia | Within-class slope | -0.013 | 0.043 | -0.314 | 0.987 | 0.907 | 1.073 | 7.53e-01 | 1.00e+00 | 47198 | 34667 |
| European | Full Cohort | TCA:doxepin | Schizophrenia | Within-class slope | -0.032 | 0.091 | -0.349 | 0.969 | 0.811 | 1.158 | 7.27e-01 | 1.00e+00 | 47198 | 34667 |
| European | Full Cohort | TCA:nortriptyline | Schizophrenia | Within-class slope | -0.018 | 0.055 | -0.317 | 0.983 | 0.882 | 1.095 | 7.51e-01 | 1.00e+00 | 47198 | 34667 |
| European | Full Cohort | NaSSA:mirtazapine | Schizophrenia | Interaction (vs SSRI:sertraline) | -0.032 | 0.067 | -0.477 | 0.968 | 0.849 | 1.105 | 6.33e-01 | 1.00e+00 | 47198 | 34667 |
| European | Full Cohort | NDRI:bupropion | Schizophrenia | Interaction (vs SSRI:sertraline) | -0.017 | 0.036 | -0.476 | 0.983 | 0.916 | 1.055 | 6.34e-01 | 1.00e+00 | 47198 | 34667 |
| European | Full Cohort | SARI:trazodone | Schizophrenia | Interaction (vs SSRI:sertraline) | -0.035 | 0.040 | -0.855 | 0.966 | 0.892 | 1.046 | 3.93e-01 | 1.00e+00 | 47198 | 34667 |
| European | Full Cohort | SNRI:duloxetine | Schizophrenia | Interaction (vs SSRI:sertraline) | 0.006 | 0.040 | 0.156 | 1.006 | 0.930 | 1.089 | 8.76e-01 | 1.00e+00 | 47198 | 34667 |
| European | Full Cohort | SNRI:venlafaxine | Schizophrenia | Interaction (vs SSRI:sertraline) | -0.016 | 0.042 | -0.372 | 0.985 | 0.907 | 1.069 | 7.10e-01 | 1.00e+00 | 47198 | 34667 |
| European | Full Cohort | SSRI:citalopram | Schizophrenia | Interaction (vs SSRI:sertraline) | -0.064 | 0.038 | -1.691 | 0.938 | 0.870 | 1.010 | 9.08e-02 | 6.36e-01 | 47198 | 34667 |
| European | Full Cohort | SSRI:escitalopram | Schizophrenia | Interaction (vs SSRI:sertraline) | -0.006 | 0.038 | -0.172 | 0.994 | 0.923 | 1.070 | 8.63e-01 | 1.00e+00 | 47198 | 34667 |
| European | Full Cohort | SSRI:fluoxetine | Schizophrenia | Interaction (vs SSRI:sertraline) | -0.048 | 0.039 | -1.225 | 0.953 | 0.883 | 1.029 | 2.21e-01 | 1.00e+00 | 47198 | 34667 |
| European | Full Cohort | SSRI:paroxetine | Schizophrenia | Interaction (vs SSRI:sertraline) | -0.035 | 0.055 | -0.639 | 0.965 | 0.866 | 1.076 | 5.23e-01 | 1.00e+00 | 47198 | 34667 |
| European | Full Cohort | TCA:amitriptyline | Schizophrenia | Interaction (vs SSRI:sertraline) | -0.058 | 0.049 | -1.187 | 0.944 | 0.858 | 1.038 | 2.35e-01 | 1.00e+00 | 47198 | 34667 |
| European | Full Cohort | TCA:doxepin | Schizophrenia | Interaction (vs SSRI:sertraline) | -0.076 | 0.094 | -0.807 | 0.927 | 0.771 | 1.114 | 4.19e-01 | 1.00e+00 | 47198 | 34667 |
| European | Full Cohort | TCA:nortriptyline | Schizophrenia | Interaction (vs SSRI:sertraline) | -0.062 | 0.060 | -1.032 | 0.940 | 0.836 | 1.057 | 3.02e-01 | 1.00e+00 | 47198 | 34667 |
| European | Full Cohort | | Schizophrenia | Joint_Interaction_Test | | | 6.320 | | | | 8.99e-01 | 1.00e+00 | 47198 | 34667 |
| European | Full Cohort | SSRI:sertraline | Bipolar Disorder | Within-class slope | 0.042 | 0.026 | 1.601 | 1.043 | 0.991 | 1.098 | 1.09e-01 | 7.66e-01 | 47198 | 34667 |
| European | Full Cohort | NaSSA:mirtazapine | Bipolar Disorder | Within-class slope | -0.036 | 0.065 | -0.555 | 0.964 | 0.849 | 1.096 | 5.79e-01 | 1.00e+00 | 47198 | 34667 |
| European | Full Cohort | NDRI:bupropion | Bipolar Disorder | Within-class slope | 0.035 | 0.029 | 1.189 | 1.035 | 0.978 | 1.096 | 2.34e-01 | 1.00e+00 | 47198 | 34667 |
| European | Full Cohort | SARI:trazodone | Bipolar Disorder | Within-class slope | 0.092 | 0.035 | 2.643 | 1.096 | 1.024 | 1.174 | 8.22e-03 | 5.76e-02 | 47198 | 34667 |
| European | Full Cohort | SNRI:duloxetine | Bipolar Disorder | Within-class slope | 0.058 | 0.034 | 1.730 | 1.060 | 0.992 | 1.132 | 8.37e-02 | 5.86e-01 | 47198 | 34667 |
| European | Full Cohort | SNRI:venlafaxine | Bipolar Disorder | Within-class slope | 0.035 | 0.036 | 0.979 | 1.036 | 0.965 | 1.112 | 3.28e-01 | 1.00e+00 | 47198 | 34667 |
| European | Full Cohort | SSRI:citalopram | Bipolar Disorder | Within-class slope | 0.062 | 0.031 | 1.972 | 1.064 | 1.000 | 1.131 | 4.86e-02 | 3.40e-01 | 47198 | 34667 |
| European | Full Cohort | SSRI:escitalopram | Bipolar Disorder | Within-class slope | 0.064 | 0.030 | 2.141 | 1.066 | 1.005 | 1.130 | 3.23e-02 | 2.26e-01 | 47198 | 34667 |
| European | Full Cohort | SSRI:fluoxetine | Bipolar Disorder | Within-class slope | -0.006 | 0.032 | -0.170 | 0.995 | 0.933 | 1.060 | 8.65e-01 | 1.00e+00 | 47198 | 34667 |
| European | Full Cohort | SSRI:paroxetine | Bipolar Disorder | Within-class slope | 0.008 | 0.052 | 0.154 | 1.008 | 0.911 | 1.116 | 8.77e-01 | 1.00e+00 | 47198 | 34667 |
| European | Full Cohort | TCA:amitriptyline | Bipolar Disorder | Within-class slope | 0.068 | 0.044 | 1.542 | 1.070 | 0.982 | 1.166 | 1.23e-01 | 8.62e-01 | 47198 | 34667 |
| European | Full Cohort | TCA:doxepin | Bipolar Disorder | Within-class slope | 0.040 | 0.096 | 0.422 | 1.041 | 0.863 | 1.257 | 6.73e-01 | 1.00e+00 | 47198 | 34667 |
| European | Full Cohort | TCA:nortriptyline | Bipolar Disorder | Within-class slope | 0.051 | 0.055 | 0.928 | 1.052 | 0.945 | 1.172 | 3.53e-01 | 1.00e+00 | 47198 | 34667 |
| European | Full Cohort | NaSSA:mirtazapine | Bipolar Disorder | Interaction (vs SSRI:sertraline) | -0.078 | 0.069 | -1.127 | 0.925 | 0.807 | 1.059 | 2.60e-01 | 1.00e+00 | 47198 | 34667 |
| European | Full Cohort | NDRI:bupropion | Bipolar Disorder | Interaction (vs SSRI:sertraline) | -0.007 | 0.038 | -0.194 | 0.993 | 0.922 | 1.069 | 8.46e-01 | 1.00e+00 | 47198 | 34667 |
| European | Full Cohort | SARI:trazodone | Bipolar Disorder | Interaction (vs SSRI:sertraline) | 0.050 | 0.043 | 1.172 | 1.051 | 0.967 | 1.143 | 2.41e-01 | 1.00e+00 | 47198 | 34667 |
| European | Full Cohort | SNRI:duloxetine | Bipolar Disorder | Interaction (vs SSRI:sertraline) | 0.016 | 0.041 | 0.392 | 1.016 | 0.937 | 1.102 | 6.95e-01 | 1.00e+00 | 47198 | 34667 |
| European | Full Cohort | SNRI:venlafaxine | Bipolar Disorder | Interaction (vs SSRI:sertraline) | -0.007 | 0.043 | -0.151 | 0.993 | 0.912 | 1.082 | 8.80e-01 | 1.00e+00 | 47198 | 34667 |
| European | Full Cohort | SSRI:citalopram | Bipolar Disorder | Interaction (vs SSRI:sertraline) | 0.020 | 0.040 | 0.506 | 1.020 | 0.944 | 1.103 | 6.13e-01 | 1.00e+00 | 47198 | 34667 |
| European | Full Cohort | SSRI:escitalopram | Bipolar Disorder | Interaction (vs SSRI:sertraline) | 0.022 | 0.039 | 0.563 | 1.022 | 0.947 | 1.103 | 5.73e-01 | 1.00e+00 | 47198 | 34667 |
| European | Full Cohort | SSRI:fluoxetine | Bipolar Disorder | Interaction (vs SSRI:sertraline) | -0.047 | 0.040 | -1.177 | 0.954 | 0.881 | 1.032 | 2.39e-01 | 1.00e+00 | 47198 | 34667 |
| European | Full Cohort | SSRI:paroxetine | Bipolar Disorder | Interaction (vs SSRI:sertraline) | -0.034 | 0.057 | -0.593 | 0.967 | 0.864 | 1.081 | 5.53e-01 | 1.00e+00 | 47198 | 34667 |
| European | Full Cohort | TCA:amitriptyline | Bipolar Disorder | Interaction (vs SSRI:sertraline) | 0.026 | 0.050 | 0.514 | 1.026 | 0.930 | 1.132 | 6.07e-01 | 1.00e+00 | 47198 | 34667 |
| European | Full Cohort | TCA:doxepin | Bipolar Disorder | Interaction (vs SSRI:sertraline) | -0.001 | 0.099 | -0.015 | 0.999 | 0.823 | 1.212 | 9.88e-01 | 1.00e+00 | 47198 | 34667 |
| European | Full Cohort | TCA:nortriptyline | Bipolar Disorder | Interaction (vs SSRI:sertraline) | 0.009 | 0.060 | 0.153 | 1.009 | 0.897 | 1.135 | 8.79e-01 | 1.00e+00 | 47198 | 34667 |
| European | Full Cohort | | Bipolar Disorder | Joint_Interaction_Test | | | 8.014 | | | | 7.84e-01 | 1.00e+00 | 47198 | 34667 |
| European | Full Cohort | SSRI:sertraline | BMI | Within-class slope | 0.031 | 0.025 | 1.262 | 1.032 | 0.983 | 1.083 | 2.07e-01 | 1.00e+00 | 47198 | 34667 |
| European | Full Cohort | NaSSA:mirtazapine | BMI | Within-class slope | 0.041 | 0.062 | 0.657 | 1.042 | 0.922 | 1.176 | 5.11e-01 | 1.00e+00 | 47198 | 34667 |
| European | Full Cohort | NDRI:bupropion | BMI | Within-class slope | -0.032 | 0.028 | -1.127 | 0.969 | 0.916 | 1.024 | 2.60e-01 | 1.00e+00 | 47198 | 34667 |
| European | Full Cohort | SARI:trazodone | BMI | Within-class slope | 0.047 | 0.033 | 1.425 | 1.048 | 0.983 | 1.118 | 1.54e-01 | 1.00e+00 | 47198 | 34667 |
| European | Full Cohort | SNRI:duloxetine | BMI | Within-class slope | 0.005 | 0.032 | 0.150 | 1.005 | 0.944 | 1.070 | 8.80e-01 | 1.00e+00 | 47198 | 34667 |
| European | Full Cohort | SNRI:venlafaxine | BMI | Within-class slope | 0.019 | 0.035 | 0.556 | 1.019 | 0.953 | 1.091 | 5.78e-01 | 1.00e+00 | 47198 | 34667 |
| European | Full Cohort | SSRI:citalopram | BMI | Within-class slope | 0.025 | 0.030 | 0.863 | 1.026 | 0.968 | 1.087 | 3.88e-01 | 1.00e+00 | 47198 | 34667 |

| | | | | | | | | | | | | | | |
|---|---|---|---|---|---|---|---|---|---|---|---|---|---|---|
| European | Full Cohort | SSRI:escitalopram | BMI | Within-class slope | 0.070 | 0.029 | 2.371 | 1.072 | 1.012 | 1.136 | 1.77e-02 | 1.24e-01 | 47198 | 34667 |
| European | Full Cohort | SSRI:fluoxetine | BMI | Within-class slope | 0.045 | 0.031 | 1.475 | 1.046 | 0.985 | 1.111 | 1.40e-01 | 9.82e-01 | 47198 | 34667 |
| European | Full Cohort | SSRI:paroxetine | BMI | Within-class slope | 0.100 | 0.048 | 2.069 | 1.105 | 1.005 | 1.215 | 3.85e-02 | 2.70e-01 | 47198 | 34667 |
| European | Full Cohort | TCA:amitriptyline | BMI | Within-class slope | 0.074 | 0.042 | 1.758 | 1.077 | 0.992 | 1.169 | 7.87e-02 | 5.51e-01 | 47198 | 34667 |
| European | Full Cohort | TCA:doxepin | BMI | Within-class slope | 0.040 | 0.098 | 0.404 | 1.040 | 0.858 | 1.261 | 6.86e-01 | 1.00e+00 | 47198 | 34667 |
| European | Full Cohort | TCA:nortriptyline | BMI | Within-class slope | 0.065 | 0.055 | 1.182 | 1.067 | 0.958 | 1.187 | 2.37e-01 | 1.00e+00 | 47198 | 34667 |
| European | Full Cohort | NaSSA:mirtazapine | BMI | Interaction (vs SSRI:sertraline) | 0.010 | 0.066 | 0.145 | 1.010 | 0.887 | 1.149 | 8.85e-01 | 1.00e+00 | 47198 | 34667 |
| European | Full Cohort | NDRI:bupropion | BMI | Interaction (vs SSRI:sertraline) | -0.063 | 0.036 | -1.754 | 0.939 | 0.875 | 1.007 | 7.95e-02 | 5.56e-01 | 47198 | 34667 |
| European | Full Cohort | SARI:trazodone | BMI | Interaction (vs SSRI:sertraline) | 0.016 | 0.040 | 0.392 | 1.016 | 0.939 | 1.099 | 6.95e-01 | 1.00e+00 | 47198 | 34667 |
| European | Full Cohort | SNRI:duloxetine | BMI | Interaction (vs SSRI:sertraline) | -0.026 | 0.039 | -0.674 | 0.974 | 0.902 | 1.051 | 5.00e-01 | 1.00e+00 | 47198 | 34667 |
| European | Full Cohort | SNRI:venlafaxine | BMI | Interaction (vs SSRI:sertraline) | -0.012 | 0.041 | -0.286 | 0.988 | 0.911 | 1.072 | 7.75e-01 | 1.00e+00 | 47198 | 34667 |
| European | Full Cohort | SSRI:citalopram | BMI | Interaction (vs SSRI:sertraline) | -0.006 | 0.037 | -0.152 | 0.994 | 0.925 | 1.069 | 8.79e-01 | 1.00e+00 | 47198 | 34667 |
| European | Full Cohort | SSRI:escitalopram | BMI | Interaction (vs SSRI:sertraline) | 0.039 | 0.037 | 1.036 | 1.040 | 0.966 | 1.119 | 3.00e-01 | 1.00e+00 | 47198 | 34667 |
| European | Full Cohort | SSRI:fluoxetine | BMI | Interaction (vs SSRI:sertraline) | 0.014 | 0.038 | 0.374 | 1.014 | 0.942 | 1.092 | 7.08e-01 | 1.00e+00 | 47198 | 34667 |
| European | Full Cohort | SSRI:paroxetine | BMI | Interaction (vs SSRI:sertraline) | 0.069 | 0.053 | 1.299 | 1.071 | 0.966 | 1.189 | 1.94e-01 | 1.00e+00 | 47198 | 34667 |
| European | Full Cohort | TCA:amitriptyline | BMI | Interaction (vs SSRI:sertraline) | 0.043 | 0.048 | 0.896 | 1.044 | 0.950 | 1.146 | 3.70e-01 | 1.00e+00 | 47198 | 34667 |
| European | Full Cohort | TCA:doxepin | BMI | Interaction (vs SSRI:sertraline) | 0.009 | 0.101 | 0.085 | 1.009 | 0.828 | 1.228 | 9.33e-01 | 1.00e+00 | 47198 | 34667 |
| European | Full Cohort | TCA:nortriptyline | BMI | Interaction (vs SSRI:sertraline) | 0.033 | 0.059 | 0.567 | 1.034 | 0.921 | 1.161 | 5.71e-01 | 1.00e+00 | 47198 | 34667 |
| European | Full Cohort | | BMI | Joint_Interaction_Test | | | 12.144 | | | | 4.34e-01 | 1.00e+00 | 47198 | 34667 |
| European | Recorded Depression | SSRI:sertraline | ADHD | Within-class slope | 0.066 | 0.032 | 2.046 | 1.069 | 1.003 | 1.139 | 4.07e-02 | 2.85e-01 | 26002 | 17379 |
| European | Recorded Depression | NaSSA:mirtazapine | ADHD | Within-class slope | 0.020 | 0.090 | 0.225 | 1.020 | 0.855 | 1.218 | 8.22e-01 | 1.00e+00 | 26002 | 17379 |
| European | Recorded Depression | NDRI:bupropion | ADHD | Within-class slope | -0.003 | 0.037 | -0.094 | 0.997 | 0.927 | 1.071 | 9.25e-01 | 1.00e+00 | 26002 | 17379 |
| European | Recorded Depression | SARI:trazodone | ADHD | Within-class slope | 0.015 | 0.057 | 0.260 | 1.015 | 0.907 | 1.136 | 7.95e-01 | 1.00e+00 | 26002 | 17379 |
| European | Recorded Depression | SNRI:duloxetine | ADHD | Within-class slope | -0.003 | 0.045 | -0.068 | 0.997 | 0.913 | 1.089 | 9.45e-01 | 1.00e+00 | 26002 | 17379 |
| European | Recorded Depression | SNRI:venlafaxine | ADHD | Within-class slope | 0.010 | 0.045 | 0.223 | 1.010 | 0.925 | 1.103 | 8.23e-01 | 1.00e+00 | 26002 | 17379 |
| European | Recorded Depression | SSRI:citalopram | ADHD | Within-class slope | 0.085 | 0.040 | 2.100 | 1.088 | 1.006 | 1.178 | 3.57e-02 | 2.50e-01 | 26002 | 17379 |
| European | Recorded Depression | SSRI:escitalopram | ADHD | Within-class slope | 0.093 | 0.039 | 2.406 | 1.097 | 1.017 | 1.183 | 1.61e-02 | 1.13e-01 | 26002 | 17379 |
| European | Recorded Depression | SSRI:fluoxetine | ADHD | Within-class slope | -0.037 | 0.039 | -0.936 | 0.964 | 0.893 | 1.041 | 3.49e-01 | 1.00e+00 | 26002 | 17379 |
| European | Recorded Depression | SSRI:paroxetine | ADHD | Within-class slope | 0.139 | 0.069 | 2.014 | 1.150 | 1.004 | 1.317 | 4.40e-02 | 3.08e-01 | 26002 | 17379 |
| European | Recorded Depression | TCA:amitriptyline | ADHD | Within-class slope | 0.038 | 0.072 | 0.529 | 1.039 | 0.902 | 1.195 | 5.97e-01 | 1.00e+00 | 26002 | 17379 |
| European | Recorded Depression | TCA:doxepin | ADHD | Within-class slope | -0.009 | 0.196 | -0.048 | 0.991 | 0.675 | 1.454 | 9.61e-01 | 1.00e+00 | 26002 | 17379 |
| European | Recorded Depression | TCA:nortriptyline | ADHD | Within-class slope | -0.132 | 0.084 | -1.559 | 0.877 | 0.743 | 1.034 | 1.19e-01 | 8.33e-01 | 26002 | 17379 |
| European | Recorded Depression | NaSSA:mirtazapine | ADHD | Interaction (vs SSRI:sertraline) | -0.046 | 0.095 | -0.483 | 0.955 | 0.793 | 1.151 | 6.29e-01 | 1.00e+00 | 26002 | 17379 |
| European | Recorded Depression | NDRI:bupropion | ADHD | Interaction (vs SSRI:sertraline) | -0.070 | 0.047 | -1.493 | 0.933 | 0.851 | 1.022 | 1.35e-01 | 9.48e-01 | 26002 | 17379 |
| European | Recorded Depression | SARI:trazodone | ADHD | Interaction (vs SSRI:sertraline) | -0.051 | 0.064 | -0.799 | 0.950 | 0.838 | 1.077 | 4.24e-01 | 1.00e+00 | 26002 | 17379 |
| European | Recorded Depression | SNRI:duloxetine | ADHD | Interaction (vs SSRI:sertraline) | -0.069 | 0.053 | -1.296 | 0.933 | 0.840 | 1.036 | 1.95e-01 | 1.00e+00 | 26002 | 17379 |
| European | Recorded Depression | SNRI:venlafaxine | ADHD | Interaction (vs SSRI:sertraline) | -0.056 | 0.054 | -1.046 | 0.945 | 0.851 | 1.050 | 2.95e-01 | 1.00e+00 | 26002 | 17379 |
| European | Recorded Depression | SSRI:citalopram | ADHD | Interaction (vs SSRI:sertraline) | 0.018 | 0.050 | 0.368 | 1.019 | 0.924 | 1.123 | 7.13e-01 | 1.00e+00 | 26002 | 17379 |
| European | Recorded Depression | SSRI:escitalopram | ADHD | Interaction (vs SSRI:sertraline) | 0.026 | 0.049 | 0.544 | 1.027 | 0.933 | 1.129 | 5.86e-01 | 1.00e+00 | 26002 | 17379 |
| European | Recorded Depression | SSRI:fluoxetine | ADHD | Interaction (vs SSRI:sertraline) | -0.103 | 0.049 | -2.091 | 0.902 | 0.819 | 0.994 | 3.65e-02 | 2.56e-01 | 26002 | 17379 |
| European | Recorded Depression | SSRI:paroxetine | ADHD | Interaction (vs SSRI:sertraline) | 0.073 | 0.075 | 0.975 | 1.076 | 0.929 | 1.247 | 3.30e-01 | 1.00e+00 | 26002 | 17379 |
| European | Recorded Depression | TCA:amitriptyline | ADHD | Interaction (vs SSRI:sertraline) | -0.028 | 0.077 | -0.366 | 0.972 | 0.835 | 1.131 | 7.14e-01 | 1.00e+00 | 26002 | 17379 |
| European | Recorded Depression | TCA:doxepin | ADHD | Interaction (vs SSRI:sertraline) | -0.076 | 0.198 | -0.382 | 0.927 | 0.629 | 1.367 | 7.02e-01 | 1.00e+00 | 26002 | 17379 |
| European | Recorded Depression | TCA:nortriptyline | ADHD | Interaction (vs SSRI:sertraline) | -0.198 | 0.089 | -2.234 | 0.821 | 0.690 | 0.976 | 2.55e-02 | 1.78e-01 | 26002 | 17379 |
| European | Recorded Depression | | ADHD | Joint_Interaction_Test | | | 17.085 | | | | 1.46e-01 | 1.00e+00 | 26002 | 17379 |
| European | Recorded Depression | SSRI:sertraline | Migraine | Within-class slope | 0.089 | 0.032 | 2.784 | 1.093 | 1.027 | 1.164 | 5.37e-03 | 3.76e-02 | 26002 | 17379 |
| European | Recorded Depression | NaSSA:mirtazapine | Migraine | Within-class slope | 0.045 | 0.088 | 0.520 | 1.047 | 0.882 | 1.242 | 6.03e-01 | 1.00e+00 | 26002 | 17379 |
| European | Recorded Depression | NDRI:bupropion | Migraine | Within-class slope | 0.127 | 0.036 | 3.522 | 1.136 | 1.058 | 1.219 | 4.28e-04 | 3.00e-03 | 26002 | 17379 |
| European | Recorded Depression | SARI:trazodone | Migraine | Within-class slope | 0.004 | 0.055 | 0.080 | 1.004 | 0.902 | 1.119 | 9.36e-01 | 1.00e+00 | 26002 | 17379 |
| European | Recorded Depression | SNRI:duloxetine | Migraine | Within-class slope | 0.007 | 0.043 | 0.170 | 1.007 | 0.925 | 1.097 | 8.65e-01 | 1.00e+00 | 26002 | 17379 |
| European | Recorded Depression | SNRI:venlafaxine | Migraine | Within-class slope | 0.038 | 0.045 | 0.848 | 1.039 | 0.951 | 1.135 | 3.96e-01 | 1.00e+00 | 26002 | 17379 |
| European | Recorded Depression | SSRI:citalopram | Migraine | Within-class slope | 0.008 | 0.039 | 0.206 | 1.008 | 0.934 | 1.088 | 8.37e-01 | 1.00e+00 | 26002 | 17379 |
| European | Recorded Depression | SSRI:escitalopram | Migraine | Within-class slope | 0.061 | 0.038 | 1.595 | 1.062 | 0.986 | 1.144 | 1.11e-01 | 7.76e-01 | 26002 | 17379 |
| European | Recorded Depression | SSRI:fluoxetine | Migraine | Within-class slope | 0.097 | 0.039 | 2.486 | 1.102 | 1.021 | 1.189 | 1.29e-02 | 9.05e-02 | 26002 | 17379 |
| European | Recorded Depression | SSRI:paroxetine | Migraine | Within-class slope | 0.111 | 0.065 | 1.701 | 1.118 | 0.983 | 1.270 | 8.89e-02 | 6.22e-01 | 26002 | 17379 |
| European | Recorded Depression | TCA:amitriptyline | Migraine | Within-class slope | 0.057 | 0.072 | 0.791 | 1.059 | 0.919 | 1.221 | 4.29e-01 | 1.00e+00 | 26002 | 17379 |
| European | Recorded Depression | TCA:doxepin | Migraine | Within-class slope | 0.402 | 0.198 | 2.033 | 1.495 | 1.015 | 2.204 | 4.21e-02 | 2.95e-01 | 26002 | 17379 |
| European | Recorded Depression | TCA:nortriptyline | Migraine | Within-class slope | -0.051 | 0.087 | -0.591 | 0.950 | 0.801 | 1.127 | 5.55e-01 | 1.00e+00 | 26002 | 17379 |
| European | Recorded Depression | NaSSA:mirtazapine | Migraine | Interaction (vs SSRI:sertraline) | -0.044 | 0.092 | -0.472 | 0.957 | 0.799 | 1.147 | 6.37e-01 | 1.00e+00 | 26002 | 17379 |

| | | | | | | | | | | | | | | |
|---|---|---|---|---|---|---|---|---|---|---|---|---|---|---|
| European | Recorded Depression | NDRI:bupropion | Migraine | Interaction (vs SSRI:sertraline) | 0.038 | 0.046 | 0.826 | 1.039 | 0.949 | 1.137 | 4.09e-01 | 1.00e+00 | 26002 | 17379 |
| European | Recorded Depression | SARI:trazodone | Migraine | Interaction (vs SSRI:sertraline) | -0.085 | 0.063 | -1.352 | 0.919 | 0.813 | 1.039 | 1.76e-01 | 1.00e+00 | 26002 | 17379 |
| European | Recorded Depression | SNRI:duloxetine | Migraine | Interaction (vs SSRI:sertraline) | -0.082 | 0.052 | -1.563 | 0.922 | 0.832 | 1.021 | 1.18e-01 | 8.27e-01 | 26002 | 17379 |
| European | Recorded Depression | SNRI:venlafaxine | Migraine | Interaction (vs SSRI:sertraline) | -0.051 | 0.054 | -0.948 | 0.950 | 0.855 | 1.056 | 3.43e-01 | 1.00e+00 | 26002 | 17379 |
| European | Recorded Depression | SSRI:citalopram | Migraine | Interaction (vs SSRI:sertraline) | -0.081 | 0.049 | -1.656 | 0.922 | 0.838 | 1.015 | 9.77e-02 | 6.84e-01 | 26002 | 17379 |
| European | Recorded Depression | SSRI:escitalopram | Migraine | Interaction (vs SSRI:sertraline) | -0.029 | 0.049 | -0.586 | 0.972 | 0.883 | 1.069 | 5.58e-01 | 1.00e+00 | 26002 | 17379 |
| European | Recorded Depression | SSRI:fluoxetine | Migraine | Interaction (vs SSRI:sertraline) | 0.008 | 0.048 | 0.160 | 1.008 | 0.917 | 1.108 | 8.73e-01 | 1.00e+00 | 26002 | 17379 |
| European | Recorded Depression | SSRI:paroxetine | Migraine | Interaction (vs SSRI:sertraline) | 0.022 | 0.072 | 0.308 | 1.022 | 0.888 | 1.176 | 7.58e-01 | 1.00e+00 | 26002 | 17379 |
| European | Recorded Depression | TCA:amitriptyline | Migraine | Interaction (vs SSRI:sertraline) | -0.032 | 0.078 | -0.407 | 0.969 | 0.831 | 1.129 | 6.84e-01 | 1.00e+00 | 26002 | 17379 |
| European | Recorded Depression | TCA:doxepin | Migraine | Interaction (vs SSRI:sertraline) | 0.313 | 0.200 | 1.564 | 1.368 | 0.924 | 2.026 | 1.18e-01 | 8.24e-01 | 26002 | 17379 |
| European | Recorded Depression | TCA:nortriptyline | Migraine | Interaction (vs SSRI:sertraline) | -0.140 | 0.092 | -1.529 | 0.869 | 0.726 | 1.040 | 1.26e-01 | 8.84e-01 | 26002 | 17379 |
| European | Recorded Depression | | Migraine | Joint_Interaction_Test | | | 15.514 | | | | 2.15e-01 | 1.00e+00 | 26002 | 17379 |
| European | Recorded Depression | SSRI:sertraline | Insomnia | Within-class slope | 0.093 | 0.033 | 2.825 | 1.097 | 1.029 | 1.170 | 4.73e-03 | 3.31e-02 | 26002 | 17379 |
| European | Recorded Depression | NaSSA:mirtazapine | Insomnia | Within-class slope | -0.021 | 0.087 | -0.242 | 0.979 | 0.825 | 1.162 | 8.09e-01 | 1.00e+00 | 26002 | 17379 |
| European | Recorded Depression | NDRI:bupropion | Insomnia | Within-class slope | 0.095 | 0.038 | 2.530 | 1.100 | 1.022 | 1.184 | 1.14e-02 | 7.98e-02 | 26002 | 17379 |
| European | Recorded Depression | SARI:trazodone | Insomnia | Within-class slope | -0.010 | 0.058 | -0.163 | 0.991 | 0.883 | 1.111 | 8.70e-01 | 1.00e+00 | 26002 | 17379 |
| European | Recorded Depression | SNRI:duloxetine | Insomnia | Within-class slope | 0.067 | 0.046 | 1.456 | 1.069 | 0.977 | 1.170 | 1.45e-01 | 1.00e+00 | 26002 | 17379 |
| European | Recorded Depression | SNRI:venlafaxine | Insomnia | Within-class slope | 0.081 | 0.049 | 1.664 | 1.085 | 0.986 | 1.194 | 9.62e-02 | 6.73e-01 | 26002 | 17379 |
| European | Recorded Depression | SSRI:citalopram | Insomnia | Within-class slope | 0.083 | 0.041 | 2.012 | 1.087 | 1.002 | 1.178 | 4.42e-02 | 3.09e-01 | 26002 | 17379 |
| European | Recorded Depression | SSRI:escitalopram | Insomnia | Within-class slope | 0.090 | 0.040 | 2.261 | 1.094 | 1.012 | 1.183 | 2.37e-02 | 1.66e-01 | 26002 | 17379 |
| European | Recorded Depression | SSRI:fluoxetine | Insomnia | Within-class slope | -0.020 | 0.041 | -0.502 | 0.980 | 0.905 | 1.061 | 6.16e-01 | 1.00e+00 | 26002 | 17379 |
| European | Recorded Depression | SSRI:paroxetine | Insomnia | Within-class slope | 0.127 | 0.070 | 1.803 | 1.135 | 0.989 | 1.303 | 7.15e-02 | 5.00e-01 | 26002 | 17379 |
| European | Recorded Depression | TCA:amitriptyline | Insomnia | Within-class slope | 0.112 | 0.073 | 1.537 | 1.118 | 0.970 | 1.289 | 1.24e-01 | 8.70e-01 | 26002 | 17379 |
| European | Recorded Depression | TCA:doxepin | Insomnia | Within-class slope | 0.068 | 0.197 | 0.347 | 1.071 | 0.728 | 1.574 | 7.29e-01 | 1.00e+00 | 26002 | 17379 |
| European | Recorded Depression | TCA:nortriptyline | Insomnia | Within-class slope | -0.006 | 0.095 | -0.067 | 0.994 | 0.825 | 1.197 | 9.47e-01 | 1.00e+00 | 26002 | 17379 |
| European | Recorded Depression | NaSSA:mirtazapine | Insomnia | Interaction (vs SSRI:sertraline) | -0.114 | 0.092 | -1.237 | 0.892 | 0.745 | 1.069 | 2.16e-01 | 1.00e+00 | 26002 | 17379 |
| European | Recorded Depression | NDRI:bupropion | Insomnia | Interaction (vs SSRI:sertraline) | 0.003 | 0.048 | 0.052 | 1.003 | 0.912 | 1.102 | 9.58e-01 | 1.00e+00 | 26002 | 17379 |
| European | Recorded Depression | SARI:trazodone | Insomnia | Interaction (vs SSRI:sertraline) | -0.102 | 0.065 | -1.569 | 0.903 | 0.794 | 1.026 | 1.17e-01 | 8.17e-01 | 26002 | 17379 |
| European | Recorded Depression | SNRI:duloxetine | Insomnia | Interaction (vs SSRI:sertraline) | -0.026 | 0.055 | -0.476 | 0.974 | 0.875 | 1.085 | 6.34e-01 | 1.00e+00 | 26002 | 17379 |
| European | Recorded Depression | SNRI:venlafaxine | Insomnia | Interaction (vs SSRI:sertraline) | -0.011 | 0.057 | -0.198 | 0.989 | 0.884 | 1.106 | 8.43e-01 | 1.00e+00 | 26002 | 17379 |
| European | Recorded Depression | SSRI:citalopram | Insomnia | Interaction (vs SSRI:sertraline) | -0.010 | 0.051 | -0.192 | 0.990 | 0.896 | 1.094 | 8.48e-01 | 1.00e+00 | 26002 | 17379 |
| European | Recorded Depression | SSRI:escitalopram | Insomnia | Interaction (vs SSRI:sertraline) | -0.003 | 0.050 | -0.053 | 0.997 | 0.904 | 1.100 | 9.58e-01 | 1.00e+00 | 26002 | 17379 |
| European | Recorded Depression | SSRI:fluoxetine | Insomnia | Interaction (vs SSRI:sertraline) | -0.113 | 0.051 | -2.212 | 0.893 | 0.808 | 0.987 | 2.69e-02 | 1.89e-01 | 26002 | 17379 |
| European | Recorded Depression | SSRI:paroxetine | Insomnia | Interaction (vs SSRI:sertraline) | 0.034 | 0.077 | 0.443 | 1.035 | 0.890 | 1.202 | 6.58e-01 | 1.00e+00 | 26002 | 17379 |
| European | Recorded Depression | TCA:amitriptyline | Insomnia | Interaction (vs SSRI:sertraline) | 0.019 | 0.078 | 0.243 | 1.019 | 0.875 | 1.187 | 8.08e-01 | 1.00e+00 | 26002 | 17379 |
| European | Recorded Depression | TCA:doxepin | Insomnia | Interaction (vs SSRI:sertraline) | -0.025 | 0.199 | -0.123 | 0.976 | 0.661 | 1.441 | 9.02e-01 | 1.00e+00 | 26002 | 17379 |
| European | Recorded Depression | TCA:nortriptyline | Insomnia | Interaction (vs SSRI:sertraline) | -0.099 | 0.100 | -0.995 | 0.906 | 0.745 | 1.101 | 3.20e-01 | 1.00e+00 | 26002 | 17379 |
| European | Recorded Depression | | Insomnia | Joint_Interaction_Test | | | 11.414 | | | | 4.94e-01 | 1.00e+00 | 26002 | 17379 |
| European | Recorded Depression | SSRI:sertraline | Depression | Within-class slope | 0.182 | 0.033 | 5.553 | 1.200 | 1.125 | 1.280 | 2.80e-08 | 1.96e-07 | 26002 | 17379 |
| European | Recorded Depression | NaSSA:mirtazapine | Depression | Within-class slope | 0.156 | 0.092 | 1.700 | 1.169 | 0.976 | 1.401 | 8.91e-02 | 6.24e-01 | 26002 | 17379 |
| European | Recorded Depression | NDRI:bupropion | Depression | Within-class slope | 0.166 | 0.037 | 4.526 | 1.181 | 1.099 | 1.269 | 6.01e-06 | 4.20e-05 | 26002 | 17379 |
| European | Recorded Depression | SARI:trazodone | Depression | Within-class slope | 0.074 | 0.058 | 1.271 | 1.076 | 0.961 | 1.206 | 2.04e-01 | 1.00e+00 | 26002 | 17379 |
| European | Recorded Depression | SNRI:duloxetine | Depression | Within-class slope | 0.120 | 0.046 | 2.624 | 1.127 | 1.031 | 1.232 | 8.70e-03 | 6.09e-02 | 26002 | 17379 |
| European | Recorded Depression | SNRI:venlafaxine | Depression | Within-class slope | 0.122 | 0.047 | 2.606 | 1.130 | 1.031 | 1.239 | 9.16e-03 | 6.41e-02 | 26002 | 17379 |
| European | Recorded Depression | SSRI:citalopram | Depression | Within-class slope | 0.136 | 0.042 | 3.229 | 1.145 | 1.055 | 1.244 | 1.24e-03 | 8.69e-03 | 26002 | 17379 |
| European | Recorded Depression | SSRI:escitalopram | Depression | Within-class slope | 0.151 | 0.040 | 3.751 | 1.163 | 1.075 | 1.259 | 1.76e-04 | 1.23e-03 | 26002 | 17379 |
| European | Recorded Depression | SSRI:fluoxetine | Depression | Within-class slope | 0.054 | 0.040 | 1.372 | 1.056 | 0.977 | 1.141 | 1.70e-01 | 1.00e+00 | 26002 | 17379 |
| European | Recorded Depression | SSRI:paroxetine | Depression | Within-class slope | 0.217 | 0.071 | 3.044 | 1.243 | 1.081 | 1.430 | 2.33e-03 | 1.63e-02 | 26002 | 17379 |
| European | Recorded Depression | TCA:amitriptyline | Depression | Within-class slope | 0.148 | 0.074 | 2.012 | 1.160 | 1.004 | 1.340 | 4.43e-02 | 3.10e-01 | 26002 | 17379 |
| European | Recorded Depression | TCA:doxepin | Depression | Within-class slope | -0.020 | 0.195 | -0.102 | 0.980 | 0.669 | 1.437 | 9.19e-01 | 1.00e+00 | 26002 | 17379 |
| European | Recorded Depression | TCA:nortriptyline | Depression | Within-class slope | 0.060 | 0.094 | 0.640 | 1.062 | 0.884 | 1.275 | 5.22e-01 | 1.00e+00 | 26002 | 17379 |
| European | Recorded Depression | NaSSA:mirtazapine | Depression | Interaction (vs SSRI:sertraline) | -0.026 | 0.097 | -0.266 | 0.975 | 0.806 | 1.179 | 7.90e-01 | 1.00e+00 | 26002 | 17379 |
| European | Recorded Depression | NDRI:bupropion | Depression | Interaction (vs SSRI:sertraline) | -0.016 | 0.047 | -0.342 | 0.984 | 0.897 | 1.079 | 7.32e-01 | 1.00e+00 | 26002 | 17379 |
| European | Recorded Depression | SARI:trazodone | Depression | Interaction (vs SSRI:sertraline) | -0.109 | 0.065 | -1.666 | 0.897 | 0.789 | 1.019 | 9.58e-02 | 6.71e-01 | 26002 | 17379 |
| European | Recorded Depression | SNRI:duloxetine | Depression | Interaction (vs SSRI:sertraline) | -0.063 | 0.054 | -1.152 | 0.939 | 0.844 | 1.045 | 2.49e-01 | 1.00e+00 | 26002 | 17379 |
| European | Recorded Depression | SNRI:venlafaxine | Depression | Interaction (vs SSRI:sertraline) | -0.060 | 0.055 | -1.080 | 0.942 | 0.845 | 1.050 | 2.80e-01 | 1.00e+00 | 26002 | 17379 |
| European | Recorded Depression | SSRI:citalopram | Depression | Interaction (vs SSRI:sertraline) | -0.047 | 0.052 | -0.903 | 0.954 | 0.863 | 1.056 | 3.66e-01 | 1.00e+00 | 26002 | 17379 |
| European | Recorded Depression | SSRI:escitalopram | Depression | Interaction (vs SSRI:sertraline) | -0.031 | 0.051 | -0.613 | 0.969 | 0.878 | 1.071 | 5.40e-01 | 1.00e+00 | 26002 | 17379 |
| European | Recorded Depression | SSRI:fluoxetine | Depression | Interaction (vs SSRI:sertraline) | -0.128 | 0.050 | -2.558 | 0.880 | 0.798 | 0.971 | 1.05e-02 | 7.36e-02 | 26002 | 17379 |

| | | | | | | | | | | | | | | |
|---|---|---|---|---|---|---|---|---|---|---|---|---|---|---|
| European | Recorded Depression | SSRI:paroxetine | Depression | Interaction (vs SSRI:sertraline) | 0.035 | 0.078 | 0.454 | 1.036 | 0.890 | 1.206 | 6.50e-01 | 1.00e+00 | 26002 | 17379 |
| European | Recorded Depression | TCA:amitriptyline | Depression | Interaction (vs SSRI:sertraline) | -0.034 | 0.079 | -0.432 | 0.966 | 0.828 | 1.128 | 6.65e-01 | 1.00e+00 | 26002 | 17379 |
| European | Recorded Depression | TCA:doxepin | Depression | Interaction (vs SSRI:sertraline) | -0.202 | 0.197 | -1.027 | 0.817 | 0.555 | 1.202 | 3.05e-01 | 1.00e+00 | 26002 | 17379 |
| European | Recorded Depression | TCA:nortriptyline | Depression | Interaction (vs SSRI:sertraline) | -0.122 | 0.098 | -1.250 | 0.885 | 0.730 | 1.072 | 2.11e-01 | 1.00e+00 | 26002 | 17379 |
| European | Recorded Depression | | Depression | Joint_Interaction_Test | | | 11.728 | | | | 4.68e-01 | 1.00e+00 | 26002 | 17379 |
| European | Recorded Depression | SSRI:sertraline | Schizophrenia | Within-class slope | 0.029 | 0.032 | 0.910 | 1.030 | 0.967 | 1.096 | 3.63e-01 | 1.00e+00 | 26002 | 17379 |
| European | Recorded Depression | NaSSA:mirtazapine | Schizophrenia | Within-class slope | 0.052 | 0.087 | 0.603 | 1.054 | 0.889 | 1.249 | 5.47e-01 | 1.00e+00 | 26002 | 17379 |
| European | Recorded Depression | NDRI:bupropion | Schizophrenia | Within-class slope | 0.034 | 0.037 | 0.919 | 1.034 | 0.963 | 1.111 | 3.58e-01 | 1.00e+00 | 26002 | 17379 |
| European | Recorded Depression | SARI:trazodone | Schizophrenia | Within-class slope | -0.099 | 0.056 | -1.768 | 0.905 | 0.811 | 1.011 | 7.71e-02 | 5.39e-01 | 26002 | 17379 |
| European | Recorded Depression | SNRI:duloxetine | Schizophrenia | Within-class slope | 0.041 | 0.045 | 0.910 | 1.042 | 0.953 | 1.139 | 3.63e-01 | 1.00e+00 | 26002 | 17379 |
| European | Recorded Depression | SNRI:venlafaxine | Schizophrenia | Within-class slope | 0.009 | 0.046 | 0.203 | 1.009 | 0.923 | 1.104 | 8.39e-01 | 1.00e+00 | 26002 | 17379 |
| European | Recorded Depression | SSRI:citalopram | Schizophrenia | Within-class slope | -0.025 | 0.039 | -0.651 | 0.975 | 0.903 | 1.052 | 5.15e-01 | 1.00e+00 | 26002 | 17379 |
| European | Recorded Depression | SSRI:escitalopram | Schizophrenia | Within-class slope | 0.033 | 0.038 | 0.855 | 1.033 | 0.959 | 1.114 | 3.93e-01 | 1.00e+00 | 26002 | 17379 |
| European | Recorded Depression | SSRI:fluoxetine | Schizophrenia | Within-class slope | 0.001 | 0.040 | 0.027 | 1.001 | 0.926 | 1.082 | 9.78e-01 | 1.00e+00 | 26002 | 17379 |
| European | Recorded Depression | SSRI:paroxetine | Schizophrenia | Within-class slope | -0.042 | 0.069 | -0.605 | 0.959 | 0.837 | 1.099 | 5.45e-01 | 1.00e+00 | 26002 | 17379 |
| European | Recorded Depression | TCA:amitriptyline | Schizophrenia | Within-class slope | -0.087 | 0.074 | -1.179 | 0.917 | 0.793 | 1.059 | 2.39e-01 | 1.00e+00 | 26002 | 17379 |
| European | Recorded Depression | TCA:doxepin | Schizophrenia | Within-class slope | -0.141 | 0.201 | -0.700 | 0.869 | 0.586 | 1.288 | 4.84e-01 | 1.00e+00 | 26002 | 17379 |
| European | Recorded Depression | TCA:nortriptyline | Schizophrenia | Within-class slope | -0.014 | 0.090 | -0.154 | 0.986 | 0.826 | 1.177 | 8.78e-01 | 1.00e+00 | 26002 | 17379 |
| European | Recorded Depression | NaSSA:mirtazapine | Schizophrenia | Interaction (vs SSRI:sertraline) | 0.023 | 0.092 | 0.251 | 1.023 | 0.855 | 1.225 | 8.02e-01 | 1.00e+00 | 26002 | 17379 |
| European | Recorded Depression | NDRI:bupropion | Schizophrenia | Interaction (vs SSRI:sertraline) | 0.004 | 0.047 | 0.096 | 1.004 | 0.917 | 1.100 | 9.23e-01 | 1.00e+00 | 26002 | 17379 |
| European | Recorded Depression | SARI:trazodone | Schizophrenia | Interaction (vs SSRI:sertraline) | -0.129 | 0.063 | -2.028 | 0.879 | 0.777 | 0.996 | 4.25e-02 | 2.98e-01 | 26002 | 17379 |
| European | Recorded Depression | SNRI:duloxetine | Schizophrenia | Interaction (vs SSRI:sertraline) | 0.012 | 0.054 | 0.224 | 1.012 | 0.910 | 1.126 | 8.23e-01 | 1.00e+00 | 26002 | 17379 |
| European | Recorded Depression | SNRI:venlafaxine | Schizophrenia | Interaction (vs SSRI:sertraline) | -0.020 | 0.054 | -0.367 | 0.980 | 0.881 | 1.090 | 7.14e-01 | 1.00e+00 | 26002 | 17379 |
| European | Recorded Depression | SSRI:citalopram | Schizophrenia | Interaction (vs SSRI:sertraline) | -0.055 | 0.049 | -1.126 | 0.947 | 0.861 | 1.041 | 2.60e-01 | 1.00e+00 | 26002 | 17379 |
| European | Recorded Depression | SSRI:escitalopram | Schizophrenia | Interaction (vs SSRI:sertraline) | 0.004 | 0.049 | 0.073 | 1.004 | 0.912 | 1.104 | 9.42e-01 | 1.00e+00 | 26002 | 17379 |
| European | Recorded Depression | SSRI:fluoxetine | Schizophrenia | Interaction (vs SSRI:sertraline) | -0.028 | 0.049 | -0.568 | 0.972 | 0.882 | 1.071 | 5.70e-01 | 1.00e+00 | 26002 | 17379 |
| European | Recorded Depression | SSRI:paroxetine | Schizophrenia | Interaction (vs SSRI:sertraline) | -0.071 | 0.075 | -0.944 | 0.931 | 0.803 | 1.080 | 3.45e-01 | 1.00e+00 | 26002 | 17379 |
| European | Recorded Depression | TCA:amitriptyline | Schizophrenia | Interaction (vs SSRI:sertraline) | -0.116 | 0.079 | -1.466 | 0.890 | 0.762 | 1.040 | 1.43e-01 | 9.98e-01 | 26002 | 17379 |
| European | Recorded Depression | TCA:doxepin | Schizophrenia | Interaction (vs SSRI:sertraline) | -0.170 | 0.203 | -0.837 | 0.844 | 0.567 | 1.256 | 4.03e-01 | 1.00e+00 | 26002 | 17379 |
| European | Recorded Depression | TCA:nortriptyline | Schizophrenia | Interaction (vs SSRI:sertraline) | -0.043 | 0.095 | -0.455 | 0.958 | 0.796 | 1.153 | 6.49e-01 | 1.00e+00 | 26002 | 17379 |
| European | Recorded Depression | | Schizophrenia | Joint_Interaction_Test | | | 9.570 | | | | 6.54e-01 | 1.00e+00 | 26002 | 17379 |
| European | Recorded Depression | SSRI:sertraline | Bipolar Disorder | Within-class slope | 0.053 | 0.033 | 1.608 | 1.054 | 0.988 | 1.125 | 1.08e-01 | 7.55e-01 | 26002 | 17379 |
| European | Recorded Depression | NaSSA:mirtazapine | Bipolar Disorder | Within-class slope | -0.061 | 0.089 | -0.682 | 0.941 | 0.790 | 1.121 | 4.95e-01 | 1.00e+00 | 26002 | 17379 |
| European | Recorded Depression | NDRI:bupropion | Bipolar Disorder | Within-class slope | -0.020 | 0.038 | -0.518 | 0.981 | 0.910 | 1.056 | 6.04e-01 | 1.00e+00 | 26002 | 17379 |
| European | Recorded Depression | SARI:trazodone | Bipolar Disorder | Within-class slope | -0.019 | 0.058 | -0.329 | 0.981 | 0.876 | 1.099 | 7.42e-01 | 1.00e+00 | 26002 | 17379 |
| European | Recorded Depression | SNRI:duloxetine | Bipolar Disorder | Within-class slope | 0.017 | 0.046 | 0.369 | 1.017 | 0.929 | 1.113 | 7.12e-01 | 1.00e+00 | 26002 | 17379 |
| European | Recorded Depression | SNRI:venlafaxine | Bipolar Disorder | Within-class slope | 0.061 | 0.047 | 1.293 | 1.062 | 0.969 | 1.165 | 1.96e-01 | 1.00e+00 | 26002 | 17379 |
| European | Recorded Depression | SSRI:citalopram | Bipolar Disorder | Within-class slope | 0.003 | 0.041 | 0.085 | 1.003 | 0.927 | 1.086 | 9.33e-01 | 1.00e+00 | 26002 | 17379 |
| European | Recorded Depression | SSRI:escitalopram | Bipolar Disorder | Within-class slope | -0.002 | 0.039 | -0.062 | 0.998 | 0.925 | 1.076 | 9.50e-01 | 1.00e+00 | 26002 | 17379 |
| European | Recorded Depression | SSRI:fluoxetine | Bipolar Disorder | Within-class slope | -0.019 | 0.040 | -0.470 | 0.981 | 0.907 | 1.062 | 6.38e-01 | 1.00e+00 | 26002 | 17379 |
| European | Recorded Depression | SSRI:paroxetine | Bipolar Disorder | Within-class slope | -0.018 | 0.072 | -0.254 | 0.982 | 0.854 | 1.130 | 8.00e-01 | 1.00e+00 | 26002 | 17379 |
| European | Recorded Depression | TCA:amitriptyline | Bipolar Disorder | Within-class slope | 0.002 | 0.075 | 0.033 | 1.002 | 0.866 | 1.161 | 9.73e-01 | 1.00e+00 | 26002 | 17379 |
| European | Recorded Depression | TCA:doxepin | Bipolar Disorder | Within-class slope | -0.160 | 0.161 | -0.989 | 0.852 | 0.621 | 1.170 | 3.23e-01 | 1.00e+00 | 26002 | 17379 |
| European | Recorded Depression | TCA:nortriptyline | Bipolar Disorder | Within-class slope | 0.026 | 0.089 | 0.297 | 1.027 | 0.863 | 1.222 | 7.67e-01 | 1.00e+00 | 26002 | 17379 |
| European | Recorded Depression | NaSSA:mirtazapine | Bipolar Disorder | Interaction (vs SSRI:sertraline) | -0.114 | 0.094 | -1.210 | 0.892 | 0.742 | 1.073 | 2.26e-01 | 1.00e+00 | 26002 | 17379 |
| European | Recorded Depression | NDRI:bupropion | Bipolar Disorder | Interaction (vs SSRI:sertraline) | -0.073 | 0.048 | -1.510 | 0.930 | 0.846 | 1.022 | 1.31e-01 | 9.17e-01 | 26002 | 17379 |
| European | Recorded Depression | SARI:trazodone | Bipolar Disorder | Interaction (vs SSRI:sertraline) | -0.072 | 0.066 | -1.099 | 0.931 | 0.818 | 1.058 | 2.72e-01 | 1.00e+00 | 26002 | 17379 |
| European | Recorded Depression | SNRI:duloxetine | Bipolar Disorder | Interaction (vs SSRI:sertraline) | -0.036 | 0.055 | -0.652 | 0.965 | 0.866 | 1.075 | 5.14e-01 | 1.00e+00 | 26002 | 17379 |
| European | Recorded Depression | SNRI:venlafaxine | Bipolar Disorder | Interaction (vs SSRI:sertraline) | 0.008 | 0.056 | 0.137 | 1.008 | 0.903 | 1.124 | 8.91e-01 | 1.00e+00 | 26002 | 17379 |
| European | Recorded Depression | SSRI:citalopram | Bipolar Disorder | Interaction (vs SSRI:sertraline) | -0.050 | 0.050 | -0.986 | 0.952 | 0.863 | 1.050 | 3.24e-01 | 1.00e+00 | 26002 | 17379 |
| European | Recorded Depression | SSRI:escitalopram | Bipolar Disorder | Interaction (vs SSRI:sertraline) | -0.055 | 0.049 | -1.119 | 0.946 | 0.859 | 1.042 | 2.63e-01 | 1.00e+00 | 26002 | 17379 |
| European | Recorded Depression | SSRI:fluoxetine | Bipolar Disorder | Interaction (vs SSRI:sertraline) | -0.072 | 0.050 | -1.432 | 0.931 | 0.844 | 1.027 | 1.52e-01 | 1.00e+00 | 26002 | 17379 |
| European | Recorded Depression | SSRI:paroxetine | Bipolar Disorder | Interaction (vs SSRI:sertraline) | -0.071 | 0.078 | -0.914 | 0.931 | 0.800 | 1.085 | 3.61e-01 | 1.00e+00 | 26002 | 17379 |
| European | Recorded Depression | TCA:amitriptyline | Bipolar Disorder | Interaction (vs SSRI:sertraline) | -0.050 | 0.080 | -0.629 | 0.951 | 0.812 | 1.113 | 5.30e-01 | 1.00e+00 | 26002 | 17379 |
| European | Recorded Depression | TCA:doxepin | Bipolar Disorder | Interaction (vs SSRI:sertraline) | -0.213 | 0.164 | -1.294 | 0.809 | 0.586 | 1.115 | 1.96e-01 | 1.00e+00 | 26002 | 17379 |
| European | Recorded Depression | TCA:nortriptyline | Bipolar Disorder | Interaction (vs SSRI:sertraline) | -0.027 | 0.094 | -0.284 | 0.974 | 0.811 | 1.170 | 7.76e-01 | 1.00e+00 | 26002 | 17379 |
| European | Recorded Depression | | Bipolar Disorder | Joint_Interaction_Test | | | 6.608 | | | | 8.82e-01 | 1.00e+00 | 26002 | 17379 |
| European | Recorded Depression | SSRI:sertraline | BMI | Within-class slope | -0.022 | 0.031 | -0.688 | 0.979 | 0.921 | 1.041 | 4.91e-01 | 1.00e+00 | 26002 | 17379 |
| European | Recorded Depression | NaSSA:mirtazapine | BMI | Within-class slope | 0.061 | 0.082 | 0.749 | 1.063 | 0.906 | 1.248 | 4.54e-01 | 1.00e+00 | 26002 | 17379 |

| | | | | | | | | | | | | | | |
|---|---|---|---|---|---|---|---|---|---|---|---|---|---|---|
| European | Recorded Depression | NDRI:bupropion | BMI | Within-class slope | -0.011 | 0.036 | -0.300 | 0.989 | 0.921 | 1.062 | 7.64e-01 | 1.00e+00 | 26002 | 17379 |
| European | Recorded Depression | SARI:trazodone | BMI | Within-class slope | -0.076 | 0.053 | -1.426 | 0.927 | 0.836 | 1.029 | 1.54e-01 | 1.00e+00 | 26002 | 17379 |
| European | Recorded Depression | SNRI:duloxetine | BMI | Within-class slope | -0.008 | 0.042 | -0.183 | 0.992 | 0.913 | 1.078 | 8.55e-01 | 1.00e+00 | 26002 | 17379 |
| European | Recorded Depression | SNRI:venlafaxine | BMI | Within-class slope | -0.032 | 0.045 | -0.708 | 0.969 | 0.887 | 1.058 | 4.79e-01 | 1.00e+00 | 26002 | 17379 |
| European | Recorded Depression | SSRI:citalopram | BMI | Within-class slope | 0.009 | 0.038 | 0.238 | 1.009 | 0.936 | 1.088 | 8.12e-01 | 1.00e+00 | 26002 | 17379 |
| European | Recorded Depression | SSRI:escitalopram | BMI | Within-class slope | 0.015 | 0.038 | 0.387 | 1.015 | 0.942 | 1.094 | 6.98e-01 | 1.00e+00 | 26002 | 17379 |
| European | Recorded Depression | SSRI:fluoxetine | BMI | Within-class slope | -0.003 | 0.039 | -0.075 | 0.997 | 0.924 | 1.076 | 9.40e-01 | 1.00e+00 | 26002 | 17379 |
| European | Recorded Depression | SSRI:paroxetine | BMI | Within-class slope | 0.063 | 0.063 | 1.008 | 1.065 | 0.942 | 1.205 | 3.13e-01 | 1.00e+00 | 26002 | 17379 |
| European | Recorded Depression | TCA:amitriptyline | BMI | Within-class slope | -0.002 | 0.071 | -0.024 | 0.998 | 0.869 | 1.146 | 9.81e-01 | 1.00e+00 | 26002 | 17379 |
| European | Recorded Depression | TCA:doxepin | BMI | Within-class slope | -0.202 | 0.180 | -1.123 | 0.817 | 0.574 | 1.163 | 2.61e-01 | 1.00e+00 | 26002 | 17379 |
| European | Recorded Depression | TCA:nortriptyline | BMI | Within-class slope | -0.035 | 0.089 | -0.398 | 0.965 | 0.811 | 1.149 | 6.91e-01 | 1.00e+00 | 26002 | 17379 |
| European | Recorded Depression | NaSSA:mirtazapine | BMI | Interaction (vs SSRI:sertraline) | 0.083 | 0.087 | 0.949 | 1.086 | 0.916 | 1.288 | 3.43e-01 | 1.00e+00 | 26002 | 17379 |
| European | Recorded Depression | NDRI:bupropion | BMI | Interaction (vs SSRI:sertraline) | 0.011 | 0.046 | 0.233 | 1.011 | 0.924 | 1.106 | 8.16e-01 | 1.00e+00 | 26002 | 17379 |
| European | Recorded Depression | SARI:trazodone | BMI | Interaction (vs SSRI:sertraline) | -0.054 | 0.060 | -0.898 | 0.947 | 0.842 | 1.066 | 3.69e-01 | 1.00e+00 | 26002 | 17379 |
| European | Recorded Depression | SNRI:duloxetine | BMI | Interaction (vs SSRI:sertraline) | 0.014 | 0.051 | 0.268 | 1.014 | 0.917 | 1.121 | 7.89e-01 | 1.00e+00 | 26002 | 17379 |
| European | Recorded Depression | SNRI:venlafaxine | BMI | Interaction (vs SSRI:sertraline) | -0.010 | 0.053 | -0.195 | 0.990 | 0.891 | 1.099 | 8.45e-01 | 1.00e+00 | 26002 | 17379 |
| European | Recorded Depression | SSRI:citalopram | BMI | Interaction (vs SSRI:sertraline) | 0.031 | 0.047 | 0.646 | 1.031 | 0.940 | 1.131 | 5.18e-01 | 1.00e+00 | 26002 | 17379 |
| European | Recorded Depression | SSRI:escitalopram | BMI | Interaction (vs SSRI:sertraline) | 0.036 | 0.048 | 0.758 | 1.037 | 0.944 | 1.139 | 4.48e-01 | 1.00e+00 | 26002 | 17379 |
| European | Recorded Depression | SSRI:fluoxetine | BMI | Interaction (vs SSRI:sertraline) | 0.019 | 0.048 | 0.389 | 1.019 | 0.928 | 1.119 | 6.98e-01 | 1.00e+00 | 26002 | 17379 |
| European | Recorded Depression | SSRI:paroxetine | BMI | Interaction (vs SSRI:sertraline) | 0.085 | 0.068 | 1.240 | 1.089 | 0.952 | 1.245 | 2.15e-01 | 1.00e+00 | 26002 | 17379 |
| European | Recorded Depression | TCA:amitriptyline | BMI | Interaction (vs SSRI:sertraline) | 0.020 | 0.076 | 0.263 | 1.020 | 0.880 | 1.183 | 7.93e-01 | 1.00e+00 | 26002 | 17379 |
| European | Recorded Depression | TCA:doxepin | BMI | Interaction (vs SSRI:sertraline) | -0.181 | 0.182 | -0.992 | 0.835 | 0.584 | 1.193 | 3.21e-01 | 1.00e+00 | 26002 | 17379 |
| European | Recorded Depression | TCA:nortriptyline | BMI | Interaction (vs SSRI:sertraline) | -0.014 | 0.093 | -0.148 | 0.986 | 0.822 | 1.184 | 8.82e-01 | 1.00e+00 | 26002 | 17379 |
| European | Recorded Depression | | BMI | Joint_Interaction_Test | | | 6.192 | | | | 9.06e-01 | 1.00e+00 | 26002 | 17379 |
| European | No Recorded Depression | SSRI:sertraline | ADHD | Within-class slope | 0.102 | 0.042 | 2.433 | 1.108 | 1.020 | 1.203 | 1.50e-02 | 1.05e-01 | 21196 | 17288 |
| European | No Recorded Depression | NaSSA:mirtazapine | ADHD | Within-class slope | 0.042 | 0.100 | 0.423 | 1.043 | 0.858 | 1.269 | 6.72e-01 | 1.00e+00 | 21196 | 17288 |
| European | No Recorded Depression | NDRI:bupropion | ADHD | Within-class slope | 0.037 | 0.050 | 0.734 | 1.038 | 0.940 | 1.145 | 4.63e-01 | 1.00e+00 | 21196 | 17288 |
| European | No Recorded Depression | SARI:trazodone | ADHD | Within-class slope | 0.051 | 0.051 | 0.989 | 1.052 | 0.952 | 1.163 | 3.23e-01 | 1.00e+00 | 21196 | 17288 |
| European | No Recorded Depression | SNRI:duloxetine | ADHD | Within-class slope | 0.063 | 0.054 | 1.165 | 1.065 | 0.958 | 1.184 | 2.44e-01 | 1.00e+00 | 21196 | 17288 |
| European | No Recorded Depression | SNRI:venlafaxine | ADHD | Within-class slope | 0.072 | 0.061 | 1.186 | 1.075 | 0.954 | 1.211 | 2.36e-01 | 1.00e+00 | 21196 | 17288 |
| European | No Recorded Depression | SSRI:citalopram | ADHD | Within-class slope | -0.080 | 0.052 | -1.527 | 0.923 | 0.833 | 1.023 | 1.27e-01 | 8.88e-01 | 21196 | 17288 |
| European | No Recorded Depression | SSRI:escitalopram | ADHD | Within-class slope | 0.157 | 0.049 | 3.206 | 1.170 | 1.063 | 1.287 | 1.34e-03 | 9.41e-03 | 21196 | 17288 |
| European | No Recorded Depression | SSRI:fluoxetine | ADHD | Within-class slope | 0.112 | 0.057 | 1.957 | 1.118 | 1.000 | 1.250 | 5.03e-02 | 3.52e-01 | 21196 | 17288 |
| European | No Recorded Depression | SSRI:paroxetine | ADHD | Within-class slope | 0.066 | 0.085 | 0.783 | 1.069 | 0.905 | 1.261 | 4.33e-01 | 1.00e+00 | 21196 | 17288 |
| European | No Recorded Depression | TCA:amitriptyline | ADHD | Within-class slope | 0.067 | 0.059 | 1.131 | 1.069 | 0.952 | 1.201 | 2.58e-01 | 1.00e+00 | 21196 | 17288 |
| European | No Recorded Depression | TCA:doxepin | ADHD | Within-class slope | -0.037 | 0.146 | -0.253 | 0.964 | 0.724 | 1.283 | 8.00e-01 | 1.00e+00 | 21196 | 17288 |
| European | No Recorded Depression | TCA:nortriptyline | ADHD | Within-class slope | 0.024 | 0.074 | 0.323 | 1.024 | 0.886 | 1.184 | 7.46e-01 | 1.00e+00 | 21196 | 17288 |
| European | No Recorded Depression | NaSSA:mirtazapine | ADHD | Interaction (vs SSRI:sertraline) | -0.060 | 0.108 | -0.557 | 0.942 | 0.762 | 1.163 | 5.77e-01 | 1.00e+00 | 21196 | 17288 |
| European | No Recorded Depression | NDRI:bupropion | ADHD | Interaction (vs SSRI:sertraline) | -0.065 | 0.064 | -1.021 | 0.937 | 0.826 | 1.062 | 3.07e-01 | 1.00e+00 | 21196 | 17288 |
| European | No Recorded Depression | SARI:trazodone | ADHD | Interaction (vs SSRI:sertraline) | -0.052 | 0.065 | -0.799 | 0.950 | 0.837 | 1.078 | 4.24e-01 | 1.00e+00 | 21196 | 17288 |
| European | No Recorded Depression | SNRI:duloxetine | ADHD | Interaction (vs SSRI:sertraline) | -0.039 | 0.067 | -0.585 | 0.962 | 0.843 | 1.097 | 5.58e-01 | 1.00e+00 | 21196 | 17288 |
| European | No Recorded Depression | SNRI:venlafaxine | ADHD | Interaction (vs SSRI:sertraline) | -0.030 | 0.073 | -0.413 | 0.970 | 0.841 | 1.120 | 6.80e-01 | 1.00e+00 | 21196 | 17288 |
| European | No Recorded Depression | SSRI:citalopram | ADHD | Interaction (vs SSRI:sertraline) | -0.182 | 0.066 | -2.766 | 0.833 | 0.732 | 0.948 | 5.67e-03 | 3.97e-02 | 21196 | 17288 |
| European | No Recorded Depression | SSRI:escitalopram | ADHD | Interaction (vs SSRI:sertraline) | 0.055 | 0.063 | 0.863 | 1.056 | 0.933 | 1.196 | 3.88e-01 | 1.00e+00 | 21196 | 17288 |
| European | No Recorded Depression | SSRI:fluoxetine | ADHD | Interaction (vs SSRI:sertraline) | 0.009 | 0.069 | 0.136 | 1.009 | 0.881 | 1.157 | 8.92e-01 | 1.00e+00 | 21196 | 17288 |
| European | No Recorded Depression | SSRI:paroxetine | ADHD | Interaction (vs SSRI:sertraline) | -0.036 | 0.094 | -0.383 | 0.965 | 0.803 | 1.160 | 7.02e-01 | 1.00e+00 | 21196 | 17288 |
| European | No Recorded Depression | TCA:amitriptyline | ADHD | Interaction (vs SSRI:sertraline) | -0.035 | 0.071 | -0.492 | 0.965 | 0.839 | 1.111 | 6.22e-01 | 1.00e+00 | 21196 | 17288 |
| European | No Recorded Depression | TCA:doxepin | ADHD | Interaction (vs SSRI:sertraline) | -0.139 | 0.151 | -0.921 | 0.870 | 0.647 | 1.170 | 3.57e-01 | 1.00e+00 | 21196 | 17288 |
| European | No Recorded Depression | TCA:nortriptyline | ADHD | Interaction (vs SSRI:sertraline) | -0.078 | 0.083 | -0.940 | 0.925 | 0.785 | 1.089 | 3.47e-01 | 1.00e+00 | 21196 | 17288 |
| European | No Recorded Depression | | ADHD | Joint_Interaction_Test | | | 14.377 | | | | 2.77e-01 | 1.00e+00 | 21196 | 17288 |
| European | No Recorded Depression | SSRI:sertraline | Migraine | Within-class slope | 0.049 | 0.042 | 1.157 | 1.050 | 0.967 | 1.141 | 2.47e-01 | 1.00e+00 | 21196 | 17288 |
| European | No Recorded Depression | NaSSA:mirtazapine | Migraine | Within-class slope | 0.044 | 0.098 | 0.452 | 1.045 | 0.863 | 1.266 | 6.52e-01 | 1.00e+00 | 21196 | 17288 |
| European | No Recorded Depression | NDRI:bupropion | Migraine | Within-class slope | 0.059 | 0.051 | 1.153 | 1.061 | 0.959 | 1.174 | 2.49e-01 | 1.00e+00 | 21196 | 17288 |
| European | No Recorded Depression | SARI:trazodone | Migraine | Within-class slope | -0.024 | 0.050 | -0.471 | 0.977 | 0.885 | 1.078 | 6.38e-01 | 1.00e+00 | 21196 | 17288 |
| European | No Recorded Depression | SNRI:duloxetine | Migraine | Within-class slope | -0.062 | 0.054 | -1.155 | 0.940 | 0.845 | 1.044 | 2.48e-01 | 1.00e+00 | 21196 | 17288 |
| European | No Recorded Depression | SNRI:venlafaxine | Migraine | Within-class slope | 0.107 | 0.061 | 1.746 | 1.113 | 0.987 | 1.255 | 8.08e-02 | 5.66e-01 | 21196 | 17288 |
| European | No Recorded Depression | SSRI:citalopram | Migraine | Within-class slope | 0.068 | 0.053 | 1.296 | 1.071 | 0.966 | 1.188 | 1.95e-01 | 1.00e+00 | 21196 | 17288 |
| European | No Recorded Depression | SSRI:escitalopram | Migraine | Within-class slope | 0.038 | 0.047 | 0.815 | 1.039 | 0.948 | 1.140 | 4.15e-01 | 1.00e+00 | 21196 | 17288 |
| European | No Recorded Depression | SSRI:fluoxetine | Migraine | Within-class slope | 0.021 | 0.056 | 0.371 | 1.021 | 0.915 | 1.139 | 7.11e-01 | 1.00e+00 | 21196 | 17288 |

| | | | | | | | | | | | | | | |
|---|---|---|---|---|---|---|---|---|---|---|---|---|---|---|
| European | No Recorded Depression | SSRI:paroxetine | Migraine | Within-class slope | -0.067 | 0.079 | -0.840 | 0.936 | 0.801 | 1.093 | 4.01e-01 | 1.00e+00 | 21196 | 17288 |
| European | No Recorded Depression | TCA:amitriptyline | Migraine | Within-class slope | 0.059 | 0.059 | 1.006 | 1.061 | 0.946 | 1.190 | 3.15e-01 | 1.00e+00 | 21196 | 17288 |
| European | No Recorded Depression | TCA:doxepin | Migraine | Within-class slope | 0.067 | 0.132 | 0.510 | 1.069 | 0.826 | 1.384 | 6.10e-01 | 1.00e+00 | 21196 | 17288 |
| European | No Recorded Depression | TCA:nortriptyline | Migraine | Within-class slope | -0.003 | 0.074 | -0.041 | 0.997 | 0.863 | 1.152 | 9.68e-01 | 1.00e+00 | 21196 | 17288 |
| European | No Recorded Depression | NaSSA:mirtazapine | Migraine | Interaction (vs SSRI:sertraline) | -0.005 | 0.106 | -0.045 | 0.995 | 0.809 | 1.225 | 9.64e-01 | 1.00e+00 | 21196 | 17288 |
| European | No Recorded Depression | NDRI:bupropion | Migraine | Interaction (vs SSRI:sertraline) | 0.010 | 0.064 | 0.164 | 1.011 | 0.891 | 1.146 | 8.70e-01 | 1.00e+00 | 21196 | 17288 |
| European | No Recorded Depression | SARI:trazodone | Migraine | Interaction (vs SSRI:sertraline) | -0.072 | 0.065 | -1.123 | 0.930 | 0.820 | 1.056 | 2.61e-01 | 1.00e+00 | 21196 | 17288 |
| European | No Recorded Depression | SNRI:duloxetine | Migraine | Interaction (vs SSRI:sertraline) | -0.111 | 0.067 | -1.658 | 0.895 | 0.785 | 1.020 | 9.73e-02 | 6.81e-01 | 21196 | 17288 |
| European | No Recorded Depression | SNRI:venlafaxine | Migraine | Interaction (vs SSRI:sertraline) | 0.058 | 0.072 | 0.807 | 1.060 | 0.920 | 1.221 | 4.20e-01 | 1.00e+00 | 21196 | 17288 |
| European | No Recorded Depression | SSRI:citalopram | Migraine | Interaction (vs SSRI:sertraline) | 0.020 | 0.066 | 0.295 | 1.020 | 0.895 | 1.161 | 7.68e-01 | 1.00e+00 | 21196 | 17288 |
| European | No Recorded Depression | SSRI:escitalopram | Migraine | Interaction (vs SSRI:sertraline) | -0.010 | 0.062 | -0.168 | 0.990 | 0.876 | 1.118 | 8.66e-01 | 1.00e+00 | 21196 | 17288 |
| European | No Recorded Depression | SSRI:fluoxetine | Migraine | Interaction (vs SSRI:sertraline) | -0.028 | 0.067 | -0.419 | 0.972 | 0.852 | 1.109 | 6.75e-01 | 1.00e+00 | 21196 | 17288 |
| European | No Recorded Depression | SSRI:paroxetine | Migraine | Interaction (vs SSRI:sertraline) | -0.115 | 0.088 | -1.307 | 0.891 | 0.749 | 1.059 | 1.91e-01 | 1.00e+00 | 21196 | 17288 |
| European | No Recorded Depression | TCA:amitriptyline | Migraine | Interaction (vs SSRI:sertraline) | 0.010 | 0.072 | 0.140 | 1.010 | 0.878 | 1.162 | 8.89e-01 | 1.00e+00 | 21196 | 17288 |
| European | No Recorded Depression | TCA:doxepin | Migraine | Interaction (vs SSRI:sertraline) | 0.018 | 0.138 | 0.132 | 1.018 | 0.777 | 1.335 | 8.95e-01 | 1.00e+00 | 21196 | 17288 |
| European | No Recorded Depression | TCA:nortriptyline | Migraine | Interaction (vs SSRI:sertraline) | -0.052 | 0.084 | -0.617 | 0.949 | 0.805 | 1.119 | 5.37e-01 | 1.00e+00 | 21196 | 17288 |
| European | No Recorded Depression | | Migraine | Joint_Interaction_Test | | | 9.005 | | | | 7.03e-01 | 1.00e+00 | 21196 | 17288 |
| European | No Recorded Depression | SSRI:sertraline | Insomnia | Within-class slope | 0.109 | 0.045 | 2.408 | 1.115 | 1.020 | 1.218 | 1.60e-02 | 1.12e-01 | 21196 | 17288 |
| European | No Recorded Depression | NaSSA:mirtazapine | Insomnia | Within-class slope | 0.062 | 0.103 | 0.607 | 1.064 | 0.870 | 1.301 | 5.44e-01 | 1.00e+00 | 21196 | 17288 |
| European | No Recorded Depression | NDRI:bupropion | Insomnia | Within-class slope | 0.070 | 0.052 | 1.360 | 1.073 | 0.969 | 1.188 | 1.74e-01 | 1.00e+00 | 21196 | 17288 |
| European | No Recorded Depression | SARI:trazodone | Insomnia | Within-class slope | 0.121 | 0.051 | 2.382 | 1.129 | 1.022 | 1.248 | 1.72e-02 | 1.21e-01 | 21196 | 17288 |
| European | No Recorded Depression | SNRI:duloxetine | Insomnia | Within-class slope | 0.112 | 0.058 | 1.937 | 1.119 | 0.999 | 1.253 | 5.27e-02 | 3.69e-01 | 21196 | 17288 |
| European | No Recorded Depression | SNRI:venlafaxine | Insomnia | Within-class slope | 0.094 | 0.064 | 1.461 | 1.099 | 0.968 | 1.246 | 1.44e-01 | 1.00e+00 | 21196 | 17288 |
| European | No Recorded Depression | SSRI:citalopram | Insomnia | Within-class slope | 0.058 | 0.055 | 1.061 | 1.060 | 0.952 | 1.181 | 2.89e-01 | 1.00e+00 | 21196 | 17288 |
| European | No Recorded Depression | SSRI:escitalopram | Insomnia | Within-class slope | 0.176 | 0.049 | 3.554 | 1.192 | 1.082 | 1.313 | 3.79e-04 | 2.65e-03 | 21196 | 17288 |
| European | No Recorded Depression | SSRI:fluoxetine | Insomnia | Within-class slope | 0.182 | 0.059 | 3.079 | 1.200 | 1.069 | 1.348 | 2.08e-03 | 1.45e-02 | 21196 | 17288 |
| European | No Recorded Depression | SSRI:paroxetine | Insomnia | Within-class slope | 0.169 | 0.089 | 1.896 | 1.185 | 0.994 | 1.412 | 5.79e-02 | 4.05e-01 | 21196 | 17288 |
| European | No Recorded Depression | TCA:amitriptyline | Insomnia | Within-class slope | 0.104 | 0.059 | 1.771 | 1.110 | 0.989 | 1.245 | 7.66e-02 | 5.36e-01 | 21196 | 17288 |
| European | No Recorded Depression | TCA:doxepin | Insomnia | Within-class slope | 0.241 | 0.150 | 1.605 | 1.273 | 0.948 | 1.709 | 1.08e-01 | 7.59e-01 | 21196 | 17288 |
| European | No Recorded Depression | TCA:nortriptyline | Insomnia | Within-class slope | 0.199 | 0.081 | 2.469 | 1.220 | 1.042 | 1.429 | 1.36e-02 | 9.49e-02 | 21196 | 17288 |
| European | No Recorded Depression | NaSSA:mirtazapine | Insomnia | Interaction (vs SSRI:sertraline) | -0.047 | 0.111 | -0.419 | 0.954 | 0.767 | 1.187 | 6.75e-01 | 1.00e+00 | 21196 | 17288 |
| European | No Recorded Depression | NDRI:bupropion | Insomnia | Interaction (vs SSRI:sertraline) | -0.038 | 0.067 | -0.575 | 0.962 | 0.844 | 1.097 | 5.65e-01 | 1.00e+00 | 21196 | 17288 |
| European | No Recorded Depression | SARI:trazodone | Insomnia | Interaction (vs SSRI:sertraline) | 0.013 | 0.067 | 0.186 | 1.013 | 0.888 | 1.155 | 8.52e-01 | 1.00e+00 | 21196 | 17288 |
| European | No Recorded Depression | SNRI:duloxetine | Insomnia | Interaction (vs SSRI:sertraline) | 0.003 | 0.073 | 0.043 | 1.003 | 0.870 | 1.157 | 9.66e-01 | 1.00e+00 | 21196 | 17288 |
| European | No Recorded Depression | SNRI:venlafaxine | Insomnia | Interaction (vs SSRI:sertraline) | -0.015 | 0.077 | -0.193 | 0.985 | 0.847 | 1.146 | 8.47e-01 | 1.00e+00 | 21196 | 17288 |
| European | No Recorded Depression | SSRI:citalopram | Insomnia | Interaction (vs SSRI:sertraline) | -0.051 | 0.070 | -0.724 | 0.951 | 0.829 | 1.090 | 4.69e-01 | 1.00e+00 | 21196 | 17288 |
| European | No Recorded Depression | SSRI:escitalopram | Insomnia | Interaction (vs SSRI:sertraline) | 0.067 | 0.065 | 1.022 | 1.069 | 0.941 | 1.215 | 3.07e-01 | 1.00e+00 | 21196 | 17288 |
| European | No Recorded Depression | SSRI:fluoxetine | Insomnia | Interaction (vs SSRI:sertraline) | 0.073 | 0.073 | 1.003 | 1.076 | 0.932 | 1.243 | 3.16e-01 | 1.00e+00 | 21196 | 17288 |
| European | No Recorded Depression | SSRI:paroxetine | Insomnia | Interaction (vs SSRI:sertraline) | 0.061 | 0.099 | 0.610 | 1.062 | 0.875 | 1.290 | 5.42e-01 | 1.00e+00 | 21196 | 17288 |
| European | No Recorded Depression | TCA:amitriptyline | Insomnia | Interaction (vs SSRI:sertraline) | -0.005 | 0.073 | -0.069 | 0.995 | 0.862 | 1.148 | 9.45e-01 | 1.00e+00 | 21196 | 17288 |
| European | No Recorded Depression | TCA:doxepin | Insomnia | Interaction (vs SSRI:sertraline) | 0.132 | 0.157 | 0.846 | 1.142 | 0.840 | 1.551 | 3.98e-01 | 1.00e+00 | 21196 | 17288 |
| European | No Recorded Depression | TCA:nortriptyline | Insomnia | Interaction (vs SSRI:sertraline) | 0.090 | 0.091 | 0.992 | 1.094 | 0.916 | 1.307 | 3.21e-01 | 1.00e+00 | 21196 | 17288 |
| European | No Recorded Depression | | Insomnia | Joint_Interaction_Test | | | 7.591 | | | | 8.16e-01 | 1.00e+00 | 21196 | 17288 |
| European | No Recorded Depression | SSRI:sertraline | Depression | Within-class slope | 0.182 | 0.044 | 4.085 | 1.199 | 1.099 | 1.308 | 4.40e-05 | 3.08e-04 | 21196 | 17288 |
| European | No Recorded Depression | NaSSA:mirtazapine | Depression | Within-class slope | -0.037 | 0.097 | -0.383 | 0.964 | 0.797 | 1.165 | 7.02e-01 | 1.00e+00 | 21196 | 17288 |
| European | No Recorded Depression | NDRI:bupropion | Depression | Within-class slope | 0.201 | 0.050 | 4.018 | 1.222 | 1.108 | 1.348 | 5.86e-05 | 4.11e-04 | 21196 | 17288 |
| European | No Recorded Depression | SARI:trazodone | Depression | Within-class slope | 0.168 | 0.051 | 3.302 | 1.183 | 1.071 | 1.307 | 9.59e-04 | 6.71e-03 | 21196 | 17288 |
| European | No Recorded Depression | SNRI:duloxetine | Depression | Within-class slope | 0.232 | 0.058 | 4.031 | 1.262 | 1.127 | 1.413 | 5.56e-05 | 3.89e-04 | 21196 | 17288 |
| European | No Recorded Depression | SNRI:venlafaxine | Depression | Within-class slope | 0.300 | 0.067 | 4.476 | 1.350 | 1.184 | 1.540 | 7.59e-06 | 5.31e-05 | 21196 | 17288 |
| European | No Recorded Depression | SSRI:citalopram | Depression | Within-class slope | 0.031 | 0.055 | 0.556 | 1.031 | 0.925 | 1.150 | 5.78e-01 | 1.00e+00 | 21196 | 17288 |
| European | No Recorded Depression | SSRI:escitalopram | Depression | Within-class slope | 0.194 | 0.050 | 3.851 | 1.214 | 1.100 | 1.339 | 1.18e-04 | 8.25e-04 | 21196 | 17288 |
| European | No Recorded Depression | SSRI:fluoxetine | Depression | Within-class slope | 0.175 | 0.059 | 2.990 | 1.192 | 1.062 | 1.337 | 2.79e-03 | 1.95e-02 | 21196 | 17288 |
| European | No Recorded Depression | SSRI:paroxetine | Depression | Within-class slope | 0.189 | 0.090 | 2.107 | 1.209 | 1.013 | 1.442 | 3.51e-02 | 2.46e-01 | 21196 | 17288 |
| European | No Recorded Depression | TCA:amitriptyline | Depression | Within-class slope | 0.202 | 0.060 | 3.371 | 1.224 | 1.088 | 1.377 | 7.48e-04 | 5.23e-03 | 21196 | 17288 |
| European | No Recorded Depression | TCA:doxepin | Depression | Within-class slope | 0.311 | 0.143 | 2.172 | 1.365 | 1.031 | 1.807 | 2.99e-02 | 2.09e-01 | 21196 | 17288 |
| European | No Recorded Depression | TCA:nortriptyline | Depression | Within-class slope | 0.058 | 0.077 | 0.756 | 1.060 | 0.911 | 1.234 | 4.50e-01 | 1.00e+00 | 21196 | 17288 |
| European | No Recorded Depression | NaSSA:mirtazapine | Depression | Interaction (vs SSRI:sertraline) | -0.219 | 0.106 | -2.060 | 0.804 | 0.653 | 0.989 | 3.94e-02 | 2.76e-01 | 21196 | 17288 |
| European | No Recorded Depression | NDRI:bupropion | Depression | Interaction (vs SSRI:sertraline) | 0.019 | 0.065 | 0.292 | 1.019 | 0.897 | 1.158 | 7.70e-01 | 1.00e+00 | 21196 | 17288 |
| European | No Recorded Depression | SARI:trazodone | Depression | Interaction (vs SSRI:sertraline) | -0.014 | 0.067 | -0.203 | 0.987 | 0.866 | 1.124 | 8.39e-01 | 1.00e+00 | 21196 | 17288 |

| | | | | | | | | | | | | | | |
|---|---|---|---|---|---|---|---|---|---|---|---|---|---|---|
| European | No Recorded Depression | SNRI:duloxetine | Depression | Interaction (vs SSRI:sertraline) | 0.051 | 0.072 | 0.708 | 1.052 | 0.914 | 1.211 | 4.79e-01 | 1.00e+00 | 21196 | 17288 |
| European | No Recorded Depression | SNRI:venlafaxine | Depression | Interaction (vs SSRI:sertraline) | 0.119 | 0.079 | 1.509 | 1.126 | 0.965 | 1.313 | 1.31e-01 | 9.19e-01 | 21196 | 17288 |
| European | No Recorded Depression | SSRI:citalopram | Depression | Interaction (vs SSRI:sertraline) | -0.151 | 0.070 | -2.168 | 0.860 | 0.750 | 0.986 | 3.02e-02 | 2.11e-01 | 21196 | 17288 |
| European | No Recorded Depression | SSRI:escitalopram | Depression | Interaction (vs SSRI:sertraline) | 0.012 | 0.065 | 0.184 | 1.012 | 0.890 | 1.151 | 8.54e-01 | 1.00e+00 | 21196 | 17288 |
| European | No Recorded Depression | SSRI:fluoxetine | Depression | Interaction (vs SSRI:sertraline) | -0.006 | 0.072 | -0.088 | 0.994 | 0.863 | 1.144 | 9.30e-01 | 1.00e+00 | 21196 | 17288 |
| European | No Recorded Depression | SSRI:paroxetine | Depression | Interaction (vs SSRI:sertraline) | 0.008 | 0.100 | 0.080 | 1.008 | 0.829 | 1.225 | 9.37e-01 | 1.00e+00 | 21196 | 17288 |
| European | No Recorded Depression | TCA:amitriptyline | Depression | Interaction (vs SSRI:sertraline) | 0.021 | 0.074 | 0.281 | 1.021 | 0.883 | 1.181 | 7.79e-01 | 1.00e+00 | 21196 | 17288 |
| European | No Recorded Depression | TCA:doxepin | Depression | Interaction (vs SSRI:sertraline) | 0.129 | 0.150 | 0.863 | 1.138 | 0.849 | 1.526 | 3.88e-01 | 1.00e+00 | 21196 | 17288 |
| European | No Recorded Depression | TCA:nortriptyline | Depression | Interaction (vs SSRI:sertraline) | -0.123 | 0.089 | -1.389 | 0.884 | 0.743 | 1.052 | 1.65e-01 | 1.00e+00 | 21196 | 17288 |
| European | No Recorded Depression | | Depression | Joint_Interaction_Test | | | 21.029 | | | | 5.00e-02 | 3.50e-01 | 21196 | 17288 |
| European | No Recorded Depression | SSRI:sertraline | Schizophrenia | Within-class slope | 0.057 | 0.041 | 1.373 | 1.059 | 0.976 | 1.148 | 1.70e-01 | 1.00e+00 | 21196 | 17288 |
| European | No Recorded Depression | NaSSA:mirtazapine | Schizophrenia | Within-class slope | -0.051 | 0.100 | -0.506 | 0.951 | 0.782 | 1.156 | 6.13e-01 | 1.00e+00 | 21196 | 17288 |
| European | No Recorded Depression | NDRI:bupropion | Schizophrenia | Within-class slope | 0.018 | 0.049 | 0.370 | 1.018 | 0.925 | 1.120 | 7.12e-01 | 1.00e+00 | 21196 | 17288 |
| European | No Recorded Depression | SARI:trazodone | Schizophrenia | Within-class slope | 0.096 | 0.052 | 1.866 | 1.101 | 0.995 | 1.218 | 6.21e-02 | 4.35e-01 | 21196 | 17288 |
| European | No Recorded Depression | SNRI:duloxetine | Schizophrenia | Within-class slope | 0.069 | 0.055 | 1.253 | 1.072 | 0.962 | 1.195 | 2.10e-01 | 1.00e+00 | 21196 | 17288 |
| European | No Recorded Depression | SNRI:venlafaxine | Schizophrenia | Within-class slope | 0.051 | 0.062 | 0.817 | 1.052 | 0.932 | 1.188 | 4.14e-01 | 1.00e+00 | 21196 | 17288 |
| European | No Recorded Depression | SSRI:citalopram | Schizophrenia | Within-class slope | 0.013 | 0.053 | 0.252 | 1.013 | 0.914 | 1.124 | 8.01e-01 | 1.00e+00 | 21196 | 17288 |
| European | No Recorded Depression | SSRI:escitalopram | Schizophrenia | Within-class slope | 0.024 | 0.048 | 0.490 | 1.024 | 0.932 | 1.125 | 6.24e-01 | 1.00e+00 | 21196 | 17288 |
| European | No Recorded Depression | SSRI:fluoxetine | Schizophrenia | Within-class slope | -0.051 | 0.057 | -0.905 | 0.950 | 0.850 | 1.062 | 3.65e-01 | 1.00e+00 | 21196 | 17288 |
| European | No Recorded Depression | SSRI:paroxetine | Schizophrenia | Within-class slope | 0.044 | 0.086 | 0.504 | 1.045 | 0.882 | 1.237 | 6.14e-01 | 1.00e+00 | 21196 | 17288 |
| European | No Recorded Depression | TCA:amitriptyline | Schizophrenia | Within-class slope | -0.006 | 0.059 | -0.107 | 0.994 | 0.885 | 1.115 | 9.15e-01 | 1.00e+00 | 21196 | 17288 |
| European | No Recorded Depression | TCA:doxepin | Schizophrenia | Within-class slope | -0.017 | 0.129 | -0.132 | 0.983 | 0.763 | 1.267 | 8.95e-01 | 1.00e+00 | 21196 | 17288 |
| European | No Recorded Depression | TCA:nortriptyline | Schizophrenia | Within-class slope | -0.080 | 0.078 | -1.021 | 0.923 | 0.791 | 1.077 | 3.07e-01 | 1.00e+00 | 21196 | 17288 |
| European | No Recorded Depression | NaSSA:mirtazapine | Schizophrenia | Interaction (vs SSRI:sertraline) | -0.107 | 0.107 | -1.001 | 0.898 | 0.728 | 1.108 | 3.17e-01 | 1.00e+00 | 21196 | 17288 |
| European | No Recorded Depression | NDRI:bupropion | Schizophrenia | Interaction (vs SSRI:sertraline) | -0.039 | 0.063 | -0.620 | 0.962 | 0.851 | 1.088 | 5.35e-01 | 1.00e+00 | 21196 | 17288 |
| European | No Recorded Depression | SARI:trazodone | Schizophrenia | Interaction (vs SSRI:sertraline) | 0.039 | 0.065 | 0.604 | 1.040 | 0.915 | 1.182 | 5.46e-01 | 1.00e+00 | 21196 | 17288 |
| European | No Recorded Depression | SNRI:duloxetine | Schizophrenia | Interaction (vs SSRI:sertraline) | 0.012 | 0.068 | 0.183 | 1.013 | 0.886 | 1.157 | 8.55e-01 | 1.00e+00 | 21196 | 17288 |
| European | No Recorded Depression | SNRI:venlafaxine | Schizophrenia | Interaction (vs SSRI:sertraline) | -0.006 | 0.074 | -0.086 | 0.994 | 0.860 | 1.148 | 9.32e-01 | 1.00e+00 | 21196 | 17288 |
| European | No Recorded Depression | SSRI:citalopram | Schizophrenia | Interaction (vs SSRI:sertraline) | -0.044 | 0.066 | -0.660 | 0.957 | 0.841 | 1.090 | 5.09e-01 | 1.00e+00 | 21196 | 17288 |
| European | No Recorded Depression | SSRI:escitalopram | Schizophrenia | Interaction (vs SSRI:sertraline) | -0.033 | 0.063 | -0.530 | 0.967 | 0.855 | 1.094 | 5.96e-01 | 1.00e+00 | 21196 | 17288 |
| European | No Recorded Depression | SSRI:fluoxetine | Schizophrenia | Interaction (vs SSRI:sertraline) | -0.108 | 0.069 | -1.568 | 0.897 | 0.784 | 1.027 | 1.17e-01 | 8.18e-01 | 21196 | 17288 |
| European | No Recorded Depression | SSRI:paroxetine | Schizophrenia | Interaction (vs SSRI:sertraline) | -0.013 | 0.094 | -0.142 | 0.987 | 0.820 | 1.187 | 8.87e-01 | 1.00e+00 | 21196 | 17288 |
| European | No Recorded Depression | TCA:amitriptyline | Schizophrenia | Interaction (vs SSRI:sertraline) | -0.063 | 0.071 | -0.894 | 0.939 | 0.817 | 1.078 | 3.71e-01 | 1.00e+00 | 21196 | 17288 |
| European | No Recorded Depression | TCA:doxepin | Schizophrenia | Interaction (vs SSRI:sertraline) | -0.074 | 0.135 | -0.546 | 0.929 | 0.712 | 1.211 | 5.85e-01 | 1.00e+00 | 21196 | 17288 |
| European | No Recorded Depression | TCA:nortriptyline | Schizophrenia | Interaction (vs SSRI:sertraline) | -0.137 | 0.088 | -1.562 | 0.872 | 0.734 | 1.036 | 1.18e-01 | 8.28e-01 | 21196 | 17288 |
| European | No Recorded Depression | | Schizophrenia | Joint_Interaction_Test | | | 8.115 | | | | 7.76e-01 | 1.00e+00 | 21196 | 17288 |
| European | No Recorded Depression | SSRI:sertraline | Bipolar Disorder | Within-class slope | 0.002 | 0.045 | 0.042 | 1.002 | 0.917 | 1.094 | 9.66e-01 | 1.00e+00 | 21196 | 17288 |
| European | No Recorded Depression | NaSSA:mirtazapine | Bipolar Disorder | Within-class slope | -0.028 | 0.111 | -0.257 | 0.972 | 0.782 | 1.208 | 7.97e-01 | 1.00e+00 | 21196 | 17288 |
| European | No Recorded Depression | NDRI:bupropion | Bipolar Disorder | Within-class slope | 0.093 | 0.050 | 1.848 | 1.098 | 0.994 | 1.212 | 6.46e-02 | 4.52e-01 | 21196 | 17288 |
| European | No Recorded Depression | SARI:trazodone | Bipolar Disorder | Within-class slope | 0.151 | 0.054 | 2.806 | 1.163 | 1.046 | 1.292 | 5.02e-03 | 3.51e-02 | 21196 | 17288 |
| European | No Recorded Depression | SNRI:duloxetine | Bipolar Disorder | Within-class slope | 0.104 | 0.057 | 1.806 | 1.109 | 0.991 | 1.241 | 7.09e-02 | 4.96e-01 | 21196 | 17288 |
| European | No Recorded Depression | SNRI:venlafaxine | Bipolar Disorder | Within-class slope | -0.057 | 0.065 | -0.879 | 0.945 | 0.832 | 1.072 | 3.79e-01 | 1.00e+00 | 21196 | 17288 |
| European | No Recorded Depression | SSRI:citalopram | Bipolar Disorder | Within-class slope | 0.132 | 0.055 | 2.406 | 1.141 | 1.025 | 1.270 | 1.61e-02 | 1.13e-01 | 21196 | 17288 |
| European | No Recorded Depression | SSRI:escitalopram | Bipolar Disorder | Within-class slope | 0.144 | 0.049 | 2.922 | 1.155 | 1.049 | 1.273 | 3.48e-03 | 2.44e-02 | 21196 | 17288 |
| European | No Recorded Depression | SSRI:fluoxetine | Bipolar Disorder | Within-class slope | -0.005 | 0.057 | -0.085 | 0.995 | 0.889 | 1.113 | 9.32e-01 | 1.00e+00 | 21196 | 17288 |
| European | No Recorded Depression | SSRI:paroxetine | Bipolar Disorder | Within-class slope | 0.003 | 0.084 | 0.037 | 1.003 | 0.850 | 1.183 | 9.71e-01 | 1.00e+00 | 21196 | 17288 |
| European | No Recorded Depression | TCA:amitriptyline | Bipolar Disorder | Within-class slope | 0.095 | 0.061 | 1.553 | 1.100 | 0.975 | 1.241 | 1.20e-01 | 8.42e-01 | 21196 | 17288 |
| European | No Recorded Depression | TCA:doxepin | Bipolar Disorder | Within-class slope | -0.042 | 0.150 | -0.279 | 0.959 | 0.715 | 1.287 | 7.81e-01 | 1.00e+00 | 21196 | 17288 |
| European | No Recorded Depression | TCA:nortriptyline | Bipolar Disorder | Within-class slope | 0.016 | 0.077 | 0.208 | 1.016 | 0.874 | 1.181 | 8.35e-01 | 1.00e+00 | 21196 | 17288 |
| European | No Recorded Depression | NaSSA:mirtazapine | Bipolar Disorder | Interaction (vs SSRI:sertraline) | -0.030 | 0.119 | -0.256 | 0.970 | 0.769 | 1.225 | 7.98e-01 | 1.00e+00 | 21196 | 17288 |
| European | No Recorded Depression | NDRI:bupropion | Bipolar Disorder | Interaction (vs SSRI:sertraline) | 0.091 | 0.066 | 1.386 | 1.096 | 0.963 | 1.247 | 1.66e-01 | 1.00e+00 | 21196 | 17288 |
| European | No Recorded Depression | SARI:trazodone | Bipolar Disorder | Interaction (vs SSRI:sertraline) | 0.149 | 0.069 | 2.149 | 1.160 | 1.013 | 1.329 | 3.17e-02 | 2.22e-01 | 21196 | 17288 |
| European | No Recorded Depression | SNRI:duloxetine | Bipolar Disorder | Interaction (vs SSRI:sertraline) | 0.102 | 0.072 | 1.423 | 1.107 | 0.962 | 1.274 | 1.55e-01 | 1.00e+00 | 21196 | 17288 |
| European | No Recorded Depression | SNRI:venlafaxine | Bipolar Disorder | Interaction (vs SSRI:sertraline) | -0.059 | 0.077 | -0.763 | 0.943 | 0.811 | 1.097 | 4.46e-01 | 1.00e+00 | 21196 | 17288 |
| European | No Recorded Depression | SSRI:citalopram | Bipolar Disorder | Interaction (vs SSRI:sertraline) | 0.130 | 0.070 | 1.864 | 1.139 | 0.993 | 1.306 | 6.23e-02 | 4.36e-01 | 21196 | 17288 |
| European | No Recorded Depression | SSRI:escitalopram | Bipolar Disorder | Interaction (vs SSRI:sertraline) | 0.143 | 0.066 | 2.170 | 1.153 | 1.014 | 1.312 | 3.00e-02 | 2.10e-01 | 21196 | 17288 |
| European | No Recorded Depression | SSRI:fluoxetine | Bipolar Disorder | Interaction (vs SSRI:sertraline) | -0.007 | 0.071 | -0.095 | 0.993 | 0.864 | 1.142 | 9.24e-01 | 1.00e+00 | 21196 | 17288 |
| European | No Recorded Depression | SSRI:paroxetine | Bipolar Disorder | Interaction (vs SSRI:sertraline) | 0.001 | 0.094 | 0.013 | 1.001 | 0.833 | 1.203 | 9.90e-01 | 1.00e+00 | 21196 | 17288 |
| European | No Recorded Depression | TCA:amitriptyline | Bipolar Disorder | Interaction (vs SSRI:sertraline) | 0.094 | 0.075 | 1.247 | 1.098 | 0.948 | 1.272 | 2.12e-01 | 1.00e+00 | 21196 | 17288 |

| | | | | | | | | | | | | | |
|---|---|---|---|---|---|---|---|---|---|---|---|---|---|
| European | No Recorded Depression | TCA:doxepin | Bipolar Disorder | Interaction (vs SSRI:sertraline) | -0.044 | 0.156 | -0.280 | 0.957 | 0.705 | 1.300 | 7.80e-01 | 1.00e+00 | 21196 | 17288 |
| European | No Recorded Depression | TCA:nortriptyline | Bipolar Disorder | Interaction (vs SSRI:sertraline) | 0.014 | 0.088 | 0.159 | 1.014 | 0.853 | 1.205 | 8.74e-01 | 1.00e+00 | 21196 | 17288 |
| European | No Recorded Depression | | Bipolar Disorder | Joint_Interaction_Test | | | 17.733 | | | | 1.24e-01 | 8.68e-01 | 21196 | 17288 |
| European | No Recorded Depression | SSRI:sertraline | BMI | Within-class slope | 0.105 | 0.042 | 2.482 | 1.110 | 1.022 | 1.206 | 1.31e-02 | 9.14e-02 | 21196 | 17288 |
| European | No Recorded Depression | NaSSA:mirtazapine | BMI | Within-class slope | -0.003 | 0.099 | -0.030 | 0.997 | 0.822 | 1.209 | 9.76e-01 | 1.00e+00 | 21196 | 17288 |
| European | No Recorded Depression | NDRI:bupropion | BMI | Within-class slope | -0.018 | 0.051 | -0.351 | 0.982 | 0.889 | 1.085 | 7.25e-01 | 1.00e+00 | 21196 | 17288 |
| European | No Recorded Depression | SARI:trazodone | BMI | Within-class slope | 0.042 | 0.051 | 0.813 | 1.043 | 0.943 | 1.153 | 4.16e-01 | 1.00e+00 | 21196 | 17288 |
| European | No Recorded Depression | SNRI:duloxetine | BMI | Within-class slope | -0.031 | 0.056 | -0.564 | 0.969 | 0.869 | 1.081 | 5.73e-01 | 1.00e+00 | 21196 | 17288 |
| European | No Recorded Depression | SNRI:venlafaxine | BMI | Within-class slope | 0.101 | 0.060 | 1.677 | 1.106 | 0.983 | 1.244 | 9.36e-02 | 6.55e-01 | 21196 | 17288 |
| European | No Recorded Depression | SSRI:citalopram | BMI | Within-class slope | 0.010 | 0.052 | 0.189 | 1.010 | 0.912 | 1.118 | 8.50e-01 | 1.00e+00 | 21196 | 17288 |
| European | No Recorded Depression | SSRI:escitalopram | BMI | Within-class slope | 0.145 | 0.050 | 2.914 | 1.156 | 1.049 | 1.275 | 3.57e-03 | 2.50e-02 | 21196 | 17288 |
| European | No Recorded Depression | SSRI:fluoxetine | BMI | Within-class slope | 0.084 | 0.054 | 1.559 | 1.088 | 0.979 | 1.208 | 1.19e-01 | 8.33e-01 | 21196 | 17288 |
| European | No Recorded Depression | SSRI:paroxetine | BMI | Within-class slope | 0.129 | 0.087 | 1.477 | 1.137 | 0.959 | 1.349 | 1.40e-01 | 9.77e-01 | 21196 | 17288 |
| European | No Recorded Depression | TCA:amitriptyline | BMI | Within-class slope | 0.022 | 0.059 | 0.368 | 1.022 | 0.910 | 1.148 | 7.13e-01 | 1.00e+00 | 21196 | 17288 |
| European | No Recorded Depression | TCA:doxepin | BMI | Within-class slope | 0.112 | 0.144 | 0.773 | 1.118 | 0.842 | 1.484 | 4.40e-01 | 1.00e+00 | 21196 | 17288 |
| European | No Recorded Depression | TCA:nortriptyline | BMI | Within-class slope | 0.131 | 0.078 | 1.673 | 1.140 | 0.978 | 1.329 | 9.44e-02 | 6.61e-01 | 21196 | 17288 |
| European | No Recorded Depression | NaSSA:mirtazapine | BMI | Interaction (vs SSRI:sertraline) | -0.108 | 0.106 | -1.013 | 0.898 | 0.729 | 1.106 | 3.11e-01 | 1.00e+00 | 21196 | 17288 |
| European | No Recorded Depression | NDRI:bupropion | BMI | Interaction (vs SSRI:sertraline) | -0.122 | 0.064 | -1.924 | 0.885 | 0.781 | 1.002 | 5.44e-02 | 3.81e-01 | 21196 | 17288 |
| European | No Recorded Depression | SARI:trazodone | BMI | Interaction (vs SSRI:sertraline) | -0.063 | 0.065 | -0.965 | 0.939 | 0.826 | 1.067 | 3.35e-01 | 1.00e+00 | 21196 | 17288 |
| European | No Recorded Depression | SNRI:duloxetine | BMI | Interaction (vs SSRI:sertraline) | -0.136 | 0.068 | -1.994 | 0.873 | 0.764 | 0.998 | 4.61e-02 | 3.23e-01 | 21196 | 17288 |
| European | No Recorded Depression | SNRI:venlafaxine | BMI | Interaction (vs SSRI:sertraline) | -0.004 | 0.073 | -0.052 | 0.996 | 0.863 | 1.150 | 9.59e-01 | 1.00e+00 | 21196 | 17288 |
| European | No Recorded Depression | SSRI:citalopram | BMI | Interaction (vs SSRI:sertraline) | -0.095 | 0.066 | -1.442 | 0.910 | 0.800 | 1.035 | 1.49e-01 | 1.00e+00 | 21196 | 17288 |
| European | No Recorded Depression | SSRI:escitalopram | BMI | Interaction (vs SSRI:sertraline) | 0.041 | 0.065 | 0.627 | 1.042 | 0.917 | 1.183 | 5.30e-01 | 1.00e+00 | 21196 | 17288 |
| European | No Recorded Depression | SSRI:fluoxetine | BMI | Interaction (vs SSRI:sertraline) | -0.021 | 0.066 | -0.314 | 0.980 | 0.861 | 1.115 | 7.54e-01 | 1.00e+00 | 21196 | 17288 |
| European | No Recorded Depression | SSRI:paroxetine | BMI | Interaction (vs SSRI:sertraline) | 0.024 | 0.096 | 0.252 | 1.024 | 0.849 | 1.236 | 8.01e-01 | 1.00e+00 | 21196 | 17288 |
| European | No Recorded Depression | TCA:amitriptyline | BMI | Interaction (vs SSRI:sertraline) | -0.083 | 0.072 | -1.143 | 0.921 | 0.799 | 1.061 | 2.53e-01 | 1.00e+00 | 21196 | 17288 |
| European | No Recorded Depression | TCA:doxepin | BMI | Interaction (vs SSRI:sertraline) | 0.007 | 0.150 | 0.047 | 1.007 | 0.751 | 1.351 | 9.63e-01 | 1.00e+00 | 21196 | 17288 |
| European | No Recorded Depression | TCA:nortriptyline | BMI | Interaction (vs SSRI:sertraline) | 0.026 | 0.088 | 0.302 | 1.027 | 0.865 | 1.219 | 7.63e-01 | 1.00e+00 | 21196 | 17288 |
| European | No Recorded Depression | | BMI | Joint_Interaction_Test | | | 14.177 | | | | 2.90e-01 | 1.00e+00 | 21196 | 17288 |
| European | Recorded Anxiety | SSRI:sertraline | ADHD | Within-class slope | 0.072 | 0.029 | 2.514 | 1.075 | 1.016 | 1.137 | 1.20e-02 | 8.37e-02 | 32515 | 22457 |
| European | Recorded Anxiety | NaSSA:mirtazapine | ADHD | Within-class slope | 0.025 | 0.074 | 0.339 | 1.025 | 0.888 | 1.184 | 7.34e-01 | 1.00e+00 | 32515 | 22457 |
| European | Recorded Anxiety | NDRI:bupropion | ADHD | Within-class slope | -0.012 | 0.036 | -0.342 | 0.988 | 0.921 | 1.060 | 7.32e-01 | 1.00e+00 | 32515 | 22457 |
| European | Recorded Anxiety | SARI:trazodone | ADHD | Within-class slope | 0.020 | 0.044 | 0.446 | 1.020 | 0.935 | 1.113 | 6.56e-01 | 1.00e+00 | 32515 | 22457 |
| European | Recorded Anxiety | SNRI:duloxetine | ADHD | Within-class slope | 0.031 | 0.042 | 0.735 | 1.031 | 0.950 | 1.119 | 4.62e-01 | 1.00e+00 | 32515 | 22457 |
| European | Recorded Anxiety | SNRI:venlafaxine | ADHD | Within-class slope | 0.023 | 0.041 | 0.562 | 1.023 | 0.944 | 1.109 | 5.74e-01 | 1.00e+00 | 32515 | 22457 |
| European | Recorded Anxiety | SSRI:citalopram | ADHD | Within-class slope | 0.028 | 0.035 | 0.793 | 1.028 | 0.960 | 1.102 | 4.28e-01 | 1.00e+00 | 32515 | 22457 |
| European | Recorded Anxiety | SSRI:escitalopram | ADHD | Within-class slope | 0.102 | 0.033 | 3.072 | 1.108 | 1.038 | 1.182 | 2.12e-03 | 1.49e-02 | 32515 | 22457 |
| European | Recorded Anxiety | SSRI:fluoxetine | ADHD | Within-class slope | -0.018 | 0.037 | -0.494 | 0.982 | 0.914 | 1.055 | 6.21e-01 | 1.00e+00 | 32515 | 22457 |
| European | Recorded Anxiety | SSRI:paroxetine | ADHD | Within-class slope | 0.125 | 0.059 | 2.119 | 1.133 | 1.009 | 1.273 | 3.41e-02 | 2.38e-01 | 32515 | 22457 |
| European | Recorded Anxiety | TCA:amitriptyline | ADHD | Within-class slope | 0.051 | 0.056 | 0.908 | 1.052 | 0.943 | 1.174 | 3.64e-01 | 1.00e+00 | 32515 | 22457 |
| European | Recorded Anxiety | TCA:doxepin | ADHD | Within-class slope | 0.071 | 0.136 | 0.521 | 1.073 | 0.823 | 1.401 | 6.02e-01 | 1.00e+00 | 32515 | 22457 |
| European | Recorded Anxiety | TCA:nortriptyline | ADHD | Within-class slope | -0.032 | 0.068 | -0.467 | 0.969 | 0.848 | 1.107 | 6.40e-01 | 1.00e+00 | 32515 | 22457 |
| European | Recorded Anxiety | NaSSA:mirtazapine | ADHD | Interaction (vs SSRI:sertraline) | -0.047 | 0.078 | -0.602 | 0.954 | 0.818 | 1.112 | 5.47e-01 | 1.00e+00 | 32515 | 22457 |
| European | Recorded Anxiety | NDRI:bupropion | ADHD | Interaction (vs SSRI:sertraline) | -0.084 | 0.044 | -1.922 | 0.919 | 0.843 | 1.002 | 5.46e-02 | 3.82e-01 | 32515 | 22457 |
| European | Recorded Anxiety | SARI:trazodone | ADHD | Interaction (vs SSRI:sertraline) | -0.052 | 0.052 | -1.013 | 0.949 | 0.858 | 1.050 | 3.11e-01 | 1.00e+00 | 32515 | 22457 |
| European | Recorded Anxiety | SNRI:duloxetine | ADHD | Interaction (vs SSRI:sertraline) | -0.041 | 0.049 | -0.843 | 0.959 | 0.871 | 1.056 | 3.99e-01 | 1.00e+00 | 32515 | 22457 |
| European | Recorded Anxiety | SNRI:venlafaxine | ADHD | Interaction (vs SSRI:sertraline) | -0.049 | 0.049 | -1.008 | 0.952 | 0.866 | 1.047 | 3.13e-01 | 1.00e+00 | 32515 | 22457 |
| European | Recorded Anxiety | SSRI:citalopram | ADHD | Interaction (vs SSRI:sertraline) | -0.044 | 0.044 | -1.000 | 0.957 | 0.878 | 1.043 | 3.17e-01 | 1.00e+00 | 32515 | 22457 |
| European | Recorded Anxiety | SSRI:escitalopram | ADHD | Interaction (vs SSRI:sertraline) | 0.030 | 0.042 | 0.710 | 1.031 | 0.948 | 1.120 | 4.78e-01 | 1.00e+00 | 32515 | 22457 |
| European | Recorded Anxiety | SSRI:fluoxetine | ADHD | Interaction (vs SSRI:sertraline) | -0.090 | 0.045 | -1.991 | 0.914 | 0.836 | 0.999 | 4.65e-02 | 3.26e-01 | 32515 | 22457 |
| European | Recorded Anxiety | SSRI:paroxetine | ADHD | Interaction (vs SSRI:sertraline) | 0.053 | 0.065 | 0.823 | 1.055 | 0.929 | 1.197 | 4.11e-01 | 1.00e+00 | 32515 | 22457 |
| European | Recorded Anxiety | TCA:amitriptyline | ADHD | Interaction (vs SSRI:sertraline) | -0.021 | 0.062 | -0.343 | 0.979 | 0.867 | 1.105 | 7.32e-01 | 1.00e+00 | 32515 | 22457 |
| European | Recorded Anxiety | TCA:doxepin | ADHD | Interaction (vs SSRI:sertraline) | -0.001 | 0.138 | -0.010 | 0.999 | 0.762 | 1.310 | 9.92e-01 | 1.00e+00 | 32515 | 22457 |
| European | Recorded Anxiety | TCA:nortriptyline | ADHD | Interaction (vs SSRI:sertraline) | -0.104 | 0.073 | -1.429 | 0.901 | 0.782 | 1.039 | 1.53e-01 | 1.00e+00 | 32515 | 22457 |
| European | Recorded Anxiety | | ADHD | Joint_Interaction_Test | | | 13.980 | | | | 3.02e-01 | 1.00e+00 | 32515 | 22457 |
| European | Recorded Anxiety | SSRI:sertraline | Migraine | Within-class slope | 0.075 | 0.028 | 2.658 | 1.077 | 1.020 | 1.138 | 7.86e-03 | 5.50e-02 | 32515 | 22457 |
| European | Recorded Anxiety | NaSSA:mirtazapine | Migraine | Within-class slope | 0.060 | 0.075 | 0.796 | 1.061 | 0.917 | 1.229 | 4.26e-01 | 1.00e+00 | 32515 | 22457 |
| European | Recorded Anxiety | NDRI:bupropion | Migraine | Within-class slope | 0.108 | 0.035 | 3.062 | 1.114 | 1.040 | 1.194 | 2.20e-03 | 1.54e-02 | 32515 | 22457 |
| European | Recorded Anxiety | SARI:trazodone | Migraine | Within-class slope | -0.006 | 0.043 | -0.142 | 0.994 | 0.913 | 1.082 | 8.87e-01 | 1.00e+00 | 32515 | 22457 |

| | | | | | | | | | | | | | | |
|---|---|---|---|---|---|---|---|---|---|---|---|---|---|---|
| European | Recorded Anxiety | SNRI:duloxetine | Migraine | Within-class slope | -0.027 | 0.040 | -0.675 | 0.973 | 0.900 | 1.052 | 5.00e-01 | 1.00e+00 | 32515 | 22457 |
| European | Recorded Anxiety | SNRI:venlafaxine | Migraine | Within-class slope | 0.081 | 0.041 | 1.977 | 1.084 | 1.001 | 1.175 | 4.80e-02 | 3.36e-01 | 32515 | 22457 |
| European | Recorded Anxiety | SSRI:citalopram | Migraine | Within-class slope | 0.041 | 0.035 | 1.184 | 1.042 | 0.973 | 1.115 | 2.36e-01 | 1.00e+00 | 32515 | 22457 |
| European | Recorded Anxiety | SSRI:escitalopram | Migraine | Within-class slope | 0.050 | 0.032 | 1.564 | 1.052 | 0.987 | 1.120 | 1.18e-01 | 8.26e-01 | 32515 | 22457 |
| European | Recorded Anxiety | SSRI:fluoxetine | Migraine | Within-class slope | 0.072 | 0.036 | 2.010 | 1.075 | 1.002 | 1.153 | 4.45e-02 | 3.11e-01 | 32515 | 22457 |
| European | Recorded Anxiety | SSRI:paroxetine | Migraine | Within-class slope | 0.029 | 0.055 | 0.525 | 1.029 | 0.924 | 1.147 | 6.00e-01 | 1.00e+00 | 32515 | 22457 |
| European | Recorded Anxiety | TCA:amitriptyline | Migraine | Within-class slope | 0.023 | 0.054 | 0.429 | 1.024 | 0.920 | 1.139 | 6.68e-01 | 1.00e+00 | 32515 | 22457 |
| European | Recorded Anxiety | TCA:doxepin | Migraine | Within-class slope | 0.214 | 0.130 | 1.654 | 1.239 | 0.961 | 1.597 | 9.81e-02 | 6.86e-01 | 32515 | 22457 |
| European | Recorded Anxiety | TCA:nortriptyline | Migraine | Within-class slope | -0.024 | 0.070 | -0.348 | 0.976 | 0.852 | 1.119 | 7.28e-01 | 1.00e+00 | 32515 | 22457 |
| European | Recorded Anxiety | NaSSA:mirtazapine | Migraine | Interaction (vs SSRI:sertraline) | -0.015 | 0.079 | -0.188 | 0.985 | 0.844 | 1.150 | 8.51e-01 | 1.00e+00 | 32515 | 22457 |
| European | Recorded Anxiety | NDRI:bupropion | Migraine | Interaction (vs SSRI:sertraline) | 0.034 | 0.043 | 0.783 | 1.034 | 0.951 | 1.125 | 4.34e-01 | 1.00e+00 | 32515 | 22457 |
| European | Recorded Anxiety | SARI:trazodone | Migraine | Interaction (vs SSRI:sertraline) | -0.081 | 0.050 | -1.598 | 0.923 | 0.836 | 1.018 | 1.10e-01 | 7.70e-01 | 32515 | 22457 |
| European | Recorded Anxiety | SNRI:duloxetine | Migraine | Interaction (vs SSRI:sertraline) | -0.101 | 0.047 | -2.135 | 0.904 | 0.823 | 0.992 | 3.27e-02 | 2.29e-01 | 32515 | 22457 |
| European | Recorded Anxiety | SNRI:venlafaxine | Migraine | Interaction (vs SSRI:sertraline) | 0.006 | 0.048 | 0.132 | 1.006 | 0.916 | 1.106 | 8.95e-01 | 1.00e+00 | 32515 | 22457 |
| European | Recorded Anxiety | SSRI:citalopram | Migraine | Interaction (vs SSRI:sertraline) | -0.033 | 0.043 | -0.773 | 0.967 | 0.888 | 1.053 | 4.39e-01 | 1.00e+00 | 32515 | 22457 |
| European | Recorded Anxiety | SSRI:escitalopram | Migraine | Interaction (vs SSRI:sertraline) | -0.024 | 0.042 | -0.580 | 0.976 | 0.899 | 1.059 | 5.62e-01 | 1.00e+00 | 32515 | 22457 |
| European | Recorded Anxiety | SSRI:fluoxetine | Migraine | Interaction (vs SSRI:sertraline) | -0.002 | 0.044 | -0.056 | 0.998 | 0.916 | 1.087 | 9.55e-01 | 1.00e+00 | 32515 | 22457 |
| European | Recorded Anxiety | SSRI:paroxetine | Migraine | Interaction (vs SSRI:sertraline) | -0.046 | 0.061 | -0.753 | 0.955 | 0.848 | 1.076 | 4.52e-01 | 1.00e+00 | 32515 | 22457 |
| European | Recorded Anxiety | TCA:amitriptyline | Migraine | Interaction (vs SSRI:sertraline) | -0.051 | 0.060 | -0.848 | 0.950 | 0.844 | 1.069 | 3.97e-01 | 1.00e+00 | 32515 | 22457 |
| European | Recorded Anxiety | TCA:doxepin | Migraine | Interaction (vs SSRI:sertraline) | 0.140 | 0.132 | 1.057 | 1.150 | 0.887 | 1.491 | 2.91e-01 | 1.00e+00 | 32515 | 22457 |
| European | Recorded Anxiety | TCA:nortriptyline | Migraine | Interaction (vs SSRI:sertraline) | -0.099 | 0.074 | -1.329 | 0.906 | 0.783 | 1.048 | 1.84e-01 | 1.00e+00 | 32515 | 22457 |
| European | Recorded Anxiety | | Migraine | Joint_Interaction_Test | | | 13.570 | | | | 3.29e-01 | 1.00e+00 | 32515 | 22457 |
| European | Recorded Anxiety | SSRI:sertraline | Insomnia | Within-class slope | 0.102 | 0.029 | 3.492 | 1.108 | 1.046 | 1.173 | 4.80e-04 | 3.36e-03 | 32515 | 22457 |
| European | Recorded Anxiety | NaSSA:mirtazapine | Insomnia | Within-class slope | 0.021 | 0.078 | 0.272 | 1.022 | 0.876 | 1.191 | 7.85e-01 | 1.00e+00 | 32515 | 22457 |
| European | Recorded Anxiety | NDRI:bupropion | Insomnia | Within-class slope | 0.091 | 0.036 | 2.506 | 1.095 | 1.020 | 1.176 | 1.22e-02 | 8.56e-02 | 32515 | 22457 |
| European | Recorded Anxiety | SARI:trazodone | Insomnia | Within-class slope | 0.014 | 0.045 | 0.308 | 1.014 | 0.928 | 1.109 | 7.58e-01 | 1.00e+00 | 32515 | 22457 |
| European | Recorded Anxiety | SNRI:duloxetine | Insomnia | Within-class slope | 0.094 | 0.043 | 2.188 | 1.098 | 1.010 | 1.194 | 2.87e-02 | 2.01e-01 | 32515 | 22457 |
| European | Recorded Anxiety | SNRI:venlafaxine | Insomnia | Within-class slope | 0.094 | 0.044 | 2.132 | 1.098 | 1.008 | 1.197 | 3.30e-02 | 2.31e-01 | 32515 | 22457 |
| European | Recorded Anxiety | SSRI:citalopram | Insomnia | Within-class slope | 0.096 | 0.036 | 2.641 | 1.101 | 1.025 | 1.182 | 8.26e-03 | 5.78e-02 | 32515 | 22457 |
| European | Recorded Anxiety | SSRI:escitalopram | Insomnia | Within-class slope | 0.140 | 0.034 | 4.130 | 1.151 | 1.077 | 1.230 | 3.63e-05 | 2.54e-04 | 32515 | 22457 |
| European | Recorded Anxiety | SSRI:fluoxetine | Insomnia | Within-class slope | 0.059 | 0.039 | 1.534 | 1.061 | 0.984 | 1.144 | 1.25e-01 | 8.76e-01 | 32515 | 22457 |
| European | Recorded Anxiety | SSRI:paroxetine | Insomnia | Within-class slope | 0.170 | 0.061 | 2.786 | 1.186 | 1.052 | 1.336 | 5.34e-03 | 3.74e-02 | 32515 | 22457 |
| European | Recorded Anxiety | TCA:amitriptyline | Insomnia | Within-class slope | 0.095 | 0.055 | 1.744 | 1.100 | 0.988 | 1.224 | 8.12e-02 | 5.69e-01 | 32515 | 22457 |
| European | Recorded Anxiety | TCA:doxepin | Insomnia | Within-class slope | 0.255 | 0.145 | 1.763 | 1.291 | 0.972 | 1.715 | 7.79e-02 | 5.46e-01 | 32515 | 22457 |
| European | Recorded Anxiety | TCA:nortriptyline | Insomnia | Within-class slope | 0.022 | 0.075 | 0.291 | 1.022 | 0.883 | 1.183 | 7.71e-01 | 1.00e+00 | 32515 | 22457 |
| European | Recorded Anxiety | NaSSA:mirtazapine | Insomnia | Interaction (vs SSRI:sertraline) | -0.081 | 0.083 | -0.980 | 0.922 | 0.784 | 1.084 | 3.27e-01 | 1.00e+00 | 32515 | 22457 |
| European | Recorded Anxiety | NDRI:bupropion | Insomnia | Interaction (vs SSRI:sertraline) | -0.011 | 0.045 | -0.251 | 0.989 | 0.905 | 1.080 | 8.02e-01 | 1.00e+00 | 32515 | 22457 |
| European | Recorded Anxiety | SARI:trazodone | Insomnia | Interaction (vs SSRI:sertraline) | -0.088 | 0.053 | -1.673 | 0.916 | 0.826 | 1.015 | 9.43e-02 | 6.60e-01 | 32515 | 22457 |
| European | Recorded Anxiety | SNRI:duloxetine | Insomnia | Interaction (vs SSRI:sertraline) | -0.008 | 0.051 | -0.167 | 0.992 | 0.898 | 1.095 | 8.67e-01 | 1.00e+00 | 32515 | 22457 |
| European | Recorded Anxiety | SNRI:venlafaxine | Insomnia | Interaction (vs SSRI:sertraline) | -0.008 | 0.051 | -0.165 | 0.992 | 0.897 | 1.096 | 8.69e-01 | 1.00e+00 | 32515 | 22457 |
| European | Recorded Anxiety | SSRI:citalopram | Insomnia | Interaction (vs SSRI:sertraline) | -0.006 | 0.045 | -0.135 | 0.994 | 0.910 | 1.086 | 8.92e-01 | 1.00e+00 | 32515 | 22457 |
| European | Recorded Anxiety | SSRI:escitalopram | Insomnia | Interaction (vs SSRI:sertraline) | 0.038 | 0.043 | 0.880 | 1.039 | 0.954 | 1.131 | 3.79e-01 | 1.00e+00 | 32515 | 22457 |
| European | Recorded Anxiety | SSRI:fluoxetine | Insomnia | Interaction (vs SSRI:sertraline) | -0.043 | 0.047 | -0.910 | 0.958 | 0.873 | 1.051 | 3.63e-01 | 1.00e+00 | 32515 | 22457 |
| European | Recorded Anxiety | SSRI:paroxetine | Insomnia | Interaction (vs SSRI:sertraline) | 0.068 | 0.067 | 1.019 | 1.070 | 0.939 | 1.220 | 3.08e-01 | 1.00e+00 | 32515 | 22457 |
| European | Recorded Anxiety | TCA:amitriptyline | Insomnia | Interaction (vs SSRI:sertraline) | -0.007 | 0.061 | -0.118 | 0.993 | 0.882 | 1.118 | 9.06e-01 | 1.00e+00 | 32515 | 22457 |
| European | Recorded Anxiety | TCA:doxepin | Insomnia | Interaction (vs SSRI:sertraline) | 0.153 | 0.147 | 1.041 | 1.165 | 0.874 | 1.555 | 2.98e-01 | 1.00e+00 | 32515 | 22457 |
| European | Recorded Anxiety | TCA:nortriptyline | Insomnia | Interaction (vs SSRI:sertraline) | -0.080 | 0.079 | -1.015 | 0.923 | 0.790 | 1.078 | 3.10e-01 | 1.00e+00 | 32515 | 22457 |
| European | Recorded Anxiety | | Insomnia | Joint_Interaction_Test | | | 10.980 | | | | 5.31e-01 | 1.00e+00 | 32515 | 22457 |
| European | Recorded Anxiety | SSRI:sertraline | Depression | Within-class slope | 0.196 | 0.029 | 6.689 | 1.217 | 1.149 | 1.289 | 2.24e-11 | 1.57e-10 | 32515 | 22457 |
| European | Recorded Anxiety | NaSSA:mirtazapine | Depression | Within-class slope | 0.086 | 0.076 | 1.132 | 1.090 | 0.939 | 1.265 | 2.58e-01 | 1.00e+00 | 32515 | 22457 |
| European | Recorded Anxiety | NDRI:bupropion | Depression | Within-class slope | 0.183 | 0.036 | 5.144 | 1.200 | 1.120 | 1.287 | 2.69e-07 | 1.88e-06 | 32515 | 22457 |
| European | Recorded Anxiety | SARI:trazodone | Depression | Within-class slope | 0.177 | 0.045 | 3.934 | 1.193 | 1.093 | 1.303 | 8.36e-05 | 5.85e-04 | 32515 | 22457 |
| European | Recorded Anxiety | SNRI:duloxetine | Depression | Within-class slope | 0.142 | 0.042 | 3.353 | 1.153 | 1.061 | 1.253 | 7.99e-04 | 5.60e-03 | 32515 | 22457 |
| European | Recorded Anxiety | SNRI:venlafaxine | Depression | Within-class slope | 0.198 | 0.043 | 4.612 | 1.218 | 1.120 | 1.325 | 4.00e-06 | 2.80e-05 | 32515 | 22457 |
| European | Recorded Anxiety | SSRI:citalopram | Depression | Within-class slope | 0.148 | 0.037 | 3.971 | 1.159 | 1.078 | 1.247 | 7.14e-05 | 5.00e-04 | 32515 | 22457 |
| European | Recorded Anxiety | SSRI:escitalopram | Depression | Within-class slope | 0.189 | 0.034 | 5.575 | 1.208 | 1.131 | 1.291 | 2.47e-08 | 1.73e-07 | 32515 | 22457 |
| European | Recorded Anxiety | SSRI:fluoxetine | Depression | Within-class slope | 0.122 | 0.037 | 3.278 | 1.130 | 1.050 | 1.216 | 1.04e-03 | 7.31e-03 | 32515 | 22457 |
| European | Recorded Anxiety | SSRI:paroxetine | Depression | Within-class slope | 0.230 | 0.063 | 3.634 | 1.258 | 1.112 | 1.424 | 2.79e-04 | 1.96e-03 | 32515 | 22457 |
| European | Recorded Anxiety | TCA:amitriptyline | Depression | Within-class slope | 0.176 | 0.054 | 3.229 | 1.192 | 1.072 | 1.326 | 1.24e-03 | 8.71e-03 | 32515 | 22457 |

| | | | | | | | | | | | | | | |
|---|---|---|---|---|---|---|---|---|---|---|---|---|---|---|
| European | Recorded Anxiety | TCA:doxepin | Depression | Within-class slope | 0.199 | 0.130 | 1.534 | 1.221 | 0.946 | 1.575 | 1.25e-01 | 8.75e-01 | 32515 | 22457 |
| European | Recorded Anxiety | TCA:nortriptyline | Depression | Within-class slope | 0.041 | 0.072 | 0.570 | 1.042 | 0.905 | 1.199 | 5.69e-01 | 1.00e+00 | 32515 | 22457 |
| European | Recorded Anxiety | NaSSA:mirtazapine | Depression | Interaction (vs SSRI:sertraline) | -0.110 | 0.081 | -1.360 | 0.896 | 0.764 | 1.050 | 1.74e-01 | 1.00e+00 | 32515 | 22457 |
| European | Recorded Anxiety | NDRI:bupropion | Depression | Interaction (vs SSRI:sertraline) | -0.014 | 0.044 | -0.311 | 0.986 | 0.905 | 1.076 | 7.56e-01 | 1.00e+00 | 32515 | 22457 |
| European | Recorded Anxiety | SARI:trazodone | Depression | Interaction (vs SSRI:sertraline) | -0.020 | 0.053 | -0.378 | 0.980 | 0.884 | 1.087 | 7.06e-01 | 1.00e+00 | 32515 | 22457 |
| European | Recorded Anxiety | SNRI:duloxetine | Depression | Interaction (vs SSRI:sertraline) | -0.054 | 0.050 | -1.070 | 0.947 | 0.858 | 1.046 | 2.85e-01 | 1.00e+00 | 32515 | 22457 |
| European | Recorded Anxiety | SNRI:venlafaxine | Depression | Interaction (vs SSRI:sertraline) | 0.001 | 0.050 | 0.021 | 1.001 | 0.907 | 1.105 | 9.83e-01 | 1.00e+00 | 32515 | 22457 |
| European | Recorded Anxiety | SSRI:citalopram | Depression | Interaction (vs SSRI:sertraline) | -0.049 | 0.046 | -1.057 | 0.953 | 0.871 | 1.042 | 2.90e-01 | 1.00e+00 | 32515 | 22457 |
| European | Recorded Anxiety | SSRI:escitalopram | Depression | Interaction (vs SSRI:sertraline) | -0.007 | 0.044 | -0.168 | 0.993 | 0.912 | 1.081 | 8.67e-01 | 1.00e+00 | 32515 | 22457 |
| European | Recorded Anxiety | SSRI:fluoxetine | Depression | Interaction (vs SSRI:sertraline) | -0.074 | 0.046 | -1.612 | 0.929 | 0.848 | 1.016 | 1.07e-01 | 7.48e-01 | 32515 | 22457 |
| European | Recorded Anxiety | SSRI:paroxetine | Depression | Interaction (vs SSRI:sertraline) | 0.033 | 0.069 | 0.481 | 1.034 | 0.903 | 1.183 | 6.30e-01 | 1.00e+00 | 32515 | 22457 |
| European | Recorded Anxiety | TCA:amitriptyline | Depression | Interaction (vs SSRI:sertraline) | -0.021 | 0.061 | -0.341 | 0.980 | 0.870 | 1.103 | 7.33e-01 | 1.00e+00 | 32515 | 22457 |
| European | Recorded Anxiety | TCA:doxepin | Depression | Interaction (vs SSRI:sertraline) | 0.003 | 0.132 | 0.022 | 1.003 | 0.774 | 1.300 | 9.83e-01 | 1.00e+00 | 32515 | 22457 |
| European | Recorded Anxiety | TCA:nortriptyline | Depression | Interaction (vs SSRI:sertraline) | -0.156 | 0.077 | -2.022 | 0.856 | 0.736 | 0.995 | 4.32e-02 | 3.02e-01 | 32515 | 22457 |
| European | Recorded Anxiety | | Depression | Joint_Interaction_Test | | | 9.901 | | | | 6.25e-01 | 1.00e+00 | 32515 | 22457 |
| European | Recorded Anxiety | SSRI:sertraline | Schizophrenia | Within-class slope | 0.046 | 0.028 | 1.638 | 1.047 | 0.991 | 1.107 | 1.01e-01 | 7.10e-01 | 32515 | 22457 |
| European | Recorded Anxiety | NaSSA:mirtazapine | Schizophrenia | Within-class slope | 0.033 | 0.079 | 0.416 | 1.033 | 0.886 | 1.205 | 6.77e-01 | 1.00e+00 | 32515 | 22457 |
| European | Recorded Anxiety | NDRI:bupropion | Schizophrenia | Within-class slope | -0.002 | 0.035 | -0.050 | 0.998 | 0.931 | 1.070 | 9.60e-01 | 1.00e+00 | 32515 | 22457 |
| European | Recorded Anxiety | SARI:trazodone | Schizophrenia | Within-class slope | 0.004 | 0.044 | 0.080 | 1.004 | 0.921 | 1.093 | 9.36e-01 | 1.00e+00 | 32515 | 22457 |
| European | Recorded Anxiety | SNRI:duloxetine | Schizophrenia | Within-class slope | 0.029 | 0.042 | 0.704 | 1.030 | 0.949 | 1.118 | 4.81e-01 | 1.00e+00 | 32515 | 22457 |
| European | Recorded Anxiety | SNRI:venlafaxine | Schizophrenia | Within-class slope | 0.063 | 0.042 | 1.509 | 1.065 | 0.981 | 1.156 | 1.31e-01 | 9.18e-01 | 32515 | 22457 |
| European | Recorded Anxiety | SSRI:citalopram | Schizophrenia | Within-class slope | 0.000 | 0.035 | -0.007 | 1.000 | 0.934 | 1.070 | 9.94e-01 | 1.00e+00 | 32515 | 22457 |
| European | Recorded Anxiety | SSRI:escitalopram | Schizophrenia | Within-class slope | 0.049 | 0.033 | 1.494 | 1.050 | 0.985 | 1.119 | 1.35e-01 | 9.46e-01 | 32515 | 22457 |
| European | Recorded Anxiety | SSRI:fluoxetine | Schizophrenia | Within-class slope | -0.021 | 0.037 | -0.567 | 0.979 | 0.910 | 1.053 | 5.71e-01 | 1.00e+00 | 32515 | 22457 |
| European | Recorded Anxiety | SSRI:paroxetine | Schizophrenia | Within-class slope | -0.043 | 0.058 | -0.742 | 0.958 | 0.854 | 1.074 | 4.58e-01 | 1.00e+00 | 32515 | 22457 |
| European | Recorded Anxiety | TCA:amitriptyline | Schizophrenia | Within-class slope | -0.060 | 0.055 | -1.089 | 0.942 | 0.845 | 1.049 | 2.76e-01 | 1.00e+00 | 32515 | 22457 |
| European | Recorded Anxiety | TCA:doxepin | Schizophrenia | Within-class slope | -0.159 | 0.123 | -1.295 | 0.853 | 0.671 | 1.085 | 1.95e-01 | 1.00e+00 | 32515 | 22457 |
| European | Recorded Anxiety | TCA:nortriptyline | Schizophrenia | Within-class slope | -0.048 | 0.073 | -0.660 | 0.953 | 0.827 | 1.099 | 5.10e-01 | 1.00e+00 | 32515 | 22457 |
| European | Recorded Anxiety | NaSSA:mirtazapine | Schizophrenia | Interaction (vs SSRI:sertraline) | -0.014 | 0.083 | -0.163 | 0.987 | 0.838 | 1.161 | 8.71e-01 | 1.00e+00 | 32515 | 22457 |
| European | Recorded Anxiety | NDRI:bupropion | Schizophrenia | Interaction (vs SSRI:sertraline) | -0.048 | 0.043 | -1.107 | 0.953 | 0.876 | 1.038 | 2.68e-01 | 1.00e+00 | 32515 | 22457 |
| European | Recorded Anxiety | SARI:trazodone | Schizophrenia | Interaction (vs SSRI:sertraline) | -0.043 | 0.051 | -0.837 | 0.958 | 0.867 | 1.059 | 4.02e-01 | 1.00e+00 | 32515 | 22457 |
| European | Recorded Anxiety | SNRI:duloxetine | Schizophrenia | Interaction (vs SSRI:sertraline) | -0.017 | 0.049 | -0.340 | 0.983 | 0.892 | 1.084 | 7.34e-01 | 1.00e+00 | 32515 | 22457 |
| European | Recorded Anxiety | SNRI:venlafaxine | Schizophrenia | Interaction (vs SSRI:sertraline) | 0.017 | 0.049 | 0.346 | 1.017 | 0.924 | 1.120 | 7.29e-01 | 1.00e+00 | 32515 | 22457 |
| European | Recorded Anxiety | SSRI:citalopram | Schizophrenia | Interaction (vs SSRI:sertraline) | -0.046 | 0.043 | -1.071 | 0.955 | 0.877 | 1.039 | 2.84e-01 | 1.00e+00 | 32515 | 22457 |
| European | Recorded Anxiety | SSRI:escitalopram | Schizophrenia | Interaction (vs SSRI:sertraline) | 0.002 | 0.042 | 0.059 | 1.002 | 0.923 | 1.089 | 9.53e-01 | 1.00e+00 | 32515 | 22457 |
| European | Recorded Anxiety | SSRI:fluoxetine | Schizophrenia | Interaction (vs SSRI:sertraline) | -0.067 | 0.045 | -1.484 | 0.935 | 0.855 | 1.022 | 1.38e-01 | 9.65e-01 | 32515 | 22457 |
| European | Recorded Anxiety | SSRI:paroxetine | Schizophrenia | Interaction (vs SSRI:sertraline) | -0.089 | 0.064 | -1.404 | 0.914 | 0.807 | 1.036 | 1.60e-01 | 1.00e+00 | 32515 | 22457 |
| European | Recorded Anxiety | TCA:amitriptyline | Schizophrenia | Interaction (vs SSRI:sertraline) | -0.106 | 0.061 | -1.744 | 0.899 | 0.798 | 1.013 | 8.11e-02 | 5.68e-01 | 32515 | 22457 |
| European | Recorded Anxiety | TCA:doxepin | Schizophrenia | Interaction (vs SSRI:sertraline) | -0.205 | 0.125 | -1.634 | 0.815 | 0.637 | 1.042 | 1.02e-01 | 7.15e-01 | 32515 | 22457 |
| European | Recorded Anxiety | TCA:nortriptyline | Schizophrenia | Interaction (vs SSRI:sertraline) | -0.094 | 0.077 | -1.221 | 0.910 | 0.782 | 1.059 | 2.22e-01 | 1.00e+00 | 32515 | 22457 |
| European | Recorded Anxiety | | Schizophrenia | Joint_Interaction_Test | | | 11.073 | | | | 5.23e-01 | 1.00e+00 | 32515 | 22457 |
| European | Recorded Anxiety | SSRI:sertraline | Bipolar Disorder | Within-class slope | 0.047 | 0.029 | 1.607 | 1.049 | 0.990 | 1.111 | 1.08e-01 | 7.56e-01 | 32515 | 22457 |
| European | Recorded Anxiety | NaSSA:mirtazapine | Bipolar Disorder | Within-class slope | -0.056 | 0.079 | -0.712 | 0.945 | 0.810 | 1.103 | 4.76e-01 | 1.00e+00 | 32515 | 22457 |
| European | Recorded Anxiety | NDRI:bupropion | Bipolar Disorder | Within-class slope | -0.017 | 0.037 | -0.462 | 0.983 | 0.915 | 1.056 | 6.44e-01 | 1.00e+00 | 32515 | 22457 |
| European | Recorded Anxiety | SARI:trazodone | Bipolar Disorder | Within-class slope | 0.085 | 0.046 | 1.861 | 1.089 | 0.995 | 1.192 | 6.27e-02 | 4.39e-01 | 32515 | 22457 |
| European | Recorded Anxiety | SNRI:duloxetine | Bipolar Disorder | Within-class slope | 0.036 | 0.043 | 0.835 | 1.036 | 0.953 | 1.127 | 4.04e-01 | 1.00e+00 | 32515 | 22457 |
| European | Recorded Anxiety | SNRI:venlafaxine | Bipolar Disorder | Within-class slope | 0.057 | 0.042 | 1.346 | 1.059 | 0.974 | 1.151 | 1.78e-01 | 1.00e+00 | 32515 | 22457 |
| European | Recorded Anxiety | SSRI:citalopram | Bipolar Disorder | Within-class slope | 0.044 | 0.036 | 1.213 | 1.044 | 0.974 | 1.121 | 2.25e-01 | 1.00e+00 | 32515 | 22457 |
| European | Recorded Anxiety | SSRI:escitalopram | Bipolar Disorder | Within-class slope | 0.079 | 0.033 | 2.367 | 1.082 | 1.014 | 1.155 | 1.79e-02 | 1.25e-01 | 32515 | 22457 |
| European | Recorded Anxiety | SSRI:fluoxetine | Bipolar Disorder | Within-class slope | -0.008 | 0.038 | -0.206 | 0.992 | 0.921 | 1.068 | 8.37e-01 | 1.00e+00 | 32515 | 22457 |
| European | Recorded Anxiety | SSRI:paroxetine | Bipolar Disorder | Within-class slope | 0.026 | 0.059 | 0.436 | 1.026 | 0.914 | 1.153 | 6.63e-01 | 1.00e+00 | 32515 | 22457 |
| European | Recorded Anxiety | TCA:amitriptyline | Bipolar Disorder | Within-class slope | 0.052 | 0.057 | 0.908 | 1.053 | 0.942 | 1.177 | 3.64e-01 | 1.00e+00 | 32515 | 22457 |
| European | Recorded Anxiety | TCA:doxepin | Bipolar Disorder | Within-class slope | 0.032 | 0.121 | 0.263 | 1.032 | 0.814 | 1.309 | 7.93e-01 | 1.00e+00 | 32515 | 22457 |
| European | Recorded Anxiety | TCA:nortriptyline | Bipolar Disorder | Within-class slope | 0.102 | 0.073 | 1.400 | 1.107 | 0.960 | 1.278 | 1.62e-01 | 1.00e+00 | 32515 | 22457 |
| European | Recorded Anxiety | NaSSA:mirtazapine | Bipolar Disorder | Interaction (vs SSRI:sertraline) | -0.103 | 0.083 | -1.246 | 0.902 | 0.766 | 1.061 | 2.13e-01 | 1.00e+00 | 32515 | 22457 |
| European | Recorded Anxiety | NDRI:bupropion | Bipolar Disorder | Interaction (vs SSRI:sertraline) | -0.064 | 0.045 | -1.426 | 0.938 | 0.858 | 1.024 | 1.54e-01 | 1.00e+00 | 32515 | 22457 |
| European | Recorded Anxiety | SARI:trazodone | Bipolar Disorder | Interaction (vs SSRI:sertraline) | 0.038 | 0.054 | 0.709 | 1.039 | 0.935 | 1.154 | 4.79e-01 | 1.00e+00 | 32515 | 22457 |
| European | Recorded Anxiety | SNRI:duloxetine | Bipolar Disorder | Interaction (vs SSRI:sertraline) | -0.012 | 0.051 | -0.230 | 0.988 | 0.895 | 1.092 | 8.18e-01 | 1.00e+00 | 32515 | 22457 |
| European | Recorded Anxiety | SNRI:venlafaxine | Bipolar Disorder | Interaction (vs SSRI:sertraline) | 0.010 | 0.050 | 0.195 | 1.010 | 0.915 | 1.114 | 8.46e-01 | 1.00e+00 | 32515 | 22457 |

| | | | | | | | | | | | | | | |
|---|---|---|---|---|---|---|---|---|---|---|---|---|---|---|
| European | Recorded Anxiety | SSRI:citalopram | Bipolar Disorder | Interaction (vs SSRI:sertraline) | -0.004 | 0.045 | -0.086 | 0.996 | 0.912 | 1.088 | 9.32e-01 | 1.00e+00 | 32515 | 22457 |
| European | Recorded Anxiety | SSRI:escitalopram | Bipolar Disorder | Interaction (vs SSRI:sertraline) | 0.032 | 0.043 | 0.730 | 1.032 | 0.948 | 1.124 | 4.66e-01 | 1.00e+00 | 32515 | 22457 |
| European | Recorded Anxiety | SSRI:fluoxetine | Bipolar Disorder | Interaction (vs SSRI:sertraline) | -0.055 | 0.046 | -1.191 | 0.946 | 0.864 | 1.036 | 2.34e-01 | 1.00e+00 | 32515 | 22457 |
| European | Recorded Anxiety | SSRI:paroxetine | Bipolar Disorder | Interaction (vs SSRI:sertraline) | -0.021 | 0.065 | -0.329 | 0.979 | 0.861 | 1.112 | 7.42e-01 | 1.00e+00 | 32515 | 22457 |
| European | Recorded Anxiety | TCA:amitriptyline | Bipolar Disorder | Interaction (vs SSRI:sertraline) | 0.004 | 0.063 | 0.068 | 1.004 | 0.888 | 1.136 | 9.46e-01 | 1.00e+00 | 32515 | 22457 |
| European | Recorded Anxiety | TCA:doxepin | Bipolar Disorder | Interaction (vs SSRI:sertraline) | -0.016 | 0.124 | -0.126 | 0.985 | 0.772 | 1.255 | 9.00e-01 | 1.00e+00 | 32515 | 22457 |
| European | Recorded Anxiety | TCA:nortriptyline | Bipolar Disorder | Interaction (vs SSRI:sertraline) | 0.055 | 0.078 | 0.706 | 1.056 | 0.907 | 1.230 | 4.80e-01 | 1.00e+00 | 32515 | 22457 |
| European | Recorded Anxiety | | Bipolar Disorder | Joint_Interaction_Test | | | 9.441 | | | | 6.65e-01 | 1.00e+00 | 32515 | 22457 |
| European | Recorded Anxiety | SSRI:sertraline | BMI | Within-class slope | 0.021 | 0.027 | 0.772 | 1.021 | 0.968 | 1.078 | 4.40e-01 | 1.00e+00 | 32515 | 22457 |
| European | Recorded Anxiety | NaSSA:mirtazapine | BMI | Within-class slope | 0.018 | 0.075 | 0.243 | 1.018 | 0.879 | 1.179 | 8.08e-01 | 1.00e+00 | 32515 | 22457 |
| European | Recorded Anxiety | NDRI:bupropion | BMI | Within-class slope | -0.018 | 0.035 | -0.519 | 0.982 | 0.916 | 1.052 | 6.03e-01 | 1.00e+00 | 32515 | 22457 |
| European | Recorded Anxiety | SARI:trazodone | BMI | Within-class slope | 0.031 | 0.043 | 0.719 | 1.031 | 0.949 | 1.121 | 4.72e-01 | 1.00e+00 | 32515 | 22457 |
| European | Recorded Anxiety | SNRI:duloxetine | BMI | Within-class slope | -0.007 | 0.041 | -0.163 | 0.993 | 0.917 | 1.076 | 8.70e-01 | 1.00e+00 | 32515 | 22457 |
| European | Recorded Anxiety | SNRI:venlafaxine | BMI | Within-class slope | 0.006 | 0.041 | 0.137 | 1.006 | 0.928 | 1.090 | 8.91e-01 | 1.00e+00 | 32515 | 22457 |
| European | Recorded Anxiety | SSRI:citalopram | BMI | Within-class slope | 0.003 | 0.033 | 0.085 | 1.003 | 0.939 | 1.071 | 9.33e-01 | 1.00e+00 | 32515 | 22457 |
| European | Recorded Anxiety | SSRI:escitalopram | BMI | Within-class slope | 0.056 | 0.033 | 1.715 | 1.057 | 0.992 | 1.127 | 8.64e-02 | 6.05e-01 | 32515 | 22457 |
| European | Recorded Anxiety | SSRI:fluoxetine | BMI | Within-class slope | 0.075 | 0.036 | 2.094 | 1.078 | 1.005 | 1.157 | 3.63e-02 | 2.54e-01 | 32515 | 22457 |
| European | Recorded Anxiety | SSRI:paroxetine | BMI | Within-class slope | 0.106 | 0.055 | 1.934 | 1.112 | 0.999 | 1.238 | 5.31e-02 | 3.72e-01 | 32515 | 22457 |
| European | Recorded Anxiety | TCA:amitriptyline | BMI | Within-class slope | 0.062 | 0.055 | 1.129 | 1.064 | 0.955 | 1.185 | 2.59e-01 | 1.00e+00 | 32515 | 22457 |
| European | Recorded Anxiety | TCA:doxepin | BMI | Within-class slope | -0.034 | 0.125 | -0.274 | 0.966 | 0.756 | 1.235 | 7.84e-01 | 1.00e+00 | 32515 | 22457 |
| European | Recorded Anxiety | TCA:nortriptyline | BMI | Within-class slope | 0.016 | 0.070 | 0.225 | 1.016 | 0.885 | 1.166 | 8.22e-01 | 1.00e+00 | 32515 | 22457 |
| European | Recorded Anxiety | NaSSA:mirtazapine | BMI | Interaction (vs SSRI:sertraline) | -0.003 | 0.079 | -0.038 | 0.997 | 0.854 | 1.164 | 9.70e-01 | 1.00e+00 | 32515 | 22457 |
| European | Recorded Anxiety | NDRI:bupropion | BMI | Interaction (vs SSRI:sertraline) | -0.039 | 0.043 | -0.924 | 0.961 | 0.884 | 1.045 | 3.55e-01 | 1.00e+00 | 32515 | 22457 |
| European | Recorded Anxiety | SARI:trazodone | BMI | Interaction (vs SSRI:sertraline) | 0.009 | 0.050 | 0.189 | 1.009 | 0.916 | 1.112 | 8.50e-01 | 1.00e+00 | 32515 | 22457 |
| European | Recorded Anxiety | SNRI:duloxetine | BMI | Interaction (vs SSRI:sertraline) | -0.028 | 0.048 | -0.584 | 0.973 | 0.886 | 1.068 | 5.59e-01 | 1.00e+00 | 32515 | 22457 |
| European | Recorded Anxiety | SNRI:venlafaxine | BMI | Interaction (vs SSRI:sertraline) | -0.016 | 0.048 | -0.325 | 0.985 | 0.896 | 1.082 | 7.45e-01 | 1.00e+00 | 32515 | 22457 |
| European | Recorded Anxiety | SSRI:citalopram | BMI | Interaction (vs SSRI:sertraline) | -0.018 | 0.042 | -0.441 | 0.982 | 0.905 | 1.065 | 6.59e-01 | 1.00e+00 | 32515 | 22457 |
| European | Recorded Anxiety | SSRI:escitalopram | BMI | Interaction (vs SSRI:sertraline) | 0.035 | 0.041 | 0.835 | 1.035 | 0.955 | 1.123 | 4.04e-01 | 1.00e+00 | 32515 | 22457 |
| European | Recorded Anxiety | SSRI:fluoxetine | BMI | Interaction (vs SSRI:sertraline) | 0.054 | 0.043 | 1.249 | 1.056 | 0.970 | 1.150 | 2.12e-01 | 1.00e+00 | 32515 | 22457 |
| European | Recorded Anxiety | SSRI:paroxetine | BMI | Interaction (vs SSRI:sertraline) | 0.085 | 0.060 | 1.417 | 1.088 | 0.968 | 1.223 | 1.57e-01 | 1.00e+00 | 32515 | 22457 |
| European | Recorded Anxiety | TCA:amitriptyline | BMI | Interaction (vs SSRI:sertraline) | 0.041 | 0.060 | 0.678 | 1.042 | 0.926 | 1.173 | 4.98e-01 | 1.00e+00 | 32515 | 22457 |
| European | Recorded Anxiety | TCA:doxepin | BMI | Interaction (vs SSRI:sertraline) | -0.056 | 0.128 | -0.435 | 0.946 | 0.737 | 1.215 | 6.63e-01 | 1.00e+00 | 32515 | 22457 |
| European | Recorded Anxiety | TCA:nortriptyline | BMI | Interaction (vs SSRI:sertraline) | -0.005 | 0.075 | -0.073 | 0.995 | 0.859 | 1.151 | 9.42e-01 | 1.00e+00 | 32515 | 22457 |
| European | Recorded Anxiety | | BMI | Joint_Interaction_Test | | | 9.139 | | | | 6.91e-01 | 1.00e+00 | 32515 | 22457 |
| European | CIDI-MDD | SSRI:sertraline | ADHD | Within-class slope | -0.027 | 0.068 | -0.401 | 0.973 | 0.851 | 1.112 | 6.88e-01 | 1.00e+00 | 6270 | 4422 |
| European | CIDI-MDD | NaSSA:mirtazapine | ADHD | Within-class slope | -0.355 | 0.207 | -1.709 | 0.701 | 0.467 | 1.053 | 8.74e-02 | 6.12e-01 | 6270 | 4422 |
| European | CIDI-MDD | NDRI:bupropion | ADHD | Within-class slope | 0.070 | 0.074 | 0.950 | 1.073 | 0.928 | 1.241 | 3.42e-01 | 1.00e+00 | 6270 | 4422 |
| European | CIDI-MDD | SARI:trazodone | ADHD | Within-class slope | 0.176 | 0.105 | 1.670 | 1.192 | 0.970 | 1.466 | 9.50e-02 | 6.65e-01 | 6270 | 4422 |
| European | CIDI-MDD | SNRI:duloxetine | ADHD | Within-class slope | 0.051 | 0.087 | 0.592 | 1.053 | 0.888 | 1.247 | 5.54e-01 | 1.00e+00 | 6270 | 4422 |
| European | CIDI-MDD | SNRI:venlafaxine | ADHD | Within-class slope | -0.077 | 0.085 | -0.905 | 0.926 | 0.784 | 1.094 | 3.65e-01 | 1.00e+00 | 6270 | 4422 |
| European | CIDI-MDD | SSRI:citalopram | ADHD | Within-class slope | 0.026 | 0.092 | 0.279 | 1.026 | 0.857 | 1.228 | 7.80e-01 | 1.00e+00 | 6270 | 4422 |
| European | CIDI-MDD | SSRI:escitalopram | ADHD | Within-class slope | 0.110 | 0.080 | 1.374 | 1.116 | 0.954 | 1.305 | 1.69e-01 | 1.00e+00 | 6270 | 4422 |
| European | CIDI-MDD | SSRI:fluoxetine | ADHD | Within-class slope | 0.030 | 0.083 | 0.358 | 1.030 | 0.876 | 1.211 | 7.21e-01 | 1.00e+00 | 6270 | 4422 |
| European | CIDI-MDD | SSRI:paroxetine | ADHD | Within-class slope | 0.170 | 0.161 | 1.052 | 1.185 | 0.864 | 1.626 | 2.93e-01 | 1.00e+00 | 6270 | 4422 |
| European | CIDI-MDD | TCA:amitriptyline | ADHD | Within-class slope | -0.042 | 0.134 | -0.312 | 0.959 | 0.738 | 1.246 | 7.55e-01 | 1.00e+00 | 6270 | 4422 |
| European | CIDI-MDD | TCA:doxepin | ADHD | Within-class slope | -0.026 | 0.270 | -0.095 | 0.975 | 0.574 | 1.654 | 9.24e-01 | 1.00e+00 | 6270 | 4422 |
| European | CIDI-MDD | TCA:nortriptyline | ADHD | Within-class slope | -0.221 | 0.144 | -1.536 | 0.801 | 0.604 | 1.063 | 1.25e-01 | 8.72e-01 | 6270 | 4422 |
| European | CIDI-MDD | NaSSA:mirtazapine | ADHD | Interaction (vs SSRI:sertraline) | -0.327 | 0.218 | -1.504 | 0.721 | 0.471 | 1.104 | 1.33e-01 | 9.29e-01 | 6270 | 4422 |
| European | CIDI-MDD | NDRI:bupropion | ADHD | Interaction (vs SSRI:sertraline) | 0.098 | 0.098 | 1.001 | 1.103 | 0.910 | 1.336 | 3.17e-01 | 1.00e+00 | 6270 | 4422 |
| European | CIDI-MDD | SARI:trazodone | ADHD | Interaction (vs SSRI:sertraline) | 0.203 | 0.122 | 1.662 | 1.225 | 0.964 | 1.557 | 9.65e-02 | 6.75e-01 | 6270 | 4422 |
| European | CIDI-MDD | SNRI:duloxetine | ADHD | Interaction (vs SSRI:sertraline) | 0.079 | 0.106 | 0.741 | 1.082 | 0.879 | 1.332 | 4.58e-01 | 1.00e+00 | 6270 | 4422 |
| European | CIDI-MDD | SNRI:venlafaxine | ADHD | Interaction (vs SSRI:sertraline) | -0.049 | 0.105 | -0.471 | 0.952 | 0.775 | 1.169 | 6.37e-01 | 1.00e+00 | 6270 | 4422 |
| European | CIDI-MDD | SSRI:citalopram | ADHD | Interaction (vs SSRI:sertraline) | 0.053 | 0.112 | 0.474 | 1.054 | 0.847 | 1.313 | 6.35e-01 | 1.00e+00 | 6270 | 4422 |
| European | CIDI-MDD | SSRI:escitalopram | ADHD | Interaction (vs SSRI:sertraline) | 0.137 | 0.102 | 1.350 | 1.147 | 0.940 | 1.400 | 1.77e-01 | 1.00e+00 | 6270 | 4422 |
| European | CIDI-MDD | SSRI:fluoxetine | ADHD | Interaction (vs SSRI:sertraline) | 0.057 | 0.104 | 0.546 | 1.059 | 0.863 | 1.299 | 5.85e-01 | 1.00e+00 | 6270 | 4422 |
| European | CIDI-MDD | SSRI:paroxetine | ADHD | Interaction (vs SSRI:sertraline) | 0.197 | 0.174 | 1.131 | 1.218 | 0.866 | 1.714 | 2.58e-01 | 1.00e+00 | 6270 | 4422 |
| European | CIDI-MDD | TCA:amitriptyline | ADHD | Interaction (vs SSRI:sertraline) | -0.014 | 0.146 | -0.098 | 0.986 | 0.741 | 1.312 | 9.22e-01 | 1.00e+00 | 6270 | 4422 |
| European | CIDI-MDD | TCA:doxepin | ADHD | Interaction (vs SSRI:sertraline) | 0.002 | 0.277 | 0.006 | 1.002 | 0.581 | 1.726 | 9.95e-01 | 1.00e+00 | 6270 | 4422 |
| European | CIDI-MDD | TCA:nortriptyline | ADHD | Interaction (vs SSRI:sertraline) | -0.194 | 0.153 | -1.265 | 0.824 | 0.610 | 1.112 | 2.06e-01 | 1.00e+00 | 6270 | 4422 |

| | | | | | | | | | | | | | | |
|---|---|---|---|---|---|---|---|---|---|---|---|---|---|---|
| European | CIDI-MDD | | ADHD | Joint_Interaction_Test | | | 13.063 | | | | 3.64e-01 | 1.00e+00 | 6270 | 4422 |
| European | CIDI-MDD | SSRI:sertraline | Migraine | Within-class slope | 0.075 | 0.065 | 1.152 | 1.078 | 0.949 | 1.224 | 2.50e-01 | 1.00e+00 | 6270 | 4422 |
| European | CIDI-MDD | NaSSA:mirtazapine | Migraine | Within-class slope | -0.094 | 0.208 | -0.453 | 0.910 | 0.606 | 1.368 | 6.51e-01 | 1.00e+00 | 6270 | 4422 |
| European | CIDI-MDD | NDRI:bupropion | Migraine | Within-class slope | 0.051 | 0.074 | 0.695 | 1.053 | 0.911 | 1.217 | 4.87e-01 | 1.00e+00 | 6270 | 4422 |
| European | CIDI-MDD | SARI:trazodone | Migraine | Within-class slope | -0.024 | 0.101 | -0.240 | 0.976 | 0.801 | 1.189 | 8.10e-01 | 1.00e+00 | 6270 | 4422 |
| European | CIDI-MDD | SNRI:duloxetine | Migraine | Within-class slope | -0.150 | 0.083 | -1.810 | 0.861 | 0.732 | 1.012 | 7.02e-02 | 4.92e-01 | 6270 | 4422 |
| European | CIDI-MDD | SNRI:venlafaxine | Migraine | Within-class slope | 0.021 | 0.082 | 0.250 | 1.021 | 0.869 | 1.199 | 8.02e-01 | 1.00e+00 | 6270 | 4422 |
| European | CIDI-MDD | SSRI:citalopram | Migraine | Within-class slope | -0.012 | 0.088 | -0.141 | 0.988 | 0.831 | 1.174 | 8.88e-01 | 1.00e+00 | 6270 | 4422 |
| European | CIDI-MDD | SSRI:escitalopram | Migraine | Within-class slope | -0.038 | 0.081 | -0.466 | 0.963 | 0.821 | 1.129 | 6.41e-01 | 1.00e+00 | 6270 | 4422 |
| European | CIDI-MDD | SSRI:fluoxetine | Migraine | Within-class slope | 0.005 | 0.079 | 0.068 | 1.005 | 0.860 | 1.175 | 9.46e-01 | 1.00e+00 | 6270 | 4422 |
| European | CIDI-MDD | SSRI:paroxetine | Migraine | Within-class slope | 0.131 | 0.124 | 1.053 | 1.139 | 0.894 | 1.453 | 2.92e-01 | 1.00e+00 | 6270 | 4422 |
| European | CIDI-MDD | TCA:amitriptyline | Migraine | Within-class slope | 0.274 | 0.125 | 2.194 | 1.315 | 1.030 | 1.680 | 2.82e-02 | 1.98e-01 | 6270 | 4422 |
| European | CIDI-MDD | TCA:doxepin | Migraine | Within-class slope | -0.075 | 0.215 | -0.347 | 0.928 | 0.609 | 1.415 | 7.29e-01 | 1.00e+00 | 6270 | 4422 |
| European | CIDI-MDD | TCA:nortriptyline | Migraine | Within-class slope | 0.046 | 0.163 | 0.282 | 1.047 | 0.761 | 1.441 | 7.78e-01 | 1.00e+00 | 6270 | 4422 |
| European | CIDI-MDD | NaSSA:mirtazapine | Migraine | Interaction (vs SSRI:sertraline) | -0.169 | 0.218 | -0.775 | 0.844 | 0.551 | 1.295 | 4.39e-01 | 1.00e+00 | 6270 | 4422 |
| European | CIDI-MDD | NDRI:bupropion | Migraine | Interaction (vs SSRI:sertraline) | -0.024 | 0.093 | -0.254 | 0.977 | 0.814 | 1.172 | 8.00e-01 | 1.00e+00 | 6270 | 4422 |
| European | CIDI-MDD | SARI:trazodone | Migraine | Interaction (vs SSRI:sertraline) | -0.099 | 0.117 | -0.849 | 0.906 | 0.720 | 1.139 | 3.96e-01 | 1.00e+00 | 6270 | 4422 |
| European | CIDI-MDD | SNRI:duloxetine | Migraine | Interaction (vs SSRI:sertraline) | -0.225 | 0.102 | -2.204 | 0.799 | 0.654 | 0.975 | 2.75e-02 | 1.93e-01 | 6270 | 4422 |
| European | CIDI-MDD | SNRI:venlafaxine | Migraine | Interaction (vs SSRI:sertraline) | -0.054 | 0.103 | -0.530 | 0.947 | 0.775 | 1.158 | 5.96e-01 | 1.00e+00 | 6270 | 4422 |
| European | CIDI-MDD | SSRI:citalopram | Migraine | Interaction (vs SSRI:sertraline) | -0.087 | 0.108 | -0.809 | 0.916 | 0.742 | 1.132 | 4.19e-01 | 1.00e+00 | 6270 | 4422 |
| European | CIDI-MDD | SSRI:escitalopram | Migraine | Interaction (vs SSRI:sertraline) | -0.113 | 0.103 | -1.098 | 0.893 | 0.731 | 1.093 | 2.72e-01 | 1.00e+00 | 6270 | 4422 |
| European | CIDI-MDD | SSRI:fluoxetine | Migraine | Interaction (vs SSRI:sertraline) | -0.070 | 0.099 | -0.702 | 0.933 | 0.768 | 1.133 | 4.83e-01 | 1.00e+00 | 6270 | 4422 |
| European | CIDI-MDD | SSRI:paroxetine | Migraine | Interaction (vs SSRI:sertraline) | 0.056 | 0.136 | 0.408 | 1.057 | 0.809 | 1.381 | 6.83e-01 | 1.00e+00 | 6270 | 4422 |
| European | CIDI-MDD | TCA:amitriptyline | Migraine | Interaction (vs SSRI:sertraline) | 0.199 | 0.139 | 1.429 | 1.220 | 0.929 | 1.603 | 1.53e-01 | 1.00e+00 | 6270 | 4422 |
| European | CIDI-MDD | TCA:doxepin | Migraine | Interaction (vs SSRI:sertraline) | -0.150 | 0.224 | -0.668 | 0.861 | 0.555 | 1.336 | 5.04e-01 | 1.00e+00 | 6270 | 4422 |
| European | CIDI-MDD | TCA:nortriptyline | Migraine | Interaction (vs SSRI:sertraline) | -0.029 | 0.173 | -0.167 | 0.972 | 0.692 | 1.364 | 8.68e-01 | 1.00e+00 | 6270 | 4422 |
| European | CIDI-MDD | | Migraine | Joint_Interaction_Test | | | 11.603 | | | | 4.78e-01 | 1.00e+00 | 6270 | 4422 |
| European | CIDI-MDD | SSRI:sertraline | Insomnia | Within-class slope | 0.020 | 0.073 | 0.271 | 1.020 | 0.884 | 1.176 | 7.87e-01 | 1.00e+00 | 6270 | 4422 |
| European | CIDI-MDD | NaSSA:mirtazapine | Insomnia | Within-class slope | 0.076 | 0.229 | 0.333 | 1.079 | 0.689 | 1.692 | 7.39e-01 | 1.00e+00 | 6270 | 4422 |
| European | CIDI-MDD | NDRI:bupropion | Insomnia | Within-class slope | -0.052 | 0.075 | -0.694 | 0.949 | 0.820 | 1.100 | 4.88e-01 | 1.00e+00 | 6270 | 4422 |
| European | CIDI-MDD | SARI:trazodone | Insomnia | Within-class slope | -0.098 | 0.103 | -0.953 | 0.906 | 0.740 | 1.109 | 3.40e-01 | 1.00e+00 | 6270 | 4422 |
| European | CIDI-MDD | SNRI:duloxetine | Insomnia | Within-class slope | -0.042 | 0.087 | -0.481 | 0.959 | 0.808 | 1.138 | 6.30e-01 | 1.00e+00 | 6270 | 4422 |
| European | CIDI-MDD | SNRI:venlafaxine | Insomnia | Within-class slope | -0.003 | 0.088 | -0.033 | 0.997 | 0.839 | 1.185 | 9.74e-01 | 1.00e+00 | 6270 | 4422 |
| European | CIDI-MDD | SSRI:citalopram | Insomnia | Within-class slope | -0.086 | 0.096 | -0.901 | 0.917 | 0.761 | 1.107 | 3.68e-01 | 1.00e+00 | 6270 | 4422 |
| European | CIDI-MDD | SSRI:escitalopram | Insomnia | Within-class slope | 0.140 | 0.082 | 1.695 | 1.150 | 0.978 | 1.352 | 9.01e-02 | 6.30e-01 | 6270 | 4422 |
| European | CIDI-MDD | SSRI:fluoxetine | Insomnia | Within-class slope | -0.040 | 0.085 | -0.469 | 0.961 | 0.813 | 1.136 | 6.39e-01 | 1.00e+00 | 6270 | 4422 |
| European | CIDI-MDD | SSRI:paroxetine | Insomnia | Within-class slope | -0.068 | 0.151 | -0.452 | 0.934 | 0.695 | 1.255 | 6.51e-01 | 1.00e+00 | 6270 | 4422 |
| European | CIDI-MDD | TCA:amitriptyline | Insomnia | Within-class slope | 0.118 | 0.131 | 0.900 | 1.125 | 0.870 | 1.455 | 3.68e-01 | 1.00e+00 | 6270 | 4422 |
| European | CIDI-MDD | TCA:doxepin | Insomnia | Within-class slope | 0.171 | 0.319 | 0.534 | 1.186 | 0.634 | 2.218 | 5.93e-01 | 1.00e+00 | 6270 | 4422 |
| European | CIDI-MDD | TCA:nortriptyline | Insomnia | Within-class slope | -0.195 | 0.168 | -1.164 | 0.822 | 0.592 | 1.143 | 2.44e-01 | 1.00e+00 | 6270 | 4422 |
| European | CIDI-MDD | NaSSA:mirtazapine | Insomnia | Interaction (vs SSRI:sertraline) | 0.057 | 0.237 | 0.239 | 1.058 | 0.665 | 1.684 | 8.11e-01 | 1.00e+00 | 6270 | 4422 |
| European | CIDI-MDD | NDRI:bupropion | Insomnia | Interaction (vs SSRI:sertraline) | -0.072 | 0.103 | -0.698 | 0.931 | 0.761 | 1.139 | 4.85e-01 | 1.00e+00 | 6270 | 4422 |
| European | CIDI-MDD | SARI:trazodone | Insomnia | Interaction (vs SSRI:sertraline) | -0.118 | 0.124 | -0.955 | 0.889 | 0.697 | 1.132 | 3.39e-01 | 1.00e+00 | 6270 | 4422 |
| European | CIDI-MDD | SNRI:duloxetine | Insomnia | Interaction (vs SSRI:sertraline) | -0.062 | 0.111 | -0.558 | 0.940 | 0.757 | 1.168 | 5.77e-01 | 1.00e+00 | 6270 | 4422 |
| European | CIDI-MDD | SNRI:venlafaxine | Insomnia | Interaction (vs SSRI:sertraline) | -0.023 | 0.112 | -0.202 | 0.978 | 0.785 | 1.218 | 8.40e-01 | 1.00e+00 | 6270 | 4422 |
| European | CIDI-MDD | SSRI:citalopram | Insomnia | Interaction (vs SSRI:sertraline) | -0.106 | 0.117 | -0.906 | 0.900 | 0.715 | 1.131 | 3.65e-01 | 1.00e+00 | 6270 | 4422 |
| European | CIDI-MDD | SSRI:escitalopram | Insomnia | Interaction (vs SSRI:sertraline) | 0.120 | 0.106 | 1.135 | 1.128 | 0.916 | 1.387 | 2.56e-01 | 1.00e+00 | 6270 | 4422 |
| European | CIDI-MDD | SSRI:fluoxetine | Insomnia | Interaction (vs SSRI:sertraline) | -0.060 | 0.110 | -0.542 | 0.942 | 0.759 | 1.169 | 5.88e-01 | 1.00e+00 | 6270 | 4422 |
| European | CIDI-MDD | SSRI:paroxetine | Insomnia | Interaction (vs SSRI:sertraline) | -0.088 | 0.166 | -0.528 | 0.916 | 0.661 | 1.269 | 5.97e-01 | 1.00e+00 | 6270 | 4422 |
| European | CIDI-MDD | TCA:amitriptyline | Insomnia | Interaction (vs SSRI:sertraline) | 0.098 | 0.144 | 0.685 | 1.103 | 0.833 | 1.462 | 4.93e-01 | 1.00e+00 | 6270 | 4422 |
| European | CIDI-MDD | TCA:doxepin | Insomnia | Interaction (vs SSRI:sertraline) | 0.151 | 0.328 | 0.461 | 1.163 | 0.612 | 2.210 | 6.45e-01 | 1.00e+00 | 6270 | 4422 |
| European | CIDI-MDD | TCA:nortriptyline | Insomnia | Interaction (vs SSRI:sertraline) | -0.215 | 0.180 | -1.194 | 0.806 | 0.566 | 1.148 | 2.33e-01 | 1.00e+00 | 6270 | 4422 |
| European | CIDI-MDD | | Insomnia | Joint_Interaction_Test | | | 8.254 | | | | 7.65e-01 | 1.00e+00 | 6270 | 4422 |
| European | CIDI-MDD | SSRI:sertraline | Depression | Within-class slope | 0.023 | 0.071 | 0.326 | 1.023 | 0.890 | 1.177 | 7.45e-01 | 1.00e+00 | 6270 | 4422 |
| European | CIDI-MDD | NaSSA:mirtazapine | Depression | Within-class slope | 0.000 | 0.233 | 0.002 | 1.000 | 0.634 | 1.578 | 9.99e-01 | 1.00e+00 | 6270 | 4422 |
| European | CIDI-MDD | NDRI:bupropion | Depression | Within-class slope | 0.121 | 0.074 | 1.647 | 1.129 | 0.977 | 1.305 | 9.95e-02 | 6.97e-01 | 6270 | 4422 |
| European | CIDI-MDD | SARI:trazodone | Depression | Within-class slope | 0.013 | 0.105 | 0.120 | 1.013 | 0.824 | 1.244 | 9.05e-01 | 1.00e+00 | 6270 | 4422 |
| European | CIDI-MDD | SNRI:duloxetine | Depression | Within-class slope | 0.088 | 0.088 | 1.002 | 1.092 | 0.919 | 1.299 | 3.16e-01 | 1.00e+00 | 6270 | 4422 |
| European | CIDI-MDD | SNRI:venlafaxine | Depression | Within-class slope | 0.183 | 0.093 | 1.966 | 1.201 | 1.001 | 1.440 | 4.92e-02 | 3.45e-01 | 6270 | 4422 |

| | | | | | | | | | | | | | | |
|---|---|---|---|---|---|---|---|---|---|---|---|---|---|---|
| European | CIDI-MDD | SSRI:citalopram | Depression | Within-class slope | 0.020 | 0.092 | 0.222 | 1.021 | 0.853 | 1.221 | 8.24e-01 | 1.00e+00 | 6270 | 4422 |
| European | CIDI-MDD | SSRI:escitalopram | Depression | Within-class slope | 0.256 | 0.085 | 3.012 | 1.292 | 1.094 | 1.526 | 2.59e-03 | 1.82e-02 | 6270 | 4422 |
| European | CIDI-MDD | SSRI:fluoxetine | Depression | Within-class slope | 0.055 | 0.082 | 0.681 | 1.057 | 0.901 | 1.240 | 4.96e-01 | 1.00e+00 | 6270 | 4422 |
| European | CIDI-MDD | SSRI:paroxetine | Depression | Within-class slope | 0.219 | 0.155 | 1.412 | 1.245 | 0.919 | 1.688 | 1.58e-01 | 1.00e+00 | 6270 | 4422 |
| European | CIDI-MDD | TCA:amitriptyline | Depression | Within-class slope | 0.083 | 0.122 | 0.678 | 1.087 | 0.855 | 1.381 | 4.98e-01 | 1.00e+00 | 6270 | 4422 |
| European | CIDI-MDD | TCA:doxepin | Depression | Within-class slope | 0.148 | 0.329 | 0.449 | 1.160 | 0.608 | 2.212 | 6.53e-01 | 1.00e+00 | 6270 | 4422 |
| European | CIDI-MDD | TCA:nortriptyline | Depression | Within-class slope | -0.185 | 0.163 | -1.135 | 0.832 | 0.605 | 1.144 | 2.56e-01 | 1.00e+00 | 6270 | 4422 |
| European | CIDI-MDD | NaSSA:mirtazapine | Depression | Interaction (vs SSRI:sertraline) | -0.023 | 0.246 | -0.093 | 0.977 | 0.603 | 1.583 | 9.26e-01 | 1.00e+00 | 6270 | 4422 |
| European | CIDI-MDD | NDRI:bupropion | Depression | Interaction (vs SSRI:sertraline) | 0.098 | 0.099 | 0.994 | 1.103 | 0.909 | 1.339 | 3.20e-01 | 1.00e+00 | 6270 | 4422 |
| European | CIDI-MDD | SARI:trazodone | Depression | Interaction (vs SSRI:sertraline) | -0.011 | 0.124 | -0.086 | 0.989 | 0.776 | 1.261 | 9.32e-01 | 1.00e+00 | 6270 | 4422 |
| European | CIDI-MDD | SNRI:duloxetine | Depression | Interaction (vs SSRI:sertraline) | 0.065 | 0.110 | 0.592 | 1.067 | 0.860 | 1.325 | 5.54e-01 | 1.00e+00 | 6270 | 4422 |
| European | CIDI-MDD | SNRI:venlafaxine | Depression | Interaction (vs SSRI:sertraline) | 0.160 | 0.113 | 1.417 | 1.173 | 0.941 | 1.463 | 1.57e-01 | 1.00e+00 | 6270 | 4422 |
| European | CIDI-MDD | SSRI:citalopram | Depression | Interaction (vs SSRI:sertraline) | -0.003 | 0.115 | -0.025 | 0.997 | 0.796 | 1.249 | 9.80e-01 | 1.00e+00 | 6270 | 4422 |
| European | CIDI-MDD | SSRI:escitalopram | Depression | Interaction (vs SSRI:sertraline) | 0.233 | 0.108 | 2.161 | 1.262 | 1.022 | 1.559 | 3.07e-02 | 2.15e-01 | 6270 | 4422 |
| European | CIDI-MDD | SSRI:fluoxetine | Depression | Interaction (vs SSRI:sertraline) | 0.032 | 0.105 | 0.308 | 1.033 | 0.841 | 1.268 | 7.58e-01 | 1.00e+00 | 6270 | 4422 |
| European | CIDI-MDD | SSRI:paroxetine | Depression | Interaction (vs SSRI:sertraline) | 0.196 | 0.168 | 1.168 | 1.217 | 0.876 | 1.691 | 2.43e-01 | 1.00e+00 | 6270 | 4422 |
| European | CIDI-MDD | TCA:amitriptyline | Depression | Interaction (vs SSRI:sertraline) | 0.060 | 0.141 | 0.424 | 1.062 | 0.805 | 1.400 | 6.72e-01 | 1.00e+00 | 6270 | 4422 |
| European | CIDI-MDD | TCA:doxepin | Depression | Interaction (vs SSRI:sertraline) | 0.125 | 0.336 | 0.371 | 1.133 | 0.586 | 2.190 | 7.11e-01 | 1.00e+00 | 6270 | 4422 |
| European | CIDI-MDD | TCA:nortriptyline | Depression | Interaction (vs SSRI:sertraline) | -0.208 | 0.176 | -1.178 | 0.812 | 0.575 | 1.148 | 2.39e-01 | 1.00e+00 | 6270 | 4422 |
| European | CIDI-MDD | | Depression | Joint_Interaction_Test | | | 11.235 | | | | 5.09e-01 | 1.00e+00 | 6270 | 4422 |
| European | CIDI-MDD | SSRI:sertraline | Schizophrenia | Within-class slope | 0.031 | 0.069 | 0.451 | 1.031 | 0.901 | 1.180 | 6.52e-01 | 1.00e+00 | 6270 | 4422 |
| European | CIDI-MDD | NaSSA:mirtazapine | Schizophrenia | Within-class slope | 0.034 | 0.214 | 0.159 | 1.035 | 0.680 | 1.573 | 8.73e-01 | 1.00e+00 | 6270 | 4422 |
| European | CIDI-MDD | NDRI:bupropion | Schizophrenia | Within-class slope | 0.033 | 0.073 | 0.454 | 1.034 | 0.896 | 1.193 | 6.50e-01 | 1.00e+00 | 6270 | 4422 |
| European | CIDI-MDD | SARI:trazodone | Schizophrenia | Within-class slope | 0.119 | 0.102 | 1.166 | 1.126 | 0.922 | 1.374 | 2.44e-01 | 1.00e+00 | 6270 | 4422 |
| European | CIDI-MDD | SNRI:duloxetine | Schizophrenia | Within-class slope | 0.180 | 0.090 | 2.009 | 1.198 | 1.004 | 1.429 | 4.46e-02 | 3.12e-01 | 6270 | 4422 |
| European | CIDI-MDD | SNRI:venlafaxine | Schizophrenia | Within-class slope | 0.033 | 0.090 | 0.368 | 1.034 | 0.866 | 1.234 | 7.13e-01 | 1.00e+00 | 6270 | 4422 |
| European | CIDI-MDD | SSRI:citalopram | Schizophrenia | Within-class slope | 0.067 | 0.090 | 0.752 | 1.070 | 0.897 | 1.275 | 4.52e-01 | 1.00e+00 | 6270 | 4422 |
| European | CIDI-MDD | SSRI:escitalopram | Schizophrenia | Within-class slope | 0.053 | 0.081 | 0.646 | 1.054 | 0.899 | 1.236 | 5.18e-01 | 1.00e+00 | 6270 | 4422 |
| European | CIDI-MDD | SSRI:fluoxetine | Schizophrenia | Within-class slope | 0.087 | 0.084 | 1.034 | 1.091 | 0.925 | 1.286 | 3.01e-01 | 1.00e+00 | 6270 | 4422 |
| European | CIDI-MDD | SSRI:paroxetine | Schizophrenia | Within-class slope | 0.085 | 0.152 | 0.561 | 1.089 | 0.808 | 1.468 | 5.75e-01 | 1.00e+00 | 6270 | 4422 |
| European | CIDI-MDD | TCA:amitriptyline | Schizophrenia | Within-class slope | 0.163 | 0.145 | 1.128 | 1.177 | 0.886 | 1.564 | 2.59e-01 | 1.00e+00 | 6270 | 4422 |
| European | CIDI-MDD | TCA:doxepin | Schizophrenia | Within-class slope | -0.418 | 0.358 | -1.167 | 0.659 | 0.326 | 1.328 | 2.43e-01 | 1.00e+00 | 6270 | 4422 |
| European | CIDI-MDD | TCA:nortriptyline | Schizophrenia | Within-class slope | 0.165 | 0.162 | 1.021 | 1.179 | 0.859 | 1.619 | 3.07e-01 | 1.00e+00 | 6270 | 4422 |
| European | CIDI-MDD | NaSSA:mirtazapine | Schizophrenia | Interaction (vs SSRI:sertraline) | 0.003 | 0.224 | 0.014 | 1.003 | 0.647 | 1.555 | 9.89e-01 | 1.00e+00 | 6270 | 4422 |
| European | CIDI-MDD | NDRI:bupropion | Schizophrenia | Interaction (vs SSRI:sertraline) | 0.002 | 0.095 | 0.023 | 1.002 | 0.832 | 1.207 | 9.82e-01 | 1.00e+00 | 6270 | 4422 |
| European | CIDI-MDD | SARI:trazodone | Schizophrenia | Interaction (vs SSRI:sertraline) | 0.088 | 0.120 | 0.731 | 1.091 | 0.863 | 1.380 | 4.65e-01 | 1.00e+00 | 6270 | 4422 |
| European | CIDI-MDD | SNRI:duloxetine | Schizophrenia | Interaction (vs SSRI:sertraline) | 0.150 | 0.111 | 1.352 | 1.161 | 0.935 | 1.442 | 1.76e-01 | 1.00e+00 | 6270 | 4422 |
| European | CIDI-MDD | SNRI:venlafaxine | Schizophrenia | Interaction (vs SSRI:sertraline) | 0.002 | 0.111 | 0.020 | 1.002 | 0.807 | 1.245 | 9.84e-01 | 1.00e+00 | 6270 | 4422 |
| European | CIDI-MDD | SSRI:citalopram | Schizophrenia | Interaction (vs SSRI:sertraline) | 0.036 | 0.110 | 0.331 | 1.037 | 0.836 | 1.287 | 7.40e-01 | 1.00e+00 | 6270 | 4422 |
| European | CIDI-MDD | SSRI:escitalopram | Schizophrenia | Interaction (vs SSRI:sertraline) | 0.022 | 0.104 | 0.207 | 1.022 | 0.833 | 1.254 | 8.36e-01 | 1.00e+00 | 6270 | 4422 |
| European | CIDI-MDD | SSRI:fluoxetine | Schizophrenia | Interaction (vs SSRI:sertraline) | 0.056 | 0.105 | 0.534 | 1.058 | 0.861 | 1.299 | 5.94e-01 | 1.00e+00 | 6270 | 4422 |
| European | CIDI-MDD | SSRI:paroxetine | Schizophrenia | Interaction (vs SSRI:sertraline) | 0.054 | 0.166 | 0.329 | 1.056 | 0.763 | 1.461 | 7.42e-01 | 1.00e+00 | 6270 | 4422 |
| European | CIDI-MDD | TCA:amitriptyline | Schizophrenia | Interaction (vs SSRI:sertraline) | 0.132 | 0.156 | 0.850 | 1.141 | 0.841 | 1.549 | 3.95e-01 | 1.00e+00 | 6270 | 4422 |
| European | CIDI-MDD | TCA:doxepin | Schizophrenia | Interaction (vs SSRI:sertraline) | -0.449 | 0.364 | -1.232 | 0.638 | 0.313 | 1.303 | 2.18e-01 | 1.00e+00 | 6270 | 4422 |
| European | CIDI-MDD | TCA:nortriptyline | Schizophrenia | Interaction (vs SSRI:sertraline) | 0.134 | 0.170 | 0.787 | 1.144 | 0.819 | 1.597 | 4.31e-01 | 1.00e+00 | 6270 | 4422 |
| European | CIDI-MDD | | Schizophrenia | Joint_Interaction_Test | | | 5.648 | | | | 9.33e-01 | 1.00e+00 | 6270 | 4422 |
| European | CIDI-MDD | SSRI:sertraline | Bipolar Disorder | Within-class slope | -0.032 | 0.071 | -0.448 | 0.969 | 0.843 | 1.113 | 6.54e-01 | 1.00e+00 | 6270 | 4422 |
| European | CIDI-MDD | NaSSA:mirtazapine | Bipolar Disorder | Within-class slope | 0.090 | 0.200 | 0.450 | 1.094 | 0.739 | 1.621 | 6.52e-01 | 1.00e+00 | 6270 | 4422 |
| European | CIDI-MDD | NDRI:bupropion | Bipolar Disorder | Within-class slope | 0.015 | 0.074 | 0.199 | 1.015 | 0.878 | 1.173 | 8.42e-01 | 1.00e+00 | 6270 | 4422 |
| European | CIDI-MDD | SARI:trazodone | Bipolar Disorder | Within-class slope | 0.083 | 0.110 | 0.750 | 1.086 | 0.875 | 1.348 | 4.53e-01 | 1.00e+00 | 6270 | 4422 |
| European | CIDI-MDD | SNRI:duloxetine | Bipolar Disorder | Within-class slope | 0.075 | 0.087 | 0.855 | 1.078 | 0.908 | 1.279 | 3.92e-01 | 1.00e+00 | 6270 | 4422 |
| European | CIDI-MDD | SNRI:venlafaxine | Bipolar Disorder | Within-class slope | 0.062 | 0.093 | 0.664 | 1.064 | 0.886 | 1.277 | 5.06e-01 | 1.00e+00 | 6270 | 4422 |
| European | CIDI-MDD | SSRI:citalopram | Bipolar Disorder | Within-class slope | 0.079 | 0.093 | 0.849 | 1.083 | 0.901 | 1.300 | 3.96e-01 | 1.00e+00 | 6270 | 4422 |
| European | CIDI-MDD | SSRI:escitalopram | Bipolar Disorder | Within-class slope | -0.008 | 0.079 | -0.105 | 0.992 | 0.849 | 1.158 | 9.16e-01 | 1.00e+00 | 6270 | 4422 |
| European | CIDI-MDD | SSRI:fluoxetine | Bipolar Disorder | Within-class slope | 0.120 | 0.085 | 1.411 | 1.127 | 0.955 | 1.330 | 1.58e-01 | 1.00e+00 | 6270 | 4422 |
| European | CIDI-MDD | SSRI:paroxetine | Bipolar Disorder | Within-class slope | 0.133 | 0.146 | 0.910 | 1.142 | 0.858 | 1.519 | 3.63e-01 | 1.00e+00 | 6270 | 4422 |
| European | CIDI-MDD | TCA:amitriptyline | Bipolar Disorder | Within-class slope | -0.101 | 0.149 | -0.679 | 0.904 | 0.675 | 1.210 | 4.97e-01 | 1.00e+00 | 6270 | 4422 |
| European | CIDI-MDD | TCA:doxepin | Bipolar Disorder | Within-class slope | -0.468 | 0.321 | -1.458 | 0.626 | 0.334 | 1.175 | 1.45e-01 | 1.00e+00 | 6270 | 4422 |
| European | CIDI-MDD | TCA:nortriptyline | Bipolar Disorder | Within-class slope | 0.136 | 0.183 | 0.742 | 1.146 | 0.800 | 1.640 | 4.58e-01 | 1.00e+00 | 6270 | 4422 |

| | | | | | | | | | | | | | | |
|---|---|---|---|---|---|---|---|---|---|---|---|---|---|---|
| European | CIDI-MDD | NaSSA:mirtazapine | Bipolar Disorder | Interaction (vs SSRI:sertraline) | 0.122 | 0.211 | 0.577 | 1.130 | 0.747 | 1.709 | 5.64e-01 | 1.00e+00 | 6270 | 4422 |
| European | CIDI-MDD | NDRI:bupropion | Bipolar Disorder | Interaction (vs SSRI:sertraline) | 0.046 | 0.098 | 0.472 | 1.048 | 0.864 | 1.270 | 6.37e-01 | 1.00e+00 | 6270 | 4422 |
| European | CIDI-MDD | SARI:trazodone | Bipolar Disorder | Interaction (vs SSRI:sertraline) | 0.114 | 0.129 | 0.887 | 1.121 | 0.871 | 1.443 | 3.75e-01 | 1.00e+00 | 6270 | 4422 |
| European | CIDI-MDD | SNRI:duloxetine | Bipolar Disorder | Interaction (vs SSRI:sertraline) | 0.106 | 0.109 | 0.976 | 1.112 | 0.898 | 1.378 | 3.29e-01 | 1.00e+00 | 6270 | 4422 |
| European | CIDI-MDD | SNRI:venlafaxine | Bipolar Disorder | Interaction (vs SSRI:sertraline) | 0.094 | 0.113 | 0.827 | 1.098 | 0.880 | 1.371 | 4.08e-01 | 1.00e+00 | 6270 | 4422 |
| European | CIDI-MDD | SSRI:citalopram | Bipolar Disorder | Interaction (vs SSRI:sertraline) | 0.111 | 0.116 | 0.961 | 1.117 | 0.891 | 1.402 | 3.37e-01 | 1.00e+00 | 6270 | 4422 |
| European | CIDI-MDD | SSRI:escitalopram | Bipolar Disorder | Interaction (vs SSRI:sertraline) | 0.023 | 0.103 | 0.226 | 1.024 | 0.836 | 1.254 | 8.21e-01 | 1.00e+00 | 6270 | 4422 |
| European | CIDI-MDD | SSRI:fluoxetine | Bipolar Disorder | Interaction (vs SSRI:sertraline) | 0.151 | 0.107 | 1.416 | 1.163 | 0.944 | 1.434 | 1.57e-01 | 1.00e+00 | 6270 | 4422 |
| European | CIDI-MDD | SSRI:paroxetine | Bipolar Disorder | Interaction (vs SSRI:sertraline) | 0.164 | 0.158 | 1.042 | 1.179 | 0.865 | 1.605 | 2.97e-01 | 1.00e+00 | 6270 | 4422 |
| European | CIDI-MDD | TCA:amitriptyline | Bipolar Disorder | Interaction (vs SSRI:sertraline) | -0.069 | 0.161 | -0.432 | 0.933 | 0.681 | 1.278 | 6.66e-01 | 1.00e+00 | 6270 | 4422 |
| European | CIDI-MDD | TCA:doxepin | Bipolar Disorder | Interaction (vs SSRI:sertraline) | -0.436 | 0.328 | -1.331 | 0.646 | 0.340 | 1.229 | 1.83e-01 | 1.00e+00 | 6270 | 4422 |
| European | CIDI-MDD | TCA:nortriptyline | Bipolar Disorder | Interaction (vs SSRI:sertraline) | 0.168 | 0.193 | 0.867 | 1.183 | 0.809 | 1.727 | 3.86e-01 | 1.00e+00 | 6270 | 4422 |
| European | CIDI-MDD | | Bipolar Disorder | Joint_Interaction_Test | | | 7.254 | | | | 8.40e-01 | 1.00e+00 | 6270 | 4422 |
| European | CIDI-MDD | SSRI:sertraline | BMI | Within-class slope | 0.100 | 0.066 | 1.507 | 1.105 | 0.970 | 1.259 | 1.32e-01 | 9.23e-01 | 6270 | 4422 |
| European | CIDI-MDD | NaSSA:mirtazapine | BMI | Within-class slope | 0.059 | 0.234 | 0.252 | 1.061 | 0.671 | 1.677 | 8.01e-01 | 1.00e+00 | 6270 | 4422 |
| European | CIDI-MDD | NDRI:bupropion | BMI | Within-class slope | -0.103 | 0.072 | -1.437 | 0.902 | 0.784 | 1.038 | 1.51e-01 | 1.00e+00 | 6270 | 4422 |
| European | CIDI-MDD | SARI:trazodone | BMI | Within-class slope | 0.076 | 0.104 | 0.730 | 1.079 | 0.880 | 1.323 | 4.65e-01 | 1.00e+00 | 6270 | 4422 |
| European | CIDI-MDD | SNRI:duloxetine | BMI | Within-class slope | 0.002 | 0.084 | 0.019 | 1.002 | 0.849 | 1.181 | 9.85e-01 | 1.00e+00 | 6270 | 4422 |
| European | CIDI-MDD | SNRI:venlafaxine | BMI | Within-class slope | 0.036 | 0.094 | 0.378 | 1.036 | 0.862 | 1.246 | 7.05e-01 | 1.00e+00 | 6270 | 4422 |
| European | CIDI-MDD | SSRI:citalopram | BMI | Within-class slope | 0.090 | 0.091 | 0.992 | 1.094 | 0.916 | 1.307 | 3.21e-01 | 1.00e+00 | 6270 | 4422 |
| European | CIDI-MDD | SSRI:escitalopram | BMI | Within-class slope | 0.052 | 0.084 | 0.621 | 1.053 | 0.894 | 1.242 | 5.34e-01 | 1.00e+00 | 6270 | 4422 |
| European | CIDI-MDD | SSRI:fluoxetine | BMI | Within-class slope | 0.037 | 0.082 | 0.458 | 1.038 | 0.885 | 1.218 | 6.47e-01 | 1.00e+00 | 6270 | 4422 |
| European | CIDI-MDD | SSRI:paroxetine | BMI | Within-class slope | 0.172 | 0.139 | 1.240 | 1.188 | 0.905 | 1.559 | 2.15e-01 | 1.00e+00 | 6270 | 4422 |
| European | CIDI-MDD | TCA:amitriptyline | BMI | Within-class slope | 0.188 | 0.139 | 1.352 | 1.207 | 0.919 | 1.586 | 1.76e-01 | 1.00e+00 | 6270 | 4422 |
| European | CIDI-MDD | TCA:doxepin | BMI | Within-class slope | 0.265 | 0.308 | 0.860 | 1.303 | 0.713 | 2.382 | 3.90e-01 | 1.00e+00 | 6270 | 4422 |
| European | CIDI-MDD | TCA:nortriptyline | BMI | Within-class slope | 0.073 | 0.161 | 0.451 | 1.075 | 0.784 | 1.474 | 6.52e-01 | 1.00e+00 | 6270 | 4422 |
| European | CIDI-MDD | NaSSA:mirtazapine | BMI | Interaction (vs SSRI:sertraline) | -0.041 | 0.241 | -0.171 | 0.960 | 0.598 | 1.540 | 8.64e-01 | 1.00e+00 | 6270 | 4422 |
| European | CIDI-MDD | NDRI:bupropion | BMI | Interaction (vs SSRI:sertraline) | -0.203 | 0.093 | -2.187 | 0.816 | 0.681 | 0.979 | 2.87e-02 | 2.01e-01 | 6270 | 4422 |
| European | CIDI-MDD | SARI:trazodone | BMI | Interaction (vs SSRI:sertraline) | -0.024 | 0.121 | -0.200 | 0.976 | 0.770 | 1.237 | 8.42e-01 | 1.00e+00 | 6270 | 4422 |
| European | CIDI-MDD | SNRI:duloxetine | BMI | Interaction (vs SSRI:sertraline) | -0.099 | 0.104 | -0.947 | 0.906 | 0.739 | 1.111 | 3.44e-01 | 1.00e+00 | 6270 | 4422 |
| European | CIDI-MDD | SNRI:venlafaxine | BMI | Interaction (vs SSRI:sertraline) | -0.065 | 0.111 | -0.582 | 0.937 | 0.754 | 1.165 | 5.60e-01 | 1.00e+00 | 6270 | 4422 |
| European | CIDI-MDD | SSRI:citalopram | BMI | Interaction (vs SSRI:sertraline) | -0.010 | 0.110 | -0.093 | 0.990 | 0.798 | 1.228 | 9.26e-01 | 1.00e+00 | 6270 | 4422 |
| European | CIDI-MDD | SSRI:escitalopram | BMI | Interaction (vs SSRI:sertraline) | -0.048 | 0.105 | -0.456 | 0.953 | 0.775 | 1.172 | 6.48e-01 | 1.00e+00 | 6270 | 4422 |
| European | CIDI-MDD | SSRI:fluoxetine | BMI | Interaction (vs SSRI:sertraline) | -0.063 | 0.101 | -0.619 | 0.939 | 0.770 | 1.146 | 5.36e-01 | 1.00e+00 | 6270 | 4422 |
| European | CIDI-MDD | SSRI:paroxetine | BMI | Interaction (vs SSRI:sertraline) | 0.072 | 0.150 | 0.481 | 1.075 | 0.801 | 1.442 | 6.31e-01 | 1.00e+00 | 6270 | 4422 |
| European | CIDI-MDD | TCA:amitriptyline | BMI | Interaction (vs SSRI:sertraline) | 0.088 | 0.153 | 0.578 | 1.092 | 0.810 | 1.473 | 5.63e-01 | 1.00e+00 | 6270 | 4422 |
| European | CIDI-MDD | TCA:doxepin | BMI | Interaction (vs SSRI:sertraline) | 0.164 | 0.314 | 0.523 | 1.179 | 0.637 | 2.183 | 6.01e-01 | 1.00e+00 | 6270 | 4422 |
| European | CIDI-MDD | TCA:nortriptyline | BMI | Interaction (vs SSRI:sertraline) | -0.028 | 0.173 | -0.160 | 0.973 | 0.693 | 1.366 | 8.73e-01 | 1.00e+00 | 6270 | 4422 |
| European | CIDI-MDD | | BMI | Joint_Interaction_Test | | | 8.448 | | | | 7.49e-01 | 1.00e+00 | 6270 | 4422 |
| European | CIDI-noMDD | SSRI:sertraline | ADHD | Within-class slope | -0.019 | 0.106 | -0.175 | 0.982 | 0.798 | 1.208 | 8.61e-01 | 1.00e+00 | 3246 | 2599 |
| European | CIDI-noMDD | NaSSA:mirtazapine | ADHD | Within-class slope | 0.111 | 0.260 | 0.426 | 1.117 | 0.671 | 1.860 | 6.70e-01 | 1.00e+00 | 3246 | 2599 |
| European | CIDI-noMDD | NDRI:bupropion | ADHD | Within-class slope | 0.130 | 0.136 | 0.957 | 1.138 | 0.873 | 1.485 | 3.39e-01 | 1.00e+00 | 3246 | 2599 |
| European | CIDI-noMDD | SARI:trazodone | ADHD | Within-class slope | 0.071 | 0.123 | 0.582 | 1.074 | 0.844 | 1.366 | 5.61e-01 | 1.00e+00 | 3246 | 2599 |
| European | CIDI-noMDD | SNRI:duloxetine | ADHD | Within-class slope | 0.069 | 0.152 | 0.452 | 1.071 | 0.796 | 1.442 | 6.51e-01 | 1.00e+00 | 3246 | 2599 |
| European | CIDI-noMDD | SNRI:venlafaxine | ADHD | Within-class slope | -0.090 | 0.161 | -0.560 | 0.914 | 0.667 | 1.252 | 5.75e-01 | 1.00e+00 | 3246 | 2599 |
| European | CIDI-noMDD | SSRI:citalopram | ADHD | Within-class slope | -0.102 | 0.127 | -0.807 | 0.903 | 0.705 | 1.157 | 4.20e-01 | 1.00e+00 | 3246 | 2599 |
| European | CIDI-noMDD | SSRI:escitalopram | ADHD | Within-class slope | 0.140 | 0.140 | 1.001 | 1.151 | 0.874 | 1.515 | 3.17e-01 | 1.00e+00 | 3246 | 2599 |
| European | CIDI-noMDD | SSRI:fluoxetine | ADHD | Within-class slope | 0.023 | 0.158 | 0.144 | 1.023 | 0.751 | 1.394 | 8.86e-01 | 1.00e+00 | 3246 | 2599 |
| European | CIDI-noMDD | SSRI:paroxetine | ADHD | Within-class slope | 0.069 | 0.202 | 0.341 | 1.071 | 0.721 | 1.591 | 7.33e-01 | 1.00e+00 | 3246 | 2599 |
| European | CIDI-noMDD | TCA:amitriptyline | ADHD | Within-class slope | 0.240 | 0.132 | 1.814 | 1.272 | 0.981 | 1.648 | 6.97e-02 | 4.88e-01 | 3246 | 2599 |
| European | CIDI-noMDD | TCA:nortriptyline | ADHD | Within-class slope | 0.148 | 0.162 | 0.914 | 1.159 | 0.845 | 1.591 | 3.61e-01 | 1.00e+00 | 3246 | 2599 |
| European | CIDI-noMDD | NaSSA:mirtazapine | ADHD | Interaction (vs SSRI:sertraline) | 0.129 | 0.280 | 0.462 | 1.138 | 0.658 | 1.969 | 6.44e-01 | 1.00e+00 | 3246 | 2599 |
| European | CIDI-noMDD | NDRI:bupropion | ADHD | Interaction (vs SSRI:sertraline) | 0.148 | 0.166 | 0.894 | 1.160 | 0.838 | 1.605 | 3.71e-01 | 1.00e+00 | 3246 | 2599 |
| European | CIDI-noMDD | SARI:trazodone | ADHD | Interaction (vs SSRI:sertraline) | 0.090 | 0.157 | 0.571 | 1.094 | 0.804 | 1.489 | 5.68e-01 | 1.00e+00 | 3246 | 2599 |
| European | CIDI-noMDD | SNRI:duloxetine | ADHD | Interaction (vs SSRI:sertraline) | 0.087 | 0.178 | 0.490 | 1.091 | 0.770 | 1.546 | 6.24e-01 | 1.00e+00 | 3246 | 2599 |
| European | CIDI-noMDD | SNRI:venlafaxine | ADHD | Interaction (vs SSRI:sertraline) | -0.071 | 0.190 | -0.375 | 0.931 | 0.641 | 1.352 | 7.08e-01 | 1.00e+00 | 3246 | 2599 |
| European | CIDI-noMDD | SSRI:citalopram | ADHD | Interaction (vs SSRI:sertraline) | -0.084 | 0.158 | -0.530 | 0.920 | 0.675 | 1.253 | 5.96e-01 | 1.00e+00 | 3246 | 2599 |
| European | CIDI-noMDD | SSRI:escitalopram | ADHD | Interaction (vs SSRI:sertraline) | 0.159 | 0.171 | 0.929 | 1.172 | 0.838 | 1.639 | 3.53e-01 | 1.00e+00 | 3246 | 2599 |
| European | CIDI-noMDD | SSRI:fluoxetine | ADHD | Interaction (vs SSRI:sertraline) | 0.041 | 0.188 | 0.219 | 1.042 | 0.721 | 1.507 | 8.26e-01 | 1.00e+00 | 3246 | 2599 |

| | | | | | | | | | | | | | | |
|---|---|---|---|---|---|---|---|---|---|---|---|---|---|---|
| European | CIDI-noMDD | SSRI:paroxetine | ADHD | Interaction (vs SSRI:sertraline) | 0.087 | 0.224 | 0.391 | 1.091 | 0.704 | 1.692 | 6.96e-01 | 1.00e+00 | 3246 | 2599 |
| European | CIDI-noMDD | TCA:amitriptyline | ADHD | Interaction (vs SSRI:sertraline) | 0.259 | 0.167 | 1.553 | 1.295 | 0.934 | 1.796 | 1.20e-01 | 8.43e-01 | 3246 | 2599 |
| European | CIDI-noMDD | TCA:nortriptyline | ADHD | Interaction (vs SSRI:sertraline) | 0.166 | 0.193 | 0.859 | 1.181 | 0.808 | 1.725 | 3.90e-01 | 1.00e+00 | 3246 | 2599 |
| European | CIDI-noMDD | | ADHD | Joint_Interaction_Test | | | 6.148 | | | | 8.63e-01 | 1.00e+00 | 3246 | 2599 |
| European | CIDI-noMDD | SSRI:sertraline | Migraine | Within-class slope | -0.003 | 0.107 | -0.030 | 0.997 | 0.808 | 1.229 | 9.76e-01 | 1.00e+00 | 3246 | 2599 |
| European | CIDI-noMDD | NaSSA:mirtazapine | Migraine | Within-class slope | 0.245 | 0.205 | 1.195 | 1.277 | 0.855 | 1.909 | 2.32e-01 | 1.00e+00 | 3246 | 2599 |
| European | CIDI-noMDD | NDRI:bupropion | Migraine | Within-class slope | 0.047 | 0.136 | 0.348 | 1.049 | 0.803 | 1.369 | 7.28e-01 | 1.00e+00 | 3246 | 2599 |
| European | CIDI-noMDD | SARI:trazodone | Migraine | Within-class slope | 0.209 | 0.128 | 1.630 | 1.232 | 0.959 | 1.584 | 1.03e-01 | 7.21e-01 | 3246 | 2599 |
| European | CIDI-noMDD | SNRI:duloxetine | Migraine | Within-class slope | -0.026 | 0.130 | -0.201 | 0.974 | 0.755 | 1.257 | 8.40e-01 | 1.00e+00 | 3246 | 2599 |
| European | CIDI-noMDD | SNRI:venlafaxine | Migraine | Within-class slope | -0.115 | 0.138 | -0.830 | 0.891 | 0.680 | 1.169 | 4.06e-01 | 1.00e+00 | 3246 | 2599 |
| European | CIDI-noMDD | SSRI:citalopram | Migraine | Within-class slope | -0.198 | 0.119 | -1.665 | 0.820 | 0.650 | 1.036 | 9.58e-02 | 6.71e-01 | 3246 | 2599 |
| European | CIDI-noMDD | SSRI:escitalopram | Migraine | Within-class slope | 0.104 | 0.128 | 0.809 | 1.109 | 0.863 | 1.427 | 4.19e-01 | 1.00e+00 | 3246 | 2599 |
| European | CIDI-noMDD | SSRI:fluoxetine | Migraine | Within-class slope | 0.019 | 0.138 | 0.137 | 1.019 | 0.777 | 1.336 | 8.91e-01 | 1.00e+00 | 3246 | 2599 |
| European | CIDI-noMDD | SSRI:paroxetine | Migraine | Within-class slope | -0.049 | 0.183 | -0.269 | 0.952 | 0.665 | 1.363 | 7.88e-01 | 1.00e+00 | 3246 | 2599 |
| European | CIDI-noMDD | TCA:amitriptyline | Migraine | Within-class slope | -0.336 | 0.133 | -2.520 | 0.715 | 0.551 | 0.928 | 1.17e-02 | 8.22e-02 | 3246 | 2599 |
| European | CIDI-noMDD | TCA:nortriptyline | Migraine | Within-class slope | -0.255 | 0.164 | -1.557 | 0.775 | 0.562 | 1.068 | 1.20e-01 | 8.37e-01 | 3246 | 2599 |
| European | CIDI-noMDD | NaSSA:mirtazapine | Migraine | Interaction (vs SSRI:sertraline) | 0.248 | 0.231 | 1.075 | 1.282 | 0.815 | 2.015 | 2.83e-01 | 1.00e+00 | 3246 | 2599 |
| European | CIDI-noMDD | NDRI:bupropion | Migraine | Interaction (vs SSRI:sertraline) | 0.051 | 0.163 | 0.309 | 1.052 | 0.764 | 1.449 | 7.57e-01 | 1.00e+00 | 3246 | 2599 |
| European | CIDI-noMDD | SARI:trazodone | Migraine | Interaction (vs SSRI:sertraline) | 0.212 | 0.166 | 1.275 | 1.236 | 0.892 | 1.713 | 2.02e-01 | 1.00e+00 | 3246 | 2599 |
| European | CIDI-noMDD | SNRI:duloxetine | Migraine | Interaction (vs SSRI:sertraline) | -0.023 | 0.164 | -0.140 | 0.977 | 0.708 | 1.348 | 8.89e-01 | 1.00e+00 | 3246 | 2599 |
| European | CIDI-noMDD | SNRI:venlafaxine | Migraine | Interaction (vs SSRI:sertraline) | -0.112 | 0.173 | -0.646 | 0.894 | 0.637 | 1.255 | 5.18e-01 | 1.00e+00 | 3246 | 2599 |
| European | CIDI-noMDD | SSRI:citalopram | Migraine | Interaction (vs SSRI:sertraline) | -0.195 | 0.150 | -1.299 | 0.823 | 0.613 | 1.104 | 1.94e-01 | 1.00e+00 | 3246 | 2599 |
| European | CIDI-noMDD | SSRI:escitalopram | Migraine | Interaction (vs SSRI:sertraline) | 0.107 | 0.167 | 0.642 | 1.113 | 0.802 | 1.544 | 5.21e-01 | 1.00e+00 | 3246 | 2599 |
| European | CIDI-noMDD | SSRI:fluoxetine | Migraine | Interaction (vs SSRI:sertraline) | 0.022 | 0.169 | 0.131 | 1.022 | 0.735 | 1.423 | 8.95e-01 | 1.00e+00 | 3246 | 2599 |
| European | CIDI-noMDD | SSRI:paroxetine | Migraine | Interaction (vs SSRI:sertraline) | -0.046 | 0.208 | -0.222 | 0.955 | 0.635 | 1.435 | 8.24e-01 | 1.00e+00 | 3246 | 2599 |
| European | CIDI-noMDD | TCA:amitriptyline | Migraine | Interaction (vs SSRI:sertraline) | -0.332 | 0.169 | -1.969 | 0.717 | 0.515 | 0.998 | 4.89e-02 | 3.42e-01 | 3246 | 2599 |
| European | CIDI-noMDD | TCA:nortriptyline | Migraine | Interaction (vs SSRI:sertraline) | -0.252 | 0.193 | -1.303 | 0.778 | 0.533 | 1.135 | 1.92e-01 | 1.00e+00 | 3246 | 2599 |
| European | CIDI-noMDD | | Migraine | Joint_Interaction_Test | | | 17.525 | | | | 9.33e-02 | 6.53e-01 | 3246 | 2599 |
| European | CIDI-noMDD | SSRI:sertraline | Insomnia | Within-class slope | 0.004 | 0.115 | 0.037 | 1.004 | 0.801 | 1.258 | 9.71e-01 | 1.00e+00 | 3246 | 2599 |
| European | CIDI-noMDD | NaSSA:mirtazapine | Insomnia | Within-class slope | 0.117 | 0.226 | 0.520 | 1.124 | 0.723 | 1.750 | 6.03e-01 | 1.00e+00 | 3246 | 2599 |
| European | CIDI-noMDD | NDRI:bupropion | Insomnia | Within-class slope | 0.222 | 0.131 | 1.696 | 1.248 | 0.966 | 1.612 | 9.00e-02 | 6.30e-01 | 3246 | 2599 |
| European | CIDI-noMDD | SARI:trazodone | Insomnia | Within-class slope | 0.224 | 0.127 | 1.766 | 1.251 | 0.976 | 1.605 | 7.74e-02 | 5.42e-01 | 3246 | 2599 |
| European | CIDI-noMDD | SNRI:duloxetine | Insomnia | Within-class slope | 0.089 | 0.153 | 0.582 | 1.093 | 0.810 | 1.476 | 5.61e-01 | 1.00e+00 | 3246 | 2599 |
| European | CIDI-noMDD | SNRI:venlafaxine | Insomnia | Within-class slope | -0.004 | 0.154 | -0.024 | 0.996 | 0.737 | 1.347 | 9.81e-01 | 1.00e+00 | 3246 | 2599 |
| European | CIDI-noMDD | SSRI:citalopram | Insomnia | Within-class slope | 0.143 | 0.131 | 1.088 | 1.153 | 0.892 | 1.491 | 2.77e-01 | 1.00e+00 | 3246 | 2599 |
| European | CIDI-noMDD | SSRI:escitalopram | Insomnia | Within-class slope | 0.118 | 0.141 | 0.838 | 1.125 | 0.854 | 1.482 | 4.02e-01 | 1.00e+00 | 3246 | 2599 |
| European | CIDI-noMDD | SSRI:fluoxetine | Insomnia | Within-class slope | -0.061 | 0.142 | -0.429 | 0.941 | 0.712 | 1.243 | 6.68e-01 | 1.00e+00 | 3246 | 2599 |
| European | CIDI-noMDD | SSRI:paroxetine | Insomnia | Within-class slope | 0.363 | 0.209 | 1.731 | 1.437 | 0.953 | 2.166 | 8.34e-02 | 5.84e-01 | 3246 | 2599 |
| European | CIDI-noMDD | TCA:amitriptyline | Insomnia | Within-class slope | 0.123 | 0.168 | 0.732 | 1.131 | 0.814 | 1.571 | 4.64e-01 | 1.00e+00 | 3246 | 2599 |
| European | CIDI-noMDD | TCA:nortriptyline | Insomnia | Within-class slope | 0.496 | 0.189 | 2.623 | 1.642 | 1.133 | 2.378 | 8.73e-03 | 6.11e-02 | 3246 | 2599 |
| European | CIDI-noMDD | NaSSA:mirtazapine | Insomnia | Interaction (vs SSRI:sertraline) | 0.113 | 0.250 | 0.452 | 1.120 | 0.686 | 1.829 | 6.51e-01 | 1.00e+00 | 3246 | 2599 |
| European | CIDI-noMDD | NDRI:bupropion | Insomnia | Interaction (vs SSRI:sertraline) | 0.217 | 0.167 | 1.302 | 1.243 | 0.896 | 1.723 | 1.93e-01 | 1.00e+00 | 3246 | 2599 |
| European | CIDI-noMDD | SARI:trazodone | Insomnia | Interaction (vs SSRI:sertraline) | 0.220 | 0.167 | 1.320 | 1.246 | 0.899 | 1.728 | 1.87e-01 | 1.00e+00 | 3246 | 2599 |
| European | CIDI-noMDD | SNRI:duloxetine | Insomnia | Interaction (vs SSRI:sertraline) | 0.085 | 0.186 | 0.455 | 1.089 | 0.756 | 1.568 | 6.49e-01 | 1.00e+00 | 3246 | 2599 |
| European | CIDI-noMDD | SNRI:venlafaxine | Insomnia | Interaction (vs SSRI:sertraline) | -0.008 | 0.188 | -0.042 | 0.992 | 0.687 | 1.434 | 9.66e-01 | 1.00e+00 | 3246 | 2599 |
| European | CIDI-noMDD | SSRI:citalopram | Insomnia | Interaction (vs SSRI:sertraline) | 0.138 | 0.163 | 0.851 | 1.148 | 0.835 | 1.580 | 3.95e-01 | 1.00e+00 | 3246 | 2599 |
| European | CIDI-noMDD | SSRI:escitalopram | Insomnia | Interaction (vs SSRI:sertraline) | 0.114 | 0.173 | 0.659 | 1.120 | 0.799 | 1.571 | 5.10e-01 | 1.00e+00 | 3246 | 2599 |
| European | CIDI-noMDD | SSRI:fluoxetine | Insomnia | Interaction (vs SSRI:sertraline) | -0.065 | 0.179 | -0.364 | 0.937 | 0.660 | 1.330 | 7.16e-01 | 1.00e+00 | 3246 | 2599 |
| European | CIDI-noMDD | SSRI:paroxetine | Insomnia | Interaction (vs SSRI:sertraline) | 0.358 | 0.239 | 1.500 | 1.431 | 0.896 | 2.286 | 1.34e-01 | 9.36e-01 | 3246 | 2599 |
| European | CIDI-noMDD | TCA:amitriptyline | Insomnia | Interaction (vs SSRI:sertraline) | 0.119 | 0.199 | 0.598 | 1.126 | 0.763 | 1.662 | 5.50e-01 | 1.00e+00 | 3246 | 2599 |
| European | CIDI-noMDD | TCA:nortriptyline | Insomnia | Interaction (vs SSRI:sertraline) | 0.491 | 0.219 | 2.241 | 1.635 | 1.064 | 2.513 | 2.50e-02 | 1.75e-01 | 3246 | 2599 |
| European | CIDI-noMDD | | Insomnia | Joint_Interaction_Test | | | 10.787 | | | | 4.61e-01 | 1.00e+00 | 3246 | 2599 |
| European | CIDI-noMDD | SSRI:sertraline | Depression | Within-class slope | 0.144 | 0.113 | 1.278 | 1.155 | 0.926 | 1.440 | 2.01e-01 | 1.00e+00 | 3246 | 2599 |
| European | CIDI-noMDD | NaSSA:mirtazapine | Depression | Within-class slope | 0.010 | 0.267 | 0.038 | 1.010 | 0.599 | 1.704 | 9.69e-01 | 1.00e+00 | 3246 | 2599 |
| European | CIDI-noMDD | NDRI:bupropion | Depression | Within-class slope | 0.198 | 0.138 | 1.440 | 1.219 | 0.931 | 1.597 | 1.50e-01 | 1.00e+00 | 3246 | 2599 |
| European | CIDI-noMDD | SARI:trazodone | Depression | Within-class slope | 0.114 | 0.128 | 0.886 | 1.121 | 0.871 | 1.441 | 3.76e-01 | 1.00e+00 | 3246 | 2599 |
| European | CIDI-noMDD | SNRI:duloxetine | Depression | Within-class slope | 0.104 | 0.151 | 0.689 | 1.110 | 0.826 | 1.491 | 4.91e-01 | 1.00e+00 | 3246 | 2599 |
| European | CIDI-noMDD | SNRI:venlafaxine | Depression | Within-class slope | -0.042 | 0.170 | -0.246 | 0.959 | 0.688 | 1.338 | 8.06e-01 | 1.00e+00 | 3246 | 2599 |
| European | CIDI-noMDD | SSRI:citalopram | Depression | Within-class slope | 0.138 | 0.134 | 1.032 | 1.148 | 0.883 | 1.492 | 3.02e-01 | 1.00e+00 | 3246 | 2599 |

| | | | | | | | | | | | | | | |
|---|---|---|---|---|---|---|---|---|---|---|---|---|---|---|
| European | CIDI-noMDD | SSRI:escitalopram | Depression | Within-class slope | -0.151 | 0.148 | -1.023 | 0.860 | 0.644 | 1.148 | 3.06e-01 | 1.00e+00 | 3246 | 2599 |
| European | CIDI-noMDD | SSRI:fluoxetine | Depression | Within-class slope | -0.063 | 0.155 | -0.403 | 0.939 | 0.693 | 1.273 | 6.87e-01 | 1.00e+00 | 3246 | 2599 |
| European | CIDI-noMDD | SSRI:paroxetine | Depression | Within-class slope | 0.214 | 0.236 | 0.904 | 1.238 | 0.779 | 1.967 | 3.66e-01 | 1.00e+00 | 3246 | 2599 |
| European | CIDI-noMDD | TCA:amitriptyline | Depression | Within-class slope | 0.450 | 0.156 | 2.882 | 1.569 | 1.155 | 2.130 | 3.96e-03 | 2.77e-02 | 3246 | 2599 |
| European | CIDI-noMDD | TCA:nortriptyline | Depression | Within-class slope | 0.111 | 0.182 | 0.610 | 1.117 | 0.782 | 1.597 | 5.42e-01 | 1.00e+00 | 3246 | 2599 |
| European | CIDI-noMDD | NaSSA:mirtazapine | Depression | Interaction (vs SSRI:sertraline) | -0.134 | 0.288 | -0.464 | 0.875 | 0.497 | 1.539 | 6.43e-01 | 1.00e+00 | 3246 | 2599 |
| European | CIDI-noMDD | NDRI:bupropion | Depression | Interaction (vs SSRI:sertraline) | 0.054 | 0.168 | 0.322 | 1.056 | 0.759 | 1.468 | 7.48e-01 | 1.00e+00 | 3246 | 2599 |
| European | CIDI-noMDD | SARI:trazodone | Depression | Interaction (vs SSRI:sertraline) | -0.030 | 0.170 | -0.177 | 0.970 | 0.695 | 1.354 | 8.59e-01 | 1.00e+00 | 3246 | 2599 |
| European | CIDI-noMDD | SNRI:duloxetine | Depression | Interaction (vs SSRI:sertraline) | -0.040 | 0.180 | -0.222 | 0.961 | 0.675 | 1.368 | 8.24e-01 | 1.00e+00 | 3246 | 2599 |
| European | CIDI-noMDD | SNRI:venlafaxine | Depression | Interaction (vs SSRI:sertraline) | -0.186 | 0.197 | -0.942 | 0.831 | 0.564 | 1.222 | 3.46e-01 | 1.00e+00 | 3246 | 2599 |
| European | CIDI-noMDD | SSRI:citalopram | Depression | Interaction (vs SSRI:sertraline) | -0.006 | 0.165 | -0.036 | 0.994 | 0.720 | 1.373 | 9.72e-01 | 1.00e+00 | 3246 | 2599 |
| European | CIDI-noMDD | SSRI:escitalopram | Depression | Interaction (vs SSRI:sertraline) | -0.295 | 0.178 | -1.655 | 0.744 | 0.525 | 1.056 | 9.79e-02 | 6.85e-01 | 3246 | 2599 |
| European | CIDI-noMDD | SSRI:fluoxetine | Depression | Interaction (vs SSRI:sertraline) | -0.207 | 0.188 | -1.098 | 0.813 | 0.563 | 1.176 | 2.72e-01 | 1.00e+00 | 3246 | 2599 |
| European | CIDI-noMDD | SSRI:paroxetine | Depression | Interaction (vs SSRI:sertraline) | 0.070 | 0.261 | 0.266 | 1.072 | 0.643 | 1.788 | 7.90e-01 | 1.00e+00 | 3246 | 2599 |
| European | CIDI-noMDD | TCA:amitriptyline | Depression | Interaction (vs SSRI:sertraline) | 0.306 | 0.190 | 1.609 | 1.358 | 0.936 | 1.972 | 1.08e-01 | 7.53e-01 | 3246 | 2599 |
| European | CIDI-noMDD | TCA:nortriptyline | Depression | Interaction (vs SSRI:sertraline) | -0.033 | 0.210 | -0.157 | 0.968 | 0.642 | 1.459 | 8.75e-01 | 1.00e+00 | 3246 | 2599 |
| European | CIDI-noMDD | | Depression | Joint_Interaction_Test | | | 11.263 | | | | 4.22e-01 | 1.00e+00 | 3246 | 2599 |
| European | CIDI-noMDD | SSRI:sertraline | Schizophrenia | Within-class slope | -0.059 | 0.105 | -0.556 | 0.943 | 0.767 | 1.159 | 5.78e-01 | 1.00e+00 | 3246 | 2599 |
| European | CIDI-noMDD | NaSSA:mirtazapine | Schizophrenia | Within-class slope | 0.161 | 0.279 | 0.575 | 1.174 | 0.679 | 2.030 | 5.65e-01 | 1.00e+00 | 3246 | 2599 |
| European | CIDI-noMDD | NDRI:bupropion | Schizophrenia | Within-class slope | 0.069 | 0.131 | 0.524 | 1.071 | 0.829 | 1.384 | 6.00e-01 | 1.00e+00 | 3246 | 2599 |
| European | CIDI-noMDD | SARI:trazodone | Schizophrenia | Within-class slope | 0.152 | 0.135 | 1.125 | 1.164 | 0.893 | 1.518 | 2.61e-01 | 1.00e+00 | 3246 | 2599 |
| European | CIDI-noMDD | SNRI:duloxetine | Schizophrenia | Within-class slope | 0.147 | 0.143 | 1.024 | 1.158 | 0.875 | 1.533 | 3.06e-01 | 1.00e+00 | 3246 | 2599 |
| European | CIDI-noMDD | SNRI:venlafaxine | Schizophrenia | Within-class slope | 0.053 | 0.170 | 0.309 | 1.054 | 0.755 | 1.471 | 7.57e-01 | 1.00e+00 | 3246 | 2599 |
| European | CIDI-noMDD | SSRI:citalopram | Schizophrenia | Within-class slope | -0.066 | 0.119 | -0.558 | 0.936 | 0.742 | 1.181 | 5.77e-01 | 1.00e+00 | 3246 | 2599 |
| European | CIDI-noMDD | SSRI:escitalopram | Schizophrenia | Within-class slope | 0.044 | 0.133 | 0.328 | 1.045 | 0.804 | 1.357 | 7.43e-01 | 1.00e+00 | 3246 | 2599 |
| European | CIDI-noMDD | SSRI:fluoxetine | Schizophrenia | Within-class slope | 0.164 | 0.145 | 1.130 | 1.178 | 0.887 | 1.566 | 2.59e-01 | 1.00e+00 | 3246 | 2599 |
| European | CIDI-noMDD | SSRI:paroxetine | Schizophrenia | Within-class slope | 0.040 | 0.213 | 0.187 | 1.041 | 0.686 | 1.579 | 8.52e-01 | 1.00e+00 | 3246 | 2599 |
| European | CIDI-noMDD | TCA:amitriptyline | Schizophrenia | Within-class slope | 0.259 | 0.159 | 1.634 | 1.296 | 0.950 | 1.768 | 1.02e-01 | 7.16e-01 | 3246 | 2599 |
| European | CIDI-noMDD | TCA:nortriptyline | Schizophrenia | Within-class slope | 0.050 | 0.198 | 0.251 | 1.051 | 0.713 | 1.550 | 8.02e-01 | 1.00e+00 | 3246 | 2599 |
| European | CIDI-noMDD | NaSSA:mirtazapine | Schizophrenia | Interaction (vs SSRI:sertraline) | 0.219 | 0.297 | 0.736 | 1.245 | 0.695 | 2.230 | 4.61e-01 | 1.00e+00 | 3246 | 2599 |
| European | CIDI-noMDD | NDRI:bupropion | Schizophrenia | Interaction (vs SSRI:sertraline) | 0.127 | 0.158 | 0.807 | 1.136 | 0.834 | 1.546 | 4.20e-01 | 1.00e+00 | 3246 | 2599 |
| European | CIDI-noMDD | SARI:trazodone | Schizophrenia | Interaction (vs SSRI:sertraline) | 0.211 | 0.171 | 1.230 | 1.235 | 0.882 | 1.727 | 2.19e-01 | 1.00e+00 | 3246 | 2599 |
| European | CIDI-noMDD | SNRI:duloxetine | Schizophrenia | Interaction (vs SSRI:sertraline) | 0.205 | 0.172 | 1.192 | 1.228 | 0.876 | 1.720 | 2.33e-01 | 1.00e+00 | 3246 | 2599 |
| European | CIDI-noMDD | SNRI:venlafaxine | Schizophrenia | Interaction (vs SSRI:sertraline) | 0.111 | 0.198 | 0.562 | 1.118 | 0.758 | 1.647 | 5.74e-01 | 1.00e+00 | 3246 | 2599 |
| European | CIDI-noMDD | SSRI:citalopram | Schizophrenia | Interaction (vs SSRI:sertraline) | -0.008 | 0.154 | -0.049 | 0.992 | 0.734 | 1.342 | 9.61e-01 | 1.00e+00 | 3246 | 2599 |
| European | CIDI-noMDD | SSRI:escitalopram | Schizophrenia | Interaction (vs SSRI:sertraline) | 0.102 | 0.167 | 0.614 | 1.108 | 0.799 | 1.536 | 5.39e-01 | 1.00e+00 | 3246 | 2599 |
| European | CIDI-noMDD | SSRI:fluoxetine | Schizophrenia | Interaction (vs SSRI:sertraline) | 0.222 | 0.179 | 1.243 | 1.249 | 0.879 | 1.774 | 2.14e-01 | 1.00e+00 | 3246 | 2599 |
| European | CIDI-noMDD | SSRI:paroxetine | Schizophrenia | Interaction (vs SSRI:sertraline) | 0.098 | 0.232 | 0.423 | 1.103 | 0.700 | 1.740 | 6.72e-01 | 1.00e+00 | 3246 | 2599 |
| European | CIDI-noMDD | TCA:amitriptyline | Schizophrenia | Interaction (vs SSRI:sertraline) | 0.318 | 0.188 | 1.691 | 1.374 | 0.951 | 1.985 | 9.08e-02 | 6.36e-01 | 3246 | 2599 |
| European | CIDI-noMDD | TCA:nortriptyline | Schizophrenia | Interaction (vs SSRI:sertraline) | 0.108 | 0.224 | 0.483 | 1.114 | 0.718 | 1.730 | 6.29e-01 | 1.00e+00 | 3246 | 2599 |
| European | CIDI-noMDD | | Schizophrenia | Joint_Interaction_Test | | | 5.617 | | | | 8.98e-01 | 1.00e+00 | 3246 | 2599 |
| European | CIDI-noMDD | SSRI:sertraline | Bipolar Disorder | Within-class slope | 0.147 | 0.110 | 1.337 | 1.159 | 0.934 | 1.438 | 1.81e-01 | 1.00e+00 | 3246 | 2599 |
| European | CIDI-noMDD | NaSSA:mirtazapine | Bipolar Disorder | Within-class slope | 0.140 | 0.259 | 0.540 | 1.150 | 0.692 | 1.911 | 5.89e-01 | 1.00e+00 | 3246 | 2599 |
| European | CIDI-noMDD | NDRI:bupropion | Bipolar Disorder | Within-class slope | 0.202 | 0.134 | 1.506 | 1.224 | 0.941 | 1.591 | 1.32e-01 | 9.25e-01 | 3246 | 2599 |
| European | CIDI-noMDD | SARI:trazodone | Bipolar Disorder | Within-class slope | 0.009 | 0.140 | 0.061 | 1.009 | 0.767 | 1.327 | 9.52e-01 | 1.00e+00 | 3246 | 2599 |
| European | CIDI-noMDD | SNRI:duloxetine | Bipolar Disorder | Within-class slope | 0.077 | 0.159 | 0.481 | 1.080 | 0.790 | 1.475 | 6.31e-01 | 1.00e+00 | 3246 | 2599 |
| European | CIDI-noMDD | SNRI:venlafaxine | Bipolar Disorder | Within-class slope | 0.011 | 0.164 | 0.067 | 1.011 | 0.733 | 1.395 | 9.47e-01 | 1.00e+00 | 3246 | 2599 |
| European | CIDI-noMDD | SSRI:citalopram | Bipolar Disorder | Within-class slope | 0.039 | 0.119 | 0.329 | 1.040 | 0.824 | 1.312 | 7.43e-01 | 1.00e+00 | 3246 | 2599 |
| European | CIDI-noMDD | SSRI:escitalopram | Bipolar Disorder | Within-class slope | -0.044 | 0.166 | -0.266 | 0.957 | 0.692 | 1.324 | 7.91e-01 | 1.00e+00 | 3246 | 2599 |
| European | CIDI-noMDD | SSRI:fluoxetine | Bipolar Disorder | Within-class slope | 0.047 | 0.151 | 0.312 | 1.048 | 0.779 | 1.411 | 7.55e-01 | 1.00e+00 | 3246 | 2599 |
| European | CIDI-noMDD | SSRI:paroxetine | Bipolar Disorder | Within-class slope | -0.330 | 0.216 | -1.525 | 0.719 | 0.470 | 1.099 | 1.27e-01 | 8.90e-01 | 3246 | 2599 |
| European | CIDI-noMDD | TCA:amitriptyline | Bipolar Disorder | Within-class slope | 0.319 | 0.167 | 1.911 | 1.375 | 0.992 | 1.908 | 5.61e-02 | 3.92e-01 | 3246 | 2599 |
| European | CIDI-noMDD | TCA:nortriptyline | Bipolar Disorder | Within-class slope | 0.105 | 0.184 | 0.568 | 1.110 | 0.774 | 1.593 | 5.70e-01 | 1.00e+00 | 3246 | 2599 |
| European | CIDI-noMDD | NaSSA:mirtazapine | Bipolar Disorder | Interaction (vs SSRI:sertraline) | -0.007 | 0.274 | -0.027 | 0.993 | 0.580 | 1.697 | 9.78e-01 | 1.00e+00 | 3246 | 2599 |
| European | CIDI-noMDD | NDRI:bupropion | Bipolar Disorder | Interaction (vs SSRI:sertraline) | 0.054 | 0.167 | 0.327 | 1.056 | 0.762 | 1.464 | 7.44e-01 | 1.00e+00 | 3246 | 2599 |
| European | CIDI-noMDD | SARI:trazodone | Bipolar Disorder | Interaction (vs SSRI:sertraline) | -0.139 | 0.175 | -0.793 | 0.870 | 0.617 | 1.227 | 4.28e-01 | 1.00e+00 | 3246 | 2599 |
| European | CIDI-noMDD | SNRI:duloxetine | Bipolar Disorder | Interaction (vs SSRI:sertraline) | -0.071 | 0.187 | -0.380 | 0.932 | 0.646 | 1.343 | 7.04e-01 | 1.00e+00 | 3246 | 2599 |
| European | CIDI-noMDD | SNRI:venlafaxine | Bipolar Disorder | Interaction (vs SSRI:sertraline) | -0.136 | 0.194 | -0.704 | 0.872 | 0.597 | 1.275 | 4.81e-01 | 1.00e+00 | 3246 | 2599 |
| European | CIDI-noMDD | SSRI:citalopram | Bipolar Disorder | Interaction (vs SSRI:sertraline) | -0.108 | 0.160 | -0.675 | 0.897 | 0.655 | 1.229 | 4.99e-01 | 1.00e+00 | 3246 | 2599 |

| | | | | | | | | | | | | | |
|---|---|---|---|---|---|---|---|---|---|---|---|---|---|
| European | CIDI-noMDD | SSRI:escitalopram | Bipolar Disorder | Interaction (vs SSRI:sertraline) | -0.191 | 0.193 | -0.994 | 0.826 | 0.566 | 1.205 | 3.20e-01 | 1.00e+00 | 3246 | 2599 |
| European | CIDI-noMDD | SSRI:fluoxetine | Bipolar Disorder | Interaction (vs SSRI:sertraline) | -0.100 | 0.182 | -0.552 | 0.905 | 0.634 | 1.291 | 5.81e-01 | 1.00e+00 | 3246 | 2599 |
| European | CIDI-noMDD | SSRI:paroxetine | Bipolar Disorder | Interaction (vs SSRI:sertraline) | -0.477 | 0.242 | -1.969 | 0.620 | 0.386 | 0.998 | 4.89e-02 | 3.43e-01 | 3246 | 2599 |
| European | CIDI-noMDD | TCA:amitriptyline | Bipolar Disorder | Interaction (vs SSRI:sertraline) | 0.171 | 0.195 | 0.880 | 1.187 | 0.810 | 1.738 | 3.79e-01 | 1.00e+00 | 3246 | 2599 |
| European | CIDI-noMDD | TCA:nortriptyline | Bipolar Disorder | Interaction (vs SSRI:sertraline) | -0.043 | 0.212 | -0.201 | 0.958 | 0.632 | 1.453 | 8.41e-01 | 1.00e+00 | 3246 | 2599 |
| European | CIDI-noMDD | | Bipolar Disorder | Joint_Interaction_Test | | | 8.587 | | | | 6.60e-01 | 1.00e+00 | 3246 | 2599 |
| European | CIDI-noMDD | SSRI:sertraline | BMI | Within-class slope | -0.046 | 0.103 | -0.447 | 0.955 | 0.781 | 1.168 | 6.55e-01 | 1.00e+00 | 3246 | 2599 |
| European | CIDI-noMDD | NaSSA:mirtazapine | BMI | Within-class slope | 0.075 | 0.228 | 0.327 | 1.077 | 0.689 | 1.685 | 7.44e-01 | 1.00e+00 | 3246 | 2599 |
| European | CIDI-noMDD | NDRI:bupropion | BMI | Within-class slope | 0.151 | 0.133 | 1.133 | 1.163 | 0.896 | 1.509 | 2.57e-01 | 1.00e+00 | 3246 | 2599 |
| European | CIDI-noMDD | SARI:trazodone | BMI | Within-class slope | 0.140 | 0.146 | 0.959 | 1.151 | 0.864 | 1.533 | 3.38e-01 | 1.00e+00 | 3246 | 2599 |
| European | CIDI-noMDD | SNRI:duloxetine | BMI | Within-class slope | -0.222 | 0.153 | -1.449 | 0.801 | 0.593 | 1.081 | 1.47e-01 | 1.00e+00 | 3246 | 2599 |
| European | CIDI-noMDD | SNRI:venlafaxine | BMI | Within-class slope | -0.108 | 0.168 | -0.641 | 0.898 | 0.646 | 1.248 | 5.22e-01 | 1.00e+00 | 3246 | 2599 |
| European | CIDI-noMDD | SSRI:citalopram | BMI | Within-class slope | 0.073 | 0.129 | 0.562 | 1.075 | 0.835 | 1.385 | 5.74e-01 | 1.00e+00 | 3246 | 2599 |
| European | CIDI-noMDD | SSRI:escitalopram | BMI | Within-class slope | -0.096 | 0.134 | -0.719 | 0.908 | 0.698 | 1.181 | 4.72e-01 | 1.00e+00 | 3246 | 2599 |
| European | CIDI-noMDD | SSRI:fluoxetine | BMI | Within-class slope | 0.096 | 0.129 | 0.746 | 1.101 | 0.855 | 1.419 | 4.56e-01 | 1.00e+00 | 3246 | 2599 |
| European | CIDI-noMDD | SSRI:paroxetine | BMI | Within-class slope | 0.105 | 0.225 | 0.464 | 1.110 | 0.714 | 1.727 | 6.42e-01 | 1.00e+00 | 3246 | 2599 |
| European | CIDI-noMDD | TCA:amitriptyline | BMI | Within-class slope | 0.182 | 0.148 | 1.229 | 1.200 | 0.897 | 1.605 | 2.19e-01 | 1.00e+00 | 3246 | 2599 |
| European | CIDI-noMDD | TCA:nortriptyline | BMI | Within-class slope | 0.037 | 0.181 | 0.203 | 1.038 | 0.727 | 1.481 | 8.39e-01 | 1.00e+00 | 3246 | 2599 |
| European | CIDI-noMDD | NaSSA:mirtazapine | BMI | Interaction (vs SSRI:sertraline) | 0.121 | 0.246 | 0.490 | 1.128 | 0.697 | 1.827 | 6.24e-01 | 1.00e+00 | 3246 | 2599 |
| European | CIDI-noMDD | NDRI:bupropion | BMI | Interaction (vs SSRI:sertraline) | 0.197 | 0.161 | 1.222 | 1.217 | 0.888 | 1.669 | 2.22e-01 | 1.00e+00 | 3246 | 2599 |
| European | CIDI-noMDD | SARI:trazodone | BMI | Interaction (vs SSRI:sertraline) | 0.186 | 0.182 | 1.026 | 1.205 | 0.844 | 1.720 | 3.05e-01 | 1.00e+00 | 3246 | 2599 |
| European | CIDI-noMDD | SNRI:duloxetine | BMI | Interaction (vs SSRI:sertraline) | -0.176 | 0.184 | -0.957 | 0.839 | 0.585 | 1.202 | 3.39e-01 | 1.00e+00 | 3246 | 2599 |
| European | CIDI-noMDD | SNRI:venlafaxine | BMI | Interaction (vs SSRI:sertraline) | -0.062 | 0.195 | -0.315 | 0.940 | 0.641 | 1.379 | 7.52e-01 | 1.00e+00 | 3246 | 2599 |
| European | CIDI-noMDD | SSRI:citalopram | BMI | Interaction (vs SSRI:sertraline) | 0.119 | 0.159 | 0.747 | 1.126 | 0.825 | 1.537 | 4.55e-01 | 1.00e+00 | 3246 | 2599 |
| European | CIDI-noMDD | SSRI:escitalopram | BMI | Interaction (vs SSRI:sertraline) | -0.050 | 0.164 | -0.306 | 0.951 | 0.689 | 1.313 | 7.60e-01 | 1.00e+00 | 3246 | 2599 |
| European | CIDI-noMDD | SSRI:fluoxetine | BMI | Interaction (vs SSRI:sertraline) | 0.142 | 0.162 | 0.878 | 1.153 | 0.839 | 1.584 | 3.80e-01 | 1.00e+00 | 3246 | 2599 |
| European | CIDI-noMDD | SSRI:paroxetine | BMI | Interaction (vs SSRI:sertraline) | 0.151 | 0.246 | 0.612 | 1.163 | 0.718 | 1.883 | 5.40e-01 | 1.00e+00 | 3246 | 2599 |
| European | CIDI-noMDD | TCA:amitriptyline | BMI | Interaction (vs SSRI:sertraline) | 0.228 | 0.178 | 1.279 | 1.256 | 0.886 | 1.783 | 2.01e-01 | 1.00e+00 | 3246 | 2599 |
| European | CIDI-noMDD | TCA:nortriptyline | BMI | Interaction (vs SSRI:sertraline) | 0.083 | 0.208 | 0.397 | 1.086 | 0.722 | 1.634 | 6.91e-01 | 1.00e+00 | 3246 | 2599 |
| European | CIDI-noMDD | | BMI | Joint_Interaction_Test | | | 8.112 | | | | 7.03e-01 | 1.00e+00 | 3246 | 2599 |

**Supplementary Table 16. Familial recurrence risk ratios for antidepressant treatment outcomes within the Lifelines full cohort.** For each outcome, the recurrence risk ratio (λR) is the outcome prevalence among relatives of affected individuals divided by the prevalence in the full cohort, estimated across three relationship types (full siblings, parent–child, grandparent–grandchild) and pooled across all pairs ("Any"). Values >1 indicate familial aggregation.

Columns: Outcome = the treatment outcome assessed (SSRI, TCA, and SNRI continuation; and pooled Switching, Discontinuation, and Continuation); relationship_type = relative-pair class (full_sibling, parent_child, grandparent_grandchild, or Any); n_relative_pairs = number of informative relative pairs; risk_in_relatives = outcome prevalence among relatives of affected individuals; n_individuals = cohort size used for the reference risk; risk_in_sample = outcome prevalence in the full cohort; risk_differences = risk_in_relatives minus risk_in_sample; recurrence_risk_ratio = risk_in_relatives divided by risk_in_sample (λR); boot_se = bootstrap standard error; ci_lower/ci_upper = 95% bootstrap confidence interval bounds, p_value = bootstrap p-value for the null of no familial aggregation (λR = 1). Confidence intervals and p-values were derived by cluster bootstrap resampling of families (B = 1,000 replicates), preserving within-family dependence; the reference prevalence was treated as fixed. Intervals are the 2.5th and 97.5th percentiles of the bootstrap distribution. P-values were obtained by inverting this distribution against the null λR = 1: $p = 2 \times \min(p_{lo}, p_{hi})$, where $p_{lo} = (1 + \#\{\lambda R^* \leq 1\})/(1 + B)$, $p_{hi} = (1 + \#\{\lambda R^* \geq 1\})/(1 + B)$, and $\lambda R^*$ denotes a bootstrap replicate. The minimum attainable p-value is therefore 0.002. Estimates in the smaller relative-pair strata are correspondingly imprecise.

| **Outcome** | **relationship_type** | **n_relative_pairs** | **risk_in_relatives** | **n_individuals** | **risk_in_sample** | **risk_differences** | **recurrence_risk_ratio** | **ci_lower** | **ci_upper** | **boot_se** | **p_value** |
|---|---|---|---|---|---|---|---|---|---|---|---|
| SSRI Continuation | full_sibling | 184 | 0.576 | 12884 | 0.424 | 0.152 | 1.360 | 1.233 | 1.491 | 0.067 | 0.001998 |
| SSRI Continuation | grandparent_grandchild | 53 | 0.491 | 12884 | 0.424 | 0.067 | 1.158 | 0.908 | 1.399 | 0.125 | 0.225774 |
| SSRI Continuation | parent_child | 296 | 0.459 | 12884 | 0.424 | 0.036 | 1.085 | 0.985 | 1.180 | 0.052 | 0.105894 |
| SSRI Continuation | Any | 533 | 0.503 | 12884 | 0.424 | 0.079 | 1.187 | 1.107 | 1.274 | 0.043 | 0.001998 |
| TCA Continuation | full_sibling | 91 | 0.253 | 12884 | 0.221 | 0.032 | 1.144 | 0.768 | 1.452 | 0.172 | 0.3996 |
| TCA Continuation | grandparent_grandchild | 30 | 0.233 | 12884 | 0.221 | 0.012 | 1.056 | 0.502 | 1.597 | 0.282 | 0.837163 |
| TCA Continuation | parent_child | 151 | 0.325 | 12884 | 0.221 | 0.104 | 1.469 | 1.180 | 1.749 | 0.142 | 0.003996 |
| TCA Continuation | Any | 272 | 0.290 | 12884 | 0.221 | 0.069 | 1.314 | 1.092 | 1.518 | 0.108 | 0.005994 |
| SNRI Continuation | full_sibling | 50 | 0.180 | 12884 | 0.104 | 0.076 | 1.738 | 0.935 | 2.492 | 0.402 | 0.075924 |
| SNRI Continuation | grandparent_grandchild | 16 | 0.375 | 12884 | 0.104 | 0.271 | 3.622 | 1.610 | 5.519 | 0.978 | 0.00999 |
| SNRI Continuation | parent_child | 65 | 0.231 | 12884 | 0.104 | 0.127 | 2.229 | 1.305 | 2.990 | 0.420 | 0.011988 |
| SNRI Continuation | Any | 131 | 0.229 | 12884 | 0.104 | 0.125 | 2.212 | 1.448 | 2.827 | 0.361 | 0.001998 |
| Switching | full_sibling | 65 | 0.169 | 12884 | 0.166 | 0.004 | 1.022 | 0.619 | 1.438 | 0.216 | 0.869131 |
| Switching | grandparent_grandchild | 23 | 0.348 | 12884 | 0.166 | 0.182 | 2.101 | 0.503 | 3.252 | 0.734 | 0.171828 |
| Switching | parent_child | 126 | 0.190 | 12884 | 0.166 | 0.025 | 1.151 | 0.827 | 1.467 | 0.166 | 0.367632 |
| Switching | Any | 214 | 0.201 | 12884 | 0.166 | 0.035 | 1.214 | 0.908 | 1.485 | 0.146 | 0.163836 |
| Discontinuation | full_sibling | 66 | 0.152 | 12884 | 0.201 | -0.049 | 0.754 | 0.383 | 1.066 | 0.168 | 0.101898 |
| Discontinuation | grandparent_grandchild | 23 | 0.174 | 12884 | 0.201 | -0.027 | 0.865 | 0.311 | 1.422 | 0.303 | 0.593407 |
| Discontinuation | parent_child | 121 | 0.207 | 12884 | 0.201 | 0.006 | 1.028 | 0.730 | 1.295 | 0.144 | 0.867133 |
| Discontinuation | Any | 210 | 0.186 | 12884 | 0.201 | -0.015 | 0.924 | 0.700 | 1.123 | 0.112 | 0.509491 |
| Continuation | full_sibling | 234 | 0.632 | 12884 | 0.556 | 0.077 | 1.138 | 1.055 | 1.228 | 0.045 | 0.001998 |
| Continuation | grandparent_grandchild | 74 | 0.676 | 12884 | 0.556 | 0.120 | 1.215 | 1.063 | 1.362 | 0.077 | 0.005994 |
| Continuation | parent_child | 366 | 0.615 | 12884 | 0.556 | 0.059 | 1.106 | 1.038 | 1.175 | 0.036 | 0.003996 |
| Continuation | Any | 674 | 0.628 | 12884 | 0.556 | 0.072 | 1.129 | 1.077 | 1.182 | 0.028 | 0.001998 |

**Supplementary Table 17. Tetrachoric correlations for antidepressant treatment outcomes within the Lifelines full cohort.** For each outcome, the tetrachoric correlation estimates the correlation in liability to the (binary) outcome between pairs of relatives, across three relationship types (full siblings, parent–child, grandparent–grandchild) and pooled across all pairs ("Any"). Values >0 indicate correlated liability within relative pairs. Columns: outcome = the treatment outcome assessed (SSRI, TCA, and SNRI continuation; and pooled Switching, Discontinuation, and Continuation); relationship_type = relative-pair class (full_sibling, parent_child, grandparent_grandchild, or Any); n_pairs = number of informative relative pairs; n_families = number of distinct families contributing pairs; tetrachoric_r = estimated tetrachoric correlation; se = standard error; ci_lower/ci_upper = 95% bootstrap confidence interval bounds, p = bootstrap p-value for the null of zero liability correlation. Confidence intervals and p-values were derived by cluster bootstrap resampling of families (B = 1,000 replicates), preserving within-family dependence. Intervals are the 2.5th and 97.5th percentiles of the bootstrap distribution. P-values were obtained by inverting this distribution against the null r = 0: p = 2 × min(p_lo, p_hi), where p_lo = (1 + #{r* ≤ 0})/(1 + B), p_hi = (1 + #{r* ≥ 0})/(1 + B), and r* denotes a bootstrap replicate. Replicates for which the estimator failed to converge were excluded and B adjusted accordingly. The minimum attainable p-value is therefore 0.002. A correlation with a confidence interval excluding 0 indicates statistically significant within-pair liability correlation.

| outcome | relationship_type | n_pairs | n_families | tetrachoric_r | se | ci_lower | ci_upper | p |
|---|---|---|---|---|---|---|---|---|
| SSRI_Continuation | full_sibling | 401 | 352 | 0.264 | 0.078 | 0.109 | 0.412 | 0.003996 |
| SSRI_Continuation | grandparent_grandchild | 127 | 118 | 0.155 | 0.145 | -0.145 | 0.426 | 0.305694 |
| SSRI_Continuation | parent_child | 667 | 552 | 0.197 | 0.063 | 0.070 | 0.323 | 0.003996 |
| SSRI_Continuation | Any | 1195 | 881 | 0.217 | 0.049 | 0.121 | 0.310 | 0.001998 |
| TCA_Continuation | full_sibling | 401 | 352 | 0.038 | 0.104 | -0.182 | 0.229 | 0.719281 |
| TCA_Continuation | grandparent_grandchild | 127 | 118 | 0.013 | 0.168 | -0.333 | 0.321 | 0.883117 |
| TCA_Continuation | parent_child | 667 | 552 | 0.117 | 0.073 | -0.029 | 0.256 | 0.113886 |
| TCA_Continuation | Any | 1195 | 881 | 0.081 | 0.057 | -0.033 | 0.191 | 0.163836 |
| SNRI_Continuation | full_sibling | 401 | 352 | 0.173 | 0.126 | -0.075 | 0.394 | 0.203796 |
| SNRI_Continuation | grandparent_grandchild | 127 | 118 | 0.373 | 0.190 | -0.025 | 0.664 | 0.067932 |
| SNRI_Continuation | parent_child | 667 | 552 | 0.300 | 0.101 | 0.071 | 0.477 | 0.013986 |
| SNRI_Continuation | Any | 1195 | 881 | 0.270 | 0.087 | 0.088 | 0.419 | 0.005994 |
| Switching | full_sibling | 401 | 352 | 0.019 | 0.114 | -0.229 | 0.224 | 0.927073 |
| Switching | grandparent_grandchild | 127 | 118 | 0.527 | 0.249 | -0.072 | 0.821 | 0.075924 |
| Switching | parent_child | 667 | 552 | 0.044 | 0.088 | -0.145 | 0.213 | 0.623377 |
| Switching | Any | 1195 | 881 | 0.093 | 0.068 | -0.040 | 0.222 | 0.175824 |
| Discontinuation | full_sibling | 401 | 352 | 0.066 | 0.119 | -0.204 | 0.276 | 0.605395 |
| Discontinuation | grandparent_grandchild | 127 | 118 | 0.094 | 0.250 | -0.453 | 0.441 | 0.657343 |

| | | | | | | | | |
|---|---|---|---|---|---|---|---|---|
| Discontinuation | parent_child | 667 | 552 | 0.076 | 0.086 | -0.105 | 0.227 | 0.411588 |
| Discontinuation | Any | 1195 | 881 | 0.077 | 0.069 | -0.067 | 0.204 | 0.2997 |
| Continuation | full_sibling | 401 | 352 | 0.103 | 0.081 | -0.058 | 0.253 | 0.21978 |
| Continuation | grandparent_grandchild | 127 | 118 | 0.208 | 0.154 | -0.112 | 0.507 | 0.181818 |
| Continuation | parent_child | 667 | 552 | 0.117 | 0.061 | -0.010 | 0.230 | 0.067932 |
| Continuation | Any | 1195 | 881 | 0.123 | 0.049 | 0.023 | 0.219 | 0.017982 |

**Supplementary Table S18. List of antidepressants considered in All of Us and Lifelines.** Classification follows the mechanism-based scheme applied in All of Us and Lifelies. ATC codes are WHO ATC/DDD fifth-level codes. ✓ indicates inclusion in the All of Us antidepressant list; tick the Lifelines column where the agent is captured in Lifelines. † Selegiline has no dedicated N06A code; it is classified under N04BD01 (anti-Parkinson) and used as an antidepressant transdermally.

| Drug (generic name) | ATC code | Class | Used in All of Us | Used in Pharmlines |
|---|---|---|---|---|
| Citalopram | N06AB04 | SSRI | ✓ | ✓ |
| Escitalopram | N06AB10 | SSRI | ✓ | ✓ |
| Fluoxetine | N06AB03 | SSRI | ✓ | ✓ |
| Fluvoxamine | N06AB08 | SSRI | ✓ | ✓ |
| Paroxetine | N06AB05 | SSRI | ✓ | ✓ |
| Sertraline | N06AB06 | SSRI | ✓ | ✓ |
| Desvenlafaxine | N06AX23 | SNRI | ✓ | |
| Duloxetine | N06AX21 | SNRI | ✓ | ✓ |
| Levomilnacipran | N06AX28 | SNRI | ✓ | |
| Milnacipran | N06AX17 | SNRI | ✓ | |
| Venlafaxine | N06AX16 | SNRI | ✓ | ✓ |
| Viloxazine | N06AX09 | NRI | ✓ | |
| Maprotiline | N06AA21 | NRI | ✓ | ✓ |
| Bupropion | N06AX12 | NDRI | ✓ | ✓ |
| Mirtazapine | N06AX11 | NaSSA | ✓ | ✓ |
| Mianserin | N06AX03 | NaSSA | ✓ | ✓ |
| Trazodone | N06AX05 | SARI | ✓ | ✓ |
| Nefazodone | N06AX06 | SARI | ✓ | ✓ |
| Vortioxetine | N06AX26 | Serotonin modulator | ✓ | ✓ |
| Vilazodone | N06AX24 | Serotonin modulator | ✓ | |
| Oxitriptan (5-HTP) | N06AX01 | Serotonin precursor | ✓ | ✓ |
| Imipramine | N06AA02 | TCA | ✓ | ✓ |
| Amitriptyline | N06AA09 | TCA | ✓ | ✓ |
| Clomipramine | N06AA04 | TCA | ✓ | ✓ |
| Desipramine | N06AA01 | TCA | ✓ | ✓ |
| Doxepin | N06AA12 | TCA | ✓ | ✓ |

| | | | | |
|---|---|---|---|---|
| Nortriptyline | N06AA10 | TCA | √ | √ |
| Protriptyline | N06AA11 | TCA | √ | |
| Trimipramine | N06AA06 | TCA | √ | √ |
| Amoxapine | N06AA17 | TCA | √ | |
| Isocarboxazid | N06AF01 | MAOI | √ | |
| Phenelzine | N06AF03 | MAOI | √ | √ |
| Selegiline | N06AX† | MAOI | √ | |
| Tranylcypromine | N06AF04 | MAOI | √ | √ |
| Esketamine | N06AX27 | NMDA antagonist | √ | |
| Moclobemide | N06AG02 | RIMA | √ | √ |
| Agomelatine | N06AX22 | Melatonin agonist | | √ |
| Hypericiherba | N06AX25 | Hypericiherba | | √ |

**Supplementary Table S19. List of atypical antipsychotics considered in All of Us and Lifelines.** ATC codes are WHO ATC/DDD fifth-level codes. ✓ indicates inclusion in the All of Us screening list; tick the Lifelines column where the agent is captured in Lifelines. Lithium is a mood stabiliser used as an augmentation agent, not an antipsychotic, and is listed separately.

| Drug (generic name) | ATC code | ATC subgroup | Used in All of Us | Used in Pharmlines |
|---|---|---|---|---|
| Clozapine | N05AH02 | Diazepines, oxazepines, thiazepines (N05AH) | ✓ | ✓ |
| Olanzapine | N05AH03 | Diazepines, oxazepines, thiazepines (N05AH) | ✓ | ✓ |
| Quetiapine | N05AH04 | Diazepines, oxazepines, thiazepines (N05AH) | ✓ | ✓ |
| Asenapine | N05AH05 | Diazepines, oxazepines, thiazepines (N05AH) | ✓ | ✓ |
| Sulpiride | N05AL01 | Benzamides (N05AL) | ✓ | |
| Amisulpride | N05AL05 | Benzamides (N05AL) | ✓ | |
| Ziprasidone | N05AE04 | Indole derivatives (N05AE) | ✓ | ✓ |
| Lurasidone | N05AE05 | Indole derivatives (N05AE) | ✓ | ✓ |
| Risperidone | N05AX08 | Other antipsychotics (N05AX) | ✓ | ✓ |
| Aripiprazole | N05AX12 | Other antipsychotics (N05AX) | ✓ | ✓ |
| Paliperidone | N05AX13 | Other antipsychotics (N05AX) | ✓ | ✓ |
| Iloperidone | N05AX14 | Other antipsychotics (N05AX) | ✓ | ✓ |
| Cariprazine | N05AX15 | Other antipsychotics (N05AX) | ✓ | ✓ |
| Brexpiprazole | N05AX16 | Other antipsychotics (N05AX) | ✓ | ✓ |
| Pimavanserin | N05AX17 | Other antipsychotics (N05AX) | ✓ | |
| Lumateperone | N05AX18 | Other antipsychotics (N05AX) | ✓ | |
| Pipamperone | N05AD05 | Butyrophenone derivatives (N05AD) | | ✓ |
| ***Augmentation agent (non-antipsychotic)*** | | | | |
| Lithium | N05AN01 | Mood stabiliser — augmentation agent (N05AN) | ✓ | |

**Supplementary Table S20. List of GWAS summary statistics from which PGS were calculated from using the SBayesRC method implemented in GCTB software.** One trait used published GWAS with the Lifelines sample metasubstracted to avoid biased PGS in the Lifelines cohort due to sample overlap. We benchmark statistical power by reporting the number of significant COJO loci, sample size, and SNP-based heritability estimated by SBayesRC.

| Trait | Lifelines Metasubstracted? | Author | Publication Link | N | N_Cases | N_Controls | SBayesRC_hsq | N_COJO_SNPs |
|---|---|---|---|---|---|---|---|---|
| ADHD | No | Demontis et al., 2023 | https://doi.org/10.1038/s41588-022-01285-8 | 225534 | 38,691 | 186,843 | 0.107 | 26 |
| Schizophrenia (SCZ) | No | Trubetskoy et al., 2022 | https://www.ncbi.nlm.nih.gov/pmc/articles/PMC9392466/ | 130644 | 53,386 | 77,258 | 0.474 | 197 |
| Migraine | No | Hautakangas et al., 2022 | https://doi.org/10.1038/s41588-021-00990-0 | 588413 | 48,975 | 540,381 | 0.037 | 38 |
| Bipolar Disorder (BIP) | No | O'Connell, et al., 2025 | https://www.nature.com/articles/s41586-024-08468-9 | 2438091 | 128,269 | 2,309,822 | 0.020 | 169 |
| Body Mass Index (BMI) | Yes | Yengo, et al., 2018 | https://doi.org/10.1093/hmg/ddy271 | 686542 | N/A | N/A | 0.205 | 1017 |
| Major Depression (MD) | No | Adams, et al., 2025 | https://pubmed.ncbi.nlm.nih.gov/39814019/ | 3857621 | 510,763 | 3,346,858 | 0.029 | 579 |
| Insomnia | No | Jiang et al., 2019 | https://pubmed.ncbi.nlm.nih.gov/34737426/ | 455744 | N/A | N/A | 0.071 | 50 |
